# Sex differences in the genetic architecture of clinical quantitative traits in the electronic health record

**DOI:** 10.64898/2026.09.02.26362069

**Authors:** Freida Blostein, Banabithi Bose, Andrew Hill, Ky’Era Actkins, Allison Lake, Naomi Freilich, Peter Straub, Maria Niarchou, Barbara E. Stranger, Lea K. Davis

## Abstract

Sex differences exist in complex diseases, but the genetic architecture of sex differences in quantitative clinical traits is understudied. Here, we performed sex-stratified genome-wide association studies of 508 clinical traits from electronic health records (EHRs) in 64,956 European-ancestry individuals in the Vanderbilt biobank, BioVU, with replication in the UK Biobank and Colorado Center for Personalized Medicine biobank. Most trait distributions differed significantly by sex. We identified a female-specific locus for erythrocyte distribution width at *PIEZO1* and sex-differentiated effect magnitudes at *APOE* (LDL cholesterol) and *SLC2A9* (uric acid) that replicated across biobanks, varied with age, and partially mediated associations with heart disease and gout. Opposite-direction sex effects were less replicable. Ascertainment bias could be addressed using inverse-probability weighting. Sex-stratified colocalization with eQTLs nominated additional candidate genes. Differential effect magnitude, not opposing effects, predominate in sex-specific genetic architecture, requiring large sample sizes for discovery that can be achieved using EHR data.

## Introduction

Sex differences in the incidence, prevalence, and etiology of complex diseases are well established^1^. Both exogenous (e.g., environmental exposures) and endogenous (e.g., hormones, genetic variants, gene expression) factors contribute to sex differences in quantitative clinical traits (hereafter referred to as labs)^1^, which are used to diagnose and monitor complex diseases. When clinical guidelines are developed from combined-sex studies, reference ranges may inadequately capture sex-specific physiology, contributing to differences in diagnosis and treatment. For example, sex differences in the sodium-adjusted model for end-stage liver disease, used for transplant allocation, persist from baseline throughout disease course and contribute to increased wait times and mortality for females on liver transplant lists^2^. Further, sex differences in cardiac troponin may contribute to underdiagnosis and undertreatment of myocardial injury in females^3^.

Moreover, many labs or traits that fluctuate with disease progression are moderately to highly heritable^4^ indicating a genetic contribution to a proportion of their observed variances. Although autosomal genetic variants occur at equal frequencies in males and females, as predicted by Mendelian segregation, their phenotypic effects need not be identical. Differences in cellular, hormonal, and physiological environments can modify genetic variants’ effects, producing gene-by-sex interactions despite shared underlying genetic architecture. Characterizing these interactions can provide mechanistic insight into the biological pathways underlying phenotypic sex differences. Understanding how sex modifies the genetic architecture of quantitative heritable labs may improve interpretation of labs and ultimately inform sex-aware precision medicine through more accurate genetic risk prediction and clinical decision-making.

Gene-by-sex interactions appear to be small in magnitude but widespread in the genetic architecture of complex traits^5–7^. For instance, genetic correlations between males and females are generally strong when averaged across the whole genome, but locus-level analyses reveal evidence of sex differences^7^. The predominant model for these differences appears to be an increase in the magnitude of genetic effects in one sex (i.e. amplification), rather than opposing directions of effect or different causal loci between sexes^6^. At the level of polygenic scores, modeling these locus-specific amplifications can improve prediction^6^, underlining how sex-aware analyses can translate to improved genomic tools for clinical risk stratification and precision medicine.

However, almost all previous investigations of sex differences in labs at scale have used data from the UK biobank (UKB), and investigations in other cohorts have analyzed limited numbers of traits^8–10^. Repeated analyses of the same data set can help establish reproducibility, but not replicability nor generalizability across populations and contexts. The UKB also is affected by selection biases, including healthy volunteer bias, which can skew associations^11^. Increasingly, academic medical centers (AMCs) that provide primary, secondary, and tertiary clinical care include biobanks which link electronic health records (EHRs) to genomic data^12,13^. EHR-linked biobanks contain a broad range of quantitative measurements, as well as rich data on potential modifiers and downstream outcomes. Although EHR-based biobanks come with their own ascertainment biases, they represent an opportunity to assess the robustness of gene-by-sex interactions across study designs. Moreover, AMCs are more representative of the environment in which sex-aware precision medicine approaches would eventually be implemented.

To our knowledge, no study has systematically investigated sex differences in the genetic architecture of a broad range of labs in EHR-linked biobanks. Here, we conducted sex-stratified genome-wide association analyses of labs in the Vanderbilt University Medical Center’s biobank, BioVU, replicating our results in the Colorado Center for Personalized Medicine biobank (CCPM) and the UKB. We identified sex-specific associations, including a female-specific signal for erythrocyte distribution width at *PIEZO1* and replicable effect magnitude differences for *APOE* on LDL cholesterol and *SLC2A9* on uric acid levels. Leveraging EHR data, we further show that these sex differences vary by age, remain after controlling for medication use and comorbidity patterns, and may mediate associations with heart disease and gout. Finally, sex stratified colocalization identified several gene-trait pairs with strong colocalization in only one sex. Our results establish EHR-linked biobanks as a powerful resource for discovering and interpreting sex-dependent genetic effects on clinically relevant traits.

## Results

### Labs exhibit small but significant sex differences in distribution

There were 64956 individuals of European ancestry in the Vanderbilt University Medical Center’s biobank, BioVU, with ≥ 1 lab(s) measured at or after age 18 (Supplementary Table 1), and 508 labs (19 measured overwhelmingly in one sex, Supplementary Figure 1, Supplementary Tables 1-2). We summarized observations across the medical record as per-person medians and, for individuals with >1 observation, per-person variances, residualized for age at median observation (three cubic splines) and inverse normalized within sex (Supplementary Figure 2, Methods).

Of the 489 labs measured in both sexes, 59% exhibited significant sex differences in mean per-person median values (Bonferroni threshold=0.05/489=1.00e-04, Figure 1). Standardized mean differences of per-person medians were typically small (standardized value<1), except for endocrine hormones (Supplementary Table 1). However, even small sex differences may be clinically meaningful: for example, LDL cholesterol exhibited a standardized mean difference of 0.31 (p=1.78e-176 raw difference of 10.05 mg/dL higher in females). Per-person variances showed sex differences less often (30% of tested labs; Figure 1) and largely overlapped with labs showing median differences (Chi Sq=51.8, *P*=6e-13); only 24 traits, e.g., QT interval, differed by sex in variance alone (Figure 1B).

**Figure 1.**
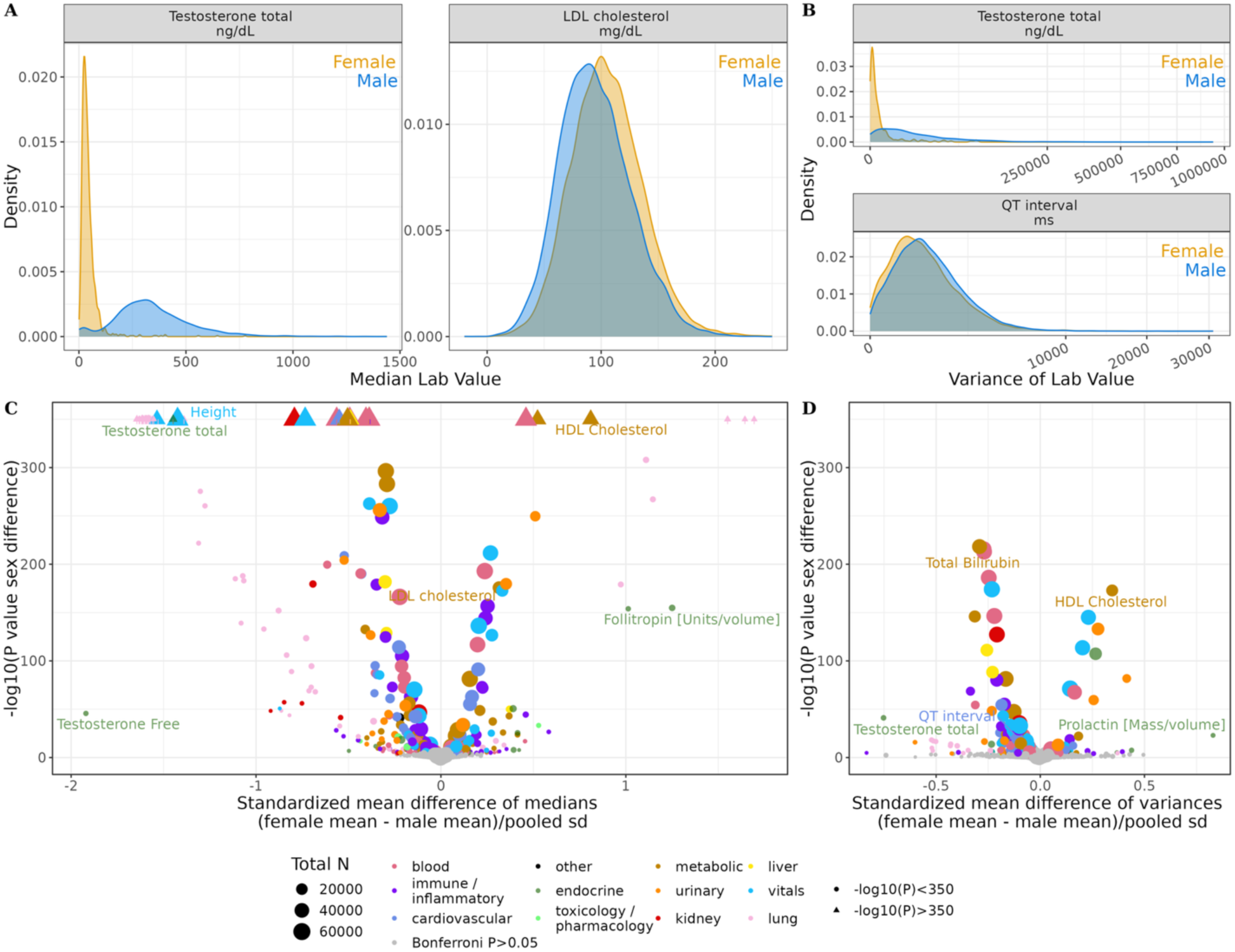
Sex differences in the distributions of clinical quantitative traits in BioVU: Distributions of A) per-person median values and B) per-person variances of 508 quantitative clinical traits with significant sex differences in per-person medians (testosterone and LDL cholesterol) or per-person variances (testosterone and QT interval) in males (blue) and females (yellow) in BioVU European ancestry participants C) Volcano plot of sex differences in 508 quantitative clinical traits per-person medians in European ancestry participants. On the x-axis is the difference between the female and male mean divided by the pooled standard error and on the y axis is the –log10 transformation of the p-value from the t-test. Quantitative clinical traits with a p-value<0.05/512 are colored according to the category of trait, those with p-value>0.05/512 are in grey. The size of the point corresponds to the sample size for that quantitative trait. Some quantitative traits had –log10(p-values)>400, these are represented as triangles at the edge of the plot. D) Volcano plot of sex differences in 463 quantitative clinical traits per-person variances in European ancestry participants in BioVU.

### Sex-stratified genome-wide association studies (GWAS) replicate known and identify novel associations

Across the 508 sex-stratified GWAS of per-person median values, we identified 2010 independent loci with p<5e-8 (1734 excluding the *MHC/HLA* region, Supplementary Tables 3-7, Supplementary Figure 3). Locus count per lab correlated with sample size (Pearson r=0.41, *P*=7.30e-07, Figure 2A).

**Figure 2.**
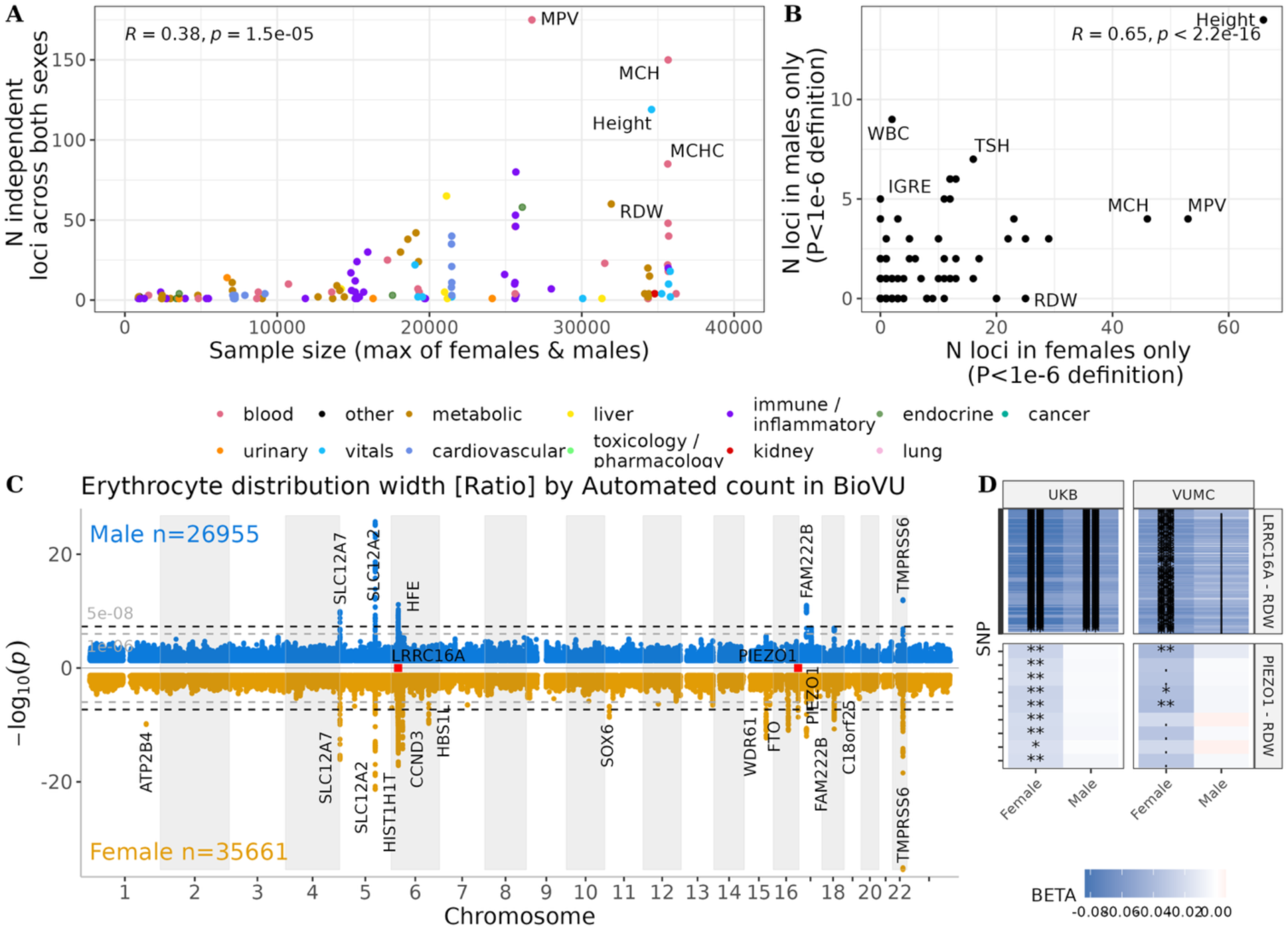
A) Count of independent loci across both sexes (excluding MHC region) per clinical quantitative trait plotted against the sample size in BioVU. B) Count of independent loci with a genome-wide significant (P<5e-8) in one sex but no variant in the locus with P<1e-6 (suggestive) in the other sex (excluding MHC region). C) Example Miami plot showing p-values for RDW - Erythrocyte distribution width for males (top, blue) and females (bottom orange). Highlighted in red are *LRRC1CA* and *PIEZO1.* The locus near *PIEZO1* was the only locus that was also sex specific (P<5e-8 in one sex, no P<0.01 for a variant in the locus in the other sex) in the UKB. The locus near *LRRC1CA* met the suggestive definition of sex specificity in BioVU (P<5e-8 in one sex, no P<1e-6 in the other sex) and showed a concordant sex difference, but not complete sex specificity, in the UK D) Heatmap of effect size estimates (betas) for association with RDW in the UK Biobank (UKB) and BioVU (BioVU) at *PIEZO1* and *LRRC1CA.* While the sex specificity of the *PIEZO1* replicated in the UKB, the locus at *LRRC1CA* was genome-wide significant in both sexes in the UKB, although the association was stronger in females.

Most significant loci reproduced known associations: a median 90% (mean 72%) of loci per lab contained variants or mapped to genes previously reported as associated with a corresponding phenotype in the GWAS catalog. However, 421 loci from 69 labs were not previously reported (Table 1, Supplementary Table 6), including a disproportionate share (53%) of the 19 X chromosome loci (Supplementary Figure 3). Additionally, 106 loci from 17 labs had no corresponding GWAS catalog results (Supplementary Table7).

**Table 1:** Identified sex-specific or sex-interaction loci in the Vanderbilt University Medical Center biobank, BioVU, and their confidence based on replicability in the UK Biobank (UKB) or University of Colorado Center for Precision Medicine Biobank (CCPM).

|  |  |  |  |  |  |  |  |  |  |
| --- | --- | --- | --- | --- | --- | --- | --- | --- | --- |
| Trait | Gene (SNP-effect allele) | Sex effect of SNP in BioVU<sup>2</sup> | Confidence<sup>1</sup> | BioVU female beta | BioVU male beta | UKB female beta | UKB male beta | CCPM female beta | CCPM male beta |
| Erythrocyte distribution width (RDW) | *PIEZO1* (rs837763-C) | Single sex effect | High | -0.04 | -0.01 | -0.03 | -0.003 | NA | NA |
| LDL cholesterol (LDL.C) | *APOE* (rs7412-T) | Differential effect | High | -0.46 | -0.30 | NA | NA | -0.54 | -0.35 |
| Cholesterol | *APOE* (rs7412-T) | Differential effect | High | -0.29 | -0.16 | -0.42 | -0.31 | -0.37 | -0.22 |
| Uric Acid (UricA) | *SLCA29* (rs9994216-G) | Differential effect | High | -0.26 | -0.12 | -0.46 | -0.24 | -0.32 | -0.13 |
| Cholesterol | *PVRL2* (rs2927472-T) | Single sex effect/<br>Differential effect | Low (sex direction replicated in UKB but not sex specificity) | -0.09 | -0.02 | -0.12 | -0.08 | NA | NA |
| White blood cell count (WBC) | *PGAP3*- (rs1495100-T) | Single sex effect/<br>Differential effect | Low (sex direction replicated in UKB but not sex specificity) | 0.04 | 0.01 | 0.04 | 0.038 | NA | NA |
| Weight | *AGBL1* (rs11073644-T) | Opposite effect | Moderate (replicated in CCPM, not UKB) | -0.03 | 0.03 | -0.002 | -0.01 | -0.02 | 0.02 |
| Height | *MYO3B* (rs11884023-T) | Opposite effect | Moderate (replicated in UKB, not CCPM) | -0.02 | 0.03 | -0.009 | 0.005 | 0.008 | -0.002 |
| aPTT in Platelet poor plasma (PTT.pt) | *RP11-12N13* (rs9819879-C) | Opposite effect | Low (nominal replication in CCPM, trait not in UKB) | -0.06 | 0.08 | NA | NA | -0.05 | 0.04 |
| Erythrocyte distribution width (RDW) | *ALDH1A2* (rs28409799-A) | Opposite effect | Low (nominal replication in both UKB and CCPM) | -0.06 | 0.06 | -0.01 | 0.01 | -0.006 | 0.03 |
| <sup>1</sup>High confidence: Replicated ($P < 0.0007$ (Bonferroni correction for sex interaction loci testing)) in all both biobanks or replicated in at least one biobank and missing trait in the other biobank. Moderate confidence: Replicated in at least one biobank (Bonferroni correction). Low confidence: Nominal replication ($P < 0.05$) in one biobank with concordant directions of effect in both sexes. Potential<br><sup>2</sup>Differential effect = SNP was significant in both sexes (same direction of effect), but had a stronger association in one sex; Single sex effect = SNP was only significant in one sex; Opposite effect = SNP was significant in both sexes, but the effect estimate was in opposite directions between sexes. Sex interaction loci from post-hoc t-test for sex differences were tested in CCPM and UKB, sex specific loci identified in BioVU were only replicated in UKB.<br>NA – Either no exact matching trait in UKB or single sex loci were only replicated in UKB, not CCPM | | | | | | | | | |

We defined sex-specific loci as loci with a significant (P<5e-8) variant in one sex but no variants in the locus reaching significance in the other sex using strict (all P>0.01) and lenient (all P>1e-6) cut-offs. Under the strict definition, most loci had effects in both sexes (91%, 179 sex-specific loci). Under the lenient definition, 52% of loci were sex-specific loci (Figure 2B, 1041 sex-specific loci), likely reflecting BioVU’s smaller male sample sizes for most traits. Sex-specific loci were less likely to be previously reported in the GWAS catalog than loci significant in both sexes under either definition (P>1e6 Chi Sq=57, P=4.70e-13, P>0.01 Chi Sq=31.32, P=1.58e-07).

To test replicability of strictly defined (P>0.01) sex-specific loci, we used sex-stratified UKB summary statistics from the Neale lab. One BioVU sex-specific locus near *PIEZO1* and associated with erythrocyte distribution width replicated in UKB, of 115 tested loci from 41 matched traits. Many sex-specific loci in BioVU reached P<5e-8 in both sexes in the larger UKB sample, however, at 60% of these loci, the same sex had a larger magnitude of effect and smaller p value across most variants in both cohorts, consistent with replicable sex differences in effect magnitude rather than true sex specificity (example of *LRRC16A* and RDW shown in Figure 2D, Supplementary Table 8, Supplementary Figures 4-9). Female effect estimates were more consistent with UKB than male estimates (Pearson r=0.72 vs 0.53, both P<2e-16), with variation by category of lab (Supplementary Figure 10).

Sex-stratified GWAS of per-person variances identified 116 loci, 68% overlapping loci for the corresponding median trait, concentrated in lipid and blood cell traits;75% were significant in both sexes (Supplementary Table 9).

### Post-hoc interaction tests identify sex-differentiated genetic effects

Even when a locus is associated with a lab in both sexes, the allelic direction (opposite-effect) or magnitude (differential-effect) of the association could differ by sex. We therefore performed post-hoc interaction tests at variants with a nominal (P<0.05) association in at least one sex (Methods), identifying 660 variants with sex interactions across 151 loci and 118 traits (Benjamini-Hochberg FDR within-trait (Supplementary Table 10. The three topmost significant such loci were near *APOE* (LDL), *DLG2* (heart rate from EKG) and *BMPER* (vitamin D, Figure 3). The number of sex interaction loci per trait associated with the standardized mean sex difference in trait value (p=0.03 restricting to traits with n>10,000).

**Figure 3:**
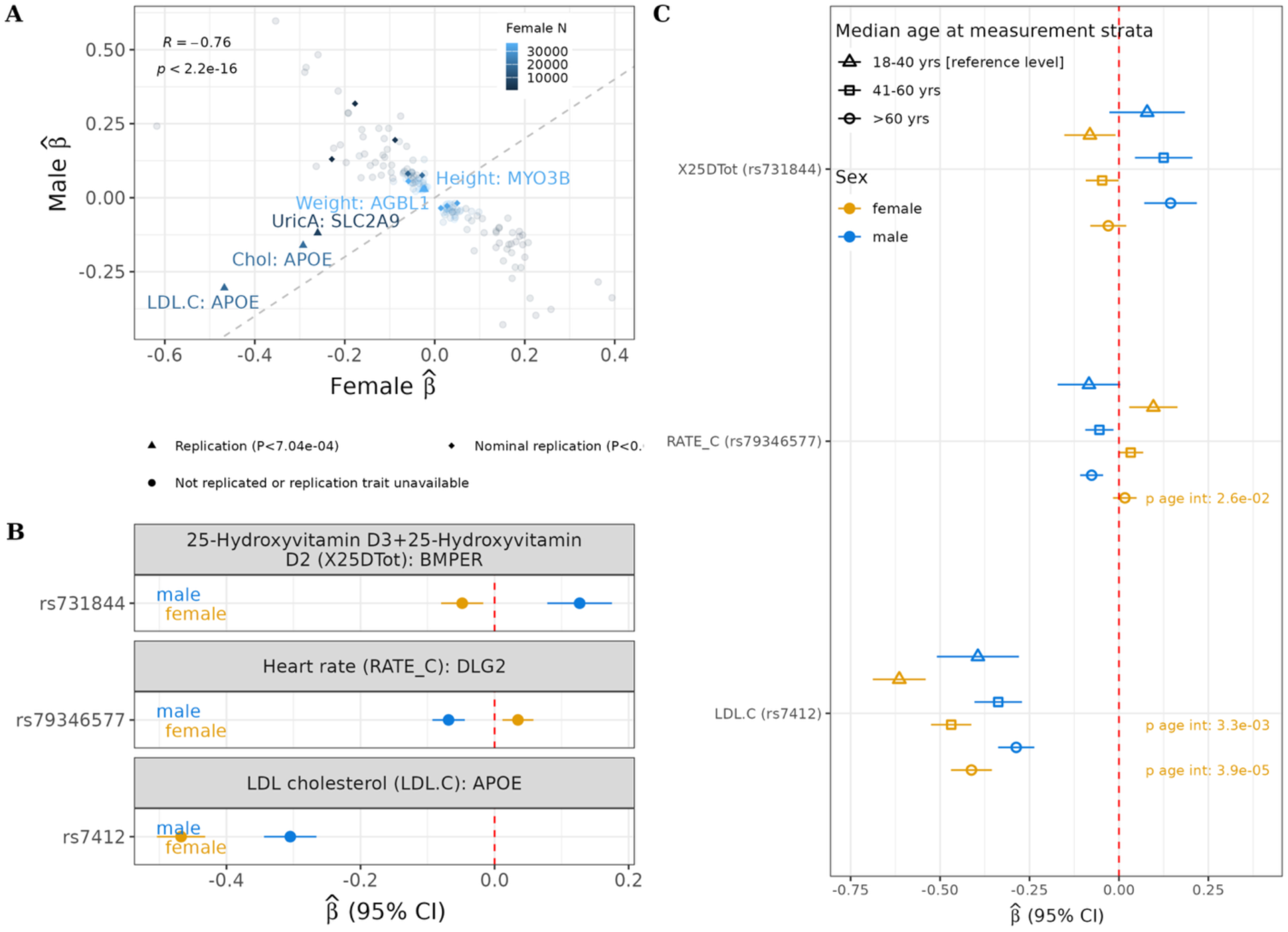
A) Sex interaction SNPs at FDR<0.05 plotted by effect estimates (beta) in males (y-axis) and females (x-axis), colored by female sample size. Sex interaction loci with at least nominal replication in UKB or CPM Biobank are highlighted, sex interaction loci that did not replicate or were from a trait without a mapped trait in a replicating biobank are greyed out. B) The top three most significant sex interaction loci in BioVU are shown with effect estimates for female (orange) and male (blue). C) The same three sex interaction loci shown in (B) further stratified by median age at that clinical trait’s measurements.

Of sex interaction loci in BioVU, 99% showed opposite allelic effects but only differential-effect loci replicated in UKB or CCPM (n=3 at P<5e-8 of 71 tested loci): a locus near *APOE* with LDL cholesterol and total cholesterol, and a locus near *SLC2A9* with uric acid ((supplementary Figure 11). Two opposite effect loci for height and weight replicated at P<0.05/71=7.04e-04 but had inconsistent effects across CCPM and UKB (Table 1). Only 11 (12% of opposite-effect loci had nominal replication (P<0.05, Table 1, Supplementary Figure 12).

Age-stratified follow-up analyses found more sex-interaction SNPs with significant age interactions (p<0.05) than expected by chance (Kolmogorov Smirnov *P*=0.04).

### Estimated trait heritability is similar between males and females

Male and female SNP-based heritability estimates converged as sample size increased and were highly concordant among well-powered labs (n>10,000 per sex) using both GCTA estimation (Pearson r=0.96, *P*=9.29e-55) and LD score regression (Pearson r=0.8, *P*=1.81e-24). Using high-definition likelihood estimation, 90/104 of the well-powered labs were significantly correlated between sexes (P<0.05). Exceptions included urinary and clotting labs, and some labs with strong sex interaction loci, notably LDL cholesterol. The between-sex correlation was lower in BioVU than UKB, likely reflecting smaller sample sizes (Supplementary Figures 13-14, Supplementary Table 11).

Traits with larger standardized differences in mean values between males and females did not have lower genetic correlation between sexes; if anything, the relationship was positive after controlling for sample size (Supplementary Figure 15).

### Sex interaction loci associate with clinical disease

We investigated relationships between the top variant in two loci with strong replicated sex-differentiated effects (rs7412-*APOE*, rs1014290-*SLC2A9*), first observations of their associated labs (LDL cholesterol, uric acid), and related diseases (ischemic heart disease, gout). Both diseases and their treatments were more common in males than females (Figure 4A). In individuals with first lab measurements at least 2 years prior to these diagnoses (Methods), the sex interaction persisted after adjusting for medication use, EHR utilization, and comorbidities (Methods and Supplementary Table 12). In these cohorts, both loci also showed strong age-by-sex-by-variant interactions (Figure 4B): females had lower LDL cholesterol than males before age 50 (most evident with 1-2 rs7412 T alleles), but higher LDL cholesterol after age 60 (most evident with 0 rs7412 T alleles).

**Figure 4:**
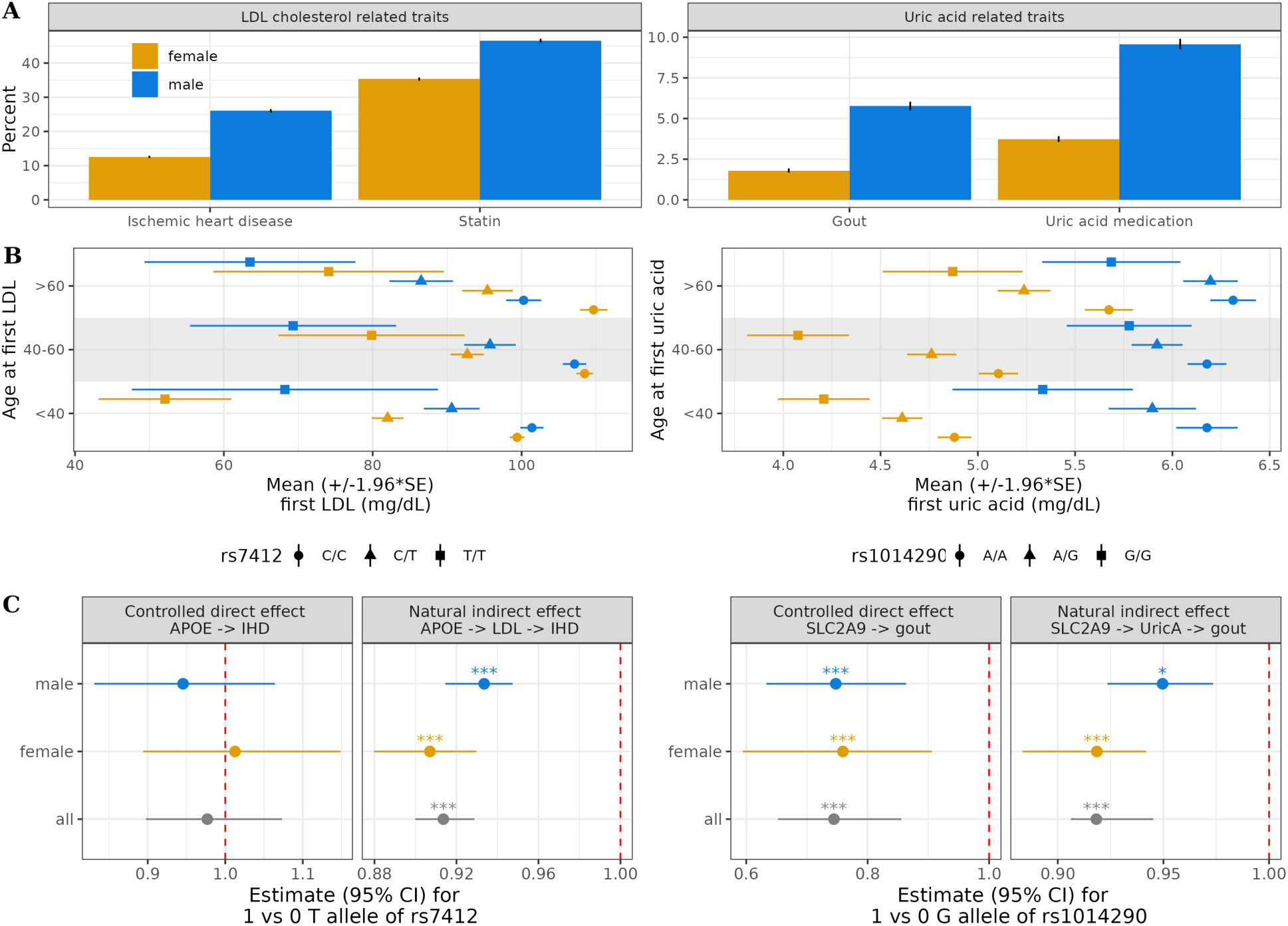
A) Males had higher prevalences of ischemic heart disease, statin use, gout, and uric acid medication use than females in the cohort. B) the sex differences in the effects of the variants in APOE and SLC2A9 on first measurement of LDL and uric acid become attenuated after age 60, even when excluding individuals with reports of statins or uric acid medications and restricting to lab measurements obtained at least two years before diagnosis of ischemic heart disease or gout or from those without those diagnoses (respectively). C) first measurement of LDL and uric acid mediate part of the effect of the variants in *APOE* and *SLC2AS* on ischemic heart disease and gout respectively, with a larger indirect effect (effect mediated through the clinical biomarker) in females than in males.

Mediation models showed significant indirect effects of rs7412 on ischemic heart disease through LDL cholesterol, and of rs1014290 on gout through uric acid (Supplementary Table 13-14, both slightly stronger in females than males (female:male indirect effect ratios, rs7412->LDL->ischemic heart disease=0.97 (95% CI: (0.94, 1), p=0.09), rs1014290->Uric acid->Gout=0.97 (95% CI: (0.92, 1.02), p=0.19)).

A phenome-wide association study (PheWAS) of lab ascertainment identified potential sources of ascertainment bias - LDL cholesterol and uric acid ascertainment tracked their associated diseases (lipid disorders, gout) in both sexes, with sex-specific enrichment for secondary conditions (e.g., osteoporosis and fatigue in females for LDL; vitamin D deficiency in males for uric acid). Among two traits with strong, non-replicating opposite-effect loci, vitamin D (locus near *BMPER*) ascertainment was more strongly associated with chronic kidney disease and renal failure in males, and heart rate from EKG (locus near *DLG2*) was more strongly associated with digestive disorders and anxiety in females and peripheral vascular disease in males (Supplementary Table 15; Supplementary Figures 16-17). Inverse-probability weighting to the entire BioVU genetic sample or general US population (Methods) left the *APOE*-LDL and *SLC2A9*-uric acid sex differences unchanged (Supplementary Figures 18-19, Supplementary Tables 16-17) but removed the vitamin D association (driven largely by reweighting to general-population chronic kidney disease prevalence); the *DLG2*-heart rate association was unaffected.

### Colocalization with sex-biased expression quantitative trait loci (eQTLs)

We performed colocalization analyses between sex-stratified GWAS statistics and 845 sex-biased eQTLs from breast tissue. We focused on breast tissue because it contained the largest number of previously identified sex-biased eQTLs, providing the greatest power for this hypothesis-generating analysis despite the diverse tissue origins of the labs. We identified 31 loci with posterior probability of shared causal variant (PP4) >90%, and 386 with PP4>50% (Figure 5A, Supplementary Table 18). Some sex-biased eQTLs colocalized with multiple traits. An eQTL signal for *ZKSCAN3*, a transcriptional repressor and autophagy regulator, colocalized more strongly with male immune/inflammatory lab GWAS signals. An eQTL signal for *SDHDPC*, a pseudogene, colocalized more strongly with female blood lab GWAS signals (Figure 5B), and an eQTL signal for *CDCA7L,* a multifunctional transcriptional regulator, colocalized more strongly with a GWAS signal for chloride in females (Figure 5E, other examples in Figure 5D and 5F). Among traits with PP4>50%, blood traits were modestly overrepresented among females and kidney traits among males (Figure 5C, Chi Sq=24, *p*=0.02).

**Figure 5:**
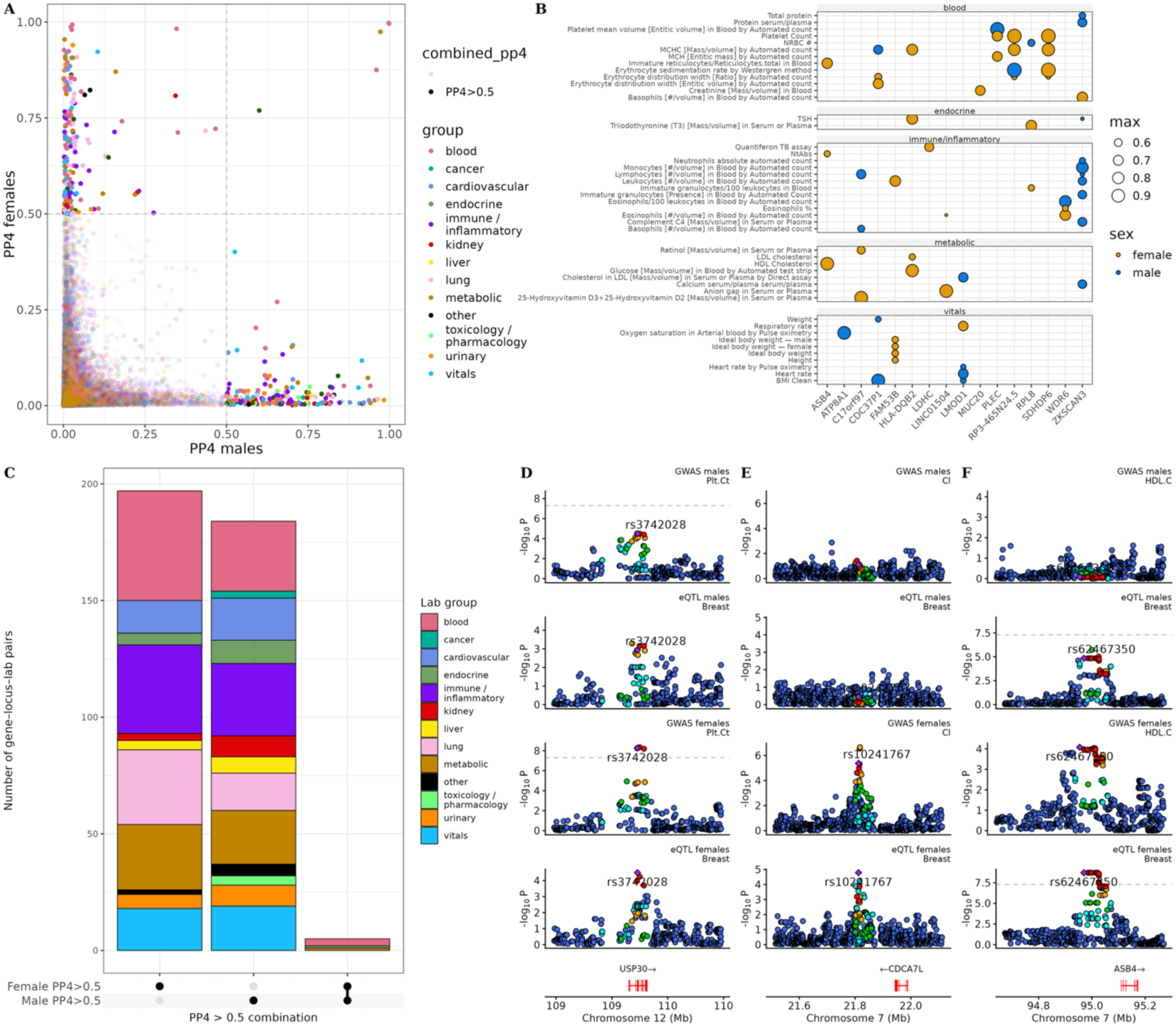
A) Posterior probability of colocalization (PP4), representing the probability that the eQTL and GWAS signals share a single causal variant, for sex-biased eQTLs in breast tissue using sex-stratified eQTL data from Oliva *et al* and BioVU sex-stratified GWASes. B) Colocalization results for genes (x-axis) and traits (y-axis) with multiple colocalization signals. Circle size corresponds to PP4, and color indicates the sex with the stronger colocalization signal. C) Upset plot of the number of colocalized loci with female, male or both sexes having PP4>0.5. D) Locus zoom plot of colocalization of USP30 and Platelet count (Plt.Ct). E) Locus zoom plot of colocalization for *CDCA7L* and chloride (Cl). F) Locus zoom plot for colocalization of *ASB4* and high density lipoprotein cholesterol (HDL.C).

## Discussion

We present the first systematic analysis of sex differences in the genetic architecture of quantitative traits in EHR-linked biobanks, demonstrating that EHR data can identify robust sex-biased genetic effects and provide insight into their biological contexts. Significant mean sex differences existed in >50% of quantitative traits. While most labs showed strong genetic correlation between the sexes, we identified individual loci with sex-biased effects, including a female-specific association for erythrocyte distribution width near *PIEZO1* and effect magnitude differences for lipids near *APOE* and uric acid near *SLC2AS*. Age-stratified analyses revealed sex differences varied across the lifespan, suggesting a role of sex hormones, and mediation analysis suggests sex-biased loci-trait relationships contribute to diseases with known sex prevalence differences. Colocalization of sex-stratified GWAS with sex-biased eQTLs implicated *ZKSCAN3* and *CDCA7L*, among others. Overall, sex differences in the genetic architecture of quantitative traits seem driven by widespread but small effect magnitude differences, which will require very large sample sizes to detect, and which EHR-based biobanks can help uncover.

Our results support differential effect magnitude, or amplification, as the predominant model for sex differences in genetic architecture^6^. We identified one replicable sex-specific locus across biobanks. Many loci originally identified as sex-specific in the discovery BioVU sample failed to replicate sex-specificity, suggesting that most sexspecific loci detected at the current sample size reflected power differences from amplified effects rather than true sex specificity. Most loci identified from post-hoc interaction tests had opposite effect directions, but only those with concordant direction and differences in magnitude replicated, suggesting opposite directions of effect are easier to detect statistically but less biologically plausible^14^. Opposite effect interaction loci may reflect disease-specific contexts, ascertainment biases, or spurious findings. Reliably cataloging sex interaction loci will require very large sample sizes and replication efforts. We believe this should be a precision medicine research priority as pinpointing sex-interactions offers invaluable insights into sex biology on disease states.

Our study demonstrates that EHR-linked biobanks are valuable for this discovery work, offering opportunities to identify sex-interactions before disease onset and during disease progression. EHR-linked biobanks recapitualated results from healthy cohorts: sex differences in associations of *APOE* with cholesterol traits and *SLC2A9* with uric acid have been previously reported^8,10,15–17^, including in the UKB^7,18^. We extended previous findings by showing these sex differences were strongest before age 60 which was previously reported for *SLC2A9* but not *APOE*^16^. We found a broader enrichment for age interactions across sex-interaction loci, underscoring that sex differences vary across the life course and likely reflect sex hormone regulation. In disease contexts, we showed that LDL cholesterol and uric acid mediated effects of *APOE* and *SLC2A9* on ischemic heart disease and gout respectively, with stronger indirect effects in females than males. Sex differences were apparent in indirect but not direct effects, which may explain why previous studies have not reported sex-*SLC2A9* interactions in gout^15,18^. These findings suggest sex differences in the genetic architecture of labs can influence diseases with known sex- and age-related prevalence differences.

Sex-biased gene expression and its hormonal regulation is one hypothesized mechanism for phenotypic sex differences^1^. Several novel loci we identified map to genes with plausible sex-differential biology. For instance, *PIEZO1,* which associated with erythorcyte distribution width^19,20^, exhibits differential expression by sex in a rat model of hypertension^21^. *PVRL2* (near a sex biased cholesterol locus), is a plasma cholesterol– responsive gene and its expression is linked to atherosclerosis^22,23^ while *PGAP3* (near a sex biased white blood cell count locus) has been associated with asthma^24^, inflammatory bowel disease^25^, and menopause age^26,27^, conditions which have sex prevalence differences or are sex specific. Colocalization analyses between our sex-stratified GWAS and sex-biased eQTL data identified other candidate genes, including *ZKSCAN3* (a master transcriptional regulator of autophagy^28^ and xist expression^29^, near loci associated with immune/inflammatory traits), and *CDCA7L* (a transcriptional regulator of cell proliferation near a locus associated with chloride and previously linked to male-pattern baldness^30^, prostate cancer^31^, and breast cancer^32^). None of the sex interaction loci from our GWASes colocalized with sex-biased eQTLs, likely reflecting limited power for sex-stratified eQTLs and GWASes^33^.

While we replicated sex differences previously reported in healthy cohorts, EHR-linked biobanks differ from healthy cohorts in important ways. EHR populations tend to be sicker, and labs may be ascertained selectively to inform diagnosis or monitor disease and treatment. Thus, differences may arise from disease-context dependent effects that drive biology and/or from ascertainment biases due to health care access or utilization differences. Both can differ by sex and may be difficult to disentangle^34–36^. For example, our GWAS of vitamin-D levels replicated known associations and identified post-hoc evidence for sex interaction with variants near *BMPER*. While several members of the BMP protein family bidirectionally interact with sex hormones^37,38^ this was the first evidence, to our knowledge, suggesting that *BMPER* may also be sensitive to sex. To thoroughly investigate this finding and control for sex differences in ascertainment biases, we applied inverse-probability weighting (IPW) and found that the result was driven by males with renal failure, a possible sex-by-disease context association. Convergent data from mouse models show that reduced expression of *BMPER*, a secreted protein expressed in the kidney, is a primary driver of renal tubular fibrosis in the context of renal disease^39^. Renal tubules are the site of hydroxylation of vitamin D to its active form, calcitriol. Thus, the IPW-adjusted finding raises the hypothesis that *BMPER* may also be involved in tubule health in human renal disease, for which vitamin D acts as a read-out. However, the *BMPER*-vitamin D association did not replicate in CCPM even when stratifying by renal disease (data not shown). While we cannot rule out winner’s curse^40^ or the possibility of a simple false positive in our original and IPW-adjusted GWAS, the lack of replication may also be confounded by environmental differences between the Nashville (Tennessee Valley) and Denver (Rocky Mountain) populations including the documented effect of increased altitude on Vitamin D production^41,42^.

Nevertheless, limited power and winner’s curse^40^ likely explains why many sex-interaction loci did not replicate and underscores the need for replication; to date, most such investigations have used the UKB, and our results suggest that EHR-linked biobank consortia could enable better-powered discovery and replication across biological contexts. Other limitations include ascertainment bias, despite our IPW correction, and unmeasured confounding. Given very limited sample sizes for other BioVU ancestry groups, we restricted to individuals of European ancestry, underscoring the need for more diverse cohorts. These limitations are offset by the EHR’s rich phenotypic depth, which enabled epidemiological follow-up.

## Online Methods

### Description of cohort

We investigated sex differences in the genetic architecture of clinical quantitative traits using data from the deidentified Vanderbilt University Medical Center (VUMC) electronic health record (EHR) (referred to as the Synthetic Derivative, or SD) and linked genetic biobank, BioVU. This analysis used data from the June 2023 data freeze of the SD and the study cohort was restricted to individuals in BioVU who were genotyped on the Illumina Expanded Multi-Ethnic Genotyping Array (MEGAex)^43^. The SD includes information on demographics, vital measurements, ICD9 and ICD10 codes, clinical quantitative measurements, medications, and clinical notes, organized into the Observational Medical Outcomes Partnership (OMOP) Common Data Model (CDM)^44,45^. The SD applies date shifting within a 1-year time frame to reduce the potential for participant reidentification. Detailed information about BioVU’s data management and quality control, ethical considerations, and continuing patient engagement have been previously published^13,46–49^. The study was approved by the VUMC IRB (IRB# 212285).

The University of Colorado’s Colorado Center for Personalized Medicine (CCPM) biobank was used for replication. The CCPM Biobank is an EHR-linked genomic resource jointly developed by the University of Colorado Anschutz Medical Campus and UCHealth and currently includes approximately 100,000 genotyped participants, of whom approximately 80% are of European ancestry. Participants were genotyped using customized versions of the MEGAex, with additional whole-exome sequencing data available for a subset of participants. Genotype data were imputed using the TOPMed reference panel, yielding approximately 49.5 million variants for downstream analyses. Clinical phenotypes are derived from longitudinal electronic health records from UCHealth and harmonized to the OMOP Common Data Model, enabling large-scale genomic and phenotypic analyses across diverse populations.

### Cleaning and preparation of quantitative clinical trait phenotype data

Quantitative clinical trait data was extracted for all participants in the OMOP CDM measurements table. For each quantitative clinical trait, units were standardized using the Unified Codes of Units of Measurement (UCUM) vocabulary and javascript library developed by the Lister Hill National Center for Biomedical Communications. Briefly, the most common unit reported for each quantitative clinical trait was selected. If this most common unit could be validated as a UCUM unit, then all values for observations in other commensurable UCUM units were converted to this unit. Observation values in non-commensurable units set to missing and excluded. If the most common unit could not be validated as a UCUM unit, then only observations in the most common unit were kept.

Subsequent to unit conversion, clinical quantitative trait data was processed using the previously described QualityLabs pipeline^4^. Only quantitative clinical traits with at least 1000 observations from at least 100 people were processed through QualityLabs. Missing and infinite quantitative clinical trait values were excluded, as were observations taken when an individuals was <18 years of age. Sample means and standard deviations were calculated, and outliers (≥ 4 standard deviations outside the mean) were excluded. Next, per-person summary measures were calculated, specifically, the per-person median value, age at per-person median, per-person number of observations, and per-person variance (for individuals with >1 observations of the quantitative clinical trait). To adjust for the effect of age on clinical quantitative traits, per-person median values and per-person variance values were residualized for three splines of age at per-person median and then inverse-normalized. Outlier filtering, per-person summary measure calculation, age-adjustment and inverse normalization were performed both across all adults and within sex. Overall measurements were used to test for sex differences in the distributions of quantitative clinical traits, while within sex measurements were used for all genetic association analyses.

Only those clinical quantitative traits with at least 1000 participants of European ancestry who also had MEGA array genotype data were included in the discovery (BioVU) analysis. Clinical quantitative traits were excluded if they could not be reliably mapped to a known clinical concept or encoded information unlikely to be related to human genetics (e.g., collection time of urine specimens, antibiotic resistance of collected specimens). Some quantitative clinical traits were exclusively or overwhelmingly measured in a single sex (n<100 individuals of one sex); we label these traits as ‘sex-specific’. We mapped clinical quantitative trait names from BioVU to terms from the experimental factor ontology (GWAS catalog) and UK Biobank traits using OMOP CDM vocabulary and otherwise mapped to the closest matching string based on Jaccard similarity, followed by manual review.

### Cleaning and preparation of genotype data

The discovery analysis included only BioVU participants with genotype data measured on the MEGAex array^43^ who were determined to cluster with spiked-in 1000 genome samples of known recent European ancestry using principal components analysis and Eigenstrat^50^.

Data were imputed using the Michigan Imputation Server with the Haplotype Reference Consortium reference panel. Imputed genotype data were subjected to several quality control filters optimized for sex difference analysis using the package GXwasR^51^ and plink^52^, namely only biallelic variants were considered, variants with imputation R^2 < 0.3 were removed, variants with minor allele frequency <0.05 or missingness >0.05 within each sex were removed, and variants in violation of Hardy-Weinberg equilibrium (p<1e-10) were removed. Samples were removed that had a missingness rate >0.10 or were outside 3 SD of heterozygosity.

### Statistical analysis of sex differences in quantitative clinical traits

We tested for differences between the male and female sample means of each quantitative clinical trait summary metric (e.g., per-person medians, per-person variances, inverse normalized and age-adjusted per-person medians and variances) using t-tests, correcting for multiple testing across traits using the Benjamini-Hochberg method. We calculated the standardized mean differences as the difference between the female and male sample mean divided by the pooled standard error. We used a Benjamini-Hochberg false discovery rate correction to assess significance, correcting for the number of tested quantitative clinical traits.

### Sex stratified genome-wide association analyses (GWASs)

We performed sex-stratified genome wide association analyses using Regenie, a computationally efficient method for fitting whole-genome regression model and testing for genomic associations^53^. Regenie uses an imputation scheme to handle missing data, and it is recommended to analyze traits in groups that have similar missingness patterns with <20% missingness. Therefore, we used hierarchical clustering and tree cutting to group quantitative clinical traits with similar patterns of missingness, iteratively reclustering within branches until no single trait in a cluster had a missingness rate > 20%. Quantitative clinical traits were processed through the Regenie pipeline in these clusters, which generally reflected clinical ascertainment patterns (e.g., complete blood count panel counts clustered together, EKG components clustered together).

Ten genomic principal components (PCs) were calculated within the set of participants in each cluster, using flashPCA and only variants from the autosomal chromosomes after excluding high linkage disequilibrium regions. The whole genome model was fit to generate leave-one-chromosome-out genomic predictors, removing any variants with a minor allele count within the cluster participants of < 100 and controlling for the 10 genomic PCs. For fitting the whole genome model in Regenie, it is recommended to use several hundred thousand common markers from a micro-array, i.e, the directy genotyped variants. We restricted the preimputation markers to just those that passed the postimputation quality control filters described above while additionally filtering out high linkage disequilibrium regions.

Then we used Regenie to perform single variant association testing between variant dosage data from post-imputation QCed genomic data and the inverse-normalized and age-residualized per-person median and variance clinical quantitative trait measures, again controlling for the 10 genetic principal components.

Genome-wide association results for each trait were clumped into loci using the plink clump functionality access through the GXwasR package^51^, a p-value threshold of 5e-8, an R^2 threshold of 0.5 and 250kb. We removed loci that had only a single variant. We defined sex-specific loci as those where there was a genome-wide significant variant (P<5e-8, lead variant) for one sex, but no variant in linkage disequilibrium with the lead variant was either nominally (all P>0.01, strict definition) or suggestively significant (all P>1e-6, lenient definition) in the other sex. In supplemental tables, we also provide a definition using all P>5e-8 (genome-wide significant).

### Post-hoc sex by genotype interaction tests

We tested for sex differences in the effect size of the association between variants and clinical quantitative traits using a post-hoc t-test as implemented in the SexDiffT function of GXwasR^51^ . The t-test is of the form 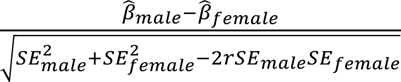 where *r* is defined as 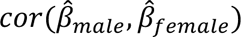.

To reduce the number of multiple comparisons, we only tested for sex differences in the effect size at variants that had a nominal (P<0.05) association with the clinical quantitative trait in at least one sex, then applied a Benjamini-Hochberg false discovery rate correction for the number of sex difference tests within-trait (sdSNPs). This approach is based on the rationale that sex differences in genetic effects are unlikely to be biologically meaningful in the absence of evidence for association in either sex. Nonetheless, we acknowledge this approach may inflate false discovery rates as the sex-stratified test for effect is not statistically independent of the post-hoc sex difference test. Therefore, we also report whether sex difference tests achieved genome-wide (P<5e-8) significance.

We used chi-square tests to test for enrichment of protein coding genes in all GWAS significant SNPs, in sex-specific SNPs, and in sdSNPs.

### Calculation of heritability and genetic correlations

Sex stratified heritability estimates were calculated in two ways: 1) using the GCTA-GREML approach and individual-level genotype and clinical quantitative trait data and 2) using LD score regression and sex-stratified summary statistics from GWASs^54,55^. We also calculated genetic correlations between sexes using the high definition likelihood inference method and sex-stratified summary statistics from GWASs^56^. All three methods were applied using the GXwasR package^51^.

For GCTA GREML heritability estimation, phenotypes were run in the same clusters as used for the sex-stratified GWAS. Related individuals were excluded from this analysis using a *π*+ threshold of 0.025.

For the LD score regression, precalculated LD scores from Thousand Genomes Project participants were used.

In addition to calculating genetic correlation between sexes, we calculated genetic correlation with publicly available sex-specific genome wide association statistics from the UK Biobank, as provided by the Neale lab^57^.

### Follow-up epidemiological analyses

To test whether sex differences in the effect estimates for sdSNPs varied across the life-course, we performed follow-up age stratified analyses for sdSNPs. We extracted the top-most significant variant per gene-quantitative trait pair from the list of sdSNPs and reran linear regression models for the inverse normalized and age adjusted median within strata of age at median measurement that approximately mirror reproductive life stages for females: 18-40 years of age, 41-60 years of age and >60 years of age. We adjusted for the cluster-specific 10 genomic PCs and the LOCO predictions from Regenie step 1. We also performed an interaction test using a linear regression with an interaction term between the sdSNP and the age strata. We used a Kolmogrov Smirnov test to test if the distribution of interaction p-values from the sdSNP-age strata interaction were significantly different from a uniform distribution.

We constructed prospective cohorts for ischemic heart disease (IHD) and gout analyses by restricting to individuals with at least one measurement of LDL cholesterol or uric acid, respectively, who lacked the relevant outcome diagnosis at the time of or within two years following their first laboratory measurement. We examined the association between sex-stratified lead variants (*APOE* rs7412 and *SLC2A9* rs1014290) and the first observation of these respective laboratory traits in linear models adjusting for age (modeled with cubic polynomials), medication use (statins or uric acid-lowering drugs), number of ICD codes as a proxy for healthcare utilization, and comorbidity burden from PheCodes (diabetes, hypertension, atrial fibrilation, anemias, chronic liver disease or cirrhosis, renal failure, depression, dementia, COPD, and asthma). We tested sex-by-genotype interaction terms to assess whether genetic effects on laboratory traits differed between sexes. Mediation analyses were conducted using the g-formula approach implemented in the CMAverse package^58^, with first observation of LDL cholesterol and uric acid as mediators of the effect of rs7412 and rs1014290 on IHD and gout, respectively. Bootstrap confidence intervals were estimated with 10000 resamples. Sex differences in natural indirect effects were tested by computing the ratio of female to male indirect effects on the log scale using the delta method.

To characterize ascertainment bias in EHR-derived laboratory data, we conducted phenome-wide association studies (PheWAS) using binary indicators of selected laboratory traits’ ascertainment as outcomes in both sex-stratified and sex-combined models with sex-by-phecode interaction terms. We then applied inverse probability weighting (IPW) to reweight the ascertained subpopulation to be representative of the full BioVU genetic cohort with respect to phecodes associated with ascertainment. We used the top 20 phecodes associated with ascertainment in sex-stratified analyses, as well as the top phecodes associated with an interaction term between sex and ascertainment, collapsing phecodes wtihin nested chapters. A second set of weights additionally reweighted to target phecode prevalences from general US population estimates from the Global Burden of Disease Study 2019^59,60^ for the top most significant ascertainment and sex by ascertainment phecodes. For LDL cholesterol, we reweighted to target prevalences for disorders of lipid metabolism (phecode 272, US prevalence 15%, BioVU prevalence 34%) and osteoporosis (phecode 743, US prevalence 4%, BioVU prevalence 12%). For uric acid, we reweighted to target prevalences for gout (phecode 274, US prevalence 3.5%, BioVU prevalence 4.3%) and vitamin deficiency (phecode 261, US prevalence 3%, BioVU prevalence 12%). For heart rate from EKG, we reweighted to target prevalences for disorders of fluid, electrolyte, and acid-base balance (phecode 276, US prevalence 7%, BIoVU prevalence 25%), anxiety disorders (phecode 300, US prevalence 7%, BioVU prevalence 19%), and peripheral vascular disease (phecode 443, prevalence 2.4%, BioVU prevalence 5%). For vitamin D, we reweighted to target prevalences for vitamin deficiency (phecode 261, US prevalence 3%, BioVU prevalence 12%) and renal failure (phecode 585, US prevalence 8%, BioVU prevalence 17%). Associations between lead variants and laboratory traits were reestimated under each weighting scheme to assess sensitivity of sex-by-genotype interaction estimates to ascertainment.

### Colocalization with sex stratified eQTL data

We performed colocalization analysis using the R package coloc^61^ and sexstratified eQTL association data from the Genotype-Tissue Expression (GTEx) Consortium^62^. We restricted to variants within 1 MB of the transcription starts site of genes that were reported as having sex-biased eQTLs in the original GTEx paper in breast tissue, the tissue with the most sex biased eQTLs^62^.

## Data and code availability

Individual-level genotype and phenotype data from BioVU and the CCPM is not publicly available due to license, privacy and ethical restrictions on patient medical data, but may be available upon request subject to approval by the Vanderbilt University Medical Center and University of Colorado respectively. Sex-stratified GWAS summary statistics from BioVU will be uploaded to the GWAS catalog upon acceptance of the manuscript. Data to reproduce most figures in the manuscript are available as Supplementary Tables.

