## Supplementary Figures for "Sex differences in the genetic architecture of clinical quantitative traits in the electronic health record"

### Supplemental Figures

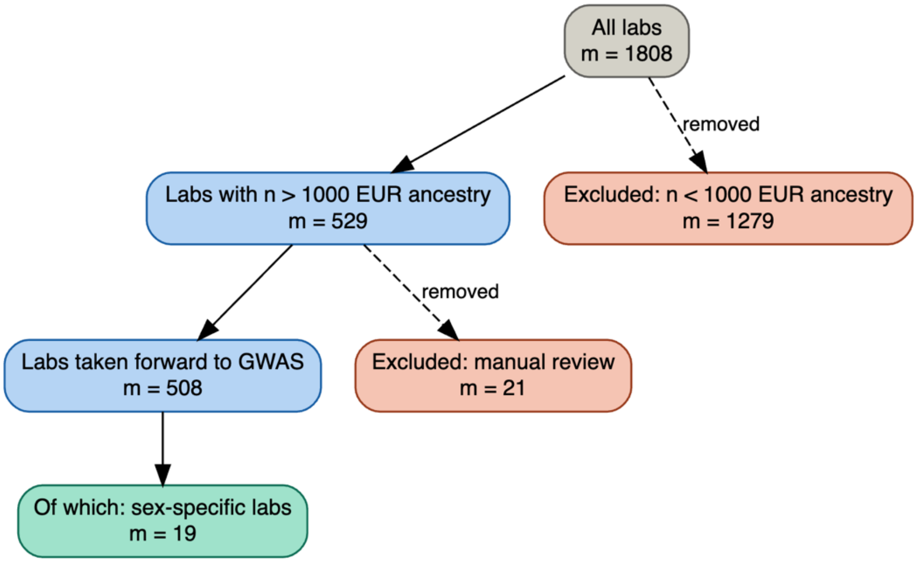


Figure S1 Flow of clinical quantitative traits, i.e. clinical laboratories through the selection for genome-wide association analysis. Manually excluded quantitative traits included those unlikely to be related to antimicrobial resistance or related to clinical lab metadata (e.g. duration).

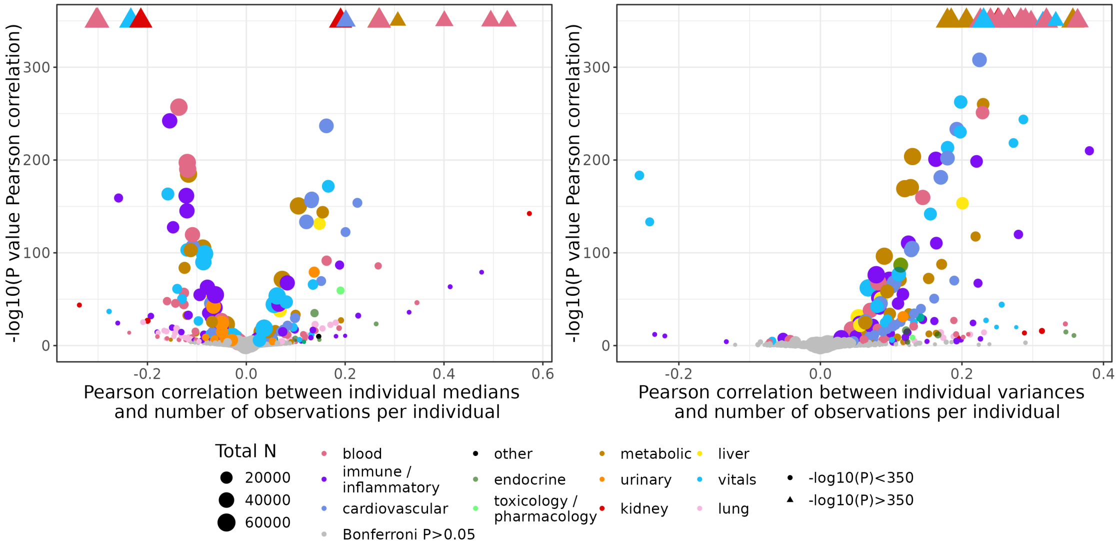


*Figure S2* Correlation between the individual per person medians and the number of measurements per individual (A) or between the per person variances and the number of measurements per individual.

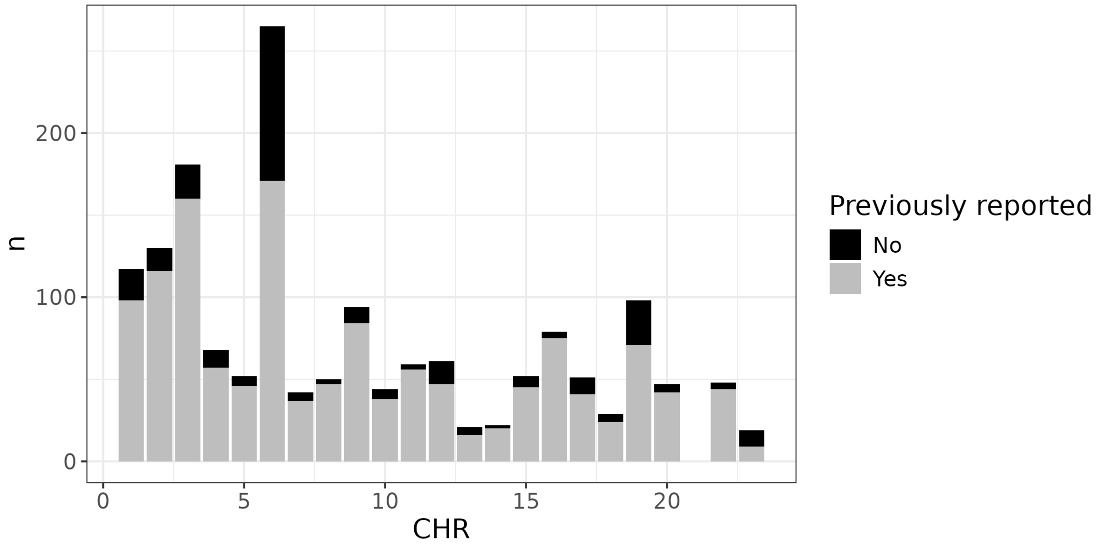


Figure S3 Count of significant loci with P<5e-8 by chromosome, annotated by whether the locus or nearest gene had been previously reported in the GWAS catalog.

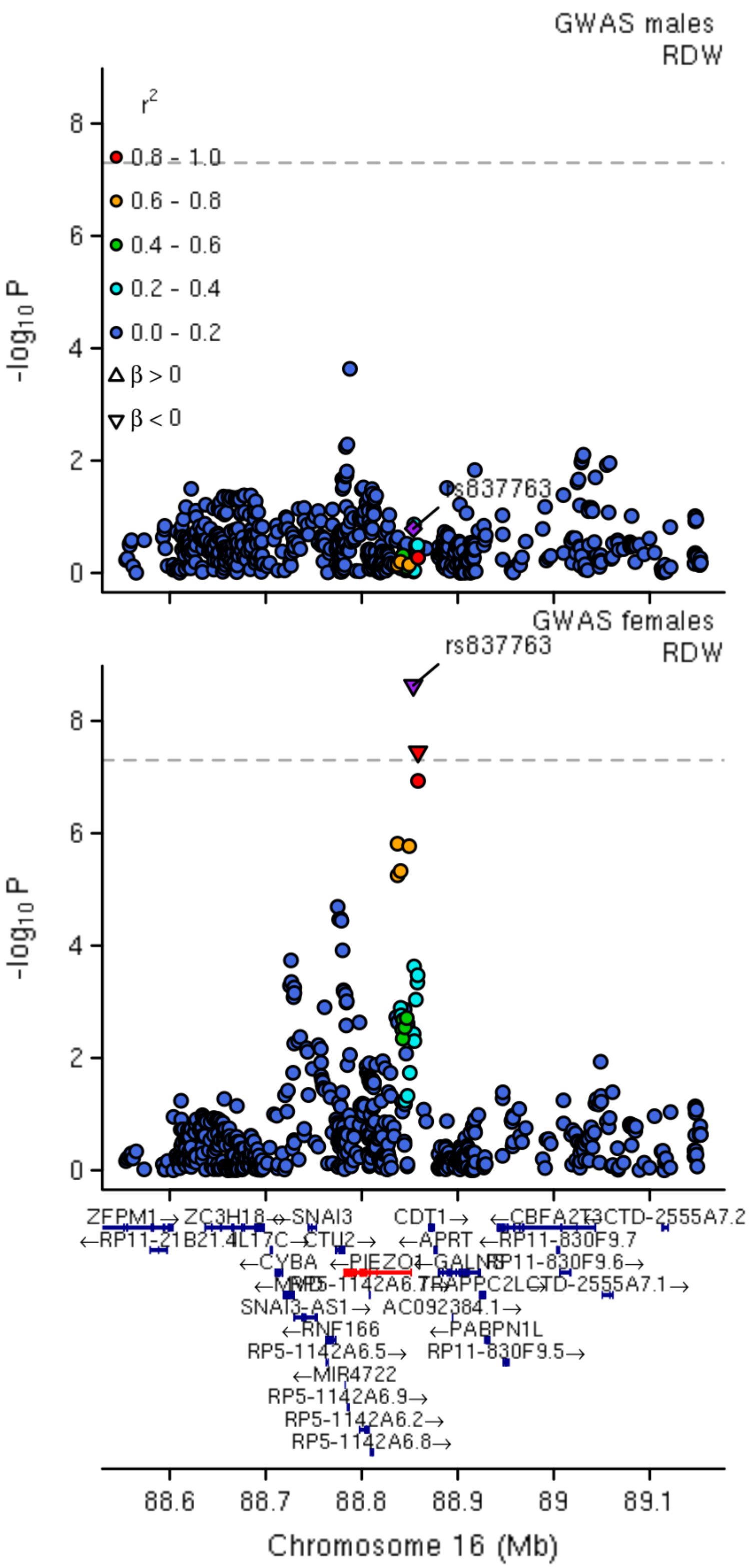


Figure S4 LocusZoom plot for *PIEZO1* and erythrocyte distribution width, the sole replicable sex-specific association across UK Biobank and BioVU.

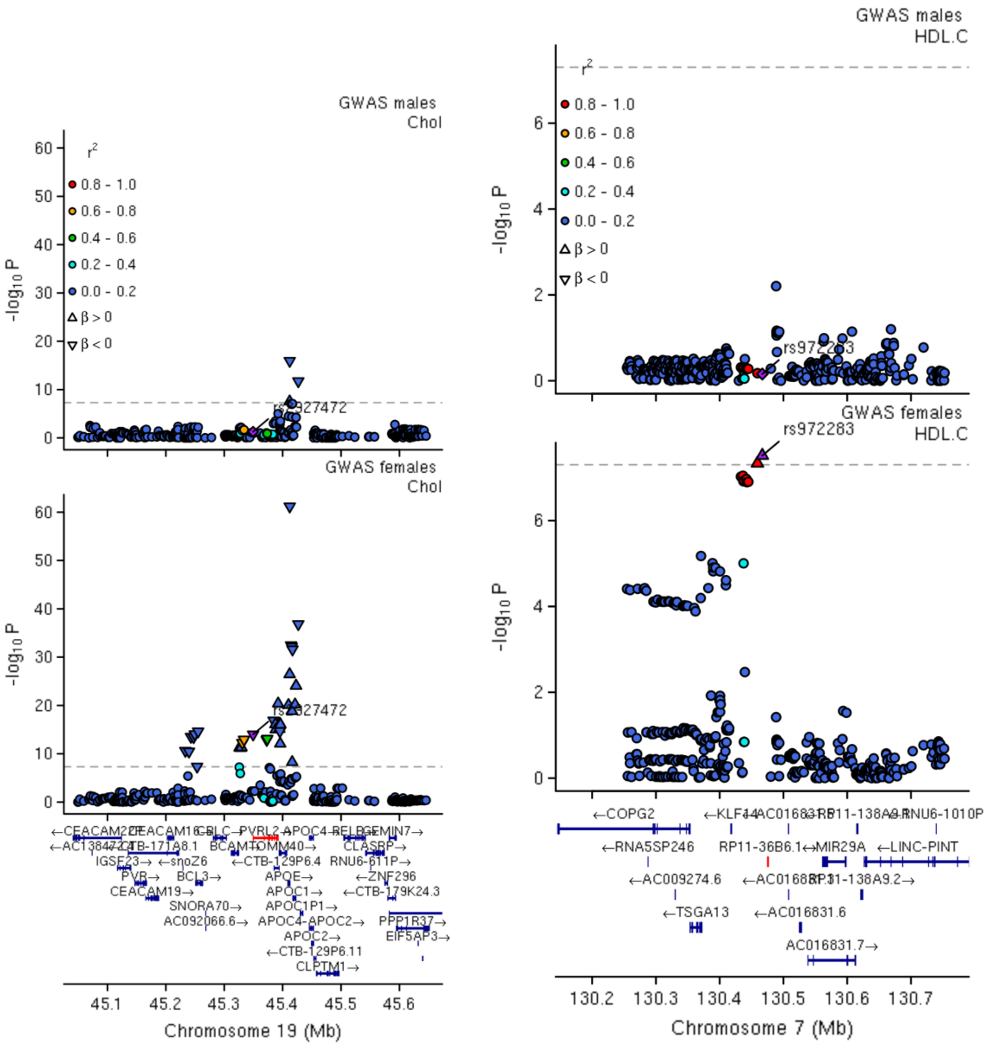


Figure S5 LocusZoom plots for two sex-specific loci in BioVU related to lipid biology.

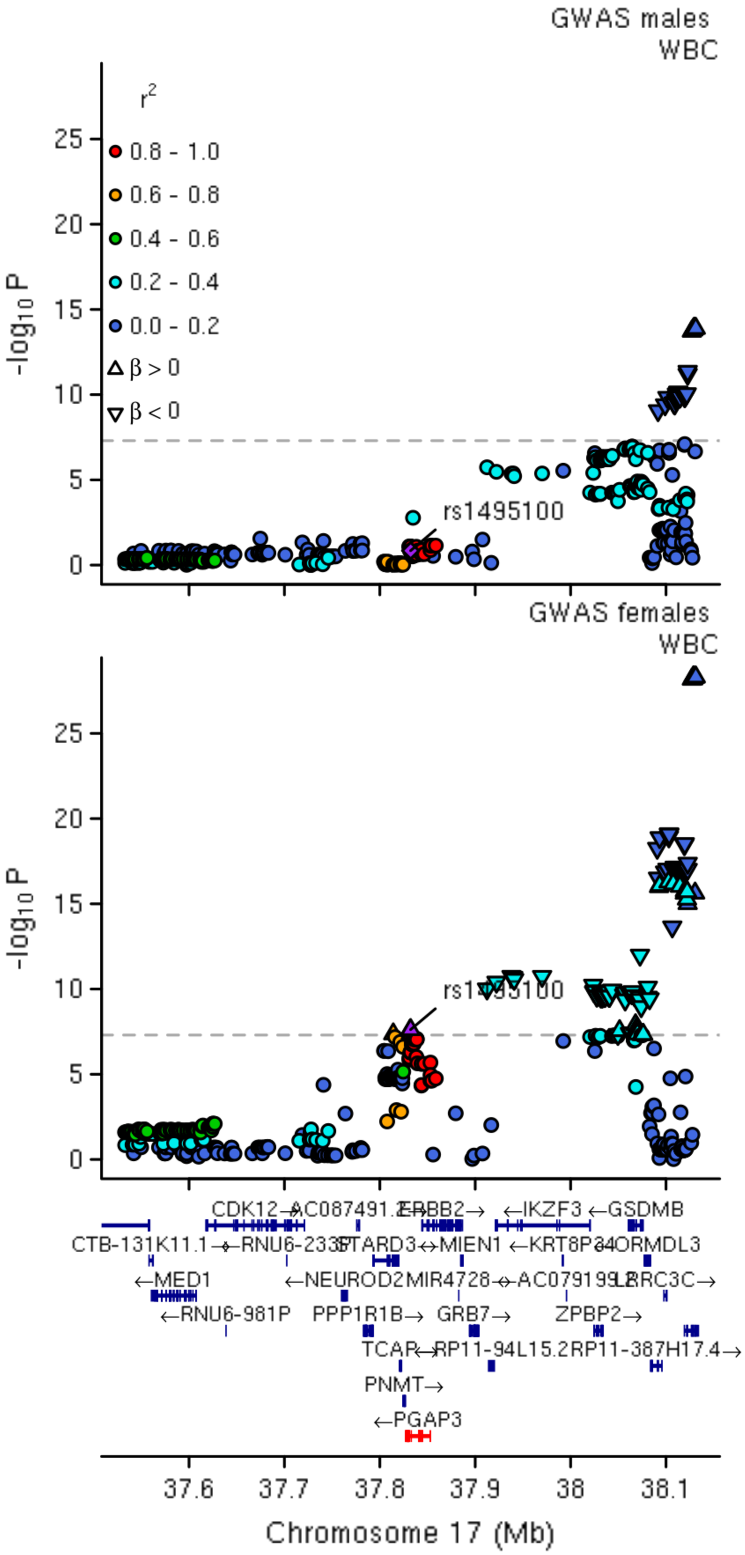


Figure S6 LocusZoom plot for a locus near *PGAP3* and white blood cell count

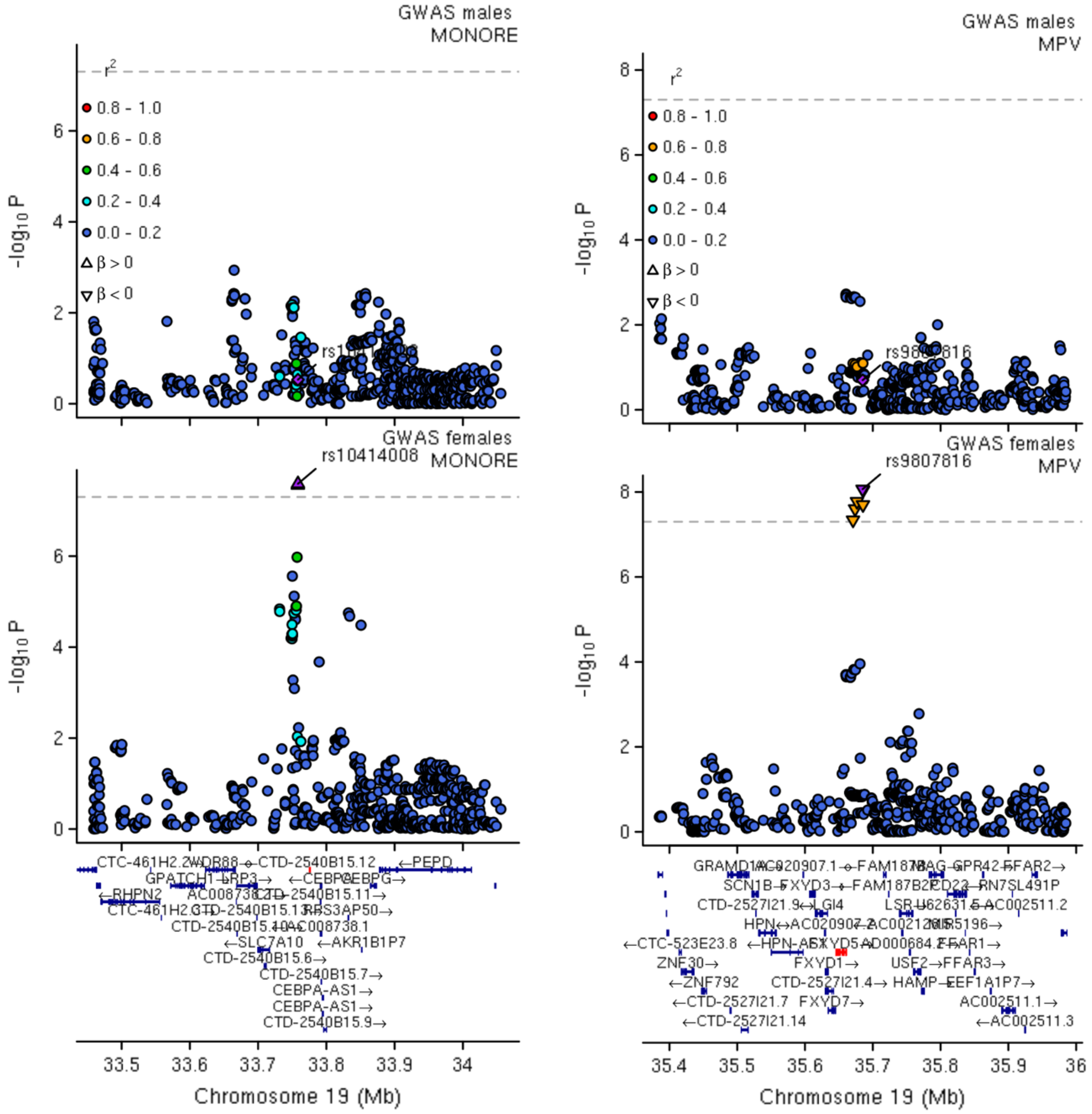


Figure S7 LocusZoom plot for two sex-specific loci on chromosome 19, one associated with monocyte percent and the other with mean platelet volume

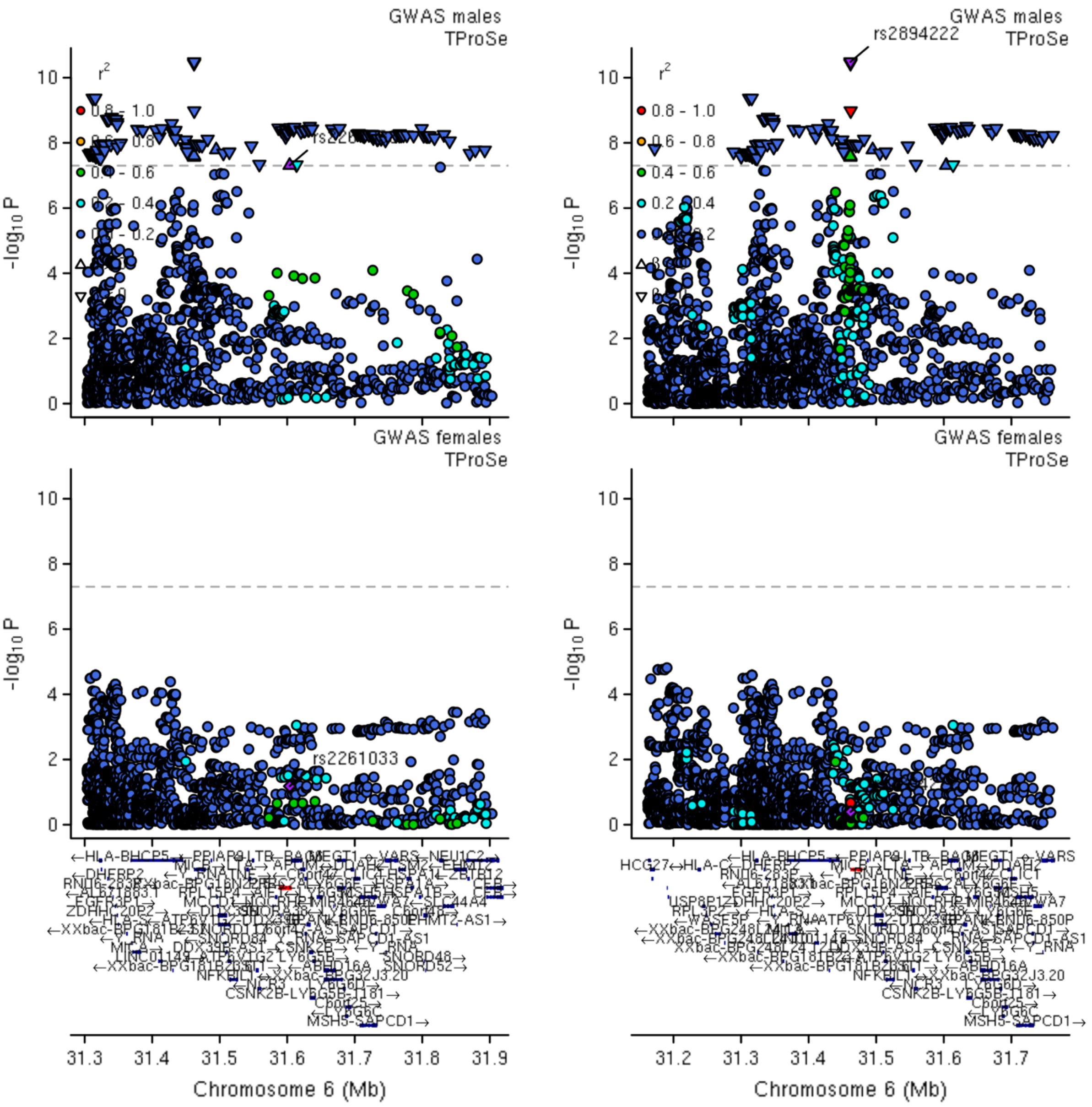


Figure S8 LocusZoom plots for two loci on chromosome 6 with sex specificity for protein in serum measurement.

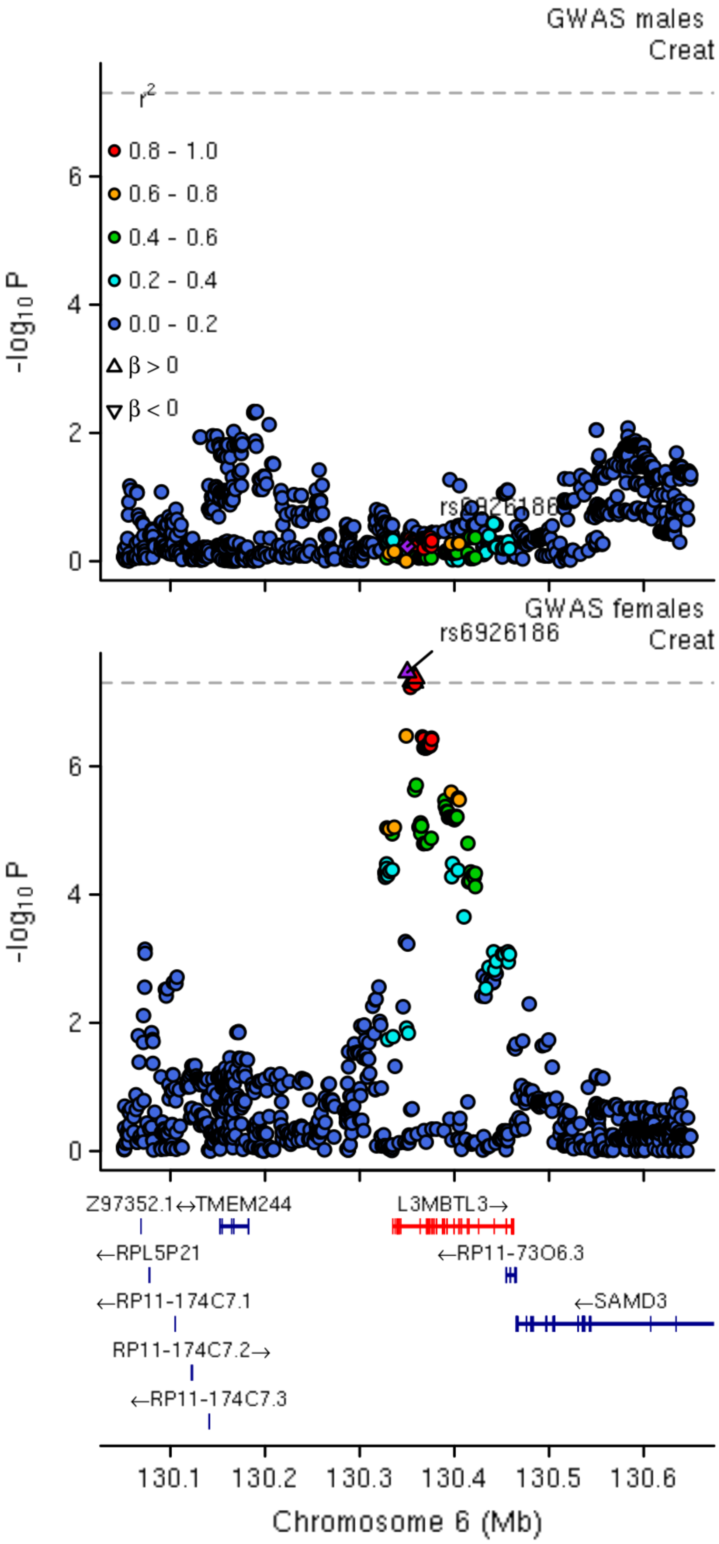


Figure S9 LocusZoom plot for a sex-specific locus on chromosome 6 associated with creatinine levels.

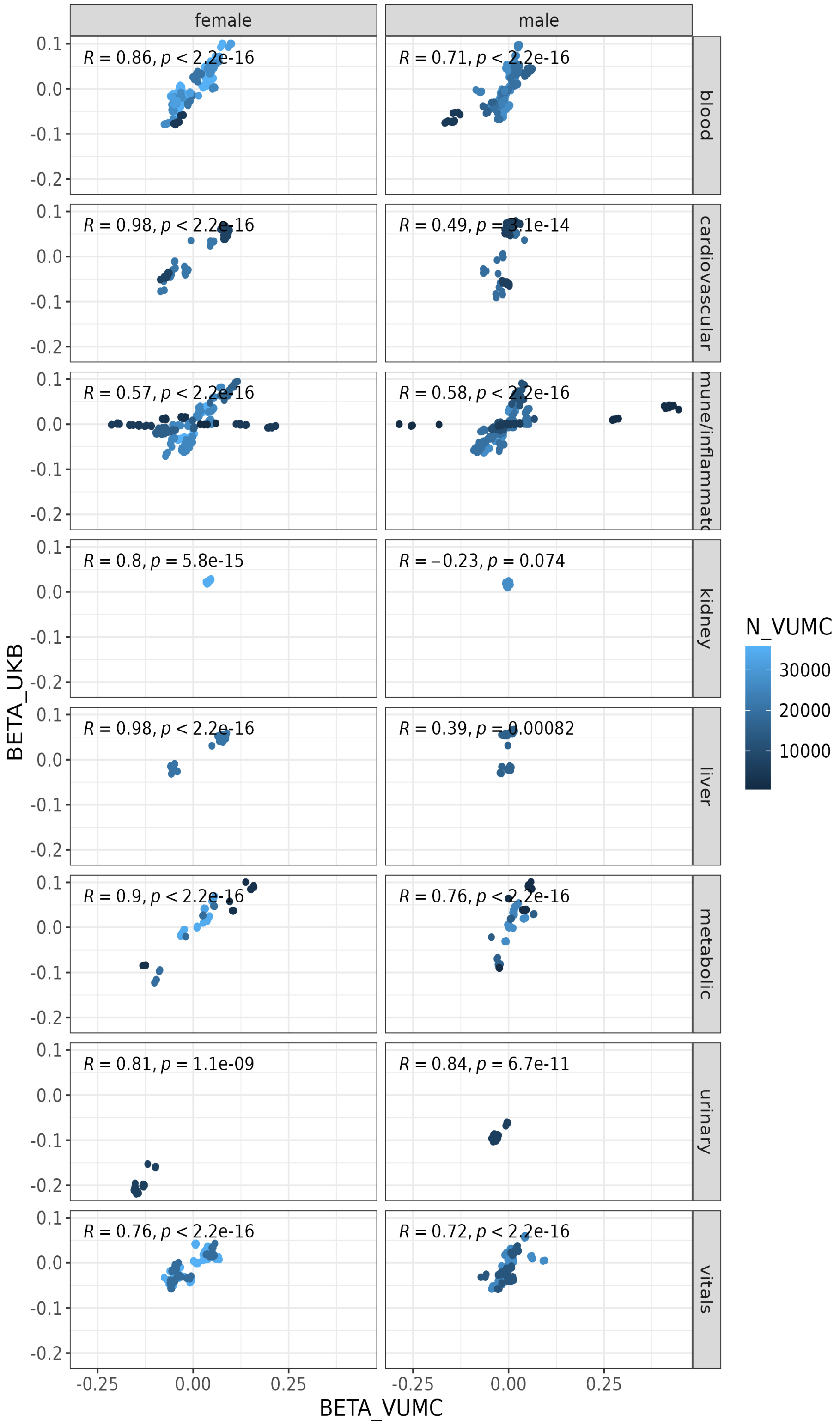


Figure S10 Correlation between beta regression estimates in BioVU (x-axis) and UKBiobank (y-axis) for sex-specific loci identified in BioVU, faceted by sex and clinical quantitative trait category

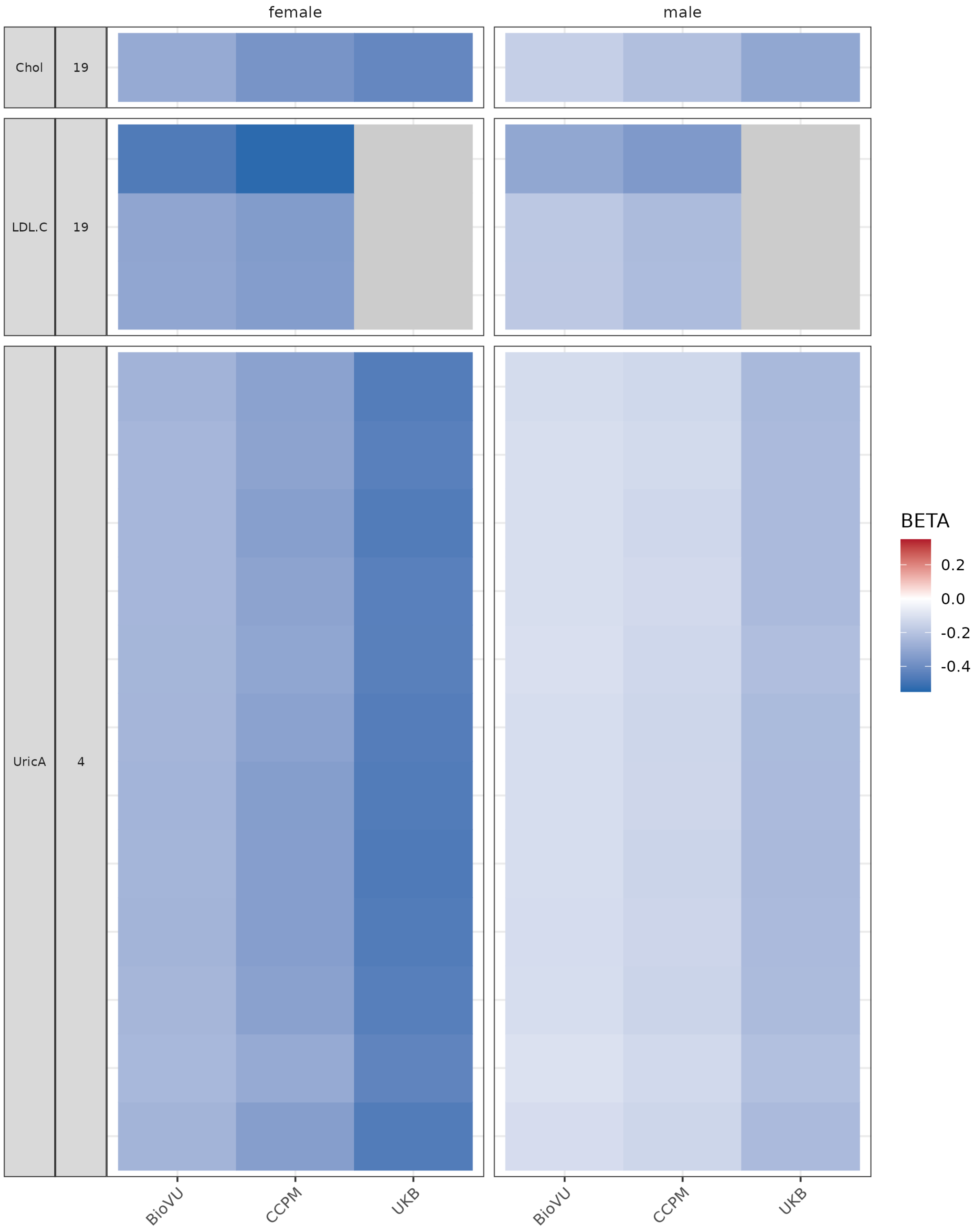


Figure S11 Comparison of beta effect estimates from sex interaction loci across biobanks that displayed replicable sex differences (P<5e-8) in the magnitude of association.

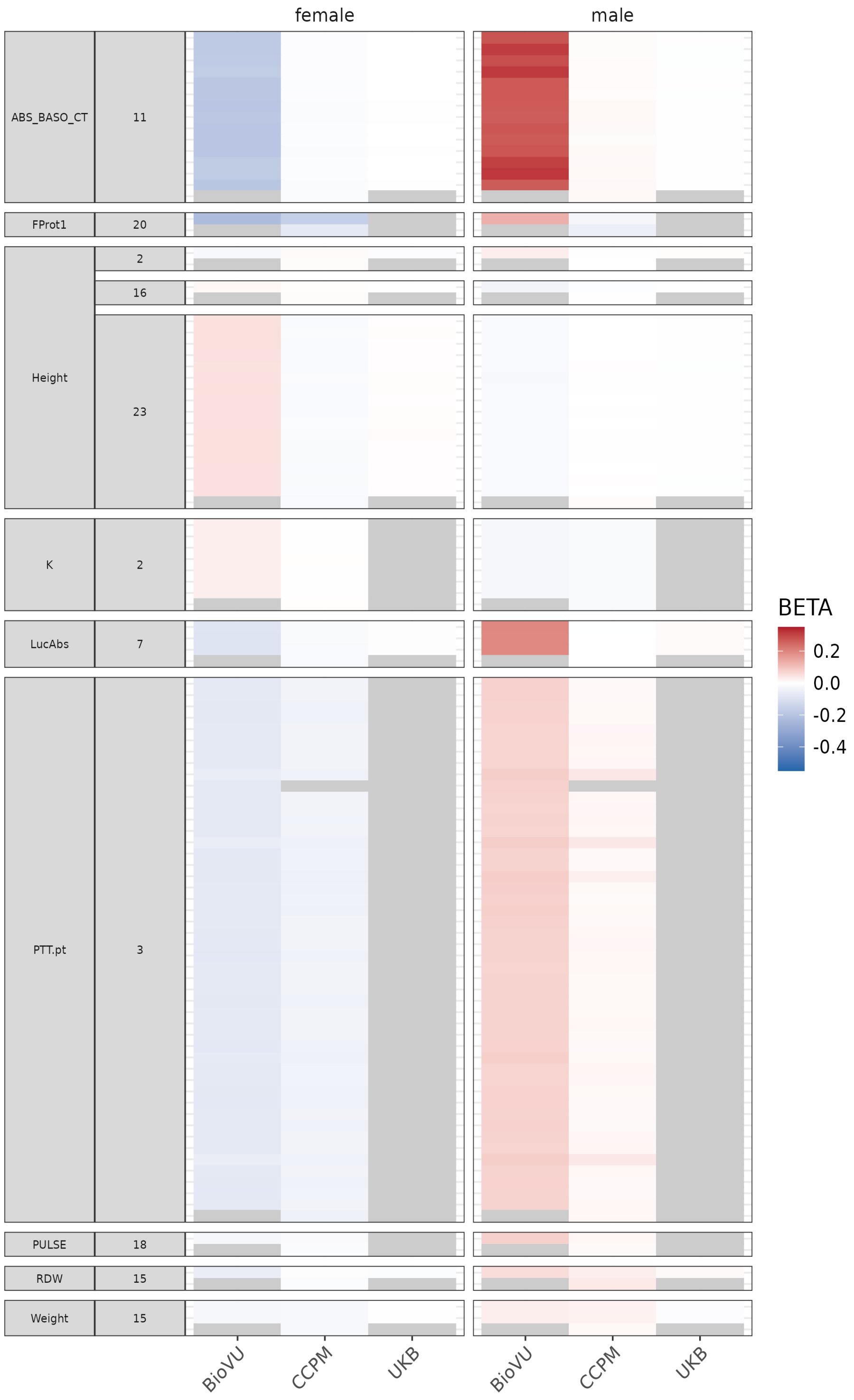


Figure S12 Comparison of beta effect estimates from sex interaction loci across biobanks that displayed nominally replicable (post-hoc t-test p<0.05 and concordant t-statistic sex differences in a majority of variants within the locus) sex differences in the magnitude or direction of association.

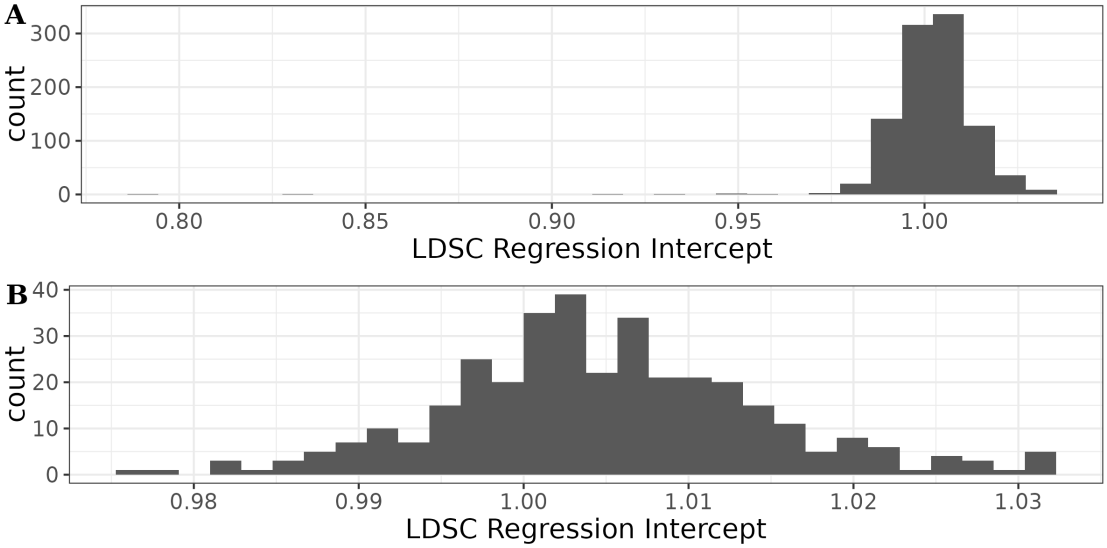


SFigure 13 LDSC regression intercepts from sex-stratified A) all traits or B) traits with sample size greater than 5,000

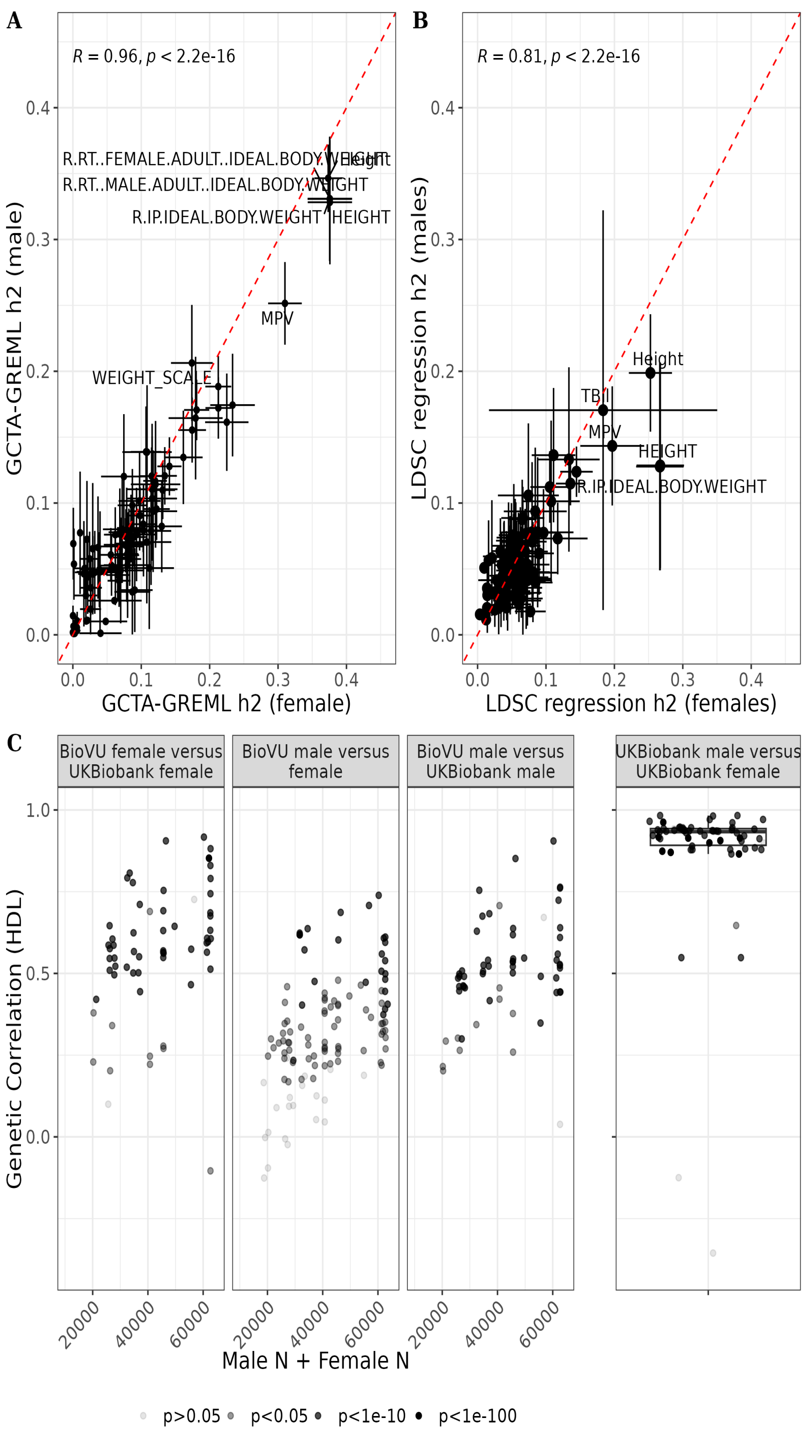


SFigure 14 SNP-based heritability (h2) of traits in females (x-axis) and males (y-axis) using either A) GCTA-GREML or B) LD score regression methods in BioVU. C) Genetic correlation (y-axis) using high-definition likelihood (HDL) method and comparing BioVU to UK Biobank females, BioVU females to males, and BioVU to UK Biobank males against sample size of BioVU traits (x-axis) as well as UKBiobank males to UKBiobank females.

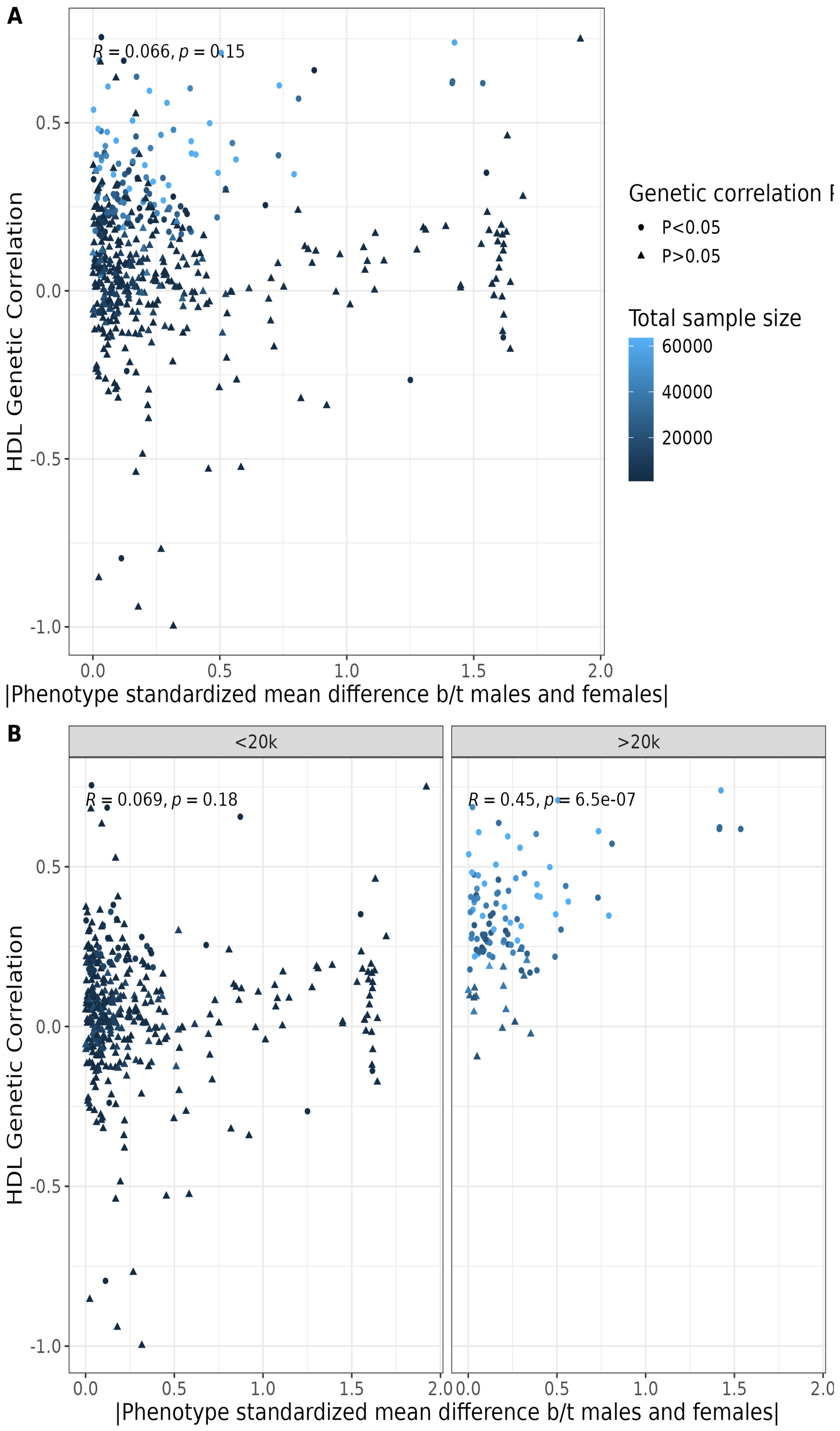


SFigure 15 A) scatterplot of the absolute value of the standardized mean difference between males and females and the high definition likelihood estimated correlation between males and females and B) the same plot but stratified by sample size of trait (greater versus less than 20,000).

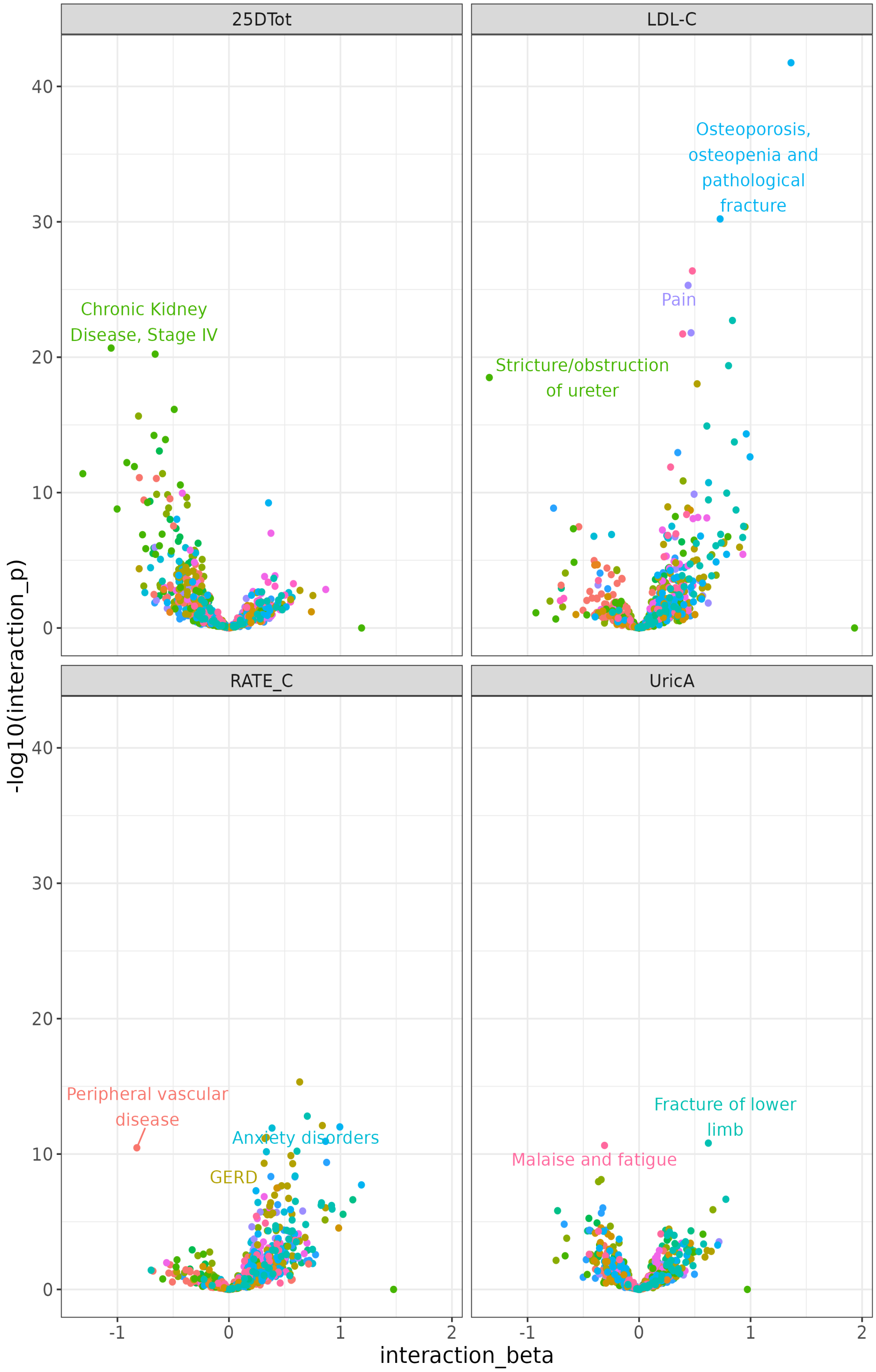


SFigure 16 Volcano plots of sex-by-ascertainment status interaction PheWAS with regression coefficients on the x-axis and -log10(p-value) on the y-axis

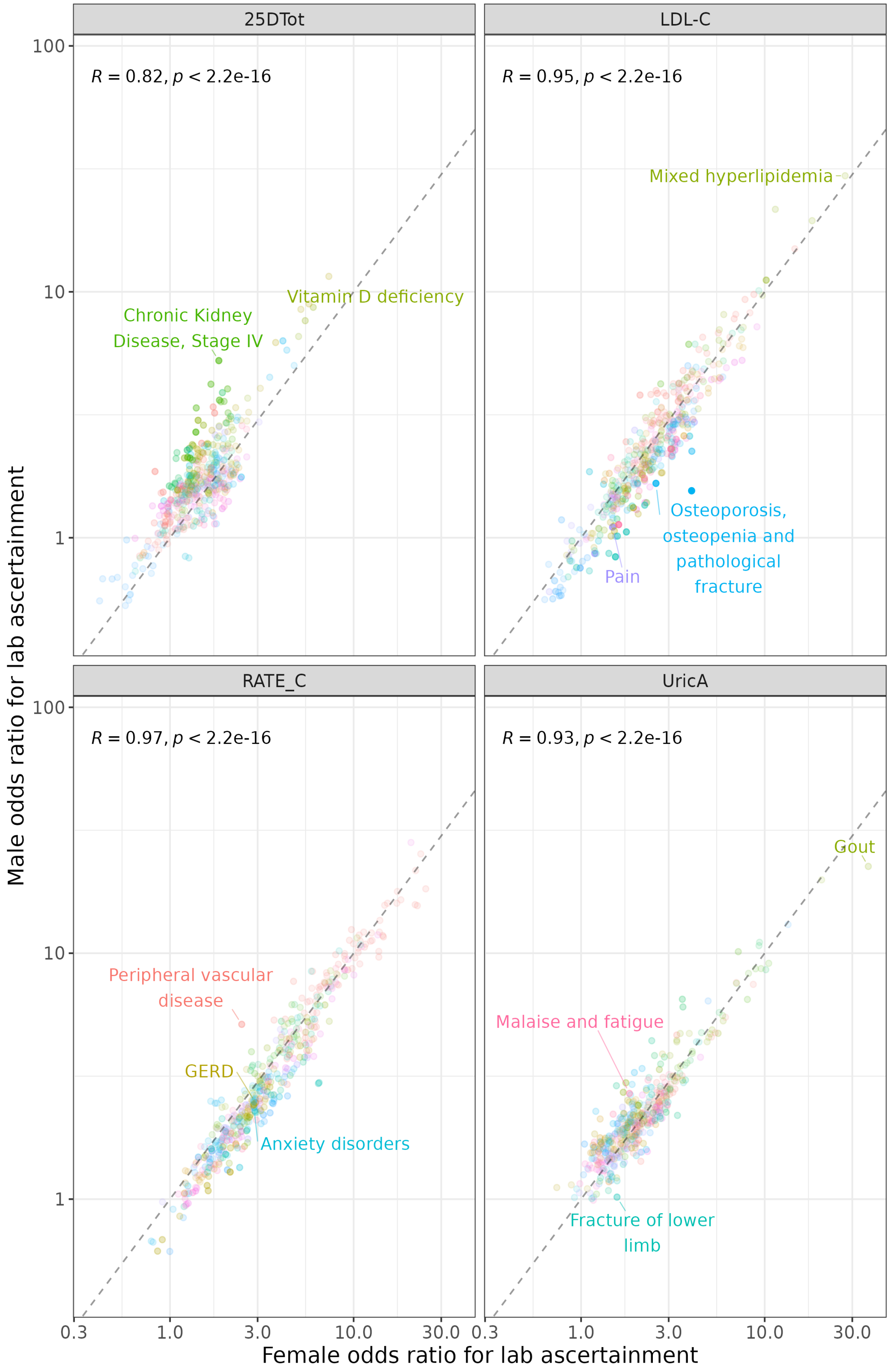


SFigure 17 Scatterplot of sex-stratified ascertainment PheWAS odds ratio for females and males across traits

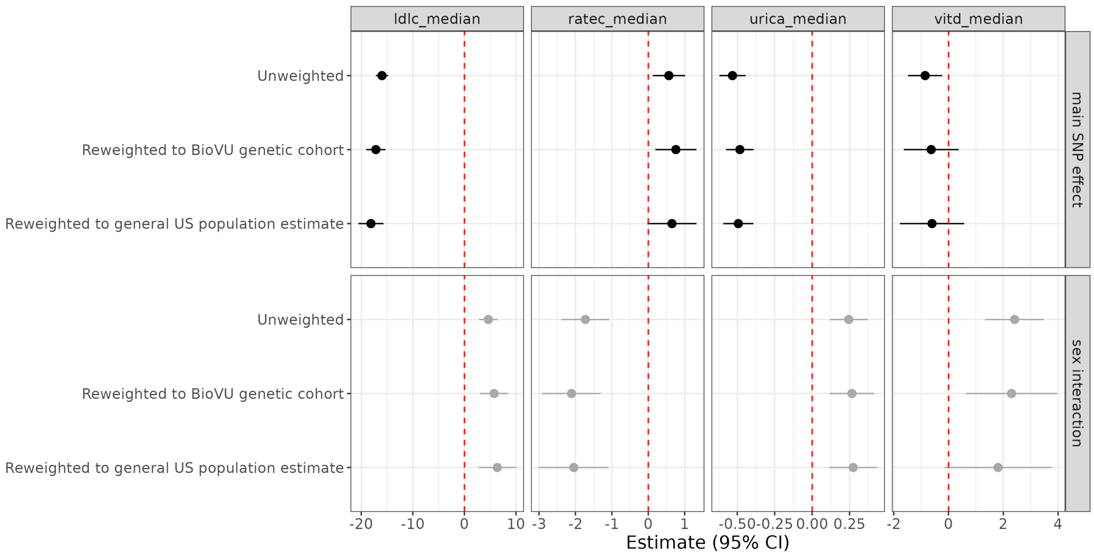


SFigure 18 Regression coefficients for the main variant effect and sex interaction coefficient for each variant on the per-person trait across various weighting schemes correcting for ascertainment phecodes selected based on statistical significance for the association between lab ascertainment and that phecode, and/or statistical significance for the sex interaction. Traits are LDL cholesterol, heart rate from EKG, uric acid, and vitamin D.

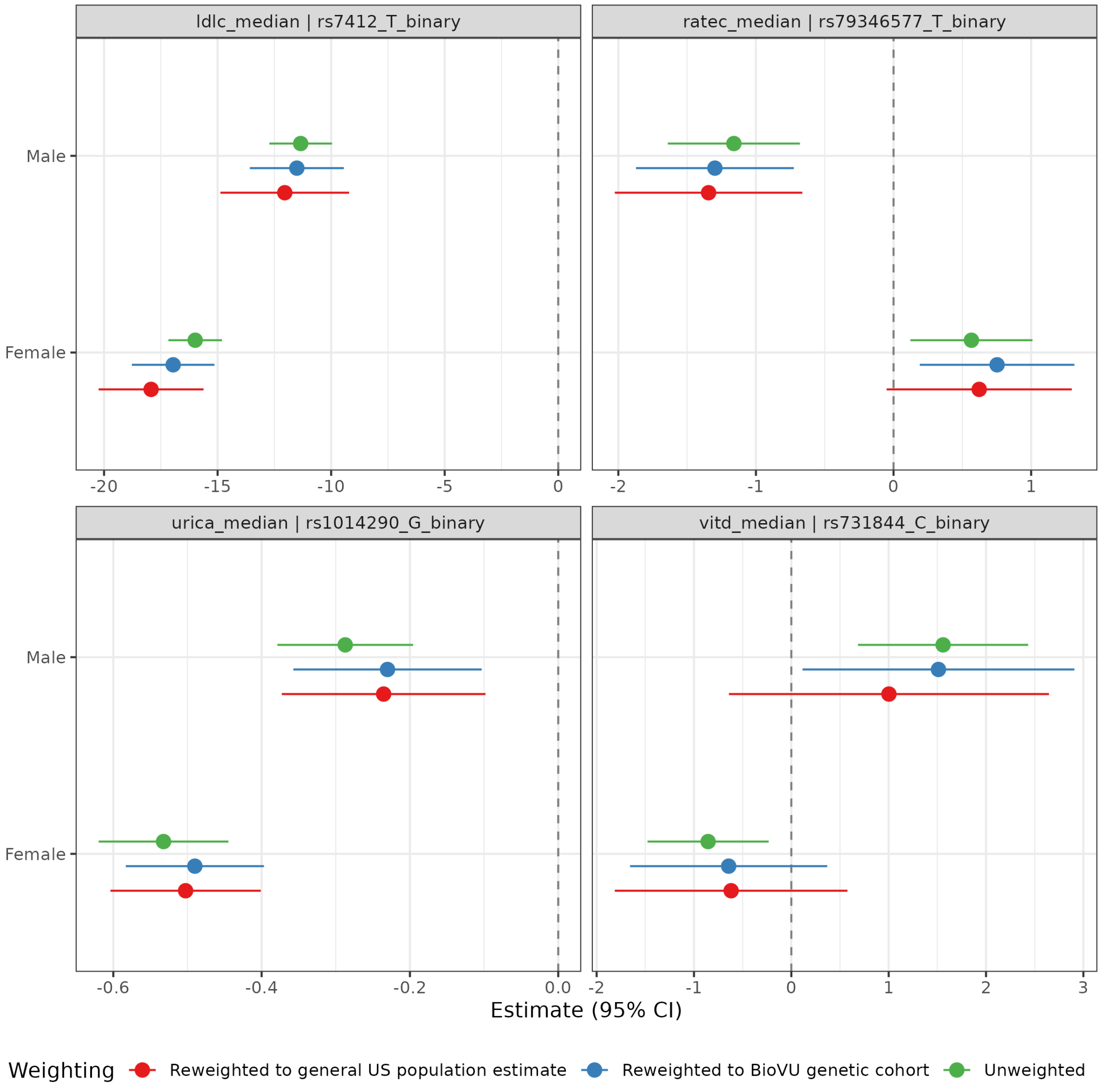


SFigure 19 Regression coefficients from sex-stratified regressions for each variant on the per-person trait across various weighting schemes correcting for ascertainment phecodes selected based on statistical significance for the association between lab ascertainment and that phecode, and/or statistical significance for the sex interaction. Traits are LDL cholesterol, heart rate from EKG, urica acid, and vitamin D.
