## Supplementary Tables for "Sex differences in the genetic architecture of clinical quantitative traits in the electronic health record"

#### Supplementary Tables — Table of Contents

| Name | Description | Notes |
|---|---|---|
| STable1 | Cohort description | |
| STable2 | Description of 508 clinical quantiative traits extracted from BioVU | Columns prefixed with "raw" have summary statistcs within males or females for the per-person median or per-person variance of lab values with no transformation. If not prefixed with raw, the per-person medians and per-person variances were residualized for cubic splines of median age at measurement and inverse normalized within the entire EHR |
| STable3 | Count of variants and independent loci identified as associated with per-person median trait values in males, females, or both sexes and count in EMBL GWAS catalog | significant=P<5e-8, suggestive=P<1e-6, noHLA indicates variants within HLA were excluded |
| STable4 | Unique loci identified as associated with per-person median sclinical quantitative trait values using plink | |
| STable5 | 'Starry" loci - e.g. loci for per-person median trait values with only a single variant - excluded from counts of significant loci | |
| STable6 | Independent loci for per-person median trait values that exist in GWAS catalog that were not previously reported (either any variant within the loci reported or gene previously reported) in GWAS catalog | |
| STable7 | Independent loci for per-person median trait values that did not have data in the GWAS catalog | |
| STable8 | Variants from loci identified as sex specific (P<5e-8 in one sex, all variants within loci with P>0.01 in other sex) in BioVU, with effect estimates for BioVU (VUMC) males and females as well as UKBiobank males and females | |
| STable9 | Unique loci identified as associated with per-person variances of clinical quantitative trait values using plink | |
| STable10 | Sex interaction loci for per-person medians of clinical quantitative traits in BioVU identified from post-hoc t test and replication status | |
| STable11 | Heritability and genetic correlation estimates of per-person median values of clinical quantitative traits in BioVU | |
| STable12 | Regression estimates from regressions of replicable sex difference trait-loci pairs (APOE and LDL cholesterol; uric acid and SLC2A9) with control for comorbidities and medication use | |
| STable13 | Causal mediation estimates from gout, uric acid, and SLC2A9 analysis | |
| STable14 | Causal mediation estimates from ischemic heart disease, LDL cholesterol, and APOE analysis | |
| STable15 | Ascertainment PheWAS of having a lab ever measured for vitamin D, LDL cholesterol, heart rate from EKG, and uric acid | cells with blank regression estimates were not run due to low sample size in phewas |
| STable16 | Sex stratified regression estimate results from reweighting via inverse probability for ascertainment weighting for vitamin D, LDL cholesterol, heart rate from EKG and uric acid | |
| STable17 | SNP and SNP*sex interaction regression estimate results from reweighting via inverse probability for ascertainment weighting for vitamin D, LDL cholesterol, heart rate from EKG and uric acid in sex combined models | |
| STable18 | Colocalization statistics from loci with PP4>0.5 for colocalization between sex-stratified and sex-biased eQTL data and sex-stratifiied GWAS statistics from BioVU | |

#### STable1: Cohort description

10 rows × 4 columns

| STable 1: Clinical characteristics of BioVU participants included in the genetic analyses, restricted to individuals with at least one clinical quantitative measurement at ≥18 years of age. | | | |
|---|---|---|---|
| Characteristic | Female | Male | p-value2 |
| | N = 37,0781 | N = 27,8781 | |
| Birth year | 1959 (1947, 1975) | 1953 (1943, 1966) | <0.001 |
| Record length | 12 (5, 18) | 10 (3, 17) | <0.001 |
| (years, date last ICD code minus date first ICD code) | | | |
| Unique days across medical record | 51 (20, 113) | 50 (19, 110) | 0.088 |
| Number of unique ICD codes | 144 (54, 336) | 161 (58, 380) | <0.001 |
| 1Median (Q1, Q3) | | | |
| 2Wilcoxon rank sum test | | | |
| ICD = International classification of disease | | | |

#### STable2: Description of 508 clinical quantiative traits extracted from BioVU

*Notes: Columns prefixed with "raw" have summary statistcs within males or females for the per-person median or per-person variance of lab values with no transformation. If not prefixed with raw, the per-person medians and per-person variances were residualized for cubic splines of median age at measurement and inverse normalized within the entire EHR*

508 rows × 26 columns

STable2 — Columns 1–23 of 26

| all_labs | full_name | group | selected_unit | male_n | female_n | total_n | sex_specific_lab | raw_median_male_mean | raw_median_female_mean | raw_median_p.value | raw_variance_female_mean | raw_variance_male_mean | raw_variance_p.value | std_mean_diff | median_male_mean | median_female_mean | median_p.value | median_p.value.fdr | std_variance_diff | variance_female_mean | variance_male_mean | variance_p.value |
|---|---|---|---|---|---|---|---|---|---|---|---|---|---|---|---|---|---|---|---|---|---|---|
| #WBC | # of WBC's | blood | | 761 | 925 | 1686 | | 100.94 | 100.62 | 0.46 | 248.08 | 518.22 | 0.1787 | -0.1427 | 0.2039 | 0.06358 | 0.003305 | 0.005038 | -0.09731 | -0.288 | -0.1944 | 0.1878 |
| %SAT | % saturation | other | % | 6545 | 8469 | 15014 | | 24.68 | 21.88 | 5.95e-42 | 106.54 | 117.12 | 0.01009 | -0.2237 | 0.09158 | -0.128 | 9.29e-42 | 3.21e-41 | -0.06705 | 0.09874 | 0.1603 | 0.004563 |
| 125DHy | Calcitriol [Mass/volume] in Serum or Plasma | metabolic | pg/mL | 897 | 2073 | 2970 | | 41.46 | 49.59 | 8.07e-22 | 321.19 | 229.66 | 0.1488 | 0.4143 | -0.4034 | -0.007011 | 2.43e-24 | 7.10e-24 | 0.03922 | 0.01702 | -0.02051 | 0.6125 |
| 25DTot | 25-Hydroxyvitamin D3+25-Hydroxyvitamin D2 [Mass/volume] in Serum or Plasma | metabolic | ng/mL | 2828 | 7048 | 9876 | | 32.44 | 35.78 | 2.78e-37 | 106.75 | 106.20 | 0.9394 | 0.2804 | -0.09755 | 0.1725 | 5.02e-38 | 1.70e-37 | 0.04262 | 0.02275 | -0.01877 | 0.2318 |
| 25HyD2 | 25-Hydroxyvitamin D2 [Mass/volume] in Serum or Plasma | metabolic | ng/mL | 811 | 2276 | 3087 | | 17.34 | 18.98 | 0.005969 | 190.58 | 160.63 | 0.1488 | 0.0816 | -0.03296 | 0.04922 | 0.04052 | 0.05385 | 0.08415 | 0.04513 | -0.04302 | 0.2043 |
| 25HyD3 | 25-Hydroxyvitamin D3 [Mass/volume] in Serum or Plasma | metabolic | ng/mL | 2839 | 7065 | 9904 | | 28.66 | 31.02 | 1.88e-17 | 66.21 | 64.74 | 0.7183 | 0.1707 | -0.04683 | 0.1201 | 5.13e-16 | 1.27e-15 | 0.03112 | 0.03746 | 0.006715 | 0.3719 |
| A-1Gg_ | Alpha 1 globulin [Mass/volume] in Serum or Plasma by Electrophoresis | liver | g/dL | 2884 | 3674 | 6558 | | 0.259 | 0.261 | 0.2772 | 0.003283 | 0.003486 | 0.5134 | 0.1004 | -0.08861 | 0.01263 | 6.36e-05 | 1.11e-04 | -0.03369 | -0.02016 | 0.0137 | 0.5174 |
| A-2Gg_ | Alpha 2 globulin [Mass/volume] in Serum or Plasma by Electrophoresis | liver | g/dL | 2892 | 3671 | 6563 | | 0.9594 | 1.025 | 4.08e-44 | 0.02151 | 0.0216 | 0.9595 | 0.3708 | -0.1405 | 0.2185 | 1.01e-50 | 3.94e-50 | -0.05273 | -0.017 | 0.03791 | 0.3099 |
| A1Anti | Alpha 1 antitrypsin [Mass/volume] in Serum or Plasma | other | mg/dL | 962 | 925 | 1887 | | 147.88 | 153.48 | 0.001063 | 550.12 | 572.16 | 0.9291 | 0.1561 | -0.07142 | 0.08244 | 6.85e-04 | 0.001098 | 0.09507 | 0.2438 | 0.1519 | 0.652 |
| ABS_BASO_CT | Basophils [#/volume] in Blood by Automated count | blood | 10*3/uL | 638 | 1059 | 1697 | | 0.04158 | 0.0407 | 0.4413 | 2.41e-04 | 3.13e-04 | 0.06381 | -0.03335 | -0.04296 | -0.07793 | 0.5043 | 0.5545 | -0.03811 | 0.1219 | 0.1611 | 0.6249 |
| ABS_EOS_CT | Eosinophils [#/volume] in Blood by Automated count | blood | 10*3/uL | 598 | 996 | 1594 | | 0.1683 | 0.1487 | 0.009692 | 0.009393 | 0.01276 | 0.07857 | -0.0904 | -0.006553 | -0.105 | 0.08618 | 0.111 | -0.05727 | 0.05437 | 0.1145 | 0.4726 |
| ABS_IMMATURE_GRANULOCYTES_CT | Immature granulocytes [#/volume] in Blood by Automated count | blood | 10*3/uL | 626 | 1042 | 1668 | | 0.04859 | 0.04052 | 0.005448 | 0.003136 | 0.002782 | 0.6728 | -0.1709 | 0.2264 | 0.04533 | 7.93e-04 | 0.001263 | 0.06887 | 0.03848 | -0.03695 | 0.3709 |
| ABS_LYMPH_CT | Lymphocytes [#/volume] in Blood by Automated count | blood | 10*3/uL | 640 | 1060 | 1700 | | 1.538 | 1.736 | 8.64e-08 | 0.2454 | 0.1976 | 0.1449 | 0.2194 | -0.3001 | -0.07235 | 9.69e-06 | 1.75e-05 | 0.07048 | 0.1486 | 0.08538 | 0.3366 |

| all_labs | full_name | group | selected_unit | male_n | female_n | total_n | sex_specific_lab | raw_median_male_mean | raw_median_female_mean | raw_median_p.value | raw_variance_female_mean | raw_variance_male_mean | raw_variance_p.value | std_mean_diff | median_male_mean | median_female_mean | median_p.value | median_p.value.fdr | std_variance_diff | variance_female_mean | variance_male_mean | variance_p.value |
|---|---|---|---|---|---|---|---|---|---|---|---|---|---|---|---|---|---|---|---|---|---|---|
| ABS_MONO_CT | Monocytes [#/volume] in Blood by Automated count | blood | 10*3/uL | 639 | 1059 | 1698 | | 0.7292 | 0.6387 | 9.94e-10 | 0.05067 | 0.05886 | 0.2576 | -0.3095 | 0.3 | -0.03153 | 8.00e-10 | 1.72e-09 | -0.0296 | 0.1085 | 0.1392 | 0.7015 |
| ABS_NEUTRO_CT | Neutrophils [#/volume] in Blood by Automated count | blood | 10*3/uL | 638 | 1060 | 1698 | | 6.125 | 5.868 | 0.1004 | 6.033 | 5.6 | 0.5313 | -0.1001 | 0.2278 | 0.1259 | 0.0449 | 0.05935 | 0.02996 | 0.1458 | 0.1167 | 0.6917 |
| ACAIgA | Cardiolipin IgA Ab [Units/volume] in Serum by Immunoassay | immune/inflammatory | | 548 | 1508 | 2056 | | 5.149 | 4.536 | 0.002849 | 7.038 | 6.846 | 0.9458 | -0.05461 | -0.09501 | -0.1506 | 0.2903 | 0.3402 | 0.02311 | 0.1107 | 0.08774 | 0.8589 |
| ACAIgG | Cardiolipin IgG Ab [Units/volume] in Serum by Immunoassay | immune/inflammatory | | 675 | 1806 | 2481 | | 7.291 | 6.802 | 0.1075 | 20.38 | 7.289 | 1.29e-04 | -0.05763 | 0.04242 | -0.01561 | 0.2092 | 0.2517 | 0.01809 | 0.4424 | 0.426 | 0.8297 |
| ACAIgM | Cardiolipin IgM Ab [Units/volume] in Serum by Immunoassay | immune/inflammatory | [MPL'U] | 675 | 1805 | 2480 | | 5.408 | 5.302 | 0.589 | 8.941 | 4.839 | 0.009977 | 0.004272 | 0.04204 | 0.0464 | 0.9246 | 0.9378 | -0.006532 | 0.4594 | 0.4655 | 0.9382 |
| ACTH | Corticotropin [Mass/volume] in Plasma | endocrine | pg/mL | 598 | 987 | 1585 | | 26.01 | 25.03 | 0.8293 | 31870.7 | 50091.8 | 0.5579 | -0.3528 | 0.208 | -0.123 | 2.01e-12 | 4.60e-12 | -0.2556 | 0.01923 | 0.2589 | 0.004848 |
| AFP-Ob | Maternal Alpha-1-Fetoprotein [Mass/volume] in Serum or Plasma | ob/gyn | ng/mL | 2 | 1035 | 1037 | Sex specific lab | 4.95 | 46.34 | 0.0209 | | | | 3.019 | -3.006 | -0.001857 | 0.006747 | 0.009719 | | | | |
| AFPTum | Alpha-1-fetoprotein.tumor marker [Mass/volume] in Serum or Plasma | cancer | ng/mL | 1721 | 1312 | 3033 | | 151.96 | 115.06 | 0.5921 | 256167.2 | 2266237.6 | 0.02299 | -0.133 | -0.02064 | -0.1452 | 2.89e-04 | 4.81e-04 | -0.2307 | -0.1403 | 0.08644 | 1.52e-06 |
| Alb | Albumin [Mass/volume] in Serum or Plasma | liver | g/dL | 24351 | 31325 | 55676 | | 4.017 | 4.049 | 1.77e-15 | 0.1182 | 0.1368 | 4.88e-27 | -0.03383 | 0.03498 | 0.005133 | 8.95e-05 | 1.54e-04 | -0.07649 | 0.1891 | 0.2564 | 5.78e-16 |
| AlbCnc | Albumin [Presence] in Urine | urinary | ug/mL | 3332 | 3319 | 6651 | | 88.35 | 52.66 | 3.51e-15 | 17869.0 | 28983.9 | 2.21e-04 | -0.249 | 0.04429 | -0.2236 | 2.08e-24 | 6.15e-24 | -0.1838 | 0.01811 | 0.2016 | 1.62e-09 |
| AlbCre | Microalbumin/Creatinine [Mass Ratio] in 24 hour Urine | liver | ug/mg | 2218 | 2156 | 4374 | | 110.37 | 73.01 | 7.88e-07 | 22829.1 | 33736.7 | 0.03276 | -0.08303 | 0.03675 | -0.05152 | 0.005911 | 0.008589 | -0.1508 | 0.08433 | 0.2317 | 1.18e-04 |
| Album | Albumin [Mass/volume] in Serum or Plasma | liver | g/dL | 3191 | 4050 | 7241 | | 3.568 | 3.576 | 0.4963 | 0.1138 | 0.1207 | 0.4957 | -0.03921 | 0.06515 | 0.02773 | 0.1014 | 0.128 | -0.04818 | -0.01389 | 0.03759 | 0.2981 |
| Aldo-P | Aldosterone [Mass/volume] in Serum or Plasma | endocrine | ng/dL | 573 | 864 | 1437 | | 16.6 | 13.91 | 0.08861 | 123466.0 | 187058.9 | 0.6623 | -0.07699 | 0.01653 | -0.05959 | 0.1495 | 0.1845 | -0.006105 | -0.1177 | -0.1122 | 0.9567 |
| AlkP | Alkaline phosphatase [Enzymatic activity/volume] in Serum or Plasma | liver | [U]/L | 15939 | 21116 | 37055 | | 88.36 | 87.04 | 0.004879 | 885.45 | 1190.91 | 5.22e-10 | 0.03223 | 0.01174 | 0.04447 | 0.002268 | 0.003479 | 0.04694 | 0.05724 | 0.008015 | 5.35e-05 |
| Amm | Ammonia [Moles/volume] in Plasma | metabolic | umol/L | 1791 | 1426 | 3217 | | 48.17 | 44.85 | 0.00302 | 891.08 | 1043.69 | 0.1915 | -0.137 | 0.1011 | -0.03371 | 1.19e-04 | 2.03e-04 | -0.101 | 0.04138 | 0.1424 | 0.09532 |

| all_labs | full_name | group | selected_unit | male_n | female_n | total_n | sex_specific_lab | raw_median_male_mean | raw_median_female_mean | raw_median_p.value | raw_variance_female_mean | raw_variance_male_mean | raw_variance_p.value | std_mean_diff | median_male_mean | median_female_mean | median_p.value | median_p.value.fdr | std_variance_diff | variance_female_mean | variance_male_mean | variance_p.value |
|---|---|---|---|---|---|---|---|---|---|---|---|---|---|---|---|---|---|---|---|---|---|---|
| Amyl | Amylase [Enzymatic activity/volume] in Serum or Plasma | metabolic | [iU]/L | 3776 | 4794 | 8570 | | 45.91 | 42.98 | 0.004874 | 2407.09 | 2764.87 | 0.4346 | -0.02467 | -0.2894 | -0.3142 | 0.2599 | 0.3091 | -0.06032 | -0.0833 | -0.0237 | 0.06537 |
| AN-GAP | Anion gap in Serum or Plasma | metabolic | | 26892 | 34129 | 61021 | | 8.96 | 9.115 | 1.19e-20 | 6.039 | 5.967 | 0.07648 | 0.05591 | -0.2345 | -0.1837 | 7.04e-12 | 1.59e-11 | -0.0123 | 0.2089 | 0.2189 | 0.1552 |
| ANESTHESIA PULSE | Heart rate by pulse oximetry | vitals | | 2556 | 3746 | 6302 | | 71.69 | 75.25 | 1.65e-31 | 51.97 | 44.32 | 2.33e-06 | 0.2726 | -0.1612 | 0.09973 | 6.33e-26 | 1.91e-25 | 0.1401 | -0.01435 | -0.1455 | 4.10e-08 |
| ANION_GAP | Anion gap | metabolic | 10*-3.eq/L | 471 | 619 | 1090 | | 11.09 | 10.83 | 0.1205 | 6.701 | 5.499 | 0.04183 | -0.1356 | 0.04778 | -0.08067 | 0.02538 | 0.03466 | 0.1589 | 0.1143 | -0.03096 | 0.05883 |
| AntDNA | DNA double strand Ab [Units/volume] in Serum | immune/inflammatory | [iU]/mL | 642 | 2079 | 2721 | | 11.88 | 13.2 | 0.1341 | 334.90 | 386.40 | 0.6056 | 0.04323 | -0.07155 | -0.02499 | 0.3398 | 0.39 | -0.05559 | 0.1634 | 0.2177 | 0.6118 |
| APBone | Alkaline phosphatase bone | other | ug/L | 285 | 1045 | 1330 | | 16.63 | 16.2 | 0.5888 | 36.24 | 98.23 | 0.2215 | 0.08953 | -0.01035 | 0.07143 | 0.1823 | 0.221 | -0.04656 | 0.05597 | 0.09939 | 0.7415 |
| AT3 | Antithrombin actual/normal in Platelet poor plasma by Chromogenic method | blood | % | 409 | 625 | 1034 | | 96.58 | 104.34 | 2.60e-08 | 261.69 | 295.05 | 0.6944 | 0.3083 | -0.1613 | 0.1588 | 9.05e-07 | 1.72e-06 | -0.3433 | 0.1138 | 0.5047 | 0.03725 |
| AThyP | Thyroperoxidase Ab | immune/inflammatory | [U]/mL | 231 | 1015 | 1246 | | 54.67 | 46.46 | 0.4558 | 6991.55 | 12352.7 | 0.5589 | -0.2377 | 0.3754 | 0.1442 | 5.62e-04 | 9.11e-04 | -0.834 | 0.063 | 0.942 | 1.40e-05 |
| ATRIAL RATE | P wave Atrium by EKG | cardiovascular | | 6314 | 7227 | 13541 | | 74.35 | 77.7 | 5.78e-19 | 491.45 | 681.24 | 5.72e-14 | 0.1559 | -0.124 | 0.02603 | 2.41e-19 | 6.48e-19 | -0.1131 | -0.157 | -0.0366 | 1.30e-07 |
| AtyLym | Variant lymphocytes/100 leukocytes in Blood by Manual count | immune/inflammatory | % | 4046 | 3888 | 7934 | | 2.6 | 2.471 | 0.03534 | 4.986 | 5.57 | 0.1999 | -0.0891 | 0.003818 | -0.07731 | 7.27e-05 | 1.26e-04 | -0.0777 | -0.0835 | -0.009908 | 0.01887 |
| AutoNt | AutoNt | blood | % | 933 | 1354 | 2287 | | 66.06 | 66.22 | 0.7332 | 73.07 | 76.71 | 0.5368 | 0.00698 | -0.1313 | -0.1249 | 0.8693 | 0.8927 | -0.09339 | -0.1454 | -0.05238 | 0.1189 |
| BAbs | Basophils [#/volume] in Blood by Automated count | immune/inflammatory | 10*3/uL | 10793 | 14842 | 25635 | | 0.03697 | 0.03659 | 0.3017 | 6.88e-04 | 8.88e-04 | 0.01109 | 0.03435 | -0.07295 | -0.04009 | 0.007113 | 0.01019 | -0.003061 | -0.05595 | -0.05297 | 0.8476 |
| Baso | Basophils [#/volume] in Blood | immune/inflammatory | % | 11083 | 15240 | 26323 | | 0.5307 | 0.5277 | 0.5124 | 0.1217 | 0.1409 | 0.01025 | 0.04548 | -0.1018 | -0.06013 | 3.07e-04 | 5.09e-04 | -0.01137 | 0.03185 | 0.04237 | 0.4683 |
| BASOA | Basophils [#/volume] in Blood | blood | 10*3/uL | 4078 | 3882 | 7960 | | 0.08449 | 0.08355 | 0.7986 | 0.02799 | 0.02556 | 0.6121 | -0.006423 | -0.1206 | -0.1271 | 0.7746 | 0.8122 | -0.02356 | 0.06015 | 0.08261 | 0.4496 |
| BASOAB | Basophils [#/volume] in Blood by Automated count | blood | 10*3/uL | 19957 | 25623 | 45580 | | 0.03555 | 0.0354 | 0.4663 | 2.88e-04 | 3.33e-04 | 1.08e-14 | 0.01423 | -0.08646 | -0.07294 | 0.1334 | 0.1662 | -0.05784 | 0.09448 | 0.1503 | 1.38e-07 |
| BASOM | Basophils/100 leukocytes in Blood by Manual count | immune/inflammatory | % | 3957 | 3717 | 7674 | | 1.052 | 1.049 | 0.889 | 1.048 | 1.09 | 0.5272 | 0.04325 | -0.1858 | -0.1395 | 0.05811 | 0.0764 | -0.009626 | 0.002791 | 0.01253 | 0.7616 |
| BASOPHILS | BASOPHILS | immune/inflammatory | % | 592 | 885 | 1477 | | 0.8637 | 0.8876 | 0.2548 | 0.1122 | 0.09574 | 0.3329 | 0.08647 | -0.02494 | 0.06478 | 0.1026 | 0.1291 | 0.03648 | -0.01549 | -0.05266 | 0.6529 |
| BASORE | Basophils/100 leukocytes in Blood by Automated count | immune/inflammatory | % | 19975 | 25659 | 45634 | | 0.4748 | 0.4725 | 0.3943 | 0.05878 | 0.06132 | 0.00557 | 0.01752 | -0.05499 | -0.03835 | 0.06463 | 0.08475 | -0.0031 | 0.1223 | 0.1253 | 0.7773 |

| all_labs | full_name | group | selected_unit | male_n | female_n | total_n | sex_specific_lab | raw_median_male_mean | raw_median_female_mean | raw_median_p.value | raw_variance_female_mean | raw_variance_male_mean | raw_variance_p.value | std_mean_diff | median_male_mean | median_female_mean | median_p.value | median_p.value.fdr | std_variance_diff | variance_female_mean | variance_male_mean | variance_p.value |
|---|---|---|---|---|---|---|---|---|---|---|---|---|---|---|---|---|---|---|---|---|---|---|
| BBTAb | Borrelia burgdorferi Ab [Units/volume] in Serum by Immunoassay | toxicology/pharmacology | | 407 | 684 | 1091 | | 0.4115 | 0.4243 | 0.4531 | 0.02281 | 0.05242 | 0.376 | 0.01167 | -0.02042 | -0.009134 | 0.8554 | 0.8826 | -0.04735 | 0.08527 | 0.1305 | 0.8561 |
| BeGlg_ | Beta globulin [Mass/volume] in Serum or Plasma by Electrophoresis | immune/inflammatory | g/dL | 2889 | 3676 | 6565 | | 0.9876 | 1.025 | 2.22e-15 | 0.0188 | 0.02258 | 0.08929 | 0.1977 | -0.1196 | 0.06836 | 2.25e-15 | 5.53e-15 | -0.1248 | -0.09122 | 0.03054 | 0.01596 |
| BeHCG | Choriogonadotropin.beta subunit [Moles/volume] in Serum or Plasma | cancer | 10*-3.[iU]/mL | 67 | 1620 | 1687 | Sex specific lab | 194.66 | 14715.2 | 8.54e-76 | 753989129.7 | 64247692.0 | 3.94e-12 | 0.1996 | -0.2063 | -0.03588 | 0.1614 | 0.1981 | 0.4597 | 0.1717 | -0.2668 | 0.1369 |
| BEVN | Base excess standard in Venous blood by calculation | metabolic | mmol/L | 3698 | 3271 | 6969 | | 0.08185 | 0.05515 | 0.8513 | 24.04 | 20.79 | 0.02973 | 0.004095 | -0.003651 | 4.51e-04 | 0.865 | 0.8901 | 0.1006 | 0.1575 | 0.06114 | 0.005797 |
| BGPIgG | Beta 2 glycoprotein 1 IgG Ab in Serum | immune/inflammatory | | 449 | 1125 | 1574 | | 2.314 | 2.275 | 0.8992 | 9.479 | 12.12 | 0.8037 | 0.01691 | -0.04297 | -0.02577 | 0.7595 | 0.7997 | 0.205 | 0.1817 | -0.07068 | 0.1158 |
| BGPIgM | Beta 2 glycoprotein 1 IgM Ab in Serum | immune/inflammatory | | 452 | 1126 | 1578 | | 6.467 | 5.569 | 0.03656 | 8.168 | 13.11 | 0.4668 | 0.09952 | -0.03575 | 0.06818 | 0.09207 | 0.117 | 0.1871 | 0.1729 | -0.05371 | 0.1558 |
| Blasts | Blasts/100 leukocytes in Blood by Manual count | blood | | 867 | 769 | 1636 | | 4.939 | 5.126 | 0.767 | 152.67 | 135.70 | 0.458 | 0.009876 | -0.1143 | -0.1046 | 0.8418 | 0.8716 | -0.0228 | 0.01871 | 0.03977 | 0.748 |
| BLOOD PRESSURE | Diastolic blood pressure | vitals | mm[Hg] | 12921 | 19427 | 32348 | | 74.86 | 72.19 | 3.02e-150 | 91.48 | 89.38 | 0.007822 | -0.387 | 0.07567 | -0.2848 | 2.08e-263 | 1.84e-262 | 0.06445 | 0.1598 | 0.1037 | 4.32e-08 |
| BMI_CLEAN | BMI Clean | vitals | kg/m2 | 26408 | 35694 | 62102 | | 28.73 | 28.8 | 0.203 | 5.616 | 3.716 | 2.03e-74 | -0.05914 | -0.003466 | -0.06288 | 6.46e-14 | 1.53e-13 | 0.1437 | 0.5757 | 0.4523 | 6.12e-72 |
| BNP | Natriuretic peptide B [Mass/volume] in Serum or Plasma | cardiovascular | pg/mL | 7214 | 7050 | 14264 | | 334.12 | 274.52 | 7.20e-12 | 175651.5 | 206987.2 | 0.01607 | -0.1005 | 0.06422 | -0.03516 | 1.88e-09 | 3.91e-09 | -0.1765 | -0.002061 | 0.1651 | 8.53e-15 |
| BUN | Urea nitrogen serum/plasma | metabolic | mg/dL | 26975 | 34401 | 61376 | | 17.62 | 14.3 | 0 | 36.32 | 58 | 1.53e-136 | -0.3886 | 0.1841 | -0.2135 | 0 | 0 | -0.1275 | 0.04311 | 0.1727 | 4.42e-48 |
| C-Gluc | Glucose spinal fluid | metabolic | mg/dL | 1209 | 1515 | 2724 | | 68.81 | 67.26 | 0.05792 | 299.29 | 298.62 | 0.9865 | -0.0149 | -0.08606 | -0.101 | 0.7025 | 0.7476 | 0.01276 | -0.01496 | -0.02798 | 0.8595 |
| C3 | Complement C3 [Mass/volume] in Serum or Plasma | immune/inflammatory | mg/dL | 1372 | 2334 | 3706 | | 123.00 | 128.35 | 6.71e-07 | 395.41 | 586.81 | 0.02935 | 0.1554 | -0.1059 | 0.04402 | 4.53e-06 | 8.40e-06 | -0.08933 | 0.1225 | 0.2064 | 0.2692 |
| C4Quan | Complement C4 [Mass/volume] in Serum or Plasma | immune/inflammatory | mg/dL | 1374 | 2347 | 3721 | | 27.73 | 27.52 | 0.5348 | 29.87 | 29.3 | 0.9159 | 0.003965 | -0.0491 | -0.04526 | 0.9078 | 0.9264 | 0.0262 | 0.1183 | 0.09431 | 0.7313 |
| all_labs | full_name | group | selected_unit | male_n | female_n | total_n | sex_specific_lab | raw_median_male_mean | raw_median_female_mean | raw_median_p.value | raw_variance_female_mean | raw_variance_male_mean | raw_variance_p.value | std_mean_diff | median_male_mean | median_female_mean | median_p.value | median_p.value.fdr | std_variance_diff | variance_female_mean | variance_male_mean | variance_p.value |
| CA-125 | Cancer antigen 125 | cancer | [U]/mL | 119 | 1247 | 1366 | | 48.91 | 88.25 | 0.001446 | 126643.8 | 2600.13 | 2.34e-08 | 0.2159 | -0.2618 | -0.05524 | 0.01948 | 0.02667 | 0.4287 | 0.05458 | -0.3668 | 5.25e-04 |
| Ca | Calcium serum/plasma serum/plasma | metabolic | mg/dL | 26934 | 34341 | 61275 | | 9.167 | 9.253 | 6.01e-96 | 0.2818 | 0.2817 | 0.9542 | 0.156 | -0.07614 | 0.06033 | 4.08e-82 | 1.88e-81 | -0.005154 | 0.1677 | 0.1722 | 0.5516 |
| CA19-9 | Cancer Ag 19-9 | cancer | [U]/mL | 749 | 758 | 1507 | | 1921.10 | 1215.72 | 0.1039 | 55321215.8 | 44693229.5 | 0.6778 | -0.05551 | 0.04806 | -0.005394 | 0.2819 | 0.3328 | -0.07269 | -0.1299 | -0.05693 | 0.3575 |
| CaIo | Calcium.ionized [Mass/volume] in Blood | metabolic | mg/dL | 9019 | 7839 | 16858 | | 4.497 | 4.498 | 0.8481 | 0.3011 | 0.2416 | 0.002058 | 0.02703 | -0.07263 | -0.04758 | 0.08236 | 0.1069 | 0.03297 | 0.0308 | 7.81e-04 | 0.0766 |

| all_labs | full_name | group | selected_unit | male_n | female_n | total_n | sex_specific_lab | raw_median_male_mean | raw_median_female_mean | raw_median_p.value | raw_variance_female_mean | raw_variance_male_mean | raw_variance_p.value | std_mean_diff | median_male_mean | median_female_mean | median_p.value | median_p.value.fdr | std_variance_diff | variance_female_mean | variance_male_mean | variance_p.value |
|---|---|---|---|---|---|---|---|---|---|---|---|---|---|---|---|---|---|---|---|---|---|---|
| CBil | Bilirubin.conjugated [Mass/volume] in Serum or Plasma | liver | mg/dL | 12855 | 14196 | 27051 | | 0.4316 | 0.31 | 5.30e-37 | 0.313 | 0.4304 | 2.30e-05 | -0.2945 | -0.05193 | -0.3631 | 8.67e-130 | 4.79e-129 | -0.1824 | -0.08725 | 0.09843 | 1.05e-32 |
| CD3# | CD3# | immune/inflammatory | mm3 | 896 | 500 | 1396 | | 1356.62 | 1244.81 | 0.009775 | 177206.9 | 157699.1 | 0.2717 | -0.1786 | 0.08895 | -0.09729 | 0.001871 | 0.002888 | -0.05629 | 0.09739 | 0.1505 | 0.5103 |
| CD3% | CD3% | immune/inflammatory | % | 926 | 570 | 1496 | | 75.56 | 76.56 | 0.1468 | 87.31 | 33.67 | 1.20e-04 | 0.09615 | 0.03726 | 0.1348 | 0.07447 | 0.0974 | 0.3167 | 0.2239 | -0.1063 | 1.89e-04 |
| CD4# | CD3+CD4+ (T4 helper) cells [#/volume] in Blood | immune/inflammatory | mm3 | 894 | 498 | 1392 | | 571.43 | 716.61 | 2.24e-08 | 55081.4 | 30373.9 | 1.28e-04 | 0.2685 | -0.0138 | 0.2515 | 7.97e-06 | 1.45e-05 | 0.2352 | 0.2927 | 0.0683 | 0.007147 |
| CD4% | CD3+CD4+ (T4 helper) cells/100 cells in Blood | immune/inflammatory | % | 945 | 582 | 1527 | | 32.49 | 42.26 | 1.25e-28 | 99.75 | 40.04 | 2.28e-05 | 0.5832 | -0.09233 | 0.4849 | 2.79e-27 | 8.61e-27 | 0.394 | 0.3346 | -0.08669 | 1.02e-06 |
| CD8_3# | CD3+CD8+ (T8 suppressor cells) cells [#/volume] in Blood | immune/inflammatory | mm3 | 895 | 500 | 1395 | | 753.91 | 501.18 | 1.54e-23 | 55915.5 | 71716.5 | 0.05685 | -0.5267 | 0.1489 | -0.4094 | 9.40e-23 | 2.64e-22 | -0.393 | -0.2542 | 0.1428 | 4.33e-06 |
| CD8_3% | CD8_3% | immune/inflammatory | % | 952 | 584 | 1536 | | 39.2 | 29.42 | 2.84e-31 | 73.22 | 37.07 | 0.001319 | -0.5662 | 0.09933 | -0.4708 | 6.84e-29 | 2.15e-28 | 0.08676 | 0.01234 | -0.07415 | 0.3077 |
| CEA | Carcinoembryonic Ag [Mass/volume] in Serum or Plasma | cancer | ng/mL | 1266 | 1255 | 2521 | | 78.17 | 54.25 | 0.0897 | 101170.0 | 78193.7 | 0.5891 | -0.05732 | -0.06176 | -0.1199 | 0.1502 | 0.1848 | -0.01928 | -0.09276 | -0.07287 | 0.7362 |
| Cerul | Ceruloplasmin [Mass/volume] in Serum or Plasma | blood | mg/dL | 698 | 676 | 1374 | | 24.81 | 28.46 | 4.70e-15 | 20.77 | 21.86 | 0.9114 | 0.4549 | -0.3192 | 0.1435 | 1.30e-17 | 3.32e-17 | -0.2415 | -0.2352 | 0.01606 | 0.278 |
| CHOL_HDL_RATIO | Cholesterol in HDL [Mass/volume] in Serum or Plasma | metabolic | | 1033 | 1151 | 2184 | | 4.123 | 3.669 | 3.82e-20 | 0.4081 | 0.4905 | 0.139 | -0.4005 | 0.1909 | -0.2031 | 4.11e-21 | 1.12e-20 | -0.1484 | -0.08317 | 0.06342 | 0.01874 |
| Chol | Cholesterol [Mass/volume] in Serum or Plasma | metabolic | mg/dL | 15405 | 19280 | 34685 | | 169.64 | 189.65 | 0 | 875.41 | 897.52 | 0.3125 | 0.5227 | -0.3189 | 0.1837 | 0 | 0 | -0.004825 | 0.3018 | 0.306 | 0.6925 |
| CKMBRe | Creatine kinase.MB [Mass/volume] in Serum or Plasma | cardiovascular | ng/mL | 7983 | 6626 | 14609 | | 7.409 | 5.701 | 9.48e-13 | 113.76 | 189.05 | 1.14e-08 | -0.2749 | 0.03205 | -0.2468 | 8.46e-62 | 3.50e-61 | -0.09611 | -0.09598 | 0.001972 | 2.29e-06 |
| Cl-WB | Chloride [Moles/volume] in Blood | metabolic | mmol/L | 4307 | 3199 | 7506 | | 104.57 | 105.05 | 0.003359 | 13.7 | 13.31 | 0.917 | 0.1407 | -0.06916 | 0.06552 | 2.37e-09 | 4.90e-09 | -0.08095 | 0.1122 | 0.1902 | 0.006582 |
| Cl | Chloride [Moles/volume] in Serum or Plasma | metabolic | mmol/L | 26963 | 34342 | 61305 | | 103.63 | 103.96 | 5.33e-40 | 10.74 | 11.62 | 1.50e-15 | 0.08117 | -0.1146 | -0.03943 | 2.89e-23 | 8.16e-23 | -0.06928 | 0.225 | 0.285 | 1.13e-15 |
| CLymph | Lymphocytes/100 leukocytes in Cerebral spinal fluid by Manual count | immune/inflammatory | % | 881 | 1015 | 1896 | | 51.46 | 58.36 | 2.48e-07 | 489.55 | 473.44 | 0.74 | 0.1927 | -0.02499 | 0.1617 | 2.72e-05 | 4.84e-05 | 0.06021 | 0.1616 | 0.09967 | 0.4294 |
| CMonMc | Monocytes+Macrophages/100 leukocytes in Cerebral spinal fluid by Manual count | immune/inflammatory | % | 862 | 980 | 1842 | | 25.39 | 21.3 | 5.32e-06 | 233.27 | 223.14 | 0.7145 | -0.1729 | 0.1962 | 0.02018 | 2.23e-04 | 3.75e-04 | 0.01429 | 0.05168 | 0.03684 | 0.8542 |

| all_labs | full_name | group | selected_unit | male_n | female_n | total_n | sex_specific_lab | raw_median_male_mean | raw_median_female_mean | raw_median_p.value | raw_variance_female_mean | raw_variance_male_mean | raw_variance_p.value | std_mean_diff | median_male_mean | median_female_mean | median_p.value | median_p.value.fdr | std_variance_diff | variance_female_mean | variance_male_mean | variance_p.value |
|---|---|---|---|---|---|---|---|---|---|---|---|---|---|---|---|---|---|---|---|---|---|---|
| CNeut | Neutrophils/100 leukocytes in Cerebral spinal fluid by Manual count | immune/inflammatory | % | 622 | 643 | 1265 | | 34.63 | 32.87 | 0.3203 | 693.50 | 681.70 | 0.882 | -0.02317 | -0.07793 | -0.1006 | 0.6809 | 0.7293 | 0.04458 | 0.1574 | 0.1108 | 0.6308 |
| CNucCl | Nucleated cells [#/volume] in Cerebral spinal fluid | immune/inflammatory | /uL | 1062 | 1294 | 2356 | | 167.05 | 93.7 | 0.08478 | 1272306.0 | 2014194.8 | 0.2773 | -0.03309 | -0.05133 | -0.08343 | 0.4249 | 0.4735 | -0.05201 | -0.06444 | -0.01112 | 0.4204 |
| CO2 | Carbon dioxide serum/plasma | blood | mmol/L | 26971 | 34362 | 61333 | | 25.94 | 25.76 | 3.64e-20 | 6.866 | 7.227 | 2.51e-12 | -0.03612 | 0.2142 | 0.1835 | 9.53e-06 | 1.73e-05 | -0.08465 | 0.1631 | 0.2334 | 7.06e-23 |
| CoHgb | Carboxyhemoglobin % in blood | toxicology/pharmacology | % | 4118 | 3020 | 7138 | | 1.708 | 1.535 | 7.17e-13 | 0.2773 | 0.2871 | 0.6774 | -0.2311 | 0.1361 | -0.1052 | 7.37e-22 | 2.03e-21 | -0.05978 | 0.08377 | 0.1441 | 0.05094 |
| Copper | Copper in Serum or Plasma | toxicology/pharmacology | ug/dL | 893 | 1198 | 2091 | | 105.77 | 122.07 | 4.28e-33 | 531.29 | 376.73 | 0.04379 | 0.5285 | -0.3347 | 0.1778 | 5.90e-34 | 1.94e-33 | 0.2174 | 0.1648 | -0.03442 | 0.03333 |
| Cort-S | Cortisol [Mass/volume] in Serum or Plasma | endocrine | ug/dL | 3588 | 4273 | 7861 | | 16.71 | 14.93 | 4.01e-08 | 160.47 | 167.00 | 0.9051 | -0.1158 | -0.01266 | -0.1294 | 3.18e-07 | 6.19e-07 | 0.1338 | -0.02763 | -0.1595 | 1.43e-05 |
| CPepti | C peptide [Mass/volume] in Serum or Plasma | endocrine | ng/mL | 476 | 545 | 1021 | | 3.768 | 3.307 | 0.01181 | 3.504 | 5.968 | 0.05871 | -0.09092 | 0.1866 | 0.09253 | 0.1496 | 0.1845 | -0.2384 | 0.003345 | 0.2501 | 0.06479 |
| CPK | CPK | cardiovascular | [iU]/L | 9195 | 9073 | 18268 | | 264.59 | 164.15 | 3.89e-21 | 290344.3 | 546507.2 | 0.002074 | -0.431 | 0.1527 | -0.2366 | 6.98e-191 | 5.03e-190 | -0.1944 | 0.01369 | 0.1878 | 2.02e-26 |
| CPKTot | Creatine kinase [Enzymatic activity/volume] in Serum or Plasma | cardiovascular | [U]/L | 4635 | 4518 | 9153 | | 285.52 | 185.20 | 6.57e-07 | 414916.2 | 924490.4 | 0.008167 | -0.3594 | 0.1891 | -0.1136 | 2.94e-67 | 1.25e-66 | -0.1501 | 0.09047 | 0.2172 | 7.60e-08 |
| CProt1 | Protein [Mass/volume] in Cerebral spinal fluid | metabolic | mg/dL | 1211 | 1540 | 2751 | | 74.9 | 59.38 | 3.19e-05 | 14516.8 | 13127.7 | 0.7259 | -0.3166 | 0.09364 | -0.2258 | 1.14e-16 | 2.83e-16 | 0.1318 | 0.03364 | -0.1025 | 0.06682 |
| CR-Day | Creatinine urine per day | kidney | mg/(24.h) | 423 | 705 | 1128 | | 1518.01 | 1018.76 | 4.03e-43 | 52858.9 | 71009.4 | 0.3897 | -0.9208 | 0.5466 | -0.385 | 5.22e-49 | 1.98e-48 | -0.2579 | -0.03387 | 0.2151 | 0.03901 |
| Cr24Hr | Creatinine [Mass/time] in 24 hour Urine | kidney | g/(24.h) | 618 | 870 | 1488 | | 1.487 | 1.032 | 4.99e-08 | 10.02 | 0.2613 | 0.3296 | -0.8478 | 0.363 | -0.4613 | 6.16e-58 | 2.53e-57 | 0.311 | 0.1275 | -0.1792 | 0.006249 |
| CRBC | Erythrocytes in Cerebral spinal fluid | other | /uL | 1072 | 1296 | 2368 | | 3766.14 | 3282.85 | 0.5856 | 654296081.0 | 692350381.3 | 0.8987 | 0.1268 | -0.2444 | -0.1228 | 0.00232 | 0.003548 | 0.1374 | -0.07418 | -0.2058 | 0.03283 |
| Creat | Creatinine [Mass/volume] in Blood Creatinine [Mass/volume] in Blood | kidney | mg/dL | 27025 | 34787 | 61812 | | 1.13 | 0.863 | 0 | 0.08941 | 0.1659 | 1.53e-74 | -0.7925 | 0.3857 | -0.4407 | 0 | 0 | -0.2082 | -0.001756 | 0.2003 | 5.51e-128 |
| CREATININE | Creatinine | kidney | mg/dL | 755 | 879 | 1634 | | 1.725 | 1.17 | 2.97e-09 | 0.4379 | 0.4778 | 0.7819 | -0.7515 | 0.5029 | -0.1965 | 8.35e-57 | 3.37e-56 | -0.2802 | 0.02625 | 0.2933 | 2.58e-04 |
| CrePOC | CrePOC | kidney | mg/dL | 3717 | 2951 | 6668 | | 1.167 | 0.9574 | 1.99e-75 | 0.04838 | 0.06898 | 0.3662 | -0.6922 | 0.2865 | -0.4061 | 3.00e-180 | 1.99e-179 | -0.1429 | -0.04438 | 0.09763 | 1.83e-04 |
| CRP | C reactive protein [Mass/volume] in Serum or Plasma | immune/inflammatory | mg/L | 8464 | 11849 | 20313 | | 38.96 | 22.41 | 1.23e-110 | 1378.61 | 2520.18 | 8.34e-51 | -0.2629 | 0.1191 | -0.1583 | 1.01e-73 | 4.55e-73 | -0.3353 | -0.1288 | 0.2168 | 2.29e-69 |
| CTo#Cl | Cells Counted Total in CSF | other | | 888 | 1019 | 1907 | | 76.15 | 74.41 | 0.2601 | 688.30 | 692.36 | 0.9609 | -0.09471 | 0.0257 | -0.062 | 0.03984 | 0.05308 | -0.04163 | 0.1291 | 0.1681 | 0.5841 |
| D25OHT | Calcitriol [Mass/volume] in Serum or Plasma | metabolic | ng/mL | 7363 | 13921 | 21284 | | 31.85 | 34.41 | 1.39e-43 | 119.34 | 102.79 | 1.27e-07 | 0.2108 | -0.1025 | 0.09311 | 2.04e-49 | 7.85e-49 | 0.1191 | 0.1074 | -0.005925 | 6.08e-11 |

| all_labs | full_name | group | selected_unit | male_n | female_n | total_n | sex_specific_lab | raw_median_male_mean | raw_median_female_mean | raw_median_p.value | raw_variance_female_mean | raw_variance_male_mean | raw_variance_p.value | std_mean_diff | median_male_mean | median_female_mean | median_p.value | median_p.value.fdr | std_variance_diff | variance_female_mean | variance_male_mean | variance_p.value |
|---|---|---|---|---|---|---|---|---|---|---|---|---|---|---|---|---|---|---|---|---|---|---|
| DHEA-S | DHEA-S in Serum or Plasma | endocrine | ug/dL | 119 | 1077 | 1196 | | 133.57 | 146.37 | 0.3079 | 5132.30 | 3362.42 | 0.4168 | -0.4977 | 0.254 | -0.1935 | 1.33e-07 | 2.63e-07 | 0.1516 | 0.06187 | -0.08355 | 0.4841 |
| DIASTOLIC | Diastolic blood pressure | vitals | mm[Hg] | 26957 | 35815 | 62772 | | 72.86 | 70.7 | 2.93e-166 | 182.30 | 178.10 | 0.6894 | -0.2775 | -0.09004 | -0.3393 | 6.76e-261 | 5.79e-260 | -0.0699 | 0.2953 | 0.3533 | 1.48e-17 |
| Digox | Digoxin [Mass/volume] in Serum or Plasma | toxicology/pharmacology | ng/mL | 794 | 551 | 1345 | | 0.889 | 1.001 | 9.05e-05 | 0.2663 | 0.2556 | 0.8036 | 0.1956 | -0.1934 | -0.01 | 4.58e-04 | 7.49e-04 | 0.25 | 0.1304 | -0.1106 | 3.34e-04 |
| Dimer | Dimer | blood | ug{FEU}/mL | 793 | 962 | 1755 | | 2.472 | 1.94 | 1.09e-04 | 4.142 | 6.602 | 0.06936 | -0.1036 | 0.119 | 0.006381 | 0.03231 | 0.04376 | -0.123 | 0.0303 | 0.1547 | 0.1834 |
| DirLDL | Cholesterol in LDL [Mass/volume] in Serum or Plasma by Direct assay | metabolic | mg/dL | 2992 | 2604 | 5596 | | 100.92 | 114.00 | 2.62e-39 | 753.47 | 622.48 | 0.006323 | 0.3688 | -0.1681 | 0.1819 | 9.94e-44 | 3.53e-43 | 0.173 | 0.1614 | -0.008705 | 2.25e-05 |
| DLCO_COR_PPR | DLCO_COR_PPR | lung | | 1268 | 1226 | 2494 | | 65.47 | 65.24 | 0.8031 | 66.21 | 81.3 | 0.1746 | -0.0683 | 0.03431 | -0.03195 | 0.08807 | 0.1128 | -0.1773 | -0.03313 | 0.1339 | 0.02893 |
| DLCO_COR_PRD | DLCO_COR_PRD | lung | | 1955 | 2056 | 4011 | | 27.23 | 21.39 | 0 | 0.2123 | 0.3396 | 9.55e-04 | -1.58 | 0.6973 | -0.704 | 0 | 0 | -0.3982 | 0.06544 | 0.4486 | 9.15e-13 |
| DLCO_COR_PRL | DLCO_COR_PRL | lung | | 1269 | 1225 | 2494 | | 19.43 | 14.91 | 3.75e-279 | 0.3823 | 0.1474 | 4.79e-04 | -1.572 | 0.7024 | -0.7114 | 0 | 0 | -0.07465 | 0.04841 | 0.1191 | 0.3656 |
| DLCO_COR_PRU | DLCO_COR_PRU | lung | | 1269 | 1225 | 2494 | | 35.43 | 28.1 | 0 | 1.222 | 0.1692 | 3.09e-04 | -1.592 | 0.6981 | -0.7656 | 0 | 0 | 0.1342 | 0.1249 | 0.004591 | 0.1043 |
| DLCO_COR_ZBD | DLCO_COR_ZBD | lung | | 1955 | 2056 | 4011 | | 18.21 | 14.34 | 8.25e-78 | 4.242 | 7.063 | 1.15e-06 | -0.7288 | 0.3771 | -0.3047 | 3.16e-124 | 1.67e-123 | -0.3994 | -0.05929 | 0.3102 | 6.41e-13 |
| DLCO_UNC_PPR | DLCO_UNC_PPR | lung | | 1269 | 1227 | 2496 | | 63.97 | 64.42 | 0.6278 | 66.1 | 81.12 | 0.1894 | -0.03639 | 0.02708 | -0.008082 | 0.3635 | 0.4143 | -0.1743 | -0.01044 | 0.1513 | 0.03314 |
| DLCO_UNC_PRD | DLCO_UNC_PRD | lung | | 1955 | 2057 | 4012 | | 27.23 | 21.38 | 0 | 0.2123 | 0.3396 | 9.55e-04 | -1.58 | 0.6973 | -0.704 | 0 | 0 | -0.3982 | 0.06544 | 0.4486 | 9.15e-13 |
| DLCO_UNC_PRL | DLCO_UNC_PRL | lung | | 1269 | 1226 | 2495 | | 19.43 | 14.91 | 2.34e-279 | 0.3823 | 0.1474 | 4.79e-04 | -1.572 | 0.7025 | -0.7113 | 0 | 0 | -0.07465 | 0.04841 | 0.1191 | 0.3656 |
| DLCO_UNC_PRU | DLCO_UNC_PRU | lung | | 1269 | 1226 | 2495 | | 35.43 | 28.1 | 0 | 1.222 | 0.1692 | 3.09e-04 | -1.592 | 0.6981 | -0.7656 | 0 | 0 | 0.1342 | 0.1249 | 0.004591 | 0.1043 |
| DLCO_UNC_ZBD | DLCO_UNC_ZBD | lung | | 1269 | 1227 | 2496 | | 17.73 | 13.99 | 1.91e-45 | 3.188 | 6.687 | 4.28e-05 | -0.6795 | 0.3255 | -0.3177 | 9.99e-69 | 4.28e-68 | -0.5204 | -0.1619 | 0.3173 | 1.25e-10 |
| DLCOOVA_PPR | DLCOOVA_PPR | lung | % | 976 | 1152 | 2128 | | 71.49 | 70.97 | 0.5603 | 79.46 | 73.82 | 0.6101 | -0.0636 | -0.007074 | -0.07009 | 0.1469 | 0.1821 | -0.05961 | -0.03784 | 0.018 | 0.4488 |
| DLCOOVA_PRD | DLCOOVA_PRD | lung | | 1954 | 2056 | 4010 | | 4.429 | 4.761 | 7.97e-113 | 0.005335 | 0.0141 | 1.32e-10 | 1.109 | -0.5791 | 0.5258 | 0 | 0 | -0.43 | 0.09889 | 0.505 | 1.19e-14 |
| DLCOOVA_ZBD | DLCOOVA_ZBD | lung | | 1954 | 2057 | 4011 | | 3.162 | 3.346 | 1.48e-08 | 0.2259 | 0.1804 | 0.05543 | 0.0993 | -0.1138 | -0.0168 | 0.001664 | 0.002601 | 0.01508 | 0.08199 | 0.06801 | 0.7875 |
| eGFR | eGFR | kidney | mL/min | 680 | 936 | 1616 | | 47.36 | 47.94 | 0.3294 | 58.89 | 59.04 | 0.9801 | 0.03334 | -0.03536 | -0.002161 | 0.5104 | 0.56 | -0.003734 | 0.0735 | 0.07721 | 0.9534 |
| EGFR | eGFR | kidney | mL/min/1.73.m2 | 27030 | 34789 | 61819 | | 83.75 | 84.01 | 0.2289 | 250.31 | 266.70 | 2.62e-07 | -0.1186 | -3.56e-04 | -0.1173 | 2.36e-47 | 8.89e-47 | -0.1013 | 0.3639 | 0.4514 | 4.41e-36 |
| eGFRcr | eGFRcr | kidney | | 1251 | 1649 | 2900 | | 62.8 | 66.99 | 1.20e-08 | 66.56 | 60.09 | 0.4023 | 0.1522 | -0.1079 | 0.04789 | 6.31e-05 | 1.10e-04 | -0.01125 | -0.08093 | -0.07042 | 0.885 |
| all_labs | full_name | group | selected_unit | male_n | female_n | total_n | sex_specific_lab | raw_median_male_mean | raw_median_female_mean | raw_median_p.value | raw_variance_female_mean | raw_variance_male_mean | raw_variance_p.value | std_mean_diff | median_male_mean | median_female_mean | median_p.value | median_p.value.fdr | std_variance_diff | variance_female_mean | variance_male_mean | variance_p.value |
| Endomy | Endomysium IgA Ab [Presence] in Serum | immune/inflammatory | [U] | 277 | 817 | 1094 | | 5.619 | 5.025 | 0.1881 | 65.68 | 86.47 | 0.7821 | -0.2181 | 0.1533 | -0.06752 | 0.001737 | 0.002706 | 0.119 | 0.1477 | 0.02239 | 0.7621 |
| EoAbs | EoAbs | immune/inflammatory | 10*3/uL | 10829 | 14884 | 25713 | | 0.1805 | 0.1604 | 1.38e-30 | 0.01313 | 0.0159 | 8.23e-06 | -0.1286 | 0.1172 | -0.004204 | 4.84e-24 | 1.40e-23 | -0.1058 | -0.04966 | 0.05419 | 2.54e-11 |
| Eos | Eos | immune/inflammatory | % | 11491 | 15677 | 27168 | | 2.475 | 2.19 | 1.85e-44 | 1.973 | 2.354 | 7.49e-10 | -0.1389 | 0.1227 | -0.01112 | 3.37e-29 | 1.07e-28 | -0.1171 | -0.02499 | 0.08805 | 1.71e-14 |

| all_labs | full_name | group | selected_unit | male_n | female_n | total_n | sex_specific_lab | raw_median_male_mean | raw_median_female_mean | raw_median_p.value | raw_variance_female_mean | raw_variance_male_mean | raw_variance_p.value | std_mean_diff | median_male_mean | median_female_mean | median_p.value | median_p.value.fdr | std_variance_diff | variance_female_mean | variance_male_mean | variance_p.value |
|---|---|---|---|---|---|---|---|---|---|---|---|---|---|---|---|---|---|---|---|---|---|---|
| EOSIA | Eosinophils [#/volume] in Blood by Automated count | immune/inflammatory | 10*3/uL | 15202 | 19735 | 34937 | | 0.1731 | 0.1535 | 1.28e-31 | 0.01652 | 0.02057 | 9.93e-08 | -0.08831 | 0.09757 | 0.009486 | 5.45e-16 | 1.35e-15 | -0.1136 | 0.01873 | 0.135 | 1.44e-19 |
| EOSIAB | Eosinophils [#/volume] in Blood by Automated count | immune/inflammatory | 10*3/uL | 12637 | 15257 | 27894 | | 0.1744 | 0.1561 | 4.21e-24 | 0.01511 | 0.01835 | 1.93e-04 | -0.1078 | 0.104 | -0.001796 | 4.25e-19 | 1.12e-18 | -0.08807 | -0.02907 | 0.06172 | 2.52e-08 |
| EOSIM | Eosinophils/100 leukocytes in Blood by Automated count | immune/inflammatory | % | 15117 | 19669 | 34786 | | 2.28 | 2.049 | 3.82e-37 | 2.053 | 2.453 | 2.07e-17 | -0.08668 | 0.1026 | 0.01696 | 2.26e-15 | 5.54e-15 | -0.1094 | 0.03985 | 0.1499 | 3.18e-18 |
| EOSINOPHILS | EOSINOPHILS | immune/inflammatory | % | 730 | 1111 | 1841 | | 2.609 | 2.354 | 0.00327 | 1.818 | 2.124 | 0.2074 | -0.08341 | -5.32e-04 | -0.08718 | 0.08612 | 0.111 | -0.008093 | 0.03514 | 0.04355 | 0.9113 |
| EOSIRE | Eosinophils % | immune/inflammatory | % | 12813 | 15398 | 28211 | | 2.304 | 2.028 | 5.10e-40 | 2.013 | 2.291 | 2.39e-06 | -0.1174 | 0.1147 | -0.001011 | 1.49e-22 | 4.15e-22 | -0.1047 | -0.00948 | 0.09622 | 2.23e-11 |
| Estrdl | Estradiol (E2) [Mass/volume] in Serum or Plasma | endocrine | pg/mL | 169 | 2960 | 3129 | | 48.19 | 94.95 | 6.35e-25 | 16881.1 | 2845.60 | 4.64e-08 | 0.3932 | -0.3871 | 0.005691 | 1.25e-08 | 2.57e-08 | 0.3475 | 0.06496 | -0.2673 | 0.05871 |
| F-Gluc | Glucose [Mass/volume] in Body fluid | metabolic | mg/dL | 975 | 716 | 1691 | | 114.57 | 114.25 | 0.9146 | 986.23 | 1164.99 | 0.4814 | 0.03046 | -0.06072 | -0.02917 | 0.5303 | 0.5779 | 0.03429 | 0.2328 | 0.2 | 0.6604 |
| Fbrngn | Fibrinogen [Mass/volume] in Platelet poor plasma by Coagulation assay | blood | mg/dL | 3225 | 2953 | 6178 | | 351.31 | 373.35 | 5.13e-08 | 9079.51 | 10752.7 | 0.005603 | 0.1173 | -0.09506 | 0.02038 | 3.92e-06 | 7.32e-06 | -0.07055 | 0.07545 | 0.1448 | 0.04343 |
| FEF25_75_AFD | FEF AFD | lung | | 810 | 1018 | 1828 | | 1.788 | 1.768 | 0.7225 | 0.1162 | 0.09955 | 0.6426 | -0.2464 | 0.07833 | -0.1524 | 1.84e-07 | 3.63e-07 | -0.222 | 0.09236 | 0.2697 | 0.0383 |
| FEF25_75_PPR | FEF PPR | lung | % | 1210 | 1559 | 2769 | | 76.54 | 80.02 | 0.02554 | 242.63 | 240.17 | 0.9445 | 0.1057 | -0.159 | -0.05156 | 0.006032 | 0.00874 | 0.08593 | -0.2534 | -0.3418 | 0.1583 |
| FEF25_75_PRD | FEF PRD | lung | | 2375 | 2730 | 5105 | | 2.834 | 2.319 | 9.11e-130 | 0.01181 | 0.03509 | 3.21e-13 | -1.53 | 0.8522 | -0.5378 | 0 | 0 | -0.1201 | 0.08998 | 0.2255 | 0.008516 |
| FEF25_75_PRL | FEF PRL | lung | | 1581 | 1686 | 3267 | | 1.345 | 1.147 | 8.48e-18 | 0.002168 | 0.02097 | 1.09e-12 | -1.449 | 0.8355 | -0.438 | 0 | 0 | -0.1804 | -0.1485 | 0.0442 | 0.003649 |
| FEF25_75_ZBD | FEF ZBD | lung | | 2375 | 2730 | 5105 | | 2.134 | 1.841 | 3.32e-19 | 0.1732 | 0.2413 | 0.008149 | -0.3642 | 0.1241 | -0.2297 | 2.96e-38 | 1.01e-37 | -0.2392 | -0.05024 | 0.1588 | 7.54e-08 |
| FEF25_75_ZPC | FEF ZPC | lung | | 497 | 576 | 1073 | | 14.89 | 15.17 | 0.8375 | 602.52 | 399.75 | 0.1104 | -0.02314 | 0.03767 | 0.01479 | 0.7051 | 0.7488 | 0.2089 | 0.2032 | 0.01259 | 0.1814 |
| FEosin | Eosinophils/100 leukocytes in Body fluid by Manual count | immune/inflammatory | % | 669 | 512 | 1181 | | 3.753 | 3.493 | 0.427 | 26.36 | 12.78 | 0.06918 | -0.06206 | 0.05237 | -0.007912 | 0.2896 | 0.3402 | 0.1806 | 0.1043 | -0.07539 | 0.1368 |
| Ferrit | Ferritin [Mass/volume] in Serum or Plasma | blood | ng/mL | 8034 | 11119 | 19153 | | 351.39 | 171.67 | 1.82e-24 | 1494663.8 | 4054734.6 | 0.1429 | -0.434 | 0.06376 | -0.3719 | 4.22e-191 | 3.09e-190 | -0.312 | -7.84e-04 | 0.3082 | 4.36e-55 |
| FET_AFD | FET AFD | lung | | 503 | 585 | 1088 | | 9.749 | 9.069 | 1.45e-04 | 3.697 | 3.114 | 0.4601 | -0.1699 | 0.1146 | -0.06087 | 0.005375 | 0.007881 | 0.1151 | 0.07814 | -0.02231 | 0.4639 |
| FET_ZBD | FET ZBD | lung | | 1595 | 1730 | 3325 | | 8.641 | 8.099 | 2.15e-09 | 3.484 | 4.006 | 0.1209 | -0.203 | 0.1235 | -0.0831 | 4.92e-09 | 1.01e-08 | -0.04731 | -0.05065 | -0.002426 | 0.431 |
| FET_ZPC | FET ZPC | lung | | 485 | 580 | 1065 | | 9.499 | 8.169 | 0.3188 | 633.99 | 285.54 | 0.03842 | -0.006903 | -0.02329 | -0.03008 | 0.9097 | 0.9265 | 0.4948 | 0.3487 | -0.08971 | 0.001435 |
| FEV1_PPR | FEV1 PPR | lung | % | 1214 | 1562 | 2776 | | 78.03 | 81.79 | 1.47e-05 | 61.46 | 63.45 | 0.7972 | 0.1526 | -0.1343 | 0.01995 | 6.76e-05 | 1.17e-04 | -0.07658 | -0.01205 | 0.05645 | 0.203 |
| FEV1_PRD | FEV1 PRD | lung | L | 1214 | 1562 | 2776 | | 3.382 | 2.487 | 5.26e-277 | 0.002882 | 0.004661 | 1.25e-04 | -1.594 | 0.8379 | -0.5612 | 0 | 0 | -0.1575 | 0.1566 | 0.3103 | 0.009672 |
| FEV1_PRL | FEV1 PRL | lung | | 1604 | 1735 | 3339 | | 2.672 | 1.946 | 2.99e-303 | 0.002043 | 0.005185 | 3.75e-05 | -1.588 | 0.8466 | -0.5631 | 0 | 0 | -0.198 | -0.05562 | 0.1476 | 0.00107 |

| all_labs | full_name | group | selected_unit | male_n | female_n | total_n | sex_specific_lab | raw_median_male_mean | raw_median_female_mean | raw_median_p.value | raw_variance_female_mean | raw_variance_male_mean | raw_variance_p.value | std_mean_diff | median_male_mean | median_female_mean | median_p.value | median_p.value.fdr | std_variance_diff | variance_female_mean | variance_male_mean | variance_p.value |
|---|---|---|---|---|---|---|---|---|---|---|---|---|---|---|---|---|---|---|---|---|---|---|
| FEV1_ZBD | FEV1 ZBD | lung | L | 1214 | 1562 | 2776 | | 2.639 | 2.039 | 6.67e-71 | 0.04141 | 0.08323 | 4.49e-06 | -0.8345 | 0.4252 | -0.3698 | 1.04e-106 | 5.35e-106 | -0.4315 | -0.06611 | 0.3162 | 6.31e-13 |
| FEV1_ZPC | FEV1 ZPC | lung | | 488 | 567 | 1055 | | 8.753 | 7.907 | 0.1137 | 83.74 | 58.8 | 0.1567 | -0.0542 | 0.08236 | 0.02782 | 0.3808 | 0.4321 | 0.1639 | 0.1405 | -0.008838 | 0.2985 |
| FEV1OFVC_AFD | FEV1/FVC AFD | lung | | 506 | 587 | 1093 | | 64.8 | 69.98 | 2.60e-09 | 15.85 | 9.706 | 0.07573 | 0.2473 | -0.2284 | 0.03171 | 4.31e-05 | 7.59e-05 | 0.04649 | 0.1088 | 0.06997 | 0.761 |
| FEV1OFVC_APP | FEV1/FVC APP | lung | | 506 | 587 | 1093 | | 85.45 | 89.13 | 7.18e-04 | 26.03 | 17.22 | 0.1093 | 0.1331 | -0.1557 | -0.01353 | 0.02877 | 0.03918 | 0.01781 | 0.1218 | 0.1071 | 0.9073 |
| FEV1OFVC_PPR | FEV1/FVC PPR | lung | | 1604 | 1735 | 3339 | | 93.36 | 94.21 | 0.1277 | 35.26 | 34.18 | 0.8972 | 0.02395 | -0.0906 | -0.06575 | 0.4911 | 0.5411 | -0.06743 | 0.03988 | 0.1086 | 0.2551 |
| FEV1OFVC_PRD | FEV1/FVC PRD | lung | | 1604 | 1735 | 3339 | | 75.74 | 78.38 | 5.03e-117 | 0.2175 | 0.7616 | 3.93e-07 | 1.146 | -0.6093 | 0.4638 | 7.28e-268 | 6.58e-267 | -0.01323 | -0.009219 | 0.005156 | 0.8281 |
| FEV1OFVC_PRL | FEV1/FVC PRL | lung | | 1604 | 1735 | 3339 | | 66.34 | 68.67 | 1.58e-89 | 0.2979 | 1.462 | 4.91e-09 | 0.9728 | -0.5288 | 0.4829 | 8.61e-180 | 5.63e-179 | -0.07073 | -0.08497 | -0.006422 | 0.247 |
| FEV1OFVC_ZBD | FEV1/FVC ZBD | lung | | 1604 | 1734 | 3338 | | 70.72 | 73.85 | 3.62e-13 | 21.86 | 19.09 | 0.5552 | 0.2264 | -0.212 | 0.02027 | 5.65e-11 | 1.25e-10 | -0.03887 | 0.1174 | 0.1533 | 0.5093 |
| FIO2 | Oxygen/Inspired gas Respiratory system --on ventilator | blood | | 4836 | 3411 | 8247 | | 15.12 | 14.94 | 0.7368 | 218.45 | 163.68 | 0.2551 | 0.03756 | -0.1833 | -0.1453 | 0.0931 | 0.118 | 0.06673 | -0.169 | -0.2266 | 0.009405 |
| FK-506 | Tacrolimus [Mass/volume] in Blood | toxicology/pharmacology | ng/mL | 960 | 673 | 1633 | | 7.398 | 7.468 | 0.764 | 15.98 | 14.29 | 0.1119 | -0.02295 | -0.05759 | -0.07934 | 0.652 | 0.7014 | 0.09151 | 0.1517 | 0.06725 | 0.09043 |
| FLymph | Lymphocytes [#/volume] in Body fluid by Manual count | immune/inflammatory | % | 1616 | 1301 | 2917 | | 29.67 | 28.98 | 0.4624 | 417.27 | 374.19 | 0.2221 | 0.01418 | -0.03336 | -0.01881 | 0.7022 | 0.7476 | 0.05088 | 0.05877 | 0.007178 | 0.3934 |
| FMoncl | Monocytes+Macrophages/100 leukocytes in Body fluid by Manual count | immune/inflammatory | % | 793 | 660 | 1453 | | 35.04 | 36.25 | 0.4185 | 496.47 | 435.72 | 0.2438 | 0.03708 | 0.009308 | 0.04652 | 0.4795 | 0.5296 | 0.02469 | 0.1642 | 0.1392 | 0.7686 |
| FMonMc | Mononuclear cells [#/volume] in Body fluid by Manual count | immune/inflammatory | % | 916 | 744 | 1660 | | 38.27 | 38.35 | 0.9576 | 518.46 | 481.13 | 0.4772 | -5.34e-04 | -0.04061 | -0.04113 | 0.9914 | 0.9914 | 0.03726 | -0.006453 | -0.04227 | 0.6423 |
| FNeut | Neutrophils/100 leukocytes in Body fluid by Manual count | immune/inflammatory | % | 1610 | 1288 | 2898 | | 34.33 | 34 | 0.7794 | 598.14 | 556.07 | 0.3424 | -0.03604 | 0.06893 | 0.03301 | 0.3331 | 0.3832 | 0.01507 | 0.1083 | 0.09418 | 0.8004 |
| FNucCl | Nucleated cells [#/volume] in Synovial fluid by Manual count | blood | /uL | 1952 | 1574 | 3526 | | 12552.9 | 7016.80 | 0.005045 | 711415700.7 | 1704204273.2 | 0.19 | -0.1403 | 0.1523 | 0.01772 | 3.37e-05 | 5.95e-05 | -0.0496 | 0.07157 | 0.1226 | 0.304 |
| FProt1 | Protein body fluid | metabolic | g/dL | 990 | 743 | 1733 | | 2.991 | 2.88 | 0.08507 | 0.4633 | 0.4542 | 0.8984 | -0.09478 | 0.1629 | 0.07942 | 0.05185 | 0.06836 | 0.02056 | 0.2115 | 0.1915 | 0.7817 |
| FRBC1 | Erythrocytes [#/volume] in Blood by Automated count | blood | /uL | 1855 | 1498 | 3353 | | 51523.1 | 44460.4 | 0.2078 | 9537469619.6 | 9708839710.0 | 0.9461 | -0.07464 | -0.002058 | -0.07829 | 0.03134 | 0.04256 | -0.07916 | 0.06691 | 0.1495 | 0.1202 |
| FRC_PPR | FRC PPR | lung | | 1145 | 1144 | 2289 | | 105.52 | 101.99 | 0.01522 | 417.30 | 236.62 | 0.3238 | -0.06891 | 0.112 | 0.04436 | 0.09906 | 0.1253 | 0.03863 | 0.05306 | 0.01319 | 0.665 |
| FRC_PRD | FRC PRD | lung | | 1144 | 1147 | 2291 | | 3.53 | 2.823 | 0 | 0.003546 | 0.01634 | 2.10e-06 | -1.39 | 0.7497 | -0.5941 | 0 | 0 | -0.05619 | -0.05991 | -0.002966 | 0.5226 |
| FRC_PRL | FRC PRL | lung | | 1145 | 1146 | 2291 | | 2.087 | 1.776 | 6.77e-96 | 0.004175 | 0.03444 | 8.58e-09 | -0.808 | 0.4613 | -0.3179 | 1.10e-89 | 5.31e-89 | -0.3697 | -0.1937 | 0.1744 | 2.36e-05 |
| FRC_PRU | FRC PRU | lung | | 1144 | 1142 | 2286 | | 4.973 | 3.87 | 0 | 0.01249 | 0.02938 | 1.33e-04 | -1.56 | 0.8336 | -0.681 | 0 | 0 | -0.1873 | -0.05005 | 0.1317 | 0.03451 |
| FRC_ZBD | FRC ZBD | lung | | 1145 | 1145 | 2290 | | 3.692 | 2.875 | 9.91e-67 | 0.3308 | 0.2863 | 0.7742 | -0.7135 | 0.4477 | -0.2397 | 8.86e-70 | 3.83e-69 | -0.169 | -0.07237 | 0.1062 | 0.05969 |

| all_labs | full_name | group | selected_unit | male_n | female_n | total_n | sex_specific_lab | raw_median_male_mean | raw_median_female_mean | raw_median_p.value | raw_variance_female_mean | raw_variance_male_mean | raw_variance_p.value | std_mean_diff | median_male_mean | median_female_mean | median_p.value | median_p.value.fdr | std_variance_diff | variance_female_mean | variance_male_mean | variance_p.value |
|---|---|---|---|---|---|---|---|---|---|---|---|---|---|---|---|---|---|---|---|---|---|---|
| FRONTAL AXIS INITIAL 40 MS | Frontal axis initial 40 ms | cardiovascular | | 6253 | 7138 | 13391 | | 31.04 | 32.93 | 0.01125 | 845.08 | 1116.62 | 8.03e-06 | 0.04438 | -0.07106 | -0.02416 | 0.01123 | 0.01576 | -0.03431 | -0.00627 | 0.0301 | 0.1131 |
| FRONTAL AXIS MEAN QRS | QRS axis | cardiovascular | | 6315 | 7218 | 13533 | | 17.84 | 24.46 | 3.11e-15 | 860.92 | 1274.05 | 3.59e-08 | 0.094 | -0.08633 | 0.01496 | 7.01e-08 | 1.40e-07 | -0.04105 | -0.09861 | -0.0537 | 0.05546 |
| FRONTAL AXIS P | P wave axis | cardiovascular | | 6062 | 7040 | 13102 | | 39.62 | 45.11 | 1.03e-28 | 629.60 | 796.60 | 7.79e-08 | 0.1873 | -0.1854 | 0.01087 | 1.20e-26 | 3.67e-26 | -0.0527 | -0.0288 | 0.02891 | 0.01721 |
| FRONTAL AXIS ST | ST segment axis | cardiovascular | | 6301 | 7193 | 13494 | | 72.47 | 75.76 | 0.006486 | 4267.92 | 3911.23 | 0.02363 | 0.1091 | -0.0519 | 0.05683 | 2.58e-10 | 5.64e-10 | 0.04652 | -0.02811 | -0.07637 | 0.03066 |
| FRONTAL AXIS TERMINAL 40 MS | Frontal axis terminal 40 ms | cardiovascular | | 6296 | 7197 | 13493 | | 55.92 | 37.93 | 3.74e-35 | 2594.81 | 3749.13 | 1.43e-11 | -0.1888 | 0.1139 | -0.08372 | 2.54e-27 | 7.90e-27 | -0.07553 | -0.1204 | -0.03891 | 4.32e-04 |
| FSH1 | Follitropin [Units/volume] in Serum or Plasma | endocrine | 10*-3.[iU]/mL | 659 | 3733 | 4392 | | 9.688 | 29.49 | 5.74e-134 | 649.07 | 54.57 | 9.19e-44 | 1.25 | -1.297 | 0.09543 | 1.44e-155 | 8.73e-155 | 0.4383 | 0.04696 | -0.4153 | 2.97e-08 |
| FT4 | Thyroxine (T4) free [Mass/volume] in Serum or Plasma | endocrine | ng/dL | 10236 | 17572 | 27808 | | 1.008 | 0.9985 | 0.343 | 0.0631 | 0.06397 | 0.8958 | 0.04456 | -0.1126 | -0.06858 | 3.42e-04 | 5.62e-04 | -0.1479 | 0.1371 | 0.2726 | 2.20e-19 |
| FTo#Cl | Cells counted in body fluid | other | | 1663 | 1347 | 3010 | | 98.46 | 98.34 | 0.7391 | 127.60 | 158.94 | 0.2944 | 0.1087 | -0.1312 | -0.02665 | 0.003359 | 0.005106 | -0.1193 | -0.02797 | 0.08995 | 0.04232 |
| FVC_PPR | FVC PPR | lung | % | 1214 | 1562 | 2776 | | 81.92 | 84.78 | 9.60e-05 | 54 | 52.88 | 0.8498 | 0.1333 | -0.09576 | 0.03538 | 4.89e-04 | 7.94e-04 | -0.01391 | -0.06587 | -0.05156 | 0.8182 |
| FVC_PRD | FVC PRD | lung | L | 1214 | 1562 | 2776 | | 4.499 | 3.189 | 0 | 0.003913 | 0.006305 | 0.001044 | -1.618 | 0.8604 | -0.5746 | 0 | 0 | -0.07659 | 0.1828 | 0.2577 | 0.2077 |
| FVC_PRL | FVC PRL | lung | | 1604 | 1736 | 3340 | | 3.607 | 2.535 | 0 | 0.003406 | 0.004231 | 0.4033 | -1.599 | 0.8403 | -0.5833 | 0 | 0 | -0.1122 | 0.06394 | 0.1706 | 0.05997 |
| FVC_ZBD | FVC ZBD | lung | L | 1214 | 1562 | 2776 | | 3.69 | 2.71 | 1.07e-135 | 0.05835 | 0.1148 | 1.63e-07 | -1.072 | 0.5839 | -0.4218 | 1.15e-188 | 8.15e-188 | -0.5138 | -0.111 | 0.359 | 6.10e-18 |
| FVC_ZPC | FVC ZPC | lung | | 485 | 550 | 1035 | | 6.94 | 6.151 | 0.1174 | 51.71 | 32.02 | 0.101 | -0.01695 | 0.07288 | 0.05559 | 0.7847 | 0.821 | 0.1991 | 0.09668 | -0.08952 | 0.2166 |

| all_labs | full_name | group | selected_unit | male_n | female_n | total_n | sex_specific_lab | raw_median_male_mean | raw_median_female_mean | raw_median_p.value | raw_variance_female_mean | raw_variance_male_mean | raw_variance_p.value | std_mean_diff | median_male_mean | median_female_mean | median_p.value | median_p.value.fdr | std_variance_diff | variance_female_mean | variance_male_mean | variance_p.value |
|---|---|---|---|---|---|---|---|---|---|---|---|---|---|---|---|---|---|---|---|---|---|---|
| GamGg_ | Gamma globulin in Serum or Plasma by electrophoresis | immune/inflammatory | g/dL | 2890 | 3670 | 6560 | | 1.195 | 1.164 | 0.006465 | 0.06395 | 0.08566 | 0.03371 | -0.06767 | -0.03692 | -0.1005 | 0.006897 | 0.009907 | -0.1778 | -0.2271 | -0.05793 | 6.23e-04 |
| GGT | Gamma glutamyl transferase activity in Serum or Plasma | liver | [iU]/L | 2733 | 2363 | 5096 | | 81.75 | 75.82 | 0.06413 | 4884.03 | 4174.75 | 0.401 | -0.07185 | -0.08256 | -0.1568 | 0.01084 | 0.01527 | 0.08532 | -0.01054 | -0.09574 | 0.04021 |
| Glob | Globulin | metabolic | g/dL | 715 | 1060 | 1775 | | 2.874 | 2.943 | 8.03e-04 | 0.06832 | 0.06495 | 0.6484 | 0.1677 | -0.1753 | -0.007887 | 6.18e-04 | 9.97e-04 | -0.01133 | 0.09655 | 0.1087 | 0.8805 |
| GluBed | Glucose [Mass/volume] in Blood by Automated test strip | metabolic | mg/dL | 13466 | 13645 | 27111 | | 138.60 | 131.66 | 1.89e-51 | 1864.33 | 1955.98 | 0.004343 | -0.1698 | 0.05087 | -0.1069 | 1.48e-44 | 5.41e-44 | -0.0633 | 0.02488 | 0.08603 | 2.85e-06 |
| Gluc | Glucose lab | metabolic | mg/dL | 26960 | 34448 | 61408 | | 110.86 | 103.24 | 4.83e-259 | 796.12 | 1033.35 | 2.91e-71 | -0.2918 | 0.0912 | -0.1787 | 8.88e-284 | 8.32e-283 | -0.1659 | -0.01266 | 0.1456 | 4.14e-82 |
| GluWB | Glucose [Mass/volume] in Blood | blood | mg/dL | 7936 | 6045 | 13981 | | 137.07 | 133.98 | 4.99e-06 | 1399.53 | 1305.94 | 0.04138 | -0.09361 | -0.001219 | -0.09025 | 4.94e-08 | 9.89e-08 | 0.01342 | 0.058 | 0.04486 | 0.5119 |
| GTT50g | GTT50g | metabolic | mg/dL | 6 | 2743 | 2749 | Sex specific lab | 116.40 | 117.90 | 0.9205 | | | | 0.5753 | -0.5333 | 0.01746 | 0.2897 | 0.3402 | | | | |

| all_labs | full_name | group | selected_unit | male_n | female_n | total_n | sex_specific_lab | raw_median_male_mean | raw_median_female_mean | raw_median_p.value | raw_variance_female_mean | raw_variance_male_mean | raw_variance_p.value | std_mean_diff | median_male_mean | median_female_mean | median_p.value | median_p.value.fdr | std_variance_diff | variance_female_mean | variance_male_mean | variance_p.value |
|---|---|---|---|---|---|---|---|---|---|---|---|---|---|---|---|---|---|---|---|---|---|---|
| Haptgl | Haptoglobin [Mass/volume] in Serum or Plasma | blood | mg/dL | 1550 | 1516 | 3066 | | 179.25 | 175.85 | 0.4099 | 3919.98 | 4599.16 | 0.2346 | 0.004707 | 0.05769 | 0.06265 | 0.8963 | 0.9165 | -0.1608 | 0.03605 | 0.187 | 0.02558 |
| HB_PRD | HB_PRD | lung | | 1215 | 1182 | 2397 | | 14.6 | 13.4 | 0 | 0 | 0 | | -1.618 | 0.6646 | -0.8897 | 0 | 0 | -0.02394 | -0.04447 | -0.02122 | 0.7756 |
| HBSA# | Hepatitis B virus surface Ab [Presence] in Serum | immune/inflammatory | 10*-3.[iU]/mL | 2001 | 2102 | 4103 | | 233.06 | 226.40 | 0.7791 | 280371.1 | 294245.4 | 0.9039 | -0.1028 | 0.2019 | 0.1095 | 9.76e-04 | 0.001549 | -0.1771 | -0.01929 | 0.1547 | 0.005016 |
| HCO3Ca | Bicarbonate [Moles/volume] in Unspecified specimen | metabolic | mmol/L | 9812 | 8404 | 18216 | | 24.93 | 24.71 | 8.83e-04 | 13.06 | 10.64 | 4.69e-12 | -0.04179 | 0.07965 | 0.04036 | 0.005242 | 0.007707 | 0.04355 | 0.1095 | 0.0671 | 0.0142 |
| HCTWB | Hematocrit | blood | % | 6990 | 5311 | 12301 | | 34.2 | 31.87 | 1.63e-96 | 17.85 | 19.36 | 0.005123 | -0.3549 | 0.1137 | -0.2217 | 4.07e-88 | 1.95e-87 | -0.09296 | 0.05657 | 0.1466 | 2.36e-05 |
| HDL-C | HDL Cholesterol | metabolic | mg/dL | 14828 | 18580 | 33408 | | 43.2 | 56.44 | 0 | 77.31 | 52.67 | 2.62e-91 | 0.8102 | -0.4579 | 0.3644 | 0 | 0 | 0.3451 | 0.5525 | 0.2541 | 1.37e-173 |
| HEART RATE | Heart rate | vitals | | 6318 | 7229 | 13547 | | 74.85 | 77.47 | 2.14e-21 | 179.66 | 184.46 | 0.4205 | 0.1543 | -0.1216 | 0.02144 | 4.10e-19 | 1.09e-18 | -0.03349 | -0.01672 | 0.01512 | 0.1182 |
| Height | Height | vitals | cm | 25698 | 34577 | 60275 | | 178.95 | 163.60 | 0 | | | | -1.424 | 0.831 | -0.5787 | 0 | 0 | -0.04557 | -0.04078 | 0.006135 | 1.95e-07 |
| HEIGHT | Height | vitals | cm | 12621 | 19052 | 31673 | | 178.56 | 163.48 | 0 | 2.65 | 3.92 | 1.85e-12 | -1.417 | 0.8019 | -0.6122 | 0 | 0 | 0.0342 | 0.141 | 0.1055 | 0.006093 |
| HEMATOCRIT | HEMATOCRIT | blood | % | 509 | 718 | 1227 | | 38.08 | 36.85 | 6.58e-04 | 9.761 | 12.87 | 0.04766 | -0.2377 | 0.1585 | -0.07212 | 8.01e-05 | 1.38e-04 | -0.2249 | 0.03414 | 0.2452 | 0.001624 |
| HEMOGLOBIN | HEMOGLOBIN | blood | g/dL | 509 | 719 | 1228 | | 12.55 | 11.97 | 7.12e-06 | 1.134 | 1.499 | 0.04132 | -0.3146 | 0.1874 | -0.1148 | 1.90e-07 | 3.74e-07 | -0.2549 | 0.04608 | 0.2852 | 3.30e-04 |
| Hgb | Hemoglobin | blood | g/dL | 26968 | 35678 | 62646 | | 13.2 | 12.47 | 0 | 1.564 | 2.214 | 8.62e-279 | -0.493 | 0.1926 | -0.272 | 0 | 0 | -0.2699 | 0.1037 | 0.356 | 1.06e-213 |
| HGB | Hemoglobin | blood | | 769 | 657 | 1426 | | 11.9 | 11.43 | 1.62e-05 | 0.9851 | 1.279 | 0.0145 | -0.2557 | 0.09083 | -0.1839 | 8.51e-07 | 1.62e-06 | -0.1474 | 0.02663 | 0.1719 | 0.0265 |
| HgbA1C | Hemoglobin A1c (Glycated) Hemoglobin A1c (Glycated) | metabolic | % | 11801 | 14409 | 26210 | | 6.234 | 5.97 | 2.51e-61 | 0.6391 | 0.7811 | 3.06e-06 | -0.1255 | -0.01187 | -0.1385 | 8.39e-24 | 2.38e-23 | -0.1645 | 0.02271 | 0.1797 | 8.64e-25 |
| HomocB | Homocysteine in Serum or Plasma | metabolic | umol/L | 538 | 829 | 1367 | | 12.2 | 10.78 | 5.49e-06 | 31 | 15.91 | 0.5189 | -0.3417 | 0.263 | -0.07403 | 4.51e-10 | 9.79e-10 | -0.5243 | -0.05101 | 0.4021 | 5.91e-04 |
| HORIZONTAL AXIS: INITIAL 40 MS | QRS axis | cardiovascular | | 6239 | 7139 | 13378 | | 37.92 | 32.67 | 6.59e-15 | 550.73 | 777.08 | 1.55e-06 | -0.1764 | 0.1459 | -0.04046 | 2.27e-24 | 6.66e-24 | 0.03829 | -0.01993 | -0.06135 | 0.07776 |
| HORIZONTAL AXIS: TERMINAL 40 MS | QRS axis | cardiovascular | | 6205 | 7074 | 13279 | | 57.76 | 20.99 | 3.82e-70 | 6908.29 | 7802.78 | 0.001947 | -0.1803 | 0.02764 | -0.1613 | 5.25e-25 | 1.56e-24 | 0.01253 | -0.08244 | -0.09544 | 0.5647 |
| HPIndx | H. pylori EIA index | immune/inflammatory | | 369 | 676 | 1045 | | 0.8497 | 0.7891 | 0.3671 | 0.3682 | 0.332 | 0.869 | 0.009789 | -0.2521 | -0.2424 | 0.8813 | 0.9031 | 0.01161 | -0.2422 | -0.2542 | 0.9597 |
| HR_INTERVAL | Heart rate | vitals | | 19279 | 21462 | 40741 | | 67.4 | 67.91 | 0.06214 | 633.26 | 573.85 | 8.24e-09 | 0.03191 | 0.2293 | 0.2567 | 0.001282 | 0.002023 | -0.02847 | 0.3319 | 0.3571 | 0.01684 |
| HSCRP | C reactive protein (high sensitivity) | immune/inflammatory | mg/L | 585 | 555 | 1140 | | 4.09 | 5.292 | 0.03178 | 72.27 | 61.21 | 0.8025 | 0.3548 | -0.1848 | 0.1789 | 1.57e-09 | 3.30e-09 | 0.2413 | 0.02654 | -0.2128 | 0.04697 |
| IC_PPR | IC PPR | lung | | 1168 | 1168 | 2336 | | 80.14 | 82.05 | 0.04345 | 147.66 | 146.40 | 0.963 | 0.04497 | -0.01212 | 0.03079 | 0.2773 | 0.3289 | 0.04746 | -0.00358 | -0.05238 | 0.5889 |
| IC_PRD | IC PRD | lung | | 1167 | 1168 | 2335 | | 3.299 | 2.34 | 0 | 0.00298 | 0.02834 | 8.92e-07 | -1.612 | 0.8641 | -0.6068 | 0 | 0 | -0.1995 | -0.1426 | 0.05738 | 0.02091 |
| IC_ZBD | IC ZBD | lung | | 1168 | 1167 | 2335 | | 2.634 | 1.924 | 2.84e-114 | 0.08349 | 0.1623 | 0.003825 | -0.9576 | 0.5345 | -0.3745 | 1.21e-133 | 6.86e-133 | -0.302 | -0.1265 | 0.1857 | 5.68e-04 |
| IctIdx | Icteric index of Serum or Plasma | metabolic | | 1967 | 2408 | 4375 | | 1.071 | 1.025 | 4.74e-13 | 0.01417 | 0.02988 | 3.28e-04 | -0.1835 | 0.2943 | 0.1312 | 1.79e-09 | 3.74e-09 | -0.00632 | 0.1207 | 0.1265 | 0.89 |
| IgA | IgA [Mass/volume] in Serum or Plasma | immune/inflammatory | mg/dL | 1980 | 4327 | 6307 | | 241.86 | 212.50 | 2.80e-13 | 1631.87 | 3745.14 | 0.08825 | -0.1751 | 0.1861 | 0.01519 | 6.96e-10 | 1.50e-09 | -0.02177 | -0.1996 | -0.1765 | 0.6327 |

| all_labs | full_name | group | selected_unit | male_n | female_n | total_n | sex_specific_lab | raw_median_male_mean | raw_median_female_mean | raw_median_p.value | raw_variance_female_mean | raw_variance_male_mean | raw_variance_p.value | std_mean_diff | median_male_mean | median_female_mean | median_p.value | median_p.value.fdr | std_variance_diff | variance_female_mean | variance_male_mean | variance_p.value |
|---|---|---|---|---|---|---|---|---|---|---|---|---|---|---|---|---|---|---|---|---|---|---|
| IGAB | Immature granulocytes [Presence] in Blood by Automated Count | immune/inflammatory | 10*3/uL | 19254 | 24902 | 44156 | | 0.03707 | 0.03188 | 3.01e-41 | 0.001842 | 0.002285 | 3.61e-07 | -0.1615 | -0.06326 | -0.2261 | 1.51e-63 | 6.29e-63 | -0.07142 | -0.08497 | -0.01027 | 2.65e-10 |
| IgEQnt | IgE [Mass/volume] in Serum | immune/inflammatory | [iU]/mL | 557 | 1008 | 1565 | | 246.11 | 126.89 | 3.84e-05 | 106749.5 | 105137.8 | 0.9814 | -0.3294 | 0.1382 | -0.1806 | 1.04e-09 | 2.22e-09 | -0.1096 | -0.1271 | -0.02029 | 0.3498 |
| IgG-Q | IgG Quantitative blood | immune/inflammatory | mg/dL | 1520 | 2071 | 3591 | | 997.43 | 1011.37 | 0.3517 | 56371.9 | 58656.3 | 0.7935 | 0.03565 | -0.05077 | -0.01819 | 0.2955 | 0.3456 | -0.0473 | -0.1537 | -0.1097 | 0.3742 |
| IgM-Q | IgM Quantitative blood | immune/inflammatory | mg/dL | 902 | 1272 | 2174 | | 119.74 | 126.55 | 0.3275 | 6139.14 | 11900.0 | 0.2324 | 0.2178 | 0.04103 | 0.2434 | 7.81e-07 | 1.49e-06 | 0.1616 | 0.05174 | -0.1235 | 0.06057 |
| IGRE | Immature granulocytes/100 leukocytes in Blood | immune/inflammatory | % | 19256 | 24933 | 44189 | | 0.409 | 0.353 | 9.86e-102 | 0.0767 | 0.09527 | 1.50e-21 | -0.2099 | -0.01335 | -0.2239 | 6.36e-106 | 3.23e-105 | -0.1124 | -0.01704 | 0.09689 | 2.35e-23 |
| Infant Apgar 5 M Hr 1 | 5 minute Apgar heart rate | vitals | | 0 | 1596 | 1596 | Sex specific lab | | | | | | | | | | | | | | | |
| Infant Apgar 5 M Hr 4 | 5 minute Apgar heart rate | vitals | | 0 | 1596 | 1596 | Sex specific lab | | | | | | | | | | | | | | | |
| Infant Apgar Color 1 M 1 | 1 minute Apgar color | vitals | | 0 | 1624 | 1624 | Sex specific lab | | | | | | | | | | | | | | | |
| Infant Apgar Color 5 M 1 | 5 minute Apgar color | vitals | | 0 | 1622 | 1622 | Sex specific lab | | | | | | | | | | | | | | | |
| Infant Apgar Muscle 1 M 1 | 1 minute Apgar muscle | vitals | | 0 | 1628 | 1628 | Sex specific lab | | | | | | | | | | | | | | | |
| Infant Apgar Muscle 5 M 1 | 5 minute Apgar muscle | vitals | | 0 | 1588 | 1588 | Sex specific lab | | | | | | | | | | | | | | | |
| Infant Apgar Resp 1 M 1 | 1 minute Apgar respiratory rate | vitals | | 0 | 1629 | 1629 | Sex specific lab | | | | | | | | | | | | | | | |
| Infant Apgar Resp 5 M 1 | 5 minute Apgar respiratory rate | vitals | | 0 | 1595 | 1595 | Sex specific lab | | | | | | | | | | | | | | | |
| Infant Apgar Rflx 1 M 1 | 1 minute Apgar reflex | vitals | | 0 | 1630 | 1630 | Sex specific lab | | | | | | | | | | | | | | | |
| Infant Apgar Rflx 5 M 1 | 5 minute Apgar reflex | vitals | | 0 | 1593 | 1593 | Sex specific lab | | | | | | | | | | | | | | | |
| Infant Apgar Score 1 M 1 | 1 minute Apgar score | vitals | | 0 | 1619 | 1619 | Sex specific lab | | | | | | | | | | | | | | | |
| Infant Apgar Score 5 M 1 | 5 minute Apgar score | vitals | | 0 | 1594 | 1594 | Sex specific lab | | | | | | | | | | | | | | | |
| INITIAL_40_MS | Initial 40 ms | cardiovascular | | 19244 | 21438 | 40682 | | 24.02 | 29.5 | 2.25e-74 | 459.10 | 592.23 | 1.13e-19 | 0.1572 | -0.1242 | 0.03392 | 5.55e-56 | 2.23e-55 | -0.08161 | 0.04767 | 0.1286 | 7.67e-12 |
| Insuln | Insulin [Units/volume] in Serum or Plasma | endocrine | | 331 | 1372 | 1703 | | 22.34 | 17.52 | 8.78e-04 | 319.36 | 589.82 | 0.06806 | -0.1824 | 0.1664 | -0.01813 | 0.005523 | 0.00805 | -0.4183 | 0.08153 | 0.4341 | 1.50e-04 |

| all_labs | full_name | group | selected_unit | male_n | female_n | total_n | sex_specific_lab | raw_median_male_mean | raw_median_female_mean | raw_median_p.value | raw_variance_female_mean | raw_variance_male_mean | raw_variance_p.value | std_mean_diff | median_male_mean | median_female_mean | median_p.value | median_p.value.fdr | std_variance_diff | variance_female_mean | variance_male_mean | variance_p.value |
|---|---|---|---|---|---|---|---|---|---|---|---|---|---|---|---|---|---|---|---|---|---|---|
| IPF | Platelets reticulated/100 platelets in Blood by Automated count | blood | % | 2273 | 2403 | 4676 | | 5.042 | 4.784 | 0.02139 | 6.255 | 7.637 | 0.1111 | -0.09116 | -0.003547 | -0.0938 | 0.001817 | 0.002822 | -0.1075 | 0.01537 | 0.1255 | 0.03679 |
| IRF | Immature reticulocytes/Reticulocytes.total in Blood | blood | % | 2822 | 3085 | 5907 | | 16.51 | 15.88 | 0.009566 | 48.84 | 47.21 | 0.6358 | -0.06104 | -0.02333 | -0.08281 | 0.01901 | 0.0261 | -0.03798 | -0.005591 | 0.0303 | 0.3771 |
| Iron | Iron [Mass/volume] in Serum or Plasma | blood | ug/dL | 7897 | 10737 | 18634 | | 73.15 | 68.69 | 2.03e-13 | 803.60 | 772.51 | 0.4682 | -0.1249 | 0.1141 | -0.00727 | 6.91e-17 | 1.73e-16 | -0.0105 | 0.3437 | 0.3532 | 0.5517 |
| K-WB | Potassium whole blood | metabolic | mmol/L | 8267 | 6320 | 14587 | | 4.118 | 3.908 | 8.65e-103 | 0.3466 | 0.3381 | 0.7079 | -0.4095 | 0.1391 | -0.2449 | 2.94e-133 | 1.64e-132 | -0.07081 | -8.71e-04 | 0.06467 | 3.71e-04 |
| K | Potassium blood | metabolic | mmol/L | 26975 | 34362 | 61337 | | 4.149 | 4.041 | 0 | 0.1875 | 0.1937 | 9.84e-06 | -0.2979 | 0.11 | -0.1522 | 5.44e-297 | 5.20e-296 | -0.03221 | 0.2067 | 0.233 | 1.85e-04 |
| KapLCQ | Kappa light chains.free [Mass/volume] in Serum | toxicology/pharmacology | mg/dL | 640 | 973 | 1613 | | 6.686 | 3.92 | 0.009438 | 244.78 | 337.07 | 0.6329 | -0.4361 | 0.1528 | -0.2703 | 1.09e-17 | 2.80e-17 | -0.02556 | -0.3312 | -0.3046 | 0.7808 |
| KLRat | Kappa light chains.free/Lambda light chains.free [Mass Ratio] in Serum | immune/inflammatory | | 617 | 949 | 1566 | | 4.047 | 2.228 | 0.2079 | 2675.30 | 3539.04 | 0.7961 | -0.09157 | -0.1586 | -0.2405 | 0.07887 | 0.1026 | -0.07433 | 0.04781 | 0.1204 | 0.421 |
| LA | Lactate [Moles/volume] in Serum or Plasma | metabolic | 10*-3.eq/L | 6279 | 6190 | 12469 | | 1.77 | 1.597 | 8.75e-20 | 1.145 | 1.291 | 0.03842 | -0.2189 | 0.06946 | -0.1292 | 1.58e-34 | 5.32e-34 | -0.05061 | 0.03248 | 0.07762 | 0.03678 |
| LAC | Lactate Whole Blood | metabolic | mmol/L | 8287 | 6477 | 14764 | | 1.485 | 1.462 | 0.2037 | 1.039 | 1.1 | 0.2093 | -0.1017 | -0.04492 | -0.1436 | 1.33e-09 | 2.81e-09 | -0.05719 | 8.62e-04 | 0.05426 | 0.003519 |
| LamLCQ | Lambda light chains.free [Mass/volume] in Serum or Plasma | immune/inflammatory | mg/dL | 639 | 971 | 1610 | | 5.159 | 3.041 | 0.00121 | 244.46 | 197.36 | 0.7572 | -0.4474 | 0.2811 | -0.1219 | 1.15e-18 | 3.03e-18 | -0.1321 | -0.02552 | 0.1034 | 0.1458 |
| LDH | LDH blood | metabolic | [iU]/L | 5881 | 6376 | 12257 | | 319.30 | 320.44 | 0.8243 | 54305.7 | 67002.8 | 0.08426 | 0.03737 | -0.4175 | -0.3833 | 0.03878 | 0.05181 | 0.003796 | -0.1704 | -0.1741 | 0.8716 |
| LDL_CALCULATED | LDL_CALCULATED | metabolic | mg/dL | 700 | 806 | 1506 | | 103.39 | 110.74 | 1.29e-05 | 519.03 | 360.35 | 0.02038 | 0.247 | -0.1835 | 0.06024 | 1.71e-06 | 3.20e-06 | 0.132 | 0.03795 | -0.09387 | 0.09217 |
| LDL_HDL_RATIO | Cholesterol.total/Cholesterol in HDL [Mass Ratio] in Serum or Plasma | metabolic | | 1029 | 1151 | 2180 | | 2.407 | 2.116 | 3.56e-14 | 0.2907 | 0.3277 | 0.3551 | -0.3325 | 0.1363 | -0.1913 | 7.24e-15 | 1.76e-14 | -0.07865 | -0.0546 | 0.02391 | 0.2162 |
| LDL-C | LDL cholesterol | metabolic | mg/dL | 14481 | 18095 | 32576 | | 95.24 | 105.29 | 5.58e-182 | 572.29 | 563.28 | 0.3237 | 0.3144 | -0.26 | 0.04115 | 1.78e-176 | 1.14e-175 | 0.03084 | 0.3046 | 0.277 | 0.01507 |
| LEUKOREDUCED_RANDOM_PLATELETS | LEUKOREDUCED_RANDOM_PLATELETS | immune/inflammatory | 10*3/uL | 1533 | 2351 | 3884 | | 212.91 | 246.85 | 2.17e-44 | 2040.39 | 1837.86 | 0.1706 | 0.4587 | -0.3452 | 0.148 | 3.73e-45 | 1.37e-44 | 0.2269 | 0.1743 | -0.05389 | 1.83e-07 |
| LH1 | Lutropin [Units/volume] in Serum or Plasma | endocrine | 10*-3.[iU]/mL | 861 | 1594 | 2455 | | 5.056 | 13.65 | 8.96e-101 | 109.18 | 14.05 | 1.60e-13 | 1.013 | -0.8155 | 0.3011 | 1.24e-154 | 7.41e-154 | 0.3679 | 0.1993 | -0.1743 | 7.00e-06 |
| Lipase | Lipase | toxicology/pharmacology | [iU]/L | 5058 | 6454 | 11512 | | 54.43 | 55.96 | 0.5543 | 54908.8 | 39972.8 | 0.2304 | 0.1262 | -0.3292 | -0.2069 | 2.31e-11 | 5.15e-11 | -0.03004 | -0.1366 | -0.1072 | 0.2698 |
| LipIdx | Lipemic index of Serum or Plasma | metabolic | | 2114 | 2762 | 4876 | | 15.73 | 14.71 | 4.72e-04 | 76.37 | 77.61 | 0.9053 | -0.1041 | 0.06503 | -0.03445 | 3.11e-04 | 5.14e-04 | -0.01417 | 0.002673 | 0.01561 | 0.7359 |

| all_labs | full_name | group | selected_unit | male_n | female_n | total_n | sex_specific_lab | raw_median_male_mean | raw_median_female_mean | raw_median_p.value | raw_variance_female_mean | raw_variance_male_mean | raw_variance_p.value | std_mean_diff | median_male_mean | median_female_mean | median_p.value | median_p.value.fdr | std_variance_diff | variance_female_mean | variance_male_mean | variance_p.value |
|---|---|---|---|---|---|---|---|---|---|---|---|---|---|---|---|---|---|---|---|---|---|---|
| Luc% | Luc% | immune/inflammatory | % | 2510 | 3541 | 6051 | | 2.364 | 2.089 | 4.94e-13 | 0.9249 | 1.031 | 0.4415 | -0.1817 | 0.1838 | 0.007559 | 4.75e-12 | 1.08e-11 | -0.02041 | 0.009679 | 0.02929 | 0.5674 |
| LucAbs | Leukocytes [#/volume] in Blood by Automated count | blood | 10*3/uL | 2514 | 3547 | 6061 | | 0.245 | 0.205 | 0.09838 | 0.3637 | 0.2048 | 0.611 | -0.06523 | 0.07222 | 0.01254 | 0.01342 | 0.01874 | 0.08637 | 0.1082 | 0.03496 | 0.01438 |
| Lym | Lymphocytes [#/volume] in Blood | immune/inflammatory | % | 11637 | 15914 | 27551 | | 23.59 | 25.94 | 8.16e-85 | 63.99 | 59.1 | 6.66e-05 | 0.2224 | -0.04221 | 0.1536 | 9.59e-74 | 4.37e-73 | 0.009234 | 0.03803 | 0.02985 | 0.5338 |
| LymAbs | Lymphocytes [#/volume] in Blood by Automated count | immune/inflammatory | 10*3/uL | 11025 | 15127 | 26152 | | 1.754 | 1.927 | 7.24e-38 | 0.4437 | 0.7253 | 0.04535 | 0.2257 | -0.0946 | 0.1167 | 8.96e-73 | 3.94e-72 | 0.1413 | -0.02641 | -0.1602 | 1.25e-19 |
| LYMPA | Lymphocytes [#/volume] in Blood | immune/inflammatory | 10*3/uL | 5327 | 5177 | 10504 | | 1.401 | 1.537 | 0.002398 | 2.565 | 3.984 | 0.05354 | 0.1972 | -0.1926 | -0.00672 | 3.97e-24 | 1.15e-23 | 0.1069 | -0.07534 | -0.1784 | 3.67e-05 |
| LYMPAB | Lymphocytes [#/volume] in Blood by Automated count | immune/inflammatory | 10*3/uL | 19927 | 25616 | 45543 | | 1.653 | 1.872 | 6.18e-191 | 0.302 | 0.2725 | 3.77e-07 | 0.2413 | -0.216 | 0.02214 | 6.42e-145 | 3.75e-144 | 0.02345 | 0.1525 | 0.1323 | 0.03089 |
| LYMPHOCYTES | LYMPHOCYTES | immune/inflammatory | % | 795 | 1237 | 2032 | | 19.37 | 22.21 | 3.00e-10 | 56.91 | 38.49 | 1.78e-05 | 0.2471 | -0.3492 | -0.103 | 3.95e-08 | 7.94e-08 | 0.256 | 0.2325 | 0.003034 | 7.67e-05 |
| LYMPM | Lymphocytes/100 leukocytes in Blood by Manual count | immune/inflammatory | % | 5485 | 5362 | 10847 | | 16.93 | 19.31 | 7.42e-17 | 157.42 | 145.18 | 0.08594 | 0.1839 | -0.1679 | 0.01081 | 8.09e-22 | 2.22e-21 | 0.04331 | 0.07146 | 0.03115 | 0.08999 |
| LYMPRE | Lymphocytes/100 leukocytes in Blood by Automated count | immune/inflammatory | % | 19940 | 25619 | 45559 | | 22.23 | 25.02 | 2.03e-205 | 59.21 | 53.9 | 1.90e-12 | 0.2523 | -0.1606 | 0.07061 | 1.90e-157 | 1.17e-156 | 0.01365 | 0.1518 | 0.1402 | 0.2078 |
| MBRat | Creatine kinase.MB/Creatine kinase.total in Serum or Plasma | cardiovascular | | 7881 | 6477 | 14358 | | 3.6 | 3.698 | 0.06295 | 6.187 | 6.875 | 0.416 | 0.06283 | 0.03731 | 0.09942 | 1.70e-04 | 2.88e-04 | -0.07027 | 0.04919 | 0.1209 | 7.49e-04 |
| MCH | MCH [Entitic mass] by Automated count | blood | pg | 26968 | 35668 | 62636 | | 30.21 | 29.73 | 1.86e-188 | 1.536 | 1.462 | 0.001278 | -0.2226 | 0.1593 | -0.05239 | 5.48e-167 | 3.40e-166 | 0.02871 | 0.3759 | 0.349 | 8.15e-04 |
| MCHC | MCHC [Mass/volume] by Automated count | blood | g/dL | 26945 | 35644 | 62589 | | 33.35 | 32.83 | 0 | 0.7789 | 0.8375 | 6.81e-18 | -0.5637 | 0.2471 | -0.2483 | 0 | 0 | -0.1015 | 0.2201 | 0.3073 | 2.70e-32 |
| MCV | MCV [Entitic volume] by Automated count | blood | fL | 26971 | 35676 | 62647 | | 1803679.2 | 90.46 | 0.3173 | | | | 0.2372 | -0.4123 | -0.1855 | 1.13e-193 | 8.37e-193 | -0.2473 | 0.1167 | 0.3498 | 8.09e-187 |
| MDALab | Atypical lymphocytes | immune/inflammatory | | 3552 | 3337 | 6889 | | 0.2357 | 0.2139 | 0.003761 | 0.08042 | 0.09426 | 0.341 | -0.09814 | 0.01524 | -0.07156 | 4.66e-05 | 8.18e-05 | -0.104 | -0.05772 | 0.04461 | 0.003992 |
| MDBlab | Blasts in Blood | immune/inflammatory | | 847 | 743 | 1590 | | 0.6779 | 0.7948 | 0.5059 | 33.22 | 31.7 | 0.8504 | 0.06825 | -0.1538 | -0.08606 | 0.1732 | 0.211 | 0.0183 | 0.02814 | 0.01121 | 0.7993 |
| MDMMab | Metamyelocytes (absolute) | immune/inflammatory | 10*3/uL | 3619 | 3474 | 7093 | | 0.3004 | 0.2936 | 0.6506 | 0.5398 | 0.5593 | 0.8432 | 0.02343 | -0.04044 | -0.01877 | 0.3239 | 0.3753 | -0.0835 | -0.02921 | 0.04566 | 0.008654 |
| MDMyab | Myelocytes (absolute) | immune/inflammatory | 10*3/uL | 3884 | 3632 | 7516 | | 0.2963 | 0.2899 | 0.7465 | 0.7776 | 0.8901 | 0.5269 | 0.04807 | -0.03607 | 0.008329 | 0.03736 | 0.0503 | -0.1036 | -5.47e-04 | 0.09214 | 7.03e-04 |
| MDPmab | Promyelocytes (absolute) | immune/inflammatory | | 1490 | 1364 | 2854 | | 0.2705 | 0.2854 | 0.6153 | 1.481 | 1.704 | 0.5814 | -0.00898 | 0.006698 | -0.00112 | 0.8109 | 0.8467 | -0.09158 | 0.0785 | 0.1636 | 0.09638 |

| all_labs | full_name | group | selected_unit | male_n | female_n | total_n | sex_specific_lab | raw_median_male_mean | raw_median_female_mean | raw_median_p.value | raw_variance_female_mean | raw_variance_male_mean | raw_variance_p.value | std_mean_diff | median_male_mean | median_female_mean | median_p.value | median_p.value.fdr | std_variance_diff | variance_female_mean | variance_male_mean | variance_p.value |
|---|---|---|---|---|---|---|---|---|---|---|---|---|---|---|---|---|---|---|---|---|---|---|
| MEAN_QRS | QRS axis | cardiovascular | deg | 19258 | 21442 | 40700 | | 20.7 | 26.74 | 4.02e-50 | 531.63 | 805.19 | 4.90e-28 | 0.1011 | -0.09705 | 0.008427 | 5.21e-24 | 1.50e-23 | -0.08936 | 0.02843 | 0.118 | 6.55e-14 |
| Meta | Metamyelocytes/100 leukocytes in Blood by Manual count | blood | % | 3764 | 3623 | 7387 | | 2.062 | 2.079 | 0.6822 | 4.026 | 3.873 | 0.4968 | 0.06662 | -0.1052 | -0.03905 | 0.004162 | 0.006249 | 0.007101 | -0.02056 | -0.02729 | 0.8189 |
| MetHgb | Methemoglobin/Hemoglobin.total in Venous blood | blood | % | 4068 | 2965 | 7033 | | 0.9555 | 0.946 | 0.5023 | 0.1357 | 0.1386 | 0.6938 | -0.04161 | 0.151 | 0.1058 | 0.08672 | 0.1114 | -0.01998 | 0.1418 | 0.1625 | 0.5128 |
| Mg | Magnesium [Mass/volume] in Serum or Plasma | metabolic | mg/dL | 11442 | 12679 | 24121 | | 2.026 | 1.998 | 2.78e-17 | 0.07391 | 0.07362 | 0.8579 | -0.09371 | 0.1215 | 0.03378 | 3.60e-13 | 8.40e-13 | -0.07205 | -0.07146 | -0.006599 | 3.82e-06 |
| MmAcid | Methylmalonic acid | metabolic | umol/L | 689 | 902 | 1591 | | 0.3023 | 0.2718 | 0.3938 | 0.09054 | 0.04611 | 0.5582 | 0.05994 | 0.05639 | 0.1178 | 0.24 | 0.286 | -0.3147 | -0.05993 | 0.2456 | 0.006498 |
| MonAbs | MonAbs | immune/inflammatory | 10*3/uL | 11006 | 15125 | 26131 | | 0.6288 | 0.5519 | 3.03e-117 | 0.0516 | 0.07741 | 1.70e-12 | -0.2994 | 0.2536 | -0.01273 | 1.87e-125 | 9.99e-125 | -0.1879 | -0.0733 | 0.1007 | 4.57e-33 |
| MONOA | Monocytes [#/volume] in Blood | immune/inflammatory | 10*3/uL | 5259 | 5055 | 10314 | | 0.663 | 0.5777 | 7.52e-12 | 0.4775 | 0.6026 | 0.1211 | -0.14 | 0.005143 | -0.1277 | 1.07e-12 | 2.45e-12 | -0.03784 | -0.03714 | -0.00163 | 0.1477 |
| MONOAB | Monocytes [#/volume] in Blood by Automated count | immune/inflammatory | 10*3/uL | 19931 | 25647 | 45578 | | 0.6886 | 0.6049 | 9.29e-236 | 0.04806 | 0.06659 | 7.29e-42 | -0.3175 | 0.1573 | -0.1432 | 2.63e-249 | 2.14e-248 | -0.2087 | 0.06341 | 0.2599 | 7.36e-81 |
| Monocy | Monocytes [#/volume] in Blood by Manual count | immune/inflammatory | % | 11630 | 15947 | 27577 | | 8.195 | 7.281 | 5.77e-191 | 5.489 | 6.87 | 1.09e-22 | -0.3482 | 0.3967 | 0.07832 | 1.14e-179 | 7.36e-179 | -0.1558 | -0.0513 | 0.09717 | 4.75e-25 |
| MONOCYTES | MONOCYTES | immune/inflammatory | % | 794 | 1238 | 2032 | | 8.618 | 7.8 | 2.20e-11 | 5.337 | 4.704 | 0.189 | -0.2783 | 0.1526 | -0.1435 | 1.06e-09 | 2.25e-09 | 0.1724 | 0.1809 | 0.00312 | 0.009484 |
| MONOM | Monocytes/100 leukocytes in Blood by Manual count | immune/inflammatory | % | 5320 | 5096 | 10416 | | 7.101 | 6.456 | 4.73e-14 | 21.74 | 24.39 | 0.01358 | -0.1413 | 0.03613 | -0.103 | 5.18e-13 | 1.20e-12 | -0.1016 | -0.04446 | 0.04832 | 9.73e-05 |
| MONORE | Monocytes/100 leukocytes in Blood by Automated count | immune/inflammatory | % | 19994 | 25672 | 45666 | | 8.758 | 7.789 | 0 | 4.634 | 5.703 | 1.35e-44 | -0.3831 | 0.2947 | -0.08012 | 0 | 0 | -0.1721 | 0.0651 | 0.2344 | 6.69e-56 |
| MPV | Platelet mean volume [Entitic volume] in Blood by Automated count Platelet mean volume [Entitic volume] in Blood by Automated count | blood | fL | 19784 | 26726 | 46510 | | 10.37 | 10.4 | 0.01169 | 0.2494 | 0.2649 | 0.4665 | 0.02412 | -0.06286 | -0.03941 | 0.0101 | 0.0143 | -0.05153 | 0.172 | 0.2179 | 3.27e-07 |
| Myelo | Myelocytes/100 leukocytes in Blood by Manual count | immune/inflammatory | % | 3995 | 3755 | 7750 | | 1.943 | 1.997 | 0.1213 | 3.254 | 3.652 | 0.04459 | 0.06478 | -0.04951 | 0.01566 | 0.004352 | 0.006516 | -0.01117 | -0.01372 | -0.003007 | 0.7094 |
| Na-WB | Sodium in Blood | metabolic | mmol/L | 7792 | 5957 | 13749 | | 135.70 | 136.10 | 2.47e-10 | 8.078 | 7.456 | 0.08707 | 0.1314 | -0.09921 | 0.02441 | 3.16e-14 | 7.56e-14 | -0.02397 | 0.0925 | 0.1147 | 0.2469 |
| Na | Sodium in Blood | metabolic | mmol/L | 26971 | 34356 | 61327 | | 138.62 | 138.89 | 4.30e-45 | 6.766 | 7.656 | 3.02e-31 | 0.09161 | -0.08022 | 0.003278 | 3.76e-29 | 1.19e-28 | -0.09998 | 0.1551 | 0.2423 | 7.59e-31 |
| NasIO2 | NasIO2 | blood | L | 1748 | 1278 | 3026 | | 327.75 | 3343.63 | 0.1896 | 21536373.3 | 3893877.5 | 0.3338 | 0.0914 | -0.1774 | -0.09059 | 0.01385 | 0.01928 | 0.02572 | -0.05445 | -0.07742 | 0.5875 |
| Neut | Neutrophils [#/volume] in Blood | immune/inflammatory | % | 11786 | 16047 | 27833 | | 64.52 | 63.55 | 9.52e-13 | 94.62 | 94.66 | 0.9822 | -0.08104 | -0.04257 | -0.113 | 3.54e-11 | 7.86e-11 | -0.04939 | -0.008956 | 0.03509 | 8.24e-04 |

| all_labs | full_name | group | selected_unit | male_n | female_n | total_n | sex_specific_lab | raw_median_male_mean | raw_median_female_mean | raw_median_p.value | raw_variance_female_mean | raw_variance_male_mean | raw_variance_p.value | std_mean_diff | median_male_mean | median_female_mean | median_p.value | median_p.value.fdr | std_variance_diff | variance_female_mean | variance_male_mean | variance_p.value |
|---|---|---|---|---|---|---|---|---|---|---|---|---|---|---|---|---|---|---|---|---|---|---|
| NEUTA | Neutrophils absolute | immune/inflammatory | 10*3/uL | 5349 | 5142 | 10491 | | 7.796 | 7.606 | 0.2386 | 47.61 | 40.29 | 0.4091 | -0.02601 | -0.005373 | -0.0298 | 0.1829 | 0.2212 | -0.07389 | 0.01419 | 0.08127 | 0.004246 |
| NEUTAB | Neutrophils absolute automated count | immune/inflammatory | 10*3/uL | 19958 | 25593 | 45551 | | 5.378 | 5.239 | 1.92e-08 | 5.426 | 6.252 | 6.55e-15 | -0.0528 | -0.0201 | -0.06761 | 2.39e-08 | 4.88e-08 | -0.115 | 0.1287 | 0.2305 | 6.18e-26 |
| NEUTM | Polymorphonuclear cells/100 leukocytes in Body fluid by Manual count | immune/inflammatory | % | 5352 | 5147 | 10499 | | 69.73 | 68.48 | 5.47e-04 | 224.68 | 219.08 | 0.5296 | -0.0733 | 0.0619 | -0.008399 | 1.72e-04 | 2.90e-04 | -0.004229 | 0.03231 | 0.03615 | 0.8705 |
| NEUTRE | Neutrophils/100 leukocytes in Blood by Automated count | immune/inflammatory | % | 20000 | 25683 | 45683 | | 65.4 | 63.99 | 2.31e-43 | 87.26 | 86.05 | 0.2779 | -0.108 | 0.06311 | -0.03537 | 3.78e-30 | 1.22e-29 | -0.04222 | 0.1309 | 0.1672 | 9.89e-05 |
| NonHDL | Cholesterol non HDL [Mass/volume] in Serum or Plasma | metabolic | mg/dL | 2171 | 1846 | 4017 | | 118.03 | 132.18 | 7.01e-29 | 887.44 | 737.50 | 0.02558 | 0.336 | -0.149 | 0.169 | 1.39e-26 | 4.23e-26 | 0.08589 | 0.05677 | -0.02265 | 0.05175 |
| NRBC | NRBC | blood | /100.{WBC} | 12556 | 16758 | 29314 | | 0.3613 | 0.2578 | 1.81e-14 | 0.9203 | 1.18 | 0.03853 | -0.008484 | -0.1176 | -0.1278 | 0.4789 | 0.5296 | -0.1142 | -0.03501 | 0.08494 | 4.04e-18 |
| NRBC# | NRBC # | blood | 10*3/uL | 23718 | 31113 | 54831 | | 0.00206 | 0.001273 | 0.01491 | 0.001346 | 0.002132 | 7.59e-04 | 0.1978 | -0.4594 | -0.2678 | 1.50e-117 | 7.85e-117 | 0.1644 | -0.1645 | -0.327 | 2.68e-68 |
| NRBCBC | Nucleated erythrocytes/100 leukocytes [Ratio] in Cerebral spinal fluid by Manual count | blood | /100.{WBC} | 1153 | 1048 | 2201 | | 2.221 | 2.196 | 0.9033 | 40.55 | 35.62 | 0.7778 | -0.01691 | 0.01926 | 0.003819 | 0.6923 | 0.7399 | -0.1208 | 0.01278 | 0.1249 | 0.0507 |
| NRBCMA | Nucleated erythrocytes [#/volume] in Blood by Automated count | blood | 10*3/mL | 1142 | 1045 | 2187 | | 21.5 | 25.28 | 0.1734 | 2520.49 | 2084.81 | 0.5922 | 0.01764 | 0.03111 | 0.04821 | 0.6807 | 0.7293 | 0.03164 | 0.06065 | 0.03163 | 0.6128 |
| Nt-PTH | NT PTH | endocrine | pg/mL | 2958 | 5143 | 8101 | | 98.98 | 74.48 | 1.62e-26 | 3988.70 | 6908.09 | 3.99e-06 | -0.2139 | 0.0353 | -0.1755 | 3.83e-19 | 1.02e-18 | -0.2341 | 0.07275 | 0.283 | 1.67e-14 |
| NtAbs | NtAbs | immune/inflammatory | 10*3/uL | 11008 | 15125 | 26133 | | 5.294 | 5.162 | 2.07e-04 | 6.44 | 7.632 | 2.63e-05 | -0.0462 | -0.07021 | -0.1101 | 2.46e-04 | 4.12e-04 | -0.1257 | -0.06122 | 0.048 | 6.38e-16 |
| NTAuto | Neutrophils | immune/inflammatory | 10*3/uL | 21662 | 27996 | 49658 | | 5.365 | 5.208 | 8.80e-11 | 6.055 | 7.242 | 2.56e-23 | -0.04902 | -0.0525 | -0.09566 | 7.03e-08 | 1.40e-07 | -0.1188 | 0.2126 | 0.3105 | 4.35e-31 |
| O2 Liter Flow | O2 liter flow | vitals | L/min | 9654 | 9833 | 19487 | | 2.755 | 3.138 | 1.16e-16 | 13.15 | 57.44 | 0.3079 | -0.0452 | 0.07693 | 0.03782 | 0.001582 | 0.00248 | 0.01084 | -0.0125 | -0.02349 | 0.4677 |
| O2Sat | Oxygen saturation Calculated from oxygen partial pressure in Arterial blood | vitals | % | 6419 | 4695 | 11114 | | 97.47 | 97.29 | 0.05819 | 18.41 | 16.75 | 0.3195 | -0.00166 | -0.08149 | -0.08304 | 0.9313 | 0.9407 | -0.05358 | 0.09781 | 0.1487 | 0.01896 |
| O2SAT | Oxygen saturation calculated from oxygen partial pressure in arterial blood | vitals | | 23729 | 30062 | 53791 | | 96.79 | 97.33 | 3.53e-234 | 266501977955.0 | 8407808251.5 | 0.1737 | 0.2682 | -0.2235 | 0.04193 | 2.33e-212 | 1.84e-211 | 0.2029 | -0.149 | -0.3575 | 2.57e-114 |

| all_labs | full_name | group | selected_unit | male_n | female_n | total_n | sex_specific_lab | raw_median_male_mean | raw_median_female_mean | raw_median_p.value | raw_variance_female_mean | raw_variance_male_mean | raw_variance_p.value | std_mean_diff | median_male_mean | median_female_mean | median_p.value | median_p.value.fdr | std_variance_diff | variance_female_mean | variance_male_mean | variance_p.value |
|---|---|---|---|---|---|---|---|---|---|---|---|---|---|---|---|---|---|---|---|---|---|---|
| O2SatC | O2SatC | vitals | % | 4606 | 3797 | 8403 | | 98.24 | 97.87 | 5.94e-06 | 11.13 | 9.438 | 0.04241 | -0.00402 | 0.1848 | 0.1812 | 0.8559 | 0.8826 | -0.05113 | -0.08761 | -0.0436 | 0.04728 |
| O2SatV | Oxygen saturation Calculated from oxygen partial pressure in Venous blood | vitals | % | 4535 | 3787 | 8322 | | 70.53 | 71 | 0.2398 | 245.30 | 259.89 | 0.1879 | 0.02284 | -0.02885 | -0.006374 | 0.3021 | 0.3525 | -0.0199 | 0.05809 | 0.07829 | 0.5195 |
| OB Infant Apgar Score 1 M 1 | 1 minute Apgar score | ob/gyn | | 0 | 1217 | 1217 | Sex specific lab | | | | | | | | | | | | | | | |
| OB Infant Apgar Score 5 M 1 | 5 minute Apgar score | ob/gyn | | 0 | 1217 | 1217 | Sex specific lab | | | | | | | | | | | | | | | |
| Osmol | Osmolality of Serum or Plasma | blood | mosm/kg{H20} | 1755 | 1780 | 3535 | | 292.15 | 288.42 | 2.88e-07 | 169.35 | 191.51 | 0.3609 | -0.1691 | -0.08602 | -0.2493 | 4.78e-07 | 9.25e-07 | 0.01039 | -0.004524 | -0.01532 | 0.8607 |
| P DURATION | P wave duration | cardiovascular | | 6307 | 7220 | 13527 | | 117.05 | 113.05 | 1.22e-36 | 335.53 | 517.18 | 2.50e-31 | -0.2378 | 0.1194 | -0.1309 | 5.55e-43 | 1.94e-42 | -0.1837 | -0.1319 | 0.06894 | 9.30e-18 |
| P HORIZONTAL AXIS | P wave axis horizontal plane | cardiovascular | | 5960 | 6926 | 12886 | | 9.548 | 8.103 | 0.01012 | 915.29 | 1384.24 | 1.77e-15 | -0.07623 | 0.03543 | -0.04517 | 1.89e-05 | 3.41e-05 | -0.1076 | -0.09432 | 0.02442 | 1.55e-06 |
| P_WAVE | P wave axis | cardiovascular | deg | 19267 | 21451 | 40718 | | 38.8 | 43.98 | 1.94e-100 | 478.42 | 526.16 | 1.63e-06 | 0.2014 | -0.1556 | 0.04221 | 5.97e-92 | 2.91e-91 | -0.04589 | 0.05694 | 0.1022 | 1.18e-04 |
| Pat-PT | Prothrombin time | blood | s | 18382 | 19238 | 37620 | | 15.49 | 14.91 | 1.97e-58 | 12.59 | 12.7 | 0.7107 | -0.2127 | 0.1701 | -0.05256 | 3.77e-95 | 1.86e-94 | 0.001502 | 0.05666 | 0.05509 | 0.9032 |
| Pat-TT | Thrombin time | blood | s | 419 | 833 | 1252 | | 17.37 | 17.14 | 0.1916 | 4.188 | 9.018 | 0.3298 | -0.00168 | -0.1024 | -0.1041 | 0.9783 | 0.9803 | 0.3273 | 0.1123 | -0.29 | 0.004448 |
| pC02-V | Carbon dioxide [Partial pressure] in Venous blood | blood | mm[Hg] | 4502 | 4142 | 8644 | | 47.17 | 47.21 | 0.8777 | 83.55 | 80.21 | 0.4834 | 0.001374 | 0.02305 | 0.02441 | 0.9492 | 0.955 | 0.03108 | 0.07451 | 0.04434 | 0.349 |
| PCAct | Protein C [Units/volume] in Platelet poor plasma by Chromogenic method | blood | % | 352 | 656 | 1008 | | 103.05 | 111.02 | 4.36e-05 | 193.95 | 356.13 | 0.2698 | 0.2892 | -0.136 | 0.1689 | 7.43e-06 | 1.36e-05 | -0.1784 | 0.08945 | 0.2661 | 0.3531 |
| pCO2 | Carbon dioxide [Partial pressure] in Arterial blood | blood | mm[Hg] | 8663 | 7132 | 15795 | | 42.1 | 41.57 | 3.26e-05 | 48.64 | 45.47 | 0.0221 | -0.1418 | 0.1019 | -0.03165 | 2.24e-18 | 5.83e-18 | -0.05023 | 0.04105 | 0.08781 | 0.007641 |
| PCR | Protein/Creatinine [Mass ratio] in Urine | kidney | g/g | 2909 | 2818 | 5727 | | 0.9375 | 0.8435 | 0.3846 | 2.754 | 21.43 | 0.1095 | -0.04542 | -0.0984 | -0.1435 | 0.08537 | 0.1105 | 0.1208 | -0.08321 | -0.2019 | 8.03e-06 |
| PCV | Hematocrit [Volume Fraction] of Blood by Automated count | blood | % | 27330 | 36195 | 63525 | | 39.41 | 37.81 | 8.40e-289 | 14.15 | 18.86 | 9.76e-215 | -0.4053 | 0.1412 | -0.2374 | 0 | 0 | -0.2207 | 0.1028 | 0.3085 | 3.36e-147 |
| PEFR_AFD | PEFR_AFD | lung | | 506 | 587 | 1093 | | 6.628 | 5.07 | 2.84e-32 | 0.51 | 0.6593 | 0.2604 | -0.8635 | 0.4167 | -0.4404 | 3.53e-49 | 1.35e-48 | -0.2201 | 0.02472 | 0.1997 | 0.1645 |
| PEFR_APP | PEFR_APP | lung | | 506 | 587 | 1093 | | 76.44 | 82.5 | 8.78e-05 | 145.19 | 79.37 | 0.02025 | 0.1693 | -0.1834 | -0.01161 | 0.004921 | 0.007279 | 0.4003 | 0.1495 | -0.2261 | 0.009705 |
| PEFR_PPR | PEFR_PPR | lung | | 1604 | 1735 | 3339 | | 83.17 | 87.22 | 1.25e-05 | 147.83 | 142.28 | 0.7241 | 0.1472 | -0.1412 | 0.009227 | 2.09e-05 | 3.76e-05 | 0.00369 | -0.07085 | -0.07465 | 0.9506 |
| PEFR_PRD | PEFR_PRD | lung | | 1604 | 1736 | 3340 | | 8.682 | 6.097 | 0 | 0.01665 | 0.04952 | 9.57e-05 | -1.616 | 0.8273 | -0.6735 | 0 | 0 | -0.1411 | -0.0702 | 0.08038 | 0.01938 |
| PEFR_PRL | PEFR_PRL | lung | | 1362 | 1540 | 2902 | | 6.399 | 4.424 | 0 | 0.004649 | 0.01204 | 7.54e-06 | -1.645 | 0.8411 | -0.5723 | 0 | 0 | -0.3463 | -0.02187 | 0.3197 | 3.23e-08 |
| PEFR_ZBD | PEFR_ZBD | lung | | 1604 | 1736 | 3340 | | 7.203 | 5.31 | 1.85e-129 | 0.5504 | 1.058 | 1.77e-08 | -0.8779 | 0.4266 | -0.4518 | 9.53e-153 | 5.64e-152 | -0.5221 | -0.1927 | 0.2873 | 5.45e-19 |

| all_labs | full_name | group | selected_unit | male_n | female_n | total_n | sex_specific_lab | raw_median_male_mean | raw_median_female_mean | raw_median_p.value | raw_variance_female_mean | raw_variance_male_mean | raw_variance_p.value | std_mean_diff | median_male_mean | median_female_mean | median_p.value | median_p.value.fdr | std_variance_diff | variance_female_mean | variance_male_mean | variance_p.value |
|---|---|---|---|---|---|---|---|---|---|---|---|---|---|---|---|---|---|---|---|---|---|---|
| PEFR_ZPC | PEFR_ZPD | lung | | 491 | 569 | 1060 | | 4.281 | 3.06 | 0.1946 | 170.85 | 178.23 | 0.8467 | -0.06162 | -0.0106 | -0.07245 | 0.3181 | 0.3693 | -0.03372 | -0.03337 | -0.00285 | 0.8346 |
| pH-Ven | Venous pH | blood | | 4714 | 4438 | 9152 | | 7.349 | 7.346 | 0.0622 | 0.004588 | 0.004004 | 0.01818 | -0.02893 | 0.002109 | -0.02589 | 0.1667 | 0.2041 | 0.02452 | 0.146 | 0.123 | 0.3808 |
| Phos | Phosphate | metabolic | mg/dL | 9880 | 10326 | 20206 | | 3.502 | 3.537 | 0.002573 | 0.6866 | 0.7845 | 7.19e-08 | 0.07745 | 0.008255 | 0.07882 | 3.82e-08 | 7.71e-08 | -0.1169 | -0.07524 | 0.03554 | 6.66e-12 |
| PIFR_AFD | PIFR_AFD | lung | | 506 | 586 | 1092 | | 4.86 | 3.667 | 1.17e-35 | 0.4455 | 0.7333 | 0.02915 | -0.8192 | 0.479 | -0.305 | 1.95e-44 | 7.04e-44 | -0.503 | -0.1164 | 0.3325 | 0.001195 |
| PIFR_APP_APP | PIFR_APP_APP | lung | | 506 | 587 | 1093 | | 82.8 | 89.2 | 1.37e-04 | 266.01 | 210.42 | 0.3714 | 0.2189 | -0.1058 | 0.1089 | 2.84e-04 | 4.73e-04 | 0.182 | 0.06598 | -0.1061 | 0.2439 |
| PIFR_PPR | PIFR_PPR | lung | | 1604 | 1734 | 3338 | | 78.56 | 86.79 | 3.20e-17 | 256.61 | 252.46 | 0.8399 | 0.2973 | -0.1562 | 0.14 | 6.02e-18 | 1.55e-17 | 0.05904 | -0.05505 | -0.1136 | 0.3231 |
| PIFR_PRD | PIFR_PRD | lung | | 1604 | 1735 | 3339 | | 5.838 | 4.089 | 0 | 0.008134 | 0.03077 | 5.27e-05 | -1.614 | 0.8505 | -0.6668 | 0 | 0 | -0.1013 | -0.09013 | 0.01839 | 0.09466 |
| PIFR_ZBD | PIFR_ZBD | lung | | 1604 | 1735 | 3339 | | 4.581 | 3.54 | 6.01e-83 | 0.4333 | 0.8301 | 8.08e-14 | -0.7 | 0.3869 | -0.2938 | 2.91e-95 | 1.45e-94 | -0.5014 | -0.2011 | 0.2413 | 2.52e-17 |
| PIFR_ZPC | PIFR_ZPC | lung | | 488 | 570 | 1058 | | 6.474 | 5.09 | 0.2721 | 474.06 | 500.06 | 0.87 | -0.02964 | -0.01512 | -0.0424 | 0.6303 | 0.6795 | 0.112 | 0.08618 | -0.01823 | 0.491 |
| Plt-Ct | Platelet Count | blood | 10*3/uL | 26981 | 35703 | 62684 | | 222.04 | 257.80 | 0 | 4010.14 | 4828.02 | 7.17e-05 | 0.4599 | -0.2972 | 0.1626 | 0 | 0 | -0.05864 | 0.2395 | 0.292 | 1.16e-11 |
| pO2_FI | Oxygen [Partial pressure] in Arterial blood | blood | mm[Hg] | 4659 | 3250 | 7909 | | 240.60 | 252.11 | 6.23e-05 | 9016.31 | 8070.43 | 0.01385 | 0.07406 | -0.03841 | 0.03461 | 0.001477 | 0.002323 | 0.009702 | 0.08156 | 0.07253 | 0.7163 |
| pO2-V | pO2 venous | blood | mm[Hg] | 4442 | 4071 | 8513 | | 46.15 | 46.88 | 0.1734 | 600.83 | 596.63 | 0.923 | -0.02069 | -0.02293 | -0.04306 | 0.3417 | 0.3913 | -0.01702 | 0.06022 | 0.07619 | 0.611 |
| pO2 | pO2 | blood | mm[Hg] | 8698 | 7210 | 15908 | | 157.51 | 157.83 | 0.8096 | 7450.58 | 8183.20 | 2.66e-05 | -0.03915 | 0.1241 | 0.08596 | 0.01517 | 0.021 | -0.09773 | 0.07909 | 0.1766 | 1.31e-07 |
| POCA1C | Hemoglobin A1c (Glycated) | blood | % | 1866 | 1988 | 3854 | | 7.252 | 7.218 | 0.4977 | 0.9737 | 0.9717 | 0.9741 | -0.1094 | 0.03403 | -0.0661 | 6.59e-04 | 0.00106 | -0.06887 | 0.0339 | 0.09734 | 0.06152 |
| PR INTERVAL | PR Interval mean | cardiovascular | | 6106 | 7089 | 13195 | | 171.70 | 159.49 | 1.08e-99 | 508.38 | 843.94 | 1.43e-19 | -0.3454 | 0.2122 | -0.1594 | 1.47e-86 | 6.95e-86 | -0.1381 | -0.136 | 0.01697 | 3.03e-10 |
| PR_INTERVAL | PR Interval mean | cardiovascular | ms | 19278 | 21460 | 40738 | | 159.10 | 154.19 | 5.42e-37 | 1182.18 | 1906.05 | 1.93e-74 | -0.2269 | 0.1193 | -0.109 | 6.71e-115 | 3.48e-114 | -0.1281 | -0.05365 | 0.07381 | 4.99e-27 |
| Pr24Hr | Pr24Hr | urinary | g/(24.h) | 529 | 1008 | 1537 | | 0.967 | 0.5286 | 8.53e-08 | 0.7645 | 0.8217 | 0.8241 | -0.2357 | 0.1301 | -0.1148 | 2.16e-05 | 3.86e-05 | -0.4249 | -0.06615 | 0.3322 | 6.43e-05 |
| Pralbu | Prealbumin [Mass/volume] in Serum or Plasma | metabolic | mg/dL | 3583 | 3276 | 6859 | | 16.8 | 17.39 | 0.003637 | 40.03 | 41.69 | 0.5252 | 0.05032 | -0.09958 | -0.05014 | 0.03753 | 0.0503 | -0.0487 | 0.033 | 0.08012 | 0.1612 |
| PRODUCT BLOOD TYPE | Product blood type | blood | | 598 | 594 | 1192 | | 5678.01 | 5715.24 | 0.7379 | 1232972.4 | 1102340.2 | 0.3768 | -0.01322 | 0.06725 | 0.0537 | 0.8196 | 0.8539 | -0.02509 | 0.05273 | 0.07473 | 0.6693 |
| Prog-R | Progesterone rapid | ob/gyn | ng/mL | 16 | 1116 | 1132 | Sex specific lab | 4.35 | 9.128 | 0.03532 | | | | 0.02787 | 0.02616 | 0.05123 | 0.9277 | 0.939 | | | | |
| Prolac | Prolactin [Mass/volume] in Serum or Plasma | endocrine | ng/mL | 964 | 2100 | 3064 | | 10.59 | 14.34 | 5.84e-12 | 553.99 | 589.82 | 0.8943 | 0.4146 | -0.4506 | -0.03055 | 3.44e-26 | 1.04e-25 | 0.8292 | -0.1302 | -1.027 | 1.12e-23 |
| Promyl | Promyelocytes/100 leukocytes in Blood by Manual count | blood | | 1537 | 1397 | 2934 | | 1.375 | 1.411 | 0.3591 | 2.46 | 2.54 | 0.8121 | 0.04604 | -0.06093 | -0.01686 | 0.2132 | 0.2559 | -0.0327 | -5.25e-04 | 0.03002 | 0.5445 |
| PSA | PSA serum/plasma | cancer | ng/mL | 9198 | 6 | 9204 | Sex specific lab | 2.572 | 2.002 | 0.7668 | | | | 0.289 | -0.1429 | 0.1409 | 0.6 | 0.6502 | | | | |
| PT-inr | INR | blood | | 18431 | 19329 | 37760 | | 1.279 | 1.211 | 5.08e-59 | 0.1826 | 0.1878 | 0.254 | -0.1985 | 0.2169 | 0.01167 | 2.60e-83 | 1.21e-82 | 0.00131 | 0.05857 | 0.0572 | 0.9153 |
| PTT-pt | aPTT in Platelet poor plasma | blood | s | 16272 | 17241 | 33513 | | 34.75 | 32.75 | 5.40e-64 | 218.54 | 224.40 | 0.3526 | -0.1977 | 0.2273 | 0.03635 | 1.63e-73 | 7.29e-73 | -0.003386 | -0.05291 | -0.04947 | 0.8049 |

| all_labs | full_name | group | selected_unit | male_n | female_n | total_n | sex_specific_lab | raw_median_male_mean | raw_median_female_mean | raw_median_p.value | raw_variance_female_mean | raw_variance_male_mean | raw_variance_p.value | std_mean_diff | median_male_mean | median_female_mean | median_p.value | median_p.value.fdr | std_variance_diff | variance_female_mean | variance_male_mean | variance_p.value |
|---|---|---|---|---|---|---|---|---|---|---|---|---|---|---|---|---|---|---|---|---|---|---|
| PTTCtr | aPTT in Platelet poor plasma by Coagulation assay | blood | s | 9959 | 10228 | 20187 | | 28.96 | 29.12 | 3.59e-05 | 3.479 | 3.102 | 6.22e-04 | -0.0495 | -0.3595 | -0.3986 | 4.32e-04 | 7.09e-04 | 0.08035 | 0.2411 | 0.151 | 1.59e-05 |
| PULSE OXIMETRY | Oxygen saturation in Arterial blood by pulse oximetry | vitals | % | 11869 | 17346 | 29215 | | 97.22 | 97.83 | 2.50e-218 | 3.376 | 3.717 | 7.52e-19 | 0.3318 | -0.1786 | 0.1686 | 1.98e-173 | 1.25e-172 | -0.08185 | 0.09283 | 0.1756 | 4.43e-11 |
| Pulse | Heart rate | vitals | | 26707 | 35247 | 61954 | | 77.02 | 79.72 | 2.56e-164 | 32174.4 | 29283.4 | 0.9026 | 0.2046 | -0.102 | 0.07999 | 4.61e-137 | 2.64e-136 | -0.2323 | 0.1973 | 0.4139 | 8.29e-175 |
| PULSE | Heart rate | vitals | | 12927 | 19242 | 32169 | | 75.01 | 78.75 | 6.01e-166 | 109.26 | 114.96 | 4.90e-06 | 0.2758 | -0.2223 | 0.02898 | 2.20e-127 | 1.19e-126 | -0.05906 | 0.1257 | 0.1745 | 7.25e-07 |
| Q ONSET | Q onset | cardiovascular | | 6313 | 7230 | 13543 | | 502.61 | 502.90 | 7.57e-08 | 9.538 | 10.43 | 0.07275 | 0.04698 | 0.005083 | 0.04726 | 0.006052 | 0.008744 | -0.008963 | 0.02402 | 0.03301 | 0.6761 |
| QMG-NL | Quantiferon Mitogen assay | immune/inflammatory | [iU]/mL | 747 | 892 | 1639 | | 5.629 | 5.84 | 0.1731 | 7.352 | 6.677 | 0.4195 | 0.04301 | -0.09823 | -0.05604 | 0.3895 | 0.441 | 0.09883 | 0.1257 | 0.02764 | 0.2712 |
| QRS DURATION | QRS duration | cardiovascular | | 6316 | 7224 | 13540 | | 108.07 | 96.54 | 4.22e-193 | 83.69 | 103.51 | 1.03e-04 | -0.5229 | 0.3938 | -0.161 | 1.66e-209 | 1.29e-208 | 0.03351 | -0.001634 | -0.03757 | 0.118 |
| QRS HORIZONTAL AXIS | QRS horizontal axis | cardiovascular | | 6237 | 7176 | 13413 | | 17.71 | -2.722 | 8.98e-55 | 3193.30 | 3870.29 | 5.80e-04 | -0.2029 | 0.07325 | -0.1487 | 2.27e-31 | 7.37e-31 | -0.008876 | -0.1373 | -0.1278 | 0.6813 |
| QRS_DURATION | QRS duration | cardiovascular | ms | 19269 | 21446 | 40715 | | 99.56 | 90.22 | 0 | 77.88 | 104.26 | 2.34e-22 | -0.5494 | 0.3361 | -0.2206 | 0 | 0 | -0.07789 | 0.009533 | 0.08841 | 6.20e-11 |
| QT INTERVAL | QT interval | cardiovascular | | 6310 | 7225 | 13535 | | 403.14 | 397.72 | 3.66e-14 | 1043.24 | 1043.62 | 0.99 | -0.06623 | 0.1375 | 0.07188 | 1.25e-04 | 2.13e-04 | 0.00618 | -0.03024 | -0.03647 | 0.7734 |
| QT_INTERVAL_C | QT interval corrected | cardiovascular | ms | 19274 | 21459 | 40733 | | 436.04 | 438.78 | 1.13e-20 | 555.10 | 715.21 | 8.09e-56 | 0.1681 | 0.03677 | 0.1957 | 1.24e-63 | 5.20e-63 | -0.1859 | 0.0041 | 0.1828 | 3.79e-55 |
| QT_INTERVAL | QT interval | cardiovascular | ms | 19277 | 21462 | 40739 | | 394.29 | 391.55 | 5.46e-13 | 1044.59 | 1184.11 | 3.98e-17 | -0.0207 | 0.06827 | 0.04845 | 0.03755 | 0.0503 | -0.1205 | 0.04013 | 0.1543 | 4.32e-24 |
| QT3 | Triiodothyronine (T3) [Mass/volume] in Serum or Plasma | endocrine | ng/dL | 1478 | 3383 | 4861 | | 107.03 | 115.37 | 1.29e-13 | 1257.74 | 988.17 | 0.03984 | 0.1463 | -0.08053 | 0.05672 | 4.89e-06 | 9.04e-06 | 0.02921 | 0.04708 | 0.02194 | 0.5516 |
| QTB-NL | Quantiferon TB assay | immune/inflammatory | [iU]/mL | 827 | 990 | 1817 | | 0.02634 | 0.01796 | 0.1099 | 0.004071 | 0.008919 | 0.4705 | 0.03635 | -0.02683 | 0.009069 | 0.4414 | 0.4897 | -0.1534 | 0.07558 | 0.2267 | 0.05663 |
| QTCB | Q-T interval corrected based on Bazett formula | cardiovascular | | 6312 | 7225 | 13537 | | 443.98 | 445.55 | 0.00576 | 621.70 | 673.98 | 0.02537 | 0.1461 | -0.02468 | 0.1217 | 4.72e-17 | 1.20e-16 | -0.05927 | -0.05139 | 0.009238 | 0.005714 |
| QTCF | Q-T interval corrected based on Fridericia formula | cardiovascular | | 6311 | 7224 | 13535 | | 429.16 | 428.40 | 0.1514 | 543.03 | 545.75 | 0.8964 | 0.08629 | 0.03912 | 0.1263 | 6.64e-07 | 1.28e-06 | 0.01442 | -0.02378 | -0.03848 | 0.5014 |
| QtfNIL | Gamma interferon background [Units/volume] in Blood by Immunoassay | immune/inflammatory | [iU]/mL | 1197 | 1447 | 2644 | | 0.04856 | 0.05209 | 0.1357 | 0.003006 | 0.003268 | 0.7833 | 0.09318 | 0.1441 | 0.2326 | 0.01725 | 0.02377 | -0.07195 | 0.06656 | 0.1344 | 0.2329 |
| Quant_TB1(-)NIL | Quant TB1(-) NIL | immune/inflammatory | [iU]/mL | 639 | 838 | 1477 | | 0.009984 | 0.01141 | 0.6802 | 1.94e-04 | 6.13e-04 | 0.344 | 0.08493 | -0.1052 | -0.02023 | 0.1063 | 0.1335 | -0.09941 | -0.09511 | 5.85e-04 | 0.2436 |
| Quant_TB2(-)NIL | Quant TB2(-) NIL | immune/inflammatory | [iU]/mL | 639 | 838 | 1477 | | 0.01082 | 0.0111 | 0.9285 | 2.39e-04 | 0.004396 | 0.2784 | 0.02754 | -0.07969 | -0.05186 | 0.6003 | 0.6502 | 0.06471 | -0.04505 | -0.1103 | 0.46 |

| all_labs | full_name | group | selected_unit | male_n | female_n | total_n | sex_specific_lab | raw_median_male_mean | raw_median_female_mean | raw_median_p.value | raw_variance_female_mean | raw_variance_male_mean | raw_variance_p.value | std_mean_diff | median_male_mean | median_female_mean | median_p.value | median_p.value.fdr | std_variance_diff | variance_female_mean | variance_male_mean | variance_p.value |
|---|---|---|---|---|---|---|---|---|---|---|---|---|---|---|---|---|---|---|---|---|---|---|
| R AN AIRWAY RESPIRATION RATE | Respiratory rate by airway flow measurement | vitals | | 5423 | 6963 | 12386 | | 13.92 | 13.81 | 0.1383 | 24.48 | 22.86 | 5.65e-06 | -0.08184 | 0.06703 | -0.01419 | 5.45e-06 | 1.00e-05 | 0.05313 | 0.2134 | 0.1629 | 0.003193 |
| R AN ANESTHESIA BLOOD PRESSURE | Diastolic blood pressure by Noninvasive | vitals | mm[Hg] | 6029 | 7924 | 13953 | | 67.57 | 63.82 | 8.19e-73 | 203.49 | 186.22 | 4.86e-12 | -0.3313 | 0.1537 | -0.1657 | 4.19e-86 | 1.97e-85 | 0.1186 | 0.09326 | -0.02246 | 3.52e-12 |
| R AN ANESTHESIA TEMPERATURE | Body temperature | vitals | Cel | 931 | 1248 | 2179 | | 36.06 | 36.09 | 0.3911 | 0.131 | 0.1344 | 0.748 | 0.008896 | -0.01861 | -0.009979 | 0.8385 | 0.87 | 0.02954 | -0.07157 | -0.1031 | 0.5051 |
| R AN CORE BODY TEMPERATURE | Body temperature - Core | vitals | Cel | 675 | 719 | 1394 | | 36.07 | 36.13 | 0.1054 | 0.2808 | 0.3377 | 0.1401 | 0.0493 | -0.03076 | 0.01731 | 0.3576 | 0.4086 | -0.05369 | -0.2015 | -0.1429 | 0.3235 |
| R AN ESOPHAGEAL TEMPERATURE | Temperature - Distal airway | vitals | Cel | 1304 | 1636 | 2940 | | 36.15 | 36.16 | 0.4807 | 0.1351 | 0.1375 | 0.786 | -0.01925 | -0.02731 | -0.04607 | 0.6051 | 0.6537 | 0.01717 | 0.007461 | -0.009446 | 0.6461 |
| R AN HEART RATE MONITOR | Heart rate | vitals | | 5668 | 7336 | 13004 | | 73.52 | 75.72 | 8.16e-26 | 88.38 | 93.23 | 0.009567 | 0.1403 | -0.1351 | -0.003466 | 3.65e-15 | 8.89e-15 | 0.02438 | 0.004867 | -0.01946 | 0.1736 |
| R AN MEAN ARTERIAL BLOOD PRESSURE | Invasive Mean blood pressure | vitals | mm[Hg] | 1168 | 878 | 2046 | | 87.98 | 91.35 | 0.03518 | 933.07 | 1012.57 | 0.1809 | 0.2465 | -0.05609 | 0.1859 | 3.40e-08 | 6.89e-08 | 0.0219 | 0.04149 | 0.01828 | 0.6223 |
| R AN NASOPHARYNGEAL TEMPERATURE | Temperature - Distal airway | vitals | Cel | 537 | 574 | 1111 | | 35.97 | 36 | 0.4503 | 0.5045 | 0.6284 | 0.2005 | 0.05894 | 0.001471 | 0.0579 | 0.3261 | 0.377 | -0.1178 | -0.1931 | -0.06788 | 0.05278 |
| R AN NIBP MEAN | Mean blood pressure by Noninvasive | vitals | mm[Hg] | 5736 | 7478 | 13214 | | 83.57 | 81.08 | 2.28e-28 | 243.45 | 232.77 | 3.14e-04 | -0.1755 | 0.04875 | -0.1214 | 7.00e-24 | 2.00e-23 | 0.1118 | 0.06918 | -0.04301 | 2.25e-10 |
| R AN PULSE SPO2 | Heart rate by Pulse oximetry | vitals | | 5626 | 7283 | 12909 | | 72.96 | 75.41 | 2.12e-31 | 88.93 | 92.86 | 0.0349 | 0.1559 | -0.1423 | 0.003317 | 2.67e-18 | 6.90e-18 | 0.02988 | -0.0155 | -0.04531 | 0.09604 |
| R AN SKIN TEMPERATURE | Temperature of Skin | vitals | Cel | 1261 | 1630 | 2891 | | 36.12 | 36.14 | 0.5552 | 0.1516 | 0.1645 | 0.3738 | 0.002882 | -0.00536 | -0.002568 | 0.9386 | 0.9462 | 0.02121 | -0.03661 | -0.05963 | 0.6086 |
| R APACHE TEMPERATURE | Body temperature | vitals | Cel | 11789 | 17589 | 29378 | | 36.69 | 36.71 | 4.91e-13 | 0.1157 | 0.1305 | 8.75e-16 | 0.0618 | -0.1067 | -0.054 | 2.19e-07 | 4.29e-07 | -0.06556 | 0.09997 | 0.1578 | 2.45e-07 |
| R ARTERIAL LINE BLOOD PRESSURE | Invasive Systolic blood pressure | vitals | mm[Hg] | 631 | 512 | 1143 | | 117.89 | 118.83 | 0.4017 | 292.98 | 273.35 | 0.2058 | 0.04985 | -0.1939 | -0.1465 | 0.4026 | 0.4506 | 0.06205 | -0.09272 | -0.1545 | 0.3051 |
| R ED PRE-ARRIVAL BP | Systolic blood pressure by Noninvasive | vitals | mm[Hg] | 1001 | 1097 | 2098 | | 74.44 | 73 | 0.02157 | 110.38 | 120.15 | 0.4487 | -0.127 | -0.1339 | -0.2594 | 0.003582 | 0.005428 | -0.06171 | 0.04199 | 0.1006 | 0.3719 |
| R ED PRE-ARRIVAL EMS O2 SAT | Oxygen saturation in Arterial blood by Pulse oximetry | vitals | % | 981 | 1061 | 2042 | | 96.59 | 96.96 | 0.007124 | 5.724 | 16.82 | 0.225 | 0.1254 | -0.1005 | 0.02968 | 0.004696 | 0.006988 | -0.1309 | 0.09327 | 0.2163 | 0.06378 |
| R ED PRE-ARRIVAL PULSE (HEART RATE) | Heart rate | vitals | | 1007 | 1100 | 2107 | | 87.7 | 88.61 | 0.2638 | 151.11 | 169.78 | 0.3468 | 0.0144 | 0.0669 | 0.08114 | 0.7415 | 0.7841 | -0.09869 | 0.1693 | 0.2641 | 0.1543 |
| R ED PRE-ARRIVAL RESP RATE | Respiratory rate | vitals | | 920 | 1028 | 1948 | | 18.7 | 18.25 | 9.08e-04 | 7.357 | 8.034 | 0.5105 | -0.1148 | 0.05055 | -0.06401 | 0.0115 | 0.0161 | -0.1027 | 0.05735 | 0.1646 | 0.1608 |
| R FN WEIGHT USED FOR EQUATIONS CALCULATIONS | Body weight | vitals | kg | 569 | 522 | 1091 | | 75.96 | 62.19 | 9.65e-47 | 32.09 | 40.86 | 0.4587 | -0.8718 | 0.2478 | -0.5984 | 1.73e-51 | 6.84e-51 | -0.1902 | -0.1443 | 0.03466 | 0.1214 |

| all_labs | full_name | group | selected_unit | male_n | female_n | total_n | sex_specific_lab | raw_median_male_mean | raw_median_female_mean | raw_median_p.value | raw_variance_female_mean | raw_variance_male_mean | raw_variance_p.value | std_mean_diff | median_male_mean | median_female_mean | median_p.value | median_p.value.fdr | std_variance_diff | variance_female_mean | variance_male_mean | variance_p.value |
|---|---|---|---|---|---|---|---|---|---|---|---|---|---|---|---|---|---|---|---|---|---|---|
| R HEART RATE MONITOR | Heart rate | vitals | | 6986 | 9190 | 16176 | | 75.82 | 78.68 | 7.98e-41 | 117.76 | 118.80 | 0.938 | 0.1823 | -0.1907 | -0.01598 | 4.71e-30 | 1.51e-29 | -0.09017 | 0.07002 | 0.1514 | 2.97e-08 |
| R IP IDEAL BODY WEIGHT | Ideal body weight | vitals | kg | 12623 | 19051 | 31674 | | 33.42 | 25.18 | 0 | 0.4675 | 0.686 | 2.47e-11 | -1.536 | 0.8676 | -0.6681 | 0 | 0 | 0.03242 | 0.1463 | 0.1128 | 0.009296 |
| R RT (FEMALE ADULT) IDEAL BODY WEIGHT | Ideal body weight — female | vitals | kg | 12624 | 19052 | 31676 | | 69.19 | 55.52 | 0 | 2.168 | 3.216 | 1.58e-12 | -1.415 | 0.8016 | -0.613 | 0 | 0 | 0.03297 | 0.1442 | 0.11 | 0.008199 |
| R RT (MALE ADULT) IDEAL BODY WEIGHT | Ideal body weight — male | vitals | kg | 12624 | 19052 | 31676 | | 73.69 | 60.02 | 0 | 2.168 | 3.216 | 1.58e-12 | -1.415 | 0.8016 | -0.613 | 0 | 0 | 0.03297 | 0.1442 | 0.11 | 0.008199 |
| R VUMC AN RESPIRATION RATE | Respiratory rate | vitals | | 4501 | 5664 | 10165 | | 13.73 | 13.43 | 6.12e-05 | 20.65 | 20.68 | 0.9396 | -0.1426 | 0.101 | -0.04005 | 6.06e-13 | 1.40e-12 | -0.03159 | 0.1576 | 0.1878 | 0.1125 |
| R VUMC OXYGEN FLOW DELIVERED | Delivered oxygen flow rate | vitals | L/min | 4330 | 5139 | 9469 | | 2.798 | 2.651 | 1.64e-05 | 6.262 | 28.81 | 0.2545 | -0.1567 | 0.1964 | 0.05509 | 2.70e-14 | 6.49e-14 | -0.1912 | 0.1762 | 0.361 | 1.31e-15 |
| R_F | Rheumatoid factor [Units/volume] in Serum or Plasma | immune/inflammatory | [iU]/mL | 1250 | 2715 | 3965 | | 38.31 | 29.03 | 0.009359 | 3023.50 | 3646.40 | 0.7204 | 0.101 | -0.2433 | -0.1428 | 0.004898 | 0.007267 | 0.07551 | -0.08204 | -0.1555 | 0.4385 |
| RATE_C | Heart rate | vitals | | 19277 | 21462 | 40739 | | 75.88 | 77.71 | 1.31e-31 | 209.38 | 245.55 | 1.62e-18 | 0.1216 | -0.0278 | 0.08661 | 2.61e-34 | 8.64e-34 | -0.1325 | 0.04965 | 0.1748 | 8.61e-29 |
| RBC | Erythrocytes [#/volume] in Blood by Automated count | blood | 10*6/uL | 26954 | 35638 | 62592 | | 4.385 | 4.217 | 2.62e-225 | 0.1603 | 0.2268 | 1.33e-257 | -0.3871 | 0.1106 | -0.2533 | 0 | 0 | -0.2721 | 0.07673 | 0.3314 | 2.59e-216 |
| RBCFol | Folate [Mass/volume] in Red Blood Cells | metabolic | ng/mL{RBC} | 2084 | 2842 | 4926 | | 924.55 | 918.57 | 0.5836 | 75600.3 | 73765.9 | 0.8355 | 0.04922 | 0.2074 | 0.2552 | 0.09008 | 0.1148 | 0.1307 | 0.1083 | -0.03024 | 0.04014 |
| RDW-SD | RDW SD | blood | fL | 2143 | 3133 | 5276 | | 45.02 | 45.06 | 0.8071 | 7.921 | 8.04 | 0.8507 | 0.1357 | 0.03275 | 0.1699 | 1.58e-06 | 2.97e-06 | 0.1174 | 0.1716 | 0.0506 | 0.001004 |
| RDW | Erythrocyte distribution width [Ratio] by Automated count Erythrocyte distribution width [Ratio] by Automated count | blood | % | 26955 | 35661 | 62616 | | 14.09 | 13.94 | 2.38e-28 | 1.289 | 1.317 | 0.1667 | -0.002563 | 0.1264 | 0.124 | 0.7523 | 0.7939 | 0.05171 | 0.2987 | 0.248 | 2.21e-09 |
| RDWSD | Erythrocyte distribution width [Entitic volume] by Automated count | blood | fL | 24073 | 31506 | 55579 | | 46.4 | 45.98 | 4.16e-19 | 12.22 | 13.4 | 7.60e-08 | 0.05607 | 0.1205 | 0.1762 | 9.58e-11 | 2.11e-10 | -0.01078 | 0.2076 | 0.2186 | 0.249 |
| RenAct | Renin [Enzymatic activity/volume] in Plasma | endocrine | ng/mL/h | 452 | 701 | 1153 | | 5.307 | 4.925 | 0.478 | 49.32 | 33.44 | 0.312 | -0.08095 | 0.1657 | 0.08354 | 0.1788 | 0.2173 | -0.05127 | 0.06557 | 0.1192 | 0.6708 |
| RESPIRATIONS | Respiratory rate | vitals | | 11399 | 16703 | 28102 | | 16.96 | 16.81 | 3.44e-14 | 6.745 | 8.288 | 1.59e-54 | -0.03561 | -0.1086 | -0.1429 | 0.004004 | 0.006049 | -0.1781 | 0.02512 | 0.2084 | 1.51e-43 |
| RespRt | Respiratory rate | vitals | | 23979 | 30952 | 54931 | | 17.33 | 17.09 | 6.75e-29 | 3053.27 | 2455.81 | 0.789 | -0.1193 | 0.006788 | -0.1016 | 5.39e-44 | 1.93e-43 | 0.2311 | -0.2207 | -0.4456 | 8.38e-146 |
| RetAbs | Reticulocytes absolute | blood | 10*6/uL | 3555 | 3978 | 7533 | | 0.07009 | 0.07033 | 0.8203 | 0.001832 | 0.004171 | 0.2783 | -0.02498 | -0.03325 | -0.05709 | 0.279 | 0.3302 | -0.1772 | 0.08007 | 0.2408 | 1.29e-06 |

| all_labs | full_name | group | selected_unit | male_n | female_n | total_n | sex_specific_lab | raw_median_male_mean | raw_median_female_mean | raw_median_p.value | raw_variance_female_mean | raw_variance_male_mean | raw_variance_p.value | std_mean_diff | median_male_mean | median_female_mean | median_p.value | median_p.value.fdr | std_variance_diff | variance_female_mean | variance_male_mean | variance_p.value |
|---|---|---|---|---|---|---|---|---|---|---|---|---|---|---|---|---|---|---|---|---|---|---|
| RETHE | Hemoglobin [Entitic mass] in Reticulocytes | blood | pg | 2403 | 2574 | 4977 | | 31.65 | 31.2 | 0.00545 | 16.01 | 15.12 | 0.4852 | -0.07614 | -0.01129 | -0.08398 | 0.007321 | 0.01046 | 0.03289 | 0.104 | 0.06894 | 0.5125 |
| RetiCt | Reticulocytes/100 erythrocytes in Blood by Automated count | blood | % | 3575 | 3964 | 7539 | | 2.006 | 2.067 | 0.1247 | 2.377 | 1.717 | 0.1632 | 0.01862 | -0.04245 | -0.02448 | 0.4194 | 0.4684 | -0.1083 | 0.02411 | 0.1206 | 0.002893 |
| RetPal | Retinyl palmitate [Mass/volume] in Serum or Plasma | metabolic | mg/L | 631 | 1135 | 1766 | | 0.03853 | 0.03806 | 0.7398 | 0.001083 | 0.001038 | 0.7891 | -0.04141 | -0.07907 | -0.1207 | 0.3998 | 0.4485 | 0.003198 | -0.007637 | -0.01093 | 0.9631 |
| RR INTERVAL | RR interval | cardiovascular | | 6317 | 7229 | 13546 | | 838.64 | 806.29 | 7.39e-31 | 15018.1 | 16999.0 | 1.18e-05 | -0.1637 | 0.1406 | -0.01452 | 2.69e-21 | 7.36e-21 | -0.1064 | -0.05993 | 0.04636 | 6.83e-07 |
| RV_PPR | RV PPR | lung | % | 869 | 1045 | 1914 | | 113.90 | 101.49 | 1.39e-10 | 738.53 | 442.25 | 0.03766 | -0.2802 | 0.1437 | -0.1216 | 1.21e-09 | 2.57e-09 | -0.05224 | 0.05299 | 0.1004 | 0.5542 |
| RV_PRD | RV PRD | lung | L | 868 | 1046 | 1914 | | 2.326 | 2.031 | 1.71e-98 | 0.001033 | 7.94e-04 | 0.2502 | -1.311 | 0.762 | -0.4025 | 1.62e-222 | 1.30e-221 | 0.004163 | 0.2262 | 0.2224 | 0.9631 |
| RV_PRL | RV PRL | lung | | 1145 | 1147 | 2292 | | 1.546 | 1.24 | 6.55e-128 | 0.002896 | 0.007371 | 0.002843 | -1.301 | 0.7004 | -0.48 | 3.97e-276 | 3.65e-275 | -0.117 | -0.03944 | 0.07144 | 0.1894 |
| RV_PRU | RV PRU | lung | | 1144 | 1143 | 2287 | | 3.06 | 2.771 | 2.22e-111 | 0.006068 | 0.002504 | 8.23e-04 | -1.276 | 0.7082 | -0.4689 | 4.23e-261 | 3.69e-260 | 0.2259 | 0.181 | -0.04453 | 0.01185 |
| RV_ZBD | RV ZBD | lung | L | 868 | 1043 | 1911 | | 2.631 | 2.069 | 1.92e-35 | 0.3336 | 0.2458 | 0.2336 | -0.5473 | 0.3157 | -0.2168 | 2.62e-33 | 8.58e-33 | -0.06195 | 0.02486 | 0.08463 | 0.4871 |
| RVOTLC_PPR | RVOTLC PPR | lung | % | 869 | 1046 | 1915 | | 119.67 | 106.29 | 4.13e-24 | 304.82 | 254.48 | 0.2892 | -0.4468 | 0.1925 | -0.235 | 1.64e-22 | 4.54e-22 | -0.1403 | -0.02921 | 0.0908 | 0.1154 |
| RVOTLC_PRD | RVOTLC PRD | lung | % | 869 | 1046 | 1915 | | 34.28 | 40.42 | 4.21e-141 | 0.5397 | 0.258 | 0.004459 | 1.694 | -1.025 | 0.9069 | 0 | 0 | 0.3705 | 0.3081 | -0.03956 | 2.61e-05 |
| RVOTLC_PRL | RVOTLC PRL | lung | | 1145 | 1147 | 2292 | | 25.28 | 29.1 | 9.88e-71 | 2.731 | 5.357 | 0.00286 | 1.55 | -0.8869 | 0.7057 | 0 | 0 | -0.0195 | 0.01252 | 0.03172 | 0.8268 |
| RVOTLC_PRU | RVOTLC PRU | lung | | 926 | 949 | 1875 | | 42.53 | 50.8 | 1.73e-212 | 0.2022 | 0.19 | 0.794 | 1.644 | -0.888 | 0.8425 | 0 | 0 | 0.05887 | 0.07915 | 0.01923 | 0.5771 |
| RVOTLC_ZBD | RVOTLC ZBD | lung | % | 869 | 1046 | 1915 | | 41.16 | 43.08 | 3.93e-04 | 47.88 | 30.06 | 0.01007 | 0.341 | -0.2132 | 0.1211 | 1.36e-13 | 3.19e-13 | 0.1391 | 0.074 | -0.05154 | 0.1181 |
| RVVT | dRVVT actual/normal (normalized LA screen) | immune/inflammatory | {ratio} | 535 | 1518 | 2053 | | 1.106 | 1.026 | 1.41e-06 | 0.03422 | 0.07969 | 0.05787 | -0.2342 | 0.2611 | 0.02698 | 4.39e-06 | 8.16e-06 | 0.2924 | 0.3517 | 0.004329 | 0.03987 |
| S-T HORIZONTAL AXIS | ST horizontal axis | cardiovascular | | 6224 | 7039 | 13263 | | 78.06 | 81.6 | 2.23e-04 | 2446.27 | 2122.11 | 0.005334 | 0.08416 | -0.08764 | 0.003101 | 1.25e-06 | 2.37e-06 | 0.1575 | 0.05291 | -0.1193 | 4.98e-13 |
| SclAb | SCL-70 extractable nuclear Ab [Units/volume] in Serum by Immunoassay | immune/inflammatory | | 237 | 772 | 1009 | | 0.3709 | 0.3266 | 0.433 | 0.02759 | 0.02676 | 0.959 | -0.1121 | 0.1431 | 0.02629 | 0.1421 | 0.1766 | -0.01427 | 0.1694 | 0.1837 | 0.9639 |
| SedRat | Erythrocyte sedimentation rate by Westergren method | blood | mm/h | 8621 | 13581 | 22202 | | 24.17 | 22.72 | 1.10e-05 | 260.69 | 386.93 | 4.00e-27 | 0.09784 | -0.1362 | -0.03853 | 8.24e-12 | 1.85e-11 | -0.1431 | -0.0587 | 0.08552 | 2.69e-13 |
| SerFol | Folate [Mass/volume] in Serum or Plasma | metabolic | ng/mL | 3545 | 4968 | 8513 | | 12.49 | 13.01 | 1.97e-06 | 17.96 | 15.56 | 0.08513 | 0.1339 | 0.0187 | 0.1558 | 1.03e-09 | 2.20e-09 | 0.06467 | 0.05775 | -0.009858 | 0.09439 |
| SGBF | Specific gravity of Body fluid | blood | | 581 | 439 | 1020 | | 1.029 | 1.028 | 0.2856 | 0.001496 | 1.49e-04 | 0.2205 | -0.1792 | 0.01114 | -0.1622 | 0.004607 | 0.006875 | -0.01589 | 0.1769 | 0.1923 | 0.8861 |
| all_labs | full_name | group | selected_unit | male_n | female_n | total_n | sex_specific_lab | raw_median_male_mean | raw_median_female_mean | raw_median_p.value | raw_variance_female_mean | raw_variance_male_mean | raw_variance_p.value | std_mean_diff | median_male_mean | median_female_mean | median_p.value | median_p.value.fdr | std_variance_diff | variance_female_mean | variance_male_mean | variance_p.value |
| SGOT | SGOT | liver | [U]/L | 15836 | 20991 | 36827 | | 29.28 | 25.56 | 4.57e-43 | 1122.39 | 1541.84 | 1.02e-04 | -0.3019 | 0.1232 | -0.1541 | 1.50e-182 | 1.02e-181 | -0.2294 | -0.0224 | 0.1979 | 4.33e-89 |
| SGPT | SGPT | liver | [U]/L | 15997 | 21170 | 37167 | | 29.93 | 23.25 | 1.71e-122 | 827.23 | 1284.36 | 1.94e-11 | -0.4893 | 0.2254 | -0.2143 | 0 | 0 | -0.2564 | 0.1142 | 0.3435 | 6.69e-112 |

| all_labs | full_name | group | selected_unit | male_n | female_n | total_n | sex_specific_lab | raw_median_male_mean | raw_median_female_mean | raw_median_p.value | raw_variance_female_mean | raw_variance_male_mean | raw_variance_p.value | std_mean_diff | median_male_mean | median_female_mean | median_p.value | median_p.value.fdr | std_variance_diff | variance_female_mean | variance_male_mean | variance_p.value |
|---|---|---|---|---|---|---|---|---|---|---|---|---|---|---|---|---|---|---|---|---|---|---|
| SSAIgG | Sjogrens syndrome-A extractable nuclear IgG Ab [Units/volume] in Serum by Immunoassay | immune/inflammatory | | 325 | 1087 | 1412 | | 0.4267 | 0.7117 | 1.27e-05 | 0.6898 | 0.1297 | 0.03504 | 0.09629 | -0.02498 | 0.07046 | 0.1192 | 0.1493 | 0.006421 | -0.0189 | -0.02543 | 0.9823 |
| SSBIgG | Sjogrens syndrome-B extractable nuclear IgG Ab [Units/volume] in Serum by Immunoassay | immune/inflammatory | | 313 | 1003 | 1316 | | 0.3353 | 0.3138 | 0.4681 | 0.09331 | 0.02774 | 0.2534 | -0.04319 | 0.05018 | 0.00582 | 0.5208 | 0.5689 | -0.3785 | -0.1232 | 0.2526 | 0.1779 |
| ST_SEGMENT | ST segment axis | cardiovascular | deg | 19279 | 21462 | 40741 | | 68.36 | 68.63 | 0.6634 | 3980.69 | 3762.09 | 0.004152 | 0.04924 | 0.04268 | 0.09127 | 6.70e-07 | 1.29e-06 | 0.03845 | 0.09298 | 0.05526 | 0.001245 |
| SU-CA | Calcium urine | urinary | mg/dL | 273 | 893 | 1166 | | 10.23 | 9.756 | 0.3511 | 26.39 | 24.36 | 0.7599 | 0.02213 | -0.08516 | -0.06447 | 0.7625 | 0.8012 | 0.3117 | 0.09917 | -0.1807 | 0.1112 |
| SU-Cre | Creatinine urine | urinary | mg/dL | 549 | 1130 | 1679 | | 90.13 | 70.37 | 8.01e-14 | 871.60 | 683.49 | 0.271 | -0.401 | 0.2265 | -0.1663 | 1.22e-14 | 2.93e-14 | -0.0227 | 0.1333 | 0.1541 | 0.8189 |
| SU-K | Potassium urine | urinary | mmol/L | 710 | 704 | 1414 | | 36.78 | 35.31 | 0.1962 | 339.27 | 261.40 | 0.2045 | -0.06535 | 0.05396 | -0.01153 | 0.2192 | 0.2625 | -0.01512 | 0.0582 | 0.07251 | 0.8958 |
| SU-Na | Sodium urine | urinary | mmol/L | 3527 | 3137 | 6664 | | 61.17 | 62.44 | 0.1446 | 994.99 | 935.43 | 0.3444 | 0.02103 | -0.05555 | -0.03547 | 0.3917 | 0.4425 | -0.039 | 0.0193 | 0.05557 | 0.342 |
| SU-Osm | Urine osmolality | urinary | mosm/kg{H20} | 1602 | 1587 | 3189 | | 431.36 | 381.72 | 1.52e-16 | 16210.0 | 17642.4 | 0.4825 | -0.3202 | 0.1315 | -0.1729 | 9.70e-20 | 2.62e-19 | -0.05752 | 0.00826 | 0.06183 | 0.3901 |
| SU-Pro | Protein urine | urinary | mg/dL | 3467 | 3560 | 7027 | | 67.26 | 51.57 | 3.16e-06 | 13568.8 | 17126.7 | 0.2933 | -0.3073 | 0.1423 | -0.1561 | 1.88e-38 | 6.44e-38 | -0.268 | 0.1024 | 0.3471 | 2.77e-17 |
| SU-Ure | Urea urine | urinary | mg/dL | 1859 | 1351 | 3210 | | 506.17 | 412.37 | 1.35e-26 | 45366.7 | 49057.9 | 0.4775 | -0.3709 | 0.152 | -0.2025 | 1.22e-25 | 3.66e-25 | -0.1101 | 0.04179 | 0.1462 | 0.08038 |
| SUCrea | Creatinine [Mass/volume] in Urine | urinary | mg/dL | 7600 | 8019 | 15619 | | 110.35 | 90.56 | 5.92e-87 | 2889.98 | 3293.98 | 4.32e-04 | -0.3804 | 0.1514 | -0.2042 | 2.04e-127 | 1.11e-126 | -0.1722 | 0.1157 | 0.264 | 3.71e-18 |
| SyMnCl | Mononuclear cells/100 leukocytes in Synovial fluid by Manual count | immune/inflammatory | % | 584 | 457 | 1041 | | 37.88 | 48.69 | 1.35e-06 | 512.74 | 495.64 | 0.8657 | 0.3168 | -0.2806 | 0.04067 | 4.23e-07 | 8.21e-07 | 0.05666 | -0.1197 | -0.1712 | 0.6361 |
| SYSTOLIC | Systolic blood pressure | vitals | mm[Hg] | 26956 | 35815 | 62771 | | 125.99 | 123.05 | 3.62e-173 | 220.48 | 240.27 | 1.04e-38 | -0.144 | -0.1049 | -0.2329 | 5.98e-71 | 2.61e-70 | -0.01526 | 0.2211 | 0.2345 | 0.0654 |
| T HORIZONTAL AXIS | T horizontal axis | cardiovascular | | 6265 | 7147 | 13412 | | 50.69 | 37.46 | 2.22e-68 | 1678.13 | 1394.15 | 0.001507 | -0.3558 | 0.1112 | -0.2581 | 9.43e-96 | 4.73e-95 | 0.1235 | 0.006569 | -0.1266 | 1.18e-08 |
| T WAVE AXIS | T wave axis | cardiovascular | | 6303 | 7207 | 13510 | | 49.24 | 47.32 | 0.009493 | 1504.23 | 1644.39 | 0.08916 | 0.02065 | -0.002801 | 0.01824 | 0.2344 | 0.2801 | 0.00389 | -0.1109 | -0.115 | 0.8563 |
| T_WAVE | T wave axis | cardiovascular | deg | 19279 | 21462 | 40741 | | 46.41 | 43.85 | 2.29e-10 | 1481.85 | 1721.12 | 7.06e-07 | -5.84e-04 | -0.004381 | -0.004962 | 0.9533 | 0.9571 | -0.05379 | -0.01842 | 0.03544 | 6.27e-06 |
| T3FRE | Triiodothyronine (T3) Free [Mass/volume] in Serum or Plasma | endocrine | pg/mL | 547 | 1107 | 1654 | | 3.035 | 2.956 | 0.4063 | 0.993 | 1.038 | 0.9296 | -0.2077 | 0.04228 | -0.1645 | 1.18e-04 | 2.02e-04 | -0.2875 | 0.01894 | 0.2795 | 9.32e-04 |
| TBil | Total Bilirubin serum/plasma | metabolic | mg/dL | 24820 | 31936 | 56756 | | 0.7859 | 0.5917 | 0 | 0.215 | 0.3799 | 5.71e-31 | -0.5053 | 0.3117 | -0.1723 | 0 | 0 | -0.2916 | -0.009349 | 0.2665 | 5.44e-219 |
| TC:HDL | TC/HDL ratio | metabolic | | 2451 | 2113 | 4564 | | 3.998 | 3.659 | 6.33e-21 | 0.4467 | 0.5143 | 0.07507 | -0.323 | 0.1482 | -0.156 | 8.88e-28 | 2.77e-27 | -0.1768 | -0.06556 | 0.0941 | 3.54e-05 |
| Temper | Body temperature | vitals | [degF] | 24959 | 32524 | 57483 | | 98.13 | 98.15 | 0.02883 | 51.69 | 43.99 | 0.5654 | 0.02617 | 0.02244 | 0.04363 | 0.002056 | 0.003163 | -0.06457 | -0.02765 | 0.0365 | 9.27e-14 |
| TEMPERATURE | Body temperature | vitals | Cel | 11789 | 17589 | 29378 | | 36.68 | 36.7 | 3.49e-22 | 0.1035 | 0.1123 | 5.00e-08 | 0.08311 | -0.1276 | -0.05757 | 2.81e-12 | 6.41e-12 | -0.04729 | 0.1232 | 0.1632 | 1.84e-04 |

| all_labs | full_name | group | selected_unit | male_n | female_n | total_n | sex_specific_lab | raw_median_male_mean | raw_median_female_mean | raw_median_p.value | raw_variance_female_mean | raw_variance_male_mean | raw_variance_p.value | std_mean_diff | median_male_mean | median_female_mean | median_p.value | median_p.value.fdr | std_variance_diff | variance_female_mean | variance_male_mean | variance_p.value |
|---|---|---|---|---|---|---|---|---|---|---|---|---|---|---|---|---|---|---|---|---|---|---|
| TERMINAL_40_MS | Terminal 40 ms | cardiovascular | | 19279 | 21462 | 40741 | | 50.25 | 38.06 | 3.12e-59 | 2147.53 | 3098.64 | 2.70e-34 | -0.1377 | 0.05434 | -0.0879 | 3.57e-43 | 1.26e-42 | -0.1102 | -0.004638 | 0.1054 | 2.05e-20 |
| TestFr | Testosterone Free [Mass/volume] in Serum or Plasma | endocrine | pg/mL | 2215 | 137 | 2352 | | 59.97 | 10.25 | 2.62e-52 | 1083.03 | 2057.11 | 0.09663 | -1.92 | 0.1028 | -1.616 | 2.19e-46 | 8.17e-46 | -0.7407 | -0.6581 | 0.08244 | 0.004403 |
| Testos | Testosterone total | endocrine | ng/dL | 3626 | 1067 | 4693 | | 343.17 | 44.77 | 0 | 3378.51 | 26109.4 | 7.78e-43 | -1.448 | 0.284 | -1.13 | 0 | 0 | -0.7523 | -0.6502 | 0.09059 | 8.71e-42 |
| TGG | Total gamma globulin | immune/inflammatory | mg/dL | 581 | 434 | 1015 | | 35.88 | 39.28 | 0.6283 | 2316.18 | 29553.1 | 0.1111 | -0.1688 | 0.02021 | -0.1376 | 0.008451 | 0.01204 | -0.1868 | -0.03576 | 0.1598 | 0.2557 |
| Thiamn | Thiamine pyrophosphate [Moles/volume] in Blood | metabolic | nmol/L | 696 | 1229 | 1925 | | 119.29 | 118.62 | 0.7776 | 1463.57 | 2190.69 | 0.01435 | 0.04795 | -0.1237 | -0.07431 | 0.3325 | 0.3832 | -0.0835 | 0.008408 | 0.09919 | 0.3217 |
| THM | Total hemoglobin | blood | g/dL | 5720 | 4099 | 9819 | | 11.27 | 10.62 | 1.04e-49 | 2.146 | 2.166 | 0.7965 | -0.2795 | 0.05328 | -0.2172 | 1.89e-44 | 6.85e-44 | -0.0463 | 0.06514 | 0.1092 | 0.06359 |
| Thyrgb | Thyroglobulin [Mass/volume] in Serum or Plasma | endocrine | ng/mL | 275 | 813 | 1088 | | 60.06 | 54.57 | 0.7457 | 51115.9 | 55514.4 | 0.8831 | 0.4227 | -0.4825 | -0.04645 | 1.63e-08 | 3.33e-08 | 0.1712 | 0.02401 | -0.1594 | 0.1433 |
| TIBC | Total iron binding capacity | blood | ug/mL | 6750 | 8759 | 15509 | | 150.18 | 160.80 | 7.83e-06 | 7848.07 | 6230.09 | 3.05e-09 | 0.04467 | -0.1817 | -0.1381 | 0.004987 | 0.007355 | 0.09085 | 0.374 | 0.2713 | 1.33e-05 |
| TIGIgG | Tissue transglutaminase IgG Ab [Units/volume] in Serum by Immunoassay | immune/inflammatory | | 266 | 757 | 1023 | | 3.077 | 3.005 | 0.5108 | 0.4144 | 7.929 | 0.3255 | -0.01575 | -0.02392 | -0.03998 | 0.8289 | 0.8619 | -0.6 | -0.1662 | 0.399 | 0.2984 |
| TLC_PPR | TLC PPR | lung | % | 869 | 1045 | 1914 | | 92.03 | 94.57 | 0.0037 | 101.22 | 62.67 | 0.009431 | 0.1126 | -0.03678 | 0.06578 | 0.01446 | 0.02007 | 0.1705 | 0.06001 | -0.096 | 0.05656 |
| TLC_PRD | TLC PRD | lung | L | 868 | 1046 | 1914 | | 6.898 | 5.027 | 0 | 0.003716 | 0.002627 | 0.4476 | -1.629 | 0.879 | -0.6323 | 0 | 0 | -0.1187 | 0.06571 | 0.1769 | 0.1853 |
| TLC_PRL | TLC PRL | lung | | 955 | 987 | 1942 | | 5.302 | 3.956 | 0 | 0.003414 | 0.02564 | 2.13e-07 | -1.553 | 0.8223 | -0.6276 | 0 | 0 | -0.5163 | -0.2271 | 0.2586 | 8.11e-08 |
| TLC_PRU | TLC PRU | lung | | 954 | 987 | 1941 | | 8.48 | 6.092 | 0 | 0.009428 | 0.01828 | 0.2686 | -1.632 | 0.8579 | -0.6932 | 0 | 0 | -0.1453 | -0.06331 | 0.07655 | 0.1381 |
| TLC_ZBD | TLC ZBD | lung | L | 869 | 1045 | 1914 | | 6.345 | 4.769 | 1.08e-133 | 0.3283 | 0.2878 | 0.5582 | -1.081 | 0.5959 | -0.3942 | 8.63e-140 | 4.99e-139 | -0.1172 | -0.05152 | 0.0576 | 0.1919 |
| TotVl | Volume in Urine collected for unspecified duration | toxicology/pharmacology | mL | 2231 | 3739 | 5970 | | 1999.17 | 1817.10 | 2.77e-13 | 296786.1 | 349521.1 | 0.05672 | -0.183 | 0.126 | -0.05691 | 1.06e-11 | 2.37e-11 | -0.1853 | 0.05054 | 0.2248 | 7.68e-06 |
| TProSe | Protein serum/plasma | metabolic | g/dL | 15800 | 20773 | 36573 | | 7.144 | 7.176 | 2.94e-07 | 0.1948 | 0.2262 | 4.37e-16 | -0.00946 | -0.06872 | -0.07725 | 0.374 | 0.4253 | -0.09327 | 0.09584 | 0.1745 | 2.76e-15 |
| TProt | Total protein | blood | g/dL | 19923 | 25624 | 45547 | | 7.031 | 7.047 | 0.002021 | 0.209 | 0.2286 | 8.61e-08 | -0.01224 | -0.0677 | -0.07838 | 0.1979 | 0.2388 | -0.05243 | 0.01467 | 0.06111 | 1.41e-06 |
| Trigs | Triglyceride [Mass/volume] in Serum or Plasma | metabolic | mg/dL | 15431 | 19112 | 34543 | | 149.54 | 135.13 | 9.48e-55 | 3213.69 | 4942.28 | 2.01e-61 | -0.1719 | 0.1403 | -0.02804 | 4.17e-57 | 1.70e-56 | -0.3143 | 0.16 | 0.4517 | 1.08e-146 |
| Trnsfr | Transferrin [Mass/volume] in Serum or Plasma | metabolic | mg/dL | 1101 | 1070 | 2171 | | 214.85 | 229.33 | 1.98e-05 | 2219.19 | 1778.83 | 0.2456 | 0.1337 | -0.02438 | 0.1087 | 0.001852 | 0.002868 | 0.0737 | 0.0871 | 0.01051 | 0.4248 |
| TropnT | Troponin T.cardiac [Mass/volume] in Serum or Plasma | cardiovascular | ng/mL | 820 | 593 | 1413 | | 0.3168 | 0.2142 | 0.00494 | 0.1265 | 0.3544 | 0.00489 | 0.03461 | -0.1498 | -0.1151 | 0.514 | 0.5627 | -0.008188 | -0.06714 | -0.05922 | 0.9072 |
| TRPI | Troponin I.cardiac [Mass/volume] in Serum or Plasma | cardiovascular | ng/mL | 7073 | 5855 | 12928 | | 1.094 | 0.6202 | 9.33e-10 | 12.45 | 20.94 | 7.64e-05 | -0.05036 | -0.01526 | -0.06421 | 0.004123 | 0.006209 | -0.02258 | 0.01081 | 0.03345 | 0.2843 |
| TSH_3RD_GENERATION | TSH 3rd gen | endocrine | 10*-6.[iU]/mL | 325 | 831 | 1156 | | 1.847 | 1.707 | 0.1364 | 1.647 | 2.376 | 0.3483 | -0.06511 | 0.06864 | 0.00128 | 0.3172 | 0.3692 | 0.004278 | 0.02043 | 0.0163 | 0.973 |

| all_labs | full_name | group | selected_unit | male_n | female_n | total_n | sex_specific_lab | raw_median_male_mean | raw_median_female_mean | raw_median_p.value | raw_variance_female_mean | raw_variance_male_mean | raw_variance_p.value | std_mean_diff | median_male_mean | median_female_mean | median_p.value | median_p.value.fdr | std_variance_diff | variance_female_mean | variance_male_mean | variance_p.value |
|---|---|---|---|---|---|---|---|---|---|---|---|---|---|---|---|---|---|---|---|---|---|---|
| TSH | TSH | endocrine | | 16812 | 26093 | 42905 | | 2.008 | 1.869 | 1.36e-16 | 4.269 | 3.582 | 0.001159 | -0.03345 | 0.04683 | 0.01348 | 7.73e-04 | 0.001235 | 0.2655 | 0.0532 | -0.2272 | 4.17e-108 |
| TT4 | Thyroxine (T4) [Mass/volume] in Serum or Plasma | endocrine | ug/dL | 2286 | 3565 | 5851 | | 7.169 | 8.101 | 1.73e-61 | 3.392 | 2.061 | 3.59e-07 | 0.3906 | -0.2474 | 0.1386 | 2.52e-51 | 9.87e-51 | 0.1514 | 0.08241 | -0.05661 | 0.00173 |
| TTAIgA | Tissue transglutaminase IgA Ab [Units/volume] in Serum by Immunoassay | immune/inflammatory | | 294 | 820 | 1114 | | 5.226 | 4.764 | 0.07324 | 17.21 | 27.55 | 0.6826 | -0.1965 | 0.1695 | -0.031 | 0.005498 | 0.008036 | -0.2462 | 0.1097 | 0.3588 | 0.522 |
| TU-Pro | 24 hour urinary protein | urinary | g/L | 784 | 1324 | 2108 | | 0.6013 | 0.4235 | 0.004682 | 1 | 0.6572 | 0.4999 | -0.07105 | -0.08321 | -0.1554 | 0.1284 | 0.1604 | -0.6013 | -0.293 | 0.2959 | 1.48e-16 |
| TUCrea | Creatinine [Mass/volume] in Urine | kidney | g/L | 1069 | 1415 | 2484 | | 0.886 | 0.7072 | 2.38e-24 | 0.4421 | 0.0807 | 0.3432 | -0.5425 | 0.2681 | -0.2427 | 4.34e-42 | 1.51e-41 | 0.192 | -0.04005 | -0.2181 | 0.00944 |
| UABM | Urinary albumin | kidney | mg/dL | 723 | 585 | 1308 | | 111.45 | 87.41 | 0.07232 | 14846.6 | 63894.2 | 0.1068 | -0.2324 | 0.1344 | -0.08262 | 2.92e-05 | 5.19e-05 | -0.08356 | -0.1388 | -0.0453 | 0.5566 |
| UAlb | Albumin [Presence] in Urine | urinary | | 602 | 569 | 1171 | | 55.93 | 54.24 | 0.3764 | 871.82 | 873.34 | 0.9883 | 0.09924 | -0.2724 | -0.1712 | 0.08917 | 0.1139 | -0.01945 | 0.003617 | 0.02012 | 0.8662 |
| UGluc | Glucose urine dipstick | urinary | mg/dL | 2730 | 2253 | 4983 | | 235.63 | 236.31 | 0.9313 | 67581.9 | 62194.2 | 0.2594 | -0.09981 | 0.2753 | 0.1829 | 4.82e-04 | 7.85e-04 | -0.08343 | 0.1896 | 0.2699 | 0.0639 |
| UGrCst | Granular casts [#/area] in Urine sediment by Microscopy high power field | urinary | /[HPF] | 673 | 489 | 1162 | | 1.813 | 1.682 | 0.1907 | 1.861 | 2.736 | 0.3033 | -0.1215 | 0.1013 | -0.0104 | 0.04237 | 0.05616 | -0.001952 | 0.06497 | 0.06681 | 0.9905 |
| UHyCst | Hyaline casts in urine | urinary | /[HPF] | 3258 | 3554 | 6812 | | 2.094 | 2.073 | 0.7269 | 9.929 | 6.769 | 0.02673 | -0.09227 | 0.071 | -0.01381 | 1.34e-04 | 2.26e-04 | 0.007235 | -0.04 | -0.04735 | 0.8584 |
| UKeto | Ketones [Mass/volume] in Urine by Test strip | urinary | mg/dL | 2477 | 3704 | 6181 | | 25.25 | 30.26 | 1.49e-13 | 817.83 | 653.36 | 0.01281 | 0.03427 | -0.01775 | 0.01433 | 0.1724 | 0.2106 | -0.04206 | 0.09874 | 0.1367 | 0.382 |
| UpH | pH of Urine by Test strip | urinary | | 18624 | 24121 | 42745 | | 5.922 | 6.045 | 9.34e-75 | 0.5374 | 0.483 | 4.93e-18 | 0.1188 | -0.04072 | 0.06325 | 1.82e-34 | 6.08e-34 | 0.08537 | 0.1186 | 0.0455 | 7.82e-14 |
| URBC | Erythrocytes [#/area] in Urine sediment by Microscopy high power field | urinary | /[HPF] | 11084 | 15324 | 26408 | | 25.95 | 18.78 | 2.39e-06 | 21728.4 | 26543.1 | 0.042 | -0.1923 | 0.2296 | 0.06592 | 2.25e-54 | 8.96e-54 | -0.2327 | 0.0873 | 0.2983 | 5.75e-49 |
| UricA | Urate [Mass/volume] in Serum or Plasma | urinary | mg/dL | 6411 | 6711 | 13122 | | 6.355 | 5.296 | 4.86e-230 | 1.61 | 1.821 | 0.004547 | -0.5234 | 0.2868 | -0.2184 | 5.96e-205 | 4.56e-204 | -0.07089 | -0.05382 | 0.01793 | 0.002188 |
| UrobUr | Urobilinogen [Units/volume] in Urine by Test strip | urinary | [EIA'U] | 2412 | 3025 | 5437 | | 0.344 | 0.306 | 6.60e-05 | 0.08266 | 0.103 | 0.04047 | -0.2376 | 0.2989 | 0.09303 | 1.75e-18 | 4.57e-18 | -0.1808 | 0.06197 | 0.2282 | 3.19e-05 |
| all_labs | full_name | group | selected_unit | male_n | female_n | total_n | sex_specific_lab | raw_median_male_mean | raw_median_female_mean | raw_median_p.value | raw_variance_female_mean | raw_variance_male_mean | raw_variance_p.value | std_mean_diff | median_male_mean | median_female_mean | median_p.value | median_p.value.fdr | std_variance_diff | variance_female_mean | variance_male_mean | variance_p.value |
| UrPro | Protein [Mass/volume] in Urine by Test strip | urinary | mg/dL | 8932 | 9898 | 18830 | | 31.65 | 29.71 | 8.53e-07 | 756.19 | 749.07 | 0.7268 | 0.001427 | -0.2641 | -0.2627 | 0.9221 | 0.9372 | 0.04267 | 0.01007 | -0.03073 | 0.03213 |
| USpGr | Specific gravity of Urine by Test strip | urinary | | 18338 | 24468 | 42806 | | 1.017 | 1.016 | 1.48e-126 | 1.80e-04 | 1.20e-04 | 0.581 | -0.3304 | 0.1478 | -0.1305 | 9.76e-257 | 8.22e-256 | 0.2771 | -0.1376 | -0.3987 | 1.05e-133 |

| all_labs | full_name | group | selected_unit | male_n | female_n | total_n | sex_specific_lab | raw_median_male_mean | raw_median_female_mean | raw_median_p.value | raw_variance_female_mean | raw_variance_male_mean | raw_variance_p.value | std_mean_diff | median_male_mean | median_female_mean | median_p.value | median_p.value.fdr | std_variance_diff | variance_female_mean | variance_male_mean | variance_p.value |
|---|---|---|---|---|---|---|---|---|---|---|---|---|---|---|---|---|---|---|---|---|---|---|
| USqEpi | Epithelial cells.squamous [#/area] in Urine sediment by Microscopy high power field | urinary | /[HPF] | 3944 | 14841 | 18785 | | 1.646 | 3.524 | 0 | 22.25 | 5.709 | 1.35e-82 | 0.5103 | -0.3252 | 0.09848 | 2.16e-250 | 1.79e-249 | 0.4149 | 0.1691 | -0.1865 | 1.61e-82 |
| UTP | Total urine protein | urinary | g/L | 808 | 869 | 1677 | | 1.203 | 1.027 | 0.4178 | 43.1 | 14.93 | 0.499 | -0.2194 | 0.1463 | -0.05945 | 6.35e-06 | 1.16e-05 | -0.1776 | -0.04771 | 0.1344 | 0.1893 |
| UUrob | Urobilinogen [Presence] in Urine by Test strip | urinary | mg/dL | 4911 | 4995 | 9906 | | 2.405 | 2.137 | 8.00e-23 | 1.37 | 1.62 | 0.01373 | -0.2838 | 0.2038 | -0.07051 | 9.04e-46 | 3.35e-45 | -0.1866 | 0.02717 | 0.2126 | 9.26e-09 |
| UWBC | Leukocytes [#/area] in Urine sediment by Microscopy high power field | urinary | /[HPF] | 11007 | 16299 | 27306 | | 15.45 | 19.46 | 3.61e-08 | 7671.90 | 5997.78 | 9.37e-05 | 0.3524 | -0.3552 | 0.0235 | 2.13e-180 | 1.43e-179 | 0.2562 | 0.04898 | -0.2311 | 3.34e-60 |
| VA_BTPS_PPR | VA BTPS PPR | lung | | 1269 | 1227 | 2496 | | 83.96 | 85.23 | 0.09012 | 72.32 | 65.38 | 0.4769 | 0.034 | 0.07078 | 0.1038 | 0.3962 | 0.4455 | 0.07824 | 0.1318 | 0.05642 | 0.3395 |
| VA_BTPS_PRD | VA BTPS PRD | lung | | 1268 | 1226 | 2494 | | 6.893 | 5.031 | 0 | 0.005922 | 0.003107 | 0.1409 | -1.601 | 0.7906 | -0.7908 | 0 | 0 | -0.002069 | 0.07195 | 0.07381 | 0.98 |
| VA_BTPS_PRL | VA BTPS PRL | lung | | 1268 | 1226 | 2494 | | 5.327 | 4.945 | 6.62e-74 | 0.003941 | 0.1629 | 8.64e-11 | -0.7018 | 0.3399 | -0.3462 | 1.93e-73 | 8.58e-73 | -0.2869 | -0.1068 | 0.167 | 4.02e-04 |
| VA_BTPS_PRU | VA BTPS PRU | lung | | 1267 | 1222 | 2489 | | 8.452 | 5.117 | 0 | 0.01084 | 0.133 | 4.79e-10 | -1.616 | 0.7992 | -0.8036 | 0 | 0 | -0.1766 | -0.04108 | 0.1251 | 0.03 |
| VA_BTPS_ZBD | VA BTPS ZBD | lung | | 1268 | 1227 | 2495 | | 5.788 | 4.296 | 8.67e-177 | 0.1777 | 0.3112 | 2.77e-04 | -1.066 | 0.5772 | -0.4346 | 1.18e-183 | 8.14e-183 | -0.2115 | -0.06663 | 0.1489 | 0.009632 |
| VC_PPR | VC PPR | lung | | 1145 | 1146 | 2291 | | 81.44 | 83.83 | 0.002957 | 67.17 | 69.21 | 0.8543 | 0.108 | 0.00498 | 0.1115 | 0.009686 | 0.01375 | 0.04262 | 0.1199 | 0.07476 | 0.6358 |
| VC_PRD | VC PRD | lung | | 1144 | 1147 | 2291 | | 4.525 | 3.161 | 0 | 0.003116 | 0.006349 | 0.01219 | -1.613 | 0.8501 | -0.6445 | 0 | 0 | -0.03383 | 0.03538 | 0.06892 | 0.7048 |
| VC_PRL | VC PRL | lung | | 1144 | 1147 | 2291 | | 3.57 | 2.473 | 0 | 0.002492 | 0.003897 | 0.07567 | -1.609 | 0.8424 | -0.6174 | 0 | 0 | -0.277 | -0.04243 | 0.2116 | 0.002106 |
| VC_ZBD | VC ZBD | lung | | 1145 | 1147 | 2292 | | 3.682 | 2.652 | 2.38e-147 | 0.0668 | 0.1565 | 4.13e-05 | -1.112 | 0.6361 | -0.4204 | 1.07e-185 | 7.46e-185 | -0.3357 | -0.07125 | 0.2862 | 1.61e-04 |
| VD25 | Calcitriol [Mass/volume] in Serum or Plasma | metabolic | ng/mL | 949 | 2437 | 3386 | | 28.72 | 29.56 | 0.07903 | 180.97 | 136.90 | 0.0118 | 0.0671 | 0.03807 | 0.1012 | 0.0769 | 0.1003 | 0.1586 | 0.03725 | -0.1225 | 0.01023 |
| Vi-B12 | Vitamin B12 | metabolic | pg/mL | 9165 | 14107 | 23272 | | 590.20 | 586.00 | 0.5098 | 178155.1 | 149100.3 | 0.0478 | 0.03214 | -0.0995 | -0.06691 | 0.01726 | 0.02377 | 0.1835 | 0.0774 | -0.112 | 8.21e-23 |
| ViARet | Retinol [Mass/volume] in Serum or Plasma | metabolic | mg/L | 795 | 1396 | 2191 | | 0.4082 | 0.4667 | 1.93e-09 | 0.01699 | 0.01565 | 0.4986 | 0.2871 | -0.152 | 0.1521 | 3.10e-10 | 6.76e-10 | 0.1634 | 0.173 | -0.005188 | 0.005948 |
| VitaB6 | Pyridoxine [Mass/volume] in Serum or Plasma | metabolic | nmol/L | 836 | 1801 | 2637 | | 73.75 | 69.9 | 0.2007 | 3944.78 | 4508.66 | 0.3792 | -0.02234 | -0.05485 | -0.07781 | 0.6005 | 0.6502 | 0.04398 | -0.09385 | -0.1431 | 0.5623 |
| VLDL | VLDL | metabolic | | 1012 | 1142 | 2154 | | 26.84 | 25.16 | 0.003741 | 70.38 | 85.76 | 0.1106 | -0.09023 | 0.05445 | -0.03784 | 0.03704 | 0.05003 | -0.1109 | -0.02491 | 0.08992 | 0.08355 |
| WBC | Leukocytes [#/volume] in Blood by Automated count | immune/inflammatory | 10*3/uL | 26968 | 35673 | 62641 | | 8.07 | 7.996 | 0.006462 | 10.17 | 14.02 | 9.21e-13 | -0.02073 | -0.06697 | -0.08594 | 0.0106 | 0.01497 | -0.1016 | 0.08284 | 0.1766 | 9.85e-32 |
| WBCCnt | Leukocytes [#/volume] in Blood by Manual count | immune/inflammatory | | 5932 | 5812 | 11744 | | 112.74 | 111.51 | 9.74e-11 | 241.19 | 251.42 | 0.5433 | -0.1547 | 0.09426 | -0.06321 | 4.98e-17 | 1.26e-16 | -0.1069 | -0.04393 | 0.05845 | 1.28e-05 |
| WBCrt | Creatinine [Mass/volume] in Blood | blood | mg/dL | 4826 | 4458 | 9284 | | 1.236 | 1.007 | 8.05e-91 | 0.07336 | 0.08842 | 0.2235 | -0.6149 | 0.3712 | -0.2608 | 2.77e-200 | 2.09e-199 | -0.1909 | 0.04086 | 0.228 | 2.63e-08 |
| WBINR | INR | blood | | 1463 | 1253 | 2716 | | 1.083 | 1.034 | 2.41e-08 | 0.03333 | 0.03636 | 0.8043 | -0.2829 | 0.07363 | -0.2174 | 1.85e-13 | 4.33e-13 | 0.08198 | -0.09275 | -0.175 | 0.4406 |

| all_labs | full_name | group | selected_unit | male_n | female_n | total_n | sex_specific_lab | raw_median_male_mean | raw_median_female_mean | raw_median_p.value | raw_variance_female_mean | raw_variance_male_mean | raw_variance_p.value | std_mean_diff | median_male_mean | median_female_mean | median_p.value | median_p.value.fdr | std_variance_diff | variance_female_mean | variance_male_mean | variance_p.value |
|---|---|---|---|---|---|---|---|---|---|---|---|---|---|---|---|---|---|---|---|---|---|---|
| WBTrpI | Troponin I.cardiac [Mass/volume] in Blood | cardiovascular | ng/mL | 1106 | 906 | 2012 | | 0.1875 | 0.1498 | 0.06966 | 0.1769 | 0.2167 | 0.6369 | -0.1453 | 0.02832 | -0.116 | 0.001175 | 0.001861 | 0.2264 | 0.1572 | -0.06858 | 0.001462 |
| WEIGHT_SCALE | Weight scale | vitals | kg | 12994 | 19541 | 32535 | | 92.24 | 77.93 | 0 | 21.81 | 22.27 | 0.5225 | -0.7309 | 0.4052 | -0.3337 | 0 | 0 | -0.1425 | 0.2603 | 0.3778 | 2.22e-34 |
| Weight | Weight | vitals | kg | 26877 | 35804 | 62681 | | 91.92 | 76.98 | 0 | 961.15 | 1059.00 | 0.8371 | -0.7343 | 0.4092 | -0.3247 | 0 | 0 | -0.0983 | 0.1414 | 0.2384 | 1.24e-33 |
| Zinc | Zinc | metabolic | ug/dL | 681 | 1051 | 1732 | | 71.96 | 72.56 | 0.4714 | 167.35 | 213.42 | 0.09103 | 0.0397 | -0.03886 | -0.001229 | 0.4314 | 0.4797 | -0.2755 | -0.02939 | 0.211 | 0.004485 |

| variance_p.value.fdr | male_variance_n | female_variance_n |
|---|---|---|
| 0.2939 | 349 | 384 |
| 0.01172 | 3111 | 4311 |
| 0.7315 | 211 | 684 |
| 0.3516 | 1083 | 3053 |
| 0.3148 | 273 | 902 |
| 0.5093 | 1092 | 3070 |
| 0.6524 | 706 | 784 |
| 0.4428 | 706 | 781 |
| 0.761 | 52 | 42 |
| 0.7423 | 282 | 450 |
| 0.6132 | 262 | 426 |
| 0.5093 | 272 | 442 |
| 0.4754 | 283 | 452 |
| 0.8006 | 283 | 453 |
| 0.7921 | 279 | 449 |
| 0.9066 | 76 | 286 |
| 0.8912 | 131 | 470 |
| 0.9597 | 131 | 472 |
| 0.01238 | 198 | 296 |
| | 0 | 193 |
| 6.12e-06 | 1044 | 731 |
| 4.39e-15 | 20267 | 25739 |
| 8.33e-09 | 2203 | 2079 |
| 3.86e-04 | 1372 | 1225 |
| 0.4316 | 879 | 995 |
| 0.9718 | 133 | 204 |
| 1.82e-04 | 13445 | 17371 |
| 0.1672 | 665 | 455 |
| 0.1194 | 1791 | 1958 |
| 0.2517 | 23854 | 29531 |
| 1.92e-07 | 2537 | 3712 |
| 0.1116 | 230 | 329 |
| 0.7315 | 98 | 548 |
| 0.8327 | 74 | 270 |
| 0.07621 | 81 | 69 |
| 5.14e-05 | 19 | 111 |
| 5.81e-07 | 4401 | 4287 |
| 0.04153 | 1959 | 1705 |
| 0.1993 | 481 | 640 |
| 0.9064 | 6930 | 9309 |
| 0.6092 | 7161 | 9628 |
| 0.5911 | 2196 | 1934 |
| 6.09e-07 | 15033 | 18885 |
| 0.8481 | 2124 | 1859 |
| 0.761 | 249 | 364 |
| 0.8573 | 15055 | 18924 |
| 0.9066 | 27 | 38 |

| variance_p.value.fdr | male_variance_n | female_variance_n |
|---|---|---|
| 0.03577 | 707 | 784 |
| 0.2264 | 19 | 633 |
| 0.01458 | 1627 | 1413 |
| 0.1969 | 86 | 264 |
| 0.2519 | 91 | 269 |
| 0.8381 | 448 | 358 |
| 2.01e-07 | 12065 | 17908 |
| 1.51e-70 | 26030 | 35202 |
| 6.01e-14 | 4109 | 3670 |
| 7.79e-47 | 24162 | 30181 |
| 0.9066 | 390 | 381 |
| 0.397 | 239 | 775 |
| 0.8269 | 233 | 740 |

| variance_p.value.fdr | male_variance_n | female_variance_n |
|---|---|---|
| 0.001577 | 37 | 546 |
| 0.6817 | 23992 | 29988 |
| 0.4965 | 331 | 313 |
| 0.1388 | 6757 | 5246 |
| 1.47e-31 | 8289 | 8721 |
| 0.6483 | 651 | 224 |
| 5.93e-04 | 654 | 228 |
| 0.01744 | 651 | 222 |
| 4.24e-06 | 668 | 241 |
| 1.68e-05 | 651 | 221 |
| 0.441 | 668 | 240 |
| 0.8286 | 679 | 569 |
| 0.4067 | 52 | 38 |
| 0.04144 | 524 | 482 |
| 0.7921 | 12149 | 15013 |
| 9.11e-06 | 5441 | 4237 |
| 0.01614 | 2911 | 1957 |
| 8.32e-15 | 24136 | 30120 |
| 0.5721 | 346 | 344 |
| 0.9066 | 336 | 327 |
| 0.7476 | 227 | 241 |
| 0.564 | 465 | 500 |
| 7.10e-22 | 24186 | 30182 |
| 0.09927 | 2713 | 1800 |
| 0.06933 | 153 | 250 |
| 5.22e-05 | 1992 | 2196 |
| 0.1192 | 113 | 128 |
| 2.37e-25 | 6291 | 5607 |
| 3.47e-07 | 2780 | 2386 |
| 0.1216 | 390 | 383 |
| 0.07915 | 107 | 187 |
| 0.01548 | 118 | 166 |
| 0.06887 | 463 | 506 |
| 2.26e-126 | 24266 | 30734 |

| variance_p.value.fdr | male_variance_n | female_variance_n |
|---|---|---|
| 8.05e-04 | 332 | 341 |
| 5.84e-04 | 1701 | 1173 |
| 5.39e-68 | 4832 | 6576 |
| 0.7093 | 349 | 344 |
| 3.33e-10 | 4406 | 9274 |
| 0.6264 | 18 | 189 |
| 1.20e-16 | 25914 | 34499 |
| 0.00103 | 490 | 344 |
| 0.2888 | 218 | 263 |
| 8.09e-05 | 1380 | 1061 |
| 0.06202 | 316 | 293 |
| 5.50e-12 | 617 | 649 |
| 0.5048 | 310 | 289 |
| 0.1804 | 310 | 289 |
| 3.95e-12 | 622 | 653 |
| 0.06924 | 310 | 289 |
| 5.50e-12 | 617 | 649 |
| 0.5048 | 310 | 289 |
| 0.1804 | 310 | 289 |
| 6.70e-10 | 306 | 285 |
| 0.5911 | 303 | 341 |
| 8.24e-14 | 617 | 649 |
| 0.8627 | 622 | 653 |
| 0.9711 | 423 | 566 |
| 7.01e-35 | 26905 | 34555 |
| 0.9216 | 327 | 335 |

| variance_p.value.fdr | male_variance_n | female_variance_n |
|---|---|---|
| 0.8481 | 12 | 39 |
| 1.42e-10 | 6966 | 9359 |
| 1.15e-13 | 7550 | 10079 |
| 1.34e-18 | 11458 | 14553 |
| 1.24e-07 | 7650 | 8422 |
| 2.80e-17 | 11402 | 14516 |
| 0.936 | 322 | 496 |
| 1.27e-10 | 7809 | 8551 |
| 0.1116 | 43 | 938 |
| 0.7679 | 394 | 279 |
| 0.08599 | 1772 | 1512 |
| 0.07803 | 148 | 201 |
| 0.2551 | 495 | 612 |
| 0.02038 | 908 | 1119 |
| 0.009671 | 506 | 621 |
| 3.47e-07 | 905 | 1119 |
| 0.2867 | 70 | 92 |
| 0.2264 | 180 | 126 |
| 8.26e-54 | 4102 | 5983 |
| 0.6066 | 72 | 93 |
| 0.5727 | 507 | 629 |

| variance_p.value.fdr | male_variance_n | female_variance_n |
|---|---|---|
| 0.003998 | 71 | 93 |
| 0.3138 | 497 | 612 |
| 0.02236 | 497 | 612 |
| 0.003031 | 512 | 630 |
| 3.94e-12 | 497 | 612 |
| 0.4316 | 72 | 91 |
| 0.8481 | 72 | 94 |
| 0.9339 | 72 | 94 |
| 0.3786 | 510 | 631 |
| 0.8912 | 509 | 628 |
| 0.369 | 509 | 628 |
| 0.6483 | 510 | 630 |
| 0.02216 | 3969 | 2613 |
| 0.1609 | 889 | 583 |
| 0.5299 | 649 | 509 |
| 0.8534 | 314 | 267 |
| 0.754 | 362 | 276 |
| 0.8731 | 648 | 511 |
| 0.4383 | 935 | 766 |
| 0.8583 | 445 | 321 |
| 0.201 | 873 | 701 |
| 0.7714 | 264 | 240 |
| 0.6539 | 265 | 240 |
| 8.38e-05 | 265 | 240 |
| 0.07148 | 265 | 240 |
| 0.1127 | 259 | 235 |
| 0.1936 | 4308 | 4211 |
| 0.1068 | 4399 | 4285 |
| 0.03823 | 4097 | 4070 |
| 0.06516 | 4387 | 4255 |
| 0.001305 | 4399 | 4265 |
| 1.42e-07 | 137 | 1138 |
| 2.01e-18 | 5218 | 10308 |
| 0.08414 | 671 | 524 |
| 0.8853 | 497 | 612 |
| 0.3181 | 497 | 612 |
| 0.1128 | 512 | 631 |
| 5.10e-17 | 497 | 612 |
| 0.3295 | 66 | 85 |
| variance_p.value.fdr | male_variance_n | female_variance_n |
| 0.00184 | 705 | 778 |
| 0.08058 | 1376 | 1023 |
| 0.9196 | 301 | 417 |
| 1.12e-05 | 11262 | 10652 |
| 1.13e-80 | 24173 | 30279 |
| 0.6483 | 5898 | 4129 |
|  | 0 | 1140 |
| 0.05531 | 405 | 369 |

| variance_p.value.fdr | male_variance_n | female_variance_n |
|---|---|---|
| 0.8573 | 297 | 272 |
| 0.01275 | 554 | 466 |
| 0.03226 | 7551 | 5779 |
| 8.38e-05 | 5067 | 3536 |
| 9.66e-172 | 11677 | 14413 |
| 0.1989 | 4414 | 4291 |
| 8.42e-07 | 22571 | 30966 |
| 0.01517 | 11301 | 16950 |
| 0.004472 | 328 | 476 |
| 0.001023 | 331 | 473 |
| 1.31e-211 | 23950 | 31311 |
| 0.05706 | 515 | 395 |
| 9.46e-24 | 7274 | 8426 |
| 0.001754 | 63 | 102 |
| 0.1404 | 4290 | 4186 |
| 0.6908 | 4278 | 4163 |
| 0.9718 | 30 | 50 |
| 0.03757 | 14101 | 14099 |
| 0.09263 | 130 | 143 |
| 0.7116 | 272 | 252 |
| 0.04541 | 272 | 251 |
| 0.001697 | 266 | 248 |
| 0.9237 | 951 | 969 |
| 0.748 | 758 | 1812 |
| 1.39e-09 | 14136 | 18072 |
| 0.4885 | 126 | 218 |
| 0.511 | 649 | 762 |
| 0.1135 | 248 | 321 |
| 2.42e-22 | 14131 | 18092 |
| | 0 | 308 |
| | 0 | 308 |
| | 0 | 314 |
| | 0 | 313 |
| | 0 | 316 |
| | 0 | 303 |
| | 0 | 316 |
| | 0 | 306 |
| | 0 | 316 |
| | 0 | 302 |
| | 0 | 312 |
| | 0 | 303 |
| 4.45e-11 | 14044 | 14071 |
| 4.86e-04 | 92 | 437 |
| 0.07558 | 751 | 757 |
| 0.5135 | 1038 | 1120 |
| 0.6817 | 5590 | 7421 |
| 0.001137 | 6259 | 4407 |
| 5.84e-04 | 24241 | 30236 |

| variance_p.value.fdr | male_variance_n | female_variance_n |
|---|---|---|
| 0.8583 | 208 | 280 |
| 0.564 | 199 | 271 |
| 0.07558 | 3546 | 3253 |
| 0.009379 | 6332 | 4527 |

| variance_p.value.fdr | male_variance_n | female_variance_n |
|---|---|---|
| 0.2388 | 207 | 278 |
| 0.9123 | 3614 | 3641 |
| 0.1635 | 326 | 325 |
| 0.3295 | 514 | 479 |
| 0.03393 | 11249 | 13887 |
| 7.97e-07 | 899 | 1357 |
| 2.67e-05 | 206 | 345 |
| 0.397 | 2475 | 2960 |
| 0.8286 | 1071 | 1197 |
| 0.6924 | 1376 | 1861 |
| 0.03252 | 1383 | 1870 |
| 0.6663 | 7791 | 10402 |
| 1.18e-18 | 7219 | 9622 |
| 1.26e-04 | 3115 | 2831 |
| 0.06535 | 14989 | 18870 |
| 2.57e-04 | 385 | 583 |
| 0.1607 | 3213 | 2940 |
| 0.3181 | 15010 | 18876 |
| 0.002185 | 5331 | 4067 |
| 0.002365 | 23938 | 31294 |
| 3.70e-31 | 23886 | 31192 |
| 7.97e-185 | 23952 | 31309 |
| 0.01053 | 1650 | 1407 |
| 0.8731 | 431 | 349 |
| 0.02061 | 2014 | 1943 |
| 0.002063 | 2213 | 2066 |
| 0.1679 | 732 | 601 |
| 4.37e-13 | 14048 | 14084 |
| 0.8853 | 2103 | 2060 |
| 0.6483 | 2699 | 1796 |
| 1.49e-05 | 8243 | 8208 |
| 0.01602 | 147 | 150 |
| 6.63e-32 | 7189 | 9618 |
| 0.2412 | 3068 | 2758 |
| 1.91e-79 | 15045 | 18904 |
| 5.32e-24 | 7760 | 10415 |
| 0.02216 | 383 | 578 |
| 3.24e-04 | 3103 | 2784 |
| 1.37e-54 | 15071 | 18945 |
| 1.39e-06 | 17241 | 22860 |
| 0.8058 | 2291 | 2150 |
| 0.369 | 5775 | 4089 |
| 9.60e-30 | 24181 | 30176 |

| variance_p.value.fdr | male_variance_n | female_variance_n |
|---|---|---|
| 0.7116 | 1172 | 769 |
| 0.002375 | 7922 | 10526 |
| 0.01114 | 3167 | 2857 |
| 7.09e-25 | 15080 | 18965 |
| 0.9123 | 3144 | 2828 |
| 3.27e-04 | 15074 | 18949 |
| 0.1004 | 1213 | 906 |
| 3.44e-17 | 10246 | 13423 |
| 6.01e-67 | 20169 | 25769 |
| 0.09919 | 572 | 482 |
| 0.7315 | 573 | 471 |
| 1.14e-13 | 1800 | 2938 |
| 4.77e-15 | 7193 | 9618 |
| 5.65e-30 | 17098 | 21744 |
| 0.6092 | 8992 | 8951 |
| 0.04155 | 4859 | 3313 |
| 9.74e-113 | 22296 | 27837 |
| variance_p.value.fdr | male_variance_n | female_variance_n |
| 0.09286 | 3589 | 2690 |
| 0.6534 | 2391 | 1863 |
|  | 0 | 290 |
|  | 0 | 290 |
| 0.9066 | 571 | 573 |
| 7.64e-17 | 4397 | 4280 |
| 6.23e-06 | 3976 | 3984 |
| 3.86e-04 | 14074 | 14078 |
| 0.9335 | 13538 | 12695 |
| 0.01154 | 121 | 265 |
| 0.4885 | 1952 | 1700 |
| 0.4917 | 44 | 75 |
| 0.01856 | 6844 | 5043 |
| 3.02e-05 | 2797 | 2679 |
| 2.07e-145 | 24595 | 32259 |
| 0.2625 | 72 | 94 |
| 0.02236 | 74 | 95 |
| 0.9703 | 512 | 632 |
| 0.04227 | 513 | 633 |
| 1.53e-07 | 458 | 588 |
| 4.89e-18 | 509 | 630 |
| 0.8945 | 70 | 90 |
| 0.5172 | 2789 | 2404 |
| 3.91e-11 | 7193 | 6617 |
| 0.003368 | 72 | 94 |
| 0.3666 | 72 | 95 |
| 0.4583 | 512 | 627 |
| 0.1667 | 512 | 630 |
| 2.00e-16 | 510 | 629 |
| 0.6321 | 70 | 89 |

| variance_p.value.fdr | male_variance_n | female_variance_n |
|---|---|---|
| 6.64e-11 | 23916 | 31226 |
| 0.8118 | 3688 | 2388 |
| 0.7315 | 1919 | 1670 |
| 5.81e-07 | 6872 | 5082 |
| 0.1149 | 1482 | 1468 |
| 1.57e-09 | 4158 | 4136 |
| 6.00e-26 | 14098 | 14098 |
| 2.17e-04 | 125 | 263 |
| 0.258 | 1806 | 1547 |
| 0.7745 | 587 | 574 |
| | 1 | 444 |
| 1.17e-22 | 232 | 557 |
| 0.6779 | 761 | 625 |
| | 6667 | 0 |
| 0.9381 | 13600 | 12782 |
| 0.876 | 11146 | 10172 |
| 5.75e-05 | 6140 | 5458 |
| 2.45e-10 | 10983 | 15722 |
| 6.81e-173 | 25334 | 33419 |
| 3.03e-06 | 12055 | 17661 |
| 0.7788 | 4408 | 4288 |
| 0.398 | 228 | 274 |
| 0.1989 | 4408 | 4286 |
| 0.783 | 4326 | 4231 |
| 3.36e-10 | 14078 | 14092 |
| 0.8568 | 4402 | 4287 |
| 7.47e-54 | 14094 | 14093 |
| 4.63e-23 | 14096 | 14097 |
| 0.6817 | 548 | 1326 |
| 0.1082 | 276 | 352 |
| variance_p.value.fdr | male_variance_n | female_variance_n |
| 0.01445 | 4405 | 4286 |
| 0.6437 | 4405 | 4287 |
| 0.3522 | 495 | 603 |
| 0.3666 | 242 | 303 |
| 0.6032 | 242 | 303 |
| 0.008556 | 5422 | 6960 |
| 2.09e-11 | 5997 | 7908 |
| 0.6467 | 907 | 1225 |
| 0.4583 | 655 | 709 |
| 0.7566 | 1290 | 1622 |
| 0.2762 | 5664 | 7324 |
| 0.7411 | 1157 | 865 |
| 0.102 | 527 | 559 |
| 1.19e-09 | 5690 | 7457 |
| 0.1679 | 5624 | 7283 |
| 0.7315 | 1053 | 1354 |
| 1.05e-06 | 10680 | 15737 |

| variance_p.value.fdr | male_variance_n | female_variance_n |
|---|---|---|
| 0.4386 | 620 | 498 |
| 0.5093 | 415 | 424 |
| 0.118 | 395 | 409 |
| 0.2511 | 410 | 425 |
| 0.258 | 361 | 385 |
| 0.2022 | 138 | 128 |
| 1.42e-07 | 6708 | 8727 |
| 0.02203 | 11302 | 16959 |
| 0.01972 | 11301 | 16955 |
| 0.01972 | 11301 | 16955 |
| 0.1933 | 4486 | 5650 |
| 9.47e-15 | 3314 | 3660 |
| 0.5812 | 162 | 348 |
| 1.06e-27 | 14098 | 14098 |
| 4.25e-214 | 23831 | 31059 |
| 0.08058 | 415 | 698 |
| 0.002861 | 1336 | 1968 |
| 1.12e-08 | 23932 | 31283 |
| 0.3709 | 20634 | 26238 |
| 0.7745 | 103 | 175 |
| 2.56e-42 | 10275 | 14879 |
| 4.13e-144 | 21568 | 27449 |
| 5.30e-06 | 1394 | 1581 |
| 0.6483 | 790 | 798 |
| 0.007795 | 1420 | 1583 |
| 0.9718 | 331 | 625 |
| 2.88e-06 | 4411 | 4291 |
| 0.6831 | 224 | 270 |
| 0.9718 | 225 | 271 |
| 0.2946 | 265 | 240 |
| 0.02705 | 265 | 238 |
| 0.6287 | 223 | 271 |
| 0.1968 | 225 | 271 |
| 9.12e-05 | 225 | 271 |
| 0.8912 | 263 | 240 |
| 0.7025 | 204 | 169 |
| 0.1989 | 225 | 271 |
| 0.08056 | 87 | 329 |
| 3.15e-12 | 4281 | 4122 |
| 0.9718 | 10 | 46 |
| 1.72e-12 | 4429 | 7356 |
| 0.1667 | 1092 | 1680 |
| 0.9216 | 203 | 147 |
| variance_p.value.fdr | male_variance_n | female_variance_n |
| 1.33e-87 | 13359 | 17288 |
| 2.36e-110 | 13482 | 17436 |
| 0.9843 | 15 | 94 |
| 0.2821 | 13 | 76 |

| variance_p.value.fdr | male_variance_n | female_variance_n |
|---|---|---|
| 0.003488 | 14102 | 14099 |
| 0.1916 | 44 | 149 |
| 0.8853 | 144 | 274 |
| 0.9278 | 146 | 154 |
| 0.4803 | 1316 | 1114 |
| 0.5269 | 429 | 467 |
| 2.17e-16 | 2098 | 1905 |
| 0.1446 | 585 | 443 |
| 3.21e-17 | 5067 | 5072 |
| 0.7485 | 169 | 127 |
| 0.1194 | 25913 | 34499 |
| 5.85e-08 | 4328 | 4192 |
| 0.9066 | 4384 | 4273 |
| 2.41e-05 | 14102 | 14099 |
| 0.002671 | 203 | 310 |
| 1.34e-216 | 20981 | 26502 |
| 1.22e-04 | 1288 | 961 |
| 6.01e-13 | 23517 | 30373 |
| 5.84e-04 | 10679 | 15737 |
| 1.98e-19 | 14102 | 14099 |
| 0.01149 | 1005 | 23 |
| 1.43e-40 | 1544 | 197 |
| 0.3786 | 88 | 63 |
| 0.4583 | 213 | 594 |
| 0.118 | 4144 | 2678 |
| 0.2355 | 112 | 295 |
| 4.95e-05 | 3927 | 5393 |
| 0.4316 | 7 | 37 |
| 0.1082 | 225 | 269 |
| 0.2909 | 225 | 271 |
| 3.67e-07 | 223 | 197 |
| 0.2277 | 223 | 197 |
| 0.2975 | 225 | 271 |
| 2.91e-05 | 903 | 1546 |
| 1.97e-14 | 12879 | 16590 |
| 5.74e-06 | 15277 | 19168 |
| 5.92e-145 | 12083 | 14828 |
| 0.5675 | 244 | 230 |
| 0.9339 | 491 | 332 |
| 0.4147 | 5075 | 3904 |
| 0.979 | 89 | 315 |
| 1.37e-106 | 11225 | 19066 |
| 0.004739 | 647 | 1008 |
| 0.6539 | 7 | 41 |
| 1.14e-15 | 266 | 402 |
| 0.02216 | 349 | 394 |
| 0.6843 | 113 | 82 |
| 0.9105 | 166 | 137 |

| variance_p.value.fdr | male_variance_n | female_variance_n |
|---|---|---|
| 0.118 | 1244 | 872 |
| 0.9905 | 99 | 58 |
| 0.9066 | 1161 | 1269 |
| 0.5174 | 653 | 1049 |
| 5.14e-13 | 13418 | 17417 |
| 1.05e-47 | 6748 | 9204 |
| 0.005926 | 3979 | 3507 |
| 1.11e-04 | 1016 | 1089 |
| **variance_p.value.fdr** | **male_variance_n** | **female_variance_n** |
| 0.06769 | 5001 | 5087 |
| 4.69e-132 | 13090 | 18000 |
| 4.68e-81 | 1605 | 8621 |
| 0.2946 | 106 | 115 |
| 4.66e-08 | 1909 | 1871 |
| 7.16e-59 | 6665 | 10146 |
| 0.4782 | 310 | 290 |
| 0.984 | 310 | 289 |
| 0.001223 | 309 | 288 |
| 0.06403 | 310 | 288 |
| 0.02236 | 306 | 285 |
| 0.7485 | 261 | 235 |
| 0.8025 | 262 | 238 |
| 0.005735 | 262 | 237 |
| 5.19e-04 | 259 | 235 |
| 0.02346 | 333 | 928 |
| 8.09e-22 | 4668 | 7686 |
| 0.01489 | 452 | 846 |
| 0.6896 | 255 | 729 |
| 0.1498 | 501 | 474 |
| 1.31e-30 | 23952 | 31286 |
| 4.77e-05 | 3448 | 3239 |
| 1.28e-07 | 1935 | 1531 |
| 0.5824 | 198 | 158 |
| 0.00405 | 441 | 340 |
| 3.42e-33 | 11884 | 17705 |
| 1.86e-32 | 26530 | 35372 |
| 0.01158 | 160 | 282 |

#### STable3: Count of variants and independent loci identified as associated with per-person median trait values in males, females, or both sexes and count in EMBL GWAS catalog

*Notes: significant=P<5e-8, suggestive=P<1e-6, noHLA indicates variants within HLA were excluded*

508 rows × 45 columns

STable3 — Columns 1–23 of 45

| all_labs | full_name | group | male.significant.variants | female.significant.variants | both.significant.variants | male.only.significant.variants | female.only.significant.variants | male.annotated.genes.significant.variants | female.annotated.genes.significant.variants | male.suggestive.variants | female.suggestive.variants | both.suggestive.variants | male.only.suggestive.variants | female.only.suggestive.variants | female.significant.x.variants | male.significant.x.variants | female.suggestive.x.variants | male.suggestive.x.variants | n_loci | n_loci_noHLA | P1e6_both | P1e2_both |
|---|---|---|---|---|---|---|---|---|---|---|---|---|---|---|---|---|---|---|---|---|---|---|
| #WBC | # of WBC’s | blood | 0 | 0 | 0 | 0 | 0 | | | 23 | 0 | 0 | 23 | 0 | 0 | 0 | 0 | 0 | 0 | | 0 | 0 |
| %SAT | % saturation | other | 111 | 337 | 102 | 9 | 235 | LRRC16A; SCGN; HIST1H2AA; SLC17A1; SLC17A3; SLC17A2; TRIM38; U91328.21; U91328.19; U91328.22; HIST1H3A; HIST1H4A; HIST1H4B; HIST1H3B; HIST1H2BB; U91328.2; HIST1H3C; HIST1H1C; HFE; HIST1H4C; HIST1H1T; HIST1H2BC; HIST1H3G; HIST1H3PS1; RNU6-1259P; BTN1A1; VN1R13P; MIR3143; TMPRSS6 | RP11-191A15.4; RP3-522P13.3; RP3-522P13.1; RP3-522P13.2; LRRC16A; SCGN; HIST1H2AA; SLC17A1; SLC17A3; SLC17A2; TRIM38; U91328.21; U91328.19; U91328.22; HIST1H3A; HIST1H4A; HIST1H4B; HIST1H3B; HIST1H2AB; HIST1H2BB; U91328.2; HIST1H3C; HIST1H1C; HFE; HIST1H4C; HIST1H1T; HIST1H2BC; HIST1H3G; HIST1H3PS1; MPST; TMPRSS6 | 128 | 435 | 118 | 10 | 317 | 0 | 0 | 0 | 0 | 15 | 15 | 10 | 13 |
| 125DHy | Calcitriol [Mass/volume] in Serum or Plasma | metabolic | 0 | 0 | 0 | 0 | 0 | | | 0 | 0 | 0 | 0 | 0 | 0 | 0 | 0 | 0 | 0 | | 0 | 0 |
| 25DTot | 25-Hydroxyvitamin D3+25-Hydroxyvitamin D2 [Mass/volume] in Serum or Plasma | metabolic | 6 | 172 | 6 | 0 | 166 | GC | GC; RP11-545L5.1; RRAS2; RP11-140L24.3; COPB1; PSMA1; PDE3B; CYP2R1; CALCB | 36 | 245 | 24 | 12 | 221 | 0 | 0 | 0 | 0 | 11 | 11 | 2 | 11 |
| 25HyD2 | 25-Hydroxyvitamin D2 [Mass/volume] in Serum or Plasma | metabolic | 0 | 0 | 0 | 0 | 0 | | | 3 | 1 | 0 | 3 | 1 | 0 | 0 | 0 | 0 | 0 | | 0 | 0 |
| 25HyD3 | 25-Hydroxyvitamin D3 [Mass/volume] in Serum or Plasma | metabolic | 11 | 83 | 11 | 0 | 72 | GC | GC; RP11-545L5.1; PDE3B; CYP2R1 | 31 | 154 | 28 | 3 | 126 | 0 | 0 | 0 | 0 | 6 | 6 | 3 | 6 |

| all_labs | full_name | group | male.significant.variants | female.significant.variants | both.significant.variants | male.only.significant.variants | female.only.significant.variants | male.annotated.genes.significant.variants | female.annotated.genes.significant.variants | male.suggestive.variants | female.suggestive.variants | both.suggestive.variants | male.only.suggestive.variants | female.only.suggestive.variants | female.significant.x.variants | male.significant.x.variants | female.suggestive.x.variants | male.suggestive.x.variants | n_loci | n_loci_noHLA | P1e6_both | P1e2_both |
|---|---|---|---|---|---|---|---|---|---|---|---|---|---|---|---|---|---|---|---|---|---|---|
| A-1Gg_ | Alpha 1 globulin [Mass/volume] in Serum or Plasma by Electrophoresis | liver | 0 | 0 | 0 | 0 | 0 | | | 0 | 1 | 0 | 0 | 1 | 0 | 0 | 0 | 0 | 0 | | 0 | 0 |
| A-2Gg_ | Alpha 2 globulin [Mass/volume] in Serum or Plasma by Electrophoresis | liver | 12 | 0 | 0 | 12 | 0 | PKD1L3; DHODH; TXNL4B; DHX38; PMFBP1 | | 22 | 6 | 0 | 22 | 6 | 0 | 0 | 0 | 0 | 2 | 2 | 0 | 2 |
| A1Anti | Alpha 1 antitrypsin [Mass/volume] in Serum or Plasma | other | 5 | 5 | 5 | 0 | 0 | SERPINA6; SERPINA2P; SERPINA1 | SERPINA6; SERPINA2P; SERPINA1 | 12 | 6 | 6 | 6 | 0 | 0 | 0 | 0 | 0 | 2 | 2 | 2 | 2 |
| ABS_BASO_CT | Basophils [#/volume] in Blood by Automated count | blood | 0 | 0 | 0 | 0 | 0 | | | 0 | 2 | 0 | 0 | 2 | 0 | 0 | 0 | 0 | 0 | | 0 | 0 |
| ABS_EOS_CT | Eosinophils [#/volume] in Blood by Automated count | blood | 0 | 0 | 0 | 0 | 0 | | | 0 | 1 | 0 | 0 | 1 | 0 | 0 | 0 | 0 | 0 | | 0 | 0 |
| ABS_IMMATURE_GRANULOCYTES_CT | Immature granulocytes [#/volume] in Blood by Automated count | blood | 0 | 0 | 0 | 0 | 0 | | | 0 | 5 | 0 | 0 | 5 | 0 | 0 | 0 | 0 | 0 | | 0 | 0 |
| ABS_LYMPH_CT | Lymphocytes [#/volume] in Blood by Automated count | blood | 0 | 0 | 0 | 0 | 0 | | | 0 | 0 | 0 | 0 | 0 | 0 | 0 | 0 | 0 | 0 | | 0 | 0 |

| all_labs | full_name | group | male.significant.variants | female.significant.variants | both.significant.variants | male.only.significant.variants | female.only.significant.variants | male.annotated.genes.significant.variants | female.annotated.genes.significant.variants | male.suggestive.variants | female.suggestive.variants | both.suggestive.variants | male.only.suggestive.variants | female.only.suggestive.variants | female.significant.x.variants | male.significant.x.variants | female.suggestive.x.variants | male.suggestive.x.variants | n_loci | n_loci_noHLA | P1e6_both | P1e2_both |
|---|---|---|---|---|---|---|---|---|---|---|---|---|---|---|---|---|---|---|---|---|---|---|
| ABS_MONO_CT | Monocytes [#/volume] in Blood by Automated count | blood | 0 | 0 | 0 | 0 | 0 | | | 0 | 0 | 0 | 0 | 0 | 0 | 0 | 0 | 0 | 0 | | 0 | 0 |
| ABS_NEUTRO_CT | Neutrophils [#/volume] in Blood by Automated count | blood | 0 | 0 | 0 | 0 | 0 | | | 0 | 7 | 0 | 0 | 7 | 0 | 0 | 0 | 0 | 0 | | 0 | 0 |
| ACAIgA | Cardiolipin IgA Ab [Units/volume] in Serum by Immunoassay | immune / inflammatory | 0 | 0 | 0 | 0 | 0 | | | 0 | 2 | 0 | 0 | 2 | 0 | 0 | 0 | 0 | 0 | | 0 | 0 |
| ACAIgG | Cardiolipin IgG Ab [Units/volume] in Serum by Immunoassay | immune / inflammatory | 0 | 0 | 0 | 0 | 0 | | | 0 | 1 | 0 | 0 | 1 | 0 | 0 | 0 | 0 | 0 | | 0 | 0 |
| ACAIgM | Cardiolipin IgM Ab [Units/volume] in Serum by Immunoassay | immune / inflammatory | 0 | 0 | 0 | 0 | 0 | | | 0 | 0 | 0 | 0 | 0 | 0 | 0 | 0 | 0 | 0 | | 0 | 0 |
| ACTH | Corticotropin [Mass/volume] in Plasma | endocrine | 1 | 0 | 0 | 1 | 0 | EPHB1 | | 6 | 0 | 0 | 6 | 0 | 0 | 0 | 0 | 0 | 1 | 1 | 0 | 0 |
| AFP-Ob | Maternal Alpha-1-Fetoprotein [Mass/volume] in Serum or Plasma | ob / gyn | | 0 | | | | | | | 7 | | | | 0 | | 0 | | 0 | | 0 | 0 |

| all_labs | full_name | group | male.significant.variants | female.significant.variants | both.significant.variants | male.only.significant.variants | female.only.significant.variants | male.annotated.genes.significant.variants | female.annotated.genes.significant.variants | male.suggestive.variants | female.suggestive.variants | both.suggestive.variants | male.only.suggestive.variants | female.only.suggestive.variants | female.significant.x.variants | male.significant.x.variants | female.suggestive.x.variants | male.suggestive.x.variants | n_loci | n_loci_noHLA | P1e6_both | P1e2_both |
|---|---|---|---|---|---|---|---|---|---|---|---|---|---|---|---|---|---|---|---|---|---|---|
| AFPTum | Alpha-1-fetoprotein.tumor marker [Mass/volume] in Serum or Plasma | cancer | 0 | 0 | 0 | 0 | 0 | | | 1 | 4 | 0 | 1 | 4 | 0 | 0 | 0 | 0 | 0 | | 0 | 0 |
| Alb | Albumin [Mass/volume] in Serum or Plasma | liver | 24 | 0 | 0 | 24 | 0 | PPM1G; GCKR; HLA-C; DHFRP2; RNU6-283P; AL671883.1; FGFR3P1; ZDHHC20P2 | | 193 | 77 | 17 | 176 | 60 | 0 | 0 | 0 | 0 | 4 | 1 | 1 | 4 |
| AlbCnc | Albumin [Presence] in Urine | urinary | 0 | 0 | 0 | 0 | 0 | | | 0 | 1 | 0 | 0 | 1 | 0 | 0 | 0 | 0 | 0 | | 0 | 0 |
| AlbCre | Microalbumin/Creatinine [Mass Ratio] in 24 hour Urine | liver | 0 | 0 | 0 | 0 | 0 | | | 1 | 2 | 0 | 1 | 2 | 0 | 0 | 0 | 0 | 0 | | 0 | 0 |
| Album | Albumin [Mass/volume] in Serum or Plasma | liver | 0 | 0 | 0 | 0 | 0 | | | 5 | 11 | 0 | 5 | 11 | 0 | 0 | 0 | 0 | 0 | | 0 | 0 |
| Aldo-P | Aldosterone [Mass/volume] in Serum or Plasma | endocrine | 0 | 0 | 0 | 0 | 0 | | | 0 | 1 | 0 | 0 | 1 | 0 | 0 | 0 | 0 | 0 | | 0 | 0 |
| AlkP | Alkaline phosphatase [Enzymatic activity/volume] in Serum or Plasma | liver | 469 | 930 | 409 | 60 | 521 | NBPF3; ALPL; RP11-63N8.3; AL359815.1; RAP1GAP; DCDC2; MRS2; GPLD1; ALDH5A1; RP11-115J16.1; GBGT1; ABO; LCN1P2; C9orf96; ADAMTS13; SLC2A6; B4GALNT3; FUT2; MAMSTR; RASIP1; IZUMO1 | ECE1; PPP1R11P1; RP11-293F5.8; NBPF3; ALPL; RP11-63N8.3; AL359815.1; RAP1GAP; MRS2; GPLD1; ALDH5A1; KIAA0319; RP11-115J16.1; GBGT1; OBP2B; LCN1P1; ABO; LCN1P2; SURF6; C9orf96; ADAMTS13; SLC2A6; NRBF2; JMJD1C; JMJD1C-AS1; RP11-351O1.3; REEP3; NYNRIN; ASGR1; FUT2; IZUMO1; AL035252.1; PYGB; ABHD12; GINS1; NINL | 652 | 1145 | 549 | 103 | 596 | 0 | 0 | 0 | 0 | 57 | 57 | 38 | 53 |
| Amm | Ammonia [Moles/volume] in Plasma | metabolic | 0 | 0 | 0 | 0 | 0 | | | 0 | 0 | 0 | 0 | 0 | 0 | 0 | 0 | 0 | 0 | | 0 | 0 |

| all_labs | full_name | group | male.significant.variants | female.significant.variants | both.significant.variants | male.only.significant.variants | female.only.significant.variants | male.annotated.genes.significant.variants | female.annotated.genes.significant.variants | male.suggestive.variants | female.suggestive.variants | both.suggestive.variants | male.only.suggestive.variants | female.only.suggestive.variants | female.significant.x.variants | male.significant.x.variants | female.suggestive.x.variants | male.suggestive.x.variants | n_loci | n_loci_noHLA | P1e6_both | P1e2_both |
|---|---|---|---|---|---|---|---|---|---|---|---|---|---|---|---|---|---|---|---|---|---|---|
| Amyl | Amylase [Enzymatic activity/volume] in Serum or Plasma | metabolic | 3 | 9 | 3 | 0 | 6 | RP11-153F1.1; AMY1C | RP11-153F1.1; RNPC3; AMY2B; AMY1C | 64 | 34 | 3 | 61 | 31 | 0 | 0 | 0 | 0 | 2 | 2 | 1 | 2 |
| AN-GAP | Anion gap in Serum or Plasma | metabolic | 36 | 13 | 9 | 27 | 4 | GCKR; AC109829.1; C2orf16; ZNF512 | PPM1G; GCKR; AC074091.13; MRPL33; COL4A2 | 36 | 15 | 9 | 27 | 6 | 0 | 0 | 0 | 0 | 4 | 4 | 1 | 3 |
| ANESTHESIA PULSE | Heart rate by pulse oximetry | vitals | 0 | 0 | 0 | 0 | 0 | | | 9 | 1 | 0 | 9 | 1 | 0 | 0 | 0 | 0 | 0 | | 0 | 0 |
| ANION_GAP | Anion gap | metabolic | 0 | 0 | 0 | 0 | 0 | | | 0 | 0 | 0 | 0 | 0 | 0 | 0 | 0 | 0 | 0 | | 0 | 0 |
| AntDNA | DNA double strand Ab [Units/volume] in Serum | immune / inflammatory | 0 | 0 | 0 | 0 | 0 | | | 0 | 0 | 0 | 0 | 0 | 0 | 0 | 0 | 0 | 0 | | 0 | 0 |
| APBone | Alkaline phosphatase bone | other | 0 | 0 | 0 | 0 | 0 | | | 0 | 0 | 0 | 0 | 0 | 0 | 0 | 0 | 0 | 0 | | 0 | 0 |
| AT3 | Antithrombin actual/normal in Platelet poor plasma by Chromogenic method | blood | 0 | 0 | 0 | 0 | 0 | | | 2 | 0 | 0 | 2 | 0 | 0 | 0 | 0 | 0 | 0 | | 0 | 0 |
| AThyP | Thyroperoxidase Ab | immune / inflammatory | 0 | 0 | 0 | 0 | 0 | | | 0 | 2 | 0 | 0 | 2 | 0 | 0 | 0 | 0 | 0 | | 0 | 0 |
| ATRIAL RATE | P wave Atrium by EKG | cardiovascular | 0 | 0 | 0 | 0 | 0 | | | 0 | 26 | 0 | 0 | 26 | 0 | 0 | 0 | 0 | 0 | | 0 | 0 |
| AtyLym | Variant lymphocytes/100 leukocytes in Blood by Manual count | immune / inflammatory | 0 | 0 | 0 | 0 | 0 | | | 1 | 1 | 0 | 1 | 1 | 0 | 0 | 1 | 0 | 0 | | 0 | 0 |

| all_labs | full_name | group | male.significant.variants | female.significant.variants | both.significant.variants | male.only.significant.variants | female.only.significant.variants | male.annotated.genes.significant.variants | female.annotated.genes.significant.variants | male.suggestive.variants | female.suggestive.variants | both.suggestive.variants | male.only.suggestive.variants | female.only.suggestive.variants | female.significant.x.variants | male.significant.x.variants | female.suggestive.x.variants | male.suggestive.x.variants | n_loci | n_loci_noHLA | P1e6_both | P1e2_both |
|---|---|---|---|---|---|---|---|---|---|---|---|---|---|---|---|---|---|---|---|---|---|---|
| AutoNt | Neutrophils by automated count | blood | 0 | 0 | 0 | 0 | 0 | | | 0 | 3 | 0 | 0 | 3 | 0 | 0 | 0 | 0 | 0 | | 0 | 0 |
| BAbs | Basophils [#/volume] in Blood by Automated count | immune / inflammatory | 51 | 244 | 49 | 2 | 195 | TMED10P2; C3orf27; RPN1; SNORA24; SLC7A10; CTD-2540B15.12 | CRBN; GATA2; RP11-475N22.4; TMED10P2; C3orf27; RPN1; SNORA24; RAB7A; PDCL3P7; RN7SKP166; HELB; GRIP1; C14orf119; CEBPE; SLC7A10; CTD-2540B15.12; C20orf203; BAK1P1; COMMD7 | 69 | 315 | 61 | 8 | 254 | 0 | 0 | 0 | 0 | 15 | 15 | 7 | 14 |
| Baso | Basophils [#/volume] in Blood | immune / inflammatory | 110 | 214 | 88 | 22 | 126 | TMED10P2; C3orf27; RPN1; SNORA24; RAB7A; AFF1; SLC7A10; CTD-2540B15.12 | CRBN; GATA2; RP11-475N22.4; TMED10P2; C3orf27; RPN1; SNORA24; RAB7A; HELB; C14orf119; CEBPE; SLC7A10; CTD-2540B15.12; C20orf112; C20orf203; BAK1P1; COMMD7 | 204 | 468 | 124 | 80 | 344 | 0 | 0 | 0 | 0 | 21 | 21 | 10 | 17 |
| BASOA | Basophils [#/volume] in Blood | blood | 0 | 0 | 0 | 0 | 0 | | | 0 | 1 | 0 | 0 | 1 | 0 | 0 | 0 | 0 | 0 | | 0 | 0 |
| BASOAB | Basophils [#/volume] in Blood by Automated count | blood | 197 | 791 | 175 | 22 | 616 | RP5-1052I5.2; LMO4; IL5RA; CRBN; TMED10P2; C3orf27; RPN1; SNORA24; RAB7A; HLA-B; XXbac-BPG248L24.12; DHFRP2; RNU6-283P; ZDHHC20P2; MICA; HCP5; C6orf183; CEBPE; SLC7A10; CTD-2540B15.12; C20orf203 | RP5-1052I5.2; LMO4; THADA; TRNT1; CRBN; GATA2; RP11-475N22.4; TMED10P2; C3orf27; RPN1; SNORA24; RAB7A; RP11-221E20.4; RP11-723O4.9; RP11-723O4.2; ACAD9; KIAA1257; AFF1; KLHL8; XXbac-BPG248L24.10; HLA-B; HLA-S; MICA; HCP5; C6orf183; RP11-425D10.1; PTCHD3P3; CCDC162P; JAK2; INSL6; RP11-140C18.2; INSL4; C11orf21; TSPAN32; RP11-335I12.2; PDCL3P7; RN7SKP166; HELB; GRIP1; C14orf119; CEBPE; SLC7A8; SLC7A10; CTD-2540B15.12; C20orf112; C20orf203; BAK1P1; COMMD7 | 532 | 1374 | 422 | 110 | 952 | 0 | 0 | 0 | 0 | 52 | 41 | 29 | 50 |
| BASOM | Basophils/100 leukocytes in Blood by Manual count | immune / inflammatory | 0 | 1 | 0 | 0 | 1 | | CMIP | 0 | 7 | 0 | 0 | 7 | 0 | 0 | 0 | 0 | 1 | 1 | 0 | 1 |
| BASOPHILS | BASOPHILS | immune / inflammatory | 1 | 0 | 0 | 1 | 0 | TENM1 | | 4 | 2 | 0 | 4 | 2 | 0 | 1 | 0 | 2 | 1 | 1 | 0 | 0 |
| BASORE | Basophils/100 leukocytes in Blood by Automated count | immune / inflammatory | 250 | 542 | 198 | 52 | 344 | RP5-1052I5.2; LMO4; TMED10P2; C3orf27; RPN1; SNORA24; RAB7A; MICA; C14orf119; CEBPE; RP11-96C21.1; SLC7A10; CTD-2540B15.12; C20orf112; C20orf203 | RP5-1052I5.2; LMO4; TRNT1; CRBN; GATA2; RP11-475N22.4; TMED10P2; C3orf27; RPN1; SNORA24; RAB7A; RP11-221E20.4; RP11-723O4.9; RP11-723O4.2; ACAD9; KIAA1257; AFF1; MICA; C6orf183; C11orf21; TSPAN32; RP11-335I12.2; PDCL3P7; RN7SKP166; HELB; GRIP1; C14orf119; CEBPE; SLC7A8; RP11-737G21.1; SS18; SLC7A10; CTD-2540B15.12; TM9SF4; C20orf112; C20orf203; BAK1P1; COMMD7 | 502 | 1015 | 314 | 188 | 701 | 0 | 0 | 0 | 0 | 43 | 42 | 23 | 41 |

| all_labs | full_name | group | male.significant.variants | female.significant.variants | both.significant.variants | male.only.significant.variants | female.only.significant.variants | male.annotated.genes.significant.variants | female.annotated.genes.significant.variants | male.suggestive.variants | female.suggestive.variants | both.suggestive.variants | male.only.suggestive.variants | female.only.suggestive.variants | female.significant.x.variants | male.significant.x.variants | female.suggestive.x.variants | male.suggestive.x.variants | n_loci | n_loci_noHLA | P1e6_both | P1e2_both |
|---|---|---|---|---|---|---|---|---|---|---|---|---|---|---|---|---|---|---|---|---|---|---|
| BBTAb | Borrelia burgdorferi Ab [Units/volume] in Serum by Immunoassay | toxicology / pharmacology | 0 | 0 | 0 | 0 | 0 | | | 0 | 0 | 0 | 0 | 0 | 0 | 0 | 0 | 0 | 0 | | 0 | 0 |
| BeGlg_ | Beta globulin [Mass/volume] in Serum or Plasma by Electrophoresis | immune / inflammatory | 0 | 0 | 0 | 0 | 0 | | | 0 | 5 | 0 | 0 | 5 | 0 | 0 | 0 | 0 | 0 | | 0 | 0 |
| BeHCG | Choriogonadotropin.beta subunit [Moles/volume] in Serum or Plasma | cancer | | 0 | | | | | | | 0 | | | | 0 | | 0 | | 0 | | 0 | 0 |
| BEVN | Base excess standard in Venous blood by calculation | metabolic | 0 | 0 | 0 | 0 | 0 | | | 1 | 0 | 0 | 1 | 0 | 0 | 0 | 0 | 0 | 0 | | 0 | 0 |
| BGPIgG | Beta 2 glycoprotein 1 IgG Ab in Serum | immune / inflammatory | 0 | 0 | 0 | 0 | 0 | | | 0 | 0 | 0 | 0 | 0 | 0 | 0 | 0 | 0 | 0 | | 0 | 0 |
| BGPIgM | Beta 2 glycoprotein 1 IgM Ab in Serum | immune / inflammatory | 0 | 0 | 0 | 0 | 0 | | | 3 | 1 | 0 | 3 | 1 | 0 | 0 | 0 | 0 | 0 | | 0 | 0 |
| Blasts | Blasts/100 leukocytes in Blood by Manual count | blood | 0 | 0 | 0 | 0 | 0 | | | 0 | 0 | 0 | 0 | 0 | 0 | 0 | 0 | 0 | 0 | | 0 | 0 |
| BLOOD PRESSURE | Diastolic blood pressure | vitals | 0 | 0 | 0 | 0 | 0 | | | 0 | 0 | 0 | 0 | 0 | 0 | 0 | 0 | 0 | 0 | | 0 | 0 |

| all_labs | full_name | group | male.significant.variants | female.significant.variants | both.significant.variants | male.only.significant.variants | female.only.significant.variants | male.annotated.genes.significant.variants | female.annotated.genes.significant.variants | male.suggestive.variants | female.suggestive.variants | both.suggestive.variants | male.only.suggestive.variants | female.only.suggestive.variants | female.significant.x.variants | male.significant.x.variants | female.suggestive.x.variants | male.suggestive.x.variants | n_loci | n_loci_noHLA | P1e6_both | P1e2_both |
|---|---|---|---|---|---|---|---|---|---|---|---|---|---|---|---|---|---|---|---|---|---|---|
| BMI_CLEAN | BMI Clean | vitals | 90 | 507 | 90 | 0 | 417 | FTO | SEC16B; AC093326.3; TMEM18; ADCY3; FTO; RNU6-567P; RP11-795H16.3; RNU4-17P; RP11-795H16.2; RP11-396N11.1 | 295 | 710 | 265 | 30 | 445 | 0 | 0 | 0 | 0 | 9 | 9 | 5 | 8 |
| BNP | Natriuretic peptide B [Mass/volume] in Serum or Plasma | cardiovascular | 0 | 0 | 0 | 0 | 0 | | | 3 | 1 | 0 | 3 | 1 | 0 | 0 | 0 | 0 | 0 | | 0 | 0 |
| BUN | Urea nitrogen serum/plasma | metabolic | 13 | 9 | 0 | 13 | 9 | UBE2Q2; FBXO22; NRG4; UMOD; PDILT | PTGER4; TTC33; PRKAA1; RPL37 | 38 | 38 | 1 | 37 | 37 | 0 | 0 | 0 | 0 | 3 | 3 | 0 | 3 |
| C-Gluc | Glucose spinal fluid | metabolic | 0 | 11 | 0 | 0 | 11 | | RN7SL571P; KCTD12 | 0 | 24 | 0 | 0 | 24 | 0 | 0 | 0 | 0 | 1 | 1 | 0 | 0 |
| C3 | Complement C3 [Mass/volume] in Serum or Plasma | immune / inflammatory | 0 | 0 | 0 | 0 | 0 | | | 1 | 1 | 0 | 1 | 1 | 0 | 0 | 0 | 0 | 0 | | 0 | 0 |

| all_labs | full_name | group | male.significant.variants | female.significant.variants | both.significant.variants | male.only.significant.variants | female.only.significant.variants | male.annotated.genes.significant.variants | female.annotated.genes.significant.variants | male.suggestive.variants | female.suggestive.variants | both.suggestive.variants | male.only.suggestive.variants | female.only.suggestive.variants | female.significant.x.variants | male.significant.x.variants | female.suggestive.x.variants | male.suggestive.x.variants | n_loci | n_loci_noHLA | P1e6_both | P1e2_both |
|---|---|---|---|---|---|---|---|---|---|---|---|---|---|---|---|---|---|---|---|---|---|---|
| C4Quan | Complement C4 [Mass/volume] in Serum or Plasma | immune / inflammatory | 824 | 2382 | 729 | 95 | 1653 | ZFP57; HLA-V; ZNRD1; RNF39; TRIM31; TRIM31-AS1; TRIM40; TRIM10; TRIM26; HCG17; TRIM39; HLA-N; HCG19P; UBQLN1P1; MICC; TMPOP1; RANP1; PPP1R18; FLOT1; RN7SL353P; LINC00243; XXbac-BPG27H4.8; RN7SKP186; DDR1-AS1; DDR1; RN7SL175P; GTF2H4; VARS2; HCG21; MUC21; MUC22; HCG22; RNU6-1133P; C6orf15; PSORS1C1; TCF19; HCG27; XXbac-BPG299F13.16; HLA-C; USP8P1; RPL3P2; XXbac-BPG248L24.10; HLA-B; XXbac-BPG248L24.12; DHFRP2; RNU6-283P; AL671883.1; FGFR3P1; ZDHHC20P2; HLA-S; XXbac-BPG181B23.7; HCP5; MICA; MICB; XXbac-BPG16N22.5; RPL15P4; ATP6V1G2-DDX39B; TNF; NCR3; PRRC2A; BAG6; APOM; GPANK1; CSNK2B; ABHD16A; XXbac-BPG32J3.20; CLIC1; MSH5; VWA7; VARS; LSM2; HSPA1L; HSPA1A; HSPA1B; C6orf48; NEU1; SLC44A4; C2; CFB; SKIV2L; STK19; TNXB; ATF6B; NOTCH4; XXbac-BPG154L12.4; C6orf10; HCG23; BTNL2; HLA-DRA; HLA-DRB9; HLA-DRB1; HLA-DQA1; HLA-DQB1; MTCO3P1; XXbac-BPG254F23.7; HLA-DQB2; HLA-DOB; TAP2; PSMB8; PSMB9; PPP1R2P1; HLA-Z | ZSCAN31; ZKSCAN3; ZSCAN12; ZSCAN23; OR2E1P; GPX6; LINC00533; RPSAP2; AL662890.1; NOL5BP; AL662890.3; XXbac-BPG308K3.5; XXbac-BPG308K3.6; ZNF90P2; HCG14; TRIM27; C6orf100; KRT18P1; RN7SL471P; HCG15; HCG16; ZNF311; OR2AD1P; OR2W1; OR2P1P; SAR1P1; OR2B3; XXbac-BCX196D17.5; OR2N1P; OR2J2; OR2J4P; OR2H4P; XXbac-BPG308J9.3; OR2B4P; OR14J1; OR5V1; OR11A1; UBDP1; MAS1LP1; MAS1L; RPS17P1; XXbac-BPG13B8.10; OR2I1P; UBD; GABBR1; SUMO2P1; ZFP57; HLA-V; HLA-A; MICD; HCG9; ZNRD1; RNF39; TRIM31; TRIM31-AS1; TRIM40; TRIM10; TRIM15; TRIM26; HCG17; TRIM39; HLA-N; HCG19P; UBQLN1P1; MICC; TMPOP1; SUCLA2P1; RANP1; XXbac-BPG249D20.9; GNL1; ABCF1; ATAT1; C6orf136; PPP1R18; TUBB; FLOT1; RN7SL353P; HCG20; LINC00243; XXbac-BPG27H4.8; RN7SKP186; DDR1-AS1; DDR1; RN7SL175P; GTF2H4; VARS2; SFTA2; DPCR1; HCG21; MUC21; MUC22; HCG22; RNU6-1133P; C6orf15; PSORS1C1; TCF19; HCG27; XXbac-BPG299F13.16; HLA-C; USP8P1; RPL3P2; WASF5P; XXbac-BPG248L24.13; XXbac-BPG248L24.10; HLA-B; XXbac-BPG248L24.12; DHFRP2; RNU6-283P; AL671883.1; FGFR3P1; ZDHHC20P2; HLA-S; XXbac-BPG181B23.7; HCP5; MICA; MICB; XXbac-BPG16N22.5; RPL15P4; MCCD1; ATP6V1G2-DDX39B; NFKBIL1; TNF; NCR3; UQCRHP1; AIF1; PRRC2A; BAG6; APOM; C6orf47; GPANK1; CSNK2B; LY6G5C; ABHD16A; XXbac-BPG32J3.20; DDAH2; CLIC1; MSH5; VWA7; VARS; LSM2; HSPA1L; HSPA1A; HSPA1B; C6orf48; NEU1; SLC44A4; EHMT2; C2; CFB; NELFE; SKIV2L; DXO; STK19; TNXB; ATF6B; FKBPL; PRRT1; RNF5; NOTCH4; XXbac-BPG154L12.4; C6orf10; HCG23; BTNL2; HLA-DRA; HLA-DRB9; HLA-DRB1; HLA-DQA1; HLA-DQB1; MTCO3P1; XXbac-BPG254F23.7; HLA-DQA2; HLA-DQB2; HLA-DOB; TAP2; PSMB8; PSMB9; PPP1R2P1; HLA-Z; HLA-DMB; XXbac-BPG181M17.5; HLA-DMA; BRD2; HLA-DOA | 1603 | 3117 | 1512 | 91 | 1605 | 0 | 0 | 0 | 0 | 124 | 5 | 66 | 119 |
| all_labs | full_name | group | male.significant.variants | female.significant.variants | both.significant.variants | male.only.significant.variants | female.only.significant.variants | male.annotated.genes.significant.variants | female.annotated.genes.significant.variants | male.suggestive.variants | female.suggestive.variants | both.suggestive.variants | male.only.suggestive.variants | female.only.suggestive.variants | female.significant.x.variants | male.significant.x.variants | female.suggestive.x.variants | male.suggestive.x.variants | n_loci | n_loci_noHLA | P1e6_both | P1e2_both |
| CA-125 | Cancer antigen 125 | cancer | 0 | 0 | 0 | 0 | 0 | | | 0 | 0 | 0 | 0 | 0 | 0 | 0 | 0 | 0 | 0 | | 0 | 0 |

| all_labs | full_name | group | male.significant.variants | female.significant.variants | both.significant.variants | male.only.significant.variants | female.only.significant.variants | male.annotated.genes.significant.variants | female.annotated.genes.significant.variants | male.suggestive.variants | female.suggestive.variants | both.suggestive.variants | male.only.suggestive.variants | female.only.suggestive.variants | female.significant.x.variants | male.significant.x.variants | female.suggestive.x.variants | male.suggestive.x.variants | n_loci | n_loci_noHLA | P1e6_both | P1e2_both |
|---|---|---|---|---|---|---|---|---|---|---|---|---|---|---|---|---|---|---|---|---|---|---|
| Ca | Calcium serum/plasma serum/plasma | metabolic | 130 | 235 | 114 | 16 | 121 | PPM1G; AC079610.1; CD86; CASR; HNRNPA1P23; CSTA; CCDC58; FAM162A; RP11-299J3.8; KPNA1; PARP9 | DGKD; CD86; CASR; HNRNPA1P23; CSTA; CCDC58; FAM162A; WDR5B; RP11-299J3.8; KPNA1; PARP9; GNAQ; RP11-205K6.1 | 221 | 294 | 159 | 62 | 135 | 0 | 0 | 0 | 0 | 19 | 19 | 11 | 16 |
| CA19-9 | Cancer Ag 19-9 | cancer | 0 | 0 | 0 | 0 | 0 | | | 0 | 0 | 0 | 0 | 0 | 0 | 0 | 0 | 0 | 0 | | 0 | 0 |
| CaIo | Calcium.ionized [Mass/volume] in Blood | metabolic | 5 | 0 | 0 | 5 | 0 | CASR | | 37 | 0 | 0 | 37 | 0 | 0 | 0 | 0 | 0 | 1 | 1 | 0 | 1 |
| CBil | Bilirubin.conjugated [Mass/volume] in Serum or Plasma | liver | 138 | 127 | 122 | 16 | 5 | UGT1A11P; UGT1A8 | UGT1A11P; UGT1A8 | 139 | 173 | 139 | 0 | 34 | 0 | 0 | 0 | 0 | 4 | 4 | 4 | 4 |
| CD3# | CD3# | immune / inflammatory | 0 | 0 | 0 | 0 | 0 | | | 0 | 1 | 0 | 0 | 1 | 0 | 0 | 0 | 0 | 0 | | 0 | 0 |
| CD3% | CD3% | immune / inflammatory | 0 | 0 | 0 | 0 | 0 | | | 0 | 0 | 0 | 0 | 0 | 0 | 0 | 0 | 0 | 0 | | 0 | 0 |
| CD4# | CD3+CD4+ (T4 helper) cells [#/volume] in Blood | immune / inflammatory | 0 | 0 | 0 | 0 | 0 | | | 0 | 1 | 0 | 0 | 1 | 0 | 0 | 0 | 0 | 0 | | 0 | 0 |
| CD4% | CD3+CD4+ (T4 helper) cells/100 cells in Blood | immune / inflammatory | 0 | 0 | 0 | 0 | 0 | | | 0 | 0 | 0 | 0 | 0 | 0 | 0 | 0 | 0 | 0 | | 0 | 0 |
| CD8_3# | CD3+CD8+(T8 suppressor cells) cells [#/volume] in Blood | immune / inflammatory | 0 | 0 | 0 | 0 | 0 | | | 0 | 8 | 0 | 0 | 8 | 0 | 0 | 0 | 0 | 0 | | 0 | 0 |
| CD8_3% | CD8_3% | immune / inflammatory | 0 | 0 | 0 | 0 | 0 | | | 0 | 7 | 0 | 0 | 7 | 0 | 0 | 7 | 0 | 0 | | 0 | 0 |
| CEA | Carcinoembryonic Ag [Mass/volume] in Serum or Plasma | cancer | 0 | 0 | 0 | 0 | 0 | | | 0 | 0 | 0 | 0 | 0 | 0 | 0 | 0 | 0 | 0 | | 0 | 0 |

| all_labs | full_name | group | male.significant.variants | female.significant.variants | both.significant.variants | male.only.significant.variants | female.only.significant.variants | male.annotated.genes.significant.variants | female.annotated.genes.significant.variants | male.suggestive.variants | female.suggestive.variants | both.suggestive.variants | male.only.suggestive.variants | female.only.suggestive.variants | female.significant.x.variants | male.significant.x.variants | female.suggestive.x.variants | male.suggestive.x.variants | n_loci | n_loci_noHLA | P1e6_both | P1e2_both |
|---|---|---|---|---|---|---|---|---|---|---|---|---|---|---|---|---|---|---|---|---|---|---|
| Cerul | Ceruloplasmin [Mass/volume] in Serum or Plasma | blood | 0 | 0 | 0 | 0 | 0 | | | 1 | 3 | 0 | 1 | 3 | 0 | 0 | 0 | 1 | 0 | | 0 | 0 |
| CHOL_HDL_RATIO | Cholesterol in HDL [Mass/volume] in Serum or Plasma | metabolic | 0 | 0 | 0 | 0 | 0 | | | 1 | 13 | 0 | 1 | 13 | 0 | 0 | 0 | 0 | 0 | | 0 | 0 |
| Chol | Cholesterol [Mass/volume] in Serum or Plasma | metabolic | 87 | 399 | 41 | 46 | 358 | SMARCA4; LDLR; TM6SF2; SUGP1; MAU2; GATAD2A; NDUFA13; CILP2; PBX4; GMIP; APOE; APOC1P1 | SARS; CELSR2; PSRC1; RP11-116D2.1; APOB; TDRD15; RP11-79J24.1; AC067959.1; GCKR; C6orf10; HCG23; BTNL2; HLA-DRA; HLA-DQA1; SMARCA4; LDLR; CTB-171A8.1; BCL3; BCAM; PVRL2; TOMM40; APOE; APOC1; APOC1P1 | 146 | 872 | 44 | 102 | 828 | 0 | 0 | 0 | 0 | 24 | 22 | 3 | 23 |
| CKMBRe | Creatine kinase.MB [Mass/volume] in Serum or Plasma | cardiovascular | 0 | 0 | 0 | 0 | 0 | | | 1 | 0 | 0 | 1 | 0 | 0 | 0 | 0 | 1 | 0 | | 0 | 0 |
| Cl-WB | Chloride [Moles/volume] in Blood | metabolic | 0 | 0 | 0 | 0 | 0 | | | 0 | 2 | 0 | 0 | 2 | 0 | 0 | 1 | 0 | 0 | | 0 | 0 |
| Cl | Chloride [Moles/volume] in Serum or Plasma | metabolic | 1 | 0 | 0 | 1 | 0 | BDNF-AS | | 8 | 4 | 0 | 8 | 4 | 0 | 0 | 0 | 0 | 1 | 1 | 0 | 1 |
| CLymph | Lymphocytes/100 leukocytes in Cerebral spinal fluid by Manual count | immune / inflammatory | 0 | 0 | 0 | 0 | 0 | | | 0 | 1 | 0 | 0 | 1 | 0 | 0 | 0 | 0 | 0 | | 0 | 0 |

| all_labs | full_name | group | male.significant.variants | female.significant.variants | both.significant.variants | male.only.significant.variants | female.only.significant.variants | male.annotated.genes.significant.variants | female.annotated.genes.significant.variants | male.suggestive.variants | female.suggestive.variants | both.suggestive.variants | male.only.suggestive.variants | female.only.suggestive.variants | female.significant.x.variants | male.significant.x.variants | female.suggestive.x.variants | male.suggestive.x.variants | n_loci | n_loci_noHLA | P1e6_both | P1e2_both |
|---|---|---|---|---|---|---|---|---|---|---|---|---|---|---|---|---|---|---|---|---|---|---|
| CMonMc | Monocytes+Macrophages/100 leukocytes in Cerebral spinal fluid by Manual count | immune / inflammatory | 0 | 0 | 0 | 0 | 0 | | | 0 | 0 | 0 | 0 | 0 | 0 | 0 | 0 | 0 | 0 | | 0 | 0 |
| CNeut | Neutrophils/100 leukocytes in Cerebral spinal fluid by Manual count | immune / inflammatory | 0 | 0 | 0 | 0 | 0 | | | 0 | 0 | 0 | 0 | 0 | 0 | 0 | 0 | 0 | 0 | | 0 | 0 |
| CNucCl | Nucleated cells [#/volume] in Cerebral spinal fluid | immune / inflammatory | 0 | 1 | 0 | 0 | 1 | | LINC01101 | 7 | 2 | 0 | 7 | 2 | 0 | 0 | 0 | 0 | 0 | | 0 | 0 |
| CO2 | Carbon dioxide serum/plasma | blood | 0 | 66 | 0 | 0 | 66 | | IL18R1; AC007248.7; IL18RAP; SLC9A4; SLC9A2 | 21 | 178 | 0 | 21 | 178 | 0 | 0 | 0 | 0 | 1 | 1 | 0 | 1 |
| CoHgb | Carboxyhemoglobin % in blood | toxicology / pharmacology | 0 | 0 | 0 | 0 | 0 | | | 0 | 0 | 0 | 0 | 0 | 0 | 0 | 0 | 0 | 0 | | 0 | 0 |
| Copper | Copper in Serum or Plasma | toxicology / pharmacology | 0 | 0 | 0 | 0 | 0 | | | 1 | 0 | 0 | 1 | 0 | 0 | 0 | 0 | 0 | 0 | | 0 | 0 |
| Cort-S | Cortisol [Mass/volume] in Serum or Plasma | endocrine | 0 | 0 | 0 | 0 | 0 | | | 0 | 4 | 0 | 0 | 4 | 0 | 0 | 0 | 0 | 0 | | 0 | 0 |
| CPepti | C peptide [Mass/volume] in Serum or Plasma | endocrine | 0 | 0 | 0 | 0 | 0 | | | 0 | 3 | 0 | 0 | 3 | 0 | 0 | 0 | 0 | 0 | | 0 | 0 |
| CPK | Creatine Kinase | cardiovascular | 0 | 7 | 0 | 0 | 7 | | CTD-2337J16.1; LILRB5; RNU6-1307P; AC010518.3 | 0 | 14 | 0 | 0 | 14 | 0 | 0 | 1 | 0 | 2 | 2 | 0 | 1 |

| all_labs | full_name | group | male.significant.variants | female.significant.variants | both.significant.variants | male.only.significant.variants | female.only.significant.variants | male.annotated.genes.significant.variants | female.annotated.genes.significant.variants | male.suggestive.variants | female.suggestive.variants | both.suggestive.variants | male.only.suggestive.variants | female.only.suggestive.variants | female.significant.x.variants | male.significant.x.variants | female.suggestive.x.variants | male.suggestive.x.variants | n_loci | n_loci_noHLA | P1e6_both | P1e2_both |
|---|---|---|---|---|---|---|---|---|---|---|---|---|---|---|---|---|---|---|---|---|---|---|
| CPKTot | Creatine kinase [Enzymatic activity/volume] in Serum or Plasma | cardiovascular | 0 | 0 | 0 | 0 | 0 | | | 11 | 0 | 0 | 11 | 0 | 0 | 0 | 0 | 0 | 0 | | 0 | 0 |
| CProt1 | Protein [Mass/volume] in Cerebral spinal fluid | metabolic | 0 | 0 | 0 | 0 | 0 | | | 1 | 8 | 0 | 1 | 8 | 0 | 0 | 0 | 0 | 0 | | 0 | 0 |
| CR-Day | Creatinine urine per day | kidney | 0 | 0 | 0 | 0 | 0 | | | 1 | 0 | 0 | 1 | 0 | 0 | 0 | 0 | 0 | 0 | | 0 | 0 |
| Cr24Hr | Creatinine [Mass/time] in 24 hour Urine | kidney | 0 | 0 | 0 | 0 | 0 | | | 1 | 0 | 0 | 1 | 0 | 0 | 0 | 0 | 0 | 0 | | 0 | 0 |
| CRBC | Erythrocytes in Cerebral spinal fluid | other | 0 | 0 | 0 | 0 | 0 | | | 1 | 0 | 0 | 1 | 0 | 0 | 0 | 0 | 0 | 0 | | 0 | 0 |
| Creat | Creatinine [Mass/volume] in Blood Creatinine [Mass/volume] in Blood | kidney | 33 | 39 | 31 | 2 | 8 | PRKAG2; UMOD; PDILT | L3MBTL3; PRKAG2; UMOD; PDILT | 39 | 80 | 35 | 4 | 45 | 0 | 0 | 1 | 0 | 3 | 3 | 2 | 2 |
| CREATININE | Creatinine | kidney | 0 | 0 | 0 | 0 | 0 | | | 2 | 0 | 0 | 2 | 0 | 0 | 0 | 0 | 0 | 0 | | 0 | 0 |
| CrePOC | Creatinine (point of care) | kidney | 0 | 0 | 0 | 0 | 0 | | | 1 | 0 | 0 | 1 | 0 | 0 | 0 | 0 | 0 | 0 | | 0 | 0 |
| CRP | C reactive protein [Mass/volume] in Serum or Plasma | immune / inflammatory | 0 | 1 | 0 | 0 | 1 | | APOE | 4 | 1 | 1 | 3 | 0 | 0 | 0 | 0 | 0 | 1 | 1 | 1 | 1 |
| CTo#Cl | Cells Counted Total in CSF | other | 0 | 0 | 0 | 0 | 0 | | | 0 | 0 | 0 | 0 | 0 | 0 | 0 | 0 | 0 | 0 | | 0 | 0 |

| all_labs | full_name | group | male.significant.variants | female.significant.variants | both.significant.variants | male.only.significant.variants | female.only.significant.variants | male.annotated.genes.significant.variants | female.annotated.genes.significant.variants | male.suggestive.variants | female.suggestive.variants | both.suggestive.variants | male.only.suggestive.variants | female.only.suggestive.variants | female.significant.x.variants | male.significant.x.variants | female.suggestive.x.variants | male.suggestive.x.variants | n_loci | n_loci_noHLA | P1e6_both | P1e2_both |
|---|---|---|---|---|---|---|---|---|---|---|---|---|---|---|---|---|---|---|---|---|---|---|
| D25OHT | Calcitriol [Mass/volume] in Serum or Plasma | metabolic | 93 | 156 | 89 | 4 | 67 | GC; RP11-140L24.3; COPB1; PSMA1; PDE3B; CYP2R1; CALCB | GC; SPON1; RP11-21L19.1; RRAS2; RP11-140L24.3; COPB1; PSMA1; PDE3B; CYP2R1; CALCB | 112 | 180 | 104 | 8 | 76 | 0 | 0 | 0 | 0 | 8 | 8 | 7 | 8 |
| DHEA-S | DHEA-S in Serum or Plasma | endocrine | 0 | 0 | 0 | 0 | 0 | | | 0 | 0 | 0 | 0 | 0 | 0 | 0 | 0 | 0 | 0 | | 0 | 0 |
| DIASTOLIC | Diastolic blood pressure | vitals | 0 | 0 | 0 | 0 | 0 | | | 0 | 2 | 0 | 0 | 2 | 0 | 0 | 0 | 0 | 0 | | 0 | 0 |
| Digox | Digoxin [Mass/volume] in Serum or Plasma | toxicology / pharmacology | 0 | 0 | 0 | 0 | 0 | | | 0 | 2 | 0 | 0 | 2 | 0 | 0 | 0 | 0 | 0 | | 0 | 0 |
| Dimer | Dimer | blood | 0 | 0 | 0 | 0 | 0 | | | 2 | 0 | 0 | 2 | 0 | 0 | 0 | 0 | 0 | 0 | | 0 | 0 |
| DirLDL | Cholesterol in LDL [Mass/volume] in Serum or Plasma by Direct assay | metabolic | 1 | 2 | 1 | 0 | 1 | APOE | APOE | 2 | 5 | 2 | 0 | 3 | 0 | 0 | 0 | 0 | 1 | 1 | 1 | 1 |
| DLCO_COR_PPR | DLCO_COR_PPR | lung | 0 | 0 | 0 | 0 | 0 | | | 0 | 0 | 0 | 0 | 0 | 0 | 0 | 0 | 0 | 0 | | 0 | 0 |
| DLCO_COR_PRD | DLCO_COR_PRD | lung | 0 | 0 | 0 | 0 | 0 | | | 3 | 0 | 0 | 3 | 0 | 0 | 0 | 0 | 0 | 0 | | 0 | 0 |
| DLCO_COR_PRL | DLCO_COR_PRL | lung | 0 | 0 | 0 | 0 | 0 | | | 2 | 0 | 0 | 2 | 0 | 0 | 0 | 0 | 0 | 0 | | 0 | 0 |
| DLCO_COR_PRU | DLCO_COR_PRU | lung | 0 | 0 | 0 | 0 | 0 | | | 0 | 0 | 0 | 0 | 0 | 0 | 0 | 0 | 0 | 0 | | 0 | 0 |
| DLCO_COR_ZBD | DLCO_COR_ZBD | lung | 0 | 0 | 0 | 0 | 0 | | | 0 | 1 | 0 | 0 | 1 | 0 | 0 | 0 | 0 | 0 | | 0 | 0 |
| DLCO_UNC_PPR | DLCO_UNC_PPR | lung | 0 | 0 | 0 | 0 | 0 | | | 1 | 0 | 0 | 1 | 0 | 0 | 0 | 0 | 0 | 0 | | 0 | 0 |
| DLCO_UNC_PRD | DLCO_UNC_PRD | lung | 0 | 0 | 0 | 0 | 0 | | | 3 | 0 | 0 | 3 | 0 | 0 | 0 | 0 | 0 | 0 | | 0 | 0 |
| DLCO_UNC_PRL | DLCO_UNC_PRL | lung | 0 | 0 | 0 | 0 | 0 | | | 2 | 0 | 0 | 2 | 0 | 0 | 0 | 0 | 0 | 0 | | 0 | 0 |

| all_labs | full_name | group | male.significant.variants | female.significant.variants | both.significant.variants | male.only.significant.variants | female.only.significant.variants | male.annotated.genes.significant.variants | female.annotated.genes.significant.variants | male.suggestive.variants | female.suggestive.variants | both.suggestive.variants | male.only.suggestive.variants | female.only.suggestive.variants | female.significant.x.variants | male.significant.x.variants | female.suggestive.x.variants | male.suggestive.x.variants | n_loci | n_loci_noHLA | P1e6_both | P1e2_both |
|---|---|---|---|---|---|---|---|---|---|---|---|---|---|---|---|---|---|---|---|---|---|---|
| DLCO_UNC_PRU | DLCO_UNC_PRU | lung | 0 | 0 | 0 | 0 | 0 | | | 0 | 0 | 0 | 0 | 0 | 0 | 0 | 0 | 0 | 0 | | 0 | 0 |
| DLCO_UNC_ZBD | DLCO_UNC_ZBD | lung | 0 | 0 | 0 | 0 | 0 | | | 1 | 0 | 0 | 1 | 0 | 0 | 0 | 0 | 0 | 0 | | 0 | 0 |
| DLCOOVA_PPR | DLCOOVA_PPR | lung | 0 | 0 | 0 | 0 | 0 | | | 0 | 0 | 0 | 0 | 0 | 0 | 0 | 0 | 0 | 0 | | 0 | 0 |
| DLCOOVA_PRD | DLCOOVA_PRD | lung | 0 | 4 | 0 | 0 | 4 | | LINC00942 | 2 | 7 | 0 | 2 | 7 | 0 | 0 | 0 | 0 | 1 | 1 | 0 | 0 |
| DLCOOVA_ZBD | DLCOOVA_ZBD | lung | 0 | 0 | 0 | 0 | 0 | | | 1 | 0 | 0 | 1 | 0 | 0 | 0 | 0 | 0 | 0 | | 0 | 0 |
| eGFR | Estimation of glomerular filtration rate | kidney | 0 | 0 | 0 | 0 | 0 | | | 0 | 4 | 0 | 0 | 4 | 0 | 0 | 0 | 0 | 0 | | 0 | 0 |
| EGFR | Estimation of glomerular filtration rate | kidney | 36 | 31 | 30 | 6 | 1 | PRKAG2; CTD-2651B20.3; CTD-2651B20.5; UMOD; PDILT | PRKAG2; UMOD; PDILT | 86 | 97 | 50 | 36 | 47 | 0 | 0 | 1 | 0 | 3 | 3 | 3 | 3 |
| eGFRcr | Estimation of glomerular filtration rate (eGFRcr) | kidney | 0 | 0 | 0 | 0 | 0 | | | 3 | 0 | 0 | 3 | 0 | 0 | 0 | 0 | 0 | 0 | | 0 | 0 |
| Endomy | Endomysium IgA Ab [Presence] in Serum | immune / inflammatory | 0 | 0 | 0 | 0 | 0 | | | 0 | 0 | 0 | 0 | 0 | 0 | 0 | 0 | 0 | 0 | | 0 | 0 |
| EoAbs | Absolute Eosinophils | immune / inflammatory | 6 | 113 | 0 | 6 | 113 | C3orf27 | AC007248.6; IL1RL1; IL18R1; IL18RAP; C3orf27; RPN1; RP11-617D20.1; AC021860.1; KLF3; CSF2; P4HA2; SLC22A4; C5orf56 | 75 | 304 | 35 | 40 | 269 | 0 | 0 | 0 | 0 | 6 | 6 | 3 | 6 |
| Eos | Eosinophils | immune / inflammatory | 28 | 102 | 10 | 18 | 92 | TMED10P2; C3orf27; RPN1 | AC007248.6; IL1RL1; IL18R1; IL18RAP; C3orf27; RPN1; SNORA24; RP11-617D20.1; AC021860.1; KLF3; CSF2 | 84 | 142 | 31 | 53 | 111 | 0 | 0 | 0 | 0 | 5 | 5 | 2 | 5 |
| EOSIA | Eosinophils [#/volume] in Blood by Automated count | immune / inflammatory | 0 | 4 | 0 | 0 | 4 | | C3orf27; RPN1 | 35 | 14 | 0 | 35 | 14 | 0 | 0 | 0 | 0 | 1 | 1 | 0 | 1 |

| all_labs | full_name | group | male.significant.variants | female.significant.variants | both.significant.variants | male.only.significant.variants | female.only.significant.variants | male.annotated.genes.significant.variants | female.annotated.genes.significant.variants | male.suggestive.variants | female.suggestive.variants | both.suggestive.variants | male.only.suggestive.variants | female.only.suggestive.variants | female.significant.x.variants | male.significant.x.variants | female.suggestive.x.variants | male.suggestive.x.variants | n_loci | n_loci_noHLA | P1e6_both | P1e2_both |
|---|---|---|---|---|---|---|---|---|---|---|---|---|---|---|---|---|---|---|---|---|---|---|
| EOSIAB | Eosinophils [#/volume] in Blood by Automated count | immune / inflammatory | 0 | 4 | 0 | 0 | 4 | | C3orf27; RPN1 | 17 | 18 | 3 | 14 | 15 | 0 | 0 | 0 | 0 | 1 | 1 | 1 | 1 |
| EOSIM | Eosinophils/100 leukocytes in Blood by Automated count | immune / inflammatory | 23 | 5 | 4 | 19 | 1 | C3orf27; RPN1; SNORA24 | TMED10P2; C3orf27; RPN1 | 52 | 139 | 14 | 38 | 125 | 0 | 0 | 0 | 0 | 1 | 1 | 1 | 1 |
| EOSINOPHILS | EOSINOPHILS | immune / inflammatory | 0 | 0 | 0 | 0 | 0 | | | 3 | 0 | 0 | 3 | 0 | 0 | 0 | 0 | 0 | 0 | | 0 | 0 |
| EOSIRE | Eosinophils % | immune / inflammatory | 17 | 4 | 0 | 17 | 4 | RAD50 | C3orf27; RPN1 | 27 | 8 | 4 | 23 | 4 | 0 | 0 | 0 | 0 | 2 | 2 | 1 | 2 |
| Estrdl | Estradiol (E2) [Mass/volume] in Serum or Plasma | endocrine | 0 | 0 | 0 | 0 | 0 | | | 0 | 0 | 0 | 0 | 0 | 0 | 0 | 0 | 0 | 0 | | 0 | 0 |
| F-Gluc | Glucose [Mass/volume] in Body fluid | metabolic | 0 | 0 | 0 | 0 | 0 | | | 0 | 2 | 0 | 0 | 2 | 0 | 0 | 0 | 0 | 0 | | 0 | 0 |
| Fbrngn | Fibrinogen [Mass/volume] in Platelet poor plasma by Coagulation assay | blood | 0 | 0 | 0 | 0 | 0 | | | 5 | 0 | 0 | 5 | 0 | 0 | 0 | 0 | 0 | 0 | | 0 | 0 |
| FEF25_75_AFD | FEF AFD | lung | 0 | 0 | 0 | 0 | 0 | | | 1 | 3 | 0 | 1 | 3 | 0 | 0 | 0 | 0 | 0 | | 0 | 0 |
| FEF25_75_PPR | FEF PPR | lung | 0 | 0 | 0 | 0 | 0 | | | 0 | 11 | 0 | 0 | 11 | 0 | 0 | 0 | 0 | 0 | | 0 | 0 |
| FEF25_75_PRD | FEF PRD | lung | 0 | 0 | 0 | 0 | 0 | | | 6 | 1 | 0 | 6 | 1 | 0 | 0 | 0 | 0 | 0 | | 0 | 0 |
| FEF25_75_PRL | FEF PRL | lung | 0 | 0 | 0 | 0 | 0 | | | 0 | 0 | 0 | 0 | 0 | 0 | 0 | 0 | 0 | 0 | | 0 | 0 |

| all_labs | full_name | group | male.significant.variants | female.significant.variants | both.significant.variants | male.only.significant.variants | female.only.significant.variants | male.annotated.genes.significant.variants | female.annotated.genes.significant.variants | male.suggestive.variants | female.suggestive.variants | both.suggestive.variants | male.only.suggestive.variants | female.only.suggestive.variants | female.significant.x.variants | male.significant.x.variants | female.suggestive.x.variants | male.suggestive.x.variants | n_loci | n_loci_noHLA | P1e6_both | P1e2_both |
|---|---|---|---|---|---|---|---|---|---|---|---|---|---|---|---|---|---|---|---|---|---|---|
| FEF25_75_ZBD | FEF ZBD | lung | 0 | 0 | 0 | 0 | 0 | | | 0 | 4 | 0 | 0 | 4 | 0 | 0 | 0 | 0 | 0 | | 0 | 0 |
| FEF25_75_ZPC | FEF ZPC | lung | 0 | 0 | 0 | 0 | 0 | | | 1 | 7 | 0 | 1 | 7 | 0 | 0 | 0 | 0 | 0 | | 0 | 0 |
| FEosin | Eosinophils/100 leukocytes in Body fluid by Manual count | immune / inflammatory | 0 | 0 | 0 | 0 | 0 | | | 0 | 5 | 0 | 0 | 5 | 0 | 0 | 0 | 0 | 0 | | 0 | 0 |
| Ferrit | Ferritin [Mass/volume] in Serum or Plasma | blood | 0 | 0 | 0 | 0 | 0 | | | 1 | 1 | 0 | 1 | 1 | 0 | 0 | 0 | 0 | 0 | | 0 | 0 |
| FET_AFD | FET AFD | lung | 0 | 0 | 0 | 0 | 0 | | | 0 | 0 | 0 | 0 | 0 | 0 | 0 | 0 | 0 | 0 | | 0 | 0 |
| FET_ZBD | FET ZBD | lung | 0 | 0 | 0 | 0 | 0 | | | 3 | 0 | 0 | 3 | 0 | 0 | 0 | 0 | 0 | 0 | | 0 | 0 |
| FET_ZPC | FET ZPC | lung | 0 | 0 | 0 | 0 | 0 | | | 0 | 0 | 0 | 0 | 0 | 0 | 0 | 0 | 0 | 0 | | 0 | 0 |
| FEV1_PPR | FEV1 PPR | lung | 0 | 0 | 0 | 0 | 0 | | | 0 | 1 | 0 | 0 | 1 | 0 | 0 | 0 | 0 | 0 | | 0 | 0 |
| FEV1_PRD | FEV1 PRD | lung | 0 | 0 | 0 | 0 | 0 | | | 0 | 1 | 0 | 0 | 1 | 0 | 0 | 0 | 0 | 0 | | 0 | 0 |
| FEV1_PRL | FEV1 PRL | lung | 0 | 0 | 0 | 0 | 0 | | | 1 | 1 | 0 | 1 | 1 | 0 | 0 | 0 | 0 | 0 | | 0 | 0 |
| FEV1_ZBD | FEV1 ZBD | lung | 0 | 0 | 0 | 0 | 0 | | | 0 | 0 | 0 | 0 | 0 | 0 | 0 | 0 | 0 | 0 | | 0 | 0 |
| FEV1_ZPC | FEV1 ZPC | lung | 0 | 0 | 0 | 0 | 0 | | | 0 | 3 | 0 | 0 | 3 | 0 | 0 | 0 | 0 | 0 | | 0 | 0 |
| FEV1OFVC_AFD | FEV1/FVC AFD | lung | 0 | 0 | 0 | 0 | 0 | | | 0 | 0 | 0 | 0 | 0 | 0 | 0 | 0 | 0 | 0 | | 0 | 0 |
| FEV1OFVC_APP | FEV1/FVC APP | lung | 0 | 0 | 0 | 0 | 0 | | | 0 | 0 | 0 | 0 | 0 | 0 | 0 | 0 | 0 | 0 | | 0 | 0 |
| FEV1OFVC_PPR | FEV1/FVC PPR | lung | 0 | 0 | 0 | 0 | 0 | | | 0 | 0 | 0 | 0 | 0 | 0 | 0 | 0 | 0 | 0 | | 0 | 0 |
| FEV1OFVC_PRD | FEV1/FVC PRD | lung | 0 | 0 | 0 | 0 | 0 | | | 14 | 22 | 0 | 14 | 22 | 0 | 0 | 0 | 0 | 0 | | 0 | 0 |
| FEV1OFVC_PRL | FEV1/FVC PRL | lung | 0 | 0 | 0 | 0 | 0 | | | 1 | 0 | 0 | 1 | 0 | 0 | 0 | 0 | 1 | 0 | | 0 | 0 |
| FEV1OFVC_ZBD | FEV1/FVC ZBD | lung | 0 | 0 | 0 | 0 | 0 | | | 0 | 1 | 0 | 0 | 1 | 0 | 0 | 0 | 0 | 0 | | 0 | 0 |

| all_labs | full_name | group | male.significant.variants | female.significant.variants | both.significant.variants | male.only.significant.variants | female.only.significant.variants | male.annotated.genes.significant.variants | female.annotated.genes.significant.variants | male.suggestive.variants | female.suggestive.variants | both.suggestive.variants | male.only.suggestive.variants | female.only.suggestive.variants | female.significant.x.variants | male.significant.x.variants | female.suggestive.x.variants | male.suggestive.x.variants | n_loci | n_loci_noHLA | P1e6_both | P1e2_both |
|---|---|---|---|---|---|---|---|---|---|---|---|---|---|---|---|---|---|---|---|---|---|---|
| FIO2 | Oxygen/Inspired gas Respiratory system--on ventilator | blood | 0 | 0 | 0 | 0 | 0 | | | 0 | 4 | 0 | 0 | 4 | 0 | 0 | 0 | 0 | 0 | | 0 | 0 |
| FK-506 | Tacrolimus [Mass/volume] in Blood | toxicology / pharmacology | 0 | 0 | 0 | 0 | 0 | | | 0 | 0 | 0 | 0 | 0 | 0 | 0 | 0 | 0 | 0 | | 0 | 0 |
| FLymph | Lymphocytes [#/volume] in Body fluid by Manual count | immune / inflammatory | 0 | 0 | 0 | 0 | 0 | | | 0 | 12 | 0 | 0 | 12 | 0 | 0 | 0 | 0 | 0 | | 0 | 0 |
| FMoncl | Monocytes+Macrophages/100 leukocytes in Body fluid by Manual count | immune / inflammatory | 0 | 0 | 0 | 0 | 0 | | | 2 | 5 | 0 | 2 | 5 | 0 | 0 | 1 | 0 | 0 | | 0 | 0 |
| FMonMc | Mononuclear cells [#/volume] in Body fluid by Manual count | immune / inflammatory | 0 | 0 | 0 | 0 | 0 | | | 6 | 0 | 0 | 6 | 0 | 0 | 0 | 0 | 0 | 0 | | 0 | 0 |
| FNeut | Neutrophils/100 leukocytes in Body fluid by Manual count | immune / inflammatory | 0 | 0 | 0 | 0 | 0 | | | 16 | 0 | 0 | 16 | 0 | 0 | 0 | 0 | 0 | 0 | | 0 | 0 |
| FNucCl | Nucleated cells [#/volume] in Synovial fluid by Manual count | blood | 0 | 0 | 0 | 0 | 0 | | | 0 | 0 | 0 | 0 | 0 | 0 | 0 | 0 | 0 | 0 | | 0 | 0 |
| FProt1 | Protein body fluid | metabolic | 0 | 0 | 0 | 0 | 0 | | | 0 | 0 | 0 | 0 | 0 | 0 | 0 | 0 | 0 | 0 | | 0 | 0 |

| all_labs | full_name | group | male.significant.variants | female.significant.variants | both.significant.variants | male.only.significant.variants | female.only.significant.variants | male.annotated.genes.significant.variants | female.annotated.genes.significant.variants | male.suggestive.variants | female.suggestive.variants | both.suggestive.variants | male.only.suggestive.variants | female.only.suggestive.variants | female.significant.x.variants | male.significant.x.variants | female.suggestive.x.variants | male.suggestive.x.variants | n_loci | n_loci_noHLA | P1e6_both | P1e2_both |
|---|---|---|---|---|---|---|---|---|---|---|---|---|---|---|---|---|---|---|---|---|---|---|
| FRBC1 | Erythrocytes [#/volume] in Blood by Automated count | blood | 0 | 0 | 0 | 0 | 0 | | | 0 | 0 | 0 | 0 | 0 | 0 | 0 | 0 | 0 | 0 | | 0 | 0 |
| FRC_PPR | FRC PPR | lung | 0 | 0 | 0 | 0 | 0 | | | 12 | 0 | 0 | 12 | 0 | 0 | 0 | 0 | 0 | 0 | | 0 | 0 |
| FRC_PRD | FRC PRD | lung | 0 | 0 | 0 | 0 | 0 | | | 1 | 0 | 0 | 1 | 0 | 0 | 0 | 0 | 0 | 0 | | 0 | 0 |
| FRC_PRL | FRC PRL | lung | 0 | 0 | 0 | 0 | 0 | | | 1 | 33 | 0 | 1 | 33 | 0 | 0 | 0 | 0 | 0 | | 0 | 0 |
| FRC_PRU | FRC PRU | lung | 0 | 0 | 0 | 0 | 0 | | | 0 | 5 | 0 | 0 | 5 | 0 | 0 | 0 | 0 | 0 | | 0 | 0 |
| FRC_ZBD | FRC ZBD | lung | 0 | 0 | 0 | 0 | 0 | | | 3 | 0 | 0 | 3 | 0 | 0 | 0 | 0 | 0 | 0 | | 0 | 0 |
| FRONTAL AXIS INITIAL 40 MS | Frontal axis initial 40 ms | cardiovascular | 0 | 0 | 0 | 0 | 0 | | | 0 | 1 | 0 | 0 | 1 | 0 | 0 | 0 | 0 | 0 | | 0 | 0 |
| FRONTAL AXIS MEAN QRS | QRS axis | cardiovascular | 0 | 0 | 0 | 0 | 0 | | | 0 | 0 | 0 | 0 | 0 | 0 | 0 | 0 | 0 | 0 | | 0 | 0 |
| FRONTAL AXIS P | P wave axis | cardiovascular | 0 | 0 | 0 | 0 | 0 | | | 0 | 0 | 0 | 0 | 0 | 0 | 0 | 0 | 0 | 0 | | 0 | 0 |
| FRONTAL AXIS ST | ST segment axis | cardiovascular | 0 | 0 | 0 | 0 | 0 | | | 0 | 0 | 0 | 0 | 0 | 0 | 0 | 0 | 0 | 0 | | 0 | 0 |
| FRONTAL AXIS TERMINAL 40 MS | Frontal axis terminal 40 ms | cardiovascular | 0 | 0 | 0 | 0 | 0 | | | 18 | 0 | 0 | 18 | 0 | 0 | 0 | 0 | 0 | 0 | | 0 | 0 |
| FSH1 | Follitropin [Units/volume] in Serum or Plasma | endocrine | 0 | 0 | 0 | 0 | 0 | | | 0 | 15 | 0 | 0 | 15 | 0 | 0 | 0 | 0 | 0 | | 0 | 0 |
| FT4 | Thyroxine (T4) free [Mass/volume] in Serum or Plasma | endocrine | 0 | 3 | 0 | 0 | 3 | | DIO1; QSOX2 | 0 | 6 | 0 | 0 | 6 | 0 | 0 | 0 | 0 | 2 | 2 | 0 | 2 |
| FTo#Cl | Cells counted in body fluid | other | 0 | 0 | 0 | 0 | 0 | | | 2 | 0 | 0 | 2 | 0 | 0 | 0 | 0 | 0 | 0 | | 0 | 0 |

| all_labs | full_name | group | male.significant.variants | female.significant.variants | both.significant.variants | male.only.significant.variants | female.only.significant.variants | male.annotated.genes.significant.variants | female.annotated.genes.significant.variants | male.suggestive.variants | female.suggestive.variants | both.suggestive.variants | male.only.suggestive.variants | female.only.suggestive.variants | female.significant.x.variants | male.significant.x.variants | female.suggestive.x.variants | male.suggestive.x.variants | n_loci | n_loci_noHLA | P1e6_both | P1e2_both |
|---|---|---|---|---|---|---|---|---|---|---|---|---|---|---|---|---|---|---|---|---|---|---|
| FVC_PPR | FVC PPR | lung | 0 | 0 | 0 | 0 | 0 | | | 17 | 14 | 0 | 17 | 14 | 0 | 0 | 0 | 17 | 0 | | 0 | 0 |
| FVC_PRD | FVC PRD | lung | 0 | 0 | 0 | 0 | 0 | | | 0 | 1 | 0 | 0 | 1 | 0 | 0 | 0 | 0 | 0 | | 0 | 0 |
| FVC_PRL | FVC PRL | lung | 0 | 0 | 0 | 0 | 0 | | | 7 | 8 | 0 | 7 | 8 | 0 | 0 | 0 | 0 | 0 | | 0 | 0 |
| FVC_ZBD | FVC ZBD | lung | 0 | 0 | 0 | 0 | 0 | | | 8 | 4 | 0 | 8 | 4 | 0 | 0 | 0 | 0 | 0 | | 0 | 0 |
| FVC_ZPC | FVC ZPC | lung | 0 | 0 | 0 | 0 | 0 | | | 3 | 0 | 0 | 3 | 0 | 0 | 0 | 0 | 0 | 0 | | 0 | 0 |

| all_labs | full_name | group | male.significant.variants | female.significant.variants | both.significant.variants | male.only.significant.variants | female.only.significant.variants | male.annotated.genes.significant.variants | female.annotated.genes.significant.variants | male.suggestive.variants | female.suggestive.variants | both.suggestive.variants | male.only.suggestive.variants | female.only.suggestive.variants | female.significant.x.variants | male.significant.x.variants | female.suggestive.x.variants | male.suggestive.x.variants | n_loci | n_loci_noHLA | P1e6_both | P1e2_both |
|---|---|---|---|---|---|---|---|---|---|---|---|---|---|---|---|---|---|---|---|---|---|---|
| GamGg_ | Gamma globulin in Serum or Plasma by electrophoresis | immune / inflammatory | 0 | 0 | 0 | 0 | 0 | | | 0 | 9 | 0 | 0 | 9 | 0 | 0 | 0 | 0 | 0 | | 0 | 0 |
| GGT | Gamma glutamyl transferase activity in Serum or Plasma | liver | 0 | 0 | 0 | 0 | 0 | | | 3 | 0 | 0 | 3 | 0 | 0 | 0 | 0 | 2 | 0 | | 0 | 0 |
| Glob | Globulin | metabolic | 0 | 0 | 0 | 0 | 0 | | | 1 | 3 | 0 | 1 | 3 | 0 | 0 | 0 | 0 | 0 | | 0 | 0 |
| GluBed | Glucose [Mass/volume] in Blood by Automated test strip | metabolic | 2 | 0 | 0 | 2 | 0 | TCF7L2 | | 19 | 12 | 12 | 7 | 0 | 0 | 0 | 0 | 0 | 1 | 1 | 1 | 1 |
| Gluc | Glucose lab | metabolic | 76 | 132 | 42 | 34 | 90 | ABCB11; ADCY5; EIF5A2; KLF7P1; RNU1-70P; SLC2A2; HLA-DQB1; GCK; YKT6; CAMK2B; TCF7L2 | SPC25; ABCB11; ADCY5; GCK; YKT6; CAMK2B; SLC30A8; TCF7L2 | 263 | 171 | 82 | 181 | 89 | 0 | 0 | 0 | 0 | 15 | 14 | 7 | 15 |
| GluWB | Glucose [Mass/volume] in Blood | blood | 0 | 0 | 0 | 0 | 0 | | | 0 | 0 | 0 | 0 | 0 | 0 | 0 | 0 | 0 | 0 | | 0 | 0 |
| GTT50g | Glucose tolerance50g (GTT50g) | metabolic | | 0 | | | | | | | 4 | | | | 0 | | 0 | | 0 | | 0 | 0 |

| all_labs | full_name | group | male.significant.variants | female.significant.variants | both.significant.variants | male.only.significant.variants | female.only.significant.variants | male.annotated.genes.significant.variants | female.annotated.genes.significant.variants | male.suggestive.variants | female.suggestive.variants | both.suggestive.variants | male.only.suggestive.variants | female.only.suggestive.variants | female.significant.x.variants | male.significant.x.variants | female.suggestive.x.variants | male.suggestive.x.variants | n_loci | n_loci_noHLA | P1e6_both | P1e2_both |
|---|---|---|---|---|---|---|---|---|---|---|---|---|---|---|---|---|---|---|---|---|---|---|
| Haptgl | Haptoglobin [Mass/volume] in Serum or Plasma | blood | 5 | 41 | 5 | 0 | 36 | DHODH; TXNL4B; DHX38; PMFBP1 | RP11-417N10.3; ATXN1L; IST1; PKD1L3; DHODH; TXNL4B; DHX38; PMFBP1 | 6 | 43 | 5 | 1 | 38 | 0 | 0 | 0 | 0 | 3 | 3 | 1 | 3 |
| HB_PRD | HB_PRD | lung | 0 | 0 | 0 | 0 | 0 | | | 0 | 15 | 0 | 0 | 15 | 0 | 0 | 0 | 0 | 0 | | 0 | 0 |
| HBSA# | Hepatitis B virus surface Ab [Presence] in Serum | immune / inflammatory | 0 | 0 | 0 | 0 | 0 | | | 0 | 1 | 0 | 0 | 1 | 0 | 0 | 0 | 0 | 0 | | 0 | 0 |
| HCO3Ca | Bicarbonate [Moles/volume] in Unspecified specimen | metabolic | 0 | 0 | 0 | 0 | 0 | | | 1 | 25 | 0 | 1 | 25 | 0 | 0 | 0 | 0 | 0 | | 0 | 0 |
| HCTWB | Hematocrit | blood | 0 | 0 | 0 | 0 | 0 | | | 1 | 0 | 0 | 1 | 0 | 0 | 0 | 0 | 1 | 0 | | 0 | 0 |
| HDL-C | HDL Cholesterol | metabolic | 258 | 348 | 244 | 14 | 104 | CSGALNACT1; LPL; AC100802.3; ALDH1A2; AC012181.1; CETP; PVRL2; APOE; APOC1 | RP11-36B6.1; LPL; AC100802.3; APOA1; SIK3; PCSK7; ALDH1A2; SLC12A3; HERPUD1; AC012181.1; CETP; NLRC5; SMUG1P1 | 323 | 461 | 271 | 52 | 190 | 0 | 0 | 0 | 0 | 30 | 30 | 17 | 28 |
| HEART RATE | Heart rate | vitals | 0 | 4 | 0 | 0 | 4 | | MTUS2 | 15 | 5 | 0 | 15 | 5 | 0 | 0 | 0 | 0 | 1 | 1 | 0 | 0 |

| all_labs | full_name | group | male.significant.variants | female.significant.variants | both.significant.variants | male.only.significant.variants | female.only.significant.variants | male.annotated.genes.significant.variants | female.annotated.genes.significant.variants | male.suggestive.variants | female.suggestive.variants | both.suggestive.variants | male.only.suggestive.variants | female.only.suggestive.variants | female.significant.x.variants | male.significant.x.variants | female.suggestive.x.variants | male.suggestive.x.variants | n_loci | n_loci_noHLA | P1e6_both | P1e2_both |
|---|---|---|---|---|---|---|---|---|---|---|---|---|---|---|---|---|---|---|---|---|---|---|
| Height | Height | vitals | 639 | 2206 | 487 | 152 | 1719 | RNA5SP56; MTMR11; AC006369.3; PNPT1; EFEMP1; NPPC; DIS3L2; ZBTB38; FNDC3B; FAM184B; DCAF16; NCAPG; LCORL; HHIP-AS1; HHIP; NPR3; BMP6; ANKS1A; MIR588; L3MBTL3; CDK6; INS-IGF2; KDM2A; SSH3; TPCN2; RP11-554A11.7; SERPINH1; HMGA2; ADAMTSL3; ATAD5; ADAP2; C17orf82; CABLES1; ADAMTS10; CDK5RAP1; PXMP4; EDEM2; UQCC1; GDF5OS; GDF5; KIF4CP; ITM2A; RP1-164L12.1 | MFAP2; RP1-37C10.3; ATP13A2; SDHB; MTMR11; DNM3; COLGALT2; EFEMP1; NPPC; DIS3L2; RP11-894J14.5; RP11-438D8.2; ZBTB38; FAM184B; DCAF16; NCAPG; LCORL; PRKG2; HHIP-AS1; HHIP; NPR3; P4HA2; SLC22A4; AC034220.3; SLC22A5; CTB-33O18.1; HIST1H2BF; HIST1H4E; HIST1H2BG; HIST1H3E; HIST1H2APS3; HIST1H1D; HIST1H4F; HIST1H3G; HIST1H2APS4; HIST1H2BI; HIST1H4H; HIST1H3PS1; RNU6-1259P; RP3-391O22.3; C6orf106; RP11-140K17.2; RP3-375P9.2; SNRPC; UHRF1BP1; SUPT3H; BCKDHB; RP11-486E2.1; LINC00577; LIN28B; L3MBTL3; GPR126; RP11-440G9.1; AMZ1; GNA12; JAZF1; CDK6; RP11-203E8.1; ADAM28; PLAG1; RP11-140I16.3; RP11-140I16.2; SDR16C5; AC105242.1; PCSK5; RNU2-36P; C9orf3; PTCH1; INS-IGF2; KDM2A; SSH3; TMEM134; SERPINH1; CCND2; CCDC91; HMGA2; DLEU1; LTBP2; MARK3; PML; SH3GL3; ADAMTSL3; ACAN; ADAMTS17; ATAD5; TEFM; ADAP2; C17orf82; CSHL1; CD79B; CABLES1; NFATC1; MYO1F; ADAMTS10; CASC20; RP5-859D4.3; EDEM2; UQCC1; GDF5OS; GDF5; CEP250; RP3-477O4.16; RP3-477O4.5; C20orf173; ERGIC3; FER1L4; SPAG4; CPNE1; RP1-309K20.6; NFS1; RBM39; RPF2P1; PHF20; SCAND1; CNBD2; RP1-164L12.1 | 1384 | 3861 | 744 | 640 | 3117 | 51 | 26 | 70 | 83 | 110 | 110 | 30 | 99 |
| HEIGHT | Height | vitals | 49 | 547 | 15 | 34 | 532 | EFEMP1; IGF2BP3; CABLES1 | MTMR11; DNM3; LTBP1; ZBTB38; FAM184B; DCAF16; NCAPG; LCORL; HHIP-AS1; HHIP; KDM2A; SSH3; CCDC91; HMGA2; DLEU1; ADAMTSL3; ACAN; ATAD5; TEFM; ADAP2; CABLES1; UQCC1; GDF5OS; GDF5 | 264 | 1195 | 144 | 120 | 1051 | 0 | 0 | 57 | 0 | 21 | 21 | 5 | 16 |
| HEMATOCRIT | HEMATOCRIT | blood | 0 | 0 | 0 | 0 | 0 | | | 4 | 0 | 0 | 4 | 0 | 0 | 0 | 0 | 0 | 0 | | 0 | 0 |
| HEMOGLOBIN | HEMOGLOBIN | blood | 0 | 0 | 0 | 0 | 0 | | | 0 | 0 | 0 | 0 | 0 | 0 | 0 | 0 | 0 | 0 | | 0 | 0 |
| Hgb | Hemoglobin | blood | 40 | 166 | 15 | 25 | 151 | LRRC16A; SCGN; HIST1H2AA; SLC17A1; SLC17A3; SLC17A2; HFE; HIST1H2BC; MIR3143; ZDHHC20P2; TMPRSS6 | SCGN; HIST1H2AA; SLC17A1; SLC17A3; SLC17A2; TRIM38; U91328.21; U91328.19; U91328.22; HIST1H3A; HIST1H4A; HIST1H4B; HIST1H3B; HIST1H2BB; U91328.2; HIST1H3C; HIST1H1C; HFE; HIST1H4C; HIST1H1T; HIST1H2BC; HIST1H2AC; HCG4P7; ABO; HK1; TST; MPST; KCTD17; TMPRSS6 | 90 | 246 | 16 | 74 | 230 | 0 | 0 | 0 | 0 | 18 | 16 | 3 | 16 |
| HGB | Hemoglobin | blood | 0 | 0 | 0 | 0 | 0 | | | 3 | 1 | 0 | 3 | 1 | 0 | 0 | 0 | 0 | 0 | | 0 | 0 |
| HgbA1C | Hemoglobin A1c (Glycated) Hemoglobin A1c (Glycated) | metabolic | 61 | 75 | 20 | 41 | 55 | XXbac-BPG16N22.5; PRRC2A; BAG6; GPANK1; RNF5; AGER; PBX2; HLA-DQA1; HLA-DQB1; MTCO3P1; HK1; TCF7L2 | GPSM3; NOTCH4; BTNL2; HLA-DRA; HLA-DRB9; HLA-DQB1; MTCO3P1; HK1; TCF7L2 | 225 | 187 | 57 | 168 | 130 | 0 | 0 | 0 | 0 | 11 | 2 | 8 | 11 |

| all_labs | full_name | group | male.significant.variants | female.significant.variants | both.significant.variants | male.only.significant.variants | female.only.significant.variants | male.annotated.genes.significant.variants | female.annotated.genes.significant.variants | male.suggestive.variants | female.suggestive.variants | both.suggestive.variants | male.only.suggestive.variants | female.only.suggestive.variants | female.significant.x.variants | male.significant.x.variants | female.suggestive.x.variants | male.suggestive.x.variants | n_loci | n_loci_noHLA | P1e6_both | P1e2_both |
|---|---|---|---|---|---|---|---|---|---|---|---|---|---|---|---|---|---|---|---|---|---|---|
| HomocB | Homocysteine in Serum or Plasma | metabolic | 0 | 0 | 0 | 0 | 0 | | | 2 | 1 | 0 | 2 | 1 | 0 | 0 | 0 | 0 | 0 | | 0 | 0 |
| HORIZONTAL AXIS: INITIAL 40 MS | QRS axis | cardiovascular | 0 | 35 | 0 | 0 | 35 | | RP4-601P9.1; RP11-25E2.1 | 5 | 46 | 4 | 1 | 42 | 0 | 0 | 0 | 0 | 1 | 1 | 1 | 1 |
| HORIZONTAL AXIS: TERMINAL 40 MS | QRS axis | cardiovascular | 0 | 0 | 0 | 0 | 0 | | | 2 | 0 | 0 | 2 | 0 | 0 | 0 | 0 | 0 | 0 | | 0 | 0 |
| HPIndx | H. pylori EIA index | immune / inflammatory | 0 | 0 | 0 | 0 | 0 | | | 1 | 0 | 0 | 1 | 0 | 0 | 0 | 0 | 0 | 0 | | 0 | 0 |
| HR_INTERVAL | Heart rate | vitals | 0 | 0 | 0 | 0 | 0 | | | 1 | 4 | 0 | 1 | 4 | 0 | 0 | 0 | 0 | 0 | | 0 | 0 |
| HSCRP | C reactive protein (high sensitivity) | immune / inflammatory | 0 | 0 | 0 | 0 | 0 | | | 0 | 24 | 0 | 0 | 24 | 0 | 0 | 0 | 0 | 0 | | 0 | 0 |
| IC_PPR | IC PPR | lung | 0 | 0 | 0 | 0 | 0 | | | 0 | 0 | 0 | 0 | 0 | 0 | 0 | 0 | 0 | 0 | | 0 | 0 |
| IC_PRD | IC PRD | lung | 0 | 0 | 0 | 0 | 0 | | | 0 | 0 | 0 | 0 | 0 | 0 | 0 | 0 | 0 | 0 | | 0 | 0 |
| IC_ZBD | IC ZBD | lung | 0 | 0 | 0 | 0 | 0 | | | 0 | 0 | 0 | 0 | 0 | 0 | 0 | 0 | 0 | 0 | | 0 | 0 |
| IctIdx | Icteric index of Serum or Plasma | metabolic | 42 | 0 | 0 | 42 | 0 | UGT1A11P; UGT1A8 | | 86 | 3 | 0 | 86 | 3 | 0 | 0 | 0 | 0 | 1 | 1 | 0 | 1 |
| IgA | IgA [Mass/volume] in Serum or Plasma | immune / inflammatory | 0 | 0 | 0 | 0 | 0 | | | 4 | 2 | 0 | 4 | 2 | 0 | 0 | 0 | 0 | 0 | | 0 | 0 |
| IGAB | Immature granulocytes [Presence] in Blood by Automated Count | immune / inflammatory | 57 | 43 | 30 | 27 | 13 | RP1-111C20.3; RP11-13P5.1; CCM2; TBC1D13; ENDOG; C9orf114; CCBL1; LRRC8A; PRTFDC1 | ITGA4; RP1-111C20.3; RP11-13P5.1; TBC1D13; ENDOG; C9orf114; CCBL1; LRRC8A; DEPDC5; C22orf24 | 129 | 75 | 47 | 82 | 28 | 0 | 0 | 0 | 1 | 9 | 9 | 6 | 9 |
| IgEQnt | IgE [Mass/volume] in Serum | immune / inflammatory | 0 | 0 | 0 | 0 | 0 | | | 0 | 0 | 0 | 0 | 0 | 0 | 0 | 0 | 0 | 0 | | 0 | 0 |

| all_labs | full_name | group | male.significant.variants | female.significant.variants | both.significant.variants | male.only.significant.variants | female.only.significant.variants | male.annotated.genes.significant.variants | female.annotated.genes.significant.variants | male.suggestive.variants | female.suggestive.variants | both.suggestive.variants | male.only.suggestive.variants | female.only.suggestive.variants | female.significant.x.variants | male.significant.x.variants | female.suggestive.x.variants | male.suggestive.x.variants | n_loci | n_loci_noHLA | P1e6_both | P1e2_both |
|---|---|---|---|---|---|---|---|---|---|---|---|---|---|---|---|---|---|---|---|---|---|---|
| IgG-Q | IgG Quantitative blood | immune / inflammatory | 0 | 0 | 0 | 0 | 0 | | | 0 | 0 | 0 | 0 | 0 | 0 | 0 | 0 | 0 | 0 | | 0 | 0 |
| IgM-Q | IgM Quantitative blood | immune / inflammatory | 0 | 38 | 0 | 0 | 38 | | PDE11A | 1 | 50 | 0 | 1 | 50 | 0 | 0 | 0 | 0 | 1 | 1 | 0 | 0 |
| IGRE | Immature granulocytes/100 leukocytes in Blood | immune / inflammatory | 195 | 71 | 61 | 134 | 10 | RP1-111C20.4; RP1-111C20.3; RP11-13P5.1; MYO1G; CCM2; NACAD; TBRG4; CHD7; ZDHHC12; TBC1D13; ENDOG; C9orf114; CCBL1; LRRC8A; PRTFDC1; DEPDC5; C22orf24 | ITGA4; RP1-111C20.3; RP11-13P5.1; CHD7; ZDHHC12; TBC1D13; ENDOG; C9orf114; CCBL1; LRRC8A; DEPDC5; C22orf24 | 241 | 91 | 82 | 159 | 9 | 0 | 0 | 0 | 3 | 15 | 15 | 10 | 15 |
| Infant Apgar 5 M Hr 1 | 5 minute Apgar heart rate | vitals | | 0 | | | | | | | 2 | | | | 0 | | 0 | | 0 | | 0 | 0 |
| Infant Apgar 5 M Hr 4 | 5 minute Apgar heart rate | vitals | | 0 | | | | | | | 2 | | | | 0 | | 0 | | 0 | | 0 | 0 |
| Infant Apgar Color 1 M 1 | 1 minute Apgar color | vitals | | 1 | | | | | METTL21C | | 3 | | | | 0 | | 0 | | 1 | 1 | 0 | 0 |
| Infant Apgar Color 5 M 1 | 5 minute Apgar color | vitals | | 0 | | | | | | | 0 | | | | 0 | | 0 | | 0 | | 0 | 0 |
| Infant Apgar Muscle 1 M 1 | 1 minute Apgar muscle | vitals | | 0 | | | | | | | 2 | | | | 0 | | 0 | | 0 | | 0 | 0 |
| Infant Apgar Muscle 5 M 1 | 5 minute Apgar muscle | vitals | | 0 | | | | | | | 0 | | | | 0 | | 0 | | 0 | | 0 | 0 |
| Infant Apgar Resp 1 M 1 | 1 minute Apgar respiratory rate | vitals | | 0 | | | | | | | 2 | | | | 0 | | 0 | | 0 | | 0 | 0 |
| Infant Apgar Resp 5 M 1 | 5 minute Apgar respiratory rate | vitals | | 5 | | | | | ARHGEF7 | | 6 | | | | 0 | | 0 | | 1 | 1 | 0 | 0 |
| Infant Apgar Rflx 1 M 1 | 1 minute Apgar reflex | vitals | | 0 | | | | | | | 0 | | | | 0 | | 0 | | 0 | | 0 | 0 |
| Infant Apgar Rflx 5 M 1 | 5 minute Apgar reflex | vitals | | 0 | | | | | | | 5 | | | | 0 | | 0 | | 0 | | 0 | 0 |

| all_labs | full_name | group | male.significant.variants | female.significant.variants | both.significant.variants | male.only.significant.variants | female.only.significant.variants | male.annotated.genes.significant.variants | female.annotated.genes.significant.variants | male.suggestive.variants | female.suggestive.variants | both.suggestive.variants | male.only.suggestive.variants | female.only.suggestive.variants | female.significant.x.variants | male.significant.x.variants | female.suggestive.x.variants | male.suggestive.x.variants | n_loci | n_loci_noHLA | P1e6_both | P1e2_both |
|---|---|---|---|---|---|---|---|---|---|---|---|---|---|---|---|---|---|---|---|---|---|---|
| Infant Apgar Score 1 M 1 | 1 minute Apgar score | vitals | | 0 | | | | | | | 0 | | | | 0 | | 0 | | 0 | | 0 | 0 |
| Infant Apgar Score 5 M 1 | 5 minute Apgar score | vitals | | 0 | | | | | | | 0 | | | | 0 | | 0 | | 0 | | 0 | 0 |
| INITIAL_40_MS | Initial 40 ms | cardiovascular | 0 | 0 | 0 | 0 | 0 | | | 1 | 6 | 0 | 1 | 6 | 0 | 0 | 0 | 0 | 0 | | 0 | 0 |
| Insuln | Insulin [Units/volume] in Serum or Plasma | endocrine | 0 | 0 | 0 | 0 | 0 | | | 0 | 0 | 0 | 0 | 0 | 0 | 0 | 0 | 0 | 0 | | 0 | 0 |
| IPF | Platelets reticulated/100 platelets in Blood by Automated count | blood | 0 | 0 | 0 | 0 | 0 | | | 3 | 10 | 0 | 3 | 10 | 0 | 0 | 0 | 0 | 0 | | 0 | 0 |
| IRF | Immature reticulocytes/Reticulocytes.total in Blood | blood | 0 | 0 | 0 | 0 | 0 | | | 1 | 0 | 0 | 1 | 0 | 0 | 0 | 0 | 0 | 0 | | 0 | 0 |
| Iron | Iron [Mass/volume] in Serum or Plasma | blood | 76 | 134 | 75 | 1 | 59 | LRRC16A; SCGN; HIST1H2AA; SLC17A1; SLC17A3; SLC17A2; U91328.21; U91328.22; HIST1H3A; HIST1H4A; HIST1H4B; HIST1H3B; HIST1H2BB; U91328.2; HIST1H3C; HIST1H1C; HFE; HIST1H2BC; TMPRSS6 | LRRC16A; SCGN; HIST1H2AA; SLC17A1; SLC17A3; SLC17A2; TRIM38; U91328.21; U91328.19; U91328.22; HIST1H3A; HIST1H4A; HIST1H4B; HIST1H3B; HIST1H2AB; HIST1H2BB; U91328.2; HIST1H3C; HIST1H1C; HFE; HIST1H4C; HIST1H1T; HIST1H2BC; HIST1H2AC; MPST; KCTD17; TMPRSS6 | 100 | 241 | 99 | 1 | 142 | 0 | 0 | 0 | 0 | 9 | 9 | 6 | 9 |
| K-WB | Potassium whole blood | metabolic | 0 | 0 | 0 | 0 | 0 | | | 1 | 21 | 0 | 1 | 21 | 0 | 0 | 21 | 0 | 0 | | 0 | 0 |
| K | Potassium blood | metabolic | 16 | 71 | 0 | 16 | 71 | HLA-DQA1; HLA-DQB1; MTCO3P1 | KLHL3; RP11-381K20.5; HNRNPA0; RP11-381K20.4; NPY6R; RP11-381K20.2; MYOT; PKD2L2; FAM13B; WNT8A | 138 | 112 | 0 | 138 | 112 | 0 | 0 | 0 | 0 | 4 | 3 | 0 | 4 |
| KapLCQ | Kappa light chains.free [Mass/volume] in Serum | toxicology / pharmacology | 0 | 0 | 0 | 0 | 0 | | | 0 | 0 | 0 | 0 | 0 | 0 | 0 | 0 | 0 | 0 | | 0 | 0 |

| all_labs | full_name | group | male.significant.variants | female.significant.variants | both.significant.variants | male.only.significant.variants | female.only.significant.variants | male.annotated.genes.significant.variants | female.annotated.genes.significant.variants | male.suggestive.variants | female.suggestive.variants | both.suggestive.variants | male.only.suggestive.variants | female.only.suggestive.variants | female.significant.x.variants | male.significant.x.variants | female.suggestive.x.variants | male.suggestive.x.variants | n_loci | n_loci_noHLA | P1e6_both | P1e2_both |
|---|---|---|---|---|---|---|---|---|---|---|---|---|---|---|---|---|---|---|---|---|---|---|
| KLRat | Kappa light chains.free/Lambda light chains.free [Mass Ratio] in Serum | immune / inflammatory | 0 | 0 | 0 | 0 | 0 | | | 1 | 0 | 0 | 1 | 0 | 0 | 0 | 0 | 0 | 0 | | 0 | 0 |
| LA | Lactate [Moles/volume] in Serum or Plasma | metabolic | 0 | 0 | 0 | 0 | 0 | | | 0 | 5 | 0 | 0 | 5 | 0 | 0 | 0 | 0 | 0 | | 0 | 0 |
| LAC | Lactate Whole Blood | metabolic | 0 | 0 | 0 | 0 | 0 | | | 3 | 0 | 0 | 3 | 0 | 0 | 0 | 0 | 0 | 0 | | 0 | 0 |

| all_labs | full_name | group | male.significant.variants | female.significant.variants | both.significant.variants | male.only.significant.variants | female.only.significant.variants | male.annotated.genes.significant.variants | female.annotated.genes.significant.variants | male.suggestive.variants | female.suggestive.variants | both.suggestive.variants | male.only.suggestive.variants | female.only.suggestive.variants | female.significant.x.variants | male.significant.x.variants | female.suggestive.x.variants | male.suggestive.x.variants | n_loci | n_loci_noHLA | P1e6_both | P1e2_both |
|---|---|---|---|---|---|---|---|---|---|---|---|---|---|---|---|---|---|---|---|---|---|---|
| LamLCQ | Lambda light chains.free [Mass/volume] in Serum or Plasma | immune / inflammatory | 0 | 0 | 0 | 0 | 0 | | | 2 | 1 | 0 | 2 | 1 | 0 | 0 | 0 | 0 | 0 | | 0 | 0 |
| LDH | Lactate dehydrogenase (LDH) | metabolic | 0 | 0 | 0 | 0 | 0 | | | 2 | 2 | 0 | 2 | 2 | 0 | 0 | 0 | 0 | 0 | | 0 | 0 |
| LDL_CALCULATED | LDL_CALCULATED | metabolic | 0 | 0 | 0 | 0 | 0 | | | 1 | 0 | 0 | 1 | 0 | 0 | 0 | 0 | 0 | 0 | | 0 | 0 |
| LDL_HDL_RATIO | Cholesterol.total/Cholesterol in HDL [Mass Ratio] in Serum or Plasma | metabolic | 0 | 0 | 0 | 0 | 0 | | | 0 | 3 | 0 | 0 | 3 | 0 | 0 | 0 | 0 | 0 | | 0 | 0 |
| LDL-C | LDL cholesterol | metabolic | 69 | 345 | 69 | 0 | 276 | CELSR2; PSRC1; SMARCA4; LDLR; BCL3; PVRL2; APOE; APOC1; APOC1P1 | SARS; CELSR2; PSRC1; RP11-116D2.1; APOB; TDRD15; RP11-79J24.1; AC067959.1; CTD-2235C13.2; LPA; RP1-81D8.4; SMARCA4; LDLR; CTB-171A8.1; BCL3; BCAM; PVRL2; TOMM40; APOE; APOC1; APOC1P1 | 84 | 439 | 77 | 7 | 362 | 0 | 0 | 0 | 0 | 29 | 29 | 9 | 28 |

| all_labs | full_name | group | male.significant.variants | female.significant.variants | both.significant.variants | male.only.significant.variants | female.only.significant.variants | male.annotated.genes.significant.variants | female.annotated.genes.significant.variants | male.suggestive.variants | female.suggestive.variants | both.suggestive.variants | male.only.suggestive.variants | female.only.suggestive.variants | female.significant.x.variants | male.significant.x.variants | female.suggestive.x.variants | male.suggestive.x.variants | n_loci | n_loci_noHLA | P1e6_both | P1e2_both |
|---|---|---|---|---|---|---|---|---|---|---|---|---|---|---|---|---|---|---|---|---|---|---|
| LEUKOREDUCED_RANDOM_PLATELETS | LEUKOREDUCED_RANDOM_PLATELETS | immune / inflammatory | 0 | 0 | 0 | 0 | 0 | | | 0 | 0 | 0 | 0 | 0 | 0 | 0 | 0 | 0 | 0 | | 0 | 0 |
| LH1 | Lutropin [Units/volume] in Serum or Plasma | endocrine | 0 | 0 | 0 | 0 | 0 | | | 1 | 0 | 0 | 1 | 0 | 0 | 0 | 0 | 0 | 0 | | 0 | 0 |
| Lipase | Lipase | toxicology / pharmacology | 0 | 0 | 0 | 0 | 0 | | | 0 | 21 | 0 | 0 | 21 | 0 | 0 | 0 | 0 | 0 | | 0 | 0 |
| LipIdx | Lipemic index of Serum or Plasma | metabolic | 4 | 0 | 0 | 4 | 0 | APOE; APOC1 | | 4 | 7 | 0 | 4 | 7 | 0 | 0 | 0 | 0 | 1 | 1 | 0 | 1 |
| Luc% | Luc% | immune / inflammatory | 2 | 2 | 2 | 0 | 0 | BZRAP1-AS1 | BZRAP1-AS1 | 2 | 2 | 2 | 0 | 0 | 0 | 0 | 0 | 0 | 1 | 1 | 1 | 1 |
| LucAbs | Leukocytes [#/volume] in Blood by Automated count | blood | 0 | 1 | 0 | 0 | 1 | | BZRAP1-AS1 | 0 | 3 | 0 | 0 | 3 | 0 | 0 | 0 | 0 | 1 | 1 | 0 | 1 |
| Lym | Lymphocytes [#/volume] in Blood | immune / inflammatory | 0 | 0 | 0 | 0 | 0 | | | 0 | 8 | 0 | 0 | 8 | 0 | 0 | 0 | 0 | 0 | | 0 | 0 |
| LymAbs | Lymphocytes [#/volume] in Blood by Automated count | immune / inflammatory | 2 | 68 | 0 | 2 | 68 | GGT5 | HCG20; HLA-C; USP8P1; RPL3P2; HLA-B; XXbac-BPG248L24.12; DHFRP2; RNU6-283P; AL671883.1; FGFR3P1; ZDHHC20P2; HLA-S; MICA; HCP5 | 41 | 226 | 0 | 41 | 226 | 0 | 0 | 0 | 0 | 6 | 1 | 0 | 5 |
| LYMPA | Lymphocytes [#/volume] in Blood | immune / inflammatory | 0 | 0 | 0 | 0 | 0 | | | 0 | 2 | 0 | 0 | 2 | 0 | 0 | 0 | 0 | 0 | | 0 | 0 |

| all_labs | full_name | group | male.significant.variants | female.significant.variants | both.significant.variants | male.only.significant.variants | female.only.significant.variants | male.annotated.genes.significant.variants | female.annotated.genes.significant.variants | male.suggestive.variants | female.suggestive.variants | both.suggestive.variants | male.only.suggestive.variants | female.only.suggestive.variants | female.significant.x.variants | male.significant.x.variants | female.suggestive.x.variants | male.suggestive.x.variants | n_loci | n_loci_noHLA | P1e6_both | P1e2_both |
|---|---|---|---|---|---|---|---|---|---|---|---|---|---|---|---|---|---|---|---|---|---|---|
| LYMPAB | Lymphocytes [#/volume] in Blood by Automated count | immune / inflammatory | 237 | 805 | 211 | 26 | 594 | RPSAP2; AL662890.1; NOL5BP; HLA-V; TRIM26; HCG17; TRIM39; HLA-N; HCG19P; UBQLN1P1; MICC; MUC22; RNU6-1133P; C6orf15; PSORS1C1; HCG27; XXbac-BPG299F13.16; HLA-C; USP8P1; RPL3P2; XXbac-BPG248L24.10; HLA-B; XXbac-BPG248L24.12; DHFRP2; RNU6-283P; AL671883.1; FGFR3P1; ZDHHC20P2; HLA-S; MICA; HCP5; MICB; XXbac-BPG16N22.5; ATP6V1G2-DDX39B; TNF; NCR3; PRRC2A; APOM; CSNK2B; ABHD16A; XXbac-BPG32J3.20; CLIC1; MSH5; VWA7; VARS; LSM2; HSPA1L; HSPA1A; C6orf48; NEU1; SLC44A4; C2; CFB; NELFE; SKIV2L; STK19; TNXB; C6orf10 | XXbac-BPGBPG34I8.1; HNRNPA1P1; XXbac-BPG34I8.3; RNU6-471P; TRNAI6; RP1-15D7.1; RP1-97D16.1; RSL24D1P1; HIST1H4PS1; HIST1H2BM; HIST1H4K; HIST1H2AK; HIST1H2BN; HIST1H2AL; HIST1H1B; HIST1H4L; RNU7-26P; OR2B2; OR2W6P; RPLP2P1; OR2W4P; IQCB2P; OR2W2P; OR2B7P; OR1F12; ZSCAN12P1; RP1-265C24.5; ZSCAN16-AS1; ZKSCAN8; ZNF192P1; AL022393.7; ZNF192P2; TOB2P1; ZSCAN9; ZKSCAN4; RP5-874C20.3; PGBD1; RP5-874C20.6; ZSCAN31; ZKSCAN3; ZSCAN12; RNU2-45P; ZSCAN23; COX11P1; RPSAP2; AL662890.1; NOL5BP; AL662890.3; XXbac-BPG308K3.5; XXbac-BPG308K3.6; C6orf100; HCG15; HCG16; ZNF311; OR2AD1P; OR2W1; OR2P1P; SAR1P1; OR2B3; XXbac-BCX196D17.5; OR2N1P; OR2J2; OR2J4P; OR2H4P; XXbac-BPG308J9.3; OR14J1; OR5V1; OR11A1; MAS1LP1; MAS1L; XXbac-BPG13B8.10; OR2I1P; UBD; GABBR1; SUMO2P1; ZFP57; HLA-V; ZNRD1; RNF39; TRIM31; TRIM31-AS1; TRIM40; TRIM10; TRIM15; TRIM26; HCG17; TRIM39; TRIM39-RPP21; HLA-N; HCG19P; UBQLN1P1; MICC; TMPOP1; SUCLA2P1; RANP1; XXbac-BPG249D20.9; GNL1; ABCF1; ATAT1; C6orf136; PPP1R18; FLOT1; RN7SL353P; HCG20; LINC00243; XXbac-BPG27H4.8; RN7SKP186; DDR1-AS1; DDR1; RN7SL175P; GTF2H4; VARS2; SFTA2; DPCR1; HCG21; MUC21; MUC22; HCG22; RNU6-1133P; C6orf15; PSORS1C1; TCF19; HCG27; XXbac-BPG299F13.16; HLA-C; USP8P1; RPL3P2; WASF5P; XXbac-BPG248L24.13; XXbac-BPG248L24.10; HLA-B; XXbac-BPG248L24.12; DHFRP2; RNU6-283P; AL671883.1; FGFR3P1; ZDHHC20P2; HLA-S; MICA; HCP5; MICB; XXbac-BPG16N22.5; RPL15P4; ATP6V1G2-DDX39B; TNF; NCR3; PRRC2A; APOM; CSNK2B; ABHD16A; XXbac-BPG32J3.20; CLIC1; MSH5; VWA7; VARS; LSM2; HSPA1L; HSPA1A; C6orf48; NEU1; SLC44A4; C2; CFB; HLA-DRA; HLA-DQA1; CD69; EPS15L1 | 564 | 1231 | 476 | 88 | 755 | 0 | 0 | 0 | 0 | 30 | 11 | 17 | 30 |
| LYMPHOCYTES | LYMPHOCYTES | immune / inflammatory | 0 | 0 | 0 | 0 | 0 | | | 0 | 0 | 0 | 0 | 0 | 0 | 0 | 0 | 0 | 0 | | 0 | 0 |
| LYMPM | Lymphocytes/100 leukocytes in Blood by Manual count | immune / inflammatory | 0 | 21 | 0 | 0 | 21 | | MTND6P12; RP11-585K14.2 | 0 | 23 | 0 | 0 | 23 | 21 | 0 | 23 | 0 | 1 | 1 | 0 | 0 |

| all_labs | full_name | group | male.significant.variants | female.significant.variants | both.significant.variants | male.only.significant.variants | female.only.significant.variants | male.annotated.genes.significant.variants | female.annotated.genes.significant.variants | male.suggestive.variants | female.suggestive.variants | both.suggestive.variants | male.only.suggestive.variants | female.only.suggestive.variants | female.significant.x.variants | male.significant.x.variants | female.suggestive.x.variants | male.suggestive.x.variants | n_loci | n_loci_noHLA | P1e6_both | P1e2_both |
|---|---|---|---|---|---|---|---|---|---|---|---|---|---|---|---|---|---|---|---|---|---|---|
| LYMPRE | Lymphocytes/100 leukocytes in Blood by Automated count | immune / inflammatory | 0 | 1 | 0 | 0 | 1 | | RP11-334L9.1 | 0 | 33 | 0 | 0 | 33 | 0 | 0 | 0 | 0 | 1 | 1 | 0 | 0 |
| MBRat | Creatine kinase.MB/Creatine kinase.total in Serum or Plasma | cardiovascular | 4 | 0 | 0 | 4 | 0 | CTD-2337J16.1; LILRB5 | | 12 | 55 | 0 | 12 | 55 | 0 | 0 | 0 | 0 | 2 | 2 | 0 | 2 |

| all_labs | full_name | group | male.significant.variants | female.significant.variants | both.significant.variants | male.only.significant.variants | female.only.significant.variants | male.annotated.genes.significant.variants | female.annotated.genes.significant.variants | male.suggestive.variants | female.suggestive.variants | both.suggestive.variants | male.only.suggestive.variants | female.only.suggestive.variants | female.significant.x.variants | male.significant.x.variants | female.suggestive.x.variants | male.suggestive.x.variants | n_loci | n_loci_noHLA | P1e6_both | P1e2_both |
|---|---|---|---|---|---|---|---|---|---|---|---|---|---|---|---|---|---|---|---|---|---|---|
| MCH | MCH [Entitic mass] by Automated count | blood | 1141 | 2476 | 957 | 184 | 1519 | TNK2; RNU2-11P; TFRC; LINC00885; RP11-545H22.1; AC006552.1; LRRC16A; SCGN; HIST1H2AA; SLC17A1; SLC17A3; HIST1H2APS2; SLC17A2; TRIM38; U91328.21; U91328.19; U91328.22; HIST1H1PS2; HIST1H3A; HIST1H4A; HIST1H4B; HIST1H3B; HIST1H2AB; HIST1H2BB; U91328.2; HIST1H3C; HIST1H1C; HFE; HIST1H4C; HIST1H1T; HIST1H2BC; HIST1H2AC; HIST1H4F; HIST1H4G; HIST1H3F; HIST1H2BH; HIST1H3G; HIST1H2APS4; HIST1H2BI; HIST1H4H; HIST1H3PS1; RNU6-1259P; BTN1A1; VN1R13P; MIR3143; HIST1H2BO; OR2W6P; OR2W4P; IQCB2P; PGBD1; ZSCAN31; ZKSCAN3; RNU2-45P; ZSCAN23; OR2E1P; GPX6; XXbac-BPG308K3.5; OR2AD1P; OR2P1P; MOG; XXbac-BPG170G13.31; HLA-G; HLA-W; ZNRD1; AL669914.1; TRIM10; TRIM15; TRIM26; HCG17; TRIM39; UBQLN1P1; MICC; HLA-B; PGC; RP11-298J23.8; FRS3; PRICKLE4; USP49; MED20; BYSL; CCND3; C6orf183; RP11-425D10.1; CCDC162P; RP3-352A20.1; ALDH8A1; HBS1L; MYB; RP11-12A2.3; IKZF1; MOSPD3; TFR2; GIGYF1; POP7; EPO; FNTB; NPRL3; C18orf25; HDGFRP2; KLF1; GCDH; TEX33; TST; MPST; KCTD17; TMPRSS6; SCO2; ODF3B | BCL11A; MIR4435-1HG; TFRC; LINC00885; RP11-231C18.2; RP11-545H22.1; AC006552.1; LRRC16A; SCGN; HIST1H2AA; SLC17A4; SLC17A1; SLC17A3; HIST1H2APS2; SLC17A2; TRIM38; U91328.21; U91328.19; U91328.22; HIST1H1PS2; HIST1H3A; HIST1H4A; HIST1H4B; HIST1H3B; HIST1H2AB; HIST1H2BB; U91328.2; HIST1H3C; HIST1H1C; HFE; HIST1H4C; HIST1H1T; HIST1H2BC; HIST1H2AC; HIST1H3G; HIST1H2APS4; HIST1H4H; HIST1H3PS1; RNU6-1259P; BTN3A2; BTN2A2; BTN3A1; BTN2A3P; BTN3A3; BTN2A1; BTN1A1; RNU6-502P; HCG11; CTA-14H9.5; HMGN4; ABT1; RP11-457M11.6; VN1R14P; RP11-457M11.2; ZNF322; RP11-457M11.5; GUSBP2; VN1R13P; MIR3143; VN1R10P; HIST1H2BO; OR2W6P; OR2W4P; IQCB2P; PGBD1; ZSCAN31; ZKSCAN3; RNU2-45P; ZSCAN23; COX11P1; OR2E1P; GPX6; XXbac-BPG308K3.5; OR2AD1P; OR2P1P; OR2J3; MOG; XXbac-BPG170G13.31; HLA-V; HLA-G; HLA-W; MICD; HCG9; DDX39BP2; MCCD1P2; ZNRD1-AS1; ZNRD1; PPP1R11; AL669914.1; TRIM31; TRIM31-AS1; TRIM40; TRIM10; TRIM15; TRIM26; HCG17; HCG19P; UBQLN1P1; MICC; TMPOP1; HLA-E; XXbac-BPG249D20.9; ATAT1; XXbac-BPG27H4.8; DDR1-AS1; DDR1; RN7SL175P; PGC; RP11-298J23.8; FRS3; PRICKLE4; USP49; MED20; BYSL; CCND3; C6orf183; RP11-425D10.1; PTCHD3P3; CCDC162P; RP3-352A20.1; ALDH8A1; HBS1L; MYB; RP11-12A2.3; IKZF1; AP4M1; PILRA; ZCWPW1; LRCH4; FBXO24; PCOLCE-AS1; MOSPD3; TFR2; ACTL6B; GNB2; GIGYF1; POP7; EPO; ZAN; RP11-307I14.4; RCL1; TMEM72-AS1; ALOX5; RP11-67C2.2; MARCH8; ZFAND4; HK1; FNTB; IGDCC4; DPP8; PTPLAD1; VWA9; SLC24A1; DENND4A; RAB11A; NPRL3; Z69890.1; C18orf25; HDGFRP2; HOOK2; MAST1; DNASE2; CTD-2265O21.7; KLF1; GCDH; SYCE2; MIR5695; FARSA; CALR; RAD23A; CTD-2540B15.12; RBM38; LL22NC01-81G9.3; TEX33; TST; MPST; KCTD17; TMPRSS6; NCAPH2; SCO2; TYMP; ODF3B | 1772 | 3527 | 1363 | 409 | 2164 | 0 | 0 | 0 | 0 | 146 | 134 | 90 | 141 |

| all_labs | full_name | group | male.significant.variants | female.significant.variants | both.significant.variants | male.only.significant.variants | female.only.significant.variants | male.annotated.genes.significant.variants | female.annotated.genes.significant.variants | male.suggestive.variants | female.suggestive.variants | both.suggestive.variants | male.only.suggestive.variants | female.only.suggestive.variants | female.significant.x.variants | male.significant.x.variants | female.suggestive.x.variants | male.suggestive.x.variants | n_loci | n_loci_noHLA | P1e6_both | P1e2_both |
|---|---|---|---|---|---|---|---|---|---|---|---|---|---|---|---|---|---|---|---|---|---|---|
| MCHC | MCHC [Mass/volume] by Automated count | blood | 540 | 1401 | 467 | 73 | 934 | SMIM1; LRRC47; CEP104; DFFB; ATP2B4; TRIM58; LINC00885; LRRC16A; SCGN; HIST1H2AA; SLC17A1; SLC17A3; HIST1H2APS2; SLC17A2; TRIM38; U91328.21; U91328.19; U91328.22; HIST1H3A; HIST1H4A; HIST1H4B; HIST1H3B; HIST1H2AB; HIST1H2BB; U91328.2; HIST1H3C; HIST1H1C; HFE; HIST1H4C; HIST1H1T; HIST1H2BC; HIST1H2AC; HIST1H3G; HIST1H3PS1; BTN1A1; VN1R13P; MIR3143; HIST1H2BO; OR2W6P; OR2W4P; IQCB2P; PGBD1; ZSCAN31; ZKSCAN3; RNU2-45P; ZSCAN23; OR2E1P; GPX6; XXbac-BPG308K3.5; OR2AD1P; OR2P1P; HLA-G; MICF; HLA-W; ZNRD1; AL669914.1; TRIM40; TRIM10; TRIM15; TRIM26; HCG17; TRIM39; HCG19P; UBQLN1P1; MICC; XXbac-BPG248L24.12; MICB; NCR3; UQCRHP1; AIF1; PRRC2A; BAG6; CSNK2B; CSNK2B-LY6G5B-1181; LY6G5B; LY6G5C; MSH5; C6orf48; SLC44A4; C2; TNXB; PRRT1; PPT2; XXbac-BPG154L12.4; HBS1L; SLC20A2; SMIM19; RP11-503E24.3; FNTB; HBZ; PIEZO1; FAM222B; RP11-116O18.3; SMG9; TST; MPST; KCTD17; TMPRSS6 | SMIM1; LRRC47; CEP104; DFFB; OR10X1; SPTA1; ATP2B4; TRIM58; IL1F10; LRRC16A; SCGN; HIST1H2AA; SLC17A4; SLC17A1; SLC17A3; HIST1H2APS2; SLC17A2; TRIM38; U91328.21; U91328.19; U91328.22; HIST1H1PS2; HIST1H3A; HIST1H4A; HIST1H4B; HIST1H3B; HIST1H2AB; HIST1H2BB; U91328.2; HIST1H3C; HIST1H1C; HFE; HIST1H4C; HIST1H1T; HIST1H2BC; HIST1H2AC; BTN1A1; VN1R13P; MIR3143; HIST1H2BO; OR2W6P; OR2W4P; IQCB2P; PGBD1; ZSCAN31; ZKSCAN3; RNU2-45P; ZSCAN23; COX11P1; OR2E1P; GPX6; XXbac-BPG308K3.5; OR2AD1P; OR2P1P; MOG; HLA-F; HLA-F-AS1; IFITM4P; XXbac-BPG170G13.32; XXbac-BPG170G13.31; HLA-V; MICG; HLA-G; MICF; HCG4P7; HLA-A; HLA-W; MICD; HCG9; DDX39BP2; MCCD1P2; ZNRD1-AS1; ZNRD1; PPP1R11; AL669914.1; TRIM31; TRIM31-AS1; TRIM40; TRIM10; TRIM15; TRIM26; HCG17; HCG19P; UBQLN1P1; MICC; XXbac-BPG249D20.9; ABCF1; DHX16; LINC00243; XXbac-BPG27H4.8; DDR1-AS1; DDR1; RN7SL175P; HCG22; RNU6-1133P; C6orf15; PSORS1C1; HLA-S; XXbac-BPG181B23.7; HCP5; C6orf183; RP11-425D10.1; PTCHD3P3; CCDC162P; HBS1L; SLC20A2; SMIM19; RP11-503E24.3; DPP8; PTPLAD1; VWA9; SLC24A1; DENND4A; RAB11A; DNAJA4; WDR61; CTU2; PIEZO1; UBTF; RP5-882C2.2; SHC1P2; SLC4A1; KLF1; GCDH; SMG9; KCNN4; ZNF283; TEX33; TST; MPST; KCTD17; TMPRSS6 | 1095 | 2422 | 684 | 411 | 1738 | 0 | 0 | 0 | 0 | 92 | 74 | 52 | 88 |
| MCV | MCV [Entitic volume] by Automated count | blood | 0 | 0 | 0 | 0 | 0 | | | 6 | 1 | 0 | 6 | 1 | 0 | 0 | 0 | 0 | 0 | | 0 | 0 |
| MDALab | Atypical lymphocytes | immune / inflammatory | 0 | 0 | 0 | 0 | 0 | | | 1 | 12 | 0 | 1 | 12 | 0 | 0 | 0 | 0 | 0 | | 0 | 0 |
| MDBlab | Blasts in Blood | immune / inflammatory | 0 | 0 | 0 | 0 | 0 | | | 0 | 3 | 0 | 0 | 3 | 0 | 0 | 0 | 0 | 0 | | 0 | 0 |
| MDMMab | Metamyelocytes (absolute) | immune / inflammatory | 0 | 0 | 0 | 0 | 0 | | | 7 | 1 | 0 | 7 | 1 | 0 | 0 | 0 | 0 | 0 | | 0 | 0 |
| MDMyab | Myelocytes (absolute) | immune / inflammatory | 0 | 0 | 0 | 0 | 0 | | | 4 | 0 | 0 | 4 | 0 | 0 | 0 | 0 | 0 | 0 | | 0 | 0 |

| all_labs | full_name | group | male.significant.variants | female.significant.variants | both.significant.variants | male.only.significant.variants | female.only.significant.variants | male.annotated.genes.significant.variants | female.annotated.genes.significant.variants | male.suggestive.variants | female.suggestive.variants | both.suggestive.variants | male.only.suggestive.variants | female.only.suggestive.variants | female.significant.x.variants | male.significant.x.variants | female.suggestive.x.variants | male.suggestive.x.variants | n_loci | n_loci_noHLA | P1e6_both | P1e2_both |
|---|---|---|---|---|---|---|---|---|---|---|---|---|---|---|---|---|---|---|---|---|---|---|
| MDPmab | Promyelocytes (absolute) | immune / inflammatory | 0 | 0 | 0 | 0 | 0 | | | 0 | 1 | 0 | 0 | 1 | 0 | 0 | 0 | 0 | 0 | | 0 | 0 |
| MEAN_QRS | QRS axis | cardiovascular | 38 | 142 | 0 | 38 | 142 | NRP1; RP11-90B22.1; RPL7P37; VTI1A | CASQ2; LIMCH1; RP4-601P9.1; RP11-25E2.1 | 101 | 223 | 0 | 101 | 223 | 0 | 0 | 0 | 0 | 8 | 8 | 0 | 6 |
| Meta | Metamyelocytes/100 leukocytes in Blood by Manual count | blood | 14 | 0 | 0 | 14 | 0 | RP5-998G20.1 | | 117 | 1 | 0 | 117 | 1 | 0 | 14 | 0 | 103 | 1 | 1 | 0 | 0 |
| MetHgb | Methemoglobin/Hemoglobin.total in Venous blood | blood | 0 | 0 | 0 | 0 | 0 | | | 0 | 13 | 0 | 0 | 13 | 0 | 0 | 0 | 0 | 0 | | 0 | 0 |
| Mg | Magnesium [Mass/volume] in Serum or Plasma | metabolic | 1 | 19 | 0 | 1 | 19 | DCAF6 | RP11-201K10.3; THBS3; MTX1; GBAP1 | 17 | 19 | 0 | 17 | 19 | 0 | 0 | 0 | 0 | 2 | 2 | 0 | 1 |
| MmAcid | Methyl malonic acid | metabolic | 209 | 68 | 68 | 141 | 0 | C2orf88; HIBCH; INPP1 | HIBCH; INPP1 | 235 | 256 | 208 | 27 | 48 | 0 | 0 | 0 | 0 | 2 | 2 | 2 | 2 |
| MonAbs | Absolute mono cytes | immune / inflammatory | 129 | 157 | 121 | 8 | 36 | ITGA4; RPN1; CCDC26; RNY4P18; RP11-202G18.1 | ITGA4; C3orf27; RPN1; CCDC26; RNY4P18; RP11-202G18.1; RP11-542M13.3 | 272 | 340 | 141 | 131 | 199 | 0 | 0 | 0 | 0 | 12 | 12 | 8 | 12 |
| MONOA | Monocytes [#/volume] in Blood | immune / inflammatory | 0 | 0 | 0 | 0 | 0 | | | 3 | 18 | 0 | 3 | 18 | 0 | 0 | 1 | 2 | 0 | | 0 | 0 |
| MONOAB | Monocytes [#/volume] in Blood by Automated count | immune / inflammatory | 308 | 530 | 226 | 82 | 304 | ITGA4; TMED10P2; C3orf27; RPN1; SNORA24; XXbac-BPGBPG34I8.1; HNRNPA1P1; XXbac-BPG34I8.3; RNU6-471P; TRNAI6; RP1-15D7.1; RP1-97D16.1; RSL24D1P1; HIST1H2BN; HIST1H1B; OR2B2; ZNF192P2; ZKSCAN4; SCAND3; RPSAP2; OR5V1; HLA-F; IFITM4P; UBQLN1P1; HCG27; XXbac-BPG248L24.10; HLA-B; RNU6-283P; AL671883.1; HCP5; TNXB; ATF6B; CCDC26; RP11-202G18.1; IRF8; RP11-542M13.3 | KIAA1107; RPAP2; GFI1; ITGA4; KLHL40; CCDC13; HIGD1A; ACKR2; KRBOX1; C3orf27; RPN1; SNORA24; CDK6; CCDC26; RNY4P18; RP11-202G18.1; RP11-266E8.2; UBASH3B; STXBP6; IRF8; RP11-542M13.3; RP11-542M13.2; RP11-410N8.4 | 1160 | 778 | 279 | 881 | 499 | 0 | 0 | 0 | 0 | 51 | 46 | 19 | 45 |
| Monocy | Monocytes [#/volume] in Blood by Manual count | immune / inflammatory | 113 | 321 | 112 | 1 | 209 | ITGA4; TMED10P2; C3orf27; RPN1; RP11-202G18.1; IRF8; RP11-542M13.3 | EPHX4; KIAA1107; RP4-621B10.8; RPAP2; EVI5; FAM69A; ITGA4; C3orf27; RPN1; SNORA24; CCDC26; RNY4P18; RP11-202G18.1; IRF8; RP11-542M13.3; RP11-290F20.3; RP11-290F20.1 | 210 | 616 | 174 | 36 | 442 | 0 | 0 | 0 | 0 | 26 | 26 | 15 | 22 |

| all_labs | full_name | group | male.significant.variants | female.significant.variants | both.significant.variants | male.only.significant.variants | female.only.significant.variants | male.annotated.genes.significant.variants | female.annotated.genes.significant.variants | male.suggestive.variants | female.suggestive.variants | both.suggestive.variants | male.only.suggestive.variants | female.only.suggestive.variants | female.significant.x.variants | male.significant.x.variants | female.suggestive.x.variants | male.suggestive.x.variants | n_loci | n_loci_noHLA | P1e6_both | P1e2_both |
|---|---|---|---|---|---|---|---|---|---|---|---|---|---|---|---|---|---|---|---|---|---|---|
| MONOCYTES | MONOCYTES | immune / inflammatory | 0 | 0 | 0 | 0 | 0 | | | 0 | 1 | 0 | 0 | 1 | 0 | 0 | 0 | 0 | 0 | | 0 | 0 |
| MONOM | Monocytes/100 leukocytes in Blood by Manual count | immune / inflammatory | 0 | 0 | 0 | 0 | 0 | | | 0 | 0 | 0 | 0 | 0 | 0 | 0 | 0 | 0 | 0 | | 0 | 0 |
| MONORE | Monocytes/100 leukocytes in Blood by Automated count | immune / inflammatory | 488 | 878 | 376 | 112 | 502 | RNA5SP106; MBD5; MYO3B; ITGA4; TMED10P2; C3orf27; RPN1; SNORA24; CCDC26; RP11-202G18.1; RN7SKP2; IRF8; RP11-542M13.3; RP11-542M13.2; GSDMA; PSMD3; RP11-387H17.6; CSF3; MED24 | TGFBR3; EPHX4; KIAA1107; RP4-621B10.8; RP4-775D17.1; C1orf146; GLMN; RPAP2; RN7SL824P; GFI1; EVI5; AC093577.1; ITGA4; HIGD1A; ACKR2; KRBOX1; TMED10P2; C3orf27; RPN1; SNORA24; CCHCR1; AC020743.3; AC020743.4; CCDC26; MIR4289; RNY4P18; RP11-202G18.1; FOXO1; MIR320D1; MRPS31; SLC25A15; TPTE2P5; ELF1; RP11-118F19.1; C16orf74; IRF8; RP11-542M13.3; RP11-542M13.2; GSDMA; PSMD3; CTD-2540B15.12; RP11-410N8.4; RP11-290F20.3; RP11-290F20.1 | 646 | 1125 | 475 | 171 | 650 | 0 | 0 | 0 | 0 | 69 | 68 | 40 | 63 |

| all_labs | full_name | group | male.significant.variants | female.significant.variants | both.significant.variants | male.only.significant.variants | female.only.significant.variants | male.annotated.genes.significant.variants | female.annotated.genes.significant.variants | male.suggestive.variants | female.suggestive.variants | both.suggestive.variants | male.only.suggestive.variants | female.only.suggestive.variants | female.significant.x.variants | male.significant.x.variants | female.suggestive.x.variants | male.suggestive.x.variants | n_loci | n_loci_noHLA | P1e6_both | P1e2_both |
|---|---|---|---|---|---|---|---|---|---|---|---|---|---|---|---|---|---|---|---|---|---|---|
| MPV | Platelet mean volume [Entitic volume] in Blood by Automated count | blood | 1977 | 3355 | 1708 | 269 | 1647 | KIF1B; RN7SL721P; PGD; DNM3; CNTN2; TMEM81; RBBP5; RP11-383G10.1; DSTYK; TMCC2; NUAK2; KLHDC8A; TRIM58; EHD3; ARHGEF3; KALRN; KIAA0232; IQGAP2; F2R; MEF2C; RNF145; CTB-111H14.1; ZFPM2; ASAP1; EGR2; RNU6-543P; NRBF2; JMJD1C; JMJD1C-AS1; RP11-351O1.3; REEP3; MRPL35P2; BET1L; RIC8A; SIRT3; PSMD13; NLRP6; TMEM258; FADS2; VWF; RP3-454B23.1; CD9; FAR2; ERGIC2; OVCH1-AS1; NFE2; COPZ1; BAZ2A; MORN3; TMEM120B; SETD1B; PSMD9; RP11-87C12.2; WDR66; RP11-87C12.5; BCL7A; RAD51B; TPM1; RPL35AP35; RP11-296K13.4; TAOK1; AP2B1; CD226; TPM4; SIRPD; RP4-576H24.4; SIRPB1; RP5-968J1.1; SIRPA; RP4-684O24.5; TUBB1; MIR1587; RP13-188A5.1; AL353698.1; KLF8; UBQLN2; RP11-622K12.1; RP11-445O16.3; SPIN3; RP11-445O16.2; SPIN2B; RP3-323P24.3; SPIN2A; PPP1R11P2; FAAH2; ZXDB; RP13-487C10.1; RP1-49C23.1; ZXDA; MYCLP2; KRT8P17 | KIF1B; RN7SL721P; PGD; DNM3; CNTN2; TMEM81; RBBP5; RP11-383G10.1; DSTYK; TMCC2; NUAK2; KLHDC8A; GCSAML; RP11-978I15.10; OR11L1; TRIM58; EHD3; CPS1; CAPN10; ARHGEF3; PDIA5; KIAA0232; CTD-2384B11.2; F2R; F2RL1; RN7SL208P; P4HA2; SLC22A4; AC034220.3; SLC22A5; C5orf56; RNF145; CTB-11I22.1; LRRC16A; SCGN; HLA-B; XXbac-BPG248L24.12; ZDHHC20P2; HLA-S; XXbac-BPG181B23.7; HCP5; MICA; UQCRHP1; AIF1; PRRC2A; BAG6; ABHD16A; XXbac-BPG32J3.20; DDAH2; MSH5; VWA7; NOTCH4; XXbac-BPG154L12.4; C6orf10; HLA-DRA; CTB-111H14.1; CCDC71L; ZFPM2; ASAP1; DOCK8; GFI1B; LINC00094; BRD3; RP11-349K21.1; EGR2; RNU6-543P; RP11-144G16.1; NRBF2; JMJD1C; JMJD1C-AS1; RP11-351O1.3; REEP3; MRPL35P2; BET1L; RIC8A; SIRT3; PSMD13; NLRP6; MYRF; TMEM258; FADS2; VWF; RP3-454B23.1; CD9; FAR2; ERGIC2; OVCH1-AS1; AC139768.1; DAZAP2; BIN2; CELA1; CBX5; NFE2; COPZ1; RP11-753H16.3; BAZ2A; ATP5B; PTGES3; NACA; PRIM1; HSD17B6; RP11-74M13.4; KDM2B; RNU6-1004P; ORAI1; MORN3; TMEM120B; SETD1B; HPD; PSMD9; RP11-87C12.2; WDR66; RP11-87C12.5; BCL7A; RSRC2; GRTP1; RP11-736N17.8; EXOC3L4; TPM1; PIF1; AC069368.3; MYO18A; RP11-22N12.2; TWF1P1; CRYBA1; NUFIP2; RPL35AP35; RP11-296K13.4; TAOK1; GIT1; PPP4R1; RP11-692N5.1; CABLES1; DOK6; CD226; FXYD5; RP4-576H24.4; SIRPB1; SIRPG; RP5-968J1.1; SIRPA; RP4-684O24.5; CTSZ; TUBB1; ATP5E; RPL5P34; ARFGAP3; PACSIN2; MIR1587; RP13-188A5.1; AL353698.1; KLF8; UBQLN2; RP11-431N15.2; RP11-622K12.1; RP11-445O16.3; SPIN3; RP11-445O16.2; SPIN2B; RP3-323P24.3; SPIN2A; PPP1R11P2; FAAH2; ZXDB; RP13-487C10.1; RP1-49C23.1; ZXDA; MYCLP2; KRT8P17 | 3276 | 4482 | 2561 | 715 | 1921 | 837 | 816 | 1188 | 1493 | 151 | 146 | 89 | 143 |
| Myelo | Myelocytes/100 leukocytes in Blood by Manual count | immune / inflammatory | 0 | 0 | 0 | 0 | 0 | | | 0 | 0 | 0 | 0 | 0 | 0 | 0 | 0 | 0 | 0 | | 0 | 0 |
| Na-WB | Sodium in Blood | metabolic | 0 | 0 | 0 | 0 | 0 | | | 5 | 2 | 0 | 5 | 2 | 0 | 0 | 0 | 0 | 0 | | 0 | 0 |
| Na | Sodium in Blood | metabolic | 0 | 12 | 0 | 0 | 12 | | PREX1 | 26 | 71 | 0 | 26 | 71 | 0 | 0 | 2 | 5 | 2 | 2 | 0 | 0 |
| NaslO2 | NaslO2 | blood | 0 | 0 | 0 | 0 | 0 | | | 0 | 0 | 0 | 0 | 0 | 0 | 0 | 0 | 0 | 0 | | 0 | 0 |

| all_labs | full_name | group | male.significant.variants | female.significant.variants | both.significant.variants | male.only.significant.variants | female.only.significant.variants | male.annotated.genes.significant.variants | female.annotated.genes.significant.variants | male.suggestive.variants | female.suggestive.variants | both.suggestive.variants | male.only.suggestive.variants | female.only.suggestive.variants | female.significant.x.variants | male.significant.x.variants | female.suggestive.x.variants | male.suggestive.x.variants | n_loci | n_loci_noHLA | P1e6_both | P1e2_both |
|---|---|---|---|---|---|---|---|---|---|---|---|---|---|---|---|---|---|---|---|---|---|---|
| Neut | Neutrophils [#/volume] in Blood | immune / inflammatory | 0 | 0 | 0 | 0 | 0 | | | 3 | 14 | 0 | 3 | 14 | 0 | 0 | 0 | 0 | 0 | | 0 | 0 |
| NEUTA | Neutrophils absolute | immune / inflammatory | 0 | 0 | 0 | 0 | 0 | | | 0 | 11 | 0 | 0 | 11 | 0 | 0 | 0 | 0 | 0 | | 0 | 0 |
| NEUTAB | Neutrophils absolute automated count | immune / inflammatory | 228 | 155 | 143 | 85 | 12 | RN7SL51P; RP11-642D6.1; RP11-94L15.2; IKZF3; ZPBP2; GSDMB; ORMDL3; RP11-387H17.4; LRRC3C; GSDMA; PSMD3; RP11-387H17.6; CSF3; MED24; THRA | CXCL3; PPBPP2; RP11-629B11.4; CXCL2; MTHFD2L; LRRC3C; GSDMA; PSMD3; RP11-387H17.6; CSF3; MED24; THRA | 281 | 198 | 160 | 121 | 38 | 0 | 0 | 0 | 0 | 7 | 7 | 3 | 6 |
| NEUTM | Polymorphonuclear cells/100 leukocytes in Body fluid by Manual count | immune / inflammatory | 0 | 21 | 0 | 0 | 21 | | MTND6P12; RP11-585K14.2 | 2 | 23 | 0 | 2 | 23 | 21 | 0 | 23 | 0 | 1 | 1 | 0 | 0 |
| NEUTRE | Neutrophils/100 leukocytes in Blood by Automated count | immune / inflammatory | 40 | 27 | 7 | 33 | 20 | PSMD3; RP11-387H17.6; CSF3; MED24 | PSMD3; RP11-387H17.6; MED24 | 63 | 89 | 48 | 15 | 41 | 0 | 0 | 0 | 0 | 3 | 3 | 1 | 3 |
| NonHDL | Cholesterol non HDL [Mass/volume] in Serum or Plasma | metabolic | 0 | 0 | 0 | 0 | 0 | | | 0 | 3 | 0 | 0 | 3 | 0 | 0 | 0 | 0 | 0 | | 0 | 0 |
| NRBC | NRBC | blood | 0 | 0 | 0 | 0 | 0 | | | 1 | 7 | 0 | 1 | 7 | 0 | 0 | 0 | 0 | 0 | | 0 | 0 |
| NRBC# | NRBC # | blood | 0 | 0 | 0 | 0 | 0 | | | 1 | 3 | 0 | 1 | 3 | 0 | 0 | 0 | 0 | 0 | | 0 | 0 |
| NRBCBC | Nucleated erythrocytes/100 leukocytes [Ratio] in Cerebral spinal fluid by Manual count | blood | 0 | 0 | 0 | 0 | 0 | | | 11 | 1 | 0 | 11 | 1 | 0 | 0 | 0 | 0 | 0 | | 0 | 0 |

| all_labs | full_name | group | male.significant.variants | female.significant.variants | both.significant.variants | male.only.significant.variants | female.only.significant.variants | male.annotated.genes.significant.variants | female.annotated.genes.significant.variants | male.suggestive.variants | female.suggestive.variants | both.suggestive.variants | male.only.suggestive.variants | female.only.suggestive.variants | female.significant.x.variants | male.significant.x.variants | female.suggestive.x.variants | male.suggestive.x.variants | n_loci | n_loci_noHLA | P1e6_both | P1e2_both |
|---|---|---|---|---|---|---|---|---|---|---|---|---|---|---|---|---|---|---|---|---|---|---|
| NRBCMA | Nucleated erythrocytes [#/volume] in Blood by Automated count | blood | 0 | 0 | 0 | 0 | 0 | | | 0 | 0 | 0 | 0 | 0 | 0 | 0 | 0 | 0 | 0 | | 0 | 0 |
| Nt-PTH | NT PTH | endocrine | 0 | 0 | 0 | 0 | 0 | | | 2 | 2 | 0 | 2 | 2 | 0 | 0 | 0 | 0 | 0 | | 0 | 0 |
| NtAbs | Absolute neutrophils | immune / inflammatory | 29 | 148 | 29 | 0 | 119 | PSMD3; RP11-387H17.6; CSF3; MED24 | LRRC3C; GSDMA; PSMD3; RP11-387H17.6; CSF3; MED24 | 97 | 166 | 76 | 21 | 90 | 0 | 0 | 0 | 0 | 3 | 3 | 2 | 3 |
| NTAuto | Neutrophils | immune / inflammatory | 150 | 173 | 150 | 0 | 23 | RP11-387H17.4; LRRC3C; GSDMA; PSMD3; RP11-387H17.6; CSF3; MED24; THRA | CXCL2; MTHFD2L; RP11-387H17.4; LRRC3C; GSDMA; PSMD3; RP11-387H17.6; CSF3; MED24; THRA | 161 | 204 | 153 | 8 | 51 | 0 | 0 | 0 | 0 | 5 | 5 | 3 | 5 |
| O2 Liter Flow | O2 liter flow | vitals | 0 | 0 | 0 | 0 | 0 | | | 0 | 6 | 0 | 0 | 6 | 0 | 0 | 0 | 0 | 0 | | 0 | 0 |
| O2Sat | Oxygen saturation Calculated from oxygen partial pressure in Arterial blood | vitals | 0 | 0 | 0 | 0 | 0 | | | 0 | 1 | 0 | 0 | 1 | 0 | 0 | 0 | 0 | 0 | | 0 | 0 |
| O2SAT | Oxygen saturation calculated from oxygen partial pressure in arterial blood | vitals | 0 | 1 | 0 | 0 | 1 | | CDK5RAP3 | 0 | 6 | 0 | 0 | 6 | 0 | 0 | 1 | 0 | 1 | 1 | 0 | 0 |
| O2SatC | O2 saturation (calculated) | vitals | 0 | 0 | 0 | 0 | 0 | | | 1 | 0 | 0 | 1 | 0 | 0 | 0 | 0 | 1 | 0 | | 0 | 0 |

| all_labs | full_name | group | male.significant.variants | female.significant.variants | both.significant.variants | male.only.significant.variants | female.only.significant.variants | male.annotated.genes.significant.variants | female.annotated.genes.significant.variants | male.suggestive.variants | female.suggestive.variants | both.suggestive.variants | male.only.suggestive.variants | female.only.suggestive.variants | female.significant.x.variants | male.significant.x.variants | female.suggestive.x.variants | male.suggestive.x.variants | n_loci | n_loci_noHLA | P1e6_both | P1e2_both |
|---|---|---|---|---|---|---|---|---|---|---|---|---|---|---|---|---|---|---|---|---|---|---|
| O2SatV | Oxygen saturation Calculated from oxygen partial pressure in Venous blood | vitals | 0 | 0 | 0 | 0 | 0 | | | 0 | 0 | 0 | 0 | 0 | 0 | 0 | 0 | 0 | 0 | | 0 | 0 |
| OB Infant Apgar Score 1 M 1 | 1 minute Apgar score | ob / gyn | | 0 | | | | | | | 1 | | | | 0 | | 0 | | 0 | | 0 | 0 |
| OB Infant Apgar Score 5 M 1 | 5 minute Apgar score | ob / gyn | | 0 | | | | | | | 2 | | | | 0 | | 0 | | 0 | | 0 | 0 |
| Osmol | Osmolality of Serum or Plasma | blood | 0 | 0 | 0 | 0 | 0 | | | 0 | 0 | 0 | 0 | 0 | 0 | 0 | 0 | 0 | 0 | | 0 | 0 |
| P DURATION | P wave duration | cardiovascular | 0 | 0 | 0 | 0 | 0 | | | 5 | 0 | 0 | 5 | 0 | 0 | 0 | 0 | 0 | 0 | | 0 | 0 |
| P HORIZONTAL AXIS | P wave axis horizontal plane | cardiovascular | 0 | 0 | 0 | 0 | 0 | | | 0 | 0 | 0 | 0 | 0 | 0 | 0 | 0 | 0 | 0 | | 0 | 0 |
| P_WAVE | P wave axis | cardiovascular | 0 | 0 | 0 | 0 | 0 | | | 0 | 0 | 0 | 0 | 0 | 0 | 0 | 0 | 0 | 0 | | 0 | 0 |
| Pat-PT | Prothrombin time | blood | 39 | 213 | 39 | 0 | 174 | MCF2L; AL137002.1; F7; F10; EDEM2 | F5; MCF2L; AL137002.1; F7; F10; GSS; MYH7B; TRPC4AP; EDEM2 | 120 | 226 | 115 | 5 | 111 | 0 | 0 | 0 | 0 | 6 | 6 | 5 | 6 |
| Pat-TT | Thrombin time | blood | 0 | 0 | 0 | 0 | 0 | | | 122 | 6 | 0 | 122 | 6 | 0 | 0 | 0 | 0 | 0 | | 0 | 0 |
| pC02-V | Carbon dioxide [Partial pressure] in Venous blood | blood | 0 | 0 | 0 | 0 | 0 | | | 4 | 0 | 0 | 4 | 0 | 0 | 0 | 0 | 0 | 0 | | 0 | 0 |
| PCAct | Protein C [Units/volume] in Platelet poor plasma by Chromogenic method | blood | 0 | 114 | 0 | 0 | 114 | | GSS; MYH7B; TRPC4AP; EDEM2 | 0 | 142 | 0 | 0 | 142 | 0 | 0 | 0 | 0 | 1 | 1 | 0 | 0 |

| all_labs | full_name | group | male.significant.variants | female.significant.variants | both.significant.variants | male.only.significant.variants | female.only.significant.variants | male.annotated.genes.significant.variants | female.annotated.genes.significant.variants | male.suggestive.variants | female.suggestive.variants | both.suggestive.variants | male.only.suggestive.variants | female.only.suggestive.variants | female.significant.x.variants | male.significant.x.variants | female.suggestive.x.variants | male.suggestive.x.variants | n_loci | n_loci_noHLA | P1e6_both | P1e2_both |
|---|---|---|---|---|---|---|---|---|---|---|---|---|---|---|---|---|---|---|---|---|---|---|
| pCO2 | Carbon dioxide [Partial pressure] in Arterial blood | blood | 0 | 6 | 0 | 0 | 6 | | CDKAL1 | 9 | 13 | 0 | 9 | 13 | 0 | 0 | 0 | 1 | 1 | 1 | 0 | 0 |
| PCR | Protein/Creatinine [Mass ratio] in Urine | kidney | 0 | 0 | 0 | 0 | 0 | | | 0 | 0 | 0 | 0 | 0 | 0 | 0 | 0 | 0 | 0 | | 0 | 0 |
| PCV | Hematocrit [Volume Fraction] of Blood by Automated count | blood | 0 | 23 | 0 | 0 | 23 | | HBS1L; MYB; HK1; TMPRSS6 | 4 | 48 | 3 | 1 | 45 | 0 | 0 | 0 | 0 | 4 | 4 | 1 | 4 |
| PEFR_AFD | PEFR_AFD | lung | 0 | 0 | 0 | 0 | 0 | | | 0 | 2 | 0 | 0 | 2 | 0 | 0 | 0 | 0 | 0 | | 0 | 0 |
| PEFR_APP | PEFR_APP | lung | 0 | 0 | 0 | 0 | 0 | | | 0 | 6 | 0 | 0 | 6 | 0 | 0 | 0 | 0 | 0 | | 0 | 0 |
| PEFR_PPR | PEFR_PPR | lung | 0 | 0 | 0 | 0 | 0 | | | 3 | 8 | 0 | 3 | 8 | 0 | 0 | 0 | 0 | 0 | | 0 | 0 |
| PEFR_PRD | PEFR_PRD | lung | 0 | 0 | 0 | 0 | 0 | | | 0 | 3 | 0 | 0 | 3 | 0 | 0 | 0 | 0 | 0 | | 0 | 0 |
| PEFR_PRL | PEFR_PRL | lung | 0 | 0 | 0 | 0 | 0 | | | 0 | 2 | 0 | 0 | 2 | 0 | 0 | 0 | 0 | 0 | | 0 | 0 |
| PEFR_ZBD | PEFR_ZBD | lung | 0 | 0 | 0 | 0 | 0 | | | 1 | 5 | 0 | 1 | 5 | 0 | 0 | 0 | 0 | 0 | | 0 | 0 |
| PEFR_ZPC | PEFR_ZPD | lung | 0 | 0 | 0 | 0 | 0 | | | 0 | 1 | 0 | 0 | 1 | 0 | 0 | 0 | 0 | 0 | | 0 | 0 |
| pH-Ven | Venous pH | blood | 0 | 0 | 0 | 0 | 0 | | | 0 | 0 | 0 | 0 | 0 | 0 | 0 | 0 | 0 | 0 | | 0 | 0 |
| Phos | Phosphate | metabolic | 0 | 0 | 0 | 0 | 0 | | | 6 | 2 | 0 | 6 | 2 | 0 | 0 | 0 | 0 | 0 | | 0 | 0 |
| PIFR_AFD | PIFR_AFD | lung | 0 | 0 | 0 | 0 | 0 | | | 19 | 0 | 0 | 19 | 0 | 0 | 0 | 0 | 0 | 0 | | 0 | 0 |
| PIFR_APP_APP | PIFR_APP_APP | lung | 0 | 0 | 0 | 0 | 0 | | | 0 | 0 | 0 | 0 | 0 | 0 | 0 | 0 | 0 | 0 | | 0 | 0 |
| PIFR_PPR | PIFR_PPR | lung | 0 | 0 | 0 | 0 | 0 | | | 1 | 0 | 0 | 1 | 0 | 0 | 0 | 0 | 0 | 0 | | 0 | 0 |
| PIFR_PRD | PIFR_PRD | lung | 0 | 0 | 0 | 0 | 0 | | | 0 | 0 | 0 | 0 | 0 | 0 | 0 | 0 | 0 | 0 | | 0 | 0 |
| PIFR_ZBD | PIFR_ZBD | lung | 0 | 0 | 0 | 0 | 0 | | | 1 | 1 | 0 | 1 | 1 | 0 | 0 | 0 | 0 | 0 | | 0 | 0 |
| PIFR_ZPC | PIFR_ZPC | lung | 0 | 0 | 0 | 0 | 0 | | | 0 | 1 | 0 | 0 | 1 | 0 | 0 | 0 | 0 | 0 | | 0 | 0 |

| all_labs | full_name | group | male.significant.variants | female.significant.variants | both.significant.variants | male.only.significant.variants | female.only.significant.variants | male.annotated.genes.significant.variants | female.annotated.genes.significant.variants | male.suggestive.variants | female.suggestive.variants | both.suggestive.variants | male.only.suggestive.variants | female.only.suggestive.variants | female.significant.x.variants | male.significant.x.variants | female.suggestive.x.variants | male.suggestive.x.variants | n_loci | n_loci_noHLA | P1e6_both | P1e2_both |
|---|---|---|---|---|---|---|---|---|---|---|---|---|---|---|---|---|---|---|---|---|---|---|
| Plt-Ct | Platelet Count | blood | 121 | 400 | 101 | 20 | 299 | ARHGEF3; BAK1; GGNBP1; HBS1L; MYB; AK3; RP11-307I14.4; PNPLA3 | ARHGEF3; EIF2B5; RN7SL26P; BAK1; GGNBP1; LINC00336; ITPR3; HBS1L; MYB; ZFPM2; AK3; RP11-307I14.4; RP11-307I14.2; RCL1; CDKN2B-AS1; NRBF2; JMJD1C; JMJD1C-AS1; RP11-351O1.3; REEP3; BET1L; RIC8A; SIRT3; PSMD13; NLRP6; USP30; ATXN2; BRAP; NAA25; TRAFD1; HECTD4; PTPN11; WDR66; ABCC4; AC069368.3; PTPRH; SIRPA; RP4-684O24.5; PNPLA3; SAMM50 | 147 | 644 | 132 | 15 | 512 | 0 | 0 | 0 | 0 | 37 | 37 | 12 | 35 |
| pO2_FI | Oxygen [Partial pressure] in Arterial blood | blood | 0 | 0 | 0 | 0 | 0 | | | 0 | 0 | 0 | 0 | 0 | 0 | 0 | 0 | 0 | 0 | | 0 | 0 |
| pO2-V | pO2 venous | blood | 0 | 0 | 0 | 0 | 0 | | | 0 | 0 | 0 | 0 | 0 | 0 | 0 | 0 | 0 | 0 | | 0 | 0 |
| pO2 | pO2 arterial | blood | 0 | 0 | 0 | 0 | 0 | | | 1 | 0 | 0 | 1 | 0 | 0 | 0 | 0 | 0 | 0 | | 0 | 0 |
| POCA1C | Hemoglobin A1c (Glycated) | blood | 0 | 0 | 0 | 0 | 0 | | | 1 | 0 | 0 | 1 | 0 | 0 | 0 | 0 | 0 | 0 | | 0 | 0 |
| PR INTERVAL | PR Interval mean | cardiovascular | 20 | 2 | 0 | 20 | 2 | SCN10A | SCN5A | 30 | 18 | 3 | 27 | 15 | 0 | 0 | 0 | 0 | 3 | 3 | 1 | 3 |
| PR_INTERVAL | PR Interval mean | cardiovascular | 181 | 340 | 154 | 27 | 186 | C1orf86; SCN5A; SCN10A; ARHGAP24; CAV1; SH3PXD2A; TBX5 | MEIS1; XYLB; EXOG; SCN5A; SCN10A; ARHGAP24; PRRC2A; C6orf47; LY6G5C; ABHD16A; CAV2; CAV1; AC006159.4; AC006159.3; KNOP1P1; RP11-615I16.1; TBX5; TBX3 | 224 | 428 | 207 | 17 | 221 | 0 | 0 | 0 | 0 | 36 | 35 | 21 | 34 |
| Pr24Hr | Protein 24 hour | urinary | 0 | 0 | 0 | 0 | 0 | | | 3 | 5 | 0 | 3 | 5 | 0 | 0 | 0 | 0 | 0 | | 0 | 0 |
| Pralbu | Prealbumin [Mass/volume] in Serum or Plasma | metabolic | 0 | 0 | 0 | 0 | 0 | | | 4 | 29 | 0 | 4 | 29 | 0 | 0 | 0 | 0 | 0 | | 0 | 0 |
| PRODUCT BLOOD TYPE | Product blood type | blood | 0 | 0 | 0 | 0 | 0 | | | 0 | 0 | 0 | 0 | 0 | 0 | 0 | 0 | 0 | 0 | | 0 | 0 |
| Prog-R | Progesterone rapid | ob / gyn | | 0 | | | | | | | 0 | | | | 0 | | 0 | | 0 | | 0 | 0 |
| Prolac | Prolactin [Mass/volume] in Serum or Plasma | endocrine | 0 | 0 | 0 | 0 | 0 | | | 1 | 1 | 0 | 1 | 1 | 0 | 0 | 1 | 0 | 0 | | 0 | 0 |

| all_labels | full_name | group | male.significant.variants | female.significant.variants | both.significant.variants | male.only.significant.variants | female.only.significant.variants | male.annotated.genes.significant.variants | female.annotated.genes.significant.variants | male.suggestive.variants | female.suggestive.variants | both.suggestive.variants | male.only.suggestive.variants | female.only.suggestive.variants | female.significant.x.variants | male.significant.x.variants | female.suggestive.x.variants | male.suggestive.x.variants | n_loci | n_loci_noHLA | P1e6_both | P1e2_both |
|---|---|---|---|---|---|---|---|---|---|---|---|---|---|---|---|---|---|---|---|---|---|---|
| Promyl | Promyelocytes/100 leukocytes in Blood by Manual count | blood | 0 | 0 | 0 | 0 | 0 | | | 0 | 1 | 0 | 0 | 1 | 0 | 0 | 0 | 0 | 0 | | 0 | 0 |
| PSA | PSA serum/plasma | cancer | 10 | | | | | AC011523.2; KLK3; KLK2 | | 21 | | | | | | 0 | | 0 | 4 | 4 | 0 | 0 |
| PT-inr | International normalized ratio | blood | 28 | 170 | 24 | 4 | 146 | MCF2L; AL137002.1; F7; F10 | MCF2L; AL137002.1; F7; F10; GSS; MYH7B; TRPC4AP; EDEM2 | 51 | 189 | 38 | 13 | 151 | 0 | 0 | 0 | 0 | 4 | 4 | 4 | 4 |
| PTT-pt | aPTT in Platelet poor plasma | blood | 100 | 211 | 90 | 10 | 121 | FETUB; RP11-134F2.2; HRG; RP11-795A2.3; RP11-573D15.8; KNG1; NSD1; RAB24; MXD3; SLC34A1; F12; GRK6 | FETUB; RP11-134F2.2; HRG; RP11-795A2.3; RP11-573D15.8; KNG1; F11; NSD1; RAB24; MXD3; LMAN2; RGS14; SLC34A1; F12; GRK6; ABO | 113 | 265 | 103 | 10 | 162 | 0 | 0 | 0 | 0 | 21 | 21 | 13 | 21 |
| PTTCtr | aPTT in Platelet poor plasma by Coagulation assay | blood | 0 | 0 | 0 | 0 | 0 | | | 0 | 0 | 0 | 0 | 0 | 0 | 0 | 0 | 0 | 0 | | 0 | 0 |
| PULSE OXIMETRY | Oxygen saturation in Arterial blood by pulse oximetry | vitals | 0 | 0 | 0 | 0 | 0 | | | 0 | 1 | 0 | 0 | 1 | 0 | 0 | 0 | 0 | 0 | | 0 | 0 |
| Pulse | Heart rate | vitals | 58 | 111 | 58 | 0 | 53 | RNU2-8P; RP11-129H15.1; RP11-129H15.4; HMGB3P18 | RNU2-8P; RP11-129H15.1; RP11-129H15.4; HMGB3P18; MYH6; MYH7 | 71 | 158 | 64 | 7 | 94 | 0 | 0 | 0 | 0 | 4 | 4 | 2 | 3 |
| PULSE | Heart rate | vitals | 0 | 44 | 0 | 0 | 44 | | RNU2-8P; RP11-129H15.1; RP11-129H15.4; HMGB3P18; MYH6 | 1 | 84 | 0 | 1 | 84 | 0 | 0 | 0 | 0 | 2 | 2 | 0 | 1 |
| Q ONSET | Q onset | cardiovascular | 0 | 0 | 0 | 0 | 0 | | | 4 | 2 | 0 | 4 | 2 | 0 | 0 | 1 | 0 | 0 | | 0 | 0 |
| QMG-NL | Quantiferon Mitogen assay | immune / inflammatory | 0 | 0 | 0 | 0 | 0 | | | 4 | 3 | 0 | 4 | 3 | 0 | 0 | 0 | 0 | 0 | | 0 | 0 |
| QRS DURATION | QRS duration | cardiovascular | 0 | 22 | 0 | 0 | 22 | | RP11-57K17.1; CEP85L | 0 | 50 | 0 | 0 | 50 | 0 | 0 | 0 | 0 | 2 | 2 | 0 | 0 |
| QRS HORIZONTAL AXIS | QRS horizontal axis | cardiovascular | 0 | 0 | 0 | 0 | 0 | | | 1 | 0 | 0 | 1 | 0 | 0 | 0 | 0 | 0 | 0 | | 0 | 0 |
| QRS_DURATION | QRS duration | cardiovascular | 55 | 453 | 28 | 27 | 425 | EXOG; SCN5A; CDKN1A; VTI1A; SETBP1 | RP11-536I6.2; EXOG; SCN5A; SCN10A; RNU1-88P; LAP3P2; CDKN1A; SLC35F1; RP11-57K17.1; CEP85L; ATP5B; PTGES3; RP4-601P9.1 | 126 | 585 | 47 | 79 | 538 | 0 | 0 | 0 | 0 | 31 | 31 | 6 | 26 |
| QT INTERVAL | QT interval | cardiovascular | 0 | 3 | 0 | 0 | 3 | | RP11-227F8.2; NOS1AP | 3 | 34 | 0 | 3 | 34 | 0 | 0 | 0 | 0 | 1 | 1 | 0 | 1 |

| all_labs | full_name | group | male.significant.variants | female.significant.variants | both.significant.variants | male.only.significant.variants | female.only.significant.variants | male.annotated.genes.significant.variants | female.annotated.genes.significant.variants | male.suggestive.variants | female.suggestive.variants | both.suggestive.variants | male.only.suggestive.variants | female.only.suggestive.variants | female.significant.x.variants | male.significant.x.variants | female.suggestive.x.variants | male.suggestive.x.variants | n_loci | n_loci_noHLA | P1e6_both | P1e2_both |
|---|---|---|---|---|---|---|---|---|---|---|---|---|---|---|---|---|---|---|---|---|---|---|
| QT_INTERVAL_C | QT interval corrected | cardiovascular | 216 | 385 | 215 | 1 | 170 | RP11-227F8.2; NOS1AP; KCNQ1; SETD6; CNOT1 | OLFML2B; RP11-227F8.2; NOS1AP; RP5-1018K9.1; ATP1B1; SLC8A1; SLC35F1; RP11-57K17.1; CEP85L; KCNH2; KCNQ1; SETD6; CNOT1 | 304 | 521 | 270 | 34 | 251 | 0 | 0 | 0 | 0 | 18 | 18 | 10 | 17 |
| QT_INTERVAL | QT interval | cardiovascular | 22 | 182 | 6 | 16 | 176 | RP11-227F8.2; NOS1AP; CHRNA5; CHRNA3; CHRNB4 | RP11-227F8.2; NOS1AP; SLC35F1; RP11-57K17.1; CEP85L | 90 | 306 | 37 | 53 | 269 | 0 | 0 | 0 | 0 | 10 | 10 | 6 | 9 |
| QT3 | Triiodothyronine (T3) [ Mass/volume] in Serum or Plasma | endocrine | 16 | 0 | 0 | 16 | 0 | SERPINA7; RP11-560L11.1 | | 47 | 4 | 0 | 47 | 4 | 0 | 16 | 0 | 47 | 1 | 1 | 0 | 1 |
| QTB-NL | Quantiferon TB assay | immune / inflammatory | 0 | 2 | 0 | 0 | 2 | | RP11-486E2.1 | 2 | 3 | 0 | 2 | 3 | 0 | 0 | 0 | 0 | 1 | 1 | 0 | 0 |
| QTCB | Q-T interval corrected based on Bazett formula | cardiovascular | 6 | 48 | 6 | 0 | 42 | RP11-227F8.2; NOS1AP | RP11-227F8.2; NOS1AP | 26 | 112 | 25 | 1 | 87 | 0 | 0 | 0 | 0 | 2 | 2 | 1 | 2 |
| QTCF | Q-T interval corrected based on Fridericia formula | cardiovascular | 23 | 107 | 23 | 0 | 84 | RP11-227F8.2; NOS1AP | RP11-227F8.2; NOS1AP | 52 | 149 | 31 | 21 | 118 | 0 | 0 | 0 | 0 | 2 | 2 | 1 | 2 |
| QtfNIL | Gamma interferon background [Units/volume] in Blood by Immunoassay | immune / inflammatory | 0 | 0 | 0 | 0 | 0 | | | 0 | 0 | 0 | 0 | 0 | 0 | 0 | 0 | 0 | 0 | | 0 | 0 |
| Quant_TB1(-)NIL | Quant TB1(-) NIL | immune / inflammatory | 0 | 0 | 0 | 0 | 0 | | | 10 | 0 | 0 | 10 | 0 | 0 | 0 | 0 | 0 | 0 | | 0 | 0 |
| Quant_TB2(-)NIL | Quant TB2(-) NIL | immune / inflammatory | 0 | 0 | 0 | 0 | 0 | | | 0 | 0 | 0 | 0 | 0 | 0 | 0 | 0 | 0 | 0 | | 0 | 0 |

| all_labs | full_name | group | male.significant.variants | female.significant.variants | both.significant.variants | male.only.significant.variants | female.only.significant.variants | male.annotated.genes.significant.variants | female.annotated.genes.significant.variants | male.suggestive.variants | female.suggestive.variants | both.suggestive.variants | male.only.suggestive.variants | female.only.suggestive.variants | female.significant.x.variants | male.significant.x.variants | female.suggestive.x.variants | male.suggestive.x.variants | n_loci | n_loci_noHLA | P1e6_both | P1e2_both |
|---|---|---|---|---|---|---|---|---|---|---|---|---|---|---|---|---|---|---|---|---|---|---|
| R AN AIRWAY RESPIRATION RATE | Respiratory rate by airway flow measurement | vitals | 0 | 0 | 0 | 0 | 0 | | | 18 | 0 | 0 | 18 | 0 | 0 | 0 | 0 | 0 | 0 | | 0 | 0 |
| R AN ANESTHESIA BLOOD PRESSURE | Diastolic blood pressure by Noninvasive | vitals | 0 | 0 | 0 | 0 | 0 | | | 21 | 1 | 0 | 21 | 1 | 0 | 0 | 0 | 0 | 0 | | 0 | 0 |
| R AN ANESTHESIA TEMPERATURE | Body temperature | vitals | 0 | 0 | 0 | 0 | 0 | | | 0 | 1 | 0 | 0 | 1 | 0 | 0 | 0 | 0 | 0 | | 0 | 0 |
| R AN CORE BODY TEMPERATURE | Body temperature - Core | vitals | 0 | 0 | 0 | 0 | 0 | | | 3 | 5 | 0 | 3 | 5 | 0 | 0 | 0 | 0 | 0 | | 0 | 0 |
| R AN ESOPHAGEAL TEMPERATURE | Temperature - Distal airway | vitals | 0 | 0 | 0 | 0 | 0 | | | 10 | 1 | 0 | 10 | 1 | 0 | 0 | 0 | 0 | 0 | | 0 | 0 |
| R AN HEART RATE MONITOR | Heart rate | vitals | 0 | 0 | 0 | 0 | 0 | | | 0 | 1 | 0 | 0 | 1 | 0 | 0 | 0 | 0 | 0 | | 0 | 0 |
| R AN MEAN ARTERIAL BLOOD PRESSURE | Invasive Mean blood pressure | vitals | 0 | 0 | 0 | 0 | 0 | | | 0 | 2 | 0 | 0 | 2 | 0 | 0 | 0 | 0 | 0 | | 0 | 0 |
| R AN NASOPHARYNGEAL TEMPERATURE | Temperature - Distal airway | vitals | 0 | 0 | 0 | 0 | 0 | | | 3 | 4 | 0 | 3 | 4 | 0 | 0 | 0 | 0 | 0 | | 0 | 0 |
| R AN NIBP MEAN | Mean blood pressure by Noninvasive | vitals | 0 | 0 | 0 | 0 | 0 | | | 1 | 4 | 0 | 1 | 4 | 0 | 0 | 0 | 0 | 0 | | 0 | 0 |

| all_labs | full_name | group | male.significant.variants | female.significant.variants | both.significant.variants | male.only.significant.variants | female.only.significant.variants | male.annotated.genes.significant.variants | female.annotated.genes.significant.variants | male.suggestive.variants | female.suggestive.variants | both.suggestive.variants | male.only.suggestive.variants | female.only.suggestive.variants | female.significant.x.variants | male.significant.x.variants | female.suggestive.x.variants | male.suggestive.x.variants | n_loci | n_loci_noHLA | P1e6_both | P1e2_both |
|---|---|---|---|---|---|---|---|---|---|---|---|---|---|---|---|---|---|---|---|---|---|---|
| R AN PULSE SPO2 | Heart rate by Pulse oximetry | vitals | 0 | 0 | 0 | 0 | 0 | | | 0 | 0 | 0 | 0 | 0 | 0 | 0 | 0 | 0 | 0 | | 0 | 0 |
| R AN SKIN TEMPERATURE | Temperature of Skin | vitals | 0 | 0 | 0 | 0 | 0 | | | 0 | 40 | 0 | 0 | 40 | 0 | 0 | 0 | 0 | 0 | | 0 | 0 |
| R APACHE TEMPERATURE | Body temperature | vitals | 15 | 0 | 0 | 15 | 0 | KLF7; RP11-560G2.1 | | 19 | 0 | 0 | 19 | 0 | 0 | 0 | 0 | 0 | 2 | 2 | 0 | 0 |
| R ARTERIAL LINE BLOOD PRESSURE | Invasive Systolic blood pressure | vitals | 0 | 0 | 0 | 0 | 0 | | | 0 | 0 | 0 | 0 | 0 | 0 | 0 | 0 | 0 | 0 | | 0 | 0 |
| R ED PRE-ARRIVAL BP | Systolic blood pressure by Noninvasive | vitals | 0 | 0 | 0 | 0 | 0 | | | 6 | 0 | 0 | 6 | 0 | 0 | 0 | 0 | 0 | 0 | | 0 | 0 |
| R ED PRE-ARRIVAL EMS O2 SAT | Oxygen saturation in Arterial blood by Pulse oximetry | vitals | 0 | 0 | 0 | 0 | 0 | | | 0 | 1 | 0 | 0 | 1 | 0 | 0 | 0 | 0 | 0 | | 0 | 0 |
| R ED PRE-ARRIVAL PULSE (HEART RATE) | Heart rate | vitals | 0 | 0 | 0 | 0 | 0 | | | 1 | 3 | 0 | 1 | 3 | 0 | 0 | 0 | 0 | 0 | | 0 | 0 |
| R ED PRE-ARRIVAL RESP RATE | Respiratory rate | vitals | 0 | 4 | 0 | 0 | 4 | | GPC5 | 2 | 6 | 0 | 2 | 6 | 0 | 0 | 0 | 0 | 1 | 1 | 0 | 0 |
| R FN WEIGHT USED FOR EQUATIONS CALCULATIONS | Body weight | vitals | 0 | 0 | 0 | 0 | 0 | | | 0 | 0 | 0 | 0 | 0 | 0 | 0 | 0 | 0 | 0 | | 0 | 0 |
| R HEART RATE MONITOR | Heart rate | vitals | 0 | 0 | 0 | 0 | 0 | | | 4 | 1 | 0 | 4 | 1 | 0 | 0 | 0 | 0 | 0 | | 0 | 0 |

| all_labs | full_name | group | male.significant.variants | female.significant.variants | both.significant.variants | male.only.significant.variants | female.only.significant.variants | male.annotated.genes.significant.variants | female.annotated.genes.significant.variants | male.suggestive.variants | female.suggestive.variants | both.suggestive.variants | male.only.suggestive.variants | female.only.suggestive.variants | female.significant.x.variants | male.significant.x.variants | female.suggestive.x.variants | male.suggestive.x.variants | n_loci | n_loci_noHLA | P1e6_both | P1e2_both |
|---|---|---|---|---|---|---|---|---|---|---|---|---|---|---|---|---|---|---|---|---|---|---|
| R IP IDEAL BODY WEIGHT | Ideal body weight | vitals | 62 | 553 | 19 | 43 | 534 | EFEMP1; ZBTB38; IGF2BP3; CABLES1; GDF5 | MTMR11; DNM3; LTBP1; ZBTB38; FAM184B; DCAF16; NCAPG; LCORL; HHIP-AS1; HHIP; RP11-486E2.1; KDM2A; SSH3; CCDC91; HMGA2; DLEU1; ADAMTSL3; ACAN; ATAD5; TEFM; ADAP2; CABLES1; UQCC1; GDF5OS; GDF5 | 272 | 1201 | 149 | 123 | 1052 | 0 | 0 | 61 | 0 | 22 | 22 | 5 | 17 |
| R RT ( FEMALE ADULT) IDEAL BODY WEIGHT | Ideal body weight — female | vitals | 62 | 553 | 19 | 43 | 534 | EFEMP1; ZBTB38; IGF2BP3; CABLES1; GDF5 | MTMR11; DNM3; LTBP1; ZBTB38; FAM184B; DCAF16; NCAPG; LCORL; HHIP-AS1; HHIP; RP11-486E2.1; KDM2A; SSH3; CCDC91; HMGA2; DLEU1; ADAMTSL3; ACAN; ATAD5; TEFM; ADAP2; CABLES1; UQCC1; GDF5OS; GDF5 | 272 | 1208 | 149 | 123 | 1059 | 0 | 0 | 61 | 0 | 23 | 23 | 5 | 17 |
| R RT (MALE ADULT) IDEAL BODY WEIGHT | Ideal body weight — male | vitals | 62 | 553 | 19 | 43 | 534 | EFEMP1; ZBTB38; IGF2BP3; CABLES1; GDF5 | MTMR11; DNM3; LTBP1; ZBTB38; FAM184B; DCAF16; NCAPG; LCORL; HHIP-AS1; HHIP; RP11-486E2.1; KDM2A; SSH3; CCDC91; HMGA2; DLEU1; ADAMTSL3; ACAN; ATAD5; TEFM; ADAP2; CABLES1; UQCC1; GDF5OS; GDF5 | 272 | 1208 | 149 | 123 | 1059 | 0 | 0 | 61 | 0 | 23 | 23 | 5 | 17 |
| R VUMC AN RESPIRATION RATE | Respiratory rate | vitals | 0 | 0 | 0 | 0 | 0 | | | 0 | 2 | 0 | 0 | 2 | 0 | 0 | 2 | 0 | 0 | | 0 | 0 |
| R VUMC OXYGEN FLOW DELIVERED | Delivered oxygen flow rate | vitals | 0 | 0 | 0 | 0 | 0 | | | 0 | 5 | 0 | 0 | 5 | 0 | 0 | 0 | 0 | 0 | | 0 | 0 |
| R_F | Rheumatoid factor [ Units/volume] in Serum or Plasma | immune / inflammatory | 14 | 0 | 0 | 14 | 0 | AC004875.1 | | 29 | 3 | 0 | 29 | 3 | 0 | 0 | 0 | 0 | 1 | 1 | 0 | 0 |
| RATE_C | Heart rate | vitals | 32 | 31 | 31 | 1 | 0 | HMGB3P18; DLG2 | HMGB3P18 | 97 | 35 | 31 | 66 | 4 | 0 | 0 | 0 | 30 | 2 | 2 | 1 | 2 |
| RBC | Erythrocytes [#/volume] in Blood by Automated count | blood | 119 | 498 | 89 | 30 | 409 | AC006552.1; HBS1L; MYB; MOSPD3; TFR2; GNB2; GIGYF1; POP7; EPO; ZAN; KLF1; GCDH; SYCE2; MIR5695; FARSA; CALR | RP11-231C18.2; RP11-545H22.1; AC006552.1; HIST1H4C; HIST1H1T; C6orf183; RP11-425D10.1; PTCHD3P3; CCDC162P; RP3-352A20.1; ALDH8A1; HBS1L; MYB; LRCH4; FBXO24; PCOLCE-AS1; MOSPD3; TFR2; GNB2; GIGYF1; POP7; EPO; ZAN | 144 | 602 | 107 | 37 | 495 | 0 | 0 | 0 | 0 | 21 | 21 | 8 | 20 |
| RBCFol | Folate [ Mass/volume] in Red Blood Cells | metabolic | 0 | 0 | 0 | 0 | 0 | | | 1 | 0 | 0 | 1 | 0 | 0 | 0 | 0 | 0 | 0 | | 0 | 0 |
| RDW-SD | RDW SD | blood | 0 | 0 | 0 | 0 | 0 | | | 0 | 2 | 0 | 0 | 2 | 0 | 0 | 0 | 0 | 0 | | 0 | 0 |

| all_labs | full_name | group | male.significant.variants | female.significant.variants | both.significant.variants | male.only.significant.variants | female.only.significant.variants | male.annotated.genes.significant.variants | female.annotated.genes.significant.variants | male.suggestive.variants | female.suggestive.variants | both.suggestive.variants | male.only.suggestive.variants | female.only.suggestive.variants | female.significant.x.variants | male.significant.x.variants | female.suggestive.x.variants | male.suggestive.x.variants | n_loci | n_loci_noHLA | P1e6_both | P1e2_both |
|---|---|---|---|---|---|---|---|---|---|---|---|---|---|---|---|---|---|---|---|---|---|---|
| RDW | Erythrocyte distribution width [Ratio] by Automated count | blood | 203 | 693 | 163 | 40 | 530 | SLC12A7; CTC-228N24.3; SLC12A2; FBN2; LRRC16A; SCGN; HIST1H2AA; SLC17A3; SLC17A2; TRIM38; U91328.21; HIST1H4B; HIST1H2AB; HIST1H2BB; HFE; HIST1H4C; HIST1H1T; HIST1H2BC; HIST1H2AC; TRAF4; FAM222B; ERAL1; MIR451B; TMPRSS6 | ATP2B4; SLC12A7; CTC-228N24.3; SLC12A2; FBN2; LRRC16A; SCGN; HIST1H2AA; SLC17A1; SLC17A3; HIST1H2APS2; SLC17A2; TRIM38; U91328.21; U91328.19; U91328.22; HIST1H3A; HIST1H4A; HIST1H4B; HIST1H3B; HIST1H2AB; HIST1H2BB; U91328.2; HIST1H3C; HIST1H1C; HFE; HIST1H4C; HIST1H1T; HIST1H2BC; HIST1H2AC; VN1R13P; MIR3143; PGBD1; GPX6; HCG4P7; PGC; RP11-298J23.8; FRS3; USP49; MED20; BYSL; CCND3; HBS1L; SOX6; ACSBG1; RP11-762H8.3; DNAJA4; WDR61; FTO; PIEZO1; TRAF4; FAM222B; ERAL1; MIR451B; C18orf25; TST; MPST; KCTD17; TMPRSS6 | 303 | 937 | 247 | 56 | 690 | 0 | 0 | 0 | 0 | 45 | 44 | 19 | 41 |
| RDWSD | Erythrocyte distribution width [Entitic volume] by Automated count | blood | 82 | 257 | 67 | 15 | 190 | ATP2B4; DPP10; CTC-228N24.3; SLC12A2; FBN2; HBS1L; FEN1; FADS2; ACSBG1; RP11-762H8.3 | ATP2B4; CTC-228N24.3; SLC12A2; FBN2; HBS1L; MYB; AVL9; LCN1P1; ABO; LCN1P2; HK1; SOX6; MYRF; TMEM258; FEN1; FADS2; ACSBG1; RP11-762H8.3; DNAJA4; WDR61; CTU2; PIEZO1; SMG9; KCNN4 | 153 | 441 | 107 | 46 | 334 | 0 | 0 | 0 | 0 | 22 | 22 | 7 | 18 |
| RenAct | Renin [Enzymatic activity/volume] in Plasma | endocrine | 0 | 0 | 0 | 0 | 0 | | | 10 | 0 | 0 | 10 | 0 | 0 | 0 | 0 | 0 | 0 | | 0 | 0 |
| RESPIRATIONS | Respiratory rate | vitals | 0 | 0 | 0 | 0 | 0 | | | 0 | 12 | 0 | 0 | 12 | 0 | 0 | 0 | 0 | 0 | | 0 | 0 |
| RespRt | Respiratory rate | vitals | 0 | 0 | 0 | 0 | 0 | | | 1 | 1 | 0 | 1 | 1 | 0 | 0 | 0 | 0 | 0 | | 0 | 0 |
| RetAbs | Reticulocytes absolute | blood | 0 | 0 | 0 | 0 | 0 | | | 0 | 6 | 0 | 0 | 6 | 0 | 0 | 0 | 0 | 0 | | 0 | 0 |
| RETHE | Hemoglobin [Entitic mass] in Reticulocytes | blood | 0 | 0 | 0 | 0 | 0 | | | 3 | 0 | 0 | 3 | 0 | 0 | 0 | 0 | 0 | 0 | | 0 | 0 |
| RetiCt | Reticulocytes/100 erythrocytes in Blood by Automated count | blood | 0 | 0 | 0 | 0 | 0 | | | 0 | 0 | 0 | 0 | 0 | 0 | 0 | 0 | 0 | 0 | | 0 | 0 |

| all_labs | full_name | group | male.significant.variants | female.significant.variants | both.significant.variants | male.only.significant.variants | female.only.significant.variants | male.annotated.genes.significant.variants | female.annotated.genes.significant.variants | male.suggestive.variants | female.suggestive.variants | both.suggestive.variants | male.only.suggestive.variants | female.only.suggestive.variants | female.significant.x.variants | male.significant.x.variants | female.suggestive.x.variants | male.suggestive.x.variants | n_loci | n_loci_noHLA | P1e6_both | P1e2_both |
|---|---|---|---|---|---|---|---|---|---|---|---|---|---|---|---|---|---|---|---|---|---|---|
| RetPal | Retinyl palmitate [Mass/volume] in Serum or Plasma | metabolic | 0 | 0 | 0 | 0 | 0 | | | 0 | 10 | 0 | 0 | 10 | 0 | 0 | 0 | 0 | 0 | | 0 | 0 |
| RR INTERVAL | RR interval | cardiovascular | 0 | 2 | 0 | 0 | 2 | | MTUS2 | 28 | 4 | 0 | 28 | 4 | 0 | 0 | 0 | 0 | 1 | 1 | 0 | 0 |
| RV_PPR | RV PPR | lung | 0 | 0 | 0 | 0 | 0 | | | 1 | 2 | 0 | 1 | 2 | 0 | 0 | 0 | 0 | 0 | | 0 | 0 |
| RV_PRD | RV PRD | lung | 0 | 0 | 0 | 0 | 0 | | | 0 | 3 | 0 | 0 | 3 | 0 | 0 | 0 | 0 | 0 | | 0 | 0 |
| RV_PRL | RV PRL | lung | 0 | 0 | 0 | 0 | 0 | | | 3 | 4 | 0 | 3 | 4 | 0 | 0 | 0 | 0 | 0 | | 0 | 0 |
| RV_PRU | RV PRU | lung | 0 | 0 | 0 | 0 | 0 | | | 0 | 17 | 0 | 0 | 17 | 0 | 0 | 0 | 0 | 0 | | 0 | 0 |
| RV_ZBD | RV ZBD | lung | 0 | 0 | 0 | 0 | 0 | | | 14 | 6 | 0 | 14 | 6 | 0 | 0 | 0 | 0 | 0 | | 0 | 0 |
| RVOTLC_PPR | RVOTLC PPR | lung | 0 | 0 | 0 | 0 | 0 | | | 0 | 0 | 0 | 0 | 0 | 0 | 0 | 0 | 0 | 0 | | 0 | 0 |
| RVOTLC_PRD | RVOTLC PRD | lung | 0 | 0 | 0 | 0 | 0 | | | 1 | 0 | 0 | 1 | 0 | 0 | 0 | 0 | 0 | 0 | | 0 | 0 |
| RVOTLC_PRL | RVOTLC PRL | lung | 0 | 0 | 0 | 0 | 0 | | | 0 | 0 | 0 | 0 | 0 | 0 | 0 | 0 | 0 | 0 | | 0 | 0 |
| RVOTLC_PRU | RVOTLC PRU | lung | 0 | 0 | 0 | 0 | 0 | | | 1 | 1 | 0 | 1 | 1 | 0 | 0 | 0 | 0 | 0 | | 0 | 0 |
| RVOTLC_ZBD | RVOTLC ZBD | lung | 0 | 0 | 0 | 0 | 0 | | | 0 | 0 | 0 | 0 | 0 | 0 | 0 | 0 | 0 | 0 | | 0 | 0 |
| RVVT | dRVVT actual/normal ( normalized LA screen) | immune / inflammatory | 0 | 0 | 0 | 0 | 0 | | | 0 | 1 | 0 | 0 | 1 | 0 | 0 | 0 | 0 | 0 | | 0 | 0 |
| S-T HORIZONTAL AXIS | ST horizontal axis | cardiovascular | 0 | 0 | 0 | 0 | 0 | | | 1 | 1 | 0 | 1 | 1 | 0 | 0 | 0 | 0 | 0 | | 0 | 0 |
| SclAb | SCL-70 extractable nuclear Ab [Units/volume] in Serum by Immunoassay | immune / inflammatory | 0 | 0 | 0 | 0 | 0 | | | 0 | 0 | 0 | 0 | 0 | 0 | 0 | 0 | 0 | 0 | | 0 | 0 |

| all_labs | full_name | group | male.significant.variants | female.significant.variants | both.significant.variants | male.only.significant.variants | female.only.significant.variants | male.annotated.genes.significant.variants | female.annotated.genes.significant.variants | male.suggestive.variants | female.suggestive.variants | both.suggestive.variants | male.only.suggestive.variants | female.only.suggestive.variants | female.significant.x.variants | male.significant.x.variants | female.suggestive.x.variants | male.suggestive.x.variants | n_loci | n_loci_noHLA | P1e6_both | P1e2_both |
|---|---|---|---|---|---|---|---|---|---|---|---|---|---|---|---|---|---|---|---|---|---|---|
| SedRat | Erythrocyte sedimentation rate by Westergren method | blood | 65 | 192 | 65 | 0 | 127 | CR1 | C1orf63; TMEM50A; RHCE; TMEM57; LDLRAP1; CR1; CR1L | 80 | 212 | 68 | 12 | 144 | 0 | 0 | 0 | 0 | 4 | 4 | 3 | 4 |
| SerFol | Folate [Mass/volume] in Serum or Plasma | metabolic | 0 | 0 | 0 | 0 | 0 | | | 0 | 0 | 0 | 0 | 0 | 0 | 0 | 0 | 0 | 0 | | 0 | 0 |
| SGBF | Specific gravity of Body fluid | blood | 0 | 0 | 0 | 0 | 0 | | | 1 | 0 | 0 | 1 | 0 | 0 | 0 | 0 | 0 | 0 | | 0 | 0 |
| all_labs | full_name | group | male.significant.variants | female.significant.variants | both.significant.variants | male.only.significant.variants | female.only.significant.variants | male.annotated.genes.significant.variants | female.annotated.genes.significant.variants | male.suggestive.variants | female.suggestive.variants | both.suggestive.variants | male.only.suggestive.variants | female.only.suggestive.variants | female.significant.x.variants | male.significant.x.variants | female.suggestive.x.variants | male.suggestive.x.variants | n_loci | n_loci_noHLA | P1e6_both | P1e2_both |
| SGOT | SGOT | liver | 74 | 67 | 64 | 10 | 3 | PNPLA3; SAMM50 | SLC39A12; CUX2; PNPLA3; SAMM50 | 83 | 103 | 77 | 6 | 26 | 0 | 0 | 0 | 0 | 5 | 5 | 3 | 4 |
| SGPT | SGPT | liver | 2 | 8 | 2 | 0 | 6 | PNPLA3 | PNPLA3 | 37 | 63 | 37 | 0 | 26 | 0 | 0 | 0 | 0 | 1 | 1 | 1 | 1 |
| SSAIgG | Sjogrens syndrome-A extractable nuclear IgG Ab [Units/volume] in Serum by Immunoassay | immune / inflammatory | 0 | 22 | 0 | 0 | 22 | | HLA-DRB1; HLA-DQA1 | 0 | 32 | 0 | 0 | 32 | 0 | 0 | 0 | 0 | 1 | | 0 | 0 |
| SSBIgG | Sjogrens syndrome-B extractable nuclear IgG Ab [Units/volume] in Serum by Immunoassay | immune / inflammatory | 0 | 0 | 0 | 0 | 0 | | | 3 | 0 | 0 | 3 | 0 | 0 | 0 | 0 | 0 | 0 | | 0 | 0 |
| ST_SEGMENT | ST segment axis | cardiovascular | 0 | 0 | 0 | 0 | 0 | | | 5 | 0 | 0 | 5 | 0 | 0 | 0 | 0 | 0 | 0 | | 0 | 0 |
| SU-CA | Calcium urine | urinary | 0 | 0 | 0 | 0 | 0 | | | 0 | 1 | 0 | 0 | 1 | 0 | 0 | 0 | 0 | 0 | | 0 | 0 |
| SU-Cre | Creatinine urine | urinary | 0 | 0 | 0 | 0 | 0 | | | 0 | 1 | 0 | 0 | 1 | 0 | 0 | 0 | 0 | 0 | | 0 | 0 |

| all_labs | full_name | group | male.significant.variants | female.significant.variants | both.significant.variants | male.only.significant.variants | female.only.significant.variants | male.annotated.genes.significant.variants | female.annotated.genes.significant.variants | male.suggestive.variants | female.suggestive.variants | both.suggestive.variants | male.only.suggestive.variants | female.only.suggestive.variants | female.significant.x.variants | male.significant.x.variants | female.suggestive.x.variants | male.suggestive.x.variants | n_loci | n_loci_noHLA | P1e6_both | P1e2_both |
|---|---|---|---|---|---|---|---|---|---|---|---|---|---|---|---|---|---|---|---|---|---|---|
| SU-K | Potassium urine | urinary | 0 | 0 | 0 | 0 | 0 | | | 0 | 0 | 0 | 0 | 0 | 0 | 0 | 0 | 0 | 0 | | 0 | 0 |
| SU-Na | Sodium urine | urinary | 0 | 0 | 0 | 0 | 0 | | | 0 | 1 | 0 | 0 | 1 | 0 | 0 | 0 | 0 | 0 | | 0 | 0 |
| SU-Osm | Urine osmolality | urinary | 0 | 0 | 0 | 0 | 0 | | | 1 | 0 | 0 | 1 | 0 | 0 | 0 | 0 | 0 | 0 | | 0 | 0 |
| SU-Pro | Protein urine | urinary | 1 | 0 | 0 | 1 | 0 | RNU6-525P | | 1 | 4 | 0 | 1 | 4 | 0 | 0 | 0 | 0 | 1 | 1 | 0 | 0 |
| SU-Ure | Urea urine | urinary | 0 | 0 | 0 | 0 | 0 | | | 0 | 0 | 0 | 0 | 0 | 0 | 0 | 0 | 0 | 0 | | 0 | 0 |
| SUCrea | Creatinine [Mass/volume] in Urine | urinary | 0 | 0 | 0 | 0 | 0 | | | 1 | 0 | 0 | 1 | 0 | 0 | 0 | 0 | 0 | 0 | | 0 | 0 |
| SyMnCl | Mononuclear cells/100 leukocytes in Synovial fluid by Manual count | immune / inflammatory | 0 | 0 | 0 | 0 | 0 | | | 0 | 0 | 0 | 0 | 0 | 0 | 0 | 0 | 0 | 0 | | 0 | 0 |
| SYSTOLIC | Systolic blood pressure | vitals | 0 | 28 | 0 | 0 | 28 | | RP11-734K2.4; RP11-981P6.1; ATP2B1 | 1 | 33 | 0 | 1 | 33 | 0 | 0 | 0 | 0 | 2 | 2 | 0 | 1 |
| T HORIZONTAL AXIS | T horizontal axis | cardiovascular | 0 | 0 | 0 | 0 | 0 | | | 0 | 4 | 0 | 0 | 4 | 0 | 0 | 0 | 0 | 0 | | 0 | 0 |
| T WAVE AXIS | T wave axis | cardiovascular | 0 | 0 | 0 | 0 | 0 | | | 1 | 0 | 0 | 1 | 0 | 0 | 0 | 0 | 0 | 0 | | 0 | 0 |
| T_WAVE | T wave axis | cardiovascular | 0 | 0 | 0 | 0 | 0 | | | 1 | 1 | 0 | 1 | 1 | 0 | 0 | 0 | 0 | 0 | | 0 | 0 |
| T3FRE | Triiodothyronine (T3) Free [Mass/volume] in Serum or Plasma | endocrine | 0 | 0 | 0 | 0 | 0 | | | 0 | 3 | 0 | 0 | 3 | 0 | 0 | 0 | 0 | 0 | | 0 | 0 |
| TBil | Total Bilirubin serum/plasma | metabolic | 605 | 790 | 589 | 16 | 201 | ATG16L1; SAG; RP11-400N9.1; DGKD; USP40; UGT1A11P; UGT1A8; MROH2A; HJURP; MSL3P1; TRPM8; SLCO1C1; SLCO1B3; RP11-125O5.2; SLCO1B1; SLCO1A2; PNPLA3 | ATG16L1; SAG; RP11-400N9.1; DGKD; USP40; UGT1A11P; UGT1A8; MROH2A; HJURP; MSL3P1; TRPM8; SLCO1C1; SLCO1B3; RP11-125O5.2; SLCO1B1; SLCO1A2 | 701 | 937 | 652 | 49 | 285 | 0 | 0 | 0 | 0 | 48 | 48 | 35 | 47 |
| TC:HDL | TC/HDL ratio | metabolic | 1 | 9 | 0 | 1 | 9 | CETP | AC012181.1; CETP | 2 | 12 | 0 | 2 | 12 | 0 | 0 | 0 | 0 | 3 | 3 | 0 | 3 |
| Temper | Body temperature | vitals | 0 | 0 | 0 | 0 | 0 | | | 2 | 0 | 0 | 2 | 0 | 0 | 0 | 0 | 0 | 0 | | 0 | 0 |

| all_labs | full_name | group | male.significant.variants | female.significant.variants | both.significant.variants | male.only.significant.variants | female.only.significant.variants | male.annotated.genes.significant.variants | female.annotated.genes.significant.variants | male.suggestive.variants | female.suggestive.variants | both.suggestive.variants | male.only.suggestive.variants | female.only.suggestive.variants | female.significant.x.variants | male.significant.x.variants | female.suggestive.x.variants | male.suggestive.x.variants | n_loci | n_loci_noHLA | P1e6_both | P1e2_both |
|---|---|---|---|---|---|---|---|---|---|---|---|---|---|---|---|---|---|---|---|---|---|---|
| TEMPERATURE | Body temperature | vitals | 0 | 0 | 0 | 0 | 0 | | | 6 | 0 | 0 | 6 | 0 | 0 | 0 | 0 | 0 | 0 | | 0 | 0 |
| TERMINAL_40_MS | Terminal 40 ms | cardiovascular | 0 | 6 | 0 | 0 | 6 | | NFIA; RPL7P6 | 1 | 43 | 0 | 1 | 43 | 0 | 0 | 0 | 0 | 3 | 3 | 0 | 1 |
| TestFr | Testosterone Free [Mass/volume] in Serum or Plasma | endocrine | 0 | 0 | 0 | 0 | 0 | | | 4 | 0 | 0 | 4 | 0 | 0 | 0 | 0 | 0 | 0 | | 0 | 0 |
| Testos | Testosterone total | endocrine | 0 | 0 | 0 | 0 | 0 | | | 2 | 0 | 0 | 2 | 0 | 0 | 0 | 0 | 0 | 0 | | 0 | 0 |
| TGG | Total gamma globulin | immune / inflammatory | 0 | 0 | 0 | 0 | 0 | | | 4 | 0 | 0 | 4 | 0 | 0 | 0 | 0 | 0 | 0 | | 0 | 0 |
| Thiamn | Thiamine pyrophosphate [Moles/volume] in Blood | metabolic | 0 | 0 | 0 | 0 | 0 | | | 1 | 1 | 0 | 1 | 1 | 0 | 0 | 0 | 0 | 0 | | 0 | 0 |
| THM | Total hemoglobin | blood | 0 | 0 | 0 | 0 | 0 | | | 1 | 6 | 0 | 1 | 6 | 0 | 0 | 0 | 0 | 0 | | 0 | 0 |
| Thyrgb | Thyroglobulin [Mass/volume] in Serum or Plasma | endocrine | 0 | 0 | 0 | 0 | 0 | | | 0 | 0 | 0 | 0 | 0 | 0 | 0 | 0 | 0 | 0 | | 0 | 0 |
| TIBC | Total iron binding capacity | blood | 67 | 0 | 0 | 67 | 0 | TFP1; TF; SRPRB | | 90 | 0 | 0 | 90 | 0 | 0 | 0 | 0 | 1 | 4 | 4 | 0 | 1 |
| TIGIgG | Tissue transglutaminase IgG Ab [Units/volume] in Serum by Immunoassay | immune / inflammatory | 0 | 0 | 0 | 0 | 0 | | | 0 | 0 | 0 | 0 | 0 | 0 | 0 | 0 | 0 | 0 | | 0 | 0 |
| TLC_PPR | TLC PPR | lung | 0 | 0 | 0 | 0 | 0 | | | 0 | 0 | 0 | 0 | 0 | 0 | 0 | 0 | 0 | 0 | | 0 | 0 |
| TLC_PRD | TLC PRD | lung | 0 | 0 | 0 | 0 | 0 | | | 0 | 1 | 0 | 0 | 1 | 0 | 0 | 0 | 0 | 0 | | 0 | 0 |
| TLC_PRL | TLC PRL | lung | 0 | 0 | 0 | 0 | 0 | | | 0 | 1 | 0 | 0 | 1 | 0 | 0 | 0 | 0 | 0 | | 0 | 0 |

| all_labs | full_name | group | male.significant.variants | female.significant.variants | both.significant.variants | male.only.significant.variants | female.only.significant.variants | male.annotated.genes.significant.variants | female.annotated.genes.significant.variants | male.suggestive.variants | female.suggestive.variants | both.suggestive.variants | male.only.suggestive.variants | female.only.suggestive.variants | female.significant.x.variants | male.significant.x.variants | female.suggestive.x.variants | male.suggestive.x.variants | n_loci | n_loci_noHLA | P1e6_both | P1e2_both |
|---|---|---|---|---|---|---|---|---|---|---|---|---|---|---|---|---|---|---|---|---|---|---|
| TLC_PRU | TLC PRU | lung | 0 | 0 | 0 | 0 | 0 | | | 3 | 0 | 0 | 3 | 0 | 0 | 0 | 0 | 0 | 0 | | 0 | 0 |
| TLC_ZBD | TLC ZBD | lung | 0 | 0 | 0 | 0 | 0 | | | 0 | 6 | 0 | 0 | 6 | 0 | 0 | 3 | 0 | 0 | | 0 | 0 |
| TotVl | Volume in Urine collected for unspecified duration | toxicology / pharmacology | 0 | 0 | 0 | 0 | 0 | | | 0 | 4 | 0 | 0 | 4 | 0 | 0 | 0 | 0 | 0 | | 0 | 0 |
| TProSe | Protein serum/plasma | metabolic | 225 | 0 | 0 | 225 | 0 | HLA-F-AS1; TRIM26; HLA-N; MUC22; HCG22; HCG27; XXbac-BPG248L24.10; HLA-B; XXbac-BPG248L24.12; DHFRP2; RNU6-283P; AL671883.1; FGFR3P1; ZDHHC20P2; HLA-S; MICA; HCP5; MICB; XXbac-BPG16N22.5; RPL15P4; ATP6V1G2-DDX39B; TNF; NCR3; PRRC2A; BAG6; APOM; GPANK1; CSNK2B; ABHD16A; XXbac-BPG32J3.20; CLIC1; MSH5; VWA7; VARS; LSM2; HSPA1L; HSPA1A; HSPA1B; C6orf48; NEU1; SLC44A4; C2; CFB; STK19; TNXB; PRRT1; PPT2; PPT2-EGFL8; AGPAT1; RNF5; GPSM3; NOTCH4; HLA-DQB1 | | 414 | 12 | 7 | 407 | 5 | 0 | 0 | 0 | 0 | 17 | | 0 | 13 |
| TProt | Total protein | blood | 557 | 40 | 13 | 544 | 27 | FCGR2A; ZNRD1; RNF39; TRIM31-AS1; TRIM26; HLA-N; XXbac-BPG249D20.9; GNL1; ABCF1; ATAT1; FLOT1; HCG20; LINC00243; XXbac-BPG27H4.8; RN7SKP186; DDR1-AS1; DDR1; RN7SL175P; GTF2H4; VARS2; SFTA2; DPCR1; HCG21; MUC21; MUC22; HCG22; RNU6-1133P; C6orf15; PSORS1C1; CCHCR1; TCF19; POU5F1; XXbac-BPG299F13.17; HCG27; XXbac-BPG299F13.16; HLA-C; XXbac-BPG248L24.10; HLA-B; XXbac-BPG248L24.12; DHFRP2; RNU6-283P; AL671883.1; FGFR3P1; ZDHHC20P2; HLA-S; MICA; HCP5; MICB; XXbac-BPG16N22.5; ATP6V1G2-DDX39B; TNF; NCR3; PRRC2A; BAG6; APOM; GPANK1; CSNK2B; ABHD16A; XXbac-BPG32J3.20; CLIC1; MSH5; VWA7; VARS; LSM2; HSPA1L; HSPA1A; HSPA1B; C6orf48; NEU1; SLC44A4; C2; CFB; STK19; TNXB; ATF6B; PRRT1; PPT2; PPT2-EGFL8; AGPAT1; RNF5; AGER; PBX2; GPSM3; NOTCH4; XXbac-BPG154L12.4; C6orf10; BTNL2; HLA-DRA; HLA-DRB9; HLA-DQA1; HLA-DQB1; MTCO3P1; NOSIP; PRR12 | HCG17; TRIM39; MICC; RN7SL353P; HCG20; LINC00243; SFTA2; DPCR1; HCG21; HLA-DQB1; FADS2; TNFRSF13B | 863 | 289 | 210 | 653 | 79 | 0 | 0 | 0 | 0 | 28 | 4 | 11 | 28 |

| all_labs | full_name | group | male.significant.variants | female.significant.variants | both.significant.variants | male.only.significant.variants | female.only.significant.variants | male.annotated.genes.significant.variants | female.annotated.genes.significant.variants | male.suggestive.variants | female.suggestive.variants | both.suggestive.variants | male.only.suggestive.variants | female.only.suggestive.variants | female.significant.x.variants | male.significant.x.variants | female.suggestive.x.variants | male.suggestive.x.variants | n_loci | n_loci_noHLA | P1e6_both | P1e2_both |
|---|---|---|---|---|---|---|---|---|---|---|---|---|---|---|---|---|---|---|---|---|---|---|
| Trigs | Triglyceride [Mass/volume] in Serum or Plasma | metabolic | 731 | 862 | 684 | 47 | 178 | USP1; DOCK7; RP11-230B22.1; GTF3C2; PPM1G; NRBP1; IFT172; GCKR; AC109829.1; C2orf16; ZNF512; SLC4A1AP; AC074091.13; HLA-B; BAZ1B; BCL7B; TBL2; MLXIPL; LPL; AC100802.3; RP11-136O12.2; AP000770.1; BUD13; AP006216.10; AP006216.11; ZNF259; APOA5; AP006216.5; APOA4; APOC3; APOA1; APOA1-AS; SIK3; AP000936.4; AP000936.5; RP11-109L13.1; PAFAH1B2; SIDT2; TAGLN; PCSK7; TM6SF2; APOE; APOC1; APOC1P1 | USP1; DOCK7; RP11-230B22.1; MPV17; GTF3C2; AC074117.10; EIF2B4; SNX17; ZNF513; PPM1G; NRBP1; KRTCAP3; IFT172; FNDC4; GCKR; AC109829.1; C2orf16; ZNF512; BAZ1B; BCL7B; TBL2; MLXIPL; LPL; AC100802.3; RP11-136O12.2; AP000770.1; BUD13; AP006216.10; AP006216.11; ZNF259; APOA5; AP006216.5; APOA4; APOC3; APOA1; APOA1-AS; SIK3; AP000936.4; AP000936.5; RP11-109L13.1; PAFAH1B2; SIDT2; PCSK7; APOE | 828 | 1094 | 730 | 98 | 364 | 0 | 0 | 0 | 0 | 38 | 37 | 30 | 38 |
| Trnsfr | Transferrin [Mass/volume] in Serum or Plasma | metabolic | 0 | 0 | 0 | 0 | 0 | | | 1 | 6 | 0 | 1 | 6 | 0 | 0 | 0 | 0 | 0 | | 0 | 0 |
| Tropn T | Troponin T.cardiac [Mass/volume] in Serum or Plasma | cardiovascular | 0 | 0 | 0 | 0 | 0 | | | 25 | 0 | 0 | 25 | 0 | 0 | 0 | 0 | 0 | 0 | | 0 | 0 |
| TRPI | Troponin I.cardiac [Mass/volume] in Serum or Plasma | cardiovascular | 0 | 0 | 0 | 0 | 0 | | | 2 | 0 | 0 | 2 | 0 | 0 | 0 | 0 | 0 | 0 | | 0 | 0 |
| TSH_3RD_GENERATION | Thyroid stimulating hormone (3rd generation) | endocrine | 0 | 0 | 0 | 0 | 0 | | | 18 | 0 | 0 | 18 | 0 | 0 | 0 | 0 | 0 | 0 | | 0 | 0 |
| TSH | Thyroid stimulating hormone | endocrine | 397 | 750 | 267 | 130 | 483 | CAPZB; RNU4-28P; RP5-1056L3.1; MINOS1-NBL1; AC007563.5; ZBED3-AS1; PDE8B; RP11-344J7.2; PDE10A; NRG1; RP11-23B15.1; FOXE1; ABO; TSHR; AC009159.1; RP11-345M22.1; INSR; RP4-788L20.3; RP11-359G22.2 | CAPZB; RNU4-28P; MINOS1-NBL1; VAV3; AC007563.5; SYN2; RP11-503L19.1; ATP5LP4; PDE8B; RP11-344J7.2; RP5-1120P11.1; PDE10A; NRG1; RP11-546O6.4; RP11-23B15.1; FOXE1; C9orf156; ABO; PRDM11; CTD-2560E9.3; SECISBP2L; RP11-325E5.1; COPS2; GALK2; FAM227B; DTWD1; RP11-353B9.1; AC009159.1; RP11-345M22.1; FOXA2; RP4-788L20.3; RP11-359G22.2 | 599 | 996 | 399 | 200 | 597 | 0 | 0 | 0 | 0 | 54 | 54 | 31 | 50 |
| TT4 | Thyroxine (T4) [Mass/volume] in Serum or Plasma | endocrine | 115 | 30 | 30 | 85 | 0 | IL1RAPL2; NRK; SERPINA7; RP11-560L11.1 | IL1RAPL2; SERPINA7; RP11-560L11.1 | 134 | 35 | 34 | 100 | 1 | 30 | 115 | 34 | 130 | 4 | 4 | 2 | 4 |

| all_labs | full_name | group | male.significant.variants | female.significant.variants | both.significant.variants | male.only.significant.variants | female.only.significant.variants | male.annotated.genes.significant.variants | female.annotated.genes.significant.variants | male.suggestive.variants | female.suggestive.variants | both.suggestive.variants | male.only.suggestive.variants | female.only.suggestive.variants | female.significant.x.variants | male.significant.x.variants | female.suggestive.x.variants | male.suggestive.x.variants | n_loci | n_loci_noHLA | P1e6_both | P1e2_both |
|---|---|---|---|---|---|---|---|---|---|---|---|---|---|---|---|---|---|---|---|---|---|---|
| TTAIgA | Tissue transglutaminase IgA Ab [Units/volume] in Serum by Immunoassay | immune / inflammatory | 0 | 0 | 0 | 0 | 0 | | | 0 | 5 | 0 | 0 | 5 | 0 | 0 | 0 | 0 | 0 | | 0 | 0 |
| TU-Pro | 24 hour urinary protein | urinary | 0 | 0 | 0 | 0 | 0 | | | 2 | 1 | 0 | 2 | 1 | 0 | 0 | 1 | 0 | 0 | | 0 | 0 |
| TUCrea | Creatinine [Mass/volume] in Urine | kidney | 0 | 0 | 0 | 0 | 0 | | | 0 | 7 | 0 | 0 | 7 | 0 | 0 | 0 | 0 | 0 | | 0 | 0 |
| UABM | Urinary albumin | kidney | 0 | 0 | 0 | 0 | 0 | | | 1 | 0 | 0 | 1 | 0 | 0 | 0 | 0 | 0 | 0 | | 0 | 0 |
| UAlb | Albumin [Presence] in Urine | urinary | 0 | 0 | 0 | 0 | 0 | | | 2 | 0 | 0 | 2 | 0 | 0 | 0 | 0 | 0 | 0 | | 0 | 0 |
| UGluc | Glucose urine dipstick | urinary | 0 | 0 | 0 | 0 | 0 | | | 1 | 0 | 0 | 1 | 0 | 0 | 0 | 0 | 1 | 0 | | 0 | 0 |
| UGrCst | Granular casts [#/area] in Urine sediment by Microscopy high power field | urinary | 0 | 0 | 0 | 0 | 0 | | | 0 | 0 | 0 | 0 | 0 | 0 | 0 | 0 | 0 | 0 | | 0 | 0 |
| UHyCst | Hyaline casts in urine | urinary | 0 | 0 | 0 | 0 | 0 | | | 0 | 0 | 0 | 0 | 0 | 0 | 0 | 0 | 0 | 0 | | 0 | 0 |
| UKeto | Ketones [Mass/volume] in Urine by Test strip | urinary | 0 | 0 | 0 | 0 | 0 | | | 0 | 0 | 0 | 0 | 0 | 0 | 0 | 0 | 0 | 0 | | 0 | 0 |
| UpH | pH of Urine by Test strip | urinary | 1 | 0 | 0 | 1 | 0 | WDR72 | | 71 | 44 | 1 | 70 | 43 | 0 | 0 | 0 | 0 | 1 | 1 | 1 | 1 |

| all_labs | full_name | group | male.significant.variants | female.significant.variants | both.significant.variants | male.only.significant.variants | female.only.significant.variants | male.annotated.genes.significant.variants | female.annotated.genes.significant.variants | male.suggestive.variants | female.suggestive.variants | both.suggestive.variants | male.only.suggestive.variants | female.only.suggestive.variants | female.significant.x.variants | male.significant.x.variants | female.suggestive.x.variants | male.suggestive.x.variants | n_loci | n_loci_noHLA | P1e6_both | P1e2_both |
|---|---|---|---|---|---|---|---|---|---|---|---|---|---|---|---|---|---|---|---|---|---|---|
| URBC | Erythrocytes [#/area] in Urine sediment by Microscopy high power field | urinary | 0 | 0 | 0 | 0 | 0 | | | 0 | 0 | 0 | 0 | 0 | 0 | 0 | 0 | 0 | 0 | | 0 | 0 |
| UricA | Urate [ Mass/volume] in Serum or Plasma | urinary | 123 | 528 | 123 | 0 | 405 | SLC2A9; RP11-448G15.3; WDR1; RP11-480G3.1; AC006499.6; AC006499.9; AC006499.4; AC006499.3; AC006499.2; AC006499.1; RP11-136I13.1 | SLC2A9; RP11-448G15.3; WDR1; RP11-480G3.1; AC006499.7; AC006499.6; AC006499.9; AC006499.8; AC006499.5; AC006499.4; AC006499.3; AC006499.2; AC006499.1; RP11-136I13.1 | 177 | 790 | 175 | 2 | 615 | 0 | 0 | 0 | 0 | 13 | 13 | 5 | 11 |
| UrobUr | Urobilinogen [Units/volume] in Urine by Test strip | urinary | 0 | 0 | 0 | 0 | 0 | | | 1 | 0 | 0 | 1 | 0 | 0 | 0 | 0 | 0 | 0 | | 0 | 0 |
| all_labs | full_name | group | male.significant.variants | female.significant.variants | both.significant.variants | male.only.significant.variants | female.only.significant.variants | male.annotated.genes.significant.variants | female.annotated.genes.significant.variants | male.suggestive.variants | female.suggestive.variants | both.suggestive.variants | male.only.suggestive.variants | female.only.suggestive.variants | female.significant.x.variants | male.significant.x.variants | female.suggestive.x.variants | male.suggestive.x.variants | n_loci | n_loci_noHLA | P1e6_both | P1e2_both |
| UrPro | Protein [Mass/volume] in Urine by Test strip | urinary | 0 | 0 | 0 | 0 | 0 | | | 0 | 0 | 0 | 0 | 0 | 0 | 0 | 0 | 0 | 0 | | 0 | 0 |
| USpGr | Specific gravity of Urine by Test strip | urinary | 0 | 0 | 0 | 0 | 0 | | | 8 | 1 | 0 | 8 | 1 | 0 | 0 | 0 | 3 | 0 | | 0 | 0 |
| USqEpi | Epithelial cells.squamous [#/area] in Urine sediment by Microscopy high power field | urinary | 0 | 0 | 0 | 0 | 0 | | | 5 | 4 | 0 | 5 | 4 | 0 | 0 | 0 | 0 | 0 | | 0 | 0 |
| UTP | Total urine protein | urinary | 0 | 0 | 0 | 0 | 0 | | | 2 | 4 | 0 | 2 | 4 | 0 | 0 | 0 | 0 | 0 | | 0 | 0 |
| UUrob | Urobilinogen [Presence] in Urine by Test strip | urinary | 0 | 0 | 0 | 0 | 0 | | | 0 | 2 | 0 | 0 | 2 | 0 | 0 | 0 | 0 | 0 | | 0 | 0 |

| all_labs | full_name | group | male.significant.variants | female.significant.variants | both.significant.variants | male.only.significant.variants | female.only.significant.variants | male.annotated.genes.significant.variants | female.annotated.genes.significant.variants | male.suggestive.variants | female.suggestive.variants | both.suggestive.variants | male.only.suggestive.variants | female.only.suggestive.variants | female.significant.x.variants | male.significant.x.variants | female.suggestive.x.variants | male.suggestive.x.variants | n_loci | n_loci_noHLA | P1e6_both | P1e2_both |
|---|---|---|---|---|---|---|---|---|---|---|---|---|---|---|---|---|---|---|---|---|---|---|
| UWBC | Leukocytes [#/area] in Urine sediment by Microscopy high power field | urinary | 1 | 0 | 0 | 1 | 0 | ENTPD4 | | 21 | 0 | 0 | 21 | 0 | 0 | 0 | 0 | 0 | 1 | 1 | 0 | 0 |
| VA_BTPS_PPR | VA BTPS PPR | lung | 0 | 0 | 0 | 0 | 0 | | | 0 | 1 | 0 | 0 | 1 | 0 | 0 | 0 | 0 | 0 | | 0 | 0 |
| VA_BTPS_PRD | VA BTPS PRD | lung | 0 | 0 | 0 | 0 | 0 | | | 7 | 0 | 0 | 7 | 0 | 0 | 0 | 0 | 0 | 0 | | 0 | 0 |
| VA_BTPS_PRL | VA BTPS PRL | lung | 0 | 0 | 0 | 0 | 0 | | | 0 | 5 | 0 | 0 | 5 | 0 | 0 | 0 | 0 | 0 | | 0 | 0 |
| VA_BTPS_PRU | VA BTPS PRU | lung | 0 | 0 | 0 | 0 | 0 | | | 0 | 0 | 0 | 0 | 0 | 0 | 0 | 0 | 0 | 0 | | 0 | 0 |
| VA_BTPS_ZBD | VA BTPS ZBD | lung | 0 | 0 | 0 | 0 | 0 | | | 0 | 0 | 0 | 0 | 0 | 0 | 0 | 0 | 0 | 0 | | 0 | 0 |
| VC_PPR | VC PPR | lung | 0 | 0 | 0 | 0 | 0 | | | 24 | 0 | 0 | 24 | 0 | 0 | 0 | 0 | 0 | 0 | | 0 | 0 |
| VC_PRD | VC PRD | lung | 0 | 0 | 0 | 0 | 0 | | | 1 | 0 | 0 | 1 | 0 | 0 | 0 | 0 | 0 | 0 | | 0 | 0 |
| VC_PRL | VC PRL | lung | 0 | 0 | 0 | 0 | 0 | | | 0 | 0 | 0 | 0 | 0 | 0 | 0 | 0 | 0 | 0 | | 0 | 0 |
| VC_ZBD | VC ZBD | lung | 0 | 0 | 0 | 0 | 0 | | | 0 | 0 | 0 | 0 | 0 | 0 | 0 | 0 | 0 | 0 | | 0 | 0 |
| VD25 | Calcitriol [Mass/volume] in Serum or Plasma | metabolic | 0 | 8 | 0 | 0 | 8 | | PDE3B; CYP2R1 | 0 | 11 | 0 | 0 | 11 | 0 | 0 | 0 | 0 | 1 | 1 | 0 | 0 |
| Vi-B12 | Vitamin B12 | metabolic | 61 | 40 | 25 | 36 | 15 | CUBN; FUT2; MAMSTR; RASIP1; IZUMO1; FGF21 | EEF1A1P42; MUT; CUBN; FUT2 | 73 | 92 | 33 | 40 | 59 | 0 | 0 | 0 | 0 | 6 | 6 | 3 | 6 |
| ViARet | Retinol [Mass/volume] in Serum or Plasma | metabolic | 0 | 0 | 0 | 0 | 0 | | | 0 | 0 | 0 | 0 | 0 | 0 | 0 | 0 | 0 | 0 | | 0 | 0 |
| VitaB6 | Pyridoxine [Mass/volume] in Serum or Plasma | metabolic | 0 | 0 | 0 | 0 | 0 | | | 2 | 3 | 0 | 2 | 3 | 0 | 0 | 0 | 0 | 0 | | 0 | 0 |
| VLDL | very-low-density lipoprotein | metabolic | 0 | 0 | 0 | 0 | 0 | | | 2 | 0 | 0 | 2 | 0 | 0 | 0 | 0 | 0 | 0 | | 0 | 0 |

| all_labs | full_name | group | male.significant.variants | female.significant.variants | both.significant.variants | male.only.significant.variants | female.only.significant.variants | male.annotated.genes.significant.variants | female.annotated.genes.significant.variants | male.suggestive.variants | female.suggestive.variants | both.suggestive.variants | male.only.suggestive.variants | female.only.suggestive.variants | female.significant.x.variants | male.significant.x.variants | female.suggestive.x.variants | male.suggestive.x.variants | n_loci | n_loci_noHLA | P1e6_both | P1e2_both |
|---|---|---|---|---|---|---|---|---|---|---|---|---|---|---|---|---|---|---|---|---|---|---|
| WBC | Leukocytes [#/volume] in Blood by Automated count | immune / inflammatory | 699 | 649 | 269 | 430 | 380 | SCD5; BTN2A2; BTN3A1; XXbac-BPGBPG34I8.1; HNRNPA1P1; XXbac-BPG34I8.3; RNU6-471P; TRNAI6; RP1-15D7.1; RP1-97D16.1; RSL24D1P1; HIST1H4PS1; HIST1H2BM; HIST1H4K; HIST1H2AK; HIST1H2BN; HIST1H2AL; HIST1H1B; HIST1H4L; RNU7-26P; OR2B2; OR2W6P; RPLP2P1; OR2W4P; IQCB2P; OR2W2P; OR2B7P; OR1F12; ZSCAN12P1; RP1-265C24.5; ZSCAN16-AS1; ZKSCAN8; ZNF192P1; AL022393.7; ZNF192P2; TOB2P1; ZSCAN9; PGBD1; RP5-874C20.6; ZSCAN31; ZKSCAN3; ZSCAN12; ZSCAN23; OR2E1P; GPX6; LINC00533; RPSAP2; AL662890.1; NOL5BP; AL662890.3; XXbac-BPG308K3.5; XXbac-BPG308K3.6; ZNF90P2; HCG14; TRIM27; C6orf100; KRT18P1; RN7SL471P; HCG15; HCG16; ZNF311; OR2AD1P; OR2W1; OR2P1P; SAR1P1; OR2B3; XXbac-BCX196D17.5; OR2N1P; OR2J4P; OR2H4P; XXbac-BPG308J9.3; OR2B4P; OR14J1; DDX6P1; OR5V1; OR11A1; UBDP1; MAS1LP1; MAS1L; RPS17P1; XXbac-BPG13B8.10; OR2I1P; UBD; GABBR1; SUMO2P1; MOG; ZFP57; HLA-F; XXbac-BPG170G13.31; HLA-V; MICF; HCG4P7; HLA-A; HLA-W; MICD; HCG9; DDX39BP2; ZNRD1-AS1; ZNRD1; RNF39; TRIM31; TRIM31-AS1; TRIM40; TRIM10; TRIM15; TRIM26; HCG17; TRIM39; HLA-N; HCG19P; UBQLN1P1; MICC; TMPOP1; SUCLA2P1; RANP1; C2; NELFE; SKIV2L; STK19; HLA-DRA; HLA-DRB9; HLA-DQB1; MTCO3P1; XXbac-BPG254F23.7; CDK6; RP11-387H17.4; LRRC3C; GSDMA; PSMD3; RP11-387H17.6; CSF3; MED24; THRA | RP11-629B11.4; CXCL2; MTHFD2L; SUMO2P1; MOG; ZFP57; ZDHHC20P1; XXbac-BPG170G13.31; HLA-V; MICF; HCG4P7; HLA-A; HLA-W; MICD; DDX39BP2; ZNRD1-AS1; ZNRD1; RNF39; TRIM31-AS1; HCG19P; UBQLN1P1; MICC; MUC21; MUC22; HCG22; RNU6-1133P; C6orf15; PSORS1C1; TCF19; HCG27; XXbac-BPG299F13.16; HLA-C; USP8P1; RPL3P2; WASF5P; XXbac-BPG248L24.13; XXbac-BPG248L24.10; HLA-B; XXbac-BPG248L24.12; DHFRP2; RNU6-283P; AL671883.1; FGFR3P1; ZDHHC20P2; HLA-S; XXbac-BPG181B23.7; MICA; HCP5; MICB; XXbac-BPG16N22.5; ATP6V1G2-DDX39B; TNF; NCR3; HLA-DRB9; HLA-DQA1; HLA-DQB1; MTCO3P1; XXbac-BPG254F23.7; STARD3; PGAP3; RP11-94L15.2; IKZF3; ZPBP2; GSDMB; ORMDL3; RP11-387H17.4; LRRC3C; GSDMA; PSMD3; RP11-387H17.6; CSF3; MED24; THRA | 1308 | 1192 | 567 | 741 | 625 | 0 | 0 | 0 | 0 | 34 | 17 | 15 | 30 |
| WBCCnt | Leukocytes [#/volume] in Blood by Manual count | immune / inflammatory | 0 | 0 | 0 | 0 | 0 | | | 0 | 1 | 0 | 0 | 1 | 0 | 0 | 0 | 0 | 0 | | 0 | 0 |
| WBCrt | Creatinine [Mass/volume] in Blood | blood | 19 | 0 | 0 | 19 | 0 | UMOD | | 29 | 2 | 0 | 29 | 2 | 0 | 0 | 0 | 0 | 1 | 1 | 0 | 0 |
| WBINR | International normalized ratio | blood | 0 | 0 | 0 | 0 | 0 | | | 7 | 0 | 0 | 7 | 0 | 0 | 0 | 0 | 0 | 0 | | 0 | 0 |
| WBTrpI | Troponin I.cardiac [Mass/volume] in Blood | cardiovascular | 0 | 0 | 0 | 0 | 0 | | | 1 | 2 | 0 | 1 | 2 | 0 | 0 | 0 | 0 | 0 | | 0 | 0 |

| all_labs | full_name | group | male.significant.variants | female.significant.variants | both.significant.variants | male.only.significant.variants | female.only.significant.variants | male.annotated.genes.significant.variants | female.annotated.genes.significant.variants | male.suggestive.variants | female.suggestive.variants | both.suggestive.variants | male.only.suggestive.variants | female.only.suggestive.variants | female.significant.x.variants | male.significant.x.variants | female.suggestive.x.variants | male.suggestive.x.variants | n_loci | n_loci_noHLA | P1e6_both | P1e2_both |
|---|---|---|---|---|---|---|---|---|---|---|---|---|---|---|---|---|---|---|---|---|---|---|
| WEIGHT_SCALE | Weight scale | vitals | 18 | 87 | 18 | 0 | 69 | FTO | FTO | 120 | 112 | 76 | 44 | 36 | 0 | 0 | 0 | 0 | 2 | 2 | 1 | 2 |
| Weight | Weight | vitals | 94 | 538 | 92 | 2 | 446 | NQO2; FTO; RP11-795H16.3 | SEC16B; AC093326.3; TMEM18; ZBTB38; EIF2B5; BDNF-AS; BDNF; FAIM2; FTO; RNU6-567P; RP11-795H16.3; RNU4-17P; RP11-795H16.2; RP11-396N11.1; RASIP1; IZUMO1; FGF21 | 154 | 709 | 143 | 11 | 566 | 0 | 0 | 0 | 0 | 17 | 17 | 5 | 14 |
| Zinc | Zinc | metabolic | 0 | 0 | 0 | 0 | 0 | | | 1 | 0 | 0 | 1 | 0 | 0 | 0 | 0 | 0 | 0 | | 0 | 0 |

| P5e8_both | P1e6_both_noHLA | P1e2_both_noHLA | P5e8_both_noHLA | P1e6_female | P1e2_female | P5e8_female | P1e6_female_noHLA | P1e2_female_noHLA | P5e8_female_noHLA | P1e6_male | P1e2_male | P5e8_male | P1e6_male_noHLA | P1e2_male_noHLA | P5e8_male_noHLA | n_in_embl_any | n_in_embl_gene | n_in_embl_any_or_gene | n_in_embl_any_noHLA | n_in_embl_gene_noHLA | n_in_embl_any_or_gene_noHLA |
|---|---|---|---|---|---|---|---|---|---|---|---|---|---|---|---|---|---|---|---|---|---|
| 0 | 0 | 0 | 0 | 0 | 0 | 0 | 0 | 0 | 0 | 0 | 0 | 0 | 0 | 0 | 0 | | | | | | |
| 9 | 10 | 13 | 9 | 4 | 2 | 5 | 4 | 2 | 5 | 1 | 0 | 1 | 1 | 0 | 1 | | | | | | |
| 0 | 0 | 0 | 0 | 0 | 0 | 0 | 0 | 0 | 0 | 0 | 0 | 0 | 0 | 0 | 0 | | | | | | |
| 1 | 2 | 11 | 1 | 9 | 0 | 10 | 9 | 0 | 10 | 0 | 0 | 0 | 0 | 0 | 0 | 8 | 9 | 11 | 8 | 9 | 11 |
| 0 | 0 | 0 | 0 | 0 | 0 | 0 | 0 | 0 | 0 | 0 | 0 | 0 | 0 | 0 | 0 | | | | | | |
| 2 | 3 | 6 | 2 | 3 | 0 | 4 | 3 | 0 | 4 | 0 | 0 | 0 | 0 | 0 | 0 | 0 | 4 | 4 | 0 | 4 | 4 |
| 0 | 0 | 0 | 0 | 0 | 0 | 0 | 0 | 0 | 0 | 0 | 0 | 0 | 0 | 0 | 0 | | | | | | |
| 0 | 0 | 2 | 0 | 0 | 0 | 0 | 0 | 0 | 0 | 2 | 0 | 2 | 2 | 0 | 2 | | | | | | |
| 2 | 2 | 2 | 2 | 0 | 0 | 0 | 0 | 0 | 0 | 0 | 0 | 0 | 0 | 0 | 0 | 1 | 1 | 2 | 1 | 1 | 2 |
| 0 | 0 | 0 | 0 | 0 | 0 | 0 | 0 | 0 | 0 | 0 | 0 | 0 | 0 | 0 | 0 | | | | | | |
| 0 | 0 | 0 | 0 | 0 | 0 | 0 | 0 | 0 | 0 | 0 | 0 | 0 | 0 | 0 | 0 | | | | | | |
| 0 | 0 | 0 | 0 | 0 | 0 | 0 | 0 | 0 | 0 | 0 | 0 | 0 | 0 | 0 | 0 | | | | | | |
| 0 | 0 | 0 | 0 | 0 | 0 | 0 | 0 | 0 | 0 | 0 | 0 | 0 | 0 | 0 | 0 | | | | | | |
| 0 | 0 | 0 | 0 | 0 | 0 | 0 | 0 | 0 | 0 | 0 | 0 | 0 | 0 | 0 | 0 | | | | | | |
| 0 | 0 | 0 | 0 | 0 | 0 | 0 | 0 | 0 | 0 | 0 | 0 | 0 | 0 | 0 | 0 | | | | | | |
| 0 | 0 | 0 | 0 | 0 | 0 | 0 | 0 | 0 | 0 | 0 | 0 | 0 | 0 | 0 | 0 | | | | | | |
| 0 | 0 | 0 | 0 | 0 | 0 | 0 | 0 | 0 | 0 | 0 | 0 | 0 | 0 | 0 | 0 | | | | | | |
| 0 | 0 | 0 | 0 | 0 | 0 | 0 | 0 | 0 | 0 | 0 | 0 | 0 | 0 | 0 | 0 | | | | | | |
| 0 | 0 | 0 | 0 | 0 | 0 | 0 | 0 | 0 | 0 | 1 | 1 | 1 | 1 | 1 | 1 | 0 | 0 | 0 | 0 | 0 | 0 |
| 0 | 0 | 0 | 0 | 0 | 0 | 0 | 0 | 0 | 0 | 0 | 0 | 0 | 0 | 0 | 0 | | | | | | |
| 0 | 0 | 0 | 0 | 0 | 0 | 0 | 0 | 0 | 0 | 0 | 0 | 0 | 0 | 0 | 0 | | | | | | |
| 0 | 0 | 1 | 0 | 0 | 0 | 0 | 0 | 0 | 0 | 3 | 0 | 4 | 1 | 0 | 1 | 1 | 3 | 4 | 1 | 0 | 1 |
| 0 | 0 | 0 | 0 | 0 | 0 | 0 | 0 | 0 | 0 | 0 | 0 | 0 | 0 | 0 | 0 | | | | | | |
| 0 | 0 | 0 | 0 | 0 | 0 | 0 | 0 | 0 | 0 | 0 | 0 | 0 | 0 | 0 | 0 | | | | | | |
| 0 | 0 | 0 | 0 | 0 | 0 | 0 | 0 | 0 | 0 | 0 | 0 | 0 | 0 | 0 | 0 | | | | | | |
| 0 | 0 | 0 | 0 | 0 | 0 | 0 | 0 | 0 | 0 | 0 | 0 | 0 | 0 | 0 | 0 | | | | | | |
| 27 | 38 | 53 | 27 | 13 | 4 | 22 | 13 | 4 | 22 | 6 | 0 | 8 | 6 | 0 | 8 | 27 | 60 | 63 | 27 | 60 | 63 |
| 0 | 0 | 0 | 0 | 0 | 0 | 0 | 0 | 0 | 0 | 0 | 0 | 0 | 0 | 0 | 0 | | | | | | |
| 1 | 1 | 2 | 1 | 1 | 0 | 1 | 1 | 0 | 1 | 0 | 0 | 0 | 0 | 0 | 0 | 0 | 1 | 1 | 0 | 1 | 1 |
| 1 | 1 | 3 | 1 | 2 | 1 | 2 | 2 | 1 | 2 | 1 | 0 | 1 | 1 | 0 | 1 | 0 | 0 | 0 | 0 | 0 | 0 |
| 0 | 0 | 0 | 0 | 0 | 0 | 0 | 0 | 0 | 0 | 0 | 0 | 0 | 0 | 0 | 0 | | | | | | |
| 0 | 0 | 0 | 0 | 0 | 0 | 0 | 0 | 0 | 0 | 0 | 0 | 0 | 0 | 0 | 0 | | | | | | |
| 0 | 0 | 0 | 0 | 0 | 0 | 0 | 0 | 0 | 0 | 0 | 0 | 0 | 0 | 0 | 0 | | | | | | |
| 0 | 0 | 0 | 0 | 0 | 0 | 0 | 0 | 0 | 0 | 0 | 0 | 0 | 0 | 0 | 0 | | | | | | |
| 0 | 0 | 0 | 0 | 0 | 0 | 0 | 0 | 0 | 0 | 0 | 0 | 0 | 0 | 0 | 0 | | | | | | |
| 0 | 0 | 0 | 0 | 0 | 0 | 0 | 0 | 0 | 0 | 0 | 0 | 0 | 0 | 0 | 0 | | | | | | |
| 0 | 0 | 0 | 0 | 0 | 0 | 0 | 0 | 0 | 0 | 0 | 0 | 0 | 0 | 0 | 0 | | | | | | |
| 0 | 0 | 0 | 0 | 0 | 0 | 0 | 0 | 0 | 0 | 0 | 0 | 0 | 0 | 0 | 0 | | | | | | |
| 0 | 0 | 0 | 0 | 0 | 0 | 0 | 0 | 0 | 0 | 0 | 0 | 0 | 0 | 0 | 0 | | | | | | |
| 5 | 7 | 14 | 5 | 8 | 1 | 9 | 8 | 1 | 9 | 0 | 0 | 1 | 0 | 0 | 1 | 7 | 10 | 12 | 7 | 10 | 12 |
| 7 | 10 | 17 | 7 | 10 | 4 | 13 | 10 | 4 | 13 | 1 | 0 | 1 | 1 | 0 | 1 | 8 | 13 | 15 | 8 | 13 | 15 |
| 0 | 0 | 0 | 0 | 0 | 0 | 0 | 0 | 0 | 0 | 0 | 0 | 0 | 0 | 0 | 0 | | | | | | |
| 20 | 24 | 40 | 18 | 19 | 2 | 24 | 16 | 1 | 20 | 4 | 0 | 8 | 1 | 0 | 3 | 15 | 36 | 39 | 12 | 27 | 30 |
| 0 | 0 | 1 | 0 | 1 | 0 | 1 | 1 | 0 | 1 | 0 | 0 | 0 | 0 | 0 | 0 | 0 | 0 | 0 | 0 | 0 | 0 |
| 0 | 0 | 0 | 0 | 0 | 0 | 0 | 0 | 0 | 0 | 1 | 1 | 1 | 1 | 1 | 1 | 0 | 0 | 0 | 0 | 0 | 0 |
| 21 | 22 | 40 | 20 | 20 | 2 | 21 | 20 | 2 | 21 | 0 | 0 | 1 | 0 | 0 | 1 | 11 | 21 | 26 | 10 | 20 | 25 |
| 0 | 0 | 0 | 0 | 0 | 0 | 0 | 0 | 0 | 0 | 0 | 0 | 0 | 0 | 0 | 0 | | | | | | |
| 0 | 0 | 0 | 0 | 0 | 0 | 0 | 0 | 0 | 0 | 0 | 0 | 0 | 0 | 0 | 0 | | | | | | |
| 0 | 0 | 0 | 0 | 0 | 0 | 0 | 0 | 0 | 0 | 0 | 0 | 0 | 0 | 0 | 0 | | | | | | |
| 0 | 0 | 0 | 0 | 0 | 0 | 0 | 0 | 0 | 0 | 0 | 0 | 0 | 0 | 0 | 0 | | | | | | |
| 0 | 0 | 0 | 0 | 0 | 0 | 0 | 0 | 0 | 0 | 0 | 0 | 0 | 0 | 0 | 0 | | | | | | |

| P5e8_both | P1e6_both_noHLA | P1e2_both_noHLA | P5e8_both_noHLA | P1e6_female | P1e2_female | P5e8_female | P1e6_female_noHLA | P1e2_female_noHLA | P5e8_female_noHLA | P1e6_male | P1e2_male | P5e8_male | P1e6_male_noHLA | P1e2_male_noHLA | P5e8_male_noHLA | n_in_embl_any | n_in_embl_gene | n_in_embl_any_or_gene | n_in_embl_any_noHLA | n_in_embl_gene_noHLA | n_in_embl_any_or_gene_noHLA |
|---|---|---|---|---|---|---|---|---|---|---|---|---|---|---|---|---|---|---|---|---|---|
| 0 | 0 | 0 | 0 | 0 | 0 | 0 | 0 | 0 | 0 | 0 | 0 | 0 | 0 | 0 | 0 | | | | | | |
| 0 | 0 | 0 | 0 | 0 | 0 | 0 | 0 | 0 | 0 | 0 | 0 | 0 | 0 | 0 | 0 | | | | | | |
| 0 | 0 | 0 | 0 | 0 | 0 | 0 | 0 | 0 | 0 | 0 | 0 | 0 | 0 | 0 | 0 | | | | | | |
| 2 | 5 | 8 | 2 | 4 | 1 | 7 | 4 | 1 | 7 | 0 | 0 | 0 | 0 | 0 | 0 | 7 | 9 | 10 | 7 | 9 | 10 |
| 0 | 0 | 0 | 0 | 0 | 0 | 0 | 0 | 0 | 0 | 0 | 0 | 0 | 0 | 0 | 0 | | | | | | |
| 0 | 0 | 3 | 0 | 1 | 0 | 1 | 1 | 0 | 1 | 2 | 0 | 2 | 2 | 0 | 2 | 4 | 4 | 4 | 4 | 4 | 4 |
| 0 | 0 | 0 | 0 | 1 | 1 | 1 | 1 | 1 | 1 | 0 | 0 | 0 | 0 | 0 | 0 | | | | | | |
| 0 | 0 | 0 | 0 | 0 | 0 | 0 | 0 | 0 | 0 | 0 | 0 | 0 | 0 | 0 | 0 | | | | | | |
| 43 | 3 | 5 | 0 | 56 | 5 | 78 | 2 | 0 | 5 | 2 | 0 | 3 | 0 | 0 | 0 | 17 | 69 | 78 | 0 | 1 | 1 |
| P5e8_both | P1e6_both_noHLA | P1e2_both_noHLA | P5e8_both_noHLA | P1e6_female | P1e2_female | P5e8_female | P1e6_female_noHLA | P1e2_female_noHLA | P5e8_female_noHLA | P1e6_male | P1e2_male | P5e8_male | P1e6_male_noHLA | P1e2_male_noHLA | P5e8_male_noHLA | n_in_embl_any | n_in_embl_gene | n_in_embl_any_or_gene | n_in_embl_any_noHLA | n_in_embl_gene_noHLA | n_in_embl_any_or_gene_noHLA |
| 0 | 0 | 0 | 0 | 0 | 0 | 0 | 0 | 0 | 0 | 0 | 0 | 0 | 0 | 0 | 0 | | | | | | |
| 9 | 11 | 16 | 9 | 6 | 2 | 8 | 6 | 2 | 8 | 2 | 1 | 2 | 2 | 1 | 2 | 7 | 17 | 18 | 7 | 17 | 18 |
| 0 | 0 | 0 | 0 | 0 | 0 | 0 | 0 | 0 | 0 | 0 | 0 | 0 | 0 | 0 | 0 | | | | | | |
| 0 | 0 | 1 | 0 | 0 | 0 | 0 | 0 | 0 | 0 | 1 | 0 | 1 | 1 | 0 | 1 | 1 | 1 | 1 | 1 | 1 | 1 |
| 3 | 4 | 4 | 3 | 0 | 0 | 1 | 0 | 0 | 1 | 0 | 0 | 0 | 0 | 0 | 0 | 6 | 7 | 7 | 6 | 7 | 7 |
| 0 | 0 | 0 | 0 | 0 | 0 | 0 | 0 | 0 | 0 | 0 | 0 | 0 | 0 | 0 | 0 | | | | | | |
| 0 | 0 | 0 | 0 | 0 | 0 | 0 | 0 | 0 | 0 | 0 | 0 | 0 | 0 | 0 | 0 | | | | | | |
| 0 | 0 | 0 | 0 | 0 | 0 | 0 | 0 | 0 | 0 | 0 | 0 | 0 | 0 | 0 | 0 | | | | | | |
| 0 | 0 | 0 | 0 | 0 | 0 | 0 | 0 | 0 | 0 | 0 | 0 | 0 | 0 | 0 | 0 | | | | | | |
| 0 | 0 | 0 | 0 | 0 | 0 | 0 | 0 | 0 | 0 | 0 | 0 | 0 | 0 | 0 | 0 | | | | | | |
| 0 | 0 | 0 | 0 | 0 | 0 | 0 | 0 | 0 | 0 | 0 | 0 | 0 | 0 | 0 | 0 | | | | | | |
| 0 | 0 | 0 | 0 | 0 | 0 | 0 | 0 | 0 | 0 | 0 | 0 | 0 | 0 | 0 | 0 | | | | | | |
| 0 | 0 | 0 | 0 | 0 | 0 | 0 | 0 | 0 | 0 | 0 | 0 | 0 | 0 | 0 | 0 | | | | | | |
| 0 | 0 | 0 | 0 | 0 | 0 | 0 | 0 | 0 | 0 | 0 | 0 | 0 | 0 | 0 | 0 | | | | | | |
| 3 | 3 | 21 | 3 | 19 | 1 | 19 | 17 | 1 | 17 | 2 | 0 | 2 | 2 | 0 | 2 | 19 | 17 | 22 | 18 | 17 | 21 |
| 0 | 0 | 0 | 0 | 0 | 0 | 0 | 0 | 0 | 0 | 0 | 0 | 0 | 0 | 0 | 0 | | | | | | |
| 0 | 0 | 0 | 0 | 0 | 0 | 0 | 0 | 0 | 0 | 0 | 0 | 0 | 0 | 0 | 0 | | | | | | |
| 0 | 0 | 1 | 0 | 0 | 0 | 0 | 0 | 0 | 0 | 1 | 0 | 1 | 1 | 0 | 1 | 1 | 1 | 1 | 1 | 1 | 1 |
| 0 | 0 | 0 | 0 | 0 | 0 | 0 | 0 | 0 | 0 | 0 | 0 | 0 | 0 | 0 | 0 | | | | | | |
| 0 | 0 | 0 | 0 | 0 | 0 | 0 | 0 | 0 | 0 | 0 | 0 | 0 | 0 | 0 | 0 | | | | | | |
| 0 | 0 | 0 | 0 | 0 | 0 | 0 | 0 | 0 | 0 | 0 | 0 | 0 | 0 | 0 | 0 | | | | | | |
| 0 | 0 | 0 | 0 | 0 | 0 | 0 | 0 | 0 | 0 | 0 | 0 | 0 | 0 | 0 | 0 | | | | | | |
| 0 | 0 | 1 | 0 | 1 | 0 | 1 | 1 | 0 | 1 | 0 | 0 | 0 | 0 | 0 | 0 | 0 | 0 | 0 | 0 | 0 | 0 |
| 0 | 0 | 0 | 0 | 0 | 0 | 0 | 0 | 0 | 0 | 0 | 0 | 0 | 0 | 0 | 0 | | | | | | |
| 0 | 0 | 0 | 0 | 0 | 0 | 0 | 0 | 0 | 0 | 0 | 0 | 0 | 0 | 0 | 0 | | | | | | |
| 0 | 0 | 0 | 0 | 0 | 0 | 0 | 0 | 0 | 0 | 0 | 0 | 0 | 0 | 0 | 0 | | | | | | |
| 0 | 0 | 0 | 0 | 0 | 0 | 0 | 0 | 0 | 0 | 0 | 0 | 0 | 0 | 0 | 0 | | | | | | |
| 0 | 0 | 1 | 0 | 2 | 1 | 2 | 2 | 1 | 2 | 0 | 0 | 0 | 0 | 0 | 0 | 2 | 2 | 3 | 2 | 2 | 3 |
| 0 | 0 | 0 | 0 | 0 | 0 | 0 | 0 | 0 | 0 | 0 | 0 | 0 | 0 | 0 | 0 | | | | | | |
| 0 | 0 | 0 | 0 | 0 | 0 | 0 | 0 | 0 | 0 | 0 | 0 | 0 | 0 | 0 | 0 | | | | | | |
| 0 | 0 | 0 | 0 | 0 | 0 | 0 | 0 | 0 | 0 | 0 | 0 | 0 | 0 | 0 | 0 | | | | | | |
| 0 | 0 | 0 | 0 | 0 | 0 | 0 | 0 | 0 | 0 | 0 | 0 | 0 | 0 | 0 | 0 | | | | | | |
| 0 | 0 | 0 | 0 | 0 | 0 | 0 | 0 | 0 | 0 | 0 | 0 | 0 | 0 | 0 | 0 | | | | | | |
| 2 | 2 | 2 | 2 | 1 | 1 | 1 | 1 | 1 | 1 | 0 | 0 | 0 | 0 | 0 | 0 | 4 | 4 | 4 | 4 | 4 | 4 |
| 0 | 0 | 0 | 0 | 0 | 0 | 0 | 0 | 0 | 0 | 0 | 0 | 0 | 0 | 0 | 0 | | | | | | |
| 0 | 0 | 0 | 0 | 0 | 0 | 0 | 0 | 0 | 0 | 0 | 0 | 0 | 0 | 0 | 0 | | | | | | |
| 0 | 1 | 1 | 0 | 0 | 0 | 1 | 0 | 0 | 1 | 0 | 0 | 0 | 0 | 0 | 0 | 1 | 1 | 1 | 1 | 1 | 1 |
| 0 | 0 | 0 | 0 | 0 | 0 | 0 | 0 | 0 | 0 | 0 | 0 | 0 | 0 | 0 | 0 | | | | | | |
| 5 | 7 | 8 | 5 | 1 | 0 | 2 | 1 | 0 | 2 | 0 | 0 | 1 | 0 | 0 | 1 | 4 | 9 | 9 | 4 | 9 | 9 |
| 0 | 0 | 0 | 0 | 0 | 0 | 0 | 0 | 0 | 0 | 0 | 0 | 0 | 0 | 0 | 0 | | | | | | |
| 0 | 0 | 0 | 0 | 0 | 0 | 0 | 0 | 0 | 0 | 0 | 0 | 0 | 0 | 0 | 0 | | | | | | |

| P5e8_both | P1e6_both_noHLA | P1e2_both_noHLA | P5e8_both_noHLA | P1e6_female | P1e2_female | P5e8_female | P1e6_female_noHLA | P1e2_female_noHLA | P5e8_female_noHLA | P1e6_male | P1e2_male | P5e8_male | P1e6_male_noHLA | P1e2_male_noHLA | P5e8_male_noHLA | n_in_embl_any | n_in_embl_gene | n_in_embl_any_or_gene | n_in_embl_any_noHLA | n_in_embl_gene_noHLA | n_in_embl_any_or_gene_noHLA |
|---|---|---|---|---|---|---|---|---|---|---|---|---|---|---|---|---|---|---|---|---|---|
| 0 | 0 | 0 | 0 | 0 | 0 | 0 | 0 | 0 | 0 | 0 | 0 | 0 | 0 | 0 | 0 | | | | | | |
| 0 | 0 | 0 | 0 | 0 | 0 | 0 | 0 | 0 | 0 | 0 | 0 | 0 | 0 | 0 | 0 | | | | | | |
| 1 | 1 | 1 | 1 | 0 | 0 | 0 | 0 | 0 | 0 | 0 | 0 | 0 | 0 | 0 | 0 | 1 | 1 | 1 | 1 | 1 | 1 |
| 0 | 0 | 0 | 0 | 0 | 0 | 0 | 0 | 0 | 0 | 0 | 0 | 0 | 0 | 0 | 0 | | | | | | |
| 0 | 0 | 0 | 0 | 0 | 0 | 0 | 0 | 0 | 0 | 0 | 0 | 0 | 0 | 0 | 0 | | | | | | |
| 0 | 0 | 0 | 0 | 0 | 0 | 0 | 0 | 0 | 0 | 0 | 0 | 0 | 0 | 0 | 0 | | | | | | |
| 0 | 0 | 0 | 0 | 0 | 0 | 0 | 0 | 0 | 0 | 0 | 0 | 0 | 0 | 0 | 0 | | | | | | |
| 0 | 0 | 0 | 0 | 0 | 0 | 0 | 0 | 0 | 0 | 0 | 0 | 0 | 0 | 0 | 0 | | | | | | |
| 0 | 0 | 0 | 0 | 0 | 0 | 0 | 0 | 0 | 0 | 0 | 0 | 0 | 0 | 0 | 0 | | | | | | |
| 0 | 0 | 0 | 0 | 0 | 0 | 0 | 0 | 0 | 0 | 0 | 0 | 0 | 0 | 0 | 0 | | | | | | |
| 0 | 0 | 0 | 0 | 0 | 0 | 0 | 0 | 0 | 0 | 0 | 0 | 0 | 0 | 0 | 0 | | | | | | |
| 0 | 0 | 0 | 0 | 0 | 0 | 0 | 0 | 0 | 0 | 0 | 0 | 0 | 0 | 0 | 0 | | | | | | |
| 0 | 0 | 0 | 0 | 0 | 0 | 0 | 0 | 0 | 0 | 0 | 0 | 0 | 0 | 0 | 0 | | | | | | |
| 0 | 0 | 0 | 0 | 0 | 0 | 0 | 0 | 0 | 0 | 0 | 0 | 0 | 0 | 0 | 0 | | | | | | |
| 0 | 0 | 0 | 0 | 1 | 1 | 1 | 1 | 1 | 1 | 0 | 0 | 0 | 0 | 0 | 0 | | | | | | |
| 0 | 0 | 0 | 0 | 0 | 0 | 0 | 0 | 0 | 0 | 0 | 0 | 0 | 0 | 0 | 0 | | | | | | |
| 0 | 0 | 0 | 0 | 0 | 0 | 0 | 0 | 0 | 0 | 0 | 0 | 0 | 0 | 0 | 0 | | | | | | |
| 2 | 3 | 3 | 2 | 0 | 0 | 0 | 0 | 0 | 0 | 0 | 0 | 1 | 0 | 0 | 1 | 4 | 3 | 4 | 4 | 3 | 4 |
| 0 | 0 | 0 | 0 | 0 | 0 | 0 | 0 | 0 | 0 | 0 | 0 | 0 | 0 | 0 | 0 | | | | | | |

| P5e8_both | P1e6_both_noHLA | P1e2_both_noHLA | P5e8_both_noHLA | P1e6_female | P1e2_female | P5e8_female | P1e6_female_noHLA | P1e2_female_noHLA | P5e8_female_noHLA | P1e6_male | P1e2_male | P5e8_male | P1e6_male_noHLA | P1e2_male_noHLA | P5e8_male_noHLA | n_in_embl_any | n_in_embl_gene | n_in_embl_any_or_gene | n_in_embl_any_noHLA | n_in_embl_gene_noHLA | n_in_embl_any_or_gene_noHLA |
|---|---|---|---|---|---|---|---|---|---|---|---|---|---|---|---|---|---|---|---|---|---|
| 0 | 0 | 0 | 0 | 0 | 0 | 0 | 0 | 0 | 0 | 0 | 0 | 0 | 0 | 0 | 0 | | | | | | |
| 0 | 3 | 6 | 0 | 3 | 0 | 5 | 3 | 0 | 5 | 0 | 0 | 1 | 0 | 0 | 1 | 6 | 5 | 6 | 6 | 5 | 6 |
| 2 | 2 | 5 | 2 | 3 | 0 | 3 | 3 | 0 | 3 | 0 | 0 | 0 | 0 | 0 | 0 | 5 | 5 | 5 | 5 | 5 | 5 |
| 0 | 0 | 1 | 0 | 1 | 0 | 1 | 1 | 0 | 1 | 0 | 0 | 0 | 0 | 0 | 0 | 1 | 1 | 1 | 1 | 1 | 1 |
| 0 | 1 | 1 | 0 | 0 | 0 | 1 | 0 | 0 | 1 | 0 | 0 | 0 | 0 | 0 | 0 | 1 | 1 | 1 | 1 | 1 | 1 |
| 1 | 1 | 1 | 1 | 0 | 0 | 0 | 0 | 0 | 0 | 0 | 0 | 0 | 0 | 0 | 0 | 1 | 1 | 1 | 1 | 1 | 1 |
| 0 | 0 | 0 | 0 | 0 | 0 | 0 | 0 | 0 | 0 | 0 | 0 | 0 | 0 | 0 | 0 | | | | | | |
| 0 | 1 | 2 | 0 | 0 | 0 | 1 | 0 | 0 | 1 | 1 | 0 | 1 | 1 | 0 | 1 | 2 | 2 | 2 | 2 | 2 | 2 |
| 0 | 0 | 0 | 0 | 0 | 0 | 0 | 0 | 0 | 0 | 0 | 0 | 0 | 0 | 0 | 0 | | | | | | |
| 0 | 0 | 0 | 0 | 0 | 0 | 0 | 0 | 0 | 0 | 0 | 0 | 0 | 0 | 0 | 0 | | | | | | |
| 0 | 0 | 0 | 0 | 0 | 0 | 0 | 0 | 0 | 0 | 0 | 0 | 0 | 0 | 0 | 0 | | | | | | |
| 0 | 0 | 0 | 0 | 0 | 0 | 0 | 0 | 0 | 0 | 0 | 0 | 0 | 0 | 0 | 0 | | | | | | |
| 0 | 0 | 0 | 0 | 0 | 0 | 0 | 0 | 0 | 0 | 0 | 0 | 0 | 0 | 0 | 0 | | | | | | |
| 0 | 0 | 0 | 0 | 0 | 0 | 0 | 0 | 0 | 0 | 0 | 0 | 0 | 0 | 0 | 0 | | | | | | |
| 0 | 0 | 0 | 0 | 0 | 0 | 0 | 0 | 0 | 0 | 0 | 0 | 0 | 0 | 0 | 0 | | | | | | |
| 0 | 0 | 0 | 0 | 0 | 0 | 0 | 0 | 0 | 0 | 0 | 0 | 0 | 0 | 0 | 0 | | | | | | |
| 0 | 0 | 0 | 0 | 0 | 0 | 0 | 0 | 0 | 0 | 0 | 0 | 0 | 0 | 0 | 0 | | | | | | |
| 0 | 0 | 0 | 0 | 0 | 0 | 0 | 0 | 0 | 0 | 0 | 0 | 0 | 0 | 0 | 0 | | | | | | |
| 0 | 0 | 0 | 0 | 0 | 0 | 0 | 0 | 0 | 0 | 0 | 0 | 0 | 0 | 0 | 0 | | | | | | |
| 0 | 0 | 0 | 0 | 0 | 0 | 0 | 0 | 0 | 0 | 0 | 0 | 0 | 0 | 0 | 0 | | | | | | |
| 0 | 0 | 0 | 0 | 0 | 0 | 0 | 0 | 0 | 0 | 0 | 0 | 0 | 0 | 0 | 0 | | | | | | |
| 0 | 0 | 0 | 0 | 0 | 0 | 0 | 0 | 0 | 0 | 0 | 0 | 0 | 0 | 0 | 0 | | | | | | |
| 0 | 0 | 0 | 0 | 0 | 0 | 0 | 0 | 0 | 0 | 0 | 0 | 0 | 0 | 0 | 0 | | | | | | |
| 0 | 0 | 0 | 0 | 0 | 0 | 0 | 0 | 0 | 0 | 0 | 0 | 0 | 0 | 0 | 0 | | | | | | |
| 0 | 0 | 0 | 0 | 0 | 0 | 0 | 0 | 0 | 0 | 0 | 0 | 0 | 0 | 0 | 0 | | | | | | |
| 0 | 0 | 0 | 0 | 0 | 0 | 0 | 0 | 0 | 0 | 0 | 0 | 0 | 0 | 0 | 0 | | | | | | |
| 0 | 0 | 0 | 0 | 0 | 0 | 0 | 0 | 0 | 0 | 0 | 0 | 0 | 0 | 0 | 0 | | | | | | |
| 0 | 0 | 0 | 0 | 0 | 0 | 0 | 0 | 0 | 0 | 0 | 0 | 0 | 0 | 0 | 0 | | | | | | |
| 0 | 0 | 0 | 0 | 0 | 0 | 0 | 0 | 0 | 0 | 0 | 0 | 0 | 0 | 0 | 0 | | | | | | |
| 0 | 0 | 0 | 0 | 0 | 0 | 0 | 0 | 0 | 0 | 0 | 0 | 0 | 0 | 0 | 0 | | | | | | |
| 0 | 0 | 0 | 0 | 0 | 0 | 0 | 0 | 0 | 0 | 0 | 0 | 0 | 0 | 0 | 0 | | | | | | |

| P5e8_both | P1e6_both_noHLA | P1e2_both_noHLA | P5e8_both_noHLA | P1e6_female | P1e2_female | P5e8_female | P1e6_female_noHLA | P1e2_female_noHLA | P5e8_female_noHLA | P1e6_male | P1e2_male | P5e8_male | P1e6_male_noHLA | P1e2_male_noHLA | P5e8_male_noHLA | n_in_embl_any | n_in_embl_gene | n_in_embl_any_or_gene | n_in_embl_any_noHLA | n_in_embl_gene_noHLA | n_in_embl_any_or_gene_noHLA |
|---|---|---|---|---|---|---|---|---|---|---|---|---|---|---|---|---|---|---|---|---|---|
| 0 | 0 | 0 | 0 | 0 | 0 | 0 | 0 | 0 | 0 | 0 | 0 | 0 | 0 | 0 | 0 | | | | | | |
| 0 | 0 | 0 | 0 | 0 | 0 | 0 | 0 | 0 | 0 | 0 | 0 | 0 | 0 | 0 | 0 | | | | | | |
| 0 | 0 | 0 | 0 | 0 | 0 | 0 | 0 | 0 | 0 | 0 | 0 | 0 | 0 | 0 | 0 | | | | | | |
| 0 | 0 | 0 | 0 | 0 | 0 | 0 | 0 | 0 | 0 | 0 | 0 | 0 | 0 | 0 | 0 | | | | | | |
| 0 | 0 | 0 | 0 | 0 | 0 | 0 | 0 | 0 | 0 | 0 | 0 | 0 | 0 | 0 | 0 | | | | | | |
| 0 | 0 | 0 | 0 | 0 | 0 | 0 | 0 | 0 | 0 | 0 | 0 | 0 | 0 | 0 | 0 | | | | | | |
| 0 | 0 | 0 | 0 | 0 | 0 | 0 | 0 | 0 | 0 | 0 | 0 | 0 | 0 | 0 | 0 | | | | | | |
| 0 | 0 | 0 | 0 | 0 | 0 | 0 | 0 | 0 | 0 | 0 | 0 | 0 | 0 | 0 | 0 | | | | | | |
| 0 | 0 | 0 | 0 | 0 | 0 | 0 | 0 | 0 | 0 | 0 | 0 | 0 | 0 | 0 | 0 | | | | | | |
| 0 | 0 | 0 | 0 | 0 | 0 | 0 | 0 | 0 | 0 | 0 | 0 | 0 | 0 | 0 | 0 | | | | | | |
| 0 | 0 | 0 | 0 | 0 | 0 | 0 | 0 | 0 | 0 | 0 | 0 | 0 | 0 | 0 | 0 | | | | | | |
| 0 | 0 | 0 | 0 | 0 | 0 | 0 | 0 | 0 | 0 | 0 | 0 | 0 | 0 | 0 | 0 | | | | | | |
| 0 | 0 | 0 | 0 | 0 | 0 | 0 | 0 | 0 | 0 | 0 | 0 | 0 | 0 | 0 | 0 | | | | | | |
| 0 | 0 | 0 | 0 | 0 | 0 | 0 | 0 | 0 | 0 | 0 | 0 | 0 | 0 | 0 | 0 | | | | | | |
| 0 | 0 | 0 | 0 | 0 | 0 | 0 | 0 | 0 | 0 | 0 | 0 | 0 | 0 | 0 | 0 | | | | | | |
| 0 | 0 | 0 | 0 | 0 | 0 | 0 | 0 | 0 | 0 | 0 | 0 | 0 | 0 | 0 | 0 | | | | | | |
| 0 | 0 | 0 | 0 | 0 | 0 | 0 | 0 | 0 | 0 | 0 | 0 | 0 | 0 | 0 | 0 | | | | | | |
| 0 | 0 | 0 | 0 | 0 | 0 | 0 | 0 | 0 | 0 | 0 | 0 | 0 | 0 | 0 | 0 | | | | | | |
| 0 | 0 | 0 | 0 | 0 | 0 | 0 | 0 | 0 | 0 | 0 | 0 | 0 | 0 | 0 | 0 | | | | | | |
| 0 | 0 | 0 | 0 | 0 | 0 | 0 | 0 | 0 | 0 | 0 | 0 | 0 | 0 | 0 | 0 | | | | | | |
| 0 | 0 | 0 | 0 | 0 | 0 | 0 | 0 | 0 | 0 | 0 | 0 | 0 | 0 | 0 | 0 | | | | | | |
| 0 | 0 | 0 | 0 | 0 | 0 | 0 | 0 | 0 | 0 | 0 | 0 | 0 | 0 | 0 | 0 | | | | | | |
| 0 | 0 | 2 | 0 | 2 | 0 | 2 | 2 | 0 | 2 | 0 | 0 | 0 | 0 | 0 | 0 | 1 | 3 | 3 | 1 | 3 | 3 |
| 0 | 0 | 0 | 0 | 0 | 0 | 0 | 0 | 0 | 0 | 0 | 0 | 0 | 0 | 0 | 0 | | | | | | |
| 0 | 0 | 0 | 0 | 0 | 0 | 0 | 0 | 0 | 0 | 0 | 0 | 0 | 0 | 0 | 0 | | | | | | |
| 0 | 0 | 0 | 0 | 0 | 0 | 0 | 0 | 0 | 0 | 0 | 0 | 0 | 0 | 0 | 0 | | | | | | |
| 0 | 0 | 0 | 0 | 0 | 0 | 0 | 0 | 0 | 0 | 0 | 0 | 0 | 0 | 0 | 0 | | | | | | |
| 0 | 0 | 0 | 0 | 0 | 0 | 0 | 0 | 0 | 0 | 0 | 0 | 0 | 0 | 0 | 0 | | | | | | |
| 0 | 0 | 0 | 0 | 0 | 0 | 0 | 0 | 0 | 0 | 0 | 0 | 0 | 0 | 0 | 0 | | | | | | |

| P5e8_both | P1e6_both_noHLA | P1e2_both_noHLA | P5e8_both_noHLA | P1e6_female | P1e2_female | P5e8_female | P1e6_female_noHLA | P1e2_female_noHLA | P5e8_female_noHLA | P1e6_male | P1e2_male | P5e8_male | P1e6_male_noHLA | P1e2_male_noHLA | P5e8_male_noHLA | n_in_embl_any | n_in_embl_gene | n_in_embl_any_or_gene | n_in_embl_any_noHLA | n_in_embl_gene_noHLA | n_in_embl_any_or_gene_noHLA |
|---|---|---|---|---|---|---|---|---|---|---|---|---|---|---|---|---|---|---|---|---|---|
| 0 | 0 | 0 | 0 | 0 | 0 | 0 | 0 | 0 | 0 | 0 | 0 | 0 | 0 | 0 | 0 | | | | | | |
| 0 | 0 | 0 | 0 | 0 | 0 | 0 | 0 | 0 | 0 | 0 | 0 | 0 | 0 | 0 | 0 | | | | | | |
| 0 | 0 | 0 | 0 | 0 | 0 | 0 | 0 | 0 | 0 | 0 | 0 | 0 | 0 | 0 | 0 | | | | | | |
| 0 | 1 | 1 | 0 | 0 | 0 | 0 | 0 | 0 | 0 | 0 | 0 | 1 | 0 | 0 | 1 | 1 | 1 | 1 | 1 | 1 | 1 |
| 6 | 7 | 14 | 6 | 5 | 0 | 6 | 5 | 0 | 6 | 3 | 0 | 3 | 2 | 0 | 2 | 11 | 16 | 16 | 11 | 15 | 15 |
| 0 | 0 | 0 | 0 | 0 | 0 | 0 | 0 | 0 | 0 | 0 | 0 | 0 | 0 | 0 | 0 | | | | | | |
| 0 | 0 | 0 | 0 | 0 | 0 | 0 | 0 | 0 | 0 | 0 | 0 | 0 | 0 | 0 | 0 | | | | | | |
| 1 | 1 | 3 | 1 | 2 | 0 | 2 | 2 | 0 | 2 | 0 | 0 | 0 | 0 | 0 | 0 | 1 | 3 | 3 | 1 | 3 | 3 |
| 0 | 0 | 0 | 0 | 0 | 0 | 0 | 0 | 0 | 0 | 0 | 0 | 0 | 0 | 0 | 0 | | | | | | |
| 0 | 0 | 0 | 0 | 0 | 0 | 0 | 0 | 0 | 0 | 0 | 0 | 0 | 0 | 0 | 0 | | | | | | |
| 0 | 0 | 0 | 0 | 0 | 0 | 0 | 0 | 0 | 0 | 0 | 0 | 0 | 0 | 0 | 0 | | | | | | |
| 0 | 0 | 0 | 0 | 0 | 0 | 0 | 0 | 0 | 0 | 0 | 0 | 0 | 0 | 0 | 0 | | | | | | |
| 13 | 17 | 28 | 13 | 11 | 1 | 15 | 11 | 1 | 15 | 2 | 1 | 2 | 2 | 1 | 2 | 31 | 29 | 36 | 31 | 29 | 36 |
| 0 | 0 | 0 | 0 | 1 | 1 | 1 | 1 | 1 | 1 | 0 | 0 | 0 | 0 | 0 | 0 | 0 | 0 | 0 | 0 | 0 | 0 |
| 21 | 30 | 99 | 21 | 66 | 10 | 73 | 66 | 10 | 73 | 14 | 1 | 16 | 14 | 1 | 16 | 95 | 111 | 115 | 95 | 111 | 115 |
| 1 | 5 | 16 | 1 | 11 | 4 | 15 | 11 | 4 | 15 | 5 | 1 | 5 | 5 | 1 | 5 | 21 | 22 | 22 | 21 | 22 | 22 |
| 0 | 0 | 0 | 0 | 0 | 0 | 0 | 0 | 0 | 0 | 0 | 0 | 0 | 0 | 0 | 0 | | | | | | |
| 0 | 0 | 0 | 0 | 0 | 0 | 0 | 0 | 0 | 0 | 0 | 0 | 0 | 0 | 0 | 0 | | | | | | |
| 3 | 3 | 14 | 3 | 11 | 2 | 11 | 10 | 2 | 10 | 4 | 0 | 4 | 3 | 0 | 3 | 15 | 17 | 19 | 14 | 16 | 18 |
| 0 | 0 | 0 | 0 | 0 | 0 | 0 | 0 | 0 | 0 | 0 | 0 | 0 | 0 | 0 | 0 | | | | | | |
| 4 | 2 | 2 | 2 | 1 | 0 | 4 | 0 | 0 | 0 | 2 | 0 | 3 | 0 | 0 | 0 | 4 | 5 | 7 | 2 | 2 | 2 |

| P5e8_both | P1e6_both_noHLA | P1e2_both_noHLA | P5e8_both_noHLA | P1e6_female | P1e2_female | P5e8_female | P1e6_female_noHLA | P1e2_female_noHLA | P5e8_female_noHLA | P1e6_male | P1e2_male | P5e8_male | P1e6_male_noHLA | P1e2_male_noHLA | P5e8_male_noHLA | n_in_embl_any | n_in_embl_gene | n_in_embl_any_or_gene | n_in_embl_any_noHLA | n_in_embl_gene_noHLA | n_in_embl_any_or_gene_noHLA |
|---|---|---|---|---|---|---|---|---|---|---|---|---|---|---|---|---|---|---|---|---|---|
| 0 | 0 | 0 | 0 | 0 | 0 | 0 | 0 | 0 | 0 | 0 | 0 | 0 | 0 | 0 | 0 | | | | | | |
| 0 | 1 | 1 | 0 | 0 | 0 | 1 | 0 | 0 | 1 | 0 | 0 | 0 | 0 | 0 | 0 | 1 | 0 | 1 | 1 | 0 | 1 |
| 0 | 0 | 0 | 0 | 0 | 0 | 0 | 0 | 0 | 0 | 0 | 0 | 0 | 0 | 0 | 0 | | | | | | |
| 0 | 0 | 0 | 0 | 0 | 0 | 0 | 0 | 0 | 0 | 0 | 0 | 0 | 0 | 0 | 0 | | | | | | |
| 0 | 0 | 0 | 0 | 0 | 0 | 0 | 0 | 0 | 0 | 0 | 0 | 0 | 0 | 0 | 0 | | | | | | |
| 0 | 0 | 0 | 0 | 0 | 0 | 0 | 0 | 0 | 0 | 0 | 0 | 0 | 0 | 0 | 0 | | | | | | |
| 0 | 0 | 0 | 0 | 0 | 0 | 0 | 0 | 0 | 0 | 0 | 0 | 0 | 0 | 0 | 0 | | | | | | |
| 0 | 0 | 0 | 0 | 0 | 0 | 0 | 0 | 0 | 0 | 0 | 0 | 0 | 0 | 0 | 0 | | | | | | |
| 0 | 0 | 0 | 0 | 0 | 0 | 0 | 0 | 0 | 0 | 0 | 0 | 0 | 0 | 0 | 0 | | | | | | |
| 0 | 0 | 1 | 0 | 0 | 0 | 0 | 0 | 0 | 0 | 1 | 0 | 1 | 1 | 0 | 1 | 0 | 0 | 0 | 0 | 0 | 0 |
| 0 | 0 | 0 | 0 | 0 | 0 | 0 | 0 | 0 | 0 | 0 | 0 | 0 | 0 | 0 | 0 | | | | | | |
| 3 | 6 | 9 | 3 | 1 | 0 | 2 | 1 | 0 | 2 | 2 | 0 | 4 | 2 | 0 | 4 | | | | | | |
| 0 | 0 | 0 | 0 | 0 | 0 | 0 | 0 | 0 | 0 | 0 | 0 | 0 | 0 | 0 | 0 | | | | | | |
| 0 | 0 | 0 | 0 | 0 | 0 | 0 | 0 | 0 | 0 | 0 | 0 | 0 | 0 | 0 | 0 | | | | | | |
| 0 | 0 | 0 | 0 | 1 | 1 | 1 | 1 | 1 | 1 | 0 | 0 | 0 | 0 | 0 | 0 | 0 | 0 | 0 | 0 | 0 | 0 |
| 8 | 10 | 15 | 8 | 0 | 0 | 1 | 0 | 0 | 1 | 5 | 0 | 6 | 5 | 0 | 6 | 1 | 3 | 3 | 1 | 3 | 3 |
| 0 | 0 | 0 | 0 | 0 | 0 | 0 | 0 | 0 | 0 | 0 | 0 | 0 | 0 | 0 | 0 | | | | | | |
| 0 | 0 | 0 | 0 | 0 | 0 | 0 | 0 | 0 | 0 | 0 | 0 | 0 | 0 | 0 | 0 | | | | | | |
| 0 | 0 | 0 | 0 | 0 | 0 | 0 | 0 | 0 | 0 | 0 | 0 | 0 | 0 | 0 | 0 | | | | | | |
| 0 | 0 | 0 | 0 | 0 | 0 | 0 | 0 | 0 | 0 | 0 | 0 | 0 | 0 | 0 | 0 | | | | | | |
| 0 | 0 | 0 | 0 | 0 | 0 | 0 | 0 | 0 | 0 | 0 | 0 | 0 | 0 | 0 | 0 | | | | | | |
| 0 | 0 | 0 | 0 | 0 | 0 | 0 | 0 | 0 | 0 | 0 | 0 | 0 | 0 | 0 | 0 | | | | | | |
| 0 | 0 | 0 | 0 | 0 | 0 | 0 | 0 | 0 | 0 | 0 | 0 | 0 | 0 | 0 | 0 | | | | | | |
| 0 | 0 | 0 | 0 | 0 | 0 | 0 | 0 | 0 | 0 | 0 | 0 | 0 | 0 | 0 | 0 | | | | | | |
| 0 | 0 | 0 | 0 | 0 | 0 | 0 | 0 | 0 | 0 | 0 | 0 | 0 | 0 | 0 | 0 | | | | | | |
| 0 | 0 | 0 | 0 | 0 | 0 | 0 | 0 | 0 | 0 | 0 | 0 | 0 | 0 | 0 | 0 | | | | | | |
| 0 | 0 | 0 | 0 | 0 | 0 | 0 | 0 | 0 | 0 | 0 | 0 | 0 | 0 | 0 | 0 | | | | | | |
| 0 | 0 | 0 | 0 | 0 | 0 | 0 | 0 | 0 | 0 | 0 | 0 | 0 | 0 | 0 | 0 | | | | | | |
| 0 | 0 | 0 | 0 | 0 | 0 | 0 | 0 | 0 | 0 | 0 | 0 | 0 | 0 | 0 | 0 | | | | | | |
| 0 | 0 | 0 | 0 | 0 | 0 | 0 | 0 | 0 | 0 | 0 | 0 | 0 | 0 | 0 | 0 | | | | | | |
| 0 | 0 | 0 | 0 | 0 | 0 | 0 | 0 | 0 | 0 | 0 | 0 | 0 | 0 | 0 | 0 | | | | | | |
| 0 | 0 | 0 | 0 | 0 | 0 | 0 | 0 | 0 | 0 | 0 | 0 | 0 | 0 | 0 | 0 | | | | | | |
| 5 | 6 | 9 | 5 | 3 | 0 | 4 | 3 | 0 | 4 | 0 | 0 | 0 | 0 | 0 | 0 | 3 | 7 | 7 | 3 | 7 | 7 |
| 0 | 0 | 0 | 0 | 0 | 0 | 0 | 0 | 0 | 0 | 0 | 0 | 0 | 0 | 0 | 0 | | | | | | |
| 0 | 0 | 3 | 0 | 3 | 0 | 3 | 3 | 0 | 3 | 1 | 0 | 1 | 0 | 0 | 0 | 1 | 2 | 3 | 1 | 2 | 3 |
| 0 | 0 | 0 | 0 | 0 | 0 | 0 | 0 | 0 | 0 | 0 | 0 | 0 | 0 | 0 | 0 | | | | | | |
| 0 | 0 | 0 | 0 | 0 | 0 | 0 | 0 | 0 | 0 | 0 | 0 | 0 | 0 | 0 | 0 | | | | | | |
| 0 | 0 | 0 | 0 | 0 | 0 | 0 | 0 | 0 | 0 | 0 | 0 | 0 | 0 | 0 | 0 | | | | | | |
| 0 | 0 | 0 | 0 | 0 | 0 | 0 | 0 | 0 | 0 | 0 | 0 | 0 | 0 | 0 | 0 | | | | | | |

| P5e8_both | P1e6_both_noHLA | P1e2_both_noHLA | P5e8_both_noHLA | P1e6_female | P1e2_female | P5e8_female | P1e6_female_noHLA | P1e2_female_noHLA | P5e8_female_noHLA | P1e6_male | P1e2_male | P5e8_male | P1e6_male_noHLA | P1e2_male_noHLA | P5e8_male_noHLA | n_in_embl_any | n_in_embl_gene | n_in_embl_any_or_gene | n_in_embl_any_noHLA | n_in_embl_gene_noHLA | n_in_embl_any_or_gene_noHLA |
|---|---|---|---|---|---|---|---|---|---|---|---|---|---|---|---|---|---|---|---|---|---|
| 0 | 0 | 0 | 0 | 0 | 0 | 0 | 0 | 0 | 0 | 0 | 0 | 0 | 0 | 0 | 0 | | | | | | |
| 0 | 0 | 0 | 0 | 0 | 0 | 0 | 0 | 0 | 0 | 0 | 0 | 0 | 0 | 0 | 0 | | | | | | |
| 0 | 0 | 0 | 0 | 0 | 0 | 0 | 0 | 0 | 0 | 0 | 0 | 0 | 0 | 0 | 0 | | | | | | |
| 0 | 0 | 0 | 0 | 0 | 0 | 0 | 0 | 0 | 0 | 0 | 0 | 0 | 0 | 0 | 0 | | | | | | |
| 6 | 9 | 28 | 6 | 20 | 1 | 23 | 20 | 1 | 23 | 0 | 0 | 0 | 0 | 0 | 0 | 23 | 26 | 30 | 23 | 26 | 30 |
| 0 | 0 | 0 | 0 | 0 | 0 | 0 | 0 | 0 | 0 | 0 | 0 | 0 | 0 | 0 | 0 | | | | | | |
| 0 | 0 | 0 | 0 | 0 | 0 | 0 | 0 | 0 | 0 | 0 | 0 | 0 | 0 | 0 | 0 | | | | | | |
| 0 | 0 | 0 | 0 | 0 | 0 | 0 | 0 | 0 | 0 | 0 | 0 | 0 | 0 | 0 | 0 | | | | | | |
| 0 | 0 | 1 | 0 | 0 | 0 | 0 | 0 | 0 | 0 | 1 | 0 | 1 | 1 | 0 | 1 | 0 | 0 | 0 | 0 | 0 | 0 |
| 1 | 1 | 1 | 1 | 0 | 0 | 0 | 0 | 0 | 0 | 0 | 0 | 0 | 0 | 0 | 0 | | | | | | |
| 0 | 0 | 1 | 0 | 1 | 0 | 1 | 1 | 0 | 1 | 0 | 0 | 0 | 0 | 0 | 0 | 0 | 0 | 0 | 0 | 0 | 0 |

| P5e8_both | P1e6_both_noHLA | P1e2_both_noHLA | P5e8_both_noHLA | P1e6_female | P1e2_female | P5e8_female | P1e6_female_noHLA | P1e2_female_noHLA | P5e8_female_noHLA | P1e6_male | P1e2_male | P5e8_male | P1e6_male_noHLA | P1e2_male_noHLA | P5e8_male_noHLA | n_in_embl_any | n_in_embl_gene | n_in_embl_any_or_gene | n_in_embl_any_noHLA | n_in_embl_gene_noHLA | n_in_embl_any_or_gene_noHLA |
|---|---|---|---|---|---|---|---|---|---|---|---|---|---|---|---|---|---|---|---|---|---|
| 0 | 0 | 0 | 0 | 0 | 0 | 0 | 0 | 0 | 0 | 0 | 0 | 0 | 0 | 0 | 0 | | | | | | |
| 0 | 0 | 0 | 0 | 5 | 0 | 5 | 0 | 0 | 0 | 1 | 1 | 1 | 1 | 1 | 1 | 3 | 6 | 6 | 0 | 1 | 1 |
| 0 | 0 | 0 | 0 | 0 | 0 | 0 | 0 | 0 | 0 | 0 | 0 | 0 | 0 | 0 | 0 | | | | | | |
| 8 | 3 | 11 | 0 | 12 | 0 | 19 | 7 | 0 | 10 | 1 | 0 | 3 | 1 | 0 | 1 | 7 | 17 | 18 | 1 | 4 | 4 |
| 0 | 0 | 0 | 0 | 0 | 0 | 0 | 0 | 0 | 0 | 0 | 0 | 0 | 0 | 0 | 0 | | | | | | |
| 0 | 0 | 0 | 0 | 1 | 1 | 1 | 1 | 1 | 1 | 0 | 0 | 0 | 0 | 0 | 0 | 0 | 0 | 0 | 0 | 0 | 0 |
| 0 | 0 | 0 | 0 | 1 | 1 | 1 | 1 | 1 | 1 | 0 | 0 | 0 | 0 | 0 | 0 | 0 | 0 | 0 | 0 | 0 | 0 |
| 0 | 0 | 2 | 0 | 0 | 0 | 0 | 0 | 0 | 0 | 2 | 0 | 2 | 2 | 0 | 2 | 3 | 2 | 3 | 3 | 2 | 3 |
| 69 | 84 | 129 | 66 | 50 | 4 | 65 | 46 | 4 | 58 | 6 | 1 | 12 | 4 | 1 | 10 | 50 | 95 | 104 | 49 | 92 | 100 |
| 37 | 42 | 70 | 32 | 35 | 4 | 46 | 29 | 4 | 37 | 5 | 0 | 9 | 3 | 0 | 5 | 29 | 53 | 61 | 28 | 53 | 60 |
| 0 | 0 | 0 | 0 | 0 | 0 | 0 | 0 | 0 | 0 | 0 | 0 | 0 | 0 | 0 | 0 | | | | | | |
| 0 | 0 | 0 | 0 | 0 | 0 | 0 | 0 | 0 | 0 | 0 | 0 | 0 | 0 | 0 | 0 | | | | | | |
| 0 | 0 | 0 | 0 | 0 | 0 | 0 | 0 | 0 | 0 | 0 | 0 | 0 | 0 | 0 | 0 | | | | | | |
| 0 | 0 | 0 | 0 | 0 | 0 | 0 | 0 | 0 | 0 | 0 | 0 | 0 | 0 | 0 | 0 | | | | | | |
| 0 | 0 | 0 | 0 | 0 | 0 | 0 | 0 | 0 | 0 | 0 | 0 | 0 | 0 | 0 | 0 | | | | | | |
| 0 | 0 | 0 | 0 | 0 | 0 | 0 | 0 | 0 | 0 | 0 | 0 | 0 | 0 | 0 | 0 | | | | | | |
| 0 | 0 | 6 | 0 | 5 | 1 | 5 | 5 | 1 | 5 | 3 | 1 | 3 | 3 | 1 | 3 | 4 | 4 | 5 | 4 | 4 | 5 |
| 0 | 0 | 0 | 0 | 0 | 0 | 0 | 0 | 0 | 0 | 1 | 1 | 1 | 1 | 1 | 1 | | | | | | |
| 0 | 0 | 0 | 0 | 0 | 0 | 0 | 0 | 0 | 0 | 0 | 0 | 0 | 0 | 0 | 0 | | | | | | |
| 0 | 0 | 1 | 0 | 1 | 0 | 1 | 1 | 0 | 1 | 1 | 1 | 1 | 1 | 1 | 1 | 1 | 0 | 1 | 1 | 0 | 1 |
| 2 | 2 | 2 | 2 | 0 | 0 | 0 | 0 | 0 | 0 | 0 | 0 | 0 | 0 | 0 | 0 | 0 | 0 | 0 | 0 | 0 | 0 |
| 7 | 8 | 12 | 7 | 4 | 0 | 5 | 4 | 0 | 5 | 0 | 0 | 0 | 0 | 0 | 0 | 10 | 12 | 12 | 10 | 12 | 12 |
| 0 | 0 | 0 | 0 | 0 | 0 | 0 | 0 | 0 | 0 | 0 | 0 | 0 | 0 | 0 | 0 | | | | | | |
| 13 | 19 | 41 | 13 | 23 | 3 | 27 | 23 | 3 | 27 | 9 | 3 | 11 | 4 | 2 | 6 | 26 | 48 | 49 | 25 | 47 | 48 |
| 9 | 15 | 22 | 9 | 11 | 4 | 17 | 11 | 4 | 17 | 0 | 0 | 0 | 0 | 0 | 0 | 14 | 28 | 29 | 14 | 28 | 29 |
| 0 | 0 | 0 | 0 | 0 | 0 | 0 | 0 | 0 | 0 | 0 | 0 | 0 | 0 | 0 | 0 | | | | | | |
| 0 | 0 | 0 | 0 | 0 | 0 | 0 | 0 | 0 | 0 | 0 | 0 | 0 | 0 | 0 | 0 | | | | | | |
| 34 | 40 | 62 | 34 | 26 | 4 | 29 | 25 | 4 | 28 | 3 | 2 | 6 | 3 | 2 | 6 | 23 | 65 | 70 | 23 | 64 | 69 |
| 64 | 89 | 138 | 64 | 58 | 8 | 78 | 53 | 8 | 73 | 4 | 0 | 9 | 4 | 0 | 9 | 59 | 144 | 146 | 57 | 142 | 144 |
| 0 | 0 | 0 | 0 | 0 | 0 | 0 | 0 | 0 | 0 | 0 | 0 | 0 | 0 | 0 | 0 | | | | | | |
| 0 | 0 | 0 | 0 | 0 | 0 | 0 | 0 | 0 | 0 | 0 | 0 | 0 | 0 | 0 | 0 | | | | | | |
| 0 | 0 | 0 | 0 | 2 | 2 | 2 | 2 | 2 | 2 | 0 | 0 | 0 | 0 | 0 | 0 | 1 | 0 | 1 | 1 | 0 | 1 |
| 0 | 0 | 0 | 0 | 0 | 0 | 0 | 0 | 0 | 0 | 0 | 0 | 0 | 0 | 0 | 0 | | | | | | |
| 0 | 0 | 0 | 0 | 0 | 0 | 0 | 0 | 0 | 0 | 0 | 0 | 0 | 0 | 0 | 0 | | | | | | |
| 0 | 0 | 0 | 0 | 0 | 0 | 0 | 0 | 0 | 0 | 0 | 0 | 0 | 0 | 0 | 0 | | | | | | |
| 3 | 3 | 6 | 3 | 1 | 0 | 1 | 1 | 0 | 1 | 3 | 1 | 3 | 3 | 1 | 3 | 5 | 9 | 9 | 5 | 9 | 9 |
| 0 | 0 | 0 | 0 | 1 | 1 | 1 | 1 | 1 | 1 | 0 | 0 | 0 | 0 | 0 | 0 | 0 | 0 | 0 | 0 | 0 | 0 |
| 1 | 1 | 3 | 1 | 1 | 0 | 1 | 1 | 0 | 1 | 1 | 0 | 1 | 1 | 0 | 1 | 2 | 3 | 3 | 2 | 3 | 3 |
| 0 | 0 | 0 | 0 | 0 | 0 | 0 | 0 | 0 | 0 | 0 | 0 | 0 | 0 | 0 | 0 | | | | | | |
| 0 | 0 | 0 | 0 | 0 | 0 | 0 | 0 | 0 | 0 | 0 | 0 | 0 | 0 | 0 | 0 | | | | | | |
| 0 | 0 | 0 | 0 | 0 | 0 | 0 | 0 | 0 | 0 | 0 | 0 | 0 | 0 | 0 | 0 | | | | | | |
| 0 | 0 | 0 | 0 | 0 | 0 | 0 | 0 | 0 | 0 | 0 | 0 | 0 | 0 | 0 | 0 | | | | | | |
| 0 | 0 | 0 | 0 | 0 | 0 | 0 | 0 | 0 | 0 | 0 | 0 | 0 | 0 | 0 | 0 | | | | | | |
| 0 | 0 | 0 | 0 | 0 | 0 | 0 | 0 | 0 | 0 | 0 | 0 | 0 | 0 | 0 | 0 | | | | | | |
| 2 | 2 | 3 | 2 | 1 | 0 | 1 | 1 | 0 | 1 | 0 | 0 | 0 | 0 | 0 | 0 | 2 | 5 | 5 | 2 | 5 | 5 |
| 3 | 3 | 5 | 3 | 2 | 0 | 2 | 2 | 0 | 2 | 0 | 0 | 0 | 0 | 0 | 0 | 3 | 7 | 7 | 3 | 7 | 7 |
| 0 | 0 | 0 | 0 | 0 | 0 | 0 | 0 | 0 | 0 | 0 | 0 | 0 | 0 | 0 | 0 | | | | | | |
| 0 | 0 | 0 | 0 | 0 | 0 | 0 | 0 | 0 | 0 | 0 | 0 | 0 | 0 | 0 | 0 | | | | | | |
| 0 | 0 | 0 | 0 | 1 | 1 | 1 | 1 | 1 | 1 | 0 | 0 | 0 | 0 | 0 | 0 | 0 | 0 | 0 | 0 | 0 | 0 |
| P5e8_both | P1e6_both_noHLA | P1e2_both_noHLA | P5e8_both_noHLA | P1e6_female | P1e2_female | P5e8_female | P1e6_female_noHLA | P1e2_female_noHLA | P5e8_female_noHLA | P1e6_male | P1e2_male | P5e8_male | P1e6_male_noHLA | P1e2_male_noHLA | P5e8_male_noHLA | n_in_embl_any | n_in_embl_gene | n_in_embl_any_or_gene | n_in_embl_any_noHLA | n_in_embl_gene_noHLA | n_in_embl_any_or_gene_noHLA |
| 0 | 0 | 0 | 0 | 0 | 0 | 0 | 0 | 0 | 0 | 0 | 0 | 0 | 0 | 0 | 0 | | | | | | |

| P5e8_both | P1e6_both_noHLA | P1e2_both_noHLA | P5e8_both_noHLA | P1e6_female | P1e2_female | P5e8_female | P1e6_female_noHLA | P1e2_female_noHLA | P5e8_female_noHLA | P1e6_male | P1e2_male | P5e8_male | P1e6_male_noHLA | P1e2_male_noHLA | P5e8_male_noHLA | n_in_embl_any | n_in_embl_gene | n_in_embl_any_or_gene | n_in_embl_any_noHLA | n_in_embl_gene_noHLA | n_in_embl_any_or_gene_noHLA |
|---|---|---|---|---|---|---|---|---|---|---|---|---|---|---|---|---|---|---|---|---|---|
| 0 | 0 | 0 | 0 | 0 | 0 | 0 | 0 | 0 | 0 | 0 | 0 | 0 | 0 | 0 | 0 | | | | | | |
| 0 | 0 | 0 | 0 | 0 | 0 | 0 | 0 | 0 | 0 | 0 | 0 | 0 | 0 | 0 | 0 | | | | | | |
| 0 | 0 | 0 | 0 | 0 | 0 | 0 | 0 | 0 | 0 | 0 | 0 | 0 | 0 | 0 | 0 | | | | | | |
| 0 | 0 | 0 | 0 | 0 | 0 | 0 | 0 | 0 | 0 | 0 | 0 | 0 | 0 | 0 | 0 | | | | | | |
| 0 | 0 | 0 | 0 | 0 | 0 | 0 | 0 | 0 | 0 | 0 | 0 | 0 | 0 | 0 | 0 | | | | | | |
| 0 | 0 | 0 | 0 | 0 | 0 | 0 | 0 | 0 | 0 | 0 | 0 | 0 | 0 | 0 | 0 | | | | | | |
| 0 | 0 | 0 | 0 | 0 | 0 | 0 | 0 | 0 | 0 | 0 | 0 | 0 | 0 | 0 | 0 | | | | | | |
| 4 | 5 | 6 | 4 | 1 | 0 | 2 | 1 | 0 | 2 | 0 | 0 | 0 | 0 | 0 | 0 | 3 | 6 | 6 | 3 | 6 | 6 |
| 0 | 0 | 0 | 0 | 0 | 0 | 0 | 0 | 0 | 0 | 0 | 0 | 0 | 0 | 0 | 0 | | | | | | |
| 0 | 0 | 0 | 0 | 0 | 0 | 0 | 0 | 0 | 0 | 0 | 0 | 0 | 0 | 0 | 0 | | | | | | |
| 0 | 0 | 0 | 0 | 1 | 1 | 1 | 1 | 1 | 1 | 0 | 0 | 0 | 0 | 0 | 0 | | | | | | |
| 0 | 0 | 0 | 0 | 1 | 1 | 1 | 1 | 1 | 1 | 0 | 0 | 0 | 0 | 0 | 0 | 0 | 0 | 0 | 0 | 0 | 0 |
| 0 | 0 | 0 | 0 | 0 | 0 | 0 | 0 | 0 | 0 | 0 | 0 | 0 | 0 | 0 | 0 | | | | | | |
| 0 | 1 | 4 | 0 | 3 | 0 | 4 | 3 | 0 | 4 | 0 | 0 | 0 | 0 | 0 | 0 | 3 | 4 | 4 | 3 | 4 | 4 |
| 0 | 0 | 0 | 0 | 0 | 0 | 0 | 0 | 0 | 0 | 0 | 0 | 0 | 0 | 0 | 0 | | | | | | |
| 0 | 0 | 0 | 0 | 0 | 0 | 0 | 0 | 0 | 0 | 0 | 0 | 0 | 0 | 0 | 0 | | | | | | |
| 0 | 0 | 0 | 0 | 0 | 0 | 0 | 0 | 0 | 0 | 0 | 0 | 0 | 0 | 0 | 0 | | | | | | |
| 0 | 0 | 0 | 0 | 0 | 0 | 0 | 0 | 0 | 0 | 0 | 0 | 0 | 0 | 0 | 0 | | | | | | |
| 0 | 0 | 0 | 0 | 0 | 0 | 0 | 0 | 0 | 0 | 0 | 0 | 0 | 0 | 0 | 0 | | | | | | |
| 0 | 0 | 0 | 0 | 0 | 0 | 0 | 0 | 0 | 0 | 0 | 0 | 0 | 0 | 0 | 0 | | | | | | |
| 0 | 0 | 0 | 0 | 0 | 0 | 0 | 0 | 0 | 0 | 0 | 0 | 0 | 0 | 0 | 0 | | | | | | |
| 0 | 0 | 0 | 0 | 0 | 0 | 0 | 0 | 0 | 0 | 0 | 0 | 0 | 0 | 0 | 0 | | | | | | |
| 0 | 0 | 0 | 0 | 0 | 0 | 0 | 0 | 0 | 0 | 0 | 0 | 0 | 0 | 0 | 0 | | | | | | |
| 0 | 0 | 0 | 0 | 0 | 0 | 0 | 0 | 0 | 0 | 0 | 0 | 0 | 0 | 0 | 0 | | | | | | |
| 0 | 0 | 0 | 0 | 0 | 0 | 0 | 0 | 0 | 0 | 0 | 0 | 0 | 0 | 0 | 0 | | | | | | |
| 0 | 0 | 0 | 0 | 0 | 0 | 0 | 0 | 0 | 0 | 0 | 0 | 0 | 0 | 0 | 0 | | | | | | |
| 0 | 0 | 0 | 0 | 0 | 0 | 0 | 0 | 0 | 0 | 0 | 0 | 0 | 0 | 0 | 0 | | | | | | |
| 0 | 0 | 0 | 0 | 0 | 0 | 0 | 0 | 0 | 0 | 0 | 0 | 0 | 0 | 0 | 0 | | | | | | |
| 0 | 0 | 0 | 0 | 0 | 0 | 0 | 0 | 0 | 0 | 0 | 0 | 0 | 0 | 0 | 0 | | | | | | |
| 10 | 12 | 35 | 10 | 25 | 2 | 27 | 25 | 2 | 27 | 0 | 0 | 0 | 0 | 0 | 0 | 25 | 39 | 39 | 25 | 39 | 39 |
| 0 | 0 | 0 | 0 | 0 | 0 | 0 | 0 | 0 | 0 | 0 | 0 | 0 | 0 | 0 | 0 | | | | | | |
| 0 | 0 | 0 | 0 | 0 | 0 | 0 | 0 | 0 | 0 | 0 | 0 | 0 | 0 | 0 | 0 | | | | | | |
| 0 | 0 | 0 | 0 | 0 | 0 | 0 | 0 | 0 | 0 | 0 | 0 | 0 | 0 | 0 | 0 | | | | | | |
| 0 | 0 | 0 | 0 | 0 | 0 | 0 | 0 | 0 | 0 | 0 | 0 | 0 | 0 | 0 | 0 | | | | | | |
| 0 | 1 | 3 | 0 | 1 | 0 | 1 | 1 | 0 | 1 | 1 | 0 | 2 | 1 | 0 | 2 | 2 | 3 | 3 | 2 | 3 | 3 |
| 15 | 21 | 34 | 15 | 14 | 2 | 19 | 13 | 1 | 18 | 1 | 0 | 2 | 1 | 0 | 2 | 14 | 38 | 40 | 14 | 38 | 40 |
| 0 | 0 | 0 | 0 | 0 | 0 | 0 | 0 | 0 | 0 | 0 | 0 | 0 | 0 | 0 | 0 | | | | | | |
| 0 | 0 | 0 | 0 | 0 | 0 | 0 | 0 | 0 | 0 | 0 | 0 | 0 | 0 | 0 | 0 | | | | | | |
| 0 | 0 | 0 | 0 | 0 | 0 | 0 | 0 | 0 | 0 | 0 | 0 | 0 | 0 | 0 | 0 | | | | | | |
| 0 | 0 | 0 | 0 | 0 | 0 | 0 | 0 | 0 | 0 | 0 | 0 | 0 | 0 | 0 | 0 | | | | | | |
| 0 | 0 | 0 | 0 | 0 | 0 | 0 | 0 | 0 | 0 | 0 | 0 | 0 | 0 | 0 | 0 | | | | | | |
| 0 | 0 | 0 | 0 | 0 | 0 | 0 | 0 | 0 | 0 | 0 | 0 | 0 | 0 | 0 | 0 | | | | | | |
| 0 | 0 | 0 | 0 | 0 | 0 | 0 | 0 | 0 | 0 | 0 | 0 | 0 | 0 | 0 | 0 | 3 | 2 | 3 | 3 | 2 | 3 |
| 2 | 4 | 4 | 2 | 0 | 0 | 1 | 0 | 0 | 1 | 0 | 0 | 1 | 0 | 0 | 1 | 2 | 5 | 5 | 2 | 5 | 5 |
| 12 | 13 | 21 | 12 | 8 | 0 | 8 | 8 | 0 | 8 | 0 | 0 | 1 | 0 | 0 | 1 | 6 | 17 | 18 | 6 | 17 | 18 |
| 0 | 0 | 0 | 0 | 0 | 0 | 0 | 0 | 0 | 0 | 0 | 0 | 0 | 0 | 0 | 0 | | | | | | |
| 0 | 0 | 0 | 0 | 0 | 0 | 0 | 0 | 0 | 0 | 0 | 0 | 0 | 0 | 0 | 0 | | | | | | |
| 1 | 2 | 3 | 1 | 2 | 1 | 3 | 2 | 1 | 3 | 0 | 0 | 0 | 0 | 0 | 0 | 2 | 4 | 4 | 2 | 4 | 4 |
| 0 | 0 | 1 | 0 | 2 | 1 | 2 | 2 | 1 | 2 | 0 | 0 | 0 | 0 | 0 | 0 | 2 | 2 | 2 | 2 | 2 | 2 |
| 0 | 0 | 0 | 0 | 0 | 0 | 0 | 0 | 0 | 0 | 0 | 0 | 0 | 0 | 0 | 0 | | | | | | |
| 0 | 0 | 0 | 0 | 0 | 0 | 0 | 0 | 0 | 0 | 0 | 0 | 0 | 0 | 0 | 0 | | | | | | |
| 0 | 0 | 0 | 0 | 2 | 2 | 2 | 2 | 2 | 2 | 0 | 0 | 0 | 0 | 0 | 0 | 1 | 1 | 2 | 1 | 1 | 2 |

| P5e8_both | P1e6_both_noHLA | P1e2_both_noHLA | P5e8_both_noHLA | P1e6_female | P1e2_female | P5e8_female | P1e6_female_noHLA | P1e2_female_noHLA | P5e8_female_noHLA | P1e6_male | P1e2_male | P5e8_male | P1e6_male_noHLA | P1e2_male_noHLA | P5e8_male_noHLA | n_in_embl_any | n_in_embl_gene | n_in_embl_any_or_gene | n_in_embl_any_noHLA | n_in_embl_gene_noHLA | n_in_embl_any_or_gene_noHLA |
|---|---|---|---|---|---|---|---|---|---|---|---|---|---|---|---|---|---|---|---|---|---|
| 0 | 0 | 0 | 0 | 0 | 0 | 0 | 0 | 0 | 0 | 0 | 0 | 0 | 0 | 0 | 0 | | | | | | |
| 4 | 6 | 26 | 4 | 22 | 4 | 23 | 22 | 4 | 23 | 3 | 1 | 4 | 3 | 1 | 4 | 8 | 31 | 32 | 8 | 31 | 32 |
| 0 | 0 | 1 | 0 | 1 | 0 | 1 | 1 | 0 | 1 | 0 | 0 | 0 | 0 | 0 | 0 | 1 | 1 | 1 | 1 | 1 | 1 |
| 7 | 10 | 17 | 7 | 8 | 1 | 11 | 8 | 1 | 11 | 0 | 0 | 0 | 0 | 0 | 0 | 11 | 17 | 19 | 11 | 17 | 19 |
| 1 | 6 | 9 | 1 | 3 | 0 | 8 | 3 | 0 | 8 | 1 | 1 | 1 | 1 | 1 | 1 | 6 | 11 | 11 | 6 | 11 | 11 |
| 0 | 0 | 1 | 0 | 0 | 0 | 0 | 0 | 0 | 0 | 1 | 0 | 1 | 1 | 0 | 1 | 1 | 1 | 1 | 1 | 1 | 1 |
| 0 | 0 | 0 | 0 | 1 | 1 | 1 | 1 | 1 | 1 | 0 | 0 | 0 | 0 | 0 | 0 | 0 | 0 | 0 | 0 | 0 | 0 |
| P5e8_both | P1e6_both_noHLA | P1e2_both_noHLA | P5e8_both_noHLA | P1e6_female | P1e2_female | P5e8_female | P1e6_female_noHLA | P1e2_female_noHLA | P5e8_female_noHLA | P1e6_male | P1e2_male | P5e8_male | P1e6_male_noHLA | P1e2_male_noHLA | P5e8_male_noHLA | n_in_embl_any | n_in_embl_gene | n_in_embl_any_or_gene | n_in_embl_any_noHLA | n_in_embl_gene_noHLA | n_in_embl_any_or_gene_noHLA |
| 1 | 1 | 2 | 1 | 1 | 0 | 1 | 1 | 0 | 1 | 0 | 0 | 0 | 0 | 0 | 0 | 2 | 3 | 3 | 2 | 3 | 3 |
| 1 | 1 | 2 | 1 | 1 | 0 | 1 | 1 | 0 | 1 | 0 | 0 | 0 | 0 | 0 | 0 | 2 | 2 | 2 | 2 | 2 | 2 |
| 0 | 0 | 0 | 0 | 0 | 0 | 0 | 0 | 0 | 0 | 0 | 0 | 0 | 0 | 0 | 0 | | | | | | |
| 0 | 0 | 0 | 0 | 0 | 0 | 0 | 0 | 0 | 0 | 0 | 0 | 0 | 0 | 0 | 0 | | | | | | |
| 0 | 0 | 0 | 0 | 0 | 0 | 0 | 0 | 0 | 0 | 0 | 0 | 0 | 0 | 0 | 0 | | | | | | |
| 0 | 0 | 0 | 0 | 0 | 0 | 0 | 0 | 0 | 0 | 0 | 0 | 0 | 0 | 0 | 0 | | | | | | |
| 0 | 0 | 0 | 0 | 0 | 0 | 0 | 0 | 0 | 0 | 0 | 0 | 0 | 0 | 0 | 0 | | | | | | |
| 0 | 0 | 0 | 0 | 0 | 0 | 0 | 0 | 0 | 0 | 0 | 0 | 0 | 0 | 0 | 0 | | | | | | |
| 0 | 0 | 0 | 0 | 0 | 0 | 0 | 0 | 0 | 0 | 0 | 0 | 0 | 0 | 0 | 0 | | | | | | |
| 0 | 0 | 0 | 0 | 0 | 0 | 0 | 0 | 0 | 0 | 0 | 0 | 0 | 0 | 0 | 0 | | | | | | |
| 0 | 0 | 0 | 0 | 0 | 0 | 0 | 0 | 0 | 0 | 0 | 0 | 0 | 0 | 0 | 0 | | | | | | |
| 0 | 0 | 0 | 0 | 0 | 0 | 0 | 0 | 0 | 0 | 0 | 0 | 0 | 0 | 0 | 0 | | | | | | |
| 0 | 0 | 0 | 0 | 0 | 0 | 0 | 0 | 0 | 0 | 0 | 0 | 0 | 0 | 0 | 0 | | | | | | |
| 0 | 0 | 0 | 0 | 0 | 0 | 0 | 0 | 0 | 0 | 0 | 0 | 0 | 0 | 0 | 0 | | | | | | |
| 0 | 0 | 0 | 0 | 0 | 0 | 0 | 0 | 0 | 0 | 0 | 0 | 0 | 0 | 0 | 0 | | | | | | |
| 0 | 0 | 0 | 0 | 0 | 0 | 0 | 0 | 0 | 0 | 0 | 0 | 0 | 0 | 0 | 0 | | | | | | |
| 0 | 0 | 0 | 0 | 0 | 0 | 0 | 0 | 0 | 0 | 2 | 2 | 2 | 2 | 2 | 2 | | | | | | |
| 0 | 0 | 0 | 0 | 0 | 0 | 0 | 0 | 0 | 0 | 0 | 0 | 0 | 0 | 0 | 0 | | | | | | |
| 0 | 0 | 0 | 0 | 0 | 0 | 0 | 0 | 0 | 0 | 0 | 0 | 0 | 0 | 0 | 0 | | | | | | |
| 0 | 0 | 0 | 0 | 0 | 0 | 0 | 0 | 0 | 0 | 0 | 0 | 0 | 0 | 0 | 0 | | | | | | |
| 0 | 0 | 0 | 0 | 0 | 0 | 0 | 0 | 0 | 0 | 0 | 0 | 0 | 0 | 0 | 0 | | | | | | |
| 0 | 0 | 0 | 0 | 1 | 1 | 1 | 1 | 1 | 1 | 0 | 0 | 0 | 0 | 0 | 0 | | | | | | |
| 0 | 0 | 0 | 0 | 0 | 0 | 0 | 0 | 0 | 0 | 0 | 0 | 0 | 0 | 0 | 0 | | | | | | |
| 0 | 0 | 0 | 0 | 0 | 0 | 0 | 0 | 0 | 0 | 0 | 0 | 0 | 0 | 0 | 0 | | | | | | |
| 3 | 5 | 17 | 3 | 12 | 4 | 14 | 12 | 4 | 14 | 5 | 1 | 5 | 5 | 1 | 5 | | | | | | |
| 3 | 5 | 17 | 3 | 12 | 4 | 14 | 12 | 4 | 14 | 6 | 2 | 6 | 6 | 2 | 6 | | | | | | |
| 3 | 5 | 17 | 3 | 12 | 4 | 14 | 12 | 4 | 14 | 6 | 2 | 6 | 6 | 2 | 6 | | | | | | |
| 0 | 0 | 0 | 0 | 0 | 0 | 0 | 0 | 0 | 0 | 0 | 0 | 0 | 0 | 0 | 0 | | | | | | |
| 0 | 0 | 0 | 0 | 0 | 0 | 0 | 0 | 0 | 0 | 0 | 0 | 0 | 0 | 0 | 0 | | | | | | |
| 0 | 0 | 0 | 0 | 0 | 0 | 0 | 0 | 0 | 0 | 1 | 1 | 1 | 1 | 1 | 1 | 0 | 0 | 0 | 0 | 0 | 0 |
| 1 | 1 | 2 | 1 | 0 | 0 | 0 | 0 | 0 | 0 | 1 | 0 | 1 | 1 | 0 | 1 | 1 | 1 | 1 | 1 | 1 | 1 |
| 5 | 8 | 20 | 5 | 12 | 1 | 15 | 12 | 1 | 15 | 1 | 0 | 1 | 1 | 0 | 1 | 13 | 15 | 19 | 13 | 15 | 19 |
| 0 | 0 | 0 | 0 | 0 | 0 | 0 | 0 | 0 | 0 | 0 | 0 | 0 | 0 | 0 | 0 | | | | | | |
| 0 | 0 | 0 | 0 | 0 | 0 | 0 | 0 | 0 | 0 | 0 | 0 | 0 | 0 | 0 | 0 | | | | | | |
| 8 | 19 | 41 | 8 | 26 | 4 | 34 | 25 | 3 | 33 | 0 | 0 | 3 | 0 | 0 | 3 | 22 | 36 | 39 | 22 | 36 | 39 |
| 6 | 7 | 18 | 6 | 13 | 3 | 14 | 13 | 3 | 14 | 2 | 1 | 2 | 2 | 1 | 2 | 11 | 18 | 19 | 11 | 18 | 19 |
| 0 | 0 | 0 | 0 | 0 | 0 | 0 | 0 | 0 | 0 | 0 | 0 | 0 | 0 | 0 | 0 | | | | | | |
| 0 | 0 | 0 | 0 | 0 | 0 | 0 | 0 | 0 | 0 | 0 | 0 | 0 | 0 | 0 | 0 | | | | | | |
| 0 | 0 | 0 | 0 | 0 | 0 | 0 | 0 | 0 | 0 | 0 | 0 | 0 | 0 | 0 | 0 | | | | | | |
| 0 | 0 | 0 | 0 | 0 | 0 | 0 | 0 | 0 | 0 | 0 | 0 | 0 | 0 | 0 | 0 | | | | | | |
| 0 | 0 | 0 | 0 | 0 | 0 | 0 | 0 | 0 | 0 | 0 | 0 | 0 | 0 | 0 | 0 | | | | | | |
| 0 | 0 | 0 | 0 | 0 | 0 | 0 | 0 | 0 | 0 | 0 | 0 | 0 | 0 | 0 | 0 | | | | | | |
| 0 | 0 | 0 | 0 | 0 | 0 | 0 | 0 | 0 | 0 | 0 | 0 | 0 | 0 | 0 | 0 | | | | | | |

| P5e8_both | P1e6_both_noHLA | P1e2_both_noHLA | P5e8_both_noHLA | P1e6_female | P1e2_female | P5e8_female | P1e6_female_noHLA | P1e2_female_noHLA | P5e8_female_noHLA | P1e6_male | P1e2_male | P5e8_male | P1e6_male_noHLA | P1e2_male_noHLA | P5e8_male_noHLA | n_in_embl_any | n_in_embl_gene | n_in_embl_any_or_gene | n_in_embl_any_noHLA | n_in_embl_gene_noHLA | n_in_embl_any_or_gene_noHLA |
|---|---|---|---|---|---|---|---|---|---|---|---|---|---|---|---|---|---|---|---|---|---|
| 0 | 0 | 0 | 0 | 1 | 1 | 1 | 1 | 1 | 1 | 0 | 0 | 0 | 0 | 0 | 0 | | | | | | |
| 0 | 0 | 0 | 0 | 0 | 0 | 0 | 0 | 0 | 0 | 0 | 0 | 0 | 0 | 0 | 0 | | | | | | |
| 0 | 0 | 0 | 0 | 0 | 0 | 0 | 0 | 0 | 0 | 0 | 0 | 0 | 0 | 0 | 0 | | | | | | |
| 0 | 0 | 0 | 0 | 0 | 0 | 0 | 0 | 0 | 0 | 0 | 0 | 0 | 0 | 0 | 0 | | | | | | |
| 0 | 0 | 0 | 0 | 0 | 0 | 0 | 0 | 0 | 0 | 0 | 0 | 0 | 0 | 0 | 0 | | | | | | |
| 0 | 0 | 0 | 0 | 0 | 0 | 0 | 0 | 0 | 0 | 0 | 0 | 0 | 0 | 0 | 0 | | | | | | |
| 0 | 0 | 0 | 0 | 0 | 0 | 0 | 0 | 0 | 0 | 0 | 0 | 0 | 0 | 0 | 0 | | | | | | |
| 0 | 0 | 0 | 0 | 0 | 0 | 0 | 0 | 0 | 0 | 0 | 0 | 0 | 0 | 0 | 0 | | | | | | |
| 0 | 0 | 0 | 0 | 0 | 0 | 0 | 0 | 0 | 0 | 0 | 0 | 0 | 0 | 0 | 0 | | | | | | |
| 0 | 0 | 0 | 0 | 0 | 0 | 0 | 0 | 0 | 0 | 0 | 0 | 0 | 0 | 0 | 0 | | | | | | |
| 0 | 0 | 0 | 0 | 0 | 0 | 0 | 0 | 0 | 0 | 0 | 0 | 0 | 0 | 0 | 0 | | | | | | |
| 0 | 0 | 0 | 0 | 0 | 0 | 0 | 0 | 0 | 0 | 0 | 0 | 0 | 0 | 0 | 0 | | | | | | |
| 0 | 0 | 0 | 0 | 0 | 0 | 0 | 0 | 0 | 0 | 0 | 0 | 0 | 0 | 0 | 0 | | | | | | |
| 0 | 0 | 0 | 0 | 0 | 0 | 0 | 0 | 0 | 0 | 0 | 0 | 0 | 0 | 0 | 0 | | | | | | |
| 2 | 3 | 4 | 2 | 1 | 0 | 2 | 1 | 0 | 2 | 0 | 0 | 0 | 0 | 0 | 0 | 5 | 5 | 5 | 5 | 5 | 5 |
| 0 | 0 | 0 | 0 | 0 | 0 | 0 | 0 | 0 | 0 | 0 | 0 | 0 | 0 | 0 | 0 | | | | | | |
| 0 | 0 | 0 | 0 | 0 | 0 | 0 | 0 | 0 | 0 | 0 | 0 | 0 | 0 | 0 | 0 | | | | | | |

| P5e8_both | P1e6_both_noHLA | P1e2_both_noHLA | P5e8_both_noHLA | P1e6_female | P1e2_female | P5e8_female | P1e6_female_noHLA | P1e2_female_noHLA | P5e8_female_noHLA | P1e6_male | P1e2_male | P5e8_male | P1e6_male_noHLA | P1e2_male_noHLA | P5e8_male_noHLA | n_in_embl_any | n_in_embl_gene | n_in_embl_any_or_gene | n_in_embl_any_noHLA | n_in_embl_gene_noHLA | n_in_embl_any_or_gene_noHLA |
|---|---|---|---|---|---|---|---|---|---|---|---|---|---|---|---|---|---|---|---|---|---|
| 2 | 3 | 4 | 2 | 2 | 1 | 2 | 2 | 1 | 2 | 0 | 0 | 1 | 0 | 0 | 1 | 3 | 5 | 5 | 3 | 5 | 5 |
| 1 | 1 | 1 | 1 | 0 | 0 | 0 | 0 | 0 | 0 | 0 | 0 | 0 | 0 | 0 | 0 | 1 | 1 | 1 | 1 | 1 | 1 |
| 0 | 0 | 0 | 0 | 1 | 1 | 1 | 0 | 0 | 0 | 0 | 0 | 0 | 0 | 0 | 0 | | | | | | |
| 0 | 0 | 0 | 0 | 0 | 0 | 0 | 0 | 0 | 0 | 0 | 0 | 0 | 0 | 0 | 0 | | | | | | |
| 0 | 0 | 0 | 0 | 0 | 0 | 0 | 0 | 0 | 0 | 0 | 0 | 0 | 0 | 0 | 0 | | | | | | |
| 0 | 0 | 0 | 0 | 0 | 0 | 0 | 0 | 0 | 0 | 0 | 0 | 0 | 0 | 0 | 0 | | | | | | |
| 0 | 0 | 0 | 0 | 0 | 0 | 0 | 0 | 0 | 0 | 0 | 0 | 0 | 0 | 0 | 0 | | | | | | |
| 0 | 0 | 0 | 0 | 0 | 0 | 0 | 0 | 0 | 0 | 0 | 0 | 0 | 0 | 0 | 0 | | | | | | |
| 0 | 0 | 0 | 0 | 0 | 0 | 0 | 0 | 0 | 0 | 0 | 0 | 0 | 0 | 0 | 0 | | | | | | |
| 0 | 0 | 0 | 0 | 0 | 0 | 0 | 0 | 0 | 0 | 0 | 0 | 0 | 0 | 0 | 0 | | | | | | |
| 0 | 0 | 0 | 0 | 0 | 0 | 0 | 0 | 0 | 0 | 1 | 1 | 1 | 1 | 1 | 1 | 0 | 0 | 0 | 0 | 0 | 0 |
| 0 | 0 | 0 | 0 | 0 | 0 | 0 | 0 | 0 | 0 | 0 | 0 | 0 | 0 | 0 | 0 | | | | | | |
| 0 | 0 | 0 | 0 | 0 | 0 | 0 | 0 | 0 | 0 | 0 | 0 | 0 | 0 | 0 | 0 | | | | | | |
| 0 | 0 | 0 | 0 | 0 | 0 | 0 | 0 | 0 | 0 | 0 | 0 | 0 | 0 | 0 | 0 | | | | | | |
| 0 | 0 | 1 | 0 | 2 | 1 | 2 | 2 | 1 | 2 | 0 | 0 | 0 | 0 | 0 | 0 | 2 | 2 | 2 | 2 | 2 | 2 |
| 0 | 0 | 0 | 0 | 0 | 0 | 0 | 0 | 0 | 0 | 0 | 0 | 0 | 0 | 0 | 0 | | | | | | |
| 0 | 0 | 0 | 0 | 0 | 0 | 0 | 0 | 0 | 0 | 0 | 0 | 0 | 0 | 0 | 0 | | | | | | |
| 0 | 0 | 0 | 0 | 0 | 0 | 0 | 0 | 0 | 0 | 0 | 0 | 0 | 0 | 0 | 0 | | | | | | |
| 0 | 0 | 0 | 0 | 0 | 0 | 0 | 0 | 0 | 0 | 0 | 0 | 0 | 0 | 0 | 0 | | | | | | |
| 29 | 35 | 47 | 29 | 12 | 1 | 18 | 12 | 1 | 18 | 1 | 0 | 1 | 1 | 0 | 1 | 23 | 56 | 56 | 23 | 56 | 56 |
| 0 | 0 | 3 | 0 | 2 | 0 | 2 | 2 | 0 | 2 | 1 | 0 | 1 | 1 | 0 | 1 | 2 | 1 | 2 | 2 | 1 | 2 |
| 0 | 0 | 0 | 0 | 0 | 0 | 0 | 0 | 0 | 0 | 0 | 0 | 0 | 0 | 0 | 0 | | | | | | |
| 0 | 0 | 0 | 0 | 0 | 0 | 0 | 0 | 0 | 0 | 0 | 0 | 0 | 0 | 0 | 0 | | | | | | |
| 0 | 0 | 1 | 0 | 3 | 2 | 3 | 3 | 2 | 3 | 0 | 0 | 0 | 0 | 0 | 0 | 2 | 2 | 2 | 2 | 2 | 2 |
| 0 | 0 | 0 | 0 | 0 | 0 | 0 | 0 | 0 | 0 | 0 | 0 | 0 | 0 | 0 | 0 | | | | | | |
| 0 | 0 | 0 | 0 | 0 | 0 | 0 | 0 | 0 | 0 | 0 | 0 | 0 | 0 | 0 | 0 | | | | | | |
| 0 | 0 | 0 | 0 | 0 | 0 | 0 | 0 | 0 | 0 | 0 | 0 | 0 | 0 | 0 | 0 | | | | | | |
| 0 | 0 | 0 | 0 | 0 | 0 | 0 | 0 | 0 | 0 | 0 | 0 | 0 | 0 | 0 | 0 | | | | | | |
| 0 | 0 | 0 | 0 | 0 | 0 | 0 | 0 | 0 | 0 | 0 | 0 | 0 | 0 | 0 | 0 | | | | | | |
| 0 | 0 | 0 | 0 | 0 | 0 | 0 | 0 | 0 | 0 | 0 | 0 | 0 | 0 | 0 | 0 | | | | | | |
| 0 | 0 | 1 | 0 | 0 | 0 | 0 | 0 | 0 | 0 | 4 | 3 | 4 | 4 | 3 | 4 | 1 | 5 | 5 | 1 | 5 | 5 |
| 0 | 0 | 0 | 0 | 0 | 0 | 0 | 0 | 0 | 0 | 0 | 0 | 0 | 0 | 0 | 0 | | | | | | |
| 0 | 0 | 0 | 0 | 0 | 0 | 0 | 0 | 0 | 0 | 0 | 0 | 0 | 0 | 0 | 0 | | | | | | |

| P5e8_both | P1e6_both_noHLA | P1e2_both_noHLA | P5e8_both_noHLA | P1e6_female | P1e2_female | P5e8_female | P1e6_female_noHLA | P1e2_female_noHLA | P5e8_female_noHLA | P1e6_male | P1e2_male | P5e8_male | P1e6_male_noHLA | P1e2_male_noHLA | P5e8_male_noHLA | n_in_embl_any | n_in_embl_gene | n_in_embl_any_or_gene | n_in_embl_any_noHLA | n_in_embl_gene_noHLA | n_in_embl_any_or_gene_noHLA |
|---|---|---|---|---|---|---|---|---|---|---|---|---|---|---|---|---|---|---|---|---|---|
| 0 | 0 | 0 | 0 | 0 | 0 | 0 | 0 | 0 | 0 | 0 | 0 | 0 | 0 | 0 | 0 | | | | | | |
| 0 | 0 | 0 | 0 | 0 | 0 | 0 | 0 | 0 | 0 | 0 | 0 | 0 | 0 | 0 | 0 | | | | | | |
| 0 | 0 | 0 | 0 | 0 | 0 | 0 | 0 | 0 | 0 | 0 | 0 | 0 | 0 | 0 | 0 | | | | | | |
| 0 | 0 | 0 | 0 | 0 | 0 | 0 | 0 | 0 | 0 | 0 | 0 | 0 | 0 | 0 | 0 | | | | | | |
| 0 | 0 | 0 | 0 | 0 | 0 | 0 | 0 | 0 | 0 | 0 | 0 | 0 | 0 | 0 | 0 | | | | | | |
| 0 | 0 | 0 | 0 | 0 | 0 | 0 | 0 | 0 | 0 | 17 | 4 | 17 | 0 | 0 | 0 | 0 | 2 | 2 | | | |
| 4 | 1 | 4 | 0 | 3 | 0 | 3 | 2 | 0 | 2 | 14 | 0 | 21 | 1 | 0 | 2 | 5 | 12 | 13 | 4 | 4 | 4 |
| 27 | 30 | 37 | 27 | 3 | 0 | 4 | 3 | 0 | 4 | 5 | 0 | 7 | 4 | 0 | 6 | 37 | 39 | 43 | 36 | 38 | 42 |
| 0 | 0 | 0 | 0 | 0 | 0 | 0 | 0 | 0 | 0 | 0 | 0 | 0 | 0 | 0 | 0 | | | | | | |
| 0 | 0 | 0 | 0 | 0 | 0 | 0 | 0 | 0 | 0 | 0 | 0 | 0 | 0 | 0 | 0 | | | | | | |
| 0 | 0 | 0 | 0 | 0 | 0 | 0 | 0 | 0 | 0 | 0 | 0 | 0 | 0 | 0 | 0 | | | | | | |
| 0 | 0 | 0 | 0 | 0 | 0 | 0 | 0 | 0 | 0 | 0 | 0 | 0 | 0 | 0 | 0 | | | | | | |
| 24 | 31 | 50 | 24 | 16 | 3 | 21 | 16 | 3 | 21 | 7 | 1 | 9 | 7 | 1 | 9 | 24 | 31 | 42 | 24 | 31 | 42 |
| 2 | 2 | 4 | 2 | 0 | 0 | 0 | 0 | 0 | 0 | 2 | 0 | 2 | 2 | 0 | 2 | 2 | 3 | 3 | 2 | 3 | 3 |
| 0 | 0 | 0 | 0 | 0 | 0 | 0 | 0 | 0 | 0 | 0 | 0 | 0 | 0 | 0 | 0 | | | | | | |
| 0 | 0 | 0 | 0 | 0 | 0 | 0 | 0 | 0 | 0 | 0 | 0 | 0 | 0 | 0 | 0 | | | | | | |
| 0 | 0 | 0 | 0 | 0 | 0 | 0 | 0 | 0 | 0 | 0 | 0 | 0 | 0 | 0 | 0 | | | | | | |
| 0 | 0 | 0 | 0 | 0 | 0 | 0 | 0 | 0 | 0 | 0 | 0 | 0 | 0 | 0 | 0 | | | | | | |
| 0 | 0 | 0 | 0 | 0 | 0 | 0 | 0 | 0 | 0 | 0 | 0 | 0 | 0 | 0 | 0 | | | | | | |
| 0 | 0 | 0 | 0 | 0 | 0 | 0 | 0 | 0 | 0 | 0 | 0 | 0 | 0 | 0 | 0 | | | | | | |
| 0 | 0 | 0 | 0 | 0 | 0 | 0 | 0 | 0 | 0 | 0 | 0 | 0 | 0 | 0 | 0 | | | | | | |
| 0 | 0 | 0 | 0 | 0 | 0 | 0 | 0 | 0 | 0 | 0 | 0 | 0 | 0 | 0 | 0 | | | | | | |
| 0 | 0 | 0 | 0 | 0 | 0 | 0 | 0 | 0 | 0 | 0 | 0 | 0 | 0 | 0 | 0 | | | | | | |
| 0 | 1 | 1 | 0 | 0 | 0 | 0 | 0 | 0 | 0 | 0 | 0 | 1 | 0 | 0 | 1 | 0 | 1 | 1 | 0 | 1 | 1 |
| 0 | 0 | 0 | 0 | 0 | 0 | 0 | 0 | 0 | 0 | 0 | 0 | 0 | 0 | 0 | 0 | | | | | | |
| 4 | 5 | 11 | 4 | 8 | 2 | 9 | 8 | 2 | 9 | 0 | 0 | 0 | 0 | 0 | 0 | 6 | 10 | 11 | 6 | 10 | 11 |
| 0 | 0 | 0 | 0 | 0 | 0 | 0 | 0 | 0 | 0 | 0 | 0 | 0 | 0 | 0 | 0 | | | | | | |
| P5e8_both | P1e6_both_noHLA | P1e2_both_noHLA | P5e8_both_noHLA | P1e6_female | P1e2_female | P5e8_female | P1e6_female_noHLA | P1e2_female_noHLA | P5e8_female_noHLA | P1e6_male | P1e2_male | P5e8_male | P1e6_male_noHLA | P1e2_male_noHLA | P5e8_male_noHLA | n_in_embl_any | n_in_embl_gene | n_in_embl_any_or_gene | n_in_embl_any_noHLA | n_in_embl_gene_noHLA | n_in_embl_any_or_gene_noHLA |
| 0 | 0 | 0 | 0 | 0 | 0 | 0 | 0 | 0 | 0 | 0 | 0 | 0 | 0 | 0 | 0 | | | | | | |
| 0 | 0 | 0 | 0 | 0 | 0 | 0 | 0 | 0 | 0 | 0 | 0 | 0 | 0 | 0 | 0 | | | | | | |
| 0 | 0 | 0 | 0 | 0 | 0 | 0 | 0 | 0 | 0 | 0 | 0 | 0 | 0 | 0 | 0 | | | | | | |
| 0 | 0 | 0 | 0 | 0 | 0 | 0 | 0 | 0 | 0 | 0 | 0 | 0 | 0 | 0 | 0 | | | | | | |
| 0 | 0 | 0 | 0 | 0 | 0 | 0 | 0 | 0 | 0 | 0 | 0 | 0 | 0 | 0 | 0 | | | | | | |
| 0 | 0 | 0 | 0 | 0 | 0 | 0 | 0 | 0 | 0 | 1 | 1 | 1 | 1 | 1 | 1 | 0 | 0 | 0 | 0 | 0 | 0 |
| 0 | 0 | 0 | 0 | 0 | 0 | 0 | 0 | 0 | 0 | 0 | 0 | 0 | 0 | 0 | 0 | | | | | | |
| 0 | 0 | 0 | 0 | 0 | 0 | 0 | 0 | 0 | 0 | 0 | 0 | 0 | 0 | 0 | 0 | | | | | | |
| 0 | 0 | 0 | 0 | 0 | 0 | 0 | 0 | 0 | 0 | 0 | 0 | 0 | 0 | 0 | 0 | | | | | | |
| 0 | 0 | 0 | 0 | 0 | 0 | 0 | 0 | 0 | 0 | 0 | 0 | 0 | 0 | 0 | 0 | | | | | | |
| 0 | 0 | 0 | 0 | 0 | 0 | 0 | 0 | 0 | 0 | 0 | 0 | 0 | 0 | 0 | 0 | | | | | | |
| 0 | 0 | 0 | 0 | 0 | 0 | 0 | 0 | 0 | 0 | 0 | 0 | 0 | 0 | 0 | 0 | | | | | | |
| 0 | 0 | 0 | 0 | 0 | 0 | 0 | 0 | 0 | 0 | 0 | 0 | 0 | 0 | 0 | 0 | | | | | | |
| 0 | 0 | 0 | 0 | 0 | 0 | 0 | 0 | 0 | 0 | 0 | 0 | 0 | 0 | 0 | 0 | | | | | | |
| 0 | 0 | 0 | 0 | 0 | 0 | 0 | 0 | 0 | 0 | 0 | 0 | 0 | 0 | 0 | 0 | | | | | | |
| 0 | 0 | 0 | 0 | 1 | 1 | 1 | 1 | 1 | 1 | 0 | 0 | 0 | 0 | 0 | 0 | 0 | 0 | 0 | 0 | 0 | 0 |
| 2 | 3 | 6 | 2 | 1 | 0 | 1 | 1 | 0 | 1 | 2 | 0 | 3 | 2 | 0 | 3 | 4 | 5 | 5 | 4 | 5 | 5 |
| 0 | 0 | 0 | 0 | 0 | 0 | 0 | 0 | 0 | 0 | 0 | 0 | 0 | 0 | 0 | 0 | | | | | | |
| 0 | 0 | 0 | 0 | 0 | 0 | 0 | 0 | 0 | 0 | 0 | 0 | 0 | 0 | 0 | 0 | | | | | | |
| 0 | 0 | 0 | 0 | 0 | 0 | 0 | 0 | 0 | 0 | 0 | 0 | 0 | 0 | 0 | 0 | | | | | | |
| 7 | 6 | 14 | 3 | 9 | 2 | 15 | 2 | 1 | 4 | 10 | 2 | 12 | 9 | 2 | 10 | 15 | 24 | 27 | 8 | 11 | 13 |
| 0 | 0 | 0 | 0 | 0 | 0 | 0 | 0 | 0 | 0 | 0 | 0 | 0 | 0 | 0 | 0 | | | | | | |
| 0 | 0 | 0 | 0 | 0 | 0 | 0 | 0 | 0 | 0 | 1 | 1 | 1 | 1 | 1 | 1 | 1 | 1 | 1 | 1 | 1 | 1 |

| P5e8_both | P1e6_both_noHLA | P1e2_both_noHLA | P5e8_both_noHLA | P1e6_female | P1e2_female | P5e8_female | P1e6_female_noHLA | P1e2_female_noHLA | P5e8_female_noHLA | P1e6_male | P1e2_male | P5e8_male | P1e6_male_noHLA | P1e2_male_noHLA | P5e8_male_noHLA | n_in_embl_any | n_in_embl_gene | n_in_embl_any_or_gene | n_in_embl_any_noHLA | n_in_embl_gene_noHLA | n_in_embl_any_or_gene_noHLA |
|---|---|---|---|---|---|---|---|---|---|---|---|---|---|---|---|---|---|---|---|---|---|
| 0 | 0 | 0 | 0 | 0 | 0 | 0 | 0 | 0 | 0 | 0 | 0 | 0 | 0 | 0 | 0 | | | | | | |
| 0 | 0 | 0 | 0 | 0 | 0 | 0 | 0 | 0 | 0 | 0 | 0 | 0 | 0 | 0 | 0 | | | | | | |
| 1 | 1 | 2 | 1 | 1 | 0 | 1 | 1 | 0 | 1 | 0 | 0 | 0 | 0 | 0 | 0 | 1 | 2 | 2 | 1 | 2 | 2 |
| 3 | 5 | 14 | 3 | 11 | 2 | 13 | 11 | 2 | 13 | 1 | 1 | 1 | 1 | 1 | 1 | 12 | 13 | 15 | 12 | 13 | 15 |
| 0 | 0 | 0 | 0 | 0 | 0 | 0 | 0 | 0 | 0 | 0 | 0 | 0 | 0 | 0 | 0 | | | | | | |

#### STable4: Unique loci identified as associated with per-person median sclinical quantitative trait values using plink

2010 rows × 18 columns

| pheno | CHR | Gene_Symbol | biotype | lead_snp | lead_BP | lead_P | TOTAL | NSIG | S05 | S01 | S001 | S0001 | independent_loci_in_sex_P1e6 | independent_loci_in_sex_P1e2 | independent_loci_in_sex_P5e8 | in_gwas_catalog | in_HLA |
|---|---|---|---|---|---|---|---|---|---|---|---|---|---|---|---|---|---|
| A.2Gg_ | 16 | PMFBP1 | protein_coding | rs12445401 | 72148419 | 1.07e-10 | 25 | 0 | 0 | 0 | 10 | 15 | Male only | Both sexes | Male only | False | False |
| A.2Gg_ | 16 | PKD1L3 | processed_transcript | rs11645582 | 72029008 | 4.10e-09 | 41 | 0 | 0 | 7 | 13 | 21 | Male only | Both sexes | Male only | False | False |
| A1Anti | 14 | SERPINA2P | pseudogene | rs926144 | 94813402 | 1.46e-14 | 11 | 3 | 3 | 0 | 0 | 5 | Both sexes | Both sexes | Both sexes | True | False |
| A1Anti | 14 | SERPINA1 | protein_coding | rs55862705 | 94866482 | 6.52e-12 | 5 | 0 | 0 | 0 | 0 | 5 | Both sexes | Both sexes | Both sexes | True | False |
| ACTH | 3 | EPHB1 | protein_coding | rs9851564 | 134924786 | 2.65e-09 | 11 | 6 | 0 | 0 | 0 | 5 | Male only | Male only | Male only | True | False |
| AN.GAP | 2 | GCKR | protein_coding | rs780094 | 27741237 | 7.50e-14 | 23 | 0 | 0 | 0 | 1 | 22 | Both sexes | Both sexes | Both sexes | True | False |
| AN.GAP | 2 | ZNF512 | protein_coding | rs3749147 | 27851918 | 2.55e-09 | 71 | 0 | 6 | 5 | 3 | 57 | Male only | Both sexes | Male only | True | False |
| AN.GAP | 2 | MRPL33 | protein_coding | rs12104449 | 27972833 | 1.43e-08 | 5 | 0 | 1 | 2 | 0 | 2 | Female only | Both sexes | Female only | True | False |
| AN.GAP | 13 | COL4A2 | protein_coding | rs2172724 | 110961409 | 1.98e-08 | 3 | 2 | 0 | 0 | 0 | 1 | Female only | Female only | Female only | True | False |
| Alb | 6 | ZDHHC20P2 | pseudogene | rs2596570 | 31348040 | 1.65e-09 | 241 | 0 | 0 | 19 | 27 | 195 | Both sexes | Both sexes | Male only | True | True |
| Alb | 6 | HLA-C | protein_coding | rs2853953 | 31235505 | 2.21e-09 | 167 | 7 | 0 | 15 | 71 | 74 | Male only | Both sexes | Male only | True | True |
| Alb | 2 | PPM1G | protein_coding | rs1728918 | 27635463 | 7.88e-09 | 7 | 0 | 0 | 0 | 1 | 6 | Male only | Both sexes | Male only | True | False |
| Alb | 6 | HLA-C | protein_coding | rs9264185 | 31219982 | 3.50e-08 | 33 | 0 | 0 | 12 | 4 | 17 | Male only | Both sexes | Male only | True | True |
| AlkP | 9 | ABO | processed_transcript | rs550057 | 136146597 | 6.16e-93 | 69 | 0 | 0 | 0 | 0 | 69 | Both sexes | Both sexes | Both sexes | True | False |
| AlkP | 9 | ABO | processed_transcript | rs11244061 | 136153981 | 7.23e-47 | 3 | 0 | 0 | 0 | 0 | 3 | Both sexes | Both sexes | Both sexes | True | False |
| AlkP | 9 | ABO | processed_transcript | rs630510 | 136149581 | 3.24e-34 | 7 | 0 | 0 | 0 | 0 | 7 | Both sexes | Both sexes | Both sexes | True | False |
| AlkP | 9 | ABO | processed_transcript | rs633862 | 136155444 | 2.12e-29 | 31 | 0 | 0 | 0 | 0 | 31 | Both sexes | Both sexes | Both sexes | True | False |
| AlkP | 1 | NBPF3 | protein_coding | rs1976403 | 21766453 | 5.10e-27 | 41 | 0 | 0 | 0 | 0 | 41 | Both sexes | Both sexes | Both sexes | True | False |
| AlkP | 9 | C9orf96 | protein_coding | rs41302673 | 136270538 | 2.17e-24 | 9 | 0 | 0 | 0 | 0 | 9 | Both sexes | Both sexes | Both sexes | True | False |
| AlkP | 9 | ABO | processed_transcript | rs2073826 | 136136963 | 6.38e-22 | 25 | 0 | 0 | 0 | 0 | 25 | Both sexes | Both sexes | Both sexes | True | False |
| AlkP | 9 | ABO | processed_transcript | rs7873416 | 136131846 | 1.11e-21 | 145 | 0 | 0 | 0 | 0 | 145 | Both sexes | Both sexes | Both sexes | True | False |
| AlkP | 1 | NBPF3 | protein_coding | rs76372215 | 21818173 | 3.91e-20 | 9 | 0 | 0 | 1 | 0 | 8 | Both sexes | Both sexes | Both sexes | True | False |
| AlkP | 1 | ALPL | protein_coding | rs145377039 | 21863467 | 5.82e-20 | 21 | 0 | 0 | 0 | 0 | 21 | Both sexes | Both sexes | Both sexes | True | False |
| AlkP | 1 | AL359815.1 | miRNA | rs72660309 | 21919370 | 1.11e-18 | 13 | 0 | 0 | 0 | 0 | 13 | Both sexes | Both sexes | Both sexes | True | False |

| pheno | CHR | Gene_Symbol | biotype | lead_snp | lead_BP | lead_P | TOTAL | NSIG | S05 | S01 | S001 | S0001 | independent_loci_in_sex_P1e6 | independent_loci_in_sex_P1e2 | independent_loci_in_sex_P5e8 | in_gwas_catalog | in_HLA |
|---|---|---|---|---|---|---|---|---|---|---|---|---|---|---|---|---|---|
| AlkP | 1 | ALPL | protein_coding | rs1780329 | 21902950 | 8.00e-18 | 73 | 0 | 1 | 0 | 0 | 72 | Both sexes | Both sexes | Both sexes | True | False |
| AlkP | 6 | GPLD1 | protein_coding | rs9295625 | 24474278 | 1.01e-17 | 103 | 0 | 0 | 2 | 2 | 99 | Both sexes | Both sexes | Both sexes | True | False |
| AlkP | 19 | FUT2 | protein_coding | rs485073 | 49207255 | 3.09e-17 | 63 | 0 | 0 | 0 | 0 | 63 | Both sexes | Both sexes | Both sexes | True | False |
| AlkP | 9 | ABO | processed_transcript | rs176694 | 136164593 | 7.31e-17 | 3 | 0 | 0 | 0 | 0 | 3 | Both sexes | Both sexes | Both sexes | True | False |
| AlkP | 6 | GPLD1 | protein_coding | rs1883415 | 24491475 | 2.10e-16 | 115 | 1 | 1 | 0 | 9 | 104 | Both sexes | Both sexes | Both sexes | True | False |
| AlkP | 6 | GPLD1 | protein_coding | rs6922313 | 24438719 | 3.03e-16 | 75 | 0 | 0 | 0 | 0 | 75 | Both sexes | Both sexes | Both sexes | True | False |
| AlkP | 1 | NBPF3 | protein_coding | rs1697421 | 21823292 | 3.37e-16 | 3 | 0 | 0 | 0 | 0 | 3 | Both sexes | Both sexes | Both sexes | True | False |
| AlkP | 9 | GBGT1 | protein_coding | rs2886307 | 136050340 | 5.57e-16 | 35 | 0 | 0 | 0 | 1 | 34 | Both sexes | Both sexes | Both sexes | True | False |
| AlkP | 1 | ALPL | protein_coding | rs1256342 | 21876492 | 6.30e-16 | 15 | 0 | 0 | 0 | 0 | 15 | Both sexes | Both sexes | Both sexes | True | False |
| AlkP | 1 | NBPF3 | protein_coding | rs6680125 | 21814390 | 1.26e-15 | 79 | 0 | 0 | 0 | 0 | 79 | Both sexes | Both sexes | Both sexes | True | False |
| AlkP | 6 | GPLD1 | protein_coding | rs793671 | 24438662 | 3.01e-15 | 17 | 0 | 0 | 1 | 3 | 13 | Both sexes | Both sexes | Both sexes | True | False |
| AlkP | 1 | ALPL | protein_coding | rs34605986 | 21904131 | 6.92e-15 | 23 | 0 | 0 | 0 | 0 | 23 | Both sexes | Both sexes | Both sexes | True | False |
| AlkP | 1 | ECE1 | protein_coding | rs7530820 | 21685431 | 1.44e-14 | 125 | 0 | 0 | 26 | 15 | 84 | Female only | Both sexes | Female only | True | False |
| AlkP | 6 | MRS2 | protein_coding | rs62401887 | 24416482 | 6.76e-14 | 7 | 0 | 0 | 0 | 0 | 7 | Both sexes | Both sexes | Both sexes | True | False |
| AlkP | 1 | ALPL | protein_coding | 1:21889143 | 21889143 | 4.84e-12 | 27 | 0 | 17 | 0 | 0 | 10 | Female only | Both sexes | Female only | True | False |
| AlkP | 1 | ECE1 | protein_coding | rs3107384 | 21690627 | 6.19e-12 | 19 | 1 | 3 | 5 | 0 | 10 | Female only | Both sexes | Female only | True | False |
| AlkP | 1 | ALPL | protein_coding | rs1536934 | 21901277 | 8.10e-12 | 11 | 0 | 1 | 1 | 4 | 5 | Both sexes | Both sexes | Both sexes | True | False |
| AlkP | 9 | SLC2A6 | protein_coding | rs7875786 | 136353663 | 4.37e-11 | 13 | 0 | 0 | 0 | 0 | 13 | Both sexes | Both sexes | Both sexes | True | False |
| AlkP | 19 | RASIP1 | protein_coding | rs546308 | 49223570 | 4.98e-11 | 13 | 0 | 0 | 0 | 3 | 10 | Both sexes | Both sexes | Male only | True | False |
| AlkP | 1 | ALPL | protein_coding | 1:21895061 | 21895061 | 1.30e-10 | 7 | 2 | 0 | 2 | 0 | 3 | Male only | Both sexes | Male only | True | False |
| AlkP | 9 | LCN1P1 | pseudogene | rs886082 | 136101999 | 1.47e-10 | 31 | 16 | 0 | 0 | 0 | 15 | Female only | Female only | Female only | True | False |
| AlkP | 1 | RAP1GAP | protein_coding | rs12759525 | 21927848 | 1.59e-10 | 3 | 0 | 0 | 0 | 0 | 3 | Both sexes | Both sexes | Male only | True | False |
| AlkP | 9 | GBGT1 | protein_coding | rs10751500 | 136057106 | 3.07e-10 | 9 | 0 | 3 | 0 | 3 | 3 | Female only | Both sexes | Female only | True | False |
| AlkP | 17 | ASGR1 | protein_coding | rs55714927 | 7080316 | 5.62e-10 | 21 | 0 | 0 | 0 | 0 | 21 | Both sexes | Both sexes | Female only | True | False |
| AlkP | 1 | RAP1GAP | protein_coding | rs79714367 | 21942971 | 9.44e-10 | 5 | 0 | 0 | 0 | 0 | 5 | Both sexes | Both sexes | Female only | True | False |
| AlkP | 9 | ABO | processed_transcript | rs8176630 | 136152722 | 9.58e-10 | 3 | 0 | 0 | 0 | 0 | 3 | Both sexes | Both sexes | Female only | True | False |

| pheno | CHR | Gene_Symbol | biotype | lead_snp | lead_BP | lead_P | TOTAL | NSIG | S05 | S01 | S001 | S0001 | independent_loci_in_sex_P1e6 | independent_loci_in_sex_P1e2 | independent_loci_in_sex_P5e8 | in_gwas_catalog | in_HLA |
|---|---|---|---|---|---|---|---|---|---|---|---|---|---|---|---|---|---|
| AlkP | 1 | NBPF3 | protein_coding | rs4430298 | 21785540 | 1.37e-09 | 5 | 0 | 0 | 0 | 3 | 2 | Female only | Both sexes | Female only | True | False |
| AlkP | 9 | LCN1P2 | pseudogene | rs623179 | 136174678 | 1.41e-09 | 3 | 0 | 0 | 0 | 0 | 3 | Both sexes | Both sexes | Female only | True | False |
| AlkP | 9 | SURF6 | protein_coding | rs17150482 | 136194595 | 1.75e-09 | 23 | 14 | 0 | 0 | 3 | 6 | Female only | Both sexes | Female only | True | False |
| AlkP | 14 | NYNRIN | protein_coding | rs11620783 | 24871530 | 1.95e-09 | 11 | 5 | 1 | 0 | 0 | 5 | Female only | Female only | Female only | True | False |
| AlkP | 6 | DCDC2 | protein_coding | rs3857542 | 24309501 | 2.01e-09 | 81 | 2 | 0 | 9 | 30 | 40 | Male only | Both sexes | Male only | True | False |
| AlkP | 10 | REEP3 | protein_coding | rs10995554 | 65295110 | 3.60e-09 | 177 | 0 | 35 | 20 | 14 | 108 | Both sexes | Both sexes | Female only | True | False |
| AlkP | 9 | LCN1P2 | pseudogene | rs1179011 | 136186383 | 4.06e-09 | 5 | 2 | 0 | 2 | 0 | 1 | Male only | Both sexes | Male only | True | False |
| AlkP | 9 | OBP2B | protein_coding | rs11244034 | 136080272 | 6.16e-09 | 9 | 5 | 0 | 0 | 0 | 4 | Female only | Female only | Female only | True | False |
| AlkP | 20 | GINS1 | protein_coding | rs761025 | 25426968 | 7.28e-09 | 689 | 0 | 0 | 25 | 68 | 596 | Female only | Both sexes | Female only | True | False |
| AlkP | 10 | JMJD1C | protein_coding | rs4454603 | 65012750 | 8.13e-09 | 141 | 0 | 0 | 0 | 1 | 140 | Both sexes | Both sexes | Female only | True | False |
| pheno | CHR | Gene_Symbol | biotype | lead_snp | lead_BP | lead_P | TOTAL | NSIG | S05 | S01 | S001 | S0001 | independent_loci_in_sex_P1e6 | independent_loci_in_sex_P1e2 | independent_loci_in_sex_P5e8 | in_gwas_catalog | in_HLA |
| AlkP | 8 | RP11-115J16.1 | lincRNA | rs11774915 | 9188762 | 1.22e-08 | 31 | 14 | 9 | 2 | 0 | 6 | Male only | Both sexes | Male only | True | False |
| AlkP | 8 | RP11-115J16.1 | lincRNA | rs4841132 | 9183596 | 1.24e-08 | 63 | 0 | 0 | 0 | 2 | 61 | Both sexes | Both sexes | Both sexes | True | False |
| AlkP | 10 | REEP3 | protein_coding | rs10740136 | 65329622 | 1.31e-08 | 37 | 0 | 0 | 2 | 0 | 35 | Both sexes | Both sexes | Female only | True | False |
| AlkP | 6 | KIAA0319 | protein_coding | rs807526 | 24543720 | 1.51e-08 | 13 | 0 | 1 | 1 | 2 | 9 | Both sexes | Both sexes | Female only | True | False |
| AlkP | 6 | GPLD1 | protein_coding | rs16901722 | 24434711 | 1.94e-08 | 167 | 1 | 2 | 71 | 19 | 74 | Female only | Both sexes | Female only | True | False |
| AlkP | 8 | RP11-115J16.1 | lincRNA | rs878336 | 9225629 | 3.01e-08 | 17 | 0 | 2 | 8 | 1 | 6 | Male only | Both sexes | Male only | True | False |
| AlkP | 1 | RAP1GAP | protein_coding | rs11591167 | 21958536 | 3.04e-08 | 19 | 1 | 8 | 4 | 2 | 4 | Female only | Both sexes | Female only | True | False |
| AlkP | 9 | ABO | processed_transcript | rs3205136 | 136126631 | 3.60e-08 | 83 | 42 | 12 | 0 | 14 | 15 | Female only | Female only | Female only | True | False |
| AlkP | 1 | RAP1GAP | protein_coding | rs72660313 | 21926600 | 3.94e-08 | 3 | 1 | 0 | 1 | 0 | 1 | Male only | Both sexes | Male only | True | False |
| AlkP | 1 | RAP1GAP | protein_coding | rs1531829 | 21953304 | 4.41e-08 | 7 | 0 | 1 | 5 | 0 | 1 | Both sexes | Both sexes | Female only | True | False |
| Amyl | 1 | AMY1C | protein_coding | rs12080068 | 104329823 | 2.44e-13 | 3 | 0 | 0 | 0 | 0 | 3 | Both sexes | Both sexes | Both sexes | True | False |
| Amyl | 1 | RNPC3 | protein_coding | rs147499264 | 104082019 | 1.39e-10 | 11 | 0 | 0 | 1 | 2 | 8 | Female only | Both sexes | Female only | True | False |
| BASOAB | 19 | CTD-2540B15.12 | lincRNA | rs78744187 | 33754548 | 3.48e-114 | 5 | 0 | 0 | 0 | 0 | 5 | Both sexes | Both sexes | Both sexes | True | False |
| BASOAB | 3 | RPN1 | protein_coding | rs6782812 | 128317997 | 1.45e-47 | 53 | 0 | 0 | 0 | 0 | 53 | Both sexes | Both sexes | Both sexes | True | False |
| BASOAB | 19 | CTD-2540B15.12 | lincRNA | rs67228209 | 33751033 | 5.61e-32 | 3 | 0 | 0 | 0 | 0 | 3 | Both sexes | Both sexes | Both sexes | True | False |

| pheno | CHR | Gene_Symbol | biotype | lead_snp | lead_BP | lead_P | TOTAL | NSIG | S05 | S01 | S001 | S0001 | independent_loci_in_sex_P1e6 | independent_loci_in_sex_P1e2 | independent_loci_in_sex_P5e8 | in_gwas_catalog | in_HLA |
|---|---|---|---|---|---|---|---|---|---|---|---|---|---|---|---|---|---|
| BASOAB | 3 | C3orf27 | protein_coding | rs4857906 | 128275459 | 1.58e-23 | 103 | 0 | 0 | 0 | 0 | 103 | Both sexes | Both sexes | Both sexes | True | False |
| BASOAB | 19 | CTD-2540B15.12 | lincRNA | rs12461296 | 33774591 | 1.20e-21 | 3 | 0 | 0 | 0 | 0 | 3 | Both sexes | Both sexes | Both sexes | True | False |
| BASOAB | 19 | CTD-2540B15.12 | lincRNA | rs4432367 | 33752920 | 1.68e-21 | 3 | 0 | 0 | 0 | 0 | 3 | Both sexes | Both sexes | Both sexes | True | False |
| BASOAB | 14 | CEBPE | protein_coding | rs8015478 | 23586018 | 3.92e-18 | 7 | 0 | 0 | 0 | 0 | 7 | Both sexes | Both sexes | Both sexes | True | False |
| BASOAB | 19 | CTD-2540B15.12 | lincRNA | rs10414008 | 33758327 | 1.36e-17 | 5 | 0 | 1 | 0 | 0 | 4 | Both sexes | Both sexes | Both sexes | True | False |
| BASOAB | 3 | RAB7A | protein_coding | rs35904610 | 128452108 | 1.56e-16 | 115 | 0 | 0 | 0 | 0 | 115 | Both sexes | Both sexes | Both sexes | True | False |
| BASOAB | 3 | C3orf27 | protein_coding | rs72987210 | 128283406 | 5.27e-14 | 37 | 0 | 0 | 0 | 0 | 37 | Both sexes | Both sexes | Both sexes | True | False |
| BASOAB | 3 | RAB7A | protein_coding | rs35151839 | 128546891 | 8.87e-14 | 45 | 0 | 0 | 0 | 0 | 45 | Both sexes | Both sexes | Female only | True | False |
| BASOAB | 1 | LMO4 | protein_coding | rs1888342 | 87751406 | 1.36e-13 | 35 | 0 | 0 | 0 | 0 | 35 | Both sexes | Both sexes | Both sexes | True | False |
| BASOAB | 20 | C20orf203 | lincRNA | rs6057620 | 31265165 | 5.62e-13 | 125 | 0 | 0 | 0 | 0 | 125 | Both sexes | Both sexes | Female only | True | False |
| BASOAB | 6 | HCP5 | sense_overlapping | rs2596528 | 31385485 | 5.81e-13 | 43 | 0 | 0 | 0 | 1 | 42 | Both sexes | Both sexes | Both sexes | True | True |
| BASOAB | 3 | CRBN | protein_coding | rs1669340 | 3198380 | 9.92e-13 | 41 | 0 | 0 | 3 | 1 | 37 | Both sexes | Both sexes | Both sexes | True | False |
| BASOAB | 20 | C20orf203 | lincRNA | rs6057610 | 31241389 | 2.97e-12 | 31 | 9 | 0 | 2 | 12 | 8 | Female only | Both sexes | Female only | True | False |
| BASOAB | 4 | AFF1 | protein_coding | rs236987 | 88009779 | 1.05e-11 | 73 | 12 | 25 | 0 | 0 | 36 | Female only | Female only | Female only | True | False |
| BASOAB | 1 | LMO4 | protein_coding | rs4655844 | 87775058 | 1.64e-11 | 17 | 0 | 0 | 0 | 0 | 17 | Both sexes | Both sexes | Both sexes | True | False |
| BASOAB | 19 | CTD-2540B15.12 | lincRNA | rs34610286 | 33754671 | 2.40e-11 | 7 | 0 | 0 | 0 | 0 | 7 | Both sexes | Both sexes | Male only | True | False |
| BASOAB | 3 | RPN1 | protein_coding | rs2712404 | 128332679 | 4.07e-11 | 169 | 31 | 1 | 0 | 0 | 137 | Both sexes | Both sexes | Both sexes | True | False |
| BASOAB | 3 | RAB7A | protein_coding | rs56373544 | 128523503 | 9.94e-11 | 25 | 0 | 0 | 0 | 3 | 22 | Female only | Both sexes | Female only | True | False |
| BASOAB | 19 | CTD-2540B15.12 | lincRNA | rs56079812 | 33755525 | 1.06e-10 | 3 | 0 | 0 | 1 | 1 | 1 | Female only | Both sexes | Female only | True | False |
| BASOAB | 12 | HELB | protein_coding | rs1168311 | 66725912 | 2.40e-10 | 143 | 0 | 0 | 2 | 38 | 103 | Female only | Both sexes | Female only | True | False |
| BASOAB | 3 | RP11-475N22.4 | antisense | rs4550835 | 128228394 | 2.70e-10 | 3 | 0 | 1 | 0 | 1 | 1 | Female only | Both sexes | Female only | True | False |
| BASOAB | 4 | AFF1 | protein_coding | rs2035403 | 88018991 | 3.31e-10 | 27 | 0 | 0 | 14 | 0 | 13 | Female only | Both sexes | Female only | True | False |
| BASOAB | 19 | CTD-2540B15.12 | lincRNA | rs4805869 | 33762602 | 3.80e-10 | 3 | 0 | 0 | 0 | 0 | 3 | Both sexes | Both sexes | Both sexes | True | False |
| BASOAB | 19 | SLC7A10 | protein_coding | rs11669923 | 33731379 | 9.01e-10 | 3 | 0 | 0 | 0 | 0 | 3 | Both sexes | Both sexes | Both sexes | True | False |
| BASOAB | 6 | HCP5 | sense_overlapping | rs3128981 | 31405620 | 1.02e-09 | 35 | 0 | 0 | 1 | 4 | 30 | Both sexes | Both sexes | Both sexes | True | True |
| BASOAB | 19 | CTD-2540B15.12 | lincRNA | rs736289 | 33757062 | 1.28e-09 | 3 | 1 | 1 | 0 | 0 | 1 | Both sexes | Both sexes | Both sexes | True | False |

| pheno | CHR | Gene_Symbol | biotype | lead_snp | lead_BP | lead_P | TOTAL | NSIG | S05 | S01 | S001 | S0001 | independent_loci_in_sex_P1e6 | independent_loci_in_sex_P1e2 | independent_loci_in_sex_P5e8 | in_gwas_catalog | in_HLA |
|---|---|---|---|---|---|---|---|---|---|---|---|---|---|---|---|---|---|
| BASOAB | 9 | JAK2 | protein_coding | rs3780367 | 5068755 | 1.54e-09 | 499 | 0 | 0 | 2 | 185 | 312 | Female only | Both sexes | Female only | True | False |
| BASOAB | 11 | C11orf21 | protein_coding | rs16928084 | 23229544 | 2.17e-09 | 21 | 0 | 0 | 0 | 0 | 21 | Female only | Both sexes | Female only | True | False |
| BASOAB | 6 | HLA-B | protein_coding | rs9405083 | 31319489 | 2.23e-09 | 19 | 3 | 5 | 1 | 3 | 7 | Female only | Both sexes | Female only | True | True |
| BASOAB | 1 | RP5-1052I5.2 | protein_coding | rs3893263 | 87711755 | 2.26e-09 | 7 | 0 | 0 | 0 | 0 | 7 | Both sexes | Both sexes | Female only | True | False |
| BASOAB | 6 | HLA-B | protein_coding | rs3819306 | 31322133 | 2.26e-09 | 5 | 0 | 0 | 1 | 3 | 1 | Male only | Both sexes | Male only | True | True |
| BASOAB | 12 | RP11-335I12.2 | antisense | rs11176115 | 66658183 | 3.29e-09 | 41 | 0 | 0 | 11 | 10 | 20 | Female only | Both sexes | Female only | True | False |
| BASOAB | 3 | TMED10P2 | pseudogene | rs7620032 | 128245104 | 3.45e-09 | 9 | 0 | 0 | 0 | 0 | 9 | Female only | Both sexes | Female only | True | False |
| BASOAB | 2 | THADA | protein_coding | rs77981966 | 43777964 | 5.46e-09 | 453 | 65 | 68 | 87 | 9 | 224 | Female only | Both sexes | Female only | True | False |
| BASOAB | 20 | C20orf112 | protein_coding | rs1535374 | 31162125 | 5.71e-09 | 29 | 0 | 3 | 0 | 0 | 26 | Both sexes | Both sexes | Female only | True | False |
| BASOAB | 6 | C6orf183 | polymorphic_pseudogene | rs9487003 | 109515666 | 5.81e-09 | 481 | 0 | 0 | 0 | 31 | 450 | Both sexes | Both sexes | Both sexes | True | False |
| BASOAB | 6 | HLA-B | protein_coding | rs5021845 | 31314866 | 1.29e-08 | 81 | 0 | 7 | 12 | 27 | 35 | Both sexes | Both sexes | Female only | True | True |
| BASOAB | 6 | DHFRP2 | pseudogene | rs2844572 | 31335575 | 1.46e-08 | 77 | 0 | 1 | 7 | 13 | 56 | Both sexes | Both sexes | Male only | True | True |
| BASOAB | 3 | KIAA1257 | protein_coding | rs71331680 | 128662940 | 1.60e-08 | 5 | 0 | 0 | 0 | 2 | 3 | Female only | Both sexes | Female only | True | False |
| BASOAB | 3 | RPN1 | protein_coding | rs7635019 | 128320361 | 1.68e-08 | 63 | 0 | 0 | 0 | 4 | 59 | Female only | Both sexes | Female only | True | False |
| BASOAB | 1 | LMO4 | protein_coding | rs4279831 | 87724930 | 2.24e-08 | 7 | 2 | 0 | 0 | 0 | 5 | Female only | Both sexes | Female only | True | False |
| BASOAB | 6 | ZDHHC20P2 | pseudogene | rs2244020 | 31347451 | 2.83e-08 | 9 | 0 | 1 | 1 | 1 | 6 | Male only | Both sexes | Male only | True | True |
| BASOAB | 6 | XXbac-BPG248L24.10 | pseudogene | rs3873388 | 31275827 | 2.88e-08 | 3 | 2 | 0 | 0 | 0 | 1 | Female only | Female only | Female only | True | True |
| BASOAB | 6 | XXbac-BPG248L24.12 | pseudogene | rs9266233 | 31325430 | 3.09e-08 | 7 | 0 | 0 | 3 | 0 | 4 | Both sexes | Both sexes | Male only | True | True |
| BASOAB | 20 | C20orf203 | lincRNA | rs12481717 | 31215664 | 3.76e-08 | 5 | 0 | 0 | 0 | 0 | 5 | Both sexes | Both sexes | Male only | True | False |

| pheno | CHR | Gene_Symbol | biotype | lead_snp | lead_BP | lead_P | TOTAL | NSIG | S05 | S01 | S001 | S0001 | independent_loci_in_sex_P1e6 | independent_loci_in_sex_P1e2 | independent_loci_in_sex_P5e8 | in_gwas_catalog | in_HLA |
|---|---|---|---|---|---|---|---|---|---|---|---|---|---|---|---|---|---|
| BASOAB | 14 | SLC7A8 | protein_coding | rs8010847 | 23594396 | 3.99e-08 | 3 | 0 | 0 | 0 | 2 | 1 | Female only | Both sexes | Female only | True | False |
| BASOAB | 3 | CRBN | protein_coding | rs1669339 | 3199170 | 4.62e-08 | 31 | 0 | 0 | 8 | 9 | 14 | Male only | Both sexes | Male only | True | False |
| BASOAB | 6 | DHFRP2 | pseudogene | rs9266382 | 31334877 | 4.65e-08 | 97 | 1 | 2 | 47 | 1 | 46 | Male only | Both sexes | Male only | True | True |
| BASOAB | 6 | HCP5 | sense_overlapping | rs7769393 | 31411312 | 4.74e-08 | 327 | 0 | 1 | 68 | 125 | 133 | Female only | Both sexes | Female only | True | True |
| BASOM | 16 | CMIP | protein_coding | rs75217481 | 81576331 | 4.89e-08 | 23 | 0 | 7 | 5 | 0 | 11 | Female only | Both sexes | Female only | True | False |

| pheno | CHR | Gene_Symbol | biotype | lead_snp | lead_BP | lead_P | TOTAL | NSIG | S05 | S01 | S001 | S0001 | independent_loci_in_sex_P1e6 | independent_loci_in_sex_P1e2 | independent_loci_in_sex_P5e8 | in_gwas_catalog | in_HLA |
|---|---|---|---|---|---|---|---|---|---|---|---|---|---|---|---|---|---|
| BASOPHILS | 23 | TENM1 | protein_coding | X:123595260:A:G | 123595260 | 1.47e-08 | 7 | 4 | 0 | 0 | 0 | 3 | Male only | Male only | Male only | True | False |
| BASORE | 19 | CTD-2540B15.12 | lincRNA | rs78744187 | 33754548 | 9.20e-119 | 5 | 0 | 0 | 0 | 0 | 5 | Both sexes | Both sexes | Both sexes | True | False |
| BASORE | 3 | C3orf27 | protein_coding | rs4857909 | 128300468 | 6.54e-66 | 49 | 0 | 0 | 0 | 0 | 49 | Both sexes | Both sexes | Both sexes | True | False |
| BASORE | 3 | RPN1 | protein_coding | rs2253125 | 128329793 | 1.96e-41 | 3 | 0 | 0 | 0 | 0 | 3 | Both sexes | Both sexes | Both sexes | True | False |
| BASORE | 19 | CTD-2540B15.12 | lincRNA | rs67228209 | 33751033 | 2.60e-26 | 3 | 0 | 0 | 0 | 0 | 3 | Both sexes | Both sexes | Both sexes | True | False |
| BASORE | 3 | C3orf27 | protein_coding | rs4857906 | 128275459 | 2.68e-24 | 103 | 0 | 0 | 0 | 0 | 103 | Both sexes | Both sexes | Both sexes | True | False |
| BASORE | 3 | RAB7A | protein_coding | rs61292315 | 128542609 | 8.50e-23 | 119 | 0 | 0 | 0 | 0 | 119 | Both sexes | Both sexes | Both sexes | True | False |
| BASORE | 19 | CTD-2540B15.12 | lincRNA | rs12461296 | 33774591 | 6.69e-18 | 3 | 0 | 0 | 0 | 0 | 3 | Both sexes | Both sexes | Both sexes | True | False |
| BASORE | 14 | CEBPE | protein_coding | rs4982731 | 23585333 | 7.06e-17 | 7 | 0 | 0 | 0 | 0 | 7 | Both sexes | Both sexes | Both sexes | True | False |
| BASORE | 19 | CTD-2540B15.12 | lincRNA | rs4432367 | 33752920 | 8.08e-16 | 3 | 0 | 0 | 0 | 0 | 3 | Both sexes | Both sexes | Both sexes | True | False |
| BASORE | 19 | CTD-2540B15.12 | lincRNA | rs10414008 | 33758327 | 2.15e-15 | 5 | 0 | 0 | 1 | 0 | 4 | Both sexes | Both sexes | Both sexes | True | False |
| BASORE | 1 | LMO4 | protein_coding | rs1888342 | 87751406 | 3.16e-15 | 35 | 0 | 0 | 0 | 0 | 35 | Both sexes | Both sexes | Both sexes | True | False |
| BASORE | 3 | RAB7A | protein_coding | rs3772157 | 128533990 | 3.75e-15 | 43 | 0 | 0 | 0 | 0 | 43 | Both sexes | Both sexes | Both sexes | True | False |
| BASORE | 19 | CTD-2540B15.12 | lincRNA | rs4805867 | 33762257 | 1.52e-13 | 3 | 0 | 0 | 0 | 0 | 3 | Both sexes | Both sexes | Both sexes | True | False |
| BASORE | 3 | CRBN | protein_coding | rs1669340 | 3198380 | 2.95e-13 | 41 | 0 | 0 | 5 | 8 | 28 | Female only | Both sexes | Female only | True | False |
| BASORE | 19 | CTD-2540B15.12 | lincRNA | rs56079812 | 33755525 | 4.59e-13 | 3 | 0 | 0 | 0 | 1 | 2 | Female only | Both sexes | Female only | True | False |
| BASORE | 3 | RAB7A | protein_coding | rs56373544 | 128523503 | 2.28e-12 | 25 | 0 | 0 | 0 | 1 | 24 | Both sexes | Both sexes | Female only | True | False |
| BASORE | 3 | C3orf27 | protein_coding | rs72987210 | 128283406 | 2.73e-12 | 37 | 0 | 0 | 0 | 0 | 37 | Both sexes | Both sexes | Both sexes | True | False |
| BASORE | 1 | LMO4 | protein_coding | rs4655844 | 87775058 | 1.87e-11 | 17 | 0 | 0 | 0 | 0 | 17 | Both sexes | Both sexes | Both sexes | True | False |
| BASORE | 1 | RP5-1052I5.2 | protein_coding | rs3893263 | 87711755 | 2.03e-11 | 7 | 0 | 0 | 0 | 0 | 7 | Both sexes | Both sexes | Both sexes | True | False |
| BASORE | 3 | RPN1 | protein_coding | rs2253144 | 128329920 | 6.65e-11 | 169 | 0 | 7 | 57 | 29 | 76 | Female only | Both sexes | Female only | True | False |
| BASORE | 3 | C3orf27 | protein_coding | rs66910450 | 128303893 | 8.73e-11 | 63 | 2 | 16 | 14 | 0 | 31 | Female only | Both sexes | Female only | True | False |
| BASORE | 15 | RP11-96C21.1 | lincRNA | rs13379715 | 70738476 | 1.21e-10 | 155 | 0 | 0 | 0 | 8 | 147 | Both sexes | Both sexes | Male only | True | False |
| BASORE | 20 | C20orf203 | lincRNA | rs6058799 | 31257732 | 1.25e-10 | 125 | 0 | 0 | 0 | 0 | 125 | Both sexes | Both sexes | Both sexes | True | False |
| BASORE | 20 | C20orf203 | lincRNA | rs6058790 | 31247317 | 1.85e-10 | 31 | 9 | 0 | 8 | 8 | 6 | Female only | Both sexes | Female only | True | False |

| pheno | CHR | Gene_Symbol | biotype | lead_snp | lead_BP | lead_P | TOTAL | NSIG | S05 | S01 | S001 | S0001 | independent_loci_in_sex_P1e6 | independent_loci_in_sex_P1e2 | independent_loci_in_sex_P5e8 | in_gwas_catalog | in_HLA |
|---|---|---|---|---|---|---|---|---|---|---|---|---|---|---|---|---|---|
| BASORE | 19 | CTD-2540B15.12 | lincRNA | rs36112953 | 33754309 | 2.12e-10 | 7 | 0 | 0 | 0 | 0 | 7 | Both sexes | Both sexes | Both sexes | True | False |
| BASORE | 18 | RP11-737G21.1 | pseudogene | rs11659727 | 23503073 | 2.13e-10 | 101 | 51 | 0 | 0 | 1 | 49 | Female only | Female only | Female only | True | False |
| BASORE | 12 | HELB | protein_coding | rs1168301 | 66736771 | 5.20e-10 | 143 | 0 | 3 | 47 | 0 | 93 | Female only | Both sexes | Female only | True | False |
| BASORE | 4 | AFF1 | protein_coding | rs60695258 | 88022709 | 1.45e-09 | 27 | 0 | 0 | 6 | 8 | 13 | Female only | Both sexes | Female only | True | False |
| BASORE | 11 | C11orf21 | protein_coding | rs74048209 | 2320224 | 1.64e-09 | 23 | 0 | 0 | 0 | 1 | 22 | Female only | Both sexes | Female only | True | False |
| BASORE | 20 | C20orf203 | lincRNA | rs1407256 | 31213647 | 1.74e-09 | 5 | 0 | 0 | 0 | 0 | 5 | Both sexes | Both sexes | Both sexes | True | False |
| BASORE | 1 | LMO4 | protein_coding | rs4279831 | 87724930 | 2.23e-09 | 7 | 2 | 0 | 0 | 0 | 5 | Female only | Both sexes | Female only | True | False |
| BASORE | 20 | C20orf112 | protein_coding | rs1535374 | 31162125 | 2.36e-09 | 29 | 0 | 0 | 0 | 3 | 26 | Both sexes | Both sexes | Both sexes | True | False |
| BASORE | 4 | AFF1 | protein_coding | rs442177 | 88030261 | 3.24e-09 | 69 | 10 | 12 | 13 | 0 | 34 | Female only | Both sexes | Female only | True | False |
| BASORE | 3 | RP11-475N22.4 | antisense | rs4550835 | 128228394 | 3.84e-09 | 3 | 0 | 1 | 0 | 0 | 2 | Female only | Both sexes | Female only | True | False |
| BASORE | 18 | RP11-737G21.1 | pseudogene | rs8097343 | 23542220 | 5.32e-09 | 91 | 46 | 0 | 0 | 0 | 45 | Female only | Female only | Female only | True | False |
| BASORE | 12 | PDCL3P7 | pseudogene | rs10748037 | 66664573 | 1.22e-08 | 41 | 0 | 0 | 13 | 8 | 20 | Female only | Both sexes | Female only | True | False |
| BASORE | 14 | SLC7A8 | protein_coding | rs28479481 | 23592517 | 1.36e-08 | 9 | 0 | 0 | 0 | 4 | 5 | Female only | Both sexes | Female only | True | False |
| BASORE | 3 | C3orf27 | protein_coding | rs4857857 | 128313880 | 1.50e-08 | 25 | 0 | 1 | 1 | 4 | 19 | Female only | Both sexes | Female only | True | False |
| BASORE | 6 | C6orf183 | polymorphic_pseudogene | rs34527629 | 109517529 | 2.45e-08 | 481 | 0 | 4 | 123 | 95 | 259 | Female only | Both sexes | Female only | True | False |
| BASORE | 6 | MICA | protein_coding | rs1882 | 31382911 | 2.87e-08 | 45 | 0 | 0 | 3 | 11 | 31 | Both sexes | Both sexes | Both sexes | True | True |
| BASORE | 18 | SS18 | protein_coding | rs2959356 | 23590203 | 2.91e-08 | 3 | 1 | 0 | 0 | 1 | 1 | Female only | Both sexes | Female only | True | False |
| BASORE | 20 | TM9SF4 | protein_coding | rs2235904 | 30721712 | 2.99e-08 | 315 | 49 | 97 | 12 | 43 | 114 | Female only | Both sexes | Female only | True | False |
| BASORE | 14 | SLC7A8 | protein_coding | rs17794251 | 23593442 | 3.15e-08 | 3 | 0 | 0 | 0 | 2 | 1 | Female only | Both sexes | Female only | True | False |
| BAbs | 19 | CTD-2540B15.12 | lincRNA | rs78744187 | 33754548 | 1.29e-59 | 5 | 0 | 0 | 0 | 0 | 5 | Both sexes | Both sexes | Both sexes | True | False |
| BAbs | 3 | C3orf27 | protein_coding | rs13089722 | 128306757 | 2.15e-22 | 53 | 0 | 0 | 0 | 0 | 53 | Both sexes | Both sexes | Both sexes | True | False |
| BAbs | 19 | CTD-2540B15.12 | lincRNA | rs67228209 | 33751033 | 1.51e-14 | 3 | 0 | 0 | 0 | 0 | 3 | Both sexes | Both sexes | Both sexes | True | False |
| BAbs | 3 | C3orf27 | protein_coding | rs8179974 | 128281701 | 3.26e-13 | 123 | 0 | 0 | 0 | 7 | 116 | Both sexes | Both sexes | Both sexes | True | False |
| BAbs | 19 | CTD-2540B15.12 | lincRNA | rs10414008 | 33758327 | 4.25e-12 | 5 | 1 | 1 | 0 | 0 | 3 | Both sexes | Both sexes | Both sexes | True | False |
| BAbs | 14 | CEBPE | protein_coding | rs4982731 | 23585333 | 6.45e-12 | 7 | 0 | 1 | 3 | 0 | 3 | Female only | Both sexes | Female only | True | False |
| BAbs | 12 | HELB | protein_coding | rs11176145 | 66719795 | 1.62e-10 | 185 | 0 | 0 | 33 | 17 | 135 | Female only | Both sexes | Female only | True | False |

| pheno | CHR | Gene_Symbol | biotype | lead_snp | lead_BP | lead_P | TOTAL | NSIG | S05 | S01 | S001 | S0001 | independent_loci_in_sex_P1e6 | independent_loci_in_sex_P1e2 | independent_loci_in_sex_P5e8 | in_gwas_catalog | in_HLA |
|---|---|---|---|---|---|---|---|---|---|---|---|---|---|---|---|---|---|
| BAbs | 20 | C20orf203 | lincRNA | rs6058807 | 31264608 | 5.01e-10 | 125 | 63 | 0 | 0 | 0 | 62 | Female only | Female only | Female only | True | False |
| BAbs | 3 | RAB7A | protein_coding | rs35904610 | 128452108 | 2.93e-09 | 115 | 0 | 0 | 0 | 29 | 86 | Female only | Both sexes | Female only | True | False |
| BAbs | 19 | CTD-2540B15.12 | lincRNA | rs4432367 | 33752920 | 4.14e-09 | 3 | 0 | 0 | 2 | 0 | 1 | Female only | Both sexes | Female only | True | False |
| BAbs | 3 | C3orf27 | protein_coding | rs1170 | 128282416 | 1.96e-08 | 13 | 2 | 0 | 2 | 2 | 7 | Female only | Both sexes | Female only | True | False |
| pheno | CHR | Gene_Symbol | biotype | lead_snp | lead_BP | lead_P | TOTAL | NSIG | S05 | S01 | S001 | S0001 | independent_loci_in_sex_P1e6 | independent_loci_in_sex_P1e2 | independent_loci_in_sex_P5e8 | in_gwas_catalog | in_HLA |
| BAbs | 3 | CRBN | protein_coding | rs1669340 | 3198380 | 1.98e-08 | 41 | 0 | 0 | 0 | 2 | 39 | Both sexes | Both sexes | Female only | True | False |
| BAbs | 19 | CTD-2540B15.12 | lincRNA | rs10420798 | 33764718 | 2.29e-08 | 3 | 0 | 0 | 0 | 0 | 3 | Both sexes | Both sexes | Male only | True | False |
| BAbs | 19 | SLC7A10 | protein_coding | rs11669923 | 33731379 | 2.92e-08 | 3 | 0 | 0 | 0 | 2 | 1 | Female only | Both sexes | Female only | True | False |
| BAbs | 19 | CTD-2540B15.12 | lincRNA | rs36112953 | 33754309 | 3.88e-08 | 7 | 0 | 0 | 0 | 0 | 7 | Female only | Both sexes | Female only | True | False |
| BMI_CLEAN | 16 | FTO | protein_coding | rs62048402 | 53803223 | 6.49e-41 | 169 | 0 | 0 | 0 | 0 | 169 | Both sexes | Both sexes | Both sexes | True | False |
| BMI_CLEAN | 16 | FTO | protein_coding | rs1477196 | 53808258 | 2.83e-18 | 9 | 0 | 0 | 0 | 0 | 9 | Both sexes | Both sexes | Both sexes | True | False |
| BMI_CLEAN | 18 | RP11-795H16.3 | lincRNA | rs12967878 | 57826570 | 3.67e-15 | 243 | 0 | 23 | 10 | 11 | 199 | Both sexes | Both sexes | Female only | True | False |
| BMI_CLEAN | 1 | SEC16B | protein_coding | rs516636 | 177855517 | 7.03e-13 | 101 | 21 | 0 | 5 | 25 | 50 | Female only | Both sexes | Female only | True | False |
| BMI_CLEAN | 2 | TMEM18 | protein_coding | rs6760274 | 652272 | 2.41e-11 | 337 | 0 | 0 | 0 | 0 | 337 | Both sexes | Both sexes | Female only | True | False |
| BMI_CLEAN | 18 | RP11-795H16.2 | pseudogene | rs12456624 | 57903086 | 5.68e-10 | 55 | 0 | 6 | 22 | 0 | 27 | Female only | Both sexes | Female only | True | False |
| BMI_CLEAN | 18 | RNU6-567P | snRNA | rs7231154 | 57738097 | 2.35e-09 | 29 | 9 | 6 | 0 | 0 | 14 | Female only | Female only | Female only | True | False |
| BMI_CLEAN | 2 | ADCY3 | protein_coding | rs2033654 | 25103108 | 4.02e-09 | 303 | 0 | 3 | 65 | 88 | 147 | Both sexes | Both sexes | Female only | True | False |
| BMI_CLEAN | 2 | TMEM18 | protein_coding | rs2867121 | 658550 | 4.12e-09 | 5 | 0 | 0 | 2 | 1 | 2 | Female only | Both sexes | Female only | True | False |
| BUN | 5 | PRKAA1 | protein_coding | rs74653842 | 40773178 | 2.02e-09 | 37 | 0 | 0 | 17 | 2 | 18 | Female only | Both sexes | Female only | True | False |
| BUN | 15 | UBE2Q2 | protein_coding | rs1976748 | 76160951 | 2.30e-08 | 25 | 0 | 0 | 11 | 2 | 12 | Male only | Both sexes | Male only | True | False |
| BUN | 16 | UMOD | protein_coding | rs36060036 | 20361950 | 2.61e-08 | 53 | 0 | 0 | 0 | 4 | 49 | Male only | Both sexes | Male only | True | False |
| Baso | 19 | CTD-2540B15.12 | lincRNA | rs78744187 | 33754548 | 5.99e-68 | 5 | 0 | 0 | 0 | 0 | 5 | Both sexes | Both sexes | Both sexes | True | False |

| pheno | CHR | Gene_Symbol | biotype | lead_snp | lead_BP | lead_P | TOTAL | NSIG | S05 | S01 | S001 | S0001 | independent_loci_in_sex_P1e6 | independent_loci_in_sex_P1e2 | independent_loci_in_sex_P5e8 | in_gwas_catalog | in_HLA |
|---|---|---|---|---|---|---|---|---|---|---|---|---|---|---|---|---|---|
| Baso | 3 | C3orf27 | protein_coding | rs4328821 | 128316435 | 3.78e-28 | 53 | 0 | 0 | 0 | 0 | 53 | Both sexes | Both sexes | Both sexes | True | False |
| Baso | 19 | CTD-2540B15.12 | lincRNA | rs67228209 | 33751033 | 7.17e-16 | 3 | 0 | 0 | 0 | 0 | 3 | Both sexes | Both sexes | Both sexes | True | False |
| Baso | 3 | C3orf27 | protein_coding | rs6806253 | 128282697 | 1.19e-14 | 81 | 0 | 0 | 0 | 0 | 81 | Both sexes | Both sexes | Both sexes | True | False |
| Baso | 19 | CTD-2540B15.12 | lincRNA | rs10412166 | 33758201 | 1.81e-12 | 5 | 1 | 1 | 0 | 0 | 3 | Both sexes | Both sexes | Both sexes | True | False |
| Baso | 19 | CTD-2540B15.12 | lincRNA | rs4805867 | 33762257 | 2.34e-12 | 3 | 0 | 0 | 0 | 0 | 3 | Both sexes | Both sexes | Female only | True | False |
| Baso | 20 | C20orf203 | lincRNA | rs6141766 | 31222769 | 1.10e-11 | 53 | 24 | 3 | 0 | 0 | 26 | Female only | Female only | Female only | True | False |
| Baso | 19 | CTD-2540B15.12 | lincRNA | rs10420798 | 33764718 | 3.94e-11 | 3 | 0 | 0 | 0 | 0 | 3 | Both sexes | Both sexes | Female only | True | False |
| Baso | 3 | RAB7A | protein_coding | rs35904610 | 128452108 | 3.06e-10 | 115 | 0 | 0 | 0 | 22 | 93 | Both sexes | Both sexes | Both sexes | True | False |
| Baso | 3 | RAB7A | protein_coding | rs56373544 | 128523503 | 3.74e-10 | 25 | 4 | 9 | 0 | 0 | 12 | Female only | Female only | Female only | True | False |
| Baso | 20 | COMMD7 | protein_coding | rs28510728 | 31287891 | 5.24e-10 | 77 | 39 | 0 | 0 | 0 | 38 | Female only | Female only | Female only | True | False |
| Baso | 3 | CRBN | protein_coding | rs1669340 | 3198380 | 7.51e-10 | 41 | 0 | 0 | 0 | 6 | 35 | Female only | Both sexes | Female only | True | False |
| Baso | 14 | CEBPE | protein_coding | rs4982731 | 23585333 | 1.16e-09 | 7 | 0 | 0 | 1 | 3 | 3 | Female only | Both sexes | Female only | True | False |
| Baso | 12 | HELB | protein_coding | rs11176145 | 66719795 | 2.99e-09 | 185 | 0 | 0 | 2 | 50 | 133 | Female only | Both sexes | Female only | True | False |
| Baso | 19 | CTD-2540B15.12 | lincRNA | rs36112953 | 33754309 | 3.09e-09 | 7 | 0 | 0 | 0 | 0 | 7 | Both sexes | Both sexes | Female only | True | False |
| Baso | 20 | C20orf112 | protein_coding | 20:31068883 | 31068883 | 8.33e-09 | 201 | 0 | 97 | 4 | 0 | 100 | Female only | Both sexes | Female only | True | False |
| Baso | 3 | TMED10P2 | pseudogene | 3:128250135 | 128250135 | 1.42e-08 | 59 | 0 | 5 | 0 | 5 | 49 | Both sexes | Both sexes | Both sexes | True | False |
| Baso | 4 | AFF1 | protein_coding | rs17605615 | 87996745 | 2.19e-08 | 27 | 0 | 0 | 0 | 8 | 19 | Male only | Both sexes | Male only | True | False |
| Baso | 3 | RP11-475N22.4 | antisense | rs13060840 | 128238682 | 2.67e-08 | 11 | 0 | 0 | 0 | 5 | 6 | Female only | Both sexes | Female only | True | False |
| Baso | 20 | C20orf112 | protein_coding | rs10875486 | 31163573 | 3.01e-08 | 5 | 2 | 1 | 0 | 0 | 2 | Female only | Female only | Female only | True | False |
| Baso | 19 | CTD-2540B15.12 | lincRNA | rs4432367 | 33752920 | 4.24e-08 | 3 | 0 | 0 | 0 | 0 | 3 | Female only | Both sexes | Female only | True | False |
| C.Gluc | 13 | RN7SL571P | misc_RNA | rs61963436 | 77061127 | 3.50e-09 | 45 | 23 | 0 | 0 | 0 | 22 | Female only | Female only | Female only | False | False |
| C4Quan | 6 | ATF6B | protein_coding | rs8283 | 32083300 | 8.97e-30 | 19 | 0 | 0 | 0 | 1 | 18 | Both sexes | Both sexes | Both sexes | True | True |
| C4Quan | 6 | STK19 | protein_coding | rs377370 | 31942514 | 1.97e-24 | 51 | 0 | 0 | 18 | 8 | 25 | Female only | Both sexes | Female only | True | True |
| C4Quan | 6 | HLA-DQB1 | protein_coding | 6:32623223 | 32623223 | 3.24e-22 | 127 | 0 | 0 | 0 | 0 | 127 | Both sexes | Both sexes | Both sexes | True | True |
| C4Quan | 6 | BTNL2 | protein_coding | rs6918317 | 32381210 | 4.33e-22 | 145 | 0 | 0 | 9 | 4 | 132 | Both sexes | Both sexes | Both sexes | True | True |
| C4Quan | 6 | SLC44A4 | protein_coding | rs515688 | 31832038 | 6.37e-22 | 163 | 0 | 0 | 0 | 0 | 163 | Both sexes | Both sexes | Both sexes | True | True |

| pheno | CHR | Gene_Symbol | biotype | lead_snp | lead_BP | lead_P | TOTAL | NSIG | S05 | S01 | S001 | S0001 | independent_loci_in_sex_P1e6 | independent_loci_in_sex_P1e2 | independent_loci_in_sex_P5e8 | in_gwas_catalog | in_HLA |
|---|---|---|---|---|---|---|---|---|---|---|---|---|---|---|---|---|---|
| C4Quan | 6 | HLA-DRB1 | protein_coding | rs2760995 | 32574358 | 1.24e-21 | 79 | 0 | 0 | 2 | 0 | 77 | Both sexes | Both sexes | Both sexes | True | True |
| C4Quan | 6 | TNXB | protein_coding | rs2269429 | 32029183 | 1.31e-21 | 43 | 0 | 1 | 0 | 0 | 42 | Female only | Both sexes | Female only | True | True |
| C4Quan | 6 | HCP5 | sense_overlapping | rs3131643 | 31442782 | 5.72e-21 | 199 | 0 | 0 | 0 | 0 | 199 | Both sexes | Both sexes | Both sexes | True | True |
| C4Quan | 6 | ATF6B | protein_coding | rs3130288 | 32096001 | 3.88e-20 | 147 | 0 | 0 | 0 | 6 | 141 | Both sexes | Both sexes | Both sexes | True | True |
| C4Quan | 6 | HLA-DQA1 | protein_coding | rs13208697 | 32583274 | 4.77e-20 | 3 | 0 | 0 | 0 | 0 | 3 | Both sexes | Both sexes | Both sexes | True | True |
| C4Quan | 6 | HLA-DQA1 | protein_coding | rs11751024 | 32586236 | 7.50e-20 | 11 | 2 | 0 | 3 | 0 | 6 | Female only | Both sexes | Female only | True | True |
| C4Quan | 6 | C6orf47 | protein_coding | rs805262 | 31628733 | 4.46e-19 | 35 | 17 | 6 | 11 | 1 | 0 | Female only | Both sexes | Female only | True | True |
| C4Quan | 6 | HCP5 | sense_overlapping | rs3094604 | 31434111 | 5.70e-19 | 67 | 0 | 0 | 0 | 0 | 67 | Both sexes | Both sexes | Both sexes | True | True |
| C4Quan | 6 | HLA-DQA1 | protein_coding | rs9271573 | 32590501 | 6.90e-19 | 7 | 0 | 0 | 0 | 1 | 6 | Both sexes | Both sexes | Female only | True | True |
| C4Quan | 6 | CSNK2B | protein_coding | rs14365 | 31635710 | 2.84e-18 | 61 | 0 | 0 | 16 | 12 | 33 | Both sexes | Both sexes | Both sexes | True | True |
| C4Quan | 6 | MICB | protein_coding | rs2855808 | 31469668 | 7.17e-18 | 135 | 1 | 0 | 0 | 3 | 131 | Both sexes | Both sexes | Both sexes | True | True |
| C4Quan | 6 | XXbac-BPG154L12.4 | antisense | rs9267995 | 32221552 | 1.34e-17 | 23 | 0 | 0 | 1 | 0 | 22 | Both sexes | Both sexes | Female only | True | True |
| C4Quan | 6 | XXbac-BPG254F23.7 | lincRNA | rs3104406 | 32682443 | 1.48e-17 | 83 | 0 | 0 | 0 | 2 | 81 | Both sexes | Both sexes | Female only | True | True |
| C4Quan | 6 | XXbac-BPG254F23.7 | lincRNA | rs2858331 | 32681277 | 2.57e-17 | 37 | 0 | 0 | 0 | 0 | 37 | Both sexes | Both sexes | Female only | True | True |
| C4Quan | 6 | HLA-DRB9 | pseudogene | rs29001517 | 32447074 | 3.05e-17 | 41 | 0 | 0 | 9 | 11 | 21 | Both sexes | Both sexes | Both sexes | True | True |
| C4Quan | 6 | BTNL2 | protein_coding | rs3129956 | 32368462 | 3.85e-17 | 139 | 0 | 0 | 0 | 0 | 139 | Both sexes | Both sexes | Both sexes | True | True |
| C4Quan | 6 | MTCO3P1 | pseudogene | rs2856725 | 32666738 | 7.74e-17 | 185 | 0 | 0 | 22 | 18 | 145 | Both sexes | Both sexes | Both sexes | True | True |

| pheno | CHR | Gene_Symbol | biotype | lead_snp | lead_BP | lead_P | TOTAL | NSIG | S05 | S01 | S001 | S0001 | independent_loci_in_sex_P1e6 | independent_loci_in_sex_P1e2 | independent_loci_in_sex_P5e8 | in_gwas_catalog | in_HLA |
|---|---|---|---|---|---|---|---|---|---|---|---|---|---|---|---|---|---|
| C4Quan | 6 | LSM2 | protein_coding | rs9461726 | 31769796 | 1.15e-16 | 17 | 1 | 1 | 0 | 0 | 15 | Both sexes | Both sexes | Female only | True | True |
| C4Quan | 6 | HLA-DQA1 | protein_coding | rs2097432 | 32590771 | 1.78e-16 | 101 | 0 | 0 | 0 | 10 | 91 | Both sexes | Both sexes | Both sexes | True | True |
| C4Quan | 6 | HCP5 | sense_overlapping | rs2263318 | 31432006 | 2.97e-16 | 11 | 0 | 0 | 3 | 1 | 7 | Both sexes | Both sexes | Both sexes | True | True |
| C4Quan | 6 | STK19 | protein_coding | rs41315816 | 31944851 | 5.15e-16 | 21 | 4 | 0 | 4 | 1 | 12 | Both sexes | Both sexes | Both sexes | True | True |
| C4Quan | 6 | HLA-S | pseudogene | rs2596560 | 31355318 | 5.31e-16 | 5 | 0 | 0 | 0 | 0 | 5 | Both sexes | Both sexes | Both sexes | True | True |
| C4Quan | 6 | MICB | protein_coding | rs2534664 | 31469591 | 6.61e-16 | 59 | 0 | 0 | 25 | 0 | 34 | Both sexes | Both sexes | Both sexes | True | True |
| C4Quan | 6 | C6orf10 | protein_coding | rs3132954 | 32311459 | 6.69e-16 | 45 | 2 | 0 | 0 | 0 | 43 | Both sexes | Both sexes | Both sexes | True | True |
| C4Quan | 6 | C6orf10 | protein_coding | rs3129942 | 32338283 | 1.04e-15 | 345 | 0 | 0 | 0 | 2 | 343 | Both sexes | Both sexes | Both sexes | True | True |

| pheno | CHR | Gene_Symbol | biotype | lead_snp | lead_BP | lead_P | TOTAL | NSIG | S05 | S01 | S001 | S0001 | independent_loci_in_sex_P1e6 | independent_loci_in_sex_P1e2 | independent_loci_in_sex_P5e8 | in_gwas_catalog | in_HLA |
|---|---|---|---|---|---|---|---|---|---|---|---|---|---|---|---|---|---|
| C4Quan | 6 | HLA-DRB1 | protein_coding | rs28366353 | 32565004 | 1.15e-15 | 267 | 0 | 2 | 60 | 11 | 194 | Both sexes | Both sexes | Female only | True | True |
| C4Quan | 6 | MTCO3P1 | pseudogene | rs2856665 | 32665597 | 1.72e-15 | 91 | 0 | 0 | 0 | 14 | 77 | Female only | Both sexes | Female only | True | True |
| C4Quan | 6 | DHFRP2 | pseudogene | rs2523562 | 31330618 | 2.22e-15 | 19 | 1 | 0 | 0 | 1 | 17 | Both sexes | Both sexes | Female only | True | True |
| C4Quan | 6 | CFB | protein_coding | rs537160 | 31916400 | 2.66e-15 | 39 | 1 | 1 | 0 | 0 | 37 | Both sexes | Both sexes | Both sexes | True | True |
| C4Quan | 6 | HLA-DRA | protein_coding | rs9268668 | 32413889 | 2.75e-15 | 47 | 0 | 0 | 0 | 0 | 47 | Both sexes | Both sexes | Both sexes | True | True |
| C4Quan | 6 | ABHD16A | protein_coding | rs1266074 | 31666551 | 4.47e-15 | 7 | 0 | 0 | 0 | 0 | 7 | Both sexes | Both sexes | Both sexes | True | True |
| C4Quan | 6 | HCG27 | protein_coding | rs3132510 | 31172151 | 6.98e-15 | 79 | 0 | 0 | 1 | 6 | 72 | Both sexes | Both sexes | Both sexes | True | True |
| C4Quan | 6 | XXbac-BPG16N22.5 | pseudogene | rs2516491 | 31485101 | 4.67e-14 | 5 | 0 | 0 | 0 | 0 | 5 | Both sexes | Both sexes | Both sexes | True | True |
| C4Quan | 6 | HLA-DRA | protein_coding | rs3129891 | 32415080 | 5.57e-14 | 119 | 0 | 0 | 0 | 0 | 119 | Both sexes | Both sexes | Both sexes | True | True |
| C4Quan | 6 | XXbac-BPG32J3.20 | protein_coding | rs805286 | 31679307 | 6.34e-14 | 33 | 22 | 1 | 3 | 0 | 7 | Female only | Both sexes | Female only | True | True |
| C4Quan | 6 | MSH5 | protein_coding | rs409558 | 31708147 | 1.09e-13 | 57 | 4 | 6 | 13 | 4 | 30 | Both sexes | Both sexes | Both sexes | True | True |
| C4Quan | 6 | SLC44A4 | protein_coding | rs521977 | 31836827 | 1.44e-13 | 39 | 11 | 0 | 0 | 12 | 16 | Both sexes | Both sexes | Both sexes | True | True |
| C4Quan | 6 | XXbac-BPG248L24.10 | pseudogene | rs9461693 | 31291932 | 2.09e-13 | 95 | 0 | 0 | 0 | 1 | 94 | Female only | Both sexes | Female only | True | True |
| C4Quan | 6 | HLA-C | protein_coding | rs2524096 | 31236467 | 2.64e-13 | 19 | 2 | 0 | 2 | 2 | 13 | Both sexes | Both sexes | Female only | True | True |
| C4Quan | 6 | HLA-B | protein_coding | rs2854004 | 31313972 | 4.47e-13 | 33 | 4 | 0 | 0 | 0 | 29 | Female only | Both sexes | Female only | True | True |
| C4Quan | 6 | VWA7 | protein_coding | rs6914412 | 31742242 | 4.61e-13 | 25 | 0 | 0 | 11 | 2 | 12 | Female only | Both sexes | Female only | True | True |
| C4Quan | 6 | AL671883.1 | miRNA | rs2844551 | 31342781 | 5.87e-13 | 117 | 0 | 1 | 28 | 20 | 68 | Both sexes | Both sexes | Both sexes | True | True |
| C4Quan | 6 | HLA-DRA | protein_coding | rs28366195 | 32393542 | 6.88e-13 | 17 | 0 | 0 | 4 | 5 | 8 | Female only | Both sexes | Female only | True | True |
| C4Quan | 6 | HLA-DQA1 | protein_coding | rs9271365 | 32586794 | 8.51e-13 | 13 | 6 | 0 | 1 | 2 | 4 | Both sexes | Both sexes | Female only | True | True |
| C4Quan | 6 | HLA-DRA | protein_coding | rs9268644 | 32408044 | 1.02e-12 | 123 | 1 | 1 | 0 | 0 | 121 | Both sexes | Both sexes | Both sexes | True | True |
| C4Quan | 6 | NOTCH4 | protein_coding | rs367398 | 32191730 | 1.15e-12 | 15 | 0 | 0 | 0 | 6 | 9 | Both sexes | Both sexes | Both sexes | True | True |
| C4Quan | 6 | HLA-DRA | protein_coding | rs732163 | 32394911 | 1.75e-12 | 11 | 0 | 0 | 0 | 2 | 9 | Female only | Both sexes | Female only | True | True |
| C4Quan | 6 | PSORS1C1 | protein_coding | rs3132561 | 31103931 | 2.24e-12 | 79 | 4 | 0 | 0 | 0 | 75 | Female only | Both sexes | Female only | True | True |
| C4Quan | 6 | TNXB | protein_coding | rs204886 | 32029415 | 2.37e-12 | 17 | 4 | 2 | 0 | 1 | 10 | Both sexes | Both sexes | Both sexes | True | True |
| C4Quan | 6 | C6orf15 | protein_coding | rs3095303 | 31070078 | 2.76e-12 | 123 | 0 | 0 | 5 | 57 | 61 | Both sexes | Both sexes | Female only | True | True |

| pheno | CHR | Gene_Symbol | biotype | lead_snp | lead_BP | lead_P | TOTAL | NSIG | S05 | S01 | S001 | S0001 | independent_loci_in_sex_P1e6 | independent_loci_in_sex_P1e2 | independent_loci_in_sex_P5e8 | in_gwas_catalog | in_HLA |
|---|---|---|---|---|---|---|---|---|---|---|---|---|---|---|---|---|---|
| C4Quan | 6 | AL671883.1 | miRNA | rs2844552 | 31342741 | 3.58e-12 | 85 | 2 | 19 | 17 | 34 | 13 | Both sexes | Both sexes | Female only | True | True |
| C4Quan | 6 | TRIM10 | protein_coding | rs1970 | 30125543 | 4.62e-12 | 333 | 0 | 0 | 1 | 13 | 319 | Both sexes | Both sexes | Both sexes | True | True |
| C4Quan | 6 | MTCO3P1 | pseudogene | rs7774434 | 32657578 | 5.37e-12 | 163 | 56 | 56 | 25 | 1 | 25 | Female only | Both sexes | Female only | True | True |
| C4Quan | 6 | HCP5 | sense_overlapping | rs2284178 | 31432125 | 6.47e-12 | 3 | 0 | 0 | 0 | 2 | 1 | Female only | Both sexes | Female only | True | True |
| C4Quan | 6 | XXbac-BPG248L24.12 | pseudogene | rs12662617 | 31328685 | 8.70e-12 | 7 | 3 | 0 | 0 | 3 | 1 | Female only | Both sexes | Female only | True | True |
| C4Quan | 6 | BRD2 | protein_coding | rs78881806 | 32952168 | 1.01e-11 | 3 | 0 | 0 | 1 | 1 | 1 | Female only | Both sexes | Female only | True | True |
| C4Quan | 6 | DHFRP2 | pseudogene | rs2844572 | 31335575 | 1.28e-11 | 33 | 7 | 1 | 4 | 0 | 21 | Both sexes | Both sexes | Female only | True | True |
| C4Quan | 6 | HLA-S | pseudogene | rs2844535 | 31350303 | 1.46e-11 | 3 | 0 | 1 | 1 | 0 | 1 | Female only | Both sexes | Female only | True | True |
| C4Quan | 6 | HLA-S | pseudogene | rs2507978 | 31351664 | 1.56e-11 | 11 | 4 | 1 | 5 | 0 | 1 | Female only | Both sexes | Female only | True | True |
| C4Quan | 6 | PSORS1C1 | protein_coding | rs3130552 | 31082127 | 1.63e-11 | 37 | 0 | 0 | 0 | 1 | 36 | Both sexes | Both sexes | Both sexes | True | True |
| C4Quan | 6 | USP8P1 | pseudogene | rs2844620 | 31242151 | 1.76e-11 | 59 | 0 | 0 | 0 | 8 | 51 | Both sexes | Both sexes | Female only | True | True |
| C4Quan | 6 | HLA-B | protein_coding | rs1634775 | 31303236 | 2.44e-11 | 87 | 2 | 0 | 0 | 5 | 80 | Both sexes | Both sexes | Female only | True | True |
| C4Quan | 6 | SUMO2P1 | pseudogene | rs3131856 | 29607101 | 2.91e-11 | 137 | 0 | 0 | 0 | 13 | 124 | Both sexes | Both sexes | Both sexes | True | True |
| C4Quan | 6 | FLOT1 | protein_coding | rs3095327 | 30699022 | 2.98e-11 | 221 | 0 | 2 | 2 | 11 | 206 | Both sexes | Both sexes | Both sexes | True | True |
| C4Quan | 6 | TRIM10 | protein_coding | rs2523734 | 30129676 | 3.38e-11 | 23 | 1 | 10 | 1 | 0 | 11 | Female only | Both sexes | Female only | True | True |
| C4Quan | 6 | HCP5 | sense_overlapping | rs116198852 | 31393014 | 3.60e-11 | 43 | 1 | 0 | 0 | 21 | 21 | Male only | Both sexes | Male only | True | True |
| C4Quan | 6 | XXbac-BPG154L12.4 | antisense | rs1894374 | 32214435 | 4.20e-11 | 57 | 1 | 14 | 0 | 0 | 42 | Female only | Both sexes | Female only | True | True |
| C4Quan | 6 | MICB | protein_coding | rs2516497 | 31465905 | 6.97e-11 | 23 | 0 | 0 | 9 | 3 | 11 | Female only | Both sexes | Female only | True | True |
| C4Quan | 6 | MTCO3P1 | pseudogene | rs4538748 | 32657505 | 6.99e-11 | 177 | 141 | 27 | 0 | 1 | 8 | Female only | Both sexes | Female only | True | True |
| C4Quan | 6 | HLA-DQB1 | protein_coding | rs9274728 | 32637772 | 7.67e-11 | 37 | 28 | 0 | 2 | 1 | 6 | Female only | Both sexes | Female only | True | True |
| C4Quan | 6 | FKBPL | protein_coding | rs28732177 | 32100877 | 9.11e-11 | 3 | 0 | 0 | 0 | 1 | 2 | Female only | Both sexes | Female only | True | True |
| C4Quan | 6 | DHFRP2 | pseudogene | rs2523558 | 31331037 | 9.94e-11 | 7 | 0 | 0 | 0 | 0 | 7 | Female only | Both sexes | Female only | True | True |
| C4Quan | 6 | HLA-DRA | protein_coding | rs3129879 | 32408907 | 1.09e-10 | 13 | 0 | 0 | 0 | 3 | 10 | Both sexes | Both sexes | Male only | True | True |
| C4Quan | 6 | XXbac-BPG32J3.20 | protein_coding | rs28366156 | 31671498 | 1.35e-10 | 17 | 8 | 1 | 0 | 0 | 8 | Female only | Female only | Female only | True | True |
| C4Quan | 6 | HLA-B | protein_coding | rs2257269 | 31323353 | 1.38e-10 | 3 | 0 | 2 | 0 | 0 | 1 | Both sexes | Both sexes | Female only | True | True |

| pheno | CHR | Gene_Symbol | biotype | lead_snp | lead_BP | lead_P | TOTAL | NSIG | S05 | S01 | S001 | S0001 | independent_loci_in_sex_P1e6 | independent_loci_in_sex_P1e2 | independent_loci_in_sex_P5e8 | in_gwas_catalog | in_HLA |
|---|---|---|---|---|---|---|---|---|---|---|---|---|---|---|---|---|---|
| C4Quan | 6 | BTNL2 | protein_coding | rs3806156 | 32373698 | 1.82e-10 | 21 | 0 | 0 | 11 | 0 | 10 | Female only | Both sexes | Female only | True | True |
| C4Quan | 6 | HLA-B | protein_coding | rs28380936 | 31320121 | 2.03e-10 | 5 | 0 | 0 | 3 | 0 | 2 | Female only | Both sexes | Female only | True | True |
| pheno | CHR | Gene_Symbol | biotype | lead_snp | lead_BP | lead_P | TOTAL | NSIG | S05 | S01 | S001 | S0001 | independent_loci_in_sex_P1e6 | independent_loci_in_sex_P1e2 | independent_loci_in_sex_P5e8 | in_gwas_catalog | in_HLA |
| C4Quan | 6 | XXbac-BPG181B23.7 | lincRNA | rs2905713 | 31356702 | 2.11e-10 | 243 | 23 | 9 | 0 | 3 | 208 | Female only | Both sexes | Female only | True | True |
| C4Quan | 6 | NOTCH4 | protein_coding | rs377763 | 32199144 | 2.66e-10 | 5 | 0 | 0 | 0 | 0 | 5 | Both sexes | Both sexes | Both sexes | True | True |
| C4Quan | 6 | HCP5 | sense_overlapping | rs1055568 | 31440006 | 4.75e-10 | 23 | 0 | 3 | 3 | 0 | 17 | Both sexes | Both sexes | Female only | True | True |
| C4Quan | 6 | SKIV2L | protein_coding | rs429608 | 31930462 | 5.41e-10 | 3 | 0 | 0 | 1 | 1 | 1 | Female only | Both sexes | Female only | True | True |
| C4Quan | 6 | HLA-Z | pseudogene | rs56098412 | 32879471 | 5.68e-10 | 35 | 0 | 0 | 0 | 0 | 35 | Both sexes | Both sexes | Both sexes | True | True |
| C4Quan | 6 | HLA-DOB | protein_coding | rs9276710 | 32757102 | 6.08e-10 | 7 | 0 | 0 | 0 | 0 | 7 | Both sexes | Both sexes | Both sexes | True | True |
| C4Quan | 6 | ZDHHC20P2 | pseudogene | rs2244027 | 31347566 | 7.59e-10 | 5 | 0 | 1 | 1 | 0 | 3 | Female only | Both sexes | Female only | True | True |
| C4Quan | 6 | NOTCH4 | protein_coding | rs3830041 | 32191339 | 1.10e-09 | 11 | 0 | 0 | 0 | 0 | 11 | Both sexes | Both sexes | Female only | True | True |
| C4Quan | 6 | HCP5 | sense_overlapping | rs2523454 | 31367865 | 1.10e-09 | 11 | 0 | 0 | 0 | 0 | 11 | Both sexes | Both sexes | Both sexes | True | True |
| C4Quan | 6 | HLA-DQB1 | protein_coding | rs9273363 | 32626272 | 1.65e-09 | 5 | 0 | 0 | 0 | 3 | 2 | Female only | Both sexes | Female only | True | True |
| C4Quan | 6 | PSORS1C1 | protein_coding | rs6906565 | 31097814 | 1.78e-09 | 119 | 1 | 12 | 31 | 15 | 60 | Female only | Both sexes | Female only | True | True |
| C4Quan | 6 | RNF5 | protein_coding | rs204996 | 32149883 | 1.96e-09 | 17 | 3 | 1 | 5 | 0 | 8 | Female only | Both sexes | Female only | True | True |
| C4Quan | 6 | BRD2 | protein_coding | rs188245 | 32955976 | 2.05e-09 | 73 | 42 | 16 | 13 | 0 | 2 | Female only | Both sexes | Female only | True | True |
| C4Quan | 6 | TMPOP1 | pseudogene | rs3132616 | 30416456 | 2.23e-09 | 19 | 0 | 0 | 1 | 1 | 17 | Both sexes | Both sexes | Both sexes | True | True |
| C4Quan | 6 | XXbac-BPG248L24.12 | pseudogene | rs115860766 | 31328860 | 2.34e-09 | 29 | 0 | 0 | 0 | 1 | 28 | Female only | Both sexes | Female only | True | True |
| C4Quan | 6 | MICA | protein_coding | rs73400361 | 31376266 | 2.61e-09 | 9 | 0 | 0 | 0 | 0 | 9 | Both sexes | Both sexes | Female only | True | True |
| C4Quan | 6 | XXbac-BPG154L12.4 | antisense | rs447459 | 32208673 | 2.75e-09 | 5 | 0 | 1 | 0 | 2 | 2 | Female only | Both sexes | Female only | True | True |
| C4Quan | 6 | MUC22 | protein_coding | rs1634718 | 30972865 | 3.34e-09 | 29 | 0 | 1 | 4 | 7 | 17 | Both sexes | Both sexes | Both sexes | True | True |
| C4Quan | 6 | HLA-A | protein_coding | rs9260085 | 29909360 | 3.79e-09 | 251 | 98 | 20 | 8 | 2 | 123 | Female only | Both sexes | Female only | True | True |
| C4Quan | 6 | C6orf100 | protein_coding | rs3131073 | 28920972 | 4.08e-09 | 323 | 3 | 0 | 11 | 18 | 291 | Both sexes | Both sexes | Female only | True | False |
| C4Quan | 6 | C6orf48 | protein_coding | rs41284523 | 31810266 | 4.85e-09 | 19 | 8 | 1 | 0 | 1 | 9 | Female only | Both sexes | Female only | True | True |
| C4Quan | 6 | OR5V1 | protein_coding | rs442694 | 29356687 | 6.21e-09 | 51 | 0 | 0 | 7 | 9 | 35 | Both sexes | Both sexes | Female only | True | False |

| pheno | CHR | Gene_Symbol | biotype | lead_snp | lead_BP | lead_P | TOTAL | NSIG | S05 | S01 | S001 | S0001 | independent_loci_in_sex_P1e6 | independent_loci_in_sex_P1e2 | independent_loci_in_sex_P5e8 | in_gwas_catalog | in_HLA |
|---|---|---|---|---|---|---|---|---|---|---|---|---|---|---|---|---|---|
| C4Quan | 6 | HCG23 | antisense | rs17208671 | 32360849 | 6.54e-09 | 209 | 104 | 104 | 1 | 0 | 0 | Female only | Female only | Female only | True | True |
| C4Quan | 6 | XXbac-BPG13B8.10 | lincRNA | rs1233478 | 29477821 | 7.26e-09 | 3 | 0 | 0 | 2 | 0 | 1 | Female only | Both sexes | Female only | True | False |
| C4Quan | 6 | HLA-S | pseudogene | rs2523484 | 31353639 | 8.48e-09 | 145 | 1 | 2 | 107 | 0 | 35 | Both sexes | Both sexes | Female only | True | True |
| C4Quan | 6 | HLA-DQA2 | protein_coding | rs9276427 | 32711857 | 8.79e-09 | 279 | 0 | 3 | 89 | 22 | 165 | Female only | Both sexes | Female only | True | True |
| C4Quan | 6 | HLA-B | protein_coding | rs9265988 | 31316387 | 8.95e-09 | 39 | 12 | 0 | 1 | 3 | 23 | Female only | Both sexes | Female only | True | True |
| C4Quan | 6 | MICB | protein_coding | rs2253044 | 31461925 | 9.95e-09 | 69 | 0 | 0 | 0 | 13 | 56 | Female only | Both sexes | Female only | True | True |
| C4Quan | 6 | HCG9 | lincRNA | rs2246199 | 29945620 | 1.02e-08 | 315 | 0 | 5 | 156 | 137 | 17 | Female only | Both sexes | Female only | True | True |
| C4Quan | 6 | ZSCAN31 | protein_coding | rs13198809 | 28323702 | 1.04e-08 | 187 | 0 | 0 | 0 | 19 | 168 | Both sexes | Both sexes | Female only | True | False |
| C4Quan | 6 | C6orf10 | protein_coding | rs3117110 | 32340176 | 1.17e-08 | 5 | 0 | 0 | 0 | 0 | 5 | Female only | Both sexes | Female only | True | True |
| C4Quan | 6 | MICB | protein_coding | rs1052416 | 31456769 | 1.33e-08 | 7 | 0 | 0 | 0 | 1 | 6 | Female only | Both sexes | Female only | True | True |
| C4Quan | 6 | HLA-S | pseudogene | rs9266772 | 31352113 | 2.43e-08 | 49 | 25 | 2 | 0 | 0 | 22 | Female only | Female only | Female only | True | True |
| C4Quan | 6 | HCG17 | lincRNA | rs3130396 | 30223490 | 2.91e-08 | 145 | 111 | 17 | 17 | 0 | 0 | Female only | Female only | Female only | True | True |
| C4Quan | 6 | HLA-DRB9 | pseudogene | rs9269171 | 32447120 | 3.10e-08 | 19 | 0 | 0 | 0 | 0 | 19 | Male only | Both sexes | Male only | True | True |
| C4Quan | 6 | LINC00533 | lincRNA | rs34724414 | 28619353 | 3.42e-08 | 17 | 0 | 0 | 6 | 6 | 5 | Female only | Both sexes | Female only | True | False |
| C4Quan | 6 | RN7SKP186 | misc_RNA | rs2844657 | 30829522 | 3.71e-08 | 21 | 0 | 0 | 1 | 1 | 19 | Female only | Both sexes | Female only | True | True |
| C4Quan | 6 | HCG17 | lincRNA | rs2516698 | 30232374 | 3.78e-08 | 63 | 14 | 18 | 13 | 0 | 18 | Female only | Female only | Female only | True | True |
| C4Quan | 6 | C6orf10 | protein_coding | rs2050190 | 32339076 | 3.87e-08 | 3 | 0 | 0 | 0 | 0 | 3 | Female only | Both sexes | Female only | True | True |
| C4Quan | 6 | HCP5 | sense_overlapping | rs2516452 | 31427095 | 4.16e-08 | 3 | 0 | 1 | 1 | 0 | 1 | Female only | Both sexes | Female only | True | True |
| C4Quan | 6 | HCG20 | lincRNA | rs4713369 | 30761129 | 4.19e-08 | 139 | 21 | 3 | 46 | 1 | 68 | Female only | Both sexes | Female only | True | True |
| C4Quan | 6 | NOTCH4 | protein_coding | rs206015 | 32182759 | 4.40e-08 | 31 | 25 | 0 | 3 | 1 | 2 | Female only | Both sexes | Female only | True | True |
| CBil | 2 | UGT1A8 | protein_coding | rs4148325 | 234673309 | 1.12e-40 | 219 | 0 | 0 | 0 | 0 | 219 | Both sexes | Both sexes | Both sexes | True | False |
| CBil | 2 | UGT1A8 | protein_coding | rs2012734 | 234622429 | 3.11e-21 | 15 | 0 | 0 | 0 | 0 | 15 | Both sexes | Both sexes | Both sexes | True | False |
| CBil | 2 | UGT1A8 | protein_coding | rs7592624 | 234602906 | 6.90e-14 | 35 | 0 | 0 | 0 | 0 | 35 | Both sexes | Both sexes | Both sexes | True | False |
| CBil | 2 | UGT1A8 | protein_coding | rs6751673 | 234604903 | 2.76e-11 | 31 | 0 | 0 | 12 | 13 | 6 | Both sexes | Both sexes | Female only | True | False |
| CO2 | 2 | SLC9A4 | protein_coding | rs76229479 | 103123912 | 2.21e-12 | 155 | 0 | 0 | 21 | 57 | 77 | Female only | Both sexes | Female only | True | False |
| CPK | 19 | LILRB5 | protein_coding | rs12975366 | 54759361 | 3.58e-15 | 7 | 0 | 2 | 1 | 1 | 3 | Female only | Both sexes | Female only | True | False |

| pheno | CHR | Gene_Symbol | biotype | lead_snp | lead_BP | lead_P | TOTAL | NSIG | S05 | S01 | S001 | S0001 | independent_loci_in_sex_P1e6 | independent_loci_in_sex_P1e2 | independent_loci_in_sex_P5e8 | in_gwas_catalog | in_HLA |
|---|---|---|---|---|---|---|---|---|---|---|---|---|---|---|---|---|---|
| CPK | 19 | RNU6-1307P | snRNA | rs12984962 | 54769205 | 1.06e-09 | 9 | 5 | 0 | 0 | 0 | 4 | Female only | Female only | Female only | True | False |
| CRP | 19 | APOE | protein_coding | rs429358 | 45411941 | 4.47e-08 | 39 | 0 | 0 | 4 | 14 | 21 | Both sexes | Both sexes | Female only | True | False |
| Ca | 3 | CASR | protein_coding | rs73186030 | 122013465 | 4.18e-49 | 95 | 0 | 0 | 0 | 0 | 95 | Both sexes | Both sexes | Both sexes | True | False |
| Ca | 3 | KPNA1 | protein_coding | rs6803240 | 122172361 | 1.26e-32 | 43 | 0 | 0 | 0 | 0 | 43 | Both sexes | Both sexes | Both sexes | True | False |
| Ca | 3 | KPNA1 | protein_coding | rs55961674 | 122196892 | 4.89e-28 | 23 | 0 | 0 | 0 | 0 | 23 | Both sexes | Both sexes | Both sexes | True | False |
| Ca | 3 | CASR | protein_coding | rs3749208 | 121980284 | 3.36e-26 | 27 | 0 | 0 | 0 | 0 | 27 | Both sexes | Both sexes | Both sexes | True | False |
| Ca | 3 | HNRNPA1P23 | pseudogene | rs77913677 | 122038460 | 5.59e-26 | 3 | 0 | 0 | 0 | 0 | 3 | Both sexes | Both sexes | Both sexes | True | False |
| Ca | 3 | CASR | protein_coding | rs3749206 | 121976547 | 5.05e-21 | 57 | 0 | 0 | 0 | 0 | 57 | Both sexes | Both sexes | Both sexes | True | False |
| Ca | 3 | CCDC58 | protein_coding | rs9863881 | 122100023 | 4.84e-17 | 71 | 37 | 9 | 0 | 0 | 25 | Both sexes | Both sexes | Female only | True | False |
| Ca | 3 | HNRNPA1P23 | pseudogene | rs4678180 | 122037797 | 2.84e-13 | 13 | 0 | 0 | 0 | 0 | 13 | Both sexes | Both sexes | Both sexes | True | False |
| Ca | 3 | HNRNPA1P23 | pseudogene | rs2001547 | 122032834 | 4.29e-13 | 71 | 2 | 0 | 0 | 0 | 69 | Both sexes | Both sexes | Female only | True | False |
| Ca | 3 | CASR | protein_coding | rs6800130 | 121873858 | 8.12e-12 | 153 | 1 | 58 | 0 | 5 | 89 | Both sexes | Both sexes | Both sexes | True | False |

| pheno | CHR | Gene_Symbol | biotype | lead_snp | lead_BP | lead_P | TOTAL | NSIG | S05 | S01 | S001 | S0001 | independent_loci_in_sex_P1e6 | independent_loci_in_sex_P1e2 | independent_loci_in_sex_P5e8 | in_gwas_catalog | in_HLA |
|---|---|---|---|---|---|---|---|---|---|---|---|---|---|---|---|---|---|
| Ca | 3 | CSTA | protein_coding | rs12054451 | 122064369 | 9.35e-12 | 5 | 0 | 0 | 0 | 0 | 5 | Both sexes | Both sexes | Both sexes | True | False |
| Ca | 9 | GNAQ | protein_coding | rs13291723 | 80510077 | 1.72e-10 | 107 | 53 | 1 | 0 | 0 | 53 | Female only | Female only | Female only | True | False |
| Ca | 2 | DGKD | protein_coding | rs838718 | 234296650 | 2.98e-09 | 27 | 0 | 0 | 4 | 10 | 13 | Female only | Both sexes | Female only | True | False |
| Ca | 2 | AC079610.1 | sense_overlapping | rs13035592 | 213733958 | 1.27e-08 | 35 | 18 | 0 | 0 | 0 | 17 | Male only | Male only | Male only | True | False |
| Ca | 9 | GNAQ | protein_coding | rs2026000 | 80334981 | 2.29e-08 | 67 | 3 | 30 | 1 | 1 | 32 | Female only | Both sexes | Female only | True | False |
| Ca | 2 | PPM1G | protein_coding | rs1728918 | 27635463 | 2.49e-08 | 7 | 0 | 0 | 1 | 2 | 4 | Male only | Both sexes | Male only | True | False |
| Ca | 3 | CASR | protein_coding | rs144486035 | 121921137 | 3.22e-08 | 37 | 0 | 0 | 11 | 8 | 18 | Female only | Both sexes | Female only | True | False |
| Ca | 3 | CASR | protein_coding | rs1042636 | 122003769 | 3.96e-08 | 69 | 0 | 0 | 1 | 35 | 33 | Female only | Both sexes | Female only | True | False |
| Ca | 9 | RP11-205K6.1 | lincRNA | rs4837096 | 129276052 | 4.67e-08 | 19 | 10 | 0 | 0 | 8 | 1 | Female only | Female only | Female only | True | False |
| Calo | 3 | CASR | protein_coding | rs73186030 | 122013465 | 2.71e-10 | 95 | 0 | 1 | 1 | 45 | 48 | Male only | Both sexes | Male only | True | False |
| Chol | 19 | APOE | protein_coding | rs7412 | 45412079 | 6.49e-62 | 9 | 0 | 0 | 0 | 0 | 9 | Both sexes | Both sexes | Both sexes | True | False |
| Chol | 19 | APOE | protein_coding | rs429358 | 45411941 | 3.49e-27 | 39 | 9 | 2 | 2 | 3 | 23 | Both sexes | Both sexes | Both sexes | True | False |
| Chol | 1 | CELSR2 | protein_coding | rs12740374 | 109817590 | 5.15e-20 | 27 | 0 | 0 | 0 | 2 | 25 | Female only | Both sexes | Female only | True | False |

| pheno | CHR | Gene_Symbol | biotype | lead_snp | lead_BP | lead_P | TOTAL | NSIG | S05 | S01 | S001 | S0001 | independent_loci_in_sex_P1e6 | independent_loci_in_sex_P1e2 | independent_loci_in_sex_P5e8 | in_gwas_catalog | in_HLA |
|---|---|---|---|---|---|---|---|---|---|---|---|---|---|---|---|---|---|
| Chol | 19 | PVRL2 | protein_coding | 19:45384405 | 45384405 | 1.28e-17 | 3 | 2 | 0 | 1 | 0 | 0 | Female only | Both sexes | Female only | True | False |
| Chol | 2 | APOB | protein_coding | rs581411 | 21289432 | 5.95e-17 | 355 | 0 | 6 | 32 | 120 | 197 | Female only | Both sexes | Female only | True | False |
| Chol | 19 | PVRL2 | protein_coding | rs34342646 | 45388130 | 9.61e-16 | 3 | 0 | 1 | 0 | 1 | 1 | Female only | Both sexes | Female only | True | False |
| Chol | 19 | TOMM40 | protein_coding | rs157580 | 45395266 | 2.15e-15 | 7 | 3 | 3 | 1 | 0 | 0 | Female only | Both sexes | Female only | True | False |
| Chol | 19 | BCL3 | protein_coding | rs62117205 | 45252666 | 2.38e-15 | 13 | 0 | 2 | 5 | 0 | 6 | Female only | Both sexes | Female only | True | False |
| Chol | 2 | TDRD15 | protein_coding | rs12997242 | 21381177 | 4.66e-15 | 103 | 11 | 28 | 11 | 2 | 51 | Female only | Both sexes | Female only | True | False |
| Chol | 19 | PVRL2 | protein_coding | rs2927472 | 45349369 | 1.00e-14 | 9 | 3 | 2 | 0 | 0 | 4 | Female only | Female only | Female only | True | False |
| Chol | 2 | APOB | protein_coding | rs934197 | 21267461 | 7.06e-14 | 15 | 7 | 0 | 0 | 4 | 4 | Female only | Both sexes | Female only | True | False |
| Chol | 19 | LDLR | protein_coding | rs77140532 | 11192831 | 1.04e-12 | 75 | 0 | 0 | 0 | 0 | 75 | Both sexes | Both sexes | Both sexes | True | False |
| Chol | 2 | RP11-116D2.1 | lincRNA | rs952275 | 21221399 | 1.73e-12 | 9 | 0 | 1 | 4 | 0 | 4 | Female only | Both sexes | Female only | True | False |
| Chol | 19 | BCAM | protein_coding | rs10412413 | 45327309 | 3.46e-12 | 13 | 0 | 1 | 6 | 0 | 6 | Female only | Both sexes | Female only | True | False |
| Chol | 19 | TM6SF2 | protein_coding | rs58542926 | 19379549 | 1.19e-11 | 51 | 0 | 0 | 7 | 16 | 28 | Male only | Both sexes | Male only | True | False |
| Chol | 19 | CILP2 | protein_coding | rs17216525 | 19662220 | 1.14e-10 | 49 | 0 | 0 | 3 | 22 | 24 | Male only | Both sexes | Male only | True | False |
| Chol | 1 | PSRC1 | protein_coding | rs11577931 | 109820884 | 1.89e-10 | 11 | 0 | 5 | 1 | 0 | 5 | Female only | Both sexes | Female only | True | False |
| Chol | 6 | BTNL2 | protein_coding | rs28732208 | 32385119 | 6.87e-10 | 597 | 1 | 62 | 233 | 3 | 298 | Female only | Both sexes | Female only | True | True |
| Chol | 2 | AC067959.1 | lincRNA | rs13387827 | 21469152 | 8.95e-10 | 15 | 1 | 1 | 6 | 0 | 7 | Female only | Both sexes | Female only | True | False |
| Chol | 2 | AC067959.1 | lincRNA | rs13420469 | 21449034 | 2.07e-09 | 43 | 4 | 14 | 4 | 3 | 18 | Female only | Both sexes | Female only | True | False |
| Chol | 2 | APOB | protein_coding | rs3791981 | 21245367 | 5.04e-09 | 15 | 3 | 4 | 1 | 0 | 7 | Female only | Both sexes | Female only | True | False |
| Chol | 2 | GCKR | protein_coding | rs1260326 | 27730940 | 5.11e-09 | 33 | 7 | 0 | 5 | 11 | 10 | Female only | Both sexes | Female only | True | False |
| Chol | 19 | APOC1 | protein_coding | rs59325138 | 45416291 | 5.36e-09 | 13 | 4 | 1 | 2 | 0 | 6 | Female only | Both sexes | Female only | True | False |
| Chol | 6 | BTNL2 | protein_coding | rs72847932 | 32362085 | 1.87e-08 | 125 | 0 | 6 | 10 | 48 | 61 | Female only | Both sexes | Female only | True | True |
| Cl | 11 | BDNF-AS | antisense | rs10742179 | 27650524 | 3.91e-08 | 115 | 4 | 0 | 36 | 32 | 43 | Male only | Both sexes | Male only | True | False |
| Creat | 16 | UMOD | protein_coding | rs36060036 | 20361950 | 5.86e-14 | 53 | 0 | 0 | 0 | 0 | 53 | Both sexes | Both sexes | Both sexes | True | False |
| Creat | 7 | PRKAG2 | protein_coding | rs10480300 | 151406005 | 9.46e-11 | 19 | 0 | 0 | 0 | 0 | 19 | Both sexes | Both sexes | Both sexes | True | False |
| Creat | 6 | L3MBTL3 | protein_coding | rs6926186 | 130350294 | 3.42e-08 | 125 | 63 | 0 | 0 | 0 | 62 | Female only | Female only | Female only | True | False |
| D25OHT | 11 | PDE3B | protein_coding | rs1037378 | 14852490 | 6.33e-22 | 125 | 0 | 0 | 0 | 17 | 108 | Both sexes | Both sexes | Both sexes | True | False |

| pheno | CHR | Gene_Symbol | biotype | lead_snp | lead_BP | lead_P | TOTAL | NSIG | S05 | S01 | S001 | S0001 | independent_loci_in_sex_P1e6 | independent_loci_in_sex_P1e2 | independent_loci_in_sex_P5e8 | in_gwas_catalog | in_HLA |
|---|---|---|---|---|---|---|---|---|---|---|---|---|---|---|---|---|---|
| D25OHT | 4 | GC | protein_coding | rs1352846 | 72617775 | 3.64e-21 | 39 | 0 | 0 | 0 | 0 | 39 | Both sexes | Both sexes | Both sexes | True | False |
| D25OHT | 11 | CALCB | protein_coding | rs10766197 | 14921880 | 3.49e-18 | 5 | 0 | 0 | 0 | 0 | 5 | Both sexes | Both sexes | Both sexes | True | False |
| D25OHT | 11 | COPB1 | protein_coding | rs12792120 | 14469443 | 4.46e-15 | 83 | 0 | 0 | 0 | 2 | 81 | Both sexes | Both sexes | Both sexes | True | False |
| D25OHT | 4 | GC | protein_coding | rs222035 | 72621674 | 7.22e-15 | 7 | 0 | 0 | 0 | 0 | 7 | Both sexes | Both sexes | Both sexes | True | False |
| D25OHT | 11 | RRAS2 | protein_coding | rs11606558 | 14328689 | 3.41e-13 | 51 | 0 | 1 | 12 | 13 | 25 | Female only | Both sexes | Female only | True | False |
| D25OHT | 11 | CYP2R1 | protein_coding | rs11023374 | 14903636 | 1.28e-09 | 3 | 0 | 0 | 0 | 0 | 3 | Both sexes | Both sexes | Female only | True | False |
| D25OHT | 4 | GC | protein_coding | rs1526692 | 72578724 | 2.44e-08 | 11 | 0 | 0 | 0 | 0 | 11 | Both sexes | Both sexes | Male only | True | False |
| DLCOOVA_PRD | 12 | LINC00942 | lincRNA | rs4765821 | 1611846 | 2.06e-08 | 43 | 20 | 2 | 0 | 1 | 20 | Female only | Female only | Female only | False | False |
| DirLDL | 19 | APOE | protein_coding | rs7412 | 45412079 | 4.97e-13 | 9 | 0 | 0 | 0 | 0 | 9 | Both sexes | Both sexes | Both sexes | True | False |
| EGFR | 16 | UMOD | protein_coding | rs36060036 | 20361950 | 2.00e-13 | 53 | 0 | 0 | 0 | 0 | 53 | Both sexes | Both sexes | Both sexes | True | False |
| EGFR | 7 | PRKAG2 | protein_coding | rs10480300 | 151406005 | 6.15e-13 | 19 | 0 | 0 | 0 | 0 | 19 | Both sexes | Both sexes | Both sexes | True | False |
| EGFR | 15 | CTD-2651B20.3 | antisense | rs8039476 | 45589161 | 1.65e-08 | 361 | 0 | 0 | 0 | 1 | 360 | Both sexes | Both sexes | Male only | True | False |
| EOSIA | 3 | RPN1 | protein_coding | rs6782812 | 128317997 | 5.98e-10 | 53 | 0 | 1 | 2 | 3 | 47 | Female only | Both sexes | Female only | True | False |
| EOSIAB | 3 | RPN1 | protein_coding | rs6782812 | 128317997 | 8.14e-10 | 53 | 0 | 2 | 1 | 11 | 39 | Both sexes | Both sexes | Female only | True | False |
| EOSIM | 3 | RPN1 | protein_coding | rs6782812 | 128317997 | 8.53e-12 | 53 | 0 | 0 | 0 | 0 | 53 | Both sexes | Both sexes | Both sexes | True | False |
| EOSIRE | 3 | RPN1 | protein_coding | rs6782812 | 128317997 | 3.45e-11 | 53 | 0 | 1 | 1 | 11 | 40 | Both sexes | Both sexes | Female only | True | False |
| EOSIRE | 5 | RAD50 | protein_coding | rs60156285 | 131955818 | 1.44e-08 | 41 | 2 | 16 | 3 | 0 | 20 | Male only | Both sexes | Male only | True | False |
| EoAbs | 3 | C3orf27 | protein_coding | rs4328821 | 128316435 | 8.57e-14 | 53 | 0 | 0 | 0 | 1 | 52 | Both sexes | Both sexes | Female only | True | False |
| EoAbs | 4 | KLF3 | protein_coding | rs4535339 | 38689228 | 9.75e-11 | 101 | 9 | 2 | 40 | 0 | 50 | Female only | Both sexes | Female only | True | False |
| EoAbs | 2 | AC007248.6 | pseudogene | rs11692065 | 102883975 | 2.01e-09 | 155 | 1 | 15 | 1 | 9 | 129 | Female only | Both sexes | Female only | True | False |
| EoAbs | 5 | C5orf56 | protein_coding | rs2522052 | 131798487 | 6.65e-09 | 89 | 0 | 0 | 0 | 2 | 87 | Both sexes | Both sexes | Female only | True | False |
| pheno | CHR | Gene_Symbol | biotype | lead_snp | lead_BP | lead_P | TOTAL | NSIG | S05 | S01 | S001 | S0001 | independent_loci_in_sex_P1e6 | independent_loci_in_sex_P1e2 | independent_loci_in_sex_P5e8 | in_gwas_catalog | in_HLA |
| EoAbs | 5 | CSF2 | protein_coding | rs25884 | 131412238 | 1.37e-08 | 71 | 0 | 9 | 9 | 10 | 43 | Female only | Both sexes | Female only | True | False |
| EoAbs | 3 | C3orf27 | protein_coding | rs4640583 | 128278418 | 2.01e-08 | 41 | 0 | 0 | 0 | 5 | 36 | Both sexes | Both sexes | Male only | True | False |
| Eos | 3 | C3orf27 | protein_coding | rs4328821 | 128316435 | 8.52e-18 | 53 | 0 | 0 | 0 | 0 | 53 | Both sexes | Both sexes | Both sexes | True | False |

| pheno | CHR | Gene_Symbol | biotype | lead_snp | lead_BP | lead_P | TOTAL | NSIG | S05 | S01 | S001 | S0001 | independent_loci_in_sex_P1e6 | independent_loci_in_sex_P1e2 | independent_loci_in_sex_P5e8 | in_gwas_catalog | in_HLA |
|---|---|---|---|---|---|---|---|---|---|---|---|---|---|---|---|---|---|
| Eos | 3 | C3orf27 | protein_coding | rs77256197 | 128277551 | 1.73e-10 | 129 | 0 | 0 | 19 | 19 | 91 | Both sexes | Both sexes | Both sexes | True | False |
| Eos | 4 | KLF3 | protein_coding | rs4535339 | 38689228 | 6.63e-10 | 101 | 10 | 1 | 37 | 4 | 49 | Female only | Both sexes | Female only | True | False |
| Eos | 2 | IL1RL1 | protein_coding | rs11690644 | 102914214 | 2.45e-09 | 141 | 0 | 9 | 1 | 31 | 100 | Female only | Both sexes | Female only | True | False |
| Eos | 5 | CSF2 | protein_coding | rs25884 | 131412238 | 3.29e-08 | 71 | 5 | 7 | 18 | 21 | 20 | Female only | Both sexes | Female only | True | False |
| FT4 | 1 | DIO1 | protein_coding | rs2235544 | 54375570 | 2.80e-12 | 51 | 0 | 2 | 4 | 25 | 20 | Female only | Both sexes | Female only | True | False |
| FT4 | 9 | QSOX2 | protein_coding | rs3780190 | 139099073 | 3.31e-08 | 11 | 0 | 0 | 2 | 3 | 6 | Female only | Both sexes | Female only | True | False |
| GluBed | 10 | TCF7L2 | protein_coding | rs7903146 | 114758349 | 2.33e-08 | 47 | 0 | 0 | 0 | 0 | 47 | Both sexes | Both sexes | Male only | True | False |
| Gluc | 2 | SPC25 | protein_coding | rs560887 | 169763148 | 7.78e-25 | 39 | 0 | 0 | 0 | 0 | 39 | Both sexes | Both sexes | Both sexes | True | False |
| Gluc | 10 | TCF7L2 | protein_coding | rs7903146 | 114758349 | 1.80e-22 | 47 | 0 | 0 | 0 | 0 | 47 | Both sexes | Both sexes | Both sexes | True | False |
| Gluc | 7 | GCK | protein_coding | rs2908293 | 44209322 | 5.72e-19 | 5 | 0 | 0 | 0 | 2 | 3 | Female only | Both sexes | Female only | True | False |
| Gluc | 7 | GCK | protein_coding | rs3757840 | 44231216 | 9.99e-19 | 11 | 0 | 0 | 0 | 0 | 11 | Both sexes | Both sexes | Both sexes | True | False |
| Gluc | 7 | CAMK2B | protein_coding | rs878521 | 44255643 | 2.35e-18 | 27 | 0 | 0 | 0 | 0 | 27 | Both sexes | Both sexes | Both sexes | True | False |
| Gluc | 2 | ABCB11 | protein_coding | rs575671 | 169780818 | 2.10e-13 | 81 | 0 | 0 | 1 | 34 | 46 | Female only | Both sexes | Female only | True | False |
| Gluc | 2 | SPC25 | protein_coding | rs34177044 | 169754485 | 3.96e-12 | 13 | 0 | 0 | 0 | 0 | 13 | Both sexes | Both sexes | Female only | True | False |
| Gluc | 6 | HLA-DQB1 | protein_coding | rs9273368 | 32626475 | 5.73e-12 | 7 | 0 | 0 | 0 | 1 | 6 | Male only | Both sexes | Male only | True | True |
| Gluc | 7 | GCK | protein_coding | rs2971672 | 44205906 | 1.14e-11 | 11 | 0 | 0 | 1 | 1 | 9 | Female only | Both sexes | Female only | True | False |
| Gluc | 10 | TCF7L2 | protein_coding | rs11196175 | 114736614 | 3.42e-11 | 5 | 0 | 1 | 0 | 0 | 4 | Male only | Both sexes | Male only | True | False |
| Gluc | 3 | SLC2A2 | protein_coding | rs5398 | 170715830 | 2.60e-10 | 65 | 0 | 7 | 5 | 17 | 36 | Male only | Both sexes | Male only | True | False |
| Gluc | 8 | SLC30A8 | protein_coding | rs13266634 | 118184783 | 3.75e-09 | 13 | 0 | 0 | 0 | 0 | 13 | Female only | Both sexes | Female only | True | False |
| Gluc | 3 | ADCY5 | protein_coding | rs72964564 | 123054770 | 7.90e-09 | 41 | 0 | 0 | 0 | 0 | 41 | Both sexes | Both sexes | Both sexes | True | False |
| Gluc | 10 | TCF7L2 | protein_coding | rs6585201 | 114768783 | 2.92e-08 | 71 | 0 | 0 | 0 | 1 | 70 | Both sexes | Both sexes | Both sexes | True | False |
| Gluc | 7 | GCK | protein_coding | rs2268573 | 44190468 | 4.65e-08 | 9 | 1 | 0 | 1 | 4 | 3 | Female only | Both sexes | Female only | True | False |
| HDL.C | 16 | AC012181.1 | miRNA | rs247617 | 56990716 | 1.27e-88 | 39 | 0 | 0 | 0 | 0 | 39 | Both sexes | Both sexes | Both sexes | True | False |
| HDL.C | 16 | CETP | protein_coding | rs7499892 | 57006590 | 3.80e-71 | 13 | 0 | 0 | 0 | 0 | 13 | Both sexes | Both sexes | Both sexes | True | False |
| HDL.C | 16 | CETP | protein_coding | rs3816117 | 56996158 | 4.56e-66 | 3 | 0 | 0 | 0 | 0 | 3 | Both sexes | Both sexes | Both sexes | True | False |
| HDL.C | 16 | AC012181.1 | miRNA | rs12923459 | 56989830 | 6.56e-52 | 5 | 0 | 0 | 0 | 0 | 5 | Both sexes | Both sexes | Both sexes | True | False |

| phen o | CHR | Gene_Sy mbol | biotype | lead_snp | lead_BP | lead_P | TOT AL | NSIG | S05 | S01 | S001 | S000 1 | independent_loci_in_sex_P1 e6 | independent_loci_in_sex_P1 e2 | independent_loci_in_sex_P5 e8 | in_gwas_catal og | in_H LA |
|---|---|---|---|---|---|---|---|---|---|---|---|---|---|---|---|---|---|
| HDL. C | 16 | AC012181. 1 | miRNA | rs1244852 8 | 5698555 5 | 6.19e-3 9 | 7 | 0 | 0 | 0 | 0 | 7 | Both sexes | Both sexes | Both sexes | True | False |
| HDL. C | 8 | AC100802. 3 | antisense | rs1158490 89 | 1991237 0 | 1.10e-2 7 | 155 | 0 | 0 | 0 | 0 | 155 | Both sexes | Both sexes | Both sexes | True | False |
| HDL. C | 16 | CETP | protein_coding | rs289719 | 5700994 1 | 2.12e-2 5 | 5 | 0 | 0 | 0 | 0 | 5 | Both sexes | Both sexes | Both sexes | True | False |
| HDL. C | 8 | LPL | protein_coding | rs15285 | 1982466 7 | 6.04e-2 5 | 157 | 0 | 0 | 0 | 0 | 157 | Both sexes | Both sexes | Both sexes | True | False |
| HDL. C | 15 | ALDH1A2 | protein_coding | rs2070895 | 5872393 9 | 2.46e-2 2 | 31 | 0 | 0 | 0 | 0 | 31 | Both sexes | Both sexes | Female only | True | False |
| HDL. C | 15 | ALDH1A2 | protein_coding | rs3585302 1 | 5868064 3 | 3.23e-1 9 | 37 | 0 | 0 | 0 | 0 | 37 | Both sexes | Both sexes | Both sexes | True | False |
| HDL. C | 16 | CETP | protein_coding | rs291043 | 5701269 9 | 1.26e-1 6 | 9 | 0 | 0 | 0 | 0 | 9 | Female only | Both sexes | Female only | True | False |
| HDL. C | 16 | SLC12A3 | protein_coding | rs1292433 1 | 5695326 1 | 1.95e-1 4 | 67 | 0 | 0 | 31 | 12 | 24 | Female only | Both sexes | Female only | True | False |
| HDL. C | 8 | LPL | protein_coding | rs256 | 1981196 7 | 2.64e-1 4 | 23 | 0 | 0 | 0 | 0 | 23 | Both sexes | Both sexes | Both sexes | True | False |
| HDL. C | 8 | LPL | protein_coding | rs894210 | 1986584 3 | 2.71e-1 3 | 9 | 0 | 0 | 0 | 0 | 9 | Both sexes | Both sexes | Female only | True | False |
| HDL. C | 15 | ALDH1A2 | protein_coding | rs723967 | 5869630 0 | 4.66e-1 1 | 7 | 0 | 0 | 0 | 0 | 7 | Both sexes | Both sexes | Both sexes | True | False |
| HDL. C | 15 | ALDH1A2 | protein_coding | rs3512888 1 | 5869214 8 | 1.27e-1 0 | 29 | 0 | 0 | 0 | 0 | 29 | Both sexes | Both sexes | Both sexes | True | False |
| HDL. C | 8 | LPL | protein_coding | rs3469328 2 | 1987356 6 | 1.39e-1 0 | 149 | 112 | 0 | 0 | 0 | 37 | Both sexes | Both sexes | Both sexes | True | False |
| HDL. C | 16 | SLC12A3 | protein_coding | rs5595862 3 | 5695511 0 | 1.56e-1 0 | 15 | 0 | 0 | 2 | 6 | 7 | Female only | Both sexes | Female only | True | False |
| HDL. C | 19 | APOE | protein_coding | rs429358 | 4541194 1 | 7.65e-1 0 | 39 | 3 | 0 | 9 | 8 | 19 | Male only | Both sexes | Male only | True | False |
| HDL. C | 8 | LPL | protein_coding | rs1441771 | 1975730 0 | 1.22e-0 9 | 33 | 0 | 0 | 0 | 0 | 33 | Both sexes | Both sexes | Female only | True | False |
| HDL. C | 16 | AC012181. 1 | miRNA | rs6499863 | 5699201 7 | 2.92e-0 9 | 5 | 0 | 0 | 0 | 0 | 5 | Both sexes | Both sexes | Female only | True | False |
| HDL. C | 11 | SIK3 | protein_coding | rs6190341 9 | 1169572 19 | 3.64e-0 9 | 297 | 1 | 0 | 9 | 65 | 222 | Female only | Both sexes | Female only | True | False |
| HDL. C | 8 | CSGALNA CT1 | protein_coding | rs2862308 8 | 1958318 2 | 5.54e-0 9 | 7 | 1 | 3 | 0 | 0 | 3 | Male only | Male only | Male only | True | False |
| HDL. C | 16 | NLRC5 | protein_coding | rs7205459 | 5702517 1 | 5.73e-0 9 | 7 | 0 | 0 | 0 | 0 | 7 | Female only | Both sexes | Female only | True | False |
| HDL. C | 11 | APOA1 | protein_coding | rs1272103 0 | 1167052 78 | 1.21e-0 8 | 15 | 5 | 0 | 4 | 3 | 3 | Female only | Both sexes | Female only | True | False |
| HDL. C | 16 | SLC12A3 | protein_coding | rs1330667 7 | 5692619 5 | 2.15e-0 8 | 47 | 0 | 0 | 8 | 17 | 22 | Female only | Both sexes | Female only | True | False |
| HDL. C | 18 | SMUG1P1 | pseudogene | rs6507940 | 4718913 8 | 2.16e-0 8 | 7 | 1 | 1 | 2 | 2 | 1 | Female only | Both sexes | Female only | True | False |
| HDL. C | 16 | CETP | protein_coding | rs4369653 | 5699755 1 | 2.72e-0 8 | 17 | 0 | 1 | 4 | 3 | 9 | Female only | Both sexes | Female only | True | False |
| HDL. C | 7 | RP11-36B 6.1 | lincRNA | rs972283 | 1304668 54 | 3.07e-0 8 | 19 | 10 | 0 | 0 | 0 | 9 | Female only | Female only | Female only | True | False |
| HDL. C | 18 | SMUG1P1 | pseudogene | rs6507939 | 4717626 1 | 3.27e-0 8 | 33 | 0 | 0 | 0 | 3 | 30 | Female only | Both sexes | Female only | True | False |

| pheno | CHR | Gene_Symbol | biotype | lead_snp | lead_BP | lead_P | TOTAL | NSIG | S05 | S01 | S001 | S0001 | independent_loci_in_sex_P1e6 | independent_loci_in_sex_P1e2 | independent_loci_in_sex_P5e8 | in_gwas_catalog | in_HLA |
|---|---|---|---|---|---|---|---|---|---|---|---|---|---|---|---|---|---|
| HEART.RATE | 13 | MTUS2 | protein_coding | rs4768988 | 29546818 | 4.69e-09 | 9 | 5 | 0 | 0 | 0 | 4 | Female only | Female only | Female only | True | False |
| HEIGHT | 4 | LCORL | protein_coding | rs112675878 | 18027696 | 3.54e-17 | 449 | 0 | 0 | 0 | 101 | 348 | Female only | Both sexes | Female only | True | False |
| HEIGHT | 20 | GDF5 | protein_coding | rs143384 | 34025756 | 9.86e-14 | 221 | 0 | 0 | 0 | 0 | 221 | Both sexes | Both sexes | Female only | True | False |
| HEIGHT | 3 | ZBTB38 | protein_coding | rs6440006 | 141142691 | 2.36e-13 | 75 | 0 | 0 | 0 | 10 | 65 | Both sexes | Both sexes | Female only | True | False |
| HEIGHT | 2 | EFEMP1 | protein_coding | rs1430203 | 56072509 | 2.87e-11 | 103 | 14 | 12 | 25 | 1 | 51 | Male only | Both sexes | Male only | True | False |
| pheno | CHR | Gene_Symbol | biotype | lead_snp | lead_BP | lead_P | TOTAL | NSIG | S05 | S01 | S001 | S0001 | independent_loci_in_sex_P1e6 | independent_loci_in_sex_P1e2 | independent_loci_in_sex_P5e8 | in_gwas_catalog | in_HLA |
| HEIGHT | 15 | ADAMTSL3 | protein_coding | rs1812707 | 84547222 | 4.50e-11 | 137 | 0 | 0 | 68 | 1 | 68 | Female only | Both sexes | Female only | True | False |
| HEIGHT | 4 | HHIP-AS1 | antisense | rs13106087 | 145566864 | 2.75e-10 | 19 | 0 | 0 | 0 | 2 | 17 | Both sexes | Both sexes | Female only | True | False |
| HEIGHT | 17 | ATAD5 | protein_coding | rs3764419 | 29164023 | 8.20e-10 | 29 | 8 | 7 | 1 | 0 | 13 | Female only | Female only | Female only | True | False |
| HEIGHT | 18 | CABLES1 | protein_coding | rs9961220 | 20733201 | 1.04e-09 | 39 | 0 | 0 | 0 | 0 | 39 | Both sexes | Both sexes | Both sexes | True | False |
| HEIGHT | 12 | HMGA2 | protein_coding | rs1351394 | 66351826 | 2.52e-09 | 31 | 0 | 0 | 0 | 1 | 30 | Female only | Both sexes | Female only | True | False |
| HEIGHT | 2 | LTBP1 | protein_coding | rs6743685 | 33389595 | 4.62e-09 | 201 | 56 | 45 | 0 | 0 | 100 | Female only | Female only | Female only | True | False |
| HEIGHT | 3 | ZBTB38 | protein_coding | rs13100711 | 141082990 | 5.30e-09 | 19 | 3 | 2 | 5 | 0 | 9 | Female only | Both sexes | Female only | True | False |
| HEIGHT | 13 | DLEU1 | protein_coding | rs2066647 | 50782074 | 5.42e-09 | 31 | 1 | 5 | 10 | 0 | 15 | Female only | Both sexes | Female only | True | False |
| HEIGHT | 12 | CCDC91 | protein_coding | rs11049356 | 28271456 | 7.32e-09 | 465 | 233 | 0 | 0 | 3 | 229 | Female only | Female only | Female only | True | False |
| HEIGHT | 2 | EFEMP1 | protein_coding | rs59985551 | 56106928 | 1.39e-08 | 17 | 1 | 0 | 7 | 1 | 8 | Male only | Both sexes | Male only | True | False |
| HEIGHT | 1 | DNM3 | protein_coding | rs678962 | 172189889 | 1.43e-08 | 37 | 0 | 0 | 17 | 2 | 18 | Female only | Both sexes | Female only | True | False |
| HEIGHT | 7 | IGF2BP3 | protein_coding | rs13247452 | 23477027 | 1.51e-08 | 49 | 0 | 0 | 2 | 24 | 23 | Male only | Both sexes | Male only | True | False |
| HEIGHT | 15 | ACAN | protein_coding | rs2280470 | 89395626 | 1.74e-08 | 23 | 12 | 0 | 0 | 5 | 6 | Female only | Female only | Female only | True | False |
| HEIGHT | 13 | DLEU1 | protein_coding | rs3116598 | 51099616 | 1.97e-08 | 109 | 0 | 0 | 2 | 1 | 106 | Both sexes | Both sexes | Female only | True | False |
| HEIGHT | 11 | SSH3 | protein_coding | rs76335321 | 67097778 | 1.99e-08 | 7 | 0 | 0 | 1 | 0 | 6 | Female only | Both sexes | Female only | True | False |
| HEIGHT | 2 | EFEMP1 | protein_coding | rs11125609 | 56115834 | 3.64e-08 | 5 | 3 | 0 | 0 | 0 | 2 | Male only | Male only | Male only | True | False |
| HEIGHT | 18 | CABLES1 | protein_coding | rs4281829 | 20705310 | 4.98e-08 | 65 | 4 | 0 | 21 | 12 | 28 | Male only | Both sexes | Male only | True | False |

| pheno | CHR | Gene_Symbol | biotype | lead_snp | lead_BP | lead_P | TOTAL | NSIG | S05 | S01 | S001 | S0001 | independent_loci_in_sex_P1e6 | independent_loci_in_sex_P1e2 | independent_loci_in_sex_P5e8 | in_gwas_catalog | in_HLA |
|---|---|---|---|---|---|---|---|---|---|---|---|---|---|---|---|---|---|
| HORIZONTAL.AXIS..INITIAL.40.MS | 12 | RP11-25E2.1 | lincRNA | rs7132327 | 115381071 | 1.10e-12 | 71 | 0 | 0 | 0 | 6 | 65 | Both sexes | Both sexes | Female only | True | False |
| Haptgl | 16 | TXNL4B | protein_coding | rs3794695 | 72097827 | 8.69e-12 | 25 | 0 | 0 | 8 | 0 | 17 | Both sexes | Both sexes | Both sexes | True | False |
| Haptgl | 16 | PKD1L3 | processed_transcript | rs12708919 | 71986995 | 2.24e-10 | 63 | 18 | 0 | 25 | 7 | 13 | Female only | Both sexes | Female only | True | False |
| Haptgl | 16 | PKD1L3 | processed_transcript | rs34972708 | 72008225 | 1.06e-09 | 41 | 7 | 1 | 6 | 9 | 18 | Female only | Both sexes | Female only | True | False |
| Height | 3 | ZBTB38 | protein_coding | rs7624084 | 141093285 | 9.77e-23 | 75 | 0 | 0 | 0 | 0 | 75 | Both sexes | Both sexes | Both sexes | True | False |
| Height | 4 | LCORL | protein_coding | rs13116230 | 18004014 | 9.88e-23 | 449 | 0 | 0 | 0 | 0 | 449 | Both sexes | Both sexes | Both sexes | True | False |
| Height | 20 | GDF5 | protein_coding | rs143384 | 34025756 | 2.28e-20 | 221 | 0 | 0 | 0 | 0 | 221 | Both sexes | Both sexes | Both sexes | True | False |
| Height | 12 | HMGA2 | protein_coding | rs7306710 | 66376091 | 3.97e-20 | 31 | 0 | 0 | 0 | 0 | 31 | Both sexes | Both sexes | Both sexes | True | False |
| Height | 15 | ADAMTSL3 | protein_coding | rs28801104 | 84552843 | 4.34e-19 | 137 | 0 | 0 | 0 | 0 | 137 | Both sexes | Both sexes | Both sexes | True | False |
| Height | 17 | ATAD5 | protein_coding | rs3764419 | 29164023 | 3.06e-17 | 29 | 0 | 1 | 0 | 0 | 28 | Both sexes | Both sexes | Both sexes | True | False |
| Height | 4 | LCORL | protein_coding | rs2061456 | 17998426 | 1.94e-15 | 47 | 0 | 0 | 0 | 0 | 47 | Both sexes | Both sexes | Both sexes | True | False |
| Height | 11 | KDM2A | protein_coding | rs12790261 | 66988048 | 2.33e-14 | 5 | 0 | 0 | 0 | 0 | 5 | Both sexes | Both sexes | Both sexes | True | False |
| Height | 4 | HHIP | protein_coding | rs7689420 | 145568352 | 2.70e-14 | 19 | 0 | 0 | 0 | 0 | 19 | Both sexes | Both sexes | Both sexes | True | False |
| Height | 2 | EFEMP1 | protein_coding | rs1344732 | 56029071 | 3.01e-14 | 123 | 18 | 2 | 0 | 2 | 101 | Both sexes | Both sexes | Both sexes | True | False |
| Height | 20 | CEP250 | protein_coding | rs224371 | 34074831 | 6.13e-14 | 195 | 0 | 0 | 1 | 15 | 179 | Both sexes | Both sexes | Female only | True | False |
| Height | 8 | PLAG1 | protein_coding | rs62515430 | 57109358 | 6.13e-14 | 41 | 1 | 7 | 13 | 0 | 20 | Female only | Both sexes | Female only | True | False |
| Height | 6 | L3MBTL3 | protein_coding | rs6569648 | 130349119 | 6.49e-14 | 95 | 0 | 0 | 0 | 19 | 76 | Both sexes | Both sexes | Both sexes | True | False |
| Height | 20 | RP5-859D4.3 | lincRNA | rs6038566 | 6615197 | 2.13e-13 | 53 | 0 | 0 | 15 | 12 | 26 | Female only | Both sexes | Female only | True | False |
| Height | 1 | ATP13A2 | protein_coding | rs4920605 | 17315425 | 5.93e-13 | 45 | 3 | 14 | 6 | 0 | 22 | Female only | Both sexes | Female only | True | False |
| Height | 8 | PLAG1 | protein_coding | rs34571768 | 57112046 | 6.02e-13 | 149 | 33 | 0 | 5 | 37 | 74 | Female only | Both sexes | Female only | True | False |
| Height | 15 | ADAMTSL3 | protein_coding | rs62025854 | 84552515 | 6.14e-13 | 63 | 0 | 1 | 0 | 3 | 59 | Female only | Both sexes | Female only | True | False |
| Height | 3 | ZBTB38 | protein_coding | rs13100711 | 141082990 | 7.18e-13 | 19 | 0 | 0 | 0 | 0 | 19 | Both sexes | Both sexes | Both sexes | True | False |
| Height | 20 | CASC20 | lincRNA | rs1884902 | 6608502 | 1.17e-12 | 59 | 0 | 0 | 3 | 27 | 29 | Female only | Both sexes | Female only | True | False |
| Height | 15 | SH3GL3 | protein_coding | rs2244838 | 84298598 | 1.39e-12 | 71 | 0 | 1 | 18 | 17 | 35 | Female only | Both sexes | Female only | True | False |

| pheno | CHR | Gene_Symbol | biotype | lead_snp | lead_BP | lead_P | TOTAL | NSIG | S05 | S01 | S001 | S0001 | independent_loci_in_sex_P1e6 | independent_loci_in_sex_P1e2 | independent_loci_in_sex_P5e8 | in_gwas_catalog | in_HLA |
|---|---|---|---|---|---|---|---|---|---|---|---|---|---|---|---|---|---|
| Height | 4 | LCORL | protein_coding | rs17526078 | 17848100 | 1.84e-12 | 7 | 0 | 0 | 2 | 0 | 5 | Female only | Both sexes | Female only | True | False |
| Height | 9 | PTCH1 | protein_coding | rs2297088 | 98242984 | 2.71e-12 | 29 | 5 | 7 | 3 | 0 | 14 | Female only | Both sexes | Female only | True | False |
| Height | 6 | LINC00577 | lincRNA | rs9322822 | 105369598 | 3.25e-12 | 105 | 0 | 0 | 1 | 18 | 86 | Female only | Both sexes | Female only | True | False |
| Height | 15 | ADAMTSL3 | protein_coding | rs35946026 | 84456496 | 7.97e-12 | 43 | 0 | 0 | 1 | 29 | 13 | Female only | Both sexes | Female only | True | False |
| Height | 18 | CABLES1 | protein_coding | rs4310973 | 20707622 | 8.73e-12 | 39 | 0 | 0 | 0 | 0 | 39 | Both sexes | Both sexes | Both sexes | True | False |
| Height | 12 | CCDC91 | protein_coding | rs10843114 | 28303296 | 1.05e-11 | 481 | 17 | 155 | 69 | 0 | 240 | Female only | Both sexes | Female only | True | False |
| Height | 12 | HMGA2 | protein_coding | rs61921611 | 66367726 | 1.52e-11 | 37 | 0 | 0 | 1 | 17 | 19 | Female only | Both sexes | Female only | True | False |
| Height | 15 | ADAMTSL3 | protein_coding | rs17370572 | 84616713 | 1.87e-11 | 15 | 0 | 8 | 5 | 0 | 2 | Female only | Female only | Female only | True | False |
| Height | 13 | DLEU1 | protein_coding | rs3116598 | 51099616 | 2.73e-11 | 109 | 0 | 0 | 2 | 14 | 93 | Female only | Both sexes | Female only | True | False |
| Height | 7 | CDK6 | protein_coding | rs3731326 | 92327026 | 2.87e-11 | 71 | 0 | 0 | 0 | 0 | 71 | Both sexes | Both sexes | Both sexes | True | False |
| Height | 4 | LCORL | protein_coding | rs2169033 | 18044357 | 3.21e-11 | 37 | 0 | 0 | 0 | 0 | 37 | Both sexes | Both sexes | Female only | True | False |
| Height | 2 | DIS3L2 | protein_coding | rs3103267 | 232988582 | 3.24e-11 | 53 | 0 | 0 | 0 | 3 | 50 | Both sexes | Both sexes | Male only | True | False |
| Height | 9 | RNU2-36P | snRNA | rs181338 | 89108161 | 3.51e-11 | 15 | 0 | 0 | 2 | 3 | 10 | Female only | Both sexes | Female only | True | False |
| Height | 23 | RP1-164L12.1 | pseudogene | X:78653645:A:G | 78653645 | 4.51e-11 | 153 | 0 | 0 | 0 | 0 | 153 | Both sexes | Both sexes | Both sexes | True | False |
| Height | 20 | SCAND1 | protein_coding | rs6060717 | 34544647 | 4.83e-11 | 555 | 0 | 35 | 85 | 136 | 299 | Female only | Both sexes | Female only | True | False |
| Height | 6 | C6orf106 | protein_coding | rs112540634 | 34623905 | 6.94e-11 | 215 | 39 | 30 | 31 | 20 | 95 | Female only | Both sexes | Female only | True | False |
| Height | 6 | LIN28B | protein_coding | rs314287 | 105434661 | 8.63e-11 | 27 | 9 | 2 | 2 | 1 | 13 | Female only | Both sexes | Female only | True | False |
| Height | 1 | DNM3 | protein_coding | rs10911323 | 172191332 | 8.65e-11 | 37 | 0 | 0 | 0 | 0 | 37 | Female only | Both sexes | Female only | True | False |
| Height | 6 | GPR126 | protein_coding | rs9389984 | 142652344 | 1.07e-10 | 161 | 0 | 0 | 2 | 15 | 144 | Female only | Both sexes | Female only | True | False |

| pheno | CHR | Gene_Symbol | biotype | lead_snp | lead_BP | lead_P | TOTAL | NSIG | S05 | S01 | S001 | S0001 | independent_loci_in_sex_P1e6 | independent_loci_in_sex_P1e2 | independent_loci_in_sex_P5e8 | in_gwas_catalog | in_HLA |
|---|---|---|---|---|---|---|---|---|---|---|---|---|---|---|---|---|---|
| Height | 6 | HIST1H4H | protein_coding | rs6456711 | 26302572 | 1.32e-10 | 85 | 0 | 36 | 4 | 3 | 42 | Female only | Both sexes | Female only | True | False |
| Height | 4 | FAM184B | protein_coding | rs740672 | 17782264 | 1.40e-10 | 9 | 0 | 0 | 0 | 0 | 9 | Female only | Both sexes | Female only | True | False |
| Height | 6 | HIST1H3PS1 | pseudogene | rs6918325 | 26327965 | 1.50e-10 | 97 | 1 | 30 | 17 | 1 | 48 | Female only | Both sexes | Female only | True | False |
| Height | 6 | HIST1H4E | protein_coding | rs9379833 | 26207175 | 1.81e-10 | 9 | 0 | 0 | 0 | 1 | 8 | Female only | Both sexes | Female only | True | False |
| Height | 20 | NFS1 | protein_coding | rs11697903 | 34272224 | 2.30e-10 | 147 | 0 | 0 | 22 | 30 | 95 | Female only | Both sexes | Female only | True | False |
| Height | 15 | ACAN | protein_coding | rs2280470 | 89395626 | 5.12e-10 | 23 | 1 | 10 | 1 | 4 | 7 | Female only | Both sexes | Female only | True | False |

| pheno | CHR | Gene_Symbol | biotype | lead_snp | lead_BP | lead_P | TOTAL | NSIG | S05 | S01 | S001 | S0001 | independent_loci_in_sex_P1e6 | independent_loci_in_sex_P1e2 | independent_loci_in_sex_P5e8 | in_gwas_catalog | in_HLA |
|---|---|---|---|---|---|---|---|---|---|---|---|---|---|---|---|---|---|
| Height | 12 | CCDC91 | protein_coding | rs2272361 | 28602970 | 5.61e-10 | 261 | 0 | 10 | 121 | 0 | 130 | Female only | Both sexes | Female only | True | False |
| Height | 7 | CDK6 | protein_coding | rs2079147 | 92332375 | 5.78e-10 | 9 | 0 | 0 | 0 | 0 | 9 | Both sexes | Both sexes | Both sexes | True | False |
| Height | 15 | ADAMTSL3 | protein_coding | rs4414460 | 84562740 | 6.11e-10 | 9 | 0 | 5 | 0 | 3 | 1 | Female only | Female only | Female only | True | False |
| Height | 6 | SUPT3H | protein_coding | rs1285027 | 45121531 | 1.02e-09 | 319 | 160 | 0 | 0 | 0 | 159 | Female only | Female only | Female only | True | False |
| Height | 15 | SH3GL3 | protein_coding | rs2585058 | 84284552 | 1.03e-09 | 5 | 0 | 0 | 1 | 2 | 2 | Female only | Both sexes | Female only | True | False |
| Height | 11 | INS-IGF2 | protein_coding | rs4320932 | 2171601 | 1.07e-09 | 7 | 0 | 0 | 0 | 0 | 7 | Both sexes | Both sexes | Both sexes | True | False |
| Height | 4 | LCORL | protein_coding | rs113921625 | 18133262 | 1.09e-09 | 3 | 0 | 0 | 0 | 2 | 1 | Female only | Both sexes | Female only | True | False |
| Height | 18 | CABLES1 | protein_coding | rs11082304 | 20720973 | 1.18e-09 | 3 | 0 | 0 | 0 | 0 | 3 | Both sexes | Both sexes | Female only | True | False |
| Height | 20 | EDEM2 | protein_coding | rs7280 | 33864484 | 1.39e-09 | 7 | 0 | 0 | 0 | 0 | 7 | Both sexes | Both sexes | Both sexes | True | False |
| Height | 3 | ZBTB38 | protein_coding | rs7636914 | 141158614 | 1.45e-09 | 53 | 0 | 0 | 0 | 8 | 45 | Female only | Both sexes | Female only | True | False |
| Height | 6 | SUPT3H | protein_coding | rs35214627 | 44790915 | 1.56e-09 | 95 | 48 | 0 | 0 | 1 | 46 | Female only | Female only | Female only | True | False |
| Height | 15 | ACAN | protein_coding | rs8042988 | 89412279 | 1.66e-09 | 5 | 0 | 0 | 3 | 0 | 2 | Female only | Both sexes | Female only | True | False |
| Height | 23 | RP1-164L12.1 | pseudogene | X:78641728:G:A | 78641728 | 1.71e-09 | 5 | 0 | 0 | 0 | 1 | 4 | Male only | Both sexes | Male only | True | False |
| Height | 15 | ADAMTSL3 | protein_coding | rs12148139 | 84405005 | 1.74e-09 | 103 | 52 | 0 | 1 | 0 | 50 | Female only | Female only | Female only | True | False |
| Height | 17 | CD79B | protein_coding | rs2003549 | 62008437 | 1.97e-09 | 127 | 0 | 0 | 0 | 0 | 127 | Both sexes | Both sexes | Female only | True | False |
| Height | 1 | DNM3 | protein_coding | rs17361955 | 172141999 | 2.12e-09 | 69 | 0 | 21 | 14 | 0 | 34 | Female only | Both sexes | Female only | True | False |
| Height | 15 | ADAMTSL3 | protein_coding | rs78015020 | 84312788 | 2.18e-09 | 5 | 3 | 0 | 0 | 0 | 2 | Female only | Female only | Female only | True | False |
| Height | 14 | MARK3 | protein_coding | rs7143609 | 103878148 | 2.49e-09 | 107 | 7 | 46 | 1 | 0 | 53 | Female only | Both sexes | Female only | True | False |
| Height | 17 | C17orf82 | protein_coding | rs2079795 | 59496649 | 2.78e-09 | 17 | 0 | 0 | 0 | 0 | 17 | Both sexes | Both sexes | Both sexes | True | False |
| Height | 2 | AC006369.3 | lincRNA | rs12615742 | 37995727 | 3.12e-09 | 3 | 1 | 0 | 1 | 0 | 1 | Male only | Both sexes | Male only | True | False |
| Height | 5 | NPR3 | protein_coding | rs3792765 | 32713836 | 3.17e-09 | 31 | 0 | 13 | 3 | 0 | 15 | Male only | Both sexes | Male only | True | False |
| Height | 23 | KIF4CP | pseudogene | X:78594876:T:G | 78594876 | 3.86e-09 | 133 | 0 | 1 | 48 | 18 | 66 | Male only | Both sexes | Male only | True | False |
| Height | 2 | EFEMP1 | protein_coding | rs1346786 | 56108333 | 4.02e-09 | 13 | 4 | 3 | 0 | 0 | 6 | Male only | Both sexes | Male only | True | False |
| Height | 1 | COLGALT2 | protein_coding | rs3814333 | 184007119 | 4.41e-09 | 65 | 0 | 9 | 0 | 3 | 53 | Female only | Both sexes | Female only | True | False |
| Height | 13 | DLEU1 | protein_coding | rs2066623 | 50753772 | 4.65e-09 | 35 | 1 | 10 | 7 | 10 | 7 | Female only | Both sexes | Female only | True | False |
| Height | 7 | JAZF1 | protein_coding | rs520161 | 28210660 | 4.78e-09 | 35 | 0 | 0 | 3 | 2 | 30 | Female only | Both sexes | Female only | True | False |

| pheno | CHR | Gene_Symbol | biotype | lead_snp | lead_BP | lead_P | TOTAL | NSIG | S05 | S01 | S001 | S0001 | independent_loci_in_sex_P1e6 | independent_loci_in_sex_P1e2 | independent_loci_in_sex_P5e8 | in_gwas_catalog | in_HLA |
|---|---|---|---|---|---|---|---|---|---|---|---|---|---|---|---|---|---|
| Height | 8 | AC105242.1 | miRNA | rs10111935 | 78131147 | 4.99e-09 | 81 | 4 | 0 | 6 | 17 | 54 | Female only | Both sexes | Female only | True | False |
| Height | 15 | ADAMTS17 | protein_coding | rs4246307 | 100763824 | 5.10e-09 | 11 | 3 | 1 | 2 | 0 | 5 | Female only | Both sexes | Female only | True | False |
| Height | 15 | ADAMTSL3 | protein_coding | rs5015595 | 84493651 | 5.63e-09 | 5 | 0 | 0 | 0 | 0 | 5 | Both sexes | Both sexes | Both sexes | True | False |
| Height | 3 | RP11-438D8.2 | antisense | rs6778010 | 141042363 | 5.86e-09 | 17 | 0 | 4 | 2 | 2 | 9 | Female only | Both sexes | Female only | True | False |
| Height | 5 | P4HA2 | protein_coding | rs10463892 | 131598705 | 6.77e-09 | 177 | 0 | 13 | 57 | 20 | 87 | Female only | Both sexes | Female only | True | False |
| Height | 7 | GNA12 | protein_coding | rs1182198 | 2862906 | 6.93e-09 | 247 | 0 | 0 | 5 | 42 | 200 | Female only | Both sexes | Female only | True | False |
| Height | 18 | NFATC1 | protein_coding | rs2055899 | 77202949 | 7.39e-09 | 141 | 51 | 17 | 3 | 16 | 54 | Female only | Both sexes | Female only | True | False |
| Height | 20 | PXMP4 | protein_coding | rs55990870 | 32302908 | 7.82e-09 | 59 | 5 | 4 | 7 | 18 | 25 | Male only | Both sexes | Male only | True | False |
| Height | 1 | RNA5SP56 | rRNA | rs59051938 | 118850501 | 7.95e-09 | 59 | 0 | 0 | 27 | 3 | 29 | Male only | Both sexes | Male only | True | False |
| Height | 4 | HHIP | protein_coding | rs2131354 | 145599908 | 8.32e-09 | 159 | 0 | 0 | 20 | 24 | 115 | Both sexes | Both sexes | Female only | True | False |
| Height | 7 | CDK6 | protein_coding | rs62466068 | 92301890 | 9.96e-09 | 3 | 2 | 0 | 0 | 0 | 1 | Female only | Female only | Female only | True | False |
| Height | 15 | ADAMTSL3 | protein_coding | rs11259926 | 84566248 | 1.03e-08 | 13 | 0 | 2 | 4 | 0 | 7 | Male only | Both sexes | Male only | True | False |
| Height | 15 | PML | protein_coding | rs16869 | 74322431 | 1.05e-08 | 57 | 28 | 1 | 0 | 0 | 28 | Female only | Female only | Female only | True | False |
| Height | 8 | ADAM28 | protein_coding | rs62501213 | 24093306 | 1.11e-08 | 77 | 19 | 19 | 4 | 8 | 27 | Female only | Both sexes | Female only | True | False |
| Height | 3 | FNDC3B | protein_coding | rs13094143 | 171973787 | 1.21e-08 | 9 | 0 | 0 | 0 | 4 | 5 | Both sexes | Both sexes | Male only | True | False |
| Height | 6 | RP11-486E2.1 | pseudogene | rs78892181 | 81305223 | 1.32e-08 | 281 | 2 | 5 | 42 | 17 | 215 | Female only | Both sexes | Female only | True | False |
| Height | 2 | DIS3L2 | protein_coding | rs3116162 | 232860077 | 1.34e-08 | 49 | 0 | 0 | 0 | 1 | 48 | Both sexes | Both sexes | Female only | True | False |
| Height | 4 | HHIP | protein_coding | rs75385537 | 145706070 | 1.40e-08 | 5 | 0 | 0 | 0 | 0 | 5 | Both sexes | Both sexes | Female only | True | False |
| Height | 3 | RP11-894J14.5 | protein_coding | rs2336723 | 53106047 | 1.55e-08 | 145 | 73 | 0 | 0 | 10 | 62 | Female only | Female only | Female only | True | False |
| Height | 11 | TPCN2 | protein_coding | rs3750963 | 68840014 | 1.60e-08 | 97 | 0 | 2 | 0 | 5 | 90 | Male only | Both sexes | Male only | True | False |
| Height | 20 | CASC20 | lincRNA | rs6085575 | 6467051 | 1.69e-08 | 103 | 0 | 0 | 11 | 25 | 67 | Female only | Both sexes | Female only | True | False |
| Height | 6 | BMP6 | protein_coding | rs56180096 | 7792320 | 1.95e-08 | 11 | 0 | 3 | 3 | 0 | 5 | Male only | Both sexes | Male only | True | False |
| Height | 4 | PRKG2 | protein_coding | rs788857 | 82138551 | 2.15e-08 | 277 | 0 | 11 | 87 | 54 | 125 | Female only | Both sexes | Female only | True | False |
| Height | 6 | BCKDHB | protein_coding | rs145101575 | 81053757 | 2.30e-08 | 17 | 0 | 2 | 6 | 2 | 7 | Female only | Both sexes | Female only | True | False |
| Height | 5 | NPR3 | protein_coding | rs1173730 | 32763846 | 2.45e-08 | 47 | 4 | 13 | 9 | 2 | 19 | Female only | Both sexes | Female only | True | False |
| Height | 6 | SUPT3H | protein_coding | rs524492 | 44796596 | 2.65e-08 | 43 | 22 | 0 | 0 | 0 | 21 | Female only | Female only | Female only | True | False |

| pheno | CHR | Gene_Symbol | biotype | lead_snp | lead_BP | lead_P | TOTAL | NSIG | S05 | S01 | S001 | S0001 | independent_loci_in_sex_P1e6 | independent_loci_in_sex_P1e2 | independent_loci_in_sex_P5e8 | in_gwas_catalog | in_HLA |
|---|---|---|---|---|---|---|---|---|---|---|---|---|---|---|---|---|---|
| Height | 9 | PCSK5 | protein_coding | rs35307904 | 78511889 | 2.93e-08 | 9 | 0 | 1 | 0 | 2 | 6 | Female only | Both sexes | Female only | True | False |
| Height | 2 | EFEMP1 | protein_coding | rs11899888 | 56102744 | 2.93e-08 | 19 | 1 | 0 | 6 | 3 | 9 | Female only | Both sexes | Female only | True | False |
| **pheno** | **CHR** | **Gene_Symbol** | **biotype** | **lead_snp** | **lead_BP** | **lead_P** | **TOTAL** | **NSIG** | **S05** | **S01** | **S001** | **S0001** | **independent_loci_in_sex_P1e6** | **independent_loci_in_sex_P1e2** | **independent_loci_in_sex_P5e8** | **in_gwas_catalog** | **in_HLA** |
| Height | 2 | DIS3L2 | protein_coding | rs6437056 | 233097174 | 3.37e-08 | 23 | 0 | 0 | 1 | 5 | 17 | Male only | Both sexes | Male only | True | False |
| Height | 5 | CTB-33O18.1 | lincRNA | rs7717406 | 172999405 | 3.39e-08 | 93 | 6 | 13 | 28 | 2 | 44 | Female only | Both sexes | Female only | True | False |
| Height | 20 | CDK5RAP1 | protein_coding | rs291700 | 31981849 | 3.70e-08 | 11 | 0 | 3 | 0 | 3 | 5 | Male only | Both sexes | Male only | True | False |
| Height | 15 | ADAMTSL3 | protein_coding | rs11638600 | 84661481 | 3.77e-08 | 47 | 0 | 0 | 5 | 21 | 21 | Female only | Both sexes | Female only | True | False |
| Height | 9 | C9orf3 | protein_coding | rs11787821 | 97660961 | 3.78e-08 | 117 | 30 | 1 | 32 | 26 | 28 | Female only | Both sexes | Female only | True | False |
| Height | 14 | LTBP2 | protein_coding | rs4899520 | 74987572 | 3.78e-08 | 43 | 0 | 1 | 8 | 10 | 24 | Female only | Both sexes | Female only | True | False |
| Height | 11 | SERPINH1 | protein_coding | rs634552 | 75282052 | 4.06e-08 | 11 | 0 | 0 | 0 | 0 | 11 | Both sexes | Both sexes | Both sexes | True | False |
| Height | 13 | DLEU1 | protein_coding | rs201801 | 50953245 | 4.12e-08 | 9 | 1 | 3 | 1 | 0 | 4 | Female only | Both sexes | Female only | True | False |
| Height | 6 | ANKS1A | protein_coding | rs13210323 | 35005084 | 4.37e-08 | 3 | 1 | 0 | 1 | 1 | 0 | Male only | Both sexes | Male only | True | False |
| Height | 17 | CD79B | protein_coding | rs2005132 | 62008232 | 4.71e-08 | 77 | 0 | 0 | 1 | 27 | 49 | Female only | Both sexes | Female only | True | False |
| Height | 6 | MIR588 | miRNA | rs1907067 | 126837663 | 4.74e-08 | 313 | 157 | 0 | 0 | 0 | 156 | Male only | Male only | Male only | True | False |
| Hgb | 22 | TMPRSS6 | protein_coding | rs4820268 | 37469591 | 1.28e-19 | 9 | 0 | 0 | 0 | 0 | 9 | Both sexes | Both sexes | Both sexes | True | False |
| Hgb | 6 | HIST1H3C | protein_coding | 6:26046744 | 26046744 | 1.37e-13 | 245 | 19 | 55 | 14 | 35 | 122 | Female only | Both sexes | Female only | True | False |
| Hgb | 6 | HFE | protein_coding | rs144861591 | 26072992 | 1.79e-12 | 19 | 0 | 0 | 0 | 0 | 19 | Both sexes | Both sexes | Both sexes | True | False |
| Hgb | 6 | HIST1H4C | protein_coding | rs198851 | 26104632 | 1.13e-10 | 39 | 0 | 7 | 3 | 9 | 20 | Female only | Both sexes | Female only | True | False |
| Hgb | 6 | HIST1H2AA | protein_coding | rs116009877 | 25715657 | 1.23e-10 | 33 | 0 | 0 | 0 | 0 | 33 | Both sexes | Both sexes | Both sexes | True | False |
| Hgb | 22 | MPST | protein_coding | rs25095 | 37434028 | 2.90e-10 | 33 | 5 | 1 | 10 | 1 | 16 | Female only | Both sexes | Female only | True | False |
| Hgb | 22 | TMPRSS6 | protein_coding | rs9607412 | 37484337 | 9.33e-10 | 57 | 23 | 6 | 0 | 0 | 28 | Female only | Female only | Female only | True | False |
| Hgb | 6 | TRIM38 | protein_coding | rs169219 | 25957392 | 9.82e-10 | 33 | 14 | 3 | 0 | 0 | 16 | Female only | Female only | Female only | True | False |
| Hgb | 6 | LRRC16A | protein_coding | rs17492120 | 25590311 | 1.18e-09 | 23 | 0 | 0 | 4 | 6 | 13 | Male only | Both sexes | Male only | True | False |
| Hgb | 6 | SLC17A1 | protein_coding | rs12191655 | 25803095 | 2.80e-09 | 297 | 44 | 100 | 4 | 7 | 142 | Female only | Both sexes | Female only | True | False |
| Hgb | 22 | TMPRSS6 | protein_coding | rs2235325 | 37481637 | 2.89e-09 | 23 | 1 | 3 | 8 | 0 | 11 | Female only | Both sexes | Female only | True | False |
| Hgb | 6 | LRRC16A | protein_coding | rs74662487 | 25453057 | 5.65e-09 | 17 | 0 | 1 | 8 | 7 | 1 | Male only | Both sexes | Male only | True | False |

| pheno | CHR | Gene_Symbol | biotype | lead_snp | lead_BP | lead_P | TOTAL | NSIG | S05 | S01 | S001 | S0001 | independent_loci_in_sex_P1e6 | independent_loci_in_sex_P1e2 | independent_loci_in_sex_P5e8 | in_gwas_catalog | in_HLA |
|---|---|---|---|---|---|---|---|---|---|---|---|---|---|---|---|---|---|
| Hgb | 6 | HCG4P7 | pseudogene | rs1611657 | 29842444 | 9.50e-09 | 155 | 2 | 74 | 2 | 0 | 77 | Female only | Both sexes | Female only | True | True |
| Hgb | 6 | ZDHHC20P2 | pseudogene | rs2596570 | 31348040 | 1.06e-08 | 241 | 1 | 16 | 31 | 77 | 116 | Male only | Both sexes | Male only | True | True |
| Hgb | 6 | SCGN | protein_coding | rs9467571 | 25700408 | 1.38e-08 | 33 | 10 | 4 | 3 | 10 | 6 | Female only | Both sexes | Female only | True | False |
| Hgb | 10 | HK1 | protein_coding | rs17476364 | 71094504 | 1.61e-08 | 11 | 0 | 0 | 5 | 1 | 5 | Female only | Both sexes | Female only | True | False |
| Hgb | 6 | MIR3143 | miRNA | rs71559014 | 27122444 | 3.72e-08 | 3 | 0 | 0 | 0 | 0 | 3 | Male only | Both sexes | Male only | True | False |
| Hgb | 9 | ABO | processed_transcript | rs532436 | 136149830 | 4.49e-08 | 21 | 1 | 9 | 1 | 0 | 10 | Female only | Both sexes | Female only | True | False |
| HgbA1C | 6 | HLA-DQB1 | protein_coding | rs9273363 | 32626272 | 2.76e-18 | 7 | 0 | 0 | 0 | 0 | 7 | Both sexes | Both sexes | Both sexes | True | True |
| HgbA1C | 10 | TCF7L2 | protein_coding | rs34872471 | 114754071 | 1.32e-13 | 47 | 0 | 0 | 0 | 0 | 47 | Both sexes | Both sexes | Both sexes | True | False |
| HgbA1C | 6 | HLA-DRB9 | pseudogene | rs9269070 | 32440451 | 7.08e-11 | 67 | 0 | 0 | 11 | 8 | 48 | Female only | Both sexes | Female only | True | True |
| HgbA1C | 6 | NOTCH4 | protein_coding | rs377763 | 32199144 | 1.86e-10 | 23 | 0 | 0 | 0 | 3 | 20 | Both sexes | Both sexes | Female only | True | True |
| HgbA1C | 6 | HLA-DQB1 | protein_coding | rs2854275 | 32628428 | 3.08e-09 | 153 | 0 | 18 | 22 | 17 | 96 | Both sexes | Both sexes | Both sexes | True | True |
| HgbA1C | 10 | HK1 | protein_coding | rs16926246 | 71093392 | 6.06e-09 | 11 | 0 | 0 | 0 | 0 | 11 | Both sexes | Both sexes | Both sexes | True | False |
| HgbA1C | 6 | NOTCH4 | protein_coding | rs3130293 | 32177263 | 8.33e-09 | 19 | 1 | 0 | 1 | 7 | 10 | Both sexes | Both sexes | Female only | True | True |
| HgbA1C | 6 | BAG6 | protein_coding | rs3130628 | 31609272 | 1.20e-08 | 127 | 0 | 0 | 1 | 45 | 81 | Male only | Both sexes | Male only | True | True |
| HgbA1C | 6 | XXbac-BPG16N22.5 | pseudogene | rs2516491 | 31485101 | 3.28e-08 | 11 | 0 | 0 | 5 | 4 | 2 | Male only | Both sexes | Male only | True | True |
| HgbA1C | 6 | AGER | protein_coding | rs3131300 | 32151934 | 3.43e-08 | 185 | 0 | 0 | 1 | 36 | 148 | Both sexes | Both sexes | Male only | True | True |
| HgbA1C | 6 | NOTCH4 | protein_coding | rs9267890 | 32202086 | 3.95e-08 | 145 | 0 | 0 | 10 | 43 | 92 | Both sexes | Both sexes | Female only | True | True |
| IGAB | 6 | RP1-111C20.3 | lincRNA | rs2451257 | 159507026 | 1.47e-26 | 15 | 0 | 0 | 0 | 0 | 15 | Both sexes | Both sexes | Both sexes | False | False |
| IGAB | 9 | LRRC8A | protein_coding | rs34305034 | 131654547 | 1.75e-24 | 3 | 0 | 0 | 0 | 0 | 3 | Both sexes | Both sexes | Both sexes | False | False |
| IGAB | 6 | RP1-111C20.3 | lincRNA | rs2451258 | 159506600 | 4.80e-12 | 21 | 0 | 3 | 2 | 0 | 16 | Male only | Both sexes | Male only | False | False |
| IGAB | 10 | PRTFDC1 | protein_coding | rs7896608 | 25206412 | 2.18e-11 | 15 | 0 | 0 | 8 | 0 | 7 | Male only | Both sexes | Male only | False | False |
| IGAB | 9 | ENDOG | protein_coding | rs4837316 | 131577450 | 4.61e-11 | 55 | 0 | 0 | 0 | 1 | 54 | Both sexes | Both sexes | Both sexes | False | False |
| IGAB | 2 | ITGA4 | protein_coding | rs6740847 | 182308352 | 6.98e-10 | 15 | 0 | 0 | 0 | 0 | 15 | Female only | Both sexes | Female only | False | False |
| IGAB | 22 | DEPDC5 | protein_coding | rs5749339 | 32315735 | 1.84e-09 | 35 | 0 | 0 | 0 | 8 | 27 | Both sexes | Both sexes | Female only | False | False |
| IGAB | 6 | RP11-13P5.1 | lincRNA | rs62431905 | 159510162 | 1.26e-08 | 7 | 0 | 0 | 0 | 0 | 7 | Both sexes | Both sexes | Male only | False | False |
| IGAB | 7 | CCM2 | protein_coding | rs2158564 | 45038881 | 1.79e-08 | 85 | 0 | 0 | 0 | 2 | 83 | Both sexes | Both sexes | Male only | False | False |

| pheno | CHR | Gene_Symbol | biotype | lead_snp | lead_BP | lead_P | TOTAL | NSIG | S05 | S01 | S001 | S0001 | independent_loci_in_sex_P1e6 | independent_loci_in_sex_P1e2 | independent_loci_in_sex_P5e8 | in_gwas_catalog | in_HLA |
|---|---|---|---|---|---|---|---|---|---|---|---|---|---|---|---|---|---|
| IGRE | 9 | LRRC8A | protein_coding | rs34305034 | 131654547 | 2.17e-45 | 3 | 0 | 0 | 0 | 0 | 3 | Both sexes | Both sexes | Both sexes | True | False |
| IGRE | 6 | RP1-111C20.3 | lincRNA | rs2451256 | 159507338 | 1.36e-33 | 15 | 0 | 0 | 0 | 0 | 15 | Both sexes | Both sexes | Both sexes | True | False |
| IGRE | 9 | ENDOG | protein_coding | rs4837316 | 131577450 | 1.43e-19 | 55 | 0 | 0 | 0 | 0 | 55 | Both sexes | Both sexes | Both sexes | True | False |
| IGRE | 6 | RP1-111C20.3 | lincRNA | rs2451258 | 159506600 | 4.72e-16 | 21 | 0 | 0 | 2 | 3 | 16 | Both sexes | Both sexes | Both sexes | True | False |
| IGRE | 10 | PRTFDC1 | protein_coding | rs10828722 | 25199951 | 3.15e-15 | 15 | 0 | 0 | 0 | 0 | 15 | Male only | Both sexes | Male only | True | False |
| IGRE | 6 | RP11-13P5.1 | lincRNA | rs62431905 | 159510162 | 3.74e-13 | 7 | 0 | 0 | 0 | 0 | 7 | Both sexes | Both sexes | Both sexes | True | False |
| IGRE | 22 | DEPDC5 | protein_coding | rs5749339 | 32315735 | 2.30e-11 | 35 | 0 | 0 | 0 | 0 | 35 | Both sexes | Both sexes | Both sexes | True | False |
| IGRE | 2 | ITGA4 | protein_coding | rs2124440 | 182328214 | 8.10e-11 | 15 | 0 | 0 | 0 | 0 | 15 | Both sexes | Both sexes | Female only | True | False |
| IGRE | 7 | MYO1G | protein_coding | rs3735485 | 45009341 | 2.64e-10 | 183 | 0 | 0 | 5 | 85 | 93 | Male only | Both sexes | Male only | True | False |
| IGRE | 9 | ZDHHC12 | protein_coding | rs10988105 | 131484526 | 4.36e-10 | 3 | 0 | 0 | 0 | 0 | 3 | Both sexes | Both sexes | Both sexes | True | False |
| IGRE | 8 | CHD7 | protein_coding | rs13256023 | 61648551 | 5.92e-10 | 7 | 0 | 0 | 1 | 1 | 5 | Both sexes | Both sexes | Both sexes | True | False |
| IGRE | 7 | CCM2 | protein_coding | rs36084700 | 45034408 | 6.16e-10 | 85 | 0 | 0 | 0 | 0 | 85 | Both sexes | Both sexes | Male only | True | False |
| IGRE | 10 | PRTFDC1 | protein_coding | rs7905553 | 25200591 | 1.09e-08 | 7 | 0 | 0 | 1 | 3 | 3 | Male only | Both sexes | Male only | True | False |
| IGRE | 7 | TBRG4 | protein_coding | rs6955660 | 45149410 | 1.40e-08 | 11 | 0 | 3 | 3 | 0 | 5 | Male only | Both sexes | Male only | True | False |
| IGRE | 6 | RP1-111C20.4 | antisense | rs394581 | 159482521 | 1.50e-08 | 9 | 0 | 0 | 2 | 0 | 7 | Male only | Both sexes | Male only | True | False |
| IctIdx | 2 | UGT1A8 | protein_coding | rs6742078 | 234672639 | 5.51e-13 | 219 | 0 | 57 | 48 | 5 | 109 | Male only | Both sexes | Male only | True | False |
| IgM.Q | 2 | PDE11A | protein_coding | rs113811979 | 178695260 | 5.13e-09 | 137 | 69 | 0 | 0 | 13 | 55 | Female only | Female only | Female only | True | False |
| Infant.Apgar.Color.1.M.1 | 13 | METTL21C | protein_coding | rs9518810 | 103343428 | 2.79e-08 | 2 | 0 | 0 | 0 | 0 | 2 | GWAS only run in females | GWAS only run in females | GWAS only run in females | False | False |
| Infant.Apgar.Resp.5.M.1 | 13 | ARHGEF7 | protein_coding | rs58934014 | 111795572 | 2.33e-08 | 5 | 0 | 0 | 0 | 0 | 5 | GWAS only run in females | GWAS only run in females | GWAS only run in females | False | False |
| Iron | 22 | TMPRSS6 | protein_coding | rs2413450 | 37470224 | 1.03e-19 | 9 | 0 | 0 | 0 | 0 | 9 | Both sexes | Both sexes | Both sexes | True | False |
| Iron | 6 | HFE | protein_coding | rs144861591 | 26072992 | 2.76e-15 | 19 | 0 | 0 | 0 | 0 | 19 | Both sexes | Both sexes | Both sexes | True | False |
| Iron | 6 | HIST1H2AA | protein_coding | rs116009877 | 25715657 | 6.04e-13 | 33 | 0 | 0 | 0 | 0 | 33 | Both sexes | Both sexes | Both sexes | True | False |

| pheno | CHR | Gene_Symbol | biotype | lead_snp | lead_BP | lead_P | TOTAL | NSIG | S05 | S01 | S001 | S0001 | independent_loci_in_sex_P1e6 | independent_loci_in_sex_P1e2 | independent_loci_in_sex_P5e8 | in_gwas_catalog | in_HLA |
|---|---|---|---|---|---|---|---|---|---|---|---|---|---|---|---|---|---|
| Iron | 6 | LRRC16A | protein_coding | rs77244967 | 25536591 | 7.63e-12 | 23 | 0 | 0 | 0 | 0 | 23 | Both sexes | Both sexes | Both sexes | True | False |
| Iron | 6 | HIST1H1T | protein_coding | rs198846 | 26107463 | 1.82e-11 | 33 | 0 | 0 | 4 | 1 | 28 | Both sexes | Both sexes | Female only | True | False |
| Iron | 6 | HIST1H3C | protein_coding | 6:26046744 | 26046744 | 2.03e-11 | 245 | 0 | 0 | 27 | 2 | 216 | Both sexes | Both sexes | Both sexes | True | False |
| Iron | 6 | LRRC16A | protein_coding | rs13203202 | 25582771 | 6.46e-10 | 9 | 1 | 0 | 1 | 3 | 4 | Female only | Both sexes | Female only | True | False |
| Iron | 22 | MPST | protein_coding | rs130610 | 37431871 | 3.01e-09 | 33 | 0 | 0 | 0 | 1 | 32 | Female only | Both sexes | Female only | True | False |
| Iron | 22 | TMPRSS6 | protein_coding | rs6000550 | 37463450 | 1.41e-08 | 15 | 4 | 0 | 3 | 3 | 5 | Female only | Both sexes | Female only | True | False |
| K | 5 | KLHL3 | protein_coding | rs56015773 | 137010528 | 5.07e-10 | 5 | 0 | 0 | 0 | 2 | 3 | Female only | Both sexes | Female only | True | False |
| K | 5 | PKD2L2 | protein_coding | rs10055169 | 137228063 | 3.54e-09 | 335 | 0 | 1 | 87 | 2 | 245 | Female only | Both sexes | Female only | True | False |
| K | 6 | HLA-DQB1 | protein_coding | rs2854275 | 32628428 | 1.02e-08 | 153 | 0 | 0 | 23 | 21 | 109 | Male only | Both sexes | Male only | True | True |
| K | 5 | KLHL3 | protein_coding | rs41298968 | 137034169 | 2.96e-08 | 3 | 0 | 0 | 1 | 1 | 1 | Female only | Both sexes | Female only | True | False |
| LDL.C | 19 | APOE | protein_coding | rs7412 | 45412079 | 3.89e-142 | 9 | 0 | 0 | 0 | 0 | 9 | Both sexes | Both sexes | Both sexes | True | False |
| LDL.C | 19 | APOE | protein_coding | rs429358 | 45411941 | 4.99e-44 | 39 | 5 | 0 | 0 | 3 | 31 | Both sexes | Both sexes | Both sexes | True | False |
| LDL.C | 1 | CELSR2 | protein_coding | rs12740374 | 109817590 | 4.13e-30 | 27 | 0 | 0 | 0 | 0 | 27 | Both sexes | Both sexes | Both sexes | True | False |
| LDL.C | 19 | BCL3 | protein_coding | rs62117205 | 45255266 | 2.50e-28 | 13 | 0 | 0 | 0 | 1 | 12 | Both sexes | Both sexes | Both sexes | True | False |
| LDL.C | 19 | PVRL2 | protein_coding | rs395908 | 45373565 | 3.08e-28 | 15 | 9 | 1 | 0 | 0 | 5 | Female only | Both sexes | Female only | True | False |
| LDL.C | 19 | PVRL2 | protein_coding | 19:45384405 | 45384405 | 5.74e-27 | 3 | 2 | 0 | 0 | 0 | 1 | Both sexes | Both sexes | Both sexes | True | False |
| LDL.C | 19 | PVRL2 | protein_coding | rs34342646 | 45388130 | 5.27e-25 | 3 | 0 | 0 | 0 | 1 | 2 | Both sexes | Both sexes | Female only | True | False |
| LDL.C | 19 | BCAM | protein_coding | rs4803760 | 45333834 | 3.08e-22 | 3 | 0 | 0 | 0 | 0 | 3 | Female only | Both sexes | Female only | True | False |
| LDL.C | 19 | BCAM | protein_coding | rs10402271 | 45329214 | 9.43e-21 | 13 | 0 | 0 | 2 | 5 | 6 | Female only | Both sexes | Female only | True | False |
| LDL.C | 19 | TOMM40 | protein_coding | rs157580 | 45395266 | 5.76e-19 | 7 | 6 | 0 | 0 | 1 | 0 | Female only | Both sexes | Female only | True | False |
| LDL.C | 19 | PVRL2 | protein_coding | rs393584 | 45377334 | 9.31e-18 | 9 | 6 | 0 | 2 | 0 | 1 | Female only | Both sexes | Female only | True | False |
| LDL.C | 19 | LDLR | protein_coding | rs73015016 | 11191300 | 3.74e-17 | 75 | 0 | 0 | 0 | 0 | 75 | Both sexes | Both sexes | Both sexes | True | False |
| LDL.C | 2 | APOB | protein_coding | rs13671155 | 21282015 | 6.72e-17 | 105 | 0 | 0 | 0 | 21 | 84 | Both sexes | Both sexes | Female only | True | False |
| LDL.C | 2 | APOB | protein_coding | rs581411 | 21289432 | 7.61e-16 | 355 | 7 | 27 | 127 | 13 | 181 | Female only | Both sexes | Female only | True | False |
| LDL.C | 2 | RP11-116D2.1 | lincRNA | rs952275 | 21221399 | 2.21e-15 | 13 | 0 | 0 | 1 | 1 | 11 | Both sexes | Both sexes | Female only | True | False |
| LDL.C | 1 | PSRC1 | protein_coding | rs11577931 | 109820884 | 4.35e-15 | 11 | 0 | 0 | 1 | 5 | 5 | Female only | Both sexes | Female only | True | False |

| pheno | CHR | Gene_Symbol | biotype | lead_snp | lead_BP | lead_P | TOTAL | NSIG | S05 | S01 | S001 | S0001 | independent_loci_in_sex_P1e6 | independent_loci_in_sex_P1e2 | independent_loci_in_sex_P5e8 | in_gwas_catalog | in_HLA |
|---|---|---|---|---|---|---|---|---|---|---|---|---|---|---|---|---|---|
| LDL.C | 19 | BCAM | protein_coding | rs8113311 | 45325738 | 6.02e-13 | 3 | 0 | 1 | 1 | 0 | 1 | Female only | Both sexes | Female only | True | False |
| LDL.C | 19 | APOE | protein_coding | rs405509 | 45408836 | 2.97e-12 | 21 | 10 | 4 | 1 | 0 | 6 | Female only | Both sexes | Female only | True | False |
| LDL.C | 19 | APOC1 | protein_coding | rs59325138 | 45416291 | 5.65e-12 | 5 | 0 | 1 | 0 | 2 | 2 | Female only | Both sexes | Female only | True | False |
| LDL.C | 2 | TDRD15 | protein_coding | rs71435594 | 21309335 | 4.14e-11 | 3 | 0 | 0 | 1 | 1 | 1 | Female only | Both sexes | Female only | True | False |
| LDL.C | 6 | RP1-81D8.4 | pseudogene | rs2315065 | 161108144 | 8.48e-11 | 7 | 0 | 0 | 3 | 1 | 3 | Female only | Both sexes | Female only | True | False |
| LDL.C | 2 | AC067959.1 | lincRNA | rs67144154 | 21448511 | 4.54e-10 | 13 | 1 | 5 | 1 | 0 | 6 | Female only | Both sexes | Female only | True | False |
| LDL.C | 19 | BCL3 | protein_coding | rs2965101 | 45237812 | 1.61e-09 | 17 | 16 | 0 | 0 | 0 | 1 | Female only | Both sexes | Female only | True | False |
| LDL.C | 2 | AC067959.1 | lincRNA | rs13420469 | 21449034 | 3.05e-09 | 43 | 0 | 1 | 8 | 10 | 24 | Female only | Both sexes | Female only | True | False |
| LDL.C | 2 | RP11-116D2.1 | lincRNA | rs4075673 | 21150787 | 4.12e-09 | 11 | 4 | 1 | 1 | 3 | 2 | Female only | Both sexes | Female only | True | False |
| LDL.C | 2 | TDRD15 | protein_coding | rs4635554 | 21389659 | 5.76e-09 | 3 | 0 | 0 | 0 | 2 | 1 | Female only | Both sexes | Female only | True | False |
| LDL.C | 2 | TDRD15 | protein_coding | rs97458 | 21327635 | 1.97e-08 | 7 | 0 | 4 | 0 | 0 | 3 | Female only | Female only | Female only | True | False |
| LDL.C | 19 | LDLR | protein_coding | rs2569550 | 11228745 | 4.38e-08 | 37 | 0 | 2 | 3 | 16 | 16 | Female only | Both sexes | Female only | True | False |
| LDL.C | 5 | CTD-2235C13.2 | antisense | rs6871667 | 74604742 | 4.40e-08 | 87 | 1 | 16 | 27 | 0 | 43 | Female only | Both sexes | Female only | True | False |
| LYMPAB | 6 | HLA-C | protein_coding | rs9264642 | 31238490 | 6.96e-16 | 83 | 0 | 0 | 0 | 0 | 83 | Both sexes | Both sexes | Both sexes | True | True |
| LYMPAB | 6 | HLA-B | protein_coding | rs2394976 | 31311912 | 1.24e-13 | 263 | 0 | 0 | 2 | 2 | 259 | Both sexes | Both sexes | Both sexes | True | True |
| LYMPAB | 6 | HLA-C | protein_coding | rs2249742 | 31240721 | 3.42e-13 | 9 | 0 | 0 | 0 | 1 | 8 | Both sexes | Both sexes | Female only | True | True |
| LYMPAB | 6 | USP8P1 | pseudogene | rs2524075 | 31243859 | 2.98e-12 | 3 | 0 | 0 | 0 | 0 | 3 | Both sexes | Both sexes | Both sexes | True | True |
| LYMPAB | 6 | RNU6-1133P | snRNA | rs3095311 | 31051675 | 2.61e-11 | 127 | 0 | 0 | 4 | 6 | 117 | Both sexes | Both sexes | Both sexes | True | True |
| LYMPAB | 6 | TRIM26 | protein_coding | rs2517611 | 30169327 | 1.35e-10 | 147 | 0 | 0 | 0 | 0 | 147 | Both sexes | Both sexes | Both sexes | True | True |
| LYMPAB | 6 | UBQLN1P1 | pseudogene | rs3132634 | 30343272 | 1.54e-10 | 175 | 0 | 0 | 0 | 2 | 173 | Both sexes | Both sexes | Both sexes | True | True |
| LYMPAB | 6 | SKIV2L | protein_coding | rs437179 | 31929014 | 3.44e-10 | 41 | 0 | 0 | 0 | 1 | 40 | Both sexes | Both sexes | Male only | True | True |
| LYMPAB | 6 | TRIM31-AS1 | antisense | rs2523978 | 30083708 | 4.86e-10 | 113 | 0 | 0 | 1 | 2 | 110 | Both sexes | Both sexes | Female only | True | True |
| LYMPAB | 6 | HCG22 | lincRNA | rs2517524 | 31025713 | 7.06e-10 | 179 | 18 | 4 | 1 | 75 | 81 | Female only | Both sexes | Female only | True | True |
| pheno | CHR | Gene_Symbol | biotype | lead_snp | lead_BP | lead_P | TOTAL | NSIG | S05 | S01 | S001 | S0001 | independent_loci_in_sex_P1e6 | independent_loci_in_sex_P1e2 | independent_loci_in_sex_P5e8 | in_gwas_catalog | in_HLA |
| LYMPAB | 6 | HIST1H1B | protein_coding | rs17695758 | 27837183 | 8.94e-10 | 195 | 0 | 1 | 4 | 14 | 176 | Female only | Both sexes | Female only | True | False |
| LYMPAB | 6 | ZFP57 | protein_coding | rs547591961 | 29660816 | 1.03e-09 | 119 | 0 | 0 | 0 | 2 | 117 | Both sexes | Both sexes | Female only | True | True |

| pheno | CHR | Gene_Symbol | biotype | lead_snp | lead_BP | lead_P | TOTAL | NSIG | S05 | S01 | S001 | S0001 | independent_loci_in_sex_P1e6 | independent_loci_in_sex_P1e2 | independent_loci_in_sex_P5e8 | in_gwas_catalog | in_HLA |
|---|---|---|---|---|---|---|---|---|---|---|---|---|---|---|---|---|---|
| LYMPAB | 6 | LINC00243 | processed_transcript | rs1264347 | 30798697 | 1.15e-09 | 155 | 0 | 0 | 3 | 5 | 147 | Both sexes | Both sexes | Female only | True | True |
| LYMPAB | 6 | ZKSCAN4 | protein_coding | rs17720293 | 28214698 | 1.42e-09 | 191 | 1 | 0 | 6 | 34 | 150 | Female only | Both sexes | Female only | True | False |
| LYMPAB | 6 | XXbac-BPGBPG34I8.1 | pseudogene | rs34573979 | 27480526 | 1.49e-09 | 67 | 0 | 0 | 0 | 1 | 66 | Female only | Both sexes | Female only | True | False |
| LYMPAB | 6 | RPSAP2 | pseudogene | rs9468413 | 28689672 | 1.71e-09 | 259 | 0 | 0 | 0 | 0 | 259 | Both sexes | Both sexes | Female only | True | False |
| LYMPAB | 19 | EPS15L1 | protein_coding | rs4808047 | 16527834 | 2.67e-09 | 21 | 0 | 0 | 2 | 7 | 12 | Both sexes | Both sexes | Female only | True | False |
| LYMPAB | 6 | OR5V1 | protein_coding | rs9257809 | 29356331 | 3.43e-09 | 75 | 0 | 0 | 0 | 0 | 75 | Both sexes | Both sexes | Female only | True | False |
| LYMPAB | 6 | CSNK2B | protein_coding | rs9267531 | 31636742 | 3.98e-09 | 129 | 0 | 0 | 0 | 0 | 129 | Both sexes | Both sexes | Both sexes | True | True |
| LYMPAB | 12 | CD69 | protein_coding | ss1388113948 | 9931347 | 4.82e-09 | 33 | 0 | 0 | 16 | 1 | 16 | Female only | Both sexes | Female only | True | False |
| LYMPAB | 6 | COX11P1 | pseudogene | rs61289879 | 28419162 | 5.72e-09 | 263 | 0 | 4 | 11 | 39 | 209 | Female only | Both sexes | Female only | True | False |
| LYMPAB | 12 | CD69 | protein_coding | ss1388113960 | 9931643 | 8.11e-09 | 35 | 1 | 12 | 5 | 1 | 16 | Female only | Both sexes | Female only | True | False |
| LYMPAB | 6 | HLA-DQA1 | protein_coding | rs9271573 | 32590501 | 1.01e-08 | 69 | 1 | 1 | 27 | 11 | 29 | Female only | Both sexes | Female only | True | True |
| LYMPAB | 6 | RPSAP2 | pseudogene | rs1233585 | 28716345 | 1.01e-08 | 29 | 1 | 0 | 0 | 5 | 23 | Male only | Both sexes | Male only | True | False |
| LYMPAB | 6 | HCP5 | sense_overlapping | rs3099840 | 31430721 | 1.14e-08 | 3 | 0 | 0 | 2 | 0 | 1 | Female only | Both sexes | Female only | True | True |
| LYMPAB | 6 | HLA-V | pseudogene | rs1633005 | 29764472 | 1.67e-08 | 525 | 0 | 113 | 23 | 153 | 236 | Both sexes | Both sexes | Male only | True | True |
| LYMPAB | 6 | HLA-B | protein_coding | rs9266071 | 31319620 | 1.91e-08 | 69 | 2 | 23 | 6 | 4 | 34 | Female only | Both sexes | Female only | True | True |
| LYMPAB | 6 | XXbac-BCX196D17.5 | lincRNA | rs3130773 | 29095908 | 2.45e-08 | 39 | 0 | 0 | 0 | 0 | 39 | Female only | Both sexes | Female only | True | False |
| LYMPAB | 6 | TNXB | protein_coding | rs1150752 | 32064726 | 3.20e-08 | 139 | 0 | 0 | 6 | 29 | 104 | Both sexes | Both sexes | Both sexes | True | True |
| LYMPAB | 6 | HLA-C | protein_coding | rs9264185 | 31219982 | 3.99e-08 | 151 | 1 | 10 | 52 | 16 | 72 | Female only | Both sexes | Female only | True | True |
| LYMPM | 23 | RP11-585K14.2 | pseudogene | X:5094786:G:A | 5094786 | 1.00e-08 | 141 | 71 | 1 | 7 | 35 | 27 | Female only | Female only | Female only | True | False |
| LYMPRE | 1 | RP11-334L9.1 | pseudogene | rs3011200 | 39079403 | 5.31e-09 | 15 | 8 | 0 | 0 | 0 | 7 | Female only | Female only | Female only | True | False |
| Lipldx | 19 | APOC1 | protein_coding | rs5117 | 45418790 | 4.80e-09 | 11 | 0 | 2 | 1 | 5 | 3 | Male only | Both sexes | Male only | True | False |
| LucAbs | 17 | BZRAP1-AS1 | antisense | rs75394768 | 56413635 | 4.42e-09 | 3 | 0 | 0 | 0 | 0 | 3 | Female only | Both sexes | Female only | True | False |
| Luc_percent_ | 17 | BZRAP1-AS1 | antisense | rs75394768 | 56413635 | 1.32e-09 | 3 | 0 | 0 | 0 | 0 | 3 | Both sexes | Both sexes | Both sexes | False | False |
| LymAbs | 6 | HLA-C | protein_coding | rs9264642 | 31238490 | 3.49e-11 | 83 | 0 | 9 | 29 | 4 | 41 | Female only | Both sexes | Female only | True | True |
| LymAbs | 6 | ZDHHC20P2 | pseudogene | rs2596570 | 31348040 | 1.15e-09 | 241 | 0 | 0 | 21 | 97 | 123 | Female only | Both sexes | Female only | True | True |

| pheno | CHR | Gene_Symbol | biotype | lead_snp | lead_BP | lead_P | TOTAL | NSIG | S05 | S01 | S001 | S0001 | independent_loci_in_sex_P1e6 | independent_loci_in_sex_P1e2 | independent_loci_in_sex_P5e8 | in_gwas_catalog | in_HLA |
|---|---|---|---|---|---|---|---|---|---|---|---|---|---|---|---|---|---|
| LymAbs | 6 | HLA-C | protein_coding | rs2249742 | 31240721 | 1.41e-09 | 9 | 2 | 0 | 3 | 1 | 3 | Female only | Both sexes | Female only | True | True |
| LymAbs | 6 | USP8P1 | pseudogene | rs2524075 | 31243859 | 6.27e-09 | 3 | 0 | 0 | 0 | 2 | 1 | Female only | Both sexes | Female only | True | True |
| LymAbs | 6 | HCG20 | lincRNA | rs3129984 | 30761572 | 3.48e-08 | 247 | 0 | 2 | 49 | 65 | 131 | Female only | Both sexes | Female only | True | True |
| LymAbs | 22 | GGT5 | protein_coding | rs2267073 | 24639069 | 4.07e-08 | 53 | 27 | 0 | 7 | 17 | 2 | Male only | Male only | Male only | True | False |
| MBRat | 19 | LILRB5 | protein_coding | rs12975366 | 54759361 | 4.61e-10 | 7 | 0 | 0 | 0 | 1 | 6 | Male only | Both sexes | Male only | True | False |
| MBRat | 19 | LILRB5 | protein_coding | rs380731 | 54759571 | 3.84e-08 | 5 | 0 | 0 | 3 | 0 | 2 | Male only | Both sexes | Male only | True | False |
| MCH | 22 | TMPRSS6 | protein_coding | rs2076085 | 37470041 | 7.45e-98 | 9 | 0 | 0 | 0 | 0 | 9 | Both sexes | Both sexes | Both sexes | True | False |
| MCH | 6 | HBS1L | protein_coding | rs9376091 | 135419636 | 7.20e-88 | 35 | 0 | 0 | 0 | 0 | 35 | Both sexes | Both sexes | Both sexes | True | False |
| MCH | 6 | HFE | protein_coding | rs1800562 | 26093141 | 1.67e-64 | 15 | 0 | 0 | 0 | 0 | 15 | Both sexes | Both sexes | Both sexes | True | False |
| MCH | 6 | SLC17A3 | protein_coding | rs1408272 | 25842951 | 1.96e-54 | 9 | 0 | 0 | 0 | 0 | 9 | Both sexes | Both sexes | Both sexes | True | False |
| MCH | 6 | HIST1H1T | protein_coding | rs198846 | 26107463 | 5.86e-54 | 33 | 0 | 0 | 0 | 0 | 33 | Both sexes | Both sexes | Both sexes | True | False |
| MCH | 22 | TMPRSS6 | protein_coding | rs5756504 | 37467270 | 5.53e-50 | 35 | 0 | 0 | 0 | 0 | 35 | Both sexes | Both sexes | Both sexes | True | False |
| MCH | 6 | HBS1L | protein_coding | rs6569992 | 135452152 | 5.83e-46 | 31 | 0 | 0 | 0 | 0 | 31 | Both sexes | Both sexes | Both sexes | True | False |
| MCH | 6 | HBS1L | protein_coding | rs1320959 | 135437386 | 9.03e-41 | 87 | 0 | 0 | 0 | 0 | 87 | Both sexes | Both sexes | Both sexes | True | False |
| MCH | 6 | LRRC16A | protein_coding | rs149359690 | 25526319 | 1.25e-38 | 45 | 0 | 0 | 0 | 0 | 45 | Both sexes | Both sexes | Both sexes | True | False |
| MCH | 6 | HIST1H3C | protein_coding | 6:26046744 | 26046744 | 2.90e-35 | 245 | 0 | 3 | 10 | 6 | 226 | Both sexes | Both sexes | Both sexes | True | False |
| MCH | 22 | TMPRSS6 | protein_coding | rs9607412 | 37484337 | 4.85e-34 | 57 | 0 | 0 | 0 | 0 | 57 | Both sexes | Both sexes | Both sexes | True | False |
| MCH | 6 | SCGN | protein_coding | rs2328891 | 25669669 | 2.03e-33 | 25 | 0 | 0 | 0 | 0 | 25 | Both sexes | Both sexes | Both sexes | True | False |
| MCH | 6 | LRRC16A | protein_coding | rs74662487 | 25453057 | 2.59e-33 | 17 | 0 | 0 | 0 | 0 | 17 | Both sexes | Both sexes | Both sexes | True | False |
| MCH | 6 | MIR3143 | miRNA | rs71559014 | 27122444 | 8.39e-31 | 3 | 0 | 0 | 0 | 0 | 3 | Both sexes | Both sexes | Both sexes | True | False |
| MCH | 7 | TFR2 | protein_coding | rs2075672 | 100240296 | 2.90e-30 | 43 | 0 | 0 | 0 | 0 | 43 | Both sexes | Both sexes | Both sexes | True | False |
| MCH | 6 | U91328.22 | lincRNA | rs9356996 | 26008487 | 1.54e-27 | 63 | 0 | 0 | 0 | 0 | 63 | Both sexes | Both sexes | Both sexes | True | False |
| MCH | 22 | TMPRSS6 | protein_coding | rs2235325 | 37481637 | 2.51e-26 | 23 | 0 | 0 | 0 | 0 | 23 | Both sexes | Both sexes | Both sexes | True | False |
| MCH | 4 | AC006552.1 | lincRNA | rs218265 | 55408999 | 1.07e-25 | 13 | 0 | 0 | 0 | 0 | 13 | Both sexes | Both sexes | Both sexes | True | False |
| MCH | 6 | HIST1H2BO | protein_coding | 6:27861670 | 27861670 | 1.71e-25 | 9 | 0 | 0 | 0 | 0 | 9 | Both sexes | Both sexes | Both sexes | True | False |
| MCH | 6 | LRRC16A | protein_coding | rs111722075 | 25374046 | 3.02e-25 | 253 | 0 | 0 | 0 | 0 | 253 | Both sexes | Both sexes | Both sexes | True | False |

| pheno | CHR | Gene_Symbol | biotype | lead_snp | lead_BP | lead_P | TOTAL | NSIG | S05 | S01 | S001 | S0001 | independent_loci_in_sex_P1e6 | independent_loci_in_sex_P1e2 | independent_loci_in_sex_P5e8 | in_gwas_catalog | in_HLA |
|---|---|---|---|---|---|---|---|---|---|---|---|---|---|---|---|---|---|
| MCH | 6 | ZSCAN31 | protein_coding | rs35814746 | 28299088 | 3.92e-24 | 41 | 0 | 0 | 0 | 0 | 41 | Both sexes | Both sexes | Both sexes | True | False |
| MCH | 22 | MPST | protein_coding | rs5756488 | 37416803 | 5.42e-24 | 7 | 0 | 0 | 0 | 0 | 7 | Both sexes | Both sexes | Both sexes | True | False |
| MCH | 6 | C6orf183 | polymorphic_pseudogene | rs9487023 | 109590004 | 8.07e-24 | 379 | 11 | 4 | 21 | 42 | 301 | Both sexes | Both sexes | Both sexes | True | False |
| MCH | 6 | HIST1H2BC | protein_coding | rs198839 | 26112620 | 9.65e-24 | 13 | 0 | 0 | 0 | 2 | 11 | Both sexes | Both sexes | Both sexes | True | False |
| MCH | 4 | AC006552.1 | lincRNA | rs170121 | 55407099 | 1.30e-23 | 15 | 0 | 0 | 0 | 0 | 15 | Both sexes | Both sexes | Both sexes | True | False |
| MCH | 6 | HIST1H2APS2 | pseudogene | rs72830103 | 25897618 | 6.50e-23 | 51 | 0 | 0 | 0 | 0 | 51 | Both sexes | Both sexes | Both sexes | True | False |
| MCH | 6 | HBS1L | protein_coding | rs41294858 | 135412636 | 7.72e-23 | 23 | 0 | 0 | 0 | 0 | 23 | Both sexes | Both sexes | Both sexes | True | False |

| pheno | CHR | Gene_Symbol | biotype | lead_snp | lead_BP | lead_P | TOTAL | NSIG | S05 | S01 | S001 | S0001 | independent_loci_in_sex_P1e6 | independent_loci_in_sex_P1e2 | independent_loci_in_sex_P5e8 | in_gwas_catalog | in_HLA |
|---|---|---|---|---|---|---|---|---|---|---|---|---|---|---|---|---|---|
| MCH | 6 | LRRC16A | protein_coding | rs181058325 | 25314597 | 2.82e-21 | 11 | 0 | 0 | 0 | 0 | 11 | Both sexes | Both sexes | Both sexes | True | False |
| MCH | 6 | LRRC16A | protein_coding | rs13203202 | 25582771 | 4.82e-21 | 9 | 0 | 0 | 0 | 0 | 9 | Both sexes | Both sexes | Both sexes | True | False |
| MCH | 6 | CCND3 | protein_coding | rs9349205 | 41925159 | 5.03e-21 | 73 | 0 | 0 | 0 | 0 | 73 | Both sexes | Both sexes | Both sexes | True | False |
| MCH | 6 | HBS1L | protein_coding | rs10484494 | 135430093 | 5.83e-21 | 3 | 0 | 0 | 0 | 0 | 3 | Both sexes | Both sexes | Both sexes | True | False |
| MCH | 6 | CCND3 | protein_coding | rs4365924 | 41995361 | 6.84e-20 | 5 | 0 | 0 | 0 | 0 | 5 | Both sexes | Both sexes | Both sexes | True | False |
| MCH | 6 | MYB | protein_coding | rs55683605 | 135487107 | 9.12e-20 | 29 | 0 | 0 | 0 | 0 | 29 | Both sexes | Both sexes | Both sexes | True | False |
| MCH | 19 | KLF1 | protein_coding | rs8110787 | 12999458 | 1.42e-19 | 75 | 0 | 0 | 0 | 1 | 74 | Both sexes | Both sexes | Both sexes | True | False |
| MCH | 6 | XXbac-BPG308K3.5 | lincRNA | rs3118362 | 28785085 | 3.41e-19 | 5 | 0 | 0 | 0 | 0 | 5 | Both sexes | Both sexes | Both sexes | True | False |
| MCH | 6 | CCND3 | protein_coding | rs3218092 | 41906661 | 1.30e-18 | 57 | 0 | 0 | 0 | 0 | 57 | Both sexes | Both sexes | Both sexes | True | False |
| MCH | 4 | RP11-545H22.1 | lincRNA | rs11133333 | 55380471 | 2.58e-18 | 55 | 3 | 1 | 2 | 7 | 42 | Female only | Both sexes | Female only | True | False |
| MCH | 22 | ODF3B | protein_coding | rs361725 | 50971047 | 3.94e-18 | 15 | 0 | 0 | 0 | 0 | 15 | Both sexes | Both sexes | Both sexes | True | False |
| MCH | 22 | TMPRSS6 | protein_coding | rs6000550 | 37463450 | 4.32e-18 | 15 | 0 | 0 | 0 | 0 | 15 | Both sexes | Both sexes | Both sexes | True | False |
| MCH | 15 | DENND4A | protein_coding | rs7497961 | 66020908 | 4.62e-18 | 135 | 66 | 0 | 2 | 0 | 67 | Female only | Both sexes | Female only | True | False |
| MCH | 6 | LRRC16A | protein_coding | rs72831267 | 25436955 | 1.82e-17 | 7 | 0 | 0 | 0 | 0 | 7 | Both sexes | Both sexes | Both sexes | True | False |
| MCH | 16 | NPRL3 | protein_coding | rs11248853 | 166960 | 6.05e-17 | 11 | 0 | 0 | 0 | 0 | 11 | Both sexes | Both sexes | Both sexes | True | False |
| MCH | 6 | LRRC16A | protein_coding | rs9358860 | 25572245 | 8.14e-17 | 53 | 46 | 0 | 0 | 0 | 7 | Both sexes | Both sexes | Both sexes | True | False |
| MCH | 6 | RP11-12A2.3 | lincRNA | rs590856 | 139844429 | 1.45e-16 | 45 | 0 | 0 | 0 | 0 | 45 | Both sexes | Both sexes | Both sexes | True | False |
| MCH | 6 | TRIM38 | protein_coding | rs199739 | 25960509 | 1.97e-16 | 47 | 22 | 0 | 0 | 1 | 24 | Both sexes | Both sexes | Female only | True | False |

| pheno | CHR | Gene_Symbol | biotype | lead_snp | lead_BP | lead_P | TOTAL | NSIG | S05 | S01 | S001 | S0001 | independent_loci_in_sex_P1e6 | independent_loci_in_sex_P1e2 | independent_loci_in_sex_P5e8 | in_gwas_catalog | in_HLA |
|---|---|---|---|---|---|---|---|---|---|---|---|---|---|---|---|---|---|
| MCH | 6 | SLC17A1 | protein_coding | rs12191655 | 25803095 | 3.17e-16 | 297 | 0 | 0 | 133 | 9 | 155 | Both sexes | Both sexes | Female only | True | False |
| MCH | 6 | UBQLN1P1 | pseudogene | rs2057726 | 30335350 | 3.36e-16 | 47 | 0 | 0 | 0 | 0 | 47 | Both sexes | Both sexes | Both sexes | True | True |
| MCH | 9 | RCL1 | protein_coding | rs10758656 | 4852599 | 4.35e-16 | 43 | 0 | 0 | 0 | 2 | 41 | Both sexes | Both sexes | Female only | True | False |
| MCH | 6 | LRRC16A | protein_coding | rs112268831 | 25348760 | 5.36e-16 | 5 | 0 | 0 | 0 | 0 | 5 | Both sexes | Both sexes | Female only | True | False |
| MCH | 6 | SCGN | protein_coding | rs4566883 | 25693581 | 8.45e-16 | 31 | 0 | 0 | 8 | 2 | 21 | Both sexes | Both sexes | Both sexes | True | False |
| MCH | 6 | MYB | protein_coding | rs9389278 | 135525927 | 1.36e-15 | 31 | 6 | 3 | 1 | 0 | 21 | Both sexes | Both sexes | Both sexes | True | False |
| MCH | 6 | HLA-W | pseudogene | rs9260707 | 29929536 | 1.83e-15 | 165 | 0 | 0 | 0 | 0 | 165 | Both sexes | Both sexes | Both sexes | True | True |
| MCH | 6 | CCND3 | protein_coding | rs12175190 | 41983371 | 2.23e-15 | 13 | 0 | 2 | 0 | 2 | 9 | Both sexes | Both sexes | Both sexes | True | False |
| MCH | 6 | HIST1H3PS1 | pseudogene | rs6932727 | 26327087 | 2.51e-15 | 95 | 0 | 0 | 0 | 0 | 95 | Both sexes | Both sexes | Both sexes | True | False |
| MCH | 7 | TFR2 | protein_coding | rs6958696 | 100237597 | 3.54e-15 | 79 | 0 | 0 | 0 | 0 | 79 | Both sexes | Both sexes | Both sexes | True | False |
| MCH | 22 | TST | protein_coding | rs5756477 | 37407527 | 4.42e-15 | 57 | 0 | 0 | 0 | 0 | 57 | Both sexes | Both sexes | Both sexes | True | False |
| MCH | 4 | AC006552.1 | lincRNA | rs80113191 | 55396420 | 5.56e-15 | 101 | 0 | 0 | 0 | 0 | 101 | Both sexes | Both sexes | Both sexes | True | False |
| MCH | 15 | RAB11A | protein_coding | rs8027781 | 66097711 | 9.91e-15 | 71 | 0 | 0 | 0 | 0 | 71 | Female only | Both sexes | Female only | True | False |
| MCH | 6 | CCDC162P | pseudogene | rs59272337 | 109638895 | 2.24e-14 | 103 | 51 | 0 | 1 | 0 | 51 | Female only | Both sexes | Female only | True | False |
| MCH | 6 | HIST1H2BC | protein_coding | rs707896 | 26116424 | 5.60e-14 | 51 | 4 | 1 | 0 | 5 | 41 | Both sexes | Both sexes | Both sexes | True | False |
| MCH | 6 | HBS1L | protein_coding | rs9494139 | 135414793 | 7.84e-14 | 11 | 0 | 0 | 0 | 0 | 11 | Both sexes | Both sexes | Both sexes | True | False |
| MCH | 14 | FNTB | protein_coding | rs11627485 | 65487694 | 1.14e-13 | 37 | 0 | 6 | 0 | 6 | 25 | Both sexes | Both sexes | Both sexes | True | False |
| MCH | 19 | HDGFRP2 | protein_coding | rs3816048 | 4475606 | 1.33e-13 | 47 | 3 | 4 | 0 | 0 | 40 | Both sexes | Both sexes | Both sexes | True | False |
| MCH | 6 | SLC17A3 | protein_coding | rs566530 | 25878361 | 2.20e-13 | 123 | 11 | 0 | 17 | 34 | 61 | Female only | Both sexes | Female only | True | False |
| MCH | 6 | HBS1L | protein_coding | rs6930223 | 135424203 | 2.52e-13 | 29 | 18 | 0 | 1 | 1 | 9 | Both sexes | Both sexes | Both sexes | True | False |
| MCH | 6 | HFE | protein_coding | rs2071303 | 26091336 | 2.80e-13 | 51 | 0 | 0 | 2 | 23 | 26 | Both sexes | Both sexes | Both sexes | True | False |
| MCH | 6 | LRRC16A | protein_coding | rs9379764 | 25414023 | 3.92e-13 | 3 | 0 | 0 | 0 | 0 | 3 | Both sexes | Both sexes | Both sexes | True | False |
| MCH | 6 | MOG | protein_coding | rs3130253 | 29634012 | 4.03e-13 | 21 | 0 | 0 | 0 | 4 | 17 | Both sexes | Both sexes | Both sexes | True | True |
| MCH | 18 | C18orf25 | protein_coding | rs6507670 | 43734409 | 5.31e-13 | 55 | 0 | 0 | 0 | 0 | 55 | Both sexes | Both sexes | Both sexes | True | False |
| MCH | 10 | MARCH8 | protein_coding | rs34654893 | 45979828 | 7.78e-13 | 431 | 0 | 0 | 0 | 5 | 426 | Female only | Both sexes | Female only | True | False |
| MCH | 22 | TMPRSS6 | protein_coding | rs11089824 | 37509186 | 9.61e-13 | 5 | 0 | 0 | 0 | 0 | 5 | Both sexes | Both sexes | Female only | True | False |

| pheno | CHR | Gene_Symbol | biotype | lead_snp | lead_BP | lead_P | TOTAL | NSIG | S05 | S01 | S001 | S0001 | independent_loci_in_sex_P1e6 | independent_loci_in_sex_P1e2 | independent_loci_in_sex_P5e8 | in_gwas_catalog | in_HLA |
|---|---|---|---|---|---|---|---|---|---|---|---|---|---|---|---|---|---|
| MCH | 15 | DPP8 | protein_coding | rs4776688 | 65795633 | 2.12e-12 | 29 | 0 | 0 | 0 | 0 | 29 | Female only | Both sexes | Female only | True | False |
| MCH | 7 | IKZF1 | protein_coding | rs12718731 | 50436948 | 2.17e-12 | 11 | 0 | 0 | 0 | 0 | 11 | Both sexes | Both sexes | Both sexes | True | False |
| MCH | 6 | RP3-352A20.1 | pseudogene | rs73547141 | 135215416 | 3.86e-12 | 57 | 0 | 0 | 0 | 0 | 57 | Both sexes | Both sexes | Both sexes | True | False |
| MCH | 6 | BTN2A3P | pseudogene | rs4320356 | 26423560 | 4.11e-12 | 73 | 0 | 4 | 17 | 17 | 35 | Female only | Both sexes | Female only | True | False |
| MCH | 19 | HOOK2 | protein_coding | rs55792045 | 12933914 | 7.60e-12 | 69 | 2 | 0 | 27 | 4 | 36 | Female only | Both sexes | Female only | True | False |
| MCH | 7 | ZAN | polymorphic_pseudogene | rs62483597 | 100363821 | 1.23e-11 | 11 | 0 | 0 | 0 | 0 | 11 | Both sexes | Both sexes | Female only | True | False |
| MCH | 16 | NPRL3 | protein_coding | rs2562189 | 187429 | 1.79e-11 | 3 | 0 | 0 | 0 | 0 | 3 | Both sexes | Both sexes | Both sexes | True | False |
| MCH | 6 | HBS1L | protein_coding | rs61738647 | 135282735 | 1.87e-11 | 287 | 0 | 0 | 54 | 40 | 193 | Both sexes | Both sexes | Both sexes | True | False |
| MCH | 6 | HMGN4 | protein_coding | rs12526680 | 26550954 | 2.06e-11 | 279 | 2 | 0 | 87 | 52 | 138 | Female only | Both sexes | Female only | True | False |
| MCH | 3 | TFRC | protein_coding | rs41298085 | 195779812 | 2.09e-11 | 49 | 0 | 0 | 0 | 0 | 49 | Both sexes | Both sexes | Male only | True | False |
| MCH | 18 | C18orf25 | protein_coding | rs8095374 | 43793488 | 2.45e-11 | 5 | 0 | 0 | 0 | 0 | 5 | Both sexes | Both sexes | Both sexes | True | False |
| MCH | 6 | C6orf183 | polymorphic_pseudogene | rs4946953 | 109583631 | 2.68e-11 | 45 | 0 | 0 | 3 | 0 | 42 | Female only | Both sexes | Female only | True | False |
| MCH | 6 | GPX6 | protein_coding | rs10428759 | 28477682 | 2.75e-11 | 9 | 0 | 0 | 0 | 0 | 9 | Female only | Both sexes | Female only | True | False |
| MCH | 3 | LINC00885 | lincRNA | rs4504190 | 195839847 | 3.28e-11 | 5 | 0 | 0 | 0 | 0 | 5 | Both sexes | Both sexes | Both sexes | True | False |
| MCH | 6 | C6orf183 | polymorphic_pseudogene | rs36052053 | 109583799 | 3.85e-11 | 77 | 0 | 0 | 0 | 9 | 68 | Both sexes | Both sexes | Female only | True | False |
| MCH | 6 | TRIM15 | protein_coding | rs2844787 | 30140913 | 4.66e-11 | 33 | 0 | 0 | 0 | 0 | 33 | Female only | Both sexes | Female only | True | True |
| pheno | CHR | Gene_Symbol | biotype | lead_snp | lead_BP | lead_P | TOTAL | NSIG | S05 | S01 | S001 | S0001 | independent_loci_in_sex_P1e6 | independent_loci_in_sex_P1e2 | independent_loci_in_sex_P5e8 | in_gwas_catalog | in_HLA |
| MCH | 6 | ATAT1 | protein_coding | rs3094106 | 30599176 | 4.97e-11 | 57 | 0 | 0 | 0 | 20 | 37 | Both sexes | Both sexes | Female only | True | True |
| MCH | 19 | CTD-2540B15.12 | lincRNA | rs78744187 | 33754548 | 5.50e-11 | 5 | 0 | 0 | 0 | 1 | 4 | Female only | Both sexes | Female only | True | False |
| MCH | 7 | GIGYF1 | protein_coding | rs221784 | 100292830 | 5.59e-11 | 49 | 0 | 0 | 12 | 3 | 34 | Female only | Both sexes | Female only | True | False |
| MCH | 10 | MARCH8 | protein_coding | rs11239550 | 46024729 | 9.03e-11 | 73 | 14 | 8 | 19 | 8 | 24 | Female only | Both sexes | Female only | True | False |
| MCH | 6 | ZSCAN23 | protein_coding | rs13190937 | 28411244 | 9.39e-11 | 71 | 35 | 9 | 13 | 0 | 14 | Female only | Both sexes | Female only | True | False |
| MCH | 6 | RP3-352A20.1 | pseudogene | rs2327576 | 135177685 | 1.12e-10 | 47 | 10 | 8 | 0 | 0 | 29 | Both sexes | Both sexes | Male only | True | False |
| MCH | 3 | TFRC | protein_coding | rs62285386 | 195817478 | 1.15e-10 | 29 | 0 | 0 | 0 | 0 | 29 | Both sexes | Both sexes | Male only | True | False |
| MCH | 6 | RP3-352A20.1 | pseudogene | rs4896115 | 135168723 | 1.17e-10 | 41 | 0 | 0 | 0 | 15 | 26 | Both sexes | Both sexes | Both sexes | True | False |
| MCH | 22 | MPST | protein_coding | rs6000536 | 37421469 | 1.68e-10 | 27 | 20 | 0 | 0 | 3 | 4 | Both sexes | Both sexes | Female only | True | False |

| pheno | CHR | Gene_Symbol | biotype | lead_snp | lead_BP | lead_P | TOTAL | NSIG | S05 | S01 | S001 | S0001 | independent_loci_in_sex_P1e6 | independent_loci_in_sex_P1e2 | independent_loci_in_sex_P5e8 | in_gwas_catalog | in_HLA |
|---|---|---|---|---|---|---|---|---|---|---|---|---|---|---|---|---|---|
| MCH | 6 | SLC17A3 | protein_coding | rs56069761 | 25878273 | 1.68e-10 | 5 | 0 | 0 | 0 | 0 | 5 | Female only | Both sexes | Female only | True | False |
| MCH | 6 | SLC17A3 | protein_coding | rs1165201 | 25874823 | 2.67e-10 | 55 | 6 | 0 | 6 | 16 | 27 | Female only | Both sexes | Female only | True | False |
| MCH | 6 | HIST1H3F | protein_coding | rs78545713 | 26248840 | 3.84e-10 | 223 | 0 | 26 | 24 | 39 | 134 | Both sexes | Both sexes | Male only | True | False |
| MCH | 2 | MIR4435-1HG | lincRNA | 2:112208126 | 112208126 | 4.34e-10 | 3 | 2 | 0 | 0 | 0 | 1 | Female only | Female only | Female only | True | False |
| MCH | 6 | OR2J3 | protein_coding | rs3129106 | 29081161 | 4.73e-10 | 3 | 0 | 0 | 0 | 1 | 2 | Female only | Both sexes | Female only | True | False |
| MCH | 22 | SCO2 | protein_coding | rs131805 | 50964153 | 5.29e-10 | 11 | 0 | 0 | 0 | 0 | 11 | Both sexes | Both sexes | Female only | True | False |
| MCH | 6 | TRIM31-AS1 | antisense | rs1419675 | 30092707 | 1.06e-09 | 5 | 1 | 0 | 2 | 0 | 2 | Female only | Both sexes | Female only | True | True |
| MCH | 4 | RP11-545H22.1 | lincRNA | rs1553663 | 55280100 | 1.20e-09 | 31 | 10 | 10 | 4 | 5 | 2 | Female only | Both sexes | Female only | True | False |
| MCH | 22 | LL22NC01-81G9.3 | antisense | rs130437 | 37369465 | 1.25e-09 | 53 | 7 | 0 | 14 | 13 | 19 | Female only | Both sexes | Female only | True | False |
| MCH | 14 | FNTB | protein_coding | rs79191167 | 65491292 | 1.27e-09 | 47 | 0 | 0 | 0 | 0 | 47 | Both sexes | Both sexes | Both sexes | True | False |
| MCH | 6 | SCGN | protein_coding | rs4712949 | 25644203 | 1.48e-09 | 179 | 0 | 2 | 1 | 0 | 176 | Female only | Both sexes | Female only | True | False |
| MCH | 6 | MYB | protein_coding | rs1013891 | 135538639 | 1.77e-09 | 3 | 2 | 0 | 0 | 0 | 1 | Female only | Both sexes | Female only | True | False |
| MCH | 2 | BCL11A | protein_coding | rs10189857 | 60713235 | 2.21e-09 | 9 | 3 | 0 | 2 | 0 | 4 | Female only | Both sexes | Female only | True | False |
| MCH | 6 | LRRC16A | protein_coding | rs9366617 | 25327270 | 2.29e-09 | 37 | 6 | 0 | 12 | 4 | 15 | Female only | Both sexes | Female only | True | False |
| MCH | 6 | CCND3 | protein_coding | rs7766960 | 41918190 | 2.36e-09 | 89 | 0 | 0 | 8 | 9 | 72 | Female only | Both sexes | Female only | True | False |
| MCH | 9 | RCL1 | protein_coding | rs1576065 | 4857802 | 2.47e-09 | 59 | 12 | 9 | 9 | 19 | 10 | Female only | Both sexes | Female only | True | False |
| MCH | 6 | HLA-B | protein_coding | rs2523616 | 31320562 | 2.67e-09 | 295 | 23 | 117 | 8 | 0 | 147 | Male only | Both sexes | Male only | True | True |
| MCH | 6 | SLC17A1 | protein_coding | rs62392759 | 25804685 | 4.16e-09 | 73 | 0 | 0 | 0 | 1 | 72 | Female only | Both sexes | Female only | True | False |
| MCH | 6 | DDR1 | protein_coding | rs3094719 | 30844972 | 4.17e-09 | 37 | 0 | 0 | 8 | 10 | 19 | Both sexes | Both sexes | Female only | True | True |
| MCH | 6 | MICD | pseudogene | rs1061539 | 29937555 | 4.82e-09 | 3 | 0 | 0 | 0 | 0 | 3 | Female only | Both sexes | Female only | True | True |
| MCH | 4 | RP11-545H22.1 | lincRNA | rs6846679 | 55305505 | 4.98e-09 | 97 | 41 | 8 | 20 | 9 | 19 | Female only | Both sexes | Female only | True | False |
| MCH | 6 | LRRC16A | protein_coding | rs12200019 | 25430896 | 6.29e-09 | 9 | 0 | 0 | 0 | 3 | 6 | Female only | Both sexes | Female only | True | False |
| MCH | 6 | TRIM40 | protein_coding | rs3132676 | 30116078 | 7.39e-09 | 15 | 0 | 0 | 0 | 0 | 15 | Female only | Both sexes | Female only | True | True |
| MCH | 6 | TRIM39 | protein_coding | rs1110464 | 30296071 | 7.61e-09 | 7 | 3 | 0 | 1 | 0 | 3 | Male only | Both sexes | Male only | True | True |
| MCH | 10 | HK1 | protein_coding | rs16926246 | 71093392 | 8.88e-09 | 11 | 0 | 0 | 0 | 1 | 10 | Female only | Both sexes | Female only | True | False |
| MCH | 6 | BTN3A1 | protein_coding | rs3799378 | 26404374 | 9.02e-09 | 21 | 0 | 7 | 2 | 6 | 6 | Female only | Both sexes | Female only | True | False |

| pheno | CHR | Gene_Symbol | biotype | lead_snp | lead_BP | lead_P | TOTAL | NSIG | S05 | S01 | S001 | S0001 | independent_loci_in_sex_P1e6 | independent_loci_in_sex_P1e2 | independent_loci_in_sex_P5e8 | in_gwas_catalog | in_HLA |
|---|---|---|---|---|---|---|---|---|---|---|---|---|---|---|---|---|---|
| MCH | 4 | RP11-545H22.1 | lincRNA | rs13151127 | 55358779 | 1.08e-08 | 33 | 0 | 0 | 0 | 9 | 24 | Female only | Both sexes | Female only | True | False |
| MCH | 3 | TNK2 | protein_coding | rs9871110 | 195625326 | 1.11e-08 | 7 | 1 | 3 | 0 | 0 | 3 | Male only | Male only | Male only | True | False |
| MCH | 6 | VN1R10P | pseudogene | rs9379980 | 27306518 | 1.18e-08 | 77 | 10 | 12 | 28 | 7 | 20 | Female only | Both sexes | Female only | True | False |
| MCH | 6 | RP11-457M11.5 | lincRNA | rs4713037 | 26758030 | 1.29e-08 | 77 | 26 | 13 | 0 | 1 | 37 | Female only | Female only | Female only | True | False |
| MCH | 19 | FARSA | protein_coding | rs2967890 | 13040647 | 1.42e-08 | 3 | 0 | 0 | 0 | 0 | 3 | Both sexes | Both sexes | Female only | True | False |
| MCH | 6 | HIST1H4F | protein_coding | rs9358913 | 26239404 | 1.73e-08 | 13 | 0 | 0 | 0 | 3 | 10 | Male only | Both sexes | Male only | True | False |
| MCH | 7 | PILRA | protein_coding | rs62482199 | 99968371 | 1.80e-08 | 9 | 1 | 1 | 1 | 2 | 4 | Female only | Both sexes | Female only | True | False |
| MCH | 3 | TFRC | protein_coding | rs34221479 | 195798600 | 1.85e-08 | 49 | 0 | 0 | 0 | 0 | 49 | Both sexes | Both sexes | Male only | True | False |
| MCH | 3 | LINC00885 | lincRNA | rs9831818 | 195878035 | 1.91e-08 | 63 | 2 | 7 | 0 | 0 | 54 | Both sexes | Both sexes | Female only | True | False |
| MCH | 4 | RP11-545H22.1 | lincRNA | rs13122088 | 55372680 | 2.17e-08 | 3 | 0 | 0 | 0 | 0 | 3 | Both sexes | Both sexes | Both sexes | True | False |
| MCH | 9 | RP11-307I14.4 | pseudogene | rs10974772 | 4767677 | 2.22e-08 | 3 | 0 | 1 | 1 | 0 | 1 | Female only | Both sexes | Female only | True | False |
| MCH | 22 | TEX33 | protein_coding | rs5756445 | 37379539 | 2.41e-08 | 85 | 0 | 2 | 6 | 26 | 51 | Female only | Both sexes | Female only | True | False |
| MCH | 6 | UBQLN1P1 | pseudogene | rs2844745 | 30343703 | 2.50e-08 | 233 | 0 | 165 | 21 | 3 | 44 | Both sexes | Both sexes | Female only | True | True |
| MCH | 15 | IGDCC4 | protein_coding | rs7171289 | 65706569 | 2.52e-08 | 17 | 9 | 0 | 0 | 0 | 8 | Female only | Female only | Female only | True | False |
| MCH | 3 | TFRC | protein_coding | rs58758084 | 195812981 | 2.61e-08 | 13 | 0 | 0 | 0 | 0 | 13 | Male only | Both sexes | Male only | True | False |
| MCH | 16 | Z69890.1 | pseudogene | rs3760047 | 281299 | 2.62e-08 | 19 | 0 | 3 | 1 | 1 | 14 | Both sexes | Both sexes | Female only | True | False |
| MCH | 10 | TMEM72-AS1 | antisense | rs4948925 | 45392455 | 2.79e-08 | 57 | 0 | 15 | 0 | 14 | 28 | Female only | Both sexes | Female only | True | False |
| MCH | 6 | CCND3 | protein_coding | rs60610471 | 41959643 | 2.86e-08 | 17 | 0 | 0 | 6 | 3 | 8 | Female only | Both sexes | Female only | True | False |
| MCH | 7 | AP4M1 | protein_coding | rs62485013 | 99703505 | 2.95e-08 | 15 | 0 | 0 | 2 | 2 | 11 | Female only | Both sexes | Female only | True | False |
| MCH | 6 | CCND3 | protein_coding | rs11756584 | 41977937 | 3.00e-08 | 23 | 4 | 0 | 0 | 0 | 19 | Both sexes | Both sexes | Male only | True | False |
| MCH | 6 | GUSBP2 | pseudogene | rs56098842 | 26833759 | 3.38e-08 | 109 | 3 | 47 | 3 | 15 | 41 | Female only | Both sexes | Female only | True | False |
| MCH | 3 | RNU2-11P | snRNA | rs6765762 | 195652851 | 3.48e-08 | 7 | 0 | 0 | 2 | 0 | 5 | Male only | Both sexes | Male only | True | False |
| MCH | 15 | SLC24A1 | protein_coding | rs16948916 | 65943893 | 3.77e-08 | 377 | 0 | 2 | 188 | 0 | 187 | Female only | Both sexes | Female only | True | False |
| MCH | 6 | BTN1A1 | protein_coding | rs12197304 | 26496733 | 4.30e-08 | 289 | 141 | 4 | 31 | 58 | 55 | Female only | Female only | Female only | True | False |
| MCHC | 22 | TMPRSS6 | protein_coding | rs4820268 | 37469591 | 4.27e-65 | 9 | 0 | 0 | 0 | 0 | 9 | Both sexes | Both sexes | Both sexes | True | False |

| pheno | CHR | Gene_Symbol | biotype | lead_snp | lead_BP | lead_P | TOTAL | NSIG | S05 | S01 | S001 | S0001 | independent_loci_in_sex_P1e6 | independent_loci_in_sex_P1e2 | independent_loci_in_sex_P5e8 | in_gwas_catalog | in_HLA |
|---|---|---|---|---|---|---|---|---|---|---|---|---|---|---|---|---|---|
| MCHC | 16 | PIEZO1 | protein_coding | rs2968478 | 88858646 | 4.79e-51 | 23 | 0 | 0 | 0 | 0 | 23 | Both sexes | Both sexes | Both sexes | True | False |
| MCHC | 1 | ATP2B4 | protein_coding | rs10900588 | 203656814 | 1.41e-37 | 3 | 0 | 0 | 0 | 0 | 3 | Both sexes | Both sexes | Both sexes | True | False |
| MCHC | 6 | HFE | protein_coding | rs1800562 | 26093141 | 3.95e-36 | 15 | 0 | 0 | 0 | 0 | 15 | Both sexes | Both sexes | Both sexes | True | False |
| MCHC | 6 | SLC17A3 | protein_coding | rs1408272 | 25842951 | 7.56e-31 | 9 | 0 | 0 | 0 | 0 | 9 | Both sexes | Both sexes | Both sexes | True | False |
| MCHC | 22 | TMPRSS6 | protein_coding | rs2076086 | 37470536 | 1.70e-30 | 35 | 0 | 0 | 0 | 0 | 35 | Both sexes | Both sexes | Both sexes | True | False |
| MCHC | 6 | HFE | protein_coding | rs2032451 | 26092170 | 5.77e-27 | 39 | 0 | 0 | 0 | 0 | 39 | Both sexes | Both sexes | Both sexes | True | False |
| MCHC | 6 | HIST1H3C | protein_coding | 6:26046744 | 26046744 | 1.18e-24 | 245 | 0 | 0 | 14 | 0 | 231 | Both sexes | Both sexes | Both sexes | True | False |
| MCHC | 6 | LRRC16A | protein_coding | rs75580845 | 25578433 | 1.10e-23 | 45 | 0 | 0 | 0 | 0 | 45 | Both sexes | Both sexes | Both sexes | True | False |
| MCHC | 22 | TMPRSS6 | protein_coding | rs9607412 | 37484337 | 3.24e-23 | 57 | 0 | 0 | 0 | 0 | 57 | Both sexes | Both sexes | Both sexes | True | False |
| MCHC | 6 | MIR3143 | miRNA | rs71559014 | 27122444 | 3.79e-22 | 3 | 0 | 0 | 0 | 0 | 3 | Both sexes | Both sexes | Both sexes | True | False |
| MCHC | 6 | ZSCAN31 | protein_coding | rs35814746 | 28299088 | 6.85e-22 | 41 | 0 | 0 | 0 | 0 | 41 | Both sexes | Both sexes | Both sexes | True | False |
| MCHC | 8 | RP11-503E24.3 | lincRNA | rs1947296 | 42452999 | 7.33e-22 | 219 | 0 | 0 | 0 | 0 | 219 | Both sexes | Both sexes | Both sexes | True | False |
| MCHC | 6 | HIST1H2BO | protein_coding | 6:27861670 | 27861670 | 8.34e-22 | 9 | 0 | 0 | 0 | 0 | 9 | Both sexes | Both sexes | Both sexes | True | False |
| MCHC | 6 | U91328.22 | lincRNA | rs9356996 | 26008487 | 2.04e-21 | 63 | 0 | 0 | 1 | 9 | 53 | Both sexes | Both sexes | Both sexes | True | False |
| MCHC | 6 | LRRC16A | protein_coding | rs74662487 | 25453057 | 2.99e-20 | 17 | 0 | 0 | 0 | 0 | 17 | Both sexes | Both sexes | Both sexes | True | False |
| MCHC | 6 | SCGN | protein_coding | rs1321250 | 25642244 | 3.20e-20 | 25 | 0 | 0 | 0 | 0 | 25 | Both sexes | Both sexes | Both sexes | True | False |
| MCHC | 1 | TRIM58 | protein_coding | rs3811444 | 248039451 | 4.58e-19 | 3 | 0 | 0 | 0 | 0 | 3 | Both sexes | Both sexes | Both sexes | True | False |
| MCHC | 16 | PIEZO1 | protein_coding | rs860762 | 88856370 | 1.12e-17 | 23 | 0 | 4 | 0 | 0 | 19 | Both sexes | Both sexes | Both sexes | True | False |
| MCHC | 19 | KCNN4 | protein_coding | rs347519 | 44273516 | 2.18e-17 | 59 | 0 | 0 | 0 | 0 | 59 | Both sexes | Both sexes | Both sexes | True | False |
| MCHC | 6 | OR2AD1P | pseudogene | rs3129796 | 28988861 | 2.50e-16 | 7 | 0 | 0 | 0 | 1 | 6 | Both sexes | Both sexes | Both sexes | True | False |
| MCHC | 6 | SCGN | protein_coding | rs9467571 | 25700408 | 6.33e-16 | 217 | 0 | 8 | 4 | 89 | 116 | Both sexes | Both sexes | Female only | True | False |
| MCHC | 6 | LRRC16A | protein_coding | rs181058325 | 25314597 | 1.84e-15 | 29 | 0 | 0 | 0 | 0 | 29 | Both sexes | Both sexes | Both sexes | True | False |
| MCHC | 16 | PIEZO1 | protein_coding | rs67237947 | 88854779 | 2.18e-15 | 29 | 2 | 0 | 0 | 2 | 25 | Both sexes | Both sexes | Female only | True | False |
| MCHC | 6 | HLA-W | pseudogene | rs9260707 | 29929536 | 2.48e-15 | 169 | 0 | 0 | 0 | 0 | 169 | Both sexes | Both sexes | Both sexes | True | True |
| MCHC | 22 | TMPRSS6 | protein_coding | rs6000550 | 37463450 | 4.74e-15 | 15 | 0 | 0 | 0 | 2 | 13 | Both sexes | Both sexes | Both sexes | True | False |
| MCHC | 6 | GPX6 | protein_coding | rs10428759 | 28477682 | 1.31e-14 | 9 | 0 | 0 | 0 | 0 | 9 | Both sexes | Both sexes | Female only | True | False |

| pheno | CHR | Gene_Symbol | biotype | lead_snp | lead_BP | lead_P | TOTAL | NSIG | S05 | S01 | S001 | S0001 | independent_loci_in_sex_P1e6 | independent_loci_in_sex_P1e2 | independent_loci_in_sex_P5e8 | in_gwas_catalog | in_HLA |
|---|---|---|---|---|---|---|---|---|---|---|---|---|---|---|---|---|---|
| MCHC | 6 | HIST1H2APS2 | pseudogene | rs79782817 | 25882678 | 4.20e-14 | 63 | 0 | 0 | 7 | 2 | 54 | Both sexes | Both sexes | Both sexes | True | False |
| MCHC | 1 | CEP104 | protein_coding | rs12134919 | 3744641 | 4.54e-14 | 37 | 0 | 0 | 0 | 0 | 37 | Both sexes | Both sexes | Both sexes | True | False |
| MCHC | 22 | TMPRSS6 | protein_coding | rs2235325 | 37481637 | 4.88e-14 | 23 | 0 | 0 | 0 | 0 | 23 | Both sexes | Both sexes | Both sexes | True | False |
| MCHC | 6 | TRIM38 | protein_coding | rs199739 | 25960509 | 5.70e-14 | 47 | 22 | 0 | 7 | 6 | 12 | Female only | Both sexes | Female only | True | False |
| MCHC | 6 | LRRC16A | protein_coding | rs111722075 | 25374046 | 6.51e-14 | 235 | 0 | 0 | 0 | 28 | 207 | Both sexes | Both sexes | Both sexes | True | False |
| MCHC | 6 | TRIM26 | protein_coding | rs3094618 | 30181621 | 1.81e-13 | 63 | 0 | 0 | 0 | 0 | 63 | Both sexes | Both sexes | Both sexes | True | True |
| MCHC | 22 | TST | protein_coding | rs5756477 | 37407527 | 5.26e-13 | 57 | 0 | 0 | 12 | 17 | 28 | Female only | Both sexes | Female only | True | False |
| MCHC | 6 | HIST1H3G | protein_coding | rs7773004 | 26267755 | 2.06e-12 | 89 | 0 | 0 | 36 | 4 | 49 | Male only | Both sexes | Male only | True | False |
| MCHC | 6 | HBS1L | protein_coding | rs9376092 | 135427144 | 2.38e-12 | 37 | 0 | 0 | 1 | 0 | 36 | Both sexes | Both sexes | Both sexes | True | False |
| MCHC | 6 | MOG | protein_coding | rs3130253 | 29634012 | 2.80e-12 | 21 | 0 | 0 | 0 | 0 | 21 | Both sexes | Both sexes | Female only | True | True |
| MCHC | 16 | PIEZO1 | protein_coding | rs7186082 | 88783903 | 2.98e-12 | 11 | 0 | 2 | 3 | 1 | 5 | Female only | Both sexes | Female only | True | False |
| MCHC | 6 | ZSCAN23 | protein_coding | rs13190937 | 28411244 | 3.13e-12 | 71 | 33 | 24 | 0 | 0 | 14 | Female only | Female only | Female only | True | False |
| MCHC | 1 | ATP2B4 | protein_coding | rs4951073 | 203658486 | 4.44e-12 | 9 | 4 | 0 | 4 | 0 | 1 | Both sexes | Both sexes | Both sexes | True | False |
| MCHC | 6 | LRRC16A | protein_coding | rs2328879 | 25583103 | 1.08e-11 | 9 | 0 | 0 | 0 | 0 | 9 | Both sexes | Both sexes | Both sexes | True | False |
| MCHC | 6 | SLC17A3 | protein_coding | rs566530 | 25878361 | 1.62e-11 | 121 | 11 | 19 | 26 | 8 | 57 | Female only | Both sexes | Female only | True | False |
| MCHC | 16 | CTU2 | protein_coding | rs8059048 | 88781040 | 1.82e-11 | 13 | 0 | 0 | 0 | 0 | 13 | Both sexes | Both sexes | Female only | True | False |
| MCHC | 15 | DENND4A | protein_coding | rs7497961 | 66020908 | 1.86e-11 | 135 | 64 | 3 | 1 | 0 | 67 | Female only | Both sexes | Female only | True | False |
| MCHC | 6 | SLC17A1 | protein_coding | rs12191655 | 25803095 | 1.98e-11 | 113 | 0 | 7 | 18 | 29 | 59 | Female only | Both sexes | Female only | True | False |
| MCHC | 6 | HIST1H2BC | protein_coding | rs198839 | 26112620 | 1.99e-11 | 13 | 0 | 3 | 3 | 0 | 7 | Both sexes | Both sexes | Female only | True | False |
| MCHC | 6 | LRRC16A | protein_coding | rs9379764 | 25414023 | 2.03e-11 | 3 | 0 | 0 | 0 | 1 | 2 | Female only | Both sexes | Female only | True | False |
| MCHC | 6 | HCG22 | lincRNA | rs2517503 | 31032407 | 3.69e-11 | 33 | 0 | 0 | 1 | 1 | 31 | Female only | Both sexes | Female only | True | True |
| MCHC | 17 | SLC4A1 | protein_coding | rs13306780 | 42329004 | 3.80e-11 | 49 | 25 | 0 | 0 | 0 | 24 | Female only | Female only | Female only | True | False |
| MCHC | 22 | TMPRSS6 | protein_coding | rs11089824 | 37509186 | 3.94e-11 | 5 | 0 | 0 | 0 | 3 | 2 | Female only | Both sexes | Female only | True | False |
| MCHC | 6 | TRIM40 | protein_coding | rs2107202 | 30105743 | 1.37e-10 | 15 | 0 | 0 | 1 | 1 | 13 | Both sexes | Both sexes | Both sexes | True | True |
| MCHC | 6 | CCDC162P | pseudogene | rs11961369 | 109624771 | 1.52e-10 | 377 | 65 | 16 | 98 | 20 | 178 | Female only | Both sexes | Female only | True | False |
| MCHC | 6 | HCG4P7 | pseudogene | rs1611657 | 29842444 | 1.80e-10 | 155 | 0 | 2 | 74 | 0 | 79 | Female only | Both sexes | Female only | True | True |

| pheno | CHR | Gene_Symbol | biotype | lead_snp | lead_BP | lead_P | TOTAL | NSIG | S05 | S01 | S001 | S0001 | independent_loci_in_sex_P1e6 | independent_loci_in_sex_P1e2 | independent_loci_in_sex_P5e8 | in_gwas_catalog | in_HLA |
|---|---|---|---|---|---|---|---|---|---|---|---|---|---|---|---|---|---|
| MCHC | 16 | PIEZO1 | protein_coding | rs2911462 | 88835914 | 2.36e-10 | 13 | 0 | 0 | 0 | 4 | 9 | Both sexes | Both sexes | Female only | True | False |
| MCHC | 6 | HBS1L | protein_coding | rs9389270 | 135431807 | 7.70e-10 | 85 | 0 | 13 | 26 | 7 | 39 | Female only | Both sexes | Female only | True | False |
| MCHC | 6 | LRRC16A | protein_coding | rs72831267 | 25436955 | 7.72e-10 | 7 | 0 | 0 | 0 | 2 | 5 | Female only | Both sexes | Female only | True | False |
| MCHC | 6 | XXbac-BPG27H4.8 | lincRNA | rs3095345 | 30822413 | 7.83e-10 | 49 | 0 | 0 | 1 | 12 | 36 | Female only | Both sexes | Female only | True | True |
| MCHC | 6 | MICC | pseudogene | rs2524205 | 30359606 | 8.01e-10 | 233 | 0 | 93 | 0 | 2 | 138 | Both sexes | Both sexes | Male only | True | True |
| MCHC | 15 | WDR61 | protein_coding | rs2867932 | 78591037 | 1.47e-09 | 53 | 0 | 0 | 4 | 4 | 45 | Both sexes | Both sexes | Female only | True | False |
| MCHC | 15 | RAB11A | protein_coding | rs8027781 | 66097711 | 1.75e-09 | 71 | 0 | 0 | 1 | 35 | 35 | Female only | Both sexes | Female only | True | False |
| MCHC | 22 | MPST | protein_coding | rs5756488 | 37416803 | 1.77e-09 | 7 | 0 | 0 | 0 | 0 | 7 | Both sexes | Both sexes | Both sexes | True | False |

| pheno | CHR | Gene_Symbol | biotype | lead_snp | lead_BP | lead_P | TOTAL | NSIG | S05 | S01 | S001 | S0001 | independent_loci_in_sex_P1e6 | independent_loci_in_sex_P1e2 | independent_loci_in_sex_P5e8 | in_gwas_catalog | in_HLA |
|---|---|---|---|---|---|---|---|---|---|---|---|---|---|---|---|---|---|
| MCHC | 6 | MICD | pseudogene | rs9260740 | 29933236 | 1.78e-09 | 15 | 0 | 0 | 0 | 0 | 15 | Both sexes | Both sexes | Both sexes | True | True |
| MCHC | 6 | XXbac-BPG249D20.9 | antisense | rs9262081 | 30484966 | 2.84e-09 | 17 | 0 | 0 | 0 | 1 | 16 | Female only | Both sexes | Female only | True | True |
| MCHC | 6 | SCGN | protein_coding | rs4712949 | 25644203 | 2.99e-09 | 179 | 2 | 0 | 63 | 22 | 92 | Female only | Both sexes | Female only | True | False |
| MCHC | 6 | HLA-G | protein_coding | rs452850 | 29799481 | 3.20e-09 | 265 | 0 | 1 | 17 | 9 | 238 | Both sexes | Both sexes | Female only | True | True |
| MCHC | 6 | LRRC16A | protein_coding | rs112268831 | 25348760 | 3.25e-09 | 5 | 0 | 0 | 1 | 1 | 3 | Female only | Both sexes | Female only | True | False |
| MCHC | 6 | MICF | pseudogene | rs2523818 | 29833840 | 3.72e-09 | 405 | 47 | 133 | 42 | 3 | 180 | Female only | Both sexes | Female only | True | True |
| MCHC | 6 | C2 | protein_coding | rs3130683 | 31888367 | 4.34e-09 | 63 | 0 | 2 | 4 | 12 | 45 | Male only | Both sexes | Male only | True | True |
| MCHC | 6 | SLC17A3 | protein_coding | rs1165201 | 25874823 | 6.19e-09 | 55 | 9 | 19 | 2 | 1 | 24 | Female only | Both sexes | Female only | True | False |
| MCHC | 15 | DPP8 | protein_coding | rs2437053 | 65781457 | 6.70e-09 | 29 | 0 | 0 | 14 | 1 | 14 | Female only | Both sexes | Female only | True | False |
| MCHC | 6 | AIF1 | protein_coding | rs2857600 | 31582287 | 7.12e-09 | 71 | 0 | 0 | 2 | 4 | 65 | Both sexes | Both sexes | Both sexes | True | True |
| MCHC | 6 | LRRC16A | protein_coding | rs9358860 | 25572245 | 9.67e-09 | 53 | 46 | 0 | 0 | 0 | 7 | Female only | Both sexes | Female only | True | False |
| MCHC | 8 | SLC20A2 | protein_coding | rs10107967 | 42342154 | 9.88e-09 | 7 | 0 | 0 | 0 | 0 | 7 | Both sexes | Both sexes | Both sexes | True | False |
| MCHC | 15 | DNAJA4 | protein_coding | rs11629630 | 78565414 | 1.08e-08 | 53 | 15 | 1 | 14 | 4 | 19 | Female only | Both sexes | Female only | True | False |
| MCHC | 6 | GPX6 | protein_coding | rs2213806 | 28457818 | 1.09e-08 | 75 | 38 | 15 | 20 | 1 | 1 | Female only | Both sexes | Female only | True | False |
| MCHC | 2 | IL1F10 | protein_coding | rs12711751 | 113837765 | 1.15e-08 | 35 | 12 | 2 | 4 | 0 | 17 | Female only | Both sexes | Female only | True | False |
| MCHC | 17 | FAM222B | protein_coding | rs9892942 | 27178761 | 1.33e-08 | 159 | 0 | 0 | 0 | 6 | 153 | Both sexes | Both sexes | Male only | True | False |

| pheno | CHR | Gene_Symbol | biotype | lead_snp | lead_BP | lead_P | TOTAL | NSIG | S05 | S01 | S001 | S0001 | independent_loci_in_sex_P1e6 | independent_loci_in_sex_P1e2 | independent_loci_in_sex_P5e8 | in_gwas_catalog | in_HLA |
|---|---|---|---|---|---|---|---|---|---|---|---|---|---|---|---|---|---|
| MCHC | 6 | TRIM15 | protein_coding | rs2844787 | 30140913 | 1.48e-08 | 3 | 0 | 0 | 0 | 1 | 2 | Both sexes | Both sexes | Female only | True | True |
| MCHC | 6 | GPX6 | protein_coding | rs2531820 | 28456719 | 1.57e-08 | 33 | 2 | 8 | 6 | 2 | 15 | Female only | Both sexes | Female only | True | False |
| MCHC | 6 | XXbac-BPG154L12.4 | antisense | rs424232 | 32208324 | 1.84e-08 | 19 | 5 | 3 | 3 | 0 | 8 | Male only | Both sexes | Male only | True | True |
| MCHC | 15 | WDR61 | protein_coding | rs7170960 | 78583818 | 1.88e-08 | 23 | 0 | 2 | 4 | 0 | 17 | Female only | Both sexes | Female only | True | False |
| MCHC | 1 | OR10X1 | protein_coding | rs857693 | 158556301 | 1.93e-08 | 109 | 0 | 1 | 31 | 23 | 54 | Female only | Both sexes | Female only | True | False |
| MCHC | 6 | HFE | protein_coding | rs2071303 | 26091336 | 2.25e-08 | 51 | 7 | 18 | 8 | 17 | 1 | Both sexes | Both sexes | Female only | True | False |
| MCHC | 19 | GCDH | protein_coding | rs62108438 | 13000247 | 2.82e-08 | 75 | 26 | 12 | 0 | 0 | 37 | Female only | Female only | Female only | True | False |
| MCHC | 14 | FNTB | protein_coding | rs11627531 | 65487796 | 3.26e-08 | 25 | 0 | 0 | 0 | 0 | 25 | Both sexes | Both sexes | Male only | True | False |
| MCHC | 19 | ZNF283 | protein_coding | rs60085895 | 44347424 | 3.29e-08 | 203 | 80 | 22 | 0 | 0 | 101 | Female only | Female only | Female only | True | False |
| MCHC | 16 | HBZ | protein_coding | rs113613943 | 201754 | 3.31e-08 | 3 | 0 | 0 | 0 | 2 | 1 | Male only | Both sexes | Male only | True | False |
| MCHC | 19 | KCNN4 | protein_coding | rs3760984 | 44286762 | 3.98e-08 | 91 | 8 | 2 | 22 | 14 | 45 | Female only | Both sexes | Female only | True | False |
| MCHC | 6 | TRIM26 | protein_coding | rs10947058 | 30176122 | 4.06e-08 | 63 | 0 | 11 | 7 | 9 | 36 | Female only | Both sexes | Female only | True | True |
| MCHC | 6 | XXbac-BPG248L24.12 | pseudogene | 6:31328756 | 31328756 | 4.23e-08 | 9 | 0 | 0 | 1 | 0 | 8 | Both sexes | Both sexes | Male only | True | True |
| MCHC | 3 | LINC00885 | lincRNA | rs9870228 | 195901514 | 4.25e-08 | 57 | 0 | 0 | 13 | 16 | 28 | Male only | Both sexes | Male only | True | False |
| MCHC | 22 | MPST | protein_coding | rs6000536 | 37421469 | 4.51e-08 | 27 | 10 | 0 | 11 | 5 | 1 | Female only | Both sexes | Female only | True | False |
| MEAN_QRS | 1 | CASQ2 | protein_coding | rs11578967 | 116301670 | 9.77e-11 | 155 | 8 | 11 | 50 | 9 | 77 | Female only | Both sexes | Female only | True | False |
| MEAN_QRS | 12 | RP11-25E2.1 | lincRNA | rs1896356 | 115367286 | 2.72e-10 | 71 | 0 | 0 | 1 | 0 | 70 | Female only | Both sexes | Female only | True | False |
| MEAN_QRS | 1 | CASQ2 | protein_coding | rs4433393 | 116332232 | 2.27e-09 | 33 | 1 | 14 | 2 | 1 | 15 | Female only | Both sexes | Female only | True | False |
| MEAN_QRS | 10 | RP11-90B22.1 | lincRNA | rs10763928 | 33632954 | 6.59e-09 | 89 | 0 | 1 | 28 | 16 | 44 | Male only | Both sexes | Male only | True | False |
| MEAN_QRS | 10 | RPL7P37 | pseudogene | rs2804817 | 36061337 | 7.06e-09 | 15 | 8 | 0 | 3 | 0 | 4 | Male only | Male only | Male only | True | False |
| MEAN_QRS | 1 | CASQ2 | protein_coding | rs12039739 | 116333111 | 1.74e-08 | 15 | 7 | 1 | 0 | 0 | 7 | Female only | Female only | Female only | True | False |
| MEAN_QRS | 10 | VTI1A | protein_coding | rs11196083 | 114500004 | 2.12e-08 | 97 | 1 | 0 | 6 | 23 | 67 | Male only | Both sexes | Male only | True | False |

| pheno | CHR | Gene_Symbol | biotype | lead_snp | lead_BP | lead_P | TOTAL | NSIG | S05 | S01 | S001 | S0001 | independent_loci_in_sex_P1e6 | independent_loci_in_sex_P1e2 | independent_loci_in_sex_P5e8 | in_gwas_catalog | in_HLA |
|---|---|---|---|---|---|---|---|---|---|---|---|---|---|---|---|---|---|
| MEAN_QRS | 4 | LIMCH1 | protein_coding | rs1377345 | 41322070 | 2.30e-08 | 131 | 0 | 9 | 56 | 14 | 52 | Female only | Both sexes | Female only | True | False |
| MONOAB | 2 | ITGA4 | protein_coding | rs1375493 | 182323766 | 6.77e-33 | 15 | 0 | 0 | 0 | 0 | 15 | Both sexes | Both sexes | Both sexes | True | False |
| MONOAB | 8 | CCDC26 | lincRNA | rs2217715 | 130615008 | 1.88e-30 | 63 | 0 | 0 | 0 | 0 | 63 | Both sexes | Both sexes | Both sexes | True | False |
| MONOAB | 9 | RP11-202G18.1 | lincRNA | rs7023923 | 113925534 | 7.13e-22 | 27 | 0 | 0 | 0 | 0 | 27 | Both sexes | Both sexes | Both sexes | True | False |
| MONOAB | 9 | RP11-202G18.1 | lincRNA | rs12348789 | 113945005 | 4.04e-20 | 51 | 0 | 0 | 0 | 0 | 51 | Both sexes | Both sexes | Both sexes | True | False |
| MONOAB | 2 | ITGA4 | protein_coding | rs1449260 | 182333447 | 1.24e-18 | 3 | 0 | 0 | 0 | 0 | 3 | Both sexes | Both sexes | Both sexes | True | False |
| MONOAB | 3 | RPN1 | protein_coding | rs34198449 | 128381270 | 4.10e-17 | 173 | 0 | 0 | 0 | 0 | 173 | Both sexes | Both sexes | Both sexes | True | False |
| MONOAB | 8 | CCDC26 | lincRNA | rs7000917 | 130590087 | 2.24e-16 | 33 | 0 | 0 | 0 | 0 | 33 | Both sexes | Both sexes | Both sexes | True | False |
| MONOAB | 3 | C3orf27 | protein_coding | rs66910450 | 128303893 | 2.26e-15 | 63 | 0 | 0 | 0 | 0 | 63 | Both sexes | Both sexes | Both sexes | True | False |
| MONOAB | 16 | RP11-542M13.3 | lincRNA | rs11642873 | 85991705 | 3.94e-15 | 7 | 0 | 0 | 0 | 0 | 7 | Female only | Both sexes | Female only | True | False |
| MONOAB | 16 | RP11-542M13.3 | lincRNA | rs76121846 | 85966548 | 4.98e-14 | 37 | 0 | 0 | 0 | 0 | 37 | Both sexes | Both sexes | Both sexes | True | False |
| MONOAB | 3 | RPN1 | protein_coding | rs2713573 | 128317273 | 2.71e-13 | 9 | 0 | 0 | 0 | 0 | 9 | Both sexes | Both sexes | Both sexes | True | False |
| MONOAB | 3 | ACKR2 | protein_coding | rs4683341 | 42877394 | 6.11e-13 | 81 | 0 | 0 | 3 | 24 | 54 | Female only | Both sexes | Female only | True | False |
| MONOAB | 3 | C3orf27 | protein_coding | rs2734047 | 128292844 | 1.99e-12 | 9 | 0 | 0 | 0 | 0 | 9 | Both sexes | Both sexes | Both sexes | True | False |
| MONOAB | 3 | C3orf27 | protein_coding | rs62273237 | 128310107 | 2.61e-12 | 3 | 0 | 0 | 0 | 0 | 3 | Both sexes | Both sexes | Both sexes | True | False |
| MONOAB | 3 | HIGD1A | protein_coding | rs6764909 | 42835889 | 2.63e-12 | 5 | 0 | 0 | 3 | 0 | 2 | Female only | Both sexes | Female only | True | False |
| MONOAB | 2 | ITGA4 | protein_coding | rs72894360 | 182318491 | 7.39e-12 | 13 | 0 | 0 | 0 | 0 | 13 | Female only | Both sexes | Female only | True | False |
| MONOAB | 3 | CCDC13 | protein_coding | rs35584330 | 42811240 | 1.08e-11 | 35 | 0 | 2 | 16 | 0 | 17 | Female only | Both sexes | Female only | True | False |
| MONOAB | 16 | IRF8 | protein_coding | rs8051462 | 85947327 | 2.02e-11 | 35 | 3 | 0 | 7 | 0 | 25 | Both sexes | Both sexes | Both sexes | True | False |
| MONOAB | 3 | SNORA24 | snoRNA | rs2001950 | 128419756 | 3.81e-11 | 17 | 0 | 0 | 0 | 0 | 17 | Both sexes | Both sexes | Female only | True | False |
| MONOAB | 16 | RP11-542M13.3 | lincRNA | rs11640143 | 85989464 | 1.44e-10 | 7 | 0 | 0 | 0 | 2 | 5 | Female only | Both sexes | Female only | True | False |
| MONOAB | 8 | CCDC26 | lincRNA | rs7840913 | 130622669 | 2.14e-10 | 49 | 0 | 0 | 0 | 2 | 47 | Both sexes | Both sexes | Female only | True | False |
| pheno | CHR | Gene_Symbol | biotype | lead_snp | lead_BP | lead_P | TOTAL | NSIG | S05 | S01 | S001 | S0001 | independent_loci_in_sex_P1e6 | independent_loci_in_sex_P1e2 | independent_loci_in_sex_P5e8 | in_gwas_catalog | in_HLA |
| MONOAB | 16 | RP11-542M13.2 | lincRNA | rs1915035 | 86015147 | 2.18e-10 | 11 | 0 | 5 | 0 | 1 | 5 | Female only | Both sexes | Female only | True | False |
| MONOAB | 11 | UBASH3B | protein_coding | rs11600178 | 122519352 | 4.29e-10 | 73 | 0 | 0 | 23 | 14 | 36 | Female only | Both sexes | Female only | True | False |

| pheno | CHR | Gene_Symbol | biotype | lead_snp | lead_BP | lead_P | TOTAL | NSIG | S05 | S01 | S001 | S0001 | independent_loci_in_sex_P1e6 | independent_loci_in_sex_P1e2 | independent_loci_in_sex_P5e8 | in_gwas_catalog | in_HLA |
|---|---|---|---|---|---|---|---|---|---|---|---|---|---|---|---|---|---|
| MONOAB | 3 | KRBOX1 | protein_coding | rs834580 | 42969592 | 4.62e-10 | 13 | 5 | 2 | 0 | 0 | 6 | Female only | Female only | Female only | True | False |
| MONOAB | 9 | RP11-202G18.1 | lincRNA | rs72757367 | 113902199 | 6.38e-10 | 157 | 0 | 0 | 5 | 14 | 138 | Female only | Both sexes | Female only | True | False |
| MONOAB | 9 | RP11-202G18.1 | lincRNA | rs1610009 | 113999400 | 1.08e-09 | 5 | 0 | 0 | 0 | 3 | 2 | Female only | Both sexes | Female only | True | False |
| MONOAB | 2 | ITGA4 | protein_coding | rs711801 | 182334873 | 1.76e-09 | 13 | 3 | 3 | 1 | 0 | 6 | Female only | Both sexes | Female only | True | False |
| MONOAB | 6 | RPSAP2 | pseudogene | rs7775835 | 28678357 | 2.04e-09 | 251 | 32 | 90 | 0 | 4 | 125 | Male only | Both sexes | Male only | True | False |
| MONOAB | 16 | RP11-542M13.3 | lincRNA | rs7202472 | 85977502 | 2.21e-09 | 21 | 0 | 9 | 1 | 9 | 2 | Female only | Both sexes | Female only | True | False |
| MONOAB | 8 | CCDC26 | lincRNA | rs4377928 | 130649867 | 2.89e-09 | 65 | 5 | 5 | 7 | 28 | 20 | Female only | Both sexes | Female only | True | False |
| MONOAB | 6 | ATF6B | protein_coding | rs204893 | 32094593 | 3.57e-09 | 17 | 5 | 4 | 0 | 0 | 8 | Male only | Male only | Male only | True | True |
| MONOAB | 2 | ITGA4 | protein_coding | rs13000914 | 182307666 | 3.69e-09 | 11 | 0 | 0 | 0 | 0 | 11 | Both sexes | Both sexes | Female only | True | False |
| MONOAB | 3 | CCDC13 | protein_coding | rs11915803 | 42800291 | 3.75e-09 | 39 | 16 | 4 | 0 | 0 | 19 | Female only | Female only | Female only | True | False |
| MONOAB | 16 | IRF8 | protein_coding | rs8064111 | 85947475 | 3.89e-09 | 35 | 2 | 2 | 0 | 1 | 30 | Both sexes | Both sexes | Male only | True | False |
| MONOAB | 6 | ZNF192P2 | pseudogene | rs735765 | 28170297 | 3.90e-09 | 349 | 175 | 0 | 0 | 0 | 174 | Male only | Male only | Male only | True | False |
| MONOAB | 2 | ITGA4 | protein_coding | rs1449264 | 182336474 | 4.40e-09 | 3 | 1 | 0 | 0 | 1 | 1 | Female only | Both sexes | Female only | True | False |
| MONOAB | 8 | CCDC26 | lincRNA | rs10105319 | 130644825 | 6.33e-09 | 5 | 0 | 0 | 3 | 0 | 2 | Female only | Both sexes | Female only | True | False |
| MONOAB | 6 | OR5V1 | protein_coding | rs9257805 | 29346329 | 6.55e-09 | 109 | 14 | 38 | 3 | 1 | 53 | Male only | Both sexes | Male only | True | False |
| MONOAB | 14 | STXBP6 | protein_coding | rs2004925 | 25445362 | 7.67e-09 | 29 | 0 | 0 | 9 | 8 | 12 | Female only | Both sexes | Female only | True | False |
| MONOAB | 6 | HLA-B | protein_coding | rs2442722 | 31320241 | 7.83e-09 | 227 | 0 | 1 | 10 | 49 | 167 | Male only | Both sexes | Male only | True | True |
| MONOAB | 3 | TMED10P2 | pseudogene | rs59855017 | 128268659 | 8.85e-09 | 3 | 0 | 0 | 0 | 0 | 3 | Both sexes | Both sexes | Male only | True | False |
| MONOAB | 1 | RPAP2 | protein_coding | rs74767605 | 92770376 | 9.71e-09 | 7 | 0 | 1 | 1 | 1 | 4 | Female only | Both sexes | Female only | True | False |
| MONOAB | 6 | RP1-97D16.1 | pseudogene | rs17693963 | 27710165 | 9.71e-09 | 169 | 44 | 41 | 0 | 0 | 84 | Male only | Male only | Male only | True | False |
| MONOAB | 20 | RP11-410N8.4 | protein_coding | rs12480732 | 31191015 | 1.03e-08 | 7 | 0 | 0 | 0 | 4 | 3 | Female only | Both sexes | Female only | True | False |
| MONOAB | 9 | RP11-202G18.1 | lincRNA | rs7850162 | 113953501 | 1.03e-08 | 97 | 0 | 3 | 15 | 24 | 55 | Female only | Both sexes | Female only | True | False |
| MONOAB | 6 | XXbac-BPG248L24.10 | pseudogene | rs9265540 | 31298164 | 1.23e-08 | 75 | 24 | 15 | 13 | 8 | 15 | Male only | Both sexes | Male only | True | True |
| MONOAB | 9 | RP11-202G18.1 | lincRNA | rs2026801 | 113905451 | 1.77e-08 | 3 | 2 | 0 | 0 | 0 | 1 | Female only | Both sexes | Female only | True | False |
| MONOAB | 6 | HLA-F | protein_coding | rs885945 | 29696852 | 2.66e-08 | 391 | 0 | 11 | 88 | 97 | 195 | Male only | Both sexes | Male only | True | True |

| pheno | CHR | Gene_Symbol | biotype | lead_snp | lead_BP | lead_P | TOTAL | NSIG | S05 | S01 | S001 | S0001 | independent_loci_in_sex_P1e6 | independent_loci_in_sex_P1e2 | independent_loci_in_sex_P5e8 | in_gwas_catalog | in_HLA |
|---|---|---|---|---|---|---|---|---|---|---|---|---|---|---|---|---|---|
| MONOAB | 16 | IRF8 | protein_coding | rs391023 | 85927814 | 3.31e-08 | 9 | 0 | 0 | 0 | 2 | 7 | Both sexes | Both sexes | Female only | True | False |
| MONOAB | 1 | GFI1 | protein_coding | rs10874656 | 92947024 | 4.25e-08 | 197 | 0 | 99 | 0 | 0 | 98 | Female only | Female only | Female only | True | False |
| MONOAB | 6 | UBQLN1P1 | pseudogene | rs3129809 | 30335621 | 4.65e-08 | 285 | 57 | 79 | 8 | 0 | 141 | Male only | Both sexes | Male only | True | True |
| MONORE | 2 | ITGA4 | protein_coding | rs1375493 | 182323766 | 7.07e-37 | 15 | 0 | 0 | 0 | 0 | 15 | Both sexes | Both sexes | Both sexes | True | False |
| MONORE | 9 | RP11-202G18.1 | lincRNA | rs10980797 | 113912553 | 5.21e-34 | 29 | 0 | 0 | 0 | 0 | 29 | Both sexes | Both sexes | Both sexes | True | False |
| MONORE | 9 | RP11-202G18.1 | lincRNA | rs12346772 | 113920599 | 1.36e-33 | 51 | 0 | 0 | 0 | 0 | 51 | Both sexes | Both sexes | Both sexes | True | False |
| MONORE | 16 | RP11-542M13.3 | lincRNA | rs76121846 | 85966548 | 1.68e-29 | 37 | 0 | 0 | 0 | 0 | 37 | Both sexes | Both sexes | Both sexes | True | False |
| MONORE | 8 | CCDC26 | lincRNA | rs35389394 | 130621254 | 8.43e-29 | 63 | 0 | 0 | 0 | 0 | 63 | Both sexes | Both sexes | Both sexes | True | False |
| MONORE | 3 | C3orf27 | protein_coding | rs9849126 | 128312069 | 7.06e-27 | 61 | 0 | 0 | 0 | 0 | 61 | Both sexes | Both sexes | Both sexes | True | False |
| MONORE | 3 | C3orf27 | protein_coding | rs2010054 | 128305302 | 6.42e-26 | 119 | 0 | 0 | 0 | 0 | 119 | Both sexes | Both sexes | Both sexes | True | False |
| MONORE | 16 | IRF8 | protein_coding | rs8051462 | 85947327 | 4.35e-24 | 35 | 0 | 0 | 1 | 5 | 29 | Both sexes | Both sexes | Both sexes | True | False |
| MONORE | 9 | RP11-202G18.1 | lincRNA | rs1610009 | 113999400 | 8.06e-24 | 5 | 0 | 0 | 0 | 0 | 5 | Both sexes | Both sexes | Both sexes | True | False |
| MONORE | 16 | IRF8 | protein_coding | rs8064111 | 85947475 | 8.24e-24 | 35 | 0 | 0 | 2 | 2 | 31 | Both sexes | Both sexes | Both sexes | True | False |
| MONORE | 2 | ITGA4 | protein_coding | rs1449260 | 182333447 | 5.56e-21 | 3 | 0 | 0 | 0 | 0 | 3 | Both sexes | Both sexes | Both sexes | True | False |
| MONORE | 3 | RPN1 | protein_coding | rs34890930 | 128329223 | 4.38e-20 | 67 | 0 | 0 | 0 | 0 | 67 | Both sexes | Both sexes | Both sexes | True | False |
| MONORE | 3 | RPN1 | protein_coding | rs2712431 | 128316890 | 2.48e-19 | 5 | 0 | 0 | 0 | 0 | 5 | Both sexes | Both sexes | Both sexes | True | False |
| MONORE | 3 | C3orf27 | protein_coding | rs2734047 | 128292844 | 2.60e-19 | 3 | 0 | 0 | 0 | 0 | 3 | Both sexes | Both sexes | Both sexes | True | False |
| MONORE | 16 | RP11-542M13.3 | lincRNA | rs11642873 | 85991705 | 5.66e-19 | 7 | 0 | 0 | 0 | 0 | 7 | Both sexes | Both sexes | Both sexes | True | False |
| MONORE | 16 | IRF8 | protein_coding | rs391023 | 85927814 | 1.13e-17 | 9 | 0 | 0 | 0 | 0 | 9 | Both sexes | Both sexes | Both sexes | True | False |
| MONORE | 9 | RNY4P18 | misc_RNA | rs56111194 | 113863138 | 3.62e-17 | 71 | 0 | 0 | 3 | 0 | 68 | Both sexes | Both sexes | Both sexes | True | False |
| MONORE | 3 | SNORA24 | snoRNA | rs11719239 | 128419054 | 1.92e-16 | 17 | 0 | 0 | 0 | 0 | 17 | Both sexes | Both sexes | Both sexes | True | False |
| MONORE | 9 | RP11-202G18.1 | lincRNA | rs10980817 | 113950581 | 2.21e-16 | 95 | 0 | 1 | 2 | 8 | 84 | Both sexes | Both sexes | Both sexes | True | False |
| MONORE | 9 | RP11-202G18.1 | lincRNA | rs2026801 | 113905451 | 9.74e-16 | 3 | 2 | 0 | 0 | 0 | 1 | Both sexes | Both sexes | Both sexes | True | False |
| MONORE | 16 | RP11-542M13.3 | lincRNA | rs11640143 | 85989464 | 1.02e-15 | 7 | 0 | 0 | 0 | 0 | 7 | Both sexes | Both sexes | Both sexes | True | False |
| MONORE | 16 | RP11-542M13.2 | lincRNA | rs1915035 | 86015147 | 4.06e-15 | 11 | 0 | 0 | 0 | 0 | 11 | Both sexes | Both sexes | Both sexes | True | False |
| MONORE | 16 | RP11-542M13.3 | lincRNA | rs7202472 | 85977502 | 3.69e-14 | 21 | 0 | 0 | 0 | 0 | 21 | Both sexes | Both sexes | Both sexes | True | False |

| pheno | CHR | Gene_Symbol | biotype | lead_snp | lead_BP | lead_P | TOTAL | NSIG | S05 | S01 | S001 | S0001 | independent_loci_in_sex_P1e6 | independent_loci_in_sex_P1e2 | independent_loci_in_sex_P5e8 | in_gwas_catalog | in_HLA |
|---|---|---|---|---|---|---|---|---|---|---|---|---|---|---|---|---|---|
| MONORE | 2 | ITGA4 | protein_coding | rs72894360 | 182318491 | 2.07e-13 | 13 | 0 | 0 | 0 | 0 | 13 | Both sexes | Both sexes | Both sexes | True | False |
| MONORE | 3 | C3orf27 | protein_coding | rs2712423 | 128290938 | 1.67e-12 | 3 | 0 | 0 | 0 | 0 | 3 | Both sexes | Both sexes | Both sexes | True | False |
| MONORE | 16 | IRF8 | protein_coding | rs11864884 | 85901039 | 2.47e-12 | 15 | 0 | 0 | 0 | 0 | 15 | Both sexes | Both sexes | Both sexes | True | False |
| MONORE | 7 | AC020743.4 | lincRNA | rs4385425 | 50307334 | 3.08e-12 | 13 | 2 | 0 | 5 | 0 | 6 | Female only | Both sexes | Female only | True | False |
| MONORE | 2 | ITGA4 | protein_coding | rs115610786 | 182334805 | 3.11e-12 | 11 | 0 | 0 | 0 | 2 | 9 | Female only | Both sexes | Female only | True | False |
| MONORE | 2 | ITGA4 | protein_coding | rs1449264 | 182336474 | 4.40e-12 | 3 | 0 | 0 | 0 | 0 | 3 | Female only | Both sexes | Female only | True | False |
| MONORE | 17 | MED24 | protein_coding | rs7502514 | 38188844 | 4.65e-12 | 139 | 0 | 0 | 0 | 3 | 136 | Both sexes | Both sexes | Both sexes | True | False |
| pheno | CHR | Gene_Symbol | biotype | lead_snp | lead_BP | lead_P | TOTAL | NSIG | S05 | S01 | S001 | S0001 | independent_loci_in_sex_P1e6 | independent_loci_in_sex_P1e2 | independent_loci_in_sex_P5e8 | in_gwas_catalog | in_HLA |
| MONORE | 9 | RNY4P18 | misc_RNA | rs10817139 | 113838746 | 6.57e-12 | 15 | 0 | 0 | 8 | 0 | 7 | Female only | Both sexes | Female only | True | False |
| MONORE | 20 | RP11-290F20.3 | lincRNA | rs6020417 | 48899352 | 7.81e-12 | 33 | 0 | 0 | 1 | 2 | 30 | Female only | Both sexes | Female only | True | False |
| MONORE | 8 | CCDC26 | lincRNA | rs7005206 | 130620813 | 9.51e-12 | 33 | 0 | 0 | 0 | 0 | 33 | Both sexes | Both sexes | Both sexes | True | False |
| MONORE | 13 | TPTE2P5 | pseudogene | rs9532600 | 41434808 | 1.36e-11 | 77 | 2 | 4 | 33 | 0 | 38 | Female only | Both sexes | Female only | True | False |
| MONORE | 8 | CCDC26 | lincRNA | rs12676406 | 130608634 | 2.13e-11 | 85 | 0 | 1 | 7 | 9 | 68 | Both sexes | Both sexes | Female only | True | False |
| MONORE | 16 | IRF8 | protein_coding | rs55764989 | 85914987 | 2.71e-11 | 13 | 0 | 0 | 0 | 0 | 13 | Both sexes | Both sexes | Both sexes | True | False |
| MONORE | 16 | IRF8 | protein_coding | rs62052470 | 85904222 | 2.95e-11 | 21 | 0 | 0 | 0 | 1 | 20 | Both sexes | Both sexes | Both sexes | True | False |
| MONORE | 9 | RP11-202G18.1 | lincRNA | rs72757365 | 113901961 | 3.12e-11 | 157 | 0 | 0 | 16 | 14 | 127 | Female only | Both sexes | Female only | True | False |
| MONORE | 3 | ACKR2 | protein_coding | rs7623543 | 42921627 | 5.77e-11 | 77 | 0 | 0 | 9 | 10 | 58 | Both sexes | Both sexes | Female only | True | False |
| MONORE | 9 | RP11-202G18.1 | lincRNA | rs113646878 | 114042695 | 8.98e-11 | 45 | 16 | 7 | 0 | 0 | 22 | Female only | Female only | Female only | True | False |
| MONORE | 9 | RP11-202G18.1 | lincRNA | rs12337468 | 114046917 | 1.27e-10 | 5 | 0 | 0 | 3 | 1 | 1 | Female only | Both sexes | Female only | True | False |
| MONORE | 16 | RP11-542M13.2 | lincRNA | rs17445836 | 86017663 | 1.94e-10 | 3 | 0 | 0 | 0 | 0 | 3 | Both sexes | Both sexes | Both sexes | True | False |
| MONORE | 1 | RPAP2 | protein_coding | rs6603963 | 92821374 | 2.76e-10 | 207 | 0 | 0 | 104 | 0 | 103 | Female only | Both sexes | Female only | True | False |
| MONORE | 13 | FOXO1 | protein_coding | rs61963264 | 41154792 | 3.23e-10 | 141 | 65 | 1 | 5 | 1 | 69 | Female only | Both sexes | Female only | True | False |
| MONORE | 1 | RPAP2 | protein_coding | rs74767605 | 92770376 | 3.43e-10 | 7 | 0 | 1 | 1 | 1 | 4 | Female only | Both sexes | Female only | True | False |
| MONORE | 3 | ACKR2 | protein_coding | rs11129979 | 42890731 | 3.65e-10 | 17 | 0 | 0 | 7 | 0 | 10 | Both sexes | Both sexes | Female only | True | False |
| MONORE | 20 | RP11-410N8.4 | protein_coding | rs11167172 | 31193045 | 6.73e-10 | 7 | 0 | 0 | 0 | 4 | 3 | Female only | Both sexes | Female only | True | False |
| MONORE | 7 | AC020743.4 | lincRNA | rs76641566 | 50284557 | 8.62e-10 | 5 | 0 | 2 | 1 | 0 | 2 | Female only | Both sexes | Female only | True | False |

| pheno | CHR | Gene_Symbol | biotype | lead_snp | lead_BP | lead_P | TOTAL | NSIG | S05 | S01 | S001 | S0001 | independent_loci_in_sex_P1e6 | independent_loci_in_sex_P1e2 | independent_loci_in_sex_P5e8 | in_gwas_catalog | in_HLA |
|---|---|---|---|---|---|---|---|---|---|---|---|---|---|---|---|---|---|
| MONORE | 17 | RP11-387H17.6 | lincRNA | rs8068260 | 38163650 | 1.24e-09 | 85 | 0 | 0 | 0 | 0 | 85 | Both sexes | Both sexes | Male only | True | False |
| MONORE | 17 | PSMD3 | protein_coding | rs8075668 | 38137623 | 1.99e-09 | 9 | 0 | 0 | 0 | 0 | 9 | Both sexes | Both sexes | Both sexes | True | False |
| MONORE | 9 | MIR4289 | miRNA | rs10780207 | 91470733 | 2.61e-09 | 15 | 0 | 0 | 0 | 7 | 8 | Female only | Both sexes | Female only | True | False |
| MONORE | 13 | RN7SKP2 | misc_RNA | rs1892548 | 41002641 | 3.02e-09 | 17 | 0 | 0 | 0 | 0 | 17 | Both sexes | Both sexes | Male only | True | False |
| MONORE | 8 | CCDC26 | lincRNA | rs4377928 | 130649867 | 6.24e-09 | 65 | 0 | 6 | 4 | 2 | 53 | Both sexes | Both sexes | Both sexes | True | False |
| MONORE | 2 | ITGA4 | protein_coding | rs711801 | 182334873 | 7.46e-09 | 13 | 0 | 0 | 2 | 3 | 8 | Both sexes | Both sexes | Both sexes | True | False |
| MONORE | 1 | EPHX4 | protein_coding | rs143792309 | 92505309 | 9.97e-09 | 5 | 0 | 0 | 2 | 1 | 2 | Female only | Both sexes | Female only | True | False |
| MONORE | 9 | RP11-202G18.1 | lincRNA | rs34273533 | 113930024 | 1.18e-08 | 7 | 4 | 0 | 0 | 0 | 3 | Female only | Female only | Female only | True | False |
| MONORE | 9 | RP11-202G18.1 | lincRNA | rs7858485 | 113948326 | 1.30e-08 | 73 | 0 | 2 | 35 | 0 | 36 | Female only | Both sexes | Female only | True | False |
| MONORE | 2 | MYO3B | protein_coding | rs7598148 | 171279752 | 1.92e-08 | 19 | 10 | 0 | 0 | 0 | 9 | Male only | Male only | Male only | True | False |
| MONORE | 13 | TPTE2P5 | pseudogene | rs4943818 | 41400473 | 2.07e-08 | 63 | 2 | 30 | 0 | 0 | 31 | Female only | Female only | Female only | True | False |
| MONORE | 9 | RNY4P18 | misc_RNA | rs62570762 | 113846061 | 2.16e-08 | 7 | 0 | 0 | 4 | 0 | 3 | Female only | Both sexes | Female only | True | False |
| MONORE | 19 | CTD-2540B15.12 | lincRNA | rs10414008 | 33758327 | 2.56e-08 | 5 | 3 | 0 | 0 | 0 | 2 | Female only | Female only | Female only | True | False |
| MONORE | 8 | CCDC26 | lincRNA | rs34976632 | 130620056 | 2.74e-08 | 3 | 0 | 0 | 0 | 0 | 3 | Both sexes | Both sexes | Male only | True | False |
| MONORE | 17 | GSDMA | protein_coding | rs7212944 | 38122686 | 2.76e-08 | 77 | 0 | 0 | 0 | 10 | 67 | Female only | Both sexes | Female only | True | False |
| MONORE | 6 | CCHCR1 | protein_coding | rs3094225 | 31113052 | 2.96e-08 | 111 | 7 | 22 | 28 | 17 | 37 | Female only | Both sexes | Female only | True | True |
| MONORE | 2 | MBD5 | protein_coding | rs12992231 | 148799710 | 4.21e-08 | 335 | 119 | 22 | 1 | 26 | 167 | Male only | Both sexes | Male only | True | False |
| MONORE | 20 | RP11-290F20.1 | lincRNA | rs1889237 | 48905978 | 4.40e-08 | 5 | 0 | 0 | 0 | 2 | 3 | Female only | Both sexes | Female only | True | False |
| MONORE | 3 | HIGD1A | protein_coding | rs6764909 | 42835889 | 4.45e-08 | 5 | 0 | 0 | 0 | 0 | 5 | Female only | Both sexes | Female only | True | False |
| MONORE | 2 | RNA5SP106 | rRNA | rs6748985 | 148470093 | 4.55e-08 | 99 | 50 | 0 | 0 | 0 | 49 | Male only | Male only | Male only | True | False |
| MONORE | 1 | AC093577.1 | miRNA | rs78663632 | 93448422 | 4.84e-08 | 201 | 6 | 93 | 2 | 0 | 100 | Female only | Both sexes | Female only | True | False |
| MPV | 7 | CTB-111H14.1 | sense_overlapping | rs342290 | 106367604 | 1.06e-83 | 123 | 0 | 0 | 0 | 0 | 123 | Both sexes | Both sexes | Both sexes | True | False |
| MPV | 3 | ARHGEF3 | protein_coding | rs1354034 | 56849749 | 1.73e-78 | 19 | 0 | 0 | 0 | 0 | 19 | Both sexes | Both sexes | Both sexes | True | False |
| MPV | 10 | JMJD1C | protein_coding | rs7098181 | 65027143 | 1.00e-66 | 157 | 0 | 0 | 0 | 0 | 157 | Both sexes | Both sexes | Both sexes | True | False |
| MPV | 1 | TMCC2 | protein_coding | rs1668868 | 205240141 | 1.44e-55 | 215 | 0 | 0 | 0 | 0 | 215 | Both sexes | Both sexes | Both sexes | True | False |
| MPV | 10 | REEP3 | protein_coding | rs7075901 | 65280994 | 2.80e-53 | 67 | 0 | 0 | 0 | 0 | 67 | Both sexes | Both sexes | Both sexes | True | False |

| pheno | CHR | Gene_Symbol | biotype | lead_snp | lead_BP | lead_P | TOTAL | NSIG | S05 | S01 | S001 | S0001 | independent_loci_in_sex_P1e6 | independent_loci_in_sex_P1e2 | independent_loci_in_sex_P5e8 | in_gwas_catalog | in_HLA |
|---|---|---|---|---|---|---|---|---|---|---|---|---|---|---|---|---|---|
| MPV | 17 | TAOK1 | protein_coding | rs6505129 | 27753618 | 4.51e-44 | 115 | 0 | 0 | 0 | 0 | 115 | Both sexes | Both sexes | Both sexes | True | False |
| MPV | 12 | TMEM120B | protein_coding | rs73227958 | 122149021 | 6.34e-43 | 51 | 0 | 0 | 0 | 0 | 51 | Both sexes | Both sexes | Both sexes | True | False |
| MPV | 12 | PSMD9 | protein_coding | rs7132055 | 122346034 | 5.77e-40 | 85 | 0 | 12 | 7 | 4 | 62 | Both sexes | Both sexes | Both sexes | True | False |
| MPV | 3 | ARHGEF3 | protein_coding | rs17288922 | 56851387 | 1.60e-37 | 11 | 0 | 0 | 0 | 0 | 11 | Both sexes | Both sexes | Both sexes | True | False |
| MPV | 3 | ARHGEF3 | protein_coding | rs13061631 | 56862390 | 2.35e-35 | 7 | 0 | 0 | 0 | 0 | 7 | Both sexes | Both sexes | Both sexes | True | False |
| MPV | 12 | TMEM120B | protein_coding | rs10840615 | 122203680 | 6.01e-32 | 11 | 0 | 0 | 0 | 0 | 11 | Both sexes | Both sexes | Both sexes | True | False |
| MPV | 11 | SIRT3 | protein_coding | rs11607019 | 234349 | 1.04e-30 | 87 | 0 | 0 | 0 | 0 | 87 | Both sexes | Both sexes | Both sexes | True | False |
| MPV | 12 | SETD1B | protein_coding | rs3741593 | 122261456 | 1.28e-30 | 7 | 0 | 0 | 0 | 0 | 7 | Both sexes | Both sexes | Both sexes | True | False |
| MPV | 17 | TAOK1 | protein_coding | rs4795512 | 27788504 | 1.60e-30 | 21 | 0 | 0 | 0 | 0 | 21 | Both sexes | Both sexes | Both sexes | True | False |
| MPV | 10 | JMJD1C | protein_coding | rs9415676 | 65010626 | 1.70e-30 | 125 | 0 | 0 | 0 | 0 | 125 | Both sexes | Both sexes | Both sexes | True | False |
| MPV | 12 | RP3-454B23.1 | lincRNA | rs10774409 | 6288322 | 1.99e-29 | 123 | 0 | 0 | 0 | 0 | 123 | Both sexes | Both sexes | Both sexes | True | False |
| MPV | 20 | SIRPA | protein_coding | rs6136492 | 1923972 | 3.13e-29 | 31 | 0 | 0 | 0 | 0 | 31 | Both sexes | Both sexes | Both sexes | True | False |
| MPV | 1 | DSTYK | protein_coding | rs10900472 | 205181526 | 4.79e-29 | 139 | 0 | 0 | 0 | 0 | 139 | Both sexes | Both sexes | Both sexes | True | False |
| MPV | 1 | DNM3 | protein_coding | rs10914127 | 171943948 | 1.12e-28 | 15 | 0 | 0 | 0 | 0 | 15 | Both sexes | Both sexes | Both sexes | True | False |
| MPV | 10 | REEP3 | protein_coding | rs10995554 | 65295110 | 1.92e-27 | 51 | 0 | 0 | 0 | 0 | 51 | Both sexes | Both sexes | Both sexes | True | False |
| MPV | 3 | ARHGEF3 | protein_coding | rs55709294 | 56874996 | 1.15e-25 | 13 | 0 | 0 | 0 | 0 | 13 | Both sexes | Both sexes | Both sexes | True | False |
| pheno | CHR | Gene_Symbol | biotype | lead_snp | lead_BP | lead_P | TOTAL | NSIG | S05 | S01 | S001 | S0001 | independent_loci_in_sex_P1e6 | independent_loci_in_sex_P1e2 | independent_loci_in_sex_P5e8 | in_gwas_catalog | in_HLA |
| MPV | 18 | CD226 | protein_coding | rs1790947 | 67527760 | 4.14e-25 | 37 | 0 | 0 | 0 | 0 | 37 | Both sexes | Both sexes | Both sexes | True | False |
| MPV | 17 | TAOK1 | protein_coding | rs8070136 | 27783295 | 1.08e-24 | 31 | 0 | 0 | 0 | 0 | 31 | Both sexes | Both sexes | Both sexes | True | False |
| MPV | 17 | TAOK1 | protein_coding | rs7210089 | 27728815 | 2.32e-24 | 5 | 0 | 0 | 0 | 0 | 5 | Both sexes | Both sexes | Both sexes | True | False |
| MPV | 12 | PSMD9 | protein_coding | rs71456717 | 122315537 | 2.69e-24 | 103 | 0 | 0 | 3 | 0 | 100 | Both sexes | Both sexes | Both sexes | True | False |
| MPV | 10 | RNU6-543P | snRNA | rs1396965 | 64839570 | 3.51e-24 | 51 | 0 | 0 | 0 | 0 | 51 | Both sexes | Both sexes | Both sexes | True | False |
| MPV | 22 | PACSIN2 | protein_coding | rs2267480 | 43362976 | 5.44e-22 | 443 | 0 | 0 | 0 | 73 | 370 | Female only | Both sexes | Female only | True | False |
| MPV | 8 | ZFPM2 | protein_coding | rs4734879 | 106583124 | 8.26e-22 | 63 | 0 | 0 | 0 | 0 | 63 | Both sexes | Both sexes | Both sexes | True | False |
| MPV | 3 | ARHGEF3 | protein_coding | rs11916112 | 56924711 | 1.31e-20 | 25 | 0 | 0 | 0 | 0 | 25 | Both sexes | Both sexes | Both sexes | True | False |
| MPV | 23 | MIR1587 | lincRNA | X:39710195:C:T | 39710195 | 1.41e-20 | 15 | 0 | 0 | 0 | 0 | 15 | Both sexes | Both sexes | Both sexes | True | False |

| pheno | CHR | Gene_Symbol | biotype | lead_snp | lead_BP | lead_P | TOTAL | NSIG | S05 | S01 | S001 | S0001 | independent_loci_in_sex_P1e6 | independent_loci_in_sex_P1e2 | independent_loci_in_sex_P5e8 | in_gwas_catalog | in_HLA |
|---|---|---|---|---|---|---|---|---|---|---|---|---|---|---|---|---|---|
| MPV | 1 | DSTYK | protein_coding | rs2369634 | 205157903 | 2.32e-20 | 117 | 0 | 0 | 0 | 0 | 117 | Both sexes | Both sexes | Both sexes | True | False |
| MPV | 2 | EHD3 | protein_coding | rs655029 | 31477838 | 5.57e-19 | 39 | 0 | 0 | 0 | 0 | 39 | Both sexes | Both sexes | Both sexes | True | False |
| MPV | 3 | ARHGEF3 | protein_coding | rs1500707 | 56876636 | 1.59e-18 | 11 | 0 | 0 | 0 | 0 | 11 | Both sexes | Both sexes | Both sexes | True | False |
| MPV | 4 | KIAA0232 | protein_coding | rs11731274 | 6891455 | 2.54e-18 | 3 | 0 | 0 | 0 | 0 | 3 | Both sexes | Both sexes | Both sexes | True | False |
| MPV | 1 | CNTN2 | protein_coding | rs10793725 | 205028915 | 4.30e-18 | 17 | 4 | 0 | 1 | 1 | 11 | Both sexes | Both sexes | Both sexes | True | False |
| MPV | 12 | COPZ1 | protein_coding | rs10876550 | 54712308 | 7.06e-18 | 9 | 0 | 0 | 0 | 0 | 9 | Both sexes | Both sexes | Both sexes | True | False |
| MPV | 1 | NUAK2 | protein_coding | rs17345997 | 205279612 | 8.04e-18 | 11 | 0 | 0 | 0 | 0 | 11 | Both sexes | Both sexes | Both sexes | True | False |
| MPV | 20 | SIRPA | protein_coding | rs6136475 | 1922237 | 1.07e-17 | 5 | 0 | 0 | 0 | 0 | 5 | Both sexes | Both sexes | Both sexes | True | False |
| MPV | 10 | JMJD1C | protein_coding | rs10740109 | 65040139 | 1.11e-17 | 179 | 0 | 81 | 8 | 1 | 89 | Female only | Both sexes | Female only | True | False |
| MPV | 5 | F2R | protein_coding | rs250721 | 76052076 | 1.12e-17 | 11 | 0 | 0 | 0 | 0 | 11 | Both sexes | Both sexes | Female only | True | False |
| MPV | 12 | BAZ2A | protein_coding | rs2950387 | 57021006 | 4.78e-17 | 125 | 0 | 0 | 0 | 0 | 125 | Both sexes | Both sexes | Both sexes | True | False |
| MPV | 11 | BET1L | protein_coding | rs4980320 | 198986 | 5.89e-17 | 7 | 0 | 2 | 1 | 0 | 4 | Both sexes | Both sexes | Female only | True | False |
| MPV | 10 | MRPL35P2 | pseudogene | rs12246748 | 65392260 | 9.93e-17 | 3 | 0 | 0 | 0 | 0 | 3 | Both sexes | Both sexes | Both sexes | True | False |
| MPV | 12 | WDR66 | protein_coding | rs1169076 | 122429484 | 5.13e-16 | 61 | 0 | 2 | 26 | 3 | 30 | Female only | Both sexes | Female only | True | False |
| MPV | 1 | KIF1B | protein_coding | rs6694081 | 10443560 | 1.79e-15 | 61 | 0 | 0 | 0 | 0 | 61 | Both sexes | Both sexes | Both sexes | True | False |
| MPV | 20 | RP4-576H24.4 | protein_coding | rs6074894 | 1597050 | 2.52e-15 | 15 | 0 | 0 | 0 | 4 | 11 | Both sexes | Both sexes | Both sexes | True | False |
| MPV | 3 | ARHGEF3 | protein_coding | rs62251088 | 56941975 | 2.65e-15 | 25 | 0 | 0 | 0 | 0 | 25 | Both sexes | Both sexes | Both sexes | True | False |
| MPV | 5 | F2RL1 | protein_coding | rs3135471 | 76123570 | 5.88e-15 | 25 | 0 | 0 | 0 | 2 | 23 | Female only | Both sexes | Female only | True | False |
| MPV | 12 | RP11-87C12.5 | lincRNA | rs745327 | 122456524 | 6.94e-15 | 9 | 0 | 0 | 0 | 0 | 9 | Both sexes | Both sexes | Both sexes | True | False |
| MPV | 6 | LRRC16A | protein_coding | rs214060 | 25537194 | 1.27e-14 | 45 | 0 | 0 | 4 | 2 | 39 | Both sexes | Both sexes | Female only | True | False |
| MPV | 11 | PSMD13 | protein_coding | rs10751643 | 253072 | 2.08e-14 | 13 | 0 | 5 | 0 | 0 | 8 | Female only | Both sexes | Female only | True | False |
| MPV | 10 | REEP3 | protein_coding | rs112252722 | 65271327 | 2.90e-14 | 5 | 0 | 0 | 0 | 0 | 5 | Both sexes | Both sexes | Both sexes | True | False |
| MPV | 1 | NUAK2 | protein_coding | rs41264875 | 205272347 | 3.25e-14 | 7 | 0 | 0 | 0 | 1 | 6 | Both sexes | Both sexes | Female only | True | False |
| MPV | 13 | GRTP1 | protein_coding | rs4907618 | 114015015 | 8.68e-14 | 11 | 0 | 0 | 0 | 3 | 8 | Female only | Both sexes | Female only | True | False |
| MPV | 22 | PACSIN2 | protein_coding | rs1071960 | 43349632 | 1.04e-13 | 15 | 0 | 0 | 0 | 0 | 15 | Female only | Both sexes | Female only | True | False |
| MPV | 20 | RP5-968J1.1 | lincRNA | rs156355 | 1818613 | 1.22e-13 | 15 | 0 | 0 | 0 | 0 | 15 | Both sexes | Both sexes | Both sexes | True | False |

| pheno | CHR | Gene_Symbol | biotype | lead_snp | lead_BP | lead_P | TOTAL | NSIG | S05 | S01 | S001 | S0001 | independent_loci_in_sex_P1e6 | independent_loci_in_sex_P1e2 | independent_loci_in_sex_P5e8 | in_gwas_catalog | in_HLA |
|---|---|---|---|---|---|---|---|---|---|---|---|---|---|---|---|---|---|
| MPV | 12 | SETD1B | protein_coding | rs1678960 | 122264635 | 1.52e-13 | 15 | 7 | 0 | 0 | 1 | 7 | Female only | Both sexes | Female only | True | False |
| MPV | 15 | AC069368.3 | protein_coding | rs1719271 | 65183801 | 5.19e-13 | 65 | 0 | 0 | 1 | 1 | 63 | Both sexes | Both sexes | Female only | True | False |
| MPV | 5 | F2R | protein_coding | rs37244 | 76015111 | 1.31e-12 | 11 | 0 | 0 | 0 | 0 | 11 | Both sexes | Both sexes | Female only | True | False |
| MPV | 12 | RP3-454B23.1 | lincRNA | rs11064058 | 6263819 | 1.42e-12 | 19 | 0 | 0 | 0 | 0 | 19 | Both sexes | Both sexes | Both sexes | True | False |
| MPV | 17 | GIT1 | protein_coding | rs894606 | 27909352 | 1.80e-12 | 3 | 0 | 0 | 0 | 0 | 3 | Female only | Both sexes | Female only | True | False |
| MPV | 10 | EGR2 | protein_coding | rs11595405 | 64763893 | 3.70e-12 | 17 | 0 | 0 | 5 | 2 | 10 | Both sexes | Both sexes | Both sexes | True | False |
| MPV | 12 | FAR2 | protein_coding | rs1006409 | 29435675 | 5.49e-12 | 189 | 0 | 0 | 0 | 0 | 189 | Both sexes | Both sexes | Both sexes | True | False |
| MPV | 5 | RNF145 | protein_coding | rs7718670 | 158593373 | 7.40e-12 | 17 | 0 | 0 | 0 | 0 | 17 | Both sexes | Both sexes | Both sexes | True | False |
| MPV | 3 | ARHGEF3 | protein_coding | rs62249773 | 56853922 | 8.78e-12 | 53 | 0 | 0 | 0 | 0 | 53 | Both sexes | Both sexes | Both sexes | True | False |
| MPV | 17 | MYO18A | protein_coding | rs8069100 | 27439527 | 9.50e-12 | 95 | 10 | 32 | 6 | 0 | 47 | Female only | Both sexes | Female only | True | False |
| MPV | 3 | ARHGEF3 | protein_coding | rs9311629 | 56934729 | 1.23e-11 | 3 | 0 | 1 | 0 | 1 | 1 | Both sexes | Both sexes | Female only | True | False |
| MPV | 20 | RP4-576H24.4 | protein_coding | rs2253427 | 1552430 | 1.26e-11 | 45 | 12 | 8 | 2 | 15 | 8 | Female only | Both sexes | Female only | True | False |
| MPV | 12 | ORAI1 | protein_coding | rs712853 | 122079668 | 1.33e-11 | 27 | 0 | 0 | 0 | 0 | 27 | Both sexes | Both sexes | Female only | True | False |
| MPV | 5 | SLC22A5 | protein_coding | rs200838 | 131710399 | 1.34e-11 | 249 | 0 | 19 | 27 | 79 | 124 | Female only | Both sexes | Female only | True | False |
| MPV | 11 | FADS2 | protein_coding | rs28456 | 61589481 | 1.83e-11 | 89 | 0 | 2 | 30 | 12 | 45 | Female only | Both sexes | Female only | True | False |
| MPV | 20 | TUBB1 | protein_coding | rs6070696 | 57597645 | 2.06e-11 | 51 | 0 | 0 | 16 | 11 | 24 | Both sexes | Both sexes | Both sexes | True | False |
| MPV | 23 | RP11-445O16.2 | pseudogene | X:57089149:G:T | 57089149 | 2.11e-11 | 823 | 0 | 0 | 0 | 0 | 823 | Both sexes | Both sexes | Both sexes | True | False |
| MPV | 9 | GFI1B | protein_coding | rs73554552 | 135861502 | 2.13e-11 | 3 | 0 | 0 | 0 | 0 | 3 | Female only | Both sexes | Female only | True | False |
| MPV | 1 | DNM3 | protein_coding | rs733189 | 172095365 | 2.64e-11 | 11 | 0 | 1 | 0 | 1 | 9 | Female only | Both sexes | Female only | True | False |
| MPV | 10 | RP11-351O1.3 | pseudogene | rs12773976 | 65264300 | 2.90e-11 | 7 | 0 | 0 | 0 | 1 | 6 | Both sexes | Both sexes | Female only | True | False |
| MPV | 2 | EHD3 | protein_coding | rs625132 | 31482300 | 3.40e-11 | 11 | 5 | 0 | 1 | 0 | 5 | Female only | Both sexes | Female only | True | False |
| MPV | 14 | RP11-736N17.8 | sense_overlapping | rs59643720 | 103564807 | 3.79e-11 | 21 | 0 | 0 | 0 | 0 | 21 | Female only | Both sexes | Female only | True | False |
| MPV | 9 | BRD3 | protein_coding | rs2810490 | 136923118 | 5.31e-11 | 55 | 2 | 26 | 0 | 0 | 27 | Female only | Female only | Female only | True | False |
| MPV | 12 | ORAI1 | protein_coding | rs6486776 | 122057293 | 5.74e-11 | 17 | 0 | 0 | 0 | 0 | 17 | Female only | Both sexes | Female only | True | False |
| MPV | 3 | ARHGEF3 | protein_coding | rs7640115 | 56862006 | 7.73e-11 | 23 | 0 | 0 | 0 | 0 | 23 | Both sexes | Both sexes | Both sexes | True | False |

| pheno | CHR | Gene_Symbol | biotype | lead_snp | lead_BP | lead_P | TOTAL | NSIG | S05 | S01 | S001 | S0001 | independent_loci_in_sex_P1e6 | independent_loci_in_sex_P1e2 | independent_loci_in_sex_P5e8 | in_gwas_catalog | in_HLA |
|---|---|---|---|---|---|---|---|---|---|---|---|---|---|---|---|---|---|
| MPV | 23 | FAAH2 | protein_coding | X:57430179:T:C | 57430179 | 8.30e-11 | 387 | 0 | 0 | 0 | 0 | 387 | Both sexes | Both sexes | Both sexes | True | False |
| MPV | 18 | PPP4R1 | protein_coding | rs428598 | 9595853 | 1.16e-10 | 57 | 7 | 5 | 2 | 15 | 28 | Female only | Both sexes | Female only | True | False |
| MPV | 10 | REEP3 | protein_coding | rs61855876 | 65357541 | 1.22e-10 | 127 | 17 | 46 | 1 | 0 | 63 | Female only | Both sexes | Female only | True | False |
| MPV | 12 | DAZAP2 | protein_coding | rs115577237 | 51661433 | 1.30e-10 | 75 | 0 | 1 | 20 | 17 | 37 | Female only | Both sexes | Female only | True | False |
| MPV | 8 | ZFPM2 | protein_coding | rs6993696 | 106581284 | 1.36e-10 | 49 | 0 | 0 | 15 | 5 | 29 | Both sexes | Both sexes | Female only | True | False |
| MPV | 20 | RP4-684O24.5 | antisense | rs6045639 | 1934548 | 1.60e-10 | 5 | 0 | 0 | 0 | 0 | 5 | Both sexes | Both sexes | Female only | True | False |
| MPV | 18 | CABLES1 | protein_coding | rs11082304 | 20720973 | 1.71e-10 | 3 | 0 | 0 | 0 | 0 | 3 | Both sexes | Both sexes | Female only | True | False |
| MPV | 1 | CNTN2 | protein_coding | rs11240351 | 205044339 | 1.75e-10 | 9 | 0 | 0 | 0 | 3 | 6 | Both sexes | Both sexes | Female only | True | False |
| MPV | 12 | HPD | protein_coding | rs2620340 | 122291261 | 1.77e-10 | 55 | 26 | 2 | 0 | 0 | 27 | Female only | Female only | Female only | True | False |
| MPV | 6 | HCP5 | sense_overlapping | rs2596468 | 31392308 | 2.02e-10 | 63 | 0 | 0 | 0 | 8 | 55 | Female only | Both sexes | Female only | True | True |
| MPV | 11 | NLRP6 | protein_coding | rs6598050 | 272010 | 2.21e-10 | 3 | 0 | 1 | 0 | 0 | 2 | Female only | Both sexes | Female only | True | False |
| MPV | 1 | DNM3 | protein_coding | rs72719086 | 172127946 | 3.27e-10 | 43 | 0 | 0 | 0 | 2 | 41 | Both sexes | Both sexes | Female only | True | False |
| MPV | 17 | TAOK1 | protein_coding | rs11651087 | 27736904 | 3.43e-10 | 19 | 0 | 0 | 0 | 0 | 19 | Both sexes | Both sexes | Female only | True | False |
| MPV | 1 | DNM3 | protein_coding | rs56125409 | 171929451 | 3.62e-10 | 5 | 0 | 0 | 0 | 0 | 5 | Both sexes | Both sexes | Both sexes | True | False |
| MPV | 23 | RP1-49C23.1 | pseudogene | X:57810911:G:A | 57810911 | 3.88e-10 | 75 | 0 | 0 | 0 | 0 | 75 | Both sexes | Both sexes | Both sexes | True | False |
| MPV | 17 | MYO18A | protein_coding | rs16964837 | 27467093 | 3.92e-10 | 59 | 6 | 24 | 0 | 0 | 29 | Female only | Female only | Female only | True | False |
| MPV | 10 | JMJD1C | protein_coding | rs12773283 | 65058224 | 4.16e-10 | 105 | 15 | 38 | 0 | 0 | 52 | Female only | Female only | Female only | True | False |
| MPV | 23 | RP11-622K12.1 | processed_transcript | X:56793966:C:T | 56793966 | 4.23e-10 | 461 | 0 | 0 | 0 | 0 | 461 | Both sexes | Both sexes | Both sexes | True | False |
| MPV | 6 | VWA7 | protein_coding | rs6914412 | 31742242 | 5.00e-10 | 31 | 0 | 0 | 2 | 14 | 15 | Female only | Both sexes | Female only | True | True |
| MPV | 6 | LRRC16A | protein_coding | rs301369 | 25520772 | 5.34e-10 | 133 | 0 | 0 | 0 | 7 | 126 | Female only | Both sexes | Female only | True | False |
| MPV | 12 | FAR2 | protein_coding | rs4931170 | 29388772 | 5.87e-10 | 117 | 0 | 0 | 3 | 3 | 111 | Both sexes | Both sexes | Male only | True | False |
| MPV | 10 | JMJD1C | protein_coding | rs138000301 | 65041324 | 5.96e-10 | 13 | 0 | 0 | 8 | 0 | 5 | Both sexes | Both sexes | Female only | True | False |
| MPV | 23 | KLF8 | protein_coding | X:56306059:T:G | 56306059 | 6.80e-10 | 531 | 0 | 0 | 0 | 0 | 531 | Both sexes | Both sexes | Both sexes | True | False |
| MPV | 9 | BRD3 | protein_coding | rs10993905 | 136927088 | 8.10e-10 | 61 | 0 | 0 | 31 | 0 | 30 | Female only | Both sexes | Female only | True | False |
| MPV | 17 | TWF1P1 | pseudogene | rs61314136 | 27545367 | 8.26e-10 | 29 | 1 | 3 | 11 | 0 | 14 | Female only | Both sexes | Female only | True | False |
| MPV | 18 | CD226 | protein_coding | rs17208112 | 67557429 | 9.14e-10 | 3 | 0 | 0 | 0 | 0 | 3 | Female only | Both sexes | Female only | True | False |

| pheno | CHR | Gene_Symbol | biotype | lead_snp | lead_BP | lead_P | TOTAL | NSIG | S05 | S01 | S001 | S0001 | independent_loci_in_sex_P1e6 | independent_loci_in_sex_P1e2 | independent_loci_in_sex_P5e8 | in_gwas_catalog | in_HLA |
|---|---|---|---|---|---|---|---|---|---|---|---|---|---|---|---|---|---|
| MPV | 3 | PDIA5 | protein_coding | rs3792378 | 122845077 | 1.16e-09 | 23 | 0 | 0 | 0 | 0 | 23 | Female only | Both sexes | Female only | True | False |
| MPV | 12 | RP11-74M13.4 | pseudogene | rs7957053 | 57214086 | 1.24e-09 | 7 | 0 | 0 | 2 | 2 | 3 | Female only | Both sexes | Female only | True | False |
| MPV | 23 | MIR1587 | lincRNA | X:39702609:G:A | 39702609 | 1.32e-09 | 5 | 0 | 0 | 0 | 0 | 5 | Both sexes | Both sexes | Both sexes | True | False |
| MPV | 2 | EHD3 | protein_coding | rs11694688 | 31471174 | 1.45e-09 | 19 | 1 | 0 | 0 | 1 | 17 | Both sexes | Both sexes | Female only | True | False |
| MPV | 5 | IQGAP2 | protein_coding | rs62362044 | 75908292 | 1.50e-09 | 9 | 0 | 4 | 1 | 0 | 4 | Male only | Both sexes | Male only | True | False |
| MPV | 12 | TMEM120B | protein_coding | rs58684387 | 122178250 | 1.75e-09 | 7 | 0 | 4 | 0 | 0 | 3 | Female only | Female only | Female only | True | False |
| MPV | 1 | TRIM58 | protein_coding | rs4925750 | 248038325 | 1.90e-09 | 31 | 0 | 1 | 0 | 10 | 20 | Both sexes | Both sexes | Both sexes | True | False |
| MPV | 15 | TPM1 | protein_coding | rs4775608 | 63331940 | 2.11e-09 | 33 | 0 | 0 | 0 | 0 | 33 | Both sexes | Both sexes | Both sexes | True | False |
| MPV | 1 | TRIM58 | protein_coding | rs10788728 | 248013181 | 2.76e-09 | 41 | 0 | 0 | 2 | 4 | 35 | Both sexes | Both sexes | Both sexes | True | False |
| MPV | 17 | TWF1P1 | pseudogene | rs7213514 | 27537242 | 3.04e-09 | 11 | 1 | 0 | 5 | 0 | 5 | Female only | Both sexes | Female only | True | False |
| MPV | 10 | RNU6-543P | snRNA | rs72824926 | 64844888 | 4.16e-09 | 17 | 0 | 0 | 0 | 6 | 11 | Both sexes | Both sexes | Female only | True | False |
| MPV | 1 | DNM3 | protein_coding | rs10910971 | 172013991 | 5.28e-09 | 9 | 7 | 1 | 0 | 0 | 1 | Both sexes | Both sexes | Female only | True | False |
| MPV | 5 | RN7SL208P | misc_RNA | rs4704366 | 76137222 | 5.37e-09 | 7 | 0 | 2 | 1 | 1 | 3 | Female only | Both sexes | Female only | True | False |
| MPV | 20 | SIRPB1 | protein_coding | rs11699591 | 1605165 | 6.40e-09 | 9 | 0 | 0 | 1 | 4 | 4 | Female only | Both sexes | Female only | True | False |
| MPV | 6 | NOTCH4 | protein_coding | rs374205 | 32196873 | 7.07e-09 | 55 | 0 | 19 | 9 | 0 | 27 | Female only | Both sexes | Female only | True | True |
| MPV | 19 | FXYD5 | protein_coding | rs9807816 | 35684944 | 8.58e-09 | 11 | 6 | 0 | 0 | 0 | 5 | Female only | Female only | Female only | True | False |
| MPV | 1 | TRIM58 | protein_coding | rs4925753 | 248040104 | 8.86e-09 | 19 | 4 | 2 | 2 | 2 | 9 | Female only | Both sexes | Female only | True | False |
| MPV | 9 | DOCK8 | protein_coding | rs16930416 | 330351 | 9.23e-09 | 51 | 14 | 8 | 22 | 0 | 7 | Female only | Both sexes | Female only | True | False |
| MPV | 17 | AP2B1 | protein_coding | rs226085 | 34019488 | 9.38e-09 | 15 | 1 | 0 | 1 | 7 | 6 | Male only | Both sexes | Male only | True | False |
| MPV | 17 | GIT1 | protein_coding | rs550818 | 27901975 | 9.45e-09 | 19 | 0 | 0 | 0 | 0 | 19 | Both sexes | Both sexes | Female only | True | False |
| MPV | 20 | ATP5E | protein_coding | rs151353 | 57604343 | 1.00e-08 | 15 | 2 | 2 | 0 | 2 | 9 | Female only | Both sexes | Female only | True | False |
| MPV | 12 | ERGIC2 | protein_coding | rs2278094 | 29534492 | 1.09e-08 | 25 | 1 | 1 | 2 | 10 | 11 | Female only | Both sexes | Female only | True | False |
| MPV | 1 | TRIM58 | protein_coding | rs61855702 | 248023069 | 1.32e-08 | 7 | 0 | 0 | 0 | 0 | 7 | Both sexes | Both sexes | Male only | True | False |
| MPV | 23 | RP13-188A5.1 | lincRNA | X:56017005:C:T | 56017005 | 1.39e-08 | 433 | 0 | 0 | 0 | 8 | 425 | Both sexes | Both sexes | Both sexes | True | False |
| MPV | 6 | LRRC16A | protein_coding | rs530145 | 25546801 | 1.53e-08 | 47 | 12 | 5 | 18 | 2 | 10 | Female only | Both sexes | Female only | True | False |
| MPV | 20 | SIRPD | protein_coding | rs11697395 | 1538266 | 1.58e-08 | 51 | 0 | 14 | 5 | 0 | 32 | Both sexes | Both sexes | Male only | True | False |

| phen o | CHR | Gene_Sy mbol | biotype | lead_snp | lead_BP | lead_P | TOT AL | NSIG | S05 | S01 | S001 | S000 1 | independent_loci_in_sex_P1 e6 | independent_loci_in_sex_P1 e2 | independent_loci_in_sex_P5 e8 | in_gwas_catal og | in_H LA |
|---|---|---|---|---|---|---|---|---|---|---|---|---|---|---|---|---|---|
| MPV | 5 | RNF145 | protein_coding | rs2163775 | 158626347 | 1.60e-08 | 9 | 0 | 0 | 1 | 1 | 7 | Female only | Both sexes | Female only | True | False |
| MPV | 1 | RP11-978I15.10 | antisense | rs12119496 | 247742899 | 1.64e-08 | 11 | 0 | 0 | 0 | 0 | 11 | Female only | Both sexes | Female only | True | False |
| MPV | 6 | XXbac-BPG248L24.12 | pseudogene | rs9266233 | 31325430 | 1.67e-08 | 9 | 4 | 3 | 1 | 0 | 1 | Female only | Both sexes | Female only | True | True |
| MPV | 14 | RAD51B | protein_coding | rs10133883 | 68513492 | 1.95e-08 | 399 | 28 | 152 | 21 | 16 | 182 | Male only | Both sexes | Male only | True | False |
| MPV | 11 | PSMD13 | protein_coding | rs2686878 | 253958 | 2.20e-08 | 63 | 0 | 0 | 6 | 27 | 30 | Female only | Both sexes | Female only | True | False |
| MPV | 17 | NUFIP2 | protein_coding | rs12452857 | 27613677 | 2.21e-08 | 17 | 2 | 0 | 7 | 0 | 8 | Female only | Both sexes | Female only | True | False |
| MPV | 11 | TMEM258 | protein_coding | rs61896141 | 61556039 | 2.36e-08 | 19 | 0 | 0 | 0 | 6 | 13 | Both sexes | Both sexes | Male only | True | False |
| MPV | 1 | DNM3 | protein_coding | rs75190610 | 172122670 | 2.38e-08 | 3 | 0 | 1 | 1 | 0 | 1 | Female only | Both sexes | Female only | True | False |
| phen o | CHR | Gene_Sy mbol | biotype | lead_snp | lead_BP | lead_P | TOT AL | NSIG | S05 | S01 | S001 | S000 1 | independent_loci_in_sex_P1 e6 | independent_loci_in_sex_P1 e2 | independent_loci_in_sex_P5 e8 | in_gwas_catal og | in_H LA |
| MPV | 8 | ASAP1 | protein_coding | rs11774659 | 131190336 | 2.40e-08 | 83 | 0 | 0 | 0 | 0 | 83 | Both sexes | Both sexes | Both sexes | True | False |
| MPV | 12 | OVCH1-AS1 | protein_coding | rs10743664 | 29558585 | 2.63e-08 | 25 | 0 | 0 | 0 | 2 | 23 | Male only | Both sexes | Male only | True | False |
| MPV | 22 | PACSIN2 | protein_coding | rs3091364 | 43414330 | 2.76e-08 | 3 | 1 | 0 | 1 | 0 | 1 | Female only | Both sexes | Female only | True | False |
| MPV | 17 | MYO18A | protein_coding | rs12601994 | 27447905 | 3.25e-08 | 31 | 8 | 8 | 0 | 0 | 15 | Female only | Female only | Female only | True | False |
| MPV | 7 | CCDC71L | protein_coding | rs2190093 | 106299404 | 3.41e-08 | 11 | 0 | 5 | 1 | 0 | 5 | Female only | Both sexes | Female only | True | False |
| MPV | 2 | CPS1 | protein_coding | rs56098529 | 211397176 | 3.48e-08 | 135 | 68 | 0 | 0 | 18 | 49 | Female only | Female only | Female only | True | False |
| MPV | 6 | ZDHHC20P2 | pseudogene | rs9266658 | 31347644 | 3.48e-08 | 23 | 0 | 0 | 1 | 11 | 11 | Female only | Both sexes | Female only | True | True |
| MPV | 3 | KALRN | protein_coding | rs7616881 | 124341152 | 3.53e-08 | 13 | 0 | 0 | 1 | 1 | 11 | Both sexes | Both sexes | Male only | True | False |
| MPV | 22 | RPL5P34 | pseudogene | rs12162975 | 43175426 | 4.24e-08 | 5 | 0 | 0 | 1 | 0 | 4 | Female only | Both sexes | Female only | True | False |
| MPV | 13 | GRTP1 | protein_coding | rs79236986 | 114020321 | 4.59e-08 | 9 | 2 | 0 | 2 | 0 | 5 | Female only | Both sexes | Female only | True | False |
| Meta | 23 | RP5-998G20.1 | pseudogene | X:87304109:T:G | 87304109 | 3.10e-08 | 289 | 145 | 0 | 0 | 4 | 140 | Male only | Male only | Male only | False | False |
| Mg | 1 | THBS3 | protein_coding | rs2066981 | 155172379 | 1.74e-10 | 77 | 0 | 0 | 6 | 9 | 62 | Female only | Both sexes | Female only | True | False |
| Mg | 1 | DCAF6 | protein_coding | rs12136645 | 167913302 | 2.85e-08 | 243 | 122 | 0 | 0 | 4 | 117 | Male only | Male only | Male only | True | False |
| MmAcid | 2 | HIBCH | protein_coding | rs291429 | 191197941 | 1.13e-14 | 69 | 0 | 0 | 0 | 0 | 69 | Both sexes | Both sexes | Both sexes | True | False |
| MmAcid | 2 | INPP1 | protein_coding | rs3791813 | 191218371 | 1.02e-10 | 439 | 0 | 0 | 0 | 1 | 438 | Both sexes | Both sexes | Both sexes | True | False |
| MonAbs | 2 | ITGA4 | protein_coding | rs1375493 | 182323766 | 2.63e-17 | 15 | 0 | 0 | 0 | 0 | 15 | Both sexes | Both sexes | Both sexes | True | False |

| pheno | CHR | Gene_Symbol | biotype | lead_snp | lead_BP | lead_P | TOTAL | NSIG | S05 | S01 | S001 | S0001 | independent_loci_in_sex_P1e6 | independent_loci_in_sex_P1e2 | independent_loci_in_sex_P5e8 | in_gwas_catalog | in_HLA |
|---|---|---|---|---|---|---|---|---|---|---|---|---|---|---|---|---|---|
| MonAbs | 9 | RP11-202G18.1 | lincRNA | rs12339649 | 113945046 | 1.64e-16 | 51 | 0 | 0 | 0 | 0 | 51 | Both sexes | Both sexes | Both sexes | True | False |
| MonAbs | 8 | CCDC26 | lincRNA | rs13277237 | 130604563 | 2.14e-14 | 61 | 0 | 0 | 0 | 1 | 60 | Both sexes | Both sexes | Both sexes | True | False |
| MonAbs | 9 | RP11-202G18.1 | lincRNA | rs10980797 | 113912553 | 1.39e-12 | 29 | 0 | 0 | 0 | 4 | 25 | Both sexes | Both sexes | Both sexes | True | False |
| MonAbs | 3 | RPN1 | protein_coding | rs35604103 | 128334852 | 1.45e-11 | 173 | 0 | 0 | 0 | 0 | 173 | Both sexes | Both sexes | Both sexes | True | False |
| MonAbs | 16 | RP11-542M13.3 | lincRNA | rs523642 | 85965595 | 3.85e-11 | 37 | 0 | 0 | 4 | 8 | 25 | Female only | Both sexes | Female only | True | False |
| MonAbs | 2 | ITGA4 | protein_coding | rs1449260 | 182333447 | 7.81e-11 | 3 | 0 | 0 | 0 | 1 | 2 | Female only | Both sexes | Female only | True | False |
| MonAbs | 3 | RPN1 | protein_coding | rs2713573 | 128317273 | 4.92e-10 | 9 | 0 | 0 | 0 | 0 | 9 | Both sexes | Both sexes | Female only | True | False |
| MonAbs | 16 | RP11-542M13.3 | lincRNA | rs11642873 | 85991705 | 2.90e-09 | 7 | 0 | 0 | 0 | 0 | 7 | Female only | Both sexes | Female only | True | False |
| MonAbs | 9 | RP11-202G18.1 | lincRNA | rs9695374 | 114004409 | 3.98e-09 | 5 | 0 | 0 | 0 | 0 | 5 | Both sexes | Both sexes | Both sexes | True | False |
| MonAbs | 9 | RNY4P18 | misc_RNA | 9:113863709 | 113863709 | 5.56e-09 | 71 | 0 | 0 | 0 | 5 | 66 | Both sexes | Both sexes | Both sexes | True | False |
| MonAbs | 8 | CCDC26 | lincRNA | rs4259387 | 130611113 | 7.61e-09 | 33 | 0 | 4 | 13 | 0 | 16 | Female only | Both sexes | Female only | True | False |
| Monocy | 2 | ITGA4 | protein_coding | rs1375493 | 182323766 | 1.10e-16 | 15 | 0 | 0 | 0 | 0 | 15 | Both sexes | Both sexes | Both sexes | True | False |
| Monocy | 9 | RP11-202G18.1 | lincRNA | rs12339649 | 113945046 | 8.78e-16 | 51 | 0 | 0 | 0 | 0 | 51 | Both sexes | Both sexes | Female only | True | False |
| Monocy | 9 | RP11-202G18.1 | lincRNA | rs10980797 | 113912553 | 1.59e-15 | 29 | 0 | 0 | 0 | 4 | 25 | Both sexes | Both sexes | Both sexes | True | False |
| Monocy | 3 | C3orf27 | protein_coding | rs9849126 | 128312069 | 1.44e-14 | 61 | 0 | 0 | 0 | 0 | 61 | Both sexes | Both sexes | Both sexes | True | False |
| Monocy | 3 | RPN1 | protein_coding | rs9864772 | 128316939 | 2.47e-14 | 119 | 0 | 0 | 0 | 0 | 119 | Both sexes | Both sexes | Both sexes | True | False |
| Monocy | 16 | RP11-542M13.3 | lincRNA | rs76121846 | 85966548 | 5.47e-14 | 37 | 0 | 0 | 0 | 0 | 37 | Both sexes | Both sexes | Both sexes | True | False |
| Monocy | 8 | CCDC26 | lincRNA | rs13280084 | 130613499 | 1.25e-13 | 63 | 0 | 0 | 2 | 3 | 58 | Both sexes | Both sexes | Female only | True | False |
| Monocy | 16 | IRF8 | protein_coding | rs8064111 | 85947475 | 5.28e-13 | 35 | 0 | 4 | 1 | 0 | 30 | Both sexes | Both sexes | Both sexes | True | False |
| Monocy | 2 | ITGA4 | protein_coding | rs1449264 | 182336474 | 1.34e-11 | 3 | 2 | 0 | 0 | 0 | 1 | Female only | Female only | Female only | True | False |
| Monocy | 16 | IRF8 | protein_coding | rs11864884 | 85901039 | 9.74e-11 | 15 | 0 | 0 | 7 | 1 | 7 | Female only | Both sexes | Female only | True | False |
| Monocy | 20 | RP11-290F20.3 | lincRNA | rs6122894 | 48863091 | 1.21e-10 | 29 | 1 | 0 | 7 | 7 | 14 | Female only | Both sexes | Female only | True | False |
| Monocy | 3 | SNORA24 | snoRNA | rs2001950 | 128419756 | 1.53e-10 | 73 | 0 | 0 | 4 | 3 | 66 | Both sexes | Both sexes | Female only | True | False |
| Monocy | 3 | RPN1 | protein_coding | rs34890930 | 128329223 | 1.87e-10 | 11 | 0 | 0 | 0 | 0 | 11 | Both sexes | Both sexes | Female only | True | False |
| Monocy | 16 | IRF8 | protein_coding | rs8051462 | 85947327 | 3.39e-10 | 35 | 3 | 3 | 3 | 6 | 20 | Both sexes | Both sexes | Both sexes | True | False |
| Monocy | 3 | C3orf27 | protein_coding | rs2713587 | 128290848 | 3.53e-10 | 7 | 0 | 0 | 0 | 0 | 7 | Both sexes | Both sexes | Both sexes | True | False |

| pheno | CHR | Gene_Symbol | biotype | lead_snp | lead_BP | lead_P | TOTAL | NSIG | S05 | S01 | S001 | S0001 | independent_loci_in_sex_P1e6 | independent_loci_in_sex_P1e2 | independent_loci_in_sex_P5e8 | in_gwas_catalog | in_HLA |
|---|---|---|---|---|---|---|---|---|---|---|---|---|---|---|---|---|---|
| Monocy | 3 | C3orf27 | protein_coding | rs2712421 | 128287743 | 4.12e-10 | 3 | 0 | 0 | 0 | 0 | 3 | Female only | Both sexes | Female only | True | False |
| Monocy | 9 | RNY4P18 | misc_RNA | 9:113863709 | 113863709 | 1.16e-09 | 71 | 0 | 0 | 3 | 6 | 62 | Female only | Both sexes | Female only | True | False |
| Monocy | 2 | ITGA4 | protein_coding | rs155149 | 182329647 | 1.29e-09 | 3 | 0 | 0 | 0 | 0 | 3 | Female only | Both sexes | Female only | True | False |
| Monocy | 1 | RPAP2 | protein_coding | rs74767605 | 92770376 | 1.98e-09 | 7 | 4 | 0 | 0 | 0 | 3 | Female only | Female only | Female only | True | False |
| Monocy | 9 | RP11-202G18.1 | lincRNA | rs11790072 | 113936306 | 5.26e-09 | 97 | 2 | 12 | 7 | 34 | 42 | Female only | Both sexes | Female only | True | False |
| Monocy | 9 | RP11-202G18.1 | lincRNA | rs10117934 | 114015405 | 5.99e-09 | 5 | 0 | 0 | 0 | 0 | 5 | Both sexes | Both sexes | Female only | True | False |
| Monocy | 16 | RP11-542M13.3 | lincRNA | rs11642873 | 85991705 | 9.48e-09 | 7 | 0 | 0 | 0 | 0 | 7 | Both sexes | Both sexes | Both sexes | True | False |
| Monocy | 16 | RP11-542M13.3 | lincRNA | rs7202472 | 85977502 | 1.27e-08 | 21 | 0 | 0 | 10 | 10 | 1 | Both sexes | Both sexes | Female only | True | False |
| Monocy | 9 | RP11-202G18.1 | lincRNA | rs2026801 | 113905451 | 2.18e-08 | 3 | 2 | 0 | 0 | 0 | 1 | Female only | Both sexes | Female only | True | False |
| Monocy | 1 | EPHX4 | protein_coding | rs143792309 | 92505309 | 2.28e-08 | 5 | 3 | 0 | 0 | 0 | 2 | Female only | Female only | Female only | True | False |
| Monocy | 1 | EVI5 | protein_coding | 1:93262946 | 93262946 | 2.61e-08 | 463 | 231 | 1 | 0 | 0 | 231 | Female only | Female only | Female only | True | False |
| NEUTAB | 17 | PSMD3 | protein_coding | rs4794822 | 38156712 | 1.59e-16 | 153 | 0 | 0 | 0 | 0 | 153 | Both sexes | Both sexes | Both sexes | True | False |
| NEUTAB | 17 | CSF3 | protein_coding | rs25645 | 38173143 | 1.08e-14 | 83 | 0 | 0 | 0 | 0 | 83 | Both sexes | Both sexes | Both sexes | True | False |
| NEUTAB | 17 | GSDMA | protein_coding | rs62068180 | 38112825 | 2.59e-12 | 91 | 0 | 0 | 0 | 0 | 91 | Both sexes | Both sexes | Both sexes | True | False |
| NEUTAB | 4 | MTHFD2L | protein_coding | rs1893319 | 74973129 | 6.05e-12 | 25 | 0 | 0 | 4 | 8 | 13 | Female only | Both sexes | Female only | True | False |
| NEUTAB | 17 | ORMDL3 | protein_coding | rs8076131 | 38080912 | 3.01e-10 | 159 | 0 | 0 | 19 | 20 | 120 | Male only | Both sexes | Male only | True | False |
| NEUTAB | 2 | RN7SL51P | misc_RNA | rs72803397 | 62530258 | 5.27e-09 | 53 | 27 | 0 | 0 | 0 | 26 | Male only | Male only | Male only | True | False |
| NEUTAB | 17 | ZPBP2 | protein_coding | rs12150079 | 38025417 | 1.71e-08 | 7 | 0 | 0 | 3 | 2 | 2 | Male only | Both sexes | Male only | True | False |
| pheno | CHR | Gene_Symbol | biotype | lead_snp | lead_BP | lead_P | TOTAL | NSIG | S05 | S01 | S001 | S0001 | independent_loci_in_sex_P1e6 | independent_loci_in_sex_P1e2 | independent_loci_in_sex_P5e8 | in_gwas_catalog | in_HLA |
| NEUTM | 23 | MTND6P12 | pseudogene | X:5080916:T:C | 5080916 | 5.55e-09 | 141 | 71 | 7 | 35 | 6 | 22 | Female only | Female only | Female only | True | False |
| NEUTRE | 17 | PSMD3 | protein_coding | rs8070444 | 38151960 | 1.13e-09 | 117 | 0 | 0 | 0 | 1 | 116 | Both sexes | Both sexes | Both sexes | True | False |
| NEUTRE | 17 | MED24 | protein_coding | rs3936197 | 38194799 | 1.46e-08 | 19 | 0 | 0 | 0 | 10 | 9 | Male only | Both sexes | Male only | True | False |
| NEUTRE | 17 | RP11-387H17.6 | lincRNA | rs2227319 | 38170845 | 4.34e-08 | 85 | 0 | 1 | 1 | 8 | 75 | Female only | Both sexes | Female only | True | False |
| NTAuto | 17 | RP11-387H17.6 | lincRNA | rs2227315 | 38169714 | 2.57e-19 | 153 | 0 | 0 | 0 | 0 | 153 | Both sexes | Both sexes | Both sexes | True | False |
| NTAuto | 17 | RP11-387H17.6 | lincRNA | rs34933056 | 38162983 | 1.57e-18 | 83 | 0 | 0 | 0 | 0 | 83 | Both sexes | Both sexes | Both sexes | True | False |
| NTAuto | 17 | GSDMA | protein_coding | rs4458030 | 38121706 | 5.38e-12 | 81 | 0 | 0 | 0 | 0 | 81 | Both sexes | Both sexes | Both sexes | True | False |

| pheno | CHR | Gene_Symbol | biotype | lead_snp | lead_BP | lead_P | TOTAL | NSIG | S05 | S01 | S001 | S0001 | independent_loci_in_sex_P1e6 | independent_loci_in_sex_P1e2 | independent_loci_in_sex_P5e8 | in_gwas_catalog | in_HLA |
|---|---|---|---|---|---|---|---|---|---|---|---|---|---|---|---|---|---|
| NTAuto | 4 | MTHFD2L | protein_coding | rs10938107 | 74990919 | 1.91e-09 | 25 | 0 | 0 | 0 | 0 | 25 | Female only | Both sexes | Female only | True | False |
| NTAuto | 17 | GSDMA | protein_coding | rs8069202 | 38122200 | 7.31e-09 | 45 | 0 | 0 | 5 | 24 | 16 | Female only | Both sexes | Female only | True | False |
| Na | 20 | PREX1 | protein_coding | rs34316626 | 47358484 | 1.46e-08 | 9 | 5 | 0 | 0 | 0 | 4 | Female only | Female only | Female only | True | False |
| Na | 20 | PREX1 | protein_coding | rs13044684 | 47454542 | 1.98e-08 | 85 | 42 | 1 | 0 | 0 | 42 | Female only | Female only | Female only | True | False |
| NtAbs | 17 | MED24 | protein_coding | rs1045929 | 38175426 | 1.36e-12 | 153 | 0 | 0 | 0 | 7 | 146 | Both sexes | Both sexes | Both sexes | True | False |
| NtAbs | 17 | PSMD3 | protein_coding | rs11078931 | 38147294 | 1.74e-10 | 83 | 0 | 0 | 0 | 1 | 82 | Both sexes | Both sexes | Both sexes | True | False |
| NtAbs | 17 | GSDMA | protein_coding | rs7218321 | 38114469 | 4.91e-09 | 91 | 0 | 5 | 9 | 8 | 69 | Female only | Both sexes | Female only | True | False |
| O2SAT | 17 | CDK5RAP3 | protein_coding | rs17617053 | 46050669 | 2.31e-08 | 5 | 3 | 0 | 0 | 0 | 2 | Female only | Female only | Female only | True | False |
| PCAct | 20 | EDEM2 | protein_coding | rs8119351 | 33754405 | 8.72e-09 | 317 | 23 | 136 | 2 | 6 | 150 | Female only | Female only | Female only | False | False |
| PCV | 6 | HBS1L | protein_coding | rs9402686 | 135427817 | 1.89e-10 | 37 | 0 | 0 | 0 | 1 | 36 | Both sexes | Both sexes | Female only | True | False |
| PCV | 10 | HK1 | protein_coding | rs17476364 | 71094504 | 2.22e-10 | 11 | 0 | 0 | 0 | 5 | 6 | Female only | Both sexes | Female only | True | False |
| PCV | 6 | MYB | protein_coding | rs62429816 | 135486022 | 4.19e-08 | 37 | 4 | 0 | 1 | 15 | 17 | Female only | Both sexes | Female only | True | False |
| PCV | 22 | TMPRSS6 | protein_coding | rs4820268 | 37469591 | 4.82e-08 | 9 | 0 | 1 | 0 | 3 | 5 | Female only | Both sexes | Female only | True | False |
| PR.INTERVAL | 3 | SCN10A | protein_coding | rs7433723 | 38784957 | 6.16e-12 | 51 | 0 | 0 | 0 | 9 | 42 | Both sexes | Both sexes | Male only | True | False |
| PR.INTERVAL | 3 | SCN10A | protein_coding | rs12638572 | 38787797 | 1.28e-08 | 79 | 13 | 11 | 17 | 16 | 22 | Male only | Both sexes | Male only | True | False |
| PR.INTERVAL | 3 | SCN5A | protein_coding | rs3922843 | 38624343 | 1.29e-08 | 25 | 0 | 1 | 12 | 0 | 12 | Female only | Both sexes | Female only | True | False |
| PR_INTERVAL | 3 | SCN10A | protein_coding | rs6801957 | 38767315 | 4.21e-32 | 51 | 0 | 0 | 0 | 0 | 51 | Both sexes | Both sexes | Both sexes | True | False |
| PR_INTERVAL | 3 | SCN5A | protein_coding | rs3922843 | 38624343 | 1.07e-29 | 25 | 0 | 0 | 0 | 0 | 25 | Both sexes | Both sexes | Both sexes | True | False |
| PR_INTERVAL | 3 | SCN5A | protein_coding | rs73070977 | 38631431 | 1.41e-22 | 7 | 0 | 0 | 2 | 2 | 3 | Both sexes | Both sexes | Both sexes | True | False |
| PR_INTERVAL | 7 | CAV1 | protein_coding | rs3807989 | 116186241 | 2.38e-22 | 27 | 0 | 0 | 0 | 0 | 27 | Both sexes | Both sexes | Both sexes | True | False |
| PR_INTERVAL | 3 | SCN5A | protein_coding | rs3924120 | 38636155 | 3.57e-17 | 9 | 0 | 0 | 0 | 0 | 9 | Both sexes | Both sexes | Both sexes | True | False |
| PR_INTERVAL | 3 | SCN10A | protein_coding | rs6599240 | 38738717 | 7.34e-17 | 3 | 0 | 0 | 0 | 0 | 3 | Both sexes | Both sexes | Both sexes | True | False |

| pheno | CHR | Gene_Symbol | biotype | lead_snp | lead_BP | lead_P | TOTAL | NSIG | S05 | S01 | S001 | S0001 | independent_loci_in_sex_P1e6 | independent_loci_in_sex_P1e2 | independent_loci_in_sex_P5e8 | in_gwas_catalog | in_HLA |
|---|---|---|---|---|---|---|---|---|---|---|---|---|---|---|---|---|---|
| PR_INTERVAL | 3 | SCN10A | protein_coding | rs6784303 | 38794915 | 1.88e-16 | 79 | 0 | 0 | 0 | 0 | 79 | Both sexes | Both sexes | Both sexes | True | False |
| PR_INTERVAL | 3 | SCN5A | protein_coding | rs9311193 | 38634659 | 7.68e-16 | 7 | 0 | 0 | 0 | 0 | 7 | Both sexes | Both sexes | Both sexes | True | False |
| PR_INTERVAL | 3 | SCN5A | protein_coding | rs9845438 | 38600456 | 9.58e-16 | 21 | 0 | 0 | 0 | 0 | 21 | Female only | Both sexes | Female only | True | False |
| PR_INTERVAL | 7 | CAV2 | protein_coding | rs76160518 | 116099441 | 1.43e-15 | 69 | 0 | 0 | 4 | 10 | 55 | Both sexes | Both sexes | Female only | True | False |
| PR_INTERVAL | 3 | SCN10A | protein_coding | rs7430439 | 38803639 | 2.09e-15 | 3 | 0 | 0 | 0 | 0 | 3 | Both sexes | Both sexes | Both sexes | True | False |
| PR_INTERVAL | 7 | CAV1 | protein_coding | rs9886216 | 116191697 | 2.49e-14 | 61 | 0 | 0 | 4 | 5 | 52 | Both sexes | Both sexes | Female only | True | False |
| PR_INTERVAL | 3 | SCN5A | protein_coding | rs9812912 | 38607229 | 2.54e-14 | 3 | 0 | 0 | 0 | 0 | 3 | Female only | Both sexes | Female only | True | False |
| PR_INTERVAL | 2 | MEIS1 | protein_coding | rs9798015 | 66768262 | 4.24e-14 | 69 | 0 | 17 | 18 | 0 | 34 | Female only | Both sexes | Female only | True | False |
| PR_INTERVAL | 4 | ARHGAP24 | protein_coding | rs11724267 | 86677076 | 4.51e-14 | 69 | 0 | 0 | 0 | 0 | 69 | Both sexes | Both sexes | Both sexes | True | False |
| PR_INTERVAL | 3 | SCN5A | protein_coding | rs9856387 | 38687226 | 4.32e-13 | 11 | 0 | 0 | 0 | 0 | 11 | Both sexes | Both sexes | Both sexes | True | False |
| PR_INTERVAL | 12 | RP11-615I16.1 | pseudogene | rs17287293 | 24770878 | 1.32e-12 | 29 | 13 | 2 | 0 | 0 | 14 | Female only | Female only | Female only | True | False |
| PR_INTERVAL | 3 | SCN5A | protein_coding | rs55742514 | 38687740 | 4.09e-12 | 13 | 0 | 0 | 0 | 0 | 13 | Both sexes | Both sexes | Both sexes | True | False |
| PR_INTERVAL | 3 | SCN10A | protein_coding | rs7611456 | 38813465 | 4.73e-12 | 3 | 0 | 0 | 0 | 0 | 3 | Both sexes | Both sexes | Both sexes | True | False |
| PR_INTERVAL | 3 | SCN5A | protein_coding | rs34519218 | 38624842 | 2.01e-11 | 25 | 2 | 5 | 5 | 0 | 13 | Both sexes | Both sexes | Female only | True | False |
| PR_INTERVAL | 12 | TBX5 | protein_coding | rs883079 | 114793240 | 3.13e-11 | 45 | 0 | 0 | 0 | 0 | 45 | Both sexes | Both sexes | Both sexes | True | False |
| PR_INTERVAL | 3 | EXOG | protein_coding | rs62241772 | 38551125 | 2.39e-10 | 13 | 4 | 0 | 3 | 4 | 2 | Female only | Both sexes | Female only | True | False |
| PR_INTERVAL | 3 | SCN5A | protein_coding | rs6790619 | 38623690 | 4.57e-10 | 13 | 4 | 0 | 3 | 0 | 6 | Female only | Both sexes | Female only | True | False |
| PR_INTERVAL | 12 | TBX5 | protein_coding | rs1895596 | 114791528 | 8.76e-10 | 39 | 0 | 0 | 0 | 1 | 38 | Both sexes | Both sexes | Both sexes | True | False |

| pheno | CHR | Gene_Symbol | biotype | lead_snp | lead_BP | lead_P | TOTAL | NSIG | S05 | S01 | S001 | S0001 | independent_loci_in_sex_P1e6 | independent_loci_in_sex_P1e2 | independent_loci_in_sex_P5e8 | in_gwas_catalog | in_HLA |
|---|---|---|---|---|---|---|---|---|---|---|---|---|---|---|---|---|---|
| PR_INTERVAL | 7 | CAV1 | protein_coding | rs3807986 | 116177825 | 9.01e-10 | 51 | 0 | 22 | 11 | 6 | 12 | Female only | Both sexes | Female only | True | False |
| PR_INTERVAL | 7 | CAV2 | protein_coding | rs55701446 | 116150077 | 1.23e-09 | 13 | 0 | 0 | 0 | 5 | 8 | Female only | Both sexes | Female only | True | False |
| PR_INTERVAL | 7 | CAV2 | protein_coding | rs11760323 | 116003062 | 1.34e-09 | 19 | 2 | 5 | 4 | 0 | 8 | Female only | Both sexes | Female only | True | False |
| PR_INTERVAL | 3 | SCN5A | protein_coding | rs6772948 | 38615316 | 1.62e-09 | 5 | 0 | 0 | 1 | 4 | 0 | Female only | Both sexes | Female only | True | False |
| PR_INTERVAL | 10 | SH3PXD2A | protein_coding | rs11191795 | 105516579 | 3.68e-09 | 31 | 0 | 7 | 1 | 13 | 10 | Male only | Both sexes | Male only | True | False |
| PR_INTERVAL | 3 | SCN10A | protein_coding | rs10428168 | 38780059 | 4.25e-09 | 29 | 1 | 1 | 1 | 7 | 19 | Both sexes | Both sexes | Female only | True | False |
| PR_INTERVAL | 3 | SCN10A | protein_coding | rs11129802 | 38750436 | 5.64e-09 | 13 | 0 | 0 | 7 | 0 | 6 | Female only | Both sexes | Female only | True | False |
| PR_INTERVAL | 3 | SCN10A | protein_coding | rs62243862 | 38741549 | 1.02e-08 | 5 | 0 | 0 | 0 | 0 | 5 | Both sexes | Both sexes | Female only | True | False |
| PR_INTERVAL | 1 | C1orf86 | protein_coding | rs2503715 | 2144107 | 1.08e-08 | 3 | 0 | 0 | 0 | 0 | 3 | Both sexes | Both sexes | Male only | True | False |
| PR_INTERVAL | 6 | PRRC2A | protein_coding | rs1046080 | 31595882 | 1.60e-08 | 21 | 5 | 6 | 0 | 5 | 5 | Female only | Female only | Female only | True | True |
| PR_INTERVAL | 12 | TBX5 | protein_coding | rs1896002 | 114787365 | 2.63e-08 | 9 | 0 | 0 | 0 | 4 | 5 | Female only | Both sexes | Female only | True | False |
| PR_INTERVAL | 3 | SCN5A | protein_coding | rs6768664 | 38684466 | 4.78e-08 | 11 | 0 | 0 | 0 | 1 | 10 | Female only | Both sexes | Female only | True | False |
| PSA | 19 | KLK3 | protein_coding | rs62113214 | 51362537 | 2.23e-12 | 4 | 0 | 0 | 0 | 0 | 4 | GWAS only run in males | GWAS only run in males | GWAS only run in males | True | False |

| pheno | CHR | Gene_Symbol | biotype | lead_snp | lead_BP | lead_P | TOTAL | NSIG | S05 | S01 | S001 | S0001 | independent_loci_in_sex_P1e6 | independent_loci_in_sex_P1e2 | independent_loci_in_sex_P5e8 | in_gwas_catalog | in_HLA |
|---|---|---|---|---|---|---|---|---|---|---|---|---|---|---|---|---|---|
| PSA | 19 | AC011523.2 | antisense | rs7251465 | 51350791 | 4.03e-10 | 3 | 0 | 0 | 0 | 0 | 3 | GWAS only run in males | GWAS only run in males | GWAS only run in males | True | False |
| PSA | 19 | AC011523.2 | antisense | rs11665748 | 51354397 | 1.09e-09 | 2 | 0 | 0 | 0 | 1 | 1 | GWAS only run in males | GWAS only run in males | GWAS only run in males | True | False |
| PSA | 19 | KLK3 | protein_coding | rs1058205 | 51363398 | 4.31e-08 | 3 | 0 | 0 | 0 | 1 | 2 | GWAS only run in males | GWAS only run in males | GWAS only run in males | True | False |
| PT.inr | 13 | F7 | protein_coding | rs561241 | 113760034 | 5.36e-26 | 37 | 0 | 0 | 0 | 0 | 37 | Both sexes | Both sexes | Both sexes | True | False |
| PT.inr | 13 | F10 | protein_coding | rs2480946 | 113784720 | 7.53e-17 | 9 | 0 | 0 | 0 | 0 | 9 | Both sexes | Both sexes | Both sexes | True | False |
| PT.inr | 20 | EDEM2 | protein_coding | rs11908683 | 33720920 | 4.87e-11 | 323 | 0 | 0 | 2 | 18 | 303 | Both sexes | Both sexes | Female only | True | False |

| pheno | CHR | Gene_Symbol | biotype | lead_snp | lead_BP | lead_P | TOTAL | NSIG | S05 | S01 | S001 | S0001 | independent_loci_in_sex_P1e6 | independent_loci_in_sex_P1e2 | independent_loci_in_sex_P5e8 | in_gwas_catalog | in_HLA |
|---|---|---|---|---|---|---|---|---|---|---|---|---|---|---|---|---|---|
| PT.inr | 13 | MCF2L | protein_coding | rs11841838 | 113748552 | 2.22e-08 | 11 | 0 | 0 | 0 | 0 | 11 | Both sexes | Both sexes | Male only | True | False |
| PTT.pt | 5 | F12 | protein_coding | rs1801020 | 176836532 | 2.75e-79 | 7 | 0 | 0 | 0 | 0 | 7 | Both sexes | Both sexes | Both sexes | True | False |
| PTT.pt | 3 | KNG1 | protein_coding | rs710446 | 186459927 | 1.53e-39 | 29 | 0 | 0 | 0 | 0 | 29 | Both sexes | Both sexes | Both sexes | True | False |
| PTT.pt | 3 | RP11-795A2.3 | pseudogene | rs56380201 | 186429367 | 9.97e-21 | 9 | 0 | 0 | 0 | 0 | 9 | Both sexes | Both sexes | Both sexes | True | False |
| PTT.pt | 5 | SLC34A1 | protein_coding | rs6556316 | 176821375 | 3.12e-18 | 11 | 0 | 0 | 0 | 0 | 11 | Both sexes | Both sexes | Both sexes | True | False |
| PTT.pt | 3 | RP11-573D15.8 | antisense | rs822373 | 186471186 | 1.02e-17 | 7 | 0 | 0 | 0 | 0 | 7 | Both sexes | Both sexes | Both sexes | True | False |
| PTT.pt | 3 | KNG1 | protein_coding | rs166479 | 186443250 | 7.28e-16 | 17 | 0 | 0 | 0 | 0 | 17 | Both sexes | Both sexes | Both sexes | True | False |
| PTT.pt | 3 | RP11-134F2.2 | antisense | rs6803759 | 186371243 | 6.17e-15 | 61 | 0 | 0 | 3 | 1 | 57 | Both sexes | Both sexes | Both sexes | True | False |
| PTT.pt | 9 | ABO | processed_transcript | rs514659 | 136142203 | 5.90e-13 | 51 | 0 | 0 | 0 | 2 | 49 | Female only | Both sexes | Female only | True | False |
| PTT.pt | 5 | MXD3 | protein_coding | rs111251222 | 176735612 | 1.55e-12 | 11 | 0 | 0 | 0 | 0 | 11 | Both sexes | Both sexes | Both sexes | True | False |
| PTT.pt | 3 | RP11-573D15.8 | antisense | rs2378065 | 186468856 | 1.72e-12 | 5 | 0 | 0 | 0 | 0 | 5 | Both sexes | Both sexes | Both sexes | True | False |
| PTT.pt | 5 | RAB24 | protein_coding | rs1128287 | 176728458 | 4.64e-12 | 7 | 0 | 0 | 0 | 0 | 7 | Both sexes | Both sexes | Both sexes | True | False |
| PTT.pt | 5 | LMAN2 | protein_coding | rs35630910 | 176757841 | 9.84e-12 | 47 | 0 | 0 | 0 | 17 | 30 | Female only | Both sexes | Female only | True | False |
| PTT.pt | 5 | NSD1 | protein_coding | rs2878686 | 176633749 | 2.22e-11 | 121 | 1 | 8 | 37 | 15 | 60 | Female only | Both sexes | Female only | True | False |
| PTT.pt | 3 | HRG | protein_coding | rs1863622 | 186396616 | 7.12e-11 | 19 | 0 | 0 | 0 | 0 | 19 | Both sexes | Both sexes | Both sexes | True | False |
| PTT.pt | 9 | ABO | processed_transcript | rs2519093 | 136141870 | 1.51e-10 | 19 | 0 | 0 | 5 | 4 | 10 | Female only | Both sexes | Female only | True | False |
| PTT.pt | 3 | KNG1 | protein_coding | rs710449 | 186452820 | 6.73e-10 | 11 | 0 | 0 | 0 | 1 | 10 | Both sexes | Both sexes | Male only | True | False |
| PTT.pt | 3 | RP11-795A2.3 | pseudogene | rs10937264 | 186428197 | 1.85e-09 | 37 | 0 | 0 | 0 | 22 | 15 | Both sexes | Both sexes | Both sexes | True | False |
| PTT.pt | 5 | RGS14 | protein_coding | rs73336260 | 176788293 | 1.94e-09 | 15 | 0 | 0 | 0 | 7 | 8 | Female only | Both sexes | Female only | True | False |
| PTT.pt | 9 | ABO | processed_transcript | rs630014 | 136149722 | 1.48e-08 | 7 | 0 | 0 | 4 | 0 | 3 | Female only | Both sexes | Female only | True | False |
| PTT.pt | 9 | ABO | processed_transcript | rs633862 | 136155444 | 1.90e-08 | 31 | 6 | 1 | 6 | 10 | 8 | Female only | Both sexes | Female only | True | False |
| PTT.pt | 4 | F11 | protein_coding | rs4253399 | 187188094 | 2.71e-08 | 15 | 0 | 0 | 5 | 6 | 4 | Female only | Both sexes | Female only | True | False |
| PULSE | 6 | HMGB3P18 | pseudogene | rs2012952 | 122124077 | 7.41e-11 | 125 | 0 | 0 | 3 | 29 | 93 | Female only | Both sexes | Female only | True | False |
| PULSE | 14 | MYH6 | protein_coding | rs422068 | 23864804 | 4.09e-08 | 37 | 9 | 10 | 0 | 0 | 18 | Female only | Female only | Female only | True | False |
| Pat.PT | 13 | F7 | protein_coding | rs3093253 | 113774092 | 1.74e-31 | 37 | 0 | 0 | 0 | 0 | 37 | Both sexes | Both sexes | Both sexes | True | False |
| Pat.PT | 13 | F10 | protein_coding | rs2480946 | 113784720 | 2.25e-20 | 9 | 0 | 0 | 0 | 0 | 9 | Both sexes | Both sexes | Both sexes | True | False |

| pheno | CHR | Gene_Symbol | biotype | lead_snp | lead_BP | lead_P | TOTAL | NSIG | S05 | S01 | S001 | S0001 | independent_loci_in_sex_P1e6 | independent_loci_in_sex_P1e2 | independent_loci_in_sex_P5e8 | in_gwas_catalog | in_HLA |
|---|---|---|---|---|---|---|---|---|---|---|---|---|---|---|---|---|---|
| Pat.PT | 13 | MCF2L | protein_coding | rs11839131 | 113748379 | 1.14e-11 | 11 | 0 | 0 | 0 | 0 | 11 | Both sexes | Both sexes | Both sexes | True | False |
| Pat.PT | 20 | EDEM2 | protein_coding | rs11907010 | 33737661 | 4.97e-11 | 317 | 0 | 0 | 2 | 8 | 307 | Both sexes | Both sexes | Both sexes | True | False |
| Pat.PT | 1 | F5 | protein_coding | rs72708008 | 169478105 | 9.40e-10 | 99 | 0 | 0 | 0 | 4 | 95 | Both sexes | Both sexes | Female only | True | False |
| Pat.PT | 13 | AL137002.1 | protein_coding | rs555212 | 113756540 | 1.20e-09 | 7 | 1 | 0 | 1 | 3 | 2 | Female only | Both sexes | Female only | True | False |
| Plt.Ct | 3 | ARHGEF3 | protein_coding | rs1354034 | 56849749 | 1.69e-25 | 19 | 0 | 0 | 0 | 0 | 19 | Both sexes | Both sexes | Both sexes | True | False |
| Plt.Ct | 9 | RP11-307I14.4 | pseudogene | rs385893 | 4763176 | 4.52e-23 | 3 | 0 | 0 | 0 | 0 | 3 | Both sexes | Both sexes | Both sexes | True | False |
| Plt.Ct | 6 | BAK1 | protein_coding | rs210140 | 33544293 | 7.05e-20 | 13 | 0 | 0 | 0 | 0 | 13 | Both sexes | Both sexes | Both sexes | True | False |
| Plt.Ct | 6 | BAK1 | protein_coding | rs511515 | 33541507 | 1.65e-19 | 7 | 0 | 0 | 0 | 0 | 7 | Both sexes | Both sexes | Both sexes | True | False |
| Plt.Ct | 9 | AK3 | protein_coding | rs409801 | 4744743 | 1.27e-17 | 3 | 0 | 0 | 0 | 0 | 3 | Both sexes | Both sexes | Both sexes | True | False |
| Plt.Ct | 6 | HBS1L | protein_coding | rs34164109 | 135421176 | 2.67e-17 | 35 | 0 | 0 | 0 | 0 | 35 | Both sexes | Both sexes | Both sexes | True | False |
| Plt.Ct | 6 | BAK1 | protein_coding | rs9296095 | 33542523 | 6.28e-17 | 63 | 0 | 0 | 9 | 11 | 43 | Both sexes | Both sexes | Both sexes | True | False |
| Plt.Ct | 22 | PNPLA3 | protein_coding | rs3747207 | 44324855 | 4.13e-15 | 113 | 0 | 0 | 0 | 0 | 113 | Both sexes | Both sexes | Both sexes | True | False |
| Plt.Ct | 9 | RP11-307I14.4 | pseudogene | rs13285743 | 4771638 | 2.42e-14 | 5 | 0 | 0 | 0 | 0 | 5 | Both sexes | Both sexes | Female only | True | False |
| Plt.Ct | 9 | RP11-307I14.4 | pseudogene | rs371506 | 4758794 | 3.14e-14 | 33 | 0 | 0 | 0 | 0 | 33 | Both sexes | Both sexes | Both sexes | True | False |
| Plt.Ct | 9 | RP11-307I14.4 | pseudogene | rs1756875 | 4770942 | 4.11e-13 | 5 | 0 | 0 | 0 | 0 | 5 | Both sexes | Both sexes | Female only | True | False |
| Plt.Ct | 9 | RCL1 | protein_coding | rs460182 | 4850613 | 1.44e-12 | 65 | 0 | 7 | 21 | 0 | 37 | Female only | Both sexes | Female only | True | False |
| Plt.Ct | 12 | BRAP | protein_coding | rs11065979 | 112059557 | 2.42e-12 | 7 | 0 | 0 | 4 | 0 | 3 | Female only | Both sexes | Female only | True | False |
| Plt.Ct | 9 | RCL1 | protein_coding | rs17803780 | 4849647 | 2.48e-12 | 41 | 0 | 0 | 3 | 8 | 30 | Female only | Both sexes | Female only | True | False |
| Plt.Ct | 8 | ZFPM2 | protein_coding | rs7832219 | 106578977 | 2.20e-11 | 63 | 22 | 1 | 5 | 6 | 29 | Female only | Both sexes | Female only | True | False |
| Plt.Ct | 3 | EIF2B5 | protein_coding | rs6141 | 184090266 | 3.16e-11 | 5 | 2 | 1 | 0 | 0 | 2 | Female only | Female only | Female only | True | False |
| Plt.Ct | 9 | RP11-307I14.4 | pseudogene | rs10758649 | 4773080 | 3.95e-11 | 13 | 1 | 4 | 0 | 3 | 5 | Female only | Both sexes | Female only | True | False |
| Plt.Ct | 6 | MYB | protein_coding | rs62429811 | 135480054 | 4.07e-11 | 37 | 0 | 0 | 0 | 0 | 37 | Both sexes | Both sexes | Both sexes | True | False |
| Plt.Ct | 9 | CDKN2B-AS1 | antisense | rs6475611 | 22151139 | 1.66e-10 | 21 | 2 | 8 | 1 | 0 | 10 | Female only | Both sexes | Female only | True | False |
| Plt.Ct | 3 | ARHGEF3 | protein_coding | rs12488986 | 56850188 | 2.03e-10 | 11 | 4 | 0 | 2 | 0 | 5 | Female only | Both sexes | Female only | True | False |
| Plt.Ct | 3 | ARHGEF3 | protein_coding | rs13061631 | 56862390 | 2.50e-10 | 7 | 0 | 1 | 3 | 0 | 3 | Female only | Both sexes | Female only | True | False |
| Plt.Ct | 9 | RCL1 | protein_coding | rs296847 | 4793798 | 2.75e-10 | 5 | 0 | 0 | 0 | 0 | 5 | Female only | Both sexes | Female only | True | False |

| pheno | CHR | Gene_Symbol | biotype | lead_snp | lead_BP | lead_P | TOTAL | NSIG | S05 | S01 | S001 | S0001 | independent_loci_in_sex_P1e6 | independent_loci_in_sex_P1e2 | independent_loci_in_sex_P5e8 | in_gwas_catalog | in_HLA |
|---|---|---|---|---|---|---|---|---|---|---|---|---|---|---|---|---|---|
| Plt.Ct | 10 | JMJD1C | protein_coding | rs10822153 | 65056813 | 7.80e-10 | 165 | 0 | 0 | 74 | 8 | 83 | Female only | Both sexes | Female only | True | False |
| Plt.Ct | 12 | TRAFD1 | protein_coding | rs17630235 | 112591686 | 1.48e-09 | 5 | 0 | 0 | 3 | 0 | 2 | Female only | Both sexes | Female only | True | False |

| pheno | CHR | Gene_Symbol | biotype | lead_snp | lead_BP | lead_P | TOTAL | NSIG | S05 | S01 | S001 | S0001 | independent_loci_in_sex_P1e6 | independent_loci_in_sex_P1e2 | independent_loci_in_sex_P5e8 | in_gwas_catalog | in_HLA |
|---|---|---|---|---|---|---|---|---|---|---|---|---|---|---|---|---|---|
| Plt.Ct | 11 | BET1L | protein_coding | rs4980320 | 198986 | 1.48e-09 | 7 | 0 | 3 | 1 | 0 | 3 | Female only | Both sexes | Female only | True | False |
| Plt.Ct | 10 | REEP3 | protein_coding | rs10740134 | 65315433 | 1.86e-09 | 55 | 0 | 0 | 28 | 0 | 27 | Female only | Both sexes | Female only | True | False |
| Plt.Ct | 10 | RP11-351O1.3 | pseudogene | rs2893923 | 65261184 | 2.28e-09 | 3 | 0 | 0 | 0 | 2 | 1 | Female only | Both sexes | Female only | True | False |
| Plt.Ct | 10 | JMJD1C | protein_coding | rs10995455 | 64936361 | 2.36e-09 | 3 | 0 | 0 | 0 | 3 | 0 | Female only | Both sexes | Female only | True | False |
| Plt.Ct | 20 | SIRPA | protein_coding | rs4814779 | 1923271 | 2.48e-09 | 35 | 0 | 0 | 2 | 9 | 24 | Female only | Both sexes | Female only | True | False |
| Plt.Ct | 3 | ARHGEF3 | protein_coding | rs55709294 | 56874996 | 4.09e-09 | 13 | 0 | 0 | 0 | 0 | 13 | Female only | Both sexes | Female only | True | False |
| Plt.Ct | 12 | USP30 | protein_coding | rs10850589 | 109499490 | 4.48e-09 | 17 | 0 | 0 | 0 | 3 | 14 | Female only | Both sexes | Female only | True | False |
| Plt.Ct | 11 | PSMD13 | protein_coding | rs17655188 | 249408 | 5.30e-09 | 87 | 0 | 1 | 43 | 0 | 43 | Female only | Both sexes | Female only | True | False |
| Plt.Ct | 15 | AC069368.3 | protein_coding | rs2013555 | 65170949 | 1.23e-08 | 65 | 20 | 11 | 2 | 2 | 30 | Female only | Both sexes | Female only | True | False |
| Plt.Ct | 6 | BAK1 | protein_coding | rs210129 | 33533341 | 1.55e-08 | 3 | 0 | 0 | 1 | 1 | 1 | Female only | Both sexes | Female only | True | False |
| Plt.Ct | 13 | ABCC4 | protein_coding | rs4148448 | 95897190 | 1.60e-08 | 19 | 0 | 0 | 1 | 7 | 11 | Female only | Both sexes | Female only | True | False |
| Plt.Ct | 19 | PTPRH | protein_coding | rs2288419 | 55693244 | 2.45e-08 | 21 | 10 | 1 | 1 | 0 | 9 | Female only | Female only | Female only | True | False |
| Plt.Ct | 6 | ITPR3 | protein_coding | rs10947421 | 33611278 | 4.51e-08 | 31 | 4 | 9 | 4 | 3 | 11 | Female only | Both sexes | Female only | True | False |
| Pulse | 14 | MYH6 | protein_coding | rs422068 | 23864804 | 1.83e-18 | 37 | 0 | 0 | 1 | 13 | 23 | Both sexes | Both sexes | Female only | True | False |
| Pulse | 6 | HMGB3P18 | pseudogene | rs763076 | 122148562 | 1.74e-15 | 125 | 0 | 0 | 0 | 0 | 125 | Both sexes | Both sexes | Both sexes | True | False |
| Pulse | 14 | MYH6 | protein_coding | rs439735 | 23868285 | 2.80e-09 | 5 | 0 | 3 | 0 | 0 | 2 | Female only | Female only | Female only | True | False |
| Pulse | 6 | HMGB3P18 | pseudogene | rs2090825 | 122271919 | 7.45e-09 | 129 | 11 | 39 | 2 | 1 | 76 | Female only | Both sexes | Female only | True | False |
| QRS.DURATION | 6 | CEP85L | protein_coding | rs55892056 | 118995297 | 2.12e-08 | 223 | 112 | 0 | 0 | 0 | 111 | Female only | Female only | Female only | True | False |
| QRS.DURATION | 6 | RP11-57K17.1 | pseudogene | rs17226667 | 118721628 | 2.61e-08 | 111 | 56 | 0 | 0 | 5 | 50 | Female only | Female only | Female only | True | False |
| QRS_DURATION | 6 | SLC35F1 | protein_coding | rs11153730 | 118667522 | 1.68e-18 | 261 | 10 | 10 | 65 | 35 | 141 | Female only | Both sexes | Female only | True | False |

| pheno | CHR | Gene_Symbol | biotype | lead_snp | lead_BP | lead_P | TOTAL | NSIG | S05 | S01 | S001 | S0001 | independent_loci_in_sex_P1e6 | independent_loci_in_sex_P1e2 | independent_loci_in_sex_P5e8 | in_gwas_catalog | in_HLA |
|---|---|---|---|---|---|---|---|---|---|---|---|---|---|---|---|---|---|
| QRS_DURATION | 6 | CEP85L | protein_coding | rs11153768 | 118988152 | 1.55e-17 | 71 | 0 | 7 | 22 | 7 | 35 | Female only | Both sexes | Female only | True | False |
| QRS_DURATION | 3 | SCN5A | protein_coding | rs73070977 | 38631431 | 9.56e-17 | 7 | 0 | 0 | 2 | 0 | 5 | Female only | Both sexes | Female only | True | False |
| QRS_DURATION | 3 | SCN10A | protein_coding | rs6801957 | 38767315 | 1.59e-16 | 51 | 0 | 0 | 3 | 0 | 48 | Female only | Both sexes | Female only | True | False |
| QRS_DURATION | 6 | SLC35F1 | protein_coding | rs4946333 | 118565665 | 4.42e-16 | 63 | 0 | 0 | 7 | 23 | 33 | Female only | Both sexes | Female only | True | False |
| QRS_DURATION | 3 | SCN5A | protein_coding | rs3922844 | 38624253 | 4.46e-16 | 13 | 0 | 3 | 4 | 0 | 6 | Female only | Both sexes | Female only | True | False |
| QRS_DURATION | 6 | CDKN1A | protein_coding | rs3176326 | 36647289 | 5.50e-16 | 35 | 0 | 0 | 0 | 1 | 34 | Both sexes | Both sexes | Both sexes | True | False |
| QRS_DURATION | 3 | SCN5A | protein_coding | rs7355944 | 38626193 | 2.02e-15 | 19 | 0 | 1 | 9 | 0 | 9 | Female only | Both sexes | Female only | True | False |
| QRS_DURATION | 3 | EXOG | protein_coding | rs6762565 | 38582191 | 4.82e-15 | 47 | 0 | 0 | 0 | 0 | 47 | Both sexes | Both sexes | Both sexes | True | False |
| QRS_DURATION | 18 | SETBP1 | protein_coding | rs11661654 | 42441443 | 5.32e-14 | 27 | 5 | 2 | 1 | 6 | 13 | Male only | Both sexes | Male only | True | False |
| QRS_DURATION | 3 | SCN5A | protein_coding | rs9845438 | 38600456 | 3.05e-12 | 3 | 0 | 0 | 0 | 0 | 3 | Both sexes | Both sexes | Female only | True | False |
| QRS_DURATION | 6 | CEP85L | protein_coding | rs9481842 | 118974798 | 4.25e-12 | 173 | 0 | 6 | 81 | 0 | 86 | Female only | Both sexes | Female only | True | False |
| QRS_DURATION | 6 | SLC35F1 | protein_coding | rs4945621 | 118658256 | 2.67e-11 | 51 | 0 | 4 | 14 | 7 | 26 | Female only | Both sexes | Female only | True | False |
| QRS_DURATION | 3 | SCN5A | protein_coding | rs4130467 | 38611704 | 7.22e-11 | 7 | 0 | 0 | 0 | 1 | 6 | Female only | Both sexes | Female only | True | False |

| pheno | CHR | Gene_Symbol | biotype | lead_snp | lead_BP | lead_P | TOTAL | NSIG | S05 | S01 | S001 | S0001 | independent_loci_in_sex_P1e6 | independent_loci_in_sex_P1e2 | independent_loci_in_sex_P5e8 | in_gwas_catalog | in_HLA |
|---|---|---|---|---|---|---|---|---|---|---|---|---|---|---|---|---|---|
| QRS_DURATION | 3 | SCN5A | protein_coding | rs9812912 | 38607229 | 9.82e-11 | 3 | 0 | 2 | 0 | 0 | 1 | Female only | Female only | Female only | True | False |
| QRS_DURATION | 3 | SCN5A | protein_coding | rs9880327 | 38682893 | 1.13e-10 | 11 | 0 | 0 | 0 | 0 | 11 | Both sexes | Both sexes | Both sexes | True | False |
| QRS_DURATION | 6 | CEP85L | protein_coding | rs2046951 | 119021888 | 1.90e-10 | 33 | 0 | 2 | 14 | 1 | 16 | Female only | Both sexes | Female only | True | False |
| QRS_DURATION | 3 | SCN10A | protein_coding | rs6599240 | 38738717 | 2.00e-10 | 3 | 0 | 0 | 0 | 2 | 1 | Female only | Both sexes | Female only | True | False |
| QRS_DURATION | 3 | SCN5A | protein_coding | rs7374540 | 38634142 | 2.22e-10 | 9 | 0 | 1 | 4 | 0 | 4 | Female only | Both sexes | Female only | True | False |
| QRS_DURATION | 18 | SETBP1 | protein_coding | rs663651 | 42456653 | 2.71e-10 | 25 | 7 | 6 | 0 | 0 | 12 | Male only | Male only | Male only | True | False |
| QRS_DURATION | 6 | LAP3P2 | pseudogene | rs6930083 | 36634156 | 2.03e-09 | 23 | 0 | 0 | 1 | 0 | 22 | Both sexes | Both sexes | Both sexes | True | False |
| QRS_DURATION | 3 | SCN10A | protein_coding | rs6764249 | 38730117 | 5.17e-09 | 13 | 6 | 1 | 0 | 5 | 1 | Female only | Female only | Female only | True | False |
| QRS_DURATION | 12 | RP4-601P9.1 | lincRNA | rs7484657 | 115353801 | 8.03e-09 | 69 | 0 | 0 | 0 | 7 | 62 | Female only | Both sexes | Female only | True | False |
| QRS_DURATION | 3 | SCN10A | protein_coding | rs7615140 | 38782026 | 1.53e-08 | 79 | 0 | 4 | 20 | 43 | 12 | Female only | Both sexes | Female only | True | False |
| QRS_DURATION | 12 | ATP5B | protein_coding | rs2950393 | 57036023 | 1.65e-08 | 125 | 0 | 0 | 48 | 15 | 62 | Female only | Both sexes | Female only | True | False |
| QRS_DURATION | 6 | SLC35F1 | protein_coding | rs7752142 | 118548830 | 2.10e-08 | 19 | 8 | 2 | 0 | 0 | 9 | Female only | Female only | Female only | True | False |
| QRS_DURATION | 3 | RP11-536I6.2 | lincRNA | rs4685095 | 14292081 | 2.15e-08 | 27 | 7 | 7 | 0 | 0 | 13 | Female only | Female only | Female only | True | False |

| pheno | CHR | Gene_Symbol | biotype | lead_snp | lead_BP | lead_P | TOTAL | NSIG | S05 | S01 | S001 | S0001 | independent_loci_in_sex_P1e6 | independent_loci_in_sex_P1e2 | independent_loci_in_sex_P5e8 | in_gwas_catalog | in_HLA |
|---|---|---|---|---|---|---|---|---|---|---|---|---|---|---|---|---|---|
| QRS_DURATION | 3 | SCN10A | protein_coding | rs7611456 | 38813465 | 2.67e-08 | 5 | 0 | 0 | 0 | 2 | 3 | Female only | Both sexes | Female only | True | False |
| QRS_DURATION | 18 | SETBP1 | protein_coding | rs659583 | 42439989 | 2.73e-08 | 19 | 0 | 0 | 7 | 3 | 9 | Male only | Both sexes | Male only | True | False |
| QRS_DURATION | 10 | VTI1A | protein_coding | rs6585173 | 114455827 | 4.60e-08 | 95 | 0 | 0 | 0 | 1 | 94 | Both sexes | Both sexes | Male only | True | False |
| QRS_DURATION | 6 | SLC35F1 | protein_coding | rs7764093 | 118692304 | 4.62e-08 | 155 | 52 | 34 | 44 | 0 | 25 | Female only | Both sexes | Female only | True | False |
| QT.INTERVAL | 1 | NOS1AP | protein_coding | rs12143842 | 162033890 | 6.20e-09 | 117 | 12 | 12 | 35 | 1 | 57 | Female only | Both sexes | Female only | True | False |
| QT3 | 23 | SERPINA7 | protein_coding | X:105278361:C:A | 105278361 | 8.34e-09 | 31 | 0 | 0 | 0 | 0 | 31 | Male only | Both sexes | Male only | True | False |
| QTB.NL | 6 | RP11-486E2.1 | pseudogene | rs80238754 | 81491052 | 5.23e-09 | 5 | 3 | 0 | 0 | 0 | 2 | Female only | Female only | Female only | True | False |
| QTCB | 1 | NOS1AP | protein_coding | rs12143842 | 162033890 | 4.26e-16 | 117 | 0 | 0 | 1 | 11 | 105 | Both sexes | Both sexes | Both sexes | True | False |
| QTCB | 1 | NOS1AP | protein_coding | rs4582765 | 162090349 | 1.29e-08 | 133 | 4 | 9 | 6 | 2 | 112 | Female only | Both sexes | Female only | True | False |
| QTCF | 1 | NOS1AP | protein_coding | rs12143842 | 162033890 | 5.16e-18 | 117 | 0 | 0 | 0 | 11 | 106 | Both sexes | Both sexes | Both sexes | True | False |
| QTCF | 1 | NOS1AP | protein_coding | rs2225845 | 162095605 | 8.20e-11 | 137 | 0 | 0 | 9 | 37 | 91 | Female only | Both sexes | Female only | True | False |
| QT_INTERVAL | 6 | SLC35F1 | protein_coding | rs11153730 | 118667522 | 8.53e-15 | 261 | 0 | 3 | 66 | 11 | 181 | Both sexes | Both sexes | Female only | True | False |
| QT_INTERVAL | 1 | NOS1AP | protein_coding | rs12143842 | 162033890 | 1.11e-14 | 117 | 0 | 0 | 1 | 0 | 116 | Both sexes | Both sexes | Both sexes | True | False |
| QT_INTERVAL | 6 | CEP85L | protein_coding | rs11756438 | 118993632 | 1.50e-12 | 101 | 0 | 0 | 15 | 7 | 79 | Both sexes | Both sexes | Female only | True | False |
| pheno | CHR | Gene_Symbol | biotype | lead_snp | lead_BP | lead_P | TOTAL | NSIG | S05 | S01 | S001 | S0001 | independent_loci_in_sex_P1e6 | independent_loci_in_sex_P1e2 | independent_loci_in_sex_P5e8 | in_gwas_catalog | in_HLA |
| QT_INTERVAL | 6 | SLC35F1 | protein_coding | rs4946333 | 118565665 | 3.32e-10 | 63 | 0 | 2 | 10 | 35 | 16 | Both sexes | Both sexes | Female only | True | False |
| QT_INTERVAL | 6 | SLC35F1 | protein_coding | rs12210733 | 118653075 | 8.71e-10 | 163 | 15 | 47 | 9 | 58 | 34 | Female only | Both sexes | Female only | True | False |
| QT_INTERVAL | 6 | SLC35F1 | protein_coding | rs4945621 | 118658256 | 1.62e-09 | 51 | 1 | 4 | 19 | 18 | 9 | Female only | Both sexes | Female only | True | False |
| QT_INTERVAL | 1 | NOS1AP | protein_coding | rs10918701 | 162090536 | 1.91e-09 | 137 | 0 | 4 | 13 | 4 | 116 | Both sexes | Both sexes | Female only | True | False |

| pheno | CHR | Gene_Symbol | biotype | lead_snp | lead_BP | lead_P | TOTAL | NSIG | S05 | S01 | S001 | S0001 | independent_loci_in_sex_P1e6 | independent_loci_in_sex_P1e2 | independent_loci_in_sex_P5e8 | in_gwas_catalog | in_HLA |
|---|---|---|---|---|---|---|---|---|---|---|---|---|---|---|---|---|---|
| QT_INTERVAL | 6 | CEP85L | protein_coding | rs2638547 | 118995471 | 1.80e-08 | 203 | 0 | 4 | 1 | 91 | 107 | Both sexes | Both sexes | Female only | True | False |
| QT_INTERVAL | 15 | CHRNB4 | protein_coding | rs2869548 | 78922638 | 2.11e-08 | 101 | 51 | 0 | 0 | 0 | 50 | Male only | Male only | Male only | True | False |
| QT_INTERVAL | 6 | SLC35F1 | protein_coding | rs7453860 | 118565620 | 4.21e-08 | 29 | 24 | 0 | 0 | 1 | 4 | Female only | Both sexes | Female only | True | False |
| QT_INTERVAL_C | 1 | RP11-227F8.2 | lincRNA | rs12042862 | 162024242 | 1.73e-39 | 99 | 0 | 0 | 0 | 0 | 99 | Both sexes | Both sexes | Both sexes | True | False |
| QT_INTERVAL_C | 1 | NOS1AP | protein_coding | rs2152761 | 162105697 | 7.24e-23 | 153 | 0 | 0 | 0 | 0 | 153 | Both sexes | Both sexes | Both sexes | True | False |
| QT_INTERVAL_C | 1 | NOS1AP | protein_coding | rs12039600 | 162159482 | 2.46e-19 | 15 | 0 | 0 | 0 | 0 | 15 | Both sexes | Both sexes | Both sexes | True | False |
| QT_INTERVAL_C | 1 | NOS1AP | protein_coding | rs10918762 | 162118984 | 1.52e-18 | 25 | 0 | 0 | 1 | 3 | 21 | Female only | Both sexes | Female only | True | False |
| QT_INTERVAL_C | 1 | NOS1AP | protein_coding | rs3934467 | 162182677 | 1.71e-18 | 105 | 0 | 0 | 0 | 1 | 104 | Both sexes | Both sexes | Both sexes | True | False |
| QT_INTERVAL_C | 1 | NOS1AP | protein_coding | rs35021332 | 162046381 | 8.88e-17 | 7 | 0 | 0 | 0 | 0 | 7 | Both sexes | Both sexes | Both sexes | True | False |
| QT_INTERVAL_C | 1 | RP11-227F8.2 | lincRNA | rs7539281 | 162008034 | 1.47e-16 | 5 | 0 | 0 | 0 | 0 | 5 | Both sexes | Both sexes | Both sexes | True | False |
| QT_INTERVAL_C | 11 | KCNQ1 | protein_coding | rs7948689 | 2480464 | 4.34e-14 | 45 | 0 | 0 | 0 | 1 | 44 | Both sexes | Both sexes | Female only | True | False |
| QT_INTERVAL_C | 11 | KCNQ1 | protein_coding | rs7928116 | 2512325 | 1.62e-10 | 3 | 0 | 0 | 0 | 0 | 3 | Female only | Both sexes | Female only | True | False |
| QT_INTERVAL_C | 6 | SLC35F1 | protein_coding | rs2093369 | 118649044 | 1.75e-10 | 233 | 110 | 5 | 2 | 0 | 116 | Female only | Both sexes | Female only | True | False |
| QT_INTERVAL_C | 1 | RP11-227F8.2 | lincRNA | rs7539367 | 162008079 | 5.75e-10 | 19 | 0 | 0 | 0 | 0 | 19 | Both sexes | Both sexes | Female only | True | False |

| pheno | CHR | Gene_Symbol | biotype | lead_snp | lead_BP | lead_P | TOTAL | NSIG | S05 | S01 | S001 | S0001 | independent_loci_in_sex_P1e6 | independent_loci_in_sex_P1e2 | independent_loci_in_sex_P5e8 | in_gwas_catalog | in_HLA |
|---|---|---|---|---|---|---|---|---|---|---|---|---|---|---|---|---|---|
| QT_INTERVAL_C | 7 | KCNH2 | protein_coding | rs34657537 | 150657808 | 7.10e-10 | 43 | 0 | 1 | 3 | 14 | 25 | Female only | Both sexes | Female only | True | False |
| QT_INTERVAL_C | 2 | SLC8A1 | protein_coding | rs35450971 | 40754314 | 2.10e-09 | 3 | 0 | 0 | 0 | 0 | 3 | Female only | Both sexes | Female only | True | False |
| QT_INTERVAL_C | 6 | SLC35F1 | protein_coding | rs89107 | 118578043 | 2.27e-09 | 75 | 27 | 7 | 7 | 2 | 32 | Female only | Both sexes | Female only | True | False |
| QT_INTERVAL_C | 6 | CEP85L | protein_coding | rs10872167 | 118988362 | 6.26e-09 | 81 | 28 | 13 | 0 | 0 | 40 | Female only | Female only | Female only | True | False |
| QT_INTERVAL_C | 16 | CNOT1 | protein_coding | rs7188697 | 58622178 | 8.32e-09 | 189 | 0 | 0 | 11 | 8 | 170 | Both sexes | Both sexes | Both sexes | True | False |
| QT_INTERVAL_C | 1 | ATP1B1 | protein_coding | rs10919062 | 169088947 | 2.96e-08 | 107 | 0 | 0 | 4 | 47 | 56 | Both sexes | Both sexes | Female only | True | False |
| QT_INTERVAL_C | 1 | RP5-1018K9.1 | antisense | rs1320977 | 169073388 | 4.50e-08 | 31 | 0 | 0 | 0 | 1 | 30 | Female only | Both sexes | Female only | True | False |
| R.APACHE.TEMPERATURE | 12 | RP11-560G2.1 | antisense | rs10879892 | 75647143 | 9.17e-09 | 11 | 6 | 0 | 0 | 0 | 5 | Male only | Male only | Male only | False | False |
| R.APACHE.TEMPERATURE | 2 | KLF7 | protein_coding | rs16839150 | 207955158 | 1.31e-08 | 29 | 15 | 0 | 0 | 0 | 14 | Male only | Male only | Male only | False | False |
| R.ED.PRE.ARRIVAL.RESP.RATE | 13 | GPC5 | protein_coding | rs67133719 | 93114447 | 4.22e-09 | 9 | 5 | 0 | 0 | 0 | 4 | Female only | Female only | Female only | False | False |
| R.IP.IDEAL.BODY.WEIGHT | 4 | LCORL | protein_coding | rs112675878 | 18027696 | 4.07e-17 | 449 | 0 | 0 | 5 | 108 | 336 | Female only | Both sexes | Female only | False | False |

| pheno | CHR | Gene_Symbol | biotype | lead_snp | lead_BP | lead_P | TOTAL | NSIG | S05 | S01 | S001 | S0001 | independent_loci_in_sex_P1e6 | independent_loci_in_sex_P1e2 | independent_loci_in_sex_P5e8 | in_gwas_catalog | in_HLA |
|---|---|---|---|---|---|---|---|---|---|---|---|---|---|---|---|---|---|
| R.IP.IDEAL.BODY.WEIGHT | 20 | GDF5 | protein_coding | rs143384 | 34025756 | 1.37e-13 | 221 | 0 | 0 | 0 | 0 | 221 | Both sexes | Both sexes | Both sexes | False | False |
| R.IP.IDEAL.BODY.WEIGHT | 3 | ZBTB38 | protein_coding | rs6440006 | 141142691 | 1.94e-13 | 75 | 0 | 0 | 0 | 1 | 74 | Both sexes | Both sexes | Both sexes | False | False |
| R.IP.IDEAL.BODY.WEIGHT | 2 | EFEMP1 | protein_coding | rs1430203 | 56072509 | 7.57e-12 | 103 | 16 | 10 | 25 | 0 | 52 | Male only | Both sexes | Male only | False | False |
| R.IP.IDEAL.BODY.WEIGHT | 15 | ADAMTSL3 | protein_coding | rs1812707 | 84547222 | 4.67e-11 | 137 | 0 | 0 | 67 | 2 | 68 | Female only | Both sexes | Female only | False | False |
| R.IP.IDEAL.BODY.WEIGHT | 4 | HHIP-AS1 | antisense | rs13106087 | 145566864 | 2.59e-10 | 19 | 0 | 0 | 0 | 2 | 17 | Both sexes | Both sexes | Female only | False | False |
| R.IP.IDEAL.BODY.WEIGHT | 18 | CABLES1 | protein_coding | rs9947743 | 20708321 | 7.32e-10 | 39 | 0 | 0 | 0 | 0 | 39 | Both sexes | Both sexes | Both sexes | False | False |
| R.IP.IDEAL.BODY.WEIGHT | 17 | ATAD5 | protein_coding | rs3764419 | 29164023 | 9.79e-10 | 29 | 7 | 8 | 1 | 0 | 13 | Female only | Female only | Female only | False | False |
| R.IP.IDEAL.BODY.WEIGHT | 12 | HMGA2 | protein_coding | rs1351394 | 66351826 | 2.29e-09 | 31 | 0 | 0 | 0 | 1 | 30 | Female only | Both sexes | Female only | False | False |
| R.IP.IDEAL.BODY.WEIGHT | 3 | ZBTB38 | protein_coding | rs13100711 | 141082990 | 4.88e-09 | 19 | 2 | 3 | 5 | 0 | 9 | Female only | Both sexes | Female only | False | False |
| R.IP.IDEAL.BODY.WEIGHT | 2 | LTBP1 | protein_coding | rs6743685 | 33389595 | 5.23e-09 | 201 | 63 | 38 | 0 | 0 | 100 | Female only | Female only | Female only | False | False |
| R.IP.IDEAL.BODY.WEIGHT | 13 | DLEU1 | protein_coding | rs2066647 | 50782074 | 6.49e-09 | 31 | 1 | 5 | 10 | 0 | 15 | Female only | Both sexes | Female only | False | False |

| pheno | CHR | Gene_Symbol | biotype | lead_snp | lead_BP | lead_P | TOTAL | NSIG | S05 | S01 | S001 | S0001 | independent_loci_in_sex_P1e6 | independent_loci_in_sex_P1e2 | independent_loci_in_sex_P5e8 | in_gwas_catalog | in_HLA |
|---|---|---|---|---|---|---|---|---|---|---|---|---|---|---|---|---|---|
| R.IP.IDEAL.BODY.WEIGHT | 2 | EFEMP1 | protein_coding | rs59985551 | 56106928 | 6.74e-09 | 17 | 1 | 0 | 6 | 2 | 8 | Male only | Both sexes | Male only | False | False |
| R.IP.IDEAL.BODY.WEIGHT | 12 | CCDC91 | protein_coding | rs11049356 | 28271456 | 6.78e-09 | 465 | 233 | 0 | 0 | 3 | 229 | Female only | Female only | Female only | False | False |
| R.IP.IDEAL.BODY.WEIGHT | 1 | DNM3 | protein_coding | rs678962 | 172189889 | 1.44e-08 | 37 | 0 | 0 | 10 | 9 | 18 | Female only | Both sexes | Female only | False | False |
| R.IP.IDEAL.BODY.WEIGHT | 11 | SSH3 | protein_coding | rs76335321 | 67097778 | 1.99e-08 | 7 | 0 | 0 | 1 | 0 | 6 | Female only | Both sexes | Female only | False | False |
| R.IP.IDEAL.BODY.WEIGHT | 13 | DLEU1 | protein_coding | rs3116598 | 51099616 | 2.00e-08 | 109 | 0 | 0 | 2 | 4 | 103 | Both sexes | Both sexes | Female only | False | False |
| R.IP.IDEAL.BODY.WEIGHT | 18 | CABLES1 | protein_coding | rs4567840 | 20705158 | 2.09e-08 | 65 | 4 | 0 | 21 | 12 | 28 | Male only | Both sexes | Male only | False | False |
| R.IP.IDEAL.BODY.WEIGHT | 15 | ACAN | protein_coding | rs2280470 | 89395626 | 2.10e-08 | 23 | 12 | 0 | 0 | 5 | 6 | Female only | Female only | Female only | False | False |
| R.IP.IDEAL.BODY.WEIGHT | 7 | IGF2BP3 | protein_coding | rs67397633 | 23491190 | 2.11e-08 | 49 | 0 | 0 | 2 | 25 | 22 | Male only | Both sexes | Male only | False | False |
| R.IP.IDEAL.BODY.WEIGHT | 2 | EFEMP1 | protein_coding | rs11125609 | 56115834 | 2.12e-08 | 5 | 3 | 0 | 0 | 0 | 2 | Male only | Male only | Male only | False | False |
| R.IP.IDEAL.BODY.WEIGHT | 6 | RP11-486E2.1 | pseudogene | rs9359426 | 81442039 | 4.53e-08 | 181 | 55 | 73 | 5 | 5 | 43 | Female only | Both sexes | Female only | False | False |

| pheno | CHR | Gene_Symbol | biotype | lead_snp | lead_BP | lead_P | TOTAL | NSIG | S05 | S01 | S001 | S0001 | independent_loci_in_sex_P1e6 | independent_loci_in_sex_P1e2 | independent_loci_in_sex_P5e8 | in_gwas_catalog | in_HLA |
|---|---|---|---|---|---|---|---|---|---|---|---|---|---|---|---|---|---|
| R.RT..FEMALE.ADULT..IDEAL.BODY.WEIGHT | 4 | LCORL | protein_coding | rs112675878 | 18027696 | 3.98e-17 | 449 | 0 | 0 | 3 | 107 | 339 | Female only | Both sexes | Female only | False | False |
| R.RT..FEMALE.ADULT..IDEAL.BODY.WEIGHT | 20 | GDF5 | protein_coding | rs143384 | 34025756 | 1.22e-13 | 221 | 0 | 0 | 0 | 0 | 221 | Both sexes | Both sexes | Both sexes | False | False |
| R.RT..FEMALE.ADULT..IDEAL.BODY.WEIGHT | 3 | ZBTB38 | protein_coding | rs6440006 | 141142691 | 2.10e-13 | 75 | 0 | 0 | 0 | 1 | 74 | Both sexes | Both sexes | Both sexes | False | False |
| R.RT..FEMALE.ADULT..IDEAL.BODY.WEIGHT | 2 | EFEMP1 | protein_coding | rs1430203 | 56072509 | 7.85e-12 | 103 | 15 | 11 | 25 | 0 | 52 | Male only | Both sexes | Male only | False | False |
| R.RT..FEMALE.ADULT..IDEAL.BODY.WEIGHT | 15 | ADAMTSL3 | protein_coding | rs1812707 | 84547222 | 4.72e-11 | 137 | 0 | 0 | 68 | 1 | 68 | Female only | Both sexes | Female only | False | False |
| R.RT..FEMALE.ADULT..IDEAL.BODY.WEIGHT | 4 | HHIP-AS1 | antisense | rs13106087 | 145566864 | 3.07e-10 | 19 | 0 | 0 | 0 | 2 | 17 | Both sexes | Both sexes | Female only | False | False |

| pheno | CHR | Gene_Symbol | biotype | lead_snp | lead_BP | lead_P | TOTAL | NSIG | S05 | S01 | S001 | S0001 | independent_loci_in_sex_P1e6 | independent_loci_in_sex_P1e2 | independent_loci_in_sex_P5e8 | in_gwas_catalog | in_HLA |
|---|---|---|---|---|---|---|---|---|---|---|---|---|---|---|---|---|---|
| R.RT..FEMALE.ADULT..IDEAL.BODY.WEIGHT | 18 | CABLES1 | protein_coding | rs9947743 | 20708321 | 7.53e-10 | 39 | 0 | 0 | 0 | 0 | 39 | Both sexes | Both sexes | Both sexes | False | False |
| R.RT..FEMALE.ADULT..IDEAL.BODY.WEIGHT | 17 | ATAD5 | protein_coding | rs3764419 | 29164023 | 8.90e-10 | 29 | 7 | 8 | 1 | 0 | 13 | Female only | Female only | Female only | False | False |
| R.RT..FEMALE.ADULT..IDEAL.BODY.WEIGHT | 12 | HMGA2 | protein_coding | rs1351394 | 66351826 | 2.67e-09 | 31 | 0 | 0 | 0 | 1 | 30 | Female only | Both sexes | Female only | False | False |
| R.RT..FEMALE.ADULT..IDEAL.BODY.WEIGHT | 3 | ZBTB38 | protein_coding | rs13100711 | 141082990 | 5.15e-09 | 19 | 2 | 3 | 5 | 0 | 9 | Female only | Both sexes | Female only | False | False |
| R.RT..FEMALE.ADULT..IDEAL.BODY.WEIGHT | 2 | LTBP1 | protein_coding | rs6743685 | 33389595 | 5.51e-09 | 201 | 63 | 38 | 0 | 0 | 100 | Female only | Female only | Female only | False | False |
| R.RT..FEMALE.ADULT..IDEAL.BODY.WEIGHT | 13 | DLEU1 | protein_coding | rs2066647 | 50782074 | 6.40e-09 | 31 | 2 | 4 | 10 | 0 | 15 | Female only | Both sexes | Female only | False | False |

| pheno | CHR | Gene_Symbol | biotype | lead_snp | lead_BP | lead_P | TOTAL | NSIG | S05 | S01 | S001 | S0001 | independent_loci_in_sex_P1e6 | independent_loci_in_sex_P1e2 | independent_loci_in_sex_P5e8 | in_gwas_catalog | in_HLA |
|---|---|---|---|---|---|---|---|---|---|---|---|---|---|---|---|---|---|
| R.RT..FEMALE.ADULT..IDEAL.BODY.WEIGHT | 2 | EFEMP1 | protein_coding | rs59985551 | 56106928 | 6.79e-09 | 17 | 1 | 0 | 6 | 2 | 8 | Male only | Both sexes | Male only | False | False |
| R.RT..FEMALE.ADULT..IDEAL.BODY.WEIGHT | 12 | CCDC91 | protein_coding | rs11049356 | 28271456 | 7.01e-09 | 465 | 233 | 0 | 0 | 3 | 229 | Female only | Female only | Female only | False | False |
| R.RT..FEMALE.ADULT..IDEAL.BODY.WEIGHT | 1 | DNM3 | protein_coding | rs678962 | 172189889 | 1.36e-08 | 37 | 0 | 0 | 10 | 9 | 18 | Female only | Both sexes | Female only | False | False |
| R.RT..FEMALE.ADULT..IDEAL.BODY.WEIGHT | 11 | SSH3 | protein_coding | rs76335321 | 67097778 | 1.75e-08 | 7 | 0 | 0 | 1 | 0 | 6 | Female only | Both sexes | Female only | False | False |
| R.RT..FEMALE.ADULT..IDEAL.BODY.WEIGHT | 15 | ACAN | protein_coding | rs2280470 | 89395626 | 1.87e-08 | 23 | 12 | 0 | 0 | 5 | 6 | Female only | Female only | Female only | False | False |
| R.RT..FEMALE.ADULT..IDEAL.BODY.WEIGHT | 13 | DLEU1 | protein_coding | rs3116598 | 51099616 | 1.97e-08 | 109 | 0 | 0 | 2 | 4 | 103 | Both sexes | Both sexes | Female only | False | False |

| pheno | CHR | Gene_Symbol | biotype | lead_snp | lead_BP | lead_P | TOTAL | NSIG | S05 | S01 | S001 | S0001 | independent_loci_in_sex_P1e6 | independent_loci_in_sex_P1e2 | independent_loci_in_sex_P5e8 | in_gwas_catalog | in_HLA |
|---|---|---|---|---|---|---|---|---|---|---|---|---|---|---|---|---|---|
| R.RT..FEMALE.ADULT..IDEAL.BODY.WEIGHT | 7 | IGF2BP3 | protein_coding | rs67397633 | 23491190 | 2.13e-08 | 49 | 0 | 0 | 2 | 26 | 21 | Male only | Both sexes | Male only | False | False |
| R.RT..FEMALE.ADULT..IDEAL.BODY.WEIGHT | 18 | CABLES1 | protein_coding | rs4567840 | 20705158 | 2.14e-08 | 65 | 4 | 0 | 21 | 12 | 28 | Male only | Both sexes | Male only | False | False |
| R.RT..FEMALE.ADULT..IDEAL.BODY.WEIGHT | 2 | EFEMP1 | protein_coding | rs11125609 | 56115834 | 2.15e-08 | 5 | 3 | 0 | 0 | 0 | 2 | Male only | Male only | Male only | False | False |
| R.RT..FEMALE.ADULT..IDEAL.BODY.WEIGHT | 6 | RP11-486E2.1 | pseudogene | rs9359426 | 81442039 | 4.35e-08 | 181 | 60 | 68 | 4 | 6 | 43 | Female only | Both sexes | Female only | False | False |
| R.RT..FEMALE.ADULT..IDEAL.BODY.WEIGHT | 2 | EFEMP1 | protein_coding | rs1346790 | 56091871 | 4.88e-08 | 65 | 44 | 2 | 1 | 0 | 18 | Male only | Male only | Male only | False | False |
| R.RT..MALE.ADULT..IDEAL.BODY.WEIGHT | 4 | LCORL | protein_coding | rs112675878 | 18027696 | 3.98e-17 | 449 | 0 | 0 | 3 | 107 | 339 | Female only | Both sexes | Female only | False | False |

| pheno | CHR | Gene_Symbol | biotype | lead_snp | lead_BP | lead_P | TOTAL | NSIG | S05 | S01 | S001 | S0001 | independent_loci_in_sex_P1e6 | independent_loci_in_sex_P1e2 | independent_loci_in_sex_P5e8 | in_gwas_catalog | in_HLA |
|---|---|---|---|---|---|---|---|---|---|---|---|---|---|---|---|---|---|
| R.RT..MALE.ADULT..IDEAL.BODY.WEIGHT | 20 | GDF5 | protein_coding | rs143384 | 34025756 | 1.22e-13 | 221 | 0 | 0 | 0 | 0 | 221 | Both sexes | Both sexes | Both sexes | False | False |
| R.RT..MALE.ADULT..IDEAL.BODY.WEIGHT | 3 | ZBTB38 | protein_coding | rs6440006 | 141142691 | 2.10e-13 | 75 | 0 | 0 | 0 | 1 | 74 | Both sexes | Both sexes | Both sexes | False | False |
| R.RT..MALE.ADULT..IDEAL.BODY.WEIGHT | 2 | EFEMP1 | protein_coding | rs1430203 | 56072509 | 7.85e-12 | 103 | 15 | 11 | 25 | 0 | 52 | Male only | Both sexes | Male only | False | False |
| R.RT..MALE.ADULT..IDEAL.BODY.WEIGHT | 15 | ADAMTSL3 | protein_coding | rs1812707 | 84547222 | 4.72e-11 | 137 | 0 | 0 | 68 | 1 | 68 | Female only | Both sexes | Female only | False | False |
| R.RT..MALE.ADULT..IDEAL.BODY.WEIGHT | 4 | HHIP-AS1 | antisense | rs13106087 | 145566864 | 3.07e-10 | 19 | 0 | 0 | 0 | 2 | 17 | Both sexes | Both sexes | Female only | False | False |
| R.RT..MALE.ADULT..IDEAL.BODY.WEIGHT | 18 | CABLES1 | protein_coding | rs9947743 | 20708321 | 7.53e-10 | 39 | 0 | 0 | 0 | 0 | 39 | Both sexes | Both sexes | Both sexes | False | False |

| pheno | CHR | Gene_Symbol | biotype | lead_snp | lead_BP | lead_P | TOTAL | NSIG | S05 | S01 | S001 | S0001 | independent_loci_in_sex_P1e6 | independent_loci_in_sex_P1e2 | independent_loci_in_sex_P5e8 | in_gwas_catalog | in_HLA |
|---|---|---|---|---|---|---|---|---|---|---|---|---|---|---|---|---|---|
| R.RT..MALE.ADULT..IDEAL.BODY.WEIGHT | 17 | ATAD5 | protein_coding | rs3764419 | 29164023 | 8.90e-10 | 29 | 7 | 8 | 1 | 0 | 13 | Female only | Female only | Female only | False | False |
| R.RT..MALE.ADULT..IDEAL.BODY.WEIGHT | 12 | HMGA2 | protein_coding | rs1351394 | 66351826 | 2.67e-09 | 31 | 0 | 0 | 0 | 1 | 30 | Female only | Both sexes | Female only | False | False |
| R.RT..MALE.ADULT..IDEAL.BODY.WEIGHT | 3 | ZBTB38 | protein_coding | rs13100711 | 141082990 | 5.15e-09 | 19 | 2 | 3 | 5 | 0 | 9 | Female only | Both sexes | Female only | False | False |
| R.RT..MALE.ADULT..IDEAL.BODY.WEIGHT | 2 | LTBP1 | protein_coding | rs6743685 | 33389595 | 5.51e-09 | 201 | 63 | 38 | 0 | 0 | 100 | Female only | Female only | Female only | False | False |
| R.RT..MALE.ADULT..IDEAL.BODY.WEIGHT | 13 | DLEU1 | protein_coding | rs2066647 | 50782074 | 6.40e-09 | 31 | 2 | 4 | 10 | 0 | 15 | Female only | Both sexes | Female only | False | False |
| R.RT..MALE.ADULT..IDEAL.BODY.WEIGHT | 2 | EFEMP1 | protein_coding | rs59985551 | 56106928 | 6.79e-09 | 17 | 1 | 0 | 6 | 2 | 8 | Male only | Both sexes | Male only | False | False |

| pheno | CHR | Gene_Symbol | biotype | lead_snp | lead_BP | lead_P | TOTAL | NSIG | S05 | S01 | S001 | S0001 | independent_loci_in_sex_P1e6 | independent_loci_in_sex_P1e2 | independent_loci_in_sex_P5e8 | in_gwas_catalog | in_HLA |
|---|---|---|---|---|---|---|---|---|---|---|---|---|---|---|---|---|---|
| R.RT..MALE.ADULT..IDEAL.BODY.WEIGHT | 12 | CCDC91 | protein_coding | rs11049356 | 28271456 | 7.01e-09 | 465 | 233 | 0 | 0 | 3 | 229 | Female only | Female only | Female only | False | False |
| R.RT..MALE.ADULT..IDEAL.BODY.WEIGHT | 1 | DNM3 | protein_coding | rs678962 | 172189889 | 1.36e-08 | 37 | 0 | 0 | 10 | 9 | 18 | Female only | Both sexes | Female only | False | False |
| R.RT..MALE.ADULT..IDEAL.BODY.WEIGHT | 11 | SSH3 | protein_coding | rs76335321 | 67097778 | 1.75e-08 | 7 | 0 | 0 | 1 | 0 | 6 | Female only | Both sexes | Female only | False | False |
| R.RT..MALE.ADULT..IDEAL.BODY.WEIGHT | 15 | ACAN | protein_coding | rs2280470 | 89395626 | 1.87e-08 | 23 | 12 | 0 | 0 | 5 | 6 | Female only | Female only | Female only | False | False |
| R.RT..MALE.ADULT..IDEAL.BODY.WEIGHT | 13 | DLEU1 | protein_coding | rs3116598 | 51099616 | 1.97e-08 | 109 | 0 | 0 | 2 | 4 | 103 | Both sexes | Both sexes | Female only | False | False |
| R.RT..MALE.ADULT..IDEAL.BODY.WEIGHT | 7 | IGF2BP3 | protein_coding | rs67397633 | 23491190 | 2.13e-08 | 49 | 0 | 0 | 2 | 26 | 21 | Male only | Both sexes | Male only | False | False |

| pheno | CHR | Gene_Symbol | biotype | lead_snp | lead_BP | lead_P | TOTAL | NSIG | S05 | S01 | S001 | S0001 | independent_loci_in_sex_P1e6 | independent_loci_in_sex_P1e2 | independent_loci_in_sex_P5e8 | in_gwas_catalog | in_HLA |
|---|---|---|---|---|---|---|---|---|---|---|---|---|---|---|---|---|---|
| R.RT..MALE.ADULT..IDEAL.BODY.WEIGHT | 18 | CABLES1 | protein_coding | rs4567840 | 20705158 | 2.14e-08 | 65 | 4 | 0 | 21 | 12 | 28 | Male only | Both sexes | Male only | False | False |
| R.RT..MALE.ADULT..IDEAL.BODY.WEIGHT | 2 | EFEMP1 | protein_coding | rs11125609 | 56115834 | 2.15e-08 | 5 | 3 | 0 | 0 | 0 | 2 | Male only | Male only | Male only | False | False |
| R.RT..MALE.ADULT..IDEAL.BODY.WEIGHT | 6 | RP11-486E2.1 | pseudogene | rs9359426 | 81442039 | 4.35e-08 | 181 | 60 | 68 | 4 | 6 | 43 | Female only | Both sexes | Female only | False | False |
| R.RT..MALE.ADULT..IDEAL.BODY.WEIGHT | 2 | EFEMP1 | protein_coding | rs1346790 | 56091871 | 4.88e-08 | 65 | 44 | 2 | 1 | 0 | 18 | Male only | Male only | Male only | False | False |
| RATE_C | 6 | HMGB3P18 | pseudogene | rs9388006 | 122133347 | 2.33e-09 | 125 | 0 | 0 | 27 | 2 | 96 | Both sexes | Both sexes | Both sexes | True | False |
| RATE_C | 11 | DLG2 | protein_coding | rs79346577 | 84989662 | 2.88e-08 | 11 | 0 | 0 | 2 | 4 | 5 | Male only | Both sexes | Male only | True | False |
| RBC | 6 | HBS1L | protein_coding | rs9376091 | 135419636 | 1.08e-53 | 35 | 0 | 0 | 0 | 0 | 35 | Both sexes | Both sexes | Both sexes | True | False |
| RBC | 6 | HBS1L | protein_coding | rs9389272 | 135459837 | 1.29e-31 | 35 | 0 | 0 | 2 | 0 | 33 | Both sexes | Both sexes | Both sexes | True | False |
| RBC | 6 | HBS1L | protein_coding | rs2210366 | 135415208 | 1.31e-21 | 69 | 0 | 0 | 0 | 2 | 67 | Both sexes | Both sexes | Female only | True | False |
| RBC | 4 | AC006552.1 | lincRNA | rs218265 | 55408999 | 8.21e-21 | 13 | 0 | 0 | 0 | 2 | 11 | Both sexes | Both sexes | Both sexes | True | False |
| RBC | 7 | TFR2 | protein_coding | rs9801017 | 100236202 | 2.31e-18 | 43 | 0 | 0 | 0 | 0 | 43 | Both sexes | Both sexes | Both sexes | True | False |
| RBC | 4 | AC006552.1 | lincRNA | rs170121 | 55407099 | 3.00e-17 | 15 | 0 | 0 | 4 | 0 | 11 | Both sexes | Both sexes | Female only | True | False |
| RBC | 6 | MYB | protein_coding | rs61593090 | 135488083 | 8.42e-17 | 25 | 0 | 4 | 9 | 0 | 12 | Female only | Both sexes | Female only | True | False |

| pheno | CHR | Gene_Symbol | biotype | lead_snp | lead_BP | lead_P | TOTAL | NSIG | S05 | S01 | S001 | S0001 | independent_loci_in_sex_P1e6 | independent_loci_in_sex_P1e2 | independent_loci_in_sex_P5e8 | in_gwas_catalog | in_HLA |
|---|---|---|---|---|---|---|---|---|---|---|---|---|---|---|---|---|---|
| RBC | 6 | MYB | protein_coding | rs9494167 | 135489069 | 3.26e-15 | 21 | 0 | 2 | 3 | 6 | 10 | Female only | Both sexes | Female only | True | False |
| RBC | 4 | AC006552.1 | lincRNA | rs80113191 | 55396420 | 1.85e-13 | 101 | 0 | 0 | 48 | 3 | 50 | Female only | Both sexes | Female only | True | False |
| RBC | 7 | TFR2 | protein_coding | rs73156234 | 100234744 | 1.21e-12 | 79 | 0 | 0 | 0 | 0 | 79 | Both sexes | Both sexes | Both sexes | True | False |
| RBC | 6 | C6orf183 | polymorphic_pseudogene | rs11964178 | 109562035 | 7.61e-12 | 441 | 42 | 48 | 131 | 0 | 220 | Female only | Both sexes | Female only | True | False |
| RBC | 6 | HBS1L | protein_coding | rs76267242 | 135417460 | 3.51e-11 | 21 | 0 | 2 | 8 | 1 | 10 | Female only | Both sexes | Female only | True | False |
| RBC | 7 | ZAN | polymorphic_pseudogene | rs3891038 | 100362411 | 1.02e-10 | 11 | 0 | 0 | 0 | 0 | 11 | Both sexes | Both sexes | Female only | True | False |
| RBC | 6 | HBS1L | protein_coding | rs11758774 | 135383789 | 1.80e-10 | 289 | 0 | 0 | 194 | 8 | 87 | Female only | Both sexes | Female only | True | False |
| RBC | 6 | HBS1L | protein_coding | rs6914564 | 135403624 | 2.54e-09 | 55 | 25 | 0 | 0 | 1 | 29 | Female only | Both sexes | Female only | True | False |
| RBC | 19 | GCDH | protein_coding | rs62108438 | 13000247 | 3.25e-09 | 75 | 7 | 7 | 22 | 2 | 37 | Male only | Both sexes | Male only | True | False |
| RBC | 6 | RP3-352A20.1 | pseudogene | rs9402659 | 135195889 | 3.48e-09 | 47 | 18 | 11 | 4 | 3 | 11 | Female only | Both sexes | Female only | True | False |
| RBC | 6 | RP3-352A20.1 | pseudogene | rs9494099 | 135193172 | 3.62e-09 | 59 | 0 | 29 | 1 | 0 | 29 | Female only | Both sexes | Female only | True | False |
| RBC | 4 | RP11-545H22.1 | lincRNA | rs11723766 | 55348554 | 7.41e-09 | 47 | 0 | 0 | 19 | 6 | 22 | Female only | Both sexes | Female only | True | False |
| RBC | 6 | HBS1L | protein_coding | rs6930223 | 135424203 | 3.32e-08 | 27 | 17 | 1 | 0 | 2 | 7 | Female only | Both sexes | Female only | True | False |
| RBC | 6 | HIST1H4C | protein_coding | rs707890 | 26105504 | 4.32e-08 | 69 | 40 | 8 | 1 | 0 | 20 | Female only | Female only | Female only | True | False |
| RDW | 22 | TMPRSS6 | protein_coding | rs4820268 | 37469591 | 3.66e-36 | 9 | 0 | 0 | 0 | 0 | 9 | Both sexes | Both sexes | Both sexes | True | False |

| pheno | CHR | Gene_Symbol | biotype | lead_snp | lead_BP | lead_P | TOTAL | NSIG | S05 | S01 | S001 | S0001 | independent_loci_in_sex_P1e6 | independent_loci_in_sex_P1e2 | independent_loci_in_sex_P5e8 | in_gwas_catalog | in_HLA |
|---|---|---|---|---|---|---|---|---|---|---|---|---|---|---|---|---|---|
| RDW | 5 | CTC-228N24.3 | lincRNA | rs6888037 | 127406259 | 1.83e-26 | 85 | 0 | 0 | 0 | 0 | 85 | Both sexes | Both sexes | Both sexes | True | False |
| RDW | 6 | HIST1H1T | protein_coding | rs198846 | 26107463 | 7.11e-18 | 33 | 0 | 0 | 0 | 0 | 33 | Both sexes | Both sexes | Both sexes | True | False |
| RDW | 5 | SLC12A7 | protein_coding | rs4580814 | 1113244 | 8.44e-17 | 29 | 0 | 0 | 0 | 0 | 29 | Both sexes | Both sexes | Both sexes | True | False |
| RDW | 6 | HIST1H3C | protein_coding | 6:26046744 | 26046744 | 1.04e-15 | 245 | 14 | 0 | 6 | 40 | 185 | Both sexes | Both sexes | Female only | True | False |
| RDW | 22 | MPST | protein_coding | rs130610 | 37431871 | 1.05e-15 | 33 | 0 | 0 | 6 | 0 | 27 | Both sexes | Both sexes | Female only | True | False |
| RDW | 6 | HFE | protein_coding | rs1800562 | 26093141 | 2.10e-15 | 15 | 0 | 0 | 0 | 0 | 15 | Both sexes | Both sexes | Both sexes | True | False |
| RDW | 6 | SLC17A1 | protein_coding | rs80176492 | 25828106 | 5.12e-15 | 9 | 0 | 0 | 0 | 0 | 9 | Both sexes | Both sexes | Both sexes | True | False |
| RDW | 6 | CCND3 | protein_coding | rs10947997 | 41921241 | 6.31e-15 | 57 | 0 | 0 | 0 | 2 | 55 | Female only | Both sexes | Female only | True | False |
| RDW | 5 | SLC12A2 | protein_coding | rs10067555 | 127427533 | 4.71e-13 | 45 | 0 | 0 | 0 | 0 | 45 | Both sexes | Both sexes | Male only | True | False |
| RDW | 15 | WDR61 | protein_coding | rs4887030 | 78599995 | 5.11e-13 | 25 | 1 | 0 | 1 | 3 | 20 | Female only | Both sexes | Female only | True | False |

| pheno | CHR | Gene_Symbol | biotype | lead_snp | lead_BP | lead_P | TOTAL | NSIG | S05 | S01 | S001 | S0001 | independent_loci_in_sex_P1e6 | independent_loci_in_sex_P1e2 | independent_loci_in_sex_P5e8 | in_gwas_catalog | in_HLA |
|---|---|---|---|---|---|---|---|---|---|---|---|---|---|---|---|---|---|
| RDW | 17 | FAM222B | protein_coding | rs9892942 | 27178761 | 1.01e-12 | 159 | 0 | 0 | 0 | 0 | 159 | Both sexes | Both sexes | Both sexes | True | False |
| RDW | 22 | TMPRSS6 | protein_coding | rs2235325 | 37481637 | 6.80e-12 | 23 | 0 | 0 | 4 | 8 | 11 | Female only | Both sexes | Female only | True | False |
| RDW | 15 | DNAJA4 | protein_coding | rs113594982 | 78557392 | 9.31e-12 | 47 | 0 | 1 | 0 | 0 | 46 | Female only | Both sexes | Female only | True | False |
| RDW | 15 | WDR61 | protein_coding | rs870731 | 78604485 | 9.94e-12 | 37 | 2 | 1 | 2 | 7 | 25 | Female only | Both sexes | Female only | True | False |
| RDW | 16 | FTO | protein_coding | rs3751812 | 53818460 | 1.07e-11 | 169 | 9 | 4 | 72 | 0 | 84 | Female only | Both sexes | Female only | True | False |
| RDW | 18 | C18orf25 | protein_coding | rs8095374 | 43793488 | 1.94e-11 | 5 | 0 | 0 | 0 | 0 | 5 | Both sexes | Both sexes | Female only | True | False |
| RDW | 6 | U91328.22 | lincRNA | rs9356996 | 26008487 | 2.30e-11 | 63 | 0 | 22 | 7 | 0 | 34 | Both sexes | Both sexes | Female only | True | False |
| RDW | 6 | LRRC16A | protein_coding | rs2274089 | 25488583 | 7.04e-11 | 49 | 0 | 0 | 0 | 0 | 49 | Both sexes | Both sexes | Both sexes | True | False |
| RDW | 1 | ATP2B4 | protein_coding | rs10900588 | 203656814 | 1.17e-10 | 3 | 0 | 0 | 2 | 0 | 1 | Female only | Both sexes | Female only | True | False |
| RDW | 15 | RP11-762H8.3 | antisense | rs8039884 | 78550081 | 1.60e-10 | 25 | 0 | 0 | 1 | 1 | 23 | Female only | Both sexes | Female only | True | False |
| RDW | 6 | HBS1L | protein_coding | rs9399136 | 135402339 | 1.81e-10 | 35 | 0 | 3 | 8 | 7 | 17 | Female only | Both sexes | Female only | True | False |
| RDW | 6 | SCGN | protein_coding | rs1321250 | 25642244 | 1.96e-10 | 25 | 0 | 0 | 0 | 0 | 25 | Both sexes | Both sexes | Male only | True | False |
| RDW | 22 | TMPRSS6 | protein_coding | rs6000550 | 37463450 | 2.17e-10 | 15 | 3 | 1 | 7 | 1 | 3 | Female only | Both sexes | Female only | True | False |
| RDW | 5 | SLC12A7 | protein_coding | rs4314449 | 1102994 | 2.55e-10 | 5 | 0 | 0 | 0 | 0 | 5 | Both sexes | Both sexes | Female only | True | False |
| RDW | 5 | SLC12A2 | protein_coding | rs13183342 | 127502389 | 4.87e-10 | 3 | 0 | 0 | 0 | 0 | 3 | Both sexes | Both sexes | Male only | True | False |
| RDW | 5 | SLC12A7 | protein_coding | rs4285270 | 1113556 | 5.88e-10 | 3 | 0 | 0 | 0 | 0 | 3 | Female only | Both sexes | Female only | True | False |
| RDW | 18 | C18orf25 | protein_coding | rs8095729 | 43741982 | 7.95e-10 | 55 | 0 | 0 | 0 | 0 | 55 | Both sexes | Both sexes | Female only | True | False |
| RDW | 6 | SLC17A2 | protein_coding | rs3799369 | 25912634 | 1.13e-09 | 51 | 0 | 0 | 0 | 3 | 48 | Both sexes | Both sexes | Female only | True | False |
| RDW | 22 | TMPRSS6 | protein_coding | rs9607412 | 37484337 | 1.42e-09 | 57 | 0 | 24 | 5 | 0 | 28 | Female only | Both sexes | Female only | True | False |
| RDW | 6 | SLC17A1 | protein_coding | rs12191655 | 25803095 | 1.64e-09 | 297 | 28 | 118 | 5 | 8 | 138 | Female only | Both sexes | Female only | True | False |
| RDW | 6 | LRRC16A | protein_coding | rs4712925 | 25389334 | 1.81e-09 | 193 | 0 | 0 | 13 | 62 | 118 | Female only | Both sexes | Female only | True | False |
| RDW | 11 | SOX6 | protein_coding | rs74979599 | 16369353 | 2.17e-09 | 39 | 20 | 0 | 0 | 0 | 19 | Female only | Female only | Female only | True | False |
| RDW | 16 | PIEZO1 | protein_coding | rs837763 | 88853729 | 2.33e-09 | 17 | 9 | 0 | 2 | 0 | 6 | Female only | Female only | Female only | True | False |
| RDW | 6 | MIR3143 | miRNA | rs71559014 | 27122444 | 4.40e-09 | 3 | 0 | 0 | 0 | 0 | 3 | Female only | Both sexes | Female only | True | False |
| RDW | 5 | SLC12A7 | protein_coding | rs13176210 | 1081668 | 6.69e-09 | 15 | 0 | 4 | 1 | 6 | 4 | Female only | Both sexes | Female only | True | False |
| RDW | 5 | FBN2 | protein_coding | rs2042327 | 127622983 | 8.04e-09 | 19 | 6 | 4 | 5 | 1 | 3 | Female only | Female only | Female only | True | False |

| pheno | CHR | Gene_Symbol | biotype | lead_snp | lead_BP | lead_P | TOTAL | NSIG | S05 | S01 | S001 | S0001 | independent_loci_in_sex_P1e6 | independent_loci_in_sex_P1e2 | independent_loci_in_sex_P5e8 | in_gwas_catalog | in_HLA |
|---|---|---|---|---|---|---|---|---|---|---|---|---|---|---|---|---|---|
| RDW | 5 | SLC12A2 | protein_coding | rs3805603 | 127451581 | 8.73e-09 | 37 | 0 | 10 | 9 | 1 | 17 | Female only | Both sexes | Female only | True | False |
| RDW | 6 | LRRC16A | protein_coding | rs116790746 | 25348745 | 1.14e-08 | 51 | 0 | 3 | 13 | 10 | 25 | Female only | Both sexes | Female only | True | False |
| RDW | 6 | HIST1H2AA | protein_coding | rs3936052 | 25715534 | 1.18e-08 | 37 | 12 | 0 | 2 | 11 | 12 | Female only | Both sexes | Female only | True | False |
| RDW | 22 | MPST | protein_coding | rs5756488 | 37416803 | 2.02e-08 | 7 | 2 | 0 | 1 | 1 | 3 | Female only | Both sexes | Female only | True | False |
| RDW | 6 | HCG4P7 | pseudogene | rs1611657 | 29842444 | 2.39e-08 | 155 | 78 | 0 | 0 | 74 | 3 | Female only | Female only | Female only | True | True |
| RDW | 6 | LRRC16A | protein_coding | rs9358860 | 25572245 | 3.28e-08 | 55 | 46 | 3 | 1 | 1 | 4 | Female only | Both sexes | Female only | True | False |
| RDW | 5 | SLC12A7 | protein_coding | rs11746711 | 1117045 | 4.27e-08 | 3 | 0 | 0 | 0 | 1 | 2 | Both sexes | Both sexes | Female only | True | False |
| RDW | 6 | GPX6 | protein_coding | rs34979126 | 28449380 | 4.28e-08 | 41 | 1 | 4 | 16 | 4 | 16 | Female only | Both sexes | Female only | True | False |
| RDWSD | 1 | ATP2B4 | protein_coding | rs10900588 | 203656814 | 1.89e-21 | 3 | 0 | 0 | 0 | 0 | 3 | Both sexes | Both sexes | Both sexes | True | False |
| RDWSD | 16 | PIEZO1 | protein_coding | rs837763 | 88853729 | 3.63e-21 | 17 | 2 | 4 | 2 | 1 | 8 | Female only | Both sexes | Female only | True | False |
| RDWSD | 5 | CTC-228N24.3 | lincRNA | rs2250127 | 127388844 | 5.28e-19 | 85 | 0 | 0 | 0 | 0 | 85 | Both sexes | Both sexes | Both sexes | True | False |
| RDWSD | 16 | PIEZO1 | protein_coding | rs2926775 | 88858246 | 9.61e-14 | 29 | 5 | 6 | 3 | 3 | 12 | Female only | Both sexes | Female only | True | False |
| RDWSD | 15 | DNAJA4 | protein_coding | rs113594982 | 78557392 | 3.46e-12 | 47 | 0 | 0 | 1 | 16 | 30 | Female only | Both sexes | Female only | True | False |
| RDWSD | 9 | ABO | processed_transcript | rs8176741 | 136131461 | 5.65e-12 | 59 | 0 | 0 | 8 | 2 | 49 | Female only | Both sexes | Female only | True | False |
| RDWSD | 11 | SOX6 | protein_coding | rs74979599 | 16369353 | 8.95e-12 | 39 | 20 | 0 | 0 | 0 | 19 | Female only | Female only | Female only | True | False |
| RDWSD | 16 | PIEZO1 | protein_coding | rs9932423 | 88846849 | 1.08e-11 | 5 | 3 | 0 | 0 | 0 | 2 | Female only | Female only | Female only | True | False |
| RDWSD | 6 | HBS1L | protein_coding | rs9376091 | 135419636 | 5.33e-11 | 35 | 0 | 0 | 0 | 0 | 35 | Both sexes | Both sexes | Both sexes | True | False |
| RDWSD | 19 | KCNN4 | protein_coding | rs11351 | 44271066 | 3.37e-10 | 59 | 0 | 0 | 2 | 23 | 34 | Female only | Both sexes | Female only | True | False |
| RDWSD | 6 | HBS1L | protein_coding | rs9389272 | 135459837 | 1.61e-09 | 35 | 2 | 0 | 8 | 8 | 17 | Female only | Both sexes | Female only | True | False |
| RDWSD | 15 | WDR61 | protein_coding | rs4887030 | 78599995 | 2.66e-09 | 25 | 0 | 2 | 0 | 0 | 23 | Both sexes | Both sexes | Both sexes | True | False |
| RDWSD | 15 | RP11-762H8.3 | antisense | rs8039884 | 78550081 | 3.24e-09 | 25 | 0 | 0 | 0 | 0 | 25 | Both sexes | Both sexes | Both sexes | True | False |
| RDWSD | 15 | WDR61 | protein_coding | rs6495287 | 78594447 | 5.34e-09 | 39 | 3 | 2 | 1 | 8 | 25 | Both sexes | Both sexes | Female only | True | False |
| RDWSD | 11 | FADS2 | protein_coding | rs174559 | 61581656 | 7.64e-09 | 95 | 0 | 0 | 0 | 5 | 90 | Both sexes | Both sexes | Both sexes | True | False |
| RDWSD | 10 | HK1 | protein_coding | rs72805692 | 71099109 | 8.14e-09 | 11 | 0 | 0 | 0 | 4 | 7 | Female only | Both sexes | Female only | True | False |
| pheno | CHR | Gene_Symbol | biotype | lead_snp | lead_BP | lead_P | TOTAL | NSIG | S05 | S01 | S001 | S0001 | independent_loci_in_sex_P1e6 | independent_loci_in_sex_P1e2 | independent_loci_in_sex_P5e8 | in_gwas_catalog | in_HLA |
| RDWSD | 5 | SLC12A2 | protein_coding | rs809841 | 127460082 | 1.02e-08 | 45 | 0 | 0 | 2 | 14 | 29 | Male only | Both sexes | Male only | True | False |

| phen o | CHR | Gene_Sy mbol | biotype | lead_snp | lead_BP | lead_P | TOT AL | NSIG | S05 | S01 | S001 | S000 1 | independent_loci_in_sex_P1 e6 | independent_loci_in_sex_P1 e2 | independent_loci_in_sex_P5 e8 | in_gwas_catal og | in_H LA |
|---|---|---|---|---|---|---|---|---|---|---|---|---|---|---|---|---|---|
| RDW SD | 7 | AVL9 | protein_coding | rs7793408 | 3300955 6 | 1.21e-0 8 | 45 | 0 | 4 | 11 | 8 | 22 | Female only | Both sexes | Female only | True | False |
| RDW SD | 9 | ABO | processed_transcript | rs1090125 6 | 1361634 47 | 1.45e-0 8 | 23 | 12 | 6 | 0 | 0 | 5 | Female only | Female only | Female only | True | False |
| RDW SD | 2 | DPP10 | protein_coding | rs4849439 | 1168578 16 | 1.93e-0 8 | 33 | 17 | 0 | 0 | 0 | 16 | Male only | Male only | Male only | True | False |
| RDW SD | 16 | CTU2 | protein_coding | rs1107670 4 | 8877799 5 | 2.20e-0 8 | 11 | 5 | 0 | 1 | 0 | 5 | Female only | Both sexes | Female only | True | False |
| RDW SD | 1 | ATP2B4 | protein_coding | rs4951073 | 2036584 86 | 3.90e-0 8 | 9 | 8 | 0 | 0 | 1 | 0 | Female only | Both sexes | Female only | True | False |
| RR.I NTE RVAL | 13 | MTUS2 | protein_coding | rs4768988 | 2954681 8 | 2.71e-0 8 | 9 | 5 | 0 | 0 | 0 | 4 | Female only | Female only | Female only | False | False |
| R_F | 7 | AC004875. 1 | miRNA | rs7371980 6 | 1211898 29 | 2.53e-0 8 | 67 | 34 | 0 | 0 | 0 | 33 | Male only | Male only | Male only | True | False |
| SGO T | 22 | PNPLA3 | protein_coding | rs3747207 | 4432485 5 | 3.27e-1 9 | 113 | 0 | 0 | 0 | 0 | 113 | Both sexes | Both sexes | Both sexes | True | False |
| SGO T | 22 | SAMM50 | protein_coding | rs2235778 | 4438951 4 | 6.08e-0 9 | 45 | 0 | 0 | 13 | 0 | 32 | Both sexes | Both sexes | Male only | True | False |
| SGO T | 22 | SAMM50 | protein_coding | rs1216784 5 | 4438076 7 | 1.85e-0 8 | 23 | 0 | 0 | 0 | 0 | 23 | Both sexes | Both sexes | Both sexes | True | False |
| SGO T | 10 | SLC39A12 | protein_coding | rs508196 | 1826387 3 | 2.00e-0 8 | 29 | 5 | 9 | 1 | 0 | 14 | Female only | Both sexes | Female only | True | False |
| SGO T | 12 | CUX2 | protein_coding | rs6490048 | 1117413 15 | 3.60e-0 8 | 29 | 15 | 0 | 0 | 0 | 14 | Female only | Female only | Female only | True | False |
| SGP T | 22 | PNPLA3 | protein_coding | rs3747207 | 4432485 5 | 3.34e-1 2 | 113 | 0 | 0 | 0 | 0 | 113 | Both sexes | Both sexes | Both sexes | True | False |
| SSAI gG | 6 | HLA-DQA1 | protein_coding | rs9272461 | 3260560 9 | 4.93e-0 9 | 135 | 68 | 0 | 0 | 0 | 67 | Female only | Female only | Female only | False | True |
| SU.Pr o | 5 | RNU6-525 P | snRNA | rs1173925 5 | 1798872 37 | 3.27e-0 8 | 3 | 2 | 0 | 0 | 1 | 0 | Male only | Male only | Male only | True | False |
| SYST OLIC | 12 | ATP2B1 | protein_coding | rs1110536 4 | 9006927 6 | 4.40e-1 2 | 39 | 0 | 8 | 12 | 0 | 19 | Female only | Both sexes | Female only | True | False |
| SYST OLIC | 12 | ATP2B1 | protein_coding | rs7136259 | 9008118 8 | 1.09e-1 0 | 17 | 4 | 5 | 0 | 0 | 8 | Female only | Female only | Female only | True | False |
| SedR at | 1 | CR1 | protein_coding | rs6540435 | 2076857 64 | 4.11e-2 7 | 107 | 0 | 0 | 0 | 0 | 107 | Both sexes | Both sexes | Both sexes | True | False |
| SedR at | 1 | CR1 | protein_coding | rs6182297 7 | 2077960 65 | 8.26e-2 5 | 23 | 0 | 0 | 0 | 0 | 23 | Both sexes | Both sexes | Both sexes | True | False |
| SedR at | 1 | TMEM57 | protein_coding | rs7266096 1 | 2581845 2 | 6.44e-1 4 | 245 | 0 | 0 | 0 | 23 | 222 | Both sexes | Both sexes | Female only | True | False |
| SedR at | 1 | CR1L | protein_coding | rs1274410 4 | 2078190 00 | 2.07e-0 8 | 101 | 65 | 4 | 18 | 0 | 14 | Female only | Both sexes | Female only | True | False |
| TBil | 2 | UGT1A11 P | pseudogene | rs2741012 | 2345089 63 | 0 | 73 | 0 | 0 | 0 | 0 | 73 | Both sexes | Both sexes | Both sexes | True | False |
| TBil | 2 | UGT1A8 | protein_coding | rs1019746 0 | 2345891 90 | 0 | 161 | 0 | 0 | 0 | 0 | 161 | Both sexes | Both sexes | Both sexes | True | False |
| TBil | 2 | UGT1A8 | protein_coding | rs7592624 | 2346029 06 | 2.12e-2 37 | 35 | 0 | 0 | 0 | 0 | 35 | Both sexes | Both sexes | Both sexes | True | False |
| TBil | 2 | UGT1A8 | protein_coding | rs6751673 | 2346049 03 | 2.13e-1 46 | 31 | 0 | 0 | 0 | 0 | 31 | Both sexes | Both sexes | Both sexes | True | False |

| pheno | CHR | Gene_Symbol | biotype | lead_snp | lead_BP | lead_P | TOTAL | NSIG | S05 | S01 | S001 | S0001 | independent_loci_in_sex_P1e6 | independent_loci_in_sex_P1e2 | independent_loci_in_sex_P5e8 | in_gwas_catalog | in_HLA |
|---|---|---|---|---|---|---|---|---|---|---|---|---|---|---|---|---|---|
| TBil | 2 | MROH2A | protein_coding | rs4663336 | 234692932 | 2.52e-93 | 5 | 0 | 0 | 0 | 0 | 5 | Both sexes | Both sexes | Both sexes | True | False |
| TBil | 2 | UGT1A8 | protein_coding | rs11123100 | 234567916 | 1.07e-80 | 25 | 0 | 0 | 0 | 0 | 25 | Both sexes | Both sexes | Both sexes | True | False |
| TBil | 2 | UGT1A8 | protein_coding | rs35203651 | 234679549 | 6.02e-68 | 25 | 0 | 0 | 0 | 0 | 25 | Both sexes | Both sexes | Both sexes | True | False |
| TBil | 2 | MROH2A | protein_coding | rs1587493 | 234689751 | 8.83e-65 | 21 | 0 | 0 | 0 | 0 | 21 | Both sexes | Both sexes | Both sexes | True | False |
| TBil | 2 | UGT1A8 | protein_coding | rs12479240 | 234570829 | 6.75e-62 | 45 | 0 | 0 | 0 | 0 | 45 | Both sexes | Both sexes | Both sexes | True | False |
| TBil | 2 | UGT1A8 | protein_coding | rs11568318 | 234665498 | 9.70e-62 | 91 | 0 | 0 | 0 | 0 | 91 | Both sexes | Both sexes | Both sexes | True | False |
| TBil | 2 | USP40 | protein_coding | rs72982316 | 234416719 | 2.18e-52 | 87 | 0 | 0 | 0 | 0 | 87 | Both sexes | Both sexes | Both sexes | True | False |
| TBil | 2 | USP40 | protein_coding | rs62192764 | 234440780 | 3.01e-38 | 13 | 0 | 0 | 0 | 0 | 13 | Both sexes | Both sexes | Both sexes | True | False |
| TBil | 12 | SLCO1B1 | protein_coding | rs4149056 | 21331549 | 1.60e-34 | 87 | 0 | 0 | 0 | 0 | 87 | Both sexes | Both sexes | Both sexes | True | False |
| TBil | 2 | ATG16L1 | protein_coding | rs62192926 | 234148737 | 1.92e-30 | 5 | 0 | 0 | 0 | 0 | 5 | Both sexes | Both sexes | Both sexes | True | False |
| TBil | 2 | DGKD | protein_coding | rs838705 | 234273242 | 6.01e-26 | 11 | 0 | 0 | 0 | 0 | 11 | Both sexes | Both sexes | Both sexes | True | False |
| TBil | 2 | SAG | protein_coding | rs3792095 | 234238949 | 1.15e-24 | 45 | 2 | 0 | 10 | 4 | 29 | Both sexes | Both sexes | Both sexes | True | False |
| TBil | 2 | MROH2A | protein_coding | rs2041651 | 234713812 | 5.71e-24 | 3 | 0 | 0 | 0 | 0 | 3 | Both sexes | Both sexes | Both sexes | True | False |
| TBil | 2 | MROH2A | protein_coding | rs6716168 | 234713429 | 8.13e-22 | 13 | 0 | 0 | 6 | 6 | 1 | Both sexes | Both sexes | Both sexes | True | False |
| TBil | 12 | SLCO1B1 | protein_coding | rs11045856 | 21350689 | 3.09e-21 | 143 | 2 | 0 | 0 | 0 | 141 | Both sexes | Both sexes | Both sexes | True | False |
| TBil | 2 | DGKD | protein_coding | rs838721 | 234303405 | 2.83e-20 | 15 | 5 | 0 | 0 | 0 | 10 | Both sexes | Both sexes | Female only | True | False |
| TBil | 2 | TRPM8 | protein_coding | rs17868378 | 234808378 | 6.21e-19 | 29 | 0 | 0 | 0 | 0 | 29 | Both sexes | Both sexes | Both sexes | True | False |
| TBil | 12 | SLCO1B1 | protein_coding | rs4149057 | 21331599 | 7.63e-18 | 5 | 4 | 0 | 0 | 0 | 1 | Both sexes | Both sexes | Female only | True | False |
| TBil | 2 | SAG | protein_coding | rs11891546 | 234232722 | 1.72e-17 | 59 | 0 | 0 | 1 | 29 | 29 | Both sexes | Both sexes | Both sexes | True | False |
| TBil | 12 | RP11-125O5.2 | protein_coding | rs10841753 | 21321370 | 9.29e-17 | 199 | 164 | 2 | 0 | 0 | 33 | Both sexes | Both sexes | Both sexes | True | False |
| TBil | 2 | MROH2A | protein_coding | rs726017 | 234725819 | 5.61e-16 | 11 | 0 | 0 | 0 | 0 | 11 | Both sexes | Both sexes | Both sexes | True | False |
| TBil | 2 | DGKD | protein_coding | rs34847903 | 234268714 | 2.86e-14 | 23 | 16 | 0 | 0 | 0 | 7 | Both sexes | Both sexes | Both sexes | True | False |
| TBil | 2 | DGKD | protein_coding | rs2305542 | 234354245 | 6.25e-14 | 29 | 0 | 0 | 0 | 0 | 29 | Both sexes | Both sexes | Female only | True | False |
| TBil | 2 | MROH2A | protein_coding | rs11674078 | 234718524 | 3.99e-13 | 9 | 8 | 0 | 0 | 0 | 1 | Both sexes | Both sexes | Female only | True | False |
| TBil | 12 | SLCO1B3 | protein_coding | rs4141475 | 20951861 | 6.38e-13 | 261 | 0 | 0 | 0 | 0 | 261 | Both sexes | Both sexes | Both sexes | True | False |
| TBil | 2 | SAG | protein_coding | rs3792094 | 234243150 | 1.51e-12 | 9 | 0 | 3 | 2 | 0 | 4 | Female only | Both sexes | Female only | True | False |

| pheno | CHR | Gene_Symbol | biotype | lead_snp | lead_BP | lead_P | TOTAL | NSIG | S05 | S01 | S001 | S0001 | independent_loci_in_sex_P1e6 | independent_loci_in_sex_P1e2 | independent_loci_in_sex_P5e8 | in_gwas_catalog | in_HLA |
|---|---|---|---|---|---|---|---|---|---|---|---|---|---|---|---|---|---|
| TBil | 2 | UGT1A8 | protein_coding | rs2012736 | 234622379 | 2.75e-12 | 131 | 27 | 4 | 0 | 9 | 91 | Female only | Both sexes | Female only | True | False |
| TBil | 2 | HJURP | protein_coding | rs566529 | 234756811 | 7.70e-12 | 105 | 2 | 0 | 0 | 1 | 102 | Both sexes | Both sexes | Female only | True | False |
| TBil | 12 | SLCO1A2 | protein_coding | rs2199686 | 21451028 | 1.52e-11 | 77 | 0 | 0 | 0 | 0 | 77 | Both sexes | Both sexes | Both sexes | True | False |
| TBil | 2 | DGKD | protein_coding | rs78447894 | 234315394 | 5.99e-11 | 43 | 7 | 2 | 9 | 1 | 24 | Both sexes | Both sexes | Female only | True | False |
| TBil | 2 | DGKD | protein_coding | rs10185174 | 234266409 | 1.03e-10 | 67 | 29 | 37 | 0 | 0 | 1 | Both sexes | Both sexes | Both sexes | True | False |
| TBil | 12 | RP11-125O5.2 | protein_coding | rs4149042 | 21328872 | 1.73e-10 | 13 | 0 | 0 | 0 | 0 | 13 | Female only | Both sexes | Female only | True | False |
| TBil | 2 | USP40 | protein_coding | rs1048603 | 234394487 | 8.19e-10 | 63 | 1 | 23 | 5 | 6 | 28 | Female only | Both sexes | Female only | True | False |
| TBil | 22 | PNPLA3 | protein_coding | rs3747207 | 44324855 | 1.10e-09 | 113 | 0 | 8 | 47 | 1 | 57 | Male only | Both sexes | Male only | True | False |
| pheno | CHR | Gene_Symbol | biotype | lead_snp | lead_BP | lead_P | TOTAL | NSIG | S05 | S01 | S001 | S0001 | independent_loci_in_sex_P1e6 | independent_loci_in_sex_P1e2 | independent_loci_in_sex_P5e8 | in_gwas_catalog | in_HLA |
| TBil | 2 | TRPM8 | protein_coding | rs4663344 | 234867009 | 2.93e-09 | 87 | 25 | 1 | 4 | 9 | 48 | Both sexes | Both sexes | Both sexes | True | False |
| TBil | 2 | MSL3P1 | pseudogene | rs2361515 | 234791098 | 4.49e-09 | 99 | 42 | 1 | 5 | 19 | 32 | Female only | Both sexes | Female only | True | False |
| TBil | 12 | SLCO1A2 | protein_coding | rs11045930 | 21432829 | 4.66e-09 | 115 | 0 | 36 | 20 | 2 | 57 | Female only | Both sexes | Female only | True | False |
| TBil | 2 | SAG | protein_coding | rs2304775 | 234227699 | 9.09e-09 | 25 | 2 | 11 | 4 | 7 | 1 | Both sexes | Both sexes | Both sexes | True | False |
| TBil | 2 | ATG16L1 | protein_coding | rs1045100 | 234203597 | 9.13e-09 | 75 | 2 | 4 | 33 | 27 | 9 | Female only | Both sexes | Female only | True | False |
| TBil | 2 | UGT1A8 | protein_coding | rs28444828 | 234557414 | 1.67e-08 | 11 | 0 | 0 | 0 | 0 | 11 | Female only | Both sexes | Female only | True | False |
| TBil | 2 | DGKD | protein_coding | rs2242102 | 234346660 | 2.56e-08 | 5 | 2 | 0 | 1 | 1 | 1 | Female only | Both sexes | Female only | True | False |
| TBil | 2 | TRPM8 | protein_coding | rs10929321 | 234871343 | 2.95e-08 | 3 | 0 | 1 | 1 | 1 | 0 | Female only | Both sexes | Female only | True | False |
| TBil | 2 | TRPM8 | protein_coding | rs17869072 | 234918905 | 4.17e-08 | 59 | 28 | 2 | 8 | 2 | 19 | Female only | Female only | Female only | True | False |
| TBil | 12 | SLCO1B1 | protein_coding | rs113830923 | 21397321 | 4.39e-08 | 3 | 2 | 0 | 0 | 0 | 1 | Female only | Both sexes | Female only | True | False |
| TC.HDL | 16 | AC012181.1 | miRNA | rs12923459 | 56989830 | 1.36e-09 | 5 | 0 | 0 | 0 | 3 | 2 | Female only | Both sexes | Female only | True | False |
| TC.HDL | 16 | AC012181.1 | miRNA | rs3764261 | 56993324 | 1.80e-08 | 41 | 0 | 0 | 11 | 19 | 11 | Female only | Both sexes | Female only | True | False |
| TC.HDL | 16 | CETP | protein_coding | rs7203984 | 56999258 | 2.37e-08 | 13 | 0 | 0 | 0 | 3 | 10 | Male only | Both sexes | Male only | True | False |
| TERMINAL_40_MS | 1 | NFIA | protein_coding | rs2474370 | 61886891 | 1.59e-08 | 15 | 0 | 8 | 0 | 0 | 7 | Female only | Female only | Female only | True | False |
| TERMINAL_40_MS | 1 | NFIA | protein_coding | rs6587924 | 61895257 | 4.16e-08 | 27 | 0 | 0 | 2 | 12 | 13 | Female only | Both sexes | Female only | True | False |

| pheno | CHR | Gene_Symbol | biotype | lead_snp | lead_BP | lead_P | TOTAL | NSIG | S05 | S01 | S001 | S0001 | independent_loci_in_sex_P1e6 | independent_loci_in_sex_P1e2 | independent_loci_in_sex_P5e8 | in_gwas_catalog | in_HLA |
|---|---|---|---|---|---|---|---|---|---|---|---|---|---|---|---|---|---|
| TERMINAL_40_MS | 12 | RPL7P6 | pseudogene | rs34900245 | 19212108 | 4.71e-08 | 35 | 18 | 0 | 0 | 0 | 17 | Female only | Female only | Female only | True | False |
| TIBC | 3 | TF | protein_coding | rs4854761 | 133498789 | 8.29e-17 | 111 | 0 | 0 | 28 | 7 | 76 | Male only | Both sexes | Male only | True | False |
| TIBC | 3 | TF | protein_coding | rs13326361 | 133500294 | 4.98e-11 | 9 | 2 | 3 | 0 | 0 | 4 | Male only | Male only | Male only | True | False |
| TIBC | 3 | TF | protein_coding | rs1880669 | 133483696 | 7.51e-11 | 33 | 11 | 6 | 0 | 0 | 16 | Male only | Male only | Male only | True | False |
| TIBC | 3 | TF | protein_coding | rs9880615 | 133470909 | 2.29e-08 | 7 | 4 | 0 | 0 | 0 | 3 | Male only | Male only | Male only | True | False |
| TProSe | 6 | MICB | protein_coding | rs2894222 | 31462035 | 3.25e-11 | 49 | 25 | 0 | 0 | 9 | 15 | Male only | Male only | Male only | True | True |
| TProSe | 6 | HLA-DQB1 | protein_coding | rs4988889 | 32634952 | 2.22e-10 | 35 | 0 | 0 | 11 | 6 | 18 | Male only | Both sexes | Male only | True | True |
| TProSe | 6 | HLA-B | protein_coding | rs9265969 | 31315706 | 4.26e-10 | 261 | 0 | 0 | 31 | 90 | 140 | Male only | Both sexes | Male only | True | True |
| TProSe | 6 | GPSM3 | protein_coding | rs204989 | 32161852 | 4.59e-10 | 35 | 0 | 0 | 6 | 12 | 17 | Male only | Both sexes | Male only | True | True |
| TProSe | 6 | HLA-N | pseudogene | rs3094061 | 30321189 | 1.77e-09 | 329 | 0 | 0 | 56 | 105 | 168 | Male only | Both sexes | Male only | True | True |
| TProSe | 6 | GPANK1 | protein_coding | rs3117579 | 31633496 | 3.12e-09 | 129 | 0 | 0 | 54 | 11 | 64 | Male only | Both sexes | Male only | True | True |
| TProSe | 6 | HLA-F-AS1 | processed_transcript | rs1633088 | 29713580 | 3.48e-09 | 403 | 66 | 130 | 8 | 8 | 191 | Male only | Both sexes | Male only | True | True |
| TProSe | 6 | HCP5 | sense_overlapping | rs3128981 | 31405620 | 3.98e-09 | 35 | 0 | 3 | 4 | 10 | 18 | Male only | Both sexes | Male only | True | True |
| TProSe | 6 | NOTCH4 | protein_coding | rs3132935 | 32171075 | 7.66e-09 | 45 | 0 | 0 | 0 | 9 | 36 | Male only | Both sexes | Male only | True | True |
| TProSe | 6 | NOTCH4 | protein_coding | rs693797 | 32204433 | 1.01e-08 | 25 | 13 | 1 | 0 | 0 | 11 | Male only | Male only | Male only | True | True |
| TProSe | 6 | TRIM26 | protein_coding | rs2517611 | 30169327 | 1.09e-08 | 39 | 0 | 0 | 19 | 1 | 19 | Male only | Both sexes | Male only | True | True |
| TProSe | 6 | HLA-B | protein_coding | rs2596498 | 31322688 | 1.13e-08 | 11 | 1 | 1 | 3 | 2 | 4 | Male only | Both sexes | Male only | True | True |
| TProSe | 6 | STK19 | protein_coding | rs389884 | 31940897 | 1.55e-08 | 39 | 0 | 5 | 5 | 10 | 19 | Male only | Both sexes | Male only | True | True |
| TProSe | 6 | RPL15P4 | pseudogene | rs2734573 | 31494738 | 1.63e-08 | 3 | 0 | 1 | 1 | 0 | 1 | Male only | Both sexes | Male only | True | True |
| TProSe | 6 | MUC22 | protein_coding | rs2517546 | 31009055 | 1.79e-08 | 171 | 0 | 1 | 33 | 80 | 57 | Male only | Both sexes | Male only | True | True |
| TProSe | 6 | MICB | protein_coding | rs2395043 | 31461966 | 2.50e-08 | 17 | 8 | 1 | 1 | 1 | 6 | Male only | Male only | Male only | True | True |
| TProSe | 6 | PRRC2A | protein_coding | rs2261033 | 31603591 | 4.88e-08 | 11 | 6 | 0 | 0 | 3 | 2 | Male only | Male only | Male only | True | True |
| TProt | 6 | HLA-DQB1 | protein_coding | rs4988889 | 32634952 | 1.41e-13 | 35 | 0 | 0 | 0 | 12 | 23 | Both sexes | Both sexes | Both sexes | True | True |
| TProt | 6 | NOTCH4 | protein_coding | rs6901158 | 32205942 | 1.07e-12 | 129 | 0 | 0 | 2 | 13 | 114 | Both sexes | Both sexes | Male only | True | True |
| TProt | 6 | GPANK1 | protein_coding | rs3117579 | 31633496 | 2.14e-12 | 129 | 0 | 0 | 0 | 0 | 129 | Both sexes | Both sexes | Male only | True | True |

| pheno | CHR | Gene_Symbol | biotype | lead_snp | lead_BP | lead_P | TOTAL | NSIG | S05 | S01 | S001 | S0001 | independent_loci_in_sex_P1e6 | independent_loci_in_sex_P1e2 | independent_loci_in_sex_P5e8 | in_gwas_catalog | in_HLA |
|---|---|---|---|---|---|---|---|---|---|---|---|---|---|---|---|---|---|
| TProt | 6 | HLA-DQB1 | protein_coding | rs9273508 | 32628439 | 6.46e-11 | 11 | 0 | 0 | 3 | 1 | 7 | Both sexes | Both sexes | Both sexes | True | True |
| TProt | 6 | HLA-C | protein_coding | rs3130444 | 31228634 | 8.35e-11 | 167 | 0 | 7 | 0 | 45 | 115 | Male only | Both sexes | Male only | True | True |
| TProt | 6 | C2 | protein_coding | rs497309 | 31892484 | 1.41e-10 | 27 | 0 | 3 | 7 | 0 | 17 | Male only | Both sexes | Male only | True | True |
| TProt | 6 | RNU6-283P | snRNA | rs2523524 | 31339996 | 1.73e-10 | 293 | 3 | 3 | 22 | 24 | 241 | Both sexes | Both sexes | Male only | True | True |
| TProt | 6 | SFTA2 | protein_coding | rs3095153 | 30899195 | 2.80e-10 | 249 | 0 | 0 | 0 | 0 | 249 | Both sexes | Both sexes | Both sexes | True | True |
| TProt | 17 | TNFRSF13B | protein_coding | rs4792800 | 16845167 | 5.76e-10 | 61 | 0 | 1 | 1 | 5 | 54 | Female only | Both sexes | Female only | True | False |
| TProt | 6 | POU5F1 | protein_coding | rs885948 | 31140532 | 8.59e-10 | 53 | 0 | 0 | 3 | 12 | 38 | Male only | Both sexes | Male only | True | True |
| TProt | 6 | HCP5 | sense_overlapping | rs3128981 | 31405620 | 1.19e-09 | 35 | 0 | 0 | 6 | 17 | 12 | Both sexes | Both sexes | Male only | True | True |
| TProt | 6 | HLA-DQB1 | protein_coding | 6:32623223 | 32623223 | 1.43e-09 | 85 | 8 | 6 | 14 | 19 | 38 | Male only | Both sexes | Male only | True | True |
| TProt | 6 | HLA-N | pseudogene | rs3130375 | 30321732 | 2.12e-09 | 329 | 0 | 0 | 0 | 11 | 318 | Both sexes | Both sexes | Both sexes | True | True |
| TProt | 6 | HLA-DRB9 | pseudogene | rs4601112 | 32426271 | 2.91e-09 | 257 | 58 | 3 | 114 | 9 | 73 | Male only | Both sexes | Male only | True | True |
| TProt | 1 | FCGR2A | protein_coding | rs7518087 | 161477816 | 3.45e-09 | 45 | 0 | 0 | 0 | 0 | 45 | Both sexes | Both sexes | Male only | True | False |
| TProt | 6 | HLA-C | protein_coding | rs9264186 | 31220017 | 4.05e-09 | 109 | 0 | 0 | 2 | 53 | 54 | Male only | Both sexes | Male only | True | True |
| TProt | 6 | TRIM26 | protein_coding | rs2517611 | 30169327 | 4.18e-09 | 39 | 0 | 0 | 0 | 1 | 38 | Both sexes | Both sexes | Male only | True | True |
| TProt | 6 | PRRT1 | protein_coding | rs204999 | 32109979 | 6.02e-09 | 37 | 0 | 0 | 0 | 0 | 37 | Male only | Both sexes | Male only | True | True |
| TProt | 6 | PSORS1C1 | protein_coding | rs528878831 | 31098832 | 7.51e-09 | 97 | 0 | 11 | 23 | 21 | 42 | Male only | Both sexes | Male only | True | True |
| TProt | 6 | NOTCH4 | protein_coding | rs3132935 | 32171075 | 1.68e-08 | 7 | 0 | 0 | 0 | 0 | 7 | Male only | Both sexes | Male only | True | True |
| TProt | 11 | FADS2 | protein_coding | rs28456 | 61589481 | 1.89e-08 | 89 | 5 | 38 | 2 | 1 | 43 | Female only | Both sexes | Female only | True | False |
| TProt | 6 | NOTCH4 | protein_coding | rs715299 | 32189841 | 2.08e-08 | 3 | 0 | 0 | 0 | 1 | 2 | Male only | Both sexes | Male only | True | True |
| TProt | 6 | RNF39 | protein_coding | rs9261290 | 30038647 | 2.89e-08 | 127 | 0 | 0 | 30 | 32 | 65 | Male only | Both sexes | Male only | True | True |
| pheno | CHR | Gene_Symbol | biotype | lead_snp | lead_BP | lead_P | TOTAL | NSIG | S05 | S01 | S001 | S0001 | independent_loci_in_sex_P1e6 | independent_loci_in_sex_P1e2 | independent_loci_in_sex_P5e8 | in_gwas_catalog | in_HLA |
| TProt | 6 | ATAT1 | protein_coding | rs9262130 | 30603519 | 3.11e-08 | 21 | 0 | 0 | 2 | 3 | 16 | Both sexes | Both sexes | Male only | True | True |
| TProt | 6 | HCG22 | lincRNA | rs2517528 | 31021078 | 3.27e-08 | 19 | 0 | 0 | 0 | 10 | 9 | Male only | Both sexes | Male only | True | True |
| TProt | 19 | NOSIP | protein_coding | rs10406941 | 50091464 | 3.35e-08 | 23 | 0 | 0 | 3 | 0 | 20 | Male only | Both sexes | Male only | True | False |
| TProt | 6 | MICC | pseudogene | rs1264544 | 30382168 | 4.06e-08 | 73 | 0 | 0 | 0 | 47 | 26 | Female only | Both sexes | Female only | True | True |

| pheno | CHR | Gene_Symbol | biotype | lead_snp | lead_BP | lead_P | TOTAL | NSIG | S05 | S01 | S001 | S0001 | independent_loci_in_sex_P1e6 | independent_loci_in_sex_P1e2 | independent_loci_in_sex_P5e8 | in_gwas_catalog | in_HLA |
|---|---|---|---|---|---|---|---|---|---|---|---|---|---|---|---|---|---|
| TProt | 6 | XXbac-BPG248L24.12 | pseudogene | rs2249935 | 31327178 | 4.40e-08 | 3 | 0 | 0 | 1 | 1 | 1 | Male only | Both sexes | Male only | True | True |
| TSH | 9 | RP11-23B15.1 | lincRNA | rs1443438 | 100550028 | 6.18e-38 | 41 | 0 | 0 | 0 | 0 | 41 | Both sexes | Both sexes | Both sexes | True | False |
| TSH | 5 | PDE8B | protein_coding | rs1382879 | 76521868 | 8.35e-34 | 29 | 0 | 0 | 0 | 0 | 29 | Both sexes | Both sexes | Both sexes | True | False |
| TSH | 16 | RP11-345M22.1 | lincRNA | rs3813579 | 79749276 | 7.31e-24 | 7 | 0 | 0 | 0 | 0 | 7 | Both sexes | Both sexes | Both sexes | True | False |
| TSH | 6 | PDE10A | protein_coding | rs1079418 | 166047034 | 3.51e-23 | 33 | 0 | 0 | 0 | 0 | 33 | Both sexes | Both sexes | Both sexes | True | False |
| TSH | 9 | C9orf156 | protein_coding | rs7021160 | 100658123 | 2.32e-22 | 113 | 0 | 0 | 0 | 0 | 113 | Both sexes | Both sexes | Both sexes | True | False |
| TSH | 9 | ABO | processed_transcript | rs657152 | 136139265 | 1.33e-21 | 51 | 0 | 0 | 0 | 0 | 51 | Both sexes | Both sexes | Both sexes | True | False |
| TSH | 16 | RP11-345M22.1 | lincRNA | rs17767491 | 79745487 | 3.99e-21 | 135 | 0 | 0 | 0 | 0 | 135 | Both sexes | Both sexes | Both sexes | True | False |
| TSH | 9 | ABO | processed_transcript | rs649129 | 136154304 | 5.50e-18 | 17 | 0 | 0 | 0 | 0 | 17 | Both sexes | Both sexes | Female only | True | False |
| TSH | 9 | RP11-23B15.1 | lincRNA | 9:100542706 | 100542706 | 1.18e-17 | 3 | 0 | 0 | 1 | 1 | 1 | Female only | Both sexes | Female only | True | False |
| TSH | 6 | RP11-344J7.2 | lincRNA | rs729761 | 43804571 | 4.91e-17 | 11 | 0 | 0 | 0 | 0 | 11 | Both sexes | Both sexes | Both sexes | True | False |
| TSH | 19 | INSR | protein_coding | rs7248104 | 7224431 | 1.70e-15 | 25 | 0 | 0 | 5 | 4 | 16 | Male only | Both sexes | Male only | True | False |
| TSH | 1 | RNU4-28P | snRNA | rs10917477 | 19861106 | 1.79e-15 | 25 | 0 | 0 | 0 | 0 | 25 | Both sexes | Both sexes | Both sexes | True | False |
| TSH | 1 | CAPZB | protein_coding | rs3913439 | 19780509 | 1.97e-15 | 91 | 0 | 0 | 0 | 0 | 91 | Both sexes | Both sexes | Both sexes | True | False |
| TSH | 5 | PDE8B | protein_coding | rs1096752 | 76549558 | 3.90e-15 | 17 | 0 | 0 | 0 | 0 | 17 | Both sexes | Both sexes | Both sexes | True | False |
| TSH | 5 | PDE8B | protein_coding | rs1962690 | 76563277 | 5.40e-15 | 5 | 0 | 0 | 0 | 0 | 5 | Both sexes | Both sexes | Both sexes | True | False |
| TSH | 16 | RP11-345M22.1 | lincRNA | rs9923437 | 79710028 | 9.77e-15 | 3 | 0 | 0 | 0 | 0 | 3 | Both sexes | Both sexes | Both sexes | True | False |
| TSH | 5 | PDE8B | protein_coding | rs12515504 | 76540021 | 8.03e-14 | 5 | 0 | 0 | 0 | 0 | 5 | Both sexes | Both sexes | Both sexes | True | False |
| TSH | 4 | ATP5LP4 | pseudogene | rs13152267 | 149621320 | 1.03e-13 | 153 | 0 | 0 | 3 | 4 | 146 | Female only | Both sexes | Female only | True | False |
| TSH | 2 | AC007563.5 | antisense | rs6724363 | 217626591 | 1.32e-13 | 9 | 0 | 0 | 0 | 0 | 9 | Both sexes | Both sexes | Both sexes | True | False |
| TSH | 6 | RP5-1120P11.1 | antisense | rs11755845 | 43904780 | 1.49e-13 | 3 | 0 | 0 | 0 | 0 | 3 | Both sexes | Both sexes | Female only | True | False |
| TSH | 6 | PDE10A | protein_coding | rs12212126 | 166043091 | 1.65e-13 | 7 | 0 | 0 | 0 | 0 | 7 | Both sexes | Both sexes | Both sexes | True | False |
| TSH | 1 | RNU4-28P | snRNA | rs10799824 | 19841174 | 1.95e-13 | 55 | 0 | 0 | 0 | 11 | 44 | Both sexes | Both sexes | Both sexes | True | False |
| TSH | 5 | PDE8B | protein_coding | rs10474491 | 76520798 | 5.29e-13 | 13 | 0 | 0 | 0 | 2 | 11 | Both sexes | Both sexes | Both sexes | True | False |
| TSH | 20 | RP4-788L20.3 | lincRNA | rs1203944 | 22596879 | 1.98e-12 | 127 | 0 | 0 | 0 | 1 | 126 | Both sexes | Both sexes | Both sexes | True | False |

| pheno | CHR | Gene_Symbol | biotype | lead_snp | lead_BP | lead_P | TOTAL | NSIG | S05 | S01 | S001 | S0001 | independent_loci_in_sex_P1e6 | independent_loci_in_sex_P1e2 | independent_loci_in_sex_P5e8 | in_gwas_catalog | in_HLA |
|---|---|---|---|---|---|---|---|---|---|---|---|---|---|---|---|---|---|
| TSH | 1 | RNU4-28P | snRNA | rs12123411 | 19862592 | 4.19e-12 | 39 | 0 | 0 | 20 | 0 | 19 | Male only | Both sexes | Male only | True | False |
| TSH | 5 | PDE8B | protein_coding | rs6885813 | 76569611 | 4.98e-12 | 23 | 3 | 0 | 9 | 4 | 7 | Both sexes | Both sexes | Male only | True | False |
| TSH | 6 | PDE10A | protein_coding | rs3008041 | 166047241 | 6.04e-12 | 11 | 0 | 0 | 0 | 0 | 11 | Both sexes | Both sexes | Both sexes | True | False |
| TSH | 5 | ZBED3-AS1 | antisense | rs12171500 | 76460134 | 1.26e-11 | 5 | 2 | 2 | 0 | 0 | 1 | Male only | Both sexes | Male only | True | False |
| TSH | 6 | RP11-344J7.2 | lincRNA | rs6933958 | 43823689 | 2.04e-11 | 5 | 0 | 0 | 0 | 0 | 5 | Both sexes | Both sexes | Both sexes | True | False |
| TSH | 1 | VAV3 | protein_coding | rs75570391 | 108355694 | 2.89e-11 | 203 | 0 | 0 | 0 | 5 | 198 | Both sexes | Both sexes | Female only | True | False |
| TSH | 15 | FAM227B | protein_coding | rs17477923 | 49711185 | 5.79e-11 | 265 | 0 | 0 | 0 | 83 | 182 | Both sexes | Both sexes | Female only | True | False |
| TSH | 9 | ABO | processed_transcript | rs8176682 | 136139297 | 1.10e-10 | 25 | 0 | 0 | 0 | 2 | 23 | Female only | Both sexes | Female only | True | False |
| TSH | 9 | ABO | processed_transcript | rs647800 | 136148000 | 1.57e-10 | 19 | 0 | 0 | 1 | 1 | 17 | Female only | Both sexes | Female only | True | False |
| TSH | 5 | PDE8B | protein_coding | rs17570031 | 76524346 | 2.03e-10 | 7 | 0 | 0 | 0 | 0 | 7 | Both sexes | Both sexes | Both sexes | True | False |
| TSH | 6 | PDE10A | protein_coding | rs56228667 | 166038575 | 2.12e-10 | 73 | 0 | 36 | 0 | 1 | 36 | Female only | Both sexes | Female only | True | False |
| TSH | 4 | RP11-503L19.1 | lincRNA | rs72735922 | 149593414 | 3.09e-10 | 25 | 13 | 0 | 0 | 0 | 12 | Female only | Female only | Female only | True | False |
| TSH | 8 | NRG1 | protein_coding | rs4733130 | 32406994 | 3.13e-10 | 47 | 2 | 3 | 2 | 0 | 40 | Female only | Both sexes | Female only | True | False |
| TSH | 2 | AC007563.5 | antisense | rs2888429 | 217624194 | 5.45e-10 | 45 | 0 | 0 | 0 | 19 | 26 | Both sexes | Both sexes | Both sexes | True | False |
| TSH | 9 | C9orf156 | protein_coding | rs13301569 | 100655780 | 9.69e-10 | 17 | 0 | 0 | 0 | 9 | 8 | Female only | Both sexes | Female only | True | False |
| TSH | 15 | GALK2 | protein_coding | rs73390343 | 49456223 | 1.88e-09 | 23 | 0 | 0 | 1 | 10 | 12 | Female only | Both sexes | Female only | True | False |
| TSH | 5 | ZBED3-AS1 | antisense | rs7708949 | 76463481 | 2.47e-09 | 9 | 0 | 0 | 5 | 0 | 4 | Male only | Both sexes | Male only | True | False |
| TSH | 8 | NRG1 | protein_coding | rs78953577 | 32429839 | 4.20e-09 | 57 | 0 | 0 | 0 | 0 | 57 | Both sexes | Both sexes | Both sexes | True | False |
| TSH | 11 | PRDM11 | protein_coding | rs7128207 | 45230061 | 6.05e-09 | 27 | 0 | 0 | 0 | 9 | 18 | Both sexes | Both sexes | Female only | True | False |
| TSH | 9 | C9orf156 | protein_coding | rs7028759 | 100677883 | 7.12e-09 | 3 | 1 | 1 | 0 | 1 | 0 | Female only | Female only | Female only | True | False |
| TSH | 6 | RP11-344J7.2 | lincRNA | rs34836069 | 43839623 | 1.20e-08 | 7 | 0 | 0 | 0 | 0 | 7 | Both sexes | Both sexes | Male only | True | False |
| TSH | 14 | TSHR | protein_coding | rs3783949 | 81448382 | 1.28e-08 | 89 | 21 | 8 | 12 | 14 | 34 | Male only | Both sexes | Male only | True | False |
| TSH | 5 | PDE8B | protein_coding | rs9686502 | 76623421 | 1.32e-08 | 7 | 0 | 0 | 4 | 0 | 3 | Male only | Both sexes | Male only | True | False |
| TSH | 8 | NRG1 | protein_coding | rs35629470 | 32207572 | 1.86e-08 | 103 | 1 | 51 | 0 | 1 | 50 | Male only | Male only | Male only | True | False |
| TSH | 15 | RP11-353B9.1 | sense_overlapping | rs17404262 | 49967619 | 3.17e-08 | 3 | 0 | 1 | 0 | 1 | 1 | Female only | Both sexes | Female only | True | False |
| TSH | 9 | RP11-23B15.1 | lincRNA | rs12352658 | 100551768 | 3.22e-08 | 7 | 0 | 0 | 4 | 0 | 3 | Female only | Both sexes | Female only | True | False |

| pheno | CHR | Gene_Symbol | biotype | lead_snp | lead_BP | lead_P | TOTAL | NSIG | S05 | S01 | S001 | S0001 | independent_loci_in_sex_P1e6 | independent_loci_in_sex_P1e2 | independent_loci_in_sex_P5e8 | in_gwas_catalog | in_HLA |
|---|---|---|---|---|---|---|---|---|---|---|---|---|---|---|---|---|---|
| TSH | 5 | PDE8B | protein_coding | rs6879904 | 76589746 | 3.61e-08 | 63 | 18 | 0 | 4 | 22 | 19 | Female only | Both sexes | Female only | True | False |
| TSH | 3 | SYN2 | processed_transcript | rs307575 | 12127827 | 3.63e-08 | 223 | 52 | 60 | 3 | 22 | 86 | Female only | Female only | Female only | True | False |
| TSH | 6 | PDE10A | protein_coding | rs13217316 | 166038893 | 3.92e-08 | 3 | 0 | 0 | 0 | 2 | 1 | Female only | Both sexes | Female only | True | False |
| TSH | 11 | PRDM11 | protein_coding | rs7940871 | 45227830 | 4.25e-08 | 11 | 0 | 0 | 0 | 3 | 8 | Female only | Both sexes | Female only | True | False |
| TT4 | 23 | SERPINA7 | protein_coding | X:105291293:G:A | 105291293 | 1.19e-31 | 31 | 0 | 0 | 0 | 0 | 31 | Both sexes | Both sexes | Both sexes | True | False |
| pheno | CHR | Gene_Symbol | biotype | lead_snp | lead_BP | lead_P | TOTAL | NSIG | S05 | S01 | S001 | S0001 | independent_loci_in_sex_P1e6 | independent_loci_in_sex_P1e2 | independent_loci_in_sex_P5e8 | in_gwas_catalog | in_HLA |
| TT4 | 23 | NRK | protein_coding | X:105122322:G:A | 105122322 | 2.05e-13 | 39 | 0 | 0 | 0 | 0 | 39 | Both sexes | Both sexes | Both sexes | True | False |
| TT4 | 23 | RP11-560L11.1 | pseudogene | X:105317946:G:T | 105317946 | 3.19e-10 | 27 | 0 | 0 | 0 | 2 | 25 | Male only | Both sexes | Male only | True | False |
| TT4 | 23 | SERPINA7 | protein_coding | X:105248928:C:T | 105248928 | 3.32e-09 | 163 | 2 | 0 | 0 | 0 | 161 | Male only | Both sexes | Male only | True | False |
| Trigs | 11 | ZNF259 | protein_coding | rs3741298 | 116657561 | 3.29e-32 | 3 | 0 | 0 | 0 | 0 | 3 | Both sexes | Both sexes | Both sexes | True | False |
| Trigs | 11 | ZNF259 | protein_coding | rs35120633 | 116655600 | 1.46e-28 | 87 | 0 | 0 | 0 | 0 | 87 | Both sexes | Both sexes | Both sexes | True | False |
| Trigs | 11 | ZNF259 | protein_coding | rs6589566 | 116652423 | 1.72e-28 | 33 | 0 | 0 | 0 | 0 | 33 | Both sexes | Both sexes | Both sexes | True | False |
| Trigs | 11 | APOA5 | protein_coding | rs10750097 | 116664040 | 6.42e-28 | 11 | 0 | 0 | 0 | 1 | 10 | Both sexes | Both sexes | Both sexes | True | False |
| Trigs | 8 | AC100802.3 | antisense | rs115849089 | 19912370 | 1.54e-27 | 155 | 0 | 0 | 0 | 0 | 155 | Both sexes | Both sexes | Both sexes | True | False |
| Trigs | 8 | LPL | protein_coding | rs15285 | 19824667 | 1.59e-24 | 157 | 0 | 0 | 0 | 0 | 157 | Both sexes | Both sexes | Both sexes | True | False |
| Trigs | 11 | APOC3 | protein_coding | rs5141 | 116702123 | 6.67e-21 | 111 | 0 | 0 | 0 | 0 | 111 | Both sexes | Both sexes | Both sexes | True | False |
| Trigs | 2 | GCKR | protein_coding | rs1260326 | 27730940 | 5.92e-20 | 33 | 0 | 0 | 0 | 0 | 33 | Both sexes | Both sexes | Both sexes | True | False |
| Trigs | 11 | AP000770.1 | lincRNA | 11:116571608 | 116571608 | 6.77e-20 | 7 | 0 | 0 | 0 | 0 | 7 | Both sexes | Both sexes | Both sexes | True | False |
| Trigs | 11 | BUD13 | protein_coding | rs11216107 | 116579663 | 1.36e-18 | 27 | 1 | 1 | 3 | 0 | 22 | Both sexes | Both sexes | Both sexes | True | False |
| Trigs | 7 | TBL2 | protein_coding | rs11974409 | 72989390 | 1.83e-18 | 185 | 0 | 0 | 0 | 0 | 185 | Both sexes | Both sexes | Both sexes | True | False |
| Trigs | 8 | LPL | protein_coding | rs256 | 19811967 | 1.98e-18 | 23 | 0 | 0 | 0 | 0 | 23 | Both sexes | Both sexes | Both sexes | True | False |
| Trigs | 11 | BUD13 | protein_coding | rs180326 | 116624703 | 4.83e-18 | 21 | 8 | 0 | 4 | 1 | 8 | Both sexes | Both sexes | Both sexes | True | False |
| Trigs | 8 | RP11-136O12.2 | lincRNA | rs2954038 | 126507389 | 1.82e-16 | 43 | 0 | 0 | 0 | 0 | 43 | Both sexes | Both sexes | Both sexes | True | False |
| Trigs | 7 | MLXIPL | protein_coding | rs7777102 | 73058017 | 6.43e-16 | 9 | 0 | 0 | 0 | 0 | 9 | Both sexes | Both sexes | Both sexes | True | False |
| Trigs | 8 | RP11-136O12.2 | lincRNA | rs2954021 | 126482077 | 1.59e-15 | 41 | 0 | 0 | 0 | 0 | 41 | Both sexes | Both sexes | Both sexes | True | False |
| Trigs | 11 | APOA1 | protein_coding | rs525028 | 116705516 | 2.00e-14 | 97 | 1 | 1 | 2 | 0 | 93 | Both sexes | Both sexes | Both sexes | True | False |

| pheno | CHR | Gene_Symbol | biotype | lead_snp | lead_BP | lead_P | TOTAL | NSIG | S05 | S01 | S001 | S0001 | independent_loci_in_sex_P1e6 | independent_loci_in_sex_P1e2 | independent_loci_in_sex_P5e8 | in_gwas_catalog | in_HLA |
|---|---|---|---|---|---|---|---|---|---|---|---|---|---|---|---|---|---|
| Trigs | 2 | ZNF512 | protein_coding | rs4666000 | 27839369 | 3.92e-14 | 71 | 0 | 0 | 0 | 0 | 71 | Both sexes | Both sexes | Both sexes | True | False |
| Trigs | 8 | LPL | protein_coding | rs1441771 | 19757300 | 1.32e-13 | 33 | 0 | 0 | 0 | 0 | 33 | Both sexes | Both sexes | Female only | True | False |
| Trigs | 8 | LPL | protein_coding | rs894210 | 19865843 | 1.55e-13 | 9 | 0 | 0 | 0 | 0 | 9 | Both sexes | Both sexes | Both sexes | True | False |
| Trigs | 11 | AP000770.1 | lincRNA | rs480823 | 116525730 | 2.17e-13 | 43 | 0 | 0 | 0 | 20 | 23 | Female only | Both sexes | Female only | True | False |
| Trigs | 11 | AP000936.5 | pseudogene | rs12366015 | 116990851 | 2.41e-13 | 9 | 0 | 0 | 0 | 0 | 9 | Both sexes | Both sexes | Both sexes | True | False |
| Trigs | 11 | RP11-109L13.1 | pseudogene | rs11216288 | 117004656 | 5.41e-13 | 41 | 0 | 0 | 0 | 0 | 41 | Both sexes | Both sexes | Both sexes | True | False |
| Trigs | 19 | APOE | protein_coding | rs72654473 | 45414399 | 1.62e-12 | 7 | 0 | 0 | 0 | 0 | 7 | Both sexes | Both sexes | Both sexes | True | False |
| Trigs | 8 | LPL | protein_coding | rs34693282 | 19873566 | 2.06e-12 | 149 | 112 | 0 | 0 | 0 | 37 | Both sexes | Both sexes | Both sexes | True | False |
| Trigs | 11 | SIK3 | protein_coding | rs672058 | 116764021 | 2.74e-11 | 21 | 0 | 0 | 0 | 0 | 21 | Both sexes | Both sexes | Both sexes | True | False |
| Trigs | 19 | APOC1 | protein_coding | rs5117 | 45418790 | 4.29e-11 | 11 | 0 | 1 | 1 | 0 | 9 | Male only | Both sexes | Male only | True | False |
| Trigs | 1 | DOCK7 | protein_coding | rs11207995 | 63049551 | 5.83e-11 | 355 | 0 | 0 | 0 | 0 | 355 | Both sexes | Both sexes | Both sexes | True | False |
| Trigs | 8 | AC100802.3 | antisense | rs13263508 | 19942181 | 2.03e-10 | 29 | 0 | 0 | 0 | 0 | 29 | Both sexes | Both sexes | Male only | True | False |
| Trigs | 11 | APOA4 | protein_coding | rs10892035 | 116694938 | 2.63e-10 | 11 | 0 | 0 | 3 | 3 | 5 | Both sexes | Both sexes | Both sexes | True | False |
| Trigs | 8 | AC100802.3 | antisense | rs28841897 | 19894504 | 5.16e-10 | 11 | 0 | 0 | 0 | 4 | 7 | Male only | Both sexes | Male only | True | False |
| Trigs | 2 | GTF3C2 | protein_coding | rs37390951 | 27556721 | 1.52e-09 | 197 | 0 | 0 | 1 | 1 | 195 | Both sexes | Both sexes | Both sexes | True | False |
| Trigs | 11 | BUD13 | protein_coding | rs1268353 | 116639692 | 3.36e-09 | 57 | 11 | 5 | 20 | 19 | 2 | Male only | Both sexes | Male only | True | False |
| Trigs | 19 | TM6SF2 | protein_coding | rs58542926 | 19379549 | 1.11e-08 | 51 | 16 | 1 | 2 | 7 | 25 | Male only | Both sexes | Male only | True | False |
| Trigs | 11 | TAGLN | protein_coding | rs494356 | 117071970 | 1.37e-08 | 83 | 0 | 2 | 23 | 25 | 33 | Both sexes | Both sexes | Male only | True | False |
| Trigs | 11 | BUD13 | protein_coding | rs1263056 | 116576415 | 3.44e-08 | 87 | 17 | 26 | 0 | 16 | 28 | Female only | Both sexes | Female only | True | False |
| Trigs | 6 | HLA-B | protein_coding | rs547630246 | 31312293 | 4.32e-08 | 79 | 0 | 6 | 10 | 27 | 36 | Male only | Both sexes | Male only | True | True |
| Trigs | 8 | LPL | protein_coding | rs1441770 | 19757319 | 4.67e-08 | 5 | 0 | 0 | 0 | 3 | 2 | Female only | Both sexes | Female only | True | False |
| UWBC | 8 | ENTPD4 | protein_coding | 8:23309914 | 23309914 | 2.21e-08 | 19 | 10 | 0 | 0 | 0 | 9 | Male only | Male only | Male only | True | False |
| UpH | 15 | WDR72 | protein_coding | rs491567 | 53946593 | 4.94e-08 | 117 | 0 | 0 | 20 | 8 | 89 | Both sexes | Both sexes | Male only | True | False |
| UricA | 4 | SLC2A9 | protein_coding | rs9994216 | 9984541 | 7.96e-41 | 203 | 0 | 0 | 0 | 0 | 203 | Both sexes | Both sexes | Both sexes | True | False |
| UricA | 4 | AC006499.3 | pseudogene | rs55766779 | 10287039 | 7.31e-31 | 139 | 0 | 0 | 0 | 0 | 139 | Both sexes | Both sexes | Both sexes | True | False |
| UricA | 4 | SLC2A9 | protein_coding | rs9291642 | 10007275 | 6.48e-29 | 31 | 0 | 0 | 0 | 0 | 31 | Both sexes | Both sexes | Both sexes | True | False |

| pheno | CHR | Gene_Symbol | biotype | lead_snp | lead_BP | lead_P | TOTAL | NSIG | S05 | S01 | S001 | S0001 | independent_loci_in_sex_P1e6 | independent_loci_in_sex_P1e2 | independent_loci_in_sex_P5e8 | in_gwas_catalog | in_HLA |
|---|---|---|---|---|---|---|---|---|---|---|---|---|---|---|---|---|---|
| UricA | 4 | SLC2A9 | protein_coding | rs10805346 | 9920347 | 2.49e-21 | 3 | 0 | 0 | 0 | 0 | 3 | Both sexes | Both sexes | Both sexes | True | False |
| UricA | 4 | SLC2A9 | protein_coding | rs1122141 | 9947278 | 2.05e-20 | 507 | 4 | 54 | 186 | 10 | 253 | Female only | Both sexes | Female only | True | False |
| UricA | 4 | SLC2A9 | protein_coding | rs7698858 | 9895070 | 1.11e-14 | 63 | 29 | 3 | 0 | 0 | 31 | Female only | Female only | Female only | True | False |
| UricA | 4 | AC006499.4 | pseudogene | rs6833292 | 10272429 | 5.47e-14 | 31 | 1 | 0 | 3 | 1 | 26 | Both sexes | Both sexes | Female only | True | False |
| UricA | 4 | SLC2A9 | protein_coding | rs11722228 | 9915741 | 1.02e-12 | 15 | 0 | 0 | 0 | 6 | 9 | Female only | Both sexes | Female only | True | False |
| UricA | 4 | SLC2A9 | protein_coding | rs56239136 | 9898003 | 2.35e-12 | 19 | 7 | 1 | 2 | 0 | 9 | Female only | Both sexes | Female only | True | False |
| UricA | 4 | SLC2A9 | protein_coding | rs1048252 | 9870535 | 1.24e-11 | 9 | 5 | 0 | 0 | 0 | 4 | Female only | Female only | Female only | True | False |
| UricA | 4 | AC006499.1 | pseudogene | rs993172 | 10324361 | 1.31e-11 | 23 | 6 | 2 | 5 | 0 | 10 | Female only | Both sexes | Female only | True | False |
| UricA | 4 | WDR1 | protein_coding | rs35289776 | 10119034 | 7.54e-11 | 33 | 7 | 4 | 7 | 3 | 12 | Female only | Both sexes | Female only | True | False |
| UricA | 4 | WDR1 | protein_coding | rs73212828 | 10078756 | 7.40e-09 | 519 | 1 | 177 | 82 | 1 | 258 | Female only | Both sexes | Female only | True | False |
| VD25 | 11 | CYP2R1 | protein_coding | rs1993116 | 14910234 | 1.94e-09 | 65 | 33 | 0 | 0 | 2 | 30 | Female only | Female only | Female only | True | False |
| Vi.B12 | 19 | FUT2 | protein_coding | rs516246 | 49206172 | 6.86e-16 | 65 | 0 | 0 | 0 | 0 | 65 | Both sexes | Both sexes | Both sexes | True | False |
| Vi.B12 | 19 | FGF21 | protein_coding | rs838133 | 49259529 | 5.96e-10 | 41 | 0 | 0 | 0 | 1 | 40 | Male only | Both sexes | Male only | True | False |
| Vi.B12 | 10 | CUBN | protein_coding | rs1801222 | 17156151 | 8.29e-10 | 7 | 0 | 0 | 0 | 0 | 7 | Both sexes | Both sexes | Both sexes | True | False |

| pheno | CHR | Gene_Symbol | biotype | lead_snp | lead_BP | lead_P | TOTAL | NSIG | S05 | S01 | S001 | S0001 | independent_loci_in_sex_P1e6 | independent_loci_in_sex_P1e2 | independent_loci_in_sex_P5e8 | in_gwas_catalog | in_HLA |
|---|---|---|---|---|---|---|---|---|---|---|---|---|---|---|---|---|---|
| Vi.B12 | 19 | RASIP1 | protein_coding | rs546308 | 49223570 | 4.00e-09 | 7 | 0 | 0 | 0 | 0 | 7 | Male only | Both sexes | Male only | True | False |
| Vi.B12 | 10 | CUBN | protein_coding | rs12243895 | 17158973 | 9.32e-09 | 89 | 0 | 0 | 19 | 23 | 47 | Both sexes | Both sexes | Male only | True | False |
| Vi.B12 | 6 | EEF1A1P42 | pseudogene | rs9473541 | 49353357 | 2.56e-08 | 203 | 0 | 0 | 22 | 8 | 173 | Female only | Both sexes | Female only | True | False |
| WBC | 17 | MED24 | protein_coding | rs1045929 | 38175426 | 1.45e-31 | 153 | 0 | 0 | 0 | 0 | 153 | Both sexes | Both sexes | Both sexes | True | False |
| WBC | 17 | MED24 | protein_coding | rs12453343 | 38181176 | 1.44e-26 | 83 | 0 | 0 | 0 | 0 | 83 | Both sexes | Both sexes | Both sexes | True | False |
| WBC | 17 | LRRC3C | protein_coding | rs62068171 | 38103242 | 7.68e-20 | 191 | 0 | 0 | 0 | 9 | 182 | Both sexes | Both sexes | Both sexes | True | False |
| WBC | 17 | GSDMA | protein_coding | 17:38122680 | 38122680 | 9.71e-16 | 3 | 0 | 0 | 0 | 0 | 3 | Female only | Both sexes | Female only | True | False |
| WBC | 6 | HLA-C | protein_coding | rs9264642 | 31238490 | 5.24e-12 | 83 | 0 | 0 | 0 | 11 | 72 | Both sexes | Both sexes | Female only | True | True |
| WBC | 17 | IKZF3 | protein_coding | rs11658993 | 37940808 | 2.74e-11 | 71 | 0 | 0 | 0 | 1 | 70 | Both sexes | Both sexes | Female only | True | False |
| WBC | 6 | HLA-B | protein_coding | rs9265994 | 31316526 | 5.80e-11 | 273 | 0 | 1 | 2 | 14 | 256 | Both sexes | Both sexes | Female only | True | True |
| WBC | 6 | HLA-DQA1 | protein_coding | rs9271586 | 32590899 | 6.15e-11 | 275 | 0 | 0 | 23 | 28 | 224 | Both sexes | Both sexes | Both sexes | True | True |

| pheno | CHR | Gene_Symbol | biotype | lead_snp | lead_BP | lead_P | TOTAL | NSIG | S05 | S01 | S001 | S0001 | independent_loci_in_sex_P1e6 | independent_loci_in_sex_P1e2 | independent_loci_in_sex_P5e8 | in_gwas_catalog | in_HLA |
|---|---|---|---|---|---|---|---|---|---|---|---|---|---|---|---|---|---|
| WBC | 6 | XXbac-BPG248L24.12 | pseudogene | rs2523579 | 31328517 | 1.04e-10 | 5 | 0 | 0 | 0 | 2 | 3 | Female only | Both sexes | Female only | True | True |
| WBC | 6 | HLA-C | protein_coding | rs2249742 | 31240721 | 1.68e-10 | 9 | 0 | 0 | 2 | 3 | 4 | Female only | Both sexes | Female only | True | True |
| WBC | 6 | HCG4P7 | pseudogene | rs1611658 | 29842451 | 2.06e-10 | 467 | 0 | 0 | 2 | 23 | 442 | Both sexes | Both sexes | Both sexes | True | True |
| WBC | 4 | CXCL2 | protein_coding | rs7667376 | 74967890 | 2.87e-10 | 25 | 0 | 0 | 0 | 0 | 25 | Both sexes | Both sexes | Female only | True | False |
| WBC | 6 | OR5V1 | protein_coding | rs9257809 | 29356331 | 3.06e-10 | 119 | 0 | 1 | 0 | 17 | 101 | Both sexes | Both sexes | Male only | True | False |
| WBC | 6 | TRIM27 | protein_coding | rs3118361 | 28898287 | 7.07e-10 | 313 | 0 | 0 | 6 | 50 | 257 | Male only | Both sexes | Male only | True | False |
| WBC | 6 | HCG17 | lincRNA | rs3094069 | 30251391 | 8.51e-10 | 325 | 0 | 0 | 9 | 18 | 298 | Both sexes | Both sexes | Both sexes | True | True |
| WBC | 6 | RP1-97D16.1 | pseudogene | rs17693963 | 27710165 | 8.73e-10 | 177 | 2 | 26 | 9 | 53 | 87 | Male only | Both sexes | Male only | True | False |
| WBC | 6 | XXbac-BPG181B23.7 | lincRNA | rs2523466 | 31363026 | 1.42e-09 | 11 | 0 | 3 | 4 | 1 | 3 | Female only | Both sexes | Female only | True | True |
| WBC | 4 | SCD5 | protein_coding | rs74532594 | 83620037 | 1.74e-09 | 35 | 0 | 0 | 0 | 18 | 17 | Male only | Both sexes | Male only | True | False |
| WBC | 6 | XXbac-BPG254F23.7 | lincRNA | rs3957147 | 32682135 | 2.94e-09 | 41 | 0 | 0 | 0 | 2 | 39 | Female only | Both sexes | Female only | True | True |
| WBC | 6 | BTN3A1 | protein_coding | rs3799378 | 26404374 | 4.23e-09 | 23 | 12 | 0 | 0 | 0 | 11 | Male only | Male only | Male only | True | False |
| WBC | 6 | MTCO3P1 | pseudogene | rs2647046 | 32668336 | 4.74e-09 | 163 | 0 | 0 | 0 | 0 | 163 | Both sexes | Both sexes | Both sexes | True | True |
| WBC | 6 | MICF | pseudogene | rs2523764 | 29817617 | 4.79e-09 | 229 | 0 | 30 | 64 | 100 | 35 | Both sexes | Both sexes | Female only | True | True |
| WBC | 6 | ZSCAN31 | protein_coding | rs35016036 | 28314880 | 5.80e-09 | 189 | 0 | 6 | 26 | 35 | 122 | Male only | Both sexes | Male only | True | False |
| WBC | 6 | SKIV2L | protein_coding | rs410851 | 31936668 | 6.34e-09 | 39 | 0 | 0 | 0 | 8 | 31 | Both sexes | Both sexes | Male only | True | True |
| WBC | 6 | LINC00533 | lincRNA | rs34724414 | 28619353 | 6.61e-09 | 7 | 0 | 0 | 0 | 2 | 5 | Male only | Both sexes | Male only | True | False |
| WBC | 6 | MUC22 | protein_coding | rs1628680 | 30988825 | 9.97e-09 | 215 | 0 | 0 | 3 | 19 | 193 | Both sexes | Both sexes | Female only | True | True |
| WBC | 6 | OR2W2P | pseudogene | rs138234416 | 27992898 | 1.41e-08 | 25 | 0 | 6 | 2 | 5 | 12 | Male only | Both sexes | Male only | True | False |
| WBC | 6 | BTN2A2 | protein_coding | rs9358938 | 26387546 | 1.72e-08 | 287 | 114 | 30 | 0 | 17 | 126 | Male only | Male only | Male only | True | False |
| WBC | 6 | SUMO2P1 | pseudogene | rs3131855 | 29610281 | 1.79e-08 | 71 | 16 | 22 | 4 | 13 | 16 | Female only | Female only | Female only | True | True |
| WBC | 17 | PGAP3 | protein_coding | rs1495100 | 37832279 | 2.40e-08 | 93 | 47 | 0 | 3 | 0 | 43 | Female only | Female only | Female only | True | False |
| WBC | 6 | AL662890.1 | miRNA | rs1233608 | 28736189 | 2.85e-08 | 13 | 0 | 2 | 1 | 4 | 6 | Male only | Both sexes | Male only | True | False |
| WBC | 6 | RNU6-1133P | snRNA | rs2517474 | 31046494 | 2.94e-08 | 183 | 21 | 40 | 44 | 1 | 77 | Female only | Both sexes | Female only | True | True |
| WBC | 6 | USP8P1 | pseudogene | rs2524075 | 31243859 | 3.16e-08 | 3 | 0 | 0 | 0 | 2 | 1 | Female only | Both sexes | Female only | True | True |

| pheno | CHR | Gene_Symbol | biotype | lead_snp | lead_BP | lead_P | TOTAL | NSIG | S05 | S01 | S001 | S0001 | independent_loci_in_sex_P1e6 | independent_loci_in_sex_P1e2 | independent_loci_in_sex_P5e8 | in_gwas_catalog | in_HLA |
|---|---|---|---|---|---|---|---|---|---|---|---|---|---|---|---|---|---|
| WBC | 6 | HLA-DRA | protein_coding | rs6937696 | 32418018 | 4.15e-08 | 11 | 0 | 0 | 0 | 5 | 6 | Male only | Both sexes | Male only | True | True |
| WBCrt | 16 | UMOD | protein_coding | rs13329952 | 20366507 | 2.90e-10 | 53 | 27 | 0 | 0 | 0 | 26 | Male only | Male only | Male only | True | False |
| WEIGHT_SCALE | 16 | FTO | protein_coding | rs56094641 | 53806453 | 9.07e-18 | 169 | 0 | 0 | 0 | 0 | 169 | Both sexes | Both sexes | Both sexes | True | False |
| WEIGHT_SCALE | 16 | FTO | protein_coding | rs1477196 | 53808258 | 5.88e-09 | 9 | 0 | 0 | 1 | 4 | 4 | Female only | Both sexes | Female only | True | False |
| Weight | 16 | FTO | protein_coding | rs1421085 | 53800954 | 5.42e-41 | 169 | 0 | 0 | 0 | 0 | 169 | Both sexes | Both sexes | Both sexes | True | False |
| Weight | 18 | RP11-795H16.3 | lincRNA | rs12967878 | 57826570 | 5.31e-20 | 243 | 0 | 0 | 10 | 23 | 210 | Both sexes | Both sexes | Both sexes | True | False |
| Weight | 16 | FTO | protein_coding | rs1477196 | 53808258 | 8.38e-16 | 9 | 0 | 0 | 0 | 0 | 9 | Both sexes | Both sexes | Both sexes | True | False |
| Weight | 18 | RP11-795H16.2 | pseudogene | rs12456624 | 57903086 | 2.55e-13 | 55 | 0 | 0 | 28 | 0 | 27 | Female only | Both sexes | Female only | True | False |
| Weight | 18 | RP11-795H16.3 | lincRNA | rs11664212 | 57739179 | 1.16e-12 | 29 | 0 | 8 | 1 | 6 | 14 | Female only | Both sexes | Female only | True | False |
| Weight | 1 | SEC16B | protein_coding | rs516636 | 177855517 | 4.02e-12 | 101 | 0 | 20 | 4 | 8 | 69 | Female only | Both sexes | Female only | True | False |
| Weight | 3 | ZBTB38 | protein_coding | rs6763931 | 141102833 | 1.93e-11 | 75 | 0 | 0 | 0 | 0 | 75 | Both sexes | Both sexes | Female only | True | False |
| Weight | 18 | RP11-795H16.3 | lincRNA | rs8086627 | 57743096 | 1.41e-10 | 33 | 0 | 1 | 14 | 2 | 16 | Female only | Both sexes | Female only | True | False |
| Weight | 3 | EIF2B5 | protein_coding | rs11546878 | 183976103 | 3.98e-10 | 5 | 2 | 1 | 0 | 0 | 2 | Female only | Female only | Female only | True | False |
| Weight | 18 | RNU4-17P | snRNA | rs502921 | 57845479 | 1.70e-09 | 45 | 0 | 0 | 23 | 0 | 22 | Female only | Both sexes | Female only | True | False |
| Weight | 2 | TMEM18 | protein_coding | rs6760274 | 652272 | 9.70e-09 | 337 | 0 | 0 | 0 | 16 | 321 | Female only | Both sexes | Female only | True | False |
| Weight | 2 | TMEM18 | protein_coding | rs2867121 | 658550 | 2.16e-08 | 5 | 0 | 1 | 2 | 1 | 1 | Female only | Both sexes | Female only | True | False |
| Weight | 19 | RASIP1 | protein_coding | rs1231281 | 49239200 | 2.91e-08 | 111 | 55 | 1 | 0 | 3 | 52 | Female only | Female only | Female only | True | False |
| Weight | 12 | FAIM2 | protein_coding | rs7132908 | 50263148 | 3.09e-08 | 21 | 1 | 4 | 0 | 3 | 13 | Both sexes | Both sexes | Female only | True | False |
| Weight | 11 | BDNF-AS | antisense | rs4923463 | 27672500 | 4.05e-08 | 139 | 36 | 33 | 10 | 2 | 58 | Female only | Both sexes | Female only | True | False |
| Weight | 16 | FTO | protein_coding | rs7203521 | 53769293 | 4.26e-08 | 3 | 0 | 0 | 2 | 0 | 1 | Female only | Both sexes | Female only | True | False |
| Weight | 6 | NQO2 | protein_coding | rs11242816 | 2995459 | 4.38e-08 | 21 | 11 | 0 | 3 | 6 | 1 | Male only | Male only | Male only | True | False |
| X25DTot | 4 | GC | protein_coding | rs3755967 | 72609398 | 1.01e-34 | 39 | 0 | 0 | 0 | 0 | 39 | Both sexes | Both sexes | Both sexes | True | False |
| X25DTot | 4 | GC | protein_coding | rs222035 | 72621674 | 6.81e-26 | 7 | 0 | 0 | 0 | 0 | 7 | Both sexes | Both sexes | Female only | True | False |
| X25DTot | 4 | GC | protein_coding | rs28889094 | 72603274 | 7.66e-14 | 45 | 0 | 0 | 19 | 4 | 22 | Female only | Both sexes | Female only | True | False |

| pheno | CHR | Gene_Symbol | biotype | lead_snp | lead_BP | lead_P | TOTAL | NSIG | S05 | S01 | S001 | S0001 | independent_loci_in_sex_P1e6 | independent_loci_in_sex_P1e2 | independent_loci_in_sex_P5e8 | in_gwas_catalog | in_HLA |
|---|---|---|---|---|---|---|---|---|---|---|---|---|---|---|---|---|---|
| X25DTot | 4 | GC | protein_coding | rs1526692 | 72578724 | 2.49e-12 | 11 | 0 | 0 | 0 | 2 | 9 | Female only | Both sexes | Female only | True | False |
| X25DTot | 4 | GC | protein_coding | rs56393564 | 72582136 | 7.86e-12 | 11 | 0 | 0 | 0 | 6 | 5 | Female only | Both sexes | Female only | True | False |
| X25DTot | 11 | CYP2R1 | protein_coding | rs10766196 | 14913131 | 3.37e-11 | 75 | 0 | 0 | 3 | 6 | 66 | Female only | Both sexes | Female only | True | False |
| X25DTot | 4 | RP11-545L5.1 | lincRNA | rs9993024 | 72723718 | 4.04e-10 | 87 | 0 | 0 | 1 | 27 | 59 | Female only | Both sexes | Female only | True | False |
| X25DTot | 11 | RP11-140L24.3 | pseudogene | rs7128011 | 14462532 | 1.93e-09 | 83 | 0 | 8 | 14 | 13 | 48 | Female only | Both sexes | Female only | True | False |
| X25DTot | 11 | RRAS2 | protein_coding | rs12292287 | 14340040 | 2.26e-09 | 51 | 12 | 13 | 0 | 1 | 25 | Female only | Both sexes | Female only | True | False |
| X25DTot | 11 | CYP2R1 | protein_coding | rs10741657 | 14914878 | 1.55e-08 | 47 | 17 | 7 | 17 | 0 | 6 | Female only | Both sexes | Female only | True | False |
| X25DTot | 11 | COPB1 | protein_coding | rs2597190 | 14510851 | 3.40e-08 | 21 | 0 | 3 | 8 | 0 | 10 | Female only | Both sexes | Female only | True | False |
| X25HyD3 | 4 | GC | protein_coding | rs4588 | 72618323 | 4.97e-27 | 39 | 0 | 0 | 0 | 0 | 39 | Both sexes | Both sexes | Both sexes | True | False |
| X25HyD3 | 4 | GC | protein_coding | rs222035 | 72621674 | 1.02e-21 | 7 | 0 | 0 | 0 | 0 | 7 | Both sexes | Both sexes | Both sexes | True | False |
| X25HyD3 | 4 | RP11-545L5.1 | lincRNA | rs6821488 | 72754943 | 1.78e-11 | 111 | 0 | 6 | 18 | 15 | 72 | Female only | Both sexes | Female only | True | False |
| X25HyD3 | 4 | GC | protein_coding | rs57897761 | 72574310 | 3.45e-11 | 11 | 0 | 0 | 0 | 0 | 11 | Both sexes | Both sexes | Female only | True | False |
| X25HyD3 | 4 | GC | protein_coding | rs28889094 | 72603274 | 7.99e-11 | 45 | 0 | 0 | 0 | 19 | 26 | Female only | Both sexes | Female only | True | False |
| X25HyD3 | 11 | CYP2R1 | protein_coding | rs10766196 | 14913131 | 5.91e-09 | 75 | 0 | 5 | 2 | 36 | 32 | Female only | Both sexes | Female only | True | False |
| X_percent_SAT | 6 | HFE | protein_coding | rs144861591 | 26072992 | 3.47e-20 | 19 | 0 | 0 | 0 | 0 | 19 | Both sexes | Both sexes | Both sexes | False | False |
| X_percent_SAT | 6 | HIST1H2AA | protein_coding | rs116009877 | 25715657 | 2.92e-18 | 33 | 0 | 0 | 0 | 0 | 33 | Both sexes | Both sexes | Both sexes | False | False |
| X_percent_SAT | 22 | TMPRSS6 | protein_coding | rs2072860 | 37470604 | 3.73e-15 | 9 | 0 | 0 | 0 | 0 | 9 | Both sexes | Both sexes | Both sexes | False | False |
| X_percent_SAT | 6 | LRRC16A | protein_coding | rs79665799 | 25408104 | 5.00e-15 | 241 | 0 | 0 | 0 | 19 | 222 | Female only | Both sexes | Female only | False | False |
| X_percent_SAT | 6 | LRRC16A | protein_coding | rs77244967 | 25536591 | 5.40e-14 | 23 | 0 | 0 | 0 | 0 | 23 | Both sexes | Both sexes | Both sexes | False | False |
| X_percent_SAT | 6 | SCGN | protein_coding | rs72838866 | 25673273 | 4.88e-13 | 19 | 0 | 0 | 0 | 0 | 19 | Both sexes | Both sexes | Both sexes | False | False |
| X_percent_SAT | 6 | HIST1H4A | protein_coding | rs72834629 | 26023032 | 3.08e-12 | 251 | 0 | 0 | 3 | 15 | 233 | Both sexes | Both sexes | Both sexes | False | False |
| X_percent_SAT | 6 | LRRC16A | protein_coding | rs116272812 | 25452783 | 4.39e-12 | 17 | 0 | 0 | 0 | 4 | 13 | Both sexes | Both sexes | Both sexes | False | False |

| pheno | CHR | Gene_Symbol | biotype | lead_snp | lead_BP | lead_P | TOTAL | NSIG | S05 | S01 | S001 | S0001 | independent_loci_in_sex_P1e6 | independent_loci_in_sex_P1e2 | independent_loci_in_sex_P5e8 | in_gwas_catalog | in_HLA |
|---|---|---|---|---|---|---|---|---|---|---|---|---|---|---|---|---|---|
| X_percent_SAT | 6 | VN1R13P | pseudogene | rs13194491 | 27037080 | 5.69e-11 | 3 | 0 | 0 | 0 | 0 | 3 | Male only | Both sexes | Male only | False | False |
| X_percent_SAT | 6 | RP3-522P13.1 | pseudogene | rs9393635 | 25265377 | 1.68e-10 | 75 | 17 | 21 | 0 | 1 | 36 | Female only | Female only | Female only | False | False |
| X_percent_SAT | 6 | LRRC16A | protein_coding | rs73730705 | 25383843 | 2.07e-10 | 27 | 0 | 0 | 0 | 5 | 22 | Female only | Both sexes | Female only | False | False |
| X_percent_SAT | 6 | HIST1H1T | protein_coding | rs198846 | 26107463 | 5.54e-10 | 33 | 0 | 0 | 4 | 3 | 26 | Both sexes | Both sexes | Both sexes | False | False |
| X_percent_SAT | 6 | SLC17A1 | protein_coding | rs12191655 | 25803095 | 2.96e-09 | 291 | 0 | 88 | 50 | 9 | 144 | Both sexes | Both sexes | Female only | False | False |
| X_percent_SAT | 6 | RP11-191A15.4 | pseudogene | rs35191161 | 25230092 | 8.30e-09 | 13 | 7 | 0 | 0 | 0 | 6 | Female only | Female only | Female only | False | False |
| X_percent_SAT | 6 | HIST1H3G | protein_coding | rs7773004 | 26267755 | 8.38e-09 | 89 | 0 | 0 | 10 | 47 | 32 | Both sexes | Both sexes | Both sexes | False | False |
| pCO2 | 6 | CDKAL1 | protein_coding | rs79529169 | 21184849 | 1.07e-08 | 49 | 25 | 0 | 0 | 1 | 23 | Female only | Female only | Female only | True | False |

#### STable5: 'Starry" loci - e.g. loci for per-person median trait values with only a single variant - excluded from counts of significant loci

241 rows × 18 columns

| pheno | CHR | Gene_Symbol | biotype | lead_snp | lead_BP | lead_P | TOTAL | NSIG | S05 | S01 | S001 | S0001 | independent_loci_in_sex_P1e6 | independent_loci_in_sex_P1e2 | independent_loci_in_sex_P5e8 | in_gwas_catalog | in_HLA |
|---|---|---|---|---|---|---|---|---|---|---|---|---|---|---|---|---|---|
| AlkP | 9 | SLC2A6 | protein_coding | rs2073935 | 136340200 | 7.98e-20 | 1 | 0 | 0 | 0 | 0 | 1 | Both sexes | Both sexes | Both sexes | True | False |
| AlkP | 6 | GPLD1 | protein_coding | rs62400470 | 24470974 | 1.96e-14 | 1 | 0 | 0 | 0 | 0 | 1 | Both sexes | Both sexes | Both sexes | True | False |
| AlkP | 1 | ALPL | protein_coding | rs111915154 | 21907893 | 3.15e-11 | 1 | 0 | 0 | 0 | 0 | 1 | Female only | Both sexes | Female only | True | False |
| AlkP | 6 | GPLD1 | protein_coding | rs145510429 | 24433782 | 2.09e-10 | 1 | 0 | 0 | 0 | 0 | 1 | Both sexes | Both sexes | Both sexes | True | False |
| AlkP | 9 | ABO | processed_transcript | rs8176694 | 136137646 | 3.57e-09 | 1 | 0 | 0 | 0 | 0 | 1 | Both sexes | Both sexes | Both sexes | True | False |
| AlkP | 12 | B4GALNT3 | protein_coding | rs7955258 | 570947 | 1.38e-08 | 1 | 0 | 0 | 1 | 0 | 0 | Male only | Both sexes | Male only | True | False |
| AlkP | 6 | MRS2 | protein_coding | rs62401885 | 24398185 | 2.22e-08 | 1 | 0 | 0 | 0 | 0 | 1 | Both sexes | Both sexes | Female only | True | False |
| AlkP | 6 | MRS2 | protein_coding | rs9393553 | 24401654 | 3.22e-08 | 1 | 0 | 0 | 1 | 0 | 0 | Female only | Both sexes | Female only | True | False |
| Amyl | 1 | RP11-153F1.1 | antisense | rs77572740 | 104059255 | 5.40e-14 | 1 | 0 | 0 | 0 | 0 | 1 | Both sexes | Both sexes | Both sexes | True | False |
| BASOAB | 19 | SLC7A10 | protein_coding | rs55779678 | 33739726 | 5.20e-36 | 1 | 0 | 0 | 0 | 0 | 1 | Both sexes | Both sexes | Both sexes | True | False |
| BASOAB | 3 | C3orf27 | protein_coding | rs2713600 | 128297068 | 4.72e-24 | 1 | 0 | 0 | 0 | 0 | 1 | Both sexes | Both sexes | Both sexes | True | False |
| BASOAB | 19 | CTD-2540B15.12 | lincRNA | rs1862465 | 33756239 | 3.25e-21 | 1 | 0 | 0 | 0 | 0 | 1 | Both sexes | Both sexes | Both sexes | True | False |
| BASOAB | 19 | SLC7A10 | protein_coding | rs11666788 | 33708069 | 2.03e-15 | 1 | 0 | 0 | 0 | 0 | 1 | Both sexes | Both sexes | Both sexes | True | False |
| BASOAB | 3 | IL5RA | protein_coding | rs340828 | 3108083 | 2.73e-08 | 1 | 0 | 0 | 0 | 0 | 1 | Both sexes | Both sexes | Male only | True | False |
| BASORE | 19 | SLC7A10 | protein_coding | rs55779678 | 33739726 | 4.83e-37 | 1 | 0 | 0 | 0 | 0 | 1 | Both sexes | Both sexes | Both sexes | True | False |
| BASORE | 3 | C3orf27 | protein_coding | rs2713600 | 128297068 | 1.22e-30 | 1 | 0 | 0 | 0 | 0 | 1 | Both sexes | Both sexes | Both sexes | True | False |
| BASORE | 19 | CTD-2540B15.12 | lincRNA | rs1862465 | 33756239 | 7.83e-17 | 1 | 0 | 0 | 0 | 0 | 1 | Both sexes | Both sexes | Both sexes | True | False |
| BASORE | 19 | SLC7A10 | protein_coding | rs11666788 | 33708069 | 1.93e-12 | 1 | 0 | 0 | 0 | 0 | 1 | Female only | Both sexes | Female only | True | False |
| BAbs | 19 | SLC7A10 | protein_coding | rs55779678 | 33739726 | 7.23e-14 | 1 | 0 | 0 | 0 | 0 | 1 | Both sexes | Both sexes | Both sexes | True | False |
| BAbs | 3 | C3orf27 | protein_coding | rs2713600 | 128297068 | 1.50e-11 | 1 | 0 | 0 | 0 | 0 | 1 | Both sexes | Both sexes | Both sexes | True | False |
| BMI_CLEAN | 16 | FTO | protein_coding | rs7187961 | 53826034 | 8.43e-09 | 1 | 0 | 0 | 0 | 0 | 1 | Both sexes | Both sexes | Female only | True | False |
| BUN | 16 | PDILT | protein_coding | rs77924615 | 20392332 | 8.56e-10 | 1 | 0 | 0 | 0 | 0 | 1 | Male only | Both sexes | Male only | True | False |
| Baso | 19 | SLC7A10 | protein_coding | rs55779678 | 33739726 | 3.32e-17 | 1 | 0 | 0 | 0 | 0 | 1 | Both sexes | Both sexes | Both sexes | True | False |
| Baso | 3 | C3orf27 | protein_coding | rs2713600 | 128297068 | 1.20e-14 | 1 | 0 | 0 | 0 | 0 | 1 | Both sexes | Both sexes | Both sexes | True | False |

| pheno | CHR | Gene_Symbol | biotype | lead_snp | lead_BP | lead_P | TOTAL | NSIG | S05 | S01 | S001 | S0001 | independent_loci_in_sex_P1e6 | independent_loci_in_sex_P1e2 | independent_loci_in_sex_P5e8 | in_gwas_catalog | in_HLA |
|---|---|---|---|---|---|---|---|---|---|---|---|---|---|---|---|---|---|
| Baso | 19 | CTD-2540B15.12 | lincRNA | rs1862465 | 33756239 | 3.84e-08 | 1 | 0 | 0 | 1 | 0 | 0 | Female only | Both sexes | Female only | True | False |
| C4Quan | 6 | SKIV2L | protein_coding | rs401775 | 31931137 | 2.36e-21 | 1 | 0 | 0 | 0 | 0 | 1 | Both sexes | Both sexes | Female only | True | True |
| C4Quan | 6 | MICB | protein_coding | rs2252937 | 31461613 | 1.17e-20 | 1 | 0 | 0 | 0 | 0 | 1 | Both sexes | Both sexes | Both sexes | True | True |
| C4Quan | 6 | NOTCH4 | protein_coding | rs3134931 | 32190620 | 5.46e-17 | 1 | 0 | 0 | 1 | 0 | 0 | Female only | Both sexes | Female only | True | True |
| C4Quan | 6 | HLA-DQB1 | protein_coding | rs12055445 | 32626739 | 1.88e-14 | 1 | 0 | 0 | 0 | 0 | 1 | Both sexes | Both sexes | Female only | True | True |
| C4Quan | 6 | BTNL2 | protein_coding | rs3135363 | 32389648 | 1.36e-13 | 1 | 0 | 0 | 0 | 0 | 1 | Both sexes | Both sexes | Both sexes | True | True |
| C4Quan | 6 | HCP5 | sense_overlapping | rs14597 | 31440014 | 6.34e-13 | 1 | 0 | 0 | 0 | 0 | 1 | Both sexes | Both sexes | Both sexes | True | True |
| C4Quan | 6 | XXbac-BPG254F23.7 | lincRNA | rs9275602 | 32682812 | 4.48e-11 | 1 | 0 | 0 | 0 | 0 | 1 | Both sexes | Both sexes | Female only | True | True |
| C4Quan | 6 | LINC00243 | processed_transcript | rs1264372 | 30769726 | 6.67e-11 | 1 | 0 | 0 | 1 | 0 | 0 | Female only | Both sexes | Female only | True | True |
| C4Quan | 6 | BAG6 | protein_coding | rs3130048 | 31613739 | 9.92e-11 | 1 | 0 | 0 | 0 | 0 | 1 | Both sexes | Both sexes | Both sexes | True | True |
| C4Quan | 6 | NOTCH4 | protein_coding | rs16869834 | 32179709 | 1.45e-10 | 1 | 0 | 0 | 0 | 0 | 1 | Both sexes | Both sexes | Female only | True | True |
| C4Quan | 6 | C2 | protein_coding | rs10947223 | 31872383 | 2.18e-10 | 1 | 0 | 1 | 0 | 0 | 0 | Female only | Female only | Female only | True | True |
| C4Quan | 6 | EHMT2 | protein_coding | rs652888 | 31851234 | 3.38e-10 | 1 | 0 | 0 | 0 | 0 | 1 | Female only | Both sexes | Female only | True | True |
| C4Quan | 6 | USP8P1 | pseudogene | rs2524084 | 31241639 | 4.85e-10 | 1 | 0 | 0 | 0 | 0 | 1 | Female only | Both sexes | Female only | True | True |
| C4Quan | 6 | TAP2 | protein_coding | rs62397687 | 32802009 | 2.85e-09 | 1 | 0 | 0 | 0 | 0 | 1 | Both sexes | Both sexes | Both sexes | True | True |
| C4Quan | 6 | BAG6 | protein_coding | rs1077394 | 31610384 | 5.07e-09 | 1 | 0 | 0 | 0 | 0 | 1 | Female only | Both sexes | Female only | True | True |
| C4Quan | 6 | PSORS1C1 | protein_coding | rs545914110 | 31091992 | 9.93e-09 | 1 | 0 | 0 | 0 | 0 | 1 | Female only | Both sexes | Female only | True | True |
| C4Quan | 6 | XXbac-BPG181B23.7 | lincRNA | rs9266791 | 31358943 | 1.11e-08 | 1 | 0 | 0 | 0 | 1 | 0 | Female only | Both sexes | Female only | True | True |
| C4Quan | 6 | HCP5 | sense_overlapping | rs2596534 | 31445345 | 2.03e-08 | 1 | 0 | 0 | 1 | 0 | 0 | Female only | Both sexes | Female only | True | True |
| C4Quan | 6 | PSORS1C1 | protein_coding | rs559951725 | 31100869 | 2.53e-08 | 1 | 0 | 0 | 1 | 0 | 0 | Female only | Both sexes | Female only | True | True |
| C4Quan | 6 | VARS | protein_coding | rs909267 | 31746548 | 3.06e-08 | 1 | 0 | 0 | 1 | 0 | 0 | Female only | Both sexes | Female only | True | True |
| CBil | 2 | UGT1A8 | protein_coding | rs7571337 | 234566418 | 2.15e-18 | 1 | 0 | 0 | 0 | 0 | 1 | Both sexes | Both sexes | Both sexes | True | False |
| CBil | 2 | UGT1A8 | protein_coding | rs28946889 | 234671462 | 5.44e-13 | 1 | 0 | 0 | 0 | 0 | 1 | Female only | Both sexes | Female only | True | False |
| CBil | 2 | UGT1A11P | pseudogene | rs2741023 | 234516714 | 1.94e-12 | 1 | 0 | 0 | 0 | 0 | 1 | Both sexes | Both sexes | Both sexes | True | False |
| CNucCl | 2 | LINC01101 | protein_coding | rs11684442 | 121228562 | 2.25e-08 | 1 | 1 | 0 | 0 | 0 | 0 | Female only | Female only | Female only | False | False |
| CPK | 19 | CTD-2337J16.1 | lincRNA | rs2361797 | 54753543 | 3.28e-09 | 1 | 0 | 1 | 0 | 0 | 0 | Female only | Female only | Female only | True | False |

| pheno | CHR | Gene_Symbol | biotype | lead_snp | lead_BP | lead_P | TOTAL | NSIG | S05 | S01 | S001 | S0001 | independent_loci_in_sex_P1e6 | independent_loci_in_sex_P1e2 | independent_loci_in_sex_P5e8 | in_gwas_catalog | in_HLA |
|---|---|---|---|---|---|---|---|---|---|---|---|---|---|---|---|---|---|
| CPK | 19 | AC010518.3 | pseudogene | rs427366 | 54795299 | 4.26e-08 | 1 | 1 | 0 | 0 | 0 | 0 | Female only | Female only | Female only | True | False |
| Ca | 3 | CASR | protein_coding | rs6808422 | 122017186 | 5.78e-13 | 1 | 0 | 0 | 0 | 0 | 1 | Female only | Both sexes | Female only | True | False |
| Chol | 2 | TDRD15 | protein_coding | rs71435594 | 21309335 | 6.96e-09 | 1 | 0 | 0 | 0 | 0 | 1 | Female only | Both sexes | Female only | True | False |
| Chol | 2 | AC067959.1 | lincRNA | rs35239705 | 21451687 | 3.51e-08 | 1 | 1 | 0 | 0 | 0 | 0 | Female only | Female only | Female only | True | False |
| Creat | 16 | PDILT | protein_coding | rs77924615 | 20392332 | 8.26e-16 | 1 | 0 | 0 | 0 | 0 | 1 | Both sexes | Both sexes | Both sexes | True | False |
| D25OHT | 11 | SPON1 | processed_transcript | rs4757244 | 14258655 | 4.25e-08 | 1 | 1 | 0 | 0 | 0 | 0 | Female only | Female only | Female only | True | False |
| EGFR | 16 | PDILT | protein_coding | rs77924615 | 20392332 | 6.57e-15 | 1 | 0 | 0 | 0 | 0 | 1 | Both sexes | Both sexes | Both sexes | True | False |
| FT4 | 1 | DIO1 | protein_coding | rs11206244 | 54375701 | 3.85e-09 | 1 | 1 | 0 | 0 | 0 | 0 | Female only | Female only | Female only | True | False |
| Gluc | 7 | GCK | protein_coding | rs12534623 | 44192203 | 3.28e-09 | 1 | 0 | 1 | 0 | 0 | 0 | Female only | Female only | Female only | True | False |
| HDL.C | 16 | CETP | protein_coding | rs4783961 | 56994894 | 1.36e-17 | 1 | 0 | 0 | 0 | 0 | 1 | Both sexes | Both sexes | Both sexes | True | False |

| pheno | CHR | Gene_Symbol | biotype | lead_snp | lead_BP | lead_P | TOTAL | NSIG | S05 | S01 | S001 | S0001 | independent_loci_in_sex_P1e6 | independent_loci_in_sex_P1e2 | independent_loci_in_sex_P5e8 | in_gwas_catalog | in_HLA |
|---|---|---|---|---|---|---|---|---|---|---|---|---|---|---|---|---|---|
| HDL.C | 8 | AC100802.3 | antisense | rs7015766 | 19939049 | 6.18e-15 | 1 | 0 | 0 | 0 | 0 | 1 | Both sexes | Both sexes | Both sexes | True | False |
| HDL.C | 16 | CETP | protein_coding | rs11076174 | 57003146 | 4.22e-13 | 1 | 0 | 0 | 0 | 0 | 1 | Both sexes | Both sexes | Both sexes | True | False |
| HDL.C | 16 | CETP | protein_coding | rs56208677 | 57010232 | 8.71e-13 | 1 | 0 | 0 | 0 | 0 | 1 | Female only | Both sexes | Female only | True | False |
| HDL.C | 8 | AC100802.3 | antisense | rs78358410 | 19889399 | 2.41e-12 | 1 | 0 | 0 | 0 | 0 | 1 | Both sexes | Both sexes | Both sexes | True | False |
| HDL.C | 16 | CETP | protein_coding | rs1801706 | 57017662 | 3.20e-09 | 1 | 0 | 0 | 1 | 0 | 0 | Female only | Both sexes | Female only | True | False |
| HDL.C | 8 | AC100802.3 | antisense | rs10103634 | 19890612 | 3.29e-09 | 1 | 0 | 0 | 0 | 0 | 1 | Both sexes | Both sexes | Male only | True | False |
| HDL.C | 11 | PCSK7 | protein_coding | rs59781045 | 117095283 | 2.37e-08 | 1 | 0 | 0 | 0 | 1 | 0 | Female only | Both sexes | Female only | True | False |
| HEIGHT | 1 | MTMR11 | protein_coding | rs11205303 | 149906413 | 8.76e-12 | 1 | 0 | 0 | 0 | 0 | 1 | Female only | Both sexes | Female only | True | False |
| Height | 1 | MTMR11 | protein_coding | rs11205303 | 149906413 | 1.48e-15 | 1 | 0 | 0 | 0 | 0 | 1 | Both sexes | Both sexes | Both sexes | True | False |
| Height | 19 | ADAMTS10 | protein_coding | rs11670030 | 8669867 | 4.74e-13 | 1 | 0 | 0 | 0 | 0 | 1 | Both sexes | Both sexes | Both sexes | True | False |
| Height | 15 | ADAMTSL3 | protein_coding | rs4843154 | 84419524 | 7.56e-11 | 1 | 0 | 1 | 0 | 0 | 0 | Female only | Female only | Female only | True | False |
| Height | 19 | MYO1F | protein_coding | rs4542783 | 8642160 | 8.91e-09 | 1 | 0 | 0 | 1 | 0 | 0 | Female only | Both sexes | Female only | True | False |
| Height | 11 | TMEM134 | protein_coding | rs12795957 | 67242216 | 1.44e-08 | 1 | 0 | 0 | 0 | 0 | 1 | Both sexes | Both sexes | Female only | True | False |
| Height | 2 | NPPC | protein_coding | rs3107179 | 232788325 | 1.50e-08 | 1 | 0 | 0 | 0 | 0 | 1 | Male only | Both sexes | Male only | True | False |
| Height | 15 | PML | protein_coding | rs5742915 | 74336633 | 1.90e-08 | 1 | 0 | 0 | 0 | 0 | 1 | Female only | Both sexes | Female only | True | False |

| pheno | CHR | Gene_Symbol | biotype | lead_snp | lead_BP | lead_P | TOTAL | NSIG | S05 | S01 | S001 | S0001 | independent_loci_in_sex_P1e6 | independent_loci_in_sex_P1e2 | independent_loci_in_sex_P5e8 | in_gwas_catalog | in_HLA |
|---|---|---|---|---|---|---|---|---|---|---|---|---|---|---|---|---|---|
| Height | 12 | CCND2 | protein_coding | rs3217833 | 4393474 | 2.40e-08 | 1 | 1 | 0 | 0 | 0 | 0 | Female only | Female only | Female only | True | False |
| Height | 2 | NPPC | protein_coding | rs2580816 | 232797966 | 3.18e-08 | 1 | 0 | 0 | 0 | 0 | 1 | Female only | Both sexes | Female only | True | False |
| Hgb | 22 | MPST | protein_coding | rs5756489 | 37417197 | 2.01e-09 | 1 | 0 | 0 | 0 | 0 | 1 | Female only | Both sexes | Female only | True | False |
| Hgb | 22 | TST | protein_coding | rs34374682 | 37408511 | 2.83e-08 | 1 | 0 | 0 | 0 | 1 | 0 | Female only | Both sexes | Female only | True | False |
| IGAB | 9 | TBC1D13 | protein_coding | rs3750323 | 131570274 | 6.51e-17 | 1 | 0 | 0 | 0 | 0 | 1 | Both sexes | Both sexes | Both sexes | False | False |
| IGRE | 9 | TBC1D13 | protein_coding | rs3750323 | 131570274 | 6.32e-30 | 1 | 0 | 0 | 0 | 0 | 1 | Both sexes | Both sexes | Both sexes | True | False |
| Iron | 22 | MPST | protein_coding | rs5756489 | 37417197 | 1.46e-12 | 1 | 0 | 0 | 0 | 0 | 1 | Both sexes | Both sexes | Female only | True | False |
| LDL.C | 19 | APOE | protein_coding | rs75627662 | 45413576 | 1.38e-14 | 1 | 0 | 0 | 0 | 0 | 1 | Both sexes | Both sexes | Both sexes | True | False |
| LYMPAB | 6 | RPL3P2 | pseudogene | rs2253491 | 31249217 | 1.76e-08 | 1 | 0 | 0 | 0 | 0 | 1 | Female only | Both sexes | Female only | True | True |
| LYMPAB | 6 | CFB | protein_coding | rs2072633 | 31919578 | 2.49e-08 | 1 | 0 | 0 | 0 | 0 | 1 | Male only | Both sexes | Male only | True | True |
| MBRat | 19 | CTD-2337J16.1 | lincRNA | rs2361797 | 54753543 | 3.23e-08 | 1 | 0 | 0 | 0 | 1 | 0 | Male only | Both sexes | Male only | True | False |
| MCH | 22 | MPST | protein_coding | rs5756489 | 37417197 | 2.84e-49 | 1 | 0 | 0 | 0 | 0 | 1 | Both sexes | Both sexes | Both sexes | True | False |
| MCH | 22 | TMPRSS6 | protein_coding | rs881144 | 37471290 | 1.27e-23 | 1 | 0 | 0 | 0 | 0 | 1 | Both sexes | Both sexes | Both sexes | True | False |
| MCH | 6 | HBS1L | protein_coding | rs13192235 | 135346351 | 2.36e-20 | 1 | 0 | 0 | 0 | 0 | 1 | Both sexes | Both sexes | Both sexes | True | False |
| MCH | 6 | LRRC16A | protein_coding | rs77042711 | 25413416 | 3.37e-19 | 1 | 0 | 0 | 0 | 0 | 1 | Both sexes | Both sexes | Both sexes | True | False |
| MCH | 22 | TMPRSS6 | protein_coding | rs4140589 | 37499019 | 8.95e-16 | 1 | 0 | 0 | 0 | 0 | 1 | Both sexes | Both sexes | Both sexes | True | False |
| MCH | 6 | BTN1A1 | protein_coding | rs13194984 | 26500563 | 9.55e-16 | 1 | 0 | 0 | 0 | 0 | 1 | Both sexes | Both sexes | Both sexes | True | False |
| MCH | 22 | TST | protein_coding | rs34374682 | 37408511 | 1.33e-15 | 1 | 0 | 0 | 0 | 0 | 1 | Both sexes | Both sexes | Female only | True | False |
| MCH | 22 | TMPRSS6 | protein_coding | rs926331 | 37510072 | 4.70e-13 | 1 | 0 | 0 | 0 | 0 | 1 | Both sexes | Both sexes | Female only | True | False |
| MCH | 2 | MIR4435-1HG | lincRNA | rs112917684 | 112137054 | 1.36e-11 | 1 | 0 | 0 | 1 | 0 | 0 | Female only | Both sexes | Female only | True | False |
| MCH | 6 | RNU6-1259P | snRNA | rs55964643 | 26340872 | 1.84e-11 | 1 | 0 | 0 | 0 | 0 | 1 | Both sexes | Both sexes | Both sexes | True | False |
| MCH | 19 | HDGFRP2 | protein_coding | rs12984070 | 4483485 | 6.14e-11 | 1 | 0 | 0 | 0 | 0 | 1 | Male only | Both sexes | Male only | True | False |
| MCH | 20 | RBM38 | protein_coding | rs99595 | 55989808 | 1.65e-09 | 1 | 0 | 0 | 0 | 0 | 1 | Female only | Both sexes | Female only | True | False |
| MCH | 6 | LRRC16A | protein_coding | rs407934 | 25504562 | 1.83e-09 | 1 | 0 | 0 | 0 | 0 | 1 | Female only | Both sexes | Female only | True | False |
| MCH | 6 | RP11-12A2.3 | lincRNA | rs12205386 | 139837171 | 1.65e-08 | 1 | 0 | 0 | 0 | 1 | 0 | Female only | Both sexes | Female only | True | False |
| MCH | 4 | AC006552.1 | lincRNA | rs1005656 | 55404602 | 2.66e-08 | 1 | 0 | 1 | 0 | 0 | 0 | Female only | Female only | Female only | True | False |

| pheno | CHR | Gene_Symbol | biotype | lead_snp | lead_BP | lead_P | TOTAL | NSIG | S05 | S01 | S001 | S0001 | independent_loci_in_sex_P1e6 | independent_loci_in_sex_P1e2 | independent_loci_in_sex_P5e8 | in_gwas_catalog | in_HLA |
|---|---|---|---|---|---|---|---|---|---|---|---|---|---|---|---|---|---|
| MCH | 4 | RP11-545H22.1 | lincRNA | rs10517334 | 55301708 | 3.35e-08 | 1 | 0 | 0 | 0 | 1 | 0 | Female only | Both sexes | Female only | True | False |
| MCHC | 22 | MPST | protein_coding | rs5756489 | 37417197 | 7.35e-25 | 1 | 0 | 0 | 0 | 0 | 1 | Both sexes | Both sexes | Both sexes | True | False |
| MCHC | 22 | TMPRSS6 | protein_coding | rs4140589 | 37499019 | 4.34e-14 | 1 | 0 | 0 | 0 | 1 | 0 | Female only | Both sexes | Female only | True | False |
| MCHC | 22 | TST | protein_coding | rs34374682 | 37408511 | 5.75e-13 | 1 | 0 | 0 | 0 | 0 | 1 | Both sexes | Both sexes | Both sexes | True | False |
| MCHC | 6 | BTN1A1 | protein_coding | rs13194984 | 26500563 | 2.11e-12 | 1 | 0 | 0 | 0 | 0 | 1 | Both sexes | Both sexes | Both sexes | True | False |
| MCHC | 6 | TRIM31 | protein_coding | rs2523990 | 30077229 | 4.55e-11 | 1 | 0 | 0 | 0 | 1 | 0 | Female only | Both sexes | Female only | True | True |
| MCHC | 6 | LRRC16A | protein_coding | rs77042711 | 25413416 | 7.59e-11 | 1 | 0 | 0 | 0 | 0 | 1 | Female only | Both sexes | Female only | True | False |
| MCHC | 22 | TMPRSS6 | protein_coding | rs881144 | 37471290 | 7.72e-11 | 1 | 0 | 0 | 0 | 0 | 1 | Both sexes | Both sexes | Both sexes | True | False |
| MCHC | 16 | PIEZO1 | protein_coding | rs8050504 | 88797568 | 1.88e-10 | 1 | 0 | 0 | 1 | 0 | 0 | Female only | Both sexes | Female only | True | False |
| MCHC | 16 | PIEZO1 | protein_coding | rs2242165 | 88807643 | 1.82e-09 | 1 | 0 | 0 | 0 | 0 | 1 | Both sexes | Both sexes | Female only | True | False |
| MCHC | 18 | RP11-116O18.3 | antisense | rs1484878 | 43303834 | 6.04e-09 | 1 | 0 | 0 | 1 | 0 | 0 | Male only | Both sexes | Male only | True | False |
| MCHC | 22 | TMPRSS6 | protein_coding | rs926331 | 37510072 | 2.24e-08 | 1 | 0 | 0 | 0 | 0 | 1 | Female only | Both sexes | Female only | True | False |
| MCHC | 6 | HIST1H2APS2 | pseudogene | rs1892247 | 25887657 | 4.01e-08 | 1 | 0 | 0 | 0 | 0 | 1 | Female only | Both sexes | Female only | True | False |
| MONOAB | 8 | CCDC26 | lincRNA | rs9649961 | 130577382 | 6.13e-12 | 1 | 0 | 0 | 0 | 0 | 1 | Female only | Both sexes | Female only | True | False |
| MONOAB | 3 | ACKR2 | protein_coding | rs2228467 | 42906116 | 2.39e-10 | 1 | 0 | 0 | 0 | 1 | 0 | Female only | Both sexes | Female only | True | False |
| MONOAB | 7 | CDK6 | protein_coding | rs445 | 92408370 | 1.47e-09 | 1 | 0 | 0 | 0 | 0 | 1 | Both sexes | Both sexes | Female only | True | False |
| MONOAB | 16 | IRF8 | protein_coding | rs12922615 | 85926695 | 5.07e-09 | 1 | 0 | 0 | 0 | 1 | 0 | Female only | Both sexes | Female only | True | False |
| MONOAB | 16 | IRF8 | protein_coding | rs4843862 | 85961459 | 1.82e-08 | 1 | 0 | 0 | 0 | 0 | 1 | Both sexes | Both sexes | Both sexes | True | False |
| MONOAB | 3 | KLHL40 | protein_coding | rs3888652 | 42726718 | 3.68e-08 | 1 | 0 | 0 | 0 | 1 | 0 | Female only | Both sexes | Female only | True | False |
| pheno | CHR | Gene_Symbol | biotype | lead_snp | lead_BP | lead_P | TOTAL | NSIG | S05 | S01 | S001 | S0001 | independent_loci_in_sex_P1e6 | independent_loci_in_sex_P1e2 | independent_loci_in_sex_P5e8 | in_gwas_catalog | in_HLA |
| MONOAB | 3 | ACKR2 | protein_coding | rs760194 | 42918238 | 4.62e-08 | 1 | 0 | 0 | 1 | 0 | 0 | Female only | Both sexes | Female only | True | False |
| MONORE | 16 | IRF8 | protein_coding | rs4843862 | 85961459 | 2.43e-20 | 1 | 0 | 0 | 0 | 0 | 1 | Both sexes | Both sexes | Both sexes | True | False |
| MONORE | 3 | C3orf27 | protein_coding | rs2713599 | 128295777 | 2.07e-19 | 1 | 0 | 0 | 0 | 0 | 1 | Both sexes | Both sexes | Both sexes | True | False |
| MONORE | 3 | ACKR2 | protein_coding | rs2228467 | 42906116 | 1.40e-13 | 1 | 0 | 0 | 0 | 0 | 1 | Both sexes | Both sexes | Female only | True | False |
| MONORE | 16 | IRF8 | protein_coding | rs12922615 | 85926695 | 8.32e-13 | 1 | 0 | 0 | 0 | 0 | 1 | Both sexes | Both sexes | Both sexes | True | False |
| MONORE | 16 | RP11-542M13.3 | lincRNA | rs908987 | 85984889 | 9.35e-13 | 1 | 0 | 0 | 1 | 0 | 0 | Female only | Both sexes | Female only | True | False |

| pheno | CHR | Gene_Symbol | biotype | lead_snp | lead_BP | lead_P | TOTAL | NSIG | S05 | S01 | S001 | S0001 | independent_loci_in_sex_P1e6 | independent_loci_in_sex_P1e2 | independent_loci_in_sex_P5e8 | in_gwas_catalog | in_HLA |
|---|---|---|---|---|---|---|---|---|---|---|---|---|---|---|---|---|---|
| MONORE | 9 | RP11-202G18.1 | lincRNA | rs62569868 | 114046613 | 1.61e-10 | 1 | 0 | 0 | 0 | 0 | 1 | Both sexes | Both sexes | Female only | True | False |
| MONORE | 8 | CCDC26 | lincRNA | rs9649961 | 130577382 | 3.42e-10 | 1 | 0 | 0 | 0 | 0 | 1 | Both sexes | Both sexes | Both sexes | True | False |
| MONORE | 16 | RP11-542M13.3 | lincRNA | rs731707 | 85980968 | 6.06e-10 | 1 | 0 | 0 | 0 | 0 | 1 | Both sexes | Both sexes | Both sexes | True | False |
| MONORE | 9 | RP11-202G18.1 | lincRNA | rs80085844 | 113923299 | 6.74e-10 | 1 | 0 | 1 | 0 | 0 | 0 | Female only | Female only | Female only | True | False |
| MONORE | 16 | RP11-118F19.1 | lincRNA | rs80025778 | 85597804 | 2.51e-09 | 1 | 1 | 0 | 0 | 0 | 0 | Female only | Female only | Female only | True | False |
| MONORE | 3 | C3orf27 | protein_coding | rs73203451 | 128284724 | 3.16e-08 | 1 | 0 | 0 | 0 | 1 | 0 | Female only | Both sexes | Female only | True | False |
| MONORE | 16 | C16orf74 | protein_coding | rs72805116 | 85792967 | 4.28e-08 | 1 | 0 | 0 | 0 | 1 | 0 | Female only | Both sexes | Female only | True | False |
| MPV | 12 | WDR66 | protein_coding | rs7961894 | 122365583 | 1.35e-136 | 1 | 0 | 0 | 0 | 0 | 1 | Both sexes | Both sexes | Both sexes | True | False |
| MPV | 12 | TMEM120B | protein_coding | rs146870349 | 122138964 | 4.44e-68 | 1 | 0 | 0 | 0 | 0 | 1 | Both sexes | Both sexes | Both sexes | True | False |
| MPV | 10 | NRBF2 | protein_coding | rs7100693 | 64920588 | 2.26e-30 | 1 | 0 | 0 | 0 | 0 | 1 | Both sexes | Both sexes | Both sexes | True | False |
| MPV | 12 | TMEM120B | protein_coding | 12:122219787 | 122219787 | 5.48e-28 | 1 | 0 | 0 | 0 | 0 | 1 | Both sexes | Both sexes | Both sexes | True | False |
| MPV | 7 | CTB-111H14.1 | sense_overlapping | rs17153388 | 106349163 | 5.24e-27 | 1 | 0 | 0 | 0 | 0 | 1 | Both sexes | Both sexes | Both sexes | True | False |
| MPV | 20 | SIRPA | protein_coding | rs3197744 | 1918487 | 5.02e-19 | 1 | 0 | 0 | 0 | 0 | 1 | Both sexes | Both sexes | Both sexes | True | False |
| MPV | 5 | F2R | protein_coding | rs2227819 | 76012745 | 2.90e-17 | 1 | 0 | 0 | 0 | 0 | 1 | Both sexes | Both sexes | Both sexes | True | False |
| MPV | 12 | PSMD9 | protein_coding | rs34149356 | 122332616 | 5.71e-17 | 1 | 0 | 0 | 0 | 0 | 1 | Both sexes | Both sexes | Both sexes | True | False |
| MPV | 11 | RIC8A | protein_coding | rs2293169 | 211015 | 6.27e-14 | 1 | 0 | 0 | 0 | 0 | 1 | Female only | Both sexes | Female only | True | False |
| MPV | 11 | PSMD13 | protein_coding | rs56257551 | 259575 | 2.36e-13 | 1 | 0 | 0 | 0 | 0 | 1 | Both sexes | Both sexes | Female only | True | False |
| MPV | 8 | ZFPM2 | protein_coding | rs16873365 | 106558235 | 1.40e-11 | 1 | 0 | 0 | 0 | 0 | 1 | Both sexes | Both sexes | Female only | True | False |
| MPV | 9 | DOCK8 | protein_coding | rs10757606 | 274108 | 2.82e-11 | 1 | 0 | 1 | 0 | 0 | 0 | Female only | Female only | Female only | True | False |
| MPV | 19 | TPM4 | protein_coding | rs11086023 | 16200378 | 4.91e-11 | 1 | 0 | 0 | 0 | 0 | 1 | Male only | Both sexes | Male only | True | False |
| MPV | 1 | DNM3 | protein_coding | rs9425824 | 171884170 | 1.11e-10 | 1 | 0 | 0 | 0 | 0 | 1 | Both sexes | Both sexes | Both sexes | True | False |
| MPV | 8 | ZFPM2 | protein_coding | rs12675041 | 106529045 | 1.66e-10 | 1 | 0 | 0 | 0 | 0 | 1 | Both sexes | Both sexes | Both sexes | True | False |
| MPV | 1 | NUAK2 | protein_coding | rs74786162 | 205271007 | 2.36e-10 | 1 | 0 | 0 | 0 | 0 | 1 | Both sexes | Both sexes | Both sexes | True | False |
| MPV | 5 | MEF2C | protein_coding | rs114694170 | 88180196 | 3.69e-10 | 1 | 0 | 0 | 0 | 0 | 1 | Both sexes | Both sexes | Male only | True | False |
| MPV | 10 | JMJD1C | protein_coding | rs117452816 | 65043795 | 4.51e-10 | 1 | 0 | 0 | 0 | 1 | 0 | Female only | Both sexes | Female only | True | False |
| MPV | 17 | TAOK1 | protein_coding | rs74925747 | 27744438 | 5.08e-10 | 1 | 0 | 0 | 0 | 0 | 1 | Female only | Both sexes | Female only | True | False |

| pheno | CHR | Gene_Symbol | biotype | lead_snp | lead_BP | lead_P | TOTAL | NSIG | S05 | S01 | S001 | S0001 | independent_loci_in_sex_P1e6 | independent_loci_in_sex_P1e2 | independent_loci_in_sex_P5e8 | in_gwas_catalog | in_HLA |
|---|---|---|---|---|---|---|---|---|---|---|---|---|---|---|---|---|---|
| MPV | 10 | JMJD1C | protein_coding | rs17813747 | 65166670 | 9.01e-10 | 1 | 0 | 0 | 0 | 0 | 1 | Both sexes | Both sexes | Female only | True | False |
| MPV | 2 | CPS1 | protein_coding | rs4673553 | 211608379 | 1.16e-09 | 1 | 1 | 0 | 0 | 0 | 0 | Female only | Female only | Female only | True | False |
| MPV | 12 | RSRC2 | protein_coding | rs78525750 | 123001692 | 1.86e-09 | 1 | 0 | 0 | 0 | 0 | 1 | Both sexes | Both sexes | Female only | True | False |
| MPV | 2 | CAPN10 | protein_coding | rs12465143 | 241537101 | 2.00e-09 | 1 | 0 | 0 | 0 | 1 | 0 | Female only | Both sexes | Female only | True | False |
| MPV | 6 | MICA | protein_coding | rs9266806 | 31378022 | 4.71e-09 | 1 | 0 | 0 | 1 | 0 | 0 | Female only | Both sexes | Female only | True | True |
| MPV | 22 | ARFGAP3 | protein_coding | rs35343665 | 43240703 | 9.77e-09 | 1 | 0 | 1 | 0 | 0 | 0 | Female only | Female only | Female only | True | False |
| MPV | 7 | CTB-111H14.1 | sense_overlapping | rs7801138 | 106285956 | 1.72e-08 | 1 | 0 | 0 | 0 | 1 | 0 | Female only | Both sexes | Female only | True | False |
| MPV | 20 | SIRPA | protein_coding | rs35726302 | 1857413 | 4.02e-08 | 1 | 0 | 0 | 0 | 0 | 1 | Female only | Both sexes | Female only | True | False |
| MPV | 12 | CBX5 | protein_coding | rs4759348 | 54630517 | 4.27e-08 | 1 | 0 | 0 | 0 | 0 | 1 | Female only | Both sexes | Female only | True | False |
| MPV | 1 | CNTN2 | protein_coding | rs7514078 | 205002356 | 4.31e-08 | 1 | 0 | 0 | 0 | 1 | 0 | Female only | Both sexes | Female only | True | False |
| MPV | 18 | DOK6 | protein_coding | rs7237652 | 67511067 | 4.37e-08 | 1 | 0 | 0 | 0 | 0 | 1 | Female only | Both sexes | Female only | True | False |
| Monocy | 3 | C3orf27 | protein_coding | rs2713599 | 128295777 | 1.45e-13 | 1 | 0 | 0 | 0 | 0 | 1 | Both sexes | Both sexes | Both sexes | True | False |
| Monocy | 16 | IRF8 | protein_coding | rs4843862 | 85961459 | 5.12e-10 | 1 | 0 | 0 | 0 | 0 | 1 | Both sexes | Both sexes | Both sexes | True | False |
| Monocy | 9 | RP11-202G18.1 | lincRNA | rs62569868 | 114046613 | 5.01e-09 | 1 | 0 | 0 | 0 | 0 | 1 | Female only | Both sexes | Female only | True | False |
| Monocy | 3 | TMED10P2 | pseudogene | rs59855017 | 128268659 | 3.47e-08 | 1 | 0 | 0 | 0 | 0 | 1 | Male only | Both sexes | Male only | True | False |
| NEUTAB | 17 | MED24 | protein_coding | rs8078692 | 38215117 | 1.71e-14 | 1 | 0 | 0 | 0 | 0 | 1 | Both sexes | Both sexes | Both sexes | True | False |
| NEUTAB | 17 | RP11-387H17.4 | lincRNA | rs35557848 | 38087439 | 3.34e-08 | 1 | 0 | 0 | 0 | 1 | 0 | Male only | Both sexes | Male only | True | False |
| NEUTAB | 17 | GSDMA | protein_coding | rs62066853 | 38130492 | 3.61e-08 | 1 | 0 | 0 | 0 | 0 | 1 | Both sexes | Both sexes | Female only | True | False |
| NTAuto | 17 | MED24 | protein_coding | rs8078692 | 38215117 | 8.81e-15 | 1 | 0 | 0 | 0 | 0 | 1 | Both sexes | Both sexes | Both sexes | True | False |
| NTAuto | 17 | GSDMA | protein_coding | rs62066853 | 38130492 | 7.14e-11 | 1 | 0 | 0 | 0 | 0 | 1 | Both sexes | Both sexes | Female only | True | False |
| NtAbs | 17 | MED24 | protein_coding | rs8078692 | 38215117 | 1.03e-09 | 1 | 0 | 0 | 0 | 0 | 1 | Female only | Both sexes | Female only | True | False |
| NtAbs | 17 | GSDMA | protein_coding | rs62066853 | 38130492 | 3.22e-08 | 1 | 0 | 0 | 1 | 0 | 0 | Female only | Both sexes | Female only | True | False |
| PR_INTERVAL | 3 | SCN5A | protein_coding | rs7430191 | 38654024 | 6.45e-14 | 1 | 0 | 0 | 0 | 1 | 0 | Female only | Both sexes | Female only | True | False |
| PR_INTERVAL | 3 | SCN10A | protein_coding | rs12490478 | 38817699 | 2.23e-12 | 1 | 0 | 0 | 0 | 0 | 1 | Both sexes | Both sexes | Both sexes | True | False |
| PR_INTERVAL | 2 | MEIS1 | protein_coding | rs6721499 | 66743591 | 1.02e-11 | 1 | 0 | 1 | 0 | 0 | 0 | Female only | Female only | Female only | True | False |
| PR_INTERVAL | 2 | MEIS1 | protein_coding | rs2300484 | 66788314 | 2.73e-09 | 1 | 0 | 0 | 1 | 0 | 0 | Female only | Both sexes | Female only | True | False |

| pheno | CHR | Gene_Symbol | biotype | lead_snp | lead_BP | lead_P | TOTAL | NSIG | S05 | S01 | S001 | S0001 | independent_loci_in_sex_P1e6 | independent_loci_in_sex_P1e2 | independent_loci_in_sex_P5e8 | in_gwas_catalog | in_HLA |
|---|---|---|---|---|---|---|---|---|---|---|---|---|---|---|---|---|---|
| PR_INTERVAL | 12 | TBX3 | protein_coding | rs7302926 | 115088313 | 3.93e-08 | 1 | 1 | 0 | 0 | 0 | 0 | Female only | Female only | Female only | True | False |
| PT.inr | 13 | F10 | protein_coding | rs3211752 | 113787459 | 2.60e-08 | 1 | 0 | 0 | 0 | 0 | 1 | Male only | Both sexes | Male only | True | False |
| PTT.pt | 3 | RP11-573D15.8 | antisense | rs9871339 | 186476598 | 4.05e-09 | 1 | 0 | 0 | 0 | 1 | 0 | Male only | Both sexes | Male only | True | False |
| PTT.pt | 3 | KNG1 | protein_coding | rs5030111 | 186455478 | 9.07e-09 | 1 | 0 | 0 | 0 | 0 | 1 | Both sexes | Both sexes | Female only | True | False |
| PTT.pt | 3 | RP11-795A2.3 | pseudogene | rs56211697 | 186429997 | 1.22e-08 | 1 | 0 | 0 | 0 | 0 | 1 | Male only | Both sexes | Male only | True | False |
| PTT.pt | 3 | KNG1 | protein_coding | rs5030102 | 186446087 | 4.71e-08 | 1 | 0 | 0 | 0 | 0 | 1 | Both sexes | Both sexes | Male only | True | False |
| Pat.PT | 13 | F10 | protein_coding | rs3211752 | 113787459 | 4.12e-10 | 1 | 0 | 0 | 0 | 0 | 1 | Both sexes | Both sexes | Both sexes | True | False |
| Plt.Ct | 9 | RP11-307I14.4 | pseudogene | rs12005199 | 4763491 | 3.57e-23 | 1 | 0 | 0 | 0 | 0 | 1 | Both sexes | Both sexes | Both sexes | True | False |
| Plt.Ct | 12 | WDR66 | protein_coding | rs7961894 | 122365583 | 3.42e-09 | 1 | 0 | 1 | 0 | 0 | 0 | Female only | Female only | Female only | True | False |
| Plt.Ct | 12 | PTPN11 | protein_coding | rs11066301 | 112871372 | 1.03e-08 | 1 | 0 | 0 | 1 | 0 | 0 | Female only | Both sexes | Female only | True | False |
| QRS_DURATION | 3 | SCN5A | protein_coding | rs7430191 | 38654024 | 3.53e-12 | 1 | 0 | 0 | 0 | 0 | 1 | Both sexes | Both sexes | Female only | True | False |
| QRS_DURATION | 6 | SLC35F1 | protein_coding | rs7747363 | 118622057 | 4.11e-12 | 1 | 0 | 0 | 1 | 0 | 0 | Female only | Both sexes | Female only | True | False |
| QRS_DURATION | 3 | SCN5A | protein_coding | rs7375123 | 38670098 | 1.86e-09 | 1 | 0 | 0 | 0 | 0 | 1 | Male only | Both sexes | Male only | True | False |
| QRS_DURATION | 3 | SCN5A | protein_coding | rs6772948 | 38615316 | 6.21e-09 | 1 | 0 | 0 | 1 | 0 | 0 | Female only | Both sexes | Female only | True | False |
| QTCB | 1 | NOS1AP | protein_coding | rs10429888 | 162024987 | 1.04e-08 | 1 | 0 | 0 | 0 | 0 | 1 | Female only | Both sexes | Female only | True | False |
| QT_INTERVAL | 6 | CEP85L | protein_coding | rs6911599 | 119009913 | 2.63e-09 | 1 | 0 | 0 | 0 | 0 | 1 | Female only | Both sexes | Female only | True | False |
| QT_INTERVAL_C | 1 | NOS1AP | protein_coding | rs10429888 | 162024987 | 1.88e-21 | 1 | 0 | 0 | 0 | 0 | 1 | Both sexes | Both sexes | Both sexes | True | False |
| QT_INTERVAL_C | 1 | NOS1AP | protein_coding | rs78694717 | 162111400 | 1.38e-11 | 1 | 0 | 0 | 0 | 0 | 1 | Both sexes | Both sexes | Female only | True | False |
| QT_INTERVAL_C | 11 | KCNQ1 | protein_coding | rs11824286 | 2472055 | 2.69e-08 | 1 | 0 | 0 | 0 | 0 | 1 | Male only | Both sexes | Male only | True | False |
| R.APACHE.TEMPERATURE | 12 | RP11-560G2.1 | antisense | rs78712959 | 75647582 | 1.08e-08 | 1 | 1 | 0 | 0 | 0 | 0 | Male only | Male only | Male only | False | False |

| pheno | CHR | Gene_Symbol | biotype | lead_snp | lead_BP | lead_P | TOTAL | NSIG | S05 | S01 | S001 | S0001 | independent_loci_in_sex_P1e6 | independent_loci_in_sex_P1e2 | independent_loci_in_sex_P5e8 | in_gwas_catalog | in_HLA |
|---|---|---|---|---|---|---|---|---|---|---|---|---|---|---|---|---|---|
| R.IP.IDEAL.BODY.WEIGHT | 1 | MTMR11 | protein_coding | rs11205303 | 149906413 | 9.62e-12 | 1 | 0 | 0 | 0 | 0 | 1 | Female only | Both sexes | Female only | False | False |
| R.RT..FEMALE.ADULT..IDEAL.BODY.WEIGHT | 1 | MTMR11 | protein_coding | rs11205303 | 149906413 | 8.68e-12 | 1 | 0 | 0 | 0 | 0 | 1 | Female only | Both sexes | Female only | False | False |
| R.RT..MALE.ADULT..IDEAL.BODY.WEIGHT | 1 | MTMR11 | protein_coding | rs11205303 | 149906413 | 8.68e-12 | 1 | 0 | 0 | 0 | 0 | 1 | Female only | Both sexes | Female only | False | False |
| RBC | 6 | HBS1L | protein_coding | rs13192235 | 135346351 | 1.51e-11 | 1 | 0 | 0 | 0 | 1 | 0 | Female only | Both sexes | Female only | True | False |
| RDW | 5 | FBN2 | protein_coding | 5:127561432 | 127561432 | 5.57e-18 | 1 | 0 | 0 | 0 | 0 | 1 | Both sexes | Both sexes | Both sexes | True | False |
| RDW | 22 | MPST | protein_coding | rs5756489 | 37417197 | 3.00e-16 | 1 | 0 | 0 | 0 | 0 | 1 | Both sexes | Both sexes | Female only | True | False |
| RDW | 22 | TMPRSS6 | protein_coding | rs9610643 | 37471385 | 2.42e-09 | 1 | 0 | 0 | 0 | 1 | 0 | Female only | Both sexes | Female only | True | False |
| RDW | 22 | TST | protein_coding | rs34374682 | 37408511 | 1.20e-08 | 1 | 0 | 1 | 0 | 0 | 0 | Female only | Female only | Female only | True | False |
| RDWSD | 5 | FBN2 | protein_coding | 5:127561432 | 127561432 | 1.87e-12 | 1 | 0 | 0 | 0 | 0 | 1 | Both sexes | Both sexes | Both sexes | True | False |
| SedRat | 1 | C1orf63 | protein_coding | rs1043879 | 25570081 | 4.44e-08 | 1 | 0 | 0 | 1 | 0 | 0 | Female only | Both sexes | Female only | True | False |
| TBil | 2 | UGT1A8 | protein_coding | rs929596 | 234674476 | 0 | 1 | 0 | 0 | 0 | 0 | 1 | Both sexes | Both sexes | Both sexes | True | False |
| TBil | 2 | UGT1A8 | protein_coding | rs7571337 | 234566418 | 1.34e-265 | 1 | 0 | 0 | 0 | 0 | 1 | Both sexes | Both sexes | Both sexes | True | False |
| TBil | 2 | UGT1A8 | protein_coding | rs28946889 | 234671462 | 5.58e-117 | 1 | 0 | 0 | 0 | 0 | 1 | Both sexes | Both sexes | Both sexes | True | False |
| TBil | 2 | UGT1A8 | protein_coding | rs2011404 | 234627937 | 1.18e-72 | 1 | 0 | 0 | 0 | 0 | 1 | Both sexes | Both sexes | Both sexes | True | False |
| TBil | 2 | MROH2A | protein_coding | rs17868346 | 234695087 | 2.17e-31 | 1 | 0 | 0 | 0 | 0 | 1 | Both sexes | Both sexes | Both sexes | True | False |
| TBil | 2 | USP40 | protein_coding | rs79031219 | 234458204 | 8.48e-28 | 1 | 0 | 0 | 0 | 0 | 1 | Both sexes | Both sexes | Both sexes | True | False |
| TBil | 2 | ATG16L1 | protein_coding | rs78583556 | 234153122 | 1.11e-17 | 1 | 0 | 0 | 0 | 0 | 1 | Both sexes | Both sexes | Both sexes | True | False |
| TBil | 12 | RP11-125O5.2 | protein_coding | rs4149033 | 21317810 | 2.14e-16 | 1 | 0 | 0 | 0 | 0 | 1 | Female only | Both sexes | Female only | True | False |
| TBil | 2 | MROH2A | protein_coding | rs879665 | 234740136 | 7.31e-15 | 1 | 0 | 0 | 0 | 0 | 1 | Both sexes | Both sexes | Both sexes | True | False |
| TBil | 2 | MROH2A | protein_coding | rs9808231 | 234708152 | 6.26e-12 | 1 | 0 | 0 | 0 | 0 | 1 | Female only | Both sexes | Female only | True | False |
| TBil | 2 | MROH2A | protein_coding | rs1500481 | 234702459 | 1.10e-10 | 1 | 0 | 0 | 0 | 0 | 1 | Female only | Both sexes | Female only | True | False |

| pheno | CHR | Gene_Symbol | biotype | lead_snp | lead_BP | lead_P | TOTAL | NSIG | S05 | S01 | S001 | S0001 | independent_loci_in_sex_P1e6 | independent_loci_in_sex_P1e2 | independent_loci_in_sex_P5e8 | in_gwas_catalog | in_HLA |
|---|---|---|---|---|---|---|---|---|---|---|---|---|---|---|---|---|---|
| TBil | 2 | MROH2A | protein_coding | rs759173 | 234704690 | 1.02e-09 | 1 | 0 | 0 | 0 | 1 | 0 | Female only | Both sexes | Female only | True | False |
| TIBC | 3 | TF | protein_coding | rs6762415 | 133478557 | 1.07e-12 | 1 | 0 | 0 | 0 | 0 | 1 | Male only | Both sexes | Male only | True | False |
| TProt | 6 | MUC22 | protein_coding | rs2523898 | 30993533 | 9.79e-09 | 1 | 0 | 0 | 1 | 0 | 0 | Male only | Both sexes | Male only | True | True |
| TSH | 9 | RP11-546O6.4 | lincRNA | rs7357631 | 100528320 | 6.65e-16 | 1 | 0 | 0 | 0 | 0 | 1 | Female only | Both sexes | Female only | True | False |
| TSH | 6 | PDE10A | protein_coding | rs3008050 | 166064041 | 1.26e-10 | 1 | 0 | 0 | 0 | 0 | 1 | Both sexes | Both sexes | Both sexes | True | False |
| TSH | 16 | AC009159.1 | miRNA | rs75380468 | 79694565 | 8.38e-10 | 1 | 0 | 0 | 0 | 0 | 1 | Both sexes | Both sexes | Both sexes | True | False |
| TSH | 6 | RP11-344J7.2 | lincRNA | rs1317840 | 43806921 | 3.66e-08 | 1 | 0 | 0 | 0 | 0 | 1 | Both sexes | Both sexes | Both sexes | True | False |
| Trigs | 8 | AC100802.3 | antisense | rs7015766 | 19939049 | 7.72e-18 | 1 | 0 | 0 | 0 | 0 | 1 | Both sexes | Both sexes | Both sexes | True | False |
| Trigs | 8 | AC100802.3 | antisense | rs78358410 | 19889399 | 4.63e-13 | 1 | 0 | 0 | 0 | 0 | 1 | Both sexes | Both sexes | Male only | True | False |
| Trigs | 8 | AC100802.3 | antisense | rs10103634 | 19890612 | 4.03e-10 | 1 | 0 | 0 | 0 | 0 | 1 | Both sexes | Both sexes | Male only | True | False |
| Trigs | 19 | APOC1P1 | pseudogene | rs141622900 | 45426792 | 2.32e-09 | 1 | 0 | 0 | 0 | 0 | 1 | Male only | Both sexes | Male only | True | False |
| Trigs | 11 | BUD13 | protein_coding | rs17440824 | 116634173 | 3.17e-08 | 1 | 0 | 0 | 0 | 0 | 1 | Female only | Both sexes | Female only | True | False |
| UricA | 4 | AC006499.2 | pseudogene | rs112042647 | 10291955 | 1.07e-12 | 1 | 0 | 0 | 1 | 0 | 0 | Female only | Both sexes | Female only | True | False |
| WBC | 17 | MED24 | protein_coding | rs8078692 | 38215117 | 2.05e-25 | 1 | 0 | 0 | 0 | 0 | 1 | Both sexes | Both sexes | Both sexes | True | False |
| WBC | 17 | GSDMA | protein_coding | rs62066853 | 38130492 | 2.35e-16 | 1 | 0 | 0 | 0 | 0 | 1 | Both sexes | Both sexes | Female only | True | False |
| WBC | 7 | CDK6 | protein_coding | rs445 | 92408370 | 1.07e-08 | 1 | 0 | 0 | 0 | 0 | 1 | Both sexes | Both sexes | Male only | True | False |
| Weight | 16 | FTO | protein_coding | rs7187961 | 53826034 | 2.00e-08 | 1 | 0 | 0 | 0 | 0 | 1 | Both sexes | Both sexes | Male only | True | False |
| X_percent_SAT | 6 | LRRC16A | protein_coding | rs181058325 | 25314597 | 1.75e-10 | 1 | 0 | 0 | 0 | 0 | 1 | Female only | Both sexes | Female only | False | False |
| X_percent_SAT | 6 | RNU6-1259P | snRNA | rs55964643 | 26340872 | 3.81e-10 | 1 | 0 | 0 | 0 | 0 | 1 | Male only | Both sexes | Male only | False | False |
| X_percent_SAT | 22 | MPST | protein_coding | rs5756489 | 37417197 | 4.32e-10 | 1 | 0 | 0 | 0 | 0 | 1 | Female only | Both sexes | Female only | False | False |
| X_percent_SAT | 6 | LRRC16A | protein_coding | rs77042711 | 25413416 | 1.53e-08 | 1 | 0 | 0 | 1 | 0 | 0 | Female only | Both sexes | Female only | False | False |


| pheno | CHR | Gene_Symbol | biotype | lead_snp | lead_BP | lead_P | TOTAL | NSIG | S05 | S01 | S001 | S0001 | independent_loci_in_sex_P1e6 | independent_loci_in_sex_P1e2 | independent_loci_in_sex_P5e8 | in_gwas_catalog | in_HLA |
|---|---|---|---|---|---|---|---|---|---|---|---|---|---|---|---|---|---|

| X_percent_SAT | 6 | BTN1A1 | protein_coding | rs13194984 | 26500563 | 2.47e-08 | 1 | 0 | 0 | 1 | 0 | 0 | Male only | Both sexes | Male only | False | False |

#### STable6: Independent loci for per-person median trait values that exist in GWAS catalog that were not previously reported (either any variant within the loci reported or gene previously reported) in GWAS catalog

421 rows × 13 columns

| lab | full_name | group | CHR | Gene_Symbol | biotype | lead_snp | lead_BP | lead_P | total_snps_in_loci | independent_loci_in_sex_P1e6 | independent_loci_in_sex_P1e2 | independent_loci_in_sex_P5e8 |
|---|---|---|---|---|---|---|---|---|---|---|---|---|
| ACTH | Corticotropin [Mass/volume] in Plasma | endocrine | 3 | EPHB1 | protein_coding | rs9851564 | 134924786 | 2.65e-09 | 11 | Male only | Male only | Male only |
| AN.GAP | Anion gap in Serum or Plasma | metabolic | 2 | GCKR | protein_coding | rs780094 | 27741237 | 7.50e-14 | 23 | Both sexes | Both sexes | Both sexes |
| AN.GAP | Anion gap in Serum or Plasma | metabolic | 2 | ZNF512 | protein_coding | rs3749147 | 27851918 | 2.55e-09 | 71 | Male only | Both sexes | Male only |
| AN.GAP | Anion gap in Serum or Plasma | metabolic | 2 | MRPL33 | protein_coding | rs12104449 | 27972833 | 1.43e-08 | 5 | Female only | Both sexes | Female only |
| AN.GAP | Anion gap in Serum or Plasma | metabolic | 13 | COL4A2 | protein_coding | rs2172724 | 110961409 | 1.98e-08 | 3 | Female only | Female only | Female only |
| AlkP | Alkaline phosphatase [Enzymatic activity/volume] in Serum or Plasma | liver | 8 | RP11-115J16.1 | lincRNA | rs11774915 | 9188762 | 1.22e-08 | 31 | Male only | Both sexes | Male only |
| AlkP | Alkaline phosphatase [Enzymatic activity/volume] in Serum or Plasma | liver | 8 | RP11-115J16.1 | lincRNA | rs878336 | 9225629 | 3.01e-08 | 17 | Male only | Both sexes | Male only |
| Amyl | Amylase [Enzymatic activity/volume] in Serum or Plasma | metabolic | 1 | RNPC3 | protein_coding | rs147499264 | 104082019 | 1.39e-10 | 11 | Female only | Both sexes | Female only |
| BASOAB | Basophils [#/volume] in Blood by Automated count | blood | 19 | CTD-2540B15.12 | lincRNA | rs67228209 | 33751033 | 5.61e-32 | 3 | Both sexes | Both sexes | Both sexes |
| BASOAB | Basophils [#/volume] in Blood by Automated count | blood | 19 | CTD-2540B15.12 | lincRNA | rs12461296 | 33774591 | 1.20e-21 | 3 | Both sexes | Both sexes | Both sexes |
| BASOAB | Basophils [#/volume] in Blood by Automated count | blood | 19 | CTD-2540B15.12 | lincRNA | rs4432367 | 33752920 | 1.68e-21 | 3 | Both sexes | Both sexes | Both sexes |
| BASOAB | Basophils [#/volume] in Blood by Automated count | blood | 19 | CTD-2540B15.12 | lincRNA | rs10414008 | 33758327 | 1.36e-17 | 5 | Both sexes | Both sexes | Both sexes |
| BASOAB | Basophils [#/volume] in Blood by Automated count | blood | 3 | RAB7A | protein_coding | rs35904610 | 128452108 | 1.56e-16 | 115 | Both sexes | Both sexes | Both sexes |
| BASOAB | Basophils [#/volume] in Blood by Automated count | blood | 3 | RAB7A | protein_coding | rs35151839 | 128546891 | 8.87e-14 | 45 | Both sexes | Both sexes | Female only |
| BASOAB | Basophils [#/volume] in Blood by Automated count | blood | 3 | RAB7A | protein_coding | rs56373544 | 128523503 | 9.94e-11 | 25 | Female only | Both sexes | Female only |
| BASOAB | Basophils [#/volume] in Blood by Automated count | blood | 19 | CTD-2540B15.12 | lincRNA | rs56079812 | 33755525 | 1.06e-10 | 3 | Female only | Both sexes | Female only |
| BASOAB | Basophils [#/volume] in Blood by Automated count | blood | 19 | CTD-2540B15.12 | lincRNA | rs4805869 | 33762602 | 3.80e-10 | 3 | Both sexes | Both sexes | Both sexes |
| BASOAB | Basophils [#/volume] in Blood by Automated count | blood | 19 | CTD-2540B15.12 | lincRNA | rs736289 | 33757062 | 1.28e-09 | 3 | Both sexes | Both sexes | Both sexes |
| BASOAB | Basophils [#/volume] in Blood by Automated count | blood | 1 | RP5-1052I5.2 | protein_coding | rs3893263 | 87711755 | 2.26e-09 | 7 | Both sexes | Both sexes | Female only |
| BASOAB | Basophils [#/volume] in Blood by Automated count | blood | 12 | RP11-335I12.2 | antisense | rs11176115 | 66658183 | 3.29e-09 | 41 | Female only | Both sexes | Female only |
| BASOAB | Basophils [#/volume] in Blood by Automated count | blood | 3 | TMED10P2 | pseudogene | rs7620032 | 128245104 | 3.45e-09 | 9 | Female only | Both sexes | Female only |
| BASOAB | Basophils [#/volume] in Blood by Automated count | blood | 3 | KIAA1257 | protein_coding | rs71331680 | 128662940 | 1.60e-08 | 5 | Female only | Both sexes | Female only |

| lab | full_name | group | CHR | Gene_Symbol | biotype | lead_snp | lead_BP | lead_P | total_snps_in_loci | independent_loci_in_sex_P 1e6 | independent_loci_in_sex_P 1e2 | independent_loci_in_sex_P 5e8 |
|---|---|---|---|---|---|---|---|---|---|---|---|---|
| BASOAB | Basophils [#/volume] in Blood by Automated count | blood | 6 | ZDHHC20P2 | pseudogene | rs2244020 | 31347451 | 2.83e-08 | 9 | Male only | Both sexes | Male only |
| BASOAB | Basophils [#/volume] in Blood by Automated count | blood | 6 | XXbac-BPG248L24.12 | pseudogene | rs9266233 | 31325430 | 3.09e-08 | 7 | Both sexes | Both sexes | Male only |
| BASOAB | Basophils [#/volume] in Blood by Automated count | blood | 14 | SLC7A8 | protein_coding | rs8010847 | 23594396 | 3.99e-08 | 3 | Female only | Both sexes | Female only |
| BASOM | Basophils/100 leukocytes in Blood by Manual count | immune/inflammatory | 16 | CMIP | protein_coding | rs75217481 | 81576331 | 4.89e-08 | 23 | Female only | Both sexes | Female only |
| BASOPHILS | BASOPHILS | immune/inflammatory | 23 | TENM1 | protein_coding | X:123595260:A:G | 123595260 | 1.47e-08 | 7 | Male only | Male only | Male only |
| BASORE | Basophils/100 leukocytes in Blood by Automated count | immune/inflammatory | 19 | CTD-2540B15.12 | lincRNA | rs67228209 | 33751033 | 2.60e-26 | 3 | Both sexes | Both sexes | Both sexes |
| BASORE | Basophils/100 leukocytes in Blood by Automated count | immune/inflammatory | 3 | RAB7A | protein_coding | rs61292315 | 128542609 | 8.50e-23 | 119 | Both sexes | Both sexes | Both sexes |
| BASORE | Basophils/100 leukocytes in Blood by Automated count | immune/inflammatory | 19 | CTD-2540B15.12 | lincRNA | rs12461296 | 33774591 | 6.69e-18 | 3 | Both sexes | Both sexes | Both sexes |
| BASORE | Basophils/100 leukocytes in Blood by Automated count | immune/inflammatory | 19 | CTD-2540B15.12 | lincRNA | rs4432367 | 33752920 | 8.08e-16 | 3 | Both sexes | Both sexes | Both sexes |
| BASORE | Basophils/100 leukocytes in Blood by Automated count | immune/inflammatory | 19 | CTD-2540B15.12 | lincRNA | rs10414008 | 33758327 | 2.15e-15 | 5 | Both sexes | Both sexes | Both sexes |
| BASORE | Basophils/100 leukocytes in Blood by Automated count | immune/inflammatory | 3 | RAB7A | protein_coding | rs3772157 | 128533990 | 3.75e-15 | 43 | Both sexes | Both sexes | Both sexes |
| BASORE | Basophils/100 leukocytes in Blood by Automated count | immune/inflammatory | 19 | CTD-2540B15.12 | lincRNA | rs4805867 | 33762257 | 1.52e-13 | 3 | Both sexes | Both sexes | Both sexes |
| BASORE | Basophils/100 leukocytes in Blood by Automated count | immune/inflammatory | 19 | CTD-2540B15.12 | lincRNA | rs56079812 | 33755525 | 4.59e-13 | 3 | Female only | Both sexes | Female only |
| BASORE | Basophils/100 leukocytes in Blood by Automated count | immune/inflammatory | 3 | RAB7A | protein_coding | rs56373544 | 128523503 | 2.28e-12 | 25 | Both sexes | Both sexes | Female only |
| BASORE | Basophils/100 leukocytes in Blood by Automated count | immune/inflammatory | 1 | LMO4 | protein_coding | rs4655844 | 87775058 | 1.87e-11 | 17 | Both sexes | Both sexes | Both sexes |
| BASORE | Basophils/100 leukocytes in Blood by Automated count | immune/inflammatory | 1 | RP5-1052I5.2 | protein_coding | rs3893263 | 87711755 | 2.03e-11 | 7 | Both sexes | Both sexes | Both sexes |
| BASORE | Basophils/100 leukocytes in Blood by Automated count | immune/inflammatory | 15 | RP11-96C21.1 | lincRNA | rs13379715 | 70738476 | 1.21e-10 | 155 | Both sexes | Both sexes | Male only |
| BASORE | Basophils/100 leukocytes in Blood by Automated count | immune/inflammatory | 18 | RP11-737G21.1 | pseudogene | rs11659727 | 23503073 | 2.13e-10 | 101 | Female only | Female only | Female only |
| BASORE | Basophils/100 leukocytes in Blood by Automated count | immune/inflammatory | 1 | LMO4 | protein_coding | rs4279831 | 87724930 | 2.23e-09 | 7 | Female only | Both sexes | Female only |
| BASORE | Basophils/100 leukocytes in Blood by Automated count | immune/inflammatory | 20 | C20orf112 | protein_coding | rs1535374 | 31162125 | 2.36e-09 | 29 | Both sexes | Both sexes | Both sexes |
| BASORE | Basophils/100 leukocytes in Blood by Automated count | immune/inflammatory | 18 | RP11-737G21.1 | pseudogene | rs8097343 | 23542220 | 5.32e-09 | 91 | Female only | Female only | Female only |
| BASORE | Basophils/100 leukocytes in Blood by Automated count | immune/inflammatory | 12 | PDCL3P7 | pseudogene | rs10748037 | 66664573 | 1.22e-08 | 41 | Female only | Both sexes | Female only |
| BASORE | Basophils/100 leukocytes in Blood by Automated count | immune/inflammatory | 18 | SS18 | protein_coding | rs2959356 | 23590203 | 2.91e-08 | 3 | Female only | Both sexes | Female only |
| BASORE | Basophils/100 leukocytes in Blood by Automated count | immune/inflammatory | 20 | TM9SF4 | protein_coding | rs2235904 | 30721712 | 2.99e-08 | 315 | Female only | Both sexes | Female only |
| BASORE | Basophils/100 leukocytes in Blood by Automated count | immune/inflammatory | 14 | SLC7A8 | protein_coding | rs17794251 | 23593442 | 3.15e-08 | 3 | Female only | Both sexes | Female only |
| BAbs | Basophils [#/volume] in Blood by Automated count | immune/inflammatory | 19 | CTD-2540B15.12 | lincRNA | rs67228209 | 33751033 | 1.51e-14 | 3 | Both sexes | Both sexes | Both sexes |

| lab | full_name | group | CHR | Gene_Symbol | biotype | lead_snp | lead_BP | lead_P | total_snps_in_loci | independent_loci_in_sex_P 1e6 | independent_loci_in_sex_P 1e2 | independent_loci_in_sex_P 5e8 |
|---|---|---|---|---|---|---|---|---|---|---|---|---|
| BAbs | Basophils [#/volume] in Blood by Automated count | immune/inflammatory | 19 | CTD-2540B15.12 | lincRNA | rs10414008 | 33758327 | 4.25e-12 | 5 | Both sexes | Both sexes | Both sexes |
| BAbs | Basophils [#/volume] in Blood by Automated count | immune/inflammatory | 3 | RAB7A | protein_coding | rs35904610 | 128452108 | 2.93e-09 | 115 | Female only | Both sexes | Female only |
| BAbs | Basophils [#/volume] in Blood by Automated count | immune/inflammatory | 19 | CTD-2540B15.12 | lincRNA | rs4432367 | 33752920 | 4.14e-09 | 3 | Female only | Both sexes | Female only |
| BAbs | Basophils [#/volume] in Blood by Automated count | immune/inflammatory | 19 | CTD-2540B15.12 | lincRNA | rs10420798 | 33764718 | 2.29e-08 | 3 | Both sexes | Both sexes | Male only |
| Baso | Basophils [#/volume] in Blood | immune/inflammatory | 19 | CTD-2540B15.12 | lincRNA | rs67228209 | 33751033 | 7.17e-16 | 3 | Both sexes | Both sexes | Both sexes |
| Baso | Basophils [#/volume] in Blood | immune/inflammatory | 19 | CTD-2540B15.12 | lincRNA | rs10412166 | 33758201 | 1.81e-12 | 5 | Both sexes | Both sexes | Both sexes |
| Baso | Basophils [#/volume] in Blood | immune/inflammatory | 19 | CTD-2540B15.12 | lincRNA | rs4805867 | 33762257 | 2.34e-12 | 3 | Both sexes | Both sexes | Female only |
| Baso | Basophils [#/volume] in Blood | immune/inflammatory | 19 | CTD-2540B15.12 | lincRNA | rs10420798 | 33764718 | 3.94e-11 | 3 | Both sexes | Both sexes | Female only |
| Baso | Basophils [#/volume] in Blood | immune/inflammatory | 3 | RAB7A | protein_coding | rs35904610 | 128452108 | 3.06e-10 | 115 | Both sexes | Both sexes | Both sexes |
| Baso | Basophils [#/volume] in Blood | immune/inflammatory | 3 | RAB7A | protein_coding | rs56373544 | 128523503 | 3.74e-10 | 25 | Female only | Female only | Female only |
| Baso | Basophils [#/volume] in Blood | immune/inflammatory | 3 | TMED10P2 | pseudogene | 3:128250135 | 128250135 | 1.42e-08 | 59 | Both sexes | Both sexes | Both sexes |
| Baso | Basophils [#/volume] in Blood | immune/inflammatory | 19 | CTD-2540B15.12 | lincRNA | rs4432367 | 33752920 | 4.24e-08 | 3 | Female only | Both sexes | Female only |

| lab | full_name | group | CHR | Gene_Symbol | biotype | lead_snp | lead_BP | lead_P | total_snps_in_loci | independent_loci_in_sex_P 1e6 | independent_loci_in_sex_P 1e2 | independent_loci_in_sex_P 5e8 |
|---|---|---|---|---|---|---|---|---|---|---|---|---|
| C4Quan | Complement C4 [Mass/volume] in Serum or Plasma | immune/inflammatory | 6 | STK19 | protein_coding | rs377370 | 31942514 | 1.97e-24 | 51 | Female only | Both sexes | Female only |
| C4Quan | Complement C4 [Mass/volume] in Serum or Plasma | immune/inflammatory | 6 | C6orf47 | protein_coding | rs805262 | 31628733 | 4.46e-19 | 35 | Female only | Both sexes | Female only |
| C4Quan | Complement C4 [Mass/volume] in Serum or Plasma | immune/inflammatory | 6 | XXbac-BPG154L12.4 | antisense | rs9267995 | 32221552 | 1.34e-17 | 23 | Both sexes | Both sexes | Female only |
| C4Quan | Complement C4 [Mass/volume] in Serum or Plasma | immune/inflammatory | 6 | XXbac-BPG254F23.7 | lincRNA | rs3104406 | 32682443 | 1.48e-17 | 83 | Both sexes | Both sexes | Female only |
| C4Quan | Complement C4 [Mass/volume] in Serum or Plasma | immune/inflammatory | 6 | BTNL2 | protein_coding | rs3129956 | 32368462 | 3.85e-17 | 139 | Both sexes | Both sexes | Both sexes |
| C4Quan | Complement C4 [Mass/volume] in Serum or Plasma | immune/inflammatory | 6 | HLA-S | pseudogene | rs2596560 | 31355318 | 5.31e-16 | 5 | Both sexes | Both sexes | Both sexes |
| C4Quan | Complement C4 [Mass/volume] in Serum or Plasma | immune/inflammatory | 6 | C6orf10 | protein_coding | rs3132954 | 32311459 | 6.69e-16 | 45 | Both sexes | Both sexes | Both sexes |
| C4Quan | Complement C4 [Mass/volume] in Serum or Plasma | immune/inflammatory | 6 | DHFRP2 | pseudogene | rs2523562 | 31330618 | 2.22e-15 | 19 | Both sexes | Both sexes | Female only |
| C4Quan | Complement C4 [Mass/volume] in Serum or Plasma | immune/inflammatory | 6 | CFB | protein_coding | rs537160 | 31916400 | 2.66e-15 | 39 | Both sexes | Both sexes | Both sexes |

| lab | full_name | group | CHR | Gene_Symbol | biotype | lead_snp | lead_BP | lead_P | total_snps_in_loci | independent_loci_in_sex_P 1e6 | independent_loci_in_sex_P 1e2 | independent_loci_in_sex_P 5e8 |
|---|---|---|---|---|---|---|---|---|---|---|---|---|
| C4Quan | Complement C4 [Mass/volume] in Serum or Plasma | immune/inflammatory | 6 | ABHD16A | protein_coding | rs1266074 | 31666551 | 4.47e-15 | 7 | Both sexes | Both sexes | Both sexes |
| C4Quan | Complement C4 [Mass/volume] in Serum or Plasma | immune/inflammatory | 6 | XXbac-BPG16N22.5 | pseudogene | rs2516491 | 31485101 | 4.67e-14 | 5 | Both sexes | Both sexes | Both sexes |
| C4Quan | Complement C4 [Mass/volume] in Serum or Plasma | immune/inflammatory | 6 | XXbac-BPG32J3.20 | protein_coding | rs805286 | 31679307 | 6.34e-14 | 33 | Female only | Both sexes | Female only |
| C4Quan | Complement C4 [Mass/volume] in Serum or Plasma | immune/inflammatory | 6 | SLC44A4 | protein_coding | rs521977 | 31836827 | 1.44e-13 | 39 | Both sexes | Both sexes | Both sexes |
| C4Quan | Complement C4 [Mass/volume] in Serum or Plasma | immune/inflammatory | 6 | XXbac-BPG248L24.10 | pseudogene | rs9461693 | 31291932 | 2.09e-13 | 95 | Female only | Both sexes | Female only |
| C4Quan | Complement C4 [Mass/volume] in Serum or Plasma | immune/inflammatory | 6 | VWA7 | protein_coding | rs6914412 | 31742242 | 4.61e-13 | 25 | Female only | Both sexes | Female only |
| C4Quan | Complement C4 [Mass/volume] in Serum or Plasma | immune/inflammatory | 6 | AL671883.1 | miRNA | rs2844551 | 31342781 | 5.87e-13 | 117 | Both sexes | Both sexes | Both sexes |
| C4Quan | Complement C4 [Mass/volume] in Serum or Plasma | immune/inflammatory | 6 | PSORS1C1 | protein_coding | rs3132561 | 31103931 | 2.24e-12 | 79 | Female only | Both sexes | Female only |
| C4Quan | Complement C4 [Mass/volume] in Serum or Plasma | immune/inflammatory | 6 | AL671883.1 | miRNA | rs2844552 | 31342741 | 3.58e-12 | 85 | Both sexes | Both sexes | Female only |
| C4Quan | Complement C4 [Mass/volume] in Serum or Plasma | immune/inflammatory | 6 | TRIM10 | protein_coding | rs1970 | 30125543 | 4.62e-12 | 333 | Both sexes | Both sexes | Both sexes |
| C4Quan | Complement C4 [Mass/volume] in Serum or Plasma | immune/inflammatory | 6 | XXbac-BPG248L24.12 | pseudogene | rs12662617 | 31328685 | 8.70e-12 | 7 | Female only | Both sexes | Female only |
| C4Quan | Complement C4 [Mass/volume] in Serum or Plasma | immune/inflammatory | 6 | BRD2 | protein_coding | rs78881806 | 32952168 | 1.01e-11 | 3 | Female only | Both sexes | Female only |
| C4Quan | Complement C4 [Mass/volume] in Serum or Plasma | immune/inflammatory | 6 | DHFRP2 | pseudogene | rs2844572 | 31335575 | 1.28e-11 | 33 | Both sexes | Both sexes | Female only |
| C4Quan | Complement C4 [Mass/volume] in Serum or Plasma | immune/inflammatory | 6 | HLA-S | pseudogene | rs2844535 | 31350303 | 1.46e-11 | 3 | Female only | Both sexes | Female only |
| C4Quan | Complement C4 [Mass/volume] in Serum or Plasma | immune/inflammatory | 6 | HLA-S | pseudogene | rs2507978 | 31351664 | 1.56e-11 | 11 | Female only | Both sexes | Female only |
| C4Quan | Complement C4 [Mass/volume] in Serum or Plasma | immune/inflammatory | 6 | SUMO2P1 | pseudogene | rs3131856 | 29607101 | 2.91e-11 | 137 | Both sexes | Both sexes | Both sexes |
| C4Quan | Complement C4 [Mass/volume] in Serum or Plasma | immune/inflammatory | 6 | FLOT1 | protein_coding | rs3095327 | 30699022 | 2.98e-11 | 221 | Both sexes | Both sexes | Both sexes |
| C4Quan | Complement C4 [Mass/volume] in Serum or Plasma | immune/inflammatory | 6 | TRIM10 | protein_coding | rs2523734 | 30129676 | 3.38e-11 | 23 | Female only | Both sexes | Female only |

| lab | full_name | group | CHR | Gene_Symbol | biotype | lead_snp | lead_BP | lead_P | total_snps_in_loci | independent_loci_in_sex_P1e6 | independent_loci_in_sex_P1e2 | independent_loci_in_sex_P5e8 |
|---|---|---|---|---|---|---|---|---|---|---|---|---|
| C4Quan | Complement C4 [Mass/volume] in Serum or Plasma | immune/inflammatory | 6 | XXbac-BPG154L12.4 | antisense | rs1894374 | 32214435 | 4.20e-11 | 57 | Female only | Both sexes | Female only |
| C4Quan | Complement C4 [Mass/volume] in Serum or Plasma | immune/inflammatory | 6 | DHFRP2 | pseudogene | rs2523558 | 31331037 | 9.94e-11 | 7 | Female only | Both sexes | Female only |
| C4Quan | Complement C4 [Mass/volume] in Serum or Plasma | immune/inflammatory | 6 | BTNL2 | protein_coding | rs3806156 | 32373698 | 1.82e-10 | 21 | Female only | Both sexes | Female only |
| C4Quan | Complement C4 [Mass/volume] in Serum or Plasma | immune/inflammatory | 6 | XXbac-BPG181B23.7 | lincRNA | rs2905713 | 31356702 | 2.11e-10 | 243 | Female only | Both sexes | Female only |
| C4Quan | Complement C4 [Mass/volume] in Serum or Plasma | immune/inflammatory | 6 | SKIV2L | protein_coding | rs429608 | 31930462 | 5.41e-10 | 3 | Female only | Both sexes | Female only |
| C4Quan | Complement C4 [Mass/volume] in Serum or Plasma | immune/inflammatory | 6 | HLA-Z | pseudogene | rs56098412 | 32879471 | 5.68e-10 | 35 | Both sexes | Both sexes | Both sexes |
| C4Quan | Complement C4 [Mass/volume] in Serum or Plasma | immune/inflammatory | 6 | HLA-DOB | protein_coding | rs9276710 | 32757102 | 6.08e-10 | 7 | Both sexes | Both sexes | Both sexes |
| C4Quan | Complement C4 [Mass/volume] in Serum or Plasma | immune/inflammatory | 6 | ZDHHC20P2 | pseudogene | rs2244027 | 31347566 | 7.59e-10 | 5 | Female only | Both sexes | Female only |
| C4Quan | Complement C4 [Mass/volume] in Serum or Plasma | immune/inflammatory | 6 | PSORS1C1 | protein_coding | rs6906565 | 31097814 | 1.78e-09 | 119 | Female only | Both sexes | Female only |
| C4Quan | Complement C4 [Mass/volume] in Serum or Plasma | immune/inflammatory | 6 | RNF5 | protein_coding | rs204996 | 32149883 | 1.96e-09 | 17 | Female only | Both sexes | Female only |
| C4Quan | Complement C4 [Mass/volume] in Serum or Plasma | immune/inflammatory | 6 | BRD2 | protein_coding | rs188245 | 32955976 | 2.05e-09 | 73 | Female only | Both sexes | Female only |
| C4Quan | Complement C4 [Mass/volume] in Serum or Plasma | immune/inflammatory | 6 | TMPOP1 | pseudogene | rs3132616 | 30416456 | 2.23e-09 | 19 | Both sexes | Both sexes | Both sexes |
| C4Quan | Complement C4 [Mass/volume] in Serum or Plasma | immune/inflammatory | 6 | XXbac-BPG248L24.12 | pseudogene | rs115860766 | 31328860 | 2.34e-09 | 29 | Female only | Both sexes | Female only |
| C4Quan | Complement C4 [Mass/volume] in Serum or Plasma | immune/inflammatory | 6 | MICA | protein_coding | rs73400361 | 31376266 | 2.61e-09 | 9 | Both sexes | Both sexes | Female only |
| C4Quan | Complement C4 [Mass/volume] in Serum or Plasma | immune/inflammatory | 6 | XXbac-BPG154L12.4 | antisense | rs447459 | 32208673 | 2.75e-09 | 5 | Female only | Both sexes | Female only |
| C4Quan | Complement C4 [Mass/volume] in Serum or Plasma | immune/inflammatory | 6 | C6orf100 | protein_coding | rs3131073 | 28920972 | 4.08e-09 | 323 | Both sexes | Both sexes | Female only |
| C4Quan | Complement C4 [Mass/volume] in Serum or Plasma | immune/inflammatory | 6 | C6orf48 | protein_coding | rs41284523 | 31810266 | 4.85e-09 | 19 | Female only | Both sexes | Female only |
| C4Quan | Complement C4 [Mass/volume] in Serum or Plasma | immune/inflammatory | 6 | OR5V1 | protein_coding | rs442694 | 29356687 | 6.21e-09 | 51 | Both sexes | Both sexes | Female only |

| lab | full_name | group | CHR | Gene_Symbol | biotype | lead_snp | lead_BP | lead_P | total_snps_in_loci | independent_loci_in_sex_P 1e6 | independent_loci_in_sex_P 1e2 | independent_loci_in_sex_P 5e8 |
|---|---|---|---|---|---|---|---|---|---|---|---|---|
| C4Quan | Complement C4 [Mass/volume] in Serum or Plasma | immune/inflammatory | 6 | HCG23 | antisense | rs17208671 | 32360849 | 6.54e-09 | 209 | Female only | Female only | Female only |
| C4Quan | Complement C4 [Mass/volume] in Serum or Plasma | immune/inflammatory | 6 | XXbac-BPG13B8.10 | lincRNA | rs1233478 | 29477821 | 7.26e-09 | 3 | Female only | Both sexes | Female only |
| C4Quan | Complement C4 [Mass/volume] in Serum or Plasma | immune/inflammatory | 6 | HLA-S | pseudogene | rs2523484 | 31353639 | 8.48e-09 | 145 | Both sexes | Both sexes | Female only |
| C4Quan | Complement C4 [Mass/volume] in Serum or Plasma | immune/inflammatory | 6 | HLA-DQA2 | protein_coding | rs9276427 | 32711857 | 8.79e-09 | 279 | Female only | Both sexes | Female only |
| C4Quan | Complement C4 [Mass/volume] in Serum or Plasma | immune/inflammatory | 6 | HCG9 | lincRNA | rs2246199 | 29945620 | 1.02e-08 | 315 | Female only | Both sexes | Female only |
| C4Quan | Complement C4 [Mass/volume] in Serum or Plasma | immune/inflammatory | 6 | ZSCAN31 | protein_coding | rs13198809 | 28323702 | 1.04e-08 | 187 | Both sexes | Both sexes | Female only |
| C4Quan | Complement C4 [Mass/volume] in Serum or Plasma | immune/inflammatory | 6 | C6orf10 | protein_coding | rs3117110 | 32340176 | 1.17e-08 | 5 | Female only | Both sexes | Female only |
| C4Quan | Complement C4 [Mass/volume] in Serum or Plasma | immune/inflammatory | 6 | HLA-S | pseudogene | rs9266772 | 31352113 | 2.43e-08 | 49 | Female only | Female only | Female only |
| C4Quan | Complement C4 [Mass/volume] in Serum or Plasma | immune/inflammatory | 6 | HCG17 | lincRNA | rs3130396 | 30223490 | 2.91e-08 | 145 | Female only | Female only | Female only |
| C4Quan | Complement C4 [Mass/volume] in Serum or Plasma | immune/inflammatory | 6 | RN7SKP186 | misc_RNA | rs2844657 | 30829522 | 3.71e-08 | 21 | Female only | Both sexes | Female only |
| C4Quan | Complement C4 [Mass/volume] in Serum or Plasma | immune/inflammatory | 6 | HCG17 | lincRNA | rs2516698 | 30232374 | 3.78e-08 | 63 | Female only | Female only | Female only |
| C4Quan | Complement C4 [Mass/volume] in Serum or Plasma | immune/inflammatory | 6 | HCG20 | lincRNA | rs4713369 | 30761129 | 4.19e-08 | 139 | Female only | Both sexes | Female only |
| CO2 | Carbon dioxide serum/plasma | blood | 2 | SLC9A4 | protein_coding | rs76229479 | 103123912 | 2.21e-12 | 155 | Female only | Both sexes | Female only |
| Ca | Calcium serum/plasma serum/plasma | metabolic | 2 | AC079610.1 | sense_overlapping | rs13035592 | 213733958 | 1.27e-08 | 35 | Male only | Male only | Male only |
| Ca | Calcium serum/plasma serum/plasma | metabolic | 9 | RP11-205K6.1 | lincRNA | rs4837096 | 129276052 | 4.67e-08 | 19 | Female only | Female only | Female only |
| lab | full_name | group | CHR | Gene_Symbol | biotype | lead_snp | lead_BP | lead_P | total_snps_in_loci | independent_loci_in_sex_P 1e6 | independent_loci_in_sex_P 1e2 | independent_loci_in_sex_P 5e8 |
| Chol | Cholesterol [Mass/volume] in Serum or Plasma | metabolic | 19 | PVRL2 | protein_coding | 19:45384405 | 45384405 | 1.28e-17 | 3 | Female only | Both sexes | Female only |
| Chol | Cholesterol [Mass/volume] in Serum or Plasma | metabolic | 2 | AC067959.1 | lincRNA | rs13420469 | 21449034 | 2.07e-09 | 43 | Female only | Both sexes | Female only |
| Chol | Cholesterol [Mass/volume] in Serum or Plasma | metabolic | 6 | BTNL2 | protein_coding | rs72847932 | 32362085 | 1.87e-08 | 125 | Female only | Both sexes | Female only |
| HEART.RATE | Heart rate | vitals | 13 | MTUS2 | protein_coding | rs4768988 | 29546818 | 4.69e-09 | 9 | Female only | Female only | Female only |

| lab | full_name | group | CHR | Gene_Symbol | biotype | lead_snp | lead_BP | lead_P | total_snps_in_loci | independent_loci_in_sex_P 1e6 | independent_loci_in_sex_P 1e2 | independent_loci_in_sex_P 5e8 |
|---|---|---|---|---|---|---|---|---|---|---|---|---|
| Height | Height | vitals | 23 | RP1-164L12.1 | pseudogene | X:78641728:G:A | 78641728 | 1.71e-09 | 5 | Male only | Both sexes | Male only |
| Height | Height | vitals | 23 | KIF4CP | pseudogene | X:78594876:T:G | 78594876 | 3.86e-09 | 133 | Male only | Both sexes | Male only |
| Height | Height | vitals | 5 | CTB-33O18.1 | lincRNA | rs7717406 | 172999405 | 3.39e-08 | 93 | Female only | Both sexes | Female only |
| Height | Height | vitals | 9 | C9orf3 | protein_coding | rs11787821 | 97660961 | 3.78e-08 | 117 | Female only | Both sexes | Female only |
| Hgb | Hemoglobin | blood | 6 | HCG4P7 | pseudogene | rs1611657 | 29842444 | 9.50e-09 | 155 | Female only | Both sexes | Female only |
| HgbA1C | Hemoglobin A1c (Glycated) Hemoglobin A1c (Glycated) | metabolic | 6 | NOTCH4 | protein_coding | rs377763 | 32199144 | 1.86e-10 | 23 | Both sexes | Both sexes | Female only |
| HgbA1C | Hemoglobin A1c (Glycated) Hemoglobin A1c (Glycated) | metabolic | 6 | NOTCH4 | protein_coding | rs3130293 | 32177263 | 8.33e-09 | 19 | Both sexes | Both sexes | Female only |
| HgbA1C | Hemoglobin A1c (Glycated) Hemoglobin A1c (Glycated) | metabolic | 6 | BAG6 | protein_coding | rs3130628 | 31609272 | 1.20e-08 | 127 | Male only | Both sexes | Male only |
| HgbA1C | Hemoglobin A1c (Glycated) Hemoglobin A1c (Glycated) | metabolic | 6 | XXbac-BPG16N22.5 | pseudogene | rs2516491 | 31485101 | 3.28e-08 | 11 | Male only | Both sexes | Male only |
| IGRE | Immature granulocytes/100 leukocytes in Blood | immune/inflammatory | 9 | LRRC8A | protein_coding | rs34305034 | 131654547 | 2.17e-45 | 3 | Both sexes | Both sexes | Both sexes |
| IGRE | Immature granulocytes/100 leukocytes in Blood | immune/inflammatory | 6 | RP1-111C20.3 | lincRNA | rs2451256 | 159507338 | 1.36e-33 | 15 | Both sexes | Both sexes | Both sexes |
| IGRE | Immature granulocytes/100 leukocytes in Blood | immune/inflammatory | 9 | ENDOG | protein_coding | rs4837316 | 131577450 | 1.43e-19 | 55 | Both sexes | Both sexes | Both sexes |
| IGRE | Immature granulocytes/100 leukocytes in Blood | immune/inflammatory | 6 | RP1-111C20.3 | lincRNA | rs2451258 | 159506600 | 4.72e-16 | 21 | Both sexes | Both sexes | Both sexes |
| IGRE | Immature granulocytes/100 leukocytes in Blood | immune/inflammatory | 10 | PRTFDC1 | protein_coding | rs10828722 | 25199951 | 3.15e-15 | 15 | Male only | Both sexes | Male only |
| IGRE | Immature granulocytes/100 leukocytes in Blood | immune/inflammatory | 6 | RP11-13P5.1 | lincRNA | rs62431905 | 159510162 | 3.74e-13 | 7 | Both sexes | Both sexes | Both sexes |
| IGRE | Immature granulocytes/100 leukocytes in Blood | immune/inflammatory | 22 | DEPDC5 | protein_coding | rs5749339 | 32315735 | 2.30e-11 | 35 | Both sexes | Both sexes | Both sexes |
| IGRE | Immature granulocytes/100 leukocytes in Blood | immune/inflammatory | 9 | ZDHHC12 | protein_coding | rs10988105 | 131484526 | 4.36e-10 | 3 | Both sexes | Both sexes | Both sexes |
| IGRE | Immature granulocytes/100 leukocytes in Blood | immune/inflammatory | 7 | CCM2 | protein_coding | rs36084700 | 45034408 | 6.16e-10 | 85 | Both sexes | Both sexes | Male only |
| IGRE | Immature granulocytes/100 leukocytes in Blood | immune/inflammatory | 10 | PRTFDC1 | protein_coding | rs7905553 | 25200591 | 1.09e-08 | 7 | Male only | Both sexes | Male only |
| IGRE | Immature granulocytes/100 leukocytes in Blood | immune/inflammatory | 7 | TBRG4 | protein_coding | rs6955660 | 45149410 | 1.40e-08 | 11 | Male only | Both sexes | Male only |
| IGRE | Immature granulocytes/100 leukocytes in Blood | immune/inflammatory | 6 | RP1-111C20.4 | antisense | rs394581 | 159482521 | 1.50e-08 | 9 | Male only | Both sexes | Male only |
| IctIdx | Icteric index of Serum or Plasma | metabolic | 2 | UGT1A8 | protein_coding | rs6742078 | 234672639 | 5.51e-13 | 219 | Male only | Both sexes | Male only |
| IgM.Q | IgM Quantitative blood | immune/inflammatory | 2 | PDE11A | protein_coding | rs113811979 | 178695260 | 5.13e-09 | 137 | Female only | Female only | Female only |
| Iron | Iron [Mass/volume] in Serum or Plasma | blood | 6 | HIST1H2AA | protein_coding | rs116009877 | 25715657 | 6.04e-13 | 33 | Both sexes | Both sexes | Both sexes |
| Iron | Iron [Mass/volume] in Serum or Plasma | blood | 6 | HIST1H1T | protein_coding | rs198846 | 26107463 | 1.82e-11 | 33 | Both sexes | Both sexes | Female only |
| Iron | Iron [Mass/volume] in Serum or Plasma | blood | 6 | HIST1H3C | protein_coding | 6:26046744 | 26046744 | 2.03e-11 | 245 | Both sexes | Both sexes | Both sexes |

| lab | full_name | group | CHR | Gene_Symbol | biotype | lead_snp | lead_BP | lead_P | total_snps_in_loci | independent_loci_in_sex_P 1e6 | independent_loci_in_sex_P 1e2 | independent_loci_in_sex_P 5e8 |
|---|---|---|---|---|---|---|---|---|---|---|---|---|
| K | Potassium blood | metabolic | 6 | HLA-DQB1 | protein_coding | rs2854275 | 32628428 | 1.02e-08 | 153 | Male only | Both sexes | Male only |
| LYMPAB | Lymphocytes [#/volume] in Blood by Automated count | immune/inflammatory | 6 | TRIM26 | protein_coding | rs2517611 | 30169327 | 1.35e-10 | 147 | Both sexes | Both sexes | Both sexes |
| LYMPAB | Lymphocytes [#/volume] in Blood by Automated count | immune/inflammatory | 6 | UBQLN1P1 | pseudogene | rs3132634 | 30343272 | 1.54e-10 | 175 | Both sexes | Both sexes | Both sexes |
| LYMPAB | Lymphocytes [#/volume] in Blood by Automated count | immune/inflammatory | 6 | SKIV2L | protein_coding | rs437179 | 31929014 | 3.44e-10 | 41 | Both sexes | Both sexes | Male only |
| LYMPAB | Lymphocytes [#/volume] in Blood by Automated count | immune/inflammatory | 6 | HIST1H1B | protein_coding | rs17695758 | 27837183 | 8.94e-10 | 195 | Female only | Both sexes | Female only |
| LYMPAB | Lymphocytes [#/volume] in Blood by Automated count | immune/inflammatory | 6 | LINC00243 | processed_transcript | rs1264347 | 30798697 | 1.15e-09 | 155 | Both sexes | Both sexes | Female only |
| LYMPAB | Lymphocytes [#/volume] in Blood by Automated count | immune/inflammatory | 6 | ZKSCAN4 | protein_coding | rs17720293 | 28214698 | 1.42e-09 | 191 | Female only | Both sexes | Female only |
| LYMPAB | Lymphocytes [#/volume] in Blood by Automated count | immune/inflammatory | 6 | XXbac-BPGBPG34I8.1 | pseudogene | rs34573979 | 27480526 | 1.49e-09 | 67 | Female only | Both sexes | Female only |
| LYMPAB | Lymphocytes [#/volume] in Blood by Automated count | immune/inflammatory | 6 | RPSAP2 | pseudogene | rs9468413 | 28689672 | 1.71e-09 | 259 | Both sexes | Both sexes | Female only |
| LYMPAB | Lymphocytes [#/volume] in Blood by Automated count | immune/inflammatory | 6 | CSNK2B | protein_coding | rs9267531 | 31636742 | 3.98e-09 | 129 | Both sexes | Both sexes | Both sexes |
| LYMPAB | Lymphocytes [#/volume] in Blood by Automated count | immune/inflammatory | 6 | COX11P1 | pseudogene | rs61289879 | 28419162 | 5.72e-09 | 263 | Female only | Both sexes | Female only |
| LYMPAB | Lymphocytes [#/volume] in Blood by Automated count | immune/inflammatory | 6 | RPSAP2 | pseudogene | rs1233585 | 28716345 | 1.01e-08 | 29 | Male only | Both sexes | Male only |
| LYMPAB | Lymphocytes [#/volume] in Blood by Automated count | immune/inflammatory | 6 | XXbac-BCX196D17.5 | lincRNA | rs3130773 | 29095908 | 2.45e-08 | 39 | Female only | Both sexes | Female only |
| LYMPAB | Lymphocytes [#/volume] in Blood by Automated count | immune/inflammatory | 6 | TNXB | protein_coding | rs1150752 | 32064726 | 3.20e-08 | 139 | Both sexes | Both sexes | Both sexes |
| LYMPM | Lymphocytes/100 leukocytes in Blood by Manual count | immune/inflammatory | 23 | RP11-585K14.2 | pseudogene | X:5094786:G:A | 5094786 | 1.00e-08 | 141 | Female only | Female only | Female only |
| LYMPRE | Lymphocytes/100 leukocytes in Blood by Automated count | immune/inflammatory | 1 | RP11-334L9.1 | pseudogene | rs3011200 | 39079403 | 5.31e-09 | 15 | Female only | Female only | Female only |
| LipIdx | Lipemic index of Serum or Plasma | metabolic | 19 | APOC1 | protein_coding | rs5117 | 45418790 | 4.80e-09 | 11 | Male only | Both sexes | Male only |
| LucAbs | Leukocytes [#/volume] in Blood by Automated count | blood | 17 | BZRAP1-AS1 | antisense | rs75394768 | 56413635 | 4.42e-09 | 3 | Female only | Both sexes | Female only |
| MCH | MCH [Entitic mass] by Automated count | blood | 6 | LRRC16A | protein_coding | rs149359690 | 25526319 | 1.25e-38 | 45 | Both sexes | Both sexes | Both sexes |
| MCH | MCH [Entitic mass] by Automated count | blood | 6 | MIR3143 | miRNA | rs71559014 | 27122444 | 8.39e-31 | 3 | Both sexes | Both sexes | Both sexes |
| MCH | MCH [Entitic mass] by Automated count | blood | 6 | U91328.22 | lincRNA | rs9356996 | 26008487 | 1.54e-27 | 63 | Both sexes | Both sexes | Both sexes |
| MCH | MCH [Entitic mass] by Automated count | blood | 6 | HIST1H2BO | protein_coding | 6:27861670 | 27861670 | 1.71e-25 | 9 | Both sexes | Both sexes | Both sexes |
| MCH | MCH [Entitic mass] by Automated count | blood | 6 | LRRC16A | protein_coding | rs111722075 | 25374046 | 3.02e-25 | 253 | Both sexes | Both sexes | Both sexes |
| MCH | MCH [Entitic mass] by Automated count | blood | 6 | ZSCAN31 | protein_coding | rs35814746 | 28299088 | 3.92e-24 | 41 | Both sexes | Both sexes | Both sexes |
| MCH | MCH [Entitic mass] by Automated count | blood | 6 | HIST1H2BC | protein_coding | rs198839 | 26112620 | 9.65e-24 | 13 | Both sexes | Both sexes | Both sexes |

| lab | full_name | group | CHR | Gene_Symbol | biotype | lead_snp | lead_BP | lead_P | total_snps_in_loci | independent_loci_in_sex_P1e6 | independent_loci_in_sex_P1e2 | independent_loci_in_sex_P5e8 |
|---|---|---|---|---|---|---|---|---|---|---|---|---|
| MCH | MCH [Entitic mass] by Automated count | blood | 6 | HIST1H2APS2 | pseudogene | rs72830103 | 25897618 | 6.50e-23 | 51 | Both sexes | Both sexes | Both sexes |
| MCH | MCH [Entitic mass] by Automated count | blood | 6 | LRRC16A | protein_coding | rs181058325 | 25314597 | 2.82e-21 | 11 | Both sexes | Both sexes | Both sexes |
| MCH | MCH [Entitic mass] by Automated count | blood | 6 | LRRC16A | protein_coding | rs13203202 | 25582771 | 4.82e-21 | 9 | Both sexes | Both sexes | Both sexes |
| MCH | MCH [Entitic mass] by Automated count | blood | 6 | XXbac-BPG308K3.5 | lincRNA | rs3118362 | 28785085 | 3.41e-19 | 5 | Both sexes | Both sexes | Both sexes |
| MCH | MCH [Entitic mass] by Automated count | blood | 6 | LRRC16A | protein_coding | rs72831267 | 25436955 | 1.82e-17 | 7 | Both sexes | Both sexes | Both sexes |
| lab | full_name | group | CHR | Gene_Symbol | biotype | lead_snp | lead_BP | lead_P | total_snps_in_loci | independent_loci_in_sex_P1e6 | independent_loci_in_sex_P1e2 | independent_loci_in_sex_P5e8 |
| MCH | MCH [Entitic mass] by Automated count | blood | 6 | LRRC16A | protein_coding | rs9358860 | 25572245 | 8.14e-17 | 53 | Both sexes | Both sexes | Both sexes |
| MCH | MCH [Entitic mass] by Automated count | blood | 6 | TRIM38 | protein_coding | rs199739 | 25960509 | 1.97e-16 | 47 | Both sexes | Both sexes | Female only |
| MCH | MCH [Entitic mass] by Automated count | blood | 6 | LRRC16A | protein_coding | rs112268831 | 25348760 | 5.36e-16 | 5 | Both sexes | Both sexes | Female only |
| MCH | MCH [Entitic mass] by Automated count | blood | 6 | HLA-W | pseudogene | rs9260707 | 29929536 | 1.83e-15 | 165 | Both sexes | Both sexes | Both sexes |
| MCH | MCH [Entitic mass] by Automated count | blood | 6 | HIST1H3PS1 | pseudogene | rs6932727 | 26327087 | 2.51e-15 | 95 | Both sexes | Both sexes | Both sexes |
| MCH | MCH [Entitic mass] by Automated count | blood | 4 | AC006552.1 | lincRNA | rs80113191 | 55396420 | 5.56e-15 | 101 | Both sexes | Both sexes | Both sexes |
| MCH | MCH [Entitic mass] by Automated count | blood | 6 | HIST1H2BC | protein_coding | rs707896 | 26116424 | 5.60e-14 | 51 | Both sexes | Both sexes | Both sexes |
| MCH | MCH [Entitic mass] by Automated count | blood | 6 | LRRC16A | protein_coding | rs9379764 | 25414023 | 3.92e-13 | 3 | Both sexes | Both sexes | Both sexes |
| MCH | MCH [Entitic mass] by Automated count | blood | 6 | MOG | protein_coding | rs3130253 | 29634012 | 4.03e-13 | 21 | Both sexes | Both sexes | Both sexes |
| MCH | MCH [Entitic mass] by Automated count | blood | 6 | RP3-352A20.1 | pseudogene | rs73547141 | 135215416 | 3.86e-12 | 57 | Both sexes | Both sexes | Both sexes |
| MCH | MCH [Entitic mass] by Automated count | blood | 6 | BTN2A3P | pseudogene | rs4320356 | 26423560 | 4.11e-12 | 73 | Female only | Both sexes | Female only |
| MCH | MCH [Entitic mass] by Automated count | blood | 6 | HMGN4 | protein_coding | rs12526680 | 26550954 | 2.06e-11 | 279 | Female only | Both sexes | Female only |
| MCH | MCH [Entitic mass] by Automated count | blood | 6 | ATAT1 | protein_coding | rs3094106 | 30599176 | 4.97e-11 | 57 | Both sexes | Both sexes | Female only |
| MCH | MCH [Entitic mass] by Automated count | blood | 7 | GIGYF1 | protein_coding | rs221784 | 100292830 | 5.59e-11 | 49 | Female only | Both sexes | Female only |
| MCH | MCH [Entitic mass] by Automated count | blood | 6 | ZSCAN23 | protein_coding | rs13190937 | 28411244 | 9.39e-11 | 71 | Female only | Both sexes | Female only |
| MCH | MCH [Entitic mass] by Automated count | blood | 6 | RP3-352A20.1 | pseudogene | rs2327576 | 135177685 | 1.12e-10 | 47 | Both sexes | Both sexes | Male only |
| MCH | MCH [Entitic mass] by Automated count | blood | 6 | RP3-352A20.1 | pseudogene | rs4896115 | 135168723 | 1.17e-10 | 41 | Both sexes | Both sexes | Both sexes |
| MCH | MCH [Entitic mass] by Automated count | blood | 6 | HIST1H3F | protein_coding | rs78545713 | 26248840 | 3.84e-10 | 223 | Both sexes | Both sexes | Male only |
| MCH | MCH [Entitic mass] by Automated count | blood | 6 | OR2J3 | protein_coding | rs3129106 | 29081161 | 4.73e-10 | 3 | Female only | Both sexes | Female only |
| MCH | MCH [Entitic mass] by Automated count | blood | 4 | RP11-545H22.1 | lincRNA | rs1553663 | 55280100 | 1.20e-09 | 31 | Female only | Both sexes | Female only |

| lab | full_name | group | CHR | Gene_Symbol | biotype | lead_snp | lead_BP | lead_P | total_snps_in_loci | independent_loci_in_sex_P 1e6 | independent_loci_in_sex_P 1e2 | independent_loci_in_sex_P 5e8 |
|---|---|---|---|---|---|---|---|---|---|---|---|---|
| MCH | MCH [Entitic mass] by Automated count | blood | 22 | LL22NC01-81G9.3 | antisense | rs130437 | 37369465 | 1.25e-09 | 53 | Female only | Both sexes | Female only |
| MCH | MCH [Entitic mass] by Automated count | blood | 6 | LRRC16A | protein_coding | rs9366617 | 25327270 | 2.29e-09 | 37 | Female only | Both sexes | Female only |
| MCH | MCH [Entitic mass] by Automated count | blood | 6 | DDR1 | protein_coding | rs3094719 | 30844972 | 4.17e-09 | 37 | Both sexes | Both sexes | Female only |
| MCH | MCH [Entitic mass] by Automated count | blood | 6 | MICD | pseudogene | rs1061539 | 29937555 | 4.82e-09 | 3 | Female only | Both sexes | Female only |
| MCH | MCH [Entitic mass] by Automated count | blood | 4 | RP11-545H22.1 | lincRNA | rs6846679 | 55305505 | 4.98e-09 | 97 | Female only | Both sexes | Female only |
| MCH | MCH [Entitic mass] by Automated count | blood | 6 | LRRC16A | protein_coding | rs12200019 | 25430896 | 6.29e-09 | 9 | Female only | Both sexes | Female only |
| MCH | MCH [Entitic mass] by Automated count | blood | 6 | TRIM40 | protein_coding | rs3132676 | 30116078 | 7.39e-09 | 15 | Female only | Both sexes | Female only |
| MCH | MCH [Entitic mass] by Automated count | blood | 6 | TRIM39 | protein_coding | rs1110464 | 30296071 | 7.61e-09 | 7 | Male only | Both sexes | Male only |
| MCH | MCH [Entitic mass] by Automated count | blood | 6 | BTN3A1 | protein_coding | rs3799378 | 26404374 | 9.02e-09 | 21 | Female only | Both sexes | Female only |
| MCH | MCH [Entitic mass] by Automated count | blood | 4 | RP11-545H22.1 | lincRNA | rs13151127 | 55358779 | 1.08e-08 | 33 | Female only | Both sexes | Female only |
| MCH | MCH [Entitic mass] by Automated count | blood | 3 | TNK2 | protein_coding | rs9871110 | 195625326 | 1.11e-08 | 7 | Male only | Male only | Male only |
| MCH | MCH [Entitic mass] by Automated count | blood | 6 | VN1R10P | pseudogene | rs9379980 | 27306518 | 1.18e-08 | 77 | Female only | Both sexes | Female only |
| MCH | MCH [Entitic mass] by Automated count | blood | 6 | RP11-457M11.5 | lincRNA | rs4713037 | 26758030 | 1.29e-08 | 77 | Female only | Female only | Female only |
| MCH | MCH [Entitic mass] by Automated count | blood | 6 | HIST1H4F | protein_coding | rs9358913 | 26239404 | 1.73e-08 | 13 | Male only | Both sexes | Male only |
| MCH | MCH [Entitic mass] by Automated count | blood | 7 | PILRA | protein_coding | rs62482199 | 99968371 | 1.80e-08 | 9 | Female only | Both sexes | Female only |
| MCH | MCH [Entitic mass] by Automated count | blood | 4 | RP11-545H22.1 | lincRNA | rs13122088 | 55372680 | 2.17e-08 | 3 | Both sexes | Both sexes | Both sexes |
| MCH | MCH [Entitic mass] by Automated count | blood | 9 | RP11-307I14.4 | pseudogene | rs10974772 | 4767677 | 2.22e-08 | 3 | Female only | Both sexes | Female only |
| MCH | MCH [Entitic mass] by Automated count | blood | 22 | TEX33 | protein_coding | rs5756445 | 37379539 | 2.41e-08 | 85 | Female only | Both sexes | Female only |
| MCH | MCH [Entitic mass] by Automated count | blood | 6 | UBQLN1P1 | pseudogene | rs2844745 | 30343703 | 2.50e-08 | 233 | Both sexes | Both sexes | Female only |
| MCH | MCH [Entitic mass] by Automated count | blood | 6 | GUSBP2 | pseudogene | rs56098842 | 26833759 | 3.38e-08 | 109 | Female only | Both sexes | Female only |
| MCHC | MCHC [Mass/volume] by Automated count | blood | 6 | MIR3143 | miRNA | rs71559014 | 27122444 | 3.79e-22 | 3 | Both sexes | Both sexes | Both sexes |
| MCHC | MCHC [Mass/volume] by Automated count | blood | 6 | ZSCAN31 | protein_coding | rs35814746 | 28299088 | 6.85e-22 | 41 | Both sexes | Both sexes | Both sexes |
| MCHC | MCHC [Mass/volume] by Automated count | blood | 6 | HIST1H2BO | protein_coding | 6:27861670 | 27861670 | 8.34e-22 | 9 | Both sexes | Both sexes | Both sexes |
| MCHC | MCHC [Mass/volume] by Automated count | blood | 6 | U91328.22 | lincRNA | rs9356996 | 26008487 | 2.04e-21 | 63 | Both sexes | Both sexes | Both sexes |
| MCHC | MCHC [Mass/volume] by Automated count | blood | 6 | LRRC16A | protein_coding | rs74662487 | 25453057 | 2.99e-20 | 17 | Both sexes | Both sexes | Both sexes |
| MCHC | MCHC [Mass/volume] by Automated count | blood | 6 | OR2AD1P | pseudogene | rs3129796 | 28988861 | 2.50e-16 | 7 | Both sexes | Both sexes | Both sexes |

| lab | full_name | group | CHR | Gene_Symbol | biotype | lead_snp | lead_BP | lead_P | total_snps_in_loci | independent_loci_in_sex_P1e6 | independent_loci_in_sex_P1e2 | independent_loci_in_sex_P5e8 |
|---|---|---|---|---|---|---|---|---|---|---|---|---|
| MCHC | MCHC [Mass/volume] by Automated count | blood | 6 | HLA-W | pseudogene | rs9260707 | 29929536 | 2.48e-15 | 169 | Both sexes | Both sexes | Both sexes |
| MCHC | MCHC [Mass/volume] by Automated count | blood | 6 | GPX6 | protein_coding | rs10428759 | 28477682 | 1.31e-14 | 9 | Both sexes | Both sexes | Female only |
| MCHC | MCHC [Mass/volume] by Automated count | blood | 1 | CEP104 | protein_coding | rs12134919 | 3744641 | 4.54e-14 | 37 | Both sexes | Both sexes | Both sexes |
| MCHC | MCHC [Mass/volume] by Automated count | blood | 6 | TRIM26 | protein_coding | rs3094618 | 30181621 | 1.81e-13 | 63 | Both sexes | Both sexes | Both sexes |
| MCHC | MCHC [Mass/volume] by Automated count | blood | 6 | HIST1H3G | protein_coding | rs7773004 | 26267755 | 2.06e-12 | 89 | Male only | Both sexes | Male only |
| MCHC | MCHC [Mass/volume] by Automated count | blood | 6 | MOG | protein_coding | rs3130253 | 29634012 | 2.80e-12 | 21 | Both sexes | Both sexes | Female only |
| MCHC | MCHC [Mass/volume] by Automated count | blood | 6 | ZSCAN23 | protein_coding | rs13190937 | 28411244 | 3.13e-12 | 71 | Female only | Female only | Female only |
| MCHC | MCHC [Mass/volume] by Automated count | blood | 6 | LRRC16A | protein_coding | rs2328879 | 25583103 | 1.08e-11 | 9 | Both sexes | Both sexes | Both sexes |
| MCHC | MCHC [Mass/volume] by Automated count | blood | 6 | SLC17A1 | protein_coding | rs12191655 | 25803095 | 1.98e-11 | 113 | Female only | Both sexes | Female only |
| MCHC | MCHC [Mass/volume] by Automated count | blood | 6 | HIST1H2BC | protein_coding | rs198839 | 26112620 | 1.99e-11 | 13 | Both sexes | Both sexes | Female only |
| MCHC | MCHC [Mass/volume] by Automated count | blood | 6 | LRRC16A | protein_coding | rs9379764 | 25414023 | 2.03e-11 | 3 | Female only | Both sexes | Female only |
| MCHC | MCHC [Mass/volume] by Automated count | blood | 6 | HCG22 | lincRNA | rs2517503 | 31032407 | 3.69e-11 | 33 | Female only | Both sexes | Female only |
| MCHC | MCHC [Mass/volume] by Automated count | blood | 6 | TRIM40 | protein_coding | rs2107202 | 30105743 | 1.37e-10 | 15 | Both sexes | Both sexes | Both sexes |
| MCHC | MCHC [Mass/volume] by Automated count | blood | 6 | HCG4P7 | pseudogene | rs1611657 | 29842444 | 1.80e-10 | 155 | Female only | Both sexes | Female only |
| lab | full_name | group | CHR | Gene_Symbol | biotype | lead_snp | lead_BP | lead_P | total_snps_in_loci | independent_loci_in_sex_P1e6 | independent_loci_in_sex_P1e2 | independent_loci_in_sex_P5e8 |
| MCHC | MCHC [Mass/volume] by Automated count | blood | 6 | LRRC16A | protein_coding | rs72831267 | 25436955 | 7.72e-10 | 7 | Female only | Both sexes | Female only |
| MCHC | MCHC [Mass/volume] by Automated count | blood | 6 | XXbac-BPG27H4.8 | lincRNA | rs3095345 | 30822413 | 7.83e-10 | 49 | Female only | Both sexes | Female only |
| MCHC | MCHC [Mass/volume] by Automated count | blood | 6 | MICC | pseudogene | rs2524205 | 30359606 | 8.01e-10 | 233 | Both sexes | Both sexes | Male only |
| MCHC | MCHC [Mass/volume] by Automated count | blood | 6 | MICD | pseudogene | rs9260740 | 29933236 | 1.78e-09 | 15 | Both sexes | Both sexes | Both sexes |
| MCHC | MCHC [Mass/volume] by Automated count | blood | 6 | XXbac-BPG249D20.9 | antisense | rs9262081 | 30484966 | 2.84e-09 | 17 | Female only | Both sexes | Female only |
| MCHC | MCHC [Mass/volume] by Automated count | blood | 6 | HLA-G | protein_coding | rs452850 | 29799481 | 3.20e-09 | 265 | Both sexes | Both sexes | Female only |
| MCHC | MCHC [Mass/volume] by Automated count | blood | 6 | LRRC16A | protein_coding | rs112268831 | 25348760 | 3.25e-09 | 5 | Female only | Both sexes | Female only |
| MCHC | MCHC [Mass/volume] by Automated count | blood | 6 | MICF | pseudogene | rs2523818 | 29833840 | 3.72e-09 | 405 | Female only | Both sexes | Female only |
| MCHC | MCHC [Mass/volume] by Automated count | blood | 6 | C2 | protein_coding | rs3130683 | 31888367 | 4.34e-09 | 63 | Male only | Both sexes | Male only |
| MCHC | MCHC [Mass/volume] by Automated count | blood | 6 | AIF1 | protein_coding | rs2857600 | 31582287 | 7.12e-09 | 71 | Both sexes | Both sexes | Both sexes |
| MCHC | MCHC [Mass/volume] by Automated count | blood | 6 | LRRC16A | protein_coding | rs9358860 | 25572245 | 9.67e-09 | 53 | Female only | Both sexes | Female only |

| lab | full_name | group | CHR | Gene_Symbol | biotype | lead_snp | lead_BP | lead_P | total_snps_in_loci | independent_loci_in_sex_P 1e6 | independent_loci_in_sex_P 1e2 | independent_loci_in_sex_P 5e8 |
|---|---|---|---|---|---|---|---|---|---|---|---|---|
| MCHC | MCHC [Mass/volume] by Automated count | blood | 6 | GPX6 | protein_coding | rs2213806 | 28457818 | 1.09e-08 | 75 | Female only | Both sexes | Female only |
| MCHC | MCHC [Mass/volume] by Automated count | blood | 6 | TRIM15 | protein_coding | rs2844787 | 30140913 | 1.48e-08 | 3 | Both sexes | Both sexes | Female only |
| MCHC | MCHC [Mass/volume] by Automated count | blood | 6 | GPX6 | protein_coding | rs2531820 | 28456719 | 1.57e-08 | 33 | Female only | Both sexes | Female only |
| MCHC | MCHC [Mass/volume] by Automated count | blood | 6 | XXbac-BPG154L12.4 | antisense | rs424232 | 32208324 | 1.84e-08 | 19 | Male only | Both sexes | Male only |
| MCHC | MCHC [Mass/volume] by Automated count | blood | 15 | WDR61 | protein_coding | rs7170960 | 78583818 | 1.88e-08 | 23 | Female only | Both sexes | Female only |
| MCHC | MCHC [Mass/volume] by Automated count | blood | 19 | ZNF283 | protein_coding | rs60085895 | 44347424 | 3.29e-08 | 203 | Female only | Female only | Female only |
| MCHC | MCHC [Mass/volume] by Automated count | blood | 6 | XXbac-BPG248L24.12 | pseudogene | 6:31328756 | 31328756 | 4.23e-08 | 9 | Both sexes | Both sexes | Male only |
| MEAN_QRS | QRS axis | cardiovascular | 10 | RP11-90B22.1 | lincRNA | rs10763928 | 33632954 | 6.59e-09 | 89 | Male only | Both sexes | Male only |
| MEAN_QRS | QRS axis | cardiovascular | 10 | RPL7P37 | pseudogene | rs2804817 | 36061337 | 7.06e-09 | 15 | Male only | Male only | Male only |
| MEAN_QRS | QRS axis | cardiovascular | 4 | LIMCH1 | protein_coding | rs1377345 | 41322070 | 2.30e-08 | 131 | Female only | Both sexes | Female only |
| MONOAB | Monocytes [#/volume] in Blood by Automated count | immune/inflammatory | 6 | RPSAP2 | pseudogene | rs7775835 | 28678357 | 2.04e-09 | 251 | Male only | Both sexes | Male only |
| MONOAB | Monocytes [#/volume] in Blood by Automated count | immune/inflammatory | 6 | ATF6B | protein_coding | rs204893 | 32094593 | 3.57e-09 | 17 | Male only | Male only | Male only |
| MONOAB | Monocytes [#/volume] in Blood by Automated count | immune/inflammatory | 6 | ZNF192P2 | pseudogene | rs735765 | 28170297 | 3.90e-09 | 349 | Male only | Male only | Male only |
| MONOAB | Monocytes [#/volume] in Blood by Automated count | immune/inflammatory | 6 | OR5V1 | protein_coding | rs9257805 | 29346329 | 6.55e-09 | 109 | Male only | Both sexes | Male only |
| MONOAB | Monocytes [#/volume] in Blood by Automated count | immune/inflammatory | 6 | RP1-97D16.1 | pseudogene | rs17693963 | 27710165 | 9.71e-09 | 169 | Male only | Male only | Male only |
| MONOAB | Monocytes [#/volume] in Blood by Automated count | immune/inflammatory | 6 | XXbac-BPG248L24.10 | pseudogene | rs9265540 | 31298164 | 1.23e-08 | 75 | Male only | Both sexes | Male only |
| MONOAB | Monocytes [#/volume] in Blood by Automated count | immune/inflammatory | 6 | HLA-F | protein_coding | rs885945 | 29696852 | 2.66e-08 | 391 | Male only | Both sexes | Male only |
| MONOAB | Monocytes [#/volume] in Blood by Automated count | immune/inflammatory | 6 | UBQLN1P1 | pseudogene | rs3129809 | 30335621 | 4.65e-08 | 285 | Male only | Both sexes | Male only |
| MONORE | Monocytes/100 leukocytes in Blood by Automated count | immune/inflammatory | 3 | SNORA24 | snoRNA | rs11719239 | 128419054 | 1.92e-16 | 17 | Both sexes | Both sexes | Both sexes |
| MONORE | Monocytes/100 leukocytes in Blood by Automated count | immune/inflammatory | 13 | TPTE2P5 | pseudogene | rs9532600 | 41434808 | 1.36e-11 | 77 | Female only | Both sexes | Female only |
| MONORE | Monocytes/100 leukocytes in Blood by Automated count | immune/inflammatory | 1 | RPAP2 | protein_coding | rs6603963 | 92821374 | 2.76e-10 | 207 | Female only | Both sexes | Female only |
| MONORE | Monocytes/100 leukocytes in Blood by Automated count | immune/inflammatory | 1 | RPAP2 | protein_coding | rs74767605 | 92770376 | 3.43e-10 | 7 | Female only | Both sexes | Female only |
| MONORE | Monocytes/100 leukocytes in Blood by Automated count | immune/inflammatory | 1 | EPHX4 | protein_coding | rs143792309 | 92505309 | 9.97e-09 | 5 | Female only | Both sexes | Female only |

| lab | full_name | group | CHR | Gene_Symbol | biotype | lead_snp | lead_BP | lead_P | total_snps_in_loci | independent_loci_in_sex_P1e6 | independent_loci_in_sex_P1e2 | independent_loci_in_sex_P5e8 |
|---|---|---|---|---|---|---|---|---|---|---|---|---|
| MONORE | Monocytes/100 leukocytes in Blood by Automated count | immune/inflammatory | 2 | MYO3B | protein_coding | rs7598148 | 171279752 | 1.92e-08 | 19 | Male only | Male only | Male only |
| MONORE | Monocytes/100 leukocytes in Blood by Automated count | immune/inflammatory | 13 | TPTE2P5 | pseudogene | rs4943818 | 41400473 | 2.07e-08 | 63 | Female only | Female only | Female only |
| MONORE | Monocytes/100 leukocytes in Blood by Automated count | immune/inflammatory | 20 | RP11-290F20.1 | lincRNA | rs1889237 | 48905978 | 4.40e-08 | 5 | Female only | Both sexes | Female only |
| MONORE | Monocytes/100 leukocytes in Blood by Automated count | immune/inflammatory | 3 | HIGD1A | protein_coding | rs6764909 | 42835889 | 4.45e-08 | 5 | Female only | Both sexes | Female only |
| MONORE | Monocytes/100 leukocytes in Blood by Automated count | immune/inflammatory | 1 | AC093577.1 | miRNA | rs78663632 | 93448422 | 4.84e-08 | 201 | Female only | Both sexes | Female only |
| MPV | Platelet mean volume [Entitic volume] in Blood by Automated count | blood | 1 | NUAK2 | protein_coding | rs17345997 | 205279612 | 8.04e-18 | 11 | Both sexes | Both sexes | Both sexes |
| MPV | Platelet mean volume [Entitic volume] in Blood by Automated count | blood | 12 | RP11-87C12.5 | lincRNA | rs745327 | 122456524 | 6.94e-15 | 9 | Both sexes | Both sexes | Both sexes |
| MPV | Platelet mean volume [Entitic volume] in Blood by Automated count | blood | 1 | NUAK2 | protein_coding | rs41264875 | 205272347 | 3.25e-14 | 7 | Both sexes | Both sexes | Female only |
| MPV | Platelet mean volume [Entitic volume] in Blood by Automated count | blood | 12 | RP3-454B23.1 | lincRNA | rs11064058 | 6263819 | 1.42e-12 | 19 | Both sexes | Both sexes | Both sexes |
| MPV | Platelet mean volume [Entitic volume] in Blood by Automated count | blood | 17 | GIT1 | protein_coding | rs894606 | 27909352 | 1.80e-12 | 3 | Female only | Both sexes | Female only |
| MPV | Platelet mean volume [Entitic volume] in Blood by Automated count | blood | 17 | MYO18A | protein_coding | rs8069100 | 27439527 | 9.50e-12 | 95 | Female only | Both sexes | Female only |
| MPV | Platelet mean volume [Entitic volume] in Blood by Automated count | blood | 12 | ORAI1 | protein_coding | rs712853 | 122079668 | 1.33e-11 | 27 | Both sexes | Both sexes | Female only |
| MPV | Platelet mean volume [Entitic volume] in Blood by Automated count | blood | 10 | RP11-351O1.3 | pseudogene | rs12773976 | 65264300 | 2.90e-11 | 7 | Both sexes | Both sexes | Female only |
| MPV | Platelet mean volume [Entitic volume] in Blood by Automated count | blood | 12 | ORAI1 | protein_coding | rs6486776 | 122057293 | 5.74e-11 | 17 | Female only | Both sexes | Female only |
| MPV | Platelet mean volume [Entitic volume] in Blood by Automated count | blood | 12 | HPD | protein_coding | rs2620340 | 122291261 | 1.77e-10 | 55 | Female only | Female only | Female only |
| MPV | Platelet mean volume [Entitic volume] in Blood by Automated count | blood | 23 | RP1-49C23.1 | pseudogene | X:57810911:G:A | 57810911 | 3.88e-10 | 75 | Both sexes | Both sexes | Both sexes |
| MPV | Platelet mean volume [Entitic volume] in Blood by Automated count | blood | 17 | MYO18A | protein_coding | rs16964837 | 27467093 | 3.92e-10 | 59 | Female only | Female only | Female only |
| MPV | Platelet mean volume [Entitic volume] in Blood by Automated count | blood | 23 | RP11-622K12.1 | processed_transcript | X:56793966:C:T | 56793966 | 4.23e-10 | 461 | Both sexes | Both sexes | Both sexes |

| lab | full_name | group | CHR | Gene_Symbol | biotype | lead_snp | lead_BP | lead_P | total_snps_in_loci | independent_loci_in_sex_P 1e6 | independent_loci_in_sex_P 1e2 | independent_loci_in_sex_P 5e8 |
|---|---|---|---|---|---|---|---|---|---|---|---|---|
| MPV | Platelet mean volume [Entitic volume] in Blood by Automated count | blood | 23 | KLF8 | protein_coding | X:56306059:T:G | 56306059 | 6.80e-10 | 531 | Both sexes | Both sexes | Both sexes |
| MPV | Platelet mean volume [Entitic volume] in Blood by Automated count | blood | 17 | TWF1P1 | pseudogene | rs61314136 | 27545367 | 8.26e-10 | 29 | Female only | Both sexes | Female only |
| MPV | Platelet mean volume [Entitic volume] in Blood by Automated count | blood | 12 | RP11-74M13.4 | pseudogene | rs7957053 | 57214086 | 1.24e-09 | 7 | Female only | Both sexes | Female only |
| MPV | Platelet mean volume [Entitic volume] in Blood by Automated count | blood | 17 | TWF1P1 | pseudogene | rs7213514 | 27537242 | 3.04e-09 | 11 | Female only | Both sexes | Female only |
| MPV | Platelet mean volume [Entitic volume] in Blood by Automated count | blood | 5 | RN7SL208P | misc_RNA | rs4704366 | 76137222 | 5.37e-09 | 7 | Female only | Both sexes | Female only |
| MPV | Platelet mean volume [Entitic volume] in Blood by Automated count | blood | 6 | NOTCH4 | protein_coding | rs374205 | 32196873 | 7.07e-09 | 55 | Female only | Both sexes | Female only |
| MPV | Platelet mean volume [Entitic volume] in Blood by Automated count | blood | 17 | GIT1 | protein_coding | rs550818 | 27901975 | 9.45e-09 | 19 | Both sexes | Both sexes | Female only |
| MPV | Platelet mean volume [Entitic volume] in Blood by Automated count | blood | 20 | ATP5E | protein_coding | rs151353 | 57604343 | 1.00e-08 | 15 | Female only | Both sexes | Female only |
| lab | full_name | group | CHR | Gene_Symbol | biotype | lead_snp | lead_BP | lead_P | total_snps_in_loci | independent_loci_in_sex_P 1e6 | independent_loci_in_sex_P 1e2 | independent_loci_in_sex_P 5e8 |
| MPV | Platelet mean volume [Entitic volume] in Blood by Automated count | blood | 12 | ERGIC2 | protein_coding | rs2278094 | 29534492 | 1.09e-08 | 25 | Female only | Both sexes | Female only |
| MPV | Platelet mean volume [Entitic volume] in Blood by Automated count | blood | 23 | RP13-188A5.1 | lincRNA | X:56017005:C:T | 56017005 | 1.39e-08 | 433 | Both sexes | Both sexes | Both sexes |
| MPV | Platelet mean volume [Entitic volume] in Blood by Automated count | blood | 1 | RP11-978I15.10 | antisense | rs12119496 | 247742899 | 1.64e-08 | 11 | Female only | Both sexes | Female only |
| MPV | Platelet mean volume [Entitic volume] in Blood by Automated count | blood | 6 | XXbac-BPG248L24.12 | pseudogene | rs9266233 | 31325430 | 1.67e-08 | 9 | Female only | Both sexes | Female only |
| MPV | Platelet mean volume [Entitic volume] in Blood by Automated count | blood | 12 | OVCH1-AS1 | protein_coding | rs10743664 | 29558585 | 2.63e-08 | 25 | Male only | Both sexes | Male only |
| MPV | Platelet mean volume [Entitic volume] in Blood by Automated count | blood | 17 | MYO18A | protein_coding | rs12601994 | 27447905 | 3.25e-08 | 31 | Female only | Female only | Female only |
| MPV | Platelet mean volume [Entitic volume] in Blood by Automated count | blood | 6 | ZDHHC20P2 | pseudogene | rs9266658 | 31347644 | 3.48e-08 | 23 | Female only | Both sexes | Female only |
| MPV | Platelet mean volume [Entitic volume] in Blood by Automated count | blood | 22 | RPL5P34 | pseudogene | rs12162975 | 43175426 | 4.24e-08 | 5 | Female only | Both sexes | Female only |
| Mg | Magnesium [Mass/volume] in Serum or Plasma | metabolic | 1 | DCAF6 | protein_coding | rs12136645 | 167913302 | 2.85e-08 | 243 | Male only | Male only | Male only |
| MmAcid | Methylmalonic acid | metabolic | 2 | HIBCH | protein_coding | rs291429 | 191197941 | 1.13e-14 | 69 | Both sexes | Both sexes | Both sexes |

| lab | full_name | group | CHR | Gene_Symbol | biotype | lead_snp | lead_BP | lead_P | total_snps_in_loci | independent_loci_in_sex_P1e6 | independent_loci_in_sex_P1e2 | independent_loci_in_sex_P5e8 |
|---|---|---|---|---|---|---|---|---|---|---|---|---|
| MmAcid | Methylmalonic acid | metabolic | 2 | INPP1 | protein_coding | rs3791813 | 191218371 | 1.02e-10 | 439 | Both sexes | Both sexes | Both sexes |
| Monocy | Monocytes [#/volume] in Blood by Manual count | immune/inflammatory | 1 | EPHX4 | protein_coding | rs143792309 | 92505309 | 2.28e-08 | 5 | Female only | Female only | Female only |
| NEUTM | Polymorphonuclear cells/100 leukocytes in Body fluid by Manual count | immune/inflammatory | 23 | MTND6P12 | pseudogene | X:5080916:T:C | 5080916 | 5.55e-09 | 141 | Female only | Female only | Female only |
| Na | Sodium in Blood | metabolic | 20 | PREX1 | protein_coding | rs34316626 | 47358484 | 1.46e-08 | 9 | Female only | Female only | Female only |
| O2SAT | Oxygen saturation calculated from oxygen partial pressure in arterial blood | vitals | 17 | CDK5RAP3 | protein_coding | rs17617053 | 46050669 | 2.31e-08 | 5 | Female only | Female only | Female only |
| PR_INTERVAL | PR Interval mean | cardiovascular | 6 | PRRC2A | protein_coding | rs1046080 | 31595882 | 1.60e-08 | 21 | Female only | Female only | Female only |
| PSA | PSA serum/plasma | cancer | 19 | AC011523.2 | antisense | rs7251465 | 51350791 | 4.03e-10 | 3 | GWAS only run in males | GWAS only run in males | GWAS only run in males |
| PTT.pt | aPTT in Platelet poor plasma | blood | 3 | RP11-795A2.3 | pseudogene | rs56380201 | 186429367 | 9.97e-21 | 9 | Both sexes | Both sexes | Both sexes |
| PTT.pt | aPTT in Platelet poor plasma | blood | 3 | RP11-573D15.8 | antisense | rs822373 | 186471186 | 1.02e-17 | 7 | Both sexes | Both sexes | Both sexes |
| PTT.pt | aPTT in Platelet poor plasma | blood | 3 | RP11-573D15.8 | antisense | rs2378065 | 186468856 | 1.72e-12 | 5 | Both sexes | Both sexes | Both sexes |
| PTT.pt | aPTT in Platelet poor plasma | blood | 5 | RAB24 | protein_coding | rs1128287 | 176728458 | 4.64e-12 | 7 | Both sexes | Both sexes | Both sexes |
| PTT.pt | aPTT in Platelet poor plasma | blood | 3 | RP11-795A2.3 | pseudogene | rs10937264 | 186428197 | 1.85e-09 | 37 | Both sexes | Both sexes | Both sexes |
| Pat.PT | Prothrombin time | blood | 13 | AL137002.1 | protein_coding | rs555212 | 113756540 | 1.20e-09 | 7 | Female only | Both sexes | Female only |
| Plt.Ct | Platelet Count | blood | 10 | RP11-351O1.3 | pseudogene | rs2893923 | 65261184 | 2.28e-09 | 3 | Female only | Both sexes | Female only |
| QRS_DURATION | QRS duration | cardiovascular | 6 | LAP3P2 | pseudogene | rs6930083 | 36634156 | 2.03e-09 | 23 | Both sexes | Both sexes | Both sexes |
| QRS_DURATION | QRS duration | cardiovascular | 12 | ATP5B | protein_coding | rs2950393 | 57036023 | 1.65e-08 | 125 | Female only | Both sexes | Female only |
| QRS_DURATION | QRS duration | cardiovascular | 3 | RP11-536I6.2 | lincRNA | rs4685095 | 14292081 | 2.15e-08 | 27 | Female only | Female only | Female only |
| QTB.NL | Quantiferon TB assay | immune/inflammatory | 6 | RP11-486E2.1 | pseudogene | rs80238754 | 81491052 | 5.23e-09 | 5 | Female only | Female only | Female only |
| QT_INTERVAL_C | QT interval corrected | cardiovascular | 1 | RP11-227F8.2 | lincRNA | rs7539281 | 162008034 | 1.47e-16 | 5 | Both sexes | Both sexes | Both sexes |
| QT_INTERVAL_C | QT interval corrected | cardiovascular | 1 | RP11-227F8.2 | lincRNA | rs7539367 | 162008079 | 5.75e-10 | 19 | Both sexes | Both sexes | Female only |
| RATE_C | Heart rate | vitals | 11 | DLG2 | protein_coding | rs79346577 | 84989662 | 2.88e-08 | 11 | Male only | Both sexes | Male only |
| RBC | Erythrocytes [#/volume] in Blood by Automated count | blood | 4 | AC006552.1 | lincRNA | rs80113191 | 55396420 | 1.85e-13 | 101 | Female only | Both sexes | Female only |

| lab | full_name | group | CHR | Gene_Symbol | biotype | lead_snp | lead_BP | lead_P | total_snps_in_loci | independent_loci_in_sex_P 1e6 | independent_loci_in_sex_P 1e2 | independent_loci_in_sex_P 5e8 |
|---|---|---|---|---|---|---|---|---|---|---|---|---|
| RBC | Erythrocytes [#/volume] in Blood by Automated count | blood | 6 | RP3-352A20.1 | pseudogene | rs9402659 | 135195889 | 3.48e-09 | 47 | Female only | Both sexes | Female only |
| RBC | Erythrocytes [#/volume] in Blood by Automated count | blood | 6 | RP3-352A20.1 | pseudogene | rs9494099 | 135193172 | 3.62e-09 | 59 | Female only | Both sexes | Female only |
| RDW | Erythrocyte distribution width [Ratio] by Automated count | blood | 6 | HIST1H3C | protein_coding | 6:26046744 | 26046744 | 1.04e-15 | 245 | Both sexes | Both sexes | Female only |
| RDW | Erythrocyte distribution width [Ratio] by Automated count | blood | 15 | WDR61 | protein_coding | rs870731 | 78604485 | 9.94e-12 | 37 | Female only | Both sexes | Female only |
| RDW | Erythrocyte distribution width [Ratio] by Automated count | blood | 15 | RP11-762H8.3 | antisense | rs8039884 | 78550081 | 1.60e-10 | 25 | Female only | Both sexes | Female only |
| RDW | Erythrocyte distribution width [Ratio] by Automated count | blood | 6 | LRRC16A | protein_coding | rs4712925 | 25389334 | 1.81e-09 | 193 | Female only | Both sexes | Female only |
| RDW | Erythrocyte distribution width [Ratio] by Automated count | blood | 6 | MIR3143 | miRNA | rs71559014 | 27122444 | 4.40e-09 | 3 | Female only | Both sexes | Female only |
| RDW | Erythrocyte distribution width [Ratio] by Automated count | blood | 6 | LRRC16A | protein_coding | rs116790746 | 25348745 | 1.14e-08 | 51 | Female only | Both sexes | Female only |
| RDW | Erythrocyte distribution width [Ratio] by Automated count | blood | 6 | HIST1H2AA | protein_coding | rs3936052 | 25715534 | 1.18e-08 | 37 | Female only | Both sexes | Female only |
| RDW | Erythrocyte distribution width [Ratio] by Automated count | blood | 6 | HCG4P7 | pseudogene | rs1611657 | 29842444 | 2.39e-08 | 155 | Female only | Female only | Female only |
| RDW | Erythrocyte distribution width [Ratio] by Automated count | blood | 6 | LRRC16A | protein_coding | rs9358860 | 25572245 | 3.28e-08 | 55 | Female only | Both sexes | Female only |
| RDW | Erythrocyte distribution width [Ratio] by Automated count | blood | 6 | GPX6 | protein_coding | rs34979126 | 28449380 | 4.28e-08 | 41 | Female only | Both sexes | Female only |
| RDWSD | Erythrocyte distribution width [Entitic volume] by Automated count | blood | 15 | RP11-762H8.3 | antisense | rs8039884 | 78550081 | 3.24e-09 | 25 | Both sexes | Both sexes | Both sexes |
| RDWSD | Erythrocyte distribution width [Entitic volume] by Automated count | blood | 15 | WDR61 | protein_coding | rs6495287 | 78594447 | 5.34e-09 | 39 | Both sexes | Both sexes | Female only |
| RDWSD | Erythrocyte distribution width [Entitic volume] by Automated count | blood | 2 | DPP10 | protein_coding | rs4849439 | 116857816 | 1.93e-08 | 33 | Male only | Male only | Male only |
| RDWSD | Erythrocyte distribution width [Entitic volume] by Automated count | blood | 16 | CTU2 | protein_coding | rs11076704 | 88777995 | 2.20e-08 | 11 | Female only | Both sexes | Female only |
| R_F | Rheumatoid factor [Units/volume] in Serum or Plasma | immune/inflammatory | 7 | AC004875.1 | miRNA | rs73719806 | 121189829 | 2.53e-08 | 67 | Male only | Male only | Male only |
| SU.Pro | Protein urine | urinary | 5 | RNU6-525P | snRNA | rs11739255 | 179887237 | 3.27e-08 | 3 | Male only | Male only | Male only |
| TBil | Total Bilirubin serum/plasma | metabolic | 12 | RP11-125O5.2 | protein_coding | rs10841753 | 21321370 | 9.29e-17 | 199 | Both sexes | Both sexes | Both sexes |

| lab | full_name | group | CHR | Gene_Symbol | biotype | lead_snp | lead_BP | lead_P | total_snps_in_loci | independent_loci_in_sex_P 1e6 | independent_loci_in_sex_P 1e2 | independent_loci_in_sex_P 5e8 |
|---|---|---|---|---|---|---|---|---|---|---|---|---|
| TBil | Total Bilirubin serum/plasma | metabolic | 12 | RP11-125O5.2 | protein_coding | rs4149042 | 21328872 | 1.73e-10 | 13 | Female only | Both sexes | Female only |
| TBil | Total Bilirubin serum/plasma | metabolic | 2 | MSL3P1 | pseudogene | rs2361515 | 234791098 | 4.49e-09 | 99 | Female only | Both sexes | Female only |
| TC.HDL | TC/HDL ratio | metabolic | 16 | AC012181.1 | miRNA | rs12923459 | 56989830 | 1.36e-09 | 5 | Female only | Both sexes | Female only |
| TERMINAL_40_MS | Terminal 40 ms | cardiovascular | 12 | RPL7P6 | pseudogene | rs34900245 | 19212108 | 4.71e-08 | 35 | Female only | Female only | Female only |
| TProSe | Protein serum/plasma | blood | 6 | MICB | protein_coding | rs2894222 | 31462035 | 3.25e-11 | 49 | Male only | Male only | Male only |
| TProSe | Protein serum/plasma | blood | 6 | HLA-B | protein_coding | rs9265969 | 31315706 | 4.26e-10 | 261 | Male only | Both sexes | Male only |
| TProSe | Protein serum/plasma | blood | 6 | GPSM3 | protein_coding | rs204989 | 32161852 | 4.59e-10 | 35 | Male only | Both sexes | Male only |
| TProSe | Protein serum/plasma | blood | 6 | HLA-N | pseudogene | rs3094061 | 30321189 | 1.77e-09 | 329 | Male only | Both sexes | Male only |
| TProSe | Protein serum/plasma | blood | 6 | GPANK1 | protein_coding | rs3117579 | 31633496 | 3.12e-09 | 129 | Male only | Both sexes | Male only |
| lab | full_name | group | CHR | Gene_Symbol | biotype | lead_snp | lead_BP | lead_P | total_snps_in_loci | independent_loci_in_sex_P 1e6 | independent_loci_in_sex_P 1e2 | independent_loci_in_sex_P 5e8 |
| TProSe | Protein serum/plasma | blood | 6 | HLA-F-AS1 | processed_transcript | rs1633088 | 29713580 | 3.48e-09 | 403 | Male only | Both sexes | Male only |
| TProSe | Protein serum/plasma | blood | 6 | HCP5 | sense_overlapping | rs3128981 | 31405620 | 3.98e-09 | 35 | Male only | Both sexes | Male only |
| TProSe | Protein serum/plasma | blood | 6 | NOTCH4 | protein_coding | rs3132935 | 32171075 | 7.66e-09 | 45 | Male only | Both sexes | Male only |
| TProSe | Protein serum/plasma | blood | 6 | NOTCH4 | protein_coding | rs693797 | 32204433 | 1.01e-08 | 25 | Male only | Male only | Male only |
| TProSe | Protein serum/plasma | blood | 6 | TRIM26 | protein_coding | rs2517611 | 30169327 | 1.09e-08 | 39 | Male only | Both sexes | Male only |
| TProSe | Protein serum/plasma | blood | 6 | HLA-B | protein_coding | rs2596498 | 31322688 | 1.13e-08 | 11 | Male only | Both sexes | Male only |
| TProSe | Protein serum/plasma | blood | 6 | STK19 | protein_coding | rs389884 | 31940897 | 1.55e-08 | 39 | Male only | Both sexes | Male only |
| TProSe | Protein serum/plasma | blood | 6 | RPL15P4 | pseudogene | rs2734573 | 31494738 | 1.63e-08 | 3 | Male only | Both sexes | Male only |
| TProSe | Protein serum/plasma | blood | 6 | MICB | protein_coding | rs2395043 | 31461966 | 2.50e-08 | 17 | Male only | Male only | Male only |
| TProSe | Protein serum/plasma | blood | 6 | PRRC2A | protein_coding | rs2261033 | 31603591 | 4.88e-08 | 11 | Male only | Male only | Male only |
| TProt | Total protein | blood | 6 | NOTCH4 | protein_coding | rs6901158 | 32205942 | 1.07e-12 | 129 | Both sexes | Both sexes | Male only |
| TProt | Total protein | blood | 6 | GPANK1 | protein_coding | rs3117579 | 31633496 | 2.14e-12 | 129 | Both sexes | Both sexes | Male only |
| TProt | Total protein | blood | 6 | HLA-C | protein_coding | rs3130444 | 31228634 | 8.35e-11 | 167 | Male only | Both sexes | Male only |
| TProt | Total protein | blood | 6 | C2 | protein_coding | rs497309 | 31892484 | 1.41e-10 | 27 | Male only | Both sexes | Male only |
| TProt | Total protein | blood | 6 | SFTA2 | protein_coding | rs3095153 | 30899195 | 2.80e-10 | 249 | Both sexes | Both sexes | Both sexes |

| lab | full_name | group | CHR | Gene_Symbol | biotype | lead_snp | lead_BP | lead_P | total_snps_in_loci | independent_loci_in_sex_P1e6 | independent_loci_in_sex_P1e2 | independent_loci_in_sex_P5e8 |
|---|---|---|---|---|---|---|---|---|---|---|---|---|
| TProt | Total protein | blood | 6 | POU5F1 | protein_coding | rs885948 | 31140532 | 8.59e-10 | 53 | Male only | Both sexes | Male only |
| TProt | Total protein | blood | 6 | HCP5 | sense_overlapping | rs3128981 | 31405620 | 1.19e-09 | 35 | Both sexes | Both sexes | Male only |
| TProt | Total protein | blood | 6 | HLA-N | pseudogene | rs3130375 | 30321732 | 2.12e-09 | 329 | Both sexes | Both sexes | Both sexes |
| TProt | Total protein | blood | 6 | HLA-C | protein_coding | rs9264186 | 31220017 | 4.05e-09 | 109 | Male only | Both sexes | Male only |
| TProt | Total protein | blood | 6 | TRIM26 | protein_coding | rs2517611 | 30169327 | 4.18e-09 | 39 | Both sexes | Both sexes | Male only |
| TProt | Total protein | blood | 6 | PSORS1C1 | protein_coding | rs528878831 | 31098832 | 7.51e-09 | 97 | Male only | Both sexes | Male only |
| TProt | Total protein | blood | 6 | NOTCH4 | protein_coding | rs3132935 | 32171075 | 1.68e-08 | 7 | Male only | Both sexes | Male only |
| TProt | Total protein | blood | 6 | NOTCH4 | protein_coding | rs715299 | 32189841 | 2.08e-08 | 3 | Male only | Both sexes | Male only |
| TProt | Total protein | blood | 6 | RNF39 | protein_coding | rs9261290 | 30038647 | 2.89e-08 | 127 | Male only | Both sexes | Male only |
| TProt | Total protein | blood | 6 | ATAT1 | protein_coding | rs9262130 | 30603519 | 3.11e-08 | 21 | Both sexes | Both sexes | Male only |
| TProt | Total protein | blood | 6 | XXbac-BPG248L24.12 | pseudogene | rs2249935 | 31327178 | 4.40e-08 | 3 | Male only | Both sexes | Male only |
| TSH | TSH | endocrine | 9 | RP11-23B15.1 | lincRNA | 9:100542706 | 100542706 | 1.18e-17 | 3 | Female only | Both sexes | Female only |
| TSH | TSH | endocrine | 6 | RP11-344J7.2 | lincRNA | rs729761 | 43804571 | 4.91e-17 | 11 | Both sexes | Both sexes | Both sexes |
| TSH | TSH | endocrine | 16 | RP11-345M22.1 | lincRNA | rs9923437 | 79710028 | 9.77e-15 | 3 | Both sexes | Both sexes | Both sexes |
| TSH | TSH | endocrine | 1 | RNU4-28P | snRNA | rs12123411 | 19862592 | 4.19e-12 | 39 | Male only | Both sexes | Male only |
| TSH | TSH | endocrine | 5 | ZBED3-AS1 | antisense | rs12171500 | 76460134 | 1.26e-11 | 5 | Male only | Both sexes | Male only |
| TSH | TSH | endocrine | 6 | RP11-344J7.2 | lincRNA | rs6933958 | 43823689 | 2.04e-11 | 5 | Both sexes | Both sexes | Both sexes |
| TSH | TSH | endocrine | 4 | RP11-503L19.1 | lincRNA | rs72735922 | 149593414 | 3.09e-10 | 25 | Female only | Female only | Female only |
| TSH | TSH | endocrine | 2 | AC007563.5 | antisense | rs2888429 | 217624194 | 5.45e-10 | 45 | Both sexes | Both sexes | Both sexes |
| TSH | TSH | endocrine | 9 | C9orf156 | protein_coding | rs13301569 | 100655780 | 9.69e-10 | 17 | Female only | Both sexes | Female only |
| TSH | TSH | endocrine | 5 | ZBED3-AS1 | antisense | rs7708949 | 76463481 | 2.47e-09 | 9 | Male only | Both sexes | Male only |
| TSH | TSH | endocrine | 9 | C9orf156 | protein_coding | rs7028759 | 100677883 | 7.12e-09 | 3 | Female only | Female only | Female only |
| TSH | TSH | endocrine | 15 | RP11-353B9.1 | sense_overlapping | rs17404262 | 49967619 | 3.17e-08 | 3 | Female only | Both sexes | Female only |
| TSH | TSH | endocrine | 9 | RP11-23B15.1 | lincRNA | rs12352658 | 100551768 | 3.22e-08 | 7 | Female only | Both sexes | Female only |
| TT4 | Thyroxine (T4) [Mass/volume] in Serum or Plasma | endocrine | 23 | RP11-560L11.1 | pseudogene | X:105317946:G:T | 105317946 | 3.19e-10 | 27 | Male only | Both sexes | Male only |

| lab | full_name | group | CHR | Gene_Symbol | biotype | lead_snp | lead_BP | lead_P | total_snps_in_loci | independent_loci_in_sex_P 1e6 | independent_loci_in_sex_P 1e2 | independent_loci_in_sex_P 5e8 |
|---|---|---|---|---|---|---|---|---|---|---|---|---|
| UWBC | Leukocytes [#/area] in Urine sediment by Microscopy high power field | urinary | 8 | ENTPD4 | protein_coding | 8:23309914 | 23309914 | 2.21e-08 | 19 | Male only | Male only | Male only |
| UricA | Urate [Mass/volume] in Serum or Plasma | urinary | 4 | AC006499.4 | pseudogene | rs6833292 | 10272429 | 5.47e-14 | 31 | Both sexes | Both sexes | Female only |
| UricA | Urate [Mass/volume] in Serum or Plasma | urinary | 4 | AC006499.1 | pseudogene | rs993172 | 10324361 | 1.31e-11 | 23 | Female only | Both sexes | Female only |
| VD25 | Calcitriol [Mass/volume] in Serum or Plasma | metabolic | 11 | CYP2R1 | protein_coding | rs1993116 | 14910234 | 1.94e-09 | 65 | Female only | Female only | Female only |
| Vi.B12 | Vitamin B12 | metabolic | 19 | FGF21 | protein_coding | rs838133 | 49259529 | 5.96e-10 | 41 | Male only | Both sexes | Male only |
| WBC | Leukocytes [#/volume] in Blood by Automated count | immune/inflammatory | 6 | TRIM27 | protein_coding | rs3118361 | 28898287 | 7.07e-10 | 313 | Male only | Both sexes | Male only |
| WBC | Leukocytes [#/volume] in Blood by Automated count | immune/inflammatory | 6 | HCG17 | lincRNA | rs3094069 | 30251391 | 8.51e-10 | 325 | Both sexes | Both sexes | Both sexes |
| WBC | Leukocytes [#/volume] in Blood by Automated count | immune/inflammatory | 6 | RP1-97D16.1 | pseudogene | rs17693963 | 27710165 | 8.73e-10 | 177 | Male only | Both sexes | Male only |
| WBC | Leukocytes [#/volume] in Blood by Automated count | immune/inflammatory | 6 | XXbac-BPG181B23.7 | lincRNA | rs2523466 | 31363026 | 1.42e-09 | 11 | Female only | Both sexes | Female only |
| WBC | Leukocytes [#/volume] in Blood by Automated count | immune/inflammatory | 6 | BTN3A1 | protein_coding | rs3799378 | 26404374 | 4.23e-09 | 23 | Male only | Male only | Male only |
| WBC | Leukocytes [#/volume] in Blood by Automated count | immune/inflammatory | 6 | ZSCAN31 | protein_coding | rs35016036 | 28314880 | 5.80e-09 | 189 | Male only | Both sexes | Male only |
| WBC | Leukocytes [#/volume] in Blood by Automated count | immune/inflammatory | 6 | SKIV2L | protein_coding | rs410851 | 31936668 | 6.34e-09 | 39 | Both sexes | Both sexes | Male only |
| WBC | Leukocytes [#/volume] in Blood by Automated count | immune/inflammatory | 6 | OR2W2P | pseudogene | rs138234416 | 27992898 | 1.41e-08 | 25 | Male only | Both sexes | Male only |
| WBC | Leukocytes [#/volume] in Blood by Automated count | immune/inflammatory | 17 | PGAP3 | protein_coding | rs1495100 | 37832279 | 2.40e-08 | 93 | Female only | Female only | Female only |
| WBC | Leukocytes [#/volume] in Blood by Automated count | immune/inflammatory | 6 | AL662890.1 | miRNA | rs1233608 | 28736189 | 2.85e-08 | 13 | Male only | Both sexes | Male only |
| Weight | Weight | vitals | 18 | RP11-795H16.3 | lincRNA | rs11664212 | 57739179 | 1.16e-12 | 29 | Female only | Both sexes | Female only |
| Weight | Weight | vitals | 18 | RP11-795H16.3 | lincRNA | rs8086627 | 57743096 | 1.41e-10 | 33 | Female only | Both sexes | Female only |
| Weight | Weight | vitals | 6 | NQO2 | protein_coding | rs11242816 | 2995459 | 4.38e-08 | 21 | Male only | Male only | Male only |
| X25HyD3 | 25-Hydroxyvitamin D3 [Mass/volume] in Serum or Plasma | metabolic | 4 | RP11-545L5.1 | lincRNA | rs6821488 | 72754943 | 1.78e-11 | 111 | Female only | Both sexes | Female only |
| X25HyD3 | 25-Hydroxyvitamin D3 [Mass/volume] in Serum or Plasma | metabolic | 11 | CYP2R1 | protein_coding | rs10766196 | 14913131 | 5.91e-09 | 75 | Female only | Both sexes | Female only |
| lab | full_name | group | CHR | Gene_Symbol | biotype | lead_snp | lead_BP | lead_P | total_snps_in_loci | independent_loci_in_sex_P 1e6 | independent_loci_in_sex_P 1e2 | independent_loci_in_sex_P 5e8 |
| pCO2 | Carbon dioxide [Partial pressure] in Arterial blood | blood | 6 | CDKAL1 | protein_coding | rs79529169 | 21184849 | 1.07e-08 | 49 | Female only | Female only | Female only |

#### STable7: Independent loci for per-person median trait values that did not have data in the GWAS catalog

106 rows × 13 columns

| lab | full_name | group | CHR | Gene_Symbol | biotype | lead_snp | lead_BP | lead_P | total_snps_in_loci | independent_loci_in_sex_P 1e6 | independent_loci_in_sex_P 1e2 | independent_loci_in_sex_P 5e8 |
|---|---|---|---|---|---|---|---|---|---|---|---|---|
| A.2Gg_ | Alpha 2 globulin [Mass/volume] in Serum or Plasma by Electrophoresis | liver | 16 | PMFBP1 | protein_coding | rs12445401 | 72148419 | 1.07e-10 | 25 | Male only | Both sexes | Male only |
| A.2Gg_ | Alpha 2 globulin [Mass/volume] in Serum or Plasma by Electrophoresis | liver | 16 | PKD1L3 | processed_transcript | rs11645582 | 72029008 | 4.10e-09 | 41 | Male only | Both sexes | Male only |
| C.Gluc | Glucose spinal fluid | metabolic | 13 | RN7SL571P | misc_RNA | rs61963436 | 77061127 | 3.50e-09 | 45 | Female only | Female only | Female only |
| DLCOOVA_PRD | DLCOOVA_PRD | lung | 12 | LINC00942 | lincRNA | rs4765821 | 1611846 | 2.06e-08 | 43 | Female only | Female only | Female only |
| IGAB | Immature granulocytes [Presence] in Blood by Automated Count | immune/inflammatory | 6 | RP1-111C20.3 | lincRNA | rs2451257 | 159507026 | 1.47e-26 | 15 | Both sexes | Both sexes | Both sexes |
| IGAB | Immature granulocytes [Presence] in Blood by Automated Count | immune/inflammatory | 9 | LRRC8A | protein_coding | rs34305034 | 131654547 | 1.75e-24 | 3 | Both sexes | Both sexes | Both sexes |
| IGAB | Immature granulocytes [Presence] in Blood by Automated Count | immune/inflammatory | 6 | RP1-111C20.3 | lincRNA | rs2451258 | 159506600 | 4.80e-12 | 21 | Male only | Both sexes | Male only |
| IGAB | Immature granulocytes [Presence] in Blood by Automated Count | immune/inflammatory | 10 | PRTFDC1 | protein_coding | rs7896608 | 25206412 | 2.18e-11 | 15 | Male only | Both sexes | Male only |
| IGAB | Immature granulocytes [Presence] in Blood by Automated Count | immune/inflammatory | 9 | ENDOG | protein_coding | rs4837316 | 131577450 | 4.61e-11 | 55 | Both sexes | Both sexes | Both sexes |
| IGAB | Immature granulocytes [Presence] in Blood by Automated Count | immune/inflammatory | 2 | ITGA4 | protein_coding | rs6740847 | 182308352 | 6.98e-10 | 15 | Female only | Both sexes | Female only |
| IGAB | Immature granulocytes [Presence] in Blood by Automated Count | immune/inflammatory | 22 | DEPDC5 | protein_coding | rs5749339 | 32315735 | 1.84e-09 | 35 | Both sexes | Both sexes | Female only |
| IGAB | Immature granulocytes [Presence] in Blood by Automated Count | immune/inflammatory | 6 | RP11-13P5.1 | lincRNA | rs62431905 | 159510162 | 1.26e-08 | 7 | Both sexes | Both sexes | Male only |
| IGAB | Immature granulocytes [Presence] in Blood by Automated Count | immune/inflammatory | 7 | CCM2 | protein_coding | rs2158564 | 45038881 | 1.79e-08 | 85 | Both sexes | Both sexes | Male only |
| Infant.Apgar.Color.1.M.1 | 1 minute Apgar color | vitals | 13 | METTL21C | protein_coding | rs9518810 | 103343428 | 2.79e-08 | 2 | GWAS only run in females | GWAS only run in females | GWAS only run in females |
| Infant.Apgar.Resp.5.M.1 | 5 minute Apgar respiratory rate | vitals | 13 | ARHGEF7 | protein_coding | rs58934014 | 111795572 | 2.33e-08 | 5 | GWAS only run in females | GWAS only run in females | GWAS only run in females |
| Luc_percent_ | Luc% | immune/inflammatory | 17 | BZRAP1-AS1 | antisense | rs75394768 | 56413635 | 1.32e-09 | 3 | Both sexes | Both sexes | Both sexes |
| Meta | Metamyelocytes/100 leukocytes in Blood by Manual count | blood | 23 | RP5-998G20.1 | pseudogene | X:87304109:T:G | 87304109 | 3.10e-08 | 289 | Male only | Male only | Male only |

| lab | full_name | group | CHR | Gene_Symbol | biotype | lead_snp | lead_BP | lead_P | total_snps_in_loci | independent_loci_in_sex_P 1e6 | independent_loci_in_sex_P 1e2 | independent_loci_in_sex_P 5e8 |
|---|---|---|---|---|---|---|---|---|---|---|---|---|
| PCAct | Protein C [Units/volume] in Platelet poor plasma by Chromogenic method | blood | 20 | EDEM2 | protein_coding | rs8119351 | 33754405 | 8.72e-09 | 317 | Female only | Female only | Female only |
| R.APACHE.TEMPERATURE | Body temperature | vitals | 12 | RP11-560G2.1 | antisense | rs10879892 | 75647143 | 9.17e-09 | 11 | Male only | Male only | Male only |
| R.APACHE.TEMPERATURE | Body temperature | vitals | 2 | KLF7 | protein_coding | rs16839150 | 207955158 | 1.31e-08 | 29 | Male only | Male only | Male only |
| R.ED.PRE.ARRIVAL.RESP.RATE | Respiratory rate | vitals | 13 | GPC5 | protein_coding | rs67133719 | 93114447 | 4.22e-09 | 9 | Female only | Female only | Female only |
| R.IP.IDEAL.BODY.WEIGHT | Ideal body weight | vitals | 4 | LCORL | protein_coding | rs112675878 | 18027696 | 4.07e-17 | 449 | Female only | Both sexes | Female only |
| R.IP.IDEAL.BODY.WEIGHT | Ideal body weight | vitals | 20 | GDF5 | protein_coding | rs143384 | 34025756 | 1.37e-13 | 221 | Both sexes | Both sexes | Both sexes |
| R.IP.IDEAL.BODY.WEIGHT | Ideal body weight | vitals | 3 | ZBTB38 | protein_coding | rs6440006 | 141142691 | 1.94e-13 | 75 | Both sexes | Both sexes | Both sexes |
| R.IP.IDEAL.BODY.WEIGHT | Ideal body weight | vitals | 2 | EFEMP1 | protein_coding | rs1430203 | 56072509 | 7.57e-12 | 103 | Male only | Both sexes | Male only |
| R.IP.IDEAL.BODY.WEIGHT | Ideal body weight | vitals | 15 | ADAMTSL3 | protein_coding | rs1812707 | 84547222 | 4.67e-11 | 137 | Female only | Both sexes | Female only |
| R.IP.IDEAL.BODY.WEIGHT | Ideal body weight | vitals | 4 | HHIP-AS1 | antisense | rs13106087 | 145566864 | 2.59e-10 | 19 | Both sexes | Both sexes | Female only |
| R.IP.IDEAL.BODY.WEIGHT | Ideal body weight | vitals | 18 | CABLES1 | protein_coding | rs9947743 | 20708321 | 7.32e-10 | 39 | Both sexes | Both sexes | Both sexes |
| R.IP.IDEAL.BODY.WEIGHT | Ideal body weight | vitals | 17 | ATAD5 | protein_coding | rs3764419 | 29164023 | 9.79e-10 | 29 | Female only | Female only | Female only |
| R.IP.IDEAL.BODY.WEIGHT | Ideal body weight | vitals | 12 | HMGA2 | protein_coding | rs1351394 | 66351826 | 2.29e-09 | 31 | Female only | Both sexes | Female only |

| lab | full_name | group | CHR | Gene_Symbol | biotype | lead_snp | lead_BP | lead_P | total_snps_in_loci | independent_loci_in_sex_P 1e6 | independent_loci_in_sex_P 1e2 | independent_loci_in_sex_P 5e8 |
|---|---|---|---|---|---|---|---|---|---|---|---|---|
| R.IP.IDEAL.BODY.WEIGHT | Ideal body weight | vitals | 3 | ZBTB38 | protein_coding | rs13100711 | 141082990 | 4.88e-09 | 19 | Female only | Both sexes | Female only |
| R.IP.IDEAL.BODY.WEIGHT | Ideal body weight | vitals | 2 | LTBP1 | protein_coding | rs6743685 | 33389595 | 5.23e-09 | 201 | Female only | Female only | Female only |
| R.IP.IDEAL.BODY.WEIGHT | Ideal body weight | vitals | 13 | DLEU1 | protein_coding | rs2066647 | 50782074 | 6.49e-09 | 31 | Female only | Both sexes | Female only |
| R.IP.IDEAL.BODY.WEIGHT | Ideal body weight | vitals | 2 | EFEMP1 | protein_coding | rs59985551 | 56106928 | 6.74e-09 | 17 | Male only | Both sexes | Male only |
| R.IP.IDEAL.BODY.WEIGHT | Ideal body weight | vitals | 12 | CCDC91 | protein_coding | rs11049356 | 28271456 | 6.78e-09 | 465 | Female only | Female only | Female only |
| R.IP.IDEAL.BODY.WEIGHT | Ideal body weight | vitals | 1 | DNM3 | protein_coding | rs678962 | 172189889 | 1.44e-08 | 37 | Female only | Both sexes | Female only |
| R.IP.IDEAL.BODY.WEIGHT | Ideal body weight | vitals | 11 | SSH3 | protein_coding | rs76335321 | 67097778 | 1.99e-08 | 7 | Female only | Both sexes | Female only |
| R.IP.IDEAL.BODY.WEIGHT | Ideal body weight | vitals | 13 | DLEU1 | protein_coding | rs3116598 | 51099616 | 2.00e-08 | 109 | Both sexes | Both sexes | Female only |
| R.IP.IDEAL.BODY.WEIGHT | Ideal body weight | vitals | 18 | CABLES1 | protein_coding | rs4567840 | 20705158 | 2.09e-08 | 65 | Male only | Both sexes | Male only |
| R.IP.IDEAL.BODY.WEIGHT | Ideal body weight | vitals | 15 | ACAN | protein_coding | rs2280470 | 89395626 | 2.10e-08 | 23 | Female only | Female only | Female only |
| R.IP.IDEAL.BODY.WEIGHT | Ideal body weight | vitals | 7 | IGF2BP3 | protein_coding | rs67397633 | 23491190 | 2.11e-08 | 49 | Male only | Both sexes | Male only |
| R.IP.IDEAL.BODY.WEIGHT | Ideal body weight | vitals | 2 | EFEMP1 | protein_coding | rs11125609 | 56115834 | 2.12e-08 | 5 | Male only | Male only | Male only |
| R.IP.IDEAL.BODY.WEIGHT | Ideal body weight | vitals | 6 | RP11-486E2.1 | pseudogene | rs9359426 | 81442039 | 4.53e-08 | 181 | Female only | Both sexes | Female only |

| lab | full_name | group | CHR | Gene_Symbol | biotype | lead_snp | lead_BP | lead_P | total_snps_in_loci | independent_loci_in_sex_P 1e6 | independent_loci_in_sex_P 1e2 | independent_loci_in_sex_P 5e8 |
|---|---|---|---|---|---|---|---|---|---|---|---|---|
| R.RT..FEMALE.ADULT..IDEAL.BODY.WEIGHT | Ideal body weight — female | vitals | 4 | LCORL | protein_coding | rs112675878 | 18027696 | 3.98e-17 | 449 | Female only | Both sexes | Female only |
| R.RT..FEMALE.ADULT..IDEAL.BODY.WEIGHT | Ideal body weight — female | vitals | 20 | GDF5 | protein_coding | rs143384 | 34025756 | 1.22e-13 | 221 | Both sexes | Both sexes | Both sexes |
| R.RT..FEMALE.ADULT..IDEAL.BODY.WEIGHT | Ideal body weight — female | vitals | 3 | ZBTB38 | protein_coding | rs6440006 | 141142691 | 2.10e-13 | 75 | Both sexes | Both sexes | Both sexes |
| R.RT..FEMALE.ADULT..IDEAL.BODY.WEIGHT | Ideal body weight — female | vitals | 2 | EFEMP1 | protein_coding | rs1430203 | 56072509 | 7.85e-12 | 103 | Male only | Both sexes | Male only |
| R.RT..FEMALE.ADULT..IDEAL.BODY.WEIGHT | Ideal body weight — female | vitals | 15 | ADAMTSL3 | protein_coding | rs1812707 | 84547222 | 4.72e-11 | 137 | Female only | Both sexes | Female only |
| R.RT..FEMALE.ADULT..IDEAL.BODY.WEIGHT | Ideal body weight — female | vitals | 4 | HHIP-AS1 | antisense | rs13106087 | 145566864 | 3.07e-10 | 19 | Both sexes | Both sexes | Female only |
| R.RT..FEMALE.ADULT..IDEAL.BODY.WEIGHT | Ideal body weight — female | vitals | 18 | CABLES1 | protein_coding | rs9947743 | 20708321 | 7.53e-10 | 39 | Both sexes | Both sexes | Both sexes |
| R.RT..FEMALE.ADULT..IDEAL.BODY.WEIGHT | Ideal body weight — female | vitals | 17 | ATAD5 | protein_coding | rs3764419 | 29164023 | 8.90e-10 | 29 | Female only | Female only | Female only |
| R.RT..FEMALE.ADULT..IDEAL.BODY.WEIGHT | Ideal body weight — female | vitals | 12 | HMGA2 | protein_coding | rs1351394 | 66351826 | 2.67e-09 | 31 | Female only | Both sexes | Female only |

| lab | full_name | group | CHR | Gene_Symbol | biotype | lead_snp | lead_BP | lead_P | total_snps_in_loci | independent_loci_in_sex_P 1e6 | independent_loci_in_sex_P 1e2 | independent_loci_in_sex_P 5e8 |
|---|---|---|---|---|---|---|---|---|---|---|---|---|
| R.RT..FEMALE.ADULT..IDEAL.BODY.WEIGHT | Ideal body weight — female | vitals | 3 | ZBTB38 | protein_coding | rs13100711 | 141082990 | 5.15e-09 | 19 | Female only | Both sexes | Female only |
| R.RT..FEMALE.ADULT..IDEAL.BODY.WEIGHT | Ideal body weight — female | vitals | 2 | LTBP1 | protein_coding | rs6743685 | 33389595 | 5.51e-09 | 201 | Female only | Female only | Female only |
| R.RT..FEMALE.ADULT..IDEAL.BODY.WEIGHT | Ideal body weight — female | vitals | 13 | DLEU1 | protein_coding | rs2066647 | 50782074 | 6.40e-09 | 31 | Female only | Both sexes | Female only |
| R.RT..FEMALE.ADULT..IDEAL.BODY.WEIGHT | Ideal body weight — female | vitals | 2 | EFEMP1 | protein_coding | rs59985551 | 56106928 | 6.79e-09 | 17 | Male only | Both sexes | Male only |
| R.RT..FEMALE.ADULT..IDEAL.BODY.WEIGHT | Ideal body weight — female | vitals | 12 | CCDC91 | protein_coding | rs11049356 | 28271456 | 7.01e-09 | 465 | Female only | Female only | Female only |
| R.RT..FEMALE.ADULT..IDEAL.BODY.WEIGHT | Ideal body weight — female | vitals | 1 | DNM3 | protein_coding | rs678962 | 172189889 | 1.36e-08 | 37 | Female only | Both sexes | Female only |
| R.RT..FEMALE.ADULT..IDEAL.BODY.WEIGHT | Ideal body weight — female | vitals | 11 | SSH3 | protein_coding | rs76335321 | 67097778 | 1.75e-08 | 7 | Female only | Both sexes | Female only |
| R.RT..FEMALE.ADULT..IDEAL.BODY.WEIGHT | Ideal body weight — female | vitals | 15 | ACAN | protein_coding | rs2280470 | 89395626 | 1.87e-08 | 23 | Female only | Female only | Female only |

| lab | full_name | group | CHR | Gene_Symbol | biotype | lead_snp | lead_BP | lead_P | total_snps_in_loci | independent_loci_in_sex_P 1e6 | independent_loci_in_sex_P 1e2 | independent_loci_in_sex_P 5e8 |

| lab | full_name | group | CHR | Gene_Symbol | biotype | lead_snp | lead_BP | lead_P | total_snps_in_loci | independent_loci_in_sex_P 1e6 | independent_loci_in_sex_P 1e2 | independent_loci_in_sex_P 5e8 |
|---|---|---|---|---|---|---|---|---|---|---|---|---|
| R.RT..FEMALE.ADULT..IDEAL.BODY.WEIGHT | Ideal body weight — female | vitals | 13 | DLEU1 | protein_coding | rs3116598 | 51099616 | 1.97e-08 | 109 | Both sexes | Both sexes | Female only |

| lab | full_name | group | CHR | Gene_Symbol | biotype | lead_snp | lead_BP | lead_P | total_snps_in_loci | independent_loci_in_sex_P 1e6 | independent_loci_in_sex_P 1e2 | independent_loci_in_sex_P 5e8 |
|---|---|---|---|---|---|---|---|---|---|---|---|---|
| R.RT..FEMALE.ADULT..IDEAL.BODY.WEIGHT | Ideal body weight — female | vitals | 7 | IGF2BP3 | protein_coding | rs67397633 | 23491190 | 2.13e-08 | 49 | Male only | Both sexes | Male only |
| R.RT..FEMALE.ADULT..IDEAL.BODY.WEIGHT | Ideal body weight — female | vitals | 18 | CABLES1 | protein_coding | rs4567840 | 20705158 | 2.14e-08 | 65 | Male only | Both sexes | Male only |
| R.RT..FEMALE.ADULT..IDEAL.BODY.WEIGHT | Ideal body weight — female | vitals | 2 | EFEMP1 | protein_coding | rs11125609 | 56115834 | 2.15e-08 | 5 | Male only | Male only | Male only |
| R.RT..FEMALE.ADULT..IDEAL.BODY.WEIGHT | Ideal body weight — female | vitals | 6 | RP11-486E2.1 | pseudogene | rs9359426 | 81442039 | 4.35e-08 | 181 | Female only | Both sexes | Female only |
| R.RT..FEMALE.ADULT..IDEAL.BODY.WEIGHT | Ideal body weight — female | vitals | 2 | EFEMP1 | protein_coding | rs1346790 | 56091871 | 4.88e-08 | 65 | Male only | Male only | Male only |
| R.RT..MALE.ADULT..IDEAL.BODY.WEIGHT | Ideal body weight — male | vitals | 4 | LCORL | protein_coding | rs112675878 | 18027696 | 3.98e-17 | 449 | Female only | Both sexes | Female only |
| R.RT..MALE.ADULT..IDEAL.BODY.WEIGHT | Ideal body weight — male | vitals | 20 | GDF5 | protein_coding | rs143384 | 34025756 | 1.22e-13 | 221 | Both sexes | Both sexes | Both sexes |
| R.RT..MALE.ADULT..IDEAL.BODY.WEIGHT | Ideal body weight — male | vitals | 3 | ZBTB38 | protein_coding | rs6440006 | 141142691 | 2.10e-13 | 75 | Both sexes | Both sexes | Both sexes |
| R.RT..MALE.ADULT..IDEAL.BODY.WEIGHT | Ideal body weight — male | vitals | 2 | EFEMP1 | protein_coding | rs1430203 | 56072509 | 7.85e-12 | 103 | Male only | Both sexes | Male only |

| lab | full_name | group | CHR | Gene_Symbol | biotype | lead_snp | lead_BP | lead_P | total_snps_in_loci | independent_loci_in_sex_P 1e6 | independent_loci_in_sex_P 1e2 | independent_loci_in_sex_P 5e8 |
|---|---|---|---|---|---|---|---|---|---|---|---|---|
| R.RT..MALE.ADULT..IDEAL.BODY.WEIGHT | Ideal body weight — male | vitals | 15 | ADAMTSL3 | protein_coding | rs1812707 | 84547222 | 4.72e-11 | 137 | Female only | Both sexes | Female only |
| R.RT..MALE.ADULT..IDEAL.BODY.WEIGHT | Ideal body weight — male | vitals | 4 | HHIP-AS1 | antisense | rs13106087 | 145566864 | 3.07e-10 | 19 | Both sexes | Both sexes | Female only |
| R.RT..MALE.ADULT..IDEAL.BODY.WEIGHT | Ideal body weight — male | vitals | 18 | CABLES1 | protein_coding | rs9947743 | 20708321 | 7.53e-10 | 39 | Both sexes | Both sexes | Both sexes |
| R.RT..MALE.ADULT..IDEAL.BODY.WEIGHT | Ideal body weight — male | vitals | 17 | ATAD5 | protein_coding | rs3764419 | 29164023 | 8.90e-10 | 29 | Female only | Female only | Female only |
| R.RT..MALE.ADULT..IDEAL.BODY.WEIGHT | Ideal body weight — male | vitals | 12 | HMGA2 | protein_coding | rs1351394 | 66351826 | 2.67e-09 | 31 | Female only | Both sexes | Female only |
| R.RT..MALE.ADULT..IDEAL.BODY.WEIGHT | Ideal body weight — male | vitals | 3 | ZBTB38 | protein_coding | rs13100711 | 141082990 | 5.15e-09 | 19 | Female only | Both sexes | Female only |
| R.RT..MALE.ADULT..IDEAL.BODY.WEIGHT | Ideal body weight — male | vitals | 2 | LTBP1 | protein_coding | rs6743685 | 33389595 | 5.51e-09 | 201 | Female only | Female only | Female only |
| R.RT..MALE.ADULT..IDEAL.BODY.WEIGHT | Ideal body weight — male | vitals | 13 | DLEU1 | protein_coding | rs2066647 | 50782074 | 6.40e-09 | 31 | Female only | Both sexes | Female only |
| R.RT..MALE.ADULT..IDEAL.BODY.WEIGHT | Ideal body weight — male | vitals | 2 | EFEMP1 | protein_coding | rs59985551 | 56106928 | 6.79e-09 | 17 | Male only | Both sexes | Male only |

| lab | full_name | group | CHR | Gene_Symbol | biotype | lead_snp | lead_BP | lead_P | total_snps_in_loci | independent_loci_in_sex_P 1e6 | independent_loci_in_sex_P 1e2 | independent_loci_in_sex_P 5e8 |
|---|---|---|---|---|---|---|---|---|---|---|---|---|
| R.RT..MALE.ADULT..IDEAL.BODY.WEIGHT | Ideal body weight — male | vitals | 12 | CCDC91 | protein_coding | rs11049356 | 28271456 | 7.01e-09 | 465 | Female only | Female only | Female only |
| R.RT..MALE.ADULT..IDEAL.BODY.WEIGHT | Ideal body weight — male | vitals | 1 | DNM3 | protein_coding | rs678962 | 172189889 | 1.36e-08 | 37 | Female only | Both sexes | Female only |
| R.RT..MALE.ADULT..IDEAL.BODY.WEIGHT | Ideal body weight — male | vitals | 11 | SSH3 | protein_coding | rs76335321 | 67097778 | 1.75e-08 | 7 | Female only | Both sexes | Female only |
| R.RT..MALE.ADULT..IDEAL.BODY.WEIGHT | Ideal body weight — male | vitals | 15 | ACAN | protein_coding | rs2280470 | 89395626 | 1.87e-08 | 23 | Female only | Female only | Female only |
| R.RT..MALE.ADULT..IDEAL.BODY.WEIGHT | Ideal body weight — male | vitals | 13 | DLEU1 | protein_coding | rs3116598 | 51099616 | 1.97e-08 | 109 | Both sexes | Both sexes | Female only |
| R.RT..MALE.ADULT..IDEAL.BODY.WEIGHT | Ideal body weight — male | vitals | 7 | IGF2BP3 | protein_coding | rs67397633 | 23491190 | 2.13e-08 | 49 | Male only | Both sexes | Male only |
| R.RT..MALE.ADULT..IDEAL.BODY.WEIGHT | Ideal body weight — male | vitals | 18 | CABLES1 | protein_coding | rs4567840 | 20705158 | 2.14e-08 | 65 | Male only | Both sexes | Male only |
| R.RT..MALE.ADULT..IDEAL.BODY.WEIGHT | Ideal body weight — male | vitals | 2 | EFEMP1 | protein_coding | rs11125609 | 56115834 | 2.15e-08 | 5 | Male only | Male only | Male only |
| R.RT..MALE.ADULT..IDEAL.BODY.WEIGHT | Ideal body weight — male | vitals | 6 | RP11-486E2.1 | pseudogene | rs9359426 | 81442039 | 4.35e-08 | 181 | Female only | Both sexes | Female only |

| lab | full_name | group | CHR | Gene_Symbol | biotype | lead_snp | lead_BP | lead_P | total_snps_in_loci | independent_loci_in_sex_P1e6 | independent_loci_in_sex_P1e2 | independent_loci_in_sex_P5e8 |
|---|---|---|---|---|---|---|---|---|---|---|---|---|
| R.RT..MALE.ADULT..IDEAL.BODY.WEIGHT | Ideal body weight — male | vitals | 2 | EFEMP1 | protein_coding | rs1346790 | 56091871 | 4.88e-08 | 65 | Male only | Male only | Male only |
| RR.INTERVAL | RR interval | cardiovascular | 13 | MTUS2 | protein_coding | rs4768988 | 29546818 | 2.71e-08 | 9 | Female only | Female only | Female only |
| SSAIgG | Sjogrens syndrome-A extractable nuclear IgG Ab [Units/volume] in Serum by Immunoassay | immune/inflammatory | 6 | HLA-DQA1 | protein_coding | rs9272461 | 32605609 | 4.93e-09 | 135 | Female only | Female only | Female only |
| X_percent_SAT | % saturation | other | 6 | HFE | protein_coding | rs144861591 | 26072992 | 3.47e-20 | 19 | Both sexes | Both sexes | Both sexes |
| X_percent_SAT | % saturation | other | 6 | HIST1H2AA | protein_coding | rs116009877 | 25715657 | 2.92e-18 | 33 | Both sexes | Both sexes | Both sexes |
| X_percent_SAT | % saturation | other | 22 | TMPRSS6 | protein_coding | rs2072860 | 37470604 | 3.73e-15 | 9 | Both sexes | Both sexes | Both sexes |
| X_percent_SAT | % saturation | other | 6 | LRRC16A | protein_coding | rs79665799 | 25408104 | 5.00e-15 | 241 | Female only | Both sexes | Female only |
| X_percent_SAT | % saturation | other | 6 | LRRC16A | protein_coding | rs77244967 | 25536591 | 5.40e-14 | 23 | Both sexes | Both sexes | Both sexes |
| X_percent_SAT | % saturation | other | 6 | SCGN | protein_coding | rs72838866 | 25673273 | 4.88e-13 | 19 | Both sexes | Both sexes | Both sexes |
| X_percent_SAT | % saturation | other | 6 | HIST1H4A | protein_coding | rs72834629 | 26023032 | 3.08e-12 | 251 | Both sexes | Both sexes | Both sexes |
| X_percent_SAT | % saturation | other | 6 | LRRC16A | protein_coding | rs116272812 | 25452783 | 4.39e-12 | 17 | Both sexes | Both sexes | Both sexes |
| X_percent_SAT | % saturation | other | 6 | VN1R13P | pseudogene | rs13194491 | 27037080 | 5.69e-11 | 3 | Male only | Both sexes | Male only |
| X_percent_SAT | % saturation | other | 6 | RP3-522P13.1 | pseudogene | rs9393635 | 25265377 | 1.68e-10 | 75 | Female only | Female only | Female only |
| X_percent_SAT | % saturation | other | 6 | LRRC16A | protein_coding | rs73730705 | 25383843 | 2.07e-10 | 27 | Female only | Both sexes | Female only |
| X_percent_SAT | % saturation | other | 6 | HIST1H1T | protein_coding | rs198846 | 26107463 | 5.54e-10 | 33 | Both sexes | Both sexes | Both sexes |
| X_percent_SAT | % saturation | other | 6 | SLC17A1 | protein_coding | rs12191655 | 25803095 | 2.96e-09 | 291 | Both sexes | Both sexes | Female only |
| X_percent_SAT | % saturation | other | 6 | RP11-191A15.4 | pseudogene | rs35191161 | 25230092 | 8.30e-09 | 13 | Female only | Female only | Female only |
| X_percent_SAT | % saturation | other | 6 | HIST1H3G | protein_coding | rs7773004 | 26267755 | 8.38e-09 | 89 | Both sexes | Both sexes | Both sexes |

### STable8: Variants from loci identified as sex specific (P<5e-8 in one sex, all variants within loci with P>0.01 in other sex) in BioVU, with effect estimates for BioVU (VUMC) males and females as well as UKBiobank males and females

4013 rows × 34 columns

STable8 — Columns 1–23 of 34

| trait | lead_snp | ID | ALLELE1 | ALLELE0 | CHR | Gene_Symbol | independent_loci_in_sex_P1e2 | in_embl_any_or_gene | A1FREQ_VUMC_female | A1FREQ_VUMC_male | A1FREQ_UKB_female | A1FREQ_UKB_male | N_VUMC_female | N_VUMC_male | N_UKB_female | N_UKB_male | BETA_VUMC_female | BETA_VUMC_male | BETA_UKB_female | BETA_UKB_male | SE_VUMC_female | SE_VUMC_male |
|---|---|---|---|---|---|---|---|---|---|---|---|---|---|---|---|---|---|---|---|---|---|---|
| Hgb | rs169219 | rs442601 | G | A | 6 | TRIM38 | Female only | True | 0.3038 | 0.2998 | 0.2822 | 0.2837 | 35678 | 26968 | 188076 | 162398 | -0.03491 | -0.004228 | -0.04138 | -0.03837 | 0.007648 | 0.009021 |
| Hgb | rs169219 | rs199736 | C | T | 6 | TRIM38 | Female only | True | 0.3043 | 0.3002 | 0.2827 | 0.2842 | 35678 | 26968 | 188076 | 162398 | -0.0341 | -0.003589 | -0.0413 | -0.03831 | 0.007643 | 0.009015 |
| Hgb | rs169219 | rs2051540 | T | C | 6 | TRIM38 | Female only | True | 0.2989 | 0.2949 | 0.2781 | 0.2799 | 35678 | 26968 | 188076 | 162398 | -0.03289 | -0.003573 | -0.0412 | -0.03848 | 0.007685 | 0.009076 |
| Hgb | rs169219 | rs199726 | G | A | 6 | TRIM38 | Female only | True | 0.3043 | 0.3001 | 0.2826 | 0.2841 | 35678 | 26968 | 188076 | 162398 | -0.03456 | -0.003652 | -0.04146 | -0.03825 | 0.007642 | 0.009016 |
| Hgb | rs169219 | rs199725 | A | G | 6 | TRIM38 | Female only | True | 0.303 | 0.2987 | 0.2829 | 0.2845 | 35678 | 26968 | 188076 | 162398 | -0.03453 | -0.003732 | -0.04147 | -0.03771 | 0.007627 | 0.008999 |
| Hgb | rs169219 | rs175319 | A | G | 6 | TRIM38 | Female only | True | 0.2922 | 0.2886 | 0.273 | 0.2749 | 35678 | 26968 | 188076 | 162398 | -0.03591 | 0.001028 | -0.041 | -0.03844 | 0.007729 | 0.009139 |
| Hgb | rs169219 | rs169219 | C | A | 6 | TRIM38 | Female only | True | 0.4025 | 0.4008 | 0.4024 | 0.4034 | 35678 | 26968 | 188076 | 162398 | -0.04379 | -0.02093 | -0.06358 | -0.05835 | 0.007165 | 0.008448 |
| Hgb | rs169219 | rs199739 | A | C | 6 | TRIM38 | Female only | True | 0.2898 | 0.2865 | 0.2713 | 0.2732 | 35678 | 26968 | 188076 | 162398 | -0.03522 | 0.002294 | -0.04099 | -0.03885 | 0.00776 | 0.009172 |
| Hgb | rs169219 | rs129129 | A | G | 6 | TRIM38 | Female only | True | 0.3043 | 0.3002 | 0.2826 | 0.2841 | 35678 | 26968 | 188076 | 162398 | -0.03483 | -0.003571 | -0.04138 | -0.03823 | 0.007647 | 0.009022 |
| Hgb | rs169219 | rs199755 | G | A | 6 | TRIM38 | Female only | True | 0.2999 | 0.2965 | 0.2814 | 0.2834 | 35678 | 26968 | 188076 | 162398 | -0.03541 | 0.002548 | -0.04167 | -0.03649 | 0.007695 | 0.009069 |
| Hgb | rs169219 | rs199754 | A | G | 6 | TRIM38 | Female only | True | 0.2979 | 0.2947 | 0.2802 | 0.2822 | 35678 | 26968 | 188076 | 162398 | -0.03605 | 0.003635 | -0.0417 | -0.03651 | 0.007715 | 0.009082 |
| Hgb | rs169219 | rs9356995 | G | A | 6 | TRIM38 | Female only | True | 0.3063 | 0.3029 | 0.2865 | 0.2881 | 35678 | 26968 | 188076 | 162398 | -0.03652 | 0.002465 | -0.04164 | -0.03616 | 0.007635 | 0.008993 |
| Hgb | rs169219 | rs6901039 | T | G | 6 | TRIM38 | Female only | True | 0.306 | 0.3027 | 0.2865 | 0.2881 | 35678 | 26968 | 188076 | 162398 | -0.03629 | 0.002609 | -0.04162 | -0.03616 | 0.007637 | 0.008996 |
| Hgb | rs169219 | rs199753 | G | A | 6 | TRIM38 | Female only | True | 0.2978 | 0.2947 | 0.2806 | 0.2825 | 35678 | 26968 | 188076 | 162398 | -0.03686 | 0.003582 | -0.04187 | -0.03657 | 0.007708 | 0.009073 |
| Hgb | rs169219 | rs9356996 | C | A | 6 | TRIM38 | Female only | True | 0.4176 | 0.4161 | 0.4182 | 0.4193 | 35678 | 26968 | 188076 | 162398 | -0.04327 | -0.02011 | -0.0647 | -0.0575 | 0.007141 | 0.008398 |
| Hgb | rs169219 | rs6942072 | A | G | 6 | TRIM38 | Female only | True | 0.3102 | 0.3066 | 0.2906 | 0.2922 | 35678 | 26968 | 188076 | 162398 | -0.03481 | 2.65e-04 | -0.04239 | -0.03638 | 0.007593 | 0.008949 |
| Hgb | rs169219 | rs1971508 | A | C | 6 | TRIM38 | Female only | True | 0.4518 | 0.4491 | 0.4545 | 0.4555 | 35678 | 26968 | 188076 | 162398 | -0.03849 | -0.01767 | -0.053 | -0.04762 | 0.007063 | 0.008304 |
| Hgb | rs9607412 | rs2111833 | T | C | 22 | TMPRSS6 | Female only | True | 0.3367 | 0.3416 | 0.3461 | 0.3453 | 35678 | 26968 | 188076 | 162398 | 0.0356 | 0.01655 | 0.05146 | 0.04888 | 0.007419 | 0.008741 |
| Hgb | rs9607412 | rs9607412 | C | T | 22 | TMPRSS6 | Female only | True | 0.4091 | 0.411 | 0.4189 | 0.4184 | 35678 | 26968 | 188076 | 162398 | 0.04396 | 0.01815 | 0.05823 | 0.05855 | 0.007182 | 0.008473 |
| Hgb | rs9607412 | rs9610646 | G | T | 22 | TMPRSS6 | Female only | True | 0.3772 | 0.3818 | 0.3897 | 0.389 | 35678 | 26968 | 188076 | 162398 | 0.03857 | 0.01589 | 0.05136 | 0.05333 | 0.007274 | 0.008577 |
| Hgb | rs9607412 | rs9610647 | A | G | 22 | TMPRSS6 | Female only | True | 0.3774 | 0.3823 | 0.3897 | 0.389 | 35678 | 26968 | 188076 | 162398 | 0.03881 | 0.01536 | 0.05124 | 0.05325 | 0.007265 | 0.008563 |
| Hgb | rs9607412 | rs2235323 | C | T | 22 | TMPRSS6 | Female only | True | 0.378 | 0.3828 | 0.3898 | 0.3891 | 35678 | 26968 | 188076 | 162398 | 0.03864 | 0.01481 | 0.05119 | 0.05315 | 0.007262 | 0.008557 |
| Hgb | rs9607412 | rs2235324 | C | T | 22 | TMPRSS6 | Female only | True | 0.3797 | 0.3845 | 0.3897 | 0.3889 | 35678 | 26968 | 188076 | 162398 | 0.03838 | 0.014 | 0.051 | 0.05309 | 0.007229 | 0.008517 |
| Hgb | rs9607412 | rs736852 | C | T | 22 | TMPRSS6 | Female only | True | 0.4106 | 0.4126 | 0.4186 | 0.4179 | 35678 | 26968 | 188076 | 162398 | 0.04327 | 0.01668 | 0.05783 | 0.05832 | 0.007141 | 0.008422 |
| Hgb | rs9607412 | rs738295 | G | A | 22 | TMPRSS6 | Female only | True | 0.3774 | 0.3822 | 0.3896 | 0.3889 | 35678 | 26968 | 188076 | 162398 | 0.03855 | 0.01431 | 0.05102 | 0.053 | 0.007265 | 0.008564 |

| trait | lead_snp | ID | ALLELE1 | ALLELE0 | CHR | Gene_Symbol | independent_loci_in_sex_P1e2 | in_embl_any_or_gene | A1FREQ_VUMC_female | A1FREQ_VUMC_male | A1FREQ_UKB_female | A1FREQ_UKB_male | N_VUMC_female | N_VUMC_male | N_UKB_female | N_UKB_male | BETA_VUMC_female | BETA_VUMC_male | BETA_UKB_female | BETA_UKB_male | SE_VUMC_female | SE_VUMC_male |
|---|---|---|---|---|---|---|---|---|---|---|---|---|---|---|---|---|---|---|---|---|---|---|
| Hgb | rs9607412 | rs4328728 | T | C | 22 | TMPRSS6 | Female only | True | 0.4085 | 0.4105 | 0.4182 | 0.4177 | 35678 | 26968 | 188076 | 162398 | 0.043 | 0.01633 | 0.05778 | 0.05826 | 0.007174 | 0.008462 |
| Hgb | rs9607412 | rs5756508 | G | A | 22 | TMPRSS6 | Female only | True | 0.4079 | 0.41 | 0.4181 | 0.4176 | 35678 | 26968 | 188076 | 162398 | 0.04305 | 0.01688 | 0.05782 | 0.0583 | 0.007176 | 0.008466 |
| Hgb | rs9607412 | rs5756509 | A | C | 22 | TMPRSS6 | Female only | True | 0.4079 | 0.41 | 0.4182 | 0.4176 | 35678 | 26968 | 188076 | 162398 | 0.04306 | 0.01694 | 0.05778 | 0.05832 | 0.007176 | 0.008467 |
| Hgb | rs9607412 | rs1421312 | G | A | 22 | TMPRSS6 | Female only | True | 0.384 | 0.388 | 0.3938 | 0.3934 | 35678 | 26968 | 188076 | 162398 | 0.03939 | 0.01691 | 0.05146 | 0.05367 | 0.007251 | 0.008542 |
| Hgb | rs9607412 | rs2743822 | T | C | 22 | TMPRSS6 | Female only | True | 0.4059 | 0.4083 | 0.4172 | 0.4166 | 35678 | 26968 | 188076 | 162398 | 0.04306 | 0.01587 | 0.05768 | 0.0579 | 0.007181 | 0.008469 |
| Hgb | rs9607412 | rs2543515 | G | A | 22 | TMPRSS6 | Female only | True | 0.4061 | 0.4085 | 0.4175 | 0.4169 | 35678 | 26968 | 188076 | 162398 | 0.04312 | 0.01604 | 0.05773 | 0.05805 | 0.007181 | 0.008469 |
| Hgb | rs9607412 | rs5750378 | T | C | 22 | TMPRSS6 | Female only | True | 0.4059 | 0.4083 | 0.4173 | 0.4166 | 35678 | 26968 | 188076 | 162398 | 0.0431 | 0.01592 | 0.05777 | 0.0578 | 0.00718 | 0.008467 |
| Hgb | rs9607412 | rs12172465 | T | G | 22 | TMPRSS6 | Female only | True | 0.4059 | 0.4083 | 0.4172 | 0.4166 | 35678 | 26968 | 188076 | 162398 | 0.0431 | 0.01591 | 0.05775 | 0.0578 | 0.00718 | 0.008467 |
| Hgb | rs9607412 | rs6000567 | T | C | 22 | TMPRSS6 | Female only | True | 0.407 | 0.4093 | 0.4176 | 0.417 | 35678 | 26968 | 188076 | 162398 | 0.04387 | 0.01653 | 0.05775 | 0.05777 | 0.007179 | 0.008467 |
| Hgb | rs9607412 | rs5750379 | G | A | 22 | TMPRSS6 | Female only | True | 0.4122 | 0.4146 | 0.4242 | 0.4238 | 35678 | 26968 | 188076 | 162398 | 0.04141 | 0.0146 | 0.05514 | 0.05638 | 0.007191 | 0.008486 |
| Hgb | rs9607412 | rs5750380 | G | A | 22 | TMPRSS6 | Female only | True | 0.4059 | 0.4083 | 0.4173 | 0.4166 | 35678 | 26968 | 188076 | 162398 | 0.04311 | 0.01591 | 0.05778 | 0.05782 | 0.00718 | 0.008467 |
| Hgb | rs9607412 | rs2543513 | G | A | 22 | TMPRSS6 | Female only | True | 0.4059 | 0.4083 | 0.4173 | 0.4166 | 35678 | 26968 | 188076 | 162398 | 0.0431 | 0.01591 | 0.05774 | 0.05789 | 0.00718 | 0.008467 |
| Hgb | rs9607412 | rs2413451 | G | A | 22 | TMPRSS6 | Female only | True | 0.4078 | 0.4102 | 0.4172 | 0.4165 | 35678 | 26968 | 188076 | 162398 | 0.04282 | 0.01595 | 0.05775 | 0.05793 | 0.007145 | 0.008428 |
| Hgb | rs9607412 | rs2251655 | T | C | 22 | TMPRSS6 | Female only | True | 0.4058 | 0.4082 | 0.4172 | 0.4165 | 35678 | 26968 | 188076 | 162398 | 0.04302 | 0.01599 | 0.05774 | 0.05792 | 0.00718 | 0.008468 |
| Hgb | rs9607412 | rs2543511 | G | A | 22 | TMPRSS6 | Female only | True | 0.4059 | 0.4083 | 0.4172 | 0.4166 | 35678 | 26968 | 188076 | 162398 | 0.04308 | 0.01592 | 0.05779 | 0.05783 | 0.00718 | 0.008467 |
| Hgb | rs9607412 | rs5756511 | T | C | 22 | TMPRSS6 | Female only | True | 0.3805 | 0.3849 | 0.3924 | 0.392 | 35678 | 26968 | 188076 | 162398 | 0.04027 | 0.01728 | 0.05178 | 0.05324 | 0.007262 | 0.008558 |
| Hgb | rs9607412 | rs2743824 | C | T | 22 | TMPRSS6 | Female only | True | 0.4048 | 0.4073 | 0.4164 | 0.4156 | 35678 | 26968 | 188076 | 162398 | 0.04326 | 0.01633 | 0.05761 | 0.05755 | 0.007185 | 0.008474 |
| Hgb | rs9607412 | rs2543510 | A | G | 22 | TMPRSS6 | Female only | True | 0.3814 | 0.3857 | 0.3926 | 0.3921 | 35678 | 26968 | 188076 | 162398 | 0.03955 | 0.01643 | 0.05171 | 0.05337 | 0.007261 | 0.008552 |
| Hgb | rs9607412 | rs2543509 | G | A | 22 | TMPRSS6 | Female only | True | 0.406 | 0.4086 | 0.4177 | 0.417 | 35678 | 26968 | 188076 | 162398 | 0.0435 | 0.01642 | 0.05728 | 0.05722 | 0.007187 | 0.008474 |
| Hgb | rs9607412 | rs2743825 | G | T | 22 | TMPRSS6 | Female only | True | 0.4061 | 0.4086 | 0.4178 | 0.4171 | 35678 | 26968 | 188076 | 162398 | 0.04352 | 0.0164 | 0.05714 | 0.05728 | 0.007187 | 0.008474 |
| Hgb | rs9607412 | rs9619658 | T | C | 22 | TMPRSS6 | Female only | True | 0.3312 | 0.3312 | 0.3385 | 0.3377 | 35678 | 26968 | 188076 | 162398 | 0.03952 | 0.01289 | 0.04817 | 0.05108 | 0.007476 | 0.008827 |
| MCH | 2:112208126 | 2:112208126 | A | G | 2 | MIR4435-1HG | Female only | True | 0.07305 | 0.07149 | | | 35668 | 26968 | | | 0.08014 | 0.02859 | | | 0.01284 | 0.01583 |
| MCH | 2:112208126 | rs1900710 | C | T | 2 | MIR4435-1HG | Female only | True | 0.07305 | 0.07166 | | | 35668 | 26968 | | | 0.07966 | 0.0284 | | | 0.01283 | 0.0158 |
| MCH | rs9871110 | rs881753 | A | G | 3 | TNK2 | Male only | False | 0.4696 | 0.472 | 0.4748 | 0.4758 | 35668 | 26968 | 188074 | 162398 | -0.01524 | -0.04501 | -0.02607 | -0.03486 | 0.006677 | 0.008167 |
| MCH | rs9871110 | rs3747672 | T | C | 3 | TNK2 | Male only | False | 0.4694 | 0.4717 | 0.4741 | 0.4752 | 35668 | 26968 | 188074 | 162398 | -0.01403 | -0.04597 | -0.02585 | -0.03525 | 0.006709 | 0.008209 |
| MCH | rs9871110 | rs9829709 | G | A | 3 | TNK2 | Male only | False | 0.4801 | 0.4831 | 0.4858 | 0.4873 | 35668 | 26968 | 188074 | 162398 | -0.01415 | -0.04587 | -0.02793 | -0.03553 | 0.006699 | 0.008199 |
| MCH | rs9871110 | rs9871110 | A | G | 3 | TNK2 | Male only | False | 0.4547 | 0.4579 | 0.4603 | 0.4612 | 35668 | 26968 | 188074 | 162398 | -0.0105 | -0.04665 | -0.0298 | -0.03683 | 0.006692 | 0.008167 |
| MCH | rs12197304 | rs6456715 | C | T | 6 | BTN1A1 | Female only | True | 0.1195 | 0.119 | 0.1112 | 0.1112 | 35668 | 26968 | 188074 | 162398 | -0.04667 | -0.01603 | -0.01484 | -0.02203 | 0.0105 | 0.01282 |
| MCH | rs12197304 | rs6456716 | G | A | 6 | BTN1A1 | Female only | True | 0.1195 | 0.119 | 0.1112 | 0.1112 | 35668 | 26968 | 188074 | 162398 | -0.04658 | -0.01588 | -0.01474 | -0.02193 | 0.0105 | 0.01282 |

| trait | lead_snp | ID | ALLELE1 | ALLELE0 | CHR | Gene_Symbol | independent_loci_in_sex_P1e2 | in_embl_any_or_gene | A1FREQ_VUMC_female | A1FREQ_VUMC_male | A1FREQ_UKB_female | A1FREQ_UKB_male | N_VUMC_female | N_VUMC_male | N_UKB_female | N_UKB_male | BETA_VUMC_female | BETA_VUMC_male | BETA_UKB_female | BETA_UKB_male | SE_VUMC_female | SE_VUMC_male |
|---|---|---|---|---|---|---|---|---|---|---|---|---|---|---|---|---|---|---|---|---|---|---|
| MCH | rs12197304 | rs9461245 | A | G | 6 | BTN1A1 | Female only | True | 0.1156 | 0.1153 | 0.1107 | 0.1107 | 35668 | 26968 | 188074 | 162398 | -0.04926 | -0.01636 | -0.01469 | -0.02225 | 0.01056 | 0.01294 |
| MCH | rs12197304 | rs72836479 | A | G | 6 | BTN1A1 | Female only | True | 0.1171 | 0.1169 | 0.1115 | 0.1114 | 35668 | 26968 | 188074 | 162398 | -0.04929 | -0.01563 | -0.01495 | -0.02215 | 0.01049 | 0.01284 |
| MCH | rs12197304 | rs116371893 | A | G | 6 | BTN1A1 | Female only | True | 0.1227 | 0.1224 | 0.1166 | 0.1163 | 35668 | 26968 | 188074 | 162398 | -0.04599 | -0.02138 | -0.01483 | -0.01816 | 0.01028 | 0.01258 |
| MCH | rs12197304 | rs12203176 | C | T | 6 | BTN1A1 | Female only | True | 0.117 | 0.1169 | 0.1115 | 0.1114 | 35668 | 26968 | 188074 | 162398 | -0.04845 | -0.01591 | -0.01495 | -0.02222 | 0.01048 | 0.01282 |
| MCH | rs12197304 | rs9688949 | T | C | 6 | BTN1A1 | Female only | True | 0.1225 | 0.1222 | 0.1165 | 0.1162 | 35668 | 26968 | 188074 | 162398 | -0.04533 | -0.02161 | -0.01473 | -0.01806 | 0.01028 | 0.01258 |
| MCH | rs12197304 | rs72841510 | A | G | 6 | BTN1A1 | Female only | True | 0.1169 | 0.1168 | 0.1114 | 0.1114 | 35668 | 26968 | 188074 | 162398 | -0.04855 | -0.0158 | -0.01496 | -0.02232 | 0.01049 | 0.01283 |
| trait | lead_snp | ID | ALLELE1 | ALLELE0 | CHR | Gene_Symbol | independent_loci_in_sex_P1e2 | in_embl_any_or_gene | A1FREQ_VUMC_female | A1FREQ_VUMC_male | A1FREQ_UKB_female | A1FREQ_UKB_male | N_VUMC_female | N_VUMC_male | N_UKB_female | N_UKB_male | BETA_VUMC_female | BETA_VUMC_male | BETA_UKB_female | BETA_UKB_male | SE_VUMC_female | SE_VUMC_male |
| MCH | rs12197304 | rs35183513 | A | G | 6 | BTN1A1 | Female only | True | 0.117 | 0.1168 | 0.1114 | 0.1114 | 35668 | 26968 | 188074 | 162398 | -0.04845 | -0.01568 | -0.01486 | -0.02249 | 0.01048 | 0.01282 |
| MCH | rs12197304 | rs55827419 | C | T | 6 | BTN1A1 | Female only | True | 0.1225 | 0.1222 | 0.1166 | 0.1162 | 35668 | 26968 | 188074 | 162398 | -0.0453 | -0.02156 | -0.01476 | -0.01802 | 0.01028 | 0.01258 |
| MCH | rs12197304 | rs56380525 | A | G | 6 | BTN1A1 | Female only | True | 0.1302 | 0.1292 | 0.1238 | 0.1231 | 35668 | 26968 | 188074 | 162398 | -0.04225 | -0.02003 | -0.01572 | -0.01716 | 0.01001 | 0.01227 |
| MCH | rs12197304 | rs12206621 | A | G | 6 | BTN1A1 | Female only | True | 0.1302 | 0.1292 | 0.1238 | 0.1231 | 35668 | 26968 | 188074 | 162398 | -0.04196 | -0.01978 | -0.0156 | -0.01729 | 0.01002 | 0.01227 |
| MCH | rs12197304 | rs9104 | A | G | 6 | BTN1A1 | Female only | True | 0.1292 | 0.1283 | 0.122 | 0.1222 | 35668 | 26968 | 188074 | 162398 | -0.05069 | -0.02272 | -0.01681 | -0.02608 | 0.01005 | 0.01236 |
| MCH | rs12197304 | rs72841540 | A | G | 6 | BTN1A1 | Female only | True | 0.1211 | 0.1203 | 0.1133 | 0.1134 | 35668 | 26968 | 188074 | 162398 | -0.04876 | -0.01987 | -0.01726 | -0.02416 | 0.01037 | 0.0127 |
| MCH | rs12197304 | rs12664899 | T | C | 6 | BTN1A1 | Female only | True | 0.1227 | 0.1215 | 0.1148 | 0.1147 | 35668 | 26968 | 188074 | 162398 | -0.04861 | -0.02097 | -0.01604 | -0.026 | 0.01028 | 0.01263 |
| MCH | rs12197304 | rs41266835 | T | C | 6 | BTN1A1 | Female only | True | 0.1229 | 0.1217 | 0.1149 | 0.1148 | 35668 | 26968 | 188074 | 162398 | -0.04797 | -0.0206 | -0.01583 | -0.02572 | 0.01025 | 0.01259 |
| MCH | rs12197304 | rs17537465 | A | C | 6 | BTN1A1 | Female only | True | 0.1229 | 0.1217 | 0.1149 | 0.1148 | 35668 | 26968 | 188074 | 162398 | -0.04798 | -0.02064 | -0.01591 | -0.02575 | 0.01025 | 0.01259 |
| MCH | rs12197304 | rs1131936 | G | A | 6 | BTN1A1 | Female only | True | 0.1455 | 0.1431 | 0.1298 | 0.1295 | 35668 | 26968 | 188074 | 162398 | -0.04203 | -0.0232 | -0.01542 | -0.02386 | 0.009563 | 0.01174 |
| MCH | rs12197304 | rs4712986 | G | A | 6 | BTN1A1 | Female only | True | 0.1474 | 0.1444 | 0.1287 | 0.1284 | 35668 | 26968 | 188074 | 162398 | -0.03856 | -0.02562 | -0.01561 | -0.02337 | 0.00954 | 0.01172 |
| MCH | rs12197304 | rs7356849 | C | T | 6 | BTN1A1 | Female only | True | 0.144 | 0.1416 | 0.1286 | 0.1284 | 35668 | 26968 | 188074 | 162398 | -0.04211 | -0.02556 | -0.0157 | -0.02346 | 0.009599 | 0.01179 |
| MCH | rs12197304 | rs12662604 | A | G | 6 | BTN1A1 | Female only | True | 0.1229 | 0.1217 | 0.1149 | 0.1148 | 35668 | 26968 | 188074 | 162398 | -0.04865 | -0.02029 | -0.01572 | -0.02564 | 0.01025 | 0.01259 |
| MCH | rs12197304 | rs9467751 | C | T | 6 | BTN1A1 | Female only | True | 0.144 | 0.1416 | 0.1285 | 0.1283 | 35668 | 26968 | 188074 | 162398 | -0.04204 | -0.02533 | -0.01574 | -0.02311 | 0.009602 | 0.0118 |
| MCH | rs12197304 | rs4712989 | C | T | 6 | BTN1A1 | Female only | True | 0.1227 | 0.1215 | 0.1148 | 0.1147 | 35668 | 26968 | 188074 | 162398 | -0.04792 | -0.02059 | -0.0159 | -0.02523 | 0.01028 | 0.01262 |
| MCH | rs12197304 | rs3902051 | G | A | 6 | BTN1A1 | Female only | True | 0.1229 | 0.1217 | 0.115 | 0.1149 | 35668 | 26968 | 188074 | 162398 | -0.04724 | -0.01994 | -0.01614 | -0.02525 | 0.01027 | 0.01261 |
| MCH | rs12197304 | rs4711108 | T | C | 6 | BTN1A1 | Female only | True | 0.1169 | 0.1159 | 0.1087 | 0.1085 | 35668 | 26968 | 188074 | 162398 | -0.05324 | -0.01563 | -0.0172 | -0.0286 | 0.01052 | 0.01292 |
| MCH | rs12197304 | rs56161420 | A | G | 6 | BTN1A1 | Female only | True | 0.1594 | 0.1588 | 0.1495 | 0.1498 | 35668 | 26968 | 188074 | 162398 | -0.02913 | 0.001741 | 0.00232 | 0.002231 | 0.009222 | 0.01128 |
| MCH | rs12197304 | rs17610161 | A | G | 6 | BTN1A1 | Female only | True | 0.1155 | 0.1145 | 0.1066 | 0.1064 | 35668 | 26968 | 188074 | 162398 | -0.05387 | -0.01782 | -0.01671 | -0.02806 | 0.01058 | 0.01299 |
| MCH | rs12197304 | rs60750900 | G | A | 6 | BTN1A1 | Female only | True | 0.1587 | 0.1578 | 0.1474 | 0.1475 | 35668 | 26968 | 188074 | 162398 | -0.03001 | 0.002894 | 0.004411 | 0.002977 | 0.009241 | 0.01133 |
| MCH | rs12197304 | rs4609015 | C | T | 6 | BTN1A1 | Female only | True | 0.1589 | 0.1582 | 0.1474 | 0.1475 | 35668 | 26968 | 188074 | 162398 | -0.03014 | 0.00241 | 0.004515 | 0.002869 | 0.009237 | 0.01133 |

| trait | lead_snp | ID | ALLELE1 | ALLELE0 | CHR | Gene_Symbol | independent_loci_in_sex_P1e2 | in_embl_any_or_gene | A1FREQ_VUMC_female | A1FREQ_VUMC_male | A1FREQ_UKB_female | A1FREQ_UKB_male | N_VUMC_female | N_VUMC_male | N_UKB_female | N_UKB_male | BETA_VUMC_female | BETA_VUMC_male | BETA_UKB_female | BETA_UKB_male | SE_VUMC_female | SE_VUMC_male |
|---|---|---|---|---|---|---|---|---|---|---|---|---|---|---|---|---|---|---|---|---|---|---|
| MCH | rs12197304 | rs55957716 | C | T | 6 | BTN1A1 | Female only | True | 0.1587 | 0.1579 | 0.1474 | 0.1475 | 35668 | 26968 | 188074 | 162398 | -0.02969 | 0.001759 | 0.004239 | 0.002925 | 0.009237 | 0.01133 |
| MCH | rs12197304 | rs7770214 | A | G | 6 | BTN1A1 | Female only | True | 0.1587 | 0.1579 | 0.1474 | 0.1475 | 35668 | 26968 | 188074 | 162398 | -0.02971 | 0.001747 | 0.004265 | 0.002934 | 0.009237 | 0.01133 |
| MCH | rs12197304 | rs57275245 | G | A | 6 | BTN1A1 | Female only | True | 0.1583 | 0.1576 | 0.1469 | 0.147 | 35668 | 26968 | 188074 | 162398 | -0.02975 | 0.002056 | 0.004186 | 0.002887 | 0.009255 | 0.01135 |
| MCH | rs12197304 | rs12214486 | A | G | 6 | BTN1A1 | Female only | True | 0.1587 | 0.1579 | 0.1474 | 0.1475 | 35668 | 26968 | 188074 | 162398 | -0.02971 | 0.001748 | 0.004243 | 0.002929 | 0.009237 | 0.01133 |
| MCH | rs12197304 | rs10946820 | C | T | 6 | BTN1A1 | Female only | True | 0.1587 | 0.1579 | 0.1474 | 0.1475 | 35668 | 26968 | 188074 | 162398 | -0.02971 | 0.001751 | 0.00431 | 0.002918 | 0.009237 | 0.01133 |
| MCH | rs12197304 | rs10946821 | T | C | 6 | BTN1A1 | Female only | True | 0.1586 | 0.1579 | 0.1474 | 0.1475 | 35668 | 26968 | 188074 | 162398 | -0.02957 | 0.001443 | 0.004243 | 0.002929 | 0.009232 | 0.01133 |
| MCH | rs12197304 | rs12214924 | C | T | 6 | BTN1A1 | Female only | True | 0.1586 | 0.1578 | 0.1474 | 0.1475 | 35668 | 26968 | 188074 | 162398 | -0.02961 | 0.001442 | 0.004155 | 0.002764 | 0.009232 | 0.01133 |
| MCH | rs12197304 | rs28571747 | C | T | 6 | BTN1A1 | Female only | True | 0.1586 | 0.1578 | 0.1474 | 0.1474 | 35668 | 26968 | 188074 | 162398 | -0.02961 | 0.001442 | 0.004293 | 0.003021 | 0.009232 | 0.01133 |
| MCH | rs12197304 | rs12194412 | G | A | 6 | BTN1A1 | Female only | True | 0.1586 | 0.1578 | 0.1474 | 0.1474 | 35668 | 26968 | 188074 | 162398 | -0.02961 | 0.001442 | 0.004295 | 0.00302 | 0.009232 | 0.01133 |
| MCH | rs12197304 | rs12207930 | T | C | 6 | BTN1A1 | Female only | True | 0.1586 | 0.1579 | 0.1473 | 0.1475 | 35668 | 26968 | 188074 | 162398 | -0.02959 | 0.001451 | 0.004413 | 0.003219 | 0.009232 | 0.01132 |
| MCH | rs12197304 | rs6900725 | C | T | 6 | BTN1A1 | Female only | True | 0.1586 | 0.1579 | 0.1475 | 0.1475 | 35668 | 26968 | 188074 | 162398 | -0.02959 | 0.001451 | 0.004095 | 0.002991 | 0.009232 | 0.01132 |
| MCH | rs12197304 | rs6920986 | T | C | 6 | BTN1A1 | Female only | True | 0.1586 | 0.1579 | 0.1473 | 0.1474 | 35668 | 26968 | 188074 | 162398 | -0.02957 | 0.001381 | 0.004257 | 0.002963 | 0.00923 | 0.01132 |
| MCH | rs12197304 | rs6901245 | G | T | 6 | BTN1A1 | Female only | True | 0.1586 | 0.1579 | 0.1473 | 0.1474 | 35668 | 26968 | 188074 | 162398 | -0.02957 | 0.001381 | 0.004295 | 0.002956 | 0.00923 | 0.01132 |
| MCH | rs12197304 | rs4712990 | A | C | 6 | BTN1A1 | Female only | True | 0.1175 | 0.1161 | 0.1094 | 0.1091 | 35668 | 26968 | 188074 | 162398 | -0.05152 | -0.01689 | -0.01632 | -0.02848 | 0.01048 | 0.01289 |
| MCH | rs12197304 | rs56190518 | T | C | 6 | BTN1A1 | Female only | True | 0.1585 | 0.1578 | 0.1473 | 0.1474 | 35668 | 26968 | 188074 | 162398 | -0.02972 | 9.64e-04 | 0.004494 | 0.002893 | 0.009235 | 0.01133 |
| MCH | rs12197304 | rs56325223 | T | G | 6 | BTN1A1 | Female only | True | 0.1158 | 0.1139 | 0.1075 | 0.1074 | 35668 | 26968 | 188074 | 162398 | -0.05022 | -0.01224 | -0.01728 | -0.02838 | 0.01056 | 0.01301 |
| MCH | rs12197304 | rs56143200 | C | T | 6 | BTN1A1 | Female only | True | 0.1585 | 0.1578 | 0.1473 | 0.1474 | 35668 | 26968 | 188074 | 162398 | -0.02972 | 0.001004 | 0.004381 | 0.0029 | 0.009235 | 0.01133 |
| MCH | rs12197304 | rs3208734 | T | C | 6 | BTN1A1 | Female only | True | 0.1585 | 0.1578 | 0.1473 | 0.1474 | 35668 | 26968 | 188074 | 162398 | -0.02972 | 0.001004 | 0.004381 | 0.0029 | 0.009235 | 0.01133 |
| MCH | rs12197304 | rs4712992 | G | A | 6 | BTN1A1 | Female only | True | 0.1584 | 0.1577 | 0.1473 | 0.1474 | 35668 | 26968 | 188074 | 162398 | -0.02948 | 9.30e-04 | 0.004399 | 0.002922 | 0.009235 | 0.01133 |
| MCH | rs12197304 | rs12213625 | A | C | 6 | BTN1A1 | Female only | True | 0.158 | 0.1572 | 0.1469 | 0.147 | 35668 | 26968 | 188074 | 162398 | -0.03008 | 0.001236 | 0.003973 | 0.002046 | 0.009244 | 0.01134 |
| MCH | rs12197304 | rs4711109 | A | G | 6 | BTN1A1 | Female only | True | 0.158 | 0.1572 | 0.1469 | 0.147 | 35668 | 26968 | 188074 | 162398 | -0.03008 | 0.001235 | 0.003918 | 0.002089 | 0.009244 | 0.01134 |
| MCH | rs12197304 | rs4712993 | T | C | 6 | BTN1A1 | Female only | True | 0.1581 | 0.1573 | 0.1469 | 0.147 | 35668 | 26968 | 188074 | 162398 | -0.03013 | 0.001174 | 0.003928 | 0.002016 | 0.009236 | 0.01134 |
| MCH | rs12197304 | rs10946823 | A | C | 6 | BTN1A1 | Female only | True | 0.158 | 0.1572 | 0.1469 | 0.147 | 35668 | 26968 | 188074 | 162398 | -0.03017 | 0.001174 | 0.003918 | 0.002089 | 0.009243 | 0.01134 |
| MCH | rs12197304 | rs10946825 | A | G | 6 | BTN1A1 | Female only | True | 0.158 | 0.1572 | 0.1469 | 0.147 | 35668 | 26968 | 188074 | 162398 | -0.03017 | 0.001176 | 0.003901 | 0.002079 | 0.009243 | 0.01134 |
| MCH | rs12197304 | rs10946826 | G | T | 6 | BTN1A1 | Female only | True | 0.158 | 0.1572 | 0.1469 | 0.147 | 35668 | 26968 | 188074 | 162398 | -0.03017 | 0.001174 | 0.00393 | 0.002126 | 0.009243 | 0.01134 |
| MCH | rs12197304 | rs34268421 | C | A | 6 | BTN1A1 | Female only | True | 0.158 | 0.1572 | 0.1469 | 0.147 | 35668 | 26968 | 188074 | 162398 | -0.03017 | 0.001174 | 0.003915 | 0.00209 | 0.009243 | 0.01134 |
| MCH | rs12197304 | rs4305717 | A | C | 6 | BTN1A1 | Female only | True | 0.158 | 0.1572 | 0.1469 | 0.147 | 35668 | 26968 | 188074 | 162398 | -0.03019 | 0.001164 | 0.003918 | 0.002089 | 0.009243 | 0.01134 |
| MCH | rs12197304 | rs56139485 | T | C | 6 | BTN1A1 | Female only | True | 0.1582 | 0.1573 | 0.147 | 0.1471 | 35668 | 26968 | 188074 | 162398 | -0.03032 | 5.24e-04 | 0.003752 | 0.002082 | 0.00924 | 0.01134 |
| MCH | rs12197304 | rs56207464 | A | C | 6 | BTN1A1 | Female only | True | 0.158 | 0.1572 | 0.1469 | 0.147 | 35668 | 26968 | 188074 | 162398 | -0.03018 | 0.001181 | 0.003903 | 0.002072 | 0.009244 | 0.01134 |

| trait | lead_snp | ID | ALLELE1 | ALLELE0 | CHR | Gene_Symbol | independent_loci_in_sex_P1e2 | in_embl_any_or_gene | A1FREQ_VUMC_female | A1FREQ_VUMC_male | A1FREQ_UKB_female | A1FREQ_UKB_male | N_VUMC_female | N_VUMC_male | N_UKB_female | N_UKB_male | BETA_VUMC_female | BETA_VUMC_male | BETA_UKB_female | BETA_UKB_male | SE_VUMC_female | SE_VUMC_male |
|---|---|---|---|---|---|---|---|---|---|---|---|---|---|---|---|---|---|---|---|---|---|---|
| MCH | rs12197304 | rs12196371 | T | C | 6 | BTN1A1 | Female only | True | 0.1161 | 0.1147 | 0.1082 | 0.1079 | 35668 | 26968 | 188074 | 162398 | -0.05411 | -0.01685 | -0.01755 | -0.03037 | 0.01053 | 0.01296 |
| MCH | rs12197304 | rs10447391 | G | T | 6 | BTN1A1 | Female only | True | 0.158 | 0.1572 | 0.1469 | 0.147 | 35668 | 26968 | 188074 | 162398 | -0.03006 | 0.001301 | 0.003915 | 0.002097 | 0.009244 | 0.01134 |
| MCH | rs12197304 | rs7744254 | C | T | 6 | BTN1A1 | Female only | True | 0.1582 | 0.1573 | 0.1471 | 0.1472 | 35668 | 26968 | 188074 | 162398 | -0.03048 | 0.001139 | 0.003827 | 0.001674 | 0.009242 | 0.01134 |
| MCH | rs12197304 | rs6456721 | G | A | 6 | BTN1A1 | Female only | True | 0.1582 | 0.1573 | 0.1469 | 0.147 | 35668 | 26968 | 188074 | 162398 | -0.03048 | 0.001139 | 0.003946 | 0.00201 | 0.009242 | 0.01134 |
| MCH | rs12197304 | rs6456722 | A | C | 6 | BTN1A1 | Female only | True | 0.1582 | 0.1573 | 0.1469 | 0.147 | 35668 | 26968 | 188074 | 162398 | -0.03048 | 0.001139 | 0.003946 | 0.00201 | 0.009242 | 0.01134 |
| MCH | rs12197304 | rs6926878 | G | A | 6 | BTN1A1 | Female only | True | 0.1582 | 0.1573 | 0.1469 | 0.147 | 35668 | 26968 | 188074 | 162398 | -0.03048 | 0.001139 | 0.003946 | 0.00201 | 0.009242 | 0.01134 |
| MCH | rs12197304 | rs3903661 | G | A | 6 | BTN1A1 | Female only | True | 0.1579 | 0.1571 | 0.1468 | 0.147 | 35668 | 26968 | 188074 | 162398 | -0.03091 | 8.04e-04 | 0.004038 | 0.001969 | 0.009249 | 0.01134 |
| MCH | rs12197304 | rs55786210 | G | A | 6 | BTN1A1 | Female only | True | 0.1574 | 0.1565 | 0.1459 | 0.146 | 35668 | 26968 | 188074 | 162398 | -0.03083 | 0.001754 | 0.004679 | 0.002978 | 0.009257 | 0.01137 |
| MCH | rs12197304 | rs56182232 | T | G | 6 | BTN1A1 | Female only | True | 0.1574 | 0.1565 | 0.1459 | 0.146 | 35668 | 26968 | 188074 | 162398 | -0.03083 | 0.001755 | 0.004682 | 0.002987 | 0.009257 | 0.01137 |
| MCH | rs12197304 | rs55994013 | C | T | 6 | BTN1A1 | Female only | True | 0.1168 | 0.1152 | 0.1079 | 0.1075 | 35668 | 26968 | 188074 | 162398 | -0.054 | -0.01528 | -0.01702 | -0.02978 | 0.01051 | 0.01295 |
| MCH | rs12197304 | rs55775018 | T | C | 6 | BTN1A1 | Female only | True | 0.1574 | 0.1565 | 0.1459 | 0.146 | 35668 | 26968 | 188074 | 162398 | -0.03083 | 0.001755 | 0.004689 | 0.002987 | 0.009257 | 0.01137 |
| MCH | rs12197304 | rs111293349 | C | T | 6 | BTN1A1 | Female only | True | 0.1574 | 0.1565 | 0.1459 | 0.146 | 35668 | 26968 | 188074 | 162398 | -0.03083 | 0.001755 | 0.004682 | 0.002987 | 0.009257 | 0.01137 |
| MCH | rs12197304 | rs12191917 | T | C | 6 | BTN1A1 | Female only | True | 0.1574 | 0.1565 | 0.1459 | 0.146 | 35668 | 26968 | 188074 | 162398 | -0.03083 | 0.001755 | 0.004686 | 0.003054 | 0.009257 | 0.01137 |
| MCH | rs12197304 | rs12198993 | C | T | 6 | BTN1A1 | Female only | True | 0.1574 | 0.1565 | 0.1459 | 0.146 | 35668 | 26968 | 188074 | 162398 | -0.03083 | 0.001755 | 0.004682 | 0.002987 | 0.009257 | 0.01137 |
| MCH | rs12197304 | rs7745238 | A | G | 6 | BTN1A1 | Female only | True | 0.1575 | 0.1566 | 0.1462 | 0.1462 | 35668 | 26968 | 188074 | 162398 | -0.03079 | 0.001755 | 0.005026 | 0.003093 | 0.00925 | 0.01136 |
| MCH | rs12197304 | rs2893844 | C | T | 6 | BTN1A1 | Female only | True | 0.1574 | 0.1565 | 0.1459 | 0.146 | 35668 | 26968 | 188074 | 162398 | -0.03083 | 0.001755 | 0.004682 | 0.002987 | 0.009257 | 0.01137 |
| MCH | rs12197304 | rs2393652 | G | A | 6 | BTN1A1 | Female only | True | 0.1574 | 0.1565 | 0.1459 | 0.146 | 35668 | 26968 | 188074 | 162398 | -0.03083 | 0.001762 | 0.004687 | 0.002996 | 0.009257 | 0.01137 |
| MCH | rs12197304 | rs2893845 | A | G | 6 | BTN1A1 | Female only | True | 0.1574 | 0.1565 | 0.1459 | 0.146 | 35668 | 26968 | 188074 | 162398 | -0.03086 | 0.001757 | 0.004679 | 0.00296 | 0.009257 | 0.01137 |
| MCH | rs12197304 | rs2893846 | G | T | 6 | BTN1A1 | Female only | True | 0.1574 | 0.1565 | 0.1459 | 0.146 | 35668 | 26968 | 188074 | 162398 | -0.03083 | 0.001762 | 0.004682 | 0.002987 | 0.009257 | 0.01137 |
| MCH | rs12197304 | rs56280732 | G | A | 6 | BTN1A1 | Female only | True | 0.1574 | 0.1565 | 0.1459 | 0.146 | 35668 | 26968 | 188074 | 162398 | -0.03083 | 0.001755 | 0.004675 | 0.002987 | 0.009257 | 0.01137 |
| MCH | rs12197304 | rs56157336 | T | C | 6 | BTN1A1 | Female only | True | 0.1156 | 0.1141 | 0.1072 | 0.1069 | 35668 | 26968 | 188074 | 162398 | -0.05505 | -0.01648 | -0.0167 | -0.0295 | 0.01055 | 0.013 |
| MCH | rs12197304 | rs12194073 | T | G | 6 | BTN1A1 | Female only | True | 0.1574 | 0.1565 | 0.1459 | 0.146 | 35668 | 26968 | 188074 | 162398 | -0.03084 | 0.001912 | 0.004682 | 0.002987 | 0.009257 | 0.01137 |
| MCH | rs12197304 | rs12194001 | T | C | 6 | BTN1A1 | Female only | True | 0.1574 | 0.1565 | 0.1459 | 0.146 | 35668 | 26968 | 188074 | 162398 | -0.03084 | 0.001905 | 0.004682 | 0.002987 | 0.009257 | 0.01137 |
| MCH | rs12197304 | rs7751280 | A | G | 6 | BTN1A1 | Female only | True | 0.1574 | 0.1565 | 0.1459 | 0.146 | 35668 | 26968 | 188074 | 162398 | -0.03084 | 0.001905 | 0.004675 | 0.002987 | 0.009257 | 0.01137 |
| MCH | rs12197304 | rs7775365 | C | T | 6 | BTN1A1 | Female only | True | 0.1574 | 0.1565 | 0.1459 | 0.146 | 35668 | 26968 | 188074 | 162398 | -0.03084 | 0.001912 | 0.004675 | 0.002987 | 0.009257 | 0.01137 |
| MCH | rs12197304 | rs7755398 | T | C | 6 | BTN1A1 | Female only | True | 0.1156 | 0.1141 | 0.1072 | 0.1069 | 35668 | 26968 | 188074 | 162398 | -0.05504 | -0.01645 | -0.0167 | -0.0295 | 0.01055 | 0.013 |
| MCH | rs12197304 | rs12203120 | C | T | 6 | BTN1A1 | Female only | True | 0.1574 | 0.1565 | 0.1459 | 0.146 | 35668 | 26968 | 188074 | 162398 | -0.03084 | 0.001912 | 0.004675 | 0.002987 | 0.009257 | 0.01137 |

| trait | lead_snp | ID | ALLELE1 | ALLELE0 | CHR | Gene_Symbol | independent_loci_in_sex_P1e2 | in_embl_any_or_gene | A1FREQ_VUMC_female | A1FREQ_VUMC_male | A1FREQ_UKB_female | A1FREQ_UKB_male | N_VUMC_female | N_VUMC_male | N_UKB_female | N_UKB_male | BETA_VUMC_female | BETA_VUMC_male | BETA_UKB_female | BETA_UKB_male | SE_VUMC_female | SE_VUMC_male |
|---|---|---|---|---|---|---|---|---|---|---|---|---|---|---|---|---|---|---|---|---|---|---|
| MCH | rs12197304 | rs10946827 | A | G | 6 | BTN1A1 | Female only | True | 0.1574 | 0.1565 | 0.1459 | 0.146 | 35668 | 26968 | 188074 | 162398 | -0.03084 | 0.001912 | 0.004675 | 0.002987 | 0.009257 | 0.01137 |
| MCH | rs12197304 | rs10946829 | A | G | 6 | BTN1A1 | Female only | True | 0.1575 | 0.1566 | 0.1459 | 0.146 | 35668 | 26968 | 188074 | 162398 | -0.03079 | 0.001917 | 0.00471 | 0.002986 | 0.00925 | 0.01136 |
| MCH | rs12197304 | rs3846843 | A | G | 6 | BTN1A1 | Female only | True | 0.1574 | 0.1565 | | | 35668 | 26968 | | | -0.03084 | 0.001912 | | | 0.009257 | 0.01137 |
| MCH | rs12197304 | rs4083469 | A | G | 6 | BTN1A1 | Female only | True | 0.1574 | 0.1565 | 0.1459 | 0.146 | 35668 | 26968 | 188074 | 162398 | -0.03084 | 0.001913 | 0.004713 | 0.002975 | 0.009257 | 0.01137 |
| MCH | rs12197304 | rs12201546 | A | G | 6 | BTN1A1 | Female only | True | 0.1574 | 0.1565 | 0.1459 | 0.146 | 35668 | 26968 | 188074 | 162398 | -0.03096 | 0.001633 | 0.004755 | 0.003069 | 0.009255 | 0.01137 |
| MCH | rs12197304 | rs4712994 | A | C | 6 | BTN1A1 | Female only | True | 0.1575 | 0.1566 | 0.146 | 0.1461 | 35668 | 26968 | 188074 | 162398 | -0.03118 | 0.001508 | 0.004545 | 0.003438 | 0.009247 | 0.01136 |
| MCH | rs12197304 | rs4712995 | G | A | 6 | BTN1A1 | Female only | True | 0.1575 | 0.1566 | 0.146 | 0.1461 | 35668 | 26968 | 188074 | 162398 | -0.03118 | 0.001539 | 0.004468 | 0.003288 | 0.009247 | 0.01135 |
| MCH | rs12197304 | rs17611220 | G | A | 6 | BTN1A1 | Female only | True | 0.1575 | 0.1566 | 0.146 | 0.1461 | 35668 | 26968 | 188074 | 162398 | -0.03117 | 0.001539 | 0.004466 | 0.003344 | 0.009247 | 0.01135 |
| MCH | rs12197304 | rs2893847 | A | C | 6 | BTN1A1 | Female only | True | 0.1574 | 0.1565 | 0.1461 | 0.1463 | 35668 | 26968 | 188074 | 162398 | -0.03185 | 0.001133 | 0.004777 | 0.003328 | 0.00925 | 0.01135 |
| MCH | rs12197304 | rs2893848 | G | A | 6 | BTN1A1 | Female only | True | 0.1575 | 0.1567 | 0.146 | 0.1461 | 35668 | 26968 | 188074 | 162398 | -0.03182 | 0.00113 | 0.004559 | 0.003175 | 0.009243 | 0.01134 |
| MCH | rs12197304 | rs45499696 | G | A | 6 | BTN1A1 | Female only | True | 0.1577 | 0.1568 | 0.1469 | 0.147 | 35668 | 26968 | 188074 | 162398 | -0.03122 | 0.001472 | 0.003013 | 0.002695 | 0.009236 | 0.01134 |
| MCH | rs12197304 | rs7770699 | G | A | 6 | BTN1A1 | Female only | True | 0.1574 | 0.1566 | 0.146 | 0.1461 | 35668 | 26968 | 188074 | 162398 | -0.03135 | 0.001608 | 0.004468 | 0.003288 | 0.009257 | 0.01137 |
| MCH | rs12197304 | rs2393655 | A | G | 6 | BTN1A1 | Female only | True | 0.1575 | 0.1566 | 0.146 | 0.1461 | 35668 | 26968 | 188074 | 162398 | -0.03101 | 0.001654 | 0.004477 | 0.003269 | 0.009256 | 0.01137 |
| MCH | rs12197304 | rs2393656 | C | A | 6 | BTN1A1 | Female only | True | 0.1155 | 0.1141 | 0.1073 | 0.107 | 35668 | 26968 | 188074 | 162398 | -0.05516 | -0.0166 | -0.01691 | -0.02916 | 0.01057 | 0.01301 |
| MCH | rs12197304 | rs4711110 | T | C | 6 | BTN1A1 | Female only | True | 0.1155 | 0.1141 | 0.1073 | 0.107 | 35668 | 26968 | 188074 | 162398 | -0.05516 | -0.01663 | -0.01691 | -0.02916 | 0.01057 | 0.01301 |
| MCH | rs12197304 | rs6910023 | T | C | 6 | BTN1A1 | Female only | True | 0.1574 | 0.1566 | 0.146 | 0.1461 | 35668 | 26968 | 188074 | 162398 | -0.03135 | 0.001608 | 0.004495 | 0.003288 | 0.009257 | 0.01137 |
| MCH | rs12197304 | rs12206839 | A | G | 6 | BTN1A1 | Female only | True | 0.1572 | 0.1565 | 0.1459 | 0.1461 | 35668 | 26968 | 188074 | 162398 | -0.03195 | 0.001261 | 0.004564 | 0.003228 | 0.009263 | 0.01137 |
| MCH | rs12197304 | rs12206812 | T | C | 6 | BTN1A1 | Female only | True | 0.1574 | 0.1566 | 0.1461 | 0.1461 | 35668 | 26968 | 188074 | 162398 | -0.03135 | 0.001608 | 0.004415 | 0.003311 | 0.009257 | 0.01137 |
| MCH | rs12197304 | rs17539219 | G | T | 6 | BTN1A1 | Female only | True | 0.1574 | 0.1566 | 0.146 | 0.1461 | 35668 | 26968 | 188074 | 162398 | -0.03135 | 0.001608 | 0.004496 | 0.003288 | 0.009257 | 0.01137 |
| MCH | rs12197304 | rs12214444 | T | C | 6 | BTN1A1 | Female only | True | 0.1156 | 0.1141 | 0.1073 | 0.107 | 35668 | 26968 | 188074 | 162398 | -0.05515 | -0.01663 | -0.01658 | -0.02944 | 0.01057 | 0.01301 |
| MCH | rs12197304 | rs4711111 | A | G | 6 | BTN1A1 | Female only | True | 0.1574 | 0.1566 | 0.146 | 0.1461 | 35668 | 26968 | 188074 | 162398 | -0.03134 | 0.001609 | 0.004483 | 0.003324 | 0.009257 | 0.01137 |
| MCH | rs12197304 | rs4712996 | C | T | 6 | BTN1A1 | Female only | True | 0.1574 | 0.1566 | 0.149 | 0.1491 | 35668 | 26968 | 188074 | 162398 | -0.03135 | 0.001609 | 0.004038 | 0.002893 | 0.009257 | 0.01137 |
| MCH | rs12197304 | rs7745767 | C | T | 6 | BTN1A1 | Female only | True | 0.1575 | 0.1567 | 0.1463 | 0.1464 | 35668 | 26968 | 188074 | 162398 | -0.0312 | 0.00144 | 0.004759 | 0.003406 | 0.009256 | 0.01137 |
| MCH | rs12197304 | rs7768108 | G | A | 6 | BTN1A1 | Female only | True | 0.1158 | 0.1144 | 0.1075 | 0.1072 | 35668 | 26968 | 188074 | 162398 | -0.05498 | -0.01551 | -0.01628 | -0.02884 | 0.01056 | 0.013 |
| MCH | rs12197304 | rs55999571 | C | A | 6 | BTN1A1 | Female only | True | 0.1158 | 0.1143 | 0.1073 | 0.107 | 35668 | 26968 | 188074 | 162398 | -0.05495 | -0.01552 | -0.01695 | -0.0291 | 0.01056 | 0.013 |
| MCH | rs12197304 | rs17611438 | A | C | 6 | BTN1A1 | Female only | True | 0.1576 | 0.1569 | 0.146 | 0.146 | 35668 | 26968 | 188074 | 162398 | -0.03104 | 0.002409 | 0.004487 | 0.00321 | 0.009255 | 0.01136 |
| MCH | rs12197304 | rs3846844 | A | G | 6 | BTN1A1 | Female only | True | 0.1576 | 0.1569 | 0.146 | 0.146 | 35668 | 26968 | 188074 | 162398 | -0.03104 | 0.002407 | 0.004508 | 0.00318 | 0.009255 | 0.01136 |
| MCH | rs12197304 | rs10946832 | G | A | 6 | BTN1A1 | Female only | True | 0.1125 | 0.1102 | 0.1048 | 0.1046 | 35668 | 26968 | 188074 | 162398 | -0.05436 | -0.01397 | -0.01746 | -0.02968 | 0.0107 | 0.01323 |
| MCH | rs12197304 | rs3846847 | T | C | 6 | BTN1A1 | Female only | True | 0.1576 | 0.1569 | 0.146 | 0.146 | 35668 | 26968 | 188074 | 162398 | -0.03104 | 0.002407 | 0.004508 | 0.00318 | 0.009255 | 0.01136 |

| trait | lead_snp | ID | ALLELE1 | ALLELE0 | CHR | Gene_Symbol | independent_loci_in_sex_P1e2 | in_embl_any_or_gene | A1FREQ_VUMC_female | A1FREQ_VUMC_male | A1FREQ_UKB_female | A1FREQ_UKB_male | N_VUMC_female | N_VUMC_male | N_UKB_female | N_UKB_male | BETA_VUMC_female | BETA_VUMC_male | BETA_UKB_female | BETA_UKB_male | SE_VUMC_female | SE_VUMC_male |
|---|---|---|---|---|---|---|---|---|---|---|---|---|---|---|---|---|---|---|---|---|---|---|
| MCH | rs12197304 | rs3846848 | T | C | 6 | BTN1A1 | Female only | True | 0.1576 | 0.1568 | 0.146 | 0.146 | 35668 | 26968 | 188074 | 162398 | -0.03124 | 0.002451 | 0.004628 | 0.003264 | 0.009257 | 0.01136 |
| MCH | rs12197304 | rs3846849 | G | A | 6 | BTN1A1 | Female only | True | 0.1571 | 0.1565 | 0.1463 | 0.1464 | 35668 | 26968 | 188074 | 162398 | -0.0314 | 0.00148 | 0.004452 | 0.002858 | 0.009279 | 0.01139 |
| MCH | rs12197304 | rs2893849 | G | A | 6 | BTN1A1 | Female only | True | 0.1419 | 0.1409 | 0.1301 | 0.1302 | 35668 | 26968 | 188074 | 162398 | -0.03399 | 0.002171 | 0.007809 | 0.005579 | 0.009739 | 0.01196 |
| MCH | rs12197304 | rs6456724 | T | C | 6 | BTN1A1 | Female only | True | 0.1447 | 0.1442 | 0.1329 | 0.133 | 35668 | 26968 | 188074 | 162398 | -0.03645 | 0.004036 | 0.004346 | 0.002992 | 0.009645 | 0.01184 |
| MCH | rs12197304 | rs6907302 | G | A | 6 | BTN1A1 | Female only | True | 0.1427 | 0.1425 | 0.1317 | 0.1319 | 35668 | 26968 | 188074 | 162398 | -0.03485 | 0.003311 | 0.005236 | 0.003124 | 0.009712 | 0.01191 |
| MCH | rs12197304 | rs6929846 | T | C | 6 | BTN1A1 | Female only | True | 0.1873 | 0.186 | 0.1674 | 0.1679 | 35668 | 26968 | 188074 | 162398 | -0.03562 | -0.008808 | -6.62e-04 | -0.002453 | 0.008696 | 0.01065 |
| MCH | rs12197304 | rs6912934 | C | T | 6 | BTN1A1 | Female only | True | 0.1447 | 0.1442 | 0.1331 | 0.1332 | 35668 | 26968 | 188074 | 162398 | -0.03668 | 0.003652 | 0.004315 | 0.003098 | 0.009651 | 0.01185 |
| MCH | rs12197304 | rs2893856 | T | C | 6 | BTN1A1 | Female only | True | 0.1447 | 0.1442 | 0.1331 | 0.1332 | 35668 | 26968 | 188074 | 162398 | -0.03668 | 0.003519 | 0.004282 | 0.003091 | 0.00965 | 0.01185 |
| MCH | rs12197304 | rs6920948 | T | C | 6 | BTN1A1 | Female only | True | 0.1445 | 0.144 | 0.1331 | 0.1332 | 35668 | 26968 | 188074 | 162398 | -0.03659 | 0.003252 | 0.004359 | 0.003119 | 0.009655 | 0.01186 |
| MCH | rs12197304 | rs6920982 | T | C | 6 | BTN1A1 | Female only | True | 0.1445 | 0.144 | 0.1331 | 0.1332 | 35668 | 26968 | 188074 | 162398 | -0.03663 | 0.003359 | 0.004337 | 0.00314 | 0.009654 | 0.01186 |
| MCH | rs12197304 | rs6941966 | A | G | 6 | BTN1A1 | Female only | True | 0.0686 | 0.06658 | 0.06294 | 0.06269 | 35668 | 26968 | 188074 | 162398 | -0.05053 | -0.01944 | -0.01186 | -0.01836 | 0.01362 | 0.01692 |
| MCH | rs12197304 | rs9467759 | T | C | 6 | BTN1A1 | Female only | True | 0.1535 | 0.1526 | 0.1405 | 0.1404 | 35668 | 26968 | 188074 | 162398 | -0.03393 | 0.001739 | 0.003536 | 0.005383 | 0.009338 | 0.01149 |
| MCH | rs12197304 | rs4712999 | G | A | 6 | BTN1A1 | Female only | True | 0.1409 | 0.1405 | 0.1325 | 0.1327 | 35668 | 26968 | 188074 | 162398 | -0.03453 | 0.002838 | 0.004946 | 0.002355 | 0.009684 | 0.01188 |
| MCH | rs12197304 | rs12211872 | G | A | 6 | BTN1A1 | Female only | True | 0.1063 | 0.1053 | 0.09963 | 0.09965 | 35668 | 26968 | 188074 | 162398 | -0.05846 | -0.01629 | -0.01582 | -0.02787 | 0.01095 | 0.01343 |
| trait | lead_snp | ID | ALLELE1 | ALLELE0 | CHR | Gene_Symbol | independent_loci_in_sex_P1e2 | in_embl_any_or_gene | A1FREQ_VUMC_female | A1FREQ_VUMC_male | A1FREQ_UKB_female | A1FREQ_UKB_male | N_VUMC_female | N_VUMC_male | N_UKB_female | N_UKB_male | BETA_VUMC_female | BETA_VUMC_male | BETA_UKB_female | BETA_UKB_male | SE_VUMC_female | SE_VUMC_male |
| MCH | rs12197304 | rs2893857 | A | C | 6 | BTN1A1 | Female only | True | 0.1061 | 0.1051 | 0.09958 | 0.09958 | 35668 | 26968 | 188074 | 162398 | -0.05858 | -0.0163 | -0.01579 | -0.02797 | 0.01096 | 0.01345 |
| MCH | rs12197304 | rs12208272 | A | G | 6 | BTN1A1 | Female only | True | 0.105 | 0.1039 | 0.09839 | 0.09838 | 35668 | 26968 | 188074 | 162398 | -0.05789 | -0.01721 | -0.01615 | -0.02836 | 0.01102 | 0.01353 |
| MCH | rs12197304 | rs12661560 | G | A | 6 | BTN1A1 | Female only | True | 0.1067 | 0.1057 | 0.1 | 0.09998 | 35668 | 26968 | 188074 | 162398 | -0.05875 | -0.01594 | -0.0163 | -0.02787 | 0.01094 | 0.01342 |
| MCH | rs12197304 | 6:26480543 | T | G | 6 | BTN1A1 | Female only | True | 0.105 | 0.1039 | | | 35668 | 26968 | | | -0.05787 | -0.0173 | | | 0.01102 | 0.01353 |
| MCH | rs12197304 | rs4713000 | G | A | 6 | BTN1A1 | Female only | True | 0.1062 | 0.1053 | 0.09987 | 0.0999 | 35668 | 26968 | 188074 | 162398 | -0.05851 | -0.01613 | -0.01581 | -0.02793 | 0.01096 | 0.01344 |
| MCH | rs12197304 | rs4713001 | A | G | 6 | BTN1A1 | Female only | True | 0.1062 | 0.1052 | 0.09958 | 0.09958 | 35668 | 26968 | 188074 | 162398 | -0.05882 | -0.01638 | -0.01608 | -0.02833 | 0.01096 | 0.01345 |
| MCH | rs12197304 | rs74917660 | A | C | 6 | BTN1A1 | Female only | True | 0.06628 | 0.06432 | 0.06303 | 0.06289 | 35668 | 26968 | 188074 | 162398 | -0.05278 | -0.01755 | -0.01089 | -0.01928 | 0.01359 | 0.01687 |
| MCH | rs12197304 | rs12193283 | A | C | 6 | BTN1A1 | Female only | True | 0.1063 | 0.1057 | 0.09995 | 0.09999 | 35668 | 26968 | 188074 | 162398 | -0.05975 | -0.0174 | -0.01641 | -0.02853 | 0.01096 | 0.01343 |
| MCH | rs12197304 | rs12197304 | T | C | 6 | BTN1A1 | Female only | True | 0.1063 | 0.1056 | 0.09993 | 0.09993 | 35668 | 26968 | 188074 | 162398 | -0.06005 | -0.0173 | -0.01658 | -0.02859 | 0.01096 | 0.01343 |
| MCH | rs12197304 | rs12215532 | A | G | 6 | BTN1A1 | Female only | True | 0.1905 | 0.1911 | 0.18 | 0.1813 | 35668 | 26968 | 188074 | 162398 | -0.04187 | -0.005587 | -0.008324 | -0.01157 | 0.008573 | 0.01043 |
| MCH | rs12197304 | rs10946833 | A | G | 6 | BTN1A1 | Female only | True | 0.1898 | 0.1901 | 0.179 | 0.1803 | 35668 | 26968 | 188074 | 162398 | -0.04292 | -0.006091 | -0.009357 | -0.01231 | 0.008599 | 0.01047 |
| MCH | rs12197304 | rs1884945 | G | A | 6 | BTN1A1 | Female only | True | 0.1063 | 0.1054 | 0.0998 | 0.09977 | 35668 | 26968 | 188074 | 162398 | -0.0587 | -0.01702 | -0.01633 | -0.02884 | 0.01097 | 0.01346 |
| MCH | rs12197304 | rs10456047 | A | G | 6 | BTN1A1 | Female only | True | 0.1061 | 0.1053 | 0.09966 | 0.09959 | 35668 | 26968 | 188074 | 162398 | -0.05813 | -0.01709 | -0.01647 | -0.02842 | 0.01098 | 0.01346 |

| trait | lead_snp | ID | ALLELE1 | ALLELE0 | CHR | Gene_Symbol | independent_loci_in_sex_P1e2 | in_embl_any_or_gene | A1FREQ_VUMC_female | A1FREQ_VUMC_male | A1FREQ_UKB_female | A1FREQ_UKB_male | N_VUMC_female | N_VUMC_male | N_UKB_female | N_UKB_male | BETA_VUMC_female | BETA_VUMC_male | BETA_UKB_female | BETA_UKB_male | SE_VUMC_female | SE_VUMC_male |
|---|---|---|---|---|---|---|---|---|---|---|---|---|---|---|---|---|---|---|---|---|---|---|
| MCH | rs12197304 | rs10456331 | C | T | 6 | BTN1A1 | Female only | True | 0.1061 | 0.1053 | 0.09967 | 0.09961 | 35668 | 26968 | 188074 | 162398 | -0.05813 | -0.0171 | -0.01652 | -0.0286 | 0.01098 | 0.01346 |
| MCH | rs12197304 | rs12208211 | G | A | 6 | BTN1A1 | Female only | True | 0.1065 | 0.1057 | 0.09975 | 0.09971 | 35668 | 26968 | 188074 | 162398 | -0.05754 | -0.01662 | -0.01642 | -0.02818 | 0.01096 | 0.01344 |
| MCH | rs12197304 | rs882010 | C | T | 6 | BTN1A1 | Female only | True | 0.1062 | 0.1051 | 0.09969 | 0.09963 | 35668 | 26968 | 188074 | 162398 | -0.05791 | -0.01667 | -0.01642 | -0.02835 | 0.01097 | 0.01347 |
| MCH | rs12197304 | rs56034653 | C | T | 6 | BTN1A1 | Female only | True | 0.1061 | 0.1049 | 0.09952 | 0.09942 | 35668 | 26968 | 188074 | 162398 | -0.05789 | -0.01614 | -0.01648 | -0.02795 | 0.01097 | 0.01347 |
| MCH | rs4713037 | rs6915555 | G | T | 6 | RP11-457M11.5 | Female only | False | 0.3736 | 0.3716 | 0.3557 | 0.3567 | 35668 | 26968 | 188074 | 162398 | -0.0335 | -0.01674 | -0.02274 | -0.03597 | 0.006988 | 0.008561 |
| MCH | rs4713037 | rs3807029 | T | C | 6 | RP11-457M11.5 | Female only | False | 0.3737 | 0.3718 | 0.3557 | 0.3567 | 35668 | 26968 | 188074 | 162398 | -0.03335 | -0.01671 | -0.02272 | -0.03594 | 0.006988 | 0.008562 |
| MCH | rs4713037 | rs3800302 | A | G | 6 | RP11-457M11.5 | Female only | False | 0.3736 | 0.3716 | 0.3557 | 0.3566 | 35668 | 26968 | 188074 | 162398 | -0.0335 | -0.01675 | -0.02275 | -0.03598 | 0.006988 | 0.008561 |
| MCH | rs4713037 | rs4591839 | C | T | 6 | RP11-457M11.5 | Female only | False | 0.3737 | 0.3717 | 0.356 | 0.357 | 35668 | 26968 | 188074 | 162398 | -0.0334 | -0.01676 | -0.02266 | -0.03585 | 0.006989 | 0.008562 |
| MCH | rs4713037 | rs6911540 | T | C | 6 | RP11-457M11.5 | Female only | False | 0.3747 | 0.3726 | 0.3561 | 0.3571 | 35668 | 26968 | 188074 | 162398 | -0.03383 | -0.01786 | -0.02277 | -0.03605 | 0.006982 | 0.008553 |
| MCH | rs4713037 | rs6931849 | A | G | 6 | RP11-457M11.5 | Female only | False | 0.3748 | 0.3728 | 0.356 | 0.357 | 35668 | 26968 | 188074 | 162398 | -0.03369 | -0.01782 | -0.02278 | -0.03626 | 0.006982 | 0.008554 |
| MCH | rs4713037 | rs9379902 | G | A | 6 | RP11-457M11.5 | Female only | False | 0.3749 | 0.3728 | 0.3561 | 0.3571 | 35668 | 26968 | 188074 | 162398 | -0.03355 | -0.01781 | -0.02274 | -0.03585 | 0.006975 | 0.008547 |
| MCH | rs4713037 | rs9379903 | T | C | 6 | RP11-457M11.5 | Female only | False | 0.3745 | 0.3725 | 0.3561 | 0.3572 | 35668 | 26968 | 188074 | 162398 | -0.03344 | -0.01769 | -0.0228 | -0.03598 | 0.006978 | 0.00855 |
| MCH | rs4713037 | rs6919391 | G | A | 6 | RP11-457M11.5 | Female only | False | 0.3731 | 0.371 | 0.3561 | 0.3571 | 35668 | 26968 | 188074 | 162398 | -0.03356 | -0.01773 | -0.02273 | -0.03594 | 0.006955 | 0.008522 |
| MCH | rs4713037 | rs2451739 | T | G | 6 | RP11-457M11.5 | Female only | False | 0.3758 | 0.3737 | 0.3568 | 0.3578 | 35668 | 26968 | 188074 | 162398 | -0.0326 | -0.01717 | -0.0231 | -0.0364 | 0.006973 | 0.008542 |
| MCH | rs4713037 | rs3999476 | G | A | 6 | RP11-457M11.5 | Female only | False | 0.3747 | 0.3728 | 0.3561 | 0.3572 | 35668 | 26968 | 188074 | 162398 | -0.03325 | -0.01628 | -0.02278 | -0.03603 | 0.006974 | 0.008541 |
| MCH | rs4713037 | rs2494697 | A | G | 6 | RP11-457M11.5 | Female only | False | 0.3747 | 0.3728 | 0.3561 | 0.3572 | 35668 | 26968 | 188074 | 162398 | -0.03325 | -0.01628 | -0.02281 | -0.03606 | 0.006974 | 0.008541 |
| MCH | rs4713037 | rs7746643 | G | A | 6 | RP11-457M11.5 | Female only | False | 0.3748 | 0.3728 | 0.3561 | 0.3572 | 35668 | 26968 | 188074 | 162398 | -0.03325 | -0.01624 | -0.02282 | -0.03606 | 0.006974 | 0.00854 |
| MCH | rs4713037 | rs6902411 | A | C | 6 | RP11-457M11.5 | Female only | False | 0.3748 | 0.3727 | 0.356 | 0.3571 | 35668 | 26968 | 188074 | 162398 | -0.03303 | -0.01625 | -0.02286 | -0.03604 | 0.006975 | 0.00854 |
| MCH | rs4713037 | rs2451742 | A | G | 6 | RP11-457M11.5 | Female only | False | 0.3748 | 0.3728 | 0.3561 | 0.3572 | 35668 | 26968 | 188074 | 162398 | -0.03314 | -0.01627 | -0.02285 | -0.03606 | 0.006975 | 0.00854 |
| MCH | rs4713037 | rs2494705 | T | C | 6 | RP11-457M11.5 | Female only | False | 0.3749 | 0.3728 | 0.3562 | 0.3572 | 35668 | 26968 | 188074 | 162398 | -0.03312 | -0.01628 | -0.02288 | -0.03609 | 0.006975 | 0.008541 |
| MCH | rs4713037 | rs2494706 | C | T | 6 | RP11-457M11.5 | Female only | False | 0.3748 | 0.3728 | 0.3561 | 0.3572 | 35668 | 26968 | 188074 | 162398 | -0.0331 | -0.01625 | -0.02286 | -0.03605 | 0.006975 | 0.008541 |

| trait | lead_snp | ID | ALLELE1 | ALLELE0 | CHR | Gene_Symbol | independent_loci_in_sex_P1e2 | in_embl_any_or_gene | A1FREQ_VUMC_female | A1FREQ_VUMC_male | A1FREQ_UKB_female | A1FREQ_UKB_male | N_VUMC_female | N_VUMC_male | N_UKB_female | N_UKB_male | BETA_VUMC_female | BETA_VUMC_male | BETA_UKB_female | BETA_UKB_male | SE_VUMC_female | SE_VUMC_male |
|---|---|---|---|---|---|---|---|---|---|---|---|---|---|---|---|---|---|---|---|---|---|---|
| MCH | rs4713037 | rs2498342 | G | A | 6 | RP11-457M11.5 | Female only | False | 0.373 | 0.371 | 0.3561 | 0.3572 | 35668 | 26968 | 188074 | 162398 | -0.033 | -0.01621 | -0.02285 | -0.03605 | 0.006955 | 0.008517 |
| MCH | rs4713037 | rs2498343 | G | A | 6 | RP11-457M11.5 | Female only | False | 0.3611 | 0.3597 | 0.3465 | 0.3476 | 35668 | 26968 | 188074 | 162398 | -0.03284 | -0.0139 | -0.02261 | -0.0334 | 0.00703 | 0.008626 |
| MCH | rs4713037 | rs2451749 | C | A | 6 | RP11-457M11.5 | Female only | False | 0.3741 | 0.3722 | 0.3555 | 0.3567 | 35668 | 26968 | 188074 | 162398 | -0.03277 | -0.01577 | -0.02285 | -0.03636 | 0.006977 | 0.008542 |
| MCH | rs4713037 | rs2494710 | A | C | 6 | RP11-457M11.5 | Female only | False | 0.3742 | 0.3722 | 0.356 | 0.3572 | 35668 | 26968 | 188074 | 162398 | -0.03285 | -0.01581 | -0.02299 | -0.03651 | 0.006977 | 0.008543 |
| MCH | rs4713037 | rs2451752 | C | A | 6 | RP11-457M11.5 | Female only | False | 0.3741 | 0.3722 | 0.3555 | 0.3567 | 35668 | 26968 | 188074 | 162398 | -0.03281 | -0.01578 | -0.02285 | -0.03633 | 0.006977 | 0.008542 |
| MCH | rs4713037 | rs2451753 | C | T | 6 | RP11-457M11.5 | Female only | False | 0.3741 | 0.3722 | 0.3555 | 0.3567 | 35668 | 26968 | 188074 | 162398 | -0.03281 | -0.01578 | -0.02285 | -0.03633 | 0.006977 | 0.008542 |
| MCH | rs4713037 | rs2494711 | T | C | 6 | RP11-457M11.5 | Female only | False | 0.3723 | 0.3703 | 0.3555 | 0.3567 | 35668 | 26968 | 188074 | 162398 | -0.03271 | -0.01575 | -0.02283 | -0.0364 | 0.006957 | 0.008518 |
| MCH | rs4713037 | rs2494714 | A | G | 6 | RP11-457M11.5 | Female only | False | 0.3742 | 0.3722 | 0.3556 | 0.3568 | 35668 | 26968 | 188074 | 162398 | -0.0328 | -0.01578 | -0.02287 | -0.03635 | 0.006978 | 0.008544 |
| MCH | rs4713037 | rs2451716 | T | G | 6 | RP11-457M11.5 | Female only | False | 0.3741 | 0.3722 | 0.3555 | 0.3567 | 35668 | 26968 | 188074 | 162398 | -0.03284 | -0.01579 | -0.02283 | -0.03635 | 0.006978 | 0.008545 |
| MCH | rs4713037 | rs2451715 | C | T | 6 | RP11-457M11.5 | Female only | False | 0.3742 | 0.3722 | 0.3577 | 0.359 | 35668 | 26968 | 188074 | 162398 | -0.03282 | -0.01578 | -0.02288 | -0.037 | 0.006978 | 0.008544 |
| MCH | rs4713037 | rs2451714 | T | C | 6 | RP11-457M11.5 | Female only | False | 0.3604 | 0.3592 | 0.346 | 0.3472 | 35668 | 26968 | 188074 | 162398 | -0.03258 | -0.01349 | -0.02261 | -0.03369 | 0.007034 | 0.008631 |
| MCH | rs4713037 | rs867796 | G | T | 6 | RP11-457M11.5 | Female only | False | 0.3723 | 0.3705 | 0.3533 | 0.3545 | 35668 | 26968 | 188074 | 162398 | -0.03412 | -0.0157 | -0.02183 | -0.03555 | 0.006991 | 0.008561 |
| MCH | rs4713037 | rs868190 | G | A | 6 | RP11-457M11.5 | Female only | False | 0.3723 | 0.3705 | 0.354 | 0.3551 | 35668 | 26968 | 188074 | 162398 | -0.03404 | -0.01572 | -0.02179 | -0.03556 | 0.006991 | 0.008562 |
| MCH | rs4713037 | rs577857202 | C | T | 6 | RP11-457M11.5 | Female only | False | 0.2637 | 0.2624 | | | 35668 | 26968 | | | -0.04241 | -0.02279 | | | 0.007652 | 0.009379 |
| MCH | rs4713037 | rs545019110 | A | G | 6 | RP11-457M11.5 | Female only | False | 0.2512 | 0.2501 | | | 35668 | 26968 | | | -0.04354 | -0.02094 | | | 0.007778 | 0.009517 |
| MCH | rs4713037 | rs558120648 | C | T | 6 | RP11-457M11.5 | Female only | False | 0.3976 | 0.3953 | | | 35668 | 26968 | | | -0.03389 | -0.01793 | | | 0.006899 | 0.008456 |
| MCH | rs4713037 | rs527347908 | G | A | 6 | RP11-457M11.5 | Female only | False | 0.2511 | 0.25 | | | 35668 | 26968 | | | -0.0436 | -0.02083 | | | 0.007778 | 0.009518 |
| MCH | rs4713037 | rs12213056 | T | C | 6 | RP11-457M11.5 | Female only | False | 0.2497 | 0.2488 | 0.2471 | 0.2475 | 35668 | 26968 | 188074 | 162398 | -0.04423 | -0.01816 | -0.02387 | -0.0421 | 0.00781 | 0.009546 |
| MCH | rs4713037 | rs4711121 | C | T | 6 | RP11-457M11.5 | Female only | False | 0.1524 | 0.1507 | 0.1437 | 0.1443 | 35668 | 26968 | 188074 | 162398 | -0.03603 | -0.01665 | -0.003137 | -0.02172 | 0.009393 | 0.01155 |
| MCH | rs4713037 | rs4713037 | G | T | 6 | RP11-457M11.5 | Female only | False | 0.2515 | 0.2504 | 0.248 | 0.2483 | 35668 | 26968 | 188074 | 162398 | -0.04412 | -0.02016 | -0.02434 | -0.0424 | 0.007756 | 0.009495 |

| trait | lead_snp | ID | ALLELE1 | ALLELE0 | CHR | Gene_Symbol | independent_loci_in_sex_P1e2 | in_embl_any_or_gene | A1FREQ_VUMC_female | A1FREQ_VUMC_male | A1FREQ_UKB_female | A1FREQ_UKB_male | N_VUMC_female | N_VUMC_male | N_UKB_female | N_UKB_male | BETA_VUMC_female | BETA_VUMC_male | BETA_UKB_female | BETA_UKB_male | SE_VUMC_female | SE_VUMC_male |
|---|---|---|---|---|---|---|---|---|---|---|---|---|---|---|---|---|---|---|---|---|---|---|
| MCH | rs4713037 | rs4711122 | T | C | 6 | RP11-457M11.5 | Female only | False | 0.2523 | 0.2513 | 0.2479 | 0.2481 | 35668 | 26968 | 188074 | 162398 | -0.04392 | -0.02007 | -0.02416 | -0.04193 | 0.00773 | 0.009464 |
| MCH | rs4713037 | rs3999429 | T | C | 6 | RP11-457M11.5 | Female only | False | 0.2514 | 0.2503 | 0.2477 | 0.248 | 35668 | 26968 | 188074 | 162398 | -0.04413 | -0.02006 | -0.02435 | -0.04242 | 0.007759 | 0.009498 |
| MCH | rs7171289 | rs6494520 | C | A | 15 | IGDCC4 | Female only | True | 0.1985 | 0.1994 | 0.2018 | 0.2011 | 35668 | 26968 | 188074 | 162398 | -0.04703 | -0.01449 | -0.05002 | -0.05562 | 0.008463 | 0.01024 |
| MCH | rs7171289 | rs16948719 | T | C | 15 | IGDCC4 | Female only | True | 0.1903 | 0.1912 | 0.1952 | 0.1944 | 35668 | 26968 | 188074 | 162398 | -0.04544 | -0.01281 | -0.05222 | -0.056 | 0.008589 | 0.01042 |
| MCH | rs7171289 | rs7171289 | T | C | 15 | IGDCC4 | Female only | True | 0.1997 | 0.2006 | 0.2018 | 0.2011 | 35668 | 26968 | 188074 | 162398 | -0.04686 | -0.01447 | -0.04996 | -0.0553 | 0.00841 | 0.01018 |
| MCH | rs7171289 | rs4492994 | A | G | 15 | IGDCC4 | Female only | True | 0.1729 | 0.1737 | 0.1761 | 0.1757 | 35668 | 26968 | 188074 | 162398 | -0.04726 | -0.01315 | -0.05618 | -0.06267 | 0.008933 | 0.01082 |

| trait | lead_snp | ID | ALLELE1 | ALLELE0 | CHR | Gene_Symbol | independent_loci_in_sex_P1e2 | in_embl_any_or_gene | A1FREQ_VUMC_female | A1FREQ_VUMC_male | A1FREQ_UKB_female | A1FREQ_UKB_male | N_VUMC_female | N_VUMC_male | N_UKB_female | N_UKB_male | BETA_VUMC_female | BETA_VUMC_male | BETA_UKB_female | BETA_UKB_male | SE_VUMC_female | SE_VUMC_male |
|---|---|---|---|---|---|---|---|---|---|---|---|---|---|---|---|---|---|---|---|---|---|---|
| MCH | rs7171289 | rs2053137 | A | G | 15 | IGDCC4 | Female only | True | 0.1726 | 0.1735 | 0.176 | 0.1755 | 35668 | 26968 | 188074 | 162398 | -0.04766 | -0.01313 | -0.05617 | -0.06251 | 0.008941 | 0.01083 |
| MCH | rs7171289 | rs28894785 | T | C | 15 | IGDCC4 | Female only | True | 0.1729 | 0.1736 | 0.1761 | 0.1757 | 35668 | 26968 | 188074 | 162398 | -0.04726 | -0.01325 | -0.05606 | -0.06246 | 0.008933 | 0.01083 |
| MCH | rs7171289 | rs9708095 | T | G | 15 | IGDCC4 | Female only | True | 0.1282 | 0.1287 | 0.1283 | 0.1283 | 35668 | 26968 | 188074 | 162398 | -0.04759 | -0.01462 | -0.05133 | -0.05368 | 0.01014 | 0.01226 |
| MCH | rs7171289 | rs8029971 | A | G | 15 | IGDCC4 | Female only | True | 0.1283 | 0.129 | 0.1283 | 0.1284 | 35668 | 26968 | 188074 | 162398 | -0.04838 | -0.01495 | -0.05127 | -0.0537 | 0.01014 | 0.01226 |
| MCH | rs7171289 | rs12916360 | T | G | 15 | IGDCC4 | Female only | True | 0.1283 | 0.129 | 0.1283 | 0.1284 | 35668 | 26968 | 188074 | 162398 | -0.04808 | -0.01472 | -0.05132 | -0.05369 | 0.01014 | 0.01226 |
| MCHC | rs13190937 | rs11752919 | C | T | 6 | ZSCAN23 | Female only | False | 0.3798 | 0.3813 | 0.4133 | 0.4147 | 35644 | 26945 | 188072 | 162396 | 0.03918 | 0.01267 | 0.01187 | 0.02045 | 0.006507 | 0.007992 |
| MCHC | rs13190937 | rs66542396 | G | A | 6 | ZSCAN23 | Female only | False | 0.3796 | 0.3811 | 0.4133 | 0.4147 | 35644 | 26945 | 188072 | 162396 | 0.03928 | 0.0127 | 0.01189 | 0.02039 | 0.00651 | 0.007995 |
| MCHC | rs13190937 | rs6917130 | G | A | 6 | ZSCAN23 | Female only | False | 0.3796 | 0.3811 | 0.4133 | 0.4147 | 35644 | 26945 | 188072 | 162396 | 0.03922 | 0.01266 | 0.01189 | 0.02039 | 0.00651 | 0.007995 |
| MCHC | rs13190937 | rs916403 | G | A | 6 | ZSCAN23 | Female only | False | 0.4602 | 0.4627 | 0.494 | 0.4948 | 35644 | 26945 | 188072 | 162396 | 0.03592 | 0.008656 | 0.006853 | 0.01364 | 0.006327 | 0.007772 |
| MCHC | rs13190937 | rs2531803 | G | T | 6 | ZSCAN23 | Female only | False | 0.4601 | 0.4626 | 0.4938 | 0.4946 | 35644 | 26945 | 188072 | 162396 | 0.03588 | 0.008612 | 0.006841 | 0.01376 | 0.006326 | 0.007772 |
| MCHC | rs13190937 | rs13190937 | A | G | 6 | ZSCAN23 | Female only | False | 0.3284 | 0.3292 | 0.3657 | 0.3668 | 35644 | 26945 | 188072 | 162396 | 0.04698 | 0.0143 | 0.01503 | 0.02024 | 0.006739 | 0.008261 |
| MCHC | rs13190937 | rs2531804 | G | A | 6 | ZSCAN23 | Female only | False | 0.3913 | 0.393 | 0.4201 | 0.4213 | 35644 | 26945 | 188072 | 162396 | 0.03912 | 0.01358 | 0.01102 | 0.01898 | 0.006472 | 0.007956 |
| MCHC | rs13190937 | rs1361387 | G | T | 6 | ZSCAN23 | Female only | False | 0.4599 | 0.4624 | 0.4938 | 0.4946 | 35644 | 26945 | 188072 | 162396 | 0.03586 | 0.008639 | 0.006819 | 0.01373 | 0.006329 | 0.007775 |
| MCHC | rs13190937 | 6:28413491 | C | T | 6 | ZSCAN23 | Female only | False | 0.3286 | 0.3294 | | | 35644 | 26945 | | | 0.04653 | 0.01378 | | | 0.006737 | 0.008259 |
| MCHC | rs13190937 | rs13207208 | C | A | 6 | ZSCAN23 | Female only | False | 0.3797 | 0.3811 | 0.4134 | 0.4148 | 35644 | 26945 | 188072 | 162396 | 0.03913 | 0.01257 | 0.01186 | 0.02038 | 0.00651 | 0.007995 |
| MCHC | rs13190937 | rs13215804 | G | A | 6 | ZSCAN23 | Female only | False | 0.3285 | 0.3293 | 0.3658 | 0.367 | 35644 | 26945 | 188072 | 162396 | 0.04671 | 0.01376 | 0.01525 | 0.01997 | 0.00674 | 0.008263 |
| MCHC | rs13190937 | rs6939966 | G | A | 6 | ZSCAN23 | Female only | False | 0.3796 | 0.3811 | 0.4133 | 0.4147 | 35644 | 26945 | 188072 | 162396 | 0.03935 | 0.01265 | 0.01175 | 0.02033 | 0.00651 | 0.007995 |
| MCHC | rs13190937 | rs2531806 | A | C | 6 | ZSCAN23 | Female only | False | 0.4659 | 0.4685 | 0.4977 | 0.4985 | 35644 | 26945 | 188072 | 162396 | 0.03625 | 0.007615 | 0.006422 | 0.0135 | 0.006325 | 0.007766 |
| MCHC | rs13190937 | rs6903535 | G | A | 6 | ZSCAN23 | Female only | False | 0.3788 | 0.3804 | 0.4126 | 0.4141 | 35644 | 26945 | 188072 | 162396 | 0.03954 | 0.01295 | 0.0123 | 0.02049 | 0.006511 | 0.007998 |
| MCHC | rs13190937 | rs2859368 | C | T | 6 | ZSCAN23 | Female only | False | 0.4599 | 0.4624 | 0.4936 | 0.4944 | 35644 | 26945 | 188072 | 162396 | 0.03597 | 0.008721 | 0.006726 | 0.01367 | 0.00633 | 0.007775 |

| trait | lead_snp | ID | ALLELE1 | ALLELE0 | CHR | Gene_Symbol | independent_loci_in_sex_P1e2 | in_embl_any_or_gene | A1FREQ_VUMC_female | A1FREQ_VUMC_male | A1FREQ_UKB_female | A1FREQ_UKB_male | N_VUMC_female | N_VUMC_male | N_UKB_female | N_UKB_male | BETA_VUMC_female | BETA_VUMC_male | BETA_UKB_female | BETA_UKB_male | SE_VUMC_female | SE_VUMC_male |
|---|---|---|---|---|---|---|---|---|---|---|---|---|---|---|---|---|---|---|---|---|---|---|
| MCHC | rs13306780 | rs2269905 | G | A | 17 | SLC4A1 | Female only | True | 0.3406 | 0.3428 | 0.3385 | 0.3373 | 35644 | 26945 | 188072 | 162396 | 0.03759 | 0.007348 | 0.05057 | 0.04781 | 0.00668 | 0.008162 |
| MCHC | rs13306780 | rs8072954 | A | G | 17 | SLC4A1 | Female only | True | 0.3401 | 0.3423 | 0.3383 | 0.3371 | 35644 | 26945 | 188072 | 162396 | 0.03777 | 0.007155 | 0.05045 | 0.04777 | 0.006678 | 0.008158 |
| MCHC | rs13306780 | rs10208 | T | C | 17 | SLC4A1 | Female only | True | 0.3131 | 0.3148 | 0.3118 | 0.3106 | 35644 | 26945 | 188072 | 162396 | 0.03523 | -0.002747 | 0.0471 | 0.04816 | 0.006822 | 0.00833 |
| MCHC | rs13306780 | rs11656758 | G | A | 17 | SLC4A1 | Female only | True | 0.343 | 0.3453 | 0.341 | 0.3394 | 35644 | 26945 | 188072 | 162396 | 0.03681 | 0.007342 | 0.0509 | 0.04812 | 0.006659 | 0.008137 |
| MCHC | rs13306780 | rs12935898 | G | A | 17 | SLC4A1 | Female only | True | 0.3431 | 0.3455 | 0.341 | 0.3395 | 35644 | 26945 | 188072 | 162396 | 0.03682 | 0.007203 | 0.05086 | 0.04809 | 0.006656 | 0.008134 |
| MCHC | rs13306780 | rs12451882 | T | C | 17 | SLC4A1 | Female only | True | 0.3147 | 0.3167 | 0.3134 | 0.3121 | 35644 | 26945 | 188072 | 162396 | 0.03462 | -0.002379 | 0.04749 | 0.04839 | 0.006807 | 0.008312 |
| MCHC | rs13306780 | rs9899127 | T | C | 17 | SLC4A1 | Female only | True | 0.3432 | 0.3455 | 0.341 | 0.3395 | 35644 | 26945 | 188072 | 162396 | 0.03679 | 0.007286 | 0.05088 | 0.0482 | 0.006656 | 0.008134 |
| MCHC | rs13306780 | rs11652651 | A | G | 17 | SLC4A1 | Female only | True | 0.3431 | 0.3455 | 0.341 | 0.3395 | 35644 | 26945 | 188072 | 162396 | 0.03682 | 0.0072 | 0.05096 | 0.04817 | 0.006656 | 0.008134 |
| MCHC | rs13306780 | rs9906807 | T | C | 17 | SLC4A1 | Female only | True | 0.3431 | 0.3454 | 0.341 | 0.3395 | 35644 | 26945 | 188072 | 162396 | 0.03684 | 0.007261 | 0.05094 | 0.0482 | 0.006656 | 0.008134 |
| MCHC | rs13306780 | rs7222349 | A | G | 17 | SLC4A1 | Female only | True | 0.3431 | 0.3455 | 0.341 | 0.3395 | 35644 | 26945 | 188072 | 162396 | 0.03682 | 0.007195 | 0.05093 | 0.0482 | 0.006656 | 0.008134 |
| MCHC | rs13306780 | rs9901595 | A | G | 17 | SLC4A1 | Female only | True | 0.3412 | 0.3433 | 0.3403 | 0.3387 | 35644 | 26945 | 188072 | 162396 | 0.03749 | 0.006804 | 0.05067 | 0.04808 | 0.006662 | 0.00814 |
| MCHC | rs13306780 | rs9906669 | G | A | 17 | SLC4A1 | Female only | True | 0.3421 | 0.3442 | 0.3402 | 0.3387 | 35644 | 26945 | 188072 | 162396 | 0.0373 | 0.00664 | 0.05064 | 0.04791 | 0.006645 | 0.008119 |
| MCHC | rs13306780 | rs8074740 | A | G | 17 | SLC4A1 | Female only | True | 0.3235 | 0.3261 | 0.3222 | 0.321 | 35644 | 26945 | 188072 | 162396 | 0.03227 | 0.003376 | 0.04794 | 0.04443 | 0.006756 | 0.008226 |
| MCHC | rs13306780 | rs4793083 | C | T | 17 | SLC4A1 | Female only | True | 0.3307 | 0.3325 | 0.325 | 0.3234 | 35644 | 26945 | 188072 | 162396 | 0.03009 | 0.003377 | 0.0483 | 0.04413 | 0.006756 | 0.008229 |
| MCHC | rs13306780 | rs9896789 | T | C | 17 | SLC4A1 | Female only | True | 0.2942 | 0.2968 | 0.2939 | 0.2928 | 35644 | 26945 | 188072 | 162396 | 0.02915 | -0.005545 | 0.04468 | 0.04494 | 0.006936 | 0.00844 |
| MCHC | rs13306780 | rs4793084 | G | T | 17 | SLC4A1 | Female only | True | 0.3012 | 0.3033 | 0.2977 | 0.2964 | 35644 | 26945 | 188072 | 162396 | 0.02873 | -0.006995 | 0.04505 | 0.04459 | 0.006925 | 0.008434 |
| MCHC | rs13306780 | rs4793085 | A | C | 17 | SLC4A1 | Female only | True | 0.3458 | 0.3476 | 0.3417 | 0.34 | 35644 | 26945 | 188072 | 162396 | 0.03634 | 0.006808 | 0.05161 | 0.04797 | 0.006686 | 0.00816 |
| MCHC | rs13306780 | rs4793086 | C | T | 17 | SLC4A1 | Female only | True | 0.3296 | 0.3315 | 0.3244 | 0.323 | 35644 | 26945 | 188072 | 162396 | 0.0297 | 0.004381 | 0.04855 | 0.04433 | 0.006769 | 0.008239 |
| MCHC | rs13306780 | rs2079008 | C | A | 17 | SLC4A1 | Female only | True | 0.329 | 0.3313 | 0.3247 | 0.3232 | 35644 | 26945 | 188072 | 162396 | 0.03019 | 0.003491 | 0.04856 | 0.04451 | 0.006769 | 0.008241 |
| MCHC | rs13306780 | rs7209801 | A | G | 17 | SLC4A1 | Female only | True | 0.2912 | 0.2942 | 0.2906 | 0.2895 | 35644 | 26945 | 188072 | 162396 | 0.03002 | -0.007195 | 0.04577 | 0.04643 | 0.006962 | 0.008472 |
| MCHC | rs13306780 | rs12937114 | G | A | 17 | SLC4A1 | Female only | True | 0.2937 | 0.2966 | 0.2935 | 0.2925 | 35644 | 26945 | 188072 | 162396 | 0.03127 | -0.008099 | 0.04574 | 0.04611 | 0.00695 | 0.00847 |
| MCHC | rs13306780 | rs2072081 | T | G | 17 | SLC4A1 | Female only | True | 0.2557 | 0.2588 | 0.2507 | 0.2505 | 35644 | 26945 | 188072 | 162396 | 0.03948 | -0.005429 | 0.05499 | 0.05366 | 0.007223 | 0.008793 |
| MCHC | rs13306780 | rs13306780 | A | C | 17 | SLC4A1 | Female only | True | 0.3063 | 0.3091 | 0.2981 | 0.2973 | 35644 | 26945 | 188072 | 162396 | 0.04557 | 0.01033 | 0.06111 | 0.05502 | 0.006892 | 0.008404 |
| MCHC | rs13306780 | rs1476512 | C | T | 17 | SLC4A1 | Female only | True | 0.4516 | 0.454 | 0.4455 | 0.4432 | 35644 | 26945 | 188072 | 162396 | 0.02911 | 7.21e-04 | 0.03634 | 0.02595 | 0.006344 | 0.007749 |
| MCHC | rs13306780 | rs2857078 | A | C | 17 | SLC4A1 | Female only | True | 0.3067 | 0.31 | 0.2994 | 0.2987 | 35644 | 26945 | 188072 | 162396 | 0.04554 | 0.007468 | 0.0609 | 0.05486 | 0.006888 | 0.008396 |
| MCHC | rs62108438 | rs8110787 | T | C | 19 | GCDH | Female only | True | 0.3791 | 0.3788 | 0.3915 | 0.3909 | 35644 | 26945 | 188072 | 162396 | 0.03597 | 0.01794 | 0.02635 | 0.02452 | 0.006493 | 0.008026 |
| MCHC | rs62108438 | rs62108438 | C | T | 19 | GCDH | Female only | True | 0.3792 | 0.379 | 0.3917 | 0.3911 | 35644 | 26945 | 188072 | 162396 | 0.03605 | 0.01818 | 0.02654 | 0.02448 | 0.006493 | 0.008026 |
| MCHC | rs62108438 | rs11085824 | G | A | 19 | GCDH | Female only | True | 0.3677 | 0.3678 | 0.3788 | 0.3779 | 35644 | 26945 | 188072 | 162396 | 0.03517 | 0.0164 | 0.02718 | 0.02465 | 0.006509 | 0.008044 |
| MCHC | rs62108438 | rs2242517 | G | T | 19 | GCDH | Female only | True | 0.4171 | 0.4174 | 0.4276 | 0.4265 | 35644 | 26945 | 188072 | 162396 | 0.03395 | 0.01684 | 0.0222 | 0.02155 | 0.006406 | 0.00789 |

| trait | lead_snp | ID | ALLELE1 | ALLELE0 | CHR | Gene_Symbol | independent_loci_in_sex_P1e2 | in_embl_any_or_gene | A1FREQ_VUMC_female | A1FREQ_VUMC_male | A1FREQ_UKB_female | A1FREQ_UKB_male | N_VUMC_female | N_VUMC_male | N_UKB_female | N_UKB_male | BETA_VUMC_female | BETA_VUMC_male | BETA_UKB_female | BETA_UKB_male | SE_VUMC_female | SE_VUMC_male |
|---|---|---|---|---|---|---|---|---|---|---|---|---|---|---|---|---|---|---|---|---|---|---|
| MCHC | rs62108438 | rs2238641 | C | T | 19 | GCDH | Female only | True | 0.3756 | 0.3755 | 0.3894 | 0.3887 | 35644 | 26945 | 188072 | 162396 | 0.03446 | 0.01834 | 0.02591 | 0.02405 | 0.006537 | 0.00807 |
| MCHC | rs62108438 | rs12151303 | A | G | 19 | GCDH | Female only | True | 0.3771 | 0.377 | 0.3903 | 0.3896 | 35644 | 26945 | 188072 | 162396 | 0.03459 | 0.01799 | 0.02577 | 0.02407 | 0.006527 | 0.008052 |
| MCHC | rs62108438 | rs11085825 | T | C | 19 | GCDH | Female only | True | 0.3644 | 0.3644 | 0.378 | 0.377 | 35644 | 26945 | 188072 | 162396 | 0.0335 | 0.01661 | 0.02667 | 0.02355 | 0.006561 | 0.008095 |
| MCHC | rs62108438 | rs1060218 | T | G | 19 | GCDH | Female only | True | 0.3672 | 0.3675 | 0.3812 | 0.3801 | 35644 | 26945 | 188072 | 162396 | 0.03303 | 0.01605 | 0.02658 | 0.02337 | 0.006567 | 0.008092 |
| MCHC | rs62108438 | rs9384 | T | G | 19 | GCDH | Female only | True | 0.3696 | 0.3696 | 0.3829 | 0.3821 | 35644 | 26945 | 188072 | 162396 | 0.03551 | 0.0144 | 0.02648 | 0.02184 | 0.006541 | 0.008078 |
| MCHC | rs62108438 | rs116374652 | C | T | 19 | GCDH | Female only | True | 0.3575 | 0.3575 | 0.3704 | 0.3693 | 35644 | 26945 | 188072 | 162396 | 0.03463 | 0.01291 | 0.0272 | 0.02183 | 0.006587 | 0.008135 |
| MCHC | rs62108438 | rs56349703 | G | A | 19 | GCDH | Female only | True | 0.3708 | 0.3706 | 0.3832 | 0.3824 | 35644 | 26945 | 188072 | 162396 | 0.03527 | 0.01446 | 0.02653 | 0.02177 | 0.006535 | 0.008066 |
| MCHC | rs62108438 | rs17706531 | G | A | 19 | GCDH | Female only | True | 0.3574 | 0.3573 | 0.3704 | 0.3693 | 35644 | 26945 | 188072 | 162396 | 0.03468 | 0.01338 | 0.02718 | 0.02177 | 0.00658 | 0.008127 |
| MCHC | rs62108438 | rs10451496 | T | C | 19 | GCDH | Female only | True | 0.3568 | 0.357 | 0.37 | 0.3688 | 35644 | 26945 | 188072 | 162396 | 0.03449 | 0.01285 | 0.02729 | 0.02147 | 0.00658 | 0.008128 |
| MCHC | rs62108438 | rs7409457 | T | C | 19 | GCDH | Female only | True | 0.3625 | 0.3627 | 0.3698 | 0.3686 | 35644 | 26945 | 188072 | 162396 | 0.03458 | 0.01307 | 0.02718 | 0.0212 | 0.006593 | 0.008144 |
| MCHC | rs62108438 | rs3115524 | G | A | 19 | GCDH | Female only | True | 0.376 | 0.3759 | 0.3827 | 0.3819 | 35644 | 26945 | 188072 | 162396 | 0.0352 | 0.01442 | 0.02664 | 0.02142 | 0.006541 | 0.008076 |

| trait | lead_snp | ID | ALLELE1 | ALLELE0 | CHR | Gene_Symbol | independent_loci_in_sex_P1e2 | in_embl_any_or_gene | A1FREQ_VUMC_female | A1FREQ_VUMC_male | A1FREQ_UKB_female | A1FREQ_UKB_male | N_VUMC_female | N_VUMC_male | N_UKB_female | N_UKB_male | BETA_VUMC_female | BETA_VUMC_male | BETA_UKB_female | BETA_UKB_male | SE_VUMC_female | SE_VUMC_male |
|---|---|---|---|---|---|---|---|---|---|---|---|---|---|---|---|---|---|---|---|---|---|---|
| MCHC | rs62108438 | rs2900660 | C | A | 19 | GCDH | Female only | True | 0.4105 | 0.4108 | 0.4154 | 0.4142 | 35644 | 26945 | 188072 | 162396 | 0.03462 | 0.01491 | 0.02264 | 0.01904 | 0.006442 | 0.007936 |
| MCHC | rs62108438 | rs741702 | A | C | 19 | GCDH | Female only | True | 0.3714 | 0.3714 | 0.3825 | 0.3816 | 35644 | 26945 | 188072 | 162396 | 0.03484 | 0.01452 | 0.02669 | 0.02124 | 0.006497 | 0.008019 |
| MCHC | rs62108438 | rs12971339 | T | C | 19 | GCDH | Female only | True | 0.3632 | 0.3635 | 0.3698 | 0.3686 | 35644 | 26945 | 188072 | 162396 | 0.03467 | 0.01371 | 0.02683 | 0.02128 | 0.00659 | 0.008139 |
| MCHC | rs62108438 | rs11667681 | A | G | 19 | GCDH | Female only | True | 0.3625 | 0.3628 | 0.3696 | 0.3685 | 35644 | 26945 | 188072 | 162396 | 0.0344 | 0.01383 | 0.02686 | 0.02135 | 0.006592 | 0.008145 |
| MCHC | rs62108438 | rs2965214 | G | A | 19 | GCDH | Female only | True | 0.3637 | 0.3639 | 0.3701 | 0.369 | 35644 | 26945 | 188072 | 162396 | 0.03485 | 0.01348 | 0.02678 | 0.0215 | 0.006588 | 0.008138 |
| MCHC | rs62108438 | rs8113575 | G | A | 19 | GCDH | Female only | True | 0.3637 | 0.3641 | 0.3699 | 0.3688 | 35644 | 26945 | 188072 | 162396 | 0.03409 | 0.01361 | 0.02678 | 0.02146 | 0.006595 | 0.008146 |
| MCHC | rs62108438 | rs2967897 | T | C | 19 | GCDH | Female only | True | 0.3638 | 0.364 | 0.3699 | 0.3688 | 35644 | 26945 | 188072 | 162396 | 0.03414 | 0.01374 | 0.02677 | 0.02152 | 0.006593 | 0.008143 |
| MCHC | rs62108438 | rs7508225 | A | G | 19 | GCDH | Female only | True | 0.3638 | 0.364 | 0.3699 | 0.3688 | 35644 | 26945 | 188072 | 162396 | 0.03413 | 0.01372 | 0.02673 | 0.02153 | 0.006593 | 0.008144 |
| MCHC | rs62108438 | rs7508226 | A | G | 19 | GCDH | Female only | True | 0.3638 | 0.364 | 0.3699 | 0.3688 | 35644 | 26945 | 188072 | 162396 | 0.03412 | 0.01374 | 0.02674 | 0.02153 | 0.006593 | 0.008144 |
| MCHC | rs62108438 | rs11673346 | T | C | 19 | GCDH | Female only | True | 0.363 | 0.3634 | 0.3698 | 0.3686 | 35644 | 26945 | 188072 | 162396 | 0.03394 | 0.01387 | 0.02676 | 0.0216 | 0.006595 | 0.008149 |
| MCHC | rs62108438 | rs2967893 | T | G | 19 | GCDH | Female only | True | 0.363 | 0.3633 | 0.3698 | 0.3686 | 35644 | 26945 | 188072 | 162396 | 0.03396 | 0.01387 | 0.02675 | 0.02162 | 0.006595 | 0.008149 |
| MCHC | rs62108438 | rs2009222 | T | C | 19 | GCDH | Female only | True | 0.3636 | 0.3639 | 0.3698 | 0.3686 | 35644 | 26945 | 188072 | 162396 | 0.03407 | 0.01368 | 0.02666 | 0.02131 | 0.006594 | 0.008143 |
| MCHC | rs62108438 | rs2009218 | A | G | 19 | GCDH | Female only | True | 0.3629 | 0.3632 | 0.3696 | 0.3685 | 35644 | 26945 | 188072 | 162396 | 0.03387 | 0.01378 | 0.02669 | 0.02136 | 0.006596 | 0.008149 |
| MCHC | rs62108438 | rs10419627 | G | A | 19 | GCDH | Female only | True | 0.4109 | 0.4114 | 0.4152 | 0.414 | 35644 | 26945 | 188072 | 162396 | 0.03406 | 0.0158 | 0.02205 | 0.01933 | 0.006448 | 0.007947 |
| MCHC | rs62108438 | rs3111316 | G | A | 19 | GCDH | Female only | True | 0.4106 | 0.4111 | 0.4152 | 0.414 | 35644 | 26945 | 188072 | 162396 | 0.03402 | 0.01626 | 0.02202 | 0.01928 | 0.006448 | 0.007946 |
| MCHC | rs62108438 | rs2974749 | A | G | 19 | GCDH | Female only | True | 0.3631 | 0.3635 | 0.3698 | 0.3687 | 35644 | 26945 | 188072 | 162396 | 0.03363 | 0.01387 | 0.02665 | 0.02165 | 0.006594 | 0.008148 |

| trait | lead_snp | ID | ALLELE1 | ALLELE0 | CHR | Gene_Symbol | independent_loci_in_sex_P1e2 | in_embl_any_or_gene | A1FREQ_VUMC_female | A1FREQ_VUMC_male | A1FREQ_UKB_female | A1FREQ_UKB_male | N_VUMC_female | N_VUMC_male | N_UKB_female | N_UKB_male | BETA_VUMC_female | BETA_VUMC_male | BETA_UKB_female | BETA_UKB_male | SE_VUMC_female | SE_VUMC_male |
|---|---|---|---|---|---|---|---|---|---|---|---|---|---|---|---|---|---|---|---|---|---|---|
| MCHC | rs62108438 | rs2293683 | A | G | 19 | GCDH | Female only | True | 0.3592 | 0.3595 | 0.3698 | 0.3687 | 35644 | 26945 | 188072 | 162396 | 0.03347 | 0.01384 | 0.02655 | 0.02159 | 0.006545 | 0.008084 |
| MCHC | rs62108438 | rs2974750 | C | A | 19 | GCDH | Female only | True | 0.364 | 0.3646 | 0.3708 | 0.3697 | 35644 | 26945 | 188072 | 162396 | 0.03356 | 0.01314 | 0.02659 | 0.02138 | 0.006595 | 0.008147 |
| MCHC | rs62108438 | rs1010222 | A | G | 19 | GCDH | Female only | True | 0.3638 | 0.3645 | 0.3709 | 0.3698 | 35644 | 26945 | 188072 | 162396 | 0.03306 | 0.01318 | 0.02654 | 0.02129 | 0.006596 | 0.008146 |
| MCHC | rs62108438 | rs2974751 | A | C | 19 | GCDH | Female only | True | 0.4079 | 0.4085 | 0.4123 | 0.4115 | 35644 | 26945 | 188072 | 162396 | 0.03321 | 0.01667 | 0.02236 | 0.01959 | 0.006463 | 0.007968 |
| MCHC | rs62108438 | rs2965220 | T | C | 19 | GCDH | Female only | True | 0.3615 | 0.3623 | 0.3683 | 0.3675 | 35644 | 26945 | 188072 | 162396 | 0.0321 | 0.01418 | 0.02699 | 0.02137 | 0.006613 | 0.008165 |
| MCHC | rs62108438 | rs1049481 | G | T | 19 | GCDH | Female only | True | 0.4091 | 0.4095 | 0.4136 | 0.4125 | 35644 | 26945 | 188072 | 162396 | 0.03314 | 0.01647 | 0.02185 | 0.01891 | 0.006457 | 0.007956 |
| MCHC | rs62108438 | rs2974752 | G | A | 19 | GCDH | Female only | True | 0.3917 | 0.3925 | 0.401 | 0.3994 | 35644 | 26945 | 188072 | 162396 | 0.03215 | 0.01457 | 0.02226 | 0.01848 | 0.006439 | 0.007938 |
| MCHC | rs60085895 | rs2356401 | T | C | 19 | ZNF283 | Female only | False | 0.3873 | 0.3867 | 0.3995 | 0.3998 | 35644 | 26945 | 188072 | 162396 | -0.03306 | -0.009547 | -0.00446 | -0.006997 | 0.00649 | 0.007936 |
| MCHC | rs60085895 | rs2356403 | A | G | 19 | ZNF283 | Female only | False | 0.3848 | 0.3842 | 0.3995 | 0.3997 | 35644 | 26945 | 188072 | 162396 | -0.03333 | -0.00988 | -0.004471 | -0.007015 | 0.006529 | 0.007984 |
| MCHC | rs60085895 | rs59503823 | C | A | 19 | ZNF283 | Female only | False | 0.3884 | 0.3882 | 0.4034 | 0.4036 | 35644 | 26945 | 188072 | 162396 | -0.03403 | -0.01066 | -0.004766 | -0.006871 | 0.006521 | 0.007975 |
| MCHC | rs60085895 | rs7249057 | T | C | 19 | ZNF283 | Female only | False | 0.3854 | 0.3848 | 0.3998 | 0.4 | 35644 | 26945 | 188072 | 162396 | -0.03352 | -0.00995 | -0.004506 | -0.006898 | 0.006532 | 0.00799 |
| MCHC | rs60085895 | rs11878787 | A | G | 19 | ZNF283 | Female only | False | 0.3839 | 0.3831 | 0.3994 | 0.3997 | 35644 | 26945 | 188072 | 162396 | -0.03345 | -0.01076 | -0.004529 | -0.00695 | 0.00654 | 0.007993 |
| MCHC | rs60085895 | 19:44330349 | A | G | 19 | ZNF283 | Female only | False | 0.3853 | 0.3847 | | | 35644 | 26945 | | | -0.0333 | -0.009872 | | | 0.006532 | 0.00799 |
| MCHC | rs60085895 | rs10408461 | G | A | 19 | ZNF283 | Female only | False | 0.4127 | 0.4112 | 0.4278 | 0.4279 | 35644 | 26945 | 188072 | 162396 | -0.03301 | -0.01175 | -0.004209 | -0.007637 | 0.006458 | 0.007895 |
| MCHC | rs60085895 | rs11879541 | T | C | 19 | ZNF283 | Female only | False | 0.4128 | 0.4112 | 0.4279 | 0.4279 | 35644 | 26945 | 188072 | 162396 | -0.03297 | -0.01174 | -0.004225 | -0.007708 | 0.006458 | 0.007895 |
| MCHC | rs60085895 | rs17712224 | C | A | 19 | ZNF283 | Female only | False | 0.4128 | 0.4112 | 0.4279 | 0.4279 | 35644 | 26945 | 188072 | 162396 | -0.03297 | -0.01174 | -0.004268 | -0.007722 | 0.006458 | 0.007895 |
| MCHC | rs60085895 | rs10500286 | C | T | 19 | ZNF283 | Female only | False | 0.4128 | 0.4112 | 0.4278 | 0.4279 | 35644 | 26945 | 188072 | 162396 | -0.03298 | -0.01174 | -0.004284 | -0.007738 | 0.006458 | 0.007896 |
| MCHC | rs60085895 | rs11880651 | C | T | 19 | ZNF283 | Female only | False | 0.4114 | 0.4098 | 0.4275 | 0.4276 | 35644 | 26945 | 188072 | 162396 | -0.03289 | -0.01275 | -0.0042 | -0.007788 | 0.006465 | 0.007897 |
| MCHC | rs60085895 | rs349030 | A | G | 19 | ZNF283 | Female only | False | 0.4362 | 0.4372 | 0.4355 | 0.4348 | 35644 | 26945 | 188072 | 162396 | 0.02593 | 0.01226 | 0.002166 | 0.006228 | 0.006397 | 0.007846 |
| MCHC | rs60085895 | rs73052639 | C | T | 19 | ZNF283 | Female only | False | 0.4107 | 0.4094 | 0.4274 | 0.4275 | 35644 | 26945 | 188072 | 162396 | -0.03357 | -0.01168 | -0.004281 | -0.007702 | 0.006472 | 0.0079 |
| MCHC | rs60085895 | rs7255083 | C | T | 19 | ZNF283 | Female only | False | 0.4262 | 0.4267 | 0.426 | 0.425 | 35644 | 26945 | 188072 | 162396 | 0.02559 | 0.01193 | 0.002309 | 0.006276 | 0.006415 | 0.007866 |
| MCHC | rs60085895 | rs62118369 | A | G | 19 | ZNF283 | Female only | False | 0.4097 | 0.4081 | 0.4238 | 0.4238 | 35644 | 26945 | 188072 | 162396 | -0.03475 | -0.01291 | -0.003424 | -0.007045 | 0.006439 | 0.007853 |
| MCHC | rs60085895 | rs7253002 | T | C | 19 | ZNF283 | Female only | False | 0.407 | 0.4054 | 0.4239 | 0.4239 | 35644 | 26945 | 188072 | 162396 | -0.0355 | -0.01303 | -0.003451 | -0.006983 | 0.006484 | 0.007909 |
| MCHC | rs60085895 | rs7253384 | T | C | 19 | ZNF283 | Female only | False | 0.407 | 0.4054 | 0.4239 | 0.4239 | 35644 | 26945 | 188072 | 162396 | -0.0355 | -0.01303 | -0.003453 | -0.006975 | 0.006484 | 0.007909 |
| MCHC | rs60085895 | rs7253606 | T | G | 19 | ZNF283 | Female only | False | 0.407 | 0.4054 | 0.4239 | 0.4239 | 35644 | 26945 | 188072 | 162396 | -0.0355 | -0.01303 | -0.003454 | -0.006972 | 0.006484 | 0.007909 |
| MCHC | rs60085895 | rs10416116 | A | C | 19 | ZNF283 | Female only | False | 0.4083 | 0.4067 | 0.4244 | 0.4243 | 35644 | 26945 | 188072 | 162396 | -0.03551 | -0.01231 | -0.003509 | -0.007027 | 0.006478 | 0.007908 |
| MCHC | rs60085895 | rs8112065 | G | T | 19 | ZNF283 | Female only | False | 0.4083 | 0.4067 | 0.4244 | 0.4243 | 35644 | 26945 | 188072 | 162396 | -0.03551 | -0.01231 | -0.003515 | -0.007012 | 0.006478 | 0.007908 |
| MCHC | rs60085895 | rs2356432 | G | A | 19 | ZNF283 | Female only | False | 0.4083 | 0.4067 | 0.4244 | 0.4243 | 35644 | 26945 | 188072 | 162396 | -0.03551 | -0.01231 | -0.003515 | -0.007011 | 0.006478 | 0.007908 |
| MCHC | rs60085895 | rs2163855 | A | G | 19 | ZNF283 | Female only | False | 0.4083 | 0.4067 | 0.4244 | 0.4243 | 35644 | 26945 | 188072 | 162396 | -0.03551 | -0.01231 | -0.003515 | -0.007011 | 0.006478 | 0.007908 |

| trait | lead_snp | ID | ALLELE1 | ALLELE0 | CHR | Gene_Symbol | independent_loci_in_sex_P1e2 | in_embl_any_or_gene | A1FREQ_VUMC_female | A1FREQ_VUMC_male | A1FREQ_UKB_female | A1FREQ_UKB_male | N_VUMC_female | N_VUMC_male | N_UKB_female | N_UKB_male | BETA_VUMC_female | BETA_VUMC_male | BETA_UKB_female | BETA_UKB_male | SE_VUMC_female | SE_VUMC_male |
|---|---|---|---|---|---|---|---|---|---|---|---|---|---|---|---|---|---|---|---|---|---|---|
| MCHC | rs60085895 | rs2356433 | T | C | 19 | ZNF283 | Female only | False | 0.4083 | 0.4067 | 0.4244 | 0.4243 | 35644 | 26945 | 188072 | 162396 | -0.03551 | -0.01231 | -0.003537 | -0.007011 | 0.006478 | 0.007908 |
| MCHC | rs60085895 | rs12608578 | A | G | 19 | ZNF283 | Female only | False | 0.4083 | 0.4066 | 0.4244 | 0.4243 | 35644 | 26945 | 188072 | 162396 | -0.03553 | -0.01223 | -0.003515 | -0.007015 | 0.006478 | 0.007909 |
| MCHC | rs60085895 | rs10405318 | C | T | 19 | ZNF283 | Female only | False | 0.4083 | 0.4067 | 0.4244 | 0.4243 | 35644 | 26945 | 188072 | 162396 | -0.03551 | -0.01231 | -0.003515 | -0.007011 | 0.006478 | 0.007908 |
| MCHC | rs60085895 | rs7247308 | G | A | 19 | ZNF283 | Female only | False | 0.4082 | 0.4065 | 0.4244 | 0.4243 | 35644 | 26945 | 188072 | 162396 | -0.03552 | -0.01232 | -0.003517 | -0.007008 | 0.00648 | 0.00791 |
| MCHC | rs60085895 | rs73052665 | C | T | 19 | ZNF283 | Female only | False | 0.4081 | 0.4065 | 0.4244 | 0.4243 | 35644 | 26945 | 188072 | 162396 | -0.03554 | -0.01224 | -0.003525 | -0.007023 | 0.00648 | 0.007911 |
| MCHC | rs60085895 | rs1994415 | T | C | 19 | ZNF283 | Female only | False | 0.4082 | 0.4065 | 0.4244 | 0.4243 | 35644 | 26945 | 188072 | 162396 | -0.03552 | -0.01232 | -0.003514 | -0.00701 | 0.00648 | 0.00791 |
| MCHC | rs60085895 | rs62118372 | G | A | 19 | ZNF283 | Female only | False | 0.4082 | 0.4065 | 0.4244 | 0.4243 | 35644 | 26945 | 188072 | 162396 | -0.03552 | -0.01232 | -0.003514 | -0.00701 | 0.00648 | 0.00791 |
| MCHC | rs60085895 | rs57133938 | C | A | 19 | ZNF283 | Female only | False | 0.4082 | 0.4065 | 0.4244 | 0.4243 | 35644 | 26945 | 188072 | 162396 | -0.03552 | -0.01232 | -0.003514 | -0.00701 | 0.00648 | 0.00791 |
| MCHC | rs60085895 | rs60085895 | T | C | 19 | ZNF283 | Female only | False | 0.4078 | 0.4062 | 0.4241 | 0.424 | 35644 | 26945 | 188072 | 162396 | -0.03582 | -0.01233 | -0.003589 | -0.007 | 0.006483 | 0.007915 |
| MCHC | rs60085895 | rs60036744 | C | T | 19 | ZNF283 | Female only | False | 0.4078 | 0.4062 | 0.4241 | 0.424 | 35644 | 26945 | 188072 | 162396 | -0.03582 | -0.01233 | -0.003539 | -0.006957 | 0.006483 | 0.007915 |
| MCHC | rs60085895 | rs13344589 | C | A | 19 | ZNF283 | Female only | False | 0.4086 | 0.4071 | 0.4246 | 0.4245 | 35644 | 26945 | 188072 | 162396 | -0.03509 | -0.01218 | -0.003613 | -0.006955 | 0.006477 | 0.007909 |
| MCHC | rs60085895 | rs4511643 | C | T | 19 | ZNF283 | Female only | False | 0.4081 | 0.4065 | 0.4244 | 0.4243 | 35644 | 26945 | 188072 | 162396 | -0.03552 | -0.01234 | -0.003543 | -0.006984 | 0.00648 | 0.007911 |
| MCHC | rs60085895 | rs8099861 | C | T | 19 | ZNF283 | Female only | False | 0.4082 | 0.4065 | 0.4244 | 0.4243 | 35644 | 26945 | 188072 | 162396 | -0.03552 | -0.01232 | -0.003521 | -0.007003 | 0.00648 | 0.007911 |
| MCHC | rs60085895 | rs8113286 | T | C | 19 | ZNF283 | Female only | False | 0.4081 | 0.4065 | 0.4244 | 0.4243 | 35644 | 26945 | 188072 | 162396 | -0.03552 | -0.01233 | -0.00352 | -0.007003 | 0.00648 | 0.007911 |
| MCHC | rs60085895 | rs1549953 | C | T | 19 | ZNF283 | Female only | False | 0.4084 | 0.4068 | 0.4246 | 0.4245 | 35644 | 26945 | 188072 | 162396 | -0.03546 | -0.01184 | -0.003439 | -0.006946 | 0.006482 | 0.007913 |
| trait | lead_snp | ID | ALLELE1 | ALLELE0 | CHR | Gene_Symbol | independent_loci_in_sex_P1e2 | in_embl_any_or_gene | A1FREQ_VUMC_female | A1FREQ_VUMC_male | A1FREQ_UKB_female | A1FREQ_UKB_male | N_VUMC_female | N_VUMC_male | N_UKB_female | N_UKB_male | BETA_VUMC_female | BETA_VUMC_male | BETA_UKB_female | BETA_UKB_male | SE_VUMC_female | SE_VUMC_male |
| MCHC | rs60085895 | rs1549954 | A | C | 19 | ZNF283 | Female only | False | 0.4084 | 0.4067 | 0.4245 | 0.4244 | 35644 | 26945 | 188072 | 162396 | -0.03558 | -0.01185 | -0.003477 | -0.006814 | 0.006481 | 0.007912 |
| MCHC | rs60085895 | rs1549955 | C | T | 19 | ZNF283 | Female only | False | 0.4082 | 0.4065 | 0.4244 | 0.4243 | 35644 | 26945 | 188072 | 162396 | -0.0355 | -0.0123 | -0.003556 | -0.006906 | 0.00648 | 0.00791 |
| MCHC | rs60085895 | rs8106802 | G | A | 19 | ZNF283 | Female only | False | 0.4081 | 0.4065 | 0.4244 | 0.4243 | 35644 | 26945 | 188072 | 162396 | -0.0355 | -0.01232 | -0.00355 | -0.006944 | 0.00648 | 0.00791 |
| MCHC | rs60085895 | rs6509120 | T | C | 19 | ZNF283 | Female only | False | 0.4081 | 0.4065 | 0.4244 | 0.4243 | 35644 | 26945 | 188072 | 162396 | -0.03553 | -0.01229 | -0.003555 | -0.00692 | 0.00648 | 0.007911 |
| MCHC | rs60085895 | rs7254659 | C | T | 19 | ZNF283 | Female only | False | 0.4081 | 0.4065 | 0.4245 | 0.4244 | 35644 | 26945 | 188072 | 162396 | -0.03551 | -0.01229 | -0.003657 | -0.006949 | 0.00648 | 0.007911 |
| MCHC | rs60085895 | rs10415344 | C | A | 19 | ZNF283 | Female only | False | 0.4113 | 0.4096 | 0.4245 | 0.4244 | 35644 | 26945 | 188072 | 162396 | -0.03514 | -0.0121 | -0.003657 | -0.006949 | 0.006432 | 0.007851 |
| MCHC | rs60085895 | rs10421861 | A | G | 19 | ZNF283 | Female only | False | 0.4128 | 0.4109 | 0.4276 | 0.4277 | 35644 | 26945 | 188072 | 162396 | -0.03302 | -0.01205 | -0.00334 | -0.006932 | 0.006465 | 0.007894 |
| MCHC | rs60085895 | rs1061769 | A | G | 19 | ZNF283 | Female only | False | 0.408 | 0.4064 | 0.4247 | 0.4247 | 35644 | 26945 | 188072 | 162396 | -0.03554 | -0.0124 | -0.003639 | -0.007217 | 0.006486 | 0.007918 |
| MCHC | rs60085895 | rs2883946 | A | G | 19 | ZNF283 | Female only | False | 0.4079 | 0.4063 | 0.4244 | 0.4243 | 35644 | 26945 | 188072 | 162396 | -0.03555 | -0.0124 | -0.003515 | -0.00696 | 0.006484 | 0.007916 |
| MCHC | rs60085895 | rs2195981 | C | T | 19 | ZNF283 | Female only | False | 0.408 | 0.4064 | 0.4244 | 0.4243 | 35644 | 26945 | 188072 | 162396 | -0.03551 | -0.01237 | -0.003514 | -0.006971 | 0.006483 | 0.007914 |
| MCHC | rs60085895 | rs2217669 | C | T | 19 | ZNF283 | Female only | False | 0.408 | 0.4064 | 0.4243 | 0.4242 | 35644 | 26945 | 188072 | 162396 | -0.03556 | -0.01233 | -0.003392 | -0.006903 | 0.006484 | 0.007914 |
| MCHC | rs60085895 | rs2356438 | A | G | 19 | ZNF283 | Female only | False | 0.408 | 0.4064 | 0.4243 | 0.4242 | 35644 | 26945 | 188072 | 162396 | -0.03556 | -0.01233 | -0.003392 | -0.006903 | 0.006484 | 0.007914 |

| trait | lead_snp | ID | ALLELE1 | ALLELE0 | CHR | Gene_Symbol | independent_loci_in_sex_P1e2 | in_embl_any_or_gene | A1FREQ_VUMC_female | A1FREQ_VUMC_male | A1FREQ_UKB_female | A1FREQ_UKB_male | N_VUMC_female | N_VUMC_male | N_UKB_female | N_UKB_male | BETA_VUMC_female | BETA_VUMC_male | BETA_UKB_female | BETA_UKB_male | SE_VUMC_female | SE_VUMC_male |
|---|---|---|---|---|---|---|---|---|---|---|---|---|---|---|---|---|---|---|---|---|---|---|
| MCHC | rs60085895 | rs2356439 | G | A | 19 | ZNF283 | Female only | False | 0.408 | 0.4064 | 0.4243 | 0.4242 | 35644 | 26945 | 188072 | 162396 | -0.03556 | -0.01233 | -0.003392 | -0.006903 | 0.006484 | 0.007914 |
| MCHC | rs60085895 | rs7259647 | C | T | 19 | ZNF283 | Female only | False | 0.408 | 0.4064 | 0.4243 | 0.4242 | 35644 | 26945 | 188072 | 162396 | -0.03556 | -0.01233 | -0.003392 | -0.006903 | 0.006484 | 0.007914 |
| MCHC | rs60085895 | rs1549956 | G | T | 19 | ZNF283 | Female only | False | 0.4081 | 0.4065 | 0.4244 | 0.4243 | 35644 | 26945 | 188072 | 162396 | -0.03525 | -0.01225 | -0.003501 | -0.006927 | 0.006485 | 0.007917 |
| MCHC | rs60085895 | rs1549957 | C | T | 19 | ZNF283 | Female only | False | 0.4081 | 0.4065 | 0.4244 | 0.4243 | 35644 | 26945 | 188072 | 162396 | -0.03525 | -0.01225 | -0.003501 | -0.006927 | 0.006485 | 0.007917 |
| MCHC | rs60085895 | rs1549959 | A | G | 19 | ZNF283 | Female only | False | 0.4081 | 0.4065 | 0.4244 | 0.4243 | 35644 | 26945 | 188072 | 162396 | -0.03526 | -0.01225 | -0.003511 | -0.006919 | 0.006485 | 0.007917 |
| MCHC | rs60085895 | rs7247863 | G | A | 19 | ZNF283 | Female only | False | 0.4081 | 0.4065 | 0.4244 | 0.4243 | 35644 | 26945 | 188072 | 162396 | -0.03526 | -0.01225 | -0.003521 | -0.006902 | 0.006485 | 0.007917 |
| MCHC | rs60085895 | rs62114569 | C | T | 19 | ZNF283 | Female only | False | 0.4081 | 0.4065 | 0.4244 | 0.4243 | 35644 | 26945 | 188072 | 162396 | -0.03526 | -0.01225 | -0.00351 | -0.006928 | 0.006485 | 0.007917 |
| MCHC | rs60085895 | rs10412955 | C | T | 19 | ZNF283 | Female only | False | 0.408 | 0.4065 | 0.4243 | 0.4242 | 35644 | 26945 | 188072 | 162396 | -0.03531 | -0.01229 | -0.00358 | -0.006889 | 0.006486 | 0.007917 |
| MCHC | rs60085895 | rs1036246 | T | C | 19 | ZNF283 | Female only | False | 0.408 | 0.4064 | 0.4243 | 0.4242 | 35644 | 26945 | 188072 | 162396 | -0.03527 | -0.01225 | -0.003579 | -0.006884 | 0.006486 | 0.007918 |
| MCHC | rs60085895 | rs1909 | T | C | 19 | ZNF283 | Female only | False | 0.4079 | 0.4064 | 0.4243 | 0.4242 | 35644 | 26945 | 188072 | 162396 | -0.0353 | -0.01231 | -0.003575 | -0.006889 | 0.006487 | 0.007918 |
| MCHC | rs60085895 | rs1964381 | T | G | 19 | ZNF283 | Female only | False | 0.4079 | 0.4064 | 0.4243 | 0.4242 | 35644 | 26945 | 188072 | 162396 | -0.03532 | -0.01228 | -0.003573 | -0.006888 | 0.006487 | 0.007918 |
| MCHC | rs60085895 | rs239949 | C | T | 19 | ZNF283 | Female only | False | 0.4139 | 0.4123 | 0.4242 | 0.4241 | 35644 | 26945 | 188072 | 162396 | -0.03526 | -0.01235 | -0.003773 | -0.006931 | 0.006467 | 0.007891 |
| MCHC | rs60085895 | rs1050054 | G | T | 19 | ZNF283 | Female only | False | 0.4979 | 0.4958 | 0.4975 | 0.496 | 35644 | 26945 | 188072 | 162396 | -0.03014 | -0.01568 | -0.008242 | -0.01034 | 0.006333 | 0.007737 |
| MCHC | rs60085895 | rs12977303 | A | G | 19 | ZNF283 | Female only | False | 0.4989 | 0.5009 | 0.4933 | 0.4917 | 35644 | 26945 | 188072 | 162396 | 0.02791 | 0.01509 | 0.007969 | 0.01035 | 0.006295 | 0.007684 |
| MCHC | rs60085895 | rs10406290 | G | T | 19 | ZNF283 | Female only | False | 0.4986 | 0.4966 | 0.497 | 0.4956 | 35644 | 26945 | 188072 | 162396 | -0.03042 | -0.01531 | -0.008148 | -0.0106 | 0.006333 | 0.007734 |
| MCHC | rs60085895 | rs10414702 | A | G | 19 | ZNF283 | Female only | False | 0.4961 | 0.4936 | 0.4977 | 0.4962 | 35644 | 26945 | 188072 | 162396 | -0.0299 | -0.01604 | -0.008298 | -0.01028 | 0.006333 | 0.00774 |
| MCHC | rs60085895 | rs10421518 | A | G | 19 | ZNF283 | Female only | False | 0.496 | 0.4936 | 0.4977 | 0.4962 | 35644 | 26945 | 188072 | 162396 | -0.02994 | -0.01606 | -0.008305 | -0.01028 | 0.006333 | 0.007739 |
| MCHC | rs60085895 | rs1978723 | T | C | 19 | ZNF283 | Female only | False | 0.4931 | 0.4907 | 0.498 | 0.4966 | 35644 | 26945 | 188072 | 162396 | -0.02982 | -0.01609 | -0.008335 | -0.01008 | 0.006297 | 0.007697 |
| MCHC | rs60085895 | rs371875 | A | G | 19 | ZNF283 | Female only | False | 0.4984 | 0.4965 | 0.4972 | 0.4957 | 35644 | 26945 | 188072 | 162396 | -0.03062 | -0.01525 | -0.00827 | -0.01043 | 0.006333 | 0.007732 |
| MCHC | rs60085895 | rs368079 | C | T | 19 | ZNF283 | Female only | False | 0.4976 | 0.4956 | 0.4977 | 0.4961 | 35644 | 26945 | 188072 | 162396 | -0.03031 | -0.01554 | -0.008367 | -0.01017 | 0.00633 | 0.007732 |
| MCHC | rs60085895 | rs425217 | T | C | 19 | ZNF283 | Female only | False | 0.4969 | 0.4989 | 0.4938 | 0.4922 | 35644 | 26945 | 188072 | 162396 | 0.02875 | 0.01486 | 0.007994 | 0.01022 | 0.006332 | 0.007725 |
| MCHC | rs60085895 | rs429027 | G | A | 19 | ZNF283 | Female only | False | 0.497 | 0.499 | 0.4939 | 0.4922 | 35644 | 26945 | 188072 | 162396 | -0.02884 | -0.0148 | -0.008069 | -0.01026 | 0.006332 | 0.007724 |
| MCHC | rs60085895 | rs397913 | A | G | 19 | ZNF283 | Female only | False | 0.4975 | 0.4955 | 0.4979 | 0.4963 | 35644 | 26945 | 188072 | 162396 | -0.03037 | -0.0154 | -0.008493 | -0.01015 | 0.00633 | 0.007731 |
| MCHC | rs60085895 | rs368089 | T | C | 19 | ZNF283 | Female only | False | 0.4975 | 0.4955 | 0.4979 | 0.4963 | 35644 | 26945 | 188072 | 162396 | -0.03037 | -0.0154 | -0.008518 | -0.01012 | 0.00633 | 0.007731 |
| MCHC | rs60085895 | rs397346 | A | G | 19 | ZNF283 | Female only | False | 0.4958 | 0.4934 | 0.498 | 0.4965 | 35644 | 26945 | 188072 | 162396 | -0.03021 | -0.01594 | -0.008553 | -0.01007 | 0.006333 | 0.007738 |
| MCHC | rs60085895 | rs396973 | A | G | 19 | ZNF283 | Female only | False | 0.4958 | 0.4934 | 0.498 | 0.4965 | 35644 | 26945 | 188072 | 162396 | -0.03019 | -0.01593 | -0.008549 | -0.01006 | 0.006333 | 0.007738 |
| MCHC | rs60085895 | rs436249 | C | T | 19 | ZNF283 | Female only | False | 0.4975 | 0.4955 | 0.4979 | 0.4963 | 35644 | 26945 | 188072 | 162396 | -0.03035 | -0.0154 | -0.008518 | -0.01012 | 0.00633 | 0.007731 |
| MCHC | rs60085895 | rs372491 | G | A | 19 | ZNF283 | Female only | False | 0.4976 | 0.4955 | 0.4977 | 0.4961 | 35644 | 26945 | 188072 | 162396 | -0.03035 | -0.0154 | -0.008294 | -0.01011 | 0.006331 | 0.007732 |
| MCHC | rs60085895 | rs453950 | G | A | 19 | ZNF283 | Female only | False | 0.4975 | 0.4955 | 0.4976 | 0.4961 | 35644 | 26945 | 188072 | 162396 | -0.03036 | -0.01542 | -0.008294 | -0.01014 | 0.00633 | 0.007731 |

| trait | lead_snp | ID | ALLELE1 | ALLELE0 | CHR | Gene_Symbol | independent_loci_in_sex_P1e2 | in_embl_any_or_gene | A1FREQ_VUMC_female | A1FREQ_VUMC_male | A1FREQ_UKB_female | A1FREQ_UKB_male | N_VUMC_female | N_VUMC_male | N_UKB_female | N_UKB_male | BETA_VUMC_female | BETA_VUMC_male | BETA_UKB_female | BETA_UKB_male | SE_VUMC_female | SE_VUMC_male |
|---|---|---|---|---|---|---|---|---|---|---|---|---|---|---|---|---|---|---|---|---|---|---|
| MCHC | rs60085895 | rs440784 | T | C | 19 | ZNF283 | Female only | False | 0.4975 | 0.4955 | 0.4979 | 0.4963 | 35644 | 26945 | 188072 | 162396 | -0.03043 | -0.01535 | -0.008474 | -0.01014 | 0.00633 | 0.007732 |
| MCHC | rs60085895 | rs374307 | G | T | 19 | ZNF283 | Female only | False | 0.4975 | 0.4955 | 0.4979 | 0.4963 | 35644 | 26945 | 188072 | 162396 | -0.03043 | -0.01535 | -0.008474 | -0.01014 | 0.00633 | 0.007732 |
| MCHC | rs60085895 | rs441344 | T | C | 19 | ZNF283 | Female only | False | 0.4975 | 0.4955 | 0.4979 | 0.4963 | 35644 | 26945 | 188072 | 162396 | -0.03043 | -0.01535 | -0.008474 | -0.01014 | 0.00633 | 0.007732 |
| MCHC | rs60085895 | rs385321 | C | T | 19 | ZNF283 | Female only | False | 0.4975 | 0.4955 | 0.4979 | 0.4963 | 35644 | 26945 | 188072 | 162396 | -0.03043 | -0.01535 | -0.008481 | -0.01012 | 0.00633 | 0.007732 |
| MCHC | rs60085895 | rs367283 | A | G | 19 | ZNF283 | Female only | False | 0.4973 | 0.4951 | 0.4979 | 0.4964 | 35644 | 26945 | 188072 | 162396 | -0.03041 | -0.01562 | -0.008466 | -0.01005 | 0.00633 | 0.007729 |
| MCHC | rs60085895 | rs448823 | T | C | 19 | ZNF283 | Female only | False | 0.4973 | 0.4952 | 0.4978 | 0.4963 | 35644 | 26945 | 188072 | 162396 | -0.03035 | -0.01548 | -0.008423 | -0.01017 | 0.00633 | 0.007728 |
| MCHC | rs60085895 | rs448829 | T | C | 19 | ZNF283 | Female only | False | 0.4973 | 0.4952 | 0.4978 | 0.4963 | 35644 | 26945 | 188072 | 162396 | -0.03035 | -0.01547 | -0.00842 | -0.01017 | 0.00633 | 0.007728 |
| MCHC | rs60085895 | rs376328 | A | G | 19 | ZNF283 | Female only | False | 0.4978 | 0.4957 | 0.4975 | 0.4959 | 35644 | 26945 | 188072 | 162396 | -0.03013 | -0.01475 | -0.008275 | -0.009963 | 0.006328 | 0.007728 |
| MCHC | rs60085895 | rs376032 | G | A | 19 | ZNF283 | Female only | False | 0.4958 | 0.498 | 0.4933 | 0.4917 | 35644 | 26945 | 188072 | 162396 | 0.028 | 0.01403 | 0.007956 | 0.009971 | 0.006329 | 0.007724 |
| MCHC | rs60085895 | rs365556 | A | G | 19 | ZNF283 | Female only | False | 0.4978 | 0.4957 | 0.4976 | 0.496 | 35644 | 26945 | 188072 | 162396 | -0.03008 | -0.01479 | -0.008284 | -0.009918 | 0.006328 | 0.007728 |
| MCHC | rs60085895 | rs451945 | T | C | 19 | ZNF283 | Female only | False | 0.4978 | 0.4957 | 0.4976 | 0.496 | 35644 | 26945 | 188072 | 162396 | -0.03009 | -0.01479 | -0.008284 | -0.009918 | 0.006328 | 0.007728 |
| MCHC | rs60085895 | rs454813 | A | C | 19 | ZNF283 | Female only | False | 0.4978 | 0.4957 | 0.4976 | 0.496 | 35644 | 26945 | 188072 | 162396 | -0.03008 | -0.01479 | -0.008284 | -0.009918 | 0.006328 | 0.007728 |
| MCHC | rs60085895 | rs417400 | A | G | 19 | ZNF283 | Female only | False | 0.4978 | 0.4957 | 0.4976 | 0.496 | 35644 | 26945 | 188072 | 162396 | -0.03008 | -0.01479 | -0.008284 | -0.009918 | 0.006328 | 0.007728 |
| MCHC | rs60085895 | rs426534 | T | G | 19 | ZNF283 | Female only | False | 0.4978 | 0.4957 | 0.4976 | 0.496 | 35644 | 26945 | 188072 | 162396 | -0.03008 | -0.01479 | -0.008287 | -0.009918 | 0.006328 | 0.007728 |
| MCHC | rs60085895 | rs430667 | T | C | 19 | ZNF283 | Female only | False | 0.4978 | 0.4957 | 0.4976 | 0.496 | 35644 | 26945 | 188072 | 162396 | -0.03013 | -0.0148 | -0.008313 | -0.009911 | 0.006328 | 0.007728 |
| MCHC | rs60085895 | rs398964 | G | A | 19 | ZNF283 | Female only | False | 0.4978 | 0.4957 | 0.4975 | 0.496 | 35644 | 26945 | 188072 | 162396 | -0.03014 | -0.0148 | -0.008301 | -0.009873 | 0.006328 | 0.007728 |
| MCHC | rs60085895 | rs423765 | C | T | 19 | ZNF283 | Female only | False | 0.4952 | 0.4931 | 0.4975 | 0.4958 | 35644 | 26945 | 188072 | 162396 | -0.03001 | -0.01467 | -0.008193 | -0.009975 | 0.006295 | 0.007689 |
| MCHC | rs60085895 | rs376069 | T | C | 19 | ZNF283 | Female only | False | 0.4978 | 0.4957 | 0.4974 | 0.4957 | 35644 | 26945 | 188072 | 162396 | -0.03031 | -0.01432 | -0.008202 | -0.01005 | 0.006327 | 0.007725 |
| MCHC | rs60085895 | rs417699 | A | G | 19 | ZNF283 | Female only | False | 0.4979 | 0.4958 | 0.4969 | 0.4952 | 35644 | 26945 | 188072 | 162396 | -0.03064 | -0.01366 | -0.008462 | -0.01022 | 0.006331 | 0.007731 |

| trait | lead_snp | ID | ALLELE1 | ALLELE0 | CHR | Gene_Symbol | independent_loci_in_sex_P1e2 | in_embl_any_or_gene | A1FREQ_VUMC_female | A1FREQ_VUMC_male | A1FREQ_UKB_female | A1FREQ_UKB_male | N_VUMC_female | N_VUMC_male | N_UKB_female | N_UKB_male | BETA_VUMC_female | BETA_VUMC_male | BETA_UKB_female | BETA_UKB_male | SE_VUMC_female | SE_VUMC_male |
|---|---|---|---|---|---|---|---|---|---|---|---|---|---|---|---|---|---|---|---|---|---|---|
| MCHC | rs60085895 | rs407731 | C | T | 19 | ZNF283 | Female only | False | 0.496 | 0.4942 | 0.4965 | 0.4948 | 35644 | 26945 | 188072 | 162396 | -0.03085 | -0.01374 | -0.008545 | -0.01021 | 0.006297 | 0.007691 |
| MCHC | rs60085895 | rs406968 | C | T | 19 | ZNF283 | Female only | False | 0.4986 | 0.4966 | 0.4968 | 0.4949 | 35644 | 26945 | 188072 | 162396 | -0.03064 | -0.01406 | -0.008654 | -0.01023 | 0.006329 | 0.00773 |
| MCHC | rs60085895 | rs425221 | A | G | 19 | ZNF283 | Female only | False | 0.4985 | 0.4966 | 0.4969 | 0.4952 | 35644 | 26945 | 188072 | 162396 | -0.03066 | -0.01362 | -0.008491 | -0.01032 | 0.00633 | 0.007731 |
| MCHC | rs60085895 | rs388706 | T | C | 19 | ZNF283 | Female only | False | 0.4959 | 0.494 | 0.4967 | 0.4949 | 35644 | 26945 | 188072 | 162396 | -0.03062 | -0.01355 | -0.008714 | -0.01037 | 0.006297 | 0.007693 |
| MCHC | rs60085895 | rs387689 | C | T | 19 | ZNF283 | Female only | False | 0.4985 | 0.4966 | 0.4968 | 0.495 | 35644 | 26945 | 188072 | 162396 | -0.03066 | -0.01362 | -0.008585 | -0.01039 | 0.00633 | 0.007731 |
| Plt.Ct | rs6141 | rs4912484 | G | T | 3 | EIF2B5 | Female only | True | 0.3677 | 0.3648 | 0.3966 | 0.3968 | 35703 | 26981 | 188075 | 162399 | -0.03761 | -0.004833 | -0.03774 | -0.04482 | 0.007609 | 0.008891 |
| Plt.Ct | rs6141 | rs4912485 | T | C | 3 | EIF2B5 | Female only | True | 0.366 | 0.3627 | 0.3731 | 0.3734 | 35703 | 26981 | 188075 | 162399 | -0.03837 | -0.004848 | -0.0381 | -0.0448 | 0.007657 | 0.008948 |
| Plt.Ct | rs6141 | rs6141 | C | T | 3 | EIF2B5 | Female only | True | 0.4561 | 0.4544 | 0.4696 | 0.4715 | 35703 | 26981 | 188075 | 162399 | -0.04901 | -0.02018 | -0.05567 | -0.05974 | 0.007382 | 0.008581 |

| trait | lead_snp | ID | ALLELE1 | ALLELE0 | CHR | Gene_Symbol | independent_loci_in_sex_P1e2 | in_embl_any_or_gene | A1FREQ_VUMC_female | A1FREQ_VUMC_male | A1FREQ_UKB_female | A1FREQ_UKB_male | N_VUMC_female | N_VUMC_male | N_UKB_female | N_UKB_male | BETA_VUMC_female | BETA_VUMC_male | BETA_UKB_female | BETA_UKB_male | SE_VUMC_female | SE_VUMC_male |
|---|---|---|---|---|---|---|---|---|---|---|---|---|---|---|---|---|---|---|---|---|---|---|
| Plt.Ct | rs2288419 | rs11667847 | G | A | 19 | PTPRH | Female only | True | 0.1735 | 0.1725 | 0.1689 | 0.17 | 35703 | 26981 | 188075 | 162399 | -0.05072 | -0.01719 | -0.02238 | -0.008091 | 0.009624 | 0.0112 |
| Plt.Ct | rs2288419 | rs891187 | T | C | 19 | PTPRH | Female only | True | 0.1782 | 0.1765 | 0.1701 | 0.1711 | 35703 | 26981 | 188075 | 162399 | -0.04891 | -0.01759 | -0.02188 | -0.007439 | 0.009482 | 0.01106 |
| Plt.Ct | rs2288419 | rs2365725 | C | T | 19 | PTPRH | Female only | True | 0.1557 | 0.1536 | 0.1453 | 0.1458 | 35703 | 26981 | 188075 | 162399 | -0.0419 | -0.007112 | -0.01404 | -0.01115 | 0.01005 | 0.01171 |
| Plt.Ct | rs2288419 | rs7260371 | A | G | 19 | PTPRH | Female only | True | 0.155 | 0.1529 | 0.1454 | 0.146 | 35703 | 26981 | 188075 | 162399 | -0.04196 | -0.007358 | -0.0142 | -0.01123 | 0.01009 | 0.01176 |
| Plt.Ct | rs2288419 | rs7260320 | G | A | 19 | PTPRH | Female only | True | 0.155 | 0.1528 | 0.1454 | 0.1459 | 35703 | 26981 | 188075 | 162399 | -0.04183 | -0.00786 | -0.01412 | -0.01126 | 0.01009 | 0.01176 |
| Plt.Ct | rs2288419 | rs3848617 | A | G | 19 | PTPRH | Female only | True | 0.155 | 0.1529 | 0.1453 | 0.1459 | 35703 | 26981 | 188075 | 162399 | -0.04222 | -0.008162 | -0.01401 | -0.01116 | 0.0101 | 0.01177 |
| Plt.Ct | rs2288419 | rs7249414 | A | C | 19 | PTPRH | Female only | True | 0.1551 | 0.1532 | 0.1454 | 0.1459 | 35703 | 26981 | 188075 | 162399 | -0.04177 | -0.008252 | -0.01466 | -0.01089 | 0.01009 | 0.01176 |
| Plt.Ct | rs2288419 | rs3786866 | A | G | 19 | PTPRH | Female only | True | 0.3033 | 0.301 | 0.289 | 0.2902 | 35703 | 26981 | 188075 | 162399 | -0.02409 | -0.01811 | -0.008585 | -0.001349 | 0.007921 | 0.009244 |
| Plt.Ct | rs2288419 | rs2288419 | C | T | 19 | PTPRH | Female only | True | 0.2036 | 0.2003 | 0.1915 | 0.1922 | 35703 | 26981 | 188075 | 162399 | -0.05039 | -0.02191 | -0.02353 | -0.009089 | 0.009036 | 0.01058 |
| Plt.Ct | rs2288419 | rs3760868 | G | T | 19 | PTPRH | Female only | True | 0.1359 | 0.1342 | 0.1275 | 0.128 | 35703 | 26981 | 188075 | 162399 | -0.04428 | -0.01522 | -0.02564 | -0.006862 | 0.01059 | 0.01239 |
| Plt.Ct | rs2288419 | rs11666987 | T | G | 19 | PTPRH | Female only | True | 0.1683 | 0.166 | 0.16 | 0.1607 | 35703 | 26981 | 188075 | 162399 | -0.04247 | -0.01708 | -0.0205 | -0.006828 | 0.009758 | 0.01138 |
| RBC | rs707890 | rs2229768 | C | T | 6 | HIST1H4C | Female only | True | 0.2267 | 0.2277 | 0.2389 | 0.239 | 35638 | 26954 | 188076 | 162399 | 0.02194 | 0.004658 | 0.007264 | 0.005286 | 0.008498 | 0.009508 |
| RBC | rs707890 | rs707890 | C | A | 6 | HIST1H4C | Female only | True | 0.1601 | 0.1611 | 0.1621 | 0.1625 | 35638 | 26954 | 188076 | 162399 | 0.05318 | 0.001514 | 0.02329 | 0.03558 | 0.00971 | 0.01083 |
| RBC | rs707890 | rs707891 | T | G | 6 | HIST1H4C | Female only | True | 0.1586 | 0.1595 | 0.1607 | 0.1611 | 35638 | 26954 | 188076 | 162399 | 0.05241 | 0.001117 | 0.02258 | 0.03557 | 0.009744 | 0.01088 |
| RBC | rs707890 | rs707892 | G | T | 6 | HIST1H4C | Female only | True | 0.1601 | 0.1611 | 0.1621 | 0.1625 | 35638 | 26954 | 188076 | 162399 | 0.0531 | 0.00161 | 0.02332 | 0.03554 | 0.009709 | 0.01083 |
| RBC | rs707890 | rs74293938 | C | T | 6 | HIST1H4C | Female only | True | 0.1563 | 0.1571 | 0.1594 | 0.1599 | 35638 | 26954 | 188076 | 162399 | 0.052 | 0.002532 | 0.02258 | 0.03533 | 0.00981 | 0.01095 |
| RBC | rs707890 | rs707893 | T | C | 6 | HIST1H4C | Female only | True | 0.1601 | 0.161 | 0.1621 | 0.1625 | 35638 | 26954 | 188076 | 162399 | 0.05303 | 0.001644 | 0.02327 | 0.03546 | 0.009709 | 0.01083 |
| RBC | rs707890 | rs707894 | C | A | 6 | HIST1H4C | Female only | True | 0.1601 | 0.1611 | 0.1622 | 0.1626 | 35638 | 26954 | 188076 | 162399 | 0.05301 | 0.00147 | 0.0231 | 0.03531 | 0.009709 | 0.01083 |
| RBC | rs707890 | rs12205050 | T | C | 6 | HIST1H4C | Female only | True | 0.1564 | 0.1571 | 0.1595 | 0.16 | 35638 | 26954 | 188076 | 162399 | 0.05199 | 0.002351 | 0.02241 | 0.03517 | 0.00981 | 0.01095 |
| RBC | rs707890 | rs707896 | A | G | 6 | HIST1H4C | Female only | True | 0.1604 | 0.1614 | 0.1621 | 0.1625 | 35638 | 26954 | 188076 | 162399 | 0.05255 | 0.001359 | 0.02333 | 0.03524 | 0.009689 | 0.01081 |
| RBC | rs707890 | rs707898 | C | T | 6 | HIST1H4C | Female only | True | 0.1588 | 0.1596 | 0.1604 | 0.1606 | 35638 | 26954 | 188076 | 162399 | 0.05079 | 0.001717 | 0.02331 | 0.03583 | 0.009736 | 0.01088 |
| RBC | rs707890 | rs16891325 | C | T | 6 | HIST1H4C | Female only | True | 0.156 | 0.1569 | 0.1587 | 0.1591 | 35638 | 26954 | 188076 | 162399 | 0.05036 | 0.002509 | 0.023 | 0.03508 | 0.009821 | 0.01097 |
| RBC | rs707890 | rs1773284 | A | C | 6 | HIST1H4C | Female only | True | 0.1585 | 0.1593 | 0.1601 | 0.1603 | 35638 | 26954 | 188076 | 162399 | 0.05078 | 0.001573 | 0.02379 | 0.03576 | 0.009753 | 0.0109 |
| RBC | rs707890 | rs74571768 | A | G | 6 | HIST1H4C | Female only | True | 0.1561 | 0.1569 | 0.1588 | 0.1591 | 35638 | 26954 | 188076 | 162399 | 0.0505 | 0.002541 | 0.02325 | 0.03518 | 0.009819 | 0.01097 |
| RBC | rs707890 | rs1773285 | T | G | 6 | HIST1H4C | Female only | True | 0.1585 | 0.1592 | 0.1601 | 0.1603 | 35638 | 26954 | 188076 | 162399 | 0.05077 | 0.001569 | 0.0237 | 0.03536 | 0.009753 | 0.0109 |
| RBC | rs707890 | rs77919048 | C | T | 6 | HIST1H4C | Female only | True | 0.1561 | 0.1569 | 0.1588 | 0.1591 | 35638 | 26954 | 188076 | 162399 | 0.0505 | 0.002543 | 0.02327 | 0.03516 | 0.009819 | 0.01097 |
| RBC | rs707890 | rs6931863 | A | G | 6 | HIST1H4C | Female only | True | 0.1566 | 0.1571 | 0.1573 | 0.1575 | 35638 | 26954 | 188076 | 162399 | 0.04962 | 0.004206 | 0.02319 | 0.03637 | 0.009836 | 0.01101 |
| RBC | rs707890 | rs6931542 | A | C | 6 | HIST1H4C | Female only | True | 0.1564 | 0.1569 | 0.1573 | 0.1575 | 35638 | 26954 | 188076 | 162399 | 0.04952 | 0.004091 | 0.02334 | 0.03628 | 0.009843 | 0.01102 |
| RBC | rs707890 | rs1150663 | G | A | 6 | HIST1H4C | Female only | True | 0.1585 | 0.1592 | 0.1601 | 0.1603 | 35638 | 26954 | 188076 | 162399 | 0.05079 | 0.001477 | 0.02359 | 0.03568 | 0.009754 | 0.0109 |

| trait | lead_snp | ID | ALLELE1 | ALLELE0 | CHR | Gene_Symbol | independent_loci_in_sex_P1e2 | in_embl_any_or_gene | A1FREQ_VUMC_female | A1FREQ_VUMC_male | A1FREQ_UKB_female | A1FREQ_UKB_male | N_VUMC_female | N_VUMC_male | N_UKB_female | N_UKB_male | BETA_VUMC_female | BETA_VUMC_male | BETA_UKB_female | BETA_UKB_male | SE_VUMC_female | SE_VUMC_male |
|---|---|---|---|---|---|---|---|---|---|---|---|---|---|---|---|---|---|---|---|---|---|---|
| RBC | rs707890 | rs806976 | T | C | 6 | HIST1H4C | Female only | True | 0.1585 | 0.1592 | 0.1601 | 0.1603 | 35638 | 26954 | 188076 | 162399 | 0.0508 | 0.001416 | 0.02352 | 0.03564 | 0.009754 | 0.0109 |
| RBC | rs707890 | rs707899 | T | C | 6 | HIST1H4C | Female only | True | 0.1588 | 0.1595 | 0.1601 | 0.1603 | 35638 | 26954 | 188076 | 162399 | 0.05071 | 0.001413 | 0.02357 | 0.03566 | 0.009737 | 0.01088 |
| RBC | rs707890 | rs707901 | T | C | 6 | HIST1H4C | Female only | True | 0.1583 | 0.159 | 0.1594 | 0.1596 | 35638 | 26954 | 188076 | 162399 | 0.05096 | 0.001138 | 0.02344 | 0.03518 | 0.009788 | 0.01094 |
| RBC | rs707890 | rs16891344 | T | G | 6 | HIST1H4C | Female only | True | 0.1559 | 0.1567 | 0.1586 | 0.159 | 35638 | 26954 | 188076 | 162399 | 0.05059 | 0.002056 | 0.02297 | 0.03514 | 0.009854 | 0.01101 |
| RDW | rs2042327 | rs255695 | G | T | 5 | FBN2 | Female only | True | 0.3093 | 0.3091 | 0.317 | 0.3174 | 35661 | 26955 | 188075 | 162398 | -0.02341 | -0.007416 | -0.01607 | -0.01581 | 0.007419 | 0.00909 |
| RDW | rs2042327 | rs190450 | A | G | 5 | FBN2 | Female only | True | 0.3074 | 0.3072 | 0.3169 | 0.3173 | 35661 | 26955 | 188075 | 162398 | -0.02356 | -0.007031 | -0.01612 | -0.01586 | 0.007398 | 0.009066 |
| RDW | rs2042327 | rs32205 | C | A | 5 | FBN2 | Female only | True | 0.3091 | 0.3089 | 0.3171 | 0.3175 | 35661 | 26955 | 188075 | 162398 | -0.02369 | -0.006968 | -0.01629 | -0.0158 | 0.007416 | 0.009087 |
| RDW | rs2042327 | rs2042327 | C | T | 5 | FBN2 | Female only | True | 0.4308 | 0.4304 | 0.4364 | 0.4357 | 35661 | 26955 | 188075 | 162398 | -0.03996 | -0.0216 | -0.02388 | -0.02773 | 0.006928 | 0.008487 |
| RDW | rs2042327 | 5:127635867 | C | A | 5 | FBN2 | Female only | True | 0.4153 | 0.4167 | | | 35661 | 26955 | | | -0.02741 | -0.01601 | | | 0.006949 | 0.0085 |
| RDW | rs2042327 | 5:127638464 | A | C | 5 | FBN2 | Female only | True | 0.4179 | 0.4194 | | | 35661 | 26955 | | | -0.02931 | -0.01684 | | | 0.006961 | 0.008515 |
| RDW | rs2042327 | rs32216 | C | T | 5 | FBN2 | Female only | True | 0.3084 | 0.3075 | 0.3162 | 0.3166 | 35661 | 26955 | 188075 | 162398 | -0.02091 | -0.01067 | -0.01558 | -0.01639 | 0.007414 | 0.009072 |
| RDW | rs2042327 | rs27713 | G | A | 5 | FBN2 | Female only | True | 0.3074 | 0.3063 | 0.3163 | 0.3167 | 35661 | 26955 | 188075 | 162398 | -0.0207 | -0.01002 | -0.01563 | -0.01648 | 0.007393 | 0.009054 |
| RDW | rs2042327 | rs27855 | A | G | 5 | FBN2 | Female only | True | 0.4139 | 0.4152 | 0.4231 | 0.4237 | 35661 | 26955 | 188075 | 162398 | -0.02741 | -0.01662 | -0.02029 | -0.02154 | 0.006931 | 0.008477 |
| RDW | rs2042327 | rs255731 | A | G | 5 | FBN2 | Female only | True | 0.4119 | 0.4134 | 0.4219 | 0.4226 | 35661 | 26955 | 188075 | 162398 | -0.02653 | -0.0179 | -0.02092 | -0.02124 | 0.006956 | 0.008516 |
| RDW | rs1611657 | rs112442053 | G | A | 6 | HCG4P7 | Female only | False | 0.39 | 0.3922 | | | 35661 | 26955 | | | -0.02682 | -0.01052 | | | 0.007021 | 0.008607 |
| RDW | rs1611657 | rs885936 | G | A | 6 | HCG4P7 | Female only | False | 0.3994 | 0.4013 | 0.391 | 0.3893 | 35661 | 26955 | 188075 | 162398 | -0.02753 | -0.009889 | -0.03028 | -0.03497 | 0.006996 | 0.00858 |
| RDW | rs1611657 | rs3094164 | G | A | 6 | HCG4P7 | Female only | False | 0.3466 | 0.3483 | 0.3347 | 0.3336 | 35661 | 26955 | 188075 | 162398 | -0.02434 | -0.01007 | -0.03012 | -0.02827 | 0.007207 | 0.008826 |
| RDW | rs1611657 | rs3132725 | C | T | 6 | HCG4P7 | Female only | False | 0.3474 | 0.3493 | | | 35661 | 26955 | | | -0.02395 | -0.009261 | | | 0.007205 | 0.008824 |
| RDW | rs1611657 | rs3115623 | T | C | 6 | HCG4P7 | Female only | False | 0.346 | 0.3477 | | | 35661 | 26955 | | | -0.0242 | -0.01008 | | | 0.0072 | 0.008817 |
| RDW | rs1611657 | rs3094162 | A | G | 6 | HCG4P7 | Female only | False | 0.3464 | 0.3481 | | | 35661 | 26955 | | | -0.02448 | -0.01028 | | | 0.007213 | 0.008832 |
| RDW | rs1611657 | rs3132724 | C | T | 6 | HCG4P7 | Female only | False | 0.3464 | 0.348 | | | 35661 | 26955 | | | -0.02449 | -0.01029 | | | 0.007213 | 0.008831 |
| RDW | rs1611657 | rs3094161 | A | G | 6 | HCG4P7 | Female only | False | 0.3466 | 0.3483 | | | 35661 | 26955 | | | -0.02426 | -0.01008 | | | 0.007207 | 0.008826 |
| RDW | rs1611657 | rs3094160 | A | G | 6 | HCG4P7 | Female only | False | 0.3466 | 0.3483 | | | 35661 | 26955 | | | -0.02426 | -0.01008 | | | 0.007207 | 0.008826 |

| trait | lead_snp | ID | ALLELE1 | ALLELE0 | CHR | Gene_Symbol | independent_loci_in_sex_P1e2 | in_embl_any_or_gene | A1FREQ_VUMC_female | A1FREQ_VUMC_male | A1FREQ_UKB_female | A1FREQ_UKB_male | N_VUMC_female | N_VUMC_male | N_UKB_female | N_UKB_male | BETA_VUMC_female | BETA_VUMC_male | BETA_UKB_female | BETA_UKB_male | SE_VUMC_female | SE_VUMC_male |
|---|---|---|---|---|---|---|---|---|---|---|---|---|---|---|---|---|---|---|---|---|---|---|
| RDW | rs1611657 | rs3132721 | C | T | 6 | HCG4P7 | Female only | False | 0.3466 | 0.3483 | | | 35661 | 26955 | | | -0.02426 | -0.01008 | | | 0.007207 | 0.008826 |
| RDW | rs1611657 | rs3094159 | G | A | 6 | HCG4P7 | Female only | False | 0.346 | 0.3477 | | | 35661 | 26955 | | | -0.0242 | -0.01008 | | | 0.0072 | 0.008817 |
| RDW | rs1611657 | rs3128898 | G | T | 6 | HCG4P7 | Female only | False | 0.3466 | 0.3483 | | | 35661 | 26955 | | | -0.02426 | -0.01008 | | | 0.007207 | 0.008826 |
| RDW | rs1611657 | rs3132720 | A | C | 6 | HCG4P7 | Female only | False | 0.346 | 0.3477 | | | 35661 | 26955 | | | -0.0242 | -0.01008 | | | 0.0072 | 0.008817 |

| trait | lead_snp | ID | ALLELE1 | ALLELE0 | CHR | Gene_Symbol | independent_loci_in_sex_P1e2 | in_embl_any_or_gene | A1FREQ_VUMC_female | A1FREQ_VUMC_male | A1FREQ_UKB_female | A1FREQ_UKB_male | N_VUMC_female | N_VUMC_male | N_UKB_female | N_UKB_male | BETA_VUMC_female | BETA_VUMC_male | BETA_UKB_female | BETA_UKB_male | SE_VUMC_female | SE_VUMC_male |
|---|---|---|---|---|---|---|---|---|---|---|---|---|---|---|---|---|---|---|---|---|---|---|
| RDW | rs1611657 | rs9258854 | G | A | 6 | HCG4P7 | Female only | False | 0.346 | 0.3477 | | | 35661 | 26955 | | | -0.0242 | -0.01008 | | | 0.0072 | 0.008817 |
| RDW | rs1611657 | rs9258855 | T | C | 6 | HCG4P7 | Female only | False | 0.3466 | 0.3483 | | | 35661 | 26955 | | | -0.02427 | -0.01007 | | | 0.007207 | 0.008826 |
| RDW | rs1611657 | rs3094158 | A | G | 6 | HCG4P7 | Female only | False | 0.3466 | 0.3483 | | | 35661 | 26955 | | | -0.02427 | -0.01007 | | | 0.007207 | 0.008826 |
| RDW | rs1611657 | rs3132719 | A | C | 6 | HCG4P7 | Female only | False | 0.3466 | 0.3483 | | | 35661 | 26955 | | | -0.02427 | -0.01007 | | | 0.007207 | 0.008826 |
| RDW | rs1611657 | rs3129011 | C | T | 6 | HCG4P7 | Female only | False | 0.346 | 0.3477 | | | 35661 | 26955 | | | -0.0242 | -0.01008 | | | 0.0072 | 0.008817 |
| RDW | rs1611657 | rs3132718 | T | C | 6 | HCG4P7 | Female only | False | 0.346 | 0.3477 | 0.3347 | 0.3336 | 35661 | 26955 | 188075 | 162398 | -0.0242 | -0.01008 | -0.03005 | -0.02817 | 0.0072 | 0.008817 |
| RDW | rs1611657 | rs1611637 | A | G | 6 | HCG4P7 | Female only | False | 0.4209 | 0.4236 | 0.4131 | 0.4124 | 35661 | 26955 | 188075 | 162398 | -0.02968 | -0.009274 | -0.03536 | -0.03845 | 0.006931 | 0.008513 |
| RDW | rs1611657 | rs3094156 | G | A | 6 | HCG4P7 | Female only | False | 0.3482 | 0.3498 | 0.3601 | 0.3588 | 35661 | 26955 | 188075 | 162398 | -0.02383 | -0.01071 | -0.02954 | -0.02751 | 0.0072 | 0.00882 |
| RDW | rs1611657 | rs9258884 | G | A | 6 | HCG4P7 | Female only | False | 0.3467 | 0.3484 | 0.3512 | 0.3501 | 35661 | 26955 | 188075 | 162398 | -0.02428 | -0.01003 | -0.02961 | -0.02762 | 0.007207 | 0.008827 |
| RDW | rs1611657 | rs9258894 | A | C | 6 | HCG4P7 | Female only | False | 0.3466 | 0.3483 | 0.3348 | 0.3337 | 35661 | 26955 | 188075 | 162398 | -0.02427 | -0.01009 | -0.03003 | -0.02814 | 0.007207 | 0.008826 |
| RDW | rs1611657 | rs9258896 | A | G | 6 | HCG4P7 | Female only | False | 0.3481 | 0.3497 | 0.3377 | 0.3368 | 35661 | 26955 | 188075 | 162398 | -0.02441 | -0.009562 | -0.02942 | -0.02759 | 0.007216 | 0.008839 |
| RDW | rs1611657 | rs9258902 | T | G | 6 | HCG4P7 | Female only | False | 0.3466 | 0.3483 | 0.3348 | 0.3336 | 35661 | 26955 | 188075 | 162398 | -0.02427 | -0.01008 | -0.03006 | -0.02815 | 0.007207 | 0.008826 |
| RDW | rs1611657 | rs2394224 | A | G | 6 | HCG4P7 | Female only | False | 0.3466 | 0.3483 | 0.3347 | 0.3336 | 35661 | 26955 | 188075 | 162398 | -0.02427 | -0.01008 | -0.03002 | -0.02815 | 0.007207 | 0.008826 |
| RDW | rs1611657 | rs2893988 | A | G | 6 | HCG4P7 | Female only | False | 0.3466 | 0.3483 | 0.3347 | 0.3336 | 35661 | 26955 | 188075 | 162398 | -0.02427 | -0.01008 | -0.03002 | -0.02815 | 0.007207 | 0.008826 |
| RDW | rs1611657 | rs2893989 | A | G | 6 | HCG4P7 | Female only | False | 0.3466 | 0.3483 | 0.3347 | 0.3336 | 35661 | 26955 | 188075 | 162398 | -0.02427 | -0.01008 | -0.03002 | -0.02815 | 0.007207 | 0.008826 |
| RDW | rs1611657 | rs3873264 | C | T | 6 | HCG4P7 | Female only | False | 0.3466 | 0.3483 | 0.3347 | 0.3336 | 35661 | 26955 | 188075 | 162398 | -0.02428 | -0.01007 | -0.03002 | -0.02815 | 0.007207 | 0.008826 |
| RDW | rs1611657 | rs2394226 | G | T | 6 | HCG4P7 | Female only | False | 0.3466 | 0.3482 | 0.3347 | 0.3336 | 35661 | 26955 | 188075 | 162398 | -0.02425 | -0.0101 | -0.03001 | -0.02815 | 0.007207 | 0.008826 |
| RDW | rs1611657 | rs9258912 | A | G | 6 | HCG4P7 | Female only | False | 0.3466 | 0.3482 | 0.3347 | 0.3336 | 35661 | 26955 | 188075 | 162398 | -0.02425 | -0.0101 | -0.03001 | -0.02815 | 0.007207 | 0.008826 |
| RDW | rs1611657 | rs9258913 | C | T | 6 | HCG4P7 | Female only | False | 0.3466 | 0.3483 | 0.3347 | 0.3336 | 35661 | 26955 | 188075 | 162398 | -0.02427 | -0.01007 | -0.03002 | -0.02815 | 0.007207 | 0.008826 |
| RDW | rs1611657 | rs9258914 | G | A | 6 | HCG4P7 | Female only | False | 0.3466 | 0.3483 | 0.3347 | 0.3336 | 35661 | 26955 | 188075 | 162398 | -0.02427 | -0.01007 | -0.03002 | -0.02815 | 0.007207 | 0.008826 |
| RDW | rs1611657 | rs9258915 | C | T | 6 | HCG4P7 | Female only | False | 0.3466 | 0.3483 | 0.3347 | 0.3336 | 35661 | 26955 | 188075 | 162398 | -0.02427 | -0.01007 | -0.03002 | -0.02815 | 0.007207 | 0.008826 |
| RDW | rs1611657 | rs9258917 | A | C | 6 | HCG4P7 | Female only | False | 0.3472 | 0.3488 | 0.3469 | 0.3457 | 35661 | 26955 | 188075 | 162398 | -0.02439 | -0.009812 | -0.03073 | -0.02856 | 0.007203 | 0.00882 |
| RDW | rs1611657 | rs3129010 | G | T | 6 | HCG4P7 | Female only | False | 0.3466 | 0.3483 | 0.3347 | 0.3336 | 35661 | 26955 | 188075 | 162398 | -0.02424 | -0.01007 | -0.03 | -0.02817 | 0.007207 | 0.008826 |
| RDW | rs1611657 | rs3129009 | C | T | 6 | HCG4P7 | Female only | False | 0.3466 | 0.3483 | 0.3347 | 0.3336 | 35661 | 26955 | 188075 | 162398 | -0.02425 | -0.01007 | -0.03 | -0.02817 | 0.007207 | 0.008826 |
| RDW | rs1611657 | rs3094154 | G | A | 6 | HCG4P7 | Female only | False | 0.3466 | 0.3483 | 0.3347 | 0.3336 | 35661 | 26955 | 188075 | 162398 | -0.02424 | -0.01007 | -0.03 | -0.02818 | 0.007207 | 0.008826 |
| RDW | rs1611657 | rs3094153 | G | A | 6 | HCG4P7 | Female only | False | 0.3466 | 0.3483 | 0.3348 | 0.3337 | 35661 | 26955 | 188075 | 162398 | -0.02424 | -0.01007 | -0.02998 | -0.02818 | 0.007207 | 0.008826 |
| RDW | rs1611657 | rs3132717 | T | C | 6 | HCG4P7 | Female only | False | 0.3466 | 0.3483 | 0.3347 | 0.3336 | 35661 | 26955 | 188075 | 162398 | -0.02424 | -0.01007 | -0.03001 | -0.02817 | 0.007207 | 0.008826 |
| RDW | rs1611657 | rs3094152 | G | A | 6 | HCG4P7 | Female only | False | 0.3466 | 0.3483 | 0.3347 | 0.3336 | 35661 | 26955 | 188075 | 162398 | -0.02427 | -0.01007 | -0.03003 | -0.02816 | 0.007207 | 0.008826 |
| RDW | rs1611657 | rs3132716 | A | C | 6 | HCG4P7 | Female only | False | 0.3466 | 0.3483 | 0.3349 | 0.3338 | 35661 | 26955 | 188075 | 162398 | -0.02427 | -0.01006 | -0.03002 | -0.0282 | 0.007207 | 0.008826 |

| trait | lead_snp | ID | ALLELE1 | ALLELE0 | CHR | Gene_Symbol | independent_loci_in_sex_P1e2 | in_embl_any_or_gene | A1FREQ_VUMC_female | A1FREQ_VUMC_male | A1FREQ_UKB_female | A1FREQ_UKB_male | N_VUMC_female | N_VUMC_male | N_UKB_female | N_UKB_male | BETA_VUMC_female | BETA_VUMC_male | BETA_UKB_female | BETA_UKB_male | SE_VUMC_female | SE_VUMC_male |
|---|---|---|---|---|---|---|---|---|---|---|---|---|---|---|---|---|---|---|---|---|---|---|
| RDW | rs1611657 | rs3128894 | A | C | 6 | HCG4P7 | Female only | False | 0.3466 | 0.3483 | 0.335 | 0.3339 | 35661 | 26955 | 188075 | 162398 | -0.02427 | -0.01007 | -0.03003 | -0.0282 | 0.007207 | 0.008826 |
| RDW | rs1611657 | rs9258940 | C | T | 6 | HCG4P7 | Female only | False | 0.3466 | 0.3483 | 0.335 | 0.3339 | 35661 | 26955 | 188075 | 162398 | -0.02427 | -0.01007 | -0.03003 | -0.02819 | 0.007207 | 0.008826 |
| RDW | rs1611657 | rs3094655 | A | G | 6 | HCG4P7 | Female only | False | 0.3466 | 0.3483 | 0.3348 | 0.3337 | 35661 | 26955 | 188075 | 162398 | -0.02427 | -0.01008 | -0.03003 | -0.02815 | 0.007207 | 0.008826 |
| RDW | rs1611657 | rs3094654 | T | C | 6 | HCG4P7 | Female only | False | 0.346 | 0.3477 | 0.3347 | 0.3336 | 35661 | 26955 | 188075 | 162398 | -0.0242 | -0.01008 | -0.03001 | -0.02817 | 0.0072 | 0.008817 |
| RDW | rs1611657 | rs3128893 | G | A | 6 | HCG4P7 | Female only | False | 0.3466 | 0.3483 | 0.3347 | 0.3336 | 35661 | 26955 | 188075 | 162398 | -0.02427 | -0.01008 | -0.03001 | -0.02815 | 0.007207 | 0.008826 |
| RDW | rs1611657 | rs3094150 | A | G | 6 | HCG4P7 | Female only | False | 0.3466 | 0.3483 | 0.3347 | 0.3336 | 35661 | 26955 | 188075 | 162398 | -0.02427 | -0.01008 | -0.03003 | -0.02816 | 0.007207 | 0.008826 |
| RDW | rs1611657 | rs3128892 | G | A | 6 | HCG4P7 | Female only | False | 0.3466 | 0.3483 | 0.3347 | 0.3336 | 35661 | 26955 | 188075 | 162398 | -0.02427 | -0.01008 | -0.03003 | -0.02815 | 0.007207 | 0.008826 |
| RDW | rs1611657 | rs3115622 | T | C | 6 | HCG4P7 | Female only | False | 0.3466 | 0.3483 | 0.3347 | 0.3336 | 35661 | 26955 | 188075 | 162398 | -0.02427 | -0.01007 | -0.03003 | -0.02815 | 0.007207 | 0.008826 |
| RDW | rs1611657 | rs3128891 | G | A | 6 | HCG4P7 | Female only | False | 0.3466 | 0.3483 | 0.3347 | 0.3336 | 35661 | 26955 | 188075 | 162398 | -0.02427 | -0.01007 | -0.03002 | -0.02815 | 0.007207 | 0.008826 |
| RDW | rs1611657 | rs9258966 | C | T | 6 | HCG4P7 | Female only | False | 0.3466 | 0.3483 | 0.3347 | 0.3336 | 35661 | 26955 | 188075 | 162398 | -0.02427 | -0.01007 | -0.02999 | -0.02817 | 0.007207 | 0.008826 |
| RDW | rs1611657 | rs3094148 | A | G | 6 | HCG4P7 | Female only | False | 0.3466 | 0.3483 | 0.3347 | 0.3336 | 35661 | 26955 | 188075 | 162398 | -0.02426 | -0.01008 | -0.03003 | -0.02815 | 0.007207 | 0.008826 |
| RDW | rs1611657 | rs3132715 | T | C | 6 | HCG4P7 | Female only | False | 0.3466 | 0.3483 | 0.3347 | 0.3336 | 35661 | 26955 | 188075 | 162398 | -0.02426 | -0.01008 | -0.02999 | -0.02815 | 0.007207 | 0.008826 |
| RDW | rs1611657 | rs3129008 | C | T | 6 | HCG4P7 | Female only | False | 0.3466 | 0.3483 | 0.3347 | 0.3336 | 35661 | 26955 | 188075 | 162398 | -0.02426 | -0.01008 | -0.03003 | -0.02815 | 0.007207 | 0.008826 |
| RDW | rs1611657 | rs3132714 | T | C | 6 | HCG4P7 | Female only | False | 0.346 | 0.3477 | 0.3347 | 0.3336 | 35661 | 26955 | 188075 | 162398 | -0.0242 | -0.01009 | -0.02999 | -0.02825 | 0.0072 | 0.008817 |
| RDW | rs1611657 | rs2394227 | G | A | 6 | HCG4P7 | Female only | False | 0.3466 | 0.3483 | 0.3347 | 0.3335 | 35661 | 26955 | 188075 | 162398 | -0.02428 | -0.01009 | -0.03003 | -0.02816 | 0.007207 | 0.008826 |
| RDW | rs1611657 | rs2893991 | C | T | 6 | HCG4P7 | Female only | False | 0.3466 | 0.3483 | 0.3347 | 0.3335 | 35661 | 26955 | 188075 | 162398 | -0.02428 | -0.01009 | -0.03003 | -0.02816 | 0.007207 | 0.008826 |
| RDW | rs1611657 | rs3132712 | A | G | 6 | HCG4P7 | Female only | False | 0.3466 | 0.3483 | 0.3347 | 0.3335 | 35661 | 26955 | 188075 | 162398 | -0.02426 | -0.01008 | -0.03003 | -0.02816 | 0.007207 | 0.008826 |
| RDW | rs1611657 | rs3132710 | A | G | 6 | HCG4P7 | Female only | False | 0.3466 | 0.3483 | 0.3347 | 0.3336 | 35661 | 26955 | 188075 | 162398 | -0.02427 | -0.01007 | -0.03001 | -0.02815 | 0.007207 | 0.008826 |
| RDW | rs1611657 | rs3132709 | A | G | 6 | HCG4P7 | Female only | False | 0.3466 | 0.3483 | 0.3347 | 0.3336 | 35661 | 26955 | 188075 | 162398 | -0.02431 | -0.01008 | -0.02999 | -0.02815 | 0.007207 | 0.008826 |
| RDW | rs1611657 | rs3129007 | C | T | 6 | HCG4P7 | Female only | False | 0.3466 | 0.3483 | 0.3347 | 0.3336 | 35661 | 26955 | 188075 | 162398 | -0.02427 | -0.01006 | -0.03002 | -0.02816 | 0.007207 | 0.008826 |
| RDW | rs1611657 | rs3132708 | A | G | 6 | HCG4P7 | Female only | False | 0.3469 | 0.3486 | 0.3347 | 0.3336 | 35661 | 26955 | 188075 | 162398 | -0.02444 | -0.009928 | -0.03002 | -0.02817 | 0.007209 | 0.008829 |
| RDW | rs1611657 | rs9258993 | T | C | 6 | HCG4P7 | Female only | False | 0.3472 | 0.3489 | 0.3346 | 0.3335 | 35661 | 26955 | 188075 | 162398 | -0.02441 | -0.009894 | -0.03004 | -0.02827 | 0.007214 | 0.008835 |
| RDW | rs1611657 | rs9258995 | G | A | 6 | HCG4P7 | Female only | False | 0.3468 | 0.3485 | 0.3347 | 0.3336 | 35661 | 26955 | 188075 | 162398 | -0.02439 | -0.009928 | -0.03001 | -0.02814 | 0.00721 | 0.008829 |
| RDW | rs1611657 | rs9258996 | C | T | 6 | HCG4P7 | Female only | False | 0.3468 | 0.3485 | 0.3347 | 0.3336 | 35661 | 26955 | 188075 | 162398 | -0.0244 | -0.009925 | -0.03002 | -0.02815 | 0.00721 | 0.00883 |
| RDW | rs1611657 | rs9258997 | G | A | 6 | HCG4P7 | Female only | False | 0.3468 | 0.3485 | 0.3347 | 0.3336 | 35661 | 26955 | 188075 | 162398 | -0.0244 | -0.009926 | -0.03002 | -0.02815 | 0.00721 | 0.00883 |
| RDW | rs1611657 | rs9258999 | G | A | 6 | HCG4P7 | Female only | False | 0.3466 | 0.3483 | 0.3347 | 0.3336 | 35661 | 26955 | 188075 | 162398 | -0.02428 | -0.01006 | -0.03001 | -0.02816 | 0.007208 | 0.008827 |
| RDW | rs1611657 | rs9259010 | C | T | 6 | HCG4P7 | Female only | False | 0.3466 | 0.3483 | 0.3347 | 0.3336 | 35661 | 26955 | 188075 | 162398 | -0.02428 | -0.01007 | -0.03 | -0.02815 | 0.007208 | 0.008826 |

| trait | lead_snp | ID | ALLELE1 | ALLELE0 | CHR | Gene_Symbol | independent_loci_in_sex_P1e2 | in_embl_any_or_gene | A1FREQ_VUMC_female | A1FREQ_VUMC_male | A1FREQ_UKB_female | A1FREQ_UKB_male | N_VUMC_female | N_VUMC_male | N_UKB_female | N_UKB_male | BETA_VUMC_female | BETA_VUMC_male | BETA_UKB_female | BETA_UKB_male | SE_VUMC_female | SE_VUMC_male |
|---|---|---|---|---|---|---|---|---|---|---|---|---|---|---|---|---|---|---|---|---|---|---|
| RDW | rs1611657 | rs1611657 | G | A | 6 | HCG4P7 | Female only | False | 0.4737 | 0.4778 | 0.4561 | 0.4553 | 35661 | 26955 | 188075 | 162398 | -0.03834 | -0.01428 | -0.04067 | -0.04168 | 0.00687 | 0.00844 |
| RDW | rs1611657 | rs3115620 | C | A | 6 | HCG4P7 | Female only | False | 0.3466 | 0.3483 | 0.3347 | 0.3336 | 35661 | 26955 | 188075 | 162398 | -0.02428 | -0.01006 | -0.03001 | -0.02815 | 0.007208 | 0.008826 |
| RDW | rs1611657 | rs3128886 | C | A | 6 | HCG4P7 | Female only | False | 0.3466 | 0.3483 | 0.3347 | 0.3336 | 35661 | 26955 | 188075 | 162398 | -0.02428 | -0.01007 | -0.03001 | -0.02815 | 0.007208 | 0.008826 |
| RDW | rs1611657 | rs3115619 | A | G | 6 | HCG4P7 | Female only | False | 0.3466 | 0.3483 | 0.3347 | 0.3336 | 35661 | 26955 | 188075 | 162398 | -0.02428 | -0.01007 | -0.02995 | -0.02818 | 0.007208 | 0.008826 |
| RDW | rs1611657 | rs9259034 | T | C | 6 | HCG4P7 | Female only | False | 0.3468 | 0.3484 | 0.3395 | 0.3385 | 35661 | 26955 | 188075 | 162398 | -0.02423 | -0.01016 | -0.02996 | -0.02807 | 0.00721 | 0.008828 |
| RDW | rs1611657 | rs3132705 | C | A | 6 | HCG4P7 | Female only | False | 0.3467 | 0.3483 | 0.335 | 0.3339 | 35661 | 26955 | 188075 | 162398 | -0.02437 | -0.01007 | -0.02978 | -0.02824 | 0.007213 | 0.008833 |
| RDW | rs1611657 | rs3132704 | A | G | 6 | HCG4P7 | Female only | False | 0.3467 | 0.3484 | 0.335 | 0.3339 | 35661 | 26955 | 188075 | 162398 | -0.02431 | -0.01006 | -0.02979 | -0.02824 | 0.007213 | 0.008833 |
| RDW | rs1611657 | rs77297191 | A | G | 6 | HCG4P7 | Female only | False | 0.3463 | 0.348 | | | 35661 | 26955 | | | -0.02454 | -0.01034 | | | 0.00722 | 0.008843 |
| RDW | rs1611657 | rs1655901 | T | C | 6 | HCG4P7 | Female only | False | 0.4805 | 0.4826 | 0.4393 | 0.4382 | 35661 | 26955 | 188075 | 162398 | -0.03278 | -0.006897 | -0.03887 | -0.04127 | 0.006895 | 0.008474 |
| RDW | rs74979599 | rs11023877 | C | T | 11 | SOX6 | Female only | True | 0.07737 | 0.07851 | 0.0845 | 0.08292 | 35661 | 26955 | 188075 | 162398 | 0.07191 | 0.02566 | 0.09229 | 0.09148 | 0.01288 | 0.01571 |
| RDW | rs74979599 | rs11023878 | A | G | 11 | SOX6 | Female only | True | 0.07736 | 0.07853 | 0.0845 | 0.08292 | 35661 | 26955 | 188075 | 162398 | 0.07183 | 0.02592 | 0.0923 | 0.09148 | 0.01288 | 0.0157 |
| RDW | rs74979599 | rs11023883 | G | A | 11 | SOX6 | Female only | True | 0.07639 | 0.07801 | 0.08352 | 0.08179 | 35661 | 26955 | 188075 | 162398 | 0.0758 | 0.02756 | 0.1003 | 0.09689 | 0.01294 | 0.01578 |
| RDW | rs74979599 | rs17539593 | A | C | 11 | SOX6 | Female only | True | 0.07674 | 0.0784 | 0.08376 | 0.08209 | 35661 | 26955 | 188075 | 162398 | 0.07642 | 0.02772 | 0.1 | 0.09675 | 0.0129 | 0.01572 |
| RDW | rs74979599 | rs11023888 | G | T | 11 | SOX6 | Female only | True | 0.07643 | 0.07806 | 0.08354 | 0.08181 | 35661 | 26955 | 188075 | 162398 | 0.07591 | 0.02685 | 0.1003 | 0.09701 | 0.01293 | 0.01577 |
| RDW | rs74979599 | rs78243491 | C | T | 11 | SOX6 | Female only | True | 0.07727 | 0.07896 | 0.08378 | 0.08211 | 35661 | 26955 | 188075 | 162398 | 0.07568 | 0.02604 | 0.1001 | 0.09688 | 0.01281 | 0.01559 |
| RDW | rs74979599 | rs11512059 | T | C | 11 | SOX6 | Female only | True | 0.07676 | 0.07842 | 0.08378 | 0.08211 | 35661 | 26955 | 188075 | 162398 | 0.07629 | 0.02652 | 0.1001 | 0.09694 | 0.01289 | 0.0157 |
| RDW | rs74979599 | rs11023895 | A | G | 11 | SOX6 | Female only | True | 0.07668 | 0.07835 | 0.08365 | 0.08199 | 35661 | 26955 | 188075 | 162398 | 0.07618 | 0.02635 | 0.1005 | 0.09703 | 0.01291 | 0.01571 |
| RDW | rs74979599 | rs74375633 | T | C | 11 | SOX6 | Female only | True | 0.07686 | 0.07845 | 0.08384 | 0.08217 | 35661 | 26955 | 188075 | 162398 | 0.07644 | 0.02612 | 0.1002 | 0.09693 | 0.01289 | 0.0157 |
| RDW | rs74979599 | rs11023905 | C | T | 11 | SOX6 | Female only | True | 0.109 | 0.109 | 0.1142 | 0.112 | 35661 | 26955 | 188075 | 162398 | 0.05759 | 0.01924 | 0.07172 | 0.07312 | 0.01096 | 0.01347 |
| RDW | rs74979599 | rs78936881 | T | C | 11 | SOX6 | Female only | True | 0.0774 | 0.07898 | 0.08393 | 0.08223 | 35661 | 26955 | 188075 | 162398 | 0.07605 | 0.02855 | 0.09952 | 0.0968 | 0.01285 | 0.01564 |
| RDW | rs74979599 | rs16932896 | T | C | 11 | SOX6 | Female only | True | 0.1066 | 0.1068 | 0.1127 | 0.1106 | 35661 | 26955 | 188075 | 162398 | 0.05815 | 0.02119 | 0.07172 | 0.07364 | 0.01117 | 0.01369 |
| RDW | rs74979599 | rs11023911 | T | G | 11 | SOX6 | Female only | True | 0.07728 | 0.07885 | 0.08397 | 0.08227 | 35661 | 26955 | 188075 | 162398 | 0.07626 | 0.029 | 0.09959 | 0.09679 | 0.01289 | 0.0157 |
| RDW | rs74979599 | rs12281323 | G | A | 11 | SOX6 | Female only | True | 0.1077 | 0.1079 | 0.1139 | 0.1118 | 35661 | 26955 | 188075 | 162398 | 0.05798 | 0.01939 | 0.07175 | 0.07382 | 0.01112 | 0.01364 |
| RDW | rs74979599 | rs75817865 | T | C | 11 | SOX6 | Female only | True | 0.0772 | 0.0788 | 0.08395 | 0.08226 | 35661 | 26955 | 188075 | 162398 | 0.0766 | 0.02907 | 0.09952 | 0.09681 | 0.01294 | 0.01575 |
| RDW | rs74979599 | rs1393923 | G | A | 11 | SOX6 | Female only | True | 0.1081 | 0.1081 | 0.1142 | 0.112 | 35661 | 26955 | 188075 | 162398 | 0.05839 | 0.02025 | 0.07175 | 0.07311 | 0.0111 | 0.01362 |
| RDW | rs74979599 | rs11023914 | C | T | 11 | SOX6 | Female only | True | 0.1081 | 0.1082 | 0.1142 | 0.112 | 35661 | 26955 | 188075 | 162398 | 0.05826 | 0.02025 | 0.0718 | 0.07309 | 0.0111 | 0.01362 |
| RDW | rs74979599 | rs11023919 | G | A | 11 | SOX6 | Female only | True | 0.077 | 0.07856 | 0.08393 | 0.08224 | 35661 | 26955 | 188075 | 162398 | 0.07763 | 0.0288 | 0.09956 | 0.09688 | 0.01307 | 0.01589 |
| RDW | rs74979599 | rs11023922 | T | C | 11 | SOX6 | Female only | True | 0.07697 | 0.07853 | 0.08386 | 0.08225 | 35661 | 26955 | 188075 | 162398 | 0.07786 | 0.02881 | 0.09954 | 0.09749 | 0.01307 | 0.01589 |
| RDW | rs74979599 | rs74979599 | G | A | 11 | SOX6 | Female only | True | 0.07694 | 0.07843 | 0.08391 | 0.08218 | 35661 | 26955 | 188075 | 162398 | 0.07841 | 0.02902 | 0.09998 | 0.09716 | 0.0131 | 0.01593 |

| trait | lead_snp | ID | ALLELE1 | ALLELE0 | CHR | Gene_Symbol | independent_loci_in_sex_P1e2 | in_embl_any_or_gene | A1FREQ_VUMC_female | A1FREQ_VUMC_male | A1FREQ_UKB_female | A1FREQ_UKB_male | N_VUMC_female | N_VUMC_male | N_UKB_female | N_UKB_male | BETA_VUMC_female | BETA_VUMC_male | BETA_UKB_female | BETA_UKB_male | SE_VUMC_female | SE_VUMC_male |
|---|---|---|---|---|---|---|---|---|---|---|---|---|---|---|---|---|---|---|---|---|---|---|
| RDW | rs837763 | rs8052231 | C | T | 16 | PIEZO1 | Female only | True | 0.3932 | 0.3923 | 0.3972 | 0.3962 | 35661 | 26955 | 188075 | 162398 | -0.03375 | -0.003519 | -0.01887 | -0.002868 | 0.007022 | 0.008594 |
| RDW | rs837763 | rs8052370 | C | T | 16 | PIEZO1 | Female only | True | 0.39 | 0.3886 | 0.3919 | 0.3908 | 35661 | 26955 | 188075 | 162398 | -0.03205 | -0.003056 | -0.02019 | -0.003108 | 0.007058 | 0.008641 |
| RDW | rs837763 | rs10445033 | G | A | 16 | PIEZO1 | Female only | True | 0.3899 | 0.3881 | 0.3926 | 0.3917 | 35661 | 26955 | 188075 | 162398 | -0.03234 | -0.004147 | -0.01945 | -0.00532 | 0.007064 | 0.008659 |
| RDW | rs837763 | rs2002833 | G | A | 16 | PIEZO1 | Female only | True | 0.3215 | 0.3202 | 0.3269 | 0.3268 | 35661 | 26955 | 188075 | 162398 | -0.02191 | 0.00605 | -0.01806 | -0.001805 | 0.007404 | 0.009022 |
| RDW | rs837763 | rs2911460 | A | G | 16 | PIEZO1 | Female only | True | 0.3097 | 0.3077 | 0.3171 | 0.3169 | 35661 | 26955 | 188075 | 162398 | -0.02124 | 0.006026 | -0.02014 | -0.004048 | 0.007482 | 0.009132 |
| RDW | rs837763 | rs2608604 | A | G | 16 | PIEZO1 | Female only | True | 0.3779 | 0.3767 | 0.3852 | 0.3841 | 35661 | 26955 | 188075 | 162398 | -0.03385 | -0.00319 | -0.02072 | -0.004885 | 0.007073 | 0.008654 |
| RDW | rs837763 | rs837763 | C | T | 16 | PIEZO1 | Female only | True | 0.4376 | 0.4384 | 0.4463 | 0.4452 | 35661 | 26955 | 188075 | 162398 | -0.04127 | -0.01175 | -0.02506 | -0.002675 | 0.006909 | 0.008418 |
| RDW | rs837763 | rs2926776 | A | G | 16 | PIEZO1 | Female only | True | 0.4159 | 0.4161 | 0.4254 | 0.4242 | 35661 | 26955 | 188075 | 162398 | -0.03829 | -0.005259 | -0.02406 | -0.002605 | 0.006947 | 0.008481 |
| RDW | rs837763 | rs2968478 | T | G | 16 | PIEZO1 | Female only | True | 0.4133 | 0.413 | 0.4195 | 0.4182 | 35661 | 26955 | 188075 | 162398 | -0.03699 | -0.00529 | -0.02515 | -0.003231 | 0.006982 | 0.008519 |
| WBC | rs9358938 | rs17598658 | C | A | 6 | BTN2A2 | Male only | True | 0.08033 | 0.08336 | 0.1053 | 0.1049 | 35673 | 26968 | 188074 | 162396 | -0.02032 | -0.05988 | -0.05148 | -0.05444 | 0.01251 | 0.01386 |
| WBC | rs9358938 | rs34961555 | T | C | 6 | BTN2A2 | Male only | True | 0.07989 | 0.083 | 0.105 | 0.1046 | 35673 | 26968 | 188074 | 162396 | -0.01964 | -0.06299 | -0.05226 | -0.05499 | 0.01259 | 0.01394 |
| WBC | rs9358938 | rs55706012 | C | A | 6 | BTN2A2 | Male only | True | 0.0816 | 0.08435 | 0.1064 | 0.1062 | 35673 | 26968 | 188074 | 162396 | -0.01963 | -0.0643 | -0.05418 | -0.05641 | 0.01251 | 0.0139 |
| WBC | rs9358938 | rs10484439 | A | G | 6 | BTN2A2 | Male only | True | 0.08452 | 0.08804 | 0.1088 | 0.1084 | 35673 | 26968 | 188074 | 162396 | -0.0183 | -0.06173 | -0.05358 | -0.05561 | 0.01221 | 0.01351 |
| WBC | rs9358938 | rs13220522 | A | G | 6 | BTN2A2 | Male only | True | 0.081 | 0.084 | 0.1064 | 0.1062 | 35673 | 26968 | 188074 | 162396 | -0.02017 | -0.06344 | -0.05421 | -0.05677 | 0.01246 | 0.01383 |
| WBC | rs9358938 | rs35162296 | T | C | 6 | BTN2A2 | Male only | True | 0.08454 | 0.088 | 0.1088 | 0.1085 | 35673 | 26968 | 188074 | 162396 | -0.01806 | -0.06203 | -0.05374 | -0.05577 | 0.01221 | 0.01353 |
| WBC | rs9358938 | rs34107459 | C | T | 6 | BTN2A2 | Male only | True | 0.08098 | 0.08401 | 0.1065 | 0.1063 | 35673 | 26968 | 188074 | 162396 | -0.02069 | -0.06429 | -0.0542 | -0.0573 | 0.01247 | 0.01383 |
| WBC | rs9358938 | rs34158769 | A | G | 6 | BTN2A2 | Male only | True | 0.08393 | 0.08709 | 0.1083 | 0.1081 | 35673 | 26968 | 188074 | 162396 | -0.02108 | -0.0601 | -0.05392 | -0.05571 | 0.0123 | 0.01365 |
| WBC | rs9358938 | rs35942569 | G | A | 6 | BTN2A2 | Male only | True | 0.08535 | 0.08819 | 0.1088 | 0.1087 | 35673 | 26968 | 188074 | 162396 | -0.02133 | -0.05893 | -0.05395 | -0.0552 | 0.01221 | 0.01356 |
| WBC | rs9358938 | rs67509210 | T | G | 6 | BTN2A2 | Male only | True | 0.1112 | 0.113 | 0.1343 | 0.1346 | 35673 | 26968 | 188074 | 162396 | -0.01802 | -0.06128 | -0.05174 | -0.05422 | 0.01078 | 0.01215 |
| WBC | rs9358938 | rs9366653 | A | G | 6 | BTN2A2 | Male only | True | 0.1113 | 0.1131 | 0.1343 | 0.1346 | 35673 | 26968 | 188074 | 162396 | -0.01794 | -0.06104 | -0.05174 | -0.0542 | 0.01077 | 0.01214 |
| WBC | rs9358938 | rs9379851 | C | A | 6 | BTN2A2 | Male only | True | 0.1113 | 0.1131 | 0.1351 | 0.1352 | 35673 | 26968 | 188074 | 162396 | -0.01807 | -0.06086 | -0.0521 | -0.05405 | 0.01077 | 0.01214 |
| WBC | rs9358938 | rs35934643 | A | G | 6 | BTN2A2 | Male only | True | 0.1112 | 0.113 | 0.1343 | 0.1346 | 35673 | 26968 | 188074 | 162396 | -0.01808 | -0.06138 | -0.05172 | -0.05413 | 0.01078 | 0.01215 |
| WBC | rs3799378 | rs6933583 | C | A | 6 | BTN3A1 | Male only | False | 0.28 | 0.2808 | 0.2942 | 0.2942 | 35673 | 26968 | 188074 | 162396 | 0.00348 | -0.03827 | -0.02095 | -0.02172 | 0.007522 | 0.008498 |
| WBC | rs3799378 | rs4712980 | G | T | 6 | BTN3A1 | Male only | False | 0.28 | 0.2808 | 0.2945 | 0.2944 | 35673 | 26968 | 188074 | 162396 | 0.003514 | -0.03827 | -0.02079 | -0.02144 | 0.007522 | 0.008498 |
| WBC | rs9358938 | rs35304979 | A | C | 6 | BTN2A2 | Male only | True | 0.1112 | 0.113 | 0.1343 | 0.1346 | 35673 | 26968 | 188074 | 162396 | -0.01807 | -0.06089 | -0.05171 | -0.05417 | 0.01078 | 0.01215 |
| WBC | rs3799378 | rs9467725 | G | A | 6 | BTN3A1 | Male only | False | 0.2797 | 0.2805 | 0.2943 | 0.2942 | 35673 | 26968 | 188074 | 162396 | 0.003497 | -0.03832 | -0.02098 | -0.02167 | 0.007529 | 0.008506 |
| WBC | rs9358938 | rs71557332 | T | C | 6 | BTN2A2 | Male only | True | 0.1112 | 0.113 | 0.1343 | 0.1346 | 35673 | 26968 | 188074 | 162396 | -0.01807 | -0.06089 | -0.05171 | -0.05416 | 0.01078 | 0.01215 |
| WBC | rs9358938 | rs9393703 | C | T | 6 | BTN2A2 | Male only | True | 0.1113 | 0.1131 | 0.1343 | 0.1346 | 35673 | 26968 | 188074 | 162396 | -0.01794 | -0.06079 | -0.05179 | -0.05419 | 0.01078 | 0.01215 |
| WBC | rs9358938 | rs9358930 | T | C | 6 | BTN2A2 | Male only | True | 0.1112 | 0.1131 | 0.1343 | 0.1346 | 35673 | 26968 | 188074 | 162396 | -0.01797 | -0.06068 | -0.05179 | -0.05419 | 0.01078 | 0.01215 |

| trait | lead_snp | ID | ALLELE1 | ALLELE0 | CHR | Gene_Symbol | independent_loci_in_sex_P1e2 | in_embl_any_or_gene | A1FREQ_VUMC_female | A1FREQ_VUMC_male | A1FREQ_UKB_female | A1FREQ_UKB_male | N_VUMC_female | N_VUMC_male | N_UKB_female | N_UKB_male | BETA_VUMC_female | BETA_VUMC_male | BETA_UKB_female | BETA_UKB_male | SE_VUMC_female | SE_VUMC_male |
|---|---|---|---|---|---|---|---|---|---|---|---|---|---|---|---|---|---|---|---|---|---|---|
| WBC | rs9358938 | rs9379852 | G | A | 6 | BTN2A2 | Male only | True | 0.1112 | 0.113 | 0.1342 | 0.1345 | 35673 | 26968 | 188074 | 162396 | -0.0181 | -0.06129 | -0.05165 | -0.05414 | 0.01078 | 0.01215 |
| WBC | rs9358938 | rs9379853 | G | A | 6 | BTN2A2 | Male only | True | 0.1112 | 0.113 | 0.1342 | 0.1345 | 35673 | 26968 | 188074 | 162396 | -0.0181 | -0.06129 | -0.05165 | -0.05414 | 0.01078 | 0.01215 |
| trait | lead_snp | ID | ALLELE1 | ALLELE0 | CHR | Gene_Symbol | independent_loci_in_sex_P1e2 | in_embl_any_or_gene | A1FREQ_VUMC_female | A1FREQ_VUMC_male | A1FREQ_UKB_female | A1FREQ_UKB_male | N_VUMC_female | N_VUMC_male | N_UKB_female | N_UKB_male | BETA_VUMC_female | BETA_VUMC_male | BETA_UKB_female | BETA_UKB_male | SE_VUMC_female | SE_VUMC_male |
| WBC | rs9358938 | rs9393704 | C | T | 6 | BTN2A2 | Male only | True | 0.1112 | 0.113 | 0.1342 | 0.1345 | 35673 | 26968 | 188074 | 162396 | -0.0181 | -0.06129 | -0.05167 | -0.05413 | 0.01078 | 0.01215 |
| WBC | rs9358938 | rs9348709 | A | G | 6 | BTN2A2 | Male only | True | 0.1112 | 0.113 | 0.1341 | 0.1345 | 35673 | 26968 | 188074 | 162396 | -0.01815 | -0.06121 | -0.05156 | -0.05412 | 0.01078 | 0.01215 |
| WBC | rs3799378 | rs9348711 | G | A | 6 | BTN3A1 | Male only | False | 0.2415 | 0.2425 | 0.2581 | 0.2576 | 35673 | 26968 | 188074 | 162396 | -0.0056 | -0.04397 | -0.02616 | -0.02893 | 0.007897 | 0.00894 |
| WBC | rs9358938 | rs9393705 | A | G | 6 | BTN2A2 | Male only | True | 0.1113 | 0.1131 | 0.134 | 0.1343 | 35673 | 26968 | 188074 | 162396 | -0.01817 | -0.06115 | -0.05122 | -0.05398 | 0.01077 | 0.01214 |
| WBC | rs3799378 | rs4712981 | T | C | 6 | BTN3A1 | Male only | False | 0.2783 | 0.2791 | 0.2929 | 0.2929 | 35673 | 26968 | 188074 | 162396 | 0.002708 | -0.03806 | -0.02086 | -0.0219 | 0.007539 | 0.008515 |
| WBC | rs9358938 | rs9393706 | A | G | 6 | BTN2A2 | Male only | True | 0.1113 | 0.1131 | 0.1341 | 0.1344 | 35673 | 26968 | 188074 | 162396 | -0.01817 | -0.06115 | -0.05156 | -0.05411 | 0.01077 | 0.01214 |
| WBC | rs9358938 | rs34550936 | G | A | 6 | BTN2A2 | Male only | True | 0.1112 | 0.1131 | 0.1341 | 0.1344 | 35673 | 26968 | 188074 | 162396 | -0.01817 | -0.06118 | -0.05156 | -0.05411 | 0.01078 | 0.01214 |
| WBC | rs9358938 | rs9393707 | C | T | 6 | BTN2A2 | Male only | True | 0.1112 | 0.1131 | 0.1341 | 0.1344 | 35673 | 26968 | 188074 | 162396 | -0.01811 | -0.06118 | -0.05156 | -0.05411 | 0.01078 | 0.01214 |
| WBC | rs9358938 | rs9393708 | C | T | 6 | BTN2A2 | Male only | True | 0.1112 | 0.1131 | 0.1341 | 0.1345 | 35673 | 26968 | 188074 | 162396 | -0.01809 | -0.06116 | -0.05152 | -0.0541 | 0.01077 | 0.01214 |
| WBC | rs9358938 | rs9358932 | T | C | 6 | BTN2A2 | Male only | True | 0.1112 | 0.1131 | 0.1341 | 0.1344 | 35673 | 26968 | 188074 | 162396 | -0.01811 | -0.06118 | -0.05156 | -0.05411 | 0.01078 | 0.01214 |
| WBC | rs9358938 | rs9379854 | C | T | 6 | BTN2A2 | Male only | True | 0.1112 | 0.1131 | 0.1341 | 0.1344 | 35673 | 26968 | 188074 | 162396 | -0.01811 | -0.06118 | -0.05156 | -0.05411 | 0.01078 | 0.01214 |
| WBC | rs3799378 | rs9358933 | T | C | 6 | BTN3A1 | Male only | False | 0.2781 | 0.2789 | 0.2929 | 0.2929 | 35673 | 26968 | 188074 | 162396 | 0.002687 | -0.03808 | -0.02096 | -0.02188 | 0.007545 | 0.008522 |
| WBC | rs9358938 | rs9357006 | C | A | 6 | BTN2A2 | Male only | True | 0.1114 | 0.1132 | 0.1342 | 0.1345 | 35673 | 26968 | 188074 | 162396 | -0.01756 | -0.06194 | -0.05159 | -0.05403 | 0.01077 | 0.01214 |
| WBC | rs9358938 | rs9379855 | C | T | 6 | BTN2A2 | Male only | True | 0.1114 | 0.1132 | 0.1343 | 0.1346 | 35673 | 26968 | 188074 | 162396 | -0.01756 | -0.06194 | -0.0515 | -0.05384 | 0.01077 | 0.01214 |
| WBC | rs9358938 | rs28362606 | C | T | 6 | BTN2A2 | Male only | True | 0.1115 | 0.1133 | 0.1341 | 0.1344 | 35673 | 26968 | 188074 | 162396 | -0.01743 | -0.06203 | -0.0514 | -0.05388 | 0.01077 | 0.01213 |
| WBC | rs9358938 | rs58825580 | G | T | 6 | BTN2A2 | Male only | True | 0.1191 | 0.1203 | 0.1414 | 0.1413 | 35673 | 26968 | 188074 | 162396 | -0.01189 | -0.05478 | -0.04621 | -0.05057 | 0.01046 | 0.01182 |
| WBC | rs9358938 | rs66827971 | C | T | 6 | BTN2A2 | Male only | True | 0.1115 | 0.1133 | 0.1341 | 0.1344 | 35673 | 26968 | 188074 | 162396 | -0.01743 | -0.06203 | -0.0514 | -0.05387 | 0.01077 | 0.01213 |
| WBC | rs9358938 | rs71557334 | T | G | 6 | BTN2A2 | Male only | True | 0.1115 | 0.1133 | 0.1341 | 0.1344 | 35673 | 26968 | 188074 | 162396 | -0.01743 | -0.06203 | -0.0514 | -0.05387 | 0.01077 | 0.01213 |
| WBC | rs9358938 | rs9379856 | C | A | 6 | BTN2A2 | Male only | True | 0.1115 | 0.1133 | 0.1342 | 0.1344 | 35673 | 26968 | 188074 | 162396 | -0.0174 | -0.06178 | -0.05131 | -0.05384 | 0.01076 | 0.01213 |
| WBC | rs9358938 | rs9379858 | C | T | 6 | BTN2A2 | Male only | True | 0.1115 | 0.1133 | 0.1342 | 0.1345 | 35673 | 26968 | 188074 | 162396 | -0.0174 | -0.06178 | -0.0513 | -0.05371 | 0.01076 | 0.01213 |
| WBC | rs9358938 | rs9393710 | G | A | 6 | BTN2A2 | Male only | True | 0.1115 | 0.1133 | 0.1342 | 0.1345 | 35673 | 26968 | 188074 | 162396 | -0.0174 | -0.06178 | -0.05131 | -0.05385 | 0.01076 | 0.01213 |
| WBC | rs9358938 | rs71557335 | A | G | 6 | BTN2A2 | Male only | True | 0.1115 | 0.1133 | 0.1344 | 0.1347 | 35673 | 26968 | 188074 | 162396 | -0.0174 | -0.06178 | -0.05102 | -0.05358 | 0.01076 | 0.01213 |
| WBC | rs9358938 | rs9379859 | T | C | 6 | BTN2A2 | Male only | True | 0.1115 | 0.1132 | 0.1341 | 0.1344 | 35673 | 26968 | 188074 | 162396 | -0.0177 | -0.06187 | -0.05125 | -0.05387 | 0.01076 | 0.01213 |
| WBC | rs9358938 | rs9358935 | G | A | 6 | BTN2A2 | Male only | True | 0.1114 | 0.1132 | 0.1341 | 0.1344 | 35673 | 26968 | 188074 | 162396 | -0.01771 | -0.0619 | -0.05126 | -0.05387 | 0.01077 | 0.01213 |
| WBC | rs9358938 | rs35307327 | T | C | 6 | BTN2A2 | Male only | True | 0.1114 | 0.1132 | 0.1341 | 0.1344 | 35673 | 26968 | 188074 | 162396 | -0.01773 | -0.06183 | -0.05126 | -0.05387 | 0.01077 | 0.01213 |

| trait | lead_snp | ID | ALLELE1 | ALLELE0 | CHR | Gene_Symbol | independent_loci_in_sex_P1e2 | in_embl_any_or_gene | A1FREQ_VUMC_female | A1FREQ_VUMC_male | A1FREQ_UKB_female | A1FREQ_UKB_male | N_VUMC_female | N_VUMC_male | N_UKB_female | N_UKB_male | BETA_VUMC_female | BETA_VUMC_male | BETA_UKB_female | BETA_UKB_male | SE_VUMC_female | SE_VUMC_male |
|---|---|---|---|---|---|---|---|---|---|---|---|---|---|---|---|---|---|---|---|---|---|---|
| WBC | rs9358938 | rs34878490 | T | C | 6 | BTN2A2 | Male only | True | 0.1114 | 0.1132 | 0.1341 | 0.1344 | 35673 | 26968 | 188074 | 162396 | -0.01773 | -0.06185 | -0.05127 | -0.05387 | 0.01077 | 0.01214 |
| WBC | rs9358938 | rs9358936 | G | A | 6 | BTN2A2 | Male only | True | 0.1114 | 0.1132 | 0.1341 | 0.1344 | 35673 | 26968 | 188074 | 162396 | -0.01773 | -0.06183 | -0.05126 | -0.05387 | 0.01077 | 0.01213 |
| WBC | rs9358938 | rs9393711 | T | C | 6 | BTN2A2 | Male only | True | 0.1111 | 0.1129 | 0.1342 | 0.1345 | 35673 | 26968 | 188074 | 162396 | -0.01718 | -0.06148 | -0.05122 | -0.05372 | 0.01081 | 0.01219 |
| WBC | rs3799378 | rs9379862 | C | T | 6 | BTN3A1 | Male only | False | 0.2415 | 0.2424 | 0.2579 | 0.2575 | 35673 | 26968 | 188074 | 162396 | -0.005755 | -0.04379 | -0.02605 | -0.02878 | 0.007893 | 0.008935 |
| WBC | rs9358938 | rs12173854 | G | A | 6 | BTN2A2 | Male only | True | 0.1114 | 0.1132 | 0.1341 | 0.1344 | 35673 | 26968 | 188074 | 162396 | -0.01773 | -0.0618 | -0.0513 | -0.05386 | 0.01077 | 0.01214 |
| WBC | rs9358938 | rs9379863 | G | A | 6 | BTN2A2 | Male only | True | 0.1108 | 0.1125 | 0.1332 | 0.1334 | 35673 | 26968 | 188074 | 162396 | -0.01676 | -0.06063 | -0.05188 | -0.05395 | 0.0108 | 0.01216 |
| WBC | rs9358938 | rs9379864 | A | G | 6 | BTN2A2 | Male only | True | 0.1108 | 0.1125 | 0.1332 | 0.1334 | 35673 | 26968 | 188074 | 162396 | -0.01676 | -0.06063 | -0.05188 | -0.05395 | 0.0108 | 0.01216 |
| WBC | rs9358938 | rs12176317 | G | A | 6 | BTN2A2 | Male only | True | 0.1108 | 0.1125 | 0.1332 | 0.1335 | 35673 | 26968 | 188074 | 162396 | -0.01681 | -0.06037 | -0.05176 | -0.05378 | 0.01079 | 0.01216 |
| WBC | rs9358938 | rs12174602 | T | C | 6 | BTN2A2 | Male only | True | 0.1107 | 0.1125 | 0.1332 | 0.1334 | 35673 | 26968 | 188074 | 162396 | -0.01679 | -0.0604 | -0.05187 | -0.05391 | 0.0108 | 0.01217 |
| WBC | rs9358938 | rs12174623 | T | C | 6 | BTN2A2 | Male only | True | 0.1108 | 0.1125 | 0.1332 | 0.1335 | 35673 | 26968 | 188074 | 162396 | -0.01657 | -0.0603 | -0.05171 | -0.0539 | 0.0108 | 0.01216 |
| WBC | rs9358938 | rs12174639 | A | G | 6 | BTN2A2 | Male only | True | 0.1103 | 0.1121 | 0.1331 | 0.1333 | 35673 | 26968 | 188074 | 162396 | -0.01687 | -0.06014 | -0.05197 | -0.05403 | 0.01081 | 0.01219 |
| WBC | rs9358938 | rs12174631 | T | C | 6 | BTN2A2 | Male only | True | 0.1107 | 0.1125 | 0.1332 | 0.1334 | 35673 | 26968 | 188074 | 162396 | -0.01681 | -0.0604 | -0.05185 | -0.05395 | 0.0108 | 0.01217 |
| WBC | rs9358938 | rs13216828 | A | G | 6 | BTN2A2 | Male only | True | 0.1107 | 0.1125 | 0.1332 | 0.1334 | 35673 | 26968 | 188074 | 162396 | -0.01681 | -0.0604 | -0.05186 | -0.05395 | 0.0108 | 0.01217 |
| WBC | rs9358938 | rs9393713 | A | G | 6 | BTN2A2 | Male only | True | 0.1108 | 0.1125 | 0.1333 | 0.1335 | 35673 | 26968 | 188074 | 162396 | -0.01681 | -0.06037 | -0.05187 | -0.05361 | 0.01079 | 0.01216 |
| WBC | rs9358938 | rs9393714 | T | G | 6 | BTN2A2 | Male only | True | 0.1108 | 0.1125 | 0.1332 | 0.1335 | 35673 | 26968 | 188074 | 162396 | -0.01681 | -0.06037 | -0.05176 | -0.05368 | 0.01079 | 0.01216 |
| WBC | rs3799378 | rs9379870 | G | A | 6 | BTN3A1 | Male only | False | 0.2774 | 0.2785 | 0.2874 | 0.2876 | 35673 | 26968 | 188074 | 162396 | 0.002662 | -0.03884 | -0.0225 | -0.02226 | 0.007551 | 0.008523 |
| WBC | rs3799378 | rs733528 | G | A | 6 | BTN3A1 | Male only | False | 0.2783 | 0.2789 | 0.2926 | 0.2926 | 35673 | 26968 | 188074 | 162396 | 0.003574 | -0.03737 | -0.02103 | -0.02178 | 0.007543 | 0.008523 |
| WBC | rs9358938 | rs2073527 | C | T | 6 | BTN2A2 | Male only | True | 0.1107 | 0.1125 | 0.1332 | 0.1334 | 35673 | 26968 | 188074 | 162396 | -0.01682 | -0.0604 | -0.0519 | -0.05398 | 0.0108 | 0.01217 |
| WBC | rs9358938 | rs2073529 | G | A | 6 | BTN2A2 | Male only | True | 0.1108 | 0.1125 | 0.1332 | 0.1334 | 35673 | 26968 | 188074 | 162396 | -0.01681 | -0.06037 | -0.05191 | -0.05397 | 0.01079 | 0.01216 |
| WBC | rs9358938 | rs2073530 | C | T | 6 | BTN2A2 | Male only | True | 0.1107 | 0.1124 | 0.133 | 0.1332 | 35673 | 26968 | 188074 | 162396 | -0.01686 | -0.06041 | -0.05196 | -0.05404 | 0.0108 | 0.01217 |
| WBC | rs9358938 | rs2073531 | G | A | 6 | BTN2A2 | Male only | True | 0.1105 | 0.1122 | 0.1328 | 0.1331 | 35673 | 26968 | 188074 | 162396 | -0.01682 | -0.06016 | -0.05204 | -0.05405 | 0.01081 | 0.01218 |
| WBC | rs9358938 | rs9393715 | A | G | 6 | BTN2A2 | Male only | True | 0.1098 | 0.1115 | 0.1307 | 0.1308 | 35673 | 26968 | 188074 | 162396 | -0.01733 | -0.0617 | -0.05223 | -0.05424 | 0.01088 | 0.01226 |
| WBC | rs9358938 | rs9348716 | A | G | 6 | BTN2A2 | Male only | True | 0.1098 | 0.1115 | 0.1308 | 0.131 | 35673 | 26968 | 188074 | 162396 | -0.01733 | -0.0619 | -0.05216 | -0.05424 | 0.01088 | 0.01226 |
| WBC | rs9358938 | rs3757138 | G | A | 6 | BTN2A2 | Male only | True | 0.1108 | 0.1124 | 0.1332 | 0.1334 | 35673 | 26968 | 188074 | 162396 | -0.01653 | -0.06047 | -0.05182 | -0.05394 | 0.0108 | 0.01217 |
| WBC | rs9358938 | rs3047288 | G | A | 6 | BTN2A2 | Male only | True | 0.1108 | 0.1124 | 0.1332 | 0.1334 | 35673 | 26968 | 188074 | 162396 | -0.01653 | -0.06047 | -0.05186 | -0.05397 | 0.0108 | 0.01217 |
| WBC | rs9358938 | rs28551159 | G | A | 6 | BTN2A2 | Male only | True | 0.1108 | 0.1124 | 0.1332 | 0.1334 | 35673 | 26968 | 188074 | 162396 | -0.01653 | -0.06047 | -0.05187 | -0.05399 | 0.0108 | 0.01217 |
| WBC | rs3799378 | rs13218591 | C | T | 6 | BTN3A1 | Male only | False | 0.2892 | 0.2896 | 0.3033 | 0.3035 | 35673 | 26968 | 188074 | 162396 | 0.002637 | -0.03647 | -0.01953 | -0.01963 | 0.007457 | 0.008427 |
| WBC | rs9358938 | rs9393716 | G | A | 6 | BTN2A2 | Male only | True | 0.1846 | 0.1859 | 0.2026 | 0.2027 | 35673 | 26968 | 188074 | 162396 | -0.01396 | -0.03589 | -0.0354 | -0.03758 | 0.008777 | 0.00992 |
| WBC | rs9358938 | rs9366654 | A | G | 6 | BTN2A2 | Male only | True | 0.11 | 0.1116 | 0.1329 | 0.1331 | 35673 | 26968 | 188074 | 162396 | -0.0172 | -0.06031 | -0.05175 | -0.05411 | 0.01087 | 0.01225 |

| trait | lead_snp | ID | ALLELE1 | ALLELE0 | CHR | Gene_Symbol | independent_loci_in_sex_P1e2 | in_embl_any_or_gene | A1FREQ_VUMC_female | A1FREQ_VUMC_male | A1FREQ_UKB_female | A1FREQ_UKB_male | N_VUMC_female | N_VUMC_male | N_UKB_female | N_UKB_male | BETA_VUMC_female | BETA_VUMC_male | BETA_UKB_female | BETA_UKB_male | SE_VUMC_female | SE_VUMC_male |
|---|---|---|---|---|---|---|---|---|---|---|---|---|---|---|---|---|---|---|---|---|---|---|
| WBC | rs9358938 | rs1977 | G | A | 6 | BTN2A2 | Male only | True | 0.1109 | 0.1126 | 0.1359 | 0.1361 | 35673 | 26968 | 188074 | 162396 | -0.01686 | -0.06018 | -0.05085 | -0.05368 | 0.01079 | 0.01215 |
| WBC | rs9358938 | rs1979 | T | G | 6 | BTN2A2 | Male only | True | 0.1108 | 0.1125 | 0.1333 | 0.1335 | 35673 | 26968 | 188074 | 162396 | -0.01701 | -0.06021 | -0.05188 | -0.05385 | 0.0108 | 0.01216 |
| WBC | rs9358938 | rs68072215 | C | T | 6 | BTN2A2 | Male only | True | 0.1084 | 0.1098 | 0.1326 | 0.1327 | 35673 | 26968 | 188074 | 162396 | -0.01657 | -0.06149 | -0.05164 | -0.05423 | 0.01102 | 0.01245 |
| WBC | rs9358938 | rs66823108 | A | G | 6 | BTN2A2 | Male only | True | 0.1084 | 0.1097 | 0.1326 | 0.1328 | 35673 | 26968 | 188074 | 162396 | -0.01657 | -0.06151 | -0.05172 | -0.05401 | 0.01102 | 0.01245 |
| WBC | rs9358938 | rs9358938 | T | C | 6 | BTN2A2 | Male only | True | 0.1125 | 0.1143 | 0.1356 | 0.1357 | 35673 | 26968 | 188074 | 162396 | -0.01797 | -0.06804 | -0.0505 | -0.05445 | 0.01073 | 0.01207 |
| WBC | rs9358938 | rs9379873 | C | T | 6 | BTN2A2 | Male only | True | 0.1094 | 0.1115 | 0.1331 | 0.1331 | 35673 | 26968 | 188074 | 162396 | -0.01947 | -0.06631 | -0.05028 | -0.05497 | 0.01086 | 0.01219 |
| trait | lead_snp | ID | ALLELE1 | ALLELE0 | CHR | Gene_Symbol | independent_loci_in_sex_P1e2 | in_embl_any_or_gene | A1FREQ_VUMC_female | A1FREQ_VUMC_male | A1FREQ_UKB_female | A1FREQ_UKB_male | N_VUMC_female | N_VUMC_male | N_UKB_female | N_UKB_male | BETA_VUMC_female | BETA_VUMC_male | BETA_UKB_female | BETA_UKB_male | SE_VUMC_female | SE_VUMC_male |
| WBC | rs3799378 | rs3799378 | G | A | 6 | BTN3A1 | Male only | False | 0.2209 | 0.2219 | 0.238 | 0.2376 | 35673 | 26968 | 188074 | 162396 | -0.004155 | -0.05443 | -0.02742 | -0.03207 | 0.008214 | 0.009265 |
| WBC | rs9358938 | rs9393718 | A | G | 6 | BTN2A2 | Male only | True | 0.1095 | 0.1115 | 0.133 | 0.1331 | 35673 | 26968 | 188074 | 162396 | -0.01878 | -0.06672 | -0.05052 | -0.05464 | 0.01086 | 0.01219 |
| WBC | rs9358938 | rs75782365 | G | T | 6 | BTN2A2 | Male only | True | 0.08439 | 0.0866 | 0.1104 | 0.1103 | 35673 | 26968 | 188074 | 162396 | -0.02569 | -0.06691 | -0.05623 | -0.06041 | 0.0122 | 0.01364 |
| WBC | rs9358938 | rs2393667 | C | T | 6 | BTN2A2 | Male only | True | 0.1099 | 0.1122 | 0.1334 | 0.1334 | 35673 | 26968 | 188074 | 162396 | -0.01936 | -0.0654 | -0.05118 | -0.05437 | 0.01085 | 0.01217 |
| WBC | rs9358938 | rs2076030 | A | G | 6 | BTN2A2 | Male only | True | 0.1101 | 0.1123 | 0.1335 | 0.1335 | 35673 | 26968 | 188074 | 162396 | -0.01999 | -0.06457 | -0.05114 | -0.05433 | 0.01084 | 0.01217 |
| WBC | rs9358938 | rs2235251 | T | C | 6 | BTN2A2 | Male only | True | 0.1097 | 0.1121 | 0.1335 | 0.1335 | 35673 | 26968 | 188074 | 162396 | -0.02012 | -0.0657 | -0.05115 | -0.05465 | 0.01086 | 0.01218 |
| WBC | rs9358938 | rs9366656 | C | T | 6 | BTN2A2 | Male only | True | 0.1097 | 0.1121 | 0.1335 | 0.1335 | 35673 | 26968 | 188074 | 162396 | -0.02023 | -0.06567 | -0.05123 | -0.05473 | 0.01086 | 0.01219 |
| WBC | rs9358938 | rs13220495 | T | C | 6 | BTN2A2 | Male only | True | 0.0847 | 0.08709 | 0.1104 | 0.1102 | 35673 | 26968 | 188074 | 162396 | -0.02582 | -0.06472 | -0.05735 | -0.06073 | 0.01221 | 0.01364 |
| WBC | rs9358938 | rs9379875 | T | C | 6 | BTN2A2 | Male only | True | 0.1097 | 0.112 | 0.1335 | 0.1335 | 35673 | 26968 | 188074 | 162396 | -0.0201 | -0.06567 | -0.05109 | -0.05473 | 0.01087 | 0.01219 |
| WBC | rs9358938 | rs34605993 | T | C | 6 | BTN2A2 | Male only | True | 0.09644 | 0.09855 | 0.1223 | 0.1219 | 35673 | 26968 | 188074 | 162396 | -0.0224 | -0.06351 | -0.0556 | -0.05774 | 0.0115 | 0.0129 |
| WBC | rs9358938 | rs66757203 | T | C | 6 | BTN2A2 | Male only | True | 0.09693 | 0.09927 | 0.1229 | 0.1225 | 35673 | 26968 | 188074 | 162396 | -0.02358 | -0.0604 | -0.05558 | -0.05766 | 0.01148 | 0.01287 |
| WBC | rs9358938 | rs35680819 | C | T | 6 | BTN2A2 | Male only | True | 0.09643 | 0.09854 | 0.1223 | 0.1219 | 35673 | 26968 | 188074 | 162396 | -0.02239 | -0.06351 | -0.05551 | -0.05778 | 0.0115 | 0.01291 |
| WBC | rs9358938 | rs36033628 | G | T | 6 | BTN2A2 | Male only | True | 0.09636 | 0.09847 | 0.1222 | 0.1219 | 35673 | 26968 | 188074 | 162396 | -0.02209 | -0.06371 | -0.05553 | -0.05789 | 0.01151 | 0.01292 |
| WBC | rs9358938 | rs35627490 | T | C | 6 | BTN2A2 | Male only | True | 0.09645 | 0.09859 | 0.1223 | 0.1219 | 35673 | 26968 | 188074 | 162396 | -0.02269 | -0.06326 | -0.0553 | -0.05766 | 0.01149 | 0.0129 |
| WBC | rs9358938 | rs13195401 | T | G | 6 | BTN2A2 | Male only | True | 0.08386 | 0.08598 | 0.1105 | 0.1102 | 35673 | 26968 | 188074 | 162396 | -0.02749 | -0.0667 | -0.05766 | -0.06092 | 0.01231 | 0.01378 |
| WBC | rs9358938 | rs13195402 | T | G | 6 | BTN2A2 | Male only | True | 0.08386 | 0.08598 | 0.1123 | 0.1121 | 35673 | 26968 | 188074 | 162396 | -0.02748 | -0.06671 | -0.05711 | -0.06021 | 0.01231 | 0.01378 |
| WBC | rs9358938 | rs13195509 | A | G | 6 | BTN2A2 | Male only | True | 0.09645 | 0.09859 | 0.1223 | 0.122 | 35673 | 26968 | 188074 | 162396 | -0.02269 | -0.06326 | -0.05526 | -0.05767 | 0.01149 | 0.0129 |
| WBC | rs3799378 | rs2273558 | C | A | 6 | BTN3A1 | Male only | False | 0.3058 | 0.3072 | 0.3114 | 0.3114 | 35673 | 26968 | 188074 | 162396 | -1.97e-04 | -0.03528 | -0.0153 | -0.01825 | 0.007356 | 0.008306 |
| WBC | rs9358938 | rs3734542 | A | G | 6 | BTN2A2 | Male only | True | 0.09694 | 0.09906 | 0.1224 | 0.1221 | 35673 | 26968 | 188074 | 162396 | -0.02285 | -0.06128 | -0.05522 | -0.05794 | 0.01146 | 0.01287 |
| WBC | rs9358938 | rs3734544 | A | G | 6 | BTN2A2 | Male only | True | 0.172 | 0.1752 | 0.1915 | 0.1922 | 35673 | 26968 | 188074 | 162396 | -0.009514 | -0.03834 | -0.0299 | -0.03062 | 0.008962 | 0.01003 |
| WBC | rs9358938 | rs9366658 | A | G | 6 | BTN2A2 | Male only | True | 0.1719 | 0.175 | 0.1914 | 0.1921 | 35673 | 26968 | 188074 | 162396 | -0.009398 | -0.03797 | -0.02996 | -0.03062 | 0.008963 | 0.01003 |

| trait | lead_snp | ID | ALLELE1 | ALLELE0 | CHR | Gene_Symbol | independent_loci_in_sex_P1e2 | in_embl_any_or_gene | A1FREQ_VUMC_female | A1FREQ_VUMC_male | A1FREQ_UKB_female | A1FREQ_UKB_male | N_VUMC_female | N_VUMC_male | N_UKB_female | N_UKB_male | BETA_VUMC_female | BETA_VUMC_male | BETA_UKB_female | BETA_UKB_male | SE_VUMC_female | SE_VUMC_male |
|---|---|---|---|---|---|---|---|---|---|---|---|---|---|---|---|---|---|---|---|---|---|---|
| WBC | rs9358938 | rs9358944 | C | A | 6 | BTN2A2 | Male only | True | 0.1719 | 0.175 | 0.1915 | 0.1922 | 35673 | 26968 | 188074 | 162396 | -0.009391 | -0.03797 | -0.02998 | -0.03058 | 0.008963 | 0.01003 |
| WBC | rs9358938 | rs9348719 | C | T | 6 | BTN2A2 | Male only | True | 0.1719 | 0.1751 | 0.1915 | 0.1922 | 35673 | 26968 | 188074 | 162396 | -0.009532 | -0.0384 | -0.02993 | -0.03063 | 0.008969 | 0.01004 |
| WBC | rs9358938 | rs9358945 | G | A | 6 | BTN2A2 | Male only | True | 0.172 | 0.1752 | 0.1915 | 0.1922 | 35673 | 26968 | 188074 | 162396 | -0.009495 | -0.03832 | -0.02992 | -0.03064 | 0.008962 | 0.01003 |
| WBC | rs9358938 | rs7773938 | T | C | 6 | BTN2A2 | Male only | True | 0.172 | 0.1752 | 0.1914 | 0.1922 | 35673 | 26968 | 188074 | 162396 | -0.009495 | -0.03832 | -0.02988 | -0.03059 | 0.008962 | 0.01003 |
| WBC | rs9358938 | rs56296968 | T | C | 6 | BTN2A2 | Male only | True | 0.1718 | 0.1751 | 0.1914 | 0.1921 | 35673 | 26968 | 188074 | 162396 | -0.009527 | -0.03838 | -0.0299 | -0.03075 | 0.008969 | 0.01004 |
| WBC | rs9358938 | rs34104395 | T | C | 6 | BTN2A2 | Male only | True | 0.09692 | 0.09903 | 0.1222 | 0.122 | 35673 | 26968 | 188074 | 162396 | -0.02302 | -0.06113 | -0.05512 | -0.0581 | 0.01146 | 0.01287 |
| WBC | rs9358938 | rs9358946 | A | G | 6 | BTN2A2 | Male only | True | 0.172 | 0.1751 | 0.1914 | 0.1921 | 35673 | 26968 | 188074 | 162396 | -0.009487 | -0.03796 | -0.02991 | -0.03071 | 0.008962 | 0.01003 |
| WBC | rs9358938 | rs9358947 | A | G | 6 | BTN2A2 | Male only | True | 0.1718 | 0.175 | 0.1914 | 0.1921 | 35673 | 26968 | 188074 | 162396 | -0.009519 | -0.03803 | -0.02983 | -0.03057 | 0.00897 | 0.01004 |
| WBC | rs9358938 | rs16891725 | T | C | 6 | BTN2A2 | Male only | True | 0.1123 | 0.1149 | 0.1349 | 0.1348 | 35673 | 26968 | 188074 | 162396 | -0.02268 | -0.0599 | -0.0509 | -0.0556 | 0.01072 | 0.01202 |
| WBC | rs9358938 | rs7756567 | G | T | 6 | BTN2A2 | Male only | True | 0.1719 | 0.1751 | 0.1915 | 0.1922 | 35673 | 26968 | 188074 | 162396 | -0.009726 | -0.03812 | -0.02979 | -0.03067 | 0.008962 | 0.01003 |
| WBC | rs9358938 | rs9348720 | A | G | 6 | BTN2A2 | Male only | True | 0.1718 | 0.1749 | 0.1915 | 0.1922 | 35673 | 26968 | 188074 | 162396 | -0.009757 | -0.03819 | -0.02979 | -0.03068 | 0.008969 | 0.01004 |
| WBC | rs9358938 | rs9358948 | A | G | 6 | BTN2A2 | Male only | True | 0.1718 | 0.1749 | 0.1915 | 0.1922 | 35673 | 26968 | 188074 | 162396 | -0.009756 | -0.03819 | -0.02979 | -0.03068 | 0.008969 | 0.01004 |
| WBC | rs9358938 | rs9348721 | G | A | 6 | BTN2A2 | Male only | True | 0.1091 | 0.1117 | 0.1326 | 0.1326 | 35673 | 26968 | 188074 | 162396 | -0.02576 | -0.06109 | -0.05128 | -0.05625 | 0.01086 | 0.01218 |
| WBC | rs9358938 | rs6900665 | T | C | 6 | BTN2A2 | Male only | True | 0.1094 | 0.1119 | 0.1327 | 0.1326 | 35673 | 26968 | 188074 | 162396 | -0.02622 | -0.06037 | -0.05146 | -0.05623 | 0.01086 | 0.01218 |
| WBC | rs9358938 | rs16891727 | A | C | 6 | BTN2A2 | Male only | True | 0.1091 | 0.1117 | 0.1326 | 0.1326 | 35673 | 26968 | 188074 | 162396 | -0.02576 | -0.06109 | -0.05148 | -0.05625 | 0.01086 | 0.01218 |
| WBC | rs9358938 | rs34313099 | T | C | 6 | BTN2A2 | Male only | True | 0.1686 | 0.1719 | 0.1892 | 0.1899 | 35673 | 26968 | 188074 | 162396 | -0.01133 | -0.03814 | -0.02993 | -0.03072 | 0.009036 | 0.01011 |
| WBC | rs9358938 | rs13195047 | T | G | 6 | BTN2A2 | Male only | True | 0.1684 | 0.1717 | 0.189 | 0.1897 | 35673 | 26968 | 188074 | 162396 | -0.01122 | -0.03813 | -0.02985 | -0.03069 | 0.009039 | 0.01011 |
| WBC | rs9358938 | rs2093169 | T | C | 6 | BTN2A2 | Male only | True | 0.1718 | 0.1751 | 0.1915 | 0.1922 | 35673 | 26968 | 188074 | 162396 | -0.009668 | -0.03795 | -0.03006 | -0.0305 | 0.008964 | 0.01003 |
| WBC | rs9358938 | rs9379880 | A | G | 6 | BTN2A2 | Male only | True | 0.1716 | 0.1748 | 0.1912 | 0.1919 | 35673 | 26968 | 188074 | 162396 | -0.009995 | -0.03789 | -0.03004 | -0.03055 | 0.008978 | 0.01005 |
| WBC | rs9358938 | rs72843784 | T | G | 6 | BTN2A2 | Male only | True | 0.08488 | 0.08722 | 0.1113 | 0.1111 | 35673 | 26968 | 188074 | 162396 | -0.02902 | -0.06553 | -0.05734 | -0.06139 | 0.01218 | 0.01361 |
| WBC | rs9358938 | rs13220261 | T | C | 6 | BTN2A2 | Male only | True | 0.09694 | 0.09909 | 0.1223 | 0.122 | 35673 | 26968 | 188074 | 162396 | -0.02391 | -0.06098 | -0.05541 | -0.05822 | 0.01147 | 0.01287 |
| WBC | rs9358938 | rs2295593 | T | C | 6 | BTN2A2 | Male only | True | 0.1187 | 0.1215 | 0.1394 | 0.1395 | 35673 | 26968 | 188074 | 162396 | -0.02035 | -0.05747 | -0.05014 | -0.05373 | 0.01048 | 0.01175 |
| WBC | rs9358938 | rs35555795 | T | C | 6 | BTN2A2 | Male only | True | 0.0968 | 0.09898 | 0.1223 | 0.122 | 35673 | 26968 | 188074 | 162396 | -0.02376 | -0.0605 | -0.05557 | -0.05817 | 0.01147 | 0.01287 |
| WBC | rs9358938 | rs3799383 | T | C | 6 | BTN2A2 | Male only | True | 0.1187 | 0.1215 | 0.1394 | 0.1394 | 35673 | 26968 | 188074 | 162396 | -0.02008 | -0.05705 | -0.0498 | -0.05349 | 0.01047 | 0.01174 |
| WBC | rs9358938 | rs9379882 | T | C | 6 | BTN2A2 | Male only | True | 0.1187 | 0.1215 | 0.1394 | 0.1394 | 35673 | 26968 | 188074 | 162396 | -0.02007 | -0.05707 | -0.05021 | -0.0539 | 0.01047 | 0.01175 |
| WBC | rs9358938 | rs12176129 | T | C | 6 | BTN2A2 | Male only | True | 0.1185 | 0.1212 | 0.1394 | 0.1394 | 35673 | 26968 | 188074 | 162396 | -0.01995 | -0.05754 | -0.0502 | -0.05392 | 0.01049 | 0.01177 |
| WBC | rs9358938 | rs35433030 | A | G | 6 | BTN2A2 | Male only | True | 0.09682 | 0.09898 | 0.1226 | 0.1223 | 35673 | 26968 | 188074 | 162396 | -0.02372 | -0.0605 | -0.05527 | -0.05781 | 0.01147 | 0.01287 |
| WBC | rs9358938 | rs66941101 | T | C | 6 | BTN2A2 | Male only | True | 0.09682 | 0.09897 | 0.1226 | 0.1223 | 35673 | 26968 | 188074 | 162396 | -0.02373 | -0.06063 | -0.05526 | -0.0578 | 0.01147 | 0.01287 |
| WBC | rs9358938 | rs67234939 | G | A | 6 | BTN2A2 | Male only | True | 0.1187 | 0.1214 | 0.1394 | 0.1394 | 35673 | 26968 | 188074 | 162396 | -0.02022 | -0.05729 | -0.0502 | -0.05391 | 0.01047 | 0.01176 |

| trait | lead_snp | ID | ALLELE1 | ALLELE0 | CHR | Gene_Symbol | independent_loci_in_sex_P1e2 | in_embl_any_or_gene | A1FREQ_VUMC_female | A1FREQ_VUMC_male | A1FREQ_UKB_female | A1FREQ_UKB_male | N_VUMC_female | N_VUMC_male | N_UKB_female | N_UKB_male | BETA_VUMC_female | BETA_VUMC_male | BETA_UKB_female | BETA_UKB_male | SE_VUMC_female | SE_VUMC_male |
|---|---|---|---|---|---|---|---|---|---|---|---|---|---|---|---|---|---|---|---|---|---|---|
| WBC | rs9358938 | rs6920256 | A | G | 6 | BTN2A2 | Male only | True | 0.1159 | 0.1185 | 0.1379 | 0.138 | 35673 | 26968 | 188074 | 162396 | -0.02062 | -0.05849 | -0.05025 | -0.05491 | 0.01059 | 0.01187 |
| WBC | rs9358938 | rs34246779 | A | G | 6 | BTN2A2 | Male only | True | 0.09678 | 0.09886 | 0.1225 | 0.1222 | 35673 | 26968 | 188074 | 162396 | -0.02372 | -0.06062 | -0.0554 | -0.05791 | 0.01147 | 0.01288 |
| WBC | rs9358938 | rs72845428 | T | C | 6 | BTN2A2 | Male only | True | 0.09678 | 0.09886 | 0.1225 | 0.1222 | 35673 | 26968 | 188074 | 162396 | -0.02372 | -0.06062 | -0.05543 | -0.05795 | 0.01147 | 0.01288 |
| WBC | rs9358938 | rs16901784 | A | C | 6 | BTN2A2 | Male only | True | 0.1185 | 0.1211 | 0.1392 | 0.1394 | 35673 | 26968 | 188074 | 162396 | -0.02029 | -0.05798 | -0.05056 | -0.05417 | 0.01049 | 0.01177 |
| WBC | rs9358938 | rs35277236 | T | G | 6 | BTN2A2 | Male only | True | 0.09679 | 0.09884 | 0.1224 | 0.1222 | 35673 | 26968 | 188074 | 162396 | -0.02381 | -0.06042 | -0.05546 | -0.05814 | 0.01147 | 0.01288 |
| WBC | rs9358938 | rs72844462 | C | A | 6 | BTN2A2 | Male only | True | 0.09679 | 0.09884 | 0.1224 | 0.1222 | 35673 | 26968 | 188074 | 162396 | -0.02382 | -0.06042 | -0.05544 | -0.05811 | 0.01147 | 0.01288 |
| WBC | rs9358938 | rs36162392 | C | T | 6 | BTN2A2 | Male only | True | 0.09743 | 0.09959 | 0.1228 | 0.1226 | 35673 | 26968 | 188074 | 162396 | -0.02369 | -0.06196 | -0.05534 | -0.0581 | 0.01147 | 0.01287 |
| WBC | rs9358938 | rs9379891 | T | C | 6 | BTN2A2 | Male only | True | 0.1263 | 0.1293 | 0.1436 | 0.1439 | 35673 | 26968 | 188074 | 162396 | -0.01567 | -0.04986 | -0.05029 | -0.05311 | 0.01019 | 0.01144 |
| WBC | rs9358938 | rs13198716 | T | C | 6 | BTN2A2 | Male only | True | 0.08485 | 0.08714 | 0.1116 | 0.1114 | 35673 | 26968 | 188074 | 162396 | -0.02836 | -0.06547 | -0.05648 | -0.06036 | 0.01217 | 0.01361 |
| WBC | rs9358938 | rs9393735 | G | A | 6 | BTN2A2 | Male only | True | 0.1236 | 0.1265 | 0.1426 | 0.1428 | 35673 | 26968 | 188074 | 162396 | -0.01552 | -0.0505 | -0.05048 | -0.05431 | 0.01029 | 0.01154 |
| trait | lead_snp | ID | ALLELE1 | ALLELE0 | CHR | Gene_Symbol | independent_loci_in_sex_P1e2 | in_embl_any_or_gene | A1FREQ_VUMC_female | A1FREQ_VUMC_male | A1FREQ_UKB_female | A1FREQ_UKB_male | N_VUMC_female | N_VUMC_male | N_UKB_female | N_UKB_male | BETA_VUMC_female | BETA_VUMC_male | BETA_UKB_female | BETA_UKB_male | SE_VUMC_female | SE_VUMC_male |
| WBC | rs9358938 | rs13190739 | T | G | 6 | BTN2A2 | Male only | True | 0.09649 | 0.09853 | 0.1221 | 0.1218 | 35673 | 26968 | 188074 | 162396 | -0.02459 | -0.0607 | -0.05698 | -0.06047 | 0.0115 | 0.01291 |
| WBC | rs9358938 | rs34229567 | T | C | 6 | BTN2A2 | Male only | True | 0.1103 | 0.1134 | 0.1337 | 0.1339 | 35673 | 26968 | 188074 | 162396 | -0.01976 | -0.0597 | -0.05236 | -0.05719 | 0.01083 | 0.01212 |
| WBC | rs9358938 | rs34781270 | G | A | 6 | BTN2A2 | Male only | True | 0.09671 | 0.09876 | 0.1221 | 0.1218 | 35673 | 26968 | 188074 | 162396 | -0.02468 | -0.06027 | -0.05687 | -0.06022 | 0.01149 | 0.01289 |
| WBC | rs9358938 | rs35400317 | T | C | 6 | BTN2A2 | Male only | True | 0.08475 | 0.08707 | 0.1112 | 0.111 | 35673 | 26968 | 188074 | 162396 | -0.02945 | -0.06525 | -0.05825 | -0.06303 | 0.01219 | 0.01362 |
| WBC | rs9358938 | rs4634439 | G | A | 6 | BTN2A2 | Male only | True | 0.1105 | 0.1137 | 0.1339 | 0.134 | 35673 | 26968 | 188074 | 162396 | -0.02005 | -0.05849 | -0.05226 | -0.05686 | 0.01081 | 0.01209 |
| WBC | rs9358938 | rs45527431 | G | A | 6 | BTN2A2 | Male only | True | 0.08457 | 0.0871 | 0.1112 | 0.111 | 35673 | 26968 | 188074 | 162396 | -0.03022 | -0.06422 | -0.05831 | -0.06302 | 0.01219 | 0.01361 |
| WBC | rs9358938 | rs9379897 | C | T | 6 | BTN2A2 | Male only | True | 0.1105 | 0.1138 | 0.134 | 0.1342 | 35673 | 26968 | 188074 | 162396 | -0.02005 | -0.05855 | -0.05219 | -0.05655 | 0.01081 | 0.01209 |
| WBC | rs9358938 | rs13208916 | A | G | 6 | BTN2A2 | Male only | True | 0.1108 | 0.1141 | 0.134 | 0.1341 | 35673 | 26968 | 188074 | 162396 | -0.01902 | -0.05828 | -0.05218 | -0.0569 | 0.0108 | 0.01208 |
| WBC | rs9358938 | rs28360517 | G | A | 6 | BTN2A2 | Male only | True | 0.1105 | 0.1137 | 0.1338 | 0.134 | 35673 | 26968 | 188074 | 162396 | -0.02005 | -0.05858 | -0.05231 | -0.05693 | 0.01081 | 0.0121 |
| WBC | rs9358938 | 6:26603811 | C | A | 6 | BTN2A2 | Male only | True | 0.1105 | 0.1137 | | | 35673 | 26968 | | | -0.02007 | -0.05859 | | | 0.01081 | 0.0121 |
| WBC | rs9358938 | rs9379901 | C | T | 6 | BTN2A2 | Male only | True | 0.1105 | 0.1137 | 0.1338 | 0.134 | 35673 | 26968 | 188074 | 162396 | -0.02007 | -0.05857 | -0.05229 | -0.05685 | 0.01081 | 0.0121 |
| WBC | rs9358938 | rs35076545 | A | C | 6 | BTN2A2 | Male only | True | 0.09662 | 0.09883 | 0.1221 | 0.1218 | 35673 | 26968 | 188074 | 162396 | -0.02482 | -0.05973 | -0.0569 | -0.06008 | 0.01148 | 0.01288 |
| WBC | rs9358938 | rs9348726 | A | G | 6 | BTN2A2 | Male only | True | 0.1105 | 0.1137 | 0.1338 | 0.134 | 35673 | 26968 | 188074 | 162396 | -0.02006 | -0.05863 | -0.05228 | -0.05688 | 0.01081 | 0.0121 |
| WBC | rs9358938 | rs66488313 | T | G | 6 | BTN2A2 | Male only | True | 0.0846 | 0.0872 | 0.1112 | 0.1109 | 35673 | 26968 | 188074 | 162396 | -0.03043 | -0.0646 | -0.05834 | -0.06295 | 0.0122 | 0.01361 |
| WBC | rs3131855 | rs1233372 | A | G | 6 | SUMO2P1 | Female only | True | 0.46 | 0.4604 | 0.4545 | 0.4548 | 35673 | 26968 | 188074 | 162396 | 0.01799 | 0.009 | 0.02566 | 0.02351 | 0.006817 | 0.007663 |
| WBC | rs3131855 | rs3131855 | A | C | 6 | SUMO2P1 | Female only | True | 0.384 | 0.3827 | 0.3953 | 0.3958 | 35673 | 26968 | 188074 | 162396 | -0.03943 | -0.01714 | -0.03548 | -0.03566 | 0.007002 | 0.007885 |
| WBC | rs3131855 | rs3129077 | T | C | 6 | SUMO2P1 | Female only | True | 0.384 | 0.3827 | 0.3954 | 0.3959 | 35673 | 26968 | 188074 | 162396 | -0.03943 | -0.01714 | -0.03544 | -0.03565 | 0.007002 | 0.007885 |

| trait | lead_snp | ID | ALLELE1 | ALLELE0 | CHR | Gene_Symbol | independent_loci_in_sex_P1e2 | in_embl_any_or_gene | A1FREQ_VUMC_female | A1FREQ_VUMC_male | A1FREQ_UKB_female | A1FREQ_UKB_male | N_VUMC_female | N_VUMC_male | N_UKB_female | N_UKB_male | BETA_VUMC_female | BETA_VUMC_male | BETA_UKB_female | BETA_UKB_male | SE_VUMC_female | SE_VUMC_male |
|---|---|---|---|---|---|---|---|---|---|---|---|---|---|---|---|---|---|---|---|---|---|---|
| WBC | rs3131855 | rs3129057 | C | T | 6 | SUMO2P1 | Female only | True | 0.4508 | 0.4522 | 0.4528 | 0.4532 | 35673 | 26968 | 188074 | 162396 | 0.01764 | 0.01195 | 0.02778 | 0.02483 | 0.006844 | 0.00768 |
| WBC | rs3131855 | rs416571 | A | G | 6 | SUMO2P1 | Female only | True | 0.4515 | 0.453 | 0.4527 | 0.4532 | 35673 | 26968 | 188074 | 162396 | 0.01774 | 0.01209 | 0.0278 | 0.02478 | 0.006855 | 0.007693 |
| WBC | rs3131855 | rs3131888 | C | T | 6 | SUMO2P1 | Female only | True | 0.4175 | 0.416 | 0.4265 | 0.4265 | 35673 | 26968 | 188074 | 162396 | -0.03772 | -0.01779 | -0.03244 | -0.0312 | 0.00691 | 0.007772 |
| WBC | rs3131855 | rs3116807 | A | G | 6 | SUMO2P1 | Female only | True | 0.4676 | 0.4693 | 0.4689 | 0.4693 | 35673 | 26968 | 188074 | 162396 | 0.02286 | 0.01508 | 0.02891 | 0.02637 | 0.006821 | 0.007674 |
| WBC | rs3131855 | rs3131886 | C | T | 6 | SUMO2P1 | Female only | True | 0.4419 | 0.4402 | 0.45 | 0.4498 | 35673 | 26968 | 188074 | 162396 | -0.03133 | -0.01751 | -0.0317 | -0.02909 | 0.006858 | 0.007713 |
| WBC | rs3131855 | rs3129083 | C | T | 6 | SUMO2P1 | Female only | True | 0.4419 | 0.4401 | 0.45 | 0.4498 | 35673 | 26968 | 188074 | 162396 | -0.03134 | -0.01752 | -0.03174 | -0.02909 | 0.006859 | 0.007714 |
| WBC | rs3131855 | rs9258051 | A | G | 6 | SUMO2P1 | Female only | True | 0.4955 | 0.4973 | 0.4901 | 0.4906 | 35673 | 26968 | 188074 | 162396 | 0.02721 | 0.01875 | 0.0335 | 0.02925 | 0.006808 | 0.007643 |
| WBC | rs3131855 | rs9258052 | G | T | 6 | SUMO2P1 | Female only | True | 0.4411 | 0.4392 | 0.4491 | 0.449 | 35673 | 26968 | 188074 | 162396 | -0.03187 | -0.01763 | -0.03191 | -0.02928 | 0.006864 | 0.007719 |
| WBC | rs3131855 | rs9258053 | C | T | 6 | SUMO2P1 | Female only | True | 0.4417 | 0.4397 | 0.4496 | 0.4495 | 35673 | 26968 | 188074 | 162396 | -0.03141 | -0.01781 | -0.03176 | -0.02912 | 0.006861 | 0.007717 |
| WBC | rs3131855 | rs3131880 | T | C | 6 | SUMO2P1 | Female only | True | 0.4418 | 0.44 | 0.4497 | 0.4496 | 35673 | 26968 | 188074 | 162396 | -0.03145 | -0.01753 | -0.03186 | -0.02898 | 0.006859 | 0.007715 |
| WBC | rs3131855 | rs3116806 | C | A | 6 | SUMO2P1 | Female only | True | 0.4418 | 0.44 | 0.4498 | 0.4497 | 35673 | 26968 | 188074 | 162396 | -0.03149 | -0.01748 | -0.03187 | -0.02892 | 0.006857 | 0.007713 |
| WBC | rs3131855 | rs3129080 | A | G | 6 | SUMO2P1 | Female only | True | 0.4418 | 0.44 | 0.4499 | 0.4497 | 35673 | 26968 | 188074 | 162396 | -0.03149 | -0.01748 | -0.03186 | -0.02892 | 0.006857 | 0.007713 |
| WBC | rs3131855 | rs3129078 | C | T | 6 | SUMO2P1 | Female only | True | 0.4417 | 0.4399 | 0.4499 | 0.4497 | 35673 | 26968 | 188074 | 162396 | -0.0315 | -0.01749 | -0.03179 | -0.0289 | 0.006858 | 0.007715 |
| WBC | rs3131855 | rs3116805 | G | A | 6 | SUMO2P1 | Female only | True | 0.4418 | 0.4399 | 0.4499 | 0.4498 | 35673 | 26968 | 188074 | 162396 | -0.03142 | -0.01749 | -0.03181 | -0.0289 | 0.006858 | 0.007715 |
| WBC | rs3131855 | rs7776082 | A | G | 6 | SUMO2P1 | Female only | True | 0.4418 | 0.44 | 0.45 | 0.4498 | 35673 | 26968 | 188074 | 162396 | -0.0314 | -0.01748 | -0.03178 | -0.02886 | 0.006857 | 0.007713 |
| WBC | rs3131855 | rs9258089 | A | G | 6 | SUMO2P1 | Female only | True | 0.4418 | 0.44 | 0.4489 | 0.4487 | 35673 | 26968 | 188074 | 162396 | -0.03139 | -0.01749 | -0.0317 | -0.02876 | 0.006857 | 0.007714 |
| WBC | rs3131855 | rs9258105 | C | T | 6 | SUMO2P1 | Female only | True | 0.4417 | 0.4399 | 0.4499 | 0.4498 | 35673 | 26968 | 188074 | 162396 | -0.03145 | -0.01749 | -0.03179 | -0.02887 | 0.006858 | 0.007715 |
| WBC | rs3131855 | rs3129064 | T | C | 6 | SUMO2P1 | Female only | True | 0.4418 | 0.44 | 0.4499 | 0.4498 | 35673 | 26968 | 188074 | 162396 | -0.03147 | -0.01748 | -0.03179 | -0.02887 | 0.006857 | 0.007713 |
| WBC | rs3131855 | rs1633090 | T | G | 6 | SUMO2P1 | Female only | True | 0.4593 | 0.4583 | 0.4645 | 0.4641 | 35673 | 26968 | 188074 | 162396 | -0.0235 | -0.01485 | -0.02871 | -0.02633 | 0.006817 | 0.00768 |
| WBC | rs3131855 | rs1633087 | A | G | 6 | SUMO2P1 | Female only | True | 0.4594 | 0.4584 | 0.4645 | 0.4641 | 35673 | 26968 | 188074 | 162396 | -0.02354 | -0.01483 | -0.02869 | -0.02634 | 0.006814 | 0.007677 |
| WBC | rs3131855 | rs1737086 | A | C | 6 | SUMO2P1 | Female only | True | 0.4593 | 0.4583 | 0.4645 | 0.4641 | 35673 | 26968 | 188074 | 162396 | -0.02355 | -0.01483 | -0.02869 | -0.02634 | 0.006817 | 0.00768 |
| WBC | rs3131855 | rs1633086 | A | G | 6 | SUMO2P1 | Female only | True | 0.4591 | 0.458 | 0.4644 | 0.464 | 35673 | 26968 | 188074 | 162396 | -0.02331 | -0.01466 | -0.02867 | -0.02648 | 0.006816 | 0.007678 |
| WBC | rs3131855 | rs1633083 | T | C | 6 | SUMO2P1 | Female only | True | 0.4584 | 0.457 | 0.4638 | 0.4635 | 35673 | 26968 | 188074 | 162396 | -0.02338 | -0.01415 | -0.02859 | -0.02631 | 0.006819 | 0.007679 |
| WBC | rs3131855 | rs1633082 | T | C | 6 | SUMO2P1 | Female only | True | 0.4585 | 0.4572 | 0.4638 | 0.4635 | 35673 | 26968 | 188074 | 162396 | -0.02337 | -0.0142 | -0.02857 | -0.02633 | 0.006817 | 0.007676 |
| WBC | rs3131855 | rs1633081 | T | G | 6 | SUMO2P1 | Female only | True | 0.4584 | 0.457 | 0.4638 | 0.4635 | 35673 | 26968 | 188074 | 162396 | -0.02338 | -0.0142 | -0.02857 | -0.02634 | 0.006819 | 0.007679 |
| WBC | rs3131855 | rs733839 | A | G | 6 | SUMO2P1 | Female only | True | 0.4582 | 0.4568 | 0.4636 | 0.4632 | 35673 | 26968 | 188074 | 162396 | -0.02341 | -0.01389 | -0.02845 | -0.02634 | 0.006817 | 0.007677 |
| WBC | rs3131855 | rs1610624 | A | G | 6 | SUMO2P1 | Female only | True | 0.4603 | 0.4592 | 0.4649 | 0.4646 | 35673 | 26968 | 188074 | 162396 | -0.02323 | -0.01381 | -0.02838 | -0.02611 | 0.006818 | 0.007678 |
| WBC | rs3131855 | rs1737080 | A | G | 6 | SUMO2P1 | Female only | True | 0.4602 | 0.4591 | 0.4649 | 0.4645 | 35673 | 26968 | 188074 | 162396 | -0.02338 | -0.01389 | -0.02836 | -0.02616 | 0.006818 | 0.007679 |
| WBC | rs3131855 | rs1737078 | T | G | 6 | SUMO2P1 | Female only | True | 0.4604 | 0.4593 | 0.4649 | 0.4645 | 35673 | 26968 | 188074 | 162396 | -0.02343 | -0.0139 | -0.02831 | -0.02608 | 0.006816 | 0.007676 |

| trait | lead_snp | ID | ALLELE1 | ALLELE0 | CHR | Gene_Symbol | independent_loci_in_sex_P1e2 | in_embl_any_or_gene | A1FREQ_VUMC_female | A1FREQ_VUMC_male | A1FREQ_UKB_female | A1FREQ_UKB_male | N_VUMC_female | N_VUMC_male | N_UKB_female | N_UKB_male | BETA_VUMC_female | BETA_VUMC_male | BETA_UKB_female | BETA_UKB_male | SE_VUMC_female | SE_VUMC_male |
|---|---|---|---|---|---|---|---|---|---|---|---|---|---|---|---|---|---|---|---|---|---|---|
| WBC | rs3131855 | rs1737076 | G | A | 6 | SUMO2P1 | Female only | True | 0.4604 | 0.4593 | 0.4649 | 0.4645 | 35673 | 26968 | 188074 | 162396 | -0.02343 | -0.0139 | -0.02844 | -0.02617 | 0.006816 | 0.007676 |
| WBC | rs3131855 | rs1633059 | C | T | 6 | SUMO2P1 | Female only | True | 0.4768 | 0.4776 | 0.4747 | 0.4755 | 35673 | 26968 | 188074 | 162396 | 0.01941 | 0.01514 | 0.03012 | 0.02647 | 0.006803 | 0.007665 |
| WBC | rs1495100 | rs9972882 | A | C | 17 | PGAP3 | Female only | False | 0.2638 | 0.2669 | 0.2507 | 0.252 | 35673 | 26968 | 188074 | 162396 | 0.0211 | 0.004008 | 0.02684 | 0.02807 | 0.007663 | 0.008691 |
| WBC | rs1495100 | rs1877031 | G | A | 17 | PGAP3 | Female only | False | 0.3454 | 0.3471 | 0.3314 | 0.3329 | 35673 | 26968 | 188074 | 162396 | 0.03895 | 5.30e-04 | 0.02672 | 0.02764 | 0.00711 | 0.008072 |
| WBC | rs1495100 | rs11658786 | G | A | 17 | PGAP3 | Female only | False | 0.3526 | 0.3543 | 0.3313 | 0.3328 | 35673 | 26968 | 188074 | 162396 | 0.03884 | 0.001081 | 0.02671 | 0.0276 | 0.007189 | 0.008161 |
| WBC | rs1495100 | rs2271308 | T | C | 17 | PGAP3 | Female only | False | 0.2888 | 0.2916 | 0.2638 | 0.2654 | 35673 | 26968 | 188074 | 162396 | 0.0245 | 5.14e-04 | 0.02577 | 0.02709 | 0.007601 | 0.008614 |
| WBC | rs1495100 | rs931992 | G | T | 17 | PGAP3 | Female only | False | 0.3529 | 0.3545 | 0.3313 | 0.3328 | 35673 | 26968 | 188074 | 162396 | 0.03799 | 8.13e-04 | 0.02678 | 0.02766 | 0.007192 | 0.008165 |
| WBC | rs1495100 | rs1053651 | A | C | 17 | PGAP3 | Female only | False | 0.2889 | 0.2915 | 0.2638 | 0.2653 | 35673 | 26968 | 188074 | 162396 | 0.02403 | 6.75e-04 | 0.02579 | 0.02722 | 0.007604 | 0.008621 |
| WBC | rs1495100 | rs3764351 | G | A | 17 | PGAP3 | Female only | False | 0.3608 | 0.3632 | 0.3382 | 0.3396 | 35673 | 26968 | 188074 | 162396 | 0.03698 | -5.83e-04 | 0.02755 | 0.02882 | 0.007167 | 0.008133 |
| WBC | rs1495100 | rs876493 | G | A | 17 | PGAP3 | Female only | False | 0.4368 | 0.4369 | 0.4099 | 0.4112 | 35673 | 26968 | 188074 | 162396 | 0.03084 | 0.001624 | 0.03591 | 0.03514 | 0.006867 | 0.007788 |
| WBC | rs1495100 | rs2941504 | A | G | 17 | PGAP3 | Female only | False | 0.32 | 0.3217 | 0.3014 | 0.3022 | 35673 | 26968 | 188074 | 162396 | 0.03541 | 0.01454 | 0.04041 | 0.03812 | 0.007339 | 0.008312 |
| WBC | rs1495100 | rs1565922 | A | G | 17 | PGAP3 | Female only | False | 0.3204 | 0.3219 | 0.3014 | 0.3023 | 35673 | 26968 | 188074 | 162396 | 0.03583 | 0.01426 | 0.04041 | 0.0381 | 0.007337 | 0.008312 |
| WBC | rs1495100 | rs1565920 | G | A | 17 | PGAP3 | Female only | False | 0.3379 | 0.3382 | 0.314 | 0.315 | 35673 | 26968 | 188074 | 162396 | 0.04032 | 0.01199 | 0.03907 | 0.03759 | 0.007248 | 0.008224 |
| WBC | rs1495100 | rs1495102 | T | C | 17 | PGAP3 | Female only | False | 0.319 | 0.3201 | 0.3004 | 0.3012 | 35673 | 26968 | 188074 | 162396 | 0.03693 | 0.01449 | 0.04033 | 0.03862 | 0.007349 | 0.008323 |

| trait | lead_snp | ID | ALLELE1 | ALLELE0 | CHR | Gene_Symbol | independent_loci_in_sex_P1e2 | in_embl_any_or_gene | A1FREQ_VUMC_female | A1FREQ_VUMC_male | A1FREQ_UKB_female | A1FREQ_UKB_male | N_VUMC_female | N_VUMC_male | N_UKB_female | N_UKB_male | BETA_VUMC_female | BETA_VUMC_male | BETA_UKB_female | BETA_UKB_male | SE_VUMC_female | SE_VUMC_male |
|---|---|---|---|---|---|---|---|---|---|---|---|---|---|---|---|---|---|---|---|---|---|---|
| WBC | rs1495100 | rs1495101 | C | A | 17 | PGAP3 | Female only | False | 0.319 | 0.3201 | 0.3004 | 0.3012 | 35673 | 26968 | 188074 | 162396 | 0.03686 | 0.01452 | 0.04032 | 0.03862 | 0.007349 | 0.008324 |
| WBC | rs1495100 | rs1495100 | C | T | 17 | PGAP3 | Female only | False | 0.3379 | 0.3381 | 0.314 | 0.3151 | 35673 | 26968 | 188074 | 162396 | 0.04044 | 0.01206 | 0.03907 | 0.03759 | 0.007247 | 0.008224 |
| WBC | rs1495100 | rs2934952 | G | A | 17 | PGAP3 | Female only | False | 0.319 | 0.3201 | 0.3004 | 0.3012 | 35673 | 26968 | 188074 | 162396 | 0.03693 | 0.01449 | 0.04036 | 0.03869 | 0.007349 | 0.008323 |
| WBC | rs1495100 | rs2941505 | A | G | 17 | PGAP3 | Female only | False | 0.336 | 0.3367 | 0.3135 | 0.3145 | 35673 | 26968 | 188074 | 162396 | 0.03976 | 0.01216 | 0.03948 | 0.03775 | 0.007252 | 0.00823 |
| WBC | rs1495100 | rs2952152 | T | C | 17 | PGAP3 | Female only | False | 0.3369 | 0.3375 | 0.3138 | 0.3149 | 35673 | 26968 | 188074 | 162396 | 0.03983 | 0.01195 | 0.03929 | 0.03771 | 0.007251 | 0.008225 |
| WBC | rs1495100 | rs2941506 | A | G | 17 | PGAP3 | Female only | False | 0.336 | 0.3367 | 0.3135 | 0.3145 | 35673 | 26968 | 188074 | 162396 | 0.03976 | 0.01216 | 0.03948 | 0.03775 | 0.007252 | 0.008229 |
| WBC | rs1495100 | rs2934951 | A | G | 17 | PGAP3 | Female only | False | 0.336 | 0.3367 | 0.3135 | 0.3145 | 35673 | 26968 | 188074 | 162396 | 0.03976 | 0.01216 | 0.03948 | 0.03775 | 0.007252 | 0.00823 |
| WBC | rs1495100 | rs907089 | G | A | 17 | PGAP3 | Female only | False | 0.3506 | 0.3515 | 0.3296 | 0.3308 | 35673 | 26968 | 188074 | 162396 | 0.03754 | 0.00931 | 0.03761 | 0.03474 | 0.007179 | 0.008159 |
| WBC | rs1495100 | rs907090 | T | C | 17 | PGAP3 | Female only | False | 0.3506 | 0.3515 | 0.3296 | 0.3308 | 35673 | 26968 | 188074 | 162396 | 0.03754 | 0.00931 | 0.03761 | 0.03474 | 0.007179 | 0.008159 |
| WBC | rs1495100 | rs9675194 | T | C | 17 | PGAP3 | Female only | False | 0.3506 | 0.3515 | 0.3295 | 0.3307 | 35673 | 26968 | 188074 | 162396 | 0.03754 | 0.009495 | 0.03752 | 0.03464 | 0.007179 | 0.008159 |
| WBC | rs1495100 | rs2313171 | T | C | 17 | PGAP3 | Female only | False | 0.3505 | 0.3515 | 0.3295 | 0.3307 | 35673 | 26968 | 188074 | 162396 | 0.03757 | 0.009502 | 0.03753 | 0.03457 | 0.00718 | 0.008158 |
| WBC | rs1495100 | rs732084 | A | C | 17 | PGAP3 | Female only | False | 0.3506 | 0.3515 | 0.3296 | 0.3308 | 35673 | 26968 | 188074 | 162396 | 0.03763 | 0.009418 | 0.03754 | 0.03478 | 0.007179 | 0.008158 |
| WBC | rs1495100 | rs732083 | G | A | 17 | PGAP3 | Female only | False | 0.3545 | 0.3552 | 0.3311 | 0.3322 | 35673 | 26968 | 188074 | 162396 | 0.03811 | 0.008331 | 0.03735 | 0.03482 | 0.00717 | 0.008144 |

| trait | lead_snp | ID | ALLELE1 | ALLELE0 | CHR | Gene_Symbol | independent_loci_in_sex_P1e2 | in_embl_any_or_gene | A1FREQ_VUMC_female | A1FREQ_VUMC_male | A1FREQ_UKB_female | A1FREQ_UKB_male | N_VUMC_female | N_VUMC_male | N_UKB_female | N_UKB_male | BETA_VUMC_female | BETA_VUMC_male | BETA_UKB_female | BETA_UKB_male | SE_VUMC_female | SE_VUMC_male |
|---|---|---|---|---|---|---|---|---|---|---|---|---|---|---|---|---|---|---|---|---|---|---|
| WBC | rs1495100 | rs12150298 | T | C | 17 | PGAP3 | Female only | False | 0.3504 | 0.3513 | 0.3295 | 0.3307 | 35673 | 26968 | 188074 | 162396 | 0.03761 | 0.009508 | 0.03753 | 0.03458 | 0.007179 | 0.008158 |
| WBC | rs1495100 | rs12150603 | G | A | 17 | PGAP3 | Female only | False | 0.3467 | 0.3472 | 0.33 | 0.3313 | 35673 | 26968 | 188074 | 162396 | 0.03812 | 0.00929 | 0.03718 | 0.03442 | 0.007115 | 0.008085 |
| WBC | rs1495100 | rs8077172 | G | T | 17 | PGAP3 | Female only | False | 0.3504 | 0.3513 | 0.3295 | 0.3307 | 35673 | 26968 | 188074 | 162396 | 0.03762 | 0.009504 | 0.03752 | 0.03456 | 0.007178 | 0.008157 |
| WBC | rs1495100 | rs8078228 | C | T | 17 | PGAP3 | Female only | False | 0.3504 | 0.3513 | 0.3295 | 0.3307 | 35673 | 26968 | 188074 | 162396 | 0.03762 | 0.009504 | 0.03752 | 0.03456 | 0.007178 | 0.008157 |
| WBC | rs1495100 | rs1018246 | C | T | 17 | PGAP3 | Female only | False | 0.3505 | 0.3514 | 0.3295 | 0.3308 | 35673 | 26968 | 188074 | 162396 | 0.03757 | 0.009454 | 0.03754 | 0.03456 | 0.007179 | 0.008157 |
| WBC | rs1495100 | rs11078919 | T | C | 17 | PGAP3 | Female only | False | 0.3447 | 0.3457 | 0.3295 | 0.3307 | 35673 | 26968 | 188074 | 162396 | 0.03734 | 0.00946 | 0.03752 | 0.03457 | 0.007118 | 0.00809 |
| WBC | rs1495100 | rs1476278 | C | T | 17 | PGAP3 | Female only | False | 0.3514 | 0.3519 | 0.3297 | 0.3308 | 35673 | 26968 | 188074 | 162396 | 0.03835 | 0.009508 | 0.03737 | 0.03448 | 0.007174 | 0.008155 |
| WBC | rs1495100 | rs9303274 | T | C | 17 | PGAP3 | Female only | False | 0.3514 | 0.3519 | 0.3297 | 0.3308 | 35673 | 26968 | 188074 | 162396 | 0.03835 | 0.009507 | 0.03737 | 0.03448 | 0.007174 | 0.008155 |
| WBC | rs1495100 | rs12940986 | G | A | 17 | PGAP3 | Female only | False | 0.3523 | 0.3527 | 0.33 | 0.3312 | 35673 | 26968 | 188074 | 162396 | 0.03843 | 0.00934 | 0.0372 | 0.03444 | 0.007173 | 0.008152 |
| WBC | rs1495100 | rs2517952 | A | G | 17 | PGAP3 | Female only | False | 0.3177 | 0.3192 | 0.3001 | 0.3009 | 35673 | 26968 | 188074 | 162396 | 0.03603 | 0.01437 | 0.04048 | 0.03832 | 0.007355 | 0.008325 |
| WBC | rs1495100 | rs2517957 | G | A | 17 | PGAP3 | Female only | False | 0.3523 | 0.3526 | 0.3302 | 0.3314 | 35673 | 26968 | 188074 | 162396 | 0.03845 | 0.009382 | 0.03723 | 0.0344 | 0.007174 | 0.008154 |
| WBC | rs1495100 | rs2517958 | G | A | 17 | PGAP3 | Female only | False | 0.3522 | 0.3525 | 0.3302 | 0.3314 | 35673 | 26968 | 188074 | 162396 | 0.03832 | 0.009527 | 0.03722 | 0.03438 | 0.007173 | 0.008154 |
| WBC | rs1495100 | rs903501 | T | C | 17 | PGAP3 | Female only | False | 0.3327 | 0.3342 | 0.3166 | 0.3175 | 35673 | 26968 | 188074 | 162396 | 0.03434 | 0.01203 | 0.03847 | 0.03525 | 0.007267 | 0.00824 |
| WBC | rs1495100 | rs2517954 | T | C | 17 | PGAP3 | Female only | False | 0.342 | 0.3429 | 0.3264 | 0.3271 | 35673 | 26968 | 188074 | 162396 | 0.0294 | 0.01229 | 0.03578 | 0.03093 | 0.007218 | 0.008181 |
| WBC | rs1495100 | rs2517955 | C | T | 17 | PGAP3 | Female only | False | 0.3616 | 0.3613 | 0.34 | 0.341 | 35673 | 26968 | 188074 | 162396 | 0.0336 | 0.009755 | 0.03467 | 0.03017 | 0.007133 | 0.0081 |
| WBC | rs1495100 | rs2517956 | G | A | 17 | PGAP3 | Female only | False | 0.3365 | 0.3374 | 0.3264 | 0.3271 | 35673 | 26968 | 188074 | 162396 | 0.0293 | 0.01225 | 0.03578 | 0.03093 | 0.007161 | 0.008117 |
| WBC | rs1495100 | rs2517960 | T | C | 17 | PGAP3 | Female only | False | 0.3616 | 0.3613 | 0.3401 | 0.341 | 35673 | 26968 | 188074 | 162396 | 0.03362 | 0.009758 | 0.03467 | 0.03017 | 0.007133 | 0.008101 |
| WBC | rs1495100 | rs2643194 | C | T | 17 | PGAP3 | Female only | False | 0.3233 | 0.3242 | 0.3076 | 0.308 | 35673 | 26968 | 188074 | 162396 | 0.03095 | 0.01482 | 0.03839 | 0.03485 | 0.007319 | 0.00827 |
| WBC | rs1495100 | rs2517951 | C | T | 17 | PGAP3 | Female only | False | 0.343 | 0.3426 | 0.321 | 0.3216 | 35673 | 26968 | 188074 | 162396 | 0.03426 | 0.0123 | 0.03752 | 0.03418 | 0.007217 | 0.008172 |
| WBC | rs1495100 | rs2643195 | A | G | 17 | PGAP3 | Female only | False | 0.3199 | 0.3204 | 0.308 | 0.3086 | 35673 | 26968 | 188074 | 162396 | 0.03188 | 0.01467 | 0.03827 | 0.03519 | 0.007254 | 0.008204 |
| WBC | rs1495100 | rs2934971 | G | T | 17 | PGAP3 | Female only | False | 0.3234 | 0.3242 | 0.3076 | 0.308 | 35673 | 26968 | 188074 | 162396 | 0.03092 | 0.01485 | 0.03839 | 0.03488 | 0.007317 | 0.008269 |
| WBC | rs1495100 | rs1565923 | A | G | 17 | PGAP3 | Female only | False | 0.3232 | 0.3241 | 0.3074 | 0.3078 | 35673 | 26968 | 188074 | 162396 | 0.03142 | 0.01489 | 0.0386 | 0.03498 | 0.007323 | 0.008273 |
| AlkP | rs11244034 | rs11244034 | T | C | 9 | OBP2B | Female only | True | 0.1638 | 0.1608 | 0.1555 | 0.1559 | 21116 | 15939 | 185016 | 159276 | 0.07467 | 0.01388 | 0.05343 | 0.06291 | 0.01285 | 0.01528 |
| AlkP | rs886082 | rs10901246 | G | T | 9 | LCN1P1 | Female only | True | 0.2036 | 0.1999 | 0.1892 | 0.1892 | 21116 | 15939 | 185016 | 159276 | 0.06492 | 0.01044 | 0.05189 | 0.05746 | 0.01182 | 0.01404 |
| AlkP | rs886082 | rs10901247 | A | C | 9 | LCN1P1 | Female only | True | 0.207 | 0.2032 | 0.1892 | 0.1891 | 21116 | 15939 | 185016 | 159276 | 0.06417 | 0.01002 | 0.05186 | 0.05746 | 0.01187 | 0.0141 |
| AlkP | rs11244034 | rs1052517 | A | G | 9 | OBP2B | Female only | True | 0.1667 | 0.1639 | 0.1553 | 0.1558 | 21116 | 15939 | 185016 | 159276 | 0.07315 | 0.01332 | 0.05334 | 0.06305 | 0.01292 | 0.01536 |
| AlkP | rs11244034 | rs2073870 | T | G | 9 | OBP2B | Female only | True | 0.1623 | 0.1595 | 0.15 | 0.1503 | 21116 | 15939 | 185016 | 159276 | 0.07342 | 0.01417 | 0.05279 | 0.06216 | 0.01312 | 0.01561 |
| AlkP | rs11244034 | rs4310274 | A | G | 9 | OBP2B | Female only | True | 0.1625 | 0.1598 | 0.1508 | 0.1511 | 21116 | 15939 | 185016 | 159276 | 0.07244 | 0.01371 | 0.05245 | 0.06206 | 0.0131 | 0.01557 |
| AlkP | rs886082 | rs3761821 | T | C | 9 | LCN1P1 | Female only | True | 0.174 | 0.1715 | 0.1603 | 0.1607 | 21116 | 15939 | 185016 | 159276 | 0.07671 | 0.01556 | 0.05365 | 0.06337 | 0.01274 | 0.01514 |

| trait | lead_snp | ID | ALLELE1 | ALLELE0 | CHR | Gene_Symbol | independent_loci_in_sex_P1e2 | in_embl_any_or_gene | A1FREQ_VUMC_female | A1FREQ_VUMC_male | A1FREQ_UKB_female | A1FREQ_UKB_male | N_VUMC_female | N_VUMC_male | N_UKB_female | N_UKB_male | BETA_VUMC_female | BETA_VUMC_male | BETA_UKB_female | BETA_UKB_male | SE_VUMC_female | SE_VUMC_male |
|---|---|---|---|---|---|---|---|---|---|---|---|---|---|---|---|---|---|---|---|---|---|---|
| AlkP | rs886082 | rs7864821 | C | T | 9 | LCN1P1 | Female only | True | 0.1701 | 0.1677 | 0.1603 | 0.1607 | 21116 | 15939 | 185016 | 159276 | 0.07673 | 0.01565 | 0.05358 | 0.06331 | 0.01267 | 0.01506 |
| AlkP | rs886082 | rs10793957 | G | T | 9 | LCN1P1 | Female only | True | 0.1744 | 0.172 | 0.1603 | 0.1607 | 21116 | 15939 | 185016 | 159276 | 0.07717 | 0.01496 | 0.05369 | 0.06337 | 0.01273 | 0.01513 |
| AlkP | rs11244034 | rs7023066 | G | A | 9 | OBP2B | Female only | True | 0.1901 | 0.1874 | 0.1754 | 0.1761 | 21116 | 15939 | 185016 | 159276 | 0.04911 | -0.001879 | 0.03097 | 0.03164 | 0.01235 | 0.01463 |
| AlkP | rs886082 | rs6597610 | G | A | 9 | LCN1P1 | Female only | True | 0.1744 | 0.172 | 0.1603 | 0.1607 | 21116 | 15939 | 185016 | 159276 | 0.07719 | 0.01499 | 0.0538 | 0.06345 | 0.01273 | 0.01513 |
| AlkP | rs886082 | rs10736860 | C | T | 9 | LCN1P1 | Female only | True | 0.1744 | 0.172 | 0.1603 | 0.1607 | 21116 | 15939 | 185016 | 159276 | 0.0772 | 0.01499 | 0.05379 | 0.06347 | 0.01273 | 0.01513 |
| AlkP | rs886082 | rs4962106 | C | A | 9 | LCN1P1 | Female only | True | 0.1748 | 0.1722 | 0.1607 | 0.161 | 21116 | 15939 | 185016 | 159276 | 0.07748 | 0.01538 | 0.0537 | 0.0633 | 0.01272 | 0.01513 |
| AlkP | rs886082 | rs4962107 | C | T | 9 | LCN1P1 | Female only | True | 0.1747 | 0.1721 | 0.1605 | 0.1608 | 21116 | 15939 | 185016 | 159276 | 0.07751 | 0.01526 | 0.05378 | 0.0633 | 0.01272 | 0.01513 |
| AlkP | rs886082 | rs10793959 | G | A | 9 | LCN1P1 | Female only | True | 0.1747 | 0.1721 | 0.1605 | 0.1608 | 21116 | 15939 | 185016 | 159276 | 0.07751 | 0.01524 | 0.05378 | 0.0633 | 0.01272 | 0.01513 |
| AlkP | rs886082 | rs4246169 | C | A | 9 | LCN1P1 | Female only | True | 0.1422 | 0.1395 | 0.1259 | 0.1259 | 21116 | 15939 | 185016 | 159276 | 0.08569 | 0.01086 | 0.05199 | 0.06207 | 0.01394 | 0.01656 |
| AlkP | rs886082 | rs11789398 | A | G | 9 | LCN1P1 | Female only | True | 0.1405 | 0.1372 | 0.1229 | 0.1231 | 21116 | 15939 | 185016 | 159276 | 0.08544 | 0.01133 | 0.05425 | 0.06268 | 0.01411 | 0.01676 |
| AlkP | rs886082 | rs4379511 | T | C | 9 | LCN1P1 | Female only | True | 0.1402 | 0.1371 | 0.1227 | 0.1229 | 21116 | 15939 | 185016 | 159276 | 0.08493 | 0.01111 | 0.05434 | 0.06292 | 0.01412 | 0.01676 |
| AlkP | rs886082 | rs886082 | T | G | 9 | LCN1P1 | Female only | True | 0.1703 | 0.1674 | 0.1553 | 0.155 | 21116 | 15939 | 185016 | 159276 | 0.0815 | 0.002642 | 0.05201 | 0.05937 | 0.01272 | 0.01506 |
| AlkP | rs886082 | rs4507838 | T | C | 9 | LCN1P1 | Female only | True | 0.1387 | 0.1359 | 0.1218 | 0.122 | 21116 | 15939 | 185016 | 159276 | 0.0853 | 0.01049 | 0.0536 | 0.06318 | 0.01417 | 0.01681 |
| AlkP | rs3205136 | rs2013075 | T | C | 9 | ABO | Female only | True | 0.06621 | 0.06536 | 0.0623 | 0.06245 | 21116 | 15939 | 185016 | 159276 | 0.08017 | -0.008527 | 0.04821 | 0.0578 | 0.01916 | 0.02262 |
| AlkP | rs3205136 | rs9411463 | T | C | 9 | ABO | Female only | True | 0.0662 | 0.0653 | 0.06237 | 0.0625 | 21116 | 15939 | 185016 | 159276 | 0.08093 | -0.008856 | 0.04839 | 0.05789 | 0.01919 | 0.02266 |
| AlkP | rs886082 | rs10793960 | A | G | 9 | LCN1P1 | Female only | True | 0.1399 | 0.137 | 0.1226 | 0.1226 | 21116 | 15939 | 185016 | 159276 | 0.086 | 0.01302 | 0.05441 | 0.06427 | 0.01418 | 0.01681 |
| AlkP | rs3205136 | rs9411464 | T | C | 9 | ABO | Female only | True | 0.06508 | 0.06395 | 0.06173 | 0.06175 | 21116 | 15939 | 185016 | 159276 | 0.08556 | -0.009724 | 0.04655 | 0.0568 | 0.01938 | 0.023 |
| AlkP | rs3205136 | rs7470777 | G | A | 9 | ABO | Female only | True | 0.06763 | 0.06619 | 0.06149 | 0.06174 | 21116 | 15939 | 185016 | 159276 | 0.07813 | -4.79e-04 | 0.04405 | 0.0581 | 0.019 | 0.02257 |
| AlkP | rs3205136 | rs11244049 | C | T | 9 | ABO | Female only | True | 0.06652 | 0.06468 | 0.05994 | 0.05993 | 21116 | 15939 | 185016 | 159276 | 0.07976 | -0.006229 | 0.04432 | 0.05583 | 0.01916 | 0.02285 |
| AlkP | rs3205136 | rs3205136 | A | C | 9 | ABO | Female only | True | 0.0994 | 0.09576 | 0.09395 | 0.09344 | 21116 | 15939 | 185016 | 159276 | 0.0879 | 0.01486 | 0.05864 | 0.06682 | 0.01595 | 0.01929 |
| AlkP | rs3205136 | rs12216891 | T | C | 9 | ABO | Female only | True | 0.06265 | 0.06085 | 0.05799 | 0.05784 | 21116 | 15939 | 185016 | 159276 | 0.07512 | -0.01354 | 0.0444 | 0.05568 | 0.0198 | 0.02363 |
| AlkP | rs3205136 | rs77693339 | C | T | 9 | ABO | Female only | True | 0.06638 | 0.06473 | 0.05972 | 0.05975 | 21116 | 15939 | 185016 | 159276 | 0.0764 | -0.005703 | 0.04237 | 0.05458 | 0.01924 | 0.02285 |
| AlkP | rs3205136 | rs8176759 | A | G | 9 | ABO | Female only | True | 0.06855 | 0.06636 | 0.06074 | 0.0607 | 21116 | 15939 | 185016 | 159276 | 0.07351 | -0.002356 | 0.0436 | 0.05412 | 0.0189 | 0.02252 |
| AlkP | rs3205136 | rs1137827 | T | C | 9 | ABO | Female only | True | 0.06855 | 0.06636 | 0.06073 | 0.06069 | 21116 | 15939 | 185016 | 159276 | 0.07351 | -0.002356 | 0.0436 | 0.05411 | 0.0189 | 0.02252 |
| AlkP | rs3205136 | rs187099314 | T | C | 9 | ABO | Female only | True | 0.06855 | 0.06635 | 0.0607 | 0.06067 | 21116 | 15939 | 185016 | 159276 | 0.07348 | -0.002268 | 0.0434 | 0.05408 | 0.0189 | 0.02252 |
| AlkP | rs3205136 | rs8176751 | T | C | 9 | ABO | Female only | True | 0.09143 | 0.08894 | 0.08348 | 0.08271 | 21116 | 15939 | 185016 | 159276 | 0.0734 | 0.01043 | 0.05414 | 0.06503 | 0.01648 | 0.01978 |
| AlkP | rs3205136 | rs8176749 | T | C | 9 | ABO | Female only | True | 0.06879 | 0.0666 | 0.06067 | 0.06064 | 21116 | 15939 | 185016 | 159276 | 0.0732 | -0.002554 | 0.04304 | 0.05444 | 0.01882 | 0.02243 |

| trait | lead_snp | ID | ALLELE1 | ALLELE0 | CHR | Gene_Symbol | independent_loci_in_sex_P1e2 | in_embl_any_or_gene | A1FREQ_VUMC_female | A1FREQ_VUMC_male | A1FREQ_UKB_female | A1FREQ_UKB_male | N_VUMC_female | N_VUMC_male | N_UKB_female | N_UKB_male | BETA_VUMC_female | BETA_VUMC_male | BETA_UKB_female | BETA_UKB_male | SE_VUMC_female | SE_VUMC_male |
|---|---|---|---|---|---|---|---|---|---|---|---|---|---|---|---|---|---|---|---|---|---|---|
| AlkP | rs3205136 | rs8176746 | T | G | 9 | ABO | Female only | True | 0.06879 | 0.06659 | 0.06076 | 0.06073 | 21116 | 15939 | 185016 | 159276 | 0.07317 | -0.002492 | 0.04329 | 0.0538 | 0.01882 | 0.02243 |
| AlkP | rs3205136 | rs8176743 | T | C | 9 | ABO | Female only | True | 0.06876 | 0.06654 | 0.06067 | 0.06066 | 21116 | 15939 | 185016 | 159276 | 0.07361 | -0.003531 | 0.04314 | 0.05394 | 0.01883 | 0.02244 |
| AlkP | rs3205136 | rs8176741 | A | G | 9 | ABO | Female only | True | 0.06886 | 0.06667 | 0.06054 | 0.06053 | 21116 | 15939 | 185016 | 159276 | 0.07334 | -0.003259 | 0.0426 | 0.05371 | 0.01882 | 0.02242 |
| AlkP | rs3205136 | rs7855255 | T | C | 9 | ABO | Female only | True | 0.09158 | 0.08906 | 0.08349 | 0.08266 | 21116 | 15939 | 185016 | 159276 | 0.07253 | 0.01029 | 0.05403 | 0.06528 | 0.01642 | 0.01971 |
| AlkP | rs3205136 | rs8176733 | C | T | 9 | ABO | Female only | True | 0.09174 | 0.08922 | 0.0835 | 0.08267 | 21116 | 15939 | 185016 | 159276 | 0.07212 | 0.01008 | 0.05401 | 0.06527 | 0.01639 | 0.01967 |
| AlkP | rs3205136 | rs2073823 | A | G | 9 | ABO | Female only | True | 0.09163 | 0.08915 | 0.08355 | 0.0827 | 21116 | 15939 | 185016 | 159276 | 0.07154 | 0.01024 | 0.054 | 0.06538 | 0.01642 | 0.0197 |
| AlkP | rs3205136 | rs8176730 | C | T | 9 | ABO | Female only | True | 0.09157 | 0.08907 | 0.08352 | 0.0827 | 21116 | 15939 | 185016 | 159276 | 0.07242 | 0.01017 | 0.05402 | 0.06538 | 0.01642 | 0.01971 |
| AlkP | rs3205136 | rs8176725 | A | G | 9 | ABO | Female only | True | 0.09234 | 0.08972 | 0.0837 | 0.08291 | 21116 | 15939 | 185016 | 159276 | 0.06882 | 0.008716 | 0.05322 | 0.06471 | 0.01633 | 0.01963 |
| AlkP | rs3205136 | rs8176722 | A | C | 9 | ABO | Female only | True | 0.0916 | 0.08913 | 0.08352 | 0.08275 | 21116 | 15939 | 185016 | 159276 | 0.07314 | 0.009537 | 0.05364 | 0.06536 | 0.0164 | 0.0197 |
| AlkP | rs3205136 | rs75179845 | C | T | 9 | ABO | Female only | True | 0.06295 | 0.06126 | 0.05828 | 0.05821 | 21116 | 15939 | 185016 | 159276 | 0.07254 | -0.01599 | 0.04298 | 0.05419 | 0.01965 | 0.02346 |
| AlkP | rs3205136 | rs8176693 | T | C | 9 | ABO | Female only | True | 0.06862 | 0.06635 | 0.06059 | 0.06059 | 21116 | 15939 | 185016 | 159276 | 0.07045 | -0.005139 | 0.04188 | 0.05335 | 0.01883 | 0.02248 |
| AlkP | rs3205136 | rs8176672 | T | C | 9 | ABO | Female only | True | 0.06836 | 0.06616 | 0.06055 | 0.06055 | 21116 | 15939 | 185016 | 159276 | 0.07108 | -0.00412 | 0.04184 | 0.05331 | 0.01889 | 0.02255 |
| AlkP | rs3205136 | rs8176671 | C | A | 9 | ABO | Female only | True | 0.06639 | 0.06449 | 0.05944 | 0.05951 | 21116 | 15939 | 185016 | 159276 | 0.07338 | -0.009117 | 0.04054 | 0.05373 | 0.01916 | 0.02284 |
| AlkP | rs3205136 | rs8176662 | G | A | 9 | ABO | Female only | True | 0.06639 | 0.06448 | 0.05944 | 0.05952 | 21116 | 15939 | 185016 | 159276 | 0.07335 | -0.009145 | 0.04054 | 0.05363 | 0.01916 | 0.02284 |
| AlkP | rs3205136 | rs79343853 | C | T | 9 | ABO | Female only | True | 0.06634 | 0.06449 | 0.05944 | 0.05951 | 21116 | 15939 | 185016 | 159276 | 0.07345 | -0.00922 | 0.04054 | 0.05363 | 0.01917 | 0.02284 |
| AlkP | rs3205136 | rs8176644 | T | C | 9 | ABO | Female only | True | 0.06509 | 0.06349 | 0.05909 | 0.05915 | 21116 | 15939 | 185016 | 159276 | 0.06849 | -0.00873 | 0.04071 | 0.05344 | 0.01934 | 0.023 |
| AlkP | rs3205136 | rs75444660 | G | A | 9 | ABO | Female only | True | 0.05323 | 0.05196 | 0.04958 | 0.04969 | 21116 | 15939 | 185016 | 159276 | 0.0779 | -0.01842 | 0.03901 | 0.05534 | 0.02134 | 0.02551 |
| AlkP | rs11620783 | rs17795034 | A | G | 14 | NYNRIN | Female only | True | 0.3239 | 0.3233 | 0.3311 | 0.331 | 21116 | 15939 | 185016 | 159276 | -0.05357 | -0.0182 | -0.01673 | -0.01559 | 0.01 | 0.01193 |
| AlkP | rs11620783 | rs11625026 | T | C | 14 | NYNRIN | Female only | True | 0.3212 | 0.3205 | 0.3305 | 0.3305 | 21116 | 15939 | 185016 | 159276 | -0.05351 | -0.01684 | -0.01654 | -0.01583 | 0.01008 | 0.01202 |
| AlkP | rs11620783 | rs11620783 | T | C | 14 | NYNRIN | Female only | True | 0.4227 | 0.4259 | 0.4308 | 0.4318 | 21116 | 15939 | 185016 | 159276 | -0.05696 | -0.01904 | -0.03128 | -0.03053 | 0.00949 | 0.0113 |
| AlkP | rs11620783 | rs36012880 | C | T | 14 | NYNRIN | Female only | True | 0.4675 | 0.4659 | 0.4782 | 0.4776 | 21116 | 15939 | 185016 | 159276 | -0.04147 | -0.02243 | -0.02638 | -0.02937 | 0.009438 | 0.0113 |
| AlkP | rs11620783 | rs2332328 | T | C | 14 | NYNRIN | Female only | True | 0.4654 | 0.4644 | 0.4759 | 0.4754 | 21116 | 15939 | 185016 | 159276 | -0.04185 | -0.02158 | -0.02574 | -0.0289 | 0.009442 | 0.0113 |
| AlkP | rs11620783 | rs8017377 | A | G | 14 | NYNRIN | Female only | True | 0.4674 | 0.4666 | 0.4757 | 0.4752 | 21116 | 15939 | 185016 | 159276 | -0.04185 | -0.022 | -0.02567 | -0.02878 | 0.0094 | 0.01125 |
| SGOT | rs6490048 | rs886126 | T | C | 12 | CUX2 | Female only | True | 0.2215 | 0.2243 | 0.2213 | 0.2215 | 20991 | 15836 | 184364 | 158626 | -0.04847 | 0.003056 | -0.009005 | -0.01407 | 0.01084 | 0.01178 |
| SGOT | rs6490048 | rs4766562 | T | C | 12 | CUX2 | Female only | True | 0.2202 | 0.2238 | 0.2202 | 0.2203 | 20991 | 15836 | 184364 | 158626 | -0.04811 | 0.003595 | -0.009454 | -0.01455 | 0.0109 | 0.01184 |
| SGOT | rs6490048 | rs11065837 | G | A | 12 | CUX2 | Female only | True | 0.2271 | 0.228 | 0.228 | 0.2282 | 20991 | 15836 | 184364 | 158626 | -0.05356 | 0.007007 | -0.01389 | -0.01667 | 0.01078 | 0.01173 |
| SGOT | rs6490048 | rs7963641 | A | G | 12 | CUX2 | Female only | True | 0.1836 | 0.1853 | 0.1824 | 0.1818 | 20991 | 15836 | 184364 | 158626 | -0.05884 | 0.005649 | -0.01446 | -0.02325 | 0.01171 | 0.01271 |
| SGOT | rs6490048 | rs4766569 | T | C | 12 | CUX2 | Female only | True | 0.1678 | 0.1683 | 0.1669 | 0.1655 | 20991 | 15836 | 184364 | 158626 | -0.0533 | 0.001892 | -0.02264 | -0.02409 | 0.01214 | 0.01326 |
| SGOT | rs6490048 | rs933305 | A | G | 12 | CUX2 | Female only | True | 0.1899 | 0.1915 | 0.1887 | 0.1867 | 20991 | 15836 | 184364 | 158626 | -0.05126 | -0.003338 | -0.01787 | -0.02183 | 0.01152 | 0.01256 |

| trait | lead_snp | ID | ALLELE1 | ALLELE0 | CHR | Gene_Symbol | independent_loci_in_sex_P1e2 | in_embl_any_or_gene | A1FREQ_VUMC_female | A1FREQ_VUMC_male | A1FREQ_UKB_female | A1FREQ_UKB_male | N_VUMC_female | N_VUMC_male | N_UKB_female | N_UKB_male | BETA_VUMC_female | BETA_VUMC_male | BETA_UKB_female | BETA_UKB_male | SE_VUMC_female | SE_VUMC_male |
|---|---|---|---|---|---|---|---|---|---|---|---|---|---|---|---|---|---|---|---|---|---|---|
| SGOT | rs6490048 | rs10849930 | C | T | 12 | CUX2 | Female only | True | 0.1627 | 0.1629 | 0.161 | 0.1596 | 20991 | 15836 | 184364 | 158626 | -0.05452 | 0.00315 | -0.02279 | -0.02305 | 0.01222 | 0.01337 |
| SGOT | rs6490048 | rs11065857 | G | A | 12 | CUX2 | Female only | True | 0.2418 | 0.244 | 0.2397 | 0.2383 | 20991 | 15836 | 184364 | 158626 | -0.05821 | 0.00648 | -0.01331 | -0.01849 | 0.01057 | 0.01151 |
| SGOT | rs6490048 | rs6490048 | C | T | 12 | CUX2 | Female only | True | 0.2416 | 0.2437 | 0.2392 | 0.2379 | 20991 | 15836 | 184364 | 158626 | -0.05822 | 0.00617 | -0.01345 | -0.01883 | 0.01057 | 0.01152 |
| SGOT | rs6490048 | rs1362006 | C | T | 12 | CUX2 | Female only | True | 0.242 | 0.244 | 0.2395 | 0.2382 | 20991 | 15836 | 184364 | 158626 | -0.05757 | 0.006209 | -0.01358 | -0.01882 | 0.01056 | 0.01152 |
| SGOT | rs6490048 | rs28403936 | G | A | 12 | CUX2 | Female only | True | 0.2216 | 0.2216 | 0.224 | 0.2221 | 20991 | 15836 | 184364 | 158626 | -0.05847 | 0.005878 | -0.01588 | -0.01606 | 0.01098 | 0.01201 |
| SGOT | rs6490048 | rs28877654 | T | G | 12 | CUX2 | Female only | True | 0.1931 | 0.1938 | 0.1983 | 0.1965 | 20991 | 15836 | 184364 | 158626 | -0.05387 | -0.002589 | -0.01758 | -0.02032 | 0.01154 | 0.01264 |
| SGOT | rs6490048 | rs28844095 | G | A | 12 | CUX2 | Female only | True | 0.2216 | 0.2216 | 0.224 | 0.2221 | 20991 | 15836 | 184364 | 158626 | -0.05847 | 0.005885 | -0.01588 | -0.01603 | 0.01098 | 0.01201 |
| SGOT | rs6490048 | rs3803167 | C | T | 12 | CUX2 | Female only | True | 0.2255 | 0.2256 | 0.224 | 0.222 | 20991 | 15836 | 184364 | 158626 | -0.05769 | 0.006019 | -0.01583 | -0.01625 | 0.01079 | 0.0118 |
| SGOT | rs6490048 | rs10219736 | T | C | 12 | CUX2 | Female only | True | 0.1968 | 0.1976 | 0.1982 | 0.1964 | 20991 | 15836 | 184364 | 158626 | -0.05237 | -0.002577 | -0.01758 | -0.02066 | 0.01131 | 0.0124 |
| TProSe | rs2395043 | rs2523655 | A | G | 6 | MICB | Male only | False | 0.182 | 0.1763 | 0.1853 | 0.1859 | 20773 | 15800 | 167977 | 146944 | 0.02257 | 0.03649 | 0.03306 | 0.03501 | 0.01109 | 0.01332 |
| TProSe | rs2395043 | rs2516507 | T | C | 6 | MICB | Male only | False | 0.4009 | 0.4 | 0.4046 | 0.4042 | 20773 | 15800 | 167977 | 146944 | 0.00211 | 0.04502 | 0.02653 | 0.03147 | 0.00875 | 0.01034 |
| TProSe | rs2395043 | rs2904788 | A | C | 6 | MICB | Male only | False | 0.4009 | 0.4 | 0.4046 | 0.4041 | 20773 | 15800 | 167977 | 146944 | 0.002122 | 0.04491 | 0.02656 | 0.03154 | 0.008751 | 0.01034 |
| TProSe | rs2395043 | rs2428500 | C | T | 6 | MICB | Male only | False | 0.3922 | 0.3913 | 0.3955 | 0.3956 | 20773 | 15800 | 167977 | 146944 | -0.001043 | 0.04147 | 0.02579 | 0.03104 | 0.008789 | 0.01037 |
| TProSe | rs2395043 | rs9267257 | A | C | 6 | MICB | Male only | False | 0.1789 | 0.1732 | 0.1887 | 0.1885 | 20773 | 15800 | 167977 | 146944 | 0.02058 | 0.04638 | 0.03131 | 0.03466 | 0.01124 | 0.01343 |
| TProSe | rs2894222 | 6:31456372 | T | C | 6 | MICB | Male only | False | 0.3222 | 0.3276 | | | 20773 | 15800 | | | -0.001174 | -0.03902 | | | 0.009262 | 0.01078 |
| TProSe | rs2894222 | rs9267260 | A | G | 6 | MICB | Male only | False | 0.3809 | 0.3807 | 0.3889 | 0.3895 | 20773 | 15800 | 167977 | 146944 | 0.004769 | 0.04691 | 0.02699 | 0.03449 | 0.008878 | 0.01047 |
| TProSe | rs2894222 | rs2905732 | G | A | 6 | MICB | Male only | False | 0.3822 | 0.3821 | 0.3887 | 0.3893 | 20773 | 15800 | 167977 | 146944 | 0.005085 | 0.04678 | 0.0271 | 0.03447 | 0.008858 | 0.01045 |
| TProSe | rs2894222 | rs2905729 | A | G | 6 | MICB | Male only | False | 0.3824 | 0.3823 | 0.3891 | 0.3897 | 20773 | 15800 | 167977 | 146944 | 0.004766 | 0.04691 | 0.02711 | 0.03447 | 0.008857 | 0.01045 |
| TProSe | rs2894222 | rs2507964 | T | C | 6 | MICB | Male only | False | 0.323 | 0.3283 | 0.3211 | 0.3206 | 20773 | 15800 | 167977 | 146944 | -0.001192 | -0.03804 | -0.01489 | -0.03061 | 0.009244 | 0.01076 |
| TProSe | rs2894222 | rs2428499 | A | G | 6 | MICB | Male only | False | 0.323 | 0.3283 | 0.3211 | 0.3206 | 20773 | 15800 | 167977 | 146944 | -0.001192 | -0.03804 | -0.0149 | -0.03061 | 0.009244 | 0.01076 |
| TProSe | rs2894222 | rs2428498 | A | G | 6 | MICB | Male only | False | 0.323 | 0.3283 | 0.3211 | 0.3206 | 20773 | 15800 | 167977 | 146944 | -0.001194 | -0.03804 | -0.01491 | -0.03059 | 0.009244 | 0.01076 |
| TProSe | rs2894222 | rs2507963 | T | C | 6 | MICB | Male only | False | 0.323 | 0.3283 | 0.3211 | 0.3206 | 20773 | 15800 | 167977 | 146944 | -0.001195 | -0.03804 | -0.01492 | -0.03059 | 0.009244 | 0.01076 |

| trait | lead_snp | ID | ALLELE1 | ALLELE0 | CHR | Gene_Symbol | independent_loci_in_sex_P1e2 | in_embl_any_or_gene | A1FREQ_VUMC_female | A1FREQ_VUMC_male | A1FREQ_UKB_female | A1FREQ_UKB_male | N_VUMC_female | N_VUMC_male | N_UKB_female | N_UKB_male | BETA_VUMC_female | BETA_VUMC_male | BETA_UKB_female | BETA_UKB_male | SE_VUMC_female | SE_VUMC_male |
|---|---|---|---|---|---|---|---|---|---|---|---|---|---|---|---|---|---|---|---|---|---|---|
| TProSe | rs2894222 | rs9267294 | G | A | 6 | MICB | Male only | False | 0.3231 | 0.3285 | 0.3212 | 0.3208 | 20773 | 15800 | 167977 | 146944 | -7.53e-04 | -0.03769 | -0.01488 | -0.03066 | 0.009244 | 0.01076 |
| TProSe | rs2894222 | rs2395033 | C | T | 6 | MICB | Male only | False | 0.3828 | 0.3826 | 0.3892 | 0.3898 | 20773 | 15800 | 167977 | 146944 | 0.00505 | 0.04775 | 0.02712 | 0.03439 | 0.00885 | 0.01045 |
| TProSe | rs2894222 | rs2905724 | A | G | 6 | MICB | Male only | False | 0.3228 | 0.3289 | 0.3214 | 0.3209 | 20773 | 15800 | 167977 | 146944 | -0.002706 | -0.03747 | -0.01421 | -0.03058 | 0.009261 | 0.01076 |
| TProSe | rs2894222 | rs2516415 | A | G | 6 | MICB | Male only | False | 0.3235 | 0.329 | 0.322 | 0.3214 | 20773 | 15800 | 167977 | 146944 | -0.001805 | -0.04036 | -0.01505 | -0.03143 | 0.009234 | 0.01076 |
| TProSe | rs2894222 | rs2904787 | A | G | 6 | MICB | Male only | False | 0.3802 | 0.3803 | 0.3866 | 0.387 | 20773 | 15800 | 167977 | 146944 | 0.00506 | 0.05084 | 0.02787 | 0.03509 | 0.00885 | 0.01046 |

| trait | lead_snp | ID | ALLELE1 | ALLELE0 | CHR | Gene_Symbol | independent_loci_in_sex_P1e2 | in_embl_any_or_gene | A1FREQ_VUMC_female | A1FREQ_VUMC_male | A1FREQ_UKB_female | A1FREQ_UKB_male | N_VUMC_female | N_VUMC_male | N_UKB_female | N_UKB_male | BETA_VUMC_female | BETA_VUMC_male | BETA_UKB_female | BETA_UKB_male | SE_VUMC_female | SE_VUMC_male |
|---|---|---|---|---|---|---|---|---|---|---|---|---|---|---|---|---|---|---|---|---|---|---|
| TProSe | rs2894222 | rs2904783 | C | T | 6 | MICB | Male only | False | 0.38 | 0.3803 | 0.3866 | 0.387 | 20773 | 15800 | 167977 | 146944 | 0.005054 | 0.05063 | 0.02788 | 0.0351 | 0.008848 | 0.01046 |
| TProSe | rs2894222 | rs2395038 | T | G | 6 | MICB | Male only | False | 0.3802 | 0.3804 | 0.3866 | 0.387 | 20773 | 15800 | 167977 | 146944 | 0.00498 | 0.05065 | 0.02789 | 0.03516 | 0.00885 | 0.01046 |
| TProSe | rs2894222 | rs2395039 | T | C | 6 | MICB | Male only | False | 0.3233 | 0.3288 | 0.3218 | 0.3212 | 20773 | 15800 | 167977 | 146944 | -0.001625 | -0.04088 | -0.01504 | -0.03142 | 0.009239 | 0.01076 |
| TProSe | rs2395043 | rs3130922 | A | G | 6 | MICB | Male only | False | 0.3326 | 0.332 | 0.3284 | 0.3291 | 20773 | 15800 | 167977 | 146944 | 0.008336 | 0.05305 | 0.03379 | 0.03804 | 0.009122 | 0.01075 |
| TProSe | rs2894222 | rs2516413 | T | G | 6 | MICB | Male only | False | 0.3236 | 0.3287 | 0.3211 | 0.3206 | 20773 | 15800 | 167977 | 146944 | -6.49e-05 | -0.04287 | -0.01492 | -0.03195 | 0.009247 | 0.01079 |
| TProSe | rs2395043 | rs3094006 | T | C | 6 | MICB | Male only | False | 0.2792 | 0.2783 | 0.2825 | 0.2828 | 20773 | 15800 | 167977 | 146944 | 0.01326 | 0.05075 | 0.0304 | 0.03431 | 0.009622 | 0.01133 |
| TProSe | rs2894222 | rs2534687 | T | C | 6 | MICB | Male only | False | 0.3236 | 0.3289 | 0.3189 | 0.3186 | 20773 | 15800 | 167977 | 146944 | -9.11e-04 | -0.04208 | -0.01511 | -0.03204 | 0.009271 | 0.01078 |
| TProSe | rs2894222 | rs2253040 | T | G | 6 | MICB | Male only | False | 0.3286 | 0.3335 | 0.3248 | 0.3242 | 20773 | 15800 | 167977 | 146944 | -8.10e-04 | -0.04444 | -0.01367 | -0.0308 | 0.009195 | 0.01074 |
| TProSe | rs2894222 | rs2395041 | C | A | 6 | MICB | Male only | False | 0.3635 | 0.3621 | 0.3574 | 0.3583 | 20773 | 15800 | 167977 | 146944 | 0.01008 | 0.05879 | 0.03408 | 0.03987 | 0.008923 | 0.01056 |
| TProSe | rs2395043 | rs2395042 | A | C | 6 | MICB | Male only | False | 0.2892 | 0.2882 | 0.2825 | 0.2832 | 20773 | 15800 | 167977 | 146944 | 0.01187 | 0.06211 | 0.03818 | 0.04436 | 0.009471 | 0.01118 |
| TProSe | rs2894222 | rs2253042 | T | C | 6 | MICB | Male only | False | 0.3285 | 0.3333 | 0.3246 | 0.324 | 20773 | 15800 | 167977 | 146944 | -0.001192 | -0.04459 | -0.01397 | -0.03084 | 0.009195 | 0.01074 |
| TProSe | rs2395043 | rs2395043 | T | C | 6 | MICB | Male only | False | 0.2891 | 0.2882 | 0.2826 | 0.2832 | 20773 | 15800 | 167977 | 146944 | 0.0118 | 0.06231 | 0.03819 | 0.04443 | 0.009471 | 0.01118 |
| TProSe | rs2894222 | rs2395044 | T | C | 6 | MICB | Male only | False | 0.4754 | 0.4782 | 0.4751 | 0.4736 | 20773 | 15800 | 167977 | 146944 | -0.007073 | -0.06707 | -0.02638 | -0.03861 | 0.00865 | 0.01013 |
| TProSe | rs2894222 | rs2253052 | T | C | 6 | MICB | Male only | False | 0.3284 | 0.3332 | 0.325 | 0.3244 | 20773 | 15800 | 167977 | 146944 | -9.00e-04 | -0.04415 | -0.01396 | -0.0307 | 0.009201 | 0.01075 |
| TProSe | rs2894222 | rs2894222 | A | G | 6 | MICB | Male only | False | 0.475 | 0.4778 | 0.4752 | 0.4736 | 20773 | 15800 | 167977 | 146944 | -0.007197 | -0.06729 | -0.02639 | -0.03854 | 0.008657 | 0.01014 |
| TProSe | rs2894222 | rs3130924 | G | T | 6 | MICB | Male only | False | 0.4556 | 0.4579 | 0.4576 | 0.4564 | 20773 | 15800 | 167977 | 146944 | -0.01083 | -0.06202 | -0.02785 | -0.04061 | 0.008676 | 0.01016 |
| TProSe | rs2261033 | rs2857698 | T | C | 6 | PRRC2A | Male only | False | 0.2958 | 0.3043 | 0.2823 | 0.2824 | 20773 | 15800 | 167977 | 146944 | 0.01154 | 0.04316 | 0.0155 | 0.02822 | 0.009428 | 0.01108 |
| TProSe | rs2261033 | rs2261033 | G | A | 6 | PRRC2A | Male only | False | 0.4546 | 0.4581 | 0.428 | 0.4276 | 20773 | 15800 | 167977 | 146944 | 0.01589 | 0.05587 | 0.03228 | 0.04669 | 0.008642 | 0.01024 |
| TProSe | rs2261033 | rs755714 | T | C | 6 | PRRC2A | Male only | False | 0.296 | 0.3039 | 0.2819 | 0.282 | 20773 | 15800 | 167977 | 146944 | 0.01151 | 0.0426 | 0.01536 | 0.02818 | 0.009409 | 0.01108 |
| TProSe | rs2261033 | rs805297 | A | C | 6 | PRRC2A | Male only | False | 0.296 | 0.3041 | 0.2819 | 0.2821 | 20773 | 15800 | 167977 | 146944 | 0.01158 | 0.04207 | 0.01526 | 0.02821 | 0.009409 | 0.01108 |
| TProSe | rs2261033 | rs707919 | G | A | 6 | PRRC2A | Male only | False | 0.2959 | 0.304 | 0.2826 | 0.2827 | 20773 | 15800 | 167977 | 146944 | 0.01224 | 0.04221 | 0.01524 | 0.02851 | 0.009417 | 0.01109 |
| TProSe | rs2261033 | rs707939 | A | C | 6 | PRRC2A | Male only | False | 0.3358 | 0.3438 | 0.3225 | 0.3232 | 20773 | 15800 | 167977 | 146944 | 0.002849 | 0.04219 | 0.02103 | 0.03348 | 0.009149 | 0.01071 |
| TProSe | rs693797 | rs7739432 | T | C | 6 | NOTCH4 | Male only | False | 0.4186 | 0.4159 | 0.4309 | 0.4312 | 20773 | 15800 | 167977 | 146944 | 0.02054 | 0.04912 | 0.03256 | 0.04502 | 0.008773 | 0.01034 |
| TProSe | rs693797 | rs404890 | A | C | 6 | NOTCH4 | Male only | False | 0.3891 | 0.3885 | 0.3969 | 0.3965 | 20773 | 15800 | 167977 | 146944 | -0.01335 | -0.05463 | -0.03639 | -0.0529 | 0.008845 | 0.01037 |
| TProSe | rs693797 | rs2849015 | A | G | 6 | NOTCH4 | Male only | False | 0.3899 | 0.3889 | 0.3969 | 0.3965 | 20773 | 15800 | 167977 | 146944 | -0.01388 | -0.05537 | -0.03635 | -0.05291 | 0.008842 | 0.01037 |
| TProSe | rs693797 | rs6913558 | G | A | 6 | NOTCH4 | Male only | False | 0.4479 | 0.4509 | 0.4473 | 0.4459 | 20773 | 15800 | 167977 | 146944 | -0.01555 | -0.0537 | -0.03289 | -0.04912 | 0.008705 | 0.01018 |
| TProSe | rs693797 | rs2395107 | A | G | 6 | NOTCH4 | Male only | False | 0.3884 | 0.3879 | 0.3972 | 0.3967 | 20773 | 15800 | 167977 | 146944 | -0.0131 | -0.05495 | -0.0362 | -0.05285 | 0.00886 | 0.01038 |
| TProSe | rs693797 | rs2395108 | C | T | 6 | NOTCH4 | Male only | False | 0.447 | 0.4505 | 0.4472 | 0.4458 | 20773 | 15800 | 167977 | 146944 | -0.01472 | -0.05311 | -0.0329 | -0.04912 | 0.008707 | 0.01017 |
| TProSe | rs693797 | rs9267904 | G | T | 6 | NOTCH4 | Male only | False | 0.4469 | 0.4505 | 0.4472 | 0.4459 | 20773 | 15800 | 167977 | 146944 | -0.01477 | -0.05311 | -0.03293 | -0.04914 | 0.008706 | 0.01017 |

| trait | lead_snp | ID | ALLELE1 | ALLELE0 | CHR | Gene_Symbol | independent_loci_in_sex_P1e2 | in_embl_any_or_gene | A1FREQ_VUMC_female | A1FREQ_VUMC_male | A1FREQ_UKB_female | A1FREQ_UKB_male | N_VUMC_female | N_VUMC_male | N_UKB_female | N_UKB_male | BETA_VUMC_female | BETA_VUMC_male | BETA_UKB_female | BETA_UKB_male | SE_VUMC_female | SE_VUMC_male |
|---|---|---|---|---|---|---|---|---|---|---|---|---|---|---|---|---|---|---|---|---|---|---|
| TProSe | rs693797 | rs693797 | C | T | 6 | NOTCH4 | Male only | False | 0.4109 | 0.4109 | 0.4171 | 0.4157 | 20773 | 15800 | 167977 | 146944 | -0.0132 | -0.05889 | -0.0358 | -0.05453 | 0.008781 | 0.01028 |
| TProSe | rs693797 | rs9267908 | A | G | 6 | NOTCH4 | Male only | False | 0.4469 | 0.4505 | 0.4472 | 0.4459 | 20773 | 15800 | 167977 | 146944 | -0.01477 | -0.05311 | -0.03294 | -0.04915 | 0.008706 | 0.01017 |
| TProSe | rs693797 | rs9267911 | C | T | 6 | NOTCH4 | Male only | False | 0.4476 | 0.4512 | 0.4472 | 0.4459 | 20773 | 15800 | 167977 | 146944 | -0.01472 | -0.05296 | -0.03292 | -0.04903 | 0.008693 | 0.01016 |
| TProSe | rs693797 | rs9267912 | G | T | 6 | NOTCH4 | Male only | False | 0.4471 | 0.4507 | 0.4472 | 0.4459 | 20773 | 15800 | 167977 | 146944 | -0.0148 | -0.0532 | -0.03289 | -0.04917 | 0.008709 | 0.01018 |
| TProSe | rs693797 | rs9267913 | T | C | 6 | NOTCH4 | Male only | False | 0.4471 | 0.4508 | 0.4472 | 0.4459 | 20773 | 15800 | 167977 | 146944 | -0.01481 | -0.05311 | -0.0329 | -0.04917 | 0.00871 | 0.01018 |
| BASOAB | rs236987 | rs17012368 | G | T | 4 | AFF1 | Female only | True | 0.375 | 0.3778 | 0.379 | 0.3795 | 25623 | 19957 | 187758 | 162098 | 0.05382 | 0.02354 | 0.002919 | 0.009433 | 0.008498 | 0.009446 |
| BASOAB | rs236987 | rs2125787 | G | A | 4 | AFF1 | Female only | True | 0.3997 | 0.4021 | 0.3991 | 0.3992 | 25623 | 19957 | 187758 | 162098 | 0.05551 | 0.02319 | 0.003264 | 0.009834 | 0.008396 | 0.00937 |
| BASOAB | rs236987 | rs9998251 | A | G | 4 | AFF1 | Female only | True | 0.3697 | 0.3728 | 0.3743 | 0.3751 | 25623 | 19957 | 187758 | 162098 | 0.05586 | 0.02425 | 0.002134 | 0.0108 | 0.008523 | 0.009466 |
| BASOAB | rs236987 | rs236996 | A | G | 4 | AFF1 | Female only | True | 0.4008 | 0.4032 | 0.3991 | 0.3991 | 25623 | 19957 | 187758 | 162098 | 0.05509 | 0.02339 | 0.003279 | 0.009854 | 0.008364 | 0.009337 |
| BASOAB | rs236987 | rs4693156 | C | T | 4 | AFF1 | Female only | True | 0.3753 | 0.3783 | 0.379 | 0.3795 | 25623 | 19957 | 187758 | 162098 | 0.05373 | 0.02342 | 0.002959 | 0.009452 | 0.008482 | 0.009427 |
| BASOAB | rs236987 | rs236984 | A | G | 4 | AFF1 | Female only | True | 0.3996 | 0.4023 | 0.399 | 0.399 | 25623 | 19957 | 187758 | 162098 | 0.05592 | 0.02265 | 0.003335 | 0.01009 | 0.008382 | 0.009355 |
| BASOAB | rs236987 | rs236985 | A | G | 4 | AFF1 | Female only | True | 0.4006 | 0.4033 | 0.399 | 0.399 | 25623 | 19957 | 187758 | 162098 | 0.05563 | 0.02236 | 0.003275 | 0.01007 | 0.008364 | 0.009337 |
| BASOAB | rs236987 | rs236987 | T | C | 4 | AFF1 | Female only | True | 0.3946 | 0.3974 | 0.3945 | 0.3947 | 25623 | 19957 | 187758 | 162098 | 0.05711 | 0.02371 | 0.002533 | 0.0112 | 0.008398 | 0.009363 |
| BASOAB | rs236987 | rs3775221 | G | T | 4 | AFF1 | Female only | True | 0.3753 | 0.3783 | 0.379 | 0.3795 | 25623 | 19957 | 187758 | 162098 | 0.05369 | 0.02326 | 0.002941 | 0.009463 | 0.008482 | 0.009427 |
| BASOAB | rs236987 | rs3775220 | G | A | 4 | AFF1 | Female only | True | 0.3753 | 0.3782 | 0.379 | 0.3795 | 25623 | 19957 | 187758 | 162098 | 0.05358 | 0.02315 | 0.002955 | 0.009453 | 0.008482 | 0.009427 |
| BASOAB | rs236987 | rs236992 | A | G | 4 | AFF1 | Female only | True | 0.4026 | 0.405 | 0.3991 | 0.3991 | 25623 | 19957 | 187758 | 162098 | 0.05519 | 0.02305 | 0.003329 | 0.009866 | 0.00839 | 0.009366 |
| BASOAB | rs236987 | rs236993 | G | T | 4 | AFF1 | Female only | True | 0.4027 | 0.4051 | 0.3992 | 0.3992 | 25623 | 19957 | 187758 | 162098 | 0.05521 | 0.02338 | 0.003377 | 0.009887 | 0.008393 | 0.009369 |
| BASOAB | rs236987 | rs342452 | A | G | 4 | AFF1 | Female only | True | 0.4024 | 0.4051 | 0.399 | 0.399 | 25623 | 19957 | 187758 | 162098 | 0.05561 | 0.02232 | 0.003291 | 0.01011 | 0.008391 | 0.009368 |
| BASOAB | rs236987 | rs3775217 | G | A | 4 | AFF1 | Female only | True | 0.3749 | 0.3779 | 0.3791 | 0.3796 | 25623 | 19957 | 187758 | 162098 | 0.05427 | 0.02251 | 0.003116 | 0.00962 | 0.008484 | 0.009432 |
| BASOAB | rs236987 | rs3775214 | G | A | 4 | AFF1 | Female only | True | 0.3983 | 0.4009 | 0.3986 | 0.3988 | 25623 | 19957 | 187758 | 162098 | 0.05553 | 0.02234 | 0.003243 | 0.00988 | 0.00837 | 0.009343 |
| BASOAB | rs236987 | rs342456 | T | C | 4 | AFF1 | Female only | True | 0.4021 | 0.4045 | 0.3989 | 0.399 | 25623 | 19957 | 187758 | 162098 | 0.05587 | 0.02192 | 0.003356 | 0.01019 | 0.00839 | 0.009368 |
| BASOAB | rs236987 | rs1674999 | G | A | 4 | AFF1 | Female only | True | 0.4024 | 0.4047 | 0.3991 | 0.3992 | 25623 | 19957 | 187758 | 162098 | 0.05591 | 0.02224 | 0.003344 | 0.01001 | 0.008383 | 0.009362 |
| BASOAB | rs236987 | rs442177 | G | T | 4 | AFF1 | Female only | True | 0.4101 | 0.4112 | 0.4068 | 0.407 | 25623 | 19957 | 187758 | 162098 | 0.05611 | 0.02064 | 0.003997 | 0.01014 | 0.008335 | 0.009304 |
| BASOAB | rs236987 | rs342438 | A | G | 4 | AFF1 | Female only | True | 0.396 | 0.3985 | 0.3932 | 0.3936 | 25623 | 19957 | 187758 | 162098 | 0.05679 | 0.02324 | 0.002322 | 0.01145 | 0.008398 | 0.009374 |
| BASOAB | rs236987 | rs342437 | A | G | 4 | AFF1 | Female only | True | 0.3936 | 0.3961 | 0.3929 | 0.3933 | 25623 | 19957 | 187758 | 162098 | 0.05587 | 0.02338 | 0.002116 | 0.01138 | 0.008382 | 0.009349 |
| BASOAB | rs236987 | rs1037814 | T | C | 4 | AFF1 | Female only | True | 0.3964 | 0.3992 | 0.3936 | 0.394 | 25623 | 19957 | 187758 | 162098 | 0.05702 | 0.02325 | 0.002357 | 0.01163 | 0.008402 | 0.009381 |
| trait | lead_snp | ID | ALLELE1 | ALLELE0 | CHR | Gene_Symbol | independent_loci_in_sex_P1e2 | in_embl_any_or_gene | A1FREQ_VUMC_female | A1FREQ_VUMC_male | A1FREQ_UKB_female | A1FREQ_UKB_male | N_VUMC_female | N_VUMC_male | N_UKB_female | N_UKB_male | BETA_VUMC_female | BETA_VUMC_male | BETA_UKB_female | BETA_UKB_male | SE_VUMC_female | SE_VUMC_male |
| BASOAB | rs236987 | rs342469 | G | A | 4 | AFF1 | Female only | True | 0.3964 | 0.3992 | 0.3936 | 0.394 | 25623 | 19957 | 187758 | 162098 | 0.05703 | 0.02324 | 0.002358 | 0.01163 | 0.008402 | 0.009381 |

| trait | lead_snp | ID | ALLELE1 | ALLELE0 | CHR | Gene_Symbol | independent_loci_in_sex_P1e2 | in_embl_any_or_gene | A1FREQ_VUMC_female | A1FREQ_VUMC_male | A1FREQ_UKB_female | A1FREQ_UKB_male | N_VUMC_female | N_VUMC_male | N_UKB_female | N_UKB_male | BETA_VUMC_female | BETA_VUMC_male | BETA_UKB_female | BETA_UKB_male | SE_VUMC_female | SE_VUMC_male |
|---|---|---|---|---|---|---|---|---|---|---|---|---|---|---|---|---|---|---|---|---|---|---|
| BASOAB | rs236987 | rs342468 | G | A | 4 | AFF1 | Female only | True | 0.3965 | 0.3993 | 0.3937 | 0.394 | 25623 | 19957 | 187758 | 162098 | 0.05693 | 0.02306 | 0.002343 | 0.01163 | 0.008402 | 0.009382 |
| BASOAB | rs236987 | rs342467 | T | C | 4 | AFF1 | Female only | True | 0.3965 | 0.3992 | 0.3936 | 0.394 | 25623 | 19957 | 187758 | 162098 | 0.05693 | 0.02305 | 0.00239 | 0.01161 | 0.008402 | 0.009382 |
| BASOAB | rs236987 | rs342466 | C | T | 4 | AFF1 | Female only | True | 0.3956 | 0.3983 | 0.3933 | 0.3937 | 25623 | 19957 | 187758 | 162098 | 0.05679 | 0.02341 | 0.002162 | 0.01153 | 0.008412 | 0.009384 |
| BASOAB | rs236987 | rs62308866 | G | A | 4 | AFF1 | Female only | True | 0.3153 | 0.3188 | 0.3242 | 0.3255 | 25623 | 19957 | 187758 | 162098 | 0.05341 | 0.01567 | 0.002713 | 0.009515 | 0.008877 | 0.009869 |
| BASOAB | rs236987 | rs12509389 | G | T | 4 | AFF1 | Female only | True | 0.3152 | 0.3188 | 0.3237 | 0.3251 | 25623 | 19957 | 187758 | 162098 | 0.05328 | 0.01525 | 0.002632 | 0.009855 | 0.008876 | 0.009867 |
| BASOAB | rs236987 | rs12505327 | T | C | 4 | AFF1 | Female only | True | 0.3143 | 0.3182 | 0.3227 | 0.3241 | 25623 | 19957 | 187758 | 162098 | 0.05371 | 0.01583 | 0.002377 | 0.009833 | 0.008888 | 0.009879 |
| BASOAB | rs236987 | rs12509445 | C | T | 4 | AFF1 | Female only | True | 0.3143 | 0.318 | 0.3227 | 0.3241 | 25623 | 19957 | 187758 | 162098 | 0.05372 | 0.0157 | 0.002355 | 0.009817 | 0.008889 | 0.00988 |
| BASOAB | rs236987 | rs12505408 | T | C | 4 | AFF1 | Female only | True | 0.3152 | 0.3188 | 0.3237 | 0.3251 | 25623 | 19957 | 187758 | 162098 | 0.05328 | 0.01529 | 0.002656 | 0.009793 | 0.008876 | 0.009866 |
| BASOAB | rs236987 | rs12505448 | T | C | 4 | AFF1 | Female only | True | 0.3152 | 0.3188 | 0.3238 | 0.3251 | 25623 | 19957 | 187758 | 162098 | 0.05329 | 0.0153 | 0.002651 | 0.009807 | 0.008876 | 0.009866 |
| BASOAB | rs236987 | rs10015288 | T | C | 4 | AFF1 | Female only | True | 0.3153 | 0.3188 | 0.3237 | 0.3251 | 25623 | 19957 | 187758 | 162098 | 0.05272 | 0.01549 | 0.002664 | 0.009847 | 0.00888 | 0.009871 |
| BASOAB | rs236987 | rs28513782 | A | G | 4 | AFF1 | Female only | True | 0.3153 | 0.3188 | 0.3237 | 0.3251 | 25623 | 19957 | 187758 | 162098 | 0.0527 | 0.01549 | 0.002652 | 0.009868 | 0.008879 | 0.00987 |
| BASOAB | rs236987 | rs12645763 | C | T | 4 | AFF1 | Female only | True | 0.315 | 0.3188 | 0.3237 | 0.3251 | 25623 | 19957 | 187758 | 162098 | 0.05308 | 0.01523 | 0.002674 | 0.009742 | 0.008878 | 0.009865 |
| BASOAB | rs236987 | rs1552145 | T | C | 4 | AFF1 | Female only | True | 0.3149 | 0.3188 | 0.3232 | 0.3246 | 25623 | 19957 | 187758 | 162098 | 0.05299 | 0.01507 | 0.002697 | 0.00958 | 0.008879 | 0.009865 |
| BASOAB | rs236987 | rs6834470 | A | C | 4 | AFF1 | Female only | True | 0.3977 | 0.4029 | 0.3999 | 0.401 | 25623 | 19957 | 187758 | 162098 | 0.05011 | 0.009142 | 0.00151 | 0.008663 | 0.008404 | 0.009327 |
| BASOAB | rs236987 | rs7683063 | A | G | 4 | AFF1 | Female only | True | 0.3966 | 0.4017 | 0.3997 | 0.4008 | 25623 | 19957 | 187758 | 162098 | 0.05044 | 0.009023 | 0.001495 | 0.008748 | 0.008428 | 0.009356 |
| BASOAB | rs3873388 | rs3873387 | A | C | 6 | XXbac-BPG248L24.10 | Female only | True | 0.2249 | 0.2244 | 0.2068 | 0.2057 | 25623 | 19957 | 187758 | 162098 | 0.05125 | 0.009797 | 5.23e-04 | 0.006871 | 0.009889 | 0.01108 |
| BASOAB | rs3873388 | rs3873388 | G | A | 6 | XXbac-BPG248L24.10 | Female only | True | 0.2969 | 0.298 | 0.2695 | 0.2675 | 25623 | 19957 | 187758 | 162098 | 0.05029 | 0.01559 | -4.51e-04 | 0.006411 | 0.009063 | 0.01015 |
| BASORE | rs11659727 | rs7241678 | C | T | 18 | RP11-737G21.1 | Female only | False | 0.324 | 0.3266 | 0.3204 | 0.3199 | 25659 | 19975 | 187760 | 162101 | -0.03739 | 0.01382 | -0.002439 | -0.006729 | 0.00884 | 0.009687 |
| BASORE | rs11659727 | rs7237696 | T | C | 18 | RP11-737G21.1 | Female only | False | 0.3283 | 0.3309 | 0.3241 | 0.3234 | 25659 | 19975 | 187760 | 162101 | -0.03849 | 0.009772 | -0.002321 | -0.006826 | 0.008805 | 0.009653 |
| BASORE | rs11659727 | rs62085600 | T | C | 18 | RP11-737G21.1 | Female only | False | 0.3255 | 0.3283 | 0.3209 | 0.3203 | 25659 | 19975 | 187760 | 162101 | -0.03934 | 0.01158 | -0.002294 | -0.006985 | 0.008825 | 0.009671 |
| BASORE | rs11659727 | rs764783 | A | G | 18 | RP11-737G21.1 | Female only | False | 0.3291 | 0.3318 | 0.324 | 0.3233 | 25659 | 19975 | 187760 | 162101 | -0.03905 | 0.009457 | -0.00215 | -0.006748 | 0.008772 | 0.009617 |
| BASORE | rs11659727 | rs3891392 | C | T | 18 | RP11-737G21.1 | Female only | False | 0.3242 | 0.3268 | 0.3203 | 0.3198 | 25659 | 19975 | 187760 | 162101 | -0.03849 | 0.01271 | -0.00236 | -0.00653 | 0.008827 | 0.009673 |
| BASORE | rs11659727 | rs11664356 | T | C | 18 | RP11-737G21.1 | Female only | False | 0.3242 | 0.3269 | 0.3203 | 0.3198 | 25659 | 19975 | 187760 | 162101 | -0.03841 | 0.01281 | -0.002321 | -0.006488 | 0.008827 | 0.009673 |
| BASORE | rs11659727 | rs11664279 | T | G | 18 | RP11-737G21.1 | Female only | False | 0.3347 | 0.3365 | 0.3313 | 0.3313 | 25659 | 19975 | 187760 | 162101 | -0.03904 | 0.01236 | -0.00247 | -0.006202 | 0.008784 | 0.009605 |
| BASORE | rs11659727 | rs7505390 | T | G | 18 | RP11-737G21.1 | Female only | False | 0.3624 | 0.3648 | 0.3577 | 0.3578 | 25659 | 19975 | 187760 | 162101 | -0.03239 | 0.01572 | -0.002785 | -0.004216 | 0.008621 | 0.009416 |
| BASORE | rs11659727 | rs8090359 | T | C | 18 | RP11-737G21.1 | Female only | False | 0.3356 | 0.3373 | 0.3309 | 0.331 | 25659 | 19975 | 187760 | 162101 | -0.03748 | 0.01122 | -0.003152 | -0.005271 | 0.008765 | 0.009589 |
| BASORE | rs11659727 | rs6508397 | A | G | 18 | RP11-737G21.1 | Female only | False | 0.3361 | 0.338 | 0.331 | 0.3311 | 25659 | 19975 | 187760 | 162101 | -0.0371 | 0.01016 | -0.003161 | -0.005131 | 0.008761 | 0.009597 |
| BASORE | rs11659727 | rs8092404 | A | G | 18 | RP11-737G21.1 | Female only | False | 0.3346 | 0.3355 | 0.3283 | 0.3287 | 25659 | 19975 | 187760 | 162101 | -0.0392 | 0.008038 | -0.00254 | -0.004127 | 0.008744 | 0.009603 |

| trait | lead_snp | ID | ALLELE1 | ALLELE0 | CHR | Gene_Symbol | independent_loci_in_sex_P1e2 | in_embl_any_or_gene | A1FREQ_VUMC_female | A1FREQ_VUMC_male | A1FREQ_UKB_female | A1FREQ_UKB_male | N_VUMC_female | N_VUMC_male | N_UKB_female | N_UKB_male | BETA_VUMC_female | BETA_VUMC_male | BETA_UKB_female | BETA_UKB_male | SE_VUMC_female | SE_VUMC_male |
|---|---|---|---|---|---|---|---|---|---|---|---|---|---|---|---|---|---|---|---|---|---|---|
| BASORE | rs11659727 | rs4800678 | T | G | 18 | RP11-737G21.1 | Female only | False | 0.3358 | 0.3366 | 0.3286 | 0.329 | 25659 | 19975 | 187760 | 162101 | -0.04013 | 0.008132 | -0.002352 | -0.003869 | 0.008737 | 0.009596 |
| BASORE | rs11659727 | rs1941236 | G | A | 18 | RP11-737G21.1 | Female only | False | 0.3354 | 0.3362 | 0.3286 | 0.3289 | 25659 | 19975 | 187760 | 162101 | -0.03995 | 0.008035 | -0.002502 | -0.003769 | 0.008742 | 0.009597 |
| BASORE | rs11659727 | rs9961677 | A | G | 18 | RP11-737G21.1 | Female only | False | 0.3353 | 0.336 | 0.3289 | 0.3292 | 25659 | 19975 | 187760 | 162101 | -0.03959 | 0.007569 | -0.002645 | -0.003854 | 0.008746 | 0.009603 |
| BASORE | rs11659727 | rs7232573 | A | C | 18 | RP11-737G21.1 | Female only | False | 0.3353 | 0.336 | 0.3289 | 0.3292 | 25659 | 19975 | 187760 | 162101 | -0.03958 | 0.007569 | -0.00262 | -0.003847 | 0.008746 | 0.009603 |
| BASORE | rs11659727 | rs8093003 | T | C | 18 | RP11-737G21.1 | Female only | False | 0.335 | 0.3355 | 0.3285 | 0.3288 | 25659 | 19975 | 187760 | 162101 | -0.03998 | 0.00782 | -0.002533 | -0.00388 | 0.008751 | 0.009617 |
| BASORE | rs11659727 | rs8093636 | G | A | 18 | RP11-737G21.1 | Female only | False | 0.3353 | 0.3359 | 0.3286 | 0.3288 | 25659 | 19975 | 187760 | 162101 | -0.04028 | 0.008136 | -0.00262 | -0.003869 | 0.008748 | 0.009615 |
| BASORE | rs11659727 | rs7239647 | C | A | 18 | RP11-737G21.1 | Female only | False | 0.3354 | 0.3359 | 0.3288 | 0.3292 | 25659 | 19975 | 187760 | 162101 | -0.03971 | 0.007885 | -0.002594 | -0.003863 | 0.008744 | 0.009606 |
| BASORE | rs11659727 | rs7239248 | T | C | 18 | RP11-737G21.1 | Female only | False | 0.3356 | 0.3362 | 0.3288 | 0.3291 | 25659 | 19975 | 187760 | 162101 | -0.03996 | 0.008142 | -0.002643 | -0.003887 | 0.008743 | 0.009609 |
| BASORE | rs11659727 | rs7228854 | G | T | 18 | RP11-737G21.1 | Female only | False | 0.3349 | 0.3356 | 0.3287 | 0.3291 | 25659 | 19975 | 187760 | 162101 | -0.03947 | 0.007924 | -0.002649 | -0.003803 | 0.008748 | 0.009611 |
| BASORE | rs11659727 | rs9676221 | G | A | 18 | RP11-737G21.1 | Female only | False | 0.3365 | 0.3369 | 0.329 | 0.3293 | 25659 | 19975 | 187760 | 162101 | -0.04013 | 0.008124 | -0.002434 | -0.003926 | 0.008738 | 0.009599 |
| BASORE | rs11659727 | rs7243966 | C | T | 18 | RP11-737G21.1 | Female only | False | 0.3357 | 0.3362 | 0.3288 | 0.3291 | 25659 | 19975 | 187760 | 162101 | -0.04008 | 0.008239 | -0.002721 | -0.003778 | 0.00874 | 0.009607 |
| BASORE | rs11659727 | rs17799466 | A | G | 18 | RP11-737G21.1 | Female only | False | 0.3339 | 0.3346 | 0.3273 | 0.3275 | 25659 | 19975 | 187760 | 162101 | -0.0384 | 0.00732 | -0.002677 | -0.003786 | 0.008716 | 0.009578 |
| BASORE | rs11659727 | rs6508399 | T | C | 18 | RP11-737G21.1 | Female only | False | 0.3373 | 0.3379 | 0.3311 | 0.3316 | 25659 | 19975 | 187760 | 162101 | -0.03954 | 0.008689 | -0.00238 | -0.003905 | 0.008755 | 0.009621 |
| BASORE | rs11659727 | rs28690394 | A | G | 18 | RP11-737G21.1 | Female only | False | 0.3354 | 0.3361 | 0.329 | 0.3293 | 25659 | 19975 | 187760 | 162101 | -0.03963 | 0.007904 | -0.002625 | -0.003845 | 0.008749 | 0.009612 |
| BASORE | rs11659727 | rs28552646 | G | A | 18 | RP11-737G21.1 | Female only | False | 0.3354 | 0.3361 | 0.329 | 0.3293 | 25659 | 19975 | 187760 | 162101 | -0.03973 | 0.007976 | -0.002621 | -0.003984 | 0.00875 | 0.009611 |
| BASORE | rs11659727 | rs9945570 | C | T | 18 | RP11-737G21.1 | Female only | False | 0.3438 | 0.3445 | 0.3395 | 0.3394 | 25659 | 19975 | 187760 | 162101 | -0.0376 | 0.006601 | -0.002977 | -0.004645 | 0.008716 | 0.009557 |
| BASORE | rs11659727 | rs8087920 | A | G | 18 | RP11-737G21.1 | Female only | False | 0.3446 | 0.3451 | 0.3404 | 0.3403 | 25659 | 19975 | 187760 | 162101 | -0.03806 | 0.006638 | -0.00275 | -0.003992 | 0.008701 | 0.009537 |
| BASORE | rs11659727 | rs4800682 | T | C | 18 | RP11-737G21.1 | Female only | False | 0.3443 | 0.3449 | 0.3404 | 0.3403 | 25659 | 19975 | 187760 | 162101 | -0.03835 | 0.005999 | -0.002761 | -0.003973 | 0.008699 | 0.009534 |
| BASORE | rs11659727 | rs17224271 | A | G | 18 | RP11-737G21.1 | Female only | False | 0.3442 | 0.3448 | 0.3403 | 0.3402 | 25659 | 19975 | 187760 | 162101 | -0.0388 | 0.005894 | -0.002866 | -0.004089 | 0.008701 | 0.009534 |
| BASORE | rs11659727 | rs9947076 | T | C | 18 | RP11-737G21.1 | Female only | False | 0.3445 | 0.3449 | 0.3405 | 0.3405 | 25659 | 19975 | 187760 | 162101 | -0.0384 | 0.006167 | -0.002875 | -0.00414 | 0.008701 | 0.009533 |
| BASORE | rs8097343 | rs1785721 | A | C | 18 | RP11-737G21.1 | Female only | False | 0.3753 | 0.3733 | 0.3789 | 0.3795 | 25659 | 19975 | 187760 | 162101 | 0.0399 | 0.01196 | 0.006345 | 0.003806 | 0.008535 | 0.009354 |
| BASORE | rs11659727 | rs4800233 | A | C | 18 | RP11-737G21.1 | Female only | False | 0.3442 | 0.3448 | 0.3404 | 0.3403 | 25659 | 19975 | 187760 | 162101 | -0.03882 | 0.005799 | -0.002845 | -0.004031 | 0.008701 | 0.009533 |
| BASORE | rs11659727 | rs4800684 | C | T | 18 | RP11-737G21.1 | Female only | False | 0.3442 | 0.3448 | 0.3404 | 0.3403 | 25659 | 19975 | 187760 | 162101 | -0.03882 | 0.005799 | -0.002844 | -0.00403 | 0.008701 | 0.009533 |
| BASORE | rs11659727 | rs4800234 | T | G | 18 | RP11-737G21.1 | Female only | False | 0.3326 | 0.3333 | 0.3278 | 0.328 | 25659 | 19975 | 187760 | 162101 | -0.04131 | 0.007453 | -0.002825 | -0.004258 | 0.00877 | 0.009624 |
| BASORE | rs11659727 | rs4800235 | A | C | 18 | RP11-737G21.1 | Female only | False | 0.3442 | 0.3447 | 0.3404 | 0.3403 | 25659 | 19975 | 187760 | 162101 | -0.03901 | 0.006114 | -0.002847 | -0.004023 | 0.008705 | 0.009539 |
| BASORE | rs8097343 | rs1599062 | A | G | 18 | RP11-737G21.1 | Female only | False | 0.3757 | 0.3736 | 0.3789 | 0.3795 | 25659 | 19975 | 187760 | 162101 | 0.03978 | 0.01172 | 0.006326 | 0.00368 | 0.008528 | 0.00935 |
| BASORE | rs8097343 | rs1785700 | A | G | 18 | RP11-737G21.1 | Female only | False | 0.3757 | 0.3736 | 0.3789 | 0.3795 | 25659 | 19975 | 187760 | 162101 | 0.03978 | 0.01175 | 0.006343 | 0.003692 | 0.008529 | 0.00935 |
| BASORE | rs11659727 | rs6508401 | C | T | 18 | RP11-737G21.1 | Female only | False | 0.3326 | 0.3334 | 0.3278 | 0.328 | 25659 | 19975 | 187760 | 162101 | -0.04114 | 0.007438 | -0.002691 | -0.004232 | 0.008768 | 0.009625 |
| BASORE | rs8097343 | rs1121183 | G | A | 18 | RP11-737G21.1 | Female only | False | 0.3756 | 0.3735 | 0.379 | 0.3796 | 25659 | 19975 | 187760 | 162101 | 0.03991 | 0.01156 | 0.006212 | 0.003583 | 0.008526 | 0.009348 |

| trait | lead_snp | ID | ALLELE1 | ALLELE0 | CHR | Gene_Symbol | independent_loci_in_sex_P1e2 | in_embl_any_or_gene | A1FREQ_VUMC_female | A1FREQ_VUMC_male | A1FREQ_UKB_female | A1FREQ_UKB_male | N_VUMC_female | N_VUMC_male | N_UKB_female | N_UKB_male | BETA_VUMC_female | BETA_VUMC_male | BETA_UKB_female | BETA_UKB_male | SE_VUMC_female | SE_VUMC_male |
|---|---|---|---|---|---|---|---|---|---|---|---|---|---|---|---|---|---|---|---|---|---|---|
| BASORE | rs8097343 | rs1674609 | T | G | 18 | RP11-737G21.1 | Female only | False | 0.3756 | 0.3736 | 0.3789 | 0.3795 | 25659 | 19975 | 187760 | 162101 | 0.04003 | 0.01161 | 0.006345 | 0.003558 | 0.008526 | 0.009347 |
| BASORE | rs11659727 | rs9953857 | C | T | 18 | RP11-737G21.1 | Female only | False | 0.3442 | 0.3447 | 0.3404 | 0.3404 | 25659 | 19975 | 187760 | 162101 | -0.0392 | 0.006255 | -0.002868 | -0.003908 | 0.008706 | 0.00954 |
| trait | lead_snp | ID | ALLELE1 | ALLELE0 | CHR | Gene_Symbol | independent_loci_in_sex_P1e2 | in_embl_any_or_gene | A1FREQ_VUMC_female | A1FREQ_VUMC_male | A1FREQ_UKB_female | A1FREQ_UKB_male | N_VUMC_female | N_VUMC_male | N_UKB_female | N_UKB_male | BETA_VUMC_female | BETA_VUMC_male | BETA_UKB_female | BETA_UKB_male | SE_VUMC_female | SE_VUMC_male |
| BASORE | rs8097343 | rs1380193 | T | C | 18 | RP11-737G21.1 | Female only | False | 0.3748 | 0.3727 | 0.3788 | 0.3794 | 25659 | 19975 | 187760 | 162101 | 0.04049 | 0.01125 | 0.006245 | 0.003385 | 0.008515 | 0.009334 |
| BASORE | rs11659727 | rs9959010 | T | C | 18 | RP11-737G21.1 | Female only | False | 0.344 | 0.3447 | 0.3403 | 0.3402 | 25659 | 19975 | 187760 | 162101 | -0.03977 | 0.006565 | -0.002858 | -0.00395 | 0.008706 | 0.009536 |
| BASORE | rs8097343 | rs1785754 | T | C | 18 | RP11-737G21.1 | Female only | False | 0.376 | 0.3736 | 0.3791 | 0.3796 | 25659 | 19975 | 187760 | 162101 | 0.04099 | 0.01126 | 0.006215 | 0.003379 | 0.008527 | 0.009346 |
| BASORE | rs8097343 | rs1893241 | A | G | 18 | RP11-737G21.1 | Female only | False | 0.3599 | 0.3566 | 0.3624 | 0.3621 | 25659 | 19975 | 187760 | 162101 | 0.04101 | 0.009077 | 0.007828 | 0.005376 | 0.008598 | 0.009464 |
| BASORE | rs11659727 | rs59490267 | T | C | 18 | RP11-737G21.1 | Female only | False | 0.3636 | 0.3642 | 0.357 | 0.3577 | 25659 | 19975 | 187760 | 162101 | -0.04269 | 0.003357 | -0.005158 | -0.005163 | 0.008575 | 0.009427 |
| BASORE | rs8097343 | rs7228892 | T | C | 18 | RP11-737G21.1 | Female only | False | 0.36 | 0.3566 | 0.3625 | 0.3621 | 25659 | 19975 | 187760 | 162101 | 0.04102 | 0.009094 | 0.007726 | 0.005323 | 0.008592 | 0.009453 |
| BASORE | rs11659727 | rs7242725 | A | G | 18 | RP11-737G21.1 | Female only | False | 0.3813 | 0.3847 | 0.376 | 0.3764 | 25659 | 19975 | 187760 | 162101 | -0.05275 | -0.002916 | -0.001631 | -0.007065 | 0.008473 | 0.009316 |
| BASORE | rs11659727 | rs17224299 | A | G | 18 | RP11-737G21.1 | Female only | False | 0.3809 | 0.3842 | 0.376 | 0.3764 | 25659 | 19975 | 187760 | 162101 | -0.05279 | -0.002992 | -0.001656 | -0.007072 | 0.008483 | 0.009329 |
| BASORE | rs8097343 | rs4800685 | T | C | 18 | RP11-737G21.1 | Female only | False | 0.3593 | 0.3561 | 0.3626 | 0.3623 | 25659 | 19975 | 187760 | 162101 | 0.04149 | 0.009144 | 0.007739 | 0.005363 | 0.008579 | 0.009441 |
| BASORE | rs11659727 | rs4800686 | G | A | 18 | RP11-737G21.1 | Female only | False | 0.377 | 0.3803 | 0.3723 | 0.3729 | 25659 | 19975 | 187760 | 162101 | -0.053 | 3.34e-04 | -0.001702 | -0.00675 | 0.008503 | 0.009342 |
| BASORE | rs11659727 | rs4800687 | T | C | 18 | RP11-737G21.1 | Female only | False | 0.3809 | 0.3843 | 0.376 | 0.3765 | 25659 | 19975 | 187760 | 162101 | -0.05328 | -0.001963 | -0.001681 | -0.006988 | 0.008484 | 0.009329 |
| BASORE | rs11659727 | rs4800688 | T | C | 18 | RP11-737G21.1 | Female only | False | 0.3776 | 0.381 | 0.3726 | 0.3732 | 25659 | 19975 | 187760 | 162101 | -0.0523 | 0.001216 | -0.001534 | -0.006642 | 0.008498 | 0.00934 |
| BASORE | rs11659727 | rs4800689 | C | T | 18 | RP11-737G21.1 | Female only | False | 0.3811 | 0.3844 | 0.3763 | 0.3767 | 25659 | 19975 | 187760 | 162101 | -0.05298 | -0.00215 | -0.001627 | -0.007064 | 0.008484 | 0.009328 |
| BASORE | rs11659727 | rs4800690 | A | C | 18 | RP11-737G21.1 | Female only | False | 0.3773 | 0.3806 | 0.3725 | 0.3731 | 25659 | 19975 | 187760 | 162101 | -0.05192 | 4.51e-04 | -0.001767 | -0.006761 | 0.008503 | 0.009337 |
| BASORE | rs11659727 | rs11659727 | T | G | 18 | RP11-737G21.1 | Female only | False | 0.3841 | 0.3875 | 0.378 | 0.3785 | 25659 | 19975 | 187760 | 162101 | -0.05385 | -0.002857 | -0.001643 | -0.006674 | 0.008479 | 0.009315 |
| BASORE | rs11659727 | rs10853659 | C | T | 18 | RP11-737G21.1 | Female only | False | 0.3799 | 0.3834 | 0.3741 | 0.3749 | 25659 | 19975 | 187760 | 162101 | -0.0531 | 2.11e-04 | -0.00157 | -0.006391 | 0.008498 | 0.009329 |
| BASORE | rs11659727 | rs7236675 | T | C | 18 | RP11-737G21.1 | Female only | False | 0.3798 | 0.3833 | 0.3741 | 0.3748 | 25659 | 19975 | 187760 | 162101 | -0.05298 | 1.66e-04 | -0.001519 | -0.006331 | 0.008499 | 0.00933 |
| BASORE | rs11659727 | rs4800691 | G | T | 18 | RP11-737G21.1 | Female only | False | 0.3837 | 0.387 | 0.3778 | 0.3784 | 25659 | 19975 | 187760 | 162101 | -0.05365 | -0.002284 | -0.001436 | -0.006632 | 0.008481 | 0.009318 |
| BASORE | rs11659727 | rs62085648 | T | C | 18 | RP11-737G21.1 | Female only | False | 0.38 | 0.3832 | 0.3741 | 0.3749 | 25659 | 19975 | 187760 | 162101 | -0.05284 | 2.04e-04 | -0.001587 | -0.006464 | 0.008501 | 0.009333 |
| BASORE | rs8097343 | rs1674611 | G | A | 18 | RP11-737G21.1 | Female only | False | 0.3546 | 0.3519 | 0.3588 | 0.3591 | 25659 | 19975 | 187760 | 162101 | 0.04094 | 0.009949 | 0.006049 | 0.004197 | 0.008645 | 0.009506 |
| BASORE | rs8097343 | rs1459099 | C | T | 18 | RP11-737G21.1 | Female only | False | 0.406 | 0.4037 | 0.4092 | 0.4089 | 25659 | 19975 | 187760 | 162101 | 0.0449 | 0.01323 | 0.006462 | 0.006791 | 0.008412 | 0.009239 |
| BASORE | rs8097343 | rs2199147 | C | T | 18 | RP11-737G21.1 | Female only | False | 0.4887 | 0.4837 | 0.4838 | 0.483 | 25659 | 19975 | 187760 | 162101 | 0.04331 | 0.006275 | 0.007849 | 0.00663 | 0.008284 | 0.009091 |
| BASORE | rs8097343 | rs2199148 | G | T | 18 | RP11-737G21.1 | Female only | False | 0.4522 | 0.4487 | 0.4489 | 0.4481 | 25659 | 19975 | 187760 | 162101 | 0.04313 | 0.006143 | 0.00584 | 0.005978 | 0.008311 | 0.009122 |
| BASORE | rs8097343 | rs2879282 | A | G | 18 | RP11-737G21.1 | Female only | False | 0.407 | 0.4046 | 0.4092 | 0.409 | 25659 | 19975 | 187760 | 162101 | 0.04503 | 0.01311 | 0.006469 | 0.006807 | 0.008425 | 0.009255 |
| BASORE | rs8097343 | rs4548960 | A | G | 18 | RP11-737G21.1 | Female only | False | 0.4071 | 0.4047 | 0.4092 | 0.409 | 25659 | 19975 | 187760 | 162101 | 0.04516 | 0.01289 | 0.006464 | 0.006807 | 0.008425 | 0.009254 |

| trait | lead_snp | ID | ALLELE1 | ALLELE0 | CHR | Gene_Symbol | independent_loci_in_sex_P1e2 | in_embl_any_or_gene | A1FREQ_VUMC_female | A1FREQ_VUMC_male | A1FREQ_UKB_female | A1FREQ_UKB_male | N_VUMC_female | N_VUMC_male | N_UKB_female | N_UKB_male | BETA_VUMC_female | BETA_VUMC_male | BETA_UKB_female | BETA_UKB_male | SE_VUMC_female | SE_VUMC_male |
|---|---|---|---|---|---|---|---|---|---|---|---|---|---|---|---|---|---|---|---|---|---|---|
| BASORE | rs8097343 | rs9950076 | A | G | 18 | RP11-737G21.1 | Female only | False | 0.4166 | 0.4133 | 0.4157 | 0.4153 | 25659 | 19975 | 187760 | 162101 | 0.0433 | 0.01008 | 0.006475 | 0.006193 | 0.008397 | 0.009208 |
| BASORE | rs8097343 | rs2338875 | A | G | 18 | RP11-737G21.1 | Female only | False | 0.4166 | 0.4132 | 0.4156 | 0.4152 | 25659 | 19975 | 187760 | 162101 | 0.04317 | 0.01023 | 0.006323 | 0.006122 | 0.008395 | 0.00921 |
| BASORE | rs8097343 | rs4638665 | A | C | 18 | RP11-737G21.1 | Female only | False | 0.3553 | 0.3524 | 0.3592 | 0.3595 | 25659 | 19975 | 187760 | 162101 | 0.04148 | 0.009735 | 0.006028 | 0.004354 | 0.00864 | 0.009501 |
| BASORE | rs8097343 | rs9956169 | T | C | 18 | RP11-737G21.1 | Female only | False | 0.4522 | 0.4486 | 0.4488 | 0.448 | 25659 | 19975 | 187760 | 162101 | 0.04303 | 0.006217 | 0.005628 | 0.005853 | 0.008308 | 0.009123 |
| BASORE | rs8097343 | rs7244203 | G | A | 18 | RP11-737G21.1 | Female only | False | 0.4166 | 0.4132 | 0.4156 | 0.4152 | 25659 | 19975 | 187760 | 162101 | 0.04324 | 0.01023 | 0.00632 | 0.006115 | 0.008395 | 0.00921 |
| BASORE | rs8097343 | rs1380192 | T | G | 18 | RP11-737G21.1 | Female only | False | 0.4166 | 0.4132 | 0.4156 | 0.4152 | 25659 | 19975 | 187760 | 162101 | 0.04323 | 0.01023 | 0.006306 | 0.006151 | 0.008395 | 0.00921 |
| BASORE | rs8097343 | rs1870431 | A | G | 18 | RP11-737G21.1 | Female only | False | 0.4173 | 0.414 | 0.4172 | 0.4167 | 25659 | 19975 | 187760 | 162101 | 0.04241 | 0.0125 | 0.006244 | 0.006513 | 0.008373 | 0.009185 |
| BASORE | rs8097343 | rs9962616 | T | C | 18 | RP11-737G21.1 | Female only | False | 0.4166 | 0.4132 | 0.4156 | 0.4152 | 25659 | 19975 | 187760 | 162101 | 0.04319 | 0.01023 | 0.006311 | 0.006162 | 0.008395 | 0.00921 |
| BASORE | rs8097343 | rs9963002 | A | G | 18 | RP11-737G21.1 | Female only | False | 0.417 | 0.4137 | 0.416 | 0.4155 | 25659 | 19975 | 187760 | 162101 | 0.04317 | 0.009246 | 0.006429 | 0.006175 | 0.008396 | 0.009201 |
| BASORE | rs8097343 | rs1459098 | T | C | 18 | RP11-737G21.1 | Female only | False | 0.4519 | 0.4484 | 0.4492 | 0.4483 | 25659 | 19975 | 187760 | 162101 | 0.043 | 0.005291 | 0.005838 | 0.005923 | 0.008294 | 0.009101 |
| BASORE | rs8097343 | rs1380191 | T | C | 18 | RP11-737G21.1 | Female only | False | 0.4171 | 0.4137 | 0.416 | 0.4155 | 25659 | 19975 | 187760 | 162101 | 0.04302 | 0.009286 | 0.006461 | 0.006102 | 0.008396 | 0.009199 |
| BASORE | rs8097343 | rs1380190 | A | G | 18 | RP11-737G21.1 | Female only | False | 0.417 | 0.4137 | 0.416 | 0.4155 | 25659 | 19975 | 187760 | 162101 | 0.04303 | 0.009207 | 0.006448 | 0.006104 | 0.008396 | 0.0092 |
| BASORE | rs8097343 | rs9957953 | C | A | 18 | RP11-737G21.1 | Female only | False | 0.4175 | 0.4141 | 0.4165 | 0.4162 | 25659 | 19975 | 187760 | 162101 | 0.0428 | 0.009032 | 0.006614 | 0.006344 | 0.008394 | 0.009203 |
| BASORE | rs8097343 | rs4800700 | T | C | 18 | RP11-737G21.1 | Female only | False | 0.4171 | 0.4135 | 0.4162 | 0.4157 | 25659 | 19975 | 187760 | 162101 | 0.04362 | 0.009034 | 0.006852 | 0.006565 | 0.008414 | 0.009214 |
| BASORE | rs8097343 | rs4800701 | G | A | 18 | RP11-737G21.1 | Female only | False | 0.4178 | 0.4142 | 0.4164 | 0.4159 | 25659 | 19975 | 187760 | 162101 | 0.0436 | 0.008077 | 0.007041 | 0.006433 | 0.008415 | 0.009222 |
| BASORE | rs8097343 | rs7235471 | C | A | 18 | RP11-737G21.1 | Female only | False | 0.4367 | 0.4334 | 0.4367 | 0.4363 | 25659 | 19975 | 187760 | 162101 | 0.04673 | 0.003228 | 0.007392 | 0.005114 | 0.008352 | 0.009144 |
| BASORE | rs8097343 | rs8082926 | T | C | 18 | RP11-737G21.1 | Female only | False | 0.344 | 0.3416 | 0.3501 | 0.3504 | 25659 | 19975 | 187760 | 162101 | 0.04525 | 0.007679 | 0.01027 | 0.005873 | 0.008712 | 0.009558 |
| BASORE | rs8097343 | rs9965696 | A | G | 18 | RP11-737G21.1 | Female only | False | 0.4722 | 0.4683 | 0.4699 | 0.4693 | 25659 | 19975 | 187760 | 162101 | 0.04822 | 6.67e-04 | 0.00703 | 0.005684 | 0.008292 | 0.009093 |
| BASORE | rs8097343 | rs609184 | C | T | 18 | RP11-737G21.1 | Female only | False | 0.3792 | 0.3764 | 0.3829 | 0.3828 | 25659 | 19975 | 187760 | 162101 | 0.04513 | 0.003707 | 0.009354 | 0.005208 | 0.008531 | 0.009362 |
| BASORE | rs8097343 | rs2338876 | C | T | 18 | RP11-737G21.1 | Female only | False | 0.3445 | 0.342 | 0.3506 | 0.3508 | 25659 | 19975 | 187760 | 162101 | 0.04537 | 0.007436 | 0.01036 | 0.005711 | 0.008702 | 0.00955 |
| BASORE | rs8097343 | rs2959359 | C | T | 18 | RP11-737G21.1 | Female only | False | 0.3444 | 0.342 | 0.3501 | 0.3503 | 25659 | 19975 | 187760 | 162101 | 0.04558 | 0.00753 | 0.01042 | 0.005361 | 0.008697 | 0.009546 |
| BASORE | rs8097343 | rs2959358 | A | C | 18 | RP11-737G21.1 | Female only | False | 0.3417 | 0.3394 | 0.3505 | 0.3506 | 25659 | 19975 | 187760 | 162101 | 0.04608 | 0.007224 | 0.0106 | 0.005501 | 0.008712 | 0.009553 |
| BASORE | rs8097343 | rs2926307 | A | G | 18 | RP11-737G21.1 | Female only | False | 0.3419 | 0.3395 | 0.3497 | 0.3499 | 25659 | 19975 | 187760 | 162101 | 0.04598 | 0.007224 | 0.01051 | 0.005405 | 0.008712 | 0.009562 |
| BASORE | rs8097343 | rs2938675 | A | C | 18 | RP11-737G21.1 | Female only | False | 0.3792 | 0.3766 | 0.3833 | 0.383 | 25659 | 19975 | 187760 | 162101 | 0.045 | 0.003667 | 0.009629 | 0.005398 | 0.008494 | 0.009321 |
| BASORE | rs8097343 | rs9960871 | T | C | 18 | RP11-737G21.1 | Female only | False | 0.4726 | 0.4688 | 0.4703 | 0.4696 | 25659 | 19975 | 187760 | 162101 | 0.0479 | 7.72e-04 | 0.006977 | 0.005847 | 0.008282 | 0.009083 |
| BASORE | rs8097343 | rs8091309 | G | A | 18 | RP11-737G21.1 | Female only | False | 0.384 | 0.3817 | 0.3874 | 0.3882 | 25659 | 19975 | 187760 | 162101 | 0.04313 | 0.002087 | 0.008789 | 0.004009 | 0.008522 | 0.00935 |
| BASORE | rs8097343 | rs8097343 | A | G | 18 | RP11-737G21.1 | Female only | False | 0.4695 | 0.4666 | 0.4696 | 0.4689 | 25659 | 19975 | 187760 | 162101 | 0.04826 | 7.35e-04 | 0.007179 | 0.005622 | 0.008268 | 0.009062 |
| BASORE | rs8097343 | rs905744 | C | A | 18 | RP11-737G21.1 | Female only | False | 0.4716 | 0.4687 | 0.4701 | 0.4693 | 25659 | 19975 | 187760 | 162101 | 0.04752 | 5.22e-04 | 0.007187 | 0.005778 | 0.008288 | 0.009084 |
| BASORE | rs8097343 | rs12607839 | C | T | 18 | RP11-737G21.1 | Female only | False | 0.4451 | 0.4428 | 0.4435 | 0.4424 | 25659 | 19975 | 187760 | 162101 | 0.04347 | 0.01313 | 0.009001 | 0.007993 | 0.008318 | 0.009075 |

| trait | lead_snp | ID | ALLELE1 | ALLELE0 | CHR | Gene_Symbol | independent_loci_in_sex_P1e2 | in_embl_any_or_gene | A1FREQ_VUMC_female | A1FREQ_VUMC_male | A1FREQ_UKB_female | A1FREQ_UKB_male | N_VUMC_female | N_VUMC_male | N_UKB_female | N_UKB_male | BETA_VUMC_female | BETA_VUMC_male | BETA_UKB_female | BETA_UKB_male | SE_VUMC_female | SE_VUMC_male |
|---|---|---|---|---|---|---|---|---|---|---|---|---|---|---|---|---|---|---|---|---|---|---|
| BASORE | rs8097343 | rs12608116 | T | C | 18 | RP11-737G21.1 | Female only | False | 0.4463 | 0.4439 | 0.4426 | 0.4417 | 25659 | 19975 | 187760 | 162101 | 0.04417 | 0.01278 | 0.008967 | 0.008014 | 0.008358 | 0.009118 |
| MONOAB | rs10874656 | rs3103173 | A | G | 1 | GFI1 | Female only | True | 0.2036 | 0.1985 | 0.1976 | 0.1972 | 25647 | 19931 | 187758 | 162098 | 0.052 | 0.02526 | 0.05924 | 0.04731 | 0.01035 | 0.01145 |
| MONOAB | rs10874656 | rs3122320 | C | T | 1 | GFI1 | Female only | True | 0.2036 | 0.1985 | 0.1976 | 0.1972 | 25647 | 19931 | 187758 | 162098 | 0.05201 | 0.02527 | 0.05924 | 0.04731 | 0.01035 | 0.01145 |
| MONOAB | rs10874656 | rs1487539 | C | T | 1 | GFI1 | Female only | True | 0.205 | 0.2004 | 0.1988 | 0.1985 | 25647 | 19931 | 187758 | 162098 | 0.05119 | 0.02442 | 0.05894 | 0.04804 | 0.01033 | 0.01143 |
| MONOAB | rs10874656 | rs3103177 | A | G | 1 | GFI1 | Female only | True | 0.2052 | 0.2006 | 0.1987 | 0.1985 | 25647 | 19931 | 187758 | 162098 | 0.05126 | 0.02432 | 0.05892 | 0.04805 | 0.01033 | 0.01142 |
| MONOAB | rs10874656 | rs2046621 | T | G | 1 | GFI1 | Female only | True | 0.205 | 0.2005 | 0.1987 | 0.1985 | 25647 | 19931 | 187758 | 162098 | 0.05119 | 0.02425 | 0.05898 | 0.04816 | 0.01031 | 0.01141 |
| trait | lead_snp | ID | ALLELE1 | ALLELE0 | CHR | Gene_Symbol | independent_loci_in_sex_P1e2 | in_embl_any_or_gene | A1FREQ_VUMC_female | A1FREQ_VUMC_male | A1FREQ_UKB_female | A1FREQ_UKB_male | N_VUMC_female | N_VUMC_male | N_UKB_female | N_UKB_male | BETA_VUMC_female | BETA_VUMC_male | BETA_UKB_female | BETA_UKB_male | SE_VUMC_female | SE_VUMC_male |
| MONOAB | rs10874656 | rs6668685 | G | T | 1 | GFI1 | Female only | True | 0.2051 | 0.2007 | 0.1987 | 0.1985 | 25647 | 19931 | 187758 | 162098 | 0.05115 | 0.02378 | 0.05898 | 0.04812 | 0.01031 | 0.0114 |
| MONOAB | rs10874656 | rs11166568 | C | T | 1 | GFI1 | Female only | True | 0.205 | 0.2005 | 0.1987 | 0.1985 | 25647 | 19931 | 187758 | 162098 | 0.05119 | 0.02425 | 0.05898 | 0.04816 | 0.01031 | 0.01141 |
| MONOAB | rs10874656 | rs11166575 | T | C | 1 | GFI1 | Female only | True | 0.205 | 0.2005 | 0.1987 | 0.1985 | 25647 | 19931 | 187758 | 162098 | 0.05119 | 0.02425 | 0.05899 | 0.04816 | 0.01031 | 0.01141 |
| MONOAB | rs10874656 | rs79491239 | T | C | 1 | GFI1 | Female only | True | 0.1982 | 0.1938 | 0.1987 | 0.1985 | 25647 | 19931 | 187758 | 162098 | 0.05077 | 0.02366 | 0.05907 | 0.04806 | 0.01033 | 0.01142 |
| MONOAB | rs10874656 | rs6604091 | C | A | 1 | GFI1 | Female only | True | 0.205 | 0.2005 | 0.1987 | 0.1985 | 25647 | 19931 | 187758 | 162098 | 0.05113 | 0.02402 | 0.05898 | 0.04816 | 0.0103 | 0.01139 |
| MONOAB | rs10874656 | rs7542747 | G | T | 1 | GFI1 | Female only | True | 0.205 | 0.2005 | 0.1987 | 0.1985 | 25647 | 19931 | 187758 | 162098 | 0.05113 | 0.02401 | 0.05898 | 0.04816 | 0.0103 | 0.01139 |
| MONOAB | rs10874656 | rs1487537 | C | T | 1 | GFI1 | Female only | True | 0.205 | 0.2005 | 0.1987 | 0.1985 | 25647 | 19931 | 187758 | 162098 | 0.05113 | 0.02401 | 0.05899 | 0.04815 | 0.0103 | 0.01139 |
| MONOAB | rs10874656 | rs7545445 | A | C | 1 | GFI1 | Female only | True | 0.2049 | 0.2004 | 0.1987 | 0.1985 | 25647 | 19931 | 187758 | 162098 | 0.05115 | 0.02414 | 0.05898 | 0.04817 | 0.0103 | 0.0114 |
| MONOAB | rs10874656 | rs1906279 | T | C | 1 | GFI1 | Female only | True | 0.205 | 0.2005 | 0.1987 | 0.1985 | 25647 | 19931 | 187758 | 162098 | 0.05146 | 0.024 | 0.05899 | 0.04815 | 0.0103 | 0.01139 |
| MONOAB | rs10874656 | rs6603963 | G | A | 1 | GFI1 | Female only | True | 0.2043 | 0.1999 | 0.1987 | 0.1985 | 25647 | 19931 | 187758 | 162098 | 0.0513 | 0.02381 | 0.05901 | 0.04815 | 0.01028 | 0.01137 |
| MONOAB | rs10874656 | rs7512301 | G | A | 1 | GFI1 | Female only | True | 0.2044 | 0.1999 | 0.1988 | 0.1986 | 25647 | 19931 | 187758 | 162098 | 0.05091 | 0.02384 | 0.05886 | 0.04825 | 0.01028 | 0.01137 |
| MONOAB | rs10874656 | rs6692060 | G | A | 1 | GFI1 | Female only | True | 0.1999 | 0.1955 | 0.1987 | 0.1985 | 25647 | 19931 | 187758 | 162098 | 0.05059 | 0.02363 | 0.05907 | 0.04818 | 0.01022 | 0.01131 |
| MONOAB | rs10874656 | rs10747411 | A | G | 1 | GFI1 | Female only | True | 0.2042 | 0.1999 | 0.1987 | 0.1985 | 25647 | 19931 | 187758 | 162098 | 0.05087 | 0.02388 | 0.05905 | 0.04816 | 0.01027 | 0.01137 |
| MONOAB | rs10874656 | rs6603967 | C | T | 1 | GFI1 | Female only | True | 0.2043 | 0.2 | 0.1987 | 0.1985 | 25647 | 19931 | 187758 | 162098 | 0.05092 | 0.0237 | 0.05909 | 0.04823 | 0.01027 | 0.01136 |
| MONOAB | rs10874656 | rs111434556 | C | T | 1 | GFI1 | Female only | True | 0.1988 | 0.1945 | 0.1988 | 0.1985 | 25647 | 19931 | 187758 | 162098 | 0.05097 | 0.02381 | 0.05912 | 0.04825 | 0.01028 | 0.01137 |
| MONOAB | rs10874656 | rs76600721 | T | C | 1 | GFI1 | Female only | True | 0.1988 | 0.1945 | 0.1988 | 0.1985 | 25647 | 19931 | 187758 | 162098 | 0.05099 | 0.02378 | 0.05913 | 0.04822 | 0.01028 | 0.01137 |
| MONOAB | rs10874656 | rs80215994 | T | C | 1 | GFI1 | Female only | True | 0.1988 | 0.1945 | 0.1988 | 0.1985 | 25647 | 19931 | 187758 | 162098 | 0.05098 | 0.02379 | 0.05909 | 0.04815 | 0.01028 | 0.01137 |
| MONOAB | rs10874656 | rs74959960 | A | G | 1 | GFI1 | Female only | True | 0.1987 | 0.1944 | 0.1987 | 0.1985 | 25647 | 19931 | 187758 | 162098 | 0.05086 | 0.0238 | 0.05913 | 0.04802 | 0.01028 | 0.01137 |
| MONOAB | rs10874656 | rs2025607 | T | C | 1 | GFI1 | Female only | True | 0.1987 | 0.1944 | 0.1987 | 0.1985 | 25647 | 19931 | 187758 | 162098 | 0.0509 | 0.02381 | 0.05918 | 0.04805 | 0.01028 | 0.01137 |
| MONOAB | rs10874656 | rs59343017 | A | G | 1 | GFI1 | Female only | True | 0.1981 | 0.1938 | 0.1987 | 0.1985 | 25647 | 19931 | 187758 | 162098 | 0.05126 | 0.02491 | 0.0592 | 0.0481 | 0.01034 | 0.01144 |
| MONOAB | rs10874656 | rs78220437 | G | T | 1 | GFI1 | Female only | True | 0.1981 | 0.1938 | 0.1986 | 0.1984 | 25647 | 19931 | 187758 | 162098 | 0.0514 | 0.02484 | 0.0594 | 0.04818 | 0.01034 | 0.01144 |

| trait | lead_snp | ID | ALLELE1 | ALLELE0 | CHR | Gene_Symbol | independent_loci_in_sex_P1e2 | in_embl_any_or_gene | A1FREQ_VUMC_female | A1FREQ_VUMC_male | A1FREQ_UKB_female | A1FREQ_UKB_male | N_VUMC_female | N_VUMC_male | N_UKB_female | N_UKB_male | BETA_VUMC_female | BETA_VUMC_male | BETA_UKB_female | BETA_UKB_male | SE_VUMC_female | SE_VUMC_male |
|---|---|---|---|---|---|---|---|---|---|---|---|---|---|---|---|---|---|---|---|---|---|---|
| MONOAB | rs10874656 | rs112032517 | T | C | 1 | GFI1 | Female only | True | 0.1975 | 0.1931 | 0.1981 | 0.1979 | 25647 | 19931 | 187758 | 162098 | 0.05066 | 0.02448 | 0.05907 | 0.04765 | 0.01037 | 0.01147 |
| MONOAB | rs10874656 | rs59679370 | T | C | 1 | GFI1 | Female only | True | 0.1983 | 0.194 | 0.1986 | 0.1985 | 25647 | 19931 | 187758 | 162098 | 0.05217 | 0.02449 | 0.0602 | 0.04841 | 0.01034 | 0.01144 |
| MONOAB | rs10874656 | 1:92925962 | G | T | 1 | GFI1 | Female only | True | 0.1993 | 0.196 | | | 25647 | 19931 | | | 0.05228 | 0.02321 | | | 0.01032 | 0.0114 |
| MONOAB | rs10874656 | rs76593661 | G | T | 1 | GFI1 | Female only | True | 0.1999 | 0.1967 | 0.2002 | 0.2002 | 25647 | 19931 | 187758 | 162098 | 0.05258 | 0.02431 | 0.05897 | 0.04699 | 0.01033 | 0.01141 |
| MONOAB | rs10874656 | rs79164177 | G | A | 1 | GFI1 | Female only | True | 0.1999 | 0.1967 | 0.2003 | 0.2003 | 25647 | 19931 | 187758 | 162098 | 0.05256 | 0.02429 | 0.05888 | 0.04702 | 0.01033 | 0.01141 |
| MONOAB | rs10874656 | rs7518357 | G | A | 1 | GFI1 | Female only | True | 0.1992 | 0.195 | 0.1987 | 0.1986 | 25647 | 19931 | 187758 | 162098 | 0.05205 | 0.02567 | 0.05989 | 0.04859 | 0.01034 | 0.01143 |
| MONOAB | rs10874656 | rs6676846 | G | A | 1 | GFI1 | Female only | True | 0.2122 | 0.2086 | 0.2064 | 0.2063 | 25647 | 19931 | 187758 | 162098 | 0.05544 | 0.02416 | 0.05984 | 0.05076 | 0.01021 | 0.01123 |
| MONOAB | rs10874656 | rs11164605 | G | A | 1 | GFI1 | Female only | True | 0.2121 | 0.2086 | 0.2064 | 0.2062 | 25647 | 19931 | 187758 | 162098 | 0.0554 | 0.02437 | 0.05984 | 0.05084 | 0.01021 | 0.01123 |
| MONOAB | rs10874656 | rs10874656 | T | C | 1 | GFI1 | Female only | True | 0.2116 | 0.208 | 0.2063 | 0.2062 | 25647 | 19931 | 187758 | 162098 | 0.05592 | 0.0238 | 0.05998 | 0.05091 | 0.0102 | 0.01123 |
| MONOAB | rs10874656 | rs6603975 | A | G | 1 | GFI1 | Female only | True | 0.2119 | 0.2083 | 0.2064 | 0.2062 | 25647 | 19931 | 187758 | 162098 | 0.05544 | 0.02409 | 0.05981 | 0.05083 | 0.0102 | 0.01122 |
| MONOAB | rs10874656 | rs7536295 | A | G | 1 | GFI1 | Female only | True | 0.212 | 0.2083 | 0.2064 | 0.2062 | 25647 | 19931 | 187758 | 162098 | 0.0558 | 0.02409 | 0.05983 | 0.05085 | 0.0102 | 0.01122 |
| MONOAB | rs10874656 | rs912795 | A | G | 1 | GFI1 | Female only | True | 0.2115 | 0.2078 | 0.2062 | 0.2061 | 25647 | 19931 | 187758 | 162098 | 0.05529 | 0.02388 | 0.06013 | 0.05094 | 0.01021 | 0.01124 |
| MONOAB | rs10874656 | rs2031494 | T | C | 1 | GFI1 | Female only | True | 0.2117 | 0.2079 | 0.2062 | 0.2062 | 25647 | 19931 | 187758 | 162098 | 0.05468 | 0.02444 | 0.05987 | 0.05078 | 0.01021 | 0.01123 |
| MONOAB | rs10874656 | rs11164654 | T | C | 1 | GFI1 | Female only | True | 0.2116 | 0.2079 | 0.2063 | 0.2063 | 25647 | 19931 | 187758 | 162098 | 0.05444 | 0.02425 | 0.05997 | 0.05068 | 0.01021 | 0.01123 |
| MONOAB | rs10874656 | rs4970696 | T | C | 1 | GFI1 | Female only | True | 0.2114 | 0.2076 | 0.2062 | 0.2062 | 25647 | 19931 | 187758 | 162098 | 0.05476 | 0.02395 | 0.06018 | 0.05078 | 0.01021 | 0.01124 |
| MONOAB | rs10874656 | rs962221 | C | T | 1 | GFI1 | Female only | True | 0.2117 | 0.2079 | 0.2063 | 0.2063 | 25647 | 19931 | 187758 | 162098 | 0.05411 | 0.02443 | 0.05991 | 0.05068 | 0.01021 | 0.01123 |
| MONOAB | rs10874656 | rs962222 | C | T | 1 | GFI1 | Female only | True | 0.2117 | 0.2079 | 0.2063 | 0.2063 | 25647 | 19931 | 187758 | 162098 | 0.05431 | 0.02438 | 0.05998 | 0.05066 | 0.01021 | 0.01123 |
| MONOAB | rs10874656 | rs2391159 | T | C | 1 | GFI1 | Female only | True | 0.2116 | 0.2078 | 0.2063 | 0.2063 | 25647 | 19931 | 187758 | 162098 | 0.05407 | 0.02475 | 0.05992 | 0.05067 | 0.0102 | 0.01123 |
| MONOAB | rs10874656 | rs1570836 | T | C | 1 | GFI1 | Female only | True | 0.2117 | 0.2079 | 0.2063 | 0.2063 | 25647 | 19931 | 187758 | 162098 | 0.05406 | 0.02478 | 0.05992 | 0.05067 | 0.0102 | 0.01123 |
| MONOAB | rs10874656 | rs6603977 | G | A | 1 | GFI1 | Female only | True | 0.2124 | 0.2085 | 0.2065 | 0.2065 | 25647 | 19931 | 187758 | 162098 | 0.05431 | 0.02486 | 0.06006 | 0.05042 | 0.0102 | 0.01122 |
| MONOAB | rs10874656 | rs4970698 | T | C | 1 | GFI1 | Female only | True | 0.2114 | 0.2076 | 0.2062 | 0.2062 | 25647 | 19931 | 187758 | 162098 | 0.05452 | 0.02445 | 0.06013 | 0.05077 | 0.01021 | 0.01124 |
| MONOAB | rs10874656 | rs4970706 | A | C | 1 | GFI1 | Female only | True | 0.2116 | 0.2078 | 0.2063 | 0.2063 | 25647 | 19931 | 187758 | 162098 | 0.05407 | 0.02475 | 0.05992 | 0.05067 | 0.0102 | 0.01123 |
| MONOAB | rs10874656 | rs10782922 | G | A | 1 | GFI1 | Female only | True | 0.2121 | 0.208 | 0.2068 | 0.2069 | 25647 | 19931 | 187758 | 162098 | 0.05426 | 0.02456 | 0.05945 | 0.05038 | 0.01019 | 0.01122 |
| MONOAB | rs10874656 | rs1886683 | T | G | 1 | GFI1 | Female only | True | 0.2121 | 0.208 | 0.208 | 0.2081 | 25647 | 19931 | 187758 | 162098 | 0.05427 | 0.02457 | 0.05899 | 0.05016 | 0.01019 | 0.01122 |
| MONOAB | rs10874656 | rs6690764 | G | A | 1 | GFI1 | Female only | True | 0.2121 | 0.208 | 0.208 | 0.2081 | 25647 | 19931 | 187758 | 162098 | 0.05427 | 0.02457 | 0.05899 | 0.05016 | 0.01019 | 0.01122 |
| MONOAB | rs10874656 | rs1886682 | A | G | 1 | GFI1 | Female only | True | 0.2121 | 0.208 | 0.208 | 0.2081 | 25647 | 19931 | 187758 | 162098 | 0.05427 | 0.02457 | 0.05898 | 0.05016 | 0.01019 | 0.01122 |
| MONOAB | rs10874656 | rs6603979 | A | G | 1 | GFI1 | Female only | True | 0.2117 | 0.2078 | 0.2075 | 0.2076 | 25647 | 19931 | 187758 | 162098 | 0.05395 | 0.02476 | 0.05945 | 0.05045 | 0.0102 | 0.01123 |
| MONOAB | rs10874656 | rs6603980 | C | T | 1 | GFI1 | Female only | True | 0.212 | 0.2081 | 0.2077 | 0.2077 | 25647 | 19931 | 187758 | 162098 | 0.05376 | 0.02426 | 0.0594 | 0.05008 | 0.0102 | 0.01122 |
| MONOAB | rs10874656 | rs6686702 | C | T | 1 | GFI1 | Female only | True | 0.2121 | 0.2082 | 0.2076 | 0.2077 | 25647 | 19931 | 187758 | 162098 | 0.05381 | 0.02497 | 0.05931 | 0.0504 | 0.01019 | 0.01122 |

| trait | lead_snp | ID | ALLELE1 | ALLELE0 | CHR | Gene_Symbol | independent_loci_in_sex_P1e2 | in_embl_any_or_gene | A1FREQ_VUMC_female | A1FREQ_VUMC_male | A1FREQ_UKB_female | A1FREQ_UKB_male | N_VUMC_female | N_VUMC_male | N_UKB_female | N_UKB_male | BETA_VUMC_female | BETA_VUMC_male | BETA_UKB_female | BETA_UKB_male | SE_VUMC_female | SE_VUMC_male |
|---|---|---|---|---|---|---|---|---|---|---|---|---|---|---|---|---|---|---|---|---|---|---|
| MONOAB | rs10874656 | rs4375281 | A | G | 1 | GFI1 | Female only | True | 0.2121 | 0.2082 | 0.2076 | 0.2077 | 25647 | 19931 | 187758 | 162098 | 0.0538 | 0.02498 | 0.05931 | 0.05039 | 0.01019 | 0.01122 |
| MONOAB | rs10874656 | rs4970709 | C | A | 1 | GFI1 | Female only | True | 0.2121 | 0.2082 | 0.2076 | 0.2077 | 25647 | 19931 | 187758 | 162098 | 0.05381 | 0.02497 | 0.05931 | 0.05039 | 0.01019 | 0.01122 |
| MONOAB | rs10874656 | rs4970710 | A | C | 1 | GFI1 | Female only | True | 0.2121 | 0.2082 | 0.2076 | 0.2077 | 25647 | 19931 | 187758 | 162098 | 0.05381 | 0.02497 | 0.0593 | 0.05039 | 0.01019 | 0.01122 |
| MONOAB | rs10874656 | rs1408916 | G | A | 1 | GFI1 | Female only | True | 0.2121 | 0.2082 | 0.2076 | 0.2077 | 25647 | 19931 | 187758 | 162098 | 0.05381 | 0.02497 | 0.0593 | 0.05038 | 0.01019 | 0.01122 |
| MONOAB | rs10874656 | rs2391160 | T | C | 1 | GFI1 | Female only | True | 0.2121 | 0.2082 | 0.2076 | 0.2077 | 25647 | 19931 | 187758 | 162098 | 0.05381 | 0.02497 | 0.0593 | 0.05038 | 0.01019 | 0.01122 |
| MONOAB | rs10874656 | rs4568842 | C | T | 1 | GFI1 | Female only | True | 0.2121 | 0.2082 | 0.2076 | 0.2077 | 25647 | 19931 | 187758 | 162098 | 0.05381 | 0.02497 | 0.0593 | 0.05038 | 0.01019 | 0.01122 |
| MONOAB | rs10874656 | rs4970712 | A | C | 1 | GFI1 | Female only | True | 0.2121 | 0.2082 | 0.2076 | 0.2077 | 25647 | 19931 | 187758 | 162098 | 0.05381 | 0.02497 | 0.0593 | 0.05038 | 0.01019 | 0.01122 |
| MONOAB | rs10874656 | rs6603981 | C | T | 1 | GFI1 | Female only | True | 0.2121 | 0.2082 | 0.2076 | 0.2077 | 25647 | 19931 | 187758 | 162098 | 0.05381 | 0.02497 | 0.0593 | 0.05038 | 0.01019 | 0.01122 |
| MONOAB | rs10874656 | rs6603982 | G | A | 1 | GFI1 | Female only | True | 0.2121 | 0.2082 | 0.2076 | 0.2077 | 25647 | 19931 | 187758 | 162098 | 0.05381 | 0.02497 | 0.0593 | 0.05038 | 0.01019 | 0.01122 |
| MONOAB | rs10874656 | rs6696173 | C | A | 1 | GFI1 | Female only | True | 0.2121 | 0.2082 | 0.2076 | 0.2077 | 25647 | 19931 | 187758 | 162098 | 0.05381 | 0.02497 | 0.0593 | 0.05038 | 0.01019 | 0.01122 |
| MONOAB | rs10874656 | rs4970701 | A | G | 1 | GFI1 | Female only | True | 0.2121 | 0.2082 | 0.2076 | 0.2077 | 25647 | 19931 | 187758 | 162098 | 0.05385 | 0.02498 | 0.0593 | 0.05038 | 0.01019 | 0.01122 |
| MONOAB | rs10874656 | rs10782935 | T | C | 1 | GFI1 | Female only | True | 0.212 | 0.2081 | 0.2076 | 0.2077 | 25647 | 19931 | 187758 | 162098 | 0.0537 | 0.02513 | 0.0593 | 0.05038 | 0.01018 | 0.01121 |
| MONOAB | rs10874656 | 1:93001143 | A | C | 1 | GFI1 | Female only | True | 0.212 | 0.2081 | | | 25647 | 19931 | | | 0.0537 | 0.02513 | | | 0.01018 | 0.01121 |
| MONOAB | rs10874656 | rs2027061 | C | T | 1 | GFI1 | Female only | True | 0.212 | 0.2081 | 0.2076 | 0.2077 | 25647 | 19931 | 187758 | 162098 | 0.0537 | 0.02513 | 0.0593 | 0.05038 | 0.01018 | 0.01121 |
| MONOAB | rs10874656 | rs989653 | A | G | 1 | GFI1 | Female only | True | 0.212 | 0.2081 | 0.2076 | 0.2077 | 25647 | 19931 | 187758 | 162098 | 0.0537 | 0.02513 | 0.0593 | 0.05037 | 0.01018 | 0.01121 |
| MONOAB | rs10874656 | rs2151577 | A | C | 1 | GFI1 | Female only | True | 0.212 | 0.2081 | 0.2076 | 0.2077 | 25647 | 19931 | 187758 | 162098 | 0.0537 | 0.02513 | 0.0593 | 0.05038 | 0.01018 | 0.01121 |
| MONOAB | rs10874656 | rs6658734 | T | C | 1 | GFI1 | Female only | True | 0.2117 | 0.2078 | 0.2075 | 0.2076 | 25647 | 19931 | 187758 | 162098 | 0.05411 | 0.02481 | 0.05951 | 0.05048 | 0.01019 | 0.01122 |
| MONOAB | rs10874656 | rs10735778 | T | C | 1 | GFI1 | Female only | True | 0.212 | 0.2081 | 0.2076 | 0.2077 | 25647 | 19931 | 187758 | 162098 | 0.0537 | 0.02513 | 0.0593 | 0.05038 | 0.01018 | 0.01121 |
| MONOAB | rs10874656 | rs1570837 | T | C | 1 | GFI1 | Female only | True | 0.212 | 0.2081 | 0.2076 | 0.2077 | 25647 | 19931 | 187758 | 162098 | 0.0537 | 0.02513 | 0.0593 | 0.05038 | 0.01018 | 0.01121 |
| MONOAB | rs10874656 | rs10735779 | G | T | 1 | GFI1 | Female only | True | 0.212 | 0.2081 | 0.2076 | 0.2077 | 25647 | 19931 | 187758 | 162098 | 0.05371 | 0.02514 | 0.05929 | 0.05036 | 0.01018 | 0.01121 |
| MONOAB | rs10874656 | rs7515577 | C | A | 1 | GFI1 | Female only | True | 0.207 | 0.203 | 0.2076 | 0.2077 | 25647 | 19931 | 187758 | 162098 | 0.05345 | 0.02508 | 0.0593 | 0.05037 | 0.01012 | 0.01114 |
| MONOAB | rs10874656 | rs6661134 | T | C | 1 | GFI1 | Female only | True | 0.212 | 0.208 | 0.2076 | 0.2077 | 25647 | 19931 | 187758 | 162098 | 0.05369 | 0.0251 | 0.0593 | 0.05037 | 0.01018 | 0.01121 |
| MONOAB | rs10874656 | rs6603984 | A | G | 1 | GFI1 | Female only | True | 0.2117 | 0.2077 | 0.2075 | 0.2076 | 25647 | 19931 | 187758 | 162098 | 0.05404 | 0.02495 | 0.05951 | 0.05047 | 0.01019 | 0.01122 |
| MONOAB | rs10874656 | rs11164773 | T | C | 1 | GFI1 | Female only | True | 0.212 | 0.208 | 0.2076 | 0.2077 | 25647 | 19931 | 187758 | 162098 | 0.05369 | 0.0251 | 0.0593 | 0.05038 | 0.01018 | 0.01121 |
| MONOAB | rs10874656 | rs78845176 | G | A | 1 | GFI1 | Female only | True | 0.2117 | 0.2078 | | | 25647 | 19931 | | | 0.0541 | 0.02479 | | | 0.01019 | 0.01122 |
| MONOAB | rs10874656 | rs12753280 | T | C | 1 | GFI1 | Female only | True | 0.212 | 0.208 | 0.2076 | 0.2077 | 25647 | 19931 | 187758 | 162098 | 0.05369 | 0.0251 | 0.0593 | 0.05039 | 0.01018 | 0.01121 |
| MONOAB | rs10874656 | rs6603987 | T | C | 1 | GFI1 | Female only | True | 0.2117 | 0.2078 | 0.2076 | 0.2077 | 25647 | 19931 | 187758 | 162098 | 0.05336 | 0.02493 | 0.0593 | 0.0504 | 0.01018 | 0.01121 |

| trait | lead_snp | ID | ALLELE1 | ALLELE0 | CHR | Gene_Symbol | independent_loci_in_sex_P1e2 | in_embl_any_or_gene | A1FREQ_VUMC_female | A1FREQ_VUMC_male | A1FREQ_UKB_female | A1FREQ_UKB_male | N_VUMC_female | N_VUMC_male | N_UKB_female | N_UKB_male | BETA_VUMC_female | BETA_VUMC_male | BETA_UKB_female | BETA_UKB_male | SE_VUMC_female | SE_VUMC_male |
|---|---|---|---|---|---|---|---|---|---|---|---|---|---|---|---|---|---|---|---|---|---|---|
| MONOAB | rs10874656 | rs10874712 | T | C | 1 | GFI1 | Female only | True | 0.2117 | 0.2078 | 0.2076 | 0.2077 | 25647 | 19931 | 187758 | 162098 | 0.05336 | 0.02493 | 0.0593 | 0.05039 | 0.01018 | 0.01121 |
| MONOAB | rs10874656 | rs6603988 | A | G | 1 | GFI1 | Female only | True | 0.2117 | 0.2078 | 0.2076 | 0.2077 | 25647 | 19931 | 187758 | 162098 | 0.05336 | 0.02493 | 0.0593 | 0.0504 | 0.01018 | 0.01121 |
| MONOAB | rs10874656 | rs4847377 | A | C | 1 | GFI1 | Female only | True | 0.2117 | 0.2078 | 0.2076 | 0.2077 | 25647 | 19931 | 187758 | 162098 | 0.05336 | 0.02493 | 0.0593 | 0.0504 | 0.01018 | 0.01121 |
| MONOAB | rs10874656 | rs6422512 | C | T | 1 | GFI1 | Female only | True | 0.2117 | 0.2078 | 0.2076 | 0.2077 | 25647 | 19931 | 187758 | 162098 | 0.05336 | 0.02493 | 0.0593 | 0.0504 | 0.01018 | 0.01121 |
| MONOAB | rs10874656 | 1:93027110 | G | A | 1 | GFI1 | Female only | True | 0.2135 | 0.2102 | | | 25647 | 19931 | | | 0.04992 | 0.02406 | | | 0.01006 | 0.01106 |
| MONOAB | rs10874656 | rs10465753 | G | A | 1 | GFI1 | Female only | True | 0.2117 | 0.2078 | 0.2076 | 0.2077 | 25647 | 19931 | 187758 | 162098 | 0.05336 | 0.02493 | 0.05927 | 0.05038 | 0.01018 | 0.01121 |
| MONOAB | rs10874656 | rs10465754 | G | A | 1 | GFI1 | Female only | True | 0.2117 | 0.2078 | 0.2076 | 0.2077 | 25647 | 19931 | 187758 | 162098 | 0.05336 | 0.02493 | 0.05927 | 0.05038 | 0.01018 | 0.01121 |
| MONOAB | rs10874656 | rs6656432 | A | G | 1 | GFI1 | Female only | True | 0.2117 | 0.2078 | 0.2077 | 0.2078 | 25647 | 19931 | 187758 | 162098 | 0.05336 | 0.02493 | 0.05921 | 0.05035 | 0.01018 | 0.01121 |
| MONOAB | rs10874656 | rs10465756 | T | G | 1 | GFI1 | Female only | True | 0.2117 | 0.2078 | 0.2077 | 0.2078 | 25647 | 19931 | 187758 | 162098 | 0.05336 | 0.02493 | 0.05918 | 0.05032 | 0.01018 | 0.01121 |
| MONOAB | rs10874656 | rs6603990 | T | C | 1 | GFI1 | Female only | True | 0.2117 | 0.2078 | | | 25647 | 19931 | | | 0.05351 | 0.02502 | | | 0.01019 | 0.01121 |
| MONOAB | rs10874656 | rs10747444 | T | C | 1 | GFI1 | Female only | True | 0.2117 | 0.2078 | | | 25647 | 19931 | | | 0.0535 | 0.02502 | | | 0.01019 | 0.01121 |
| MONOAB | rs10874656 | rs6422513 | G | A | 1 | GFI1 | Female only | True | 0.2117 | 0.2079 | | | 25647 | 19931 | | | 0.05322 | 0.0251 | | | 0.01019 | 0.01121 |
| MONOAB | rs10874656 | rs6422514 | T | C | 1 | GFI1 | Female only | True | 0.2117 | 0.2079 | | | 25647 | 19931 | | | 0.05322 | 0.02506 | | | 0.01019 | 0.01121 |
| MONOAB | rs10874656 | rs6603991 | T | C | 1 | GFI1 | Female only | True | 0.2117 | 0.2079 | | | 25647 | 19931 | | | 0.05322 | 0.02506 | | | 0.01019 | 0.01121 |
| MONOAB | rs10874656 | rs1556562 | G | T | 1 | GFI1 | Female only | True | 0.2117 | 0.2079 | | | 25647 | 19931 | | | 0.05322 | 0.02508 | | | 0.01019 | 0.01121 |
| MONOAB | rs10874656 | rs1556563 | G | A | 1 | GFI1 | Female only | True | 0.2117 | 0.2079 | | | 25647 | 19931 | | | 0.05321 | 0.02509 | | | 0.01019 | 0.01121 |
| MONOAB | rs10874656 | rs6692187 | C | T | 1 | GFI1 | Female only | True | 0.2117 | 0.2079 | | | 25647 | 19931 | | | 0.0534 | 0.02507 | | | 0.01019 | 0.01121 |
| MONOAB | rs11915803 | rs62250114 | A | G | 3 | CCDC13 | Female only | True | 0.2749 | 0.2801 | 0.276 | 0.277 | 25647 | 19931 | 187758 | 162098 | 0.05298 | 0.01706 | 0.034 | 0.04161 | 0.009286 | 0.0101 |
| MONOAB | rs11915803 | rs55884426 | T | C | 3 | CCDC13 | Female only | True | 0.2747 | 0.28 | 0.2753 | 0.2764 | 25647 | 19931 | 187758 | 162098 | 0.05295 | 0.01698 | 0.03445 | 0.04176 | 0.009269 | 0.01009 |
| MONOAB | rs11915803 | rs11713172 | T | C | 3 | CCDC13 | Female only | True | 0.2747 | 0.2799 | 0.2753 | 0.2764 | 25647 | 19931 | 187758 | 162098 | 0.05294 | 0.01713 | 0.03442 | 0.04179 | 0.009269 | 0.01009 |
| MONOAB | rs11915803 | rs17238314 | T | C | 3 | CCDC13 | Female only | True | 0.2747 | 0.2799 | 0.2753 | 0.2764 | 25647 | 19931 | 187758 | 162098 | 0.05303 | 0.01686 | 0.03446 | 0.04177 | 0.009266 | 0.01008 |
| MONOAB | rs11915803 | rs59713146 | C | T | 3 | CCDC13 | Female only | True | 0.2747 | 0.2799 | 0.2753 | 0.2764 | 25647 | 19931 | 187758 | 162098 | 0.05303 | 0.01686 | 0.03444 | 0.04177 | 0.009267 | 0.01008 |
| MONOAB | rs11915803 | rs59478814 | G | T | 3 | CCDC13 | Female only | True | 0.2747 | 0.2799 | 0.2753 | 0.2764 | 25647 | 19931 | 187758 | 162098 | 0.05271 | 0.01674 | 0.03446 | 0.04175 | 0.00926 | 0.01008 |
| MONOAB | rs11915803 | rs10865921 | T | C | 3 | CCDC13 | Female only | True | 0.2777 | 0.2833 | 0.2774 | 0.2783 | 25647 | 19931 | 187758 | 162098 | 0.05242 | 0.01631 | 0.03319 | 0.04124 | 0.00922 | 0.01004 |
| MONOAB | rs11915803 | rs11129969 | T | C | 3 | CCDC13 | Female only | True | 0.2742 | 0.2796 | 0.275 | 0.2761 | 25647 | 19931 | 187758 | 162098 | 0.05271 | 0.01617 | 0.03425 | 0.04184 | 0.009262 | 0.01008 |
| MONOAB | rs11915803 | rs11129970 | T | C | 3 | CCDC13 | Female only | True | 0.2742 | 0.2796 | 0.275 | 0.276 | 25647 | 19931 | 187758 | 162098 | 0.05272 | 0.01617 | 0.03419 | 0.04185 | 0.009262 | 0.01008 |
| MONOAB | rs11915803 | rs17074778 | A | G | 3 | CCDC13 | Female only | True | 0.2777 | 0.2833 | 0.2774 | 0.2783 | 25647 | 19931 | 187758 | 162098 | 0.05243 | 0.01631 | 0.0332 | 0.04126 | 0.00922 | 0.01004 |
| MONOAB | rs11915803 | rs62246603 | T | G | 3 | CCDC13 | Female only | True | 0.2743 | 0.2793 | 0.2751 | 0.2762 | 25647 | 19931 | 187758 | 162098 | 0.05296 | 0.01662 | 0.03382 | 0.04199 | 0.009261 | 0.01007 |
| MONOAB | rs11915803 | rs11709500 | A | G | 3 | CCDC13 | Female only | True | 0.2743 | 0.2793 | 0.2751 | 0.2762 | 25647 | 19931 | 187758 | 162098 | 0.05296 | 0.01662 | 0.0338 | 0.04199 | 0.009261 | 0.01007 |

| trait | lead_snp | ID | ALLELE1 | ALLELE0 | CHR | Gene_Symbol | independent_loci_in_sex_P1e2 | in_embl_any_or_gene | A1FREQ_VUMC_female | A1FREQ_VUMC_male | A1FREQ_UKB_female | A1FREQ_UKB_male | N_VUMC_female | N_VUMC_male | N_UKB_female | N_UKB_male | BETA_VUMC_female | BETA_VUMC_male | BETA_UKB_female | BETA_UKB_male | SE_VUMC_female | SE_VUMC_male |
|---|---|---|---|---|---|---|---|---|---|---|---|---|---|---|---|---|---|---|---|---|---|---|
| MONOAB | rs11915803 | rs62246606 | C | T | 3 | CCDC13 | Female only | True | 0.2747 | 0.2798 | 0.2751 | 0.2762 | 25647 | 19931 | 187758 | 162098 | 0.05211 | 0.0171 | 0.03399 | 0.04227 | 0.009257 | 0.01006 |
| MONOAB | rs11915803 | rs11709054 | A | G | 3 | CCDC13 | Female only | True | 0.2742 | 0.2793 | 0.2749 | 0.276 | 25647 | 19931 | 187758 | 162098 | 0.05223 | 0.01726 | 0.03404 | 0.04219 | 0.009261 | 0.01007 |
| MONOAB | rs11915803 | rs34110933 | A | G | 3 | CCDC13 | Female only | True | 0.2742 | 0.2793 | 0.2749 | 0.276 | 25647 | 19931 | 187758 | 162098 | 0.05223 | 0.01727 | 0.03405 | 0.0422 | 0.009261 | 0.01007 |
| MONOAB | rs11915803 | rs6442057 | A | G | 3 | CCDC13 | Female only | True | 0.276 | 0.2814 | 0.2773 | 0.2786 | 25647 | 19931 | 187758 | 162098 | 0.05229 | 0.01923 | 0.03461 | 0.04336 | 0.009254 | 0.01007 |
| MONOAB | rs11915803 | rs17238798 | A | G | 3 | CCDC13 | Female only | True | 0.2764 | 0.2822 | 0.2779 | 0.2794 | 25647 | 19931 | 187758 | 162098 | 0.05447 | 0.02033 | 0.03527 | 0.04429 | 0.009248 | 0.01005 |
| MONOAB | rs11915803 | rs11915803 | C | T | 3 | CCDC13 | Female only | True | 0.277 | 0.2829 | 0.2783 | 0.2799 | 25647 | 19931 | 187758 | 162098 | 0.05447 | 0.02059 | 0.03548 | 0.04403 | 0.00924 | 0.01005 |
| MONOAB | rs11915803 | rs62246629 | C | T | 3 | CCDC13 | Female only | True | 0.2947 | 0.2989 | 0.2931 | 0.2948 | 25647 | 19931 | 187758 | 162098 | 0.05219 | 0.02047 | 0.03628 | 0.04428 | 0.009046 | 0.009888 |
| MONOAB | rs11915803 | rs4682856 | A | C | 3 | CCDC13 | Female only | True | 0.274 | 0.2793 | 0.2723 | 0.2738 | 25647 | 19931 | 187758 | 162098 | 0.05336 | 0.02228 | 0.03669 | 0.04535 | 0.009223 | 0.01006 |
| MONOAB | rs834580 | rs2081811 | G | A | 3 | KRBOX1 | Female only | True | 0.2479 | 0.2475 | 0.2513 | 0.2501 | 25647 | 19931 | 187758 | 162098 | -0.0593 | -0.02119 | -0.04373 | -0.0496 | 0.009572 | 0.01045 |
| MONOAB | rs834580 | rs704901 | A | G | 3 | KRBOX1 | Female only | True | 0.2601 | 0.261 | 0.2643 | 0.263 | 25647 | 19931 | 187758 | 162098 | -0.05843 | -0.01939 | -0.04173 | -0.04718 | 0.009401 | 0.01023 |
| MONOAB | rs834580 | rs834580 | A | G | 3 | KRBOX1 | Female only | True | 0.2603 | 0.2611 | 0.2644 | 0.2631 | 25647 | 19931 | 187758 | 162098 | -0.05847 | -0.01998 | -0.04168 | -0.04715 | 0.009384 | 0.01022 |
| MONOAB | rs834580 | rs704891 | T | C | 3 | KRBOX1 | Female only | True | 0.2596 | 0.2601 | 0.2642 | 0.2628 | 25647 | 19931 | 187758 | 162098 | -0.05839 | -0.01965 | -0.04127 | -0.04731 | 0.009393 | 0.01024 |
| MONOAB | rs834580 | rs704892 | T | C | 3 | KRBOX1 | Female only | True | 0.1718 | 0.1704 | 0.1788 | 0.1775 | 25647 | 19931 | 187758 | 162098 | -0.05688 | -0.02479 | -0.03338 | -0.04164 | 0.01087 | 0.01193 |
| MONOAB | rs834580 | rs704893 | C | T | 3 | KRBOX1 | Female only | True | 0.2596 | 0.2601 | 0.2641 | 0.2628 | 25647 | 19931 | 187758 | 162098 | -0.05839 | -0.01962 | -0.04123 | -0.0473 | 0.009395 | 0.01024 |

| trait | lead_snp | ID | ALLELE1 | ALLELE0 | CHR | Gene_Symbol | independent_loci_in_sex_P1e2 | in_embl_any_or_gene | A1FREQ_VUMC_female | A1FREQ_VUMC_male | A1FREQ_UKB_female | A1FREQ_UKB_male | N_VUMC_female | N_VUMC_male | N_UKB_female | N_UKB_male | BETA_VUMC_female | BETA_VUMC_male | BETA_UKB_female | BETA_UKB_male | SE_VUMC_female | SE_VUMC_male |
|---|---|---|---|---|---|---|---|---|---|---|---|---|---|---|---|---|---|---|---|---|---|---|
| MONOAB | rs834580 | rs2559650 | T | C | 3 | KRBOX1 | Female only | True | 0.2597 | 0.2603 | 0.2642 | 0.2629 | 25647 | 19931 | 187758 | 162098 | -0.05849 | -0.02001 | -0.0412 | -0.04711 | 0.009396 | 0.01025 |
| MONOAB | rs17693963 | rs58616630 | G | A | 6 | RP1-97D16.1 | Male only | False | 0.09114 | 0.0923 | 0.1166 | 0.1162 | 25647 | 19931 | 187758 | 162098 | -0.02641 | -0.08623 | -0.05444 | -0.05463 | 0.01431 | 0.01569 |
| MONOAB | rs17693963 | rs34573979 | T | C | 6 | RP1-97D16.1 | Male only | False | 0.09114 | 0.09235 | 0.1169 | 0.1165 | 25647 | 19931 | 187758 | 162098 | -0.02641 | -0.08631 | -0.05439 | -0.05417 | 0.01431 | 0.01569 |
| MONOAB | rs17693963 | rs6904596 | A | G | 6 | RP1-97D16.1 | Male only | False | 0.1024 | 0.1037 | 0.1285 | 0.1278 | 25647 | 19931 | 187758 | 162098 | -0.02931 | -0.081 | -0.05254 | -0.04916 | 0.01357 | 0.01484 |
| MONOAB | rs17693963 | rs13195636 | C | A | 6 | RP1-97D16.1 | Male only | False | 0.09224 | 0.09353 | 0.1186 | 0.1184 | 25647 | 19931 | 187758 | 162098 | -0.02826 | -0.08772 | -0.05589 | -0.0534 | 0.01424 | 0.01559 |
| MONOAB | rs17693963 | rs35848276 | T | C | 6 | RP1-97D16.1 | Male only | False | 0.08771 | 0.08941 | 0.115 | 0.115 | 25647 | 19931 | 187758 | 162098 | -0.02252 | -0.08849 | -0.05791 | -0.05511 | 0.01458 | 0.01589 |
| MONOAB | rs17693963 | rs34965299 | G | A | 6 | RP1-97D16.1 | Male only | False | 0.09197 | 0.09328 | 0.1186 | 0.1185 | 25647 | 19931 | 187758 | 162098 | -0.02859 | -0.08721 | -0.05589 | -0.05336 | 0.01425 | 0.01559 |
| MONOAB | rs17693963 | rs66785117 | G | T | 6 | RP1-97D16.1 | Male only | False | 0.09197 | 0.09328 | 0.1186 | 0.1185 | 25647 | 19931 | 187758 | 162098 | -0.02859 | -0.08721 | -0.05589 | -0.05336 | 0.01425 | 0.01559 |
| MONOAB | rs17693963 | rs140365013 | A | G | 6 | RP1-97D16.1 | Male only | False | 0.09197 | 0.09328 | 0.1186 | 0.1185 | 25647 | 19931 | 187758 | 162098 | -0.02859 | -0.08721 | -0.05589 | -0.05337 | 0.01425 | 0.01559 |
| MONOAB | rs17693963 | rs10484399 | G | A | 6 | RP1-97D16.1 | Male only | False | 0.09208 | 0.09337 | 0.1187 | 0.1185 | 25647 | 19931 | 187758 | 162098 | -0.02896 | -0.08734 | -0.0558 | -0.05333 | 0.01423 | 0.01557 |
| MONOAB | rs17693963 | rs34105070 | T | C | 6 | RP1-97D16.1 | Male only | False | 0.09196 | 0.09317 | 0.1186 | 0.1184 | 25647 | 19931 | 187758 | 162098 | -0.02905 | -0.08835 | -0.05583 | -0.05382 | 0.01425 | 0.0156 |
| MONOAB | rs17693963 | rs13202775 | C | T | 6 | RP1-97D16.1 | Male only | False | 0.09197 | 0.09318 | 0.1185 | 0.1183 | 25647 | 19931 | 187758 | 162098 | -0.029 | -0.08836 | -0.05574 | -0.0541 | 0.01425 | 0.0156 |
| MONOAB | rs17693963 | 6:27586220 | T | C | 6 | RP1-97D16.1 | Male only | False | 0.09209 | 0.09326 | | | 25647 | 19931 | | | -0.02948 | -0.08794 | | | 0.01425 | 0.0156 |

| trait | lead_snp | ID | ALLELE1 | ALLELE0 | CHR | Gene_Symbol | independent_loci_in_sex_P1e2 | in_embl_any_or_gene | A1FREQ_VUMC_female | A1FREQ_VUMC_male | A1FREQ_UKB_female | A1FREQ_UKB_male | N_VUMC_female | N_VUMC_male | N_UKB_female | N_UKB_male | BETA_VUMC_female | BETA_VUMC_male | BETA_UKB_female | BETA_UKB_male | SE_VUMC_female | SE_VUMC_male |
|---|---|---|---|---|---|---|---|---|---|---|---|---|---|---|---|---|---|---|---|---|---|---|
| MONOAB | rs17693963 | rs35715914 | T | C | 6 | RP1-97D16.1 | Male only | False | 0.0877 | 0.08925 | 0.1138 | 0.1138 | 25647 | 19931 | 187758 | 162098 | -0.02346 | -0.08886 | -0.05883 | -0.05664 | 0.0146 | 0.01591 |
| MONOAB | rs17693963 | rs13212093 | T | C | 6 | RP1-97D16.1 | Male only | False | 0.09211 | 0.09329 | 0.1186 | 0.1185 | 25647 | 19931 | 187758 | 162098 | -0.02951 | -0.08784 | -0.05574 | -0.05415 | 0.01425 | 0.01559 |
| MONOAB | rs17693963 | rs34038546 | A | C | 6 | RP1-97D16.1 | Male only | False | 0.09212 | 0.09326 | 0.1186 | 0.1185 | 25647 | 19931 | 187758 | 162098 | -0.02968 | -0.08725 | -0.05573 | -0.05415 | 0.01425 | 0.0156 |
| MONOAB | rs17693963 | rs144969912 | T | C | 6 | RP1-97D16.1 | Male only | False | 0.09175 | 0.09287 | 0.117 | 0.117 | 25647 | 19931 | 187758 | 162098 | -0.0301 | -0.08751 | -0.05591 | -0.0548 | 0.0143 | 0.01566 |
| MONOAB | rs17693963 | rs141342723 | C | T | 6 | RP1-97D16.1 | Male only | False | 0.09213 | 0.09326 | 0.1186 | 0.1185 | 25647 | 19931 | 187758 | 162098 | -0.02971 | -0.08711 | -0.05574 | -0.05419 | 0.01425 | 0.0156 |
| MONOAB | rs17693963 | rs34543938 | G | A | 6 | RP1-97D16.1 | Male only | False | 0.09215 | 0.09327 | 0.1186 | 0.1185 | 25647 | 19931 | 187758 | 162098 | -0.02992 | -0.08712 | -0.05573 | -0.05424 | 0.01424 | 0.0156 |
| MONOAB | rs17693963 | rs56405707 | A | G | 6 | RP1-97D16.1 | Male only | False | 0.09214 | 0.09326 | 0.1186 | 0.1185 | 25647 | 19931 | 187758 | 162098 | -0.02985 | -0.087 | -0.05574 | -0.05424 | 0.01424 | 0.0156 |
| MONOAB | rs17693963 | rs13210634 | G | A | 6 | RP1-97D16.1 | Male only | False | 0.09217 | 0.09328 | 0.1186 | 0.1185 | 25647 | 19931 | 187758 | 162098 | -0.02985 | -0.08697 | -0.05584 | -0.0542 | 0.01424 | 0.0156 |
| MONOAB | rs17693963 | rs13215275 | T | G | 6 | RP1-97D16.1 | Male only | False | 0.09217 | 0.09328 | 0.1186 | 0.1185 | 25647 | 19931 | 187758 | 162098 | -0.02985 | -0.08697 | -0.05583 | -0.05419 | 0.01424 | 0.0156 |
| MONOAB | rs17693963 | rs13217620 | T | C | 6 | RP1-97D16.1 | Male only | False | 0.08503 | 0.08624 | 0.1115 | 0.1115 | 25647 | 19931 | 187758 | 162098 | -0.01662 | -0.09156 | -0.05863 | -0.05784 | 0.01492 | 0.0163 |
| MONOAB | rs17693963 | rs17749927 | G | A | 6 | RP1-97D16.1 | Male only | False | 0.09221 | 0.09324 | 0.1187 | 0.1186 | 25647 | 19931 | 187758 | 162098 | -0.02997 | -0.08645 | -0.05593 | -0.05425 | 0.01423 | 0.0156 |
| MONOAB | rs17693963 | rs13192965 | A | G | 6 | RP1-97D16.1 | Male only | False | 0.09224 | 0.09327 | 0.1187 | 0.1186 | 25647 | 19931 | 187758 | 162098 | -0.03003 | -0.08645 | -0.05599 | -0.054 | 0.01423 | 0.01559 |
| MONOAB | rs17693963 | rs34409918 | G | A | 6 | RP1-97D16.1 | Male only | False | 0.09214 | 0.09321 | 0.1187 | 0.1186 | 25647 | 19931 | 187758 | 162098 | -0.02948 | -0.0861 | -0.05624 | -0.0542 | 0.01423 | 0.01559 |
| MONOAB | rs17693963 | rs34432857 | A | G | 6 | RP1-97D16.1 | Male only | False | 0.09216 | 0.09324 | 0.1187 | 0.1186 | 25647 | 19931 | 187758 | 162098 | -0.0294 | -0.08556 | -0.05624 | -0.0542 | 0.01423 | 0.01559 |
| MONOAB | rs17693963 | rs34064842 | T | G | 6 | RP1-97D16.1 | Male only | False | 0.09217 | 0.09324 | 0.1187 | 0.1186 | 25647 | 19931 | 187758 | 162098 | -0.02943 | -0.08553 | -0.05636 | -0.05416 | 0.01422 | 0.01559 |
| MONOAB | rs17693963 | rs13212318 | C | A | 6 | RP1-97D16.1 | Male only | False | 0.09215 | 0.09321 | 0.1187 | 0.1186 | 25647 | 19931 | 187758 | 162098 | -0.02951 | -0.08607 | -0.05638 | -0.05416 | 0.01423 | 0.01559 |
| MONOAB | rs17693963 | rs2056923 | C | T | 6 | RP1-97D16.1 | Male only | False | 0.164 | 0.164 | 0.1798 | 0.1803 | 25647 | 19931 | 187758 | 162098 | -0.004817 | -0.05337 | -0.03714 | -0.03235 | 0.01109 | 0.01221 |
| MONOAB | rs17693963 | rs13202295 | T | C | 6 | RP1-97D16.1 | Male only | False | 0.08806 | 0.08938 | 0.1146 | 0.1146 | 25647 | 19931 | 187758 | 162098 | -0.02297 | -0.08711 | -0.05893 | -0.05618 | 0.01455 | 0.0159 |
| MONOAB | rs17693963 | rs13202291 | G | A | 6 | RP1-97D16.1 | Male only | False | 0.09218 | 0.09325 | 0.1188 | 0.1186 | 25647 | 19931 | 187758 | 162098 | -0.02956 | -0.08513 | -0.05649 | -0.05427 | 0.01422 | 0.01559 |
| MONOAB | rs17693963 | rs17750424 | C | T | 6 | RP1-97D16.1 | Male only | False | 0.09223 | 0.0933 | 0.1188 | 0.1186 | 25647 | 19931 | 187758 | 162098 | -0.02959 | -0.08505 | -0.05651 | -0.05427 | 0.01422 | 0.01558 |
| MONOAB | rs17693963 | rs13193542 | T | G | 6 | RP1-97D16.1 | Male only | False | 0.09218 | 0.09325 | 0.1188 | 0.1186 | 25647 | 19931 | 187758 | 162098 | -0.02956 | -0.08513 | -0.05652 | -0.05426 | 0.01422 | 0.01559 |
| MONOAB | rs17693963 | rs13193480 | G | A | 6 | RP1-97D16.1 | Male only | False | 0.09216 | 0.09322 | 0.1188 | 0.1186 | 25647 | 19931 | 187758 | 162098 | -0.02964 | -0.08567 | -0.05652 | -0.05426 | 0.01422 | 0.01559 |
| MONOAB | rs17693963 | rs6940116 | G | A | 6 | RP1-97D16.1 | Male only | False | 0.1637 | 0.1637 | 0.1801 | 0.1808 | 25647 | 19931 | 187758 | 162098 | -0.003947 | -0.05069 | -0.0373 | -0.03154 | 0.01109 | 0.01221 |
| MONOAB | rs17693963 | rs17693963 | C | A | 6 | RP1-97D16.1 | Male only | False | 0.09901 | 0.1003 | 0.125 | 0.1247 | 25647 | 19931 | 187758 | 162098 | -0.02394 | -0.08614 | -0.05533 | -0.05217 | 0.01377 | 0.01502 |
| MONOAB | rs17693963 | rs72847313 | T | C | 6 | RP1-97D16.1 | Male only | False | 0.0922 | 0.09326 | 0.1188 | 0.1187 | 25647 | 19931 | 187758 | 162098 | -0.03008 | -0.08542 | -0.0566 | -0.05442 | 0.01423 | 0.01559 |
| MONOAB | rs17693963 | rs17750747 | C | T | 6 | RP1-97D16.1 | Male only | False | 0.0922 | 0.09326 | 0.1188 | 0.1187 | 25647 | 19931 | 187758 | 162098 | -0.03007 | -0.08542 | -0.05658 | -0.05442 | 0.01423 | 0.01559 |
| MONOAB | rs17693963 | rs13195728 | C | T | 6 | RP1-97D16.1 | Male only | False | 0.09211 | 0.09308 | 0.1188 | 0.1187 | 25647 | 19931 | 187758 | 162098 | -0.0303 | -0.08484 | -0.05677 | -0.05437 | 0.01423 | 0.01561 |
| MONOAB | rs17693963 | rs200482 | A | G | 6 | RP1-97D16.1 | Male only | False | 0.1126 | 0.1128 | 0.1383 | 0.1381 | 25647 | 19931 | 187758 | 162098 | -0.01705 | -0.07317 | -0.04942 | -0.04793 | 0.01304 | 0.01428 |
| MONOAB | rs17693963 | rs200483 | A | G | 6 | RP1-97D16.1 | Male only | False | 0.1122 | 0.1125 | 0.1381 | 0.1378 | 25647 | 19931 | 187758 | 162098 | -0.01655 | -0.07395 | -0.04959 | -0.04801 | 0.01305 | 0.01429 |

| trait | lead_snp | ID | ALLELE1 | ALLELE0 | CHR | Gene_Symbol | independent_loci_in_sex_P1e2 | in_embl_any_or_gene | A1FREQ_VUMC_female | A1FREQ_VUMC_male | A1FREQ_UKB_female | A1FREQ_UKB_male | N_VUMC_female | N_VUMC_male | N_UKB_female | N_UKB_male | BETA_VUMC_female | BETA_VUMC_male | BETA_UKB_female | BETA_UKB_male | SE_VUMC_female | SE_VUMC_male |
|---|---|---|---|---|---|---|---|---|---|---|---|---|---|---|---|---|---|---|---|---|---|---|
| MONOAB | rs17693963 | rs17751184 | T | C | 6 | RP1-97D16.1 | Male only | False | 0.09219 | 0.0932 | 0.1188 | 0.1187 | 25647 | 19931 | 187758 | 162098 | -0.0305 | -0.08477 | -0.05686 | -0.05427 | 0.01422 | 0.01559 |
| MONOAB | rs17693963 | rs200484 | G | A | 6 | RP1-97D16.1 | Male only | False | 0.1121 | 0.1125 | 0.1381 | 0.1379 | 25647 | 19931 | 187758 | 162098 | -0.01721 | -0.07432 | -0.04956 | -0.048 | 0.01306 | 0.01429 |
| MONOAB | rs17693963 | rs370155 | G | T | 6 | RP1-97D16.1 | Male only | False | 0.1126 | 0.1128 | 0.1384 | 0.1381 | 25647 | 19931 | 187758 | 162098 | -0.0169 | -0.0731 | -0.04979 | -0.04819 | 0.01304 | 0.01427 |
| MONOAB | rs17693963 | rs401763 | C | T | 6 | RP1-97D16.1 | Male only | False | 0.1121 | 0.1125 | 0.1317 | 0.1315 | 25647 | 19931 | 187758 | 162098 | -0.01721 | -0.07428 | -0.05144 | -0.04999 | 0.01306 | 0.0143 |
| MONOAB | rs17693963 | rs2747054 | G | A | 6 | RP1-97D16.1 | Male only | False | 0.1121 | 0.1125 | 0.1382 | 0.1379 | 25647 | 19931 | 187758 | 162098 | -0.01705 | -0.07419 | -0.04967 | -0.04812 | 0.01306 | 0.01429 |
| MONOAB | rs17693963 | rs34718920 | T | C | 6 | RP1-97D16.1 | Male only | False | 0.09219 | 0.09322 | 0.1189 | 0.1188 | 25647 | 19931 | 187758 | 162098 | -0.03028 | -0.08477 | -0.05697 | -0.05439 | 0.01422 | 0.01559 |
| MONOAB | rs17693963 | rs200501 | T | C | 6 | RP1-97D16.1 | Male only | False | 0.1124 | 0.1128 | 0.1384 | 0.1381 | 25647 | 19931 | 187758 | 162098 | -0.01692 | -0.07393 | -0.0499 | -0.04776 | 0.01305 | 0.01427 |
| MONOAB | rs17693963 | rs200497 | T | C | 6 | RP1-97D16.1 | Male only | False | 0.1119 | 0.1123 | 0.1381 | 0.1378 | 25647 | 19931 | 187758 | 162098 | -0.0165 | -0.07269 | -0.04973 | -0.0482 | 0.01307 | 0.0143 |
| MONOAB | rs17693963 | rs200490 | T | G | 6 | RP1-97D16.1 | Male only | False | 0.1123 | 0.1126 | 0.1382 | 0.138 | 25647 | 19931 | 187758 | 162098 | -0.01591 | -0.07246 | -0.04961 | -0.04806 | 0.01305 | 0.01429 |
| MONOAB | rs17693963 | 6:27797416 | A | C | 6 | RP1-97D16.1 | Male only | False | 0.09235 | 0.09342 | | | 25647 | 19931 | | | -0.03044 | -0.08491 | | | 0.01423 | 0.01559 |
| MONOAB | rs17693963 | rs200489 | C | T | 6 | RP1-97D16.1 | Male only | False | 0.1118 | 0.1122 | 0.138 | 0.1378 | 25647 | 19931 | 187758 | 162098 | -0.01636 | -0.07362 | -0.04975 | -0.04819 | 0.01308 | 0.01431 |
| MONOAB | rs17693963 | rs34706883 | C | A | 6 | RP1-97D16.1 | Male only | False | 0.09216 | 0.09322 | 0.1189 | 0.1188 | 25647 | 19931 | 187758 | 162098 | -0.02999 | -0.08458 | -0.05694 | -0.05423 | 0.01422 | 0.01559 |
| MONOAB | rs17693963 | rs13212651 | G | A | 6 | RP1-97D16.1 | Male only | False | 0.09218 | 0.09322 | 0.1189 | 0.1188 | 25647 | 19931 | 187758 | 162098 | -0.02984 | -0.08458 | -0.05692 | -0.05451 | 0.01422 | 0.01559 |
| MONOAB | rs17693963 | rs201002 | G | A | 6 | RP1-97D16.1 | Male only | False | 0.1124 | 0.1127 | 0.1386 | 0.1384 | 25647 | 19931 | 187758 | 162098 | -0.01588 | -0.0714 | -0.04993 | -0.04788 | 0.01305 | 0.01428 |
| MONOAB | rs17693963 | rs35819751 | G | A | 6 | RP1-97D16.1 | Male only | False | 0.09211 | 0.09318 | 0.1188 | 0.1188 | 25647 | 19931 | 187758 | 162098 | -0.02989 | -0.08465 | -0.05691 | -0.05454 | 0.01423 | 0.0156 |
| MONOAB | rs17693963 | rs175597 | C | T | 6 | RP1-97D16.1 | Male only | False | 0.1119 | 0.1123 | 0.1382 | 0.1379 | 25647 | 19931 | 187758 | 162098 | -0.0164 | -0.07294 | -0.04964 | -0.04823 | 0.01307 | 0.0143 |
| MONOAB | rs17693963 | rs200996 | A | G | 6 | RP1-97D16.1 | Male only | False | 0.112 | 0.1124 | 0.138 | 0.1378 | 25647 | 19931 | 187758 | 162098 | -0.01584 | -0.07229 | -0.04974 | -0.04838 | 0.01307 | 0.0143 |
| MONOAB | rs17693963 | rs200995 | C | T | 6 | RP1-97D16.1 | Male only | False | 0.1121 | 0.1125 | 0.1382 | 0.138 | 25647 | 19931 | 187758 | 162098 | -0.0153 | -0.07338 | -0.04992 | -0.04853 | 0.01306 | 0.01429 |
| trait | lead_snp | ID | ALLELE1 | ALLELE0 | CHR | Gene_Symbol | independent_loci_in_sex_P1e2 | in_embl_any_or_gene | A1FREQ_VUMC_female | A1FREQ_VUMC_male | A1FREQ_UKB_female | A1FREQ_UKB_male | N_VUMC_female | N_VUMC_male | N_UKB_female | N_UKB_male | BETA_VUMC_female | BETA_VUMC_male | BETA_UKB_female | BETA_UKB_male | SE_VUMC_female | SE_VUMC_male |
| MONOAB | rs17693963 | rs200992 | G | A | 6 | RP1-97D16.1 | Male only | False | 0.1092 | 0.1095 | 0.1291 | 0.1289 | 25647 | 19931 | 187758 | 162098 | -0.01553 | -0.07364 | -0.05213 | -0.0513 | 0.0134 | 0.01465 |
| MONOAB | rs17693963 | rs200991 | A | C | 6 | RP1-97D16.1 | Male only | False | 0.1541 | 0.1544 | 0.1745 | 0.1743 | 25647 | 19931 | 187758 | 162098 | -0.004483 | -0.06001 | -0.04227 | -0.04033 | 0.0114 | 0.01248 |
| MONOAB | rs17693963 | rs13194781 | G | A | 6 | RP1-97D16.1 | Male only | False | 0.09212 | 0.09325 | 0.1188 | 0.1187 | 25647 | 19931 | 187758 | 162098 | -0.03009 | -0.08521 | -0.05701 | -0.055 | 0.01423 | 0.01559 |
| MONOAB | rs17693963 | rs200990 | G | T | 6 | RP1-97D16.1 | Male only | False | 0.1124 | 0.1127 | 0.1382 | 0.138 | 25647 | 19931 | 187758 | 162098 | -0.01552 | -0.07223 | -0.04992 | -0.0486 | 0.01305 | 0.01428 |
| MONOAB | rs17693963 | rs200989 | G | A | 6 | RP1-97D16.1 | Male only | False | 0.1119 | 0.1124 | 0.1381 | 0.1378 | 25647 | 19931 | 187758 | 162098 | -0.01598 | -0.07331 | -0.04986 | -0.04859 | 0.01307 | 0.0143 |
| MONOAB | rs17693963 | rs34194357 | G | A | 6 | RP1-97D16.1 | Male only | False | 0.09207 | 0.0932 | 0.1188 | 0.1187 | 25647 | 19931 | 187758 | 162098 | -0.03009 | -0.08528 | -0.05693 | -0.05488 | 0.01423 | 0.0156 |
| MONOAB | rs17693963 | rs200983 | T | C | 6 | RP1-97D16.1 | Male only | False | 0.1119 | 0.1124 | 0.138 | 0.1377 | 25647 | 19931 | 187758 | 162098 | -0.01613 | -0.07295 | -0.04976 | -0.04863 | 0.01308 | 0.01431 |
| MONOAB | rs17693963 | rs200981 | G | A | 6 | RP1-97D16.1 | Male only | False | 0.1125 | 0.1128 | 0.1381 | 0.1379 | 25647 | 19931 | 187758 | 162098 | -0.0152 | -0.0722 | -0.04983 | -0.0486 | 0.01305 | 0.01428 |
| MONOAB | rs17693963 | rs13199772 | G | A | 6 | RP1-97D16.1 | Male only | False | 0.0923 | 0.09342 | 0.1188 | 0.1187 | 25647 | 19931 | 187758 | 162098 | -0.02968 | -0.08489 | -0.05695 | -0.05519 | 0.01422 | 0.01558 |

| trait | lead_snp | ID | ALLELE1 | ALLELE0 | CHR | Gene_Symbol | independent_loci_in_sex_P1e2 | in_embl_any_or_gene | A1FREQ_VUMC_female | A1FREQ_VUMC_male | A1FREQ_UKB_female | A1FREQ_UKB_male | N_VUMC_female | N_VUMC_male | N_UKB_female | N_UKB_male | BETA_VUMC_female | BETA_VUMC_male | BETA_UKB_female | BETA_UKB_male | SE_VUMC_female | SE_VUMC_male |
|---|---|---|---|---|---|---|---|---|---|---|---|---|---|---|---|---|---|---|---|---|---|---|
| MONOAB | rs17693963 | rs17763089 | A | G | 6 | RP1-97D16.1 | Male only | False | 0.09224 | 0.09336 | 0.1192 | 0.1191 | 25647 | 19931 | 187758 | 162098 | -0.02969 | -0.08505 | -0.05621 | -0.0547 | 0.01423 | 0.01559 |
| MONOAB | rs17693963 | rs200948 | C | T | 6 | RP1-97D16.1 | Male only | False | 0.1121 | 0.1125 | 0.138 | 0.1377 | 25647 | 19931 | 187758 | 162098 | -0.01565 | -0.07331 | -0.04977 | -0.04865 | 0.01307 | 0.0143 |
| MONOAB | rs17693963 | rs200949 | G | A | 6 | RP1-97D16.1 | Male only | False | 0.1207 | 0.1202 | 0.1439 | 0.1439 | 25647 | 19931 | 187758 | 162098 | -0.0114 | -0.06818 | -0.04656 | -0.04612 | 0.01267 | 0.0139 |
| MONOAB | rs17693963 | rs200950 | G | A | 6 | RP1-97D16.1 | Male only | False | 0.1121 | 0.1125 | 0.138 | 0.1377 | 25647 | 19931 | 187758 | 162098 | -0.01563 | -0.07334 | -0.04977 | -0.04865 | 0.01307 | 0.0143 |
| MONOAB | rs17693963 | rs200952 | T | C | 6 | RP1-97D16.1 | Male only | False | 0.1121 | 0.1125 | 0.138 | 0.1377 | 25647 | 19931 | 187758 | 162098 | -0.01563 | -0.07334 | -0.04977 | -0.04865 | 0.01307 | 0.0143 |
| MONOAB | rs17693963 | rs17695758 | C | T | 6 | RP1-97D16.1 | Male only | False | 0.09223 | 0.09335 | 0.1187 | 0.1186 | 25647 | 19931 | 187758 | 162098 | -0.02967 | -0.08519 | -0.05688 | -0.05505 | 0.01423 | 0.01559 |
| MONOAB | rs17693963 | rs200953 | C | T | 6 | RP1-97D16.1 | Male only | False | 0.1126 | 0.1128 | 0.1381 | 0.1378 | 25647 | 19931 | 187758 | 162098 | -0.01518 | -0.07219 | -0.0498 | -0.04859 | 0.01305 | 0.01427 |
| MONOAB | rs17693963 | rs200956 | C | T | 6 | RP1-97D16.1 | Male only | False | 0.1612 | 0.1604 | 0.1795 | 0.1797 | 25647 | 19931 | 187758 | 162098 | -0.001459 | -0.05869 | -0.03997 | -0.0385 | 0.01119 | 0.01228 |
| MONOAB | rs17693963 | rs45509595 | C | T | 6 | RP1-97D16.1 | Male only | False | 0.08796 | 0.08928 | 0.1146 | 0.1146 | 25647 | 19931 | 187758 | 162098 | -0.02249 | -0.08603 | -0.05851 | -0.05635 | 0.01454 | 0.01588 |
| MONOAB | rs17693963 | rs557042 | T | C | 6 | RP1-97D16.1 | Male only | False | 0.1077 | 0.1083 | 0.1337 | 0.1336 | 25647 | 19931 | 187758 | 162098 | -0.009083 | -0.07365 | -0.05121 | -0.04973 | 0.01331 | 0.01454 |
| MONOAB | rs17693963 | rs200977 | C | T | 6 | RP1-97D16.1 | Male only | False | 0.1078 | 0.1085 | 0.1338 | 0.1337 | 25647 | 19931 | 187758 | 162098 | -0.01033 | -0.07417 | -0.05129 | -0.04969 | 0.01333 | 0.01456 |
| MONOAB | rs17693963 | rs13199649 | T | C | 6 | RP1-97D16.1 | Male only | False | 0.08885 | 0.09057 | 0.1162 | 0.1165 | 25647 | 19931 | 187758 | 162098 | -0.02391 | -0.08536 | -0.05926 | -0.05638 | 0.01448 | 0.0158 |
| MONOAB | rs17693963 | rs34788973 | A | C | 6 | RP1-97D16.1 | Male only | False | 0.08988 | 0.09166 | 0.1164 | 0.1166 | 25647 | 19931 | 187758 | 162098 | -0.02413 | -0.08567 | -0.05586 | -0.05427 | 0.01437 | 0.0157 |
| MONOAB | rs17693963 | rs61742093 | G | A | 6 | RP1-97D16.1 | Male only | False | 0.08989 | 0.09167 | 0.1171 | 0.1173 | 25647 | 19931 | 187758 | 162098 | -0.02411 | -0.08565 | -0.05788 | -0.05628 | 0.01437 | 0.0157 |
| MONOAB | rs17693963 | rs13218875 | T | C | 6 | RP1-97D16.1 | Male only | False | 0.08884 | 0.09054 | 0.1165 | 0.1167 | 25647 | 19931 | 187758 | 162098 | -0.02376 | -0.08484 | -0.0589 | -0.05669 | 0.01445 | 0.01579 |
| MONOAB | rs17693963 | rs71559054 | C | A | 6 | RP1-97D16.1 | Male only | False | 0.08891 | 0.09052 | 0.1164 | 0.1167 | 25647 | 19931 | 187758 | 162098 | -0.02342 | -0.08424 | -0.05913 | -0.0569 | 0.01445 | 0.0158 |
| MONOAB | rs17693963 | rs67040724 | C | T | 6 | RP1-97D16.1 | Male only | False | 0.0889 | 0.0905 | 0.1164 | 0.1167 | 25647 | 19931 | 187758 | 162098 | -0.02343 | -0.08412 | -0.05916 | -0.05682 | 0.01445 | 0.0158 |
| MONOAB | rs735765 | rs67662114 | A | G | 6 | ZNF192P2 | Male only | False | 0.08889 | 0.0905 | 0.1164 | 0.1167 | 25647 | 19931 | 187758 | 162098 | -0.02344 | -0.08411 | -0.05914 | -0.05669 | 0.01445 | 0.0158 |
| MONOAB | rs735765 | rs13216117 | G | A | 6 | ZNF192P2 | Male only | False | 0.08894 | 0.09054 | 0.1164 | 0.1167 | 25647 | 19931 | 187758 | 162098 | -0.02347 | -0.08418 | -0.0592 | -0.05669 | 0.01445 | 0.0158 |
| MONOAB | rs735765 | rs36101351 | T | C | 6 | ZNF192P2 | Male only | False | 0.08888 | 0.0905 | 0.1164 | 0.1167 | 25647 | 19931 | 187758 | 162098 | -0.02356 | -0.0841 | -0.05917 | -0.0567 | 0.01445 | 0.0158 |
| MONOAB | rs735765 | rs28360499 | A | G | 6 | ZNF192P2 | Male only | False | 0.08889 | 0.09051 | 0.1164 | 0.1167 | 25647 | 19931 | 187758 | 162098 | -0.02355 | -0.0841 | -0.05917 | -0.05672 | 0.01445 | 0.0158 |
| MONOAB | rs735765 | rs156743 | C | T | 6 | ZNF192P2 | Male only | False | 0.1255 | 0.1264 | 0.1489 | 0.1492 | 25647 | 19931 | 187758 | 162098 | -0.01465 | -0.06838 | -0.04295 | -0.04137 | 0.0124 | 0.01361 |
| MONOAB | rs735765 | rs71537572 | C | T | 6 | ZNF192P2 | Male only | False | 0.08827 | 0.08986 | 0.1146 | 0.1149 | 25647 | 19931 | 187758 | 162098 | -0.02405 | -0.08483 | -0.06002 | -0.05725 | 0.01454 | 0.0159 |
| MONOAB | rs735765 | rs138234416 | A | G | 6 | ZNF192P2 | Male only | False | 0.08973 | 0.09161 | 0.1168 | 0.1171 | 25647 | 19931 | 187758 | 162098 | -0.02403 | -0.08498 | -0.05841 | -0.05652 | 0.01439 | 0.01572 |
| MONOAB | rs735765 | rs149990 | A | G | 6 | ZNF192P2 | Male only | False | 0.1252 | 0.1263 | 0.1485 | 0.1489 | 25647 | 19931 | 187758 | 162098 | -0.01477 | -0.06929 | -0.04306 | -0.03984 | 0.01241 | 0.01362 |
| MONOAB | rs735765 | rs149943 | A | G | 6 | ZNF192P2 | Male only | False | 0.1252 | 0.1263 | 0.1493 | 0.1496 | 25647 | 19931 | 187758 | 162098 | -0.01477 | -0.06929 | -0.04319 | -0.04126 | 0.01241 | 0.01362 |
| MONOAB | rs735765 | rs149949 | C | T | 6 | ZNF192P2 | Male only | False | 0.1265 | 0.1276 | 0.1502 | 0.1505 | 25647 | 19931 | 187758 | 162098 | -0.01437 | -0.06994 | -0.0438 | -0.04151 | 0.01236 | 0.01357 |
| MONOAB | rs735765 | rs202906 | C | T | 6 | ZNF192P2 | Male only | False | 0.1267 | 0.1277 | 0.1503 | 0.1506 | 25647 | 19931 | 187758 | 162098 | -0.01486 | -0.07036 | -0.04371 | -0.04138 | 0.01235 | 0.01356 |
| MONOAB | rs735765 | rs13200214 | T | C | 6 | ZNF192P2 | Male only | False | 0.08884 | 0.09063 | 0.1164 | 0.1167 | 25647 | 19931 | 187758 | 162098 | -0.02432 | -0.0837 | -0.05944 | -0.05671 | 0.01446 | 0.01579 |

| trait | lead_snp | ID | ALLELE1 | ALLELE0 | CHR | Gene_Symbol | independent_loci_in_sex_P1e2 | in_embl_any_or_gene | A1FREQ_VUMC_female | A1FREQ_VUMC_male | A1FREQ_UKB_female | A1FREQ_UKB_male | N_VUMC_female | N_VUMC_male | N_UKB_female | N_UKB_male | BETA_VUMC_female | BETA_VUMC_male | BETA_UKB_female | BETA_UKB_male | SE_VUMC_female | SE_VUMC_male |
|---|---|---|---|---|---|---|---|---|---|---|---|---|---|---|---|---|---|---|---|---|---|---|
| MONOAB | rs735765 | rs35949109 | C | T | 6 | ZNF192P2 | Male only | False | 0.1255 | 0.1265 | 0.1494 | 0.1497 | 25647 | 19931 | 187758 | 162098 | -0.01398 | -0.06814 | -0.0437 | -0.04165 | 0.01241 | 0.01362 |
| MONOAB | rs735765 | rs71559070 | A | G | 6 | ZNF192P2 | Male only | False | 0.08869 | 0.09046 | 0.1165 | 0.1169 | 25647 | 19931 | 187758 | 162098 | -0.02377 | -0.08174 | -0.06048 | -0.05742 | 0.01447 | 0.01581 |
| MONOAB | rs735765 | rs4713135 | A | G | 6 | ZNF192P2 | Male only | False | 0.2167 | 0.2169 | 0.2392 | 0.2405 | 25647 | 19931 | 187758 | 162098 | -7.45e-04 | -0.04344 | -0.02823 | -0.02821 | 0.009942 | 0.01096 |
| MONOAB | rs735765 | rs34712084 | G | A | 6 | ZNF192P2 | Male only | False | 0.1279 | 0.1294 | 0.1531 | 0.154 | 25647 | 19931 | 187758 | 162098 | -0.009324 | -0.06848 | -0.04578 | -0.0426 | 0.01231 | 0.01345 |
| MONOAB | rs735765 | rs1321505 | C | A | 6 | ZNF192P2 | Male only | False | 0.1279 | 0.1293 | 0.1531 | 0.154 | 25647 | 19931 | 187758 | 162098 | -0.009269 | -0.06868 | -0.04586 | -0.04255 | 0.01231 | 0.01346 |
| MONOAB | rs735765 | rs13197574 | C | T | 6 | ZNF192P2 | Male only | False | 0.08865 | 0.09039 | 0.1166 | 0.117 | 25647 | 19931 | 187758 | 162098 | -0.02355 | -0.08151 | -0.06033 | -0.05725 | 0.01447 | 0.01581 |
| MONOAB | rs735765 | rs34166054 | C | T | 6 | ZNF192P2 | Male only | False | 0.08863 | 0.09037 | 0.1165 | 0.1169 | 25647 | 19931 | 187758 | 162098 | -0.02354 | -0.08152 | -0.06054 | -0.05738 | 0.01447 | 0.01581 |
| MONOAB | rs735765 | rs13437444 | T | C | 6 | ZNF192P2 | Male only | False | 0.1872 | 0.1887 | 0.217 | 0.2178 | 25647 | 19931 | 187758 | 162098 | -0.005038 | -0.04964 | -0.03048 | -0.03155 | 0.01052 | 0.01154 |
| MONOAB | rs735765 | rs34662244 | A | G | 6 | ZNF192P2 | Male only | False | 0.08863 | 0.09037 | 0.1165 | 0.1169 | 25647 | 19931 | 187758 | 162098 | -0.02354 | -0.08151 | -0.06056 | -0.05746 | 0.01447 | 0.01581 |
| MONOAB | rs735765 | rs13203816 | C | T | 6 | ZNF192P2 | Male only | False | 0.08863 | 0.09037 | 0.1165 | 0.1169 | 25647 | 19931 | 187758 | 162098 | -0.02354 | -0.08151 | -0.06056 | -0.05748 | 0.01447 | 0.01581 |
| MONOAB | rs735765 | rs9468287 | A | C | 6 | ZNF192P2 | Male only | False | 0.1279 | 0.1292 | 0.1534 | 0.1544 | 25647 | 19931 | 187758 | 162098 | -0.009386 | -0.0693 | -0.0454 | -0.04314 | 0.0123 | 0.01346 |
| MONOAB | rs735765 | rs34588114 | A | C | 6 | ZNF192P2 | Male only | False | 0.08863 | 0.09037 | 0.1166 | 0.1169 | 25647 | 19931 | 187758 | 162098 | -0.02354 | -0.08151 | -0.06059 | -0.0575 | 0.01447 | 0.01581 |
| MONOAB | rs735765 | rs68188794 | C | T | 6 | ZNF192P2 | Male only | False | 0.08863 | 0.09037 | 0.1166 | 0.117 | 25647 | 19931 | 187758 | 162098 | -0.02354 | -0.08151 | -0.06057 | -0.05751 | 0.01447 | 0.01581 |
| MONOAB | rs735765 | rs34371502 | C | T | 6 | ZNF192P2 | Male only | False | 0.08863 | 0.09037 | 0.1166 | 0.1169 | 25647 | 19931 | 187758 | 162098 | -0.02354 | -0.08151 | -0.06059 | -0.0575 | 0.01447 | 0.01581 |
| MONOAB | rs735765 | rs9461432 | C | T | 6 | ZNF192P2 | Male only | False | 0.1325 | 0.1337 | 0.1578 | 0.1586 | 25647 | 19931 | 187758 | 162098 | -0.01593 | -0.06922 | -0.04462 | -0.04174 | 0.01211 | 0.01328 |
| MONOAB | rs735765 | rs71559072 | T | C | 6 | ZNF192P2 | Male only | False | 0.08864 | 0.09038 | 0.1166 | 0.117 | 25647 | 19931 | 187758 | 162098 | -0.02354 | -0.0815 | -0.06054 | -0.0575 | 0.01447 | 0.01581 |
| MONOAB | rs735765 | 6:28089731 | T | C | 6 | ZNF192P2 | Male only | False | 0.08863 | 0.09037 | | | 25647 | 19931 | | | -0.02354 | -0.0815 | | | 0.01447 | 0.01581 |
| MONOAB | rs735765 | rs35345226 | G | A | 6 | ZNF192P2 | Male only | False | 0.08863 | 0.09037 | 0.1166 | 0.117 | 25647 | 19931 | 187758 | 162098 | -0.02354 | -0.08149 | -0.06054 | -0.05752 | 0.01447 | 0.01581 |
| MONOAB | rs735765 | rs4713139 | T | C | 6 | ZNF192P2 | Male only | False | 0.1326 | 0.1337 | 0.1578 | 0.1586 | 25647 | 19931 | 187758 | 162098 | -0.01593 | -0.06921 | -0.0445 | -0.04175 | 0.01211 | 0.01327 |
| MONOAB | rs735765 | rs4713140 | A | G | 6 | ZNF192P2 | Male only | False | 0.1326 | 0.1337 | 0.1578 | 0.1587 | 25647 | 19931 | 187758 | 162098 | -0.01589 | -0.06924 | -0.04468 | -0.0416 | 0.01211 | 0.01327 |
| MONOAB | rs735765 | rs34130214 | T | C | 6 | ZNF192P2 | Male only | False | 0.1325 | 0.1336 | 0.1578 | 0.1586 | 25647 | 19931 | 187758 | 162098 | -0.016 | -0.06919 | -0.04459 | -0.0417 | 0.01211 | 0.01328 |
| MONOAB | rs735765 | rs3757188 | C | T | 6 | ZNF192P2 | Male only | False | 0.1325 | 0.1336 | 0.1586 | 0.1596 | 25647 | 19931 | 187758 | 162098 | -0.0164 | -0.06927 | -0.04375 | -0.04173 | 0.01211 | 0.01328 |

| trait | lead_snp | ID | ALLELE1 | ALLELE0 | CHR | Gene_Symbol | independent_loci_in_sex_P1e2 | in_embl_any_or_gene | A1FREQ_VUMC_female | A1FREQ_VUMC_male | A1FREQ_UKB_female | A1FREQ_UKB_male | N_VUMC_female | N_VUMC_male | N_UKB_female | N_UKB_male | BETA_VUMC_female | BETA_VUMC_male | BETA_UKB_female | BETA_UKB_male | SE_VUMC_female | SE_VUMC_male |
|---|---|---|---|---|---|---|---|---|---|---|---|---|---|---|---|---|---|---|---|---|---|---|
| MONOAB | rs735765 | rs1904841 | T | C | 6 | ZNF192P2 | Male only | False | 0.2181 | 0.2182 | 0.2419 | 0.2431 | 25647 | 19931 | 187758 | 162098 | -0.001999 | -0.04279 | -0.02818 | -0.02717 | 0.009913 | 0.01091 |
| MONOAB | rs735765 | rs1904840 | T | C | 6 | ZNF192P2 | Male only | False | 0.2178 | 0.218 | 0.2417 | 0.243 | 25647 | 19931 | 187758 | 162098 | -0.001928 | -0.04292 | -0.02808 | -0.02716 | 0.009919 | 0.01092 |
| MONOAB | rs735765 | rs9368555 | C | T | 6 | ZNF192P2 | Male only | False | 0.2178 | 0.2178 | 0.2417 | 0.2429 | 25647 | 19931 | 187758 | 162098 | -0.001929 | -0.043 | -0.02808 | -0.02734 | 0.009919 | 0.01092 |
| MONOAB | rs735765 | rs9393893 | C | T | 6 | ZNF192P2 | Male only | False | 0.2178 | 0.2178 | 0.2417 | 0.2429 | 25647 | 19931 | 187758 | 162098 | -0.001921 | -0.04301 | -0.02805 | -0.02735 | 0.00992 | 0.01092 |
| MONOAB | rs735765 | rs868987 | T | C | 6 | ZNF192P2 | Male only | False | 0.2178 | 0.2178 | 0.2416 | 0.2429 | 25647 | 19931 | 187758 | 162098 | -0.001932 | -0.04301 | -0.02802 | -0.02742 | 0.009919 | 0.01092 |

| trait | lead_snp | ID | ALLELE1 | ALLELE0 | CHR | Gene_Symbol | independent_loci_in_sex_P1e2 | in_embl_any_or_gene | A1FREQ_VUMC_female | A1FREQ_VUMC_male | A1FREQ_UKB_female | A1FREQ_UKB_male | N_VUMC_female | N_VUMC_male | N_UKB_female | N_UKB_male | BETA_VUMC_female | BETA_VUMC_male | BETA_UKB_female | BETA_UKB_male | SE_VUMC_female | SE_VUMC_male |
|---|---|---|---|---|---|---|---|---|---|---|---|---|---|---|---|---|---|---|---|---|---|---|
| MONOAB | rs735765 | rs9393894 | G | A | 6 | ZNF192P2 | Male only | False | 0.2178 | 0.2178 | 0.2416 | 0.2429 | 25647 | 19931 | 187758 | 162098 | -0.001958 | -0.043 | -0.02804 | -0.02736 | 0.009919 | 0.01092 |
| MONOAB | rs735765 | rs9468295 | C | T | 6 | ZNF192P2 | Male only | False | 0.2178 | 0.2178 | 0.2416 | 0.2429 | 25647 | 19931 | 187758 | 162098 | -0.001958 | -0.043 | -0.02803 | -0.02736 | 0.009919 | 0.01092 |
| MONOAB | rs735765 | rs9468296 | G | A | 6 | ZNF192P2 | Male only | False | 0.2178 | 0.2178 | 0.2417 | 0.243 | 25647 | 19931 | 187758 | 162098 | -0.001958 | -0.043 | -0.02795 | -0.02728 | 0.009919 | 0.01092 |
| MONOAB | rs735765 | rs35749575 | T | G | 6 | ZNF192P2 | Male only | False | 0.08864 | 0.09037 | 0.1166 | 0.117 | 25647 | 19931 | 187758 | 162098 | -0.02356 | -0.08152 | -0.06049 | -0.05756 | 0.01447 | 0.01581 |
| MONOAB | rs735765 | rs4711164 | T | C | 6 | ZNF192P2 | Male only | False | 0.2178 | 0.2178 | 0.2418 | 0.243 | 25647 | 19931 | 187758 | 162098 | -0.00196 | -0.04301 | -0.02811 | -0.02739 | 0.00992 | 0.01092 |
| MONOAB | rs735765 | rs4711165 | G | T | 6 | ZNF192P2 | Male only | False | 0.2178 | 0.2178 | 0.244 | 0.2451 | 25647 | 19931 | 187758 | 162098 | -0.00188 | -0.043 | -0.02735 | -0.02655 | 0.009921 | 0.01092 |
| MONOAB | rs735765 | rs4713148 | A | G | 6 | ZNF192P2 | Male only | False | 0.2178 | 0.2178 | 0.2416 | 0.2429 | 25647 | 19931 | 187758 | 162098 | -0.001963 | -0.04299 | -0.02811 | -0.02733 | 0.00992 | 0.01092 |
| MONOAB | rs735765 | rs9468297 | T | G | 6 | ZNF192P2 | Male only | False | 0.2178 | 0.2178 | 0.2416 | 0.2429 | 25647 | 19931 | 187758 | 162098 | -0.001963 | -0.04299 | -0.02811 | -0.02732 | 0.00992 | 0.01092 |
| MONOAB | rs735765 | rs72846780 | C | T | 6 | ZNF192P2 | Male only | False | 0.08863 | 0.09036 | 0.1166 | 0.117 | 25647 | 19931 | 187758 | 162098 | -0.02353 | -0.08152 | -0.06051 | -0.05756 | 0.01447 | 0.01581 |
| MONOAB | rs735765 | rs17774663 | A | G | 6 | ZNF192P2 | Male only | False | 0.2178 | 0.2178 | 0.2416 | 0.2429 | 25647 | 19931 | 187758 | 162098 | -0.001923 | -0.04299 | -0.02811 | -0.02731 | 0.00992 | 0.01092 |
| MONOAB | rs735765 | rs9468298 | G | A | 6 | ZNF192P2 | Male only | False | 0.2178 | 0.2178 | 0.2416 | 0.2429 | 25647 | 19931 | 187758 | 162098 | -0.001962 | -0.04299 | -0.0281 | -0.02731 | 0.00992 | 0.01092 |
| MONOAB | rs735765 | rs13205911 | T | C | 6 | ZNF192P2 | Male only | False | 0.08865 | 0.09036 | 0.1164 | 0.1168 | 25647 | 19931 | 187758 | 162098 | -0.02354 | -0.08151 | -0.06094 | -0.05656 | 0.01447 | 0.01581 |
| MONOAB | rs735765 | rs9295759 | T | C | 6 | ZNF192P2 | Male only | False | 0.2178 | 0.2178 | 0.2416 | 0.2429 | 25647 | 19931 | 187758 | 162098 | -0.001964 | -0.04299 | -0.0281 | -0.02732 | 0.00992 | 0.01092 |
| MONOAB | rs735765 | rs9348796 | G | A | 6 | ZNF192P2 | Male only | False | 0.2178 | 0.2178 | 0.2416 | 0.2429 | 25647 | 19931 | 187758 | 162098 | -0.001964 | -0.04299 | -0.02813 | -0.02732 | 0.00992 | 0.01092 |
| MONOAB | rs735765 | rs11552219 | T | C | 6 | ZNF192P2 | Male only | False | 0.2178 | 0.2178 | 0.2416 | 0.2429 | 25647 | 19931 | 187758 | 162098 | -0.001964 | -0.04299 | -0.02812 | -0.02734 | 0.00992 | 0.01092 |
| MONOAB | rs735765 | rs9468300 | T | C | 6 | ZNF192P2 | Male only | False | 0.1325 | 0.1335 | 0.1578 | 0.1586 | 25647 | 19931 | 187758 | 162098 | -0.0164 | -0.06982 | -0.04445 | -0.04192 | 0.01211 | 0.01328 |
| MONOAB | rs735765 | rs9380058 | C | T | 6 | ZNF192P2 | Male only | False | 0.2181 | 0.2179 | 0.2418 | 0.243 | 25647 | 19931 | 187758 | 162098 | -0.00204 | -0.04282 | -0.0282 | -0.02735 | 0.009914 | 0.01091 |
| MONOAB | rs735765 | rs9393895 | C | T | 6 | ZNF192P2 | Male only | False | 0.2178 | 0.2177 | 0.2416 | 0.2429 | 25647 | 19931 | 187758 | 162098 | -0.001946 | -0.04298 | -0.02811 | -0.02733 | 0.00992 | 0.01092 |
| MONOAB | rs735765 | rs9393896 | T | C | 6 | ZNF192P2 | Male only | False | 0.2185 | 0.2184 | 0.2426 | 0.2438 | 25647 | 19931 | 187758 | 162098 | -0.001693 | -0.04309 | -0.02833 | -0.02734 | 0.009919 | 0.01092 |
| MONOAB | rs735765 | rs9393897 | T | G | 6 | ZNF192P2 | Male only | False | 0.2185 | 0.2184 | 0.243 | 0.2443 | 25647 | 19931 | 187758 | 162098 | -0.00151 | -0.04349 | -0.02841 | -0.02743 | 0.009917 | 0.01092 |
| MONOAB | rs735765 | rs13197175 | T | C | 6 | ZNF192P2 | Male only | False | 0.08864 | 0.09035 | 0.1166 | 0.117 | 25647 | 19931 | 187758 | 162098 | -0.02355 | -0.08154 | -0.06045 | -0.05773 | 0.01447 | 0.01581 |
| MONOAB | rs735765 | rs13197176 | T | C | 6 | ZNF192P2 | Male only | False | 0.08832 | 0.09004 | 0.1153 | 0.1158 | 25647 | 19931 | 187758 | 162098 | -0.02369 | -0.08195 | -0.06094 | -0.05812 | 0.01452 | 0.01586 |
| MONOAB | rs735765 | rs9357065 | C | T | 6 | ZNF192P2 | Male only | False | 0.2177 | 0.2176 | 0.2416 | 0.2429 | 25647 | 19931 | 187758 | 162098 | -0.001869 | -0.04268 | -0.02807 | -0.02732 | 0.009922 | 0.01092 |
| MONOAB | rs735765 | rs9357066 | C | T | 6 | ZNF192P2 | Male only | False | 0.2178 | 0.2177 | 0.2416 | 0.2429 | 25647 | 19931 | 187758 | 162098 | -0.00197 | -0.04298 | -0.02814 | -0.02736 | 0.00992 | 0.01092 |
| MONOAB | rs735765 | rs13201308 | T | C | 6 | ZNF192P2 | Male only | False | 0.08864 | 0.09034 | 0.1166 | 0.117 | 25647 | 19931 | 187758 | 162098 | -0.02355 | -0.08154 | -0.06045 | -0.05773 | 0.01447 | 0.01581 |
| MONOAB | rs735765 | rs9393898 | T | G | 6 | ZNF192P2 | Male only | False | 0.2181 | 0.2179 | 0.2418 | 0.243 | 25647 | 19931 | 187758 | 162098 | -0.002046 | -0.04282 | -0.02822 | -0.02739 | 0.009914 | 0.01091 |
| MONOAB | rs735765 | rs34765154 | A | G | 6 | ZNF192P2 | Male only | False | 0.08864 | 0.09034 | 0.1166 | 0.117 | 25647 | 19931 | 187758 | 162098 | -0.02355 | -0.08155 | -0.06045 | -0.05773 | 0.01447 | 0.01581 |
| MONOAB | rs735765 | rs9368556 | C | T | 6 | ZNF192P2 | Male only | False | 0.2178 | 0.2177 | 0.2416 | 0.2429 | 25647 | 19931 | 187758 | 162098 | -0.001974 | -0.04297 | -0.02816 | -0.02739 | 0.00992 | 0.01092 |
| MONOAB | rs735765 | rs9368557 | C | T | 6 | ZNF192P2 | Male only | False | 0.2178 | 0.2177 | 0.2416 | 0.2429 | 25647 | 19931 | 187758 | 162098 | -0.001974 | -0.04297 | -0.02817 | -0.0274 | 0.00992 | 0.01092 |

| trait | lead_snp | ID | ALLELE1 | ALLELE0 | CHR | Gene_Symbol | independent_loci_in_sex_P1e2 | in_embl_any_or_gene | A1FREQ_VUMC_female | A1FREQ_VUMC_male | A1FREQ_UKB_female | A1FREQ_UKB_male | N_VUMC_female | N_VUMC_male | N_UKB_female | N_UKB_male | BETA_VUMC_female | BETA_VUMC_male | BETA_UKB_female | BETA_UKB_male | SE_VUMC_female | SE_VUMC_male |
|---|---|---|---|---|---|---|---|---|---|---|---|---|---|---|---|---|---|---|---|---|---|---|
| MONOAB | rs735765 | rs9380059 | C | A | 6 | ZNF192P2 | Male only | False | 0.2178 | 0.2177 | 0.2417 | 0.2429 | 25647 | 19931 | 187758 | 162098 | -0.001977 | -0.04298 | -0.02817 | -0.02751 | 0.00992 | 0.01092 |
| MONOAB | rs735765 | rs9380060 | A | G | 6 | ZNF192P2 | Male only | False | 0.2178 | 0.2177 | 0.2416 | 0.2429 | 25647 | 19931 | 187758 | 162098 | -0.001973 | -0.04296 | -0.02816 | -0.02742 | 0.00992 | 0.01092 |
| MONOAB | rs735765 | rs35227624 | G | A | 6 | ZNF192P2 | Male only | False | 0.2178 | 0.2177 | 0.2416 | 0.2428 | 25647 | 19931 | 187758 | 162098 | -0.001973 | -0.04296 | -0.02814 | -0.02741 | 0.00992 | 0.01092 |
| MONOAB | rs735765 | rs9380061 | C | T | 6 | ZNF192P2 | Male only | False | 0.2178 | 0.2177 | 0.2416 | 0.2429 | 25647 | 19931 | 187758 | 162098 | -0.001973 | -0.04296 | -0.02816 | -0.02742 | 0.00992 | 0.01092 |
| MONOAB | rs735765 | rs9368558 | G | A | 6 | ZNF192P2 | Male only | False | 0.2178 | 0.2177 | 0.2416 | 0.2429 | 25647 | 19931 | 187758 | 162098 | -0.00197 | -0.04296 | -0.02816 | -0.02742 | 0.00992 | 0.01092 |
| MONOAB | rs735765 | rs35098436 | C | T | 6 | ZNF192P2 | Male only | False | 0.08864 | 0.09034 | 0.1166 | 0.117 | 25647 | 19931 | 187758 | 162098 | -0.02357 | -0.08157 | -0.06046 | -0.05775 | 0.01447 | 0.01581 |
| MONOAB | rs735765 | rs4713150 | A | G | 6 | ZNF192P2 | Male only | False | 0.2178 | 0.2177 | 0.2416 | 0.2429 | 25647 | 19931 | 187758 | 162098 | -0.001972 | -0.04295 | -0.02818 | -0.02741 | 0.00992 | 0.01092 |
| MONOAB | rs735765 | rs4713151 | A | G | 6 | ZNF192P2 | Male only | False | 0.2178 | 0.2177 | 0.2416 | 0.2428 | 25647 | 19931 | 187758 | 162098 | -0.001971 | -0.04296 | -0.02819 | -0.02741 | 0.00992 | 0.01092 |
| MONOAB | rs735765 | rs9393901 | G | A | 6 | ZNF192P2 | Male only | False | 0.2178 | 0.2177 | 0.2416 | 0.2428 | 25647 | 19931 | 187758 | 162098 | -0.001971 | -0.04296 | -0.0282 | -0.0274 | 0.00992 | 0.01092 |
| MONOAB | rs735765 | rs3173443 | G | T | 6 | ZNF192P2 | Male only | False | 0.2178 | 0.2177 | 0.2416 | 0.2428 | 25647 | 19931 | 187758 | 162098 | -0.001971 | -0.04296 | -0.02827 | -0.02728 | 0.00992 | 0.01092 |
| MONOAB | rs735765 | rs4713152 | A | G | 6 | ZNF192P2 | Male only | False | 0.2178 | 0.2177 | 0.2416 | 0.2429 | 25647 | 19931 | 187758 | 162098 | -0.001971 | -0.04296 | -0.02822 | -0.02726 | 0.00992 | 0.01092 |
| MONOAB | rs735765 | rs9380062 | A | G | 6 | ZNF192P2 | Male only | False | 0.2178 | 0.2177 | 0.2416 | 0.2428 | 25647 | 19931 | 187758 | 162098 | -0.001973 | -0.04295 | -0.02819 | -0.02738 | 0.00992 | 0.01092 |
| MONOAB | rs735765 | rs9380063 | G | A | 6 | ZNF192P2 | Male only | False | 0.2178 | 0.2177 | 0.2416 | 0.2428 | 25647 | 19931 | 187758 | 162098 | -0.001969 | -0.04296 | -0.0282 | -0.02739 | 0.00992 | 0.01092 |
| MONOAB | rs735765 | 6:28141253 | T | G | 6 | ZNF192P2 | Male only | False | 0.08864 | 0.09035 | | | 25647 | 19931 | | | -0.02361 | -0.0816 | | | 0.01447 | 0.01581 |
| MONOAB | rs735765 | rs8180562 | A | G | 6 | ZNF192P2 | Male only | False | 0.1326 | 0.1335 | 0.1579 | 0.1587 | 25647 | 19931 | 187758 | 162098 | -0.01672 | -0.06994 | -0.04439 | -0.04185 | 0.01211 | 0.01328 |
| MONOAB | rs735765 | rs12332979 | G | T | 6 | ZNF192P2 | Male only | False | 0.2178 | 0.2177 | 0.2416 | 0.2428 | 25647 | 19931 | 187758 | 162098 | -0.001979 | -0.04295 | -0.02817 | -0.02739 | 0.00992 | 0.01092 |
| MONOAB | rs735765 | rs9348798 | A | G | 6 | ZNF192P2 | Male only | False | 0.2178 | 0.2177 | 0.2416 | 0.2428 | 25647 | 19931 | 187758 | 162098 | -0.001944 | -0.04299 | -0.02817 | -0.0274 | 0.00992 | 0.01092 |
| MONOAB | rs735765 | rs9380064 | G | A | 6 | ZNF192P2 | Male only | False | 0.1325 | 0.1335 | 0.1578 | 0.1586 | 25647 | 19931 | 187758 | 162098 | -0.0164 | -0.06983 | -0.04439 | -0.04199 | 0.01211 | 0.01328 |
| MONOAB | rs735765 | rs9380065 | G | A | 6 | ZNF192P2 | Male only | False | 0.2178 | 0.2177 | 0.2416 | 0.2428 | 25647 | 19931 | 187758 | 162098 | -0.001946 | -0.04299 | -0.02817 | -0.02742 | 0.00992 | 0.01092 |
| MONOAB | rs735765 | rs35781323 | C | T | 6 | ZNF192P2 | Male only | False | 0.08864 | 0.09034 | 0.1165 | 0.117 | 25647 | 19931 | 187758 | 162098 | -0.02359 | -0.0816 | -0.06046 | -0.05783 | 0.01447 | 0.01581 |
| MONOAB | rs735765 | rs9295761 | G | A | 6 | ZNF192P2 | Male only | False | 0.2176 | 0.2176 | 0.2418 | 0.243 | 25647 | 19931 | 187758 | 162098 | -0.001874 | -0.04333 | -0.02824 | -0.02719 | 0.009921 | 0.01092 |
| MONOAB | rs735765 | rs34832585 | T | C | 6 | ZNF192P2 | Male only | False | 0.08859 | 0.09027 | 0.1162 | 0.1167 | 25647 | 19931 | 187758 | 162098 | -0.02343 | -0.08227 | -0.06092 | -0.05814 | 0.01448 | 0.01582 |
| MONOAB | rs735765 | rs6901017 | C | T | 6 | ZNF192P2 | Male only | False | 0.2242 | 0.2235 | 0.2454 | 0.2465 | 25647 | 19931 | 187758 | 162098 | -0.003896 | -0.04499 | -0.02842 | -0.02707 | 0.009819 | 0.01083 |
| MONOAB | rs735765 | rs9295762 | C | T | 6 | ZNF192P2 | Male only | False | 0.2242 | 0.2235 | 0.2454 | 0.2465 | 25647 | 19931 | 187758 | 162098 | -0.003955 | -0.04529 | -0.02841 | -0.02707 | 0.009819 | 0.01083 |
| MONOAB | rs735765 | rs9368560 | C | T | 6 | ZNF192P2 | Male only | False | 0.2105 | 0.2104 | 0.2333 | 0.2342 | 25647 | 19931 | 187758 | 162098 | -0.002805 | -0.04829 | -0.02846 | -0.02818 | 0.01005 | 0.01109 |
| MONOAB | rs735765 | rs1225598 | G | T | 6 | ZNF192P2 | Male only | False | 0.2246 | 0.224 | 0.2457 | 0.2468 | 25647 | 19931 | 187758 | 162098 | -0.003239 | -0.04624 | -0.02831 | -0.02693 | 0.009808 | 0.01081 |
| trait | lead_snp | ID | ALLELE1 | ALLELE0 | CHR | Gene_Symbol | independent_loci_in_sex_P1e2 | in_embl_any_or_gene | A1FREQ_VUMC_female | A1FREQ_VUMC_male | A1FREQ_UKB_female | A1FREQ_UKB_male | N_VUMC_female | N_VUMC_male | N_UKB_female | N_UKB_male | BETA_VUMC_female | BETA_VUMC_male | BETA_UKB_female | BETA_UKB_male | SE_VUMC_female | SE_VUMC_male |
| MONOAB | rs735765 | rs1225596 | C | T | 6 | ZNF192P2 | Male only | False | 0.2245 | 0.2239 | 0.2457 | 0.2468 | 25647 | 19931 | 187758 | 162098 | -0.003253 | -0.04626 | -0.02829 | -0.02692 | 0.009812 | 0.01082 |

| trait | lead_snp | ID | ALLELE1 | ALLELE0 | CHR | Gene_Symbol | independent_loci_in_sex_P1e2 | in_embl_any_or_gene | A1FREQ_VUMC_female | A1FREQ_VUMC_male | A1FREQ_UKB_female | A1FREQ_UKB_male | N_VUMC_female | N_VUMC_male | N_UKB_female | N_UKB_male | BETA_VUMC_female | BETA_VUMC_male | BETA_UKB_female | BETA_UKB_male | SE_VUMC_female | SE_VUMC_male |
|---|---|---|---|---|---|---|---|---|---|---|---|---|---|---|---|---|---|---|---|---|---|---|
| MONOAB | rs735765 | rs1150689 | C | T | 6 | ZNF192P2 | Male only | False | 0.2246 | 0.224 | 0.2457 | 0.2468 | 25647 | 19931 | 187758 | 162098 | -0.003238 | -0.04624 | -0.02842 | -0.02684 | 0.009808 | 0.01081 |
| MONOAB | rs735765 | rs1225599 | T | C | 6 | ZNF192P2 | Male only | False | 0.2246 | 0.224 | 0.2457 | 0.2468 | 25647 | 19931 | 187758 | 162098 | -0.003238 | -0.04624 | -0.02828 | -0.02691 | 0.009808 | 0.01081 |
| MONOAB | rs735765 | rs1233704 | T | C | 6 | ZNF192P2 | Male only | False | 0.2246 | 0.224 | 0.2459 | 0.247 | 25647 | 19931 | 187758 | 162098 | -0.003238 | -0.04624 | -0.02805 | -0.02699 | 0.009808 | 0.01081 |
| MONOAB | rs735765 | rs9368561 | T | C | 6 | ZNF192P2 | Male only | False | 0.1278 | 0.1291 | 0.1531 | 0.154 | 25647 | 19931 | 187758 | 162098 | -0.009678 | -0.07098 | -0.04575 | -0.04302 | 0.01231 | 0.01347 |
| MONOAB | rs735765 | rs13195291 | A | G | 6 | ZNF192P2 | Male only | False | 0.08867 | 0.09036 | 0.1166 | 0.1171 | 25647 | 19931 | 187758 | 162098 | -0.02335 | -0.0826 | -0.06042 | -0.0578 | 0.01447 | 0.01581 |
| MONOAB | rs735765 | rs735765 | A | G | 6 | ZNF192P2 | Male only | False | 0.1369 | 0.1382 | 0.1601 | 0.161 | 25647 | 19931 | 187758 | 162098 | -0.02229 | -0.07722 | -0.04399 | -0.04037 | 0.01194 | 0.01311 |
| MONOAB | rs735765 | rs1233697 | A | G | 6 | ZNF192P2 | Male only | False | 0.2246 | 0.2239 | 0.2457 | 0.2468 | 25647 | 19931 | 187758 | 162098 | -0.003168 | -0.04626 | -0.02824 | -0.02694 | 0.009812 | 0.01082 |
| MONOAB | rs735765 | rs1233707 | A | G | 6 | ZNF192P2 | Male only | False | 0.2245 | 0.2239 | 0.2457 | 0.2468 | 25647 | 19931 | 187758 | 162098 | -0.00325 | -0.04627 | -0.02825 | -0.02692 | 0.009812 | 0.01082 |
| MONOAB | rs735765 | rs1237875 | T | C | 6 | ZNF192P2 | Male only | False | 0.2246 | 0.224 | 0.2457 | 0.2468 | 25647 | 19931 | 187758 | 162098 | -0.00323 | -0.04624 | -0.02818 | -0.02696 | 0.009808 | 0.01081 |
| MONOAB | rs735765 | rs1150692 | G | A | 6 | ZNF192P2 | Male only | False | 0.2244 | 0.2236 | 0.2455 | 0.2467 | 25647 | 19931 | 187758 | 162098 | -0.003163 | -0.04653 | -0.0283 | -0.02704 | 0.00982 | 0.01083 |
| MONOAB | rs735765 | rs1150693 | C | T | 6 | ZNF192P2 | Male only | False | 0.2254 | 0.2247 | 0.2463 | 0.2474 | 25647 | 19931 | 187758 | 162098 | -0.003011 | -0.04953 | -0.0286 | -0.02684 | 0.009809 | 0.0108 |
| MONOAB | rs735765 | rs13197633 | A | G | 6 | ZNF192P2 | Male only | False | 0.0885 | 0.09018 | 0.1163 | 0.1167 | 25647 | 19931 | 187758 | 162098 | -0.02323 | -0.08284 | -0.06066 | -0.05819 | 0.0145 | 0.01584 |
| MONOAB | rs735765 | rs35001169 | G | T | 6 | ZNF192P2 | Male only | False | 0.08865 | 0.09056 | 0.1165 | 0.117 | 25647 | 19931 | 187758 | 162098 | -0.0244 | -0.07939 | -0.0607 | -0.05893 | 0.01449 | 0.0158 |
| MONOAB | rs735765 | rs9366717 | T | C | 6 | ZNF192P2 | Male only | False | 0.1266 | 0.1278 | 0.1523 | 0.1532 | 25647 | 19931 | 187758 | 162098 | -0.007944 | -0.06908 | -0.04587 | -0.04276 | 0.01237 | 0.01351 |
| MONOAB | rs735765 | rs35656932 | T | G | 6 | ZNF192P2 | Male only | False | 0.08873 | 0.09055 | 0.1169 | 0.1174 | 25647 | 19931 | 187758 | 162098 | -0.02431 | -0.0795 | -0.06057 | -0.05884 | 0.01448 | 0.01578 |
| MONOAB | rs735765 | rs34243448 | A | C | 6 | ZNF192P2 | Male only | False | 0.08873 | 0.09055 | 0.1169 | 0.1174 | 25647 | 19931 | 187758 | 162098 | -0.02431 | -0.0795 | -0.0606 | -0.05883 | 0.01448 | 0.01578 |
| MONOAB | rs735765 | rs34787248 | T | C | 6 | ZNF192P2 | Male only | False | 0.1744 | 0.1741 | 0.1949 | 0.196 | 25647 | 19931 | 187758 | 162098 | -0.004271 | -0.04849 | -0.03694 | -0.03226 | 0.01084 | 0.01195 |
| MONOAB | rs735765 | rs9468317 | C | T | 6 | ZNF192P2 | Male only | False | 0.1751 | 0.1751 | 0.1953 | 0.1964 | 25647 | 19931 | 187758 | 162098 | -0.00432 | -0.04924 | -0.03647 | -0.03209 | 0.01081 | 0.01192 |
| MONOAB | rs735765 | rs2299029 | A | G | 6 | ZNF192P2 | Male only | False | 0.1751 | 0.175 | 0.1951 | 0.1961 | 25647 | 19931 | 187758 | 162098 | -0.004341 | -0.04927 | -0.03645 | -0.0321 | 0.01081 | 0.01192 |
| MONOAB | rs735765 | rs11967137 | G | A | 6 | ZNF192P2 | Male only | False | 0.1756 | 0.1756 | 0.1951 | 0.1962 | 25647 | 19931 | 187758 | 162098 | -0.004772 | -0.04994 | -0.03659 | -0.03207 | 0.0108 | 0.01191 |
| MONOAB | rs735765 | rs13204012 | A | G | 6 | ZNF192P2 | Male only | False | 0.0887 | 0.09051 | 0.1168 | 0.1173 | 25647 | 19931 | 187758 | 162098 | -0.02499 | -0.07961 | -0.06075 | -0.05874 | 0.01448 | 0.01578 |
| MONOAB | rs735765 | rs13205211 | G | T | 6 | ZNF192P2 | Male only | False | 0.0887 | 0.09052 | 0.1168 | 0.1173 | 25647 | 19931 | 187758 | 162098 | -0.02499 | -0.07959 | -0.06077 | -0.05873 | 0.01448 | 0.01578 |
| MONOAB | rs735765 | rs9380069 | G | A | 6 | ZNF192P2 | Male only | False | 0.1301 | 0.1317 | 0.1543 | 0.1552 | 25647 | 19931 | 187758 | 162098 | -0.008899 | -0.06868 | -0.04541 | -0.04201 | 0.01222 | 0.01336 |
| MONOAB | rs735765 | rs9393909 | T | C | 6 | ZNF192P2 | Male only | False | 0.1751 | 0.1751 | 0.1949 | 0.1961 | 25647 | 19931 | 187758 | 162098 | -0.004299 | -0.04933 | -0.03682 | -0.03178 | 0.01081 | 0.01192 |
| MONOAB | rs735765 | rs9368563 | G | A | 6 | ZNF192P2 | Male only | False | 0.1751 | 0.1751 | 0.1951 | 0.1961 | 25647 | 19931 | 187758 | 162098 | -0.0043 | -0.04933 | -0.03647 | -0.03207 | 0.01081 | 0.01192 |
| MONOAB | rs735765 | rs9295768 | A | G | 6 | ZNF192P2 | Male only | False | 0.1752 | 0.1751 | 0.1951 | 0.1962 | 25647 | 19931 | 187758 | 162098 | -0.004275 | -0.04931 | -0.03651 | -0.03215 | 0.01081 | 0.01192 |
| MONOAB | rs735765 | rs967005 | T | C | 6 | ZNF192P2 | Male only | False | 0.1751 | 0.1749 | 0.1949 | 0.1959 | 25647 | 19931 | 187758 | 162098 | -0.004485 | -0.04916 | -0.03683 | -0.0319 | 0.01081 | 0.01192 |
| MONOAB | rs735765 | rs17720293 | T | C | 6 | ZNF192P2 | Male only | False | 0.1079 | 0.1098 | 0.1362 | 0.1365 | 25647 | 19931 | 187758 | 162098 | -0.02476 | -0.07427 | -0.05465 | -0.05222 | 0.01325 | 0.01448 |
| MONOAB | rs735765 | rs13200462 | C | A | 6 | ZNF192P2 | Male only | False | 0.1401 | 0.1406 | 0.1629 | 0.1635 | 25647 | 19931 | 187758 | 162098 | -0.01625 | -0.06722 | -0.04397 | -0.04001 | 0.01184 | 0.01302 |

| trait | lead_snp | ID | ALLELE1 | ALLELE0 | CHR | Gene_Symbol | independent_loci_in_sex_P1e2 | in_embl_any_or_gene | A1FREQ_VUMC_female | A1FREQ_VUMC_male | A1FREQ_UKB_female | A1FREQ_UKB_male | N_VUMC_female | N_VUMC_male | N_UKB_female | N_UKB_male | BETA_VUMC_female | BETA_VUMC_male | BETA_UKB_female | BETA_UKB_male | SE_VUMC_female | SE_VUMC_male |
|---|---|---|---|---|---|---|---|---|---|---|---|---|---|---|---|---|---|---|---|---|---|---|
| MONOAB | rs735765 | rs1736904 | A | G | 6 | ZNF192P2 | Male only | False | 0.1937 | 0.1921 | 0.2078 | 0.209 | 25647 | 19931 | 187758 | 162098 | -0.006707 | -0.04899 | -0.03367 | -0.02942 | 0.01039 | 0.01149 |
| MONOAB | rs735765 | rs9986596 | A | G | 6 | ZNF192P2 | Male only | False | 0.1401 | 0.1406 | 0.1629 | 0.1635 | 25647 | 19931 | 187758 | 162098 | -0.01626 | -0.06722 | -0.04398 | -0.03999 | 0.01184 | 0.01302 |
| MONOAB | rs735765 | rs1679732 | G | A | 6 | ZNF192P2 | Male only | False | 0.1425 | 0.1428 | 0.1639 | 0.1645 | 25647 | 19931 | 187758 | 162098 | -0.01537 | -0.06889 | -0.04369 | -0.03947 | 0.01179 | 0.01297 |
| MONOAB | rs735765 | rs10456362 | A | G | 6 | ZNF192P2 | Male only | False | 0.1404 | 0.1407 | 0.1625 | 0.1633 | 25647 | 19931 | 187758 | 162098 | -0.01662 | -0.07223 | -0.04334 | -0.03986 | 0.01183 | 0.01301 |
| MONOAB | rs735765 | rs1679709 | A | G | 6 | ZNF192P2 | Male only | False | 0.1425 | 0.1429 | 0.164 | 0.1647 | 25647 | 19931 | 187758 | 162098 | -0.0152 | -0.06928 | -0.04383 | -0.03954 | 0.01179 | 0.01297 |
| MONOAB | rs735765 | rs1778511 | G | A | 6 | ZNF192P2 | Male only | False | 0.1425 | 0.1428 | 0.1641 | 0.1647 | 25647 | 19931 | 187758 | 162098 | -0.01514 | -0.06926 | -0.0439 | -0.03938 | 0.01179 | 0.01297 |
| MONOAB | rs735765 | rs1778508 | C | T | 6 | ZNF192P2 | Male only | False | 0.1417 | 0.1421 | 0.1641 | 0.1647 | 25647 | 19931 | 187758 | 162098 | -0.01515 | -0.06908 | -0.04397 | -0.0393 | 0.01178 | 0.01296 |
| MONOAB | rs735765 | rs2799077 | T | C | 6 | ZNF192P2 | Male only | False | 0.1418 | 0.1421 | 0.1647 | 0.1652 | 25647 | 19931 | 187758 | 162098 | -0.01533 | -0.06915 | -0.04368 | -0.03921 | 0.01178 | 0.01295 |
| MONOAB | rs735765 | rs2799079 | C | A | 6 | ZNF192P2 | Male only | False | 0.1417 | 0.1421 | 0.1643 | 0.1649 | 25647 | 19931 | 187758 | 162098 | -0.01533 | -0.06917 | -0.04368 | -0.03948 | 0.01178 | 0.01296 |
| MONOAB | rs735765 | rs67998226 | C | T | 6 | ZNF192P2 | Male only | False | 0.08993 | 0.09197 | 0.1184 | 0.1187 | 25647 | 19931 | 187758 | 162098 | -0.02343 | -0.07576 | -0.06124 | -0.05821 | 0.01438 | 0.01566 |
| MONOAB | rs735765 | rs11965538 | A | G | 6 | ZNF192P2 | Male only | False | 0.1418 | 0.1421 | 0.1642 | 0.1648 | 25647 | 19931 | 187758 | 162098 | -0.01537 | -0.06915 | -0.04366 | -0.03953 | 0.01178 | 0.01295 |
| MONOAB | rs735765 | rs2743555 | A | G | 6 | ZNF192P2 | Male only | False | 0.1417 | 0.1421 | 0.1641 | 0.1647 | 25647 | 19931 | 187758 | 162098 | -0.01533 | -0.06916 | -0.0438 | -0.0395 | 0.01178 | 0.01296 |
| MONOAB | rs735765 | rs1419183 | C | A | 6 | ZNF192P2 | Male only | False | 0.1418 | 0.1421 | 0.1665 | 0.1671 | 25647 | 19931 | 187758 | 162098 | -0.01508 | -0.06895 | -0.04443 | -0.03952 | 0.01178 | 0.01295 |
| MONOAB | rs735765 | rs34878803 | C | T | 6 | ZNF192P2 | Male only | False | 0.08991 | 0.09199 | 0.1183 | 0.1187 | 25647 | 19931 | 187758 | 162098 | -0.02346 | -0.07607 | -0.06127 | -0.0584 | 0.01438 | 0.01566 |
| MONOAB | rs735765 | rs34396849 | C | A | 6 | ZNF192P2 | Male only | False | 0.0897 | 0.09174 | 0.1183 | 0.1186 | 25647 | 19931 | 187758 | 162098 | -0.02362 | -0.07635 | -0.06121 | -0.05839 | 0.01441 | 0.01569 |
| MONOAB | rs735765 | rs6901575 | A | G | 6 | ZNF192P2 | Male only | False | 0.1418 | 0.1421 | 0.164 | 0.1646 | 25647 | 19931 | 187758 | 162098 | -0.01567 | -0.06935 | -0.04407 | -0.03989 | 0.01178 | 0.01297 |
| MONOAB | rs735765 | rs13211507 | C | T | 6 | ZNF192P2 | Male only | False | 0.08994 | 0.09207 | 0.1186 | 0.1189 | 25647 | 19931 | 187758 | 162098 | -0.02318 | -0.07663 | -0.06089 | -0.05853 | 0.01438 | 0.01565 |
| MONOAB | rs735765 | rs34691223 | A | C | 6 | ZNF192P2 | Male only | False | 0.0899 | 0.09206 | 0.1184 | 0.1187 | 25647 | 19931 | 187758 | 162098 | -0.02283 | -0.07666 | -0.06134 | -0.05848 | 0.01439 | 0.01565 |
| MONOAB | rs735765 | rs6905391 | A | G | 6 | ZNF192P2 | Male only | False | 0.144 | 0.1441 | 0.164 | 0.1646 | 25647 | 19931 | 187758 | 162098 | -0.01366 | -0.06523 | -0.04098 | -0.03776 | 0.01168 | 0.01289 |
| MONOAB | rs735765 | rs13207345 | T | C | 6 | ZNF192P2 | Male only | False | 0.08877 | 0.09081 | 0.1171 | 0.1175 | 25647 | 19931 | 187758 | 162098 | -0.02477 | -0.08218 | -0.06104 | -0.05892 | 0.01448 | 0.01576 |
| MONOAB | rs735765 | rs33932084 | G | A | 6 | ZNF192P2 | Male only | False | 0.08985 | 0.09195 | 0.1179 | 0.1183 | 25647 | 19931 | 187758 | 162098 | -0.02385 | -0.08159 | -0.06069 | -0.05808 | 0.0144 | 0.01567 |
| MONOAB | rs735765 | rs56189111 | A | C | 6 | ZNF192P2 | Male only | False | 0.08879 | 0.09083 | 0.1171 | 0.1175 | 25647 | 19931 | 187758 | 162098 | -0.02448 | -0.08223 | -0.06104 | -0.05877 | 0.01447 | 0.01575 |
| MONOAB | rs735765 | rs13191786 | G | T | 6 | ZNF192P2 | Male only | False | 0.08879 | 0.09083 | 0.1171 | 0.1175 | 25647 | 19931 | 187758 | 162098 | -0.02448 | -0.08223 | -0.061 | -0.05881 | 0.01447 | 0.01575 |
| MONOAB | rs735765 | rs34950484 | A | G | 6 | ZNF192P2 | Male only | False | 0.08998 | 0.09218 | 0.1185 | 0.1188 | 25647 | 19931 | 187758 | 162098 | -0.02216 | -0.07704 | -0.06128 | -0.05832 | 0.01438 | 0.01565 |
| MONOAB | rs735765 | rs35072899 | T | C | 6 | ZNF192P2 | Male only | False | 0.08879 | 0.09082 | 0.1171 | 0.1175 | 25647 | 19931 | 187758 | 162098 | -0.0245 | -0.08223 | -0.061 | -0.05881 | 0.01447 | 0.01575 |
| MONOAB | rs735765 | rs34661125 | A | G | 6 | ZNF192P2 | Male only | False | 0.08998 | 0.09218 | 0.1185 | 0.1188 | 25647 | 19931 | 187758 | 162098 | -0.02216 | -0.07704 | -0.06129 | -0.05832 | 0.01438 | 0.01565 |
| MONOAB | rs735765 | rs13190888 | A | G | 6 | ZNF192P2 | Male only | False | 0.08999 | 0.0922 | 0.1185 | 0.1188 | 25647 | 19931 | 187758 | 162098 | -0.02216 | -0.07706 | -0.06127 | -0.05838 | 0.01438 | 0.01565 |
| MONOAB | rs735765 | rs853685 | T | C | 6 | ZNF192P2 | Male only | False | 0.1445 | 0.145 | 0.1647 | 0.1653 | 25647 | 19931 | 187758 | 162098 | -0.01381 | -0.06359 | -0.04085 | -0.03738 | 0.01165 | 0.01285 |
| MONOAB | rs735765 | rs56075693 | G | T | 6 | ZNF192P2 | Male only | False | 0.0888 | 0.09084 | 0.1171 | 0.1175 | 25647 | 19931 | 187758 | 162098 | -0.02456 | -0.08222 | -0.06095 | -0.05889 | 0.01447 | 0.01575 |

| trait | lead_snp | ID | ALLELE1 | ALLELE0 | CHR | Gene_Symbol | independent_loci_in_sex_P1e2 | in_embl_any_or_gene | A1FREQ_VUMC_female | A1FREQ_VUMC_male | A1FREQ_UKB_female | A1FREQ_UKB_male | N_VUMC_female | N_VUMC_male | N_UKB_female | N_UKB_male | BETA_VUMC_female | BETA_VUMC_male | BETA_UKB_female | BETA_UKB_male | SE_VUMC_female | SE_VUMC_male |
|---|---|---|---|---|---|---|---|---|---|---|---|---|---|---|---|---|---|---|---|---|---|---|
| MONOAB | rs735765 | rs34218844 | C | T | 6 | ZNF192P2 | Male only | False | 0.0888 | 0.09084 | 0.1171 | 0.1175 | 25647 | 19931 | 187758 | 162098 | -0.02456 | -0.08222 | -0.06094 | -0.0589 | 0.01447 | 0.01575 |
| trait | lead_snp | ID | ALLELE1 | ALLELE0 | CHR | Gene_Symbol | independent_loci_in_sex_P1e2 | in_embl_any_or_gene | A1FREQ_VUMC_female | A1FREQ_VUMC_male | A1FREQ_UKB_female | A1FREQ_UKB_male | N_VUMC_female | N_VUMC_male | N_UKB_female | N_UKB_male | BETA_VUMC_female | BETA_VUMC_male | BETA_UKB_female | BETA_UKB_male | SE_VUMC_female | SE_VUMC_male |
| MONOAB | rs735765 | rs853682 | C | T | 6 | ZNF192P2 | Male only | False | 0.1445 | 0.145 | 0.1646 | 0.1651 | 25647 | 19931 | 187758 | 162098 | -0.01383 | -0.06387 | -0.04112 | -0.03742 | 0.01165 | 0.01286 |
| MONOAB | rs735765 | rs853681 | A | C | 6 | ZNF192P2 | Male only | False | 0.1446 | 0.145 | 0.1646 | 0.1651 | 25647 | 19931 | 187758 | 162098 | -0.01384 | -0.06388 | -0.04117 | -0.03748 | 0.01165 | 0.01286 |
| MONOAB | rs735765 | rs853679 | A | C | 6 | ZNF192P2 | Male only | False | 0.1445 | 0.145 | 0.1645 | 0.1651 | 25647 | 19931 | 187758 | 162098 | -0.01384 | -0.06387 | -0.0412 | -0.03751 | 0.01165 | 0.01286 |
| MONOAB | rs735765 | rs853676 | T | C | 6 | ZNF192P2 | Male only | False | 0.1453 | 0.1458 | 0.1651 | 0.1657 | 25647 | 19931 | 187758 | 162098 | -0.01404 | -0.06297 | -0.04151 | -0.03727 | 0.01163 | 0.01283 |
| MONOAB | rs735765 | rs7752448 | G | A | 6 | ZNF192P2 | Male only | False | 0.1044 | 0.1062 | 0.1284 | 0.1283 | 25647 | 19931 | 187758 | 162098 | -0.01739 | -0.07132 | -0.05416 | -0.05266 | 0.01345 | 0.01472 |
| MONOAB | rs735765 | rs67340775 | C | A | 6 | ZNF192P2 | Male only | False | 0.09006 | 0.09197 | 0.1187 | 0.1191 | 25647 | 19931 | 187758 | 162098 | -0.02358 | -0.07827 | -0.06122 | -0.05873 | 0.01437 | 0.01566 |
| MONOAB | rs735765 | rs35030260 | A | G | 6 | ZNF192P2 | Male only | False | 0.08898 | 0.09091 | 0.1174 | 0.1179 | 25647 | 19931 | 187758 | 162098 | -0.02577 | -0.08127 | -0.06093 | -0.0593 | 0.01445 | 0.01575 |
| MONOAB | rs735765 | rs13217619 | C | T | 6 | ZNF192P2 | Male only | False | 0.08898 | 0.09091 | 0.1174 | 0.1179 | 25647 | 19931 | 187758 | 162098 | -0.02577 | -0.08127 | -0.06096 | -0.0593 | 0.01446 | 0.01575 |
| MONOAB | rs735765 | rs35016036 | T | C | 6 | ZNF192P2 | Male only | False | 0.08844 | 0.09058 | 0.1163 | 0.1168 | 25647 | 19931 | 187758 | 162098 | -0.02597 | -0.08079 | -0.06121 | -0.06009 | 0.0145 | 0.0158 |
| MONOAB | rs735765 | rs35744819 | T | G | 6 | ZNF192P2 | Male only | False | 0.08846 | 0.09059 | 0.1163 | 0.1168 | 25647 | 19931 | 187758 | 162098 | -0.02581 | -0.08094 | -0.06126 | -0.06019 | 0.0145 | 0.0158 |
| MONOAB | rs735765 | rs13198809 | A | G | 6 | ZNF192P2 | Male only | False | 0.08881 | 0.09072 | 0.1168 | 0.1173 | 25647 | 19931 | 187758 | 162098 | -0.02493 | -0.0821 | -0.06107 | -0.06131 | 0.01447 | 0.0158 |
| MONOAB | rs735765 | rs35353359 | T | C | 6 | ZNF192P2 | Male only | False | 0.08882 | 0.09072 | 0.1168 | 0.1174 | 25647 | 19931 | 187758 | 162098 | -0.02493 | -0.0821 | -0.06105 | -0.06147 | 0.01447 | 0.0158 |
| MONOAB | rs735765 | rs13214023 | A | G | 6 | ZNF192P2 | Male only | False | 0.08877 | 0.09065 | 0.1169 | 0.1175 | 25647 | 19931 | 187758 | 162098 | -0.02448 | -0.0828 | -0.06082 | -0.06159 | 0.01447 | 0.0158 |
| MONOAB | rs735765 | rs35017208 | T | C | 6 | ZNF192P2 | Male only | False | 0.08904 | 0.0909 | 0.1171 | 0.1177 | 25647 | 19931 | 187758 | 162098 | -0.02387 | -0.08116 | -0.06046 | -0.06149 | 0.01444 | 0.01578 |
| MONOAB | rs735765 | rs13213152 | G | A | 6 | ZNF192P2 | Male only | False | 0.08882 | 0.09064 | 0.1169 | 0.1175 | 25647 | 19931 | 187758 | 162098 | -0.02476 | -0.08275 | -0.06072 | -0.06163 | 0.01446 | 0.0158 |
| MONOAB | rs735765 | rs36092177 | T | C | 6 | ZNF192P2 | Male only | False | 0.08903 | 0.09091 | 0.1171 | 0.1177 | 25647 | 19931 | 187758 | 162098 | -0.0239 | -0.08121 | -0.06044 | -0.0615 | 0.01444 | 0.01578 |
| MONOAB | rs735765 | rs2232429 | G | T | 6 | ZNF192P2 | Male only | False | 0.08903 | 0.09091 | 0.1171 | 0.1177 | 25647 | 19931 | 187758 | 162098 | -0.0239 | -0.08121 | -0.06042 | -0.06151 | 0.01444 | 0.01578 |
| MONOAB | rs735765 | rs34546986 | A | G | 6 | ZNF192P2 | Male only | False | 0.0888 | 0.09064 | 0.1169 | 0.1175 | 25647 | 19931 | 187758 | 162098 | -0.0248 | -0.08271 | -0.06051 | -0.06174 | 0.01446 | 0.0158 |
| MONOAB | rs735765 | rs34871267 | A | G | 6 | ZNF192P2 | Male only | False | 0.0888 | 0.09064 | 0.1169 | 0.1175 | 25647 | 19931 | 187758 | 162098 | -0.0248 | -0.08271 | -0.0605 | -0.06174 | 0.01446 | 0.0158 |
| MONOAB | rs735765 | rs2232423 | G | A | 6 | ZNF192P2 | Male only | False | 0.08905 | 0.09094 | 0.1171 | 0.1177 | 25647 | 19931 | 187758 | 162098 | -0.02394 | -0.08115 | -0.06024 | -0.06158 | 0.01444 | 0.01577 |
| MONOAB | rs735765 | rs13201681 | T | C | 6 | ZNF192P2 | Male only | False | 0.0888 | 0.09065 | 0.117 | 0.1176 | 25647 | 19931 | 187758 | 162098 | -0.02466 | -0.08265 | -0.0603 | -0.06201 | 0.01446 | 0.0158 |
| MONOAB | rs204893 | rs2856448 | G | A | 6 | ATF6B | Male only | False | 0.4863 | 0.4862 | 0.4609 | 0.46 | 25647 | 19931 | 187758 | 162098 | 0.01396 | 0.04899 | 0.03656 | 0.0336 | 0.008227 | 0.009109 |
| MONOAB | rs204893 | rs6937318 | T | C | 6 | ATF6B | Male only | False | 0.4716 | 0.4733 | 0.4962 | 0.4975 | 25647 | 19931 | 187758 | 162098 | -0.01851 | -0.0426 | -0.03463 | -0.03202 | 0.00823 | 0.009093 |
| MONOAB | rs204893 | rs204886 | T | C | 6 | ATF6B | Male only | False | 0.4716 | 0.4732 | 0.4962 | 0.4975 | 25647 | 19931 | 187758 | 162098 | -0.01863 | -0.04259 | -0.03464 | -0.03208 | 0.008228 | 0.009092 |
| MONOAB | rs204893 | rs204883 | A | G | 6 | ATF6B | Male only | False | 0.3983 | 0.3983 | 0.3805 | 0.3799 | 25647 | 19931 | 187758 | 162098 | 0.01389 | 0.04468 | 0.02616 | 0.02468 | 0.008417 | 0.009318 |
| MONOAB | rs204893 | rs185819 | C | T | 6 | ATF6B | Male only | False | 0.4881 | 0.487 | 0.4627 | 0.462 | 25647 | 19931 | 187758 | 162098 | 0.01523 | 0.05057 | 0.03665 | 0.03314 | 0.008243 | 0.009124 |

| trait | lead_snp | ID | ALLELE1 | ALLELE0 | CHR | Gene_Symbol | independent_loci_in_sex_P1e2 | in_embl_any_or_gene | A1FREQ_VUMC_female | A1FREQ_VUMC_male | A1FREQ_UKB_female | A1FREQ_UKB_male | N_VUMC_female | N_VUMC_male | N_UKB_female | N_UKB_male | BETA_VUMC_female | BETA_VUMC_male | BETA_UKB_female | BETA_UKB_male | SE_VUMC_female | SE_VUMC_male |
|---|---|---|---|---|---|---|---|---|---|---|---|---|---|---|---|---|---|---|---|---|---|---|
| MONOAB | rs204893 | rs4711286 | C | T | 6 | ATF6B | Male only | False | 0.3976 | 0.3978 | 0.3797 | 0.3793 | 25647 | 19931 | 187758 | 162098 | 0.01461 | 0.04292 | 0.0258 | 0.02505 | 0.008432 | 0.009332 |
| MONOAB | rs204893 | rs429150 | C | T | 6 | ATF6B | Male only | False | 0.466 | 0.4634 | 0.442 | 0.4391 | 25647 | 19931 | 187758 | 162098 | 0.01538 | 0.04991 | 0.0356 | 0.03031 | 0.008242 | 0.009142 |
| MONOAB | rs204893 | rs2269426 | A | G | 6 | ATF6B | Male only | False | 0.3713 | 0.3705 | 0.3592 | 0.3572 | 25647 | 19931 | 187758 | 162098 | 0.02148 | 0.04517 | 0.02871 | 0.02726 | 0.008517 | 0.009434 |
| MONOAB | rs204893 | rs204893 | C | T | 6 | ATF6B | Male only | False | 0.4549 | 0.4539 | 0.4304 | 0.4292 | 25647 | 19931 | 187758 | 162098 | 0.01838 | 0.05415 | 0.03965 | 0.03438 | 0.00827 | 0.009174 |
| MONORE | rs6748985 | rs12474024 | C | T | 2 | RNA5SP106 | Male only | True | 0.2842 | 0.278 | 0.2818 | 0.2798 | 25672 | 19994 | 187760 | 162101 | 0.01065 | 0.04669 | 0.01854 | 0.01072 | 0.009476 | 0.01057 |
| MONORE | rs6748985 | rs1528837 | G | A | 2 | RNA5SP106 | Male only | True | 0.2842 | 0.278 | 0.2817 | 0.2798 | 25672 | 19994 | 187760 | 162101 | 0.01071 | 0.04677 | 0.01866 | 0.0108 | 0.009476 | 0.01057 |
| MONORE | rs6748985 | rs12467429 | G | A | 2 | RNA5SP106 | Male only | True | 0.2841 | 0.278 | 0.2817 | 0.2797 | 25672 | 19994 | 187760 | 162101 | 0.01072 | 0.04682 | 0.01869 | 0.01077 | 0.009474 | 0.01057 |
| MONORE | rs6748985 | rs12464985 | C | T | 2 | RNA5SP106 | Male only | True | 0.2841 | 0.2779 | 0.2817 | 0.2797 | 25672 | 19994 | 187760 | 162101 | 0.01072 | 0.04682 | 0.01868 | 0.01079 | 0.009474 | 0.01057 |
| MONORE | rs6748985 | rs35905220 | G | A | 2 | RNA5SP106 | Male only | True | 0.3073 | 0.3001 | 0.3009 | 0.2985 | 25672 | 19994 | 187760 | 162101 | 0.01112 | 0.04484 | 0.01739 | 0.01174 | 0.009238 | 0.01027 |
| MONORE | rs6748985 | rs35826590 | A | C | 2 | RNA5SP106 | Male only | True | 0.2835 | 0.2771 | 0.2806 | 0.2786 | 25672 | 19994 | 187760 | 162101 | 0.01077 | 0.0474 | 0.0186 | 0.01103 | 0.009443 | 0.01054 |
| MONORE | rs6748985 | rs6748985 | T | C | 2 | RNA5SP106 | Male only | True | 0.4214 | 0.4148 | 0.4127 | 0.4114 | 25672 | 19994 | 187760 | 162101 | 0.01318 | 0.0525 | 0.01419 | 0.006962 | 0.008581 | 0.009601 |
| MONORE | rs6748985 | rs16827722 | C | A | 2 | RNA5SP106 | Male only | True | 0.3081 | 0.3009 | 0.302 | 0.2996 | 25672 | 19994 | 187760 | 162101 | 0.01099 | 0.04458 | 0.01755 | 0.01162 | 0.00922 | 0.01025 |
| MONORE | rs6748985 | rs12464921 | T | C | 2 | RNA5SP106 | Male only | True | 0.2842 | 0.2778 | 0.2817 | 0.2797 | 25672 | 19994 | 187760 | 162101 | 0.01066 | 0.0471 | 0.01878 | 0.01091 | 0.009423 | 0.01052 |
| MONORE | rs6748985 | rs12996302 | T | C | 2 | RNA5SP106 | Male only | True | 0.2856 | 0.2791 | 0.2817 | 0.2798 | 25672 | 19994 | 187760 | 162101 | 0.01062 | 0.04697 | 0.01877 | 0.01072 | 0.009379 | 0.01047 |
| MONORE | rs6748985 | rs12996303 | T | C | 2 | RNA5SP106 | Male only | True | 0.2842 | 0.2778 | 0.2817 | 0.2797 | 25672 | 19994 | 187760 | 162101 | 0.01061 | 0.0471 | 0.01878 | 0.01091 | 0.009423 | 0.01052 |
| MONORE | rs6748985 | rs12467064 | T | C | 2 | RNA5SP106 | Male only | True | 0.2842 | 0.2778 | 0.2817 | 0.2797 | 25672 | 19994 | 187760 | 162101 | 0.01064 | 0.04711 | 0.01883 | 0.01089 | 0.009422 | 0.01052 |
| MONORE | rs6748985 | rs6721628 | C | T | 2 | RNA5SP106 | Male only | True | 0.3081 | 0.3009 | 0.302 | 0.2996 | 25672 | 19994 | 187760 | 162101 | 0.011 | 0.04457 | 0.01761 | 0.01161 | 0.00922 | 0.01025 |
| MONORE | rs6748985 | rs6706259 | T | C | 2 | RNA5SP106 | Male only | True | 0.3081 | 0.3009 | 0.302 | 0.2996 | 25672 | 19994 | 187760 | 162101 | 0.011 | 0.04457 | 0.01761 | 0.0116 | 0.00922 | 0.01025 |
| MONORE | rs6748985 | rs1113958 | A | G | 2 | RNA5SP106 | Male only | True | 0.2842 | 0.2779 | 0.2817 | 0.2798 | 25672 | 19994 | 187760 | 162101 | 0.01065 | 0.04717 | 0.01884 | 0.01096 | 0.009423 | 0.01052 |
| MONORE | rs6748985 | rs12464481 | T | C | 2 | RNA5SP106 | Male only | True | 0.2842 | 0.2778 | 0.2817 | 0.2797 | 25672 | 19994 | 187760 | 162101 | 0.01064 | 0.04711 | 0.01883 | 0.01089 | 0.009422 | 0.01052 |
| MONORE | rs6748985 | rs12691762 | T | C | 2 | RNA5SP106 | Male only | True | 0.2841 | 0.2777 | 0.2817 | 0.2797 | 25672 | 19994 | 187760 | 162101 | 0.01074 | 0.04715 | 0.0188 | 0.0109 | 0.009426 | 0.01052 |
| MONORE | rs6748985 | rs9653190 | T | C | 2 | RNA5SP106 | Male only | True | 0.3082 | 0.3012 | 0.302 | 0.2997 | 25672 | 19994 | 187760 | 162101 | 0.0111 | 0.04462 | 0.01754 | 0.01157 | 0.009219 | 0.01025 |
| MONORE | rs6748985 | rs13004499 | C | T | 2 | RNA5SP106 | Male only | True | 0.2996 | 0.2934 | 0.2947 | 0.2925 | 25672 | 19994 | 187760 | 162101 | 0.00998 | 0.04607 | 0.01871 | 0.01209 | 0.009293 | 0.01034 |
| MONORE | rs6748985 | rs4662292 | T | C | 2 | RNA5SP106 | Male only | True | 0.2996 | 0.2934 | 0.2947 | 0.2925 | 25672 | 19994 | 187760 | 162101 | 0.009981 | 0.04607 | 0.01872 | 0.01209 | 0.009293 | 0.01034 |
| MONORE | rs6748985 | rs1406447 | G | A | 2 | RNA5SP106 | Male only | True | 0.3561 | 0.3492 | 0.3511 | 0.3492 | 25672 | 19994 | 187760 | 162101 | 0.01586 | 0.05048 | 0.01787 | 0.009362 | 0.008895 | 0.00993 |
| MONORE | rs6748985 | rs4662548 | C | A | 2 | RNA5SP106 | Male only | True | 0.3561 | 0.3492 | 0.3511 | 0.3492 | 25672 | 19994 | 187760 | 162101 | 0.01582 | 0.05019 | 0.01787 | 0.009367 | 0.008895 | 0.00993 |
| MONORE | rs6748985 | rs12691766 | G | A | 2 | RNA5SP106 | Male only | True | 0.2996 | 0.2934 | 0.2947 | 0.2925 | 25672 | 19994 | 187760 | 162101 | 0.009897 | 0.046 | 0.01872 | 0.01209 | 0.009294 | 0.01034 |
| MONORE | rs6748985 | rs7581116 | A | C | 2 | RNA5SP106 | Male only | True | 0.3561 | 0.3492 | 0.3511 | 0.3491 | 25672 | 19994 | 187760 | 162101 | 0.01586 | 0.05016 | 0.01788 | 0.009355 | 0.008895 | 0.00993 |
| MONORE | rs6748985 | rs12691767 | C | T | 2 | RNA5SP106 | Male only | True | 0.3577 | 0.3508 | 0.3511 | 0.3491 | 25672 | 19994 | 187760 | 162101 | 0.01573 | 0.04993 | 0.01788 | 0.009356 | 0.008855 | 0.009886 |

| trait | lead_snp | ID | ALLELE1 | ALLELE0 | CHR | Gene_Symbol | independent_loci_in_sex_P1e2 | in_embl_any_or_gene | A1FREQ_VUMC_female | A1FREQ_VUMC_male | A1FREQ_UKB_female | A1FREQ_UKB_male | N_VUMC_female | N_VUMC_male | N_UKB_female | N_UKB_male | BETA_VUMC_female | BETA_VUMC_male | BETA_UKB_female | BETA_UKB_male | SE_VUMC_female | SE_VUMC_male |
|---|---|---|---|---|---|---|---|---|---|---|---|---|---|---|---|---|---|---|---|---|---|---|
| MONORE | rs6748985 | rs7595433 | C | T | 2 | RNA5SP106 | Male only | True | 0.3561 | 0.3492 | 0.3511 | 0.3491 | 25672 | 19994 | 187760 | 162101 | 0.01583 | 0.05021 | 0.01785 | 0.009329 | 0.008896 | 0.009931 |
| MONORE | rs6748985 | rs12988144 | A | C | 2 | RNA5SP106 | Male only | True | 0.2995 | 0.2932 | 0.2947 | 0.2925 | 25672 | 19994 | 187760 | 162101 | 0.008847 | 0.04678 | 0.01868 | 0.01205 | 0.009301 | 0.01034 |
| MONORE | rs6748985 | rs13035656 | C | T | 2 | RNA5SP106 | Male only | True | 0.2995 | 0.2932 | 0.2947 | 0.2925 | 25672 | 19994 | 187760 | 162101 | 0.008796 | 0.04675 | 0.01869 | 0.01205 | 0.009301 | 0.01035 |
| MONORE | rs6748985 | rs13022125 | A | G | 2 | RNA5SP106 | Male only | True | 0.3004 | 0.2937 | 0.2955 | 0.2932 | 25672 | 19994 | 187760 | 162101 | 0.008539 | 0.04644 | 0.01855 | 0.0119 | 0.009294 | 0.01034 |
| MONORE | rs6748985 | rs7568718 | G | A | 2 | RNA5SP106 | Male only | True | 0.3021 | 0.2954 | 0.2965 | 0.2941 | 25672 | 19994 | 187760 | 162101 | 0.00788 | 0.04759 | 0.01878 | 0.01196 | 0.00928 | 0.01032 |
| trait | lead_snp | ID | ALLELE1 | ALLELE0 | CHR | Gene_Symbol | independent_loci_in_sex_P1e2 | in_embl_any_or_gene | A1FREQ_VUMC_female | A1FREQ_VUMC_male | A1FREQ_UKB_female | A1FREQ_UKB_male | N_VUMC_female | N_VUMC_male | N_UKB_female | N_UKB_male | BETA_VUMC_female | BETA_VUMC_male | BETA_UKB_female | BETA_UKB_male | SE_VUMC_female | SE_VUMC_male |
| MONORE | rs6748985 | rs4662293 | G | A | 2 | RNA5SP106 | Male only | True | 0.3035 | 0.2968 | 0.2965 | 0.2941 | 25672 | 19994 | 187760 | 162101 | 0.008414 | 0.04723 | 0.01883 | 0.01194 | 0.009235 | 0.01027 |
| MONORE | rs6748985 | rs12468864 | C | T | 2 | RNA5SP106 | Male only | True | 0.302 | 0.2953 | 0.2964 | 0.294 | 25672 | 19994 | 187760 | 162101 | 0.008684 | 0.04733 | 0.01886 | 0.01198 | 0.009282 | 0.01032 |
| MONORE | rs6748985 | rs13026223 | C | T | 2 | RNA5SP106 | Male only | True | 0.3027 | 0.2962 | 0.2962 | 0.2938 | 25672 | 19994 | 187760 | 162101 | 0.008694 | 0.04657 | 0.01881 | 0.01216 | 0.009241 | 0.01027 |
| MONORE | rs6748985 | rs7605932 | T | C | 2 | RNA5SP106 | Male only | True | 0.302 | 0.2954 | 0.2964 | 0.294 | 25672 | 19994 | 187760 | 162101 | 0.008684 | 0.04756 | 0.01885 | 0.01197 | 0.009282 | 0.01032 |
| MONORE | rs6748985 | rs13014274 | A | C | 2 | RNA5SP106 | Male only | True | 0.3011 | 0.2946 | 0.2961 | 0.2938 | 25672 | 19994 | 187760 | 162101 | 0.008731 | 0.04682 | 0.01882 | 0.01214 | 0.009291 | 0.01033 |
| MONORE | rs6748985 | rs1919441 | G | A | 2 | RNA5SP106 | Male only | True | 0.3019 | 0.2953 | 0.2963 | 0.294 | 25672 | 19994 | 187760 | 162101 | 0.008691 | 0.04763 | 0.01887 | 0.01196 | 0.009286 | 0.01032 |
| MONORE | rs6748985 | rs1919440 | G | A | 2 | RNA5SP106 | Male only | True | 0.3009 | 0.2945 | 0.296 | 0.2937 | 25672 | 19994 | 187760 | 162101 | 0.008642 | 0.04709 | 0.01882 | 0.01221 | 0.009295 | 0.01033 |
| MONORE | rs6748985 | rs7564409 | T | C | 2 | RNA5SP106 | Male only | True | 0.3004 | 0.2938 | 0.295 | 0.2928 | 25672 | 19994 | 187760 | 162101 | 0.009363 | 0.04736 | 0.01879 | 0.0121 | 0.009329 | 0.01036 |
| MONORE | rs6748985 | rs7590862 | G | A | 2 | RNA5SP106 | Male only | True | 0.3003 | 0.2937 | 0.2949 | 0.2927 | 25672 | 19994 | 187760 | 162101 | 0.009323 | 0.04726 | 0.01884 | 0.01207 | 0.009331 | 0.01037 |
| MONORE | rs6748985 | rs10181608 | T | C | 2 | RNA5SP106 | Male only | True | 0.3011 | 0.2947 | 0.2961 | 0.2938 | 25672 | 19994 | 187760 | 162101 | 0.00869 | 0.04684 | 0.01886 | 0.01218 | 0.009292 | 0.01033 |
| MONORE | rs6748985 | rs6728992 | C | T | 2 | RNA5SP106 | Male only | True | 0.3019 | 0.2953 | 0.2963 | 0.294 | 25672 | 19994 | 187760 | 162101 | 0.008679 | 0.04762 | 0.01888 | 0.01196 | 0.009286 | 0.01032 |
| MONORE | rs6748985 | rs12465277 | A | G | 2 | RNA5SP106 | Male only | True | 0.3006 | 0.2941 | 0.2957 | 0.2935 | 25672 | 19994 | 187760 | 162101 | 0.008669 | 0.04722 | 0.01892 | 0.0123 | 0.009296 | 0.01033 |
| MONORE | rs6748985 | rs10170800 | T | C | 2 | RNA5SP106 | Male only | True | 0.3005 | 0.294 | 0.2959 | 0.2936 | 25672 | 19994 | 187760 | 162101 | 0.008772 | 0.04725 | 0.01888 | 0.01219 | 0.009297 | 0.01033 |
| MONORE | rs6748985 | rs10183294 | C | T | 2 | RNA5SP106 | Male only | True | 0.3005 | 0.294 | 0.2957 | 0.2934 | 25672 | 19994 | 187760 | 162101 | 0.008777 | 0.04725 | 0.01895 | 0.01233 | 0.009297 | 0.01033 |
| MONORE | rs6748985 | rs10497022 | T | C | 2 | RNA5SP106 | Male only | True | 0.2996 | 0.2933 | 0.2957 | 0.2934 | 25672 | 19994 | 187760 | 162101 | 0.009855 | 0.04937 | 0.01901 | 0.01233 | 0.009354 | 0.0104 |
| MONORE | rs6748985 | rs1528839 | C | A | 2 | RNA5SP106 | Male only | True | 0.3001 | 0.2937 | 0.296 | 0.2937 | 25672 | 19994 | 187760 | 162101 | 0.00992 | 0.04904 | 0.01886 | 0.01214 | 0.009349 | 0.01039 |
| MONORE | rs6748985 | rs10497023 | T | C | 2 | RNA5SP106 | Male only | True | 0.2998 | 0.2935 | 0.2959 | 0.2937 | 25672 | 19994 | 187760 | 162101 | 0.009926 | 0.04882 | 0.01902 | 0.01234 | 0.00935 | 0.01039 |
| MONORE | rs6748985 | rs12988552 | C | A | 2 | RNA5SP106 | Male only | True | 0.2998 | 0.2934 | 0.2958 | 0.2936 | 25672 | 19994 | 187760 | 162101 | 0.01004 | 0.0488 | 0.01905 | 0.01256 | 0.009352 | 0.0104 |
| MONORE | rs6748985 | rs1424953 | G | A | 2 | RNA5SP106 | Male only | True | 0.3022 | 0.2956 | 0.2974 | 0.2951 | 25672 | 19994 | 187760 | 162101 | 0.009421 | 0.0507 | 0.01956 | 0.0123 | 0.009329 | 0.01037 |
| MONORE | rs6748985 | rs1424952 | T | C | 2 | RNA5SP106 | Male only | True | 0.2979 | 0.2919 | 0.2935 | 0.2912 | 25672 | 19994 | 187760 | 162101 | 0.01092 | 0.04801 | 0.01989 | 0.01351 | 0.00937 | 0.01041 |
| MONORE | rs7598148 | rs7579463 | C | A | 2 | MYO3B | Male only | False | 0.4521 | 0.4496 | 0.4459 | 0.445 | 25672 | 19994 | 187760 | 162101 | -0.009151 | 0.04018 | -5.87e-04 | 0.001095 | 0.008519 | 0.009536 |
| MONORE | rs7598148 | rs7561268 | A | C | 2 | MYO3B | Male only | False | 0.4409 | 0.4393 | 0.4351 | 0.4342 | 25672 | 19994 | 187760 | 162101 | -0.008483 | 0.04049 | -7.20e-04 | 0.001065 | 0.008528 | 0.009534 |

| trait | lead_snp | ID | ALLELE1 | ALLELE0 | CHR | Gene_Symbol | independent_loci_in_sex_P1e2 | in_embl_any_or_gene | A1FREQ_VUMC_female | A1FREQ_VUMC_male | A1FREQ_UKB_female | A1FREQ_UKB_male | N_VUMC_female | N_VUMC_male | N_UKB_female | N_UKB_male | BETA_VUMC_female | BETA_VUMC_male | BETA_UKB_female | BETA_UKB_male | SE_VUMC_female | SE_VUMC_male |
|---|---|---|---|---|---|---|---|---|---|---|---|---|---|---|---|---|---|---|---|---|---|---|
| MONORE | rs7598148 | rs7608448 | T | C | 2 | MYO3B | Male only | False | 0.4408 | 0.4392 | 0.4349 | 0.4339 | 25672 | 19994 | 187760 | 162101 | -0.00754 | 0.04094 | -7.05e-04 | 0.00116 | 0.008548 | 0.009563 |
| MONORE | rs7598148 | rs1344940 | C | T | 2 | MYO3B | Male only | False | 0.4408 | 0.4416 | 0.437 | 0.4368 | 25672 | 19994 | 187760 | 162101 | -0.01167 | 0.04357 | 0.001739 | 9.09e-04 | 0.008512 | 0.009526 |
| MONORE | rs7598148 | rs10497368 | A | G | 2 | MYO3B | Male only | False | 0.3866 | 0.39 | 0.3814 | 0.3817 | 25672 | 19994 | 187760 | 162101 | -0.008479 | 0.0403 | 0.002314 | 0.001571 | 0.008659 | 0.009694 |
| MONORE | rs7598148 | rs11885105 | G | A | 2 | MYO3B | Male only | False | 0.3862 | 0.3895 | 0.3816 | 0.3819 | 25672 | 19994 | 187760 | 162101 | -0.008642 | 0.04037 | 0.0022 | 0.001524 | 0.008673 | 0.00971 |
| MONORE | rs7598148 | rs6728783 | G | A | 2 | MYO3B | Male only | False | 0.4276 | 0.4299 | 0.4232 | 0.4233 | 25672 | 19994 | 187760 | 162101 | -0.003055 | 0.04619 | 0.003324 | 8.93e-04 | 0.008594 | 0.009571 |
| MONORE | rs7598148 | rs1075481 | G | A | 2 | MYO3B | Male only | False | 0.4649 | 0.4654 | 0.4626 | 0.4627 | 25672 | 19994 | 187760 | 162101 | -0.009629 | 0.04781 | 0.002755 | 8.00e-05 | 0.008505 | 0.009491 |
| MONORE | rs7598148 | rs10930429 | T | C | 2 | MYO3B | Male only | False | 0.4308 | 0.4323 | 0.4263 | 0.4266 | 25672 | 19994 | 187760 | 162101 | -0.002931 | 0.04738 | 0.003273 | 0.001047 | 0.00855 | 0.009526 |
| MONORE | rs7598148 | rs7598148 | C | T | 2 | MYO3B | Male only | False | 0.4582 | 0.4586 | 0.4569 | 0.4568 | 25672 | 19994 | 187760 | 162101 | -0.008864 | 0.05335 | 0.001933 | 2.28e-05 | 0.00852 | 0.009494 |
| MONORE | rs34273533 | rs7861175 | C | T | 9 | RP11-202G18.1 | Female only | True | 0.1981 | 0.1958 | 0.1998 | 0.1996 | 25672 | 19994 | 187760 | 162101 | -0.05554 | -0.01146 | -0.05 | -0.04515 | 0.01065 | 0.01188 |
| MONORE | rs34273533 | rs13290486 | A | G | 9 | RP11-202G18.1 | Female only | True | 0.1303 | 0.1289 | 0.1321 | 0.1321 | 25672 | 19994 | 187760 | 162101 | -0.06657 | -0.02399 | -0.06275 | -0.05503 | 0.01271 | 0.01419 |
| MONORE | rs34273533 | rs36030865 | T | C | 9 | RP11-202G18.1 | Female only | True | 0.1301 | 0.1284 | 0.1313 | 0.1311 | 25672 | 19994 | 187760 | 162101 | -0.07097 | -0.02455 | -0.06577 | -0.05912 | 0.01258 | 0.01406 |
| MONORE | rs34273533 | rs34273533 | T | C | 9 | RP11-202G18.1 | Female only | True | 0.1309 | 0.1286 | 0.1334 | 0.1333 | 25672 | 19994 | 187760 | 162101 | -0.07228 | -0.02467 | -0.07099 | -0.05973 | 0.01268 | 0.01417 |
| MONORE | rs113646878 | rs73656204 | G | A | 9 | RP11-202G18.1 | Female only | True | 0.1089 | 0.1113 | 0.1106 | 0.1099 | 25672 | 19994 | 187760 | 162101 | 0.07351 | 0.02525 | 0.08167 | 0.05935 | 0.01363 | 0.01497 |
| MONORE | rs113646878 | rs12684515 | T | C | 9 | RP11-202G18.1 | Female only | True | 0.1088 | 0.1113 | 0.1106 | 0.1099 | 25672 | 19994 | 187760 | 162101 | 0.07338 | 0.0241 | 0.08172 | 0.05944 | 0.0136 | 0.01493 |
| MONORE | rs113646878 | rs12685721 | A | G | 9 | RP11-202G18.1 | Female only | True | 0.1077 | 0.1104 | 0.1102 | 0.1095 | 25672 | 19994 | 187760 | 162101 | 0.07398 | 0.0252 | 0.08222 | 0.05974 | 0.01365 | 0.01498 |
| MONORE | rs113646878 | rs57955452 | A | G | 9 | RP11-202G18.1 | Female only | True | 0.1083 | 0.1108 | 0.1102 | 0.1096 | 25672 | 19994 | 187760 | 162101 | 0.07509 | 0.02513 | 0.08148 | 0.05986 | 0.01363 | 0.01495 |
| MONORE | rs113646878 | rs7859205 | A | G | 9 | RP11-202G18.1 | Female only | True | 0.1129 | 0.115 | 0.1123 | 0.1117 | 25672 | 19994 | 187760 | 162101 | 0.07231 | 0.03042 | 0.08064 | 0.06022 | 0.01339 | 0.01472 |
| MONORE | rs113646878 | rs12686563 | A | C | 9 | RP11-202G18.1 | Female only | True | 0.1083 | 0.1108 | 0.1102 | 0.1096 | 25672 | 19994 | 187760 | 162101 | 0.0751 | 0.02513 | 0.08148 | 0.05986 | 0.01363 | 0.01495 |
| MONORE | rs113646878 | rs7024184 | T | C | 9 | RP11-202G18.1 | Female only | True | 0.1082 | 0.1107 | 0.1104 | 0.1097 | 25672 | 19994 | 187760 | 162101 | 0.07473 | 0.0253 | 0.08107 | 0.05986 | 0.01362 | 0.01494 |
| MONORE | rs113646878 | rs7861100 | T | C | 9 | RP11-202G18.1 | Female only | True | 0.1081 | 0.1107 | 0.1103 | 0.1096 | 25672 | 19994 | 187760 | 162101 | 0.07479 | 0.02514 | 0.08132 | 0.06003 | 0.0136 | 0.01493 |
| MONORE | rs113646878 | rs57710164 | A | C | 9 | RP11-202G18.1 | Female only | True | 0.1126 | 0.1148 | 0.1124 | 0.1117 | 25672 | 19994 | 187760 | 162101 | 0.07176 | 0.03051 | 0.08046 | 0.0604 | 0.01338 | 0.01471 |
| MONORE | rs113646878 | rs11788404 | A | C | 9 | RP11-202G18.1 | Female only | True | 0.1131 | 0.1152 | 0.1125 | 0.1118 | 25672 | 19994 | 187760 | 162101 | 0.06911 | 0.02935 | 0.0811 | 0.05997 | 0.01335 | 0.01469 |
| MONORE | rs113646878 | rs7041630 | A | G | 9 | RP11-202G18.1 | Female only | True | 0.1091 | 0.1116 | 0.1107 | 0.11 | 25672 | 19994 | 187760 | 162101 | 0.07031 | 0.02425 | 0.08136 | 0.05951 | 0.01357 | 0.01488 |
| MONORE | rs113646878 | rs7022946 | T | C | 9 | RP11-202G18.1 | Female only | True | 0.109 | 0.1113 | 0.1102 | 0.1095 | 25672 | 19994 | 187760 | 162101 | 0.07164 | 0.0253 | 0.0811 | 0.06028 | 0.01357 | 0.01491 |
| MONORE | rs113646878 | rs7023493 | T | C | 9 | RP11-202G18.1 | Female only | True | 0.1086 | 0.1108 | 0.1095 | 0.1088 | 25672 | 19994 | 187760 | 162101 | 0.07075 | 0.02961 | 0.08068 | 0.0605 | 0.01361 | 0.01498 |
| MONORE | rs113646878 | rs7023715 | A | G | 9 | RP11-202G18.1 | Female only | True | 0.1086 | 0.1108 | 0.1095 | 0.1088 | 25672 | 19994 | 187760 | 162101 | 0.07099 | 0.0296 | 0.08067 | 0.06049 | 0.01361 | 0.01498 |
| MONORE | rs113646878 | rs17813616 | G | T | 9 | RP11-202G18.1 | Female only | True | 0.1081 | 0.1109 | 0.1102 | 0.1096 | 25672 | 19994 | 187760 | 162101 | 0.07284 | 0.02827 | 0.08111 | 0.05969 | 0.01362 | 0.01492 |
| MONORE | rs113646878 | rs7850054 | A | G | 9 | RP11-202G18.1 | Female only | True | 0.1089 | 0.1112 | 0.1102 | 0.1095 | 25672 | 19994 | 187760 | 162101 | 0.07224 | 0.0275 | 0.08071 | 0.06012 | 0.01358 | 0.0149 |
| MONORE | rs113646878 | rs11793808 | T | C | 9 | RP11-202G18.1 | Female only | True | 0.1089 | 0.1111 | 0.1102 | 0.1095 | 25672 | 19994 | 187760 | 162101 | 0.07259 | 0.02829 | 0.08089 | 0.06002 | 0.01358 | 0.01491 |

| trait | lead_snp | ID | ALLELE1 | ALLELE0 | CHR | Gene_Symbol | independent_loci_in_sex_P1e2 | in_embl_any_or_gene | A1FREQ_VUMC_female | A1FREQ_VUMC_male | A1FREQ_UKB_female | A1FREQ_UKB_male | N_VUMC_female | N_VUMC_male | N_UKB_female | N_UKB_male | BETA_VUMC_female | BETA_VUMC_male | BETA_UKB_female | BETA_UKB_male | SE_VUMC_female | SE_VUMC_male |
|---|---|---|---|---|---|---|---|---|---|---|---|---|---|---|---|---|---|---|---|---|---|---|
| MONORE | rs113646878 | rs10759494 | G | A | 9 | RP11-202G18.1 | Female only | True | 0.1096 | 0.1117 | 0.1105 | 0.1098 | 25672 | 19994 | 187760 | 162101 | 0.07292 | 0.02852 | 0.08083 | 0.05988 | 0.01354 | 0.01488 |
| MONORE | rs113646878 | rs11795120 | A | C | 9 | RP11-202G18.1 | Female only | True | 0.1062 | 0.109 | 0.1077 | 0.1069 | 25672 | 19994 | 187760 | 162101 | 0.07228 | 0.02858 | 0.08128 | 0.06072 | 0.01376 | 0.01504 |
| MONORE | rs113646878 | rs78161076 | A | G | 9 | RP11-202G18.1 | Female only | True | 0.1057 | 0.1086 | 0.1077 | 0.1069 | 25672 | 19994 | 187760 | 162101 | 0.0743 | 0.0297 | 0.08147 | 0.06057 | 0.01379 | 0.01506 |
| MONORE | rs113646878 | rs76981484 | C | T | 9 | RP11-202G18.1 | Female only | True | 0.1059 | 0.1088 | 0.1077 | 0.107 | 25672 | 19994 | 187760 | 162101 | 0.07483 | 0.0303 | 0.08133 | 0.06036 | 0.01378 | 0.01505 |
| MONORE | rs113646878 | rs11793520 | G | A | 9 | RP11-202G18.1 | Female only | True | 0.1067 | 0.1094 | 0.108 | 0.1072 | 25672 | 19994 | 187760 | 162101 | 0.07342 | 0.02816 | 0.0812 | 0.06021 | 0.01374 | 0.01502 |
| MONORE | rs113646878 | rs113646878 | C | A | 9 | RP11-202G18.1 | Female only | True | 0.06415 | 0.06721 | 0.0651 | 0.06464 | 25672 | 19994 | 187760 | 162101 | 0.1129 | 0.02473 | 0.09346 | 0.07126 | 0.01742 | 0.01886 |
| MONORE | rs4943818 | rs9577112 | T | C | 13 | TPTE2P5 | Female only | False | 0.294 | 0.2908 | 0.2975 | 0.2972 | 25672 | 19994 | 187760 | 162101 | -0.05059 | -0.02525 | -0.03158 | -0.0218 | 0.009283 | 0.01039 |
| MONORE | rs4943818 | rs7982072 | T | C | 13 | TPTE2P5 | Female only | False | 0.2996 | 0.2962 | 0.301 | 0.3004 | 25672 | 19994 | 187760 | 162101 | -0.05128 | -0.02158 | -0.03173 | -0.02168 | 0.009241 | 0.01033 |
| MONORE | rs4943818 | rs9577115 | C | T | 13 | TPTE2P5 | Female only | False | 0.2996 | 0.2962 | 0.301 | 0.3004 | 25672 | 19994 | 187760 | 162101 | -0.05128 | -0.02161 | -0.03171 | -0.02167 | 0.009241 | 0.01033 |

| trait | lead_snp | ID | ALLELE1 | ALLELE0 | CHR | Gene_Symbol | independent_loci_in_sex_P1e2 | in_embl_any_or_gene | A1FREQ_VUMC_female | A1FREQ_VUMC_male | A1FREQ_UKB_female | A1FREQ_UKB_male | N_VUMC_female | N_VUMC_male | N_UKB_female | N_UKB_male | BETA_VUMC_female | BETA_VUMC_male | BETA_UKB_female | BETA_UKB_male | SE_VUMC_female | SE_VUMC_male |
|---|---|---|---|---|---|---|---|---|---|---|---|---|---|---|---|---|---|---|---|---|---|---|
| MONORE | rs4943818 | rs4245401 | T | G | 13 | TPTE2P5 | Female only | False | 0.2996 | 0.2962 | 0.301 | 0.3004 | 25672 | 19994 | 187760 | 162101 | -0.05134 | -0.02165 | -0.03171 | -0.02166 | 0.009241 | 0.01033 |
| MONORE | rs4943818 | rs8000965 | T | C | 13 | TPTE2P5 | Female only | False | 0.2996 | 0.2962 | 0.301 | 0.3004 | 25672 | 19994 | 187760 | 162101 | -0.05133 | -0.02164 | -0.03171 | -0.02167 | 0.009241 | 0.01033 |
| MONORE | rs4943818 | rs4943807 | C | A | 13 | TPTE2P5 | Female only | False | 0.2996 | 0.2962 | 0.3009 | 0.3003 | 25672 | 19994 | 187760 | 162101 | -0.05134 | -0.02171 | -0.03172 | -0.02166 | 0.009241 | 0.01033 |
| MONORE | rs4943818 | rs7990957 | G | A | 13 | TPTE2P5 | Female only | False | 0.294 | 0.2908 | 0.2975 | 0.2971 | 25672 | 19994 | 187760 | 162101 | -0.05058 | -0.02522 | -0.03156 | -0.02176 | 0.009283 | 0.01039 |
| MONORE | rs4943818 | rs7996228 | T | C | 13 | TPTE2P5 | Female only | False | 0.3001 | 0.2966 | 0.301 | 0.3004 | 25672 | 19994 | 187760 | 162101 | -0.05129 | -0.02144 | -0.03167 | -0.0216 | 0.009234 | 0.01032 |
| MONORE | rs4943818 | rs9549270 | T | G | 13 | TPTE2P5 | Female only | False | 0.2932 | 0.2905 | 0.2972 | 0.2968 | 25672 | 19994 | 187760 | 162101 | -0.05029 | -0.02599 | -0.03139 | -0.02175 | 0.009289 | 0.01039 |
| MONORE | rs4943818 | rs4245402 | T | C | 13 | TPTE2P5 | Female only | False | 0.301 | 0.2976 | 0.3009 | 0.3003 | 25672 | 19994 | 187760 | 162101 | -0.05104 | -0.02147 | -0.03174 | -0.02166 | 0.009199 | 0.01028 |
| MONORE | rs4943818 | rs9635107 | C | A | 13 | TPTE2P5 | Female only | False | 0.2996 | 0.2962 | 0.301 | 0.3004 | 25672 | 19994 | 187760 | 162101 | -0.05148 | -0.02148 | -0.03175 | -0.02164 | 0.009239 | 0.01032 |
| MONORE | rs4943818 | rs12184871 | A | C | 13 | TPTE2P5 | Female only | False | 0.2996 | 0.2962 | 0.301 | 0.3004 | 25672 | 19994 | 187760 | 162101 | -0.05142 | -0.02151 | -0.03174 | -0.02157 | 0.00924 | 0.01032 |
| MONORE | rs4943818 | rs4943808 | T | C | 13 | TPTE2P5 | Female only | False | 0.2996 | 0.2962 | 0.3009 | 0.3004 | 25672 | 19994 | 187760 | 162101 | -0.05128 | -0.02158 | -0.03168 | -0.02158 | 0.009241 | 0.01033 |
| MONORE | rs4943818 | rs9549271 | C | T | 13 | TPTE2P5 | Female only | False | 0.2944 | 0.2911 | 0.2976 | 0.2973 | 25672 | 19994 | 187760 | 162101 | -0.05034 | -0.02486 | -0.03159 | -0.02163 | 0.009281 | 0.01039 |
| MONORE | rs4943818 | rs6563844 | C | T | 13 | TPTE2P5 | Female only | False | 0.2933 | 0.2902 | 0.2972 | 0.2969 | 25672 | 19994 | 187760 | 162101 | -0.05033 | -0.02601 | -0.03144 | -0.02156 | 0.009291 | 0.0104 |
| MONORE | rs4943818 | rs61963345 | C | T | 13 | TPTE2P5 | Female only | False | 0.2935 | 0.2905 | 0.2973 | 0.297 | 25672 | 19994 | 187760 | 162101 | -0.04994 | -0.02605 | -0.03143 | -0.02158 | 0.009288 | 0.01039 |
| MONORE | rs4943818 | rs4941989 | C | T | 13 | TPTE2P5 | Female only | False | 0.2998 | 0.2963 | 0.301 | 0.3005 | 25672 | 19994 | 187760 | 162101 | -0.05091 | -0.02171 | -0.03179 | -0.02146 | 0.009239 | 0.01033 |
| MONORE | rs4943818 | rs4941990 | C | A | 13 | TPTE2P5 | Female only | False | 0.2998 | 0.2963 | 0.301 | 0.3005 | 25672 | 19994 | 187760 | 162101 | -0.05091 | -0.02171 | -0.03179 | -0.02146 | 0.009239 | 0.01033 |
| MONORE | rs4943818 | rs9532588 | A | G | 13 | TPTE2P5 | Female only | False | 0.2942 | 0.2909 | 0.2975 | 0.2972 | 25672 | 19994 | 187760 | 162101 | -0.05019 | -0.02538 | -0.03169 | -0.02159 | 0.009283 | 0.01039 |
| MONORE | rs4943818 | rs9566574 | G | A | 13 | TPTE2P5 | Female only | False | 0.2998 | 0.2963 | 0.301 | 0.3005 | 25672 | 19994 | 187760 | 162101 | -0.05092 | -0.02182 | -0.03177 | -0.02147 | 0.009242 | 0.01033 |
| MONORE | rs4943818 | rs7337330 | T | C | 13 | TPTE2P5 | Female only | False | 0.2942 | 0.2909 | 0.2976 | 0.2973 | 25672 | 19994 | 187760 | 162101 | -0.05022 | -0.0254 | -0.03162 | -0.02161 | 0.009284 | 0.01039 |

| trait | lead_snp | ID | ALLELE1 | ALLELE0 | CHR | Gene_Symbol | independent_loci_in_sex_P1e2 | in_embl_any_or_gene | A1FREQ_VUMC_female | A1FREQ_VUMC_male | A1FREQ_UKB_female | A1FREQ_UKB_male | N_VUMC_female | N_VUMC_male | N_UKB_female | N_UKB_male | BETA_VUMC_female | BETA_VUMC_male | BETA_UKB_female | BETA_UKB_male | SE_VUMC_female | SE_VUMC_male |
|---|---|---|---|---|---|---|---|---|---|---|---|---|---|---|---|---|---|---|---|---|---|---|
| MONORE | rs4943818 | rs9603786 | A | G | 13 | TPTE2P5 | Female only | False | 0.2998 | 0.2963 | 0.301 | 0.3005 | 25672 | 19994 | 187760 | 162101 | -0.05092 | -0.02182 | -0.03175 | -0.02152 | 0.009242 | 0.01033 |
| MONORE | rs4943818 | rs6563845 | G | A | 13 | TPTE2P5 | Female only | False | 0.2994 | 0.2959 | 0.3009 | 0.3004 | 25672 | 19994 | 187760 | 162101 | -0.05069 | -0.02203 | -0.03195 | -0.02159 | 0.009251 | 0.01034 |
| MONORE | rs4943818 | rs9577136 | C | T | 13 | TPTE2P5 | Female only | False | 0.3091 | 0.3046 | 0.3106 | 0.3104 | 25672 | 19994 | 187760 | 162101 | -0.048 | -0.0202 | -0.03075 | -0.0207 | 0.009169 | 0.01024 |
| MONORE | rs4943818 | rs9549280 | C | T | 13 | TPTE2P5 | Female only | False | 0.2933 | 0.2904 | 0.2974 | 0.2971 | 25672 | 19994 | 187760 | 162101 | -0.04976 | -0.02626 | -0.03175 | -0.02167 | 0.009292 | 0.0104 |
| MONORE | rs4943818 | rs9315785 | C | T | 13 | TPTE2P5 | Female only | False | 0.3092 | 0.3047 | 0.3108 | 0.3106 | 25672 | 19994 | 187760 | 162101 | -0.04808 | -0.02029 | -0.03092 | -0.02043 | 0.009164 | 0.01024 |
| MONORE | rs4943818 | rs7998724 | T | G | 13 | TPTE2P5 | Female only | False | 0.2998 | 0.2962 | 0.3013 | 0.3007 | 25672 | 19994 | 187760 | 162101 | -0.05073 | -0.0219 | -0.03214 | -0.02157 | 0.009244 | 0.01033 |
| MONORE | rs4943818 | rs4941991 | C | T | 13 | TPTE2P5 | Female only | False | 0.2933 | 0.2903 | 0.2974 | 0.2971 | 25672 | 19994 | 187760 | 162101 | -0.04942 | -0.02613 | -0.03174 | -0.02165 | 0.009296 | 0.0104 |
| MONORE | rs4943818 | rs4943812 | C | T | 13 | TPTE2P5 | Female only | False | 0.2997 | 0.2961 | 0.3012 | 0.3006 | 25672 | 19994 | 187760 | 162101 | -0.0507 | -0.02182 | -0.03211 | -0.02148 | 0.009247 | 0.01034 |
| MONORE | rs4943818 | rs7986077 | G | T | 13 | TPTE2P5 | Female only | False | 0.2997 | 0.2961 | 0.3012 | 0.3007 | 25672 | 19994 | 187760 | 162101 | -0.05057 | -0.02201 | -0.0321 | -0.02145 | 0.009248 | 0.01034 |
| MONORE | rs4943818 | rs9549296 | C | T | 13 | TPTE2P5 | Female only | False | 0.2964 | 0.2902 | 0.293 | 0.2933 | 25672 | 19994 | 187760 | 162101 | -0.05164 | -0.01288 | -0.028 | -0.01884 | 0.009291 | 0.01041 |
| MONORE | rs4943818 | rs4943818 | C | A | 13 | TPTE2P5 | Female only | False | 0.2911 | 0.2851 | 0.29 | 0.2902 | 25672 | 19994 | 187760 | 162101 | -0.05233 | -0.01513 | -0.02845 | -0.01849 | 0.009335 | 0.01047 |
| MONORE | rs10414008 | rs10424624 | T | C | 19 | CTD-2540B15.12 | Female only | True | 0.3137 | 0.3145 | 0.3131 | 0.3116 | 25672 | 19994 | 187760 | 162101 | 0.04009 | 0.01539 | 0.02736 | 0.02468 | 0.009177 | 0.01019 |
| MONORE | rs10414008 | rs10412166 | A | G | 19 | CTD-2540B15.12 | Female only | True | 0.4323 | 0.4339 | 0.4302 | 0.4292 | 25672 | 19994 | 187760 | 162101 | 0.04782 | 0.009621 | 0.0281 | 0.01794 | 0.008606 | 0.009534 |
| MONORE | rs10414008 | rs10414008 | A | C | 19 | CTD-2540B15.12 | Female only | True | 0.4324 | 0.434 | 0.4302 | 0.4292 | 25672 | 19994 | 187760 | 162101 | 0.04793 | 0.009978 | 0.02822 | 0.01788 | 0.008607 | 0.009532 |
| NEUTAB | rs72803397 | rs4315507 | G | T | 2 | RN7SL51P | Male only | True | 0.1274 | 0.1281 | 0.1323 | 0.134 | 25593 | 19958 | 187758 | 162098 | -0.01137 | -0.06532 | -0.001356 | -0.01267 | 0.01217 | 0.01313 |
| NEUTAB | rs72803397 | rs116029002 | C | T | 2 | RN7SL51P | Male only | True | 0.1451 | 0.1442 | 0.1484 | 0.1502 | 25593 | 19958 | 187758 | 162098 | -0.0116 | -0.06415 | -0.002986 | -0.01192 | 0.01138 | 0.01231 |
| NEUTAB | rs72803397 | rs72803383 | A | G | 2 | RN7SL51P | Male only | True | 0.1451 | 0.1443 | 0.1485 | 0.1502 | 25593 | 19958 | 187758 | 162098 | -0.01219 | -0.06411 | -0.003076 | -0.01211 | 0.01139 | 0.01232 |
| NEUTAB | rs72803397 | rs72803385 | T | C | 2 | RN7SL51P | Male only | True | 0.1436 | 0.143 | 0.1457 | 0.1474 | 25593 | 19958 | 187758 | 162098 | -0.01173 | -0.06501 | -0.003972 | -0.0113 | 0.01143 | 0.01235 |
| NEUTAB | rs72803397 | rs72803386 | G | A | 2 | RN7SL51P | Male only | True | 0.1451 | 0.1442 | 0.1485 | 0.1502 | 25593 | 19958 | 187758 | 162098 | -0.01184 | -0.0647 | -0.003031 | -0.0123 | 0.0114 | 0.01233 |
| NEUTAB | rs72803397 | rs72803388 | T | C | 2 | RN7SL51P | Male only | True | 0.1451 | 0.1441 | 0.1485 | 0.1502 | 25593 | 19958 | 187758 | 162098 | -0.01196 | -0.06484 | -0.002985 | -0.01231 | 0.0114 | 0.01233 |
| NEUTAB | rs72803397 | rs6733675 | T | C | 2 | RN7SL51P | Male only | True | 0.1451 | 0.1442 | 0.1485 | 0.1502 | 25593 | 19958 | 187758 | 162098 | -0.01199 | -0.06484 | -0.002981 | -0.01233 | 0.0114 | 0.01233 |
| NEUTAB | rs72803397 | rs6748901 | C | T | 2 | RN7SL51P | Male only | True | 0.1452 | 0.1442 | 0.1486 | 0.1502 | 25593 | 19958 | 187758 | 162098 | -0.01187 | -0.06473 | -0.002943 | -0.01224 | 0.0114 | 0.01233 |
| NEUTAB | rs72803397 | rs72803390 | T | C | 2 | RN7SL51P | Male only | True | 0.145 | 0.1439 | 0.147 | 0.1486 | 25593 | 19958 | 187758 | 162098 | -0.01323 | -0.06845 | -0.003432 | -0.01392 | 0.01143 | 0.01237 |
| NEUTAB | rs72803397 | rs72803391 | G | A | 2 | RN7SL51P | Male only | True | 0.1442 | 0.1429 | 0.147 | 0.1486 | 25593 | 19958 | 187758 | 162098 | -0.01187 | -0.06875 | -0.004023 | -0.01515 | 0.01147 | 0.0124 |
| NEUTAB | rs72803397 | rs73933355 | G | A | 2 | RN7SL51P | Male only | True | 0.1448 | 0.1437 | 0.1471 | 0.1487 | 25593 | 19958 | 187758 | 162098 | -0.01266 | -0.06803 | -0.004079 | -0.01512 | 0.01145 | 0.01237 |
| NEUTAB | rs72803397 | rs7424671 | C | T | 2 | RN7SL51P | Male only | True | 0.1449 | 0.1437 | 0.1471 | 0.1487 | 25593 | 19958 | 187758 | 162098 | -0.01262 | -0.06806 | -0.004121 | -0.01515 | 0.01145 | 0.01237 |
| NEUTAB | rs72803397 | rs56057411 | T | C | 2 | RN7SL51P | Male only | True | 0.1442 | 0.1428 | 0.1468 | 0.1484 | 25593 | 19958 | 187758 | 162098 | -0.01196 | -0.06889 | -0.004223 | -0.01525 | 0.01147 | 0.0124 |

| trait | lead_snp | ID | ALLELE1 | ALLELE0 | CHR | Gene_Symbol | independent_loci_in_sex_P1e2 | in_embl_any_or_gene | A1FREQ_VUMC_female | A1FREQ_VUMC_male | A1FREQ_UKB_female | A1FREQ_UKB_male | N_VUMC_female | N_VUMC_male | N_UKB_female | N_UKB_male | BETA_VUMC_female | BETA_VUMC_male | BETA_UKB_female | BETA_UKB_male | SE_VUMC_female | SE_VUMC_male |
|---|---|---|---|---|---|---|---|---|---|---|---|---|---|---|---|---|---|---|---|---|---|---|
| NEUTAB | rs72803397 | rs12622051 | A | G | 2 | RN7SL51P | Male only | True | 0.1446 | 0.1434 | 0.1468 | 0.1485 | 25593 | 19958 | 187758 | 162098 | -0.01314 | -0.06908 | -0.004067 | -0.0154 | 0.01145 | 0.01238 |
| NEUTAB | rs72803397 | rs56162641 | T | C | 2 | RN7SL51P | Male only | True | 0.1446 | 0.1435 | 0.1469 | 0.1485 | 25593 | 19958 | 187758 | 162098 | -0.0131 | -0.06906 | -0.00407 | -0.01537 | 0.01145 | 0.01238 |
| NEUTAB | rs72803397 | rs56216216 | A | G | 2 | RN7SL51P | Male only | True | 0.1446 | 0.1434 | 0.1468 | 0.1484 | 25593 | 19958 | 187758 | 162098 | -0.01289 | -0.0697 | -0.004185 | -0.01539 | 0.01144 | 0.01237 |
| NEUTAB | rs72803397 | rs72803397 | A | G | 2 | RN7SL51P | Male only | True | 0.1406 | 0.1393 | 0.1423 | 0.1439 | 25593 | 19958 | 187758 | 162098 | -0.01244 | -0.07318 | -0.002274 | -0.0163 | 0.0116 | 0.01253 |
| NEUTAB | rs72803397 | rs56718216 | T | C | 2 | RN7SL51P | Male only | True | 0.1425 | 0.1416 | 0.1451 | 0.1466 | 25593 | 19958 | 187758 | 162098 | -0.01262 | -0.0704 | -0.003904 | -0.01529 | 0.01152 | 0.01245 |
| NEUTAB | rs72803397 | rs7426219 | T | C | 2 | RN7SL51P | Male only | True | 0.1426 | 0.1417 | 0.1451 | 0.1466 | 25593 | 19958 | 187758 | 162098 | -0.01286 | -0.07022 | -0.004037 | -0.01558 | 0.01151 | 0.01245 |
| NEUTAB | rs72803397 | rs4630755 | C | T | 2 | RN7SL51P | Male only | True | 0.142 | 0.141 | 0.1449 | 0.1465 | 25593 | 19958 | 187758 | 162098 | -0.01237 | -0.07086 | -0.004005 | -0.01586 | 0.01153 | 0.01247 |
| NEUTAB | rs72803397 | rs58294090 | G | T | 2 | RN7SL51P | Male only | True | 0.1415 | 0.1407 | 0.1442 | 0.1457 | 25593 | 19958 | 187758 | 162098 | -0.009297 | -0.06954 | -0.003706 | -0.01564 | 0.01154 | 0.01247 |
| NEUTAB | rs72803397 | rs146513501 | T | G | 2 | RN7SL51P | Male only | True | 0.1417 | 0.1408 | 0.1444 | 0.1458 | 25593 | 19958 | 187758 | 162098 | -0.007917 | -0.06939 | -0.003226 | -0.01559 | 0.01154 | 0.01248 |
| NEUTAB | rs72803397 | rs57902549 | A | G | 2 | RN7SL51P | Male only | True | 0.1422 | 0.1412 | 0.1444 | 0.1458 | 25593 | 19958 | 187758 | 162098 | -0.00722 | -0.06925 | -0.003103 | -0.01544 | 0.01153 | 0.01247 |
| NEUTAB | rs72803397 | rs55893750 | T | G | 2 | RN7SL51P | Male only | True | 0.1418 | 0.1408 | 0.1442 | 0.1457 | 25593 | 19958 | 187758 | 162098 | -0.007053 | -0.06965 | -0.003319 | -0.01543 | 0.01154 | 0.01248 |
| NEUTAB | rs72803397 | rs56124198 | G | A | 2 | RN7SL51P | Male only | True | 0.1418 | 0.1408 | 0.1443 | 0.1458 | 25593 | 19958 | 187758 | 162098 | -0.007157 | -0.06963 | -0.003267 | -0.0154 | 0.01154 | 0.01248 |
| NEUTAB | rs72803397 | rs12621417 | C | T | 2 | RN7SL51P | Male only | True | 0.1419 | 0.1409 | 0.1443 | 0.1459 | 25593 | 19958 | 187758 | 162098 | -0.006607 | -0.06957 | -0.003254 | -0.01587 | 0.01154 | 0.01248 |
| NEUTAB | rs72803397 | rs113710006 | G | A | 2 | RN7SL51P | Male only | True | 0.1551 | 0.1542 | 0.1537 | 0.1555 | 25593 | 19958 | 187758 | 162098 | -0.01464 | -0.06083 | -0.003508 | -0.01322 | 0.01114 | 0.01202 |
| BAbs | rs6058807 | rs6141766 | G | A | 20 | C20orf203 | Female only | True | 0.1532 | 0.1534 | 0.1505 | 0.1506 | 14842 | 10793 | 187758 | 162098 | -0.09 | -0.02799 | -0.008861 | 0.003563 | 0.01508 | 0.01724 |
| BAbs | rs6058807 | rs12624715 | G | A | 20 | C20orf203 | Female only | True | 0.1526 | 0.1532 | 0.1504 | 0.1504 | 14842 | 10793 | 187758 | 162098 | -0.08569 | -0.02519 | -0.009111 | 0.003237 | 0.01508 | 0.01722 |
| BAbs | rs6058807 | rs6141328 | A | G | 20 | C20orf203 | Female only | True | 0.1536 | 0.1538 | 0.1508 | 0.1508 | 14842 | 10793 | 187758 | 162098 | -0.08949 | -0.02457 | -0.009259 | 0.003469 | 0.01505 | 0.01719 |
| BAbs | rs6058807 | rs73107865 | T | C | 20 | C20orf203 | Female only | True | 0.1544 | 0.1561 | 0.1525 | 0.1525 | 14842 | 10793 | 187758 | 162098 | -0.08775 | -0.02177 | -0.01086 | 0.004366 | 0.01501 | 0.01705 |
| BAbs | rs6058807 | rs6141768 | A | G | 20 | C20orf203 | Female only | True | 0.156 | 0.1575 | 0.153 | 0.153 | 14842 | 10793 | 187758 | 162098 | -0.09084 | -0.02074 | -0.01075 | 0.004569 | 0.01492 | 0.01696 |
| BAbs | rs6058807 | rs6087964 | A | G | 20 | C20orf203 | Female only | True | 0.2325 | 0.2336 | 0.2259 | 0.2257 | 14842 | 10793 | 187758 | 162098 | -0.07977 | -0.02733 | -0.007723 | 0.001969 | 0.01298 | 0.01467 |
| BAbs | rs6058807 | rs6058799 | A | G | 20 | C20orf203 | Female only | True | 0.2328 | 0.2339 | 0.2261 | 0.226 | 14842 | 10793 | 187758 | 162098 | -0.07884 | -0.02666 | -0.008055 | 0.002064 | 0.01297 | 0.01464 |
| BAbs | rs6058807 | rs6579029 | T | C | 20 | C20orf203 | Female only | True | 0.2326 | 0.2337 | 0.2261 | 0.226 | 14842 | 10793 | 187758 | 162098 | -0.0789 | -0.0273 | -0.008073 | 0.002024 | 0.01297 | 0.01465 |
| BAbs | rs6058807 | rs6057617 | A | G | 20 | C20orf203 | Female only | True | 0.2333 | 0.2344 | 0.2261 | 0.226 | 14842 | 10793 | 187758 | 162098 | -0.07836 | -0.02537 | -0.008121 | 0.002068 | 0.01296 | 0.01462 |
| BAbs | rs6058807 | rs12480157 | A | G | 20 | C20orf203 | Female only | True | 0.2301 | 0.2313 | 0.2262 | 0.2262 | 14842 | 10793 | 187758 | 162098 | -0.0779 | -0.02553 | -0.00799 | 0.00216 | 0.0129 | 0.01457 |
| BAbs | rs6058807 | rs6119932 | G | T | 20 | C20orf203 | Female only | True | 0.2341 | 0.2351 | 0.2264 | 0.2264 | 14842 | 10793 | 187758 | 162098 | -0.07836 | -0.02561 | -0.008312 | 0.001889 | 0.01294 | 0.01462 |
| BAbs | rs6058807 | rs6057618 | A | G | 20 | C20orf203 | Female only | True | 0.2321 | 0.2326 | 0.2251 | 0.2252 | 14842 | 10793 | 187758 | 162098 | -0.07894 | -0.02704 | -0.008395 | 0.001796 | 0.01298 | 0.01466 |
| BAbs | rs6058807 | rs6057619 | T | C | 20 | C20orf203 | Female only | True | 0.2285 | 0.2304 | 0.2245 | 0.2245 | 14842 | 10793 | 187758 | 162098 | -0.08024 | -0.02461 | -0.008334 | 0.001546 | 0.01294 | 0.0146 |

| trait | lead_snp | ID | ALLELE1 | ALLELE0 | CHR | Gene_Symbol | independent_loci_in_sex_P1e2 | in_embl_any_or_gene | A1FREQ_VUMC_female | A1FREQ_VUMC_male | A1FREQ_UKB_female | A1FREQ_UKB_male | N_VUMC_female | N_VUMC_male | N_UKB_female | N_UKB_male | BETA_VUMC_female | BETA_VUMC_male | BETA_UKB_female | BETA_UKB_male | SE_VUMC_female | SE_VUMC_male |
|---|---|---|---|---|---|---|---|---|---|---|---|---|---|---|---|---|---|---|---|---|---|---|
| BAbs | rs6058807 | rs6058807 | T | C | 20 | C20orf203 | Female only | True | 0.2325 | 0.2336 | 0.2253 | 0.2252 | 14842 | 10793 | 187758 | 162098 | -0.08068 | -0.02375 | -0.008215 | 0.001496 | 0.01297 | 0.01465 |
| BAbs | rs6058807 | rs6057620 | A | G | 20 | C20orf203 | Female only | True | 0.2326 | 0.2336 | 0.2253 | 0.2253 | 14842 | 10793 | 187758 | 162098 | -0.0806 | -0.02472 | -0.008515 | 0.00149 | 0.01297 | 0.01465 |
| BAbs | rs6058807 | rs6057621 | A | G | 20 | C20orf203 | Female only | True | 0.2317 | 0.2321 | 0.2246 | 0.2247 | 14842 | 10793 | 187758 | 162098 | -0.07993 | -0.02489 | -0.008907 | 0.001357 | 0.01299 | 0.01466 |
| BAbs | rs6058807 | rs6058809 | T | C | 20 | C20orf203 | Female only | True | 0.2306 | 0.2323 | 0.224 | 0.2241 | 14842 | 10793 | 187758 | 162098 | -0.07824 | -0.02499 | -0.009296 | 0.001061 | 0.01302 | 0.01468 |
| BAbs | rs6058807 | rs6119935 | C | A | 20 | C20orf203 | Female only | True | 0.2314 | 0.2337 | 0.2243 | 0.2243 | 14842 | 10793 | 187758 | 162098 | -0.07915 | -0.02425 | -0.008967 | 0.001127 | 0.013 | 0.01466 |
| BAbs | rs6058807 | rs6058812 | A | G | 20 | C20orf203 | Female only | True | 0.2312 | 0.233 | 0.2242 | 0.2242 | 14842 | 10793 | 187758 | 162098 | -0.0784 | -0.02523 | -0.00902 | 0.001076 | 0.01303 | 0.0147 |
| BAbs | rs6058807 | rs6058813 | C | T | 20 | C20orf203 | Female only | True | 0.2316 | 0.2336 | 0.2243 | 0.2243 | 14842 | 10793 | 187758 | 162098 | -0.07894 | -0.0253 | -0.008989 | 0.001081 | 0.01302 | 0.01469 |
| BAbs | rs6058807 | rs11905501 | C | A | 20 | C20orf203 | Female only | True | 0.2316 | 0.2336 | 0.2244 | 0.2243 | 14842 | 10793 | 187758 | 162098 | -0.07895 | -0.0253 | -0.008948 | 0.00107 | 0.01302 | 0.01469 |
| BAbs | rs6058807 | rs28456308 | C | T | 20 | C20orf203 | Female only | True | 0.2364 | 0.238 | 0.2295 | 0.2298 | 14842 | 10793 | 187758 | 162098 | -0.0745 | -0.01978 | -0.007959 | 8.28e-04 | 0.01292 | 0.01455 |
| BAbs | rs6058807 | rs28402080 | A | G | 20 | C20orf203 | Female only | True | 0.2367 | 0.2383 | 0.2296 | 0.2298 | 14842 | 10793 | 187758 | 162098 | -0.07391 | -0.01993 | -0.007991 | 7.43e-04 | 0.01292 | 0.01456 |
| BAbs | rs6058807 | rs28583637 | G | A | 20 | C20orf203 | Female only | True | 0.2367 | 0.2382 | 0.2296 | 0.2299 | 14842 | 10793 | 187758 | 162098 | -0.07377 | -0.02008 | -0.00801 | 8.25e-04 | 0.01292 | 0.01456 |
| BAbs | rs6058807 | rs28698192 | T | C | 20 | C20orf203 | Female only | True | 0.2372 | 0.2389 | 0.2296 | 0.2299 | 14842 | 10793 | 187758 | 162098 | -0.07268 | -0.02061 | -0.008079 | 7.98e-04 | 0.01291 | 0.01455 |
| BAbs | rs6058807 | rs28605309 | A | G | 20 | C20orf203 | Female only | True | 0.2372 | 0.2389 | 0.2297 | 0.23 | 14842 | 10793 | 187758 | 162098 | -0.07268 | -0.02062 | -0.008078 | 7.97e-04 | 0.01291 | 0.01455 |
| BAbs | rs6058807 | rs28565102 | T | G | 20 | C20orf203 | Female only | True | 0.2372 | 0.2389 | 0.2296 | 0.23 | 14842 | 10793 | 187758 | 162098 | -0.07268 | -0.02061 | -0.008034 | 7.62e-04 | 0.01291 | 0.01455 |
| BAbs | rs6058807 | rs28510728 | G | A | 20 | C20orf203 | Female only | True | 0.2376 | 0.2393 | 0.2297 | 0.2301 | 14842 | 10793 | 187758 | 162098 | -0.07302 | -0.02081 | -0.008072 | 5.36e-04 | 0.01292 | 0.01455 |
| BAbs | rs6058807 | rs28707468 | T | C | 20 | C20orf203 | Female only | True | 0.2377 | 0.2395 | 0.2297 | 0.2301 | 14842 | 10793 | 187758 | 162098 | -0.07283 | -0.02019 | -0.008045 | 5.66e-04 | 0.01291 | 0.01455 |
| BAbs | rs6058807 | rs28369623 | T | C | 20 | C20orf203 | Female only | True | 0.2377 | 0.2395 | 0.2297 | 0.2301 | 14842 | 10793 | 187758 | 162098 | -0.07283 | -0.02019 | -0.008068 | 5.75e-04 | 0.01291 | 0.01455 |
| BAbs | rs6058807 | rs9421 | C | T | 20 | C20orf203 | Female only | True | 0.2376 | 0.2392 | 0.229 | 0.2293 | 14842 | 10793 | 187758 | 162098 | -0.07248 | -0.01976 | -0.00818 | 9.32e-04 | 0.01292 | 0.01456 |
| BAbs | rs6058807 | rs28548540 | A | G | 20 | C20orf203 | Female only | True | 0.2377 | 0.2393 | 0.2289 | 0.2292 | 14842 | 10793 | 187758 | 162098 | -0.07236 | -0.01888 | -0.008028 | 8.62e-04 | 0.01292 | 0.01455 |
| BAbs | rs6058807 | rs28730896 | A | G | 20 | C20orf203 | Female only | True | 0.2373 | 0.2388 | 0.2289 | 0.2292 | 14842 | 10793 | 187758 | 162098 | -0.07322 | -0.01909 | -0.008014 | 8.68e-04 | 0.01292 | 0.01457 |
| BAbs | rs6058807 | rs28612490 | G | A | 20 | C20orf203 | Female only | True | 0.2372 | 0.2387 | 0.2288 | 0.2291 | 14842 | 10793 | 187758 | 162098 | -0.07326 | -0.01947 | -0.007991 | 9.05e-04 | 0.01293 | 0.01457 |
| BAbs | rs6058807 | rs28695612 | G | T | 20 | C20orf203 | Female only | True | 0.2373 | 0.2387 | 0.2288 | 0.2291 | 14842 | 10793 | 187758 | 162098 | -0.07307 | -0.01936 | -0.007986 | 8.80e-04 | 0.01292 | 0.01457 |
| BAbs | rs6058807 | rs28421806 | A | G | 20 | C20orf203 | Female only | True | 0.2373 | 0.2387 | 0.2288 | 0.2291 | 14842 | 10793 | 187758 | 162098 | -0.07307 | -0.01936 | -0.007981 | 8.84e-04 | 0.01292 | 0.01457 |
| BAbs | rs6058807 | rs28456361 | T | C | 20 | C20orf203 | Female only | True | 0.2373 | 0.2387 | 0.2288 | 0.2291 | 14842 | 10793 | 187758 | 162098 | -0.07307 | -0.01936 | -0.007969 | 8.86e-04 | 0.01292 | 0.01457 |
| BAbs | rs6058807 | rs28439340 | G | A | 20 | C20orf203 | Female only | True | 0.2373 | 0.2387 | 0.2288 | 0.2291 | 14842 | 10793 | 187758 | 162098 | -0.07306 | -0.01936 | -0.007968 | 8.89e-04 | 0.01292 | 0.01457 |
| BAbs | rs6058807 | rs28450123 | C | T | 20 | C20orf203 | Female only | True | 0.2373 | 0.2387 | 0.2288 | 0.2291 | 14842 | 10793 | 187758 | 162098 | -0.07306 | -0.01936 | -0.007968 | 8.89e-04 | 0.01292 | 0.01457 |
| BAbs | rs6058807 | rs28437232 | C | T | 20 | C20orf203 | Female only | True | 0.2373 | 0.2387 | 0.2288 | 0.2291 | 14842 | 10793 | 187758 | 162098 | -0.07308 | -0.0194 | -0.00797 | 8.92e-04 | 0.01292 | 0.01457 |
| BAbs | rs6058807 | rs28523897 | C | T | 20 | C20orf203 | Female only | True | 0.2373 | 0.2387 | 0.2288 | 0.2291 | 14842 | 10793 | 187758 | 162098 | -0.07308 | -0.0194 | -0.007971 | 8.97e-04 | 0.01292 | 0.01457 |
| BAbs | rs6058807 | rs28542091 | T | C | 20 | C20orf203 | Female only | True | 0.2373 | 0.2388 | 0.2288 | 0.2291 | 14842 | 10793 | 187758 | 162098 | -0.07294 | -0.01943 | -0.007978 | 8.79e-04 | 0.01292 | 0.01457 |

| trait | lead_snp | ID | ALLELE1 | ALLELE0 | CHR | Gene_Symbol | independent_loci_in_sex_P1e2 | in_embl_any_or_gene | A1FREQ_VUMC_female | A1FREQ_VUMC_male | A1FREQ_UKB_female | A1FREQ_UKB_male | N_VUMC_female | N_VUMC_male | N_UKB_female | N_UKB_male | BETA_VUMC_female | BETA_VUMC_male | BETA_UKB_female | BETA_UKB_male | SE_VUMC_female | SE_VUMC_male |
|---|---|---|---|---|---|---|---|---|---|---|---|---|---|---|---|---|---|---|---|---|---|---|
| BAbs | rs6058807 | rs28548407 | T | C | 20 | C20orf203 | Female only | True | 0.2378 | 0.2393 | 0.2288 | 0.2291 | 14842 | 10793 | 187758 | 162098 | -0.07248 | -0.01842 | -0.007969 | 8.98e-04 | 0.01291 | 0.01454 |
| BAbs | rs6058807 | rs28481782 | T | C | 20 | C20orf203 | Female only | True | 0.2378 | 0.2393 | 0.2288 | 0.2291 | 14842 | 10793 | 187758 | 162098 | -0.07248 | -0.01842 | -0.007967 | 8.99e-04 | 0.01291 | 0.01454 |
| BAbs | rs6058807 | rs28536730 | A | C | 20 | C20orf203 | Female only | True | 0.2378 | 0.2393 | 0.2288 | 0.2291 | 14842 | 10793 | 187758 | 162098 | -0.07249 | -0.01842 | -0.007974 | 9.18e-04 | 0.01291 | 0.01454 |
| BAbs | rs6058807 | rs6058819 | T | C | 20 | C20orf203 | Female only | True | 0.2379 | 0.2394 | 0.2288 | 0.2291 | 14842 | 10793 | 187758 | 162098 | -0.07241 | -0.01876 | -0.007967 | 9.38e-04 | 0.01291 | 0.01454 |
| BAbs | rs6058807 | rs6058820 | T | C | 20 | C20orf203 | Female only | True | 0.2378 | 0.2393 | 0.2288 | 0.2291 | 14842 | 10793 | 187758 | 162098 | -0.07224 | -0.01859 | -0.007945 | 9.48e-04 | 0.01291 | 0.01454 |
| BAbs | rs6058807 | rs6058821 | C | T | 20 | C20orf203 | Female only | True | 0.2378 | 0.2391 | 0.2285 | 0.2288 | 14842 | 10793 | 187758 | 162098 | -0.07247 | -0.01841 | -0.007811 | 8.36e-04 | 0.01291 | 0.01454 |
| BAbs | rs6058807 | rs6057623 | G | T | 20 | C20orf203 | Female only | True | 0.2377 | 0.2391 | 0.2285 | 0.2288 | 14842 | 10793 | 187758 | 162098 | -0.07241 | -0.01851 | -0.007849 | 8.41e-04 | 0.01291 | 0.01454 |
| BAbs | rs6058807 | rs6057625 | G | A | 20 | C20orf203 | Female only | True | 0.2378 | 0.2391 | 0.2285 | 0.2288 | 14842 | 10793 | 187758 | 162098 | -0.07227 | -0.0185 | -0.00785 | 8.38e-04 | 0.01291 | 0.01454 |
| BAbs | rs6058807 | rs6057626 | T | C | 20 | C20orf203 | Female only | True | 0.2378 | 0.2391 | 0.2285 | 0.2288 | 14842 | 10793 | 187758 | 162098 | -0.07227 | -0.01851 | -0.007839 | 8.70e-04 | 0.01291 | 0.01454 |
| BAbs | rs6058807 | rs6058825 | G | A | 20 | C20orf203 | Female only | True | 0.2382 | 0.2394 | 0.2288 | 0.2292 | 14842 | 10793 | 187758 | 162098 | -0.07191 | -0.01764 | -0.007723 | 8.49e-04 | 0.01291 | 0.01455 |
| BAbs | rs6058807 | rs6058828 | A | G | 20 | C20orf203 | Female only | True | 0.2377 | 0.2391 | 0.2285 | 0.2288 | 14842 | 10793 | 187758 | 162098 | -0.0724 | -0.01851 | -0.007827 | 8.82e-04 | 0.01291 | 0.01454 |
| BAbs | rs6058807 | rs6057628 | C | T | 20 | C20orf203 | Female only | True | 0.2377 | 0.2391 | 0.2285 | 0.2288 | 14842 | 10793 | 187758 | 162098 | -0.07241 | -0.0185 | -0.007822 | 8.56e-04 | 0.01291 | 0.01454 |
| BAbs | rs6058807 | rs2377665 | T | C | 20 | C20orf203 | Female only | True | 0.24 | 0.2413 | 0.2305 | 0.2309 | 14842 | 10793 | 187758 | 162098 | -0.07289 | -0.01664 | -0.007667 | 0.001142 | 0.01288 | 0.01452 |
| BAbs | rs6058807 | rs6058829 | G | A | 20 | C20orf203 | Female only | True | 0.2378 | 0.2391 | 0.2285 | 0.2288 | 14842 | 10793 | 187758 | 162098 | -0.07227 | -0.0185 | -0.007822 | 8.59e-04 | 0.01291 | 0.01454 |
| BAbs | rs6058807 | rs4425162 | A | G | 20 | C20orf203 | Female only | True | 0.2378 | 0.2391 | 0.2285 | 0.2288 | 14842 | 10793 | 187758 | 162098 | -0.07227 | -0.0185 | -0.007826 | 8.72e-04 | 0.01291 | 0.01454 |
| BAbs | rs6058807 | rs6058832 | A | G | 20 | C20orf203 | Female only | True | 0.2375 | 0.2388 | 0.2283 | 0.2287 | 14842 | 10793 | 187758 | 162098 | -0.0728 | -0.01939 | -0.007826 | 7.28e-04 | 0.01292 | 0.01454 |
| BAbs | rs6058807 | rs2064933 | A | G | 20 | C20orf203 | Female only | True | 0.2328 | 0.2342 | 0.2283 | 0.2287 | 14842 | 10793 | 187758 | 162098 | -0.07243 | -0.01899 | -0.007841 | 6.74e-04 | 0.01284 | 0.01446 |
| BAbs | rs6058807 | rs2064935 | G | A | 20 | C20orf203 | Female only | True | 0.2375 | 0.2388 | 0.2283 | 0.2287 | 14842 | 10793 | 187758 | 162098 | -0.07302 | -0.0194 | -0.00783 | 7.28e-04 | 0.01291 | 0.01454 |
| BAbs | rs6058807 | rs6119939 | T | C | 20 | C20orf203 | Female only | True | 0.2375 | 0.2388 | 0.2283 | 0.2287 | 14842 | 10793 | 187758 | 162098 | -0.0729 | -0.01942 | -0.007833 | 7.44e-04 | 0.01291 | 0.01454 |
| trait | lead_snp | ID | ALLELE1 | ALLELE0 | CHR | Gene_Symbol | independent_loci_in_sex_P1e2 | in_embl_any_or_gene | A1FREQ_VUMC_female | A1FREQ_VUMC_male | A1FREQ_UKB_female | A1FREQ_UKB_male | N_VUMC_female | N_VUMC_male | N_UKB_female | N_UKB_male | BETA_VUMC_female | BETA_VUMC_male | BETA_UKB_female | BETA_UKB_male | SE_VUMC_female | SE_VUMC_male |
| BAbs | rs6058807 | rs7360824 | A | G | 20 | C20orf203 | Female only | True | 0.2377 | 0.2389 | 0.2284 | 0.2288 | 14842 | 10793 | 187758 | 162098 | -0.07273 | -0.01937 | -0.007936 | 6.39e-04 | 0.01291 | 0.01454 |
| BAbs | rs6058807 | rs6087976 | G | A | 20 | C20orf203 | Female only | True | 0.2375 | 0.2388 | 0.2283 | 0.2287 | 14842 | 10793 | 187758 | 162098 | -0.07302 | -0.0194 | -0.007834 | 7.27e-04 | 0.01291 | 0.01454 |
| LymAbs | rs2267073 | rs9612575 | T | C | 22 | GGT5 | Male only | True | 0.503 | 0.501 | 0.4919 | 0.4918 | 15127 | 11025 | 187758 | 162098 | -0.001179 | 0.06725 | 0.008405 | 0.01157 | 0.01061 | 0.01239 |
| LymAbs | rs2267073 | rs2267073 | T | C | 22 | GGT5 | Male only | True | 0.4976 | 0.4953 | 0.4917 | 0.4917 | 15127 | 11025 | 187758 | 162098 | -0.001641 | 0.06727 | 0.008722 | 0.01167 | 0.01051 | 0.01226 |
| LymAbs | rs2267073 | rs2267075 | A | C | 22 | GGT5 | Male only | True | 0.5024 | 0.5002 | 0.4917 | 0.4916 | 15127 | 11025 | 187758 | 162098 | -0.001548 | 0.06778 | 0.00859 | 0.01157 | 0.01061 | 0.01237 |
| LymAbs | rs2267073 | rs2000472 | G | A | 22 | GGT5 | Male only | True | 0.4619 | 0.4652 | 0.4736 | 0.4742 | 15127 | 11025 | 187758 | 162098 | 0.00298 | -0.04117 | -0.006878 | -0.006908 | 0.01063 | 0.01241 |
| LymAbs | rs2267073 | rs2298380 | G | T | 22 | GGT5 | Male only | True | 0.4904 | 0.4948 | 0.4998 | 0.4994 | 15127 | 11025 | 187758 | 162098 | 8.94e-04 | -0.04383 | -0.008954 | -0.01009 | 0.01059 | 0.01237 |
| LymAbs | rs2267073 | rs5760351 | T | C | 22 | GGT5 | Male only | True | 0.489 | 0.4929 | 0.4987 | 0.499 | 15127 | 11025 | 187758 | 162098 | 0.001728 | -0.04535 | -0.009058 | -0.01017 | 0.01059 | 0.01237 |

| trait | lead_snp | ID | ALLELE1 | ALLELE0 | CHR | Gene_Symbol | independent_loci_in_sex_P1e2 | in_embl_any_or_gene | A1FREQ_VUMC_female | A1FREQ_VUMC_male | A1FREQ_UKB_female | A1FREQ_UKB_male | N_VUMC_female | N_VUMC_male | N_UKB_female | N_UKB_male | BETA_VUMC_female | BETA_VUMC_male | BETA_UKB_female | BETA_UKB_male | SE_VUMC_female | SE_VUMC_male |
|---|---|---|---|---|---|---|---|---|---|---|---|---|---|---|---|---|---|---|---|---|---|---|
| LymAbs | rs2267073 | rs738811 | C | A | 22 | GGT5 | Male only | True | 0.4629 | 0.4663 | 0.4747 | 0.4754 | 15127 | 11025 | 187758 | 162098 | 0.002124 | -0.03846 | -0.006721 | -0.007011 | 0.01063 | 0.0124 |
| LymAbs | rs2267073 | rs11090319 | T | C | 22 | GGT5 | Male only | True | 0.4881 | 0.4922 | 0.4992 | 0.4995 | 15127 | 11025 | 187758 | 162098 | 0.001076 | -0.04386 | -0.009113 | -0.01005 | 0.01057 | 0.01236 |
| LymAbs | rs2267073 | rs4820585 | C | T | 22 | GGT5 | Male only | True | 0.494 | 0.4983 | 0.5 | 0.4997 | 15127 | 11025 | 187758 | 162098 | 5.27e-04 | -0.04256 | -0.009032 | -0.01021 | 0.01049 | 0.01226 |
| LymAbs | rs2267073 | rs2330782 | T | C | 22 | GGT5 | Male only | True | 0.4615 | 0.4649 | 0.4738 | 0.4744 | 15127 | 11025 | 187758 | 162098 | 0.002863 | -0.0401 | -0.006876 | -0.006748 | 0.01062 | 0.01239 |
| LymAbs | rs2267073 | rs4822487 | C | T | 22 | GGT5 | Male only | True | 0.4899 | 0.4943 | 0.4998 | 0.4999 | 15127 | 11025 | 187758 | 162098 | 4.68e-04 | -0.04285 | -0.009024 | -0.01036 | 0.01059 | 0.01237 |
| LymAbs | rs2267073 | rs11912259 | C | T | 22 | GGT5 | Male only | True | 0.4902 | 0.4946 | 0.4998 | 0.4996 | 15127 | 11025 | 187758 | 162098 | 6.68e-04 | -0.04309 | -0.008968 | -0.01017 | 0.01059 | 0.01236 |
| LymAbs | rs2267073 | rs7289364 | G | A | 22 | GGT5 | Male only | True | 0.4628 | 0.4663 | 0.4749 | 0.4755 | 15127 | 11025 | 187758 | 162098 | 0.002298 | -0.03847 | -0.006995 | -0.006923 | 0.01062 | 0.01239 |
| LymAbs | rs2267073 | rs2196263 | A | G | 22 | GGT5 | Male only | True | 0.4873 | 0.4915 | 0.4987 | 0.499 | 15127 | 11025 | 187758 | 162098 | 0.001276 | -0.04368 | -0.009154 | -0.01023 | 0.01058 | 0.01236 |
| LymAbs | rs2267073 | rs5751856 | T | C | 22 | GGT5 | Male only | True | 0.4613 | 0.4649 | 0.4737 | 0.4744 | 15127 | 11025 | 187758 | 162098 | 0.003337 | -0.04016 | -0.00674 | -0.007001 | 0.01063 | 0.0124 |
| LymAbs | rs2267073 | rs5760382 | G | A | 22 | GGT5 | Male only | True | 0.4612 | 0.4648 | 0.4737 | 0.4743 | 15127 | 11025 | 187758 | 162098 | 0.003294 | -0.04003 | -0.006873 | -0.006955 | 0.01063 | 0.0124 |
| LymAbs | rs2267073 | rs5760387 | C | T | 22 | GGT5 | Male only | True | 0.4878 | 0.492 | 0.4991 | 0.4993 | 15127 | 11025 | 187758 | 162098 | 0.001944 | -0.04387 | -0.008829 | -0.01022 | 0.0106 | 0.01238 |
| LymAbs | rs2267073 | rs4820586 | A | G | 22 | GGT5 | Male only | True | 0.4878 | 0.4919 | 0.4994 | 0.4997 | 15127 | 11025 | 187758 | 162098 | 0.00127 | -0.04223 | -0.008955 | -0.01038 | 0.0106 | 0.01238 |
| LymAbs | rs2267073 | rs1003774 | T | G | 22 | GGT5 | Male only | True | 0.4871 | 0.4911 | 0.4991 | 0.4993 | 15127 | 11025 | 187758 | 162098 | 0.001439 | -0.04341 | -0.009041 | -0.01025 | 0.01059 | 0.01238 |
| LymAbs | rs2267073 | rs5751867 | T | C | 22 | GGT5 | Male only | True | 0.4607 | 0.4643 | 0.4743 | 0.475 | 15127 | 11025 | 187758 | 162098 | 0.00324 | -0.03967 | -0.006779 | -0.007019 | 0.01064 | 0.01242 |
| LymAbs | rs2267073 | rs743363 | A | G | 22 | GGT5 | Male only | True | 0.4607 | 0.4642 | 0.4741 | 0.4747 | 15127 | 11025 | 187758 | 162098 | 0.003338 | -0.03967 | -0.006884 | -0.006977 | 0.01064 | 0.01242 |
| LymAbs | rs2267073 | rs5760401 | A | G | 22 | GGT5 | Male only | True | 0.4871 | 0.4912 | 0.4993 | 0.4995 | 15127 | 11025 | 187758 | 162098 | 0.001549 | -0.04346 | -0.009098 | -0.01029 | 0.01059 | 0.01238 |
| LymAbs | rs2267073 | rs5751868 | A | G | 22 | GGT5 | Male only | True | 0.4872 | 0.4912 | 0.499 | 0.4992 | 15127 | 11025 | 187758 | 162098 | 0.001359 | -0.04335 | -0.009085 | -0.01024 | 0.0106 | 0.01238 |
| LymAbs | rs2267073 | rs3747114 | A | G | 22 | GGT5 | Male only | True | 0.4881 | 0.4924 | 0.4991 | 0.4994 | 15127 | 11025 | 187758 | 162098 | 0.001206 | -0.043 | -0.009248 | -0.01035 | 0.01061 | 0.01239 |
| LymAbs | rs2267073 | rs3747115 | C | T | 22 | GGT5 | Male only | True | 0.4877 | 0.4921 | 0.499 | 0.4992 | 15127 | 11025 | 187758 | 162098 | 0.001577 | -0.04348 | -0.00906 | -0.01021 | 0.0106 | 0.01238 |
| LymAbs | rs2267073 | rs5751869 | A | G | 22 | GGT5 | Male only | True | 0.4851 | 0.4896 | 0.498 | 0.4982 | 15127 | 11025 | 187758 | 162098 | 0.001819 | -0.04303 | -0.009417 | -0.01042 | 0.0106 | 0.01238 |
| LymAbs | rs2267073 | rs5760410 | A | G | 22 | GGT5 | Male only | True | 0.4926 | 0.4968 | 0.4965 | 0.4965 | 15127 | 11025 | 187758 | 162098 | 0.001527 | -0.04207 | -0.008945 | -0.01016 | 0.01048 | 0.01223 |
| MPV | rs56098529 | rs28485712 | C | T | 2 | CPS1 | Female only | True | 0.2542 | 0.2516 | 0.2478 | 0.2453 | 26726 | 19784 | 188072 | 162398 | -0.04915 | -0.008095 | -0.01951 | -0.007279 | 0.009063 | 0.01082 |
| MPV | rs56098529 | rs72932418 | G | A | 2 | CPS1 | Female only | True | 0.2543 | 0.2518 | 0.248 | 0.2455 | 26726 | 19784 | 188072 | 162398 | -0.0495 | -0.007669 | -0.01947 | -0.007294 | 0.00906 | 0.01082 |
| MPV | rs56098529 | rs72932421 | G | A | 2 | CPS1 | Female only | True | 0.2543 | 0.2518 | 0.2479 | 0.2454 | 26726 | 19784 | 188072 | 162398 | -0.04925 | -0.007671 | -0.01946 | -0.007232 | 0.009062 | 0.01082 |
| MPV | rs56098529 | rs72932424 | A | G | 2 | CPS1 | Female only | True | 0.2543 | 0.2518 | 0.2479 | 0.2454 | 26726 | 19784 | 188072 | 162398 | -0.04928 | -0.007669 | -0.01943 | -0.007247 | 0.009061 | 0.01082 |
| MPV | rs56098529 | rs12463895 | A | G | 2 | CPS1 | Female only | True | 0.2543 | 0.2518 | 0.2479 | 0.2454 | 26726 | 19784 | 188072 | 162398 | -0.04928 | -0.00767 | -0.01943 | -0.007219 | 0.009061 | 0.01082 |
| MPV | rs56098529 | rs12468052 | C | T | 2 | CPS1 | Female only | True | 0.2543 | 0.2518 | 0.2479 | 0.2454 | 26726 | 19784 | 188072 | 162398 | -0.04928 | -0.00767 | -0.01943 | -0.00722 | 0.009061 | 0.01082 |
| MPV | rs56098529 | rs72932427 | T | C | 2 | CPS1 | Female only | True | 0.2543 | 0.2518 | 0.2479 | 0.2454 | 26726 | 19784 | 188072 | 162398 | -0.04928 | -0.007661 | -0.01942 | -0.007226 | 0.009061 | 0.01082 |
| MPV | rs56098529 | rs72932430 | G | A | 2 | CPS1 | Female only | True | 0.2543 | 0.2518 | 0.2479 | 0.2454 | 26726 | 19784 | 188072 | 162398 | -0.04928 | -0.00767 | -0.01942 | -0.007221 | 0.009061 | 0.01082 |

| trait | lead_snp | ID | ALLELE1 | ALLELE0 | CHR | Gene_Symbol | independent_loci_in_sex_P1e2 | in_embl_any_or_gene | A1FREQ_VUMC_female | A1FREQ_VUMC_male | A1FREQ_UKB_female | A1FREQ_UKB_male | N_VUMC_female | N_VUMC_male | N_UKB_female | N_UKB_male | BETA_VUMC_female | BETA_VUMC_male | BETA_UKB_female | BETA_UKB_male | SE_VUMC_female | SE_VUMC_male |
|---|---|---|---|---|---|---|---|---|---|---|---|---|---|---|---|---|---|---|---|---|---|---|
| MPV | rs56098529 | rs17771664 | A | G | 2 | CPS1 | Female only | True | 0.2546 | 0.2522 | 0.2478 | 0.2454 | 26726 | 19784 | 188072 | 162398 | -0.04866 | -0.008561 | -0.01942 | -0.007242 | 0.009023 | 0.01078 |
| MPV | rs56098529 | rs72932437 | C | A | 2 | CPS1 | Female only | True | 0.2546 | 0.2522 | 0.2479 | 0.2454 | 26726 | 19784 | 188072 | 162398 | -0.04866 | -0.00856 | -0.01941 | -0.007223 | 0.009023 | 0.01078 |
| MPV | rs56098529 | rs72932439 | T | C | 2 | CPS1 | Female only | True | 0.2546 | 0.2522 | 0.2479 | 0.2454 | 26726 | 19784 | 188072 | 162398 | -0.04866 | -0.00856 | -0.01941 | -0.007223 | 0.009023 | 0.01078 |
| MPV | rs56098529 | rs16844534 | G | A | 2 | CPS1 | Female only | True | 0.2546 | 0.2522 | 0.2478 | 0.2454 | 26726 | 19784 | 188072 | 162398 | -0.04866 | -0.00856 | -0.01941 | -0.007223 | 0.009023 | 0.01078 |
| MPV | rs56098529 | rs6725303 | A | C | 2 | CPS1 | Female only | True | 0.2572 | 0.2544 | 0.2487 | 0.2462 | 26726 | 19784 | 188072 | 162398 | -0.04888 | -0.00876 | -0.01984 | -0.007278 | 0.008993 | 0.01075 |
| MPV | rs56098529 | rs6725770 | A | G | 2 | CPS1 | Female only | True | 0.2546 | 0.2522 | 0.2478 | 0.2454 | 26726 | 19784 | 188072 | 162398 | -0.04866 | -0.00856 | -0.01941 | -0.007223 | 0.009023 | 0.01078 |
| MPV | rs56098529 | rs6725979 | T | C | 2 | CPS1 | Female only | True | 0.2546 | 0.2522 | 0.2478 | 0.2454 | 26726 | 19784 | 188072 | 162398 | -0.04866 | -0.00856 | -0.01941 | -0.007244 | 0.009023 | 0.01078 |
| MPV | rs56098529 | rs6729280 | A | C | 2 | CPS1 | Female only | True | 0.2546 | 0.2522 | 0.2478 | 0.2454 | 26726 | 19784 | 188072 | 162398 | -0.04866 | -0.008559 | -0.01941 | -0.007242 | 0.009023 | 0.01078 |
| MPV | rs56098529 | rs72932447 | A | G | 2 | CPS1 | Female only | True | 0.2546 | 0.2522 | 0.2478 | 0.2454 | 26726 | 19784 | 188072 | 162398 | -0.04866 | -0.008559 | -0.0194 | -0.007234 | 0.009023 | 0.01078 |
| MPV | rs56098529 | rs17822981 | G | A | 2 | CPS1 | Female only | True | 0.2564 | 0.254 | 0.2478 | 0.2454 | 26726 | 19784 | 188072 | 162398 | -0.0482 | -0.008227 | -0.0194 | -0.007219 | 0.008961 | 0.01071 |
| MPV | rs56098529 | rs10804184 | A | G | 2 | CPS1 | Female only | True | 0.2567 | 0.2538 | 0.2489 | 0.2464 | 26726 | 19784 | 188072 | 162398 | -0.04898 | -0.008678 | -0.01987 | -0.007015 | 0.008998 | 0.01076 |
| MPV | rs56098529 | rs10932337 | G | T | 2 | CPS1 | Female only | True | 0.3484 | 0.3443 | 0.3318 | 0.3301 | 26726 | 19784 | 188072 | 162398 | -0.02844 | -0.01243 | -0.01856 | -0.003381 | 0.008283 | 0.009908 |
| MPV | rs56098529 | rs72934320 | C | T | 2 | CPS1 | Female only | True | 0.255 | 0.2526 | 0.2489 | 0.2465 | 26726 | 19784 | 188072 | 162398 | -0.04832 | -0.008777 | -0.01897 | -0.006958 | 0.009031 | 0.01079 |
| MPV | rs56098529 | rs10191395 | A | G | 2 | CPS1 | Female only | True | 0.3739 | 0.373 | 0.365 | 0.3643 | 26726 | 19784 | 188072 | 162398 | -0.03199 | -0.01816 | -0.009425 | -0.009899 | 0.00818 | 0.009762 |
| MPV | rs56098529 | rs12619519 | A | G | 2 | CPS1 | Female only | True | 0.2547 | 0.2525 | 0.249 | 0.2466 | 26726 | 19784 | 188072 | 162398 | -0.0489 | -0.0084 | -0.01899 | -0.007024 | 0.009036 | 0.01079 |
| MPV | rs56098529 | rs150841467 | A | G | 2 | CPS1 | Female only | True | 0.2539 | 0.2511 | 0.2481 | 0.2456 | 26726 | 19784 | 188072 | 162398 | -0.04887 | -0.008498 | -0.01948 | -0.006995 | 0.009037 | 0.0108 |
| MPV | rs56098529 | rs78421188 | C | T | 2 | CPS1 | Female only | True | 0.2521 | 0.2495 | 0.2463 | 0.2439 | 26726 | 19784 | 188072 | 162398 | -0.04922 | -0.008306 | -0.01973 | -0.007137 | 0.009082 | 0.01085 |
| MPV | rs56098529 | rs56103208 | G | T | 2 | CPS1 | Female only | True | 0.254 | 0.2512 | 0.2482 | 0.2457 | 26726 | 19784 | 188072 | 162398 | -0.04878 | -0.008318 | -0.01938 | -0.007029 | 0.009035 | 0.0108 |
| MPV | rs56098529 | rs918233 | C | T | 2 | CPS1 | Female only | True | 0.3453 | 0.3406 | 0.3321 | 0.3303 | 26726 | 19784 | 188072 | 162398 | -0.02844 | -0.01225 | -0.01935 | -0.003232 | 0.008201 | 0.009809 |
| MPV | rs56098529 | rs918232 | A | G | 2 | CPS1 | Female only | True | 0.3506 | 0.346 | 0.3321 | 0.3302 | 26726 | 19784 | 188072 | 162398 | -0.02845 | -0.012 | -0.01932 | -0.003266 | 0.008265 | 0.009886 |
| MPV | rs56098529 | rs62203707 | T | G | 2 | CPS1 | Female only | True | 0.3506 | 0.346 | 0.3321 | 0.3303 | 26726 | 19784 | 188072 | 162398 | -0.02841 | -0.012 | -0.01936 | -0.003203 | 0.008265 | 0.009886 |
| MPV | rs56098529 | rs1588365 | A | G | 2 | CPS1 | Female only | True | 0.3506 | 0.346 | 0.3321 | 0.3302 | 26726 | 19784 | 188072 | 162398 | -0.02853 | -0.01215 | -0.01935 | -0.003138 | 0.008267 | 0.009886 |
| MPV | rs56098529 | rs72934339 | G | A | 2 | CPS1 | Female only | True | 0.2539 | 0.2512 | 0.2482 | 0.2458 | 26726 | 19784 | 188072 | 162398 | -0.04931 | -0.008561 | -0.01954 | -0.007002 | 0.009039 | 0.0108 |
| trait | lead_snp | ID | ALLELE1 | ALLELE0 | CHR | Gene_Symbol | independent_loci_in_sex_P1e2 | in_embl_any_or_gene | A1FREQ_VUMC_female | A1FREQ_VUMC_male | A1FREQ_UKB_female | A1FREQ_UKB_male | N_VUMC_female | N_VUMC_male | N_UKB_female | N_UKB_male | BETA_VUMC_female | BETA_VUMC_male | BETA_UKB_female | BETA_UKB_male | SE_VUMC_female | SE_VUMC_male |
| MPV | rs56098529 | rs13399079 | G | A | 2 | CPS1 | Female only | True | 0.3507 | 0.3462 | 0.3321 | 0.3303 | 26726 | 19784 | 188072 | 162398 | -0.0286 | -0.01229 | -0.01952 | -0.002922 | 0.008265 | 0.009884 |
| MPV | rs56098529 | rs7594883 | C | T | 2 | CPS1 | Female only | True | 0.2542 | 0.2514 | 0.2483 | 0.2459 | 26726 | 19784 | 188072 | 162398 | -0.04866 | -0.008769 | -0.01957 | -0.00699 | 0.009035 | 0.0108 |
| MPV | rs56098529 | rs3915695 | C | T | 2 | CPS1 | Female only | True | 0.3508 | 0.3464 | 0.3322 | 0.3304 | 26726 | 19784 | 188072 | 162398 | -0.02872 | -0.01217 | -0.01954 | -0.002901 | 0.008263 | 0.009884 |
| MPV | rs56098529 | rs4372818 | C | T | 2 | CPS1 | Female only | True | 0.2542 | 0.2514 | 0.2482 | 0.2458 | 26726 | 19784 | 188072 | 162398 | -0.04866 | -0.008868 | -0.01966 | -0.006905 | 0.009035 | 0.0108 |

| trait | lead_snp | ID | ALLELE1 | ALLELE0 | CHR | Gene_Symbol | independent_loci_in_sex_P1e2 | in_embl_any_or_gene | A1FREQ_VUMC_female | A1FREQ_VUMC_male | A1FREQ_UKB_female | A1FREQ_UKB_male | N_VUMC_female | N_VUMC_male | N_UKB_female | N_UKB_male | BETA_VUMC_female | BETA_VUMC_male | BETA_UKB_female | BETA_UKB_male | SE_VUMC_female | SE_VUMC_male |
|---|---|---|---|---|---|---|---|---|---|---|---|---|---|---|---|---|---|---|---|---|---|---|
| MPV | rs56098529 | rs3915694 | A | G | 2 | CPS1 | Female only | True | 0.3507 | 0.3462 | 0.3319 | 0.3301 | 26726 | 19784 | 188072 | 162398 | -0.02874 | -0.01256 | -0.01965 | -0.002694 | 0.008265 | 0.009886 |
| MPV | rs56098529 | rs3915693 | C | T | 2 | CPS1 | Female only | True | 0.3507 | 0.3462 | 0.3319 | 0.3301 | 26726 | 19784 | 188072 | 162398 | -0.02873 | -0.01256 | -0.01966 | -0.002692 | 0.008265 | 0.009886 |
| MPV | rs56098529 | rs1509819 | A | G | 2 | CPS1 | Female only | True | 0.3507 | 0.3462 | 0.3319 | 0.3302 | 26726 | 19784 | 188072 | 162398 | -0.02861 | -0.01205 | -0.01942 | -0.00304 | 0.008262 | 0.009883 |
| MPV | rs56098529 | rs59337803 | T | C | 2 | CPS1 | Female only | True | 0.3507 | 0.3462 | 0.3319 | 0.3301 | 26726 | 19784 | 188072 | 162398 | -0.0286 | -0.01205 | -0.01945 | -0.003048 | 0.008262 | 0.009883 |
| MPV | rs56098529 | rs13405091 | G | A | 2 | CPS1 | Female only | True | 0.3507 | 0.3462 | 0.332 | 0.3302 | 26726 | 19784 | 188072 | 162398 | -0.0286 | -0.01205 | -0.01941 | -0.003042 | 0.008262 | 0.009883 |
| MPV | rs56098529 | rs10165084 | A | C | 2 | CPS1 | Female only | True | 0.3507 | 0.3462 | 0.332 | 0.3302 | 26726 | 19784 | 188072 | 162398 | -0.02883 | -0.012 | -0.01951 | -0.00304 | 0.008262 | 0.009883 |
| MPV | rs56098529 | rs2664238 | T | C | 2 | CPS1 | Female only | True | 0.3506 | 0.346 | 0.3318 | 0.33 | 26726 | 19784 | 188072 | 162398 | -0.02849 | -0.012 | -0.01936 | -0.003019 | 0.008265 | 0.009886 |
| MPV | rs56098529 | rs2544312 | A | C | 2 | CPS1 | Female only | True | 0.3729 | 0.3724 | 0.3656 | 0.3648 | 26726 | 19784 | 188072 | 162398 | -0.033 | -0.01836 | -0.008448 | -0.009882 | 0.008171 | 0.009738 |
| MPV | rs56098529 | rs12611623 | A | G | 2 | CPS1 | Female only | True | 0.2535 | 0.2508 | 0.2479 | 0.2455 | 26726 | 19784 | 188072 | 162398 | -0.0498 | -0.008171 | -0.01941 | -0.006895 | 0.009048 | 0.01081 |
| MPV | rs56098529 | rs2544311 | G | A | 2 | CPS1 | Female only | True | 0.3727 | 0.3722 | 0.3656 | 0.3648 | 26726 | 19784 | 188072 | 162398 | -0.03302 | -0.01831 | -0.008472 | -0.009885 | 0.008174 | 0.009742 |
| MPV | rs56098529 | rs2664237 | A | G | 2 | CPS1 | Female only | True | 0.3785 | 0.3785 | 0.3708 | 0.3697 | 26726 | 19784 | 188072 | 162398 | -0.03244 | -0.017 | -0.008502 | -0.01025 | 0.00815 | 0.009715 |
| MPV | rs56098529 | rs2247029 | A | G | 2 | CPS1 | Female only | True | 0.3782 | 0.3782 | 0.3709 | 0.3699 | 26726 | 19784 | 188072 | 162398 | -0.03198 | -0.01742 | -0.008542 | -0.01011 | 0.008151 | 0.009713 |
| MPV | rs56098529 | rs2664236 | A | G | 2 | CPS1 | Female only | True | 0.3786 | 0.3785 | 0.371 | 0.37 | 26726 | 19784 | 188072 | 162398 | -0.03222 | -0.01752 | -0.008512 | -0.01011 | 0.00815 | 0.009714 |
| MPV | rs56098529 | rs12472153 | C | T | 2 | CPS1 | Female only | True | 0.3786 | 0.3785 | 0.371 | 0.37 | 26726 | 19784 | 188072 | 162398 | -0.03231 | -0.01721 | -0.008502 | -0.01011 | 0.00815 | 0.009713 |
| MPV | rs56098529 | rs981024 | A | G | 2 | CPS1 | Female only | True | 0.3812 | 0.381 | 0.3709 | 0.3699 | 26726 | 19784 | 188072 | 162398 | -0.03157 | -0.01672 | -0.008566 | -0.01015 | 0.008092 | 0.009645 |
| MPV | rs56098529 | rs56098529 | T | C | 2 | CPS1 | Female only | True | 0.2531 | 0.2505 | 0.2478 | 0.2454 | 26726 | 19784 | 188072 | 162398 | -0.04998 | -0.008428 | -0.01943 | -0.006938 | 0.009062 | 0.01083 |
| MPV | rs56098529 | rs2664232 | A | G | 2 | CPS1 | Female only | True | 0.3781 | 0.378 | 0.3709 | 0.3699 | 26726 | 19784 | 188072 | 162398 | -0.03202 | -0.01691 | -0.008527 | -0.01027 | 0.008154 | 0.009721 |
| MPV | rs56098529 | rs2544310 | G | A | 2 | CPS1 | Female only | True | 0.3778 | 0.3778 | 0.3708 | 0.3698 | 26726 | 19784 | 188072 | 162398 | -0.03193 | -0.01748 | -0.008493 | -0.0103 | 0.008153 | 0.00972 |
| MPV | rs56098529 | rs6711129 | A | G | 2 | CPS1 | Female only | True | 0.2535 | 0.2507 | 0.248 | 0.2456 | 26726 | 19784 | 188072 | 162398 | -0.04925 | -0.008968 | -0.01941 | -0.006916 | 0.009059 | 0.01083 |
| MPV | rs56098529 | rs2664229 | A | C | 2 | CPS1 | Female only | True | 0.3774 | 0.3774 | 0.3706 | 0.3696 | 26726 | 19784 | 188072 | 162398 | -0.03113 | -0.0174 | -0.008553 | -0.0104 | 0.008159 | 0.009728 |
| MPV | rs56098529 | rs2544308 | T | C | 2 | CPS1 | Female only | True | 0.3774 | 0.3774 | 0.3708 | 0.3698 | 26726 | 19784 | 188072 | 162398 | -0.03092 | -0.01781 | -0.00856 | -0.0103 | 0.008157 | 0.009726 |
| MPV | rs56098529 | rs2544307 | A | G | 2 | CPS1 | Female only | True | 0.3792 | 0.3791 | 0.3715 | 0.3704 | 26726 | 19784 | 188072 | 162398 | -0.03155 | -0.01697 | -0.008803 | -0.01027 | 0.008151 | 0.009717 |
| MPV | rs56098529 | rs2012564 | G | A | 2 | CPS1 | Female only | True | 0.3818 | 0.3816 | 0.3726 | 0.3716 | 26726 | 19784 | 188072 | 162398 | -0.03194 | -0.01685 | -0.009165 | -0.01029 | 0.008139 | 0.009711 |
| MPV | rs56098529 | rs2544306 | A | G | 2 | CPS1 | Female only | True | 0.3778 | 0.3777 | 0.3712 | 0.3702 | 26726 | 19784 | 188072 | 162398 | -0.03105 | -0.01731 | -0.008761 | -0.01038 | 0.008155 | 0.009724 |
| MPV | rs56098529 | rs2250976 | G | T | 2 | CPS1 | Female only | True | 0.385 | 0.3848 | 0.3725 | 0.3715 | 26726 | 19784 | 188072 | 162398 | -0.03141 | -0.01662 | -0.009156 | -0.01027 | 0.008073 | 0.009633 |
| MPV | rs56098529 | rs72934370 | T | C | 2 | CPS1 | Female only | True | 0.2543 | 0.2513 | 0.2489 | 0.2465 | 26726 | 19784 | 188072 | 162398 | -0.04972 | -0.008839 | -0.01915 | -0.006762 | 0.009051 | 0.01081 |
| MPV | rs56098529 | rs3914067 | T | C | 2 | CPS1 | Female only | True | 0.3783 | 0.378 | 0.3712 | 0.3702 | 26726 | 19784 | 188072 | 162398 | -0.03202 | -0.01743 | -0.00852 | -0.01029 | 0.00815 | 0.009721 |
| MPV | rs56098529 | rs11904031 | T | C | 2 | CPS1 | Female only | True | 0.3797 | 0.3796 | 0.3716 | 0.3706 | 26726 | 19784 | 188072 | 162398 | -0.03236 | -0.01706 | -0.008539 | -0.01021 | 0.008144 | 0.009714 |
| MPV | rs56098529 | rs10932342 | T | G | 2 | CPS1 | Female only | True | 0.3819 | 0.3816 | 0.3725 | 0.3715 | 26726 | 19784 | 188072 | 162398 | -0.03223 | -0.01692 | -0.00893 | -0.01031 | 0.008136 | 0.009714 |

| trait | lead_snp | ID | ALLELE1 | ALLELE0 | CHR | Gene_Symbol | independent_loci_in_sex_P1e2 | in_embl_any_or_gene | A1FREQ_VUMC_female | A1FREQ_VUMC_male | A1FREQ_UKB_female | A1FREQ_UKB_male | N_VUMC_female | N_VUMC_male | N_UKB_female | N_UKB_male | BETA_VUMC_female | BETA_VUMC_male | BETA_UKB_female | BETA_UKB_male | SE_VUMC_female | SE_VUMC_male |
|---|---|---|---|---|---|---|---|---|---|---|---|---|---|---|---|---|---|---|---|---|---|---|
| MPV | rs56098529 | rs12475210 | T | G | 2 | CPS1 | Female only | True | 0.3797 | 0.3795 | 0.3716 | 0.3705 | 26726 | 19784 | 188072 | 162398 | -0.03206 | -0.0174 | -0.008533 | -0.01026 | 0.008144 | 0.009711 |
| MPV | rs56098529 | rs7588072 | A | C | 2 | CPS1 | Female only | True | 0.3737 | 0.3726 | 0.3737 | 0.3715 | 26726 | 19784 | 188072 | 162398 | -0.03584 | 3.05e-04 | -0.02015 | -0.002785 | 0.008095 | 0.009678 |
| MPV | rs56098529 | rs11684318 | T | C | 2 | CPS1 | Female only | True | 0.3737 | 0.3726 | 0.3736 | 0.3714 | 26726 | 19784 | 188072 | 162398 | -0.03598 | 1.33e-04 | -0.02013 | -0.002674 | 0.008096 | 0.009679 |
| MPV | rs56098529 | rs9288418 | A | G | 2 | CPS1 | Female only | True | 0.3736 | 0.3726 | 0.3736 | 0.3714 | 26726 | 19784 | 188072 | 162398 | -0.03594 | 1.10e-04 | -0.02017 | -0.002867 | 0.008097 | 0.009681 |
| MPV | rs2810490 | rs441233 | A | G | 9 | BRD3 | Female only | True | 0.3046 | 0.3014 | 0.291 | 0.2904 | 26726 | 19784 | 188072 | 162398 | -0.04895 | -0.0202 | -0.04855 | -0.0323 | 0.008498 | 0.01018 |
| MPV | rs2810490 | rs10993894 | T | C | 9 | BRD3 | Female only | True | 0.2792 | 0.276 | 0.264 | 0.2632 | 26726 | 19784 | 188072 | 162398 | -0.05265 | -0.02312 | -0.04995 | -0.03427 | 0.008761 | 0.01049 |
| MPV | rs2810490 | rs3094339 | A | G | 9 | BRD3 | Female only | True | 0.2791 | 0.276 | 0.2639 | 0.2631 | 26726 | 19784 | 188072 | 162398 | -0.0527 | -0.0231 | -0.05001 | -0.0343 | 0.008761 | 0.01049 |
| MPV | rs2810490 | rs448918 | A | G | 9 | BRD3 | Female only | True | 0.2799 | 0.2768 | 0.2643 | 0.2635 | 26726 | 19784 | 188072 | 162398 | -0.05327 | -0.02347 | -0.04996 | -0.03447 | 0.008739 | 0.01048 |
| MPV | rs2810490 | rs10993895 | C | T | 9 | BRD3 | Female only | True | 0.2795 | 0.2766 | 0.2636 | 0.2627 | 26726 | 19784 | 188072 | 162398 | -0.05293 | -0.02167 | -0.04961 | -0.0348 | 0.008738 | 0.01047 |
| MPV | rs2810490 | rs379471 | A | G | 9 | BRD3 | Female only | True | 0.2806 | 0.2778 | 0.265 | 0.2641 | 26726 | 19784 | 188072 | 162398 | -0.05376 | -0.02199 | -0.04937 | -0.03502 | 0.008728 | 0.01045 |
| MPV | rs2810490 | rs379417 | A | G | 9 | BRD3 | Female only | True | 0.2802 | 0.2774 | 0.2649 | 0.264 | 26726 | 19784 | 188072 | 162398 | -0.05334 | -0.02258 | -0.0496 | -0.03504 | 0.008731 | 0.01046 |
| MPV | rs2810490 | rs378740 | A | C | 9 | BRD3 | Female only | True | 0.2877 | 0.2844 | 0.2719 | 0.2708 | 26726 | 19784 | 188072 | 162398 | -0.05174 | -0.02125 | -0.04765 | -0.03337 | 0.008708 | 0.01042 |
| MPV | rs2810490 | rs417142 | T | G | 9 | BRD3 | Female only | True | 0.282 | 0.2793 | 0.265 | 0.2641 | 26726 | 19784 | 188072 | 162398 | -0.05342 | -0.02171 | -0.04939 | -0.03502 | 0.008678 | 0.01039 |
| MPV | rs2810490 | rs408307 | T | C | 9 | BRD3 | Female only | True | 0.2805 | 0.2776 | 0.2647 | 0.2638 | 26726 | 19784 | 188072 | 162398 | -0.05361 | -0.02319 | -0.05014 | -0.03506 | 0.008737 | 0.01047 |
| MPV | rs2810490 | rs410876 | A | C | 9 | BRD3 | Female only | True | 0.2811 | 0.2783 | 0.2657 | 0.2647 | 26726 | 19784 | 188072 | 162398 | -0.05308 | -0.02143 | -0.04943 | -0.03476 | 0.008729 | 0.01044 |
| MPV | rs2810490 | rs433402 | G | A | 9 | BRD3 | Female only | True | 0.2809 | 0.2782 | 0.2661 | 0.265 | 26726 | 19784 | 188072 | 162398 | -0.05366 | -0.02185 | -0.04952 | -0.03479 | 0.008735 | 0.01045 |
| MPV | rs2810490 | rs467303 | T | C | 9 | BRD3 | Female only | True | 0.2807 | 0.2778 | 0.2651 | 0.2643 | 26726 | 19784 | 188072 | 162398 | -0.05458 | -0.02165 | -0.04981 | -0.03533 | 0.008732 | 0.01044 |
| MPV | rs2810490 | rs455381 | A | C | 9 | BRD3 | Female only | True | 0.2807 | 0.2778 | 0.2651 | 0.2642 | 26726 | 19784 | 188072 | 162398 | -0.0546 | -0.02166 | -0.04969 | -0.03536 | 0.008732 | 0.01044 |
| MPV | rs2810490 | rs467387 | A | G | 9 | BRD3 | Female only | True | 0.2776 | 0.2745 | 0.2621 | 0.2617 | 26726 | 19784 | 188072 | 162398 | -0.05452 | -0.02208 | -0.05 | -0.03571 | 0.008762 | 0.0105 |
| MPV | rs2810490 | rs392134 | A | G | 9 | BRD3 | Female only | True | 0.2777 | 0.2744 | 0.2624 | 0.2618 | 26726 | 19784 | 188072 | 162398 | -0.05318 | -0.02091 | -0.04988 | -0.03481 | 0.008747 | 0.01048 |
| MPV | rs2810490 | rs460154 | A | G | 9 | BRD3 | Female only | True | 0.2836 | 0.2803 | 0.265 | 0.2642 | 26726 | 19784 | 188072 | 162398 | -0.05392 | -0.02049 | -0.04951 | -0.03545 | 0.008664 | 0.01036 |
| MPV | rs2810490 | rs465724 | T | C | 9 | BRD3 | Female only | True | 0.2829 | 0.2795 | 0.2652 | 0.2644 | 26726 | 19784 | 188072 | 162398 | -0.05426 | -0.0211 | -0.04947 | -0.03511 | 0.008703 | 0.01041 |
| MPV | rs2810490 | rs465535 | T | C | 9 | BRD3 | Female only | True | 0.2825 | 0.2792 | 0.2653 | 0.2645 | 26726 | 19784 | 188072 | 162398 | -0.05406 | -0.02127 | -0.04955 | -0.03544 | 0.008705 | 0.01041 |
| MPV | rs2810490 | rs456207 | A | G | 9 | BRD3 | Female only | True | 0.2818 | 0.2787 | 0.2648 | 0.264 | 26726 | 19784 | 188072 | 162398 | -0.05443 | -0.02121 | -0.04977 | -0.03508 | 0.00872 | 0.01043 |
| MPV | rs2810490 | rs456205 | A | G | 9 | BRD3 | Female only | True | 0.2765 | 0.2733 | 0.261 | 0.2605 | 26726 | 19784 | 188072 | 162398 | -0.05418 | -0.02123 | -0.05037 | -0.03484 | 0.008767 | 0.0105 |
| MPV | rs2810490 | rs461462 | T | C | 9 | BRD3 | Female only | True | 0.2824 | 0.2792 | 0.2652 | 0.2644 | 26726 | 19784 | 188072 | 162398 | -0.05423 | -0.02115 | -0.04954 | -0.03535 | 0.008705 | 0.01041 |
| MPV | rs2810490 | rs459571 | T | C | 9 | BRD3 | Female only | True | 0.2743 | 0.271 | 0.2595 | 0.259 | 26726 | 19784 | 188072 | 162398 | -0.05339 | -0.01983 | -0.04974 | -0.03464 | 0.008821 | 0.01057 |

| trait | lead_snp | ID | ALLELE1 | ALLELE0 | CHR | Gene_Symbol | independent_loci_in_sex_P1e2 | in_embl_any_or_gene | A1FREQ_VUMC_female | A1FREQ_VUMC_male | A1FREQ_UKB_female | A1FREQ_UKB_male | N_VUMC_female | N_VUMC_male | N_UKB_female | N_UKB_male | BETA_VUMC_female | BETA_VUMC_male | BETA_UKB_female | BETA_UKB_male | SE_VUMC_female | SE_VUMC_male |
|---|---|---|---|---|---|---|---|---|---|---|---|---|---|---|---|---|---|---|---|---|---|---|
| MPV | rs2810490 | rs2073818 | A | C | 9 | BRD3 | Female only | True | 0.2783 | 0.275 | 0.2625 | 0.262 | 26726 | 19784 | 188072 | 162398 | -0.05338 | -0.02118 | -0.04986 | -0.03525 | 0.008748 | 0.01048 |
| MPV | rs2810490 | rs2520095 | G | A | 9 | BRD3 | Female only | True | 0.2781 | 0.2747 | 0.2626 | 0.2622 | 26726 | 19784 | 188072 | 162398 | -0.05385 | -0.02119 | -0.04964 | -0.03515 | 0.008749 | 0.01049 |
| MPV | rs2810490 | rs2810489 | A | G | 9 | BRD3 | Female only | True | 0.2718 | 0.269 | 0.2573 | 0.2563 | 26726 | 19784 | 188072 | 162398 | -0.05154 | -0.02003 | -0.04903 | -0.03306 | 0.008834 | 0.01058 |
| MPV | rs2810490 | rs2810490 | A | G | 9 | BRD3 | Female only | True | 0.2994 | 0.2972 | 0.2836 | 0.2836 | 26726 | 19784 | 188072 | 162398 | -0.0561 | -0.02357 | -0.04678 | -0.0349 | 0.00855 | 0.01019 |
| MPV | rs2810490 | rs2810494 | A | G | 9 | BRD3 | Female only | True | 0.3643 | 0.3618 | 0.3484 | 0.3475 | 26726 | 19784 | 188072 | 162398 | -0.04157 | -0.01964 | -0.03202 | -0.0233 | 0.00809 | 0.009717 |
| MPV | rs12773283 | rs4545434 | C | T | 10 | JMJD1C | Female only | True | 0.1848 | 0.183 | 0.1788 | 0.1798 | 26726 | 19784 | 188072 | 162398 | 0.04555 | 0.007489 | 0.03405 | 0.02441 | 0.01008 | 0.01209 |
| MPV | rs12773283 | rs7084396 | A | G | 10 | JMJD1C | Female only | True | 0.2024 | 0.1999 | 0.1847 | 0.1859 | 26726 | 19784 | 188072 | 162398 | 0.04643 | 0.008438 | 0.03517 | 0.02545 | 0.009851 | 0.01186 |
| MPV | rs12773283 | rs10822138 | G | A | 10 | JMJD1C | Female only | True | 0.2024 | 0.2 | 0.1848 | 0.1859 | 26726 | 19784 | 188072 | 162398 | 0.04654 | 0.00833 | 0.03512 | 0.02545 | 0.00985 | 0.01185 |
| MPV | rs12773283 | rs1396964 | G | A | 10 | JMJD1C | Female only | True | 0.2024 | 0.2002 | 0.1848 | 0.186 | 26726 | 19784 | 188072 | 162398 | 0.04635 | 0.008423 | 0.03503 | 0.02568 | 0.009852 | 0.01185 |
| MPV | rs12773283 | rs6479878 | A | G | 10 | JMJD1C | Female only | True | 0.202 | 0.1997 | 0.1839 | 0.185 | 26726 | 19784 | 188072 | 162398 | 0.04596 | 0.008722 | 0.03524 | 0.02578 | 0.009862 | 0.01186 |
| MPV | rs12773283 | rs6479879 | G | T | 10 | JMJD1C | Female only | True | 0.2021 | 0.1998 | 0.1839 | 0.1851 | 26726 | 19784 | 188072 | 162398 | 0.04616 | 0.008614 | 0.03517 | 0.02573 | 0.00986 | 0.01186 |
| MPV | rs12773283 | rs1553789 | C | T | 10 | JMJD1C | Female only | True | 0.2304 | 0.2288 | 0.2154 | 0.2164 | 26726 | 19784 | 188072 | 162398 | 0.0477 | 0.01491 | 0.0413 | 0.03435 | 0.009391 | 0.01124 |
| MPV | rs12773283 | rs10995447 | A | G | 10 | JMJD1C | Female only | True | 0.1849 | 0.1832 | 0.1789 | 0.1799 | 26726 | 19784 | 188072 | 162398 | 0.04534 | 0.007275 | 0.03411 | 0.02476 | 0.01007 | 0.01209 |
| MPV | rs12773283 | rs4601646 | G | A | 10 | JMJD1C | Female only | True | 0.2304 | 0.2288 | 0.2155 | 0.2164 | 26726 | 19784 | 188072 | 162398 | 0.04769 | 0.01502 | 0.0413 | 0.03441 | 0.009391 | 0.01124 |
| MPV | rs12773283 | rs4746951 | G | A | 10 | JMJD1C | Female only | True | 0.18 | 0.1782 | 0.1746 | 0.1757 | 26726 | 19784 | 188072 | 162398 | 0.04222 | 0.005206 | 0.03344 | 0.0234 | 0.01019 | 0.01223 |
| MPV | rs12773283 | rs61853335 | T | C | 10 | JMJD1C | Female only | True | 0.1795 | 0.1779 | 0.1743 | 0.1754 | 26726 | 19784 | 188072 | 162398 | 0.04281 | 0.004606 | 0.03349 | 0.02364 | 0.01021 | 0.01224 |
| MPV | rs12773283 | rs72824948 | G | A | 10 | JMJD1C | Female only | True | 0.1469 | 0.1467 | 0.1423 | 0.1432 | 26726 | 19784 | 188072 | 162398 | 0.06759 | 0.02509 | 0.06046 | 0.05057 | 0.01111 | 0.01326 |
| MPV | rs12773283 | rs4746974 | T | G | 10 | JMJD1C | Female only | True | 0.1467 | 0.1464 | 0.1424 | 0.1433 | 26726 | 19784 | 188072 | 162398 | 0.06721 | 0.02609 | 0.06055 | 0.0506 | 0.01111 | 0.01328 |
| MPV | rs12773283 | rs868798 | G | A | 10 | JMJD1C | Female only | True | 0.1466 | 0.1464 | 0.1424 | 0.1433 | 26726 | 19784 | 188072 | 162398 | 0.06742 | 0.02579 | 0.06047 | 0.0501 | 0.01111 | 0.01328 |
| MPV | rs12773283 | rs61853336 | A | G | 10 | JMJD1C | Female only | True | 0.1466 | 0.1464 | 0.1424 | 0.1433 | 26726 | 19784 | 188072 | 162398 | 0.06742 | 0.02579 | 0.06048 | 0.05009 | 0.01111 | 0.01328 |
| MPV | rs12773283 | rs12415984 | A | G | 10 | JMJD1C | Female only | True | 0.1467 | 0.1463 | 0.1424 | 0.1433 | 26726 | 19784 | 188072 | 162398 | 0.06703 | 0.02625 | 0.06046 | 0.05009 | 0.01109 | 0.01325 |
| MPV | rs12773283 | rs11814792 | T | C | 10 | JMJD1C | Female only | True | 0.1467 | 0.1463 | 0.1422 | 0.1432 | 26726 | 19784 | 188072 | 162398 | 0.06688 | 0.02618 | 0.06039 | 0.04998 | 0.01109 | 0.01325 |
| MPV | rs12773283 | rs61853339 | C | T | 10 | JMJD1C | Female only | True | 0.1475 | 0.1469 | 0.1432 | 0.1443 | 26726 | 19784 | 188072 | 162398 | 0.06748 | 0.0274 | 0.06004 | 0.04928 | 0.01107 | 0.01323 |
| MPV | rs12773283 | rs12413415 | C | T | 10 | JMJD1C | Female only | True | 0.1465 | 0.1462 | 0.1422 | 0.1432 | 26726 | 19784 | 188072 | 162398 | 0.06782 | 0.02628 | 0.06038 | 0.04993 | 0.01109 | 0.01325 |
| MPV | rs12773283 | rs11817689 | C | T | 10 | JMJD1C | Female only | True | 0.1464 | 0.146 | 0.1422 | 0.1432 | 26726 | 19784 | 188072 | 162398 | 0.06792 | 0.02607 | 0.0604 | 0.04988 | 0.01109 | 0.01325 |
| MPV | rs12773283 | rs4747045 | T | C | 10 | JMJD1C | Female only | True | 0.1464 | 0.1461 | 0.1423 | 0.1432 | 26726 | 19784 | 188072 | 162398 | 0.06815 | 0.02581 | 0.06029 | 0.04998 | 0.01109 | 0.01325 |
| MPV | rs12773283 | rs72829170 | G | A | 10 | JMJD1C | Female only | True | 0.1463 | 0.1459 | 0.1423 | 0.1432 | 26726 | 19784 | 188072 | 162398 | 0.06771 | 0.02622 | 0.0604 | 0.04974 | 0.01109 | 0.01325 |
| MPV | rs12773283 | rs7099425 | C | T | 10 | JMJD1C | Female only | True | 0.1517 | 0.151 | 0.1423 | 0.1433 | 26726 | 19784 | 188072 | 162398 | 0.0671 | 0.02652 | 0.06058 | 0.04969 | 0.01104 | 0.01321 |
| MPV | rs12773283 | rs12413730 | T | C | 10 | JMJD1C | Female only | True | 0.1461 | 0.1457 | 0.1422 | 0.1432 | 26726 | 19784 | 188072 | 162398 | 0.06809 | 0.02711 | 0.06053 | 0.04978 | 0.01107 | 0.01323 |

| trait | lead_snp | ID | ALLELE1 | ALLELE0 | CHR | Gene_Symbol | independent_loci_in_sex_P1e2 | in_embl_any_or_gene | A1FREQ_VUMC_female | A1FREQ_VUMC_male | A1FREQ_UKB_female | A1FREQ_UKB_male | N_VUMC_female | N_VUMC_male | N_UKB_female | N_UKB_male | BETA_VUMC_female | BETA_VUMC_male | BETA_UKB_female | BETA_UKB_male | SE_VUMC_female | SE_VUMC_male |
|---|---|---|---|---|---|---|---|---|---|---|---|---|---|---|---|---|---|---|---|---|---|---|
| MPV | rs12773283 | rs11819167 | A | G | 10 | JMJD1C | Female only | True | 0.1462 | 0.1458 | 0.1422 | 0.1432 | 26726 | 19784 | 188072 | 162398 | 0.0681 | 0.0273 | 0.0605 | 0.04975 | 0.01107 | 0.01323 |
| MPV | rs12773283 | rs1904295 | G | A | 10 | JMJD1C | Female only | True | 0.1511 | 0.1508 | 0.1422 | 0.1432 | 26726 | 19784 | 188072 | 162398 | 0.06729 | 0.02681 | 0.06062 | 0.04972 | 0.01105 | 0.01321 |
| MPV | rs12773283 | rs6415875 | A | G | 10 | JMJD1C | Female only | True | 0.1511 | 0.1508 | 0.1422 | 0.1432 | 26726 | 19784 | 188072 | 162398 | 0.06731 | 0.02689 | 0.06059 | 0.04971 | 0.01105 | 0.01321 |
| MPV | rs12773283 | rs4281380 | C | A | 10 | JMJD1C | Female only | True | 0.1511 | 0.1508 | 0.1423 | 0.1432 | 26726 | 19784 | 188072 | 162398 | 0.06744 | 0.02704 | 0.06046 | 0.0498 | 0.01105 | 0.01321 |
| MPV | rs12773283 | rs7082090 | A | G | 10 | JMJD1C | Female only | True | 0.1511 | 0.1507 | 0.1422 | 0.1432 | 26726 | 19784 | 188072 | 162398 | 0.06738 | 0.02698 | 0.06061 | 0.04968 | 0.01104 | 0.01321 |
| MPV | rs12773283 | rs9414780 | A | C | 10 | JMJD1C | Female only | True | 0.151 | 0.1507 | 0.1422 | 0.1432 | 26726 | 19784 | 188072 | 162398 | 0.06757 | 0.027 | 0.06064 | 0.04971 | 0.01105 | 0.01321 |
| MPV | rs12773283 | rs6479891 | T | C | 10 | JMJD1C | Female only | True | 0.1471 | 0.1467 | 0.1422 | 0.1432 | 26726 | 19784 | 188072 | 162398 | 0.06767 | 0.02715 | 0.06044 | 0.04973 | 0.011 | 0.01315 |
| MPV | rs12773283 | rs10761730 | C | T | 10 | JMJD1C | Female only | True | 0.1509 | 0.1505 | 0.1422 | 0.1432 | 26726 | 19784 | 188072 | 162398 | 0.06748 | 0.02685 | 0.06052 | 0.04981 | 0.01104 | 0.0132 |
| MPV | rs12773283 | rs9415680 | A | G | 10 | JMJD1C | Female only | True | 0.1509 | 0.1505 | 0.1422 | 0.1432 | 26726 | 19784 | 188072 | 162398 | 0.06746 | 0.02687 | 0.06054 | 0.04988 | 0.01104 | 0.0132 |
| MPV | rs12773283 | rs9415682 | G | T | 10 | JMJD1C | Female only | True | 0.1508 | 0.1505 | 0.1422 | 0.1432 | 26726 | 19784 | 188072 | 162398 | 0.06742 | 0.02688 | 0.06051 | 0.0498 | 0.01104 | 0.0132 |
| MPV | rs12773283 | rs10740110 | A | G | 10 | JMJD1C | Female only | True | 0.1512 | 0.1506 | 0.1423 | 0.1433 | 26726 | 19784 | 188072 | 162398 | 0.06733 | 0.02693 | 0.06049 | 0.04986 | 0.01103 | 0.0132 |
| MPV | rs12773283 | rs10761735 | A | G | 10 | JMJD1C | Female only | True | 0.1508 | 0.1505 | 0.1423 | 0.1433 | 26726 | 19784 | 188072 | 162398 | 0.06743 | 0.02704 | 0.06056 | 0.04978 | 0.01104 | 0.0132 |
| MPV | rs12773283 | rs6479892 | A | G | 10 | JMJD1C | Female only | True | 0.1508 | 0.1504 | 0.1422 | 0.1432 | 26726 | 19784 | 188072 | 162398 | 0.06743 | 0.02691 | 0.06044 | 0.04982 | 0.01104 | 0.0132 |
| MPV | rs12773283 | rs9414788 | A | G | 10 | JMJD1C | Female only | True | 0.1508 | 0.1504 | 0.1423 | 0.1433 | 26726 | 19784 | 188072 | 162398 | 0.06726 | 0.02687 | 0.06044 | 0.04977 | 0.01104 | 0.0132 |
| MPV | rs12773283 | rs12773283 | C | T | 10 | JMJD1C | Female only | True | 0.1536 | 0.1526 | 0.1473 | 0.1482 | 26726 | 19784 | 188072 | 162398 | 0.06749 | 0.02954 | 0.06103 | 0.05148 | 0.0108 | 0.01293 |
| MPV | rs12773283 | rs7920768 | C | T | 10 | JMJD1C | Female only | True | 0.1508 | 0.1504 | 0.1422 | 0.1432 | 26726 | 19784 | 188072 | 162398 | 0.06716 | 0.02658 | 0.06053 | 0.04986 | 0.01104 | 0.0132 |
| MPV | rs12773283 | rs7100372 | T | C | 10 | JMJD1C | Female only | True | 0.1508 | 0.1503 | 0.1422 | 0.1432 | 26726 | 19784 | 188072 | 162398 | 0.06753 | 0.02689 | 0.06042 | 0.0498 | 0.01104 | 0.0132 |
| MPV | rs12773283 | rs7085869 | C | T | 10 | JMJD1C | Female only | True | 0.1509 | 0.1504 | 0.1422 | 0.1432 | 26726 | 19784 | 188072 | 162398 | 0.06675 | 0.02664 | 0.06047 | 0.04986 | 0.01103 | 0.0132 |
| MPV | rs12773283 | rs9299455 | A | G | 10 | JMJD1C | Female only | True | 0.1508 | 0.1504 | 0.1422 | 0.1432 | 26726 | 19784 | 188072 | 162398 | 0.06665 | 0.02663 | 0.0603 | 0.04978 | 0.01103 | 0.0132 |
| MPV | rs12773283 | rs9414793 | G | A | 10 | JMJD1C | Female only | True | 0.1508 | 0.1504 | 0.1422 | 0.1432 | 26726 | 19784 | 188072 | 162398 | 0.06664 | 0.02667 | 0.0603 | 0.04982 | 0.01103 | 0.0132 |
| MPV | rs12773283 | rs7899715 | T | C | 10 | JMJD1C | Female only | True | 0.151 | 0.1505 | 0.1424 | 0.1434 | 26726 | 19784 | 188072 | 162398 | 0.06667 | 0.02655 | 0.06018 | 0.04977 | 0.01103 | 0.0132 |
| MPV | rs12773283 | rs4525102 | C | T | 10 | JMJD1C | Female only | True | 0.1509 | 0.1505 | 0.1423 | 0.1432 | 26726 | 19784 | 188072 | 162398 | 0.06672 | 0.02656 | 0.06035 | 0.04978 | 0.01103 | 0.0132 |
| MPV | rs12773283 | rs7092504 | A | G | 10 | JMJD1C | Female only | True | 0.151 | 0.1505 | 0.1423 | 0.1432 | 26726 | 19784 | 188072 | 162398 | 0.06688 | 0.02669 | 0.06034 | 0.04978 | 0.01103 | 0.0132 |
| MPV | rs12773283 | rs4133250 | G | A | 10 | JMJD1C | Female only | True | 0.1512 | 0.1507 | 0.1422 | 0.1432 | 26726 | 19784 | 188072 | 162398 | 0.06679 | 0.02655 | 0.06027 | 0.04978 | 0.01104 | 0.0132 |
| MPV | rs12773283 | rs10995506 | A | G | 10 | JMJD1C | Female only | True | 0.1511 | 0.1507 | 0.1422 | 0.1432 | 26726 | 19784 | 188072 | 162398 | 0.06655 | 0.02642 | 0.06025 | 0.04978 | 0.01104 | 0.0132 |
| MPV | rs12773283 | rs10740117 | G | A | 10 | JMJD1C | Female only | True | 0.1512 | 0.1507 | 0.1422 | 0.1432 | 26726 | 19784 | 188072 | 162398 | 0.06639 | 0.0263 | 0.06031 | 0.04978 | 0.01104 | 0.0132 |
| MPV | rs12773283 | rs6479894 | T | C | 10 | JMJD1C | Female only | True | 0.1512 | 0.1507 | 0.1422 | 0.1432 | 26726 | 19784 | 188072 | 162398 | 0.06639 | 0.0263 | 0.06031 | 0.04978 | 0.01104 | 0.0132 |
| MPV | rs12773283 | rs7091664 | G | A | 10 | JMJD1C | Female only | True | 0.1513 | 0.1508 | 0.1422 | 0.1432 | 26726 | 19784 | 188072 | 162398 | 0.0664 | 0.02616 | 0.06031 | 0.04977 | 0.01104 | 0.0132 |
| MPV | rs12773283 | rs7081275 | G | T | 10 | JMJD1C | Female only | True | 0.1514 | 0.1508 | 0.1427 | 0.1436 | 26726 | 19784 | 188072 | 162398 | 0.06634 | 0.02619 | 0.06003 | 0.04971 | 0.01104 | 0.0132 |

| trait | lead_snp | ID | ALLELE1 | ALLELE0 | CHR | Gene_Symbol | independent_loci_in_sex_P1e2 | in_embl_any_or_gene | A1FREQ_VUMC_female | A1FREQ_VUMC_male | A1FREQ_UKB_female | A1FREQ_UKB_male | N_VUMC_female | N_VUMC_male | N_UKB_female | N_UKB_male | BETA_VUMC_female | BETA_VUMC_male | BETA_UKB_female | BETA_UKB_male | SE_VUMC_female | SE_VUMC_male |
|---|---|---|---|---|---|---|---|---|---|---|---|---|---|---|---|---|---|---|---|---|---|---|
| MPV | rs58684387 | rs77775794 | T | G | 12 | TMEM120B | Female only | True | 0.1164 | 0.1196 | 0.1181 | 0.1179 | 26726 | 19784 | 188072 | 162398 | -0.05279 | -0.03694 | -0.06757 | -0.04485 | 0.01233 | 0.01451 |
| MPV | rs58684387 | rs75405518 | T | C | 12 | TMEM120B | Female only | True | 0.1166 | 0.1197 | 0.1181 | 0.1179 | 26726 | 19784 | 188072 | 162398 | -0.05247 | -0.03731 | -0.06761 | -0.04489 | 0.01233 | 0.0145 |
| trait | lead_snp | ID | ALLELE1 | ALLELE0 | CHR | Gene_Symbol | independent_loci_in_sex_P1e2 | in_embl_any_or_gene | A1FREQ_VUMC_female | A1FREQ_VUMC_male | A1FREQ_UKB_female | A1FREQ_UKB_male | N_VUMC_female | N_VUMC_male | N_UKB_female | N_UKB_male | BETA_VUMC_female | BETA_VUMC_male | BETA_UKB_female | BETA_UKB_male | SE_VUMC_female | SE_VUMC_male |
| MPV | rs58684387 | rs111327817 | A | G | 12 | TMEM120B | Female only | True | 0.1637 | 0.1657 | 0.163 | 0.1634 | 26726 | 19784 | 188072 | 162398 | -0.06382 | -0.02919 | -0.0755 | -0.05337 | 0.01076 | 0.01275 |
| MPV | rs58684387 | rs58684387 | T | C | 12 | TMEM120B | Female only | True | 0.1627 | 0.165 | 0.162 | 0.1622 | 26726 | 19784 | 188072 | 162398 | -0.06499 | -0.02915 | -0.07544 | -0.05287 | 0.0108 | 0.01281 |
| MPV | rs2620340 | rs2620340 | G | A | 12 | HPD | Female only | False | 0.1383 | 0.1393 | 0.1307 | 0.1294 | 26726 | 19784 | 188072 | 162398 | -0.07489 | -0.0259 | -0.07898 | -0.0659 | 0.01174 | 0.01387 |
| MPV | rs2620340 | rs2620332 | T | C | 12 | HPD | Female only | False | 0.1383 | 0.1393 | 0.1307 | 0.1294 | 26726 | 19784 | 188072 | 162398 | -0.07408 | -0.02576 | -0.07885 | -0.06607 | 0.01171 | 0.01383 |
| MPV | rs2620340 | rs2596144 | T | C | 12 | HPD | Female only | False | 0.1331 | 0.1341 | 0.1307 | 0.1294 | 26726 | 19784 | 188072 | 162398 | -0.07342 | -0.025 | -0.07891 | -0.06607 | 0.01164 | 0.01375 |
| MPV | rs2620340 | rs9668191 | A | G | 12 | HPD | Female only | False | 0.1383 | 0.1393 | 0.1307 | 0.1294 | 26726 | 19784 | 188072 | 162398 | -0.07416 | -0.02572 | -0.07882 | -0.066 | 0.0117 | 0.01382 |
| MPV | rs2620340 | rs1947826 | T | C | 12 | HPD | Female only | False | 0.1381 | 0.139 | 0.1308 | 0.1295 | 26726 | 19784 | 188072 | 162398 | -0.07414 | -0.02615 | -0.07876 | -0.066 | 0.01169 | 0.01382 |
| MPV | rs2620340 | rs2101017 | C | T | 12 | HPD | Female only | False | 0.1381 | 0.139 | 0.1308 | 0.1295 | 26726 | 19784 | 188072 | 162398 | -0.07414 | -0.02615 | -0.07876 | -0.06599 | 0.01169 | 0.01382 |
| MPV | rs2620340 | rs4760099 | A | G | 12 | HPD | Female only | False | 0.133 | 0.1336 | 0.1262 | 0.1248 | 26726 | 19784 | 188072 | 162398 | -0.07356 | -0.02564 | -0.07935 | -0.06726 | 0.01187 | 0.01404 |
| MPV | rs2620340 | rs895959 | C | T | 12 | HPD | Female only | False | 0.1334 | 0.1338 | 0.1262 | 0.1248 | 26726 | 19784 | 188072 | 162398 | -0.07276 | -0.02578 | -0.0792 | -0.06736 | 0.01186 | 0.01403 |
| MPV | rs2620340 | rs2230681 | T | C | 12 | HPD | Female only | False | 0.133 | 0.1334 | 0.1262 | 0.1248 | 26726 | 19784 | 188072 | 162398 | -0.07365 | -0.02527 | -0.07915 | -0.06708 | 0.01187 | 0.01404 |
| MPV | rs2620340 | rs2596137 | C | T | 12 | HPD | Female only | False | 0.136 | 0.1367 | 0.1304 | 0.129 | 26726 | 19784 | 188072 | 162398 | -0.07393 | -0.02785 | -0.07823 | -0.06578 | 0.01174 | 0.01392 |
| MPV | rs2620340 | rs1678967 | A | G | 12 | HPD | Female only | False | 0.1307 | 0.1311 | 0.1247 | 0.1234 | 26726 | 19784 | 188072 | 162398 | -0.071 | -0.02337 | -0.07802 | -0.06753 | 0.01196 | 0.01416 |
| MPV | rs2620340 | rs1720067 | T | C | 12 | HPD | Female only | False | 0.131 | 0.1313 | 0.1248 | 0.1235 | 26726 | 19784 | 188072 | 162398 | -0.07118 | -0.02308 | -0.07826 | -0.06793 | 0.01197 | 0.01417 |
| MPV | rs2620340 | rs1667583 | T | C | 12 | HPD | Female only | False | 0.1264 | 0.1268 | 0.1246 | 0.1233 | 26726 | 19784 | 188072 | 162398 | -0.07013 | -0.02243 | -0.07805 | -0.06774 | 0.0119 | 0.01409 |
| MPV | rs2620340 | rs1678969 | T | C | 12 | HPD | Female only | False | 0.1309 | 0.1313 | 0.1247 | 0.1234 | 26726 | 19784 | 188072 | 162398 | -0.07104 | -0.02338 | -0.07812 | -0.06783 | 0.01197 | 0.01417 |
| MPV | rs2620340 | rs1667580 | G | A | 12 | HPD | Female only | False | 0.131 | 0.1315 | 0.1246 | 0.1233 | 26726 | 19784 | 188072 | 162398 | -0.07059 | -0.02328 | -0.0781 | -0.06766 | 0.01197 | 0.01417 |
| MPV | rs2620340 | rs895958 | G | A | 12 | HPD | Female only | False | 0.131 | 0.1315 | 0.1246 | 0.1233 | 26726 | 19784 | 188072 | 162398 | -0.07064 | -0.02331 | -0.07805 | -0.0678 | 0.01197 | 0.01417 |
| MPV | rs2620340 | rs2247139 | A | G | 12 | HPD | Female only | False | 0.1311 | 0.1317 | 0.1246 | 0.1232 | 26726 | 19784 | 188072 | 162398 | -0.07012 | -0.02359 | -0.0781 | -0.06802 | 0.01197 | 0.01417 |
| MPV | rs2620340 | rs1678951 | T | C | 12 | HPD | Female only | False | 0.1313 | 0.1318 | 0.1246 | 0.1233 | 26726 | 19784 | 188072 | 162398 | -0.06994 | -0.02227 | -0.07825 | -0.06791 | 0.01196 | 0.01416 |
| MPV | rs2620340 | rs693277 | A | G | 12 | HPD | Female only | False | 0.1307 | 0.131 | 0.1239 | 0.1227 | 26726 | 19784 | 188072 | 162398 | -0.07083 | -0.0255 | -0.07844 | -0.06765 | 0.012 | 0.01422 |
| MPV | rs2620340 | rs1667584 | A | G | 12 | HPD | Female only | False | 0.1307 | 0.131 | 0.1239 | 0.1227 | 26726 | 19784 | 188072 | 162398 | -0.07083 | -0.02551 | -0.07844 | -0.06766 | 0.012 | 0.01422 |
| MPV | rs2620340 | rs1185260 | A | G | 12 | HPD | Female only | False | 0.1307 | 0.131 | 0.1239 | 0.1227 | 26726 | 19784 | 188072 | 162398 | -0.07082 | -0.0255 | -0.07844 | -0.06766 | 0.012 | 0.01422 |
| MPV | rs2620340 | rs1154513 | A | G | 12 | HPD | Female only | False | 0.131 | 0.1313 | 0.1238 | 0.1225 | 26726 | 19784 | 188072 | 162398 | -0.07104 | -0.02584 | -0.07844 | -0.06764 | 0.012 | 0.01422 |
| MPV | rs2620340 | rs830124 | A | G | 12 | HPD | Female only | False | 0.1271 | 0.1274 | 0.1246 | 0.1234 | 26726 | 19784 | 188072 | 162398 | -0.07215 | -0.02607 | -0.07852 | -0.06763 | 0.01187 | 0.01406 |

| trait | lead_snp | ID | ALLELE1 | ALLELE0 | CHR | Gene_Symbol | independent_loci_in_sex_P1e2 | in_embl_any_or_gene | A1FREQ_VUMC_female | A1FREQ_VUMC_male | A1FREQ_UKB_female | A1FREQ_UKB_male | N_VUMC_female | N_VUMC_male | N_UKB_female | N_UKB_male | BETA_VUMC_female | BETA_VUMC_male | BETA_UKB_female | BETA_UKB_male | SE_VUMC_female | SE_VUMC_male |
|---|---|---|---|---|---|---|---|---|---|---|---|---|---|---|---|---|---|---|---|---|---|---|
| MPV | rs2620340 | rs493519 | T | C | 12 | HPD | Female only | False | 0.1321 | 0.1325 | 0.1247 | 0.1235 | 26726 | 19784 | 188072 | 162398 | -0.07315 | -0.02641 | -0.07848 | -0.06746 | 0.01196 | 0.01416 |
| MPV | rs2620340 | rs830126 | A | G | 12 | HPD | Female only | False | 0.1321 | 0.1325 | 0.1247 | 0.1235 | 26726 | 19784 | 188072 | 162398 | -0.07315 | -0.02641 | -0.07842 | -0.06742 | 0.01196 | 0.01416 |
| MPV | rs2620340 | rs1169080 | G | T | 12 | HPD | Female only | False | 0.1325 | 0.1328 | 0.1243 | 0.123 | 26726 | 19784 | 188072 | 162398 | -0.07428 | -0.02715 | -0.07809 | -0.068 | 0.01201 | 0.01423 |
| MPV | rs2620340 | rs830116 | A | G | 12 | HPD | Female only | False | 0.1325 | 0.1328 | 0.1243 | 0.123 | 26726 | 19784 | 188072 | 162398 | -0.07453 | -0.02717 | -0.0782 | -0.06793 | 0.01202 | 0.01424 |
| MPV | rs2620340 | rs1169075 | A | C | 12 | HPD | Female only | False | 0.1275 | 0.1283 | 0.119 | 0.1177 | 26726 | 19784 | 188072 | 162398 | -0.07618 | -0.02921 | -0.07873 | -0.06758 | 0.01227 | 0.01451 |
| MPV | rs12601994 | rs60445180 | A | G | 17 | MYO18A | Female only | False | 0.1455 | 0.1471 | 0.1446 | 0.1448 | 26726 | 19784 | 188072 | 162398 | 0.05344 | 0.02953 | 0.05751 | 0.05219 | 0.01121 | 0.01326 |
| MPV | rs12601994 | rs12601736 | C | T | 17 | MYO18A | Female only | False | 0.1511 | 0.153 | 0.1494 | 0.1497 | 26726 | 19784 | 188072 | 162398 | 0.04948 | 0.02826 | 0.05361 | 0.04832 | 0.011 | 0.01299 |
| MPV | rs12601994 | rs4795488 | A | G | 17 | MYO18A | Female only | False | 0.1812 | 0.1817 | 0.1811 | 0.182 | 26726 | 19784 | 188072 | 162398 | 0.04191 | 0.01429 | 0.05146 | 0.04798 | 0.0102 | 0.01207 |
| MPV | rs12601994 | rs2015918 | A | G | 17 | MYO18A | Female only | False | 0.1825 | 0.1829 | 0.1814 | 0.1824 | 26726 | 19784 | 188072 | 162398 | 0.04159 | 0.01467 | 0.05149 | 0.04813 | 0.01013 | 0.012 |
| MPV | rs12601994 | rs997996 | G | A | 17 | MYO18A | Female only | False | 0.1812 | 0.1818 | 0.1812 | 0.1821 | 26726 | 19784 | 188072 | 162398 | 0.04136 | 0.01411 | 0.05138 | 0.04777 | 0.0102 | 0.01208 |
| MPV | rs12601994 | rs12450143 | T | C | 17 | MYO18A | Female only | False | 0.1811 | 0.1816 | 0.1811 | 0.1821 | 26726 | 19784 | 188072 | 162398 | 0.04204 | 0.01576 | 0.0517 | 0.04818 | 0.0102 | 0.01208 |
| MPV | rs12601994 | rs4794845 | T | C | 17 | MYO18A | Female only | False | 0.1816 | 0.182 | 0.181 | 0.182 | 26726 | 19784 | 188072 | 162398 | 0.0416 | 0.01363 | 0.05154 | 0.04811 | 0.0102 | 0.01207 |
| MPV | rs12601994 | rs4794846 | A | G | 17 | MYO18A | Female only | False | 0.1814 | 0.1819 | 0.181 | 0.182 | 26726 | 19784 | 188072 | 162398 | 0.04176 | 0.01395 | 0.05149 | 0.04799 | 0.0102 | 0.01208 |
| MPV | rs12601994 | rs59130946 | A | G | 17 | MYO18A | Female only | False | 0.1457 | 0.147 | 0.1441 | 0.1445 | 26726 | 19784 | 188072 | 162398 | 0.05382 | 0.02783 | 0.05801 | 0.05311 | 0.01116 | 0.01321 |
| MPV | rs12601994 | rs112751772 | T | G | 17 | MYO18A | Female only | False | 0.1511 | 0.1532 | 0.1516 | 0.1518 | 26726 | 19784 | 188072 | 162398 | 0.05249 | 0.02382 | 0.05158 | 0.04858 | 0.01099 | 0.01301 |
| MPV | rs12601994 | rs68136852 | A | C | 17 | MYO18A | Female only | False | 0.1522 | 0.1542 | 0.1518 | 0.1523 | 26726 | 19784 | 188072 | 162398 | 0.0513 | 0.0219 | 0.05188 | 0.04904 | 0.01088 | 0.01291 |
| MPV | rs12601994 | rs60616188 | T | C | 17 | MYO18A | Female only | False | 0.1515 | 0.1548 | 0.1518 | 0.1522 | 26726 | 19784 | 188072 | 162398 | 0.0591 | 0.02566 | 0.05613 | 0.0523 | 0.01094 | 0.01298 |
| MPV | rs12601994 | rs16964783 | A | G | 17 | MYO18A | Female only | False | 0.1518 | 0.1554 | 0.1516 | 0.1519 | 26726 | 19784 | 188072 | 162398 | 0.05923 | 0.0255 | 0.05541 | 0.05211 | 0.01089 | 0.01291 |
| MPV | rs12601994 | rs8067806 | A | G | 17 | MYO18A | Female only | False | 0.1511 | 0.1546 | 0.1514 | 0.1518 | 26726 | 19784 | 188072 | 162398 | 0.05985 | 0.02578 | 0.05547 | 0.05186 | 0.01095 | 0.01298 |
| MPV | rs12601994 | rs67561831 | A | G | 17 | MYO18A | Female only | False | 0.1495 | 0.1529 | 0.15 | 0.1503 | 26726 | 19784 | 188072 | 162398 | 0.06007 | 0.02677 | 0.05548 | 0.05268 | 0.01102 | 0.01308 |
| MPV | rs12601994 | rs12601994 | T | C | 17 | MYO18A | Female only | False | 0.1509 | 0.1543 | 0.1518 | 0.1521 | 26726 | 19784 | 188072 | 162398 | 0.06073 | 0.02743 | 0.05674 | 0.05361 | 0.01099 | 0.01303 |
| MPV | rs16964837 | rs16964837 | G | A | 17 | MYO18A | Female only | False | 0.2713 | 0.2779 | 0.2684 | 0.2694 | 26726 | 19784 | 188072 | 162398 | 0.05513 | 0.02217 | 0.04626 | 0.03783 | 0.00881 | 0.01048 |
| MPV | rs16964837 | rs7210510 | T | C | 17 | MYO18A | Female only | False | 0.3172 | 0.3252 | 0.3159 | 0.3163 | 26726 | 19784 | 188072 | 162398 | 0.04604 | 0.02077 | 0.04712 | 0.0376 | 0.008413 | 0.01004 |
| MPV | rs16964837 | rs8067041 | A | G | 17 | MYO18A | Female only | False | 0.3266 | 0.3351 | 0.3244 | 0.3248 | 26726 | 19784 | 188072 | 162398 | 0.04669 | 0.01932 | 0.04712 | 0.03761 | 0.008358 | 0.009958 |
| MPV | rs16964837 | rs2172096 | C | T | 17 | MYO18A | Female only | False | 0.3642 | 0.3733 | 0.3619 | 0.3628 | 26726 | 19784 | 188072 | 162398 | 0.03783 | 0.01656 | 0.04404 | 0.03122 | 0.008135 | 0.009694 |
| MPV | rs16964837 | rs7216760 | T | G | 17 | MYO18A | Female only | False | 0.2693 | 0.2759 | 0.2672 | 0.2681 | 26726 | 19784 | 188072 | 162398 | 0.05482 | 0.02377 | 0.04672 | 0.03828 | 0.008825 | 0.0105 |
| MPV | rs16964837 | rs8079438 | C | A | 17 | MYO18A | Female only | False | 0.3281 | 0.3367 | 0.3258 | 0.326 | 26726 | 19784 | 188072 | 162398 | 0.0461 | 0.01898 | 0.04653 | 0.03715 | 0.008347 | 0.009951 |
| MPV | rs16964837 | rs12949480 | A | G | 17 | MYO18A | Female only | False | 0.3173 | 0.3253 | 0.316 | 0.3164 | 26726 | 19784 | 188072 | 162398 | 0.04611 | 0.02076 | 0.04715 | 0.03757 | 0.008415 | 0.01004 |
| MPV | rs16964837 | rs12939545 | A | G | 17 | MYO18A | Female only | False | 0.2698 | 0.2757 | 0.267 | 0.2679 | 26726 | 19784 | 188072 | 162398 | 0.05446 | 0.02498 | 0.04674 | 0.03859 | 0.00882 | 0.01052 |

| trait | lead_snp | ID | ALLELE1 | ALLELE0 | CHR | Gene_Symbol | independent_loci_in_sex_P1e2 | in_embl_any_or_gene | A1FREQ_VUMC_female | A1FREQ_VUMC_male | A1FREQ_UKB_female | A1FREQ_UKB_male | N_VUMC_female | N_VUMC_male | N_UKB_female | N_UKB_male | BETA_VUMC_female | BETA_VUMC_male | BETA_UKB_female | BETA_UKB_male | SE_VUMC_female | SE_VUMC_male |
|---|---|---|---|---|---|---|---|---|---|---|---|---|---|---|---|---|---|---|---|---|---|---|
| MPV | rs16964837 | rs56334637 | T | C | 17 | MYO18A | Female only | False | 0.2686 | 0.2747 | 0.2671 | 0.2679 | 26726 | 19784 | 188072 | 162398 | 0.05243 | 0.0258 | 0.04701 | 0.03901 | 0.00882 | 0.01051 |
| MPV | rs16964837 | rs10438797 | T | G | 17 | MYO18A | Female only | False | 0.3144 | 0.3214 | 0.3132 | 0.3144 | 26726 | 19784 | 188072 | 162398 | 0.04267 | 0.02164 | 0.04301 | 0.03146 | 0.00846 | 0.01005 |
| MPV | rs16964837 | rs11650603 | T | C | 17 | MYO18A | Female only | False | 0.2699 | 0.2759 | 0.2671 | 0.2679 | 26726 | 19784 | 188072 | 162398 | 0.05216 | 0.02571 | 0.04704 | 0.039 | 0.008779 | 0.01047 |
| MPV | rs16964837 | rs6505111 | C | T | 17 | MYO18A | Female only | False | 0.3616 | 0.3699 | 0.3617 | 0.3625 | 26726 | 19784 | 188072 | 162398 | 0.0367 | 0.01934 | 0.04449 | 0.03201 | 0.00817 | 0.009742 |
| MPV | rs16964837 | rs6505112 | C | T | 17 | MYO18A | Female only | False | 0.3609 | 0.3693 | 0.3615 | 0.3622 | 26726 | 19784 | 188072 | 162398 | 0.03614 | 0.01976 | 0.04464 | 0.03199 | 0.008177 | 0.009743 |
| MPV | rs16964837 | rs11654782 | C | T | 17 | MYO18A | Female only | False | 0.3628 | 0.371 | 0.3623 | 0.3631 | 26726 | 19784 | 188072 | 162398 | 0.03728 | 0.01917 | 0.04516 | 0.03257 | 0.008163 | 0.009733 |
| MPV | rs16964837 | rs55731015 | A | G | 17 | MYO18A | Female only | False | 0.2701 | 0.2762 | 0.2685 | 0.2691 | 26726 | 19784 | 188072 | 162398 | 0.05209 | 0.02488 | 0.04649 | 0.03892 | 0.008806 | 0.0105 |
| MPV | rs16964837 | rs12949975 | T | C | 17 | MYO18A | Female only | False | 0.2701 | 0.2762 | 0.2685 | 0.2691 | 26726 | 19784 | 188072 | 162398 | 0.05206 | 0.02488 | 0.04648 | 0.03894 | 0.008806 | 0.0105 |
| MPV | rs16964837 | rs11080088 | A | G | 17 | MYO18A | Female only | False | 0.2684 | 0.2746 | 0.2667 | 0.2672 | 26726 | 19784 | 188072 | 162398 | 0.05173 | 0.02503 | 0.0463 | 0.03884 | 0.008851 | 0.01055 |
| MPV | rs16964837 | rs11080089 | A | G | 17 | MYO18A | Female only | False | 0.2682 | 0.2744 | 0.2666 | 0.2671 | 26726 | 19784 | 188072 | 162398 | 0.05162 | 0.02512 | 0.04626 | 0.03883 | 0.008856 | 0.01055 |
| MPV | rs16964837 | rs56220857 | T | G | 17 | MYO18A | Female only | False | 0.315 | 0.3224 | 0.3153 | 0.3156 | 26726 | 19784 | 188072 | 162398 | 0.04402 | 0.02305 | 0.04773 | 0.03847 | 0.008428 | 0.01005 |
| MPV | rs16964837 | rs35377455 | A | G | 17 | MYO18A | Female only | False | 0.2687 | 0.2747 | 0.2672 | 0.268 | 26726 | 19784 | 188072 | 162398 | 0.05289 | 0.02563 | 0.04728 | 0.03919 | 0.008825 | 0.01052 |
| MPV | rs16964837 | rs9906043 | A | G | 17 | MYO18A | Female only | False | 0.364 | 0.3724 | 0.3623 | 0.363 | 26726 | 19784 | 188072 | 162398 | 0.03721 | 0.01889 | 0.04521 | 0.03253 | 0.008131 | 0.009692 |
| MPV | rs16964837 | rs11657942 | T | C | 17 | MYO18A | Female only | False | 0.2752 | 0.282 | 0.2746 | 0.2754 | 26726 | 19784 | 188072 | 162398 | 0.05085 | 0.02385 | 0.04544 | 0.03694 | 0.008759 | 0.01043 |
| MPV | rs16964837 | rs60886130 | A | G | 17 | MYO18A | Female only | False | 0.2752 | 0.282 | 0.2744 | 0.2752 | 26726 | 19784 | 188072 | 162398 | 0.05086 | 0.02385 | 0.04549 | 0.03673 | 0.008759 | 0.01043 |
| MPV | rs16964837 | rs2063139 | G | T | 17 | MYO18A | Female only | False | 0.2742 | 0.2808 | 0.2734 | 0.2742 | 26726 | 19784 | 188072 | 162398 | 0.04987 | 0.0235 | 0.04559 | 0.03678 | 0.008787 | 0.01047 |
| MPV | rs16964837 | rs2063138 | A | G | 17 | MYO18A | Female only | False | 0.2834 | 0.2908 | 0.2819 | 0.2827 | 26726 | 19784 | 188072 | 162398 | 0.04997 | 0.02223 | 0.04533 | 0.03652 | 0.008706 | 0.01037 |
| MPV | rs16964837 | rs12941511 | A | G | 17 | MYO18A | Female only | False | 0.2733 | 0.2799 | 0.2729 | 0.274 | 26726 | 19784 | 188072 | 162398 | 0.05067 | 0.02465 | 0.04622 | 0.0372 | 0.008787 | 0.01047 |
| MPV | rs16964837 | rs16963476 | T | C | 17 | MYO18A | Female only | False | 0.2728 | 0.2795 | 0.2728 | 0.2738 | 26726 | 19784 | 188072 | 162398 | 0.05013 | 0.02463 | 0.0463 | 0.03729 | 0.008801 | 0.01049 |
| MPV | rs16964837 | rs34299662 | A | G | 17 | MYO18A | Female only | False | 0.2728 | 0.2794 | 0.2728 | 0.2738 | 26726 | 19784 | 188072 | 162398 | 0.05017 | 0.02456 | 0.04631 | 0.03731 | 0.008801 | 0.01049 |
| MPV | rs16964837 | rs55667136 | C | A | 17 | MYO18A | Female only | False | 0.354 | 0.357 | 0.3574 | 0.3578 | 26726 | 19784 | 188072 | 162398 | 0.04858 | 0.01797 | 0.04031 | 0.0333 | 0.008197 | 0.009765 |
| MPV | rs16964837 | rs34904195 | C | T | 17 | MYO18A | Female only | False | 0.3499 | 0.3533 | 0.3541 | 0.3545 | 26726 | 19784 | 188072 | 162398 | 0.04884 | 0.01449 | 0.03784 | 0.03225 | 0.008227 | 0.009795 |
| MPV | rs9807816 | rs8112139 | A | G | 19 | FXYD5 | Female only | True | 0.3009 | 0.3039 | 0.2784 | 0.2784 | 26726 | 19784 | 188072 | 162398 | -0.04754 | -0.01808 | -0.02585 | -0.01534 | 0.008695 | 0.01037 |
| MPV | rs9807816 | rs10412914 | T | C | 19 | FXYD5 | Female only | True | 0.3023 | 0.3054 | 0.279 | 0.2791 | 26726 | 19784 | 188072 | 162398 | -0.04842 | -0.01774 | -0.02611 | -0.0152 | 0.008687 | 0.01035 |
| MPV | rs9807816 | rs11084804 | T | C | 19 | FXYD5 | Female only | True | 0.3019 | 0.3051 | 0.2794 | 0.2794 | 26726 | 19784 | 188072 | 162398 | -0.04897 | -0.01713 | -0.02602 | -0.01498 | 0.008679 | 0.01034 |
| MPV | rs9807816 | rs9807816 | T | G | 19 | FXYD5 | Female only | True | 0.2038 | 0.2044 | 0.1922 | 0.193 | 26726 | 19784 | 188072 | 162398 | -0.05592 | -0.0148 | -0.03505 | -0.02759 | 0.009714 | 0.01157 |
| MPV | rs9807816 | rs9807905 | C | T | 19 | FXYD5 | Female only | True | 0.2939 | 0.297 | 0.2803 | 0.2804 | 26726 | 19784 | 188072 | 162398 | -0.04813 | -0.01785 | -0.02619 | -0.0151 | 0.008576 | 0.01021 |

| trait | lead_snp | ID | ALLELE1 | ALLELE0 | CHR | Gene_Symbol | independent_loci_in_sex_P1e2 | in_embl_any_or_gene | A1FREQ_VUMC_female | A1FREQ_VUMC_male | A1FREQ_UKB_female | A1FREQ_UKB_male | N_VUMC_female | N_VUMC_male | N_UKB_female | N_UKB_male | BETA_VUMC_female | BETA_VUMC_male | BETA_UKB_female | BETA_UKB_male | SE_VUMC_female | SE_VUMC_male |
|---|---|---|---|---|---|---|---|---|---|---|---|---|---|---|---|---|---|---|---|---|---|---|
| MPV | rs9807816 | rs10425064 | A | C | 19 | FXYD5 | Female only | True | 0.2126 | 0.2134 | 0.1922 | 0.193 | 26726 | 19784 | 188072 | 162398 | -0.05641 | -0.01503 | -0.03502 | -0.02754 | 0.009812 | 0.01169 |
| R_F | rs73719806 | rs77692810 | C | T | 7 | AC004875.1 | Male only | False | 0.1018 | 0.1059 | 0.1024 | 0.1036 | 2715 | 1250 | 17000 | 13565 | -0.0706 | 0.2775 | 0.01093 | 0.01043 | 0.04258 | 0.0631 |
| R_F | rs73719806 | rs7455326 | A | G | 7 | AC004875.1 | Male only | False | 0.1029 | 0.1069 | 0.1028 | 0.104 | 2715 | 1250 | 17000 | 13565 | -0.07145 | 0.2718 | 0.0128 | 0.009189 | 0.04222 | 0.06268 |
| R_F | rs73719806 | rs11977460 | G | A | 7 | AC004875.1 | Male only | False | 0.1037 | 0.1084 | 0.1039 | 0.1048 | 2715 | 1250 | 17000 | 13565 | -0.08018 | 0.2877 | 0.0112 | 0.01225 | 0.04226 | 0.06211 |
| R_F | rs73719806 | rs73225231 | C | T | 7 | AC004875.1 | Male only | False | 0.1019 | 0.106 | 0.1026 | 0.1036 | 2715 | 1250 | 17000 | 13565 | -0.07209 | 0.2737 | 0.01023 | 0.01058 | 0.04267 | 0.06315 |
| R_F | rs73719806 | rs11979236 | G | A | 7 | AC004875.1 | Male only | False | 0.1042 | 0.1091 | 0.1037 | 0.1045 | 2715 | 1250 | 17000 | 13565 | -0.08057 | 0.2811 | 0.01182 | 0.01192 | 0.04215 | 0.06202 |
| R_F | rs73719806 | rs73717387 | G | A | 7 | AC004875.1 | Male only | False | 0.06648 | 0.06646 | 0.05817 | 0.05967 | 2715 | 1250 | 17000 | 13565 | -0.03019 | 0.4164 | 0.01452 | 0.03713 | 0.05148 | 0.07694 |
| R_F | rs73719806 | rs73717390 | A | G | 7 | AC004875.1 | Male only | False | 0.06563 | 0.06562 | 0.05806 | 0.05964 | 2715 | 1250 | 17000 | 13565 | -0.02373 | 0.4112 | 0.01551 | 0.03522 | 0.05206 | 0.07759 |
| R_F | rs73719806 | rs7792771 | T | C | 7 | AC004875.1 | Male only | False | 0.06598 | 0.06603 | 0.05813 | 0.05973 | 2715 | 1250 | 17000 | 13565 | -0.02885 | 0.4203 | 0.01486 | 0.03697 | 0.05179 | 0.07738 |
| R_F | rs73719806 | rs73717391 | C | T | 7 | AC004875.1 | Male only | False | 0.06024 | 0.06085 | 0.05294 | 0.054 | 2715 | 1250 | 17000 | 13565 | -0.02438 | 0.445 | 0.01265 | 0.03264 | 0.05426 | 0.08069 |
| R_F | rs73719806 | rs56263246 | T | C | 7 | AC004875.1 | Male only | False | 0.06604 | 0.06635 | 0.05808 | 0.05985 | 2715 | 1250 | 17000 | 13565 | -0.02946 | 0.4232 | 0.0146 | 0.03993 | 0.05179 | 0.07726 |
| R_F | rs73719806 | rs73717393 | C | T | 7 | AC004875.1 | Male only | False | 0.06604 | 0.06635 | 0.05808 | 0.05985 | 2715 | 1250 | 17000 | 13565 | -0.02946 | 0.4232 | 0.01461 | 0.03995 | 0.05179 | 0.07726 |
| R_F | rs73719806 | rs1406193 | C | T | 7 | AC004875.1 | Male only | False | 0.06586 | 0.06615 | 0.05799 | 0.05974 | 2715 | 1250 | 17000 | 13565 | -0.0297 | 0.426 | 0.01434 | 0.04103 | 0.05187 | 0.07741 |
| R_F | rs73719806 | rs1830308 | G | T | 7 | AC004875.1 | Male only | False | 0.0658 | 0.06616 | 0.05805 | 0.0598 | 2715 | 1250 | 17000 | 13565 | -0.03089 | 0.4262 | 0.01458 | 0.04084 | 0.05187 | 0.0774 |
| R_F | rs73719806 | rs74818858 | G | T | 7 | AC004875.1 | Male only | False | 0.06584 | 0.06619 | 0.05809 | 0.05988 | 2715 | 1250 | 17000 | 13565 | -0.03079 | 0.4256 | 0.01466 | 0.03975 | 0.05184 | 0.07736 |
| R_F | rs73719806 | rs77186372 | A | G | 7 | AC004875.1 | Male only | False | 0.06584 | 0.06619 | 0.05808 | 0.05987 | 2715 | 1250 | 17000 | 13565 | -0.03079 | 0.4256 | 0.01465 | 0.03981 | 0.05184 | 0.07736 |
| R_F | rs73719806 | rs59320957 | C | T | 7 | AC004875.1 | Male only | False | 0.06584 | 0.06619 | 0.05808 | 0.05987 | 2715 | 1250 | 17000 | 13565 | -0.03079 | 0.4256 | 0.01465 | 0.03981 | 0.05184 | 0.07736 |
| R_F | rs73719806 | rs6970940 | A | G | 7 | AC004875.1 | Male only | False | 0.06584 | 0.0662 | 0.05809 | 0.05988 | 2715 | 1250 | 17000 | 13565 | -0.03082 | 0.4255 | 0.01509 | 0.04038 | 0.05184 | 0.07736 |
| R_F | rs73719806 | rs6952513 | T | C | 7 | AC004875.1 | Male only | False | 0.06582 | 0.06578 | 0.05803 | 0.05984 | 2715 | 1250 | 17000 | 13565 | -0.03096 | 0.4308 | 0.01498 | 0.04089 | 0.05185 | 0.07747 |
| R_F | rs73719806 | rs73717402 | G | T | 7 | AC004875.1 | Male only | False | 0.06571 | 0.06569 | 0.05803 | 0.05987 | 2715 | 1250 | 17000 | 13565 | -0.03165 | 0.4315 | 0.01514 | 0.04083 | 0.05184 | 0.07746 |
| R_F | rs73719806 | rs73719804 | A | G | 7 | AC004875.1 | Male only | False | 0.06593 | 0.066 | 0.05804 | 0.05988 | 2715 | 1250 | 17000 | 13565 | -0.03123 | 0.4284 | 0.01536 | 0.04091 | 0.05167 | 0.07707 |
| R_F | rs73719806 | rs6964469 | A | C | 7 | AC004875.1 | Male only | False | 0.06571 | 0.06569 | 0.05802 | 0.05987 | 2715 | 1250 | 17000 | 13565 | -0.03165 | 0.4315 | 0.01538 | 0.04103 | 0.05184 | 0.07746 |
| R_F | rs73719806 | rs73719806 | T | G | 7 | AC004875.1 | Male only | False | 0.06571 | 0.0657 | 0.05802 | 0.05988 | 2715 | 1250 | 17000 | 13565 | -0.03168 | 0.4315 | 0.01584 | 0.04135 | 0.05184 | 0.07746 |
| R_F | rs73719806 | rs73719807 | C | A | 7 | AC004875.1 | Male only | False | 0.06571 | 0.0653 | 0.058 | 0.05988 | 2715 | 1250 | 17000 | 13565 | -0.03168 | 0.4212 | 0.01554 | 0.04147 | 0.05184 | 0.07763 |
| R_F | rs73719806 | rs73719809 | C | A | 7 | AC004875.1 | Male only | False | 0.06362 | 0.06264 | 0.05701 | 0.05919 | 2715 | 1250 | 17000 | 13565 | -0.01958 | 0.4079 | 0.01595 | 0.03887 | 0.05274 | 0.0795 |
| R_F | rs73719806 | rs73719811 | C | T | 7 | AC004875.1 | Male only | False | 0.0657 | 0.06542 | 0.05802 | 0.05993 | 2715 | 1250 | 17000 | 13565 | -0.0317 | 0.4208 | 0.01605 | 0.04189 | 0.05189 | 0.0777 |
| R_F | rs73719806 | rs17143495 | G | T | 7 | AC004875.1 | Male only | False | 0.0657 | 0.06542 | 0.05801 | 0.05994 | 2715 | 1250 | 17000 | 13565 | -0.0317 | 0.4208 | 0.01606 | 0.04195 | 0.05189 | 0.0777 |
| R_F | rs73719806 | rs113713539 | T | C | 7 | AC004875.1 | Male only | False | 0.0657 | 0.06542 | 0.05801 | 0.05994 | 2715 | 1250 | 17000 | 13565 | -0.0317 | 0.4208 | 0.01607 | 0.04193 | 0.05189 | 0.0777 |
| R_F | rs73719806 | rs117577850 | A | G | 7 | AC004875.1 | Male only | False | 0.06354 | 0.06275 | 0.05712 | 0.05933 | 2715 | 1250 | 17000 | 13565 | -0.0214 | 0.4077 | 0.01516 | 0.04011 | 0.05282 | 0.07955 |

| trait | lead_snp | ID | ALLELE1 | ALLELE0 | CHR | Gene_Symbol | independent_loci_in_sex_P1e2 | in_embl_any_or_gene | A1FREQ_VUMC_female | A1FREQ_VUMC_male | A1FREQ_UKB_female | A1FREQ_UKB_male | N_VUMC_female | N_VUMC_male | N_UKB_female | N_UKB_male | BETA_VUMC_female | BETA_VUMC_male | BETA_UKB_female | BETA_UKB_male | SE_VUMC_female | SE_VUMC_male |
|---|---|---|---|---|---|---|---|---|---|---|---|---|---|---|---|---|---|---|---|---|---|---|
| R_F | rs73719806 | rs59102661 | C | T | 7 | AC004875.1 | Male only | False | 0.06352 | 0.0629 | 0.0571 | 0.05923 | 2715 | 1250 | 17000 | 13565 | -0.02015 | 0.4072 | 0.01559 | 0.04001 | 0.05285 | 0.07962 |
| R_F | rs73719806 | rs112732165 | T | C | 7 | AC004875.1 | Male only | False | 0.06571 | 0.06554 | 0.05804 | 0.05994 | 2715 | 1250 | 17000 | 13565 | -0.03152 | 0.4202 | 0.01591 | 0.04185 | 0.05193 | 0.07779 |
| R_F | rs73719806 | rs76065308 | A | G | 7 | AC004875.1 | Male only | False | 0.06569 | 0.06555 | 0.058 | 0.05997 | 2715 | 1250 | 17000 | 13565 | -0.03141 | 0.42 | 0.01633 | 0.04196 | 0.05195 | 0.07781 |
| R_F | rs73719806 | rs17132763 | A | G | 7 | AC004875.1 | Male only | False | 0.06554 | 0.06563 | 0.05802 | 0.06001 | 2715 | 1250 | 17000 | 13565 | -0.02941 | 0.4198 | 0.01607 | 0.0417 | 0.05201 | 0.07781 |
| R_F | rs73719806 | rs10488048 | G | A | 7 | AC004875.1 | Male only | False | 0.06503 | 0.06523 | 0.05779 | 0.05973 | 2715 | 1250 | 17000 | 13565 | -0.02178 | 0.4106 | 0.01616 | 0.03902 | 0.05235 | 0.078 |
| R_F | rs73719806 | rs6466798 | A | G | 7 | AC004875.1 | Male only | False | 0.06556 | 0.06564 | 0.05804 | 0.06002 | 2715 | 1250 | 17000 | 13565 | -0.02939 | 0.4197 | 0.01596 | 0.04168 | 0.052 | 0.0778 |
| BMI_CLEAN | rs7231154 | rs9959028 | G | T | 18 | RNU6-567P | Female only | True | 0.3329 | 0.334 | 0.3353 | 0.335 | 35694 | 26408 | 193570 | 166413 | 0.04542 | 0.02088 | 0.03543 | 0.03507 | 0.007802 | 0.009173 |
| BMI_CLEAN | rs7231154 | rs11664406 | C | T | 18 | RNU6-567P | Female only | True | 0.33 | 0.3309 | 0.3342 | 0.3338 | 35694 | 26408 | 193570 | 166413 | 0.04623 | 0.02243 | 0.0357 | 0.03515 | 0.007773 | 0.009144 |
| BMI_CLEAN | rs7231154 | rs4940925 | C | T | 18 | RNU6-567P | Female only | True | 0.4652 | 0.4666 | 0.469 | 0.469 | 35694 | 26408 | 193570 | 166413 | 0.03536 | 0.01486 | 0.02582 | 0.02578 | 0.007359 | 0.008676 |
| BMI_CLEAN | rs7231154 | rs8085349 | G | A | 18 | RNU6-567P | Female only | True | 0.4648 | 0.466 | 0.4703 | 0.4703 | 35694 | 26408 | 193570 | 166413 | 0.03524 | 0.01421 | 0.02573 | 0.02519 | 0.007353 | 0.008665 |
| BMI_CLEAN | rs7231154 | rs7234273 | G | T | 18 | RNU6-567P | Female only | True | 0.4647 | 0.466 | 0.4702 | 0.4702 | 35694 | 26408 | 193570 | 166413 | 0.03514 | 0.01419 | 0.02569 | 0.02522 | 0.007353 | 0.008665 |
| BMI_CLEAN | rs7231154 | rs7233720 | T | C | 18 | RNU6-567P | Female only | True | 0.4644 | 0.4656 | 0.4699 | 0.4699 | 35694 | 26408 | 193570 | 166413 | 0.03539 | 0.01419 | 0.02581 | 0.02545 | 0.007353 | 0.008665 |
| BMI_CLEAN | rs7231154 | rs7238389 | C | T | 18 | RNU6-567P | Female only | True | 0.4647 | 0.466 | 0.4703 | 0.4703 | 35694 | 26408 | 193570 | 166413 | 0.03523 | 0.01423 | 0.02574 | 0.02521 | 0.007353 | 0.008665 |
| BMI_CLEAN | rs7231154 | rs7240152 | C | A | 18 | RNU6-567P | Female only | True | 0.4648 | 0.4661 | 0.4703 | 0.4703 | 35694 | 26408 | 193570 | 166413 | 0.03519 | 0.01434 | 0.0257 | 0.02518 | 0.007352 | 0.008663 |
| BMI_CLEAN | rs7231154 | rs9964646 | G | A | 18 | RNU6-567P | Female only | True | 0.4648 | 0.4662 | 0.4703 | 0.4703 | 35694 | 26408 | 193570 | 166413 | 0.03516 | 0.0142 | 0.02574 | 0.0251 | 0.007352 | 0.008662 |
| BMI_CLEAN | rs7231154 | rs12957506 | G | T | 18 | RNU6-567P | Female only | True | 0.3312 | 0.332 | 0.335 | 0.3346 | 35694 | 26408 | 193570 | 166413 | 0.04616 | 0.02172 | 0.03556 | 0.0347 | 0.007761 | 0.009132 |
| BMI_CLEAN | rs7231154 | rs7232886 | G | A | 18 | RNU6-567P | Female only | True | 0.4657 | 0.4669 | 0.4703 | 0.4704 | 35694 | 26408 | 193570 | 166413 | 0.03488 | 0.01397 | 0.02579 | 0.02506 | 0.007338 | 0.008646 |
| BMI_CLEAN | rs7231154 | rs7231154 | A | G | 18 | RNU6-567P | Female only | True | 0.3304 | 0.3312 | 0.3348 | 0.3344 | 35694 | 26408 | 193570 | 166413 | 0.04643 | 0.02126 | 0.03548 | 0.03474 | 0.007775 | 0.009147 |
| BMI_CLEAN | rs7231154 | rs11664212 | G | T | 18 | RNU6-567P | Female only | True | 0.3307 | 0.3314 | 0.3344 | 0.334 | 35694 | 26408 | 193570 | 166413 | 0.04625 | 0.02133 | 0.03555 | 0.03478 | 0.007766 | 0.009136 |
| BMI_CLEAN | rs7231154 | rs11664327 | C | T | 18 | RNU6-567P | Female only | True | 0.3323 | 0.3327 | 0.3363 | 0.3358 | 35694 | 26408 | 193570 | 166413 | 0.04577 | 0.02067 | 0.0352 | 0.03392 | 0.007755 | 0.009128 |
| BMI_CLEAN | rs7231154 | rs4603671 | A | G | 18 | RNU6-567P | Female only | True | 0.4511 | 0.4522 | 0.4553 | 0.4551 | 35694 | 26408 | 193570 | 166413 | 0.03331 | 0.01196 | 0.02596 | 0.02417 | 0.007393 | 0.008716 |
| Ca | rs13035592 | rs6713063 | A | G | 2 | AC079610.1 | Male only | False | 0.305 | 0.3027 | 0.3082 | 0.3092 | 34341 | 26934 | 168077 | 147076 | 0.01079 | 0.04416 | 0.002062 | 0.02101 | 0.007285 | 0.007947 |
| Ca | rs13035592 | rs10202665 | T | G | 2 | AC079610.1 | Male only | False | 0.3046 | 0.3022 | 0.308 | 0.3091 | 34341 | 26934 | 168077 | 147076 | 0.0107 | 0.04408 | 0.002034 | 0.02104 | 0.007289 | 0.007949 |
| Ca | rs13035592 | rs4132518 | T | G | 2 | AC079610.1 | Male only | False | 0.2904 | 0.289 | 0.2875 | 0.2878 | 34341 | 26934 | 168077 | 147076 | 0.009915 | 0.03976 | -3.19e-04 | 0.01917 | 0.007429 | 0.008094 |
| Ca | rs13035592 | rs12987688 | C | T | 2 | AC079610.1 | Male only | False | 0.2905 | 0.289 | 0.2875 | 0.2878 | 34341 | 26934 | 168077 | 147076 | 0.00996 | 0.03976 | -3.08e-04 | 0.0192 | 0.007428 | 0.008094 |
| Ca | rs13035592 | rs10168619 | T | C | 2 | AC079610.1 | Male only | False | 0.2847 | 0.2828 | 0.2871 | 0.2875 | 34341 | 26934 | 168077 | 147076 | 0.009978 | 0.03942 | -4.54e-04 | 0.01939 | 0.007423 | 0.008094 |
| Ca | rs13035592 | rs11900001 | C | A | 2 | AC079610.1 | Male only | False | 0.3103 | 0.3083 | 0.3085 | 0.3094 | 34341 | 26934 | 168077 | 147076 | 0.01089 | 0.04487 | 0.002108 | 0.02085 | 0.007292 | 0.007952 |

| trait | lead_snp | ID | ALLELE1 | ALLELE0 | CHR | Gene_Symbol | independent_loci_in_sex_P1e2 | in_embl_any_or_gene | A1FREQ_VUMC_female | A1FREQ_VUMC_male | A1FREQ_UKB_female | A1FREQ_UKB_male | N_VUMC_female | N_VUMC_male | N_UKB_female | N_UKB_male | BETA_VUMC_female | BETA_VUMC_male | BETA_UKB_female | BETA_UKB_male | SE_VUMC_female | SE_VUMC_male |
|---|---|---|---|---|---|---|---|---|---|---|---|---|---|---|---|---|---|---|---|---|---|---|
| Ca | rs13035592 | rs11884431 | T | C | 2 | AC079610.1 | Male only | False | 0.2902 | 0.2886 | 0.2874 | 0.2877 | 34341 | 26934 | 168077 | 147076 | 0.009721 | 0.03984 | -3.78e-04 | 0.01913 | 0.007431 | 0.008096 |
| Ca | rs13035592 | rs4592794 | T | C | 2 | AC079610.1 | Male only | False | 0.2902 | 0.2886 | 0.2874 | 0.2877 | 34341 | 26934 | 168077 | 147076 | 0.009742 | 0.03984 | -3.78e-04 | 0.01914 | 0.007431 | 0.008096 |
| Ca | rs13035592 | rs11694194 | T | C | 2 | AC079610.1 | Male only | False | 0.2849 | 0.2829 | 0.2872 | 0.2875 | 34341 | 26934 | 168077 | 147076 | 0.01014 | 0.03903 | -4.77e-04 | 0.01955 | 0.007418 | 0.008088 |
| Ca | rs13035592 | rs7581321 | A | G | 2 | AC079610.1 | Male only | False | 0.2847 | 0.2827 | 0.2871 | 0.2874 | 34341 | 26934 | 168077 | 147076 | 0.009909 | 0.03897 | -5.36e-04 | 0.01946 | 0.00742 | 0.008091 |
| Ca | rs13035592 | rs13035592 | G | T | 2 | AC079610.1 | Male only | False | 0.3069 | 0.3051 | 0.3086 | 0.3095 | 34341 | 26934 | 168077 | 147076 | 0.01039 | 0.04498 | 0.002027 | 0.02112 | 0.007251 | 0.007906 |
| Ca | rs13035592 | rs7585560 | A | G | 2 | AC079610.1 | Male only | False | 0.2845 | 0.2825 | 0.287 | 0.2873 | 34341 | 26934 | 168077 | 147076 | 0.01028 | 0.03834 | -7.26e-04 | 0.01939 | 0.007426 | 0.008099 |
| Ca | rs13035592 | rs10182856 | T | C | 2 | AC079610.1 | Male only | False | 0.2897 | 0.2882 | 0.2872 | 0.2875 | 34341 | 26934 | 168077 | 147076 | 0.00977 | 0.039 | -5.44e-04 | 0.01918 | 0.00743 | 0.008098 |
| Ca | rs13035592 | rs7606842 | T | C | 2 | AC079610.1 | Male only | False | 0.2865 | 0.285 | 0.2875 | 0.2878 | 34341 | 26934 | 168077 | 147076 | 0.009711 | 0.03906 | -3.99e-04 | 0.01931 | 0.007393 | 0.008055 |
| Ca | rs13035592 | rs10183789 | T | C | 2 | AC079610.1 | Male only | False | 0.2844 | 0.2821 | 0.2871 | 0.2874 | 34341 | 26934 | 168077 | 147076 | 0.01017 | 0.03808 | -5.74e-04 | 0.01949 | 0.007428 | 0.0081 |
| Ca | rs13035592 | rs13387639 | A | C | 2 | AC079610.1 | Male only | False | 0.2846 | 0.2825 | 0.2872 | 0.2876 | 34341 | 26934 | 168077 | 147076 | 0.009844 | 0.03803 | -4.91e-04 | 0.01947 | 0.007428 | 0.008099 |
| Ca | rs13035592 | rs4627506 | G | A | 2 | AC079610.1 | Male only | False | 0.2899 | 0.2883 | 0.2875 | 0.2878 | 34341 | 26934 | 168077 | 147076 | 0.01004 | 0.0387 | -4.46e-04 | 0.01914 | 0.00743 | 0.008095 |
| Ca | rs13035592 | rs4514826 | A | G | 2 | AC079610.1 | Male only | False | 0.2891 | 0.2872 | 0.2873 | 0.2877 | 34341 | 26934 | 168077 | 147076 | 0.0101 | 0.03869 | -4.70e-04 | 0.01939 | 0.007437 | 0.008107 |
| Ca | rs13291723 | rs10781458 | C | T | 9 | GNAQ | Female only | True | 0.4013 | 0.3981 | 0.3964 | 0.3966 | 34341 | 26934 | 168077 | 147076 | 0.03539 | 0.01546 | 0.02106 | 0.02799 | 0.006833 | 0.007432 |
| Ca | rs13291723 | rs11793689 | T | C | 9 | GNAQ | Female only | True | 0.3068 | 0.3055 | 0.2979 | 0.2979 | 34341 | 26934 | 168077 | 147076 | 0.04067 | 0.01455 | 0.02127 | 0.03491 | 0.007353 | 0.008017 |
| Ca | rs13291723 | rs10869978 | G | A | 9 | GNAQ | Female only | True | 0.3086 | 0.3068 | 0.2999 | 0.3001 | 34341 | 26934 | 168077 | 147076 | 0.04067 | 0.01469 | 0.02155 | 0.03475 | 0.007334 | 0.008005 |
| Ca | rs13291723 | rs7048811 | G | T | 9 | GNAQ | Female only | True | 0.2991 | 0.297 | 0.2982 | 0.2983 | 34341 | 26934 | 168077 | 147076 | 0.04012 | 0.01407 | 0.0223 | 0.03466 | 0.00728 | 0.007948 |
| Ca | rs13291723 | rs1436450 | G | A | 9 | GNAQ | Female only | True | 0.3062 | 0.3042 | 0.2988 | 0.2988 | 34341 | 26934 | 168077 | 147076 | 0.04004 | 0.01462 | 0.02192 | 0.03503 | 0.007338 | 0.008006 |
| Ca | rs13291723 | rs10521480 | G | A | 9 | GNAQ | Female only | True | 0.3057 | 0.3036 | 0.2983 | 0.2984 | 34341 | 26934 | 168077 | 147076 | 0.03984 | 0.01419 | 0.02233 | 0.03456 | 0.007341 | 0.00801 |
| Ca | rs13291723 | rs11145592 | G | T | 9 | GNAQ | Female only | True | 0.3091 | 0.3074 | 0.3004 | 0.3004 | 34341 | 26934 | 168077 | 147076 | 0.04068 | 0.01445 | 0.02178 | 0.03514 | 0.007316 | 0.007982 |
| Ca | rs13291723 | rs11145594 | C | T | 9 | GNAQ | Female only | True | 0.309 | 0.3073 | 0.3004 | 0.3003 | 34341 | 26934 | 168077 | 147076 | 0.04081 | 0.01462 | 0.0218 | 0.03525 | 0.007316 | 0.007982 |
| Ca | rs13291723 | rs2378087 | C | T | 9 | GNAQ | Female only | True | 0.309 | 0.3071 | 0.3004 | 0.3004 | 34341 | 26934 | 168077 | 147076 | 0.04021 | 0.0142 | 0.02166 | 0.03523 | 0.007315 | 0.007985 |
| Ca | rs13291723 | rs2378088 | A | G | 9 | GNAQ | Female only | True | 0.309 | 0.3071 | 0.3004 | 0.3004 | 34341 | 26934 | 168077 | 147076 | 0.04011 | 0.01418 | 0.02165 | 0.03523 | 0.007315 | 0.007985 |
| Ca | rs13291723 | rs4384049 | G | A | 9 | GNAQ | Female only | True | 0.3081 | 0.3066 | 0.3006 | 0.3006 | 34341 | 26934 | 168077 | 147076 | 0.04008 | 0.0142 | 0.02188 | 0.03523 | 0.007333 | 0.008003 |
| Ca | rs13291723 | rs11145595 | G | T | 9 | GNAQ | Female only | True | 0.3084 | 0.3068 | 0.3007 | 0.3007 | 34341 | 26934 | 168077 | 147076 | 0.04024 | 0.01459 | 0.02154 | 0.03521 | 0.00733 | 0.008002 |
| Ca | rs13291723 | rs10869979 | G | A | 9 | GNAQ | Female only | True | 0.3086 | 0.307 | 0.3007 | 0.3006 | 34341 | 26934 | 168077 | 147076 | 0.04048 | 0.01465 | 0.02159 | 0.03537 | 0.007329 | 0.008 |
| Ca | rs13291723 | rs34039885 | A | C | 9 | GNAQ | Female only | True | 0.4545 | 0.4538 | 0.4606 | 0.46 | 34341 | 26934 | 168077 | 147076 | -0.03262 | -0.006761 | -0.01922 | -0.0297 | 0.006761 | 0.007382 |
| Ca | rs13291723 | rs4745672 | T | C | 9 | GNAQ | Female only | True | 0.4549 | 0.4543 | 0.4606 | 0.4601 | 34341 | 26934 | 168077 | 147076 | -0.0325 | -0.007297 | -0.01973 | -0.03013 | 0.006769 | 0.007388 |
| Ca | rs13291723 | rs4745673 | C | T | 9 | GNAQ | Female only | True | 0.4314 | 0.4319 | 0.4247 | 0.4254 | 34341 | 26934 | 168077 | 147076 | 0.04231 | 0.01233 | 0.02496 | 0.03752 | 0.006782 | 0.007436 |
| Ca | rs13291723 | rs1410552 | T | C | 9 | GNAQ | Female only | True | 0.3147 | 0.3125 | 0.3078 | 0.3076 | 34341 | 26934 | 168077 | 147076 | 0.03877 | 0.01427 | 0.02139 | 0.03477 | 0.007259 | 0.007949 |

| trait | lead_snp | ID | ALLELE1 | ALLELE0 | CHR | Gene_Symbol | independent_loci_in_sex_P1e2 | in_embl_any_or_gene | A1FREQ_VUMC_female | A1FREQ_VUMC_male | A1FREQ_UKB_female | A1FREQ_UKB_male | N_VUMC_female | N_VUMC_male | N_UKB_female | N_UKB_male | BETA_VUMC_female | BETA_VUMC_male | BETA_UKB_female | BETA_UKB_male | SE_VUMC_female | SE_VUMC_male |
|---|---|---|---|---|---|---|---|---|---|---|---|---|---|---|---|---|---|---|---|---|---|---|
| Ca | rs13291723 | rs9695575 | A | G | 9 | GNAQ | Female only | True | 0.3153 | 0.313 | 0.3087 | 0.3084 | 34341 | 26934 | 168077 | 147076 | 0.03961 | 0.01333 | 0.02109 | 0.03483 | 0.007255 | 0.007948 |
| Ca | rs13291723 | rs1059033 | G | A | 9 | GNAQ | Female only | True | 0.316 | 0.3139 | 0.309 | 0.3087 | 34341 | 26934 | 168077 | 147076 | 0.03952 | 0.01304 | 0.02131 | 0.03445 | 0.007252 | 0.007942 |
| Ca | rs13291723 | rs13291723 | G | A | 9 | GNAQ | Female only | True | 0.4334 | 0.4341 | 0.426 | 0.4266 | 34341 | 26934 | 168077 | 147076 | 0.04331 | 0.01157 | 0.02477 | 0.03746 | 0.006785 | 0.007441 |
| Ca | rs13291723 | rs2378100 | C | T | 9 | GNAQ | Female only | True | 0.4318 | 0.4322 | 0.4248 | 0.4253 | 34341 | 26934 | 168077 | 147076 | 0.04266 | 0.0122 | 0.02479 | 0.0377 | 0.006789 | 0.007444 |
| Ca | rs13291723 | rs10781470 | A | G | 9 | GNAQ | Female only | True | 0.315 | 0.3128 | 0.3082 | 0.308 | 34341 | 26934 | 168077 | 147076 | 0.03945 | 0.01283 | 0.02136 | 0.03453 | 0.007256 | 0.007955 |
| Ca | rs13291723 | rs10781471 | G | T | 9 | GNAQ | Female only | True | 0.3088 | 0.3065 | 0.3082 | 0.308 | 34341 | 26934 | 168077 | 147076 | 0.03932 | 0.01279 | 0.02135 | 0.03453 | 0.007192 | 0.007885 |
| Ca | rs13291723 | rs4745674 | C | A | 9 | GNAQ | Female only | True | 0.3154 | 0.3133 | 0.3084 | 0.3082 | 34341 | 26934 | 168077 | 147076 | 0.03968 | 0.01302 | 0.02132 | 0.03455 | 0.007255 | 0.007951 |
| Ca | rs13291723 | rs4745675 | A | G | 9 | GNAQ | Female only | True | 0.3154 | 0.3133 | 0.3084 | 0.3082 | 34341 | 26934 | 168077 | 147076 | 0.03964 | 0.01302 | 0.02132 | 0.03455 | 0.007256 | 0.007951 |
| Ca | rs13291723 | rs10869985 | C | T | 9 | GNAQ | Female only | True | 0.3168 | 0.3148 | 0.3092 | 0.3089 | 34341 | 26934 | 168077 | 147076 | 0.03962 | 0.01385 | 0.02103 | 0.03482 | 0.007251 | 0.007937 |
| Ca | rs13291723 | rs4392969 | A | G | 9 | GNAQ | Female only | True | 0.3172 | 0.3153 | 0.3095 | 0.3092 | 34341 | 26934 | 168077 | 147076 | 0.04028 | 0.01316 | 0.02102 | 0.03476 | 0.00725 | 0.007937 |
| Ca | rs13291723 | rs10869986 | T | C | 9 | GNAQ | Female only | True | 0.3174 | 0.3153 | 0.3096 | 0.3093 | 34341 | 26934 | 168077 | 147076 | 0.04011 | 0.01378 | 0.02104 | 0.03479 | 0.00725 | 0.007938 |
| Ca | rs13291723 | rs10869987 | C | A | 9 | GNAQ | Female only | True | 0.3173 | 0.3153 | 0.3096 | 0.3093 | 34341 | 26934 | 168077 | 147076 | 0.03976 | 0.01375 | 0.02101 | 0.03473 | 0.007249 | 0.007936 |
| Ca | rs13291723 | rs4443727 | G | A | 9 | GNAQ | Female only | True | 0.3161 | 0.3142 | 0.3087 | 0.3085 | 34341 | 26934 | 168077 | 147076 | 0.04005 | 0.01372 | 0.02111 | 0.0346 | 0.007253 | 0.007947 |
| Ca | rs13291723 | rs17196333 | A | G | 9 | GNAQ | Female only | True | 0.3143 | 0.3121 | 0.3075 | 0.3074 | 34341 | 26934 | 168077 | 147076 | 0.03898 | 0.01422 | 0.0208 | 0.03443 | 0.007263 | 0.00796 |
| trait | lead_snp | ID | ALLELE1 | ALLELE0 | CHR | Gene_Symbol | independent_loci_in_sex_P1e2 | in_embl_any_or_gene | A1FREQ_VUMC_female | A1FREQ_VUMC_male | A1FREQ_UKB_female | A1FREQ_UKB_male | N_VUMC_female | N_VUMC_male | N_UKB_female | N_UKB_male | BETA_VUMC_female | BETA_VUMC_male | BETA_UKB_female | BETA_UKB_male | SE_VUMC_female | SE_VUMC_male |
| Ca | rs13291723 | rs4367623 | G | A | 9 | GNAQ | Female only | True | 0.3157 | 0.3136 | 0.3085 | 0.3083 | 34341 | 26934 | 168077 | 147076 | 0.03952 | 0.01351 | 0.02102 | 0.03451 | 0.007257 | 0.007953 |
| Ca | rs13291723 | rs11145614 | C | T | 9 | GNAQ | Female only | True | 0.4685 | 0.4685 | 0.4744 | 0.474 | 34341 | 26934 | 168077 | 147076 | -0.03232 | -0.008672 | -0.01807 | -0.03133 | 0.006754 | 0.00736 |
| Ca | rs13291723 | rs7020256 | T | C | 9 | GNAQ | Female only | True | 0.3154 | 0.3133 | 0.3078 | 0.3077 | 34341 | 26934 | 168077 | 147076 | 0.03904 | 0.01406 | 0.02095 | 0.0344 | 0.007261 | 0.007958 |
| Ca | rs13291723 | rs7028873 | T | C | 9 | GNAQ | Female only | True | 0.3151 | 0.313 | 0.3079 | 0.3078 | 34341 | 26934 | 168077 | 147076 | 0.03839 | 0.01443 | 0.02059 | 0.03459 | 0.007271 | 0.007963 |
| Ca | rs13291723 | rs4744855 | C | T | 9 | GNAQ | Female only | True | 0.4669 | 0.4665 | 0.4729 | 0.4725 | 34341 | 26934 | 168077 | 147076 | -0.03223 | -0.008776 | -0.01808 | -0.03091 | 0.006773 | 0.007385 |
| Ca | rs13291723 | rs11145634 | G | A | 9 | GNAQ | Female only | True | 0.4668 | 0.4664 | 0.4729 | 0.4725 | 34341 | 26934 | 168077 | 147076 | -0.03197 | -0.008929 | -0.01818 | -0.03094 | 0.006774 | 0.007385 |
| Ca | rs13291723 | rs35297051 | A | G | 9 | GNAQ | Female only | True | 0.4602 | 0.4595 | 0.4675 | 0.4672 | 34341 | 26934 | 168077 | 147076 | -0.03307 | -0.00925 | -0.0172 | -0.03065 | 0.006787 | 0.007402 |
| Ca | rs13291723 | rs60501721 | T | C | 9 | GNAQ | Female only | True | 0.4651 | 0.4645 | 0.4725 | 0.4722 | 34341 | 26934 | 168077 | 147076 | -0.03195 | -0.009525 | -0.01841 | -0.03083 | 0.006787 | 0.007397 |
| Ca | rs13291723 | rs17787178 | C | T | 9 | GNAQ | Female only | True | 0.4634 | 0.4626 | 0.4665 | 0.4665 | 34341 | 26934 | 168077 | 147076 | -0.03281 | -0.00914 | -0.01797 | -0.03174 | 0.00674 | 0.007348 |
| Ca | rs13291723 | rs4745686 | G | A | 9 | GNAQ | Female only | True | 0.4678 | 0.467 | 0.4667 | 0.4667 | 34341 | 26934 | 168077 | 147076 | -0.03264 | -0.008908 | -0.01789 | -0.03168 | 0.006677 | 0.007279 |
| Ca | rs13291723 | rs10870005 | C | T | 9 | GNAQ | Female only | True | 0.4659 | 0.4654 | 0.4688 | 0.4687 | 34341 | 26934 | 168077 | 147076 | -0.03244 | -0.00881 | -0.01826 | -0.03133 | 0.006741 | 0.007344 |
| Ca | rs13291723 | rs7028424 | A | G | 9 | GNAQ | Female only | True | 0.4662 | 0.4655 | 0.4686 | 0.4684 | 34341 | 26934 | 168077 | 147076 | -0.03223 | -0.008969 | -0.01832 | -0.03133 | 0.006741 | 0.007349 |
| Ca | rs13291723 | rs11145656 | T | C | 9 | GNAQ | Female only | True | 0.4629 | 0.4627 | 0.4652 | 0.4652 | 34341 | 26934 | 168077 | 147076 | -0.03041 | -0.00969 | -0.01846 | -0.03066 | 0.006747 | 0.007351 |

| trait | lead_snp | ID | ALLELE1 | ALLELE0 | CHR | Gene_Symbol | independent_loci_in_sex_P1e2 | in_embl_any_or_gene | A1FREQ_VUMC_female | A1FREQ_VUMC_male | A1FREQ_UKB_female | A1FREQ_UKB_male | N_VUMC_female | N_VUMC_male | N_UKB_female | N_UKB_male | BETA_VUMC_female | BETA_VUMC_male | BETA_UKB_female | BETA_UKB_male | SE_VUMC_female | SE_VUMC_male |
|---|---|---|---|---|---|---|---|---|---|---|---|---|---|---|---|---|---|---|---|---|---|---|
| Ca | rs13291723 | rs914787 | T | C | 9 | GNAQ | Female only | True | 0.4625 | 0.4627 | 0.4667 | 0.4668 | 34341 | 26934 | 168077 | 147076 | -0.03064 | -0.007739 | -0.01727 | -0.03054 | 0.006738 | 0.007337 |
| Ca | rs13291723 | rs914788 | G | A | 9 | GNAQ | Female only | True | 0.3158 | 0.313 | 0.312 | 0.3115 | 34341 | 26934 | 168077 | 147076 | 0.04083 | 0.01076 | 0.02064 | 0.03282 | 0.007231 | 0.007946 |
| Ca | rs13291723 | rs4745695 | C | A | 9 | GNAQ | Female only | True | 0.466 | 0.4658 | 0.4689 | 0.4687 | 34341 | 26934 | 168077 | 147076 | -0.03158 | -0.008061 | -0.01728 | -0.03117 | 0.006733 | 0.00733 |
| Ca | rs13291723 | rs10870020 | A | G | 9 | GNAQ | Female only | True | 0.4666 | 0.4663 | 0.4686 | 0.4685 | 34341 | 26934 | 168077 | 147076 | -0.03126 | -0.007483 | -0.01716 | -0.03121 | 0.006729 | 0.007324 |
| Ca | rs13291723 | rs62573096 | A | G | 9 | GNAQ | Female only | True | 0.4658 | 0.4658 | 0.4676 | 0.4675 | 34341 | 26934 | 168077 | 147076 | -0.0312 | -0.007464 | -0.01698 | -0.03113 | 0.006727 | 0.007315 |
| Ca | rs13291723 | rs13294734 | T | C | 9 | GNAQ | Female only | True | 0.4657 | 0.4658 | 0.4676 | 0.4675 | 34341 | 26934 | 168077 | 147076 | -0.03107 | -0.007127 | -0.01708 | -0.03103 | 0.006727 | 0.007317 |
| Ca | rs13291723 | rs1930548 | C | A | 9 | GNAQ | Female only | True | 0.4674 | 0.4672 | 0.4682 | 0.4679 | 34341 | 26934 | 168077 | 147076 | -0.0321 | -0.007211 | -0.01695 | -0.03134 | 0.006718 | 0.00731 |
| Ca | rs13291723 | rs958968 | C | A | 9 | GNAQ | Female only | True | 0.4695 | 0.4698 | 0.4667 | 0.4665 | 34341 | 26934 | 168077 | 147076 | -0.03177 | -0.006549 | -0.01691 | -0.03161 | 0.006676 | 0.00726 |
| Ca | rs13291723 | rs10512069 | T | C | 9 | GNAQ | Female only | True | 0.4712 | 0.4711 | 0.4685 | 0.4681 | 34341 | 26934 | 168077 | 147076 | -0.03134 | -0.006257 | -0.017 | -0.03128 | 0.006671 | 0.00726 |
| Ca | rs13291723 | rs7038083 | G | A | 9 | GNAQ | Female only | True | 0.4688 | 0.4687 | 0.4689 | 0.4686 | 34341 | 26934 | 168077 | 147076 | -0.03159 | -0.005764 | -0.0169 | -0.03155 | 0.006716 | 0.007309 |
| Ca | rs4837096 | rs10760436 | G | A | 9 | RP11-205K6.1 | Female only | False | 0.4326 | 0.4326 | 0.4349 | 0.4353 | 34341 | 26934 | 168077 | 147076 | 0.0259 | 0.002507 | 0.009779 | 0.005164 | 0.006771 | 0.007289 |
| Ca | rs4837096 | rs1548787 | A | G | 9 | RP11-205K6.1 | Female only | False | 0.4813 | 0.4777 | 0.4834 | 0.4815 | 34341 | 26934 | 168077 | 147076 | -0.0241 | 2.59e-04 | -0.004949 | -4.49e-04 | 0.006736 | 0.007228 |
| Ca | rs4837096 | rs10987305 | G | A | 9 | RP11-205K6.1 | Female only | False | 0.4802 | 0.4765 | 0.4832 | 0.4815 | 34341 | 26934 | 168077 | 147076 | -0.02245 | 0.001165 | -0.004351 | -7.66e-05 | 0.006723 | 0.007219 |
| Ca | rs4837096 | rs28619010 | C | T | 9 | RP11-205K6.1 | Female only | False | 0.4845 | 0.4807 | 0.4871 | 0.4853 | 34341 | 26934 | 168077 | 147076 | -0.02433 | 0.001504 | -0.004471 | -2.03e-04 | 0.006734 | 0.007218 |
| Ca | rs4837096 | rs9697125 | C | T | 9 | RP11-205K6.1 | Female only | False | 0.4818 | 0.4781 | 0.4851 | 0.4832 | 34341 | 26934 | 168077 | 147076 | -0.02444 | 0.002249 | -0.0051 | 1.09e-04 | 0.006736 | 0.007222 |
| Ca | rs4837096 | rs10987308 | C | T | 9 | RP11-205K6.1 | Female only | False | 0.4816 | 0.4779 | 0.4851 | 0.4832 | 34341 | 26934 | 168077 | 147076 | -0.02446 | 0.002163 | -0.005068 | 9.53e-05 | 0.006737 | 0.007221 |
| Ca | rs4837096 | rs28641758 | T | C | 9 | RP11-205K6.1 | Female only | False | 0.4766 | 0.4734 | 0.4794 | 0.4776 | 34341 | 26934 | 168077 | 147076 | -0.02475 | 0.002386 | -0.006739 | -2.71e-04 | 0.006736 | 0.007226 |
| Ca | rs4837096 | rs45520335 | A | G | 9 | RP11-205K6.1 | Female only | False | 0.4768 | 0.4736 | 0.4795 | 0.4777 | 34341 | 26934 | 168077 | 147076 | -0.02469 | 0.002371 | -0.006716 | -2.51e-04 | 0.006736 | 0.007225 |
| Ca | rs4837096 | rs2417023 | A | G | 9 | RP11-205K6.1 | Female only | False | 0.3888 | 0.3889 | 0.3963 | 0.3952 | 34341 | 26934 | 168077 | 147076 | -0.02916 | 0.0121 | -0.01219 | -0.001495 | 0.006871 | 0.007424 |
| Ca | rs4837096 | rs4837096 | C | T | 9 | RP11-205K6.1 | Female only | False | 0.4386 | 0.4407 | 0.4379 | 0.4387 | 34341 | 26934 | 168077 | 147076 | 0.03692 | -0.002856 | 0.01377 | 0.00568 | 0.006757 | 0.007264 |
| Creat | rs6926186 | rs6904762 | G | A | 6 | L3MBTL3 | Female only | True | 0.3051 | 0.3065 | 0.3091 | 0.3088 | 34787 | 27025 | 184912 | 159192 | 0.03717 | 0.001414 | 0.02007 | 0.02171 | 0.008378 | 0.00965 |
| Creat | rs6926186 | rs7754426 | A | G | 6 | L3MBTL3 | Female only | True | 0.2948 | 0.2963 | 0.2984 | 0.2985 | 34787 | 27025 | 184912 | 159192 | 0.03773 | 0.00309 | 0.02301 | 0.02234 | 0.00852 | 0.009811 |
| Creat | rs6926186 | rs1337776 | A | G | 6 | L3MBTL3 | Female only | True | 0.3061 | 0.3079 | 0.3078 | 0.3076 | 34787 | 27025 | 184912 | 159192 | 0.03696 | 0.002337 | 0.02046 | 0.02147 | 0.008418 | 0.009694 |
| Creat | rs6926186 | rs6569646 | T | C | 6 | L3MBTL3 | Female only | True | 0.295 | 0.2966 | 0.2988 | 0.2989 | 34787 | 27025 | 184912 | 159192 | 0.03786 | 0.003665 | 0.02294 | 0.02225 | 0.008523 | 0.009813 |
| Creat | rs6926186 | rs6569648 | C | T | 6 | L3MBTL3 | Female only | True | 0.2312 | 0.2314 | 0.2405 | 0.241 | 34787 | 27025 | 184912 | 159192 | 0.04665 | 8.53e-05 | 0.02878 | 0.02438 | 0.009144 | 0.01061 |
| Creat | rs6926186 | rs6926186 | G | A | 6 | L3MBTL3 | Female only | True | 0.2986 | 0.2998 | 0.3046 | 0.3043 | 34787 | 27025 | 184912 | 159192 | 0.04694 | -0.005358 | 0.02703 | 0.02159 | 0.008505 | 0.009842 |
| Creat | rs6926186 | rs9402211 | T | C | 6 | L3MBTL3 | Female only | True | 0.3019 | 0.3036 | 0.3046 | 0.3042 | 34787 | 27025 | 184912 | 159192 | 0.04591 | -0.007482 | 0.027 | 0.02149 | 0.008466 | 0.009789 |
| Creat | rs6926186 | rs9388766 | T | C | 6 | L3MBTL3 | Female only | True | 0.2985 | 0.3004 | 0.303 | 0.3026 | 34787 | 27025 | 184912 | 159192 | 0.04643 | -0.005816 | 0.02693 | 0.02127 | 0.0085 | 0.009823 |
| Creat | rs6926186 | rs9375694 | G | A | 6 | L3MBTL3 | Female only | True | 0.3006 | 0.3029 | 0.3035 | 0.3029 | 34787 | 27025 | 184912 | 159192 | 0.04631 | -0.007052 | 0.02694 | 0.02099 | 0.008468 | 0.009785 |

| trait | lead_snp | ID | ALLELE1 | ALLELE0 | CHR | Gene_Symbol | independent_loci_in_sex_P1e2 | in_embl_any_or_gene | A1FREQ_VUMC_female | A1FREQ_VUMC_male | A1FREQ_UKB_female | A1FREQ_UKB_male | N_VUMC_female | N_VUMC_male | N_UKB_female | N_UKB_male | BETA_VUMC_female | BETA_VUMC_male | BETA_UKB_female | BETA_UKB_male | SE_VUMC_female | SE_VUMC_male |
|---|---|---|---|---|---|---|---|---|---|---|---|---|---|---|---|---|---|---|---|---|---|---|
| Creat | rs6926186 | rs9388767 | G | A | 6 | L3MBTL3 | Female only | True | 0.3007 | 0.303 | 0.3035 | 0.303 | 34787 | 27025 | 184912 | 159192 | 0.04618 | -0.006605 | 0.02688 | 0.02106 | 0.008468 | 0.009785 |
| Creat | rs6926186 | rs59084494 | G | A | 6 | L3MBTL3 | Female only | True | 0.4205 | 0.4251 | 0.4193 | 0.4184 | 34787 | 27025 | 184912 | 159192 | 0.03704 | -0.001706 | 0.0223 | 0.0172 | 0.007845 | 0.00908 |
| Creat | rs6926186 | rs6899976 | G | A | 6 | L3MBTL3 | Female only | True | 0.2978 | 0.3001 | 0.3035 | 0.3029 | 34787 | 27025 | 184912 | 159192 | 0.04593 | -0.00652 | 0.0269 | 0.02107 | 0.008435 | 0.009746 |
| Creat | rs6926186 | rs9375695 | G | T | 6 | L3MBTL3 | Female only | True | 0.3006 | 0.3028 | 0.3036 | 0.303 | 34787 | 27025 | 184912 | 159192 | 0.04644 | -0.005811 | 0.02683 | 0.02099 | 0.008472 | 0.009788 |
| Creat | rs6926186 | rs9375696 | C | T | 6 | L3MBTL3 | Female only | True | 0.4205 | 0.4251 | 0.4194 | 0.4185 | 34787 | 27025 | 184912 | 159192 | 0.03732 | -0.001645 | 0.02226 | 0.01713 | 0.007846 | 0.009081 |
| Creat | rs6926186 | rs11154519 | G | A | 6 | L3MBTL3 | Female only | True | 0.4306 | 0.4339 | 0.4284 | 0.4275 | 34787 | 27025 | 184912 | 159192 | 0.03479 | -0.001235 | 0.02219 | 0.01658 | 0.007831 | 0.009057 |
| Creat | rs6926186 | rs4897361 | G | T | 6 | L3MBTL3 | Female only | True | 0.4304 | 0.4339 | 0.4284 | 0.4275 | 34787 | 27025 | 184912 | 159192 | 0.03505 | -0.001498 | 0.02216 | 0.01656 | 0.007833 | 0.00906 |
| Creat | rs6926186 | rs4895864 | C | T | 6 | L3MBTL3 | Female only | True | 0.43 | 0.4335 | 0.4279 | 0.4271 | 34787 | 27025 | 184912 | 159192 | 0.03443 | -0.001422 | 0.02236 | 0.0165 | 0.007838 | 0.009063 |
| Creat | rs6926186 | rs9398934 | A | G | 6 | L3MBTL3 | Female only | True | 0.4303 | 0.4339 | 0.4283 | 0.4274 | 34787 | 27025 | 184912 | 159192 | 0.03487 | -0.001561 | 0.02209 | 0.01655 | 0.007833 | 0.009061 |
| Creat | rs6926186 | rs9375697 | C | T | 6 | L3MBTL3 | Female only | True | 0.4303 | 0.4338 | 0.4283 | 0.4274 | 34787 | 27025 | 184912 | 159192 | 0.03486 | -0.00161 | 0.02212 | 0.01656 | 0.007833 | 0.009061 |
| Creat | rs6926186 | rs9398935 | G | A | 6 | L3MBTL3 | Female only | True | 0.4304 | 0.4339 | 0.4284 | 0.4274 | 34787 | 27025 | 184912 | 159192 | 0.03488 | -0.001679 | 0.02216 | 0.01658 | 0.007833 | 0.00906 |
| Creat | rs6926186 | rs4594967 | A | G | 6 | L3MBTL3 | Female only | True | 0.3263 | 0.328 | 0.327 | 0.3261 | 34787 | 27025 | 184912 | 159192 | 0.0422 | -0.007402 | 0.02337 | 0.01858 | 0.008283 | 0.009592 |
| Creat | rs6926186 | rs4419687 | C | T | 6 | L3MBTL3 | Female only | True | 0.4299 | 0.4333 | 0.4279 | 0.4271 | 34787 | 27025 | 184912 | 159192 | 0.03379 | -0.001439 | 0.02203 | 0.01661 | 0.007834 | 0.009066 |
| Creat | rs6926186 | rs7744830 | T | G | 6 | L3MBTL3 | Female only | True | 0.3082 | 0.3092 | 0.3119 | 0.3115 | 34787 | 27025 | 184912 | 159192 | 0.04283 | -0.004672 | 0.02652 | 0.02012 | 0.008425 | 0.009743 |
| Creat | rs6926186 | rs7763108 | G | T | 6 | L3MBTL3 | Female only | True | 0.3263 | 0.3281 | 0.3271 | 0.3262 | 34787 | 27025 | 184912 | 159192 | 0.04159 | -0.007439 | 0.02341 | 0.01842 | 0.008282 | 0.009594 |
| Creat | rs6926186 | rs6928313 | C | T | 6 | L3MBTL3 | Female only | True | 0.4299 | 0.4333 | 0.4279 | 0.427 | 34787 | 27025 | 184912 | 159192 | 0.03381 | -0.00144 | 0.02203 | 0.01663 | 0.007834 | 0.009066 |
| Creat | rs6926186 | rs9375698 | A | G | 6 | L3MBTL3 | Female only | True | 0.3263 | 0.3281 | 0.3271 | 0.3263 | 34787 | 27025 | 184912 | 159192 | 0.04159 | -0.007381 | 0.02342 | 0.01844 | 0.008282 | 0.009592 |
| Creat | rs6926186 | rs4404788 | G | A | 6 | L3MBTL3 | Female only | True | 0.3263 | 0.3281 | 0.3271 | 0.3263 | 34787 | 27025 | 184912 | 159192 | 0.04162 | -0.00741 | 0.02346 | 0.01841 | 0.008281 | 0.009591 |
| trait | lead_snp | ID | ALLELE1 | ALLELE0 | CHR | Gene_Symbol | independent_loci_in_sex_P1e2 | in_embl_any_or_gene | A1FREQ_VUMC_female | A1FREQ_VUMC_male | A1FREQ_UKB_female | A1FREQ_UKB_male | N_VUMC_female | N_VUMC_male | N_UKB_female | N_UKB_male | BETA_VUMC_female | BETA_VUMC_male | BETA_UKB_female | BETA_UKB_male | SE_VUMC_female | SE_VUMC_male |
| Creat | rs6926186 | rs7755652 | T | G | 6 | L3MBTL3 | Female only | True | 0.4295 | 0.433 | 0.4279 | 0.427 | 34787 | 27025 | 184912 | 159192 | 0.03381 | -0.001601 | 0.02209 | 0.01664 | 0.007834 | 0.00907 |
| Creat | rs6926186 | rs4279458 | T | C | 6 | L3MBTL3 | Female only | True | 0.3262 | 0.328 | 0.3271 | 0.3263 | 34787 | 27025 | 184912 | 159192 | 0.04157 | -0.00747 | 0.02346 | 0.01843 | 0.008282 | 0.009591 |
| Creat | rs6926186 | rs4321845 | C | T | 6 | L3MBTL3 | Female only | True | 0.3262 | 0.328 | 0.3271 | 0.3263 | 34787 | 27025 | 184912 | 159192 | 0.04157 | -0.00747 | 0.02346 | 0.01843 | 0.008282 | 0.009591 |
| Creat | rs6926186 | rs3890746 | C | T | 6 | L3MBTL3 | Female only | True | 0.4296 | 0.4331 | 0.4279 | 0.427 | 34787 | 27025 | 184912 | 159192 | 0.03381 | -0.001388 | 0.0221 | 0.01664 | 0.007833 | 0.009069 |
| Creat | rs6926186 | rs9385532 | T | C | 6 | L3MBTL3 | Female only | True | 0.3263 | 0.3281 | 0.3271 | 0.3263 | 34787 | 27025 | 184912 | 159192 | 0.04161 | -0.007557 | 0.02348 | 0.01843 | 0.008281 | 0.009592 |
| Creat | rs6926186 | rs7746589 | G | A | 6 | L3MBTL3 | Female only | True | 0.3263 | 0.3281 | 0.3271 | 0.3263 | 34787 | 27025 | 184912 | 159192 | 0.04162 | -0.007406 | 0.02346 | 0.01844 | 0.008281 | 0.009591 |
| Creat | rs6926186 | rs6914670 | A | G | 6 | L3MBTL3 | Female only | True | 0.3103 | 0.3116 | 0.3124 | 0.312 | 34787 | 27025 | 184912 | 159192 | 0.04243 | -0.005755 | 0.02655 | 0.02009 | 0.008402 | 0.009713 |
| Creat | rs6926186 | rs9402213 | A | G | 6 | L3MBTL3 | Female only | True | 0.3263 | 0.3281 | 0.3271 | 0.3263 | 34787 | 27025 | 184912 | 159192 | 0.04169 | -0.007486 | 0.02347 | 0.01844 | 0.008279 | 0.009589 |
| Creat | rs6926186 | rs9375699 | A | G | 6 | L3MBTL3 | Female only | True | 0.3263 | 0.3281 | 0.3271 | 0.3263 | 34787 | 27025 | 184912 | 159192 | 0.04169 | -0.007482 | 0.02348 | 0.01844 | 0.008279 | 0.009589 |

| trait | lead_snp | ID | ALLELE1 | ALLELE0 | CHR | Gene_Symbol | independent_loci_in_sex_P1e2 | in_embl_any_or_gene | A1FREQ_VUMC_female | A1FREQ_VUMC_male | A1FREQ_UKB_female | A1FREQ_UKB_male | N_VUMC_female | N_VUMC_male | N_UKB_female | N_UKB_male | BETA_VUMC_female | BETA_VUMC_male | BETA_UKB_female | BETA_UKB_male | SE_VUMC_female | SE_VUMC_male |
|---|---|---|---|---|---|---|---|---|---|---|---|---|---|---|---|---|---|---|---|---|---|---|
| Creat | rs6926186 | rs9385533 | T | C | 6 | L3MBTL3 | Female only | True | 0.3263 | 0.3281 | 0.3271 | 0.3263 | 34787 | 27025 | 184912 | 159192 | 0.04169 | -0.007486 | 0.02347 | 0.01844 | 0.008279 | 0.009589 |
| Creat | rs6926186 | rs9372954 | T | C | 6 | L3MBTL3 | Female only | True | 0.3263 | 0.3281 | 0.3271 | 0.3263 | 34787 | 27025 | 184912 | 159192 | 0.04169 | -0.007486 | 0.02347 | 0.01844 | 0.008279 | 0.009589 |
| Creat | rs6926186 | rs9388768 | C | A | 6 | L3MBTL3 | Female only | True | 0.3234 | 0.3253 | 0.3272 | 0.3264 | 34787 | 27025 | 184912 | 159192 | 0.0415 | -0.007511 | 0.02351 | 0.01851 | 0.008246 | 0.009551 |
| Creat | rs6926186 | rs4373379 | G | A | 6 | L3MBTL3 | Female only | True | 0.3262 | 0.328 | 0.327 | 0.3262 | 34787 | 27025 | 184912 | 159192 | 0.04172 | -0.007594 | 0.02345 | 0.0185 | 0.008281 | 0.009592 |
| Creat | rs6926186 | rs9398937 | A | G | 6 | L3MBTL3 | Female only | True | 0.4296 | 0.4329 | 0.4277 | 0.4268 | 34787 | 27025 | 184912 | 159192 | 0.03411 | -0.00132 | 0.02201 | 0.01647 | 0.007832 | 0.009069 |
| Creat | rs6926186 | rs6900473 | A | G | 6 | L3MBTL3 | Female only | True | 0.3102 | 0.3114 | 0.3122 | 0.3117 | 34787 | 27025 | 184912 | 159192 | 0.04274 | -0.005344 | 0.02649 | 0.01983 | 0.008402 | 0.009715 |
| Creat | rs6926186 | rs6923819 | T | C | 6 | L3MBTL3 | Female only | True | 0.3261 | 0.3279 | 0.327 | 0.3261 | 34787 | 27025 | 184912 | 159192 | 0.04205 | -0.006752 | 0.0234 | 0.01815 | 0.00828 | 0.009594 |
| Creat | rs6926186 | rs4364506 | A | G | 6 | L3MBTL3 | Female only | True | 0.2486 | 0.2487 | 0.2602 | 0.2611 | 34787 | 27025 | 184912 | 159192 | 0.04187 | 0.003522 | 0.02433 | 0.01607 | 0.009014 | 0.0104 |
| Creat | rs6926186 | rs9492440 | C | T | 6 | L3MBTL3 | Female only | True | 0.2487 | 0.2487 | 0.2602 | 0.2612 | 34787 | 27025 | 184912 | 159192 | 0.04143 | 0.003835 | 0.02428 | 0.01575 | 0.009008 | 0.01039 |
| Creat | rs6926186 | rs7759381 | A | G | 6 | L3MBTL3 | Female only | True | 0.2487 | 0.249 | 0.2603 | 0.2612 | 34787 | 27025 | 184912 | 159192 | 0.0411 | 0.004323 | 0.02444 | 0.01618 | 0.009007 | 0.01039 |
| Creat | rs6926186 | rs9492441 | T | C | 6 | L3MBTL3 | Female only | True | 0.2487 | 0.249 | 0.2602 | 0.2612 | 34787 | 27025 | 184912 | 159192 | 0.04106 | 0.004327 | 0.02441 | 0.01615 | 0.009007 | 0.01039 |
| Creat | rs6926186 | rs7769599 | G | A | 6 | L3MBTL3 | Female only | True | 0.2492 | 0.2495 | 0.2604 | 0.2614 | 34787 | 27025 | 184912 | 159192 | 0.04058 | 0.003865 | 0.02434 | 0.01589 | 0.008984 | 0.01036 |
| Creat | rs6926186 | rs7764762 | A | G | 6 | L3MBTL3 | Female only | True | 0.2487 | 0.2489 | 0.2602 | 0.2612 | 34787 | 27025 | 184912 | 159192 | 0.04065 | 0.004665 | 0.02438 | 0.01612 | 0.008988 | 0.01037 |
| Creat | rs6926186 | rs13209890 | G | T | 6 | L3MBTL3 | Female only | True | 0.2492 | 0.2495 | 0.2604 | 0.2613 | 34787 | 27025 | 184912 | 159192 | 0.04058 | 0.003586 | 0.02429 | 0.01605 | 0.008983 | 0.01036 |
| Creat | rs6926186 | rs9492442 | A | G | 6 | L3MBTL3 | Female only | True | 0.311 | 0.3137 | 0.3199 | 0.3192 | 34787 | 27025 | 184912 | 159192 | 0.03936 | -0.005974 | 0.0198 | 0.01262 | 0.008365 | 0.009667 |
| Creat | rs6926186 | rs4548027 | A | G | 6 | L3MBTL3 | Female only | True | 0.2499 | 0.2502 | 0.2604 | 0.2613 | 34787 | 27025 | 184912 | 159192 | 0.04038 | 0.00312 | 0.02435 | 0.01576 | 0.00895 | 0.01032 |
| Creat | rs6926186 | rs9492443 | T | C | 6 | L3MBTL3 | Female only | True | 0.2492 | 0.2495 | 0.2604 | 0.2613 | 34787 | 27025 | 184912 | 159192 | 0.04044 | 0.003559 | 0.02432 | 0.01581 | 0.008974 | 0.01035 |
| Creat | rs6926186 | rs12198331 | T | C | 6 | L3MBTL3 | Female only | True | 0.2492 | 0.2494 | 0.2604 | 0.2613 | 34787 | 27025 | 184912 | 159192 | 0.04041 | 0.003519 | 0.02431 | 0.01583 | 0.008981 | 0.01036 |
| Creat | rs6926186 | rs9492445 | A | G | 6 | L3MBTL3 | Female only | True | 0.2494 | 0.2496 | 0.2606 | 0.2615 | 34787 | 27025 | 184912 | 159192 | 0.04054 | 0.003507 | 0.02415 | 0.01593 | 0.008979 | 0.01036 |
| Creat | rs6926186 | rs9492446 | T | C | 6 | L3MBTL3 | Female only | True | 0.2493 | 0.2495 | 0.2605 | 0.2615 | 34787 | 27025 | 184912 | 159192 | 0.04063 | 0.003547 | 0.02419 | 0.01591 | 0.008979 | 0.01036 |
| Creat | rs6926186 | rs7762724 | G | A | 6 | L3MBTL3 | Female only | True | 0.2493 | 0.2495 | 0.2605 | 0.2615 | 34787 | 27025 | 184912 | 159192 | 0.04059 | 0.003469 | 0.02416 | 0.01592 | 0.008979 | 0.01036 |
| Creat | rs6926186 | rs6569651 | C | T | 6 | L3MBTL3 | Female only | True | 0.3101 | 0.3125 | 0.3189 | 0.3183 | 34787 | 27025 | 184912 | 159192 | 0.03909 | -0.006155 | 0.01965 | 0.01297 | 0.008384 | 0.009681 |
| Creat | rs6926186 | rs7774174 | G | A | 6 | L3MBTL3 | Female only | True | 0.3101 | 0.3124 | 0.3189 | 0.3183 | 34787 | 27025 | 184912 | 159192 | 0.03899 | -0.006113 | 0.01965 | 0.01298 | 0.008385 | 0.009682 |
| Creat | rs6926186 | rs7756339 | G | A | 6 | L3MBTL3 | Female only | True | 0.3022 | 0.3044 | 0.3129 | 0.3124 | 34787 | 27025 | 184912 | 159192 | 0.0364 | -0.003194 | 0.01782 | 0.009368 | 0.008434 | 0.009759 |
| Creat | rs6926186 | rs9492452 | T | C | 6 | L3MBTL3 | Female only | True | 0.2837 | 0.2846 | 0.2977 | 0.2976 | 34787 | 27025 | 184912 | 159192 | 0.03512 | -0.001094 | 0.02073 | 0.0102 | 0.008601 | 0.009931 |
| Creat | rs6926186 | rs7755589 | A | G | 6 | L3MBTL3 | Female only | True | 0.2841 | 0.2851 | 0.2976 | 0.2975 | 34787 | 27025 | 184912 | 159192 | 0.03485 | -0.001127 | 0.02074 | 0.0101 | 0.008584 | 0.009913 |
| Creat | rs6926186 | rs9492454 | C | T | 6 | L3MBTL3 | Female only | True | 0.2835 | 0.2845 | 0.2976 | 0.2976 | 34787 | 27025 | 184912 | 159192 | 0.03506 | -0.001551 | 0.0207 | 0.01007 | 0.008611 | 0.009944 |
| Height | rs2336723 | rs62253602 | T | C | 3 | RP11-894J14.5 | Female only | True | 0.3327 | 0.3327 | 0.3413 | 0.3407 | 34577 | 25698 | 193785 | 166603 | 0.02862 | -0.002392 | 0.02152 | 0.02123 | 0.007401 | 0.008655 |
| Height | rs2336723 | rs62253603 | A | G | 3 | RP11-894J14.5 | Female only | True | 0.3327 | 0.3328 | 0.3414 | 0.3408 | 34577 | 25698 | 193785 | 166603 | 0.02851 | -0.002112 | 0.02157 | 0.02138 | 0.007402 | 0.008652 |

| trait | lead_snp | ID | ALLELE1 | ALLELE0 | CHR | Gene_Symbol | independent_loci_in_sex_P1e2 | in_embl_any_or_gene | A1FREQ_VUMC_female | A1FREQ_VUMC_male | A1FREQ_UKB_female | A1FREQ_UKB_male | N_VUMC_female | N_VUMC_male | N_UKB_female | N_UKB_male | BETA_VUMC_female | BETA_VUMC_male | BETA_UKB_female | BETA_UKB_male | SE_VUMC_female | SE_VUMC_male |
|---|---|---|---|---|---|---|---|---|---|---|---|---|---|---|---|---|---|---|---|---|---|---|
| Height | rs2336723 | rs62253604 | T | C | 3 | RP11-894J14.5 | Female only | True | 0.3327 | 0.3328 | 0.3414 | 0.3409 | 34577 | 25698 | 193785 | 166603 | 0.02872 | -0.002433 | 0.02151 | 0.02128 | 0.0074 | 0.008652 |
| Height | rs2336723 | rs62253606 | G | A | 3 | RP11-894J14.5 | Female only | True | 0.3328 | 0.3329 | 0.3414 | 0.3408 | 34577 | 25698 | 193785 | 166603 | 0.02832 | -0.002213 | 0.02144 | 0.02111 | 0.007399 | 0.008647 |
| Height | rs2336723 | rs62253608 | T | C | 3 | RP11-894J14.5 | Female only | True | 0.3328 | 0.3329 | 0.3414 | 0.3408 | 34577 | 25698 | 193785 | 166603 | 0.02852 | -0.002566 | 0.02149 | 0.02117 | 0.007397 | 0.008646 |
| Height | rs2336723 | rs62253649 | G | A | 3 | RP11-894J14.5 | Female only | True | 0.3328 | 0.3329 | 0.3415 | 0.3409 | 34577 | 25698 | 193785 | 166603 | 0.02864 | -0.002245 | 0.02144 | 0.02117 | 0.007401 | 0.008648 |
| Height | rs2336723 | rs57916015 | G | T | 3 | RP11-894J14.5 | Female only | True | 0.3327 | 0.3328 | 0.3415 | 0.341 | 34577 | 25698 | 193785 | 166603 | 0.02884 | -0.002736 | 0.02145 | 0.02109 | 0.007402 | 0.00865 |
| Height | rs2336723 | rs35671032 | C | T | 3 | RP11-894J14.5 | Female only | True | 0.3327 | 0.3328 | 0.3415 | 0.341 | 34577 | 25698 | 193785 | 166603 | 0.0288 | -0.002803 | 0.02137 | 0.02111 | 0.007402 | 0.008649 |
| Height | rs2336723 | rs11130333 | G | A | 3 | RP11-894J14.5 | Female only | True | 0.3328 | 0.3329 | 0.3415 | 0.341 | 34577 | 25698 | 193785 | 166603 | 0.0287 | -0.002353 | 0.02147 | 0.02111 | 0.007401 | 0.008648 |
| Height | rs2336723 | rs11130335 | T | C | 3 | RP11-894J14.5 | Female only | True | 0.3296 | 0.3297 | 0.3377 | 0.3375 | 34577 | 25698 | 193785 | 166603 | 0.02951 | -0.003938 | 0.02218 | 0.02041 | 0.007421 | 0.008669 |
| Height | rs2336723 | rs12053904 | T | C | 3 | RP11-894J14.5 | Female only | True | 0.3303 | 0.3302 | 0.3379 | 0.3376 | 34577 | 25698 | 193785 | 166603 | 0.02935 | -0.003986 | 0.02221 | 0.0204 | 0.007416 | 0.008665 |
| Height | rs2336723 | rs2115779 | C | A | 3 | RP11-894J14.5 | Female only | True | 0.3302 | 0.3301 | 0.3379 | 0.3377 | 34577 | 25698 | 193785 | 166603 | 0.02948 | -0.004017 | 0.02227 | 0.02045 | 0.007418 | 0.008666 |
| Height | rs2336723 | rs56129941 | T | G | 3 | RP11-894J14.5 | Female only | True | 0.3292 | 0.3291 | 0.3373 | 0.3371 | 34577 | 25698 | 193785 | 166603 | 0.02958 | -0.003661 | 0.02215 | 0.0203 | 0.007423 | 0.008671 |
| Height | rs2336723 | rs56155510 | T | C | 3 | RP11-894J14.5 | Female only | True | 0.3293 | 0.3292 | 0.3373 | 0.3371 | 34577 | 25698 | 193785 | 166603 | 0.02958 | -0.00369 | 0.0221 | 0.0203 | 0.00742 | 0.00867 |
| Height | rs2336723 | rs55962821 | T | C | 3 | RP11-894J14.5 | Female only | True | 0.3291 | 0.3289 | 0.337 | 0.3368 | 34577 | 25698 | 193785 | 166603 | 0.02932 | -0.004089 | 0.0221 | 0.02023 | 0.007422 | 0.00867 |
| Height | rs2336723 | rs12635933 | A | G | 3 | RP11-894J14.5 | Female only | True | 0.3293 | 0.3291 | 0.3374 | 0.3372 | 34577 | 25698 | 193785 | 166603 | 0.02939 | -0.004273 | 0.02218 | 0.02022 | 0.007422 | 0.00867 |
| Height | rs2336723 | rs17304694 | A | G | 3 | RP11-894J14.5 | Female only | True | 0.3528 | 0.3529 | 0.3619 | 0.3615 | 34577 | 25698 | 193785 | 166603 | 0.02556 | -0.01038 | 0.01805 | 0.01722 | 0.007303 | 0.008536 |
| Height | rs2336723 | rs12633064 | C | T | 3 | RP11-894J14.5 | Female only | True | 0.3263 | 0.3258 | 0.3346 | 0.3345 | 34577 | 25698 | 193785 | 166603 | 0.0289 | -0.002644 | 0.02302 | 0.02133 | 0.007457 | 0.00872 |
| Height | rs2336723 | rs62255895 | C | T | 3 | RP11-894J14.5 | Female only | True | 0.3321 | 0.3319 | 0.3405 | 0.3404 | 34577 | 25698 | 193785 | 166603 | 0.02954 | -0.00424 | 0.02337 | 0.01997 | 0.007392 | 0.008642 |
| Height | rs2336723 | rs62255897 | G | A | 3 | RP11-894J14.5 | Female only | True | 0.3323 | 0.3323 | 0.3408 | 0.3408 | 34577 | 25698 | 193785 | 166603 | 0.02946 | -0.004405 | 0.02304 | 0.02007 | 0.007399 | 0.00865 |
| Height | rs2336723 | rs62255898 | A | G | 3 | RP11-894J14.5 | Female only | True | 0.3319 | 0.3317 | 0.3406 | 0.3405 | 34577 | 25698 | 193785 | 166603 | 0.0296 | -0.004416 | 0.02329 | 0.01982 | 0.007399 | 0.008648 |
| Height | rs2336723 | rs6781896 | C | T | 3 | RP11-894J14.5 | Female only | True | 0.3321 | 0.332 | 0.3406 | 0.3406 | 34577 | 25698 | 193785 | 166603 | 0.02933 | -0.004394 | 0.02294 | 0.0201 | 0.007394 | 0.008644 |
| Height | rs2336723 | rs9758988 | C | T | 3 | RP11-894J14.5 | Female only | True | 0.3326 | 0.3317 | 0.3403 | 0.3403 | 34577 | 25698 | 193785 | 166603 | 0.02865 | -0.005272 | 0.02332 | 0.02017 | 0.007386 | 0.008642 |
| Height | rs2336723 | rs1811351 | G | A | 3 | RP11-894J14.5 | Female only | True | 0.3318 | 0.3313 | 0.3401 | 0.3401 | 34577 | 25698 | 193785 | 166603 | 0.02888 | -0.005953 | 0.02312 | 0.02026 | 0.007391 | 0.008643 |
| trait | lead_snp | ID | ALLELE1 | ALLELE0 | CHR | Gene_Symbol | independent_loci_in_sex_P1e2 | in_embl_any_or_gene | A1FREQ_VUMC_female | A1FREQ_VUMC_male | A1FREQ_UKB_female | A1FREQ_UKB_male | N_VUMC_female | N_VUMC_male | N_UKB_female | N_UKB_male | BETA_VUMC_female | BETA_VUMC_male | BETA_UKB_female | BETA_UKB_male | SE_VUMC_female | SE_VUMC_male |
| Height | rs2336723 | rs2564924 | C | T | 3 | RP11-894J14.5 | Female only | True | 0.4419 | 0.4395 | 0.4437 | 0.4444 | 34577 | 25698 | 193785 | 166603 | 0.03855 | 0.006392 | 0.03554 | 0.03172 | 0.007037 | 0.008237 |
| Height | rs2336723 | rs2564923 | A | G | 3 | RP11-894J14.5 | Female only | True | 0.4409 | 0.4383 | 0.4433 | 0.444 | 34577 | 25698 | 193785 | 166603 | 0.03839 | 0.005781 | 0.0356 | 0.03152 | 0.007043 | 0.008247 |
| Height | rs2336723 | rs34530577 | G | T | 3 | RP11-894J14.5 | Female only | True | 0.4401 | 0.4372 | 0.4418 | 0.4425 | 34577 | 25698 | 193785 | 166603 | 0.03772 | 0.00574 | 0.03536 | 0.03085 | 0.007047 | 0.008259 |
| Height | rs2336723 | rs73082047 | T | G | 3 | RP11-894J14.5 | Female only | True | 0.3282 | 0.3265 | 0.336 | 0.336 | 34577 | 25698 | 193785 | 166603 | 0.03087 | -0.00569 | 0.02287 | 0.02027 | 0.007414 | 0.008697 |
| Height | rs2336723 | rs2336723 | T | C | 3 | RP11-894J14.5 | Female only | True | 0.4359 | 0.4328 | 0.4383 | 0.4388 | 34577 | 25698 | 193785 | 166603 | 0.0399 | 0.005413 | 0.03455 | 0.03039 | 0.007054 | 0.008254 |

| trait | lead_snp | ID | ALLELE1 | ALLELE0 | CHR | Gene_Symbol | independent_loci_in_sex_P1e2 | in_embl_any_or_gene | A1FREQ_VUMC_female | A1FREQ_VUMC_male | A1FREQ_UKB_female | A1FREQ_UKB_male | N_VUMC_female | N_VUMC_male | N_UKB_female | N_UKB_male | BETA_VUMC_female | BETA_VUMC_male | BETA_UKB_female | BETA_UKB_male | SE_VUMC_female | SE_VUMC_male |
|---|---|---|---|---|---|---|---|---|---|---|---|---|---|---|---|---|---|---|---|---|---|---|
| Height | rs2336723 | rs2564939 | T | C | 3 | RP11-894J14.5 | Female only | True | 0.3325 | 0.3302 | 0.3361 | 0.3361 | 34577 | 25698 | 193785 | 166603 | 0.03124 | -0.00533 | 0.02284 | 0.02012 | 0.00742 | 0.008708 |
| Height | rs2336723 | rs2564940 | G | A | 3 | RP11-894J14.5 | Female only | True | 0.4389 | 0.4358 | 0.441 | 0.4417 | 34577 | 25698 | 193785 | 166603 | 0.03891 | 0.006107 | 0.03492 | 0.03093 | 0.007044 | 0.008254 |
| Height | rs2336723 | rs2581798 | T | C | 3 | RP11-894J14.5 | Female only | True | 0.3341 | 0.3313 | 0.3371 | 0.3372 | 34577 | 25698 | 193785 | 166603 | 0.03031 | -0.005322 | 0.02348 | 0.02074 | 0.007406 | 0.008706 |
| Height | rs2336723 | rs2581800 | A | G | 3 | RP11-894J14.5 | Female only | True | 0.4363 | 0.4335 | 0.4391 | 0.4397 | 34577 | 25698 | 193785 | 166603 | 0.03933 | 0.006047 | 0.03429 | 0.03055 | 0.007061 | 0.008266 |
| Height | rs2336723 | rs1864367 | T | C | 3 | RP11-894J14.5 | Female only | True | 0.438 | 0.4345 | 0.4399 | 0.4404 | 34577 | 25698 | 193785 | 166603 | 0.03839 | 0.005245 | 0.03525 | 0.03054 | 0.007048 | 0.008255 |
| Height | rs2336723 | rs56386026 | T | C | 3 | RP11-894J14.5 | Female only | True | 0.3282 | 0.3267 | 0.336 | 0.336 | 34577 | 25698 | 193785 | 166603 | 0.02951 | -0.005886 | 0.02284 | 0.02031 | 0.007408 | 0.008698 |
| Height | rs2336723 | rs2253209 | A | C | 3 | RP11-894J14.5 | Female only | True | 0.4393 | 0.4362 | 0.441 | 0.4416 | 34577 | 25698 | 193785 | 166603 | 0.03795 | 0.006062 | 0.03495 | 0.03087 | 0.007043 | 0.008255 |
| Height | rs2336723 | rs2336724 | T | C | 3 | RP11-894J14.5 | Female only | True | 0.3368 | 0.3342 | 0.3385 | 0.3384 | 34577 | 25698 | 193785 | 166603 | 0.02928 | -0.00665 | 0.02356 | 0.02042 | 0.007392 | 0.008688 |
| Height | rs2336723 | rs2564929 | T | C | 3 | RP11-894J14.5 | Female only | True | 0.3296 | 0.3274 | 0.3361 | 0.3361 | 34577 | 25698 | 193785 | 166603 | 0.0306 | -0.00558 | 0.02289 | 0.0204 | 0.007384 | 0.008674 |
| Height | rs2336723 | rs2336725 | C | T | 3 | RP11-894J14.5 | Female only | True | 0.4368 | 0.4339 | 0.4416 | 0.4422 | 34577 | 25698 | 193785 | 166603 | 0.03773 | 0.006142 | 0.03526 | 0.0304 | 0.007007 | 0.008215 |
| Height | rs2336723 | rs930122 | T | C | 3 | RP11-894J14.5 | Female only | True | 0.3354 | 0.3331 | 0.3375 | 0.3373 | 34577 | 25698 | 193785 | 166603 | 0.02929 | -0.006829 | 0.02287 | 0.01975 | 0.0074 | 0.008689 |
| Height | rs2336723 | rs2253675 | T | C | 3 | RP11-894J14.5 | Female only | True | 0.3369 | 0.3344 | 0.3387 | 0.3386 | 34577 | 25698 | 193785 | 166603 | 0.02919 | -0.006494 | 0.02372 | 0.02027 | 0.007389 | 0.008684 |
| Height | rs2336723 | rs2253685 | G | A | 3 | RP11-894J14.5 | Female only | True | 0.4369 | 0.4339 | 0.4413 | 0.4419 | 34577 | 25698 | 193785 | 166603 | 0.03766 | 0.006121 | 0.0352 | 0.0307 | 0.007007 | 0.008215 |
| Height | rs2336723 | rs2253796 | C | T | 3 | RP11-894J14.5 | Female only | True | 0.4394 | 0.4363 | 0.4412 | 0.4417 | 34577 | 25698 | 193785 | 166603 | 0.03777 | 0.006084 | 0.03512 | 0.03086 | 0.007042 | 0.008254 |
| Height | rs2336723 | rs11130338 | C | T | 3 | RP11-894J14.5 | Female only | True | 0.4395 | 0.4365 | 0.4413 | 0.4419 | 34577 | 25698 | 193785 | 166603 | 0.03773 | 0.006214 | 0.03514 | 0.03066 | 0.007041 | 0.008253 |
| Height | rs2336723 | rs62255925 | A | G | 3 | RP11-894J14.5 | Female only | True | 0.3281 | 0.3264 | 0.336 | 0.3359 | 34577 | 25698 | 193785 | 166603 | 0.03032 | -0.005987 | 0.02286 | 0.02047 | 0.007407 | 0.008698 |
| Height | rs2336723 | rs4687701 | A | G | 3 | RP11-894J14.5 | Female only | True | 0.4395 | 0.4365 | 0.441 | 0.4416 | 34577 | 25698 | 193785 | 166603 | 0.03773 | 0.006214 | 0.03516 | 0.03082 | 0.007041 | 0.008253 |
| Height | rs2336723 | rs2581782 | T | C | 3 | RP11-894J14.5 | Female only | True | 0.4384 | 0.4351 | 0.4401 | 0.4405 | 34577 | 25698 | 193785 | 166603 | 0.03843 | 0.005268 | 0.0354 | 0.03035 | 0.007045 | 0.008253 |
| Height | rs2336723 | rs2581783 | G | A | 3 | RP11-894J14.5 | Female only | True | 0.4395 | 0.4365 | 0.4412 | 0.4418 | 34577 | 25698 | 193785 | 166603 | 0.03805 | 0.006223 | 0.03514 | 0.03069 | 0.007042 | 0.008253 |
| Height | rs2336723 | rs2581784 | A | G | 3 | RP11-894J14.5 | Female only | True | 0.4395 | 0.4366 | 0.4411 | 0.4417 | 34577 | 25698 | 193785 | 166603 | 0.03799 | 0.00621 | 0.03516 | 0.03065 | 0.007042 | 0.008252 |
| Height | rs2336723 | rs2082093 | G | A | 3 | RP11-894J14.5 | Female only | True | 0.4395 | 0.4366 | 0.4412 | 0.4418 | 34577 | 25698 | 193785 | 166603 | 0.03799 | 0.006213 | 0.03513 | 0.03066 | 0.007042 | 0.008253 |
| Height | rs2336723 | rs2099162 | C | T | 3 | RP11-894J14.5 | Female only | True | 0.3347 | 0.3322 | 0.3374 | 0.3374 | 34577 | 25698 | 193785 | 166603 | 0.02986 | -0.004928 | 0.02368 | 0.02078 | 0.007402 | 0.008706 |
| Height | rs2336723 | rs891368 | A | G | 3 | RP11-894J14.5 | Female only | True | 0.4394 | 0.4363 | 0.441 | 0.4416 | 34577 | 25698 | 193785 | 166603 | 0.0381 | 0.006057 | 0.03509 | 0.03086 | 0.007043 | 0.008254 |
| Height | rs2336723 | rs1078968 | G | A | 3 | RP11-894J14.5 | Female only | True | 0.4368 | 0.4339 | 0.4413 | 0.4419 | 34577 | 25698 | 193785 | 166603 | 0.03794 | 0.006143 | 0.03514 | 0.03071 | 0.007007 | 0.008215 |
| Height | rs2336723 | rs2564922 | G | A | 3 | RP11-894J14.5 | Female only | True | 0.3371 | 0.3345 | 0.3389 | 0.3388 | 34577 | 25698 | 193785 | 166603 | 0.02941 | -0.006595 | 0.02353 | 0.0206 | 0.007392 | 0.008691 |
| Height | rs2336723 | rs2564921 | T | C | 3 | RP11-894J14.5 | Female only | True | 0.4356 | 0.4322 | 0.4401 | 0.4404 | 34577 | 25698 | 193785 | 166603 | 0.03851 | 0.004964 | 0.03534 | 0.03057 | 0.007011 | 0.008217 |
| Height | rs2336723 | rs11242 | T | C | 3 | RP11-894J14.5 | Female only | True | 0.4367 | 0.4337 | 0.4411 | 0.4417 | 34577 | 25698 | 193785 | 166603 | 0.03813 | 0.005911 | 0.03511 | 0.0309 | 0.007008 | 0.008216 |
| Height | rs2336723 | rs2581786 | G | A | 3 | RP11-894J14.5 | Female only | True | 0.4395 | 0.4364 | 0.4411 | 0.4417 | 34577 | 25698 | 193785 | 166603 | 0.0381 | 0.00601 | 0.03508 | 0.03091 | 0.007043 | 0.008254 |
| Height | rs2336723 | rs2581787 | G | T | 3 | RP11-894J14.5 | Female only | True | 0.4399 | 0.4368 | 0.4412 | 0.4418 | 34577 | 25698 | 193785 | 166603 | 0.03789 | 0.00605 | 0.0351 | 0.03105 | 0.007044 | 0.008252 |

| trait | lead_snp | ID | ALLELE1 | ALLELE0 | CHR | Gene_Symbol | independent_loci_in_sex_P1e2 | in_embl_any_or_gene | A1FREQ_VUMC_female | A1FREQ_VUMC_male | A1FREQ_UKB_female | A1FREQ_UKB_male | N_VUMC_female | N_VUMC_male | N_UKB_female | N_UKB_male | BETA_VUMC_female | BETA_VUMC_male | BETA_UKB_female | BETA_UKB_male | SE_VUMC_female | SE_VUMC_male |
|---|---|---|---|---|---|---|---|---|---|---|---|---|---|---|---|---|---|---|---|---|---|---|
| Height | rs2336723 | rs56058643 | C | T | 3 | RP11-894J14.5 | Female only | True | 0.3281 | 0.3265 | 0.3359 | 0.3359 | 34577 | 25698 | 193785 | 166603 | 0.03032 | -0.005784 | 0.02287 | 0.02054 | 0.00741 | 0.008701 |
| Height | rs2336723 | rs55950520 | A | G | 3 | RP11-894J14.5 | Female only | True | 0.328 | 0.3263 | 0.3359 | 0.3359 | 34577 | 25698 | 193785 | 166603 | 0.03013 | -0.00581 | 0.02286 | 0.02048 | 0.007413 | 0.008703 |
| Height | rs2336723 | rs62255929 | T | C | 3 | RP11-894J14.5 | Female only | True | 0.328 | 0.3264 | 0.3358 | 0.3358 | 34577 | 25698 | 193785 | 166603 | 0.03024 | -0.006055 | 0.02289 | 0.02051 | 0.007412 | 0.008703 |
| Height | rs2336723 | rs80132892 | A | G | 3 | RP11-894J14.5 | Female only | True | 0.3279 | 0.3263 | 0.3359 | 0.3358 | 34577 | 25698 | 193785 | 166603 | 0.03012 | -0.005843 | 0.02286 | 0.02049 | 0.007414 | 0.008704 |
| Height | rs2336723 | rs7640294 | C | A | 3 | RP11-894J14.5 | Female only | True | 0.4392 | 0.4362 | 0.4408 | 0.4413 | 34577 | 25698 | 193785 | 166603 | 0.03786 | 0.006231 | 0.03517 | 0.03108 | 0.007044 | 0.008255 |
| Height | rs2336723 | rs2163167 | T | C | 3 | RP11-894J14.5 | Female only | True | 0.4396 | 0.4366 | 0.4409 | 0.4414 | 34577 | 25698 | 193785 | 166603 | 0.03801 | 0.005001 | 0.03523 | 0.03105 | 0.007051 | 0.008266 |
| Height | rs2336723 | rs2115782 | G | A | 3 | RP11-894J14.5 | Female only | True | 0.4397 | 0.4366 | 0.4411 | 0.4415 | 34577 | 25698 | 193785 | 166603 | 0.038 | 0.004942 | 0.03519 | 0.03102 | 0.007051 | 0.008266 |
| Height | rs2336723 | rs62255931 | G | A | 3 | RP11-894J14.5 | Female only | True | 0.327 | 0.3257 | 0.3352 | 0.3351 | 34577 | 25698 | 193785 | 166603 | 0.03076 | -0.007676 | 0.02293 | 0.02044 | 0.00743 | 0.00872 |
| Height | rs2336723 | rs2581829 | A | G | 3 | RP11-894J14.5 | Female only | True | 0.406 | 0.4027 | 0.4067 | 0.4069 | 34577 | 25698 | 193785 | 166603 | 0.03826 | 0.003884 | 0.03693 | 0.03062 | 0.007137 | 0.00835 |
| Height | rs2336723 | rs2581830 | T | C | 3 | RP11-894J14.5 | Female only | True | 0.4029 | 0.3997 | 0.4067 | 0.4068 | 34577 | 25698 | 193785 | 166603 | 0.03822 | 0.004258 | 0.03687 | 0.03058 | 0.007102 | 0.008311 |
| Height | rs2336723 | rs35783750 | T | C | 3 | RP11-894J14.5 | Female only | True | 0.4059 | 0.4026 | 0.4066 | 0.4068 | 34577 | 25698 | 193785 | 166603 | 0.03781 | 0.004038 | 0.03656 | 0.03061 | 0.007138 | 0.008351 |
| Height | rs2336723 | rs34454739 | A | G | 3 | RP11-894J14.5 | Female only | True | 0.4056 | 0.4023 | 0.4065 | 0.4067 | 34577 | 25698 | 193785 | 166603 | 0.03802 | 0.004179 | 0.03668 | 0.03061 | 0.007138 | 0.00835 |
| Height | rs2336723 | rs4395392 | T | C | 3 | RP11-894J14.5 | Female only | True | 0.407 | 0.404 | 0.4078 | 0.4081 | 34577 | 25698 | 193785 | 166603 | 0.03784 | 0.005331 | 0.03641 | 0.03098 | 0.007134 | 0.008347 |
| Height | rs2336723 | rs2564961 | G | A | 3 | RP11-894J14.5 | Female only | True | 0.407 | 0.4041 | 0.4079 | 0.4083 | 34577 | 25698 | 193785 | 166603 | 0.03774 | 0.00534 | 0.03653 | 0.03101 | 0.007134 | 0.008347 |
| Height | rs2336723 | rs891369 | A | G | 3 | RP11-894J14.5 | Female only | True | 0.293 | 0.2926 | 0.3008 | 0.3006 | 34577 | 25698 | 193785 | 166603 | 0.03392 | -0.005192 | 0.02651 | 0.02299 | 0.007657 | 0.008942 |
| Height | rs35214627 | rs227819 | C | T | 6 | SUPT3H | Female only | True | 0.2023 | 0.1999 | 0.1963 | 0.1964 | 34577 | 25698 | 193785 | 166603 | -0.03259 | -0.007384 | -0.00982 | -0.008831 | 0.008733 | 0.01024 |
| Height | rs524492 | rs227832 | C | T | 6 | SUPT3H | Female only | True | 0.2562 | 0.254 | 0.2495 | 0.2501 | 34577 | 25698 | 193785 | 166603 | -0.04253 | -0.001839 | -0.02693 | -0.01552 | 0.007951 | 0.009325 |
| Height | rs524492 | rs227835 | C | T | 6 | SUPT3H | Female only | True | 0.2544 | 0.2523 | 0.2492 | 0.2498 | 34577 | 25698 | 193785 | 166603 | -0.04322 | -0.001436 | -0.02687 | -0.01575 | 0.007977 | 0.009354 |
| Height | rs35214627 | rs35376827 | A | G | 6 | SUPT3H | Female only | True | 0.1574 | 0.1565 | 0.155 | 0.1554 | 34577 | 25698 | 193785 | 166603 | -0.04887 | -0.002264 | -0.01581 | -0.00904 | 0.009534 | 0.01116 |
| Height | rs35214627 | rs13206206 | C | T | 6 | SUPT3H | Female only | True | 0.1552 | 0.1542 | 0.1524 | 0.1526 | 34577 | 25698 | 193785 | 166603 | -0.05035 | -0.003164 | -0.01507 | -0.008283 | 0.009585 | 0.01122 |
| Height | rs524492 | rs227839 | G | A | 6 | SUPT3H | Female only | True | 0.2454 | 0.2427 | 0.2389 | 0.2398 | 34577 | 25698 | 193785 | 166603 | -0.04354 | -0.003057 | -0.02783 | -0.01511 | 0.00808 | 0.009463 |
| Height | rs35214627 | rs79355103 | G | T | 6 | SUPT3H | Female only | True | 0.1564 | 0.1556 | 0.1526 | 0.1527 | 34577 | 25698 | 193785 | 166603 | -0.04987 | -0.003247 | -0.01468 | -0.007539 | 0.009541 | 0.01117 |
| Height | rs524492 | rs227844 | C | A | 6 | SUPT3H | Female only | True | 0.248 | 0.2454 | 0.2422 | 0.2432 | 34577 | 25698 | 193785 | 166603 | -0.04154 | -0.003591 | -0.02798 | -0.01606 | 0.008059 | 0.009441 |
| Height | rs524492 | rs227845 | A | C | 6 | SUPT3H | Female only | True | 0.25 | 0.2473 | 0.2427 | 0.2438 | 34577 | 25698 | 193785 | 166603 | -0.0414 | -0.003013 | -0.02764 | -0.01596 | 0.008032 | 0.009419 |
| Height | rs35214627 | rs34577161 | G | A | 6 | SUPT3H | Female only | True | 0.2053 | 0.2022 | 0.1987 | 0.1988 | 34577 | 25698 | 193785 | 166603 | -0.04299 | -0.005083 | -0.02569 | -0.01533 | 0.008617 | 0.01011 |
| Height | rs524492 | rs227852 | T | C | 6 | SUPT3H | Female only | True | 0.2502 | 0.2475 | 0.2425 | 0.2435 | 34577 | 25698 | 193785 | 166603 | -0.04106 | -0.003726 | -0.02757 | -0.01575 | 0.00803 | 0.009424 |
| trait | lead_snp | ID | ALLELE1 | ALLELE0 | CHR | Gene_Symbol | independent_loci_in_sex_P1e2 | in_embl_any_or_gene | A1FREQ_VUMC_female | A1FREQ_VUMC_male | A1FREQ_UKB_female | A1FREQ_UKB_male | N_VUMC_female | N_VUMC_male | N_UKB_female | N_UKB_male | BETA_VUMC_female | BETA_VUMC_male | BETA_UKB_female | BETA_UKB_male | SE_VUMC_female | SE_VUMC_male |
| Height | rs524492 | rs227853 | C | T | 6 | SUPT3H | Female only | True | 0.2503 | 0.2475 | 0.2425 | 0.2435 | 34577 | 25698 | 193785 | 166603 | -0.04137 | -0.00334 | -0.02759 | -0.01576 | 0.00803 | 0.009423 |

| trait | lead_snp | ID | ALLELE1 | ALLELE0 | CHR | Gene_Symbol | independent_loci_in_sex_P1e2 | in_embl_any_or_gene | A1FREQ_VUMC_female | A1FREQ_VUMC_male | A1FREQ_UKB_female | A1FREQ_UKB_male | N_VUMC_female | N_VUMC_male | N_UKB_female | N_UKB_male | BETA_VUMC_female | BETA_VUMC_male | BETA_UKB_female | BETA_UKB_male | SE_VUMC_female | SE_VUMC_male |
|---|---|---|---|---|---|---|---|---|---|---|---|---|---|---|---|---|---|---|---|---|---|---|
| Height | rs35214627 | rs4714812 | A | G | 6 | SUPT3H | Female only | True | 0.205 | 0.2019 | 0.1987 | 0.1988 | 34577 | 25698 | 193785 | 166603 | -0.04291 | -0.005136 | -0.02556 | -0.01506 | 0.00862 | 0.01011 |
| Height | rs524492 | rs227854 | G | A | 6 | SUPT3H | Female only | True | 0.2693 | 0.2664 | 0.2581 | 0.259 | 34577 | 25698 | 193785 | 166603 | -0.03688 | -0.002268 | -0.0263 | -0.01759 | 0.007862 | 0.009188 |
| Height | rs524492 | rs609699 | A | G | 6 | SUPT3H | Female only | True | 0.252 | 0.2488 | 0.2446 | 0.2457 | 34577 | 25698 | 193785 | 166603 | -0.03992 | -0.003003 | -0.02767 | -0.01632 | 0.008014 | 0.009395 |
| Height | rs524492 | rs609643 | A | G | 6 | SUPT3H | Female only | True | 0.252 | 0.2488 | 0.2448 | 0.2458 | 34577 | 25698 | 193785 | 166603 | -0.03965 | -0.00285 | -0.02766 | -0.01633 | 0.008011 | 0.009393 |
| Height | rs524492 | rs687878 | C | T | 6 | SUPT3H | Female only | True | 0.2712 | 0.2679 | 0.2604 | 0.2613 | 34577 | 25698 | 193785 | 166603 | -0.03545 | -0.001709 | -0.02643 | -0.01815 | 0.00785 | 0.009163 |
| Height | rs524492 | rs3997503 | A | G | 6 | SUPT3H | Female only | True | 0.2516 | 0.2482 | 0.2447 | 0.2458 | 34577 | 25698 | 193785 | 166603 | -0.0394 | -0.002878 | -0.0276 | -0.01628 | 0.008018 | 0.009396 |
| Height | rs524492 | rs4714813 | A | C | 6 | SUPT3H | Female only | True | 0.2738 | 0.2705 | 0.2609 | 0.2619 | 34577 | 25698 | 193785 | 166603 | -0.03555 | -0.001845 | -0.02671 | -0.0184 | 0.007824 | 0.009139 |
| Height | rs35214627 | rs9472335 | A | G | 6 | SUPT3H | Female only | True | 0.2084 | 0.2049 | 0.2011 | 0.2012 | 34577 | 25698 | 193785 | 166603 | -0.04126 | -0.005936 | -0.02584 | -0.01591 | 0.008562 | 0.01005 |
| Height | rs35214627 | rs9463017 | A | C | 6 | SUPT3H | Female only | True | 0.2087 | 0.2052 | 0.2011 | 0.2013 | 34577 | 25698 | 193785 | 166603 | -0.04128 | -0.005916 | -0.02584 | -0.01603 | 0.008549 | 0.01004 |
| Height | rs35214627 | rs1418435 | C | A | 6 | SUPT3H | Female only | True | 0.2083 | 0.2048 | 0.201 | 0.2012 | 34577 | 25698 | 193785 | 166603 | -0.04132 | -0.005971 | -0.02591 | -0.01616 | 0.008564 | 0.01005 |
| Height | rs35214627 | rs9463019 | G | A | 6 | SUPT3H | Female only | True | 0.2084 | 0.2049 | 0.2011 | 0.2013 | 34577 | 25698 | 193785 | 166603 | -0.04124 | -0.005925 | -0.02587 | -0.01609 | 0.008563 | 0.01005 |
| Height | rs35214627 | rs9463020 | A | G | 6 | SUPT3H | Female only | True | 0.2212 | 0.219 | 0.213 | 0.2129 | 34577 | 25698 | 193785 | 166603 | -0.03846 | -0.006512 | -0.02665 | -0.01665 | 0.00838 | 0.009824 |
| Height | rs35214627 | rs143438084 | A | G | 6 | SUPT3H | Female only | True | 0.2084 | 0.2049 | 0.2011 | 0.2012 | 34577 | 25698 | 193785 | 166603 | -0.04129 | -0.005938 | -0.02573 | -0.01597 | 0.008563 | 0.01005 |
| Height | rs35214627 | rs35265671 | G | A | 6 | SUPT3H | Female only | True | 0.2048 | 0.2015 | 0.1973 | 0.1974 | 34577 | 25698 | 193785 | 166603 | -0.04168 | -0.004862 | -0.02623 | -0.0157 | 0.008662 | 0.01017 |
| Height | rs35214627 | rs7382444 | G | A | 6 | SUPT3H | Female only | True | 0.2083 | 0.2048 | 0.2011 | 0.2013 | 34577 | 25698 | 193785 | 166603 | -0.04124 | -0.0063 | -0.026 | -0.01603 | 0.008576 | 0.01007 |
| Height | rs35214627 | rs7381290 | G | T | 6 | SUPT3H | Female only | True | 0.2082 | 0.2047 | 0.2011 | 0.2013 | 34577 | 25698 | 193785 | 166603 | -0.04125 | -0.006305 | -0.026 | -0.01611 | 0.008576 | 0.01007 |
| Height | rs35214627 | rs72664272 | C | A | 6 | SUPT3H | Female only | True | 0.221 | 0.2189 | 0.213 | 0.2128 | 34577 | 25698 | 193785 | 166603 | -0.03826 | -0.007208 | -0.02673 | -0.01663 | 0.008393 | 0.009838 |
| Height | rs35214627 | rs9463039 | T | C | 6 | SUPT3H | Female only | True | 0.2086 | 0.2055 | 0.2015 | 0.2017 | 34577 | 25698 | 193785 | 166603 | -0.0413 | -0.007697 | -0.02621 | -0.01626 | 0.008574 | 0.01006 |
| Height | rs35214627 | rs9463040 | A | G | 6 | SUPT3H | Female only | True | 0.2087 | 0.2055 | 0.2015 | 0.2017 | 34577 | 25698 | 193785 | 166603 | -0.04127 | -0.007648 | -0.02621 | -0.01624 | 0.008572 | 0.01005 |
| Height | rs35214627 | rs2396326 | T | C | 6 | SUPT3H | Female only | True | 0.235 | 0.2334 | 0.225 | 0.2247 | 34577 | 25698 | 193785 | 166603 | -0.04522 | -0.0179 | -0.03246 | -0.02041 | 0.008207 | 0.009605 |
| Height | rs35214627 | rs2894599 | A | G | 6 | SUPT3H | Female only | True | 0.235 | 0.2335 | 0.225 | 0.2247 | 34577 | 25698 | 193785 | 166603 | -0.04539 | -0.01831 | -0.03244 | -0.02041 | 0.008207 | 0.009605 |
| Height | rs35214627 | rs66642874 | C | T | 6 | SUPT3H | Female only | True | 0.235 | 0.2335 | 0.225 | 0.2248 | 34577 | 25698 | 193785 | 166603 | -0.04534 | -0.01827 | -0.03255 | -0.02052 | 0.008207 | 0.009604 |
| Height | rs35214627 | rs4714825 | T | C | 6 | SUPT3H | Female only | True | 0.2109 | 0.21 | 0.2035 | 0.2037 | 34577 | 25698 | 193785 | 166603 | -0.04832 | -0.01345 | -0.03111 | -0.01843 | 0.008515 | 0.009977 |
| Height | rs35214627 | rs1935549 | T | C | 6 | SUPT3H | Female only | True | 0.2108 | 0.2099 | 0.2035 | 0.2037 | 34577 | 25698 | 193785 | 166603 | -0.04818 | -0.01334 | -0.03102 | -0.01852 | 0.008514 | 0.009975 |
| Height | rs35214627 | rs720989 | G | T | 6 | SUPT3H | Female only | True | 0.1988 | 0.1965 | 0.1919 | 0.1925 | 34577 | 25698 | 193785 | 166603 | -0.05229 | -0.01257 | -0.0307 | -0.01875 | 0.008704 | 0.01021 |
| Height | rs35214627 | rs720988 | T | C | 6 | SUPT3H | Female only | True | 0.1988 | 0.1966 | 0.1916 | 0.1921 | 34577 | 25698 | 193785 | 166603 | -0.05218 | -0.01259 | -0.03067 | -0.01844 | 0.008692 | 0.01019 |
| Height | rs35214627 | rs34817919 | T | C | 6 | SUPT3H | Female only | True | 0.2351 | 0.2335 | 0.2251 | 0.2248 | 34577 | 25698 | 193785 | 166603 | -0.04546 | -0.01791 | -0.03264 | -0.02064 | 0.008204 | 0.009602 |
| Height | rs35214627 | rs16872574 | T | G | 6 | SUPT3H | Female only | True | 0.2352 | 0.2338 | 0.225 | 0.2247 | 34577 | 25698 | 193785 | 166603 | -0.04491 | -0.018 | -0.03271 | -0.02021 | 0.008193 | 0.009585 |
| Height | rs35214627 | rs72664273 | A | G | 6 | SUPT3H | Female only | True | 0.2347 | 0.2333 | 0.2249 | 0.2246 | 34577 | 25698 | 193785 | 166603 | -0.04528 | -0.01843 | -0.03268 | -0.02021 | 0.00821 | 0.009605 |

| trait | lead_snp | ID | ALLELE1 | ALLELE0 | CHR | Gene_Symbol | independent_loci_in_sex_P1e2 | in_embl_any_or_gene | A1FREQ_VUMC_female | A1FREQ_VUMC_male | A1FREQ_UKB_female | A1FREQ_UKB_male | N_VUMC_female | N_VUMC_male | N_UKB_female | N_UKB_male | BETA_VUMC_female | BETA_VUMC_male | BETA_UKB_female | BETA_UKB_male | SE_VUMC_female | SE_VUMC_male |
|---|---|---|---|---|---|---|---|---|---|---|---|---|---|---|---|---|---|---|---|---|---|---|
| Height | rs35214627 | rs67597621 | G | T | 6 | SUPT3H | Female only | True | 0.2347 | 0.2333 | 0.2249 | 0.2246 | 34577 | 25698 | 193785 | 166603 | -0.04531 | -0.01843 | -0.03267 | -0.02024 | 0.008209 | 0.009604 |
| Height | rs35214627 | rs72664274 | G | A | 6 | SUPT3H | Female only | True | 0.2347 | 0.2333 | 0.2249 | 0.2246 | 34577 | 25698 | 193785 | 166603 | -0.04532 | -0.01838 | -0.03265 | -0.02025 | 0.008209 | 0.009604 |
| Height | rs35214627 | rs562016 | G | A | 6 | SUPT3H | Female only | True | 0.2162 | 0.2155 | 0.2041 | 0.2042 | 34577 | 25698 | 193785 | 166603 | -0.04862 | -0.01349 | -0.03126 | -0.01824 | 0.008522 | 0.009984 |
| Height | rs35214627 | rs7769218 | G | A | 6 | SUPT3H | Female only | True | 0.1983 | 0.1963 | 0.1917 | 0.1922 | 34577 | 25698 | 193785 | 166603 | -0.05195 | -0.01248 | -0.03084 | -0.01818 | 0.008713 | 0.01021 |
| Height | rs35214627 | rs35214627 | A | G | 6 | SUPT3H | Female only | True | 0.1386 | 0.139 | 0.1386 | 0.139 | 34577 | 25698 | 193785 | 166603 | -0.0611 | -0.01442 | -0.01849 | -0.01269 | 0.01012 | 0.01181 |
| Height | rs35214627 | rs2139863 | G | A | 6 | SUPT3H | Female only | True | 0.1974 | 0.1951 | 0.1906 | 0.191 | 34577 | 25698 | 193785 | 166603 | -0.05036 | -0.01111 | -0.03175 | -0.01868 | 0.00873 | 0.01024 |
| Height | rs524492 | rs524492 | T | C | 6 | SUPT3H | Female only | True | 0.2757 | 0.2742 | 0.2692 | 0.27 | 34577 | 25698 | 193785 | 166603 | -0.04327 | -0.01421 | -0.03498 | -0.01959 | 0.007777 | 0.009124 |
| Height | rs35214627 | rs3799965 | T | G | 6 | SUPT3H | Female only | True | 0.1961 | 0.1944 | 0.1901 | 0.1905 | 34577 | 25698 | 193785 | 166603 | -0.05115 | -0.01116 | -0.03203 | -0.01868 | 0.008749 | 0.01025 |
| Height | rs35214627 | rs3799966 | C | T | 6 | SUPT3H | Female only | True | 0.1962 | 0.1945 | 0.1901 | 0.1906 | 34577 | 25698 | 193785 | 166603 | -0.05127 | -0.01141 | -0.03201 | -0.01875 | 0.008747 | 0.01025 |
| Height | rs524492 | rs1402599 | A | G | 6 | SUPT3H | Female only | True | 0.281 | 0.2791 | 0.2698 | 0.2706 | 34577 | 25698 | 193785 | 166603 | -0.04192 | -0.01251 | -0.03493 | -0.01925 | 0.007795 | 0.009145 |
| Height | rs524492 | rs542444 | C | T | 6 | SUPT3H | Female only | True | 0.2089 | 0.2069 | 0.2019 | 0.2033 | 34577 | 25698 | 193785 | 166603 | -0.04557 | -0.006787 | -0.03309 | -0.01889 | 0.008527 | 0.01 |
| Height | rs524492 | rs9472376 | G | A | 6 | SUPT3H | Female only | True | 0.2463 | 0.2455 | 0.238 | 0.2376 | 34577 | 25698 | 193785 | 166603 | -0.03998 | -0.01622 | -0.03422 | -0.01972 | 0.008053 | 0.009447 |
| Height | rs35214627 | rs3799969 | G | T | 6 | SUPT3H | Female only | True | 0.176 | 0.1751 | 0.1691 | 0.1692 | 34577 | 25698 | 193785 | 166603 | -0.04679 | -0.01127 | -0.03299 | -0.01963 | 0.009118 | 0.01069 |
| Height | rs35214627 | rs3823252 | C | T | 6 | SUPT3H | Female only | True | 0.1772 | 0.1759 | 0.1696 | 0.1698 | 34577 | 25698 | 193785 | 166603 | -0.0455 | -0.01071 | -0.03269 | -0.0195 | 0.009098 | 0.01068 |
| Height | rs524492 | rs16869119 | G | A | 6 | SUPT3H | Female only | True | 0.1718 | 0.1708 | 0.1644 | 0.1644 | 34577 | 25698 | 193785 | 166603 | -0.04518 | -0.01098 | -0.0329 | -0.02067 | 0.009227 | 0.01083 |
| Height | rs35214627 | rs13202672 | T | C | 6 | SUPT3H | Female only | True | 0.1759 | 0.175 | 0.1691 | 0.1692 | 34577 | 25698 | 193785 | 166603 | -0.0466 | -0.01126 | -0.03289 | -0.01972 | 0.009118 | 0.01069 |
| Height | rs35214627 | rs13206526 | A | C | 6 | SUPT3H | Female only | True | 0.1772 | 0.1759 | 0.1697 | 0.1698 | 34577 | 25698 | 193785 | 166603 | -0.04546 | -0.009991 | -0.03266 | -0.01957 | 0.009097 | 0.01067 |
| Height | rs35214627 | rs3799972 | T | C | 6 | SUPT3H | Female only | True | 0.176 | 0.175 | 0.1693 | 0.1695 | 34577 | 25698 | 193785 | 166603 | -0.04662 | -0.01099 | -0.033 | -0.01962 | 0.009118 | 0.01069 |
| Height | rs35214627 | rs3799974 | C | T | 6 | SUPT3H | Female only | True | 0.1757 | 0.1748 | 0.1692 | 0.1694 | 34577 | 25698 | 193785 | 166603 | -0.04612 | -0.01045 | -0.03307 | -0.01964 | 0.009125 | 0.0107 |
| Height | rs35214627 | rs34243039 | A | C | 6 | SUPT3H | Female only | True | 0.1758 | 0.1749 | 0.1692 | 0.1694 | 34577 | 25698 | 193785 | 166603 | -0.04627 | -0.01067 | -0.03303 | -0.01958 | 0.009123 | 0.0107 |
| Height | rs35214627 | rs35856781 | A | G | 6 | SUPT3H | Female only | True | 0.1758 | 0.1749 | 0.1692 | 0.1694 | 34577 | 25698 | 193785 | 166603 | -0.04628 | -0.01067 | -0.03306 | -0.01959 | 0.009123 | 0.0107 |
| Height | rs35214627 | rs12663530 | C | T | 6 | SUPT3H | Female only | True | 0.1235 | 0.1244 | 0.1197 | 0.1197 | 34577 | 25698 | 193785 | 166603 | -0.05658 | -0.01313 | -0.01919 | -0.01248 | 0.01054 | 0.0123 |
| Height | rs35214627 | 6:44852734 | A | G | 6 | SUPT3H | Female only | True | 0.1245 | 0.125 | | | 34577 | 25698 | | | -0.05438 | -0.0115 | | | 0.01051 | 0.01227 |
| Height | rs35214627 | rs67911131 | T | C | 6 | SUPT3H | Female only | True | 0.1235 | 0.1243 | 0.1197 | 0.1197 | 34577 | 25698 | 193785 | 166603 | -0.05633 | -0.01281 | -0.01921 | -0.01249 | 0.01054 | 0.0123 |
| Height | rs35214627 | rs12529522 | T | C | 6 | SUPT3H | Female only | True | 0.1228 | 0.1235 | 0.1191 | 0.1191 | 34577 | 25698 | 193785 | 166603 | -0.05756 | -0.01303 | -0.01891 | -0.01278 | 0.01057 | 0.01233 |
| Height | rs524492 | rs12530016 | A | G | 6 | SUPT3H | Female only | True | 0.2109 | 0.2097 | 0.2039 | 0.2035 | 34577 | 25698 | 193785 | 166603 | -0.04124 | -0.01667 | -0.03465 | -0.02111 | 0.008526 | 0.009997 |
| Height | rs1285027 | rs116368192 | C | T | 6 | SUPT3H | Female only | True | 0.1228 | 0.1234 | 0.119 | 0.119 | 34577 | 25698 | 193785 | 166603 | -0.0572 | -0.01297 | -0.01894 | -0.0128 | 0.01057 | 0.01234 |
| Height | rs1285027 | rs6904584 | T | C | 6 | SUPT3H | Female only | True | 0.1233 | 0.1241 | 0.1196 | 0.1196 | 34577 | 25698 | 193785 | 166603 | -0.05588 | -0.0127 | -0.0192 | -0.01274 | 0.01055 | 0.01231 |
| Height | rs1285027 | rs6919161 | T | C | 6 | SUPT3H | Female only | True | 0.123 | 0.1238 | 0.1194 | 0.1194 | 34577 | 25698 | 193785 | 166603 | -0.05602 | -0.01298 | -0.01923 | -0.01251 | 0.01056 | 0.01233 |

| trait | lead_snp | ID | ALLELE1 | ALLELE0 | CHR | Gene_Symbol | independent_loci_in_sex_P1e2 | in_embl_any_or_gene | A1FREQ_VUMC_female | A1FREQ_VUMC_male | A1FREQ_UKB_female | A1FREQ_UKB_male | N_VUMC_female | N_VUMC_male | N_UKB_female | N_UKB_male | BETA_VUMC_female | BETA_VUMC_male | BETA_UKB_female | BETA_UKB_male | SE_VUMC_female | SE_VUMC_male |
|---|---|---|---|---|---|---|---|---|---|---|---|---|---|---|---|---|---|---|---|---|---|---|
| Height | rs1285027 | rs35704584 | T | C | 6 | SUPT3H | Female only | True | 0.1229 | 0.1238 | 0.1193 | 0.1193 | 34577 | 25698 | 193785 | 166603 | -0.05618 | -0.01321 | -0.01928 | -0.01242 | 0.01056 | 0.01232 |
| trait | lead_snp | ID | ALLELE1 | ALLELE0 | CHR | Gene_Symbol | independent_loci_in_sex_P1e2 | in_embl_any_or_gene | A1FREQ_VUMC_female | A1FREQ_VUMC_male | A1FREQ_UKB_female | A1FREQ_UKB_male | N_VUMC_female | N_VUMC_male | N_UKB_female | N_UKB_male | BETA_VUMC_female | BETA_VUMC_male | BETA_UKB_female | BETA_UKB_male | SE_VUMC_female | SE_VUMC_male |
| Height | rs1285027 | rs3857589 | A | G | 6 | SUPT3H | Female only | True | 0.1191 | 0.1207 | 0.1176 | 0.1176 | 34577 | 25698 | 193785 | 166603 | -0.05602 | -0.01413 | -0.01894 | -0.01282 | 0.01075 | 0.01251 |
| Height | rs1285027 | rs1329715 | T | C | 6 | SUPT3H | Female only | True | 0.1223 | 0.123 | 0.1187 | 0.1187 | 34577 | 25698 | 193785 | 166603 | -0.05703 | -0.01342 | -0.01912 | -0.01249 | 0.01059 | 0.01236 |
| Height | rs1285027 | rs35103634 | A | G | 6 | SUPT3H | Female only | True | 0.1224 | 0.1231 | 0.1187 | 0.1187 | 34577 | 25698 | 193785 | 166603 | -0.05727 | -0.01344 | -0.01912 | -0.01249 | 0.01059 | 0.01236 |
| Height | rs524492 | rs12525773 | G | T | 6 | SUPT3H | Female only | True | 0.2116 | 0.2108 | 0.2047 | 0.2042 | 34577 | 25698 | 193785 | 166603 | -0.03902 | -0.016 | -0.03445 | -0.02037 | 0.008519 | 0.009979 |
| Height | rs1285027 | rs4475320 | C | T | 6 | SUPT3H | Female only | True | 0.1228 | 0.1237 | 0.1193 | 0.1193 | 34577 | 25698 | 193785 | 166603 | -0.05569 | -0.01263 | -0.01941 | -0.01237 | 0.01057 | 0.01233 |
| Height | rs1285027 | rs3799987 | A | G | 6 | SUPT3H | Female only | True | 0.1227 | 0.1236 | 0.1193 | 0.1193 | 34577 | 25698 | 193785 | 166603 | -0.0557 | -0.01298 | -0.01938 | -0.01236 | 0.01057 | 0.01233 |
| Height | rs1285027 | rs12663798 | T | C | 6 | SUPT3H | Female only | True | 0.1228 | 0.1236 | 0.1193 | 0.1193 | 34577 | 25698 | 193785 | 166603 | -0.05572 | -0.01327 | -0.01935 | -0.01233 | 0.01056 | 0.01233 |
| Height | rs1285027 | rs12526755 | G | T | 6 | SUPT3H | Female only | True | 0.1227 | 0.1236 | 0.1192 | 0.1193 | 34577 | 25698 | 193785 | 166603 | -0.05562 | -0.01274 | -0.01922 | -0.01243 | 0.01056 | 0.01233 |
| Height | rs1285027 | rs35908738 | T | C | 6 | SUPT3H | Female only | True | 0.1227 | 0.1236 | 0.1192 | 0.1193 | 34577 | 25698 | 193785 | 166603 | -0.05562 | -0.01273 | -0.0193 | -0.01234 | 0.01056 | 0.01233 |
| Height | rs1285027 | rs4714829 | A | G | 6 | SUPT3H | Female only | True | 0.1228 | 0.1236 | 0.1193 | 0.1193 | 34577 | 25698 | 193785 | 166603 | -0.05564 | -0.0127 | -0.0192 | -0.01243 | 0.01056 | 0.01233 |
| Height | rs1285027 | rs12661902 | T | G | 6 | SUPT3H | Female only | True | 0.1236 | 0.1244 | 0.1198 | 0.1198 | 34577 | 25698 | 193785 | 166603 | -0.05526 | -0.01307 | -0.01944 | -0.01223 | 0.01055 | 0.01231 |
| Height | rs1285027 | rs34286542 | T | C | 6 | SUPT3H | Female only | True | 0.1228 | 0.1236 | 0.1193 | 0.1193 | 34577 | 25698 | 193785 | 166603 | -0.05582 | -0.01308 | -0.01921 | -0.01237 | 0.01056 | 0.01233 |
| Height | rs524492 | rs9296450 | C | T | 6 | SUPT3H | Female only | True | 0.2118 | 0.211 | 0.2049 | 0.2044 | 34577 | 25698 | 193785 | 166603 | -0.03869 | -0.01595 | -0.03442 | -0.02049 | 0.008511 | 0.00997 |
| Height | rs1285027 | rs35687006 | C | T | 6 | SUPT3H | Female only | True | 0.1227 | 0.1236 | 0.1193 | 0.1193 | 34577 | 25698 | 193785 | 166603 | -0.05581 | -0.01333 | -0.01931 | -0.01232 | 0.01056 | 0.01233 |
| Height | rs524492 | rs9885796 | C | A | 6 | SUPT3H | Female only | True | 0.2118 | 0.211 | 0.205 | 0.2045 | 34577 | 25698 | 193785 | 166603 | -0.03849 | -0.01613 | -0.03438 | -0.0206 | 0.008513 | 0.009972 |
| Height | rs1285027 | rs1023091 | T | C | 6 | SUPT3H | Female only | True | 0.1223 | 0.1229 | 0.1188 | 0.1189 | 34577 | 25698 | 193785 | 166603 | -0.05665 | -0.01391 | -0.01925 | -0.01299 | 0.01057 | 0.01236 |
| Height | rs1285027 | rs4714834 | T | C | 6 | SUPT3H | Female only | True | 0.1223 | 0.1229 | 0.1188 | 0.1189 | 34577 | 25698 | 193785 | 166603 | -0.05688 | -0.01392 | -0.01924 | -0.01304 | 0.01057 | 0.01236 |
| Height | rs1285027 | rs4711809 | T | C | 6 | SUPT3H | Female only | True | 0.1223 | 0.1229 | 0.1188 | 0.1189 | 34577 | 25698 | 193785 | 166603 | -0.05686 | -0.01394 | -0.01925 | -0.01298 | 0.01058 | 0.01236 |
| Height | rs1285027 | rs12528232 | A | G | 6 | SUPT3H | Female only | True | 0.1223 | 0.1229 | 0.1188 | 0.1189 | 34577 | 25698 | 193785 | 166603 | -0.05691 | -0.01393 | -0.01924 | -0.01299 | 0.01058 | 0.01236 |
| Height | rs1285027 | rs1028944 | T | C | 6 | SUPT3H | Female only | True | 0.1207 | 0.1214 | 0.1166 | 0.1169 | 34577 | 25698 | 193785 | 166603 | -0.05619 | -0.01265 | -0.01911 | -0.01309 | 0.01065 | 0.01244 |
| Height | rs1285027 | rs13209827 | C | T | 6 | SUPT3H | Female only | True | 0.121 | 0.1217 | 0.1168 | 0.117 | 34577 | 25698 | 193785 | 166603 | -0.05688 | -0.01263 | -0.019 | -0.01287 | 0.01064 | 0.01242 |
| Height | rs1285027 | rs13205113 | T | C | 6 | SUPT3H | Female only | True | 0.1211 | 0.1218 | 0.1168 | 0.1171 | 34577 | 25698 | 193785 | 166603 | -0.05728 | -0.01284 | -0.01903 | -0.01292 | 0.01063 | 0.01242 |
| Height | rs1285027 | rs12528762 | A | G | 6 | SUPT3H | Female only | True | 0.1211 | 0.1218 | 0.1168 | 0.1171 | 34577 | 25698 | 193785 | 166603 | -0.05742 | -0.01317 | -0.01896 | -0.01294 | 0.01063 | 0.01243 |
| Height | rs1285027 | rs12527668 | G | A | 6 | SUPT3H | Female only | True | 0.1205 | 0.1211 | 0.1163 | 0.1165 | 34577 | 25698 | 193785 | 166603 | -0.05866 | -0.01301 | -0.01872 | -0.01316 | 0.01066 | 0.01245 |
| Height | rs1285027 | rs34152708 | A | G | 6 | SUPT3H | Female only | True | 0.1211 | 0.1216 | 0.1166 | 0.1167 | 34577 | 25698 | 193785 | 166603 | -0.0576 | -0.01202 | -0.01847 | -0.01324 | 0.01064 | 0.01243 |
| Height | rs1285027 | rs13198471 | G | A | 6 | SUPT3H | Female only | True | 0.1211 | 0.1216 | 0.1166 | 0.1167 | 34577 | 25698 | 193785 | 166603 | -0.05762 | -0.01204 | -0.01843 | -0.01325 | 0.01064 | 0.01243 |

| trait | lead_snp | ID | ALLELE1 | ALLELE0 | CHR | Gene_Symbol | independent_loci_in_sex_P1e2 | in_embl_any_or_gene | A1FREQ_VUMC_female | A1FREQ_VUMC_male | A1FREQ_UKB_female | A1FREQ_UKB_male | N_VUMC_female | N_VUMC_male | N_UKB_female | N_UKB_male | BETA_VUMC_female | BETA_VUMC_male | BETA_UKB_female | BETA_UKB_male | SE_VUMC_female | SE_VUMC_male |
|---|---|---|---|---|---|---|---|---|---|---|---|---|---|---|---|---|---|---|---|---|---|---|
| Height | rs1285027 | rs7745506 | G | T | 6 | SUPT3H | Female only | True | 0.1211 | 0.1216 | 0.1166 | 0.1167 | 34577 | 25698 | 193785 | 166603 | -0.05782 | -0.01172 | -0.01843 | -0.01326 | 0.01063 | 0.01243 |
| Height | rs1285027 | rs4714838 | C | T | 6 | SUPT3H | Female only | True | 0.1207 | 0.1212 | 0.1165 | 0.1167 | 34577 | 25698 | 193785 | 166603 | -0.05783 | -0.01234 | -0.01851 | -0.01329 | 0.01065 | 0.01245 |
| Height | rs1285027 | rs34251367 | T | C | 6 | SUPT3H | Female only | True | 0.1207 | 0.1212 | 0.1165 | 0.1167 | 34577 | 25698 | 193785 | 166603 | -0.05727 | -0.01254 | -0.01851 | -0.01325 | 0.01065 | 0.01245 |
| Height | rs1285027 | rs35134594 | A | G | 6 | SUPT3H | Female only | True | 0.1208 | 0.1214 | 0.1165 | 0.1166 | 34577 | 25698 | 193785 | 166603 | -0.05772 | -0.01181 | -0.01856 | -0.01328 | 0.01064 | 0.01244 |
| Height | rs1285027 | rs12526649 | T | G | 6 | SUPT3H | Female only | True | 0.1209 | 0.1214 | 0.1164 | 0.1165 | 34577 | 25698 | 193785 | 166603 | -0.05756 | -0.01189 | -0.01857 | -0.0132 | 0.01064 | 0.01244 |
| Height | rs1285027 | rs12529990 | C | A | 6 | SUPT3H | Female only | True | 0.1207 | 0.1211 | 0.1165 | 0.1166 | 34577 | 25698 | 193785 | 166603 | -0.05745 | -0.01224 | -0.0186 | -0.01324 | 0.01065 | 0.01245 |
| Height | rs1285027 | rs16873131 | A | G | 6 | SUPT3H | Female only | True | 0.1209 | 0.1214 | 0.1165 | 0.1166 | 34577 | 25698 | 193785 | 166603 | -0.0575 | -0.01178 | -0.01855 | -0.01321 | 0.01064 | 0.01244 |
| Height | rs1285027 | rs1284986 | C | T | 6 | SUPT3H | Female only | True | 0.1258 | 0.1261 | 0.1165 | 0.1166 | 34577 | 25698 | 193785 | 166603 | -0.05749 | -0.01158 | -0.01858 | -0.01319 | 0.01066 | 0.01246 |
| Height | rs1285027 | rs1284989 | G | A | 6 | SUPT3H | Female only | True | 0.1258 | 0.1261 | 0.1165 | 0.1166 | 34577 | 25698 | 193785 | 166603 | -0.05749 | -0.01157 | -0.0185 | -0.01316 | 0.01066 | 0.01246 |
| Height | rs1285027 | rs1797156 | C | T | 6 | SUPT3H | Female only | True | 0.1258 | 0.1261 | 0.1164 | 0.1164 | 34577 | 25698 | 193785 | 166603 | -0.05739 | -0.01155 | -0.0183 | -0.01302 | 0.01066 | 0.01247 |
| Height | rs1285027 | rs2396381 | A | G | 6 | SUPT3H | Female only | True | 0.1256 | 0.126 | 0.1165 | 0.1166 | 34577 | 25698 | 193785 | 166603 | -0.05762 | -0.01177 | -0.01848 | -0.01312 | 0.01066 | 0.01247 |
| Height | rs1285027 | rs1284979 | A | C | 6 | SUPT3H | Female only | True | 0.1255 | 0.1259 | 0.1164 | 0.1165 | 34577 | 25698 | 193785 | 166603 | -0.05757 | -0.01163 | -0.01851 | -0.0131 | 0.01066 | 0.01246 |
| Height | rs1285027 | rs80164710 | C | T | 6 | SUPT3H | Female only | True | 0.1206 | 0.121 | 0.1164 | 0.1165 | 34577 | 25698 | 193785 | 166603 | -0.05728 | -0.01168 | -0.01857 | -0.01313 | 0.01065 | 0.01246 |
| Height | rs1285027 | rs12530207 | C | T | 6 | SUPT3H | Female only | True | 0.1206 | 0.121 | 0.1164 | 0.1164 | 34577 | 25698 | 193785 | 166603 | -0.05728 | -0.01169 | -0.01858 | -0.01312 | 0.01065 | 0.01246 |
| Height | rs1285027 | rs1284999 | C | A | 6 | SUPT3H | Female only | True | 0.1216 | 0.122 | 0.1165 | 0.1165 | 34577 | 25698 | 193785 | 166603 | -0.05686 | -0.009534 | -0.01838 | -0.01311 | 0.0106 | 0.01239 |
| Height | rs1285027 | rs1285000 | T | C | 6 | SUPT3H | Female only | True | 0.1254 | 0.1259 | 0.1165 | 0.1165 | 34577 | 25698 | 193785 | 166603 | -0.05741 | -0.01035 | -0.01846 | -0.01308 | 0.01066 | 0.01246 |
| Height | rs1285027 | rs9463069 | A | G | 6 | SUPT3H | Female only | True | 0.1254 | 0.1259 | 0.1165 | 0.1165 | 34577 | 25698 | 193785 | 166603 | -0.05741 | -0.01035 | -0.01847 | -0.01308 | 0.01066 | 0.01246 |
| Height | rs1285027 | rs9463070 | G | A | 6 | SUPT3H | Female only | True | 0.1254 | 0.1259 | 0.1165 | 0.1165 | 34577 | 25698 | 193785 | 166603 | -0.05741 | -0.01035 | -0.01847 | -0.01308 | 0.01066 | 0.01246 |
| Height | rs1285027 | rs1797148 | G | T | 6 | SUPT3H | Female only | True | 0.1246 | 0.1251 | 0.1157 | 0.1157 | 34577 | 25698 | 193785 | 166603 | -0.05735 | -0.0107 | -0.01852 | -0.01336 | 0.01071 | 0.01252 |
| Height | rs1285027 | rs1285005 | G | A | 6 | SUPT3H | Female only | True | 0.1256 | 0.126 | 0.1165 | 0.1165 | 34577 | 25698 | 193785 | 166603 | -0.05722 | -0.01003 | -0.01844 | -0.01309 | 0.01065 | 0.01245 |
| Height | rs1285027 | rs1285006 | C | T | 6 | SUPT3H | Female only | True | 0.1256 | 0.126 | 0.1165 | 0.1165 | 34577 | 25698 | 193785 | 166603 | -0.05722 | -0.01003 | -0.01845 | -0.01309 | 0.01065 | 0.01245 |
| Height | rs1285027 | rs1285007 | A | C | 6 | SUPT3H | Female only | True | 0.1411 | 0.1403 | 0.1369 | 0.1372 | 34577 | 25698 | 193785 | 166603 | -0.06064 | -0.01131 | -0.01805 | -0.01169 | 0.009951 | 0.01166 |
| Height | rs1285027 | rs1285008 | A | G | 6 | SUPT3H | Female only | True | 0.1253 | 0.1259 | 0.1165 | 0.1165 | 34577 | 25698 | 193785 | 166603 | -0.05738 | -0.01031 | -0.01851 | -0.01309 | 0.01065 | 0.01246 |
| Height | rs1285027 | rs9472442 | C | T | 6 | SUPT3H | Female only | True | 0.1255 | 0.126 | 0.1165 | 0.1165 | 34577 | 25698 | 193785 | 166603 | -0.05724 | -0.009962 | -0.01846 | -0.01309 | 0.01065 | 0.01245 |
| Height | rs1285027 | rs1285010 | A | C | 6 | SUPT3H | Female only | True | 0.1255 | 0.126 | 0.1165 | 0.1165 | 34577 | 25698 | 193785 | 166603 | -0.05724 | -0.009962 | -0.01846 | -0.01309 | 0.01065 | 0.01245 |
| Height | rs1285027 | rs1285011 | A | G | 6 | SUPT3H | Female only | True | 0.1214 | 0.1219 | 0.1165 | 0.1165 | 34577 | 25698 | 193785 | 166603 | -0.05696 | -0.009978 | -0.01846 | -0.01309 | 0.0106 | 0.0124 |
| Height | rs1285027 | rs6917594 | T | G | 6 | SUPT3H | Female only | True | 0.1253 | 0.1257 | 0.1164 | 0.1165 | 34577 | 25698 | 193785 | 166603 | -0.05745 | -0.01141 | -0.01858 | -0.01309 | 0.01066 | 0.01247 |
| Height | rs1285027 | rs1285020 | T | C | 6 | SUPT3H | Female only | True | 0.1253 | 0.1257 | 0.1164 | 0.1165 | 34577 | 25698 | 193785 | 166603 | -0.05736 | -0.0115 | -0.01855 | -0.01307 | 0.01065 | 0.01247 |
| Height | rs1285027 | rs1285021 | A | G | 6 | SUPT3H | Female only | True | 0.1253 | 0.1257 | 0.1164 | 0.1165 | 34577 | 25698 | 193785 | 166603 | -0.05736 | -0.0115 | -0.01855 | -0.01307 | 0.01065 | 0.01247 |

| trait | lead_snp | ID | ALLELE1 | ALLELE0 | CHR | Gene_Symbol | independent_loci_in_sex_P1e2 | in_embl_any_or_gene | A1FREQ_VUMC_female | A1FREQ_VUMC_male | A1FREQ_UKB_female | A1FREQ_UKB_male | N_VUMC_female | N_VUMC_male | N_UKB_female | N_UKB_male | BETA_VUMC_female | BETA_VUMC_male | BETA_UKB_female | BETA_UKB_male | SE_VUMC_female | SE_VUMC_male |
|---|---|---|---|---|---|---|---|---|---|---|---|---|---|---|---|---|---|---|---|---|---|---|
| Height | rs1285027 | rs73449282 | A | G | 6 | SUPT3H | Female only | True | 0.1207 | 0.121 | 0.1164 | 0.1164 | 34577 | 25698 | 193785 | 166603 | -0.05676 | -0.01173 | -0.01851 | -0.01313 | 0.01064 | 0.01245 |
| Height | rs1285027 | rs1324535 | A | G | 6 | SUPT3H | Female only | True | 0.1253 | 0.1257 | 0.1164 | 0.1165 | 34577 | 25698 | 193785 | 166603 | -0.05737 | -0.01153 | -0.01852 | -0.0131 | 0.01065 | 0.01247 |
| Height | rs1285027 | rs1285025 | G | A | 6 | SUPT3H | Female only | True | 0.1256 | 0.1259 | 0.1164 | 0.1165 | 34577 | 25698 | 193785 | 166603 | -0.05723 | -0.01077 | -0.01844 | -0.01317 | 0.01064 | 0.01246 |
| Height | rs1285027 | rs1285027 | A | G | 6 | SUPT3H | Female only | True | 0.1452 | 0.1443 | 0.1369 | 0.1372 | 34577 | 25698 | 193785 | 166603 | -0.06104 | -0.01247 | -0.01814 | -0.01189 | 0.009995 | 0.01172 |
| Height | rs1285027 | rs1285028 | A | C | 6 | SUPT3H | Female only | True | 0.1255 | 0.1258 | 0.1165 | 0.1166 | 34577 | 25698 | 193785 | 166603 | -0.05721 | -0.01126 | -0.01851 | -0.01328 | 0.01065 | 0.01246 |

| trait | lead_snp | ID | ALLELE1 | ALLELE0 | CHR | Gene_Symbol | independent_loci_in_sex_P1e2 | in_embl_any_or_gene | A1FREQ_VUMC_female | A1FREQ_VUMC_male | A1FREQ_UKB_female | A1FREQ_UKB_male | N_VUMC_female | N_VUMC_male | N_UKB_female | N_UKB_male | BETA_VUMC_female | BETA_VUMC_male | BETA_UKB_female | BETA_UKB_male | SE_VUMC_female | SE_VUMC_male |
|---|---|---|---|---|---|---|---|---|---|---|---|---|---|---|---|---|---|---|---|---|---|---|
| Height | rs1285027 | rs1285029 | C | T | 6 | SUPT3H | Female only | True | 0.1214 | 0.1217 | 0.1164 | 0.1165 | 34577 | 25698 | 193785 | 166603 | -0.05695 | -0.01125 | -0.01844 | -0.01319 | 0.0106 | 0.01241 |
| Height | rs1285027 | rs1285030 | A | G | 6 | SUPT3H | Female only | True | 0.1253 | 0.1257 | 0.1164 | 0.1165 | 34577 | 25698 | 193785 | 166603 | -0.05733 | -0.01157 | -0.01852 | -0.01312 | 0.01065 | 0.01247 |
| Height | rs1285027 | rs12662272 | A | C | 6 | SUPT3H | Female only | True | 0.1206 | 0.1209 | 0.1164 | 0.1164 | 34577 | 25698 | 193785 | 166603 | -0.05716 | -0.01196 | -0.01858 | -0.0131 | 0.01065 | 0.01246 |
| Height | rs1285027 | rs1797159 | A | G | 6 | SUPT3H | Female only | True | 0.1448 | 0.1438 | 0.1362 | 0.1366 | 34577 | 25698 | 193785 | 166603 | -0.05988 | -0.0125 | -0.01826 | -0.01166 | 0.01 | 0.01173 |
| Height | rs1285027 | rs1324530 | G | A | 6 | SUPT3H | Female only | True | 0.1407 | 0.1397 | 0.1363 | 0.1366 | 34577 | 25698 | 193785 | 166603 | -0.05958 | -0.0125 | -0.01833 | -0.01174 | 0.009959 | 0.01168 |
| Height | rs1285027 | rs1748236 | G | A | 6 | SUPT3H | Female only | True | 0.1248 | 0.1252 | 0.1158 | 0.1159 | 34577 | 25698 | 193785 | 166603 | -0.05647 | -0.01139 | -0.01875 | -0.01293 | 0.01067 | 0.01249 |
| Height | rs1285027 | rs1748238 | G | A | 6 | SUPT3H | Female only | True | 0.1448 | 0.1438 | 0.1363 | 0.1366 | 34577 | 25698 | 193785 | 166603 | -0.0599 | -0.01224 | -0.01829 | -0.0117 | 0.01001 | 0.01173 |
| Height | rs1285027 | rs7740542 | C | T | 6 | SUPT3H | Female only | True | 0.1449 | 0.1439 | 0.1364 | 0.1367 | 34577 | 25698 | 193785 | 166603 | -0.05955 | -0.0135 | -0.01801 | -0.01171 | 0.01 | 0.01172 |
| Height | rs1285027 | rs6936475 | C | T | 6 | SUPT3H | Female only | True | 0.1205 | 0.1209 | 0.1159 | 0.116 | 34577 | 25698 | 193785 | 166603 | -0.05577 | -0.01279 | -0.01846 | -0.01288 | 0.01065 | 0.01247 |
| Height | rs1285027 | rs1748240 | G | A | 6 | SUPT3H | Female only | True | 0.1246 | 0.1251 | 0.1159 | 0.116 | 34577 | 25698 | 193785 | 166603 | -0.05627 | -0.01211 | -0.0189 | -0.01307 | 0.01067 | 0.01249 |
| Height | rs1285027 | rs4714842 | G | A | 6 | SUPT3H | Female only | True | 0.1201 | 0.1204 | 0.1159 | 0.116 | 34577 | 25698 | 193785 | 166603 | -0.05583 | -0.01348 | -0.01856 | -0.01288 | 0.01067 | 0.01248 |
| Height | rs1285027 | rs1284962 | A | G | 6 | SUPT3H | Female only | True | 0.1246 | 0.1251 | 0.1159 | 0.116 | 34577 | 25698 | 193785 | 166603 | -0.05592 | -0.01323 | -0.01858 | -0.01285 | 0.01067 | 0.01249 |
| Height | rs1285027 | rs1284964 | C | T | 6 | SUPT3H | Female only | True | 0.1208 | 0.1212 | 0.1159 | 0.116 | 34577 | 25698 | 193785 | 166603 | -0.05556 | -0.0128 | -0.01858 | -0.01295 | 0.01062 | 0.01243 |
| Height | rs1285027 | rs16873257 | T | C | 6 | SUPT3H | Female only | True | 0.1201 | 0.1204 | 0.1159 | 0.1159 | 34577 | 25698 | 193785 | 166603 | -0.05592 | -0.0133 | -0.01867 | -0.01276 | 0.01067 | 0.01248 |
| Height | rs1285027 | rs2815263 | C | T | 6 | SUPT3H | Female only | True | 0.1249 | 0.1252 | 0.1159 | 0.116 | 34577 | 25698 | 193785 | 166603 | -0.05552 | -0.01268 | -0.01855 | -0.0128 | 0.01066 | 0.01248 |
| Height | rs1285027 | rs9369549 | T | G | 6 | SUPT3H | Female only | True | 0.125 | 0.1253 | 0.1159 | 0.116 | 34577 | 25698 | 193785 | 166603 | -0.05588 | -0.01246 | -0.01858 | -0.01267 | 0.01066 | 0.01248 |
| Height | rs1285027 | rs9395081 | G | T | 6 | SUPT3H | Female only | True | 0.1449 | 0.1439 | 0.1364 | 0.1367 | 34577 | 25698 | 193785 | 166603 | -0.0596 | -0.01348 | -0.0182 | -0.01137 | 0.01 | 0.01173 |
| Height | rs1285027 | rs6917352 | T | C | 6 | SUPT3H | Female only | True | 0.1245 | 0.1249 | 0.1159 | 0.116 | 34577 | 25698 | 193785 | 166603 | -0.05547 | -0.01367 | -0.01855 | -0.01262 | 0.01069 | 0.0125 |
| Height | rs1285027 | rs6458431 | T | C | 6 | SUPT3H | Female only | True | 0.1248 | 0.1252 | 0.1159 | 0.116 | 34577 | 25698 | 193785 | 166603 | -0.05596 | -0.01299 | -0.0186 | -0.0126 | 0.01067 | 0.01249 |
| Height | rs1285027 | rs2057553 | G | A | 6 | SUPT3H | Female only | True | 0.1444 | 0.1434 | 0.1361 | 0.1365 | 34577 | 25698 | 193785 | 166603 | -0.0593 | -0.01372 | -0.01802 | -0.0113 | 0.01003 | 0.01176 |
| Height | rs1285027 | rs1853652 | A | G | 6 | SUPT3H | Female only | True | 0.1246 | 0.1249 | 0.1158 | 0.1159 | 34577 | 25698 | 193785 | 166603 | -0.05568 | -0.01325 | -0.01855 | -0.01261 | 0.01069 | 0.01251 |
| Height | rs1285027 | rs6458432 | T | C | 6 | SUPT3H | Female only | True | 0.1245 | 0.1249 | 0.1158 | 0.1159 | 34577 | 25698 | 193785 | 166603 | -0.05595 | -0.01294 | -0.0186 | -0.01242 | 0.0107 | 0.01251 |

| trait | lead_snp | ID | ALLELE1 | ALLELE0 | CHR | Gene_Symbol | independent_loci_in_sex_P1e2 | in_embl_any_or_gene | A1FREQ_VUMC_female | A1FREQ_VUMC_male | A1FREQ_UKB_female | A1FREQ_UKB_male | N_VUMC_female | N_VUMC_male | N_UKB_female | N_UKB_male | BETA_VUMC_female | BETA_VUMC_male | BETA_UKB_female | BETA_UKB_male | SE_VUMC_female | SE_VUMC_male |
|---|---|---|---|---|---|---|---|---|---|---|---|---|---|---|---|---|---|---|---|---|---|---|
| Height | rs1285027 | rs12527743 | C | T | 6 | SUPT3H | Female only | True | 0.12 | 0.1204 | 0.1159 | 0.116 | 34577 | 25698 | 193785 | 166603 | -0.05569 | -0.0134 | -0.01863 | -0.01256 | 0.01068 | 0.01249 |
| Height | rs1285027 | rs6458433 | C | T | 6 | SUPT3H | Female only | True | 0.1254 | 0.1255 | 0.116 | 0.1161 | 34577 | 25698 | 193785 | 166603 | -0.05503 | -0.01306 | -0.01861 | -0.01269 | 0.01066 | 0.01248 |
| Height | rs1285027 | rs9296452 | T | C | 6 | SUPT3H | Female only | True | 0.1448 | 0.1439 | 0.1364 | 0.1367 | 34577 | 25698 | 193785 | 166603 | -0.05897 | -0.01408 | -0.01822 | -0.01111 | 0.01001 | 0.01174 |
| Height | rs1285027 | rs2057552 | C | T | 6 | SUPT3H | Female only | True | 0.1252 | 0.1256 | 0.1161 | 0.1162 | 34577 | 25698 | 193785 | 166603 | -0.05573 | -0.01306 | -0.01846 | -0.01264 | 0.01067 | 0.01249 |
| Height | rs1285027 | rs6458434 | T | C | 6 | SUPT3H | Female only | True | 0.1452 | 0.1442 | 0.1364 | 0.1367 | 34577 | 25698 | 193785 | 166603 | -0.05901 | -0.01392 | -0.01817 | -0.01131 | 0.01001 | 0.01174 |
| Height | rs1285027 | rs6901161 | A | G | 6 | SUPT3H | Female only | True | 0.1254 | 0.1257 | 0.116 | 0.1161 | 34577 | 25698 | 193785 | 166603 | -0.05555 | -0.01287 | -0.01857 | -0.01244 | 0.01067 | 0.01249 |
| Height | rs1285027 | rs6458435 | A | G | 6 | SUPT3H | Female only | True | 0.1252 | 0.1256 | 0.1159 | 0.116 | 34577 | 25698 | 193785 | 166603 | -0.05578 | -0.01298 | -0.01861 | -0.01249 | 0.01068 | 0.01249 |
| Height | rs1285027 | rs2763134 | C | T | 6 | SUPT3H | Female only | True | 0.1253 | 0.1256 | 0.116 | 0.116 | 34577 | 25698 | 193785 | 166603 | -0.05542 | -0.01297 | -0.01854 | -0.01254 | 0.01067 | 0.01249 |
| Height | rs1285027 | rs34167737 | T | C | 6 | SUPT3H | Female only | True | 0.1404 | 0.1394 | 0.1364 | 0.1367 | 34577 | 25698 | 193785 | 166603 | -0.05939 | -0.01377 | -0.01815 | -0.01109 | 0.00999 | 0.01172 |
| Height | rs1285027 | rs75833179 | T | C | 6 | SUPT3H | Female only | True | 0.1404 | 0.1394 | 0.1364 | 0.1367 | 34577 | 25698 | 193785 | 166603 | -0.05939 | -0.01379 | -0.0181 | -0.01116 | 0.00999 | 0.01172 |
| Height | rs1285027 | rs4714846 | C | T | 6 | SUPT3H | Female only | True | 0.1404 | 0.1394 | 0.1364 | 0.1367 | 34577 | 25698 | 193785 | 166603 | -0.05939 | -0.01379 | -0.0181 | -0.01116 | 0.00999 | 0.01172 |
| Height | rs1285027 | rs56175185 | C | T | 6 | SUPT3H | Female only | True | 0.1204 | 0.1208 | 0.1159 | 0.116 | 34577 | 25698 | 193785 | 166603 | -0.05601 | -0.01305 | -0.01861 | -0.01249 | 0.01066 | 0.01248 |
| Height | rs1285027 | rs73735337 | C | T | 6 | SUPT3H | Female only | True | 0.1404 | 0.1394 | 0.1364 | 0.1367 | 34577 | 25698 | 193785 | 166603 | -0.05983 | -0.01367 | -0.0181 | -0.01114 | 0.00999 | 0.01172 |
| Height | rs1285027 | rs73735338 | C | T | 6 | SUPT3H | Female only | True | 0.1205 | 0.1208 | 0.1159 | 0.116 | 34577 | 25698 | 193785 | 166603 | -0.05601 | -0.01289 | -0.01859 | -0.01252 | 0.01066 | 0.01248 |
| Height | rs1285027 | rs12526807 | T | C | 6 | SUPT3H | Female only | True | 0.1204 | 0.1208 | 0.1159 | 0.116 | 34577 | 25698 | 193785 | 166603 | -0.05614 | -0.01289 | -0.01861 | -0.01249 | 0.01066 | 0.01248 |
| Height | rs1285027 | rs4521586 | C | T | 6 | SUPT3H | Female only | True | 0.1204 | 0.1208 | 0.116 | 0.116 | 34577 | 25698 | 193785 | 166603 | -0.05593 | -0.01307 | -0.01863 | -0.01263 | 0.01066 | 0.01247 |
| Height | rs1285027 | rs4521587 | C | T | 6 | SUPT3H | Female only | True | 0.1204 | 0.1208 | 0.116 | 0.116 | 34577 | 25698 | 193785 | 166603 | -0.05593 | -0.01304 | -0.01862 | -0.01262 | 0.01066 | 0.01247 |
| Height | rs1285027 | rs4339452 | C | T | 6 | SUPT3H | Female only | True | 0.1403 | 0.1393 | 0.1364 | 0.1367 | 34577 | 25698 | 193785 | 166603 | -0.05942 | -0.01372 | -0.01809 | -0.01117 | 0.009993 | 0.01172 |
| Height | rs1285027 | rs55959525 | C | T | 6 | SUPT3H | Female only | True | 0.1403 | 0.1393 | 0.1364 | 0.1367 | 34577 | 25698 | 193785 | 166603 | -0.05942 | -0.01379 | -0.01806 | -0.0112 | 0.009993 | 0.01172 |
| Height | rs1285027 | rs75674202 | A | C | 6 | SUPT3H | Female only | True | 0.1204 | 0.1207 | 0.1159 | 0.116 | 34577 | 25698 | 193785 | 166603 | -0.05602 | -0.01321 | -0.0186 | -0.0125 | 0.01067 | 0.01248 |
| Height | rs1285027 | rs151275720 | G | T | 6 | SUPT3H | Female only | True | 0.1205 | 0.1208 | 0.116 | 0.1161 | 34577 | 25698 | 193785 | 166603 | -0.05572 | -0.01328 | -0.0186 | -0.01265 | 0.01066 | 0.01247 |
| Height | rs1285027 | rs6905959 | A | C | 6 | SUPT3H | Female only | True | 0.1204 | 0.1208 | 0.116 | 0.116 | 34577 | 25698 | 193785 | 166603 | -0.05569 | -0.0133 | -0.0186 | -0.01264 | 0.01066 | 0.01247 |
| Height | rs1285027 | rs142343743 | T | C | 6 | SUPT3H | Female only | True | 0.138 | 0.1368 | 0.1337 | 0.134 | 34577 | 25698 | 193785 | 166603 | -0.06032 | -0.01246 | -0.01818 | -0.01122 | 0.01013 | 0.01189 |
| Height | rs1285027 | rs12529907 | T | C | 6 | SUPT3H | Female only | True | 0.1204 | 0.1208 | 0.116 | 0.116 | 34577 | 25698 | 193785 | 166603 | -0.05568 | -0.0133 | -0.01861 | -0.01267 | 0.01066 | 0.01247 |
| Height | rs1285027 | rs77435067 | G | A | 6 | SUPT3H | Female only | True | 0.12 | 0.1204 | 0.1159 | 0.116 | 34577 | 25698 | 193785 | 166603 | -0.05591 | -0.01392 | -0.01873 | -0.01263 | 0.01068 | 0.01249 |
| Height | rs1285027 | rs4312985 | T | C | 6 | SUPT3H | Female only | True | 0.1403 | 0.1394 | 0.1364 | 0.1367 | 34577 | 25698 | 193785 | 166603 | -0.05925 | -0.01384 | -0.0181 | -0.01126 | 0.009993 | 0.01171 |
| Height | rs1285027 | rs75242452 | C | T | 6 | SUPT3H | Female only | True | 0.1204 | 0.1208 | 0.116 | 0.1161 | 34577 | 25698 | 193785 | 166603 | -0.05572 | -0.01315 | -0.01862 | -0.01258 | 0.01066 | 0.01247 |
| Height | rs1285027 | rs4341001 | T | C | 6 | SUPT3H | Female only | True | 0.1404 | 0.1394 | 0.1364 | 0.1368 | 34577 | 25698 | 193785 | 166603 | -0.05914 | -0.01369 | -0.01809 | -0.01124 | 0.009989 | 0.01171 |
| Height | rs1285027 | rs74851310 | C | T | 6 | SUPT3H | Female only | True | 0.1404 | 0.1394 | 0.1365 | 0.1368 | 34577 | 25698 | 193785 | 166603 | -0.05917 | -0.01371 | -0.01794 | -0.01135 | 0.009989 | 0.01171 |

| trait | lead_snp | ID | ALLELE1 | ALLELE0 | CHR | Gene_Symbol | independent_loci_in_sex_P1e2 | in_embl_any_or_gene | A1FREQ_VUMC_female | A1FREQ_VUMC_male | A1FREQ_UKB_female | A1FREQ_UKB_male | N_VUMC_female | N_VUMC_male | N_UKB_female | N_UKB_male | BETA_VUMC_female | BETA_VUMC_male | BETA_UKB_female | BETA_UKB_male | SE_VUMC_female | SE_VUMC_male |
|---|---|---|---|---|---|---|---|---|---|---|---|---|---|---|---|---|---|---|---|---|---|---|
| Height | rs1285027 | rs4236086 | T | C | 6 | SUPT3H | Female only | True | 0.1404 | 0.1394 | 0.1365 | 0.1368 | 34577 | 25698 | 193785 | 166603 | -0.05917 | -0.01365 | -0.0179 | -0.01122 | 0.009989 | 0.01171 |
| Height | rs1285027 | rs4711813 | T | C | 6 | SUPT3H | Female only | True | 0.1405 | 0.1394 | 0.1365 | 0.1368 | 34577 | 25698 | 193785 | 166603 | -0.05912 | -0.01398 | -0.01789 | -0.01121 | 0.009985 | 0.01171 |
| Height | rs1285027 | rs12524270 | C | A | 6 | SUPT3H | Female only | True | 0.1404 | 0.1394 | 0.1365 | 0.1369 | 34577 | 25698 | 193785 | 166603 | -0.05929 | -0.01409 | -0.01792 | -0.01133 | 0.009986 | 0.01171 |
| Height | rs1285027 | rs79798635 | G | A | 6 | SUPT3H | Female only | True | 0.1404 | 0.1395 | 0.1365 | 0.1368 | 34577 | 25698 | 193785 | 166603 | -0.05922 | -0.0141 | -0.0179 | -0.0113 | 0.009987 | 0.01171 |
| Height | rs1285027 | rs60346641 | T | G | 6 | SUPT3H | Female only | True | 0.1405 | 0.1394 | 0.1365 | 0.1368 | 34577 | 25698 | 193785 | 166603 | -0.05912 | -0.01398 | -0.01788 | -0.0112 | 0.009985 | 0.01171 |
| Height | rs1285027 | rs56243649 | T | G | 6 | SUPT3H | Female only | True | 0.1405 | 0.1394 | 0.1365 | 0.1368 | 34577 | 25698 | 193785 | 166603 | -0.05912 | -0.01398 | -0.01788 | -0.0112 | 0.009985 | 0.01171 |
| Height | rs1285027 | rs73735354 | G | T | 6 | SUPT3H | Female only | True | 0.1729 | 0.1732 | 0.1729 | 0.173 | 34577 | 25698 | 193785 | 166603 | -0.04576 | -0.01463 | -0.01309 | -0.01028 | 0.009246 | 0.01079 |
| Height | rs1285027 | rs75518301 | A | G | 6 | SUPT3H | Female only | True | 0.1725 | 0.1729 | 0.1729 | 0.173 | 34577 | 25698 | 193785 | 166603 | -0.04623 | -0.01511 | -0.01315 | -0.01025 | 0.00926 | 0.01079 |
| Height | rs1285027 | rs12662685 | A | G | 6 | SUPT3H | Female only | True | 0.1404 | 0.1394 | 0.1365 | 0.1368 | 34577 | 25698 | 193785 | 166603 | -0.059 | -0.01403 | -0.01806 | -0.0112 | 0.009986 | 0.01171 |
| trait | lead_snp | ID | ALLELE1 | ALLELE0 | CHR | Gene_Symbol | independent_loci_in_sex_P1e2 | in_embl_any_or_gene | A1FREQ_VUMC_female | A1FREQ_VUMC_male | A1FREQ_UKB_female | A1FREQ_UKB_male | N_VUMC_female | N_VUMC_male | N_UKB_female | N_UKB_male | BETA_VUMC_female | BETA_VUMC_male | BETA_UKB_female | BETA_UKB_male | SE_VUMC_female | SE_VUMC_male |
| Height | rs1285027 | rs4714850 | T | C | 6 | SUPT3H | Female only | True | 0.1405 | 0.1395 | 0.1366 | 0.1369 | 34577 | 25698 | 193785 | 166603 | -0.05902 | -0.01423 | -0.01809 | -0.01105 | 0.009985 | 0.01171 |
| Height | rs1285027 | rs10948225 | A | G | 6 | SUPT3H | Female only | True | 0.1405 | 0.1394 | 0.1365 | 0.1368 | 34577 | 25698 | 193785 | 166603 | -0.05899 | -0.01423 | -0.01811 | -0.01121 | 0.009982 | 0.01171 |
| Height | rs1285027 | rs12525603 | G | A | 6 | SUPT3H | Female only | True | 0.1426 | 0.1411 | 0.139 | 0.1391 | 34577 | 25698 | 193785 | 166603 | -0.05854 | -0.0119 | -0.01763 | -0.01114 | 0.009924 | 0.01166 |
| Height | rs1285027 | rs80250874 | T | C | 6 | SUPT3H | Female only | True | 0.1406 | 0.1395 | 0.1365 | 0.1368 | 34577 | 25698 | 193785 | 166603 | -0.059 | -0.01419 | -0.0181 | -0.01119 | 0.009982 | 0.01171 |
| Height | rs1285027 | rs4401652 | A | C | 6 | SUPT3H | Female only | True | 0.1407 | 0.1396 | 0.1366 | 0.1369 | 34577 | 25698 | 193785 | 166603 | -0.05871 | -0.0139 | -0.01795 | -0.01105 | 0.00998 | 0.01171 |
| Height | rs1285027 | rs73735369 | G | A | 6 | SUPT3H | Female only | True | 0.1426 | 0.1411 | 0.139 | 0.1391 | 34577 | 25698 | 193785 | 166603 | -0.05826 | -0.01193 | -0.01758 | -0.01116 | 0.009926 | 0.01166 |
| Height | rs1285027 | rs4590263 | A | G | 6 | SUPT3H | Female only | True | 0.1405 | 0.1394 | 0.1365 | 0.1369 | 34577 | 25698 | 193785 | 166603 | -0.05878 | -0.01421 | -0.01791 | -0.01119 | 0.009982 | 0.01171 |
| Height | rs1285027 | rs12526462 | T | C | 6 | SUPT3H | Female only | True | 0.1205 | 0.1208 | 0.116 | 0.1161 | 34577 | 25698 | 193785 | 166603 | -0.05547 | -0.01385 | -0.0186 | -0.01238 | 0.01065 | 0.01247 |
| Height | rs1285027 | rs12526484 | A | C | 6 | SUPT3H | Female only | True | 0.1405 | 0.1394 | 0.1365 | 0.1368 | 34577 | 25698 | 193785 | 166603 | -0.05889 | -0.01419 | -0.01807 | -0.01107 | 0.009982 | 0.01171 |
| Height | rs1285027 | rs59983488 | T | G | 6 | SUPT3H | Female only | True | 0.1409 | 0.1398 | 0.1365 | 0.1369 | 34577 | 25698 | 193785 | 166603 | -0.05871 | -0.01416 | -0.01789 | -0.01121 | 0.009953 | 0.01168 |
| Height | rs1285027 | rs16873345 | C | T | 6 | SUPT3H | Female only | True | 0.1405 | 0.1394 | 0.1365 | 0.1368 | 34577 | 25698 | 193785 | 166603 | -0.0587 | -0.01426 | -0.01801 | -0.01123 | 0.009982 | 0.01171 |
| Height | rs1285027 | rs28360524 | G | A | 6 | SUPT3H | Female only | True | 0.1405 | 0.1394 | 0.1365 | 0.1368 | 34577 | 25698 | 193785 | 166603 | -0.05884 | -0.01427 | -0.01807 | -0.01119 | 0.009981 | 0.01171 |
| Height | rs1285027 | rs7757676 | T | G | 6 | SUPT3H | Female only | True | 0.1205 | 0.1208 | 0.116 | 0.1161 | 34577 | 25698 | 193785 | 166603 | -0.05542 | -0.01369 | -0.01858 | -0.01237 | 0.01065 | 0.01247 |
| Height | rs1285027 | rs10948228 | C | T | 6 | SUPT3H | Female only | True | 0.1425 | 0.1411 | 0.139 | 0.1391 | 34577 | 25698 | 193785 | 166603 | -0.0584 | -0.01202 | -0.01756 | -0.011 | 0.009924 | 0.01166 |
| Height | rs1285027 | rs967588 | T | C | 6 | SUPT3H | Female only | True | 0.1425 | 0.1411 | 0.139 | 0.1391 | 34577 | 25698 | 193785 | 166603 | -0.0584 | -0.01203 | -0.01756 | -0.01102 | 0.009923 | 0.01166 |
| Height | rs1285027 | rs10948230 | G | A | 6 | SUPT3H | Female only | True | 0.1206 | 0.121 | 0.1161 | 0.1162 | 34577 | 25698 | 193785 | 166603 | -0.05521 | -0.01381 | -0.01853 | -0.01247 | 0.01065 | 0.01247 |
| Height | rs1285027 | rs73735378 | C | T | 6 | SUPT3H | Female only | True | 0.1426 | 0.1411 | 0.1391 | 0.1392 | 34577 | 25698 | 193785 | 166603 | -0.05855 | -0.01187 | -0.01753 | -0.01104 | 0.009924 | 0.01166 |
| Height | rs1285027 | rs7760112 | G | T | 6 | SUPT3H | Female only | True | 0.1416 | 0.1406 | 0.1376 | 0.1378 | 34577 | 25698 | 193785 | 166603 | -0.05801 | -0.01457 | -0.01804 | -0.01109 | 0.009954 | 0.01168 |

| trait | lead_snp | ID | ALLELE1 | ALLELE0 | CHR | Gene_Symbol | independent_loci_in_sex_P1e2 | in_embl_any_or_gene | A1FREQ_VUMC_female | A1FREQ_VUMC_male | A1FREQ_UKB_female | A1FREQ_UKB_male | N_VUMC_female | N_VUMC_male | N_UKB_female | N_UKB_male | BETA_VUMC_female | BETA_VUMC_male | BETA_UKB_female | BETA_UKB_male | SE_VUMC_female | SE_VUMC_male |
|---|---|---|---|---|---|---|---|---|---|---|---|---|---|---|---|---|---|---|---|---|---|---|
| Height | rs1285027 | rs16873348 | C | T | 6 | SUPT3H | Female only | True | 0.1221 | 0.1221 | 0.1185 | 0.1184 | 34577 | 25698 | 193785 | 166603 | -0.0547 | -0.01169 | -0.01799 | -0.01217 | 0.0106 | 0.01242 |
| Height | rs1285027 | rs12529295 | C | T | 6 | SUPT3H | Female only | True | 0.1426 | 0.1412 | 0.1391 | 0.1392 | 34577 | 25698 | 193785 | 166603 | -0.05787 | -0.01152 | -0.01762 | -0.01107 | 0.009923 | 0.01165 |
| Height | rs1285027 | rs2024786 | G | A | 6 | SUPT3H | Female only | True | 0.1226 | 0.1225 | 0.1186 | 0.1185 | 34577 | 25698 | 193785 | 166603 | -0.05445 | -0.01113 | -0.0179 | -0.01228 | 0.01058 | 0.0124 |
| Height | rs1285027 | rs6926638 | C | T | 6 | SUPT3H | Female only | True | 0.1225 | 0.1225 | 0.1185 | 0.1185 | 34577 | 25698 | 193785 | 166603 | -0.05468 | -0.01136 | -0.01749 | -0.01184 | 0.01058 | 0.0124 |
| Height | rs1285027 | rs6458440 | T | C | 6 | SUPT3H | Female only | True | 0.1405 | 0.1395 | 0.1364 | 0.1368 | 34577 | 25698 | 193785 | 166603 | -0.05865 | -0.01401 | -0.0178 | -0.01084 | 0.009983 | 0.01171 |
| Height | rs1285027 | rs4711814 | C | T | 6 | SUPT3H | Female only | True | 0.1222 | 0.1221 | 0.1186 | 0.1185 | 34577 | 25698 | 193785 | 166603 | -0.05461 | -0.01193 | -0.01768 | -0.0119 | 0.0106 | 0.01242 |
| Height | rs1285027 | rs1406848 | C | T | 6 | SUPT3H | Female only | True | 0.1425 | 0.1411 | 0.1391 | 0.1392 | 34577 | 25698 | 193785 | 166603 | -0.05804 | -0.01166 | -0.01723 | -0.01065 | 0.009923 | 0.01165 |
| Height | rs1285027 | rs16873366 | G | A | 6 | SUPT3H | Female only | True | 0.1226 | 0.1226 | 0.1187 | 0.1186 | 34577 | 25698 | 193785 | 166603 | -0.05423 | -0.01125 | -0.01763 | -0.01204 | 0.01058 | 0.0124 |
| Height | rs1285027 | rs1852985 | T | C | 6 | SUPT3H | Female only | True | 0.1233 | 0.1238 | 0.1188 | 0.1186 | 34577 | 25698 | 193785 | 166603 | -0.0554 | -0.0105 | -0.01771 | -0.01189 | 0.01055 | 0.01235 |
| Height | rs1285027 | rs6932926 | G | A | 6 | SUPT3H | Female only | True | 0.1206 | 0.1209 | 0.1161 | 0.1163 | 34577 | 25698 | 193785 | 166603 | -0.05508 | -0.01357 | -0.01846 | -0.01226 | 0.01066 | 0.01247 |
| Height | rs1285027 | rs2295296 | T | C | 6 | SUPT3H | Female only | True | 0.1405 | 0.1395 | 0.1366 | 0.1369 | 34577 | 25698 | 193785 | 166603 | -0.05853 | -0.0137 | -0.01787 | -0.01081 | 0.009984 | 0.01171 |
| Height | rs1285027 | rs16873373 | G | T | 6 | SUPT3H | Female only | True | 0.1225 | 0.1225 | 0.1187 | 0.1185 | 34577 | 25698 | 193785 | 166603 | -0.05444 | -0.0114 | -0.01758 | -0.01183 | 0.01059 | 0.01241 |
| Height | rs1285027 | rs16873379 | C | T | 6 | SUPT3H | Female only | True | 0.1226 | 0.1226 | 0.1187 | 0.1186 | 34577 | 25698 | 193785 | 166603 | -0.05414 | -0.01149 | -0.01758 | -0.01198 | 0.01059 | 0.01241 |
| Height | rs1285027 | rs4714853 | C | T | 6 | SUPT3H | Female only | True | 0.1425 | 0.1411 | 0.1392 | 0.1393 | 34577 | 25698 | 193785 | 166603 | -0.05789 | -0.0118 | -0.01723 | -0.01059 | 0.009941 | 0.01167 |
| Height | rs1285027 | rs2295295 | G | T | 6 | SUPT3H | Female only | True | 0.1226 | 0.1226 | 0.1187 | 0.1186 | 34577 | 25698 | 193785 | 166603 | -0.05414 | -0.01157 | -0.01761 | -0.01185 | 0.0106 | 0.01242 |
| Height | rs1285027 | rs3763190 | A | G | 6 | SUPT3H | Female only | True | 0.1221 | 0.122 | 0.1188 | 0.1186 | 34577 | 25698 | 193785 | 166603 | -0.05451 | -0.01251 | -0.01793 | -0.01188 | 0.01062 | 0.01245 |
| Height | rs1285027 | rs35565233 | A | G | 6 | SUPT3H | Female only | True | 0.1224 | 0.1223 | 0.119 | 0.1189 | 34577 | 25698 | 193785 | 166603 | -0.05553 | -0.01169 | -0.01784 | -0.01188 | 0.01061 | 0.01244 |
| Height | rs1285027 | rs76451089 | T | G | 6 | SUPT3H | Female only | True | 0.1239 | 0.1239 | 0.1207 | 0.1205 | 34577 | 25698 | 193785 | 166603 | -0.05395 | -0.01313 | -0.01875 | -0.01221 | 0.01059 | 0.0124 |
| Height | rs1285027 | rs12525950 | G | A | 6 | SUPT3H | Female only | True | 0.1223 | 0.1223 | 0.1187 | 0.1186 | 34577 | 25698 | 193785 | 166603 | -0.05593 | -0.01159 | -0.01771 | -0.01194 | 0.01063 | 0.01245 |
| Height | rs1285027 | rs10948231 | C | T | 6 | SUPT3H | Female only | True | 0.1224 | 0.1223 | 0.1187 | 0.1186 | 34577 | 25698 | 193785 | 166603 | -0.05587 | -0.01142 | -0.01778 | -0.01214 | 0.01062 | 0.01245 |
| Height | rs1285027 | rs717987 | G | A | 6 | SUPT3H | Female only | True | 0.1229 | 0.1229 | 0.1192 | 0.1192 | 34577 | 25698 | 193785 | 166603 | -0.05475 | -0.01215 | -0.01774 | -0.01277 | 0.01061 | 0.01243 |
| Height | rs1907067 | rs9388486 | C | T | 6 | MIR588 | Male only | True | 0.46 | 0.4612 | 0.4514 | 0.4486 | 34577 | 25698 | 193785 | 166603 | 0.006748 | 0.04307 | 0.04114 | 0.05963 | 0.007017 | 0.008217 |
| Height | rs1907067 | rs9375435 | T | C | 6 | MIR588 | Male only | True | 0.4626 | 0.4639 | 0.4531 | 0.4505 | 34577 | 25698 | 193785 | 166603 | 0.006227 | 0.04288 | 0.04115 | 0.05933 | 0.007017 | 0.008209 |
| Height | rs1907067 | rs17754780 | T | C | 6 | MIR588 | Male only | True | 0.461 | 0.462 | 0.452 | 0.4492 | 34577 | 25698 | 193785 | 166603 | 0.006456 | 0.04312 | 0.04125 | 0.05941 | 0.007004 | 0.00821 |
| Height | rs1907067 | rs6907898 | T | C | 6 | MIR588 | Male only | True | 0.4605 | 0.4613 | 0.4516 | 0.4487 | 34577 | 25698 | 193785 | 166603 | 0.006898 | 0.04355 | 0.04116 | 0.05944 | 0.007007 | 0.008213 |
| Height | rs1907067 | rs9401876 | A | C | 6 | MIR588 | Male only | True | 0.4608 | 0.4617 | 0.4519 | 0.4491 | 34577 | 25698 | 193785 | 166603 | 0.006403 | 0.04293 | 0.04123 | 0.05944 | 0.006995 | 0.008202 |
| Height | rs1907067 | rs2045258 | G | A | 6 | MIR588 | Male only | True | 0.4627 | 0.4636 | 0.452 | 0.4492 | 34577 | 25698 | 193785 | 166603 | 0.00644 | 0.04252 | 0.0413 | 0.05944 | 0.006966 | 0.008169 |
| Height | rs1907067 | 6:126678268 | T | G | 6 | MIR588 | Male only | True | 0.4786 | 0.4802 | | | 34577 | 25698 | | | 0.006114 | 0.04289 | | | 0.006995 | 0.00818 |
| Height | rs1907067 | rs2326387 | A | G | 6 | MIR588 | Male only | True | 0.4618 | 0.463 | 0.4523 | 0.4494 | 34577 | 25698 | 193785 | 166603 | 0.005958 | 0.04166 | 0.04131 | 0.05937 | 0.006992 | 0.008197 |

| trait | lead_snp | ID | ALLELE1 | ALLELE0 | CHR | Gene_Symbol | independent_loci_in_sex_P1e2 | in_embl_any_or_gene | A1FREQ_VUMC_female | A1FREQ_VUMC_male | A1FREQ_UKB_female | A1FREQ_UKB_male | N_VUMC_female | N_VUMC_male | N_UKB_female | N_UKB_male | BETA_VUMC_female | BETA_VUMC_male | BETA_UKB_female | BETA_UKB_male | SE_VUMC_female | SE_VUMC_male |
|---|---|---|---|---|---|---|---|---|---|---|---|---|---|---|---|---|---|---|---|---|---|---|
| Height | rs1907067 | rs9398803 | G | A | 6 | MIR588 | Male only | True | 0.479 | 0.4806 | 0.4713 | 0.4685 | 34577 | 25698 | 193785 | 166603 | 0.006453 | 0.04294 | 0.03928 | 0.05632 | 0.006998 | 0.008185 |
| Height | rs1907067 | rs9388489 | G | A | 6 | MIR588 | Male only | True | 0.4627 | 0.4635 | 0.4519 | 0.4491 | 34577 | 25698 | 193785 | 166603 | 0.006581 | 0.04255 | 0.04128 | 0.0594 | 0.006966 | 0.008169 |
| Height | rs1907067 | rs1361262 | C | T | 6 | MIR588 | Male only | True | 0.4607 | 0.4617 | 0.452 | 0.4492 | 34577 | 25698 | 193785 | 166603 | 0.006503 | 0.04265 | 0.04143 | 0.05936 | 0.006996 | 0.008205 |
| Height | rs1907067 | rs9388490 | T | C | 6 | MIR588 | Male only | True | 0.448 | 0.4489 | 0.4389 | 0.4362 | 34577 | 25698 | 193785 | 166603 | 0.005932 | 0.04302 | 0.04308 | 0.05981 | 0.007043 | 0.008242 |
| Height | rs1907067 | rs1591805 | A | G | 6 | MIR588 | Male only | True | 0.4964 | 0.4955 | 0.4975 | 0.4947 | 34577 | 25698 | 193785 | 166603 | -0.004297 | -0.04191 | -0.03588 | -0.0515 | 0.006987 | 0.008197 |
| Height | rs1907067 | rs9398805 | T | C | 6 | MIR588 | Male only | True | 0.4652 | 0.4658 | 0.4562 | 0.4533 | 34577 | 25698 | 193785 | 166603 | 0.005659 | 0.04282 | 0.04157 | 0.05939 | 0.006985 | 0.008195 |
| Height | rs1907067 | rs4897179 | A | G | 6 | MIR588 | Male only | True | 0.4651 | 0.4658 | 0.456 | 0.4531 | 34577 | 25698 | 193785 | 166603 | 0.005763 | 0.04277 | 0.04165 | 0.05953 | 0.006987 | 0.008199 |
| Height | rs1907067 | rs9321065 | A | G | 6 | MIR588 | Male only | True | 0.4664 | 0.4668 | 0.456 | 0.4531 | 34577 | 25698 | 193785 | 166603 | 0.005135 | 0.04342 | 0.0417 | 0.05964 | 0.006962 | 0.008172 |
| Height | rs1907067 | rs2039735 | C | T | 6 | MIR588 | Male only | True | 0.465 | 0.4656 | 0.456 | 0.4531 | 34577 | 25698 | 193785 | 166603 | 0.005146 | 0.04308 | 0.04168 | 0.0596 | 0.006985 | 0.008198 |
| Height | rs1907067 | rs1538172 | G | A | 6 | MIR588 | Male only | True | 0.4649 | 0.4653 | 0.456 | 0.4531 | 34577 | 25698 | 193785 | 166603 | 0.005394 | 0.04407 | 0.04169 | 0.05957 | 0.00698 | 0.008194 |
| Height | rs1907067 | rs9401881 | T | C | 6 | MIR588 | Male only | True | 0.4659 | 0.4664 | 0.4557 | 0.4528 | 34577 | 25698 | 193785 | 166603 | 0.00528 | 0.04405 | 0.0416 | 0.05956 | 0.006963 | 0.00817 |
| Height | rs1907067 | rs4565329 | T | C | 6 | MIR588 | Male only | True | 0.466 | 0.4664 | 0.4558 | 0.4529 | 34577 | 25698 | 193785 | 166603 | 0.005423 | 0.04394 | 0.04168 | 0.05959 | 0.006961 | 0.008169 |
| Height | rs1907067 | rs1538170 | T | C | 6 | MIR588 | Male only | True | 0.466 | 0.4664 | 0.4558 | 0.4529 | 34577 | 25698 | 193785 | 166603 | 0.005423 | 0.04394 | 0.04168 | 0.05959 | 0.006961 | 0.008169 |
| trait | lead_snp | ID | ALLELE1 | ALLELE0 | CHR | Gene_Symbol | independent_loci_in_sex_P1e2 | in_embl_any_or_gene | A1FREQ_VUMC_female | A1FREQ_VUMC_male | A1FREQ_UKB_female | A1FREQ_UKB_male | N_VUMC_female | N_VUMC_male | N_UKB_female | N_UKB_male | BETA_VUMC_female | BETA_VUMC_male | BETA_UKB_female | BETA_UKB_male | SE_VUMC_female | SE_VUMC_male |
| Height | rs1907067 | rs4897181 | T | C | 6 | MIR588 | Male only | True | 0.4665 | 0.4669 | 0.4559 | 0.453 | 34577 | 25698 | 193785 | 166603 | 0.00529 | 0.04356 | 0.04173 | 0.05951 | 0.006962 | 0.008171 |
| Height | rs1907067 | rs9401882 | G | A | 6 | MIR588 | Male only | True | 0.4662 | 0.4666 | 0.456 | 0.4531 | 34577 | 25698 | 193785 | 166603 | 0.005407 | 0.04422 | 0.04169 | 0.0596 | 0.00696 | 0.008169 |
| Height | rs1907067 | rs4897182 | G | T | 6 | MIR588 | Male only | True | 0.4653 | 0.4657 | 0.4562 | 0.4532 | 34577 | 25698 | 193785 | 166603 | 0.005248 | 0.04399 | 0.04172 | 0.05959 | 0.00698 | 0.008193 |
| Height | rs1907067 | rs9398808 | T | G | 6 | MIR588 | Male only | True | 0.4661 | 0.4664 | 0.4559 | 0.453 | 34577 | 25698 | 193785 | 166603 | 0.005229 | 0.04397 | 0.04174 | 0.05953 | 0.00696 | 0.00817 |
| Height | rs1907067 | rs9385399 | T | G | 6 | MIR588 | Male only | True | 0.4664 | 0.4668 | 0.456 | 0.4532 | 34577 | 25698 | 193785 | 166603 | 0.005337 | 0.04401 | 0.0417 | 0.05958 | 0.006959 | 0.00817 |
| Height | rs1907067 | rs1415671 | G | T | 6 | MIR588 | Male only | True | 0.4666 | 0.4671 | 0.4561 | 0.4532 | 34577 | 25698 | 193785 | 166603 | 0.005193 | 0.04399 | 0.04177 | 0.0595 | 0.00696 | 0.00817 |
| Height | rs1907067 | rs2184968 | C | T | 6 | MIR588 | Male only | True | 0.4655 | 0.4663 | 0.4538 | 0.4509 | 34577 | 25698 | 193785 | 166603 | 0.005702 | 0.04357 | 0.04162 | 0.05929 | 0.006962 | 0.008167 |
| Height | rs1907067 | rs2152876 | A | G | 6 | MIR588 | Male only | True | 0.4666 | 0.4671 | 0.4561 | 0.4532 | 34577 | 25698 | 193785 | 166603 | 0.005328 | 0.04402 | 0.04175 | 0.05951 | 0.006962 | 0.00817 |
| Height | rs1907067 | rs9385400 | G | T | 6 | MIR588 | Male only | True | 0.4662 | 0.4666 | 0.4559 | 0.4531 | 34577 | 25698 | 193785 | 166603 | 0.005529 | 0.04422 | 0.04175 | 0.05955 | 0.00696 | 0.008168 |
| Height | rs1907067 | rs1361107 | C | A | 6 | MIR588 | Male only | True | 0.4661 | 0.4664 | 0.4559 | 0.4531 | 34577 | 25698 | 193785 | 166603 | 0.005133 | 0.04368 | 0.04159 | 0.05946 | 0.00696 | 0.008168 |
| Height | rs1907067 | rs1361108 | T | C | 6 | MIR588 | Male only | True | 0.4663 | 0.4668 | 0.4558 | 0.4528 | 34577 | 25698 | 193785 | 166603 | 0.005099 | 0.04338 | 0.04185 | 0.05953 | 0.006961 | 0.008169 |
| Height | rs1907067 | rs1361109 | T | C | 6 | MIR588 | Male only | True | 0.4668 | 0.4678 | 0.4556 | 0.4527 | 34577 | 25698 | 193785 | 166603 | 0.004357 | 0.04245 | 0.04184 | 0.05931 | 0.006961 | 0.008168 |
| Height | rs1907067 | rs1572569 | A | G | 6 | MIR588 | Male only | True | 0.4667 | 0.4676 | 0.4554 | 0.4525 | 34577 | 25698 | 193785 | 166603 | 0.00458 | 0.04244 | 0.04175 | 0.05939 | 0.00696 | 0.008167 |
| Height | rs1907067 | rs2184967 | T | C | 6 | MIR588 | Male only | True | 0.4653 | 0.4659 | 0.455 | 0.4521 | 34577 | 25698 | 193785 | 166603 | 0.00498 | 0.04373 | 0.0418 | 0.05954 | 0.006959 | 0.00817 |

| trait | lead_snp | ID | ALLELE1 | ALLELE0 | CHR | Gene_Symbol | independent_loci_in_sex_P1e2 | in_embl_any_or_gene | A1FREQ_VUMC_female | A1FREQ_VUMC_male | A1FREQ_UKB_female | A1FREQ_UKB_male | N_VUMC_female | N_VUMC_male | N_UKB_female | N_UKB_male | BETA_VUMC_female | BETA_VUMC_male | BETA_UKB_female | BETA_UKB_male | SE_VUMC_female | SE_VUMC_male |
|---|---|---|---|---|---|---|---|---|---|---|---|---|---|---|---|---|---|---|---|---|---|---|
| Height | rs1907067 | rs4559102 | A | G | 6 | MIR588 | Male only | True | 0.4657 | 0.4664 | 0.4551 | 0.4523 | 34577 | 25698 | 193785 | 166603 | 0.004928 | 0.04328 | 0.04182 | 0.05949 | 0.006959 | 0.008172 |
| Height | rs1907067 | rs9388494 | T | G | 6 | MIR588 | Male only | True | 0.4657 | 0.4664 | 0.4552 | 0.4523 | 34577 | 25698 | 193785 | 166603 | 0.004848 | 0.0434 | 0.04173 | 0.05957 | 0.006959 | 0.008172 |
| Height | rs1907067 | rs9375441 | A | G | 6 | MIR588 | Male only | True | 0.4644 | 0.4651 | 0.4552 | 0.4523 | 34577 | 25698 | 193785 | 166603 | 0.005038 | 0.04339 | 0.0417 | 0.05953 | 0.006976 | 0.008194 |
| Height | rs1907067 | rs4895808 | T | C | 6 | MIR588 | Male only | True | 0.4658 | 0.4664 | 0.4552 | 0.4523 | 34577 | 25698 | 193785 | 166603 | 0.004995 | 0.04364 | 0.04174 | 0.05953 | 0.006959 | 0.008175 |
| Height | rs1907067 | rs9388495 | A | G | 6 | MIR588 | Male only | True | 0.4657 | 0.4666 | 0.4555 | 0.4526 | 34577 | 25698 | 193785 | 166603 | 0.004864 | 0.04294 | 0.04177 | 0.05957 | 0.006976 | 0.00819 |
| Height | rs1907067 | rs9388496 | G | A | 6 | MIR588 | Male only | True | 0.4662 | 0.467 | 0.4553 | 0.4524 | 34577 | 25698 | 193785 | 166603 | 0.005415 | 0.04364 | 0.04178 | 0.05954 | 0.006959 | 0.008174 |
| Height | rs1907067 | rs1844594 | A | G | 6 | MIR588 | Male only | True | 0.4658 | 0.4666 | 0.4554 | 0.4525 | 34577 | 25698 | 193785 | 166603 | 0.005141 | 0.04312 | 0.04174 | 0.05934 | 0.006977 | 0.008191 |
| Height | rs1907067 | rs9398809 | T | C | 6 | MIR588 | Male only | True | 0.4659 | 0.4664 | 0.4552 | 0.4523 | 34577 | 25698 | 193785 | 166603 | 0.005257 | 0.04395 | 0.04177 | 0.05955 | 0.006959 | 0.008174 |
| Height | rs1907067 | rs9385401 | T | C | 6 | MIR588 | Male only | True | 0.4646 | 0.4651 | 0.4552 | 0.4523 | 34577 | 25698 | 193785 | 166603 | 0.005219 | 0.04409 | 0.04177 | 0.05954 | 0.006977 | 0.008194 |
| Height | rs1907067 | rs9401883 | G | A | 6 | MIR588 | Male only | True | 0.466 | 0.4665 | 0.4552 | 0.4524 | 34577 | 25698 | 193785 | 166603 | 0.005551 | 0.04415 | 0.0418 | 0.05953 | 0.006958 | 0.008173 |
| Height | rs1907067 | rs1159619 | A | C | 6 | MIR588 | Male only | True | 0.4637 | 0.4644 | 0.4534 | 0.4505 | 34577 | 25698 | 193785 | 166603 | 0.004694 | 0.04342 | 0.04156 | 0.05938 | 0.006958 | 0.008176 |
| Height | rs1907067 | rs9375442 | C | A | 6 | MIR588 | Male only | True | 0.4672 | 0.4681 | 0.4556 | 0.4526 | 34577 | 25698 | 193785 | 166603 | 0.005215 | 0.04339 | 0.04181 | 0.05932 | 0.006959 | 0.008171 |
| Height | rs1907067 | rs2050644 | G | A | 6 | MIR588 | Male only | True | 0.4659 | 0.4664 | 0.455 | 0.4522 | 34577 | 25698 | 193785 | 166603 | 0.005424 | 0.04406 | 0.04169 | 0.05957 | 0.006959 | 0.008172 |
| Height | rs1907067 | rs4422634 | C | T | 6 | MIR588 | Male only | True | 0.466 | 0.4665 | 0.4549 | 0.4521 | 34577 | 25698 | 193785 | 166603 | 0.00534 | 0.04417 | 0.04177 | 0.05947 | 0.006961 | 0.008172 |
| Height | rs1907067 | rs1120786 | G | T | 6 | MIR588 | Male only | True | 0.4659 | 0.4664 | 0.455 | 0.4522 | 34577 | 25698 | 193785 | 166603 | 0.005395 | 0.0443 | 0.04164 | 0.05956 | 0.006959 | 0.008171 |
| Height | rs1907067 | rs9372839 | A | G | 6 | MIR588 | Male only | True | 0.4657 | 0.4662 | 0.4547 | 0.4519 | 34577 | 25698 | 193785 | 166603 | 0.005187 | 0.04411 | 0.04177 | 0.05959 | 0.00696 | 0.008172 |
| Height | rs1907067 | rs9398810 | A | C | 6 | MIR588 | Male only | True | 0.4648 | 0.4654 | 0.4552 | 0.4525 | 34577 | 25698 | 193785 | 166603 | 0.005198 | 0.0438 | 0.04159 | 0.05961 | 0.006978 | 0.008196 |
| Height | rs1907067 | rs4418209 | G | T | 6 | MIR588 | Male only | True | 0.466 | 0.4665 | 0.4552 | 0.4525 | 34577 | 25698 | 193785 | 166603 | 0.005318 | 0.0445 | 0.04151 | 0.05963 | 0.006959 | 0.008172 |
| Height | rs1907067 | rs9372840 | C | A | 6 | MIR588 | Male only | True | 0.4664 | 0.4669 | 0.4553 | 0.4525 | 34577 | 25698 | 193785 | 166603 | 0.005544 | 0.04435 | 0.04149 | 0.05952 | 0.00696 | 0.008173 |
| Height | rs1907067 | rs2130603 | C | A | 6 | MIR588 | Male only | True | 0.465 | 0.4656 | 0.4553 | 0.4525 | 34577 | 25698 | 193785 | 166603 | 0.005645 | 0.04468 | 0.04149 | 0.05951 | 0.00698 | 0.008195 |
| Height | rs1907067 | rs7738135 | A | G | 6 | MIR588 | Male only | True | 0.4657 | 0.4662 | 0.4551 | 0.4524 | 34577 | 25698 | 193785 | 166603 | 0.005326 | 0.04448 | 0.04142 | 0.05954 | 0.00696 | 0.008171 |
| Height | rs1907067 | rs4053271 | C | T | 6 | MIR588 | Male only | True | 0.4645 | 0.465 | 0.4552 | 0.4524 | 34577 | 25698 | 193785 | 166603 | 0.005444 | 0.0447 | 0.0414 | 0.05956 | 0.006979 | 0.008194 |
| Height | rs1907067 | rs1490387 | A | C | 6 | MIR588 | Male only | True | 0.4657 | 0.4662 | 0.4551 | 0.4524 | 34577 | 25698 | 193785 | 166603 | 0.005294 | 0.04431 | 0.04138 | 0.05958 | 0.006961 | 0.008172 |
| Height | rs1907067 | rs9375446 | A | G | 6 | MIR588 | Male only | True | 0.4657 | 0.4662 | 0.4551 | 0.4524 | 34577 | 25698 | 193785 | 166603 | 0.005221 | 0.04432 | 0.0414 | 0.05957 | 0.00696 | 0.008172 |
| Height | rs1907067 | rs1490388 | T | C | 6 | MIR588 | Male only | True | 0.4656 | 0.4662 | 0.455 | 0.4523 | 34577 | 25698 | 193785 | 166603 | 0.005141 | 0.04427 | 0.04138 | 0.05947 | 0.00696 | 0.008171 |
| Height | rs1907067 | rs1907067 | C | A | 6 | MIR588 | Male only | True | 0.4659 | 0.4664 | 0.455 | 0.4522 | 34577 | 25698 | 193785 | 166603 | 0.005509 | 0.04462 | 0.04137 | 0.05952 | 0.00696 | 0.008171 |
| Height | rs1907067 | 6:126838605 | C | A | 6 | MIR588 | Male only | True | 0.4659 | 0.4667 | | | 34577 | 25698 | | | 0.004979 | 0.0438 | | | 0.00698 | 0.00819 |
| Height | rs1907067 | rs1490384 | C | T | 6 | MIR588 | Male only | True | 0.493 | 0.4928 | 0.4995 | 0.4967 | 34577 | 25698 | 193785 | 166603 | -0.008395 | -0.04449 | -0.04426 | -0.05798 | 0.006961 | 0.008154 |
| Height | rs1907067 | rs6925689 | T | C | 6 | MIR588 | Male only | True | 0.4892 | 0.4897 | 0.4937 | 0.4965 | 34577 | 25698 | 193785 | 166603 | -0.009606 | -0.04086 | -0.04105 | -0.05613 | 0.006986 | 0.008167 |

| trait | lead_snp | ID | ALLELE1 | ALLELE0 | CHR | Gene_Symbol | independent_loci_in_sex_P1e2 | in_embl_any_or_gene | A1FREQ_VUMC_female | A1FREQ_VUMC_male | A1FREQ_UKB_female | A1FREQ_UKB_male | N_VUMC_female | N_VUMC_male | N_UKB_female | N_UKB_male | BETA_VUMC_female | BETA_VUMC_male | BETA_UKB_female | BETA_UKB_male | SE_VUMC_female | SE_VUMC_male |
|---|---|---|---|---|---|---|---|---|---|---|---|---|---|---|---|---|---|---|---|---|---|---|
| Height | rs1907067 | rs1155939 | A | C | 6 | MIR588 | Male only | True | 0.4982 | 0.4978 | 0.4928 | 0.4899 | 34577 | 25698 | 193785 | 166603 | 0.007052 | 0.04427 | 0.04272 | 0.05687 | 0.006963 | 0.008154 |
| Height | rs1907067 | rs1155938 | A | C | 6 | MIR588 | Male only | True | 0.4986 | 0.4985 | 0.493 | 0.49 | 34577 | 25698 | 193785 | 166603 | 0.007401 | 0.04326 | 0.0428 | 0.05694 | 0.006962 | 0.008156 |
| Height | rs1907067 | rs9375447 | G | A | 6 | MIR588 | Male only | True | 0.498 | 0.4977 | 0.4924 | 0.4894 | 34577 | 25698 | 193785 | 166603 | 0.007406 | 0.04443 | 0.04282 | 0.05681 | 0.00696 | 0.008155 |
| Height | rs1907067 | rs1490385 | C | T | 6 | MIR588 | Male only | True | 0.498 | 0.4981 | 0.4928 | 0.4897 | 34577 | 25698 | 193785 | 166603 | -0.007372 | -0.043 | -0.04288 | -0.05668 | 0.006978 | 0.008174 |
| Height | rs1907067 | rs9401888 | G | A | 6 | MIR588 | Male only | True | 0.4982 | 0.498 | 0.4928 | 0.4899 | 34577 | 25698 | 193785 | 166603 | 0.007385 | 0.04356 | 0.04276 | 0.05675 | 0.00696 | 0.008156 |
| Height | rs1907067 | rs9401890 | G | A | 6 | MIR588 | Male only | True | 0.497 | 0.4968 | 0.4928 | 0.4898 | 34577 | 25698 | 193785 | 166603 | 0.006925 | 0.04419 | 0.04282 | 0.05664 | 0.006979 | 0.00818 |
| Height | rs1907067 | rs6931369 | T | C | 6 | MIR588 | Male only | True | 0.4965 | 0.4962 | 0.4926 | 0.4896 | 34577 | 25698 | 193785 | 166603 | 0.006958 | 0.0442 | 0.04283 | 0.05667 | 0.006979 | 0.008178 |
| Height | rs1907067 | rs9401891 | T | G | 6 | MIR588 | Male only | True | 0.4978 | 0.4975 | 0.4926 | 0.4897 | 34577 | 25698 | 193785 | 166603 | 0.007219 | 0.04396 | 0.04267 | 0.05664 | 0.006961 | 0.008157 |
| Height | rs1907067 | rs9401892 | A | G | 6 | MIR588 | Male only | True | 0.4988 | 0.499 | 0.4929 | 0.49 | 34577 | 25698 | 193785 | 166603 | 0.006669 | 0.04291 | 0.04254 | 0.05637 | 0.006961 | 0.008155 |
| Height | rs1907067 | rs4621655 | A | G | 6 | MIR588 | Male only | True | 0.4989 | 0.499 | 0.493 | 0.49 | 34577 | 25698 | 193785 | 166603 | 0.00677 | 0.043 | 0.04248 | 0.05633 | 0.00696 | 0.008155 |
| Height | rs1907067 | rs4621657 | A | G | 6 | MIR588 | Male only | True | 0.4989 | 0.499 | 0.493 | 0.49 | 34577 | 25698 | 193785 | 166603 | 0.00677 | 0.043 | 0.04247 | 0.05633 | 0.00696 | 0.008155 |
| Height | rs1907067 | rs1844593 | A | G | 6 | MIR588 | Male only | True | 0.499 | 0.4991 | 0.493 | 0.49 | 34577 | 25698 | 193785 | 166603 | 0.007111 | 0.04326 | 0.04249 | 0.05633 | 0.00696 | 0.008155 |
| Height | rs1907067 | rs3861455 | C | A | 6 | MIR588 | Male only | True | 0.498 | 0.4978 | 0.4927 | 0.4898 | 34577 | 25698 | 193785 | 166603 | 0.007392 | 0.04441 | 0.04253 | 0.05649 | 0.006959 | 0.008156 |
| Height | rs1907067 | rs9372841 | G | A | 6 | MIR588 | Male only | True | 0.4983 | 0.4982 | 0.4928 | 0.4899 | 34577 | 25698 | 193785 | 166603 | 0.007607 | 0.04385 | 0.0425 | 0.05655 | 0.006959 | 0.008155 |
| Height | rs1907067 | rs3861456 | G | A | 6 | MIR588 | Male only | True | 0.4983 | 0.4982 | 0.4928 | 0.4899 | 34577 | 25698 | 193785 | 166603 | 0.007607 | 0.04385 | 0.0425 | 0.05655 | 0.006959 | 0.008155 |
| Height | rs1907067 | rs3861457 | T | C | 6 | MIR588 | Male only | True | 0.5007 | 0.5007 | 0.493 | 0.49 | 34577 | 25698 | 193785 | 166603 | -0.007139 | -0.0437 | -0.04254 | -0.05638 | 0.00696 | 0.008155 |
| Height | rs1907067 | rs9388501 | T | C | 6 | MIR588 | Male only | True | 0.5006 | 0.5004 | 0.493 | 0.49 | 34577 | 25698 | 193785 | 166603 | -0.007211 | -0.04277 | -0.04253 | -0.05646 | 0.00696 | 0.008155 |

| trait | lead_snp | ID | ALLELE1 | ALLELE0 | CHR | Gene_Symbol | independent_loci_in_sex_P1e2 | in_embl_any_or_gene | A1FREQ_VUMC_female | A1FREQ_VUMC_male | A1FREQ_UKB_female | A1FREQ_UKB_male | N_VUMC_female | N_VUMC_male | N_UKB_female | N_UKB_male | BETA_VUMC_female | BETA_VUMC_male | BETA_UKB_female | BETA_UKB_male | SE_VUMC_female | SE_VUMC_male |
|---|---|---|---|---|---|---|---|---|---|---|---|---|---|---|---|---|---|---|---|---|---|---|
| Height | rs1907067 | rs9401893 | A | G | 6 | MIR588 | Male only | True | 0.4981 | 0.4983 | 0.4932 | 0.4902 | 34577 | 25698 | 193785 | 166603 | -0.007434 | -0.04287 | -0.04247 | -0.05638 | 0.006977 | 0.008176 |
| Height | rs1907067 | rs9388502 | A | G | 6 | MIR588 | Male only | True | 0.4975 | 0.4976 | 0.4929 | 0.4899 | 34577 | 25698 | 193785 | 166603 | 0.006808 | 0.04285 | 0.04246 | 0.05635 | 0.006979 | 0.008175 |
| Height | rs1907067 | rs3861458 | A | G | 6 | MIR588 | Male only | True | 0.4978 | 0.4976 | 0.4926 | 0.4897 | 34577 | 25698 | 193785 | 166603 | 0.007538 | 0.04398 | 0.04243 | 0.05652 | 0.00696 | 0.008157 |
| Height | rs1907067 | rs9372842 | T | C | 6 | MIR588 | Male only | True | 0.4986 | 0.4984 | 0.494 | 0.4911 | 34577 | 25698 | 193785 | 166603 | -0.008227 | -0.04235 | -0.04202 | -0.05628 | 0.006976 | 0.00818 |
| Height | rs1907067 | rs1826189 | T | C | 6 | MIR588 | Male only | True | 0.4992 | 0.4988 | 0.4938 | 0.4909 | 34577 | 25698 | 193785 | 166603 | 0.00802 | 0.0423 | 0.04196 | 0.05637 | 0.006961 | 0.008159 |
| Height | rs1907067 | rs9482757 | A | C | 6 | MIR588 | Male only | True | 0.4996 | 0.4995 | 0.4942 | 0.4913 | 34577 | 25698 | 193785 | 166603 | -0.007609 | -0.04156 | -0.04197 | -0.05618 | 0.00696 | 0.008158 |
| Height | rs1907067 | rs12208811 | G | T | 6 | MIR588 | Male only | True | 0.499 | 0.4987 | 0.4936 | 0.4908 | 34577 | 25698 | 193785 | 166603 | 0.007743 | 0.04308 | 0.042 | 0.05631 | 0.00696 | 0.00816 |
| Height | rs1907067 | rs7743860 | G | T | 6 | MIR588 | Male only | True | 0.4991 | 0.4989 | 0.4937 | 0.4909 | 34577 | 25698 | 193785 | 166603 | 0.007794 | 0.04234 | 0.04198 | 0.05634 | 0.00696 | 0.008159 |
| Height | rs1907067 | rs4584000 | C | T | 6 | MIR588 | Male only | True | 0.498 | 0.4981 | 0.4934 | 0.4905 | 34577 | 25698 | 193785 | 166603 | 0.007857 | 0.04277 | 0.04196 | 0.05622 | 0.006979 | 0.008185 |
| Height | rs1907067 | rs6914367 | T | C | 6 | MIR588 | Male only | True | 0.4982 | 0.498 | 0.4938 | 0.4909 | 34577 | 25698 | 193785 | 166603 | 0.007526 | 0.04296 | 0.04205 | 0.05622 | 0.006975 | 0.00818 |

| trait | lead_snp | ID | ALLELE1 | ALLELE0 | CHR | Gene_Symbol | independent_loci_in_sex_P1e2 | in_embl_any_or_gene | A1FREQ_VUMC_female | A1FREQ_VUMC_male | A1FREQ_UKB_female | A1FREQ_UKB_male | N_VUMC_female | N_VUMC_male | N_UKB_female | N_UKB_male | BETA_VUMC_female | BETA_VUMC_male | BETA_UKB_female | BETA_UKB_male | SE_VUMC_female | SE_VUMC_male |
|---|---|---|---|---|---|---|---|---|---|---|---|---|---|---|---|---|---|---|---|---|---|---|
| Height | rs1907067 | rs7738836 | A | G | 6 | MIR588 | Male only | True | 0.4982 | 0.4981 | 0.4938 | 0.491 | 34577 | 25698 | 193785 | 166603 | -0.007483 | -0.04302 | -0.04205 | -0.05627 | 0.006974 | 0.008181 |
| Height | rs1907067 | rs4339479 | A | G | 6 | MIR588 | Male only | True | 0.4981 | 0.4979 | 0.4938 | 0.4909 | 34577 | 25698 | 193785 | 166603 | 0.007461 | 0.04276 | 0.04205 | 0.05628 | 0.006975 | 0.00818 |
| Height | rs1907067 | rs4629707 | T | C | 6 | MIR588 | Male only | True | 0.4997 | 0.4995 | 0.4941 | 0.4912 | 34577 | 25698 | 193785 | 166603 | -0.007495 | -0.04152 | -0.04198 | -0.05625 | 0.00696 | 0.008159 |
| Height | rs1907067 | rs4243491 | C | T | 6 | MIR588 | Male only | True | 0.4992 | 0.499 | 0.4937 | 0.4909 | 34577 | 25698 | 193785 | 166603 | 0.00809 | 0.04242 | 0.04202 | 0.05644 | 0.00696 | 0.008161 |
| Height | rs1907067 | rs9401897 | G | A | 6 | MIR588 | Male only | True | 0.4992 | 0.499 | 0.4938 | 0.491 | 34577 | 25698 | 193785 | 166603 | 0.008163 | 0.0424 | 0.04196 | 0.05639 | 0.006959 | 0.008161 |
| Height | rs1907067 | rs9491653 | A | G | 6 | MIR588 | Male only | True | 0.5006 | 0.5005 | 0.4953 | 0.4925 | 34577 | 25698 | 193785 | 166603 | -0.008591 | -0.04222 | -0.04233 | -0.05653 | 0.006959 | 0.008164 |
| Height | rs1907067 | rs1340952 | T | C | 6 | MIR588 | Male only | True | 0.4991 | 0.4991 | 0.4936 | 0.4908 | 34577 | 25698 | 193785 | 166603 | 0.007518 | 0.04265 | 0.04225 | 0.05625 | 0.00696 | 0.008163 |
| Height | rs1907067 | rs1340953 | G | A | 6 | MIR588 | Male only | True | 0.4992 | 0.4993 | 0.4937 | 0.4908 | 34577 | 25698 | 193785 | 166603 | 0.007214 | 0.04276 | 0.04219 | 0.05619 | 0.006959 | 0.008164 |
| Height | rs1907067 | rs1417572 | T | C | 6 | MIR588 | Male only | True | 0.5005 | 0.5002 | 0.4935 | 0.4907 | 34577 | 25698 | 193785 | 166603 | -0.008209 | -0.04229 | -0.04232 | -0.05626 | 0.006958 | 0.008164 |
| Height | rs1907067 | rs9375449 | C | T | 6 | MIR588 | Male only | True | 0.4989 | 0.499 | 0.4933 | 0.4905 | 34577 | 25698 | 193785 | 166603 | 0.008446 | 0.04228 | 0.0423 | 0.05635 | 0.006959 | 0.008161 |
| Height | rs1907067 | rs1538956 | T | G | 6 | MIR588 | Male only | True | 0.4985 | 0.4985 | 0.4932 | 0.4904 | 34577 | 25698 | 193785 | 166603 | 0.008307 | 0.0429 | 0.04238 | 0.05636 | 0.006959 | 0.008162 |
| Height | rs1907067 | rs4626447 | A | G | 6 | MIR588 | Male only | True | 0.4972 | 0.4973 | 0.4933 | 0.4905 | 34577 | 25698 | 193785 | 166603 | 0.008201 | 0.04331 | 0.04238 | 0.0563 | 0.006976 | 0.008183 |
| Height | rs1907067 | rs4549631 | C | T | 6 | MIR588 | Male only | True | 0.4992 | 0.4994 | 0.4935 | 0.4906 | 34577 | 25698 | 193785 | 166603 | 0.007934 | 0.04215 | 0.04244 | 0.05572 | 0.006958 | 0.008165 |
| Height | rs1907067 | rs9388507 | A | C | 6 | MIR588 | Male only | True | 0.4984 | 0.4985 | 0.4932 | 0.4904 | 34577 | 25698 | 193785 | 166603 | 0.008409 | 0.04223 | 0.04248 | 0.05622 | 0.006958 | 0.008163 |
| Height | rs1907067 | rs3905044 | G | A | 6 | MIR588 | Male only | True | 0.4986 | 0.4987 | 0.4932 | 0.4904 | 34577 | 25698 | 193785 | 166603 | 0.008108 | 0.04225 | 0.04248 | 0.0562 | 0.006958 | 0.008163 |
| Height | rs1907067 | rs6911485 | G | A | 6 | MIR588 | Male only | True | 0.4974 | 0.4975 | 0.4931 | 0.4904 | 34577 | 25698 | 193785 | 166603 | 0.008159 | 0.04171 | 0.04251 | 0.05623 | 0.006975 | 0.008184 |
| Height | rs1907067 | rs4895813 | T | G | 6 | MIR588 | Male only | True | 0.4971 | 0.4972 | 0.4932 | 0.4904 | 34577 | 25698 | 193785 | 166603 | 0.00832 | 0.04245 | 0.04247 | 0.05624 | 0.006975 | 0.008184 |
| Height | rs1907067 | rs6913430 | T | G | 6 | MIR588 | Male only | True | 0.498 | 0.4984 | 0.4935 | 0.4907 | 34577 | 25698 | 193785 | 166603 | -0.008191 | -0.04167 | -0.04243 | -0.05617 | 0.006974 | 0.008185 |
| Height | rs1907067 | rs9375450 | A | G | 6 | MIR588 | Male only | True | 0.4983 | 0.4985 | 0.493 | 0.4903 | 34577 | 25698 | 193785 | 166603 | 0.008294 | 0.04239 | 0.04236 | 0.0564 | 0.006958 | 0.008165 |
| Height | rs1907067 | rs9388509 | G | A | 6 | MIR588 | Male only | True | 0.5004 | 0.4998 | 0.4935 | 0.4907 | 34577 | 25698 | 193785 | 166603 | -0.007569 | -0.0415 | -0.04227 | -0.05617 | 0.006957 | 0.008163 |
| Height | rs1907067 | rs6918725 | T | G | 6 | MIR588 | Male only | True | 0.4782 | 0.4761 | 0.4807 | 0.4833 | 34577 | 25698 | 193785 | 166603 | -0.007143 | -0.03615 | -0.04068 | -0.05441 | 0.006953 | 0.008165 |
| Height | rs1907067 | rs8180649 | C | T | 6 | MIR588 | Male only | True | 0.4976 | 0.4981 | 0.4934 | 0.4906 | 34577 | 25698 | 193785 | 166603 | 0.008274 | 0.04178 | 0.04229 | 0.05629 | 0.006976 | 0.008189 |
| Height | rs1907067 | rs6925607 | T | C | 6 | MIR588 | Male only | True | 0.4976 | 0.498 | 0.4933 | 0.4905 | 34577 | 25698 | 193785 | 166603 | 0.008024 | 0.04187 | 0.04226 | 0.05626 | 0.006976 | 0.008188 |
| Height | rs1907067 | rs75169667 | G | A | 6 | MIR588 | Male only | True | 0.4971 | 0.4974 | | | 34577 | 25698 | | | 0.008029 | 0.04181 | | | 0.006978 | 0.008187 |
| Height | rs1907067 | 6:127001534 | G | T | 6 | MIR588 | Male only | True | 0.497 | 0.4972 | | | 34577 | 25698 | | | 0.008136 | 0.04273 | | | 0.006978 | 0.00819 |
| Height | rs1907067 | rs853961 | T | G | 6 | MIR588 | Male only | True | 0.5004 | 0.4999 | 0.4935 | 0.4907 | 34577 | 25698 | 193785 | 166603 | -0.007186 | -0.04117 | -0.04222 | -0.05613 | 0.006958 | 0.008163 |
| Height | rs1907067 | rs6569469 | C | T | 6 | MIR588 | Male only | True | 0.4989 | 0.4992 | 0.4933 | 0.4906 | 34577 | 25698 | 193785 | 166603 | 0.007647 | 0.04167 | 0.04227 | 0.05619 | 0.006958 | 0.008166 |
| Height | rs1907067 | rs864937 | A | G | 6 | MIR588 | Male only | True | 0.5005 | 0.5001 | 0.4935 | 0.4907 | 34577 | 25698 | 193785 | 166603 | -0.007577 | -0.04128 | -0.04215 | -0.05606 | 0.006958 | 0.008163 |
| Height | rs1907067 | rs853985 | C | T | 6 | MIR588 | Male only | True | 0.4983 | 0.4984 | 0.4931 | 0.4904 | 34577 | 25698 | 193785 | 166603 | 0.007931 | 0.04185 | 0.04224 | 0.05624 | 0.006959 | 0.008163 |

| trait | lead_snp | ID | ALLELE1 | ALLELE0 | CHR | Gene_Symbol | independent_loci_in_sex_P1e2 | in_embl_any_or_gene | A1FREQ_VUMC_female | A1FREQ_VUMC_male | A1FREQ_UKB_female | A1FREQ_UKB_male | N_VUMC_female | N_VUMC_male | N_UKB_female | N_UKB_male | BETA_VUMC_female | BETA_VUMC_male | BETA_UKB_female | BETA_UKB_male | SE_VUMC_female | SE_VUMC_male |
|---|---|---|---|---|---|---|---|---|---|---|---|---|---|---|---|---|---|---|---|---|---|---|
| Height | rs1907067 | rs853987 | A | G | 6 | MIR588 | Male only | True | 0.499 | 0.4991 | 0.4934 | 0.4906 | 34577 | 25698 | 193785 | 166603 | 0.007916 | 0.04201 | 0.04222 | 0.05618 | 0.006958 | 0.008166 |
| Height | rs1907067 | rs853962 | G | T | 6 | MIR588 | Male only | True | 0.5001 | 0.5001 | 0.4932 | 0.4904 | 34577 | 25698 | 193785 | 166603 | 0.007896 | 0.04176 | 0.04224 | 0.05625 | 0.006979 | 0.008187 |
| Height | rs1907067 | rs4460221 | A | G | 6 | MIR588 | Male only | True | 0.5005 | 0.5001 | 0.4935 | 0.4907 | 34577 | 25698 | 193785 | 166603 | -0.007272 | -0.04066 | -0.04222 | -0.05608 | 0.006958 | 0.008162 |
| Height | rs1907067 | rs1269195 | C | T | 6 | MIR588 | Male only | True | 0.4984 | 0.4984 | 0.4932 | 0.4904 | 34577 | 25698 | 193785 | 166603 | 0.00787 | 0.04172 | 0.04225 | 0.05624 | 0.006958 | 0.008162 |
| Height | rs1907067 | rs1262543 | G | A | 6 | MIR588 | Male only | True | 0.4989 | 0.4991 | 0.4933 | 0.4906 | 34577 | 25698 | 193785 | 166603 | 0.007632 | 0.04155 | 0.04227 | 0.05618 | 0.006957 | 0.008165 |
| Height | rs1907067 | rs853965 | C | T | 6 | MIR588 | Male only | True | 0.4983 | 0.4984 | 0.4931 | 0.4904 | 34577 | 25698 | 193785 | 166603 | 0.007881 | 0.04178 | 0.04223 | 0.05627 | 0.006958 | 0.008162 |
| Height | rs1907067 | rs853966 | G | T | 6 | MIR588 | Male only | True | 0.4615 | 0.4606 | 0.4634 | 0.4657 | 34577 | 25698 | 193785 | 166603 | -0.005202 | -0.03446 | -0.03784 | -0.05195 | 0.006979 | 0.008171 |
| Height | rs1907067 | rs853981 | T | C | 6 | MIR588 | Male only | True | 0.4967 | 0.497 | 0.4963 | 0.4938 | 34577 | 25698 | 193785 | 166603 | -0.005146 | -0.0386 | -0.04054 | -0.05431 | 0.006955 | 0.00815 |
| Height | rs1907067 | rs853982 | T | C | 6 | MIR588 | Male only | True | 0.4955 | 0.4956 | 0.4967 | 0.494 | 34577 | 25698 | 193785 | 166603 | -0.004608 | -0.03742 | -0.04052 | -0.05414 | 0.006955 | 0.00815 |
| Height | rs1907067 | rs853983 | A | G | 6 | MIR588 | Male only | True | 0.4967 | 0.4971 | 0.4963 | 0.4938 | 34577 | 25698 | 193785 | 166603 | -0.005156 | -0.03845 | -0.04054 | -0.05432 | 0.006955 | 0.00815 |
| Height | rs1907067 | rs1101558 | A | G | 6 | MIR588 | Male only | True | 0.4734 | 0.4723 | 0.4817 | 0.484 | 34577 | 25698 | 193785 | 166603 | -0.005036 | -0.03698 | -0.03817 | -0.05089 | 0.006997 | 0.008184 |
| Height | rs1907067 | rs1101559 | T | C | 6 | MIR588 | Male only | True | 0.4733 | 0.4723 | 0.4816 | 0.4839 | 34577 | 25698 | 193785 | 166603 | -0.004994 | -0.03701 | -0.03817 | -0.05088 | 0.006998 | 0.008184 |
| Height | rs1907067 | rs1101561 | A | G | 6 | MIR588 | Male only | True | 0.494 | 0.494 | 0.4967 | 0.494 | 34577 | 25698 | 193785 | 166603 | -0.004765 | -0.03759 | -0.04052 | -0.05415 | 0.006976 | 0.008176 |
| Height | rs1907067 | rs1101563 | C | T | 6 | MIR588 | Male only | True | 0.4948 | 0.4952 | 0.4963 | 0.4938 | 34577 | 25698 | 193785 | 166603 | -0.005313 | -0.03828 | -0.04052 | -0.0543 | 0.006979 | 0.008177 |
| Height | rs1907067 | rs1313697 | A | G | 6 | MIR588 | Male only | True | 0.495 | 0.4955 | 0.4963 | 0.4938 | 34577 | 25698 | 193785 | 166603 | -0.005378 | -0.03819 | -0.04054 | -0.05433 | 0.006978 | 0.008176 |
| Height | rs1907067 | rs1262555 | A | G | 6 | MIR588 | Male only | True | 0.4938 | 0.4938 | 0.4967 | 0.4941 | 34577 | 25698 | 193785 | 166603 | -0.004976 | -0.03739 | -0.0405 | -0.05417 | 0.006978 | 0.008177 |
| Height | rs1907067 | rs1262557 | T | C | 6 | MIR588 | Male only | True | 0.4754 | 0.4746 | 0.4817 | 0.4841 | 34577 | 25698 | 193785 | 166603 | -0.005 | -0.03645 | -0.03807 | -0.05095 | 0.006976 | 0.008155 |
| Height | rs1907067 | rs1022573 | A | C | 6 | MIR588 | Male only | True | 0.495 | 0.4954 | 0.4963 | 0.4938 | 34577 | 25698 | 193785 | 166603 | -0.005253 | -0.03812 | -0.04053 | -0.05433 | 0.006977 | 0.008175 |
| Height | rs1907067 | rs853976 | T | C | 6 | MIR588 | Male only | True | 0.4734 | 0.4723 | 0.4817 | 0.484 | 34577 | 25698 | 193785 | 166603 | -0.005206 | -0.03647 | -0.0381 | -0.05088 | 0.006999 | 0.008184 |
| Height | rs1907067 | rs853977 | C | T | 6 | MIR588 | Male only | True | 0.4951 | 0.4954 | 0.4963 | 0.4937 | 34577 | 25698 | 193785 | 166603 | -0.005563 | -0.0382 | -0.0405 | -0.05431 | 0.006978 | 0.008176 |
| Height | rs1907067 | rs853969 | G | A | 6 | MIR588 | Male only | True | 0.4957 | 0.4956 | 0.4966 | 0.494 | 34577 | 25698 | 193785 | 166603 | -0.005085 | -0.037 | -0.04044 | -0.05414 | 0.006954 | 0.008149 |
| Height | rs1907067 | rs853970 | G | A | 6 | MIR588 | Male only | True | 0.4957 | 0.4956 | 0.4965 | 0.4939 | 34577 | 25698 | 193785 | 166603 | -0.005288 | -0.0372 | -0.04038 | -0.05411 | 0.006956 | 0.00815 |
| Height | rs1907067 | rs853971 | C | T | 6 | MIR588 | Male only | True | 0.4969 | 0.4971 | 0.4962 | 0.4936 | 34577 | 25698 | 193785 | 166603 | -0.005711 | -0.03818 | -0.0404 | -0.05427 | 0.006955 | 0.00815 |
| Height | rs1907067 | rs1101556 | G | T | 6 | MIR588 | Male only | True | 0.4941 | 0.4939 | 0.4965 | 0.4939 | 34577 | 25698 | 193785 | 166603 | -0.00522 | -0.03727 | -0.04037 | -0.05411 | 0.006977 | 0.008175 |
| Height | rs1907067 | rs1101557 | C | T | 6 | MIR588 | Male only | True | 0.4952 | 0.4954 | 0.4962 | 0.4936 | 34577 | 25698 | 193785 | 166603 | -0.005687 | -0.03829 | -0.0404 | -0.05428 | 0.006978 | 0.008176 |
| Height | rs1907067 | rs853973 | G | A | 6 | MIR588 | Male only | True | 0.4942 | 0.494 | 0.4965 | 0.4939 | 34577 | 25698 | 193785 | 166603 | -0.005118 | -0.03718 | -0.04037 | -0.0541 | 0.006978 | 0.008176 |
| Height | rs1907067 | rs853975 | T | C | 6 | MIR588 | Male only | True | 0.4951 | 0.4954 | 0.4962 | 0.4937 | 34577 | 25698 | 193785 | 166603 | -0.005612 | -0.03823 | -0.04038 | -0.05427 | 0.006979 | 0.008176 |

| trait | lead_snp | ID | ALLELE1 | ALLELE0 | CHR | Gene_Symbol | independent_loci_in_sex_P1e2 | in_embl_any_or_gene | A1FREQ_VUMC_female | A1FREQ_VUMC_male | A1FREQ_UKB_female | A1FREQ_UKB_male | N_VUMC_female | N_VUMC_male | N_UKB_female | N_UKB_male | BETA_VUMC_female | BETA_VUMC_male | BETA_UKB_female | BETA_UKB_male | SE_VUMC_female | SE_VUMC_male |
|---|---|---|---|---|---|---|---|---|---|---|---|---|---|---|---|---|---|---|---|---|---|---|
| Height | rs1907067 | rs853968 | T | C | 6 | MIR588 | Male only | True | 0.4951 | 0.4953 | 0.4962 | 0.4937 | 34577 | 25698 | 193785 | 166603 | -0.0056 | -0.03824 | -0.04038 | -0.05427 | 0.006979 | 0.008176 |
| Height | rs1907067 | rs861237 | T | C | 6 | MIR588 | Male only | True | 0.4939 | 0.4938 | 0.4966 | 0.4939 | 34577 | 25698 | 193785 | 166603 | -0.004854 | -0.03693 | -0.04036 | -0.0541 | 0.006979 | 0.008177 |
| Height | rs1907067 | rs1262545 | T | C | 6 | MIR588 | Male only | True | 0.4951 | 0.4953 | 0.4962 | 0.4937 | 34577 | 25698 | 193785 | 166603 | -0.005641 | -0.03826 | -0.04039 | -0.05427 | 0.006979 | 0.008176 |
| Height | rs1907067 | rs1262546 | C | T | 6 | MIR588 | Male only | True | 0.4948 | 0.495 | 0.4962 | 0.4937 | 34577 | 25698 | 193785 | 166603 | -0.005614 | -0.03842 | -0.04039 | -0.05426 | 0.007002 | 0.008197 |
| Height | rs1907067 | rs979197 | T | C | 6 | MIR588 | Male only | True | 0.4936 | 0.4935 | 0.4965 | 0.4939 | 34577 | 25698 | 193785 | 166603 | -0.005077 | -0.03739 | -0.04038 | -0.0541 | 0.007002 | 0.008197 |
| Height | rs1907067 | rs1262550 | C | A | 6 | MIR588 | Male only | True | 0.4937 | 0.4935 | 0.4966 | 0.494 | 34577 | 25698 | 193785 | 166603 | -0.005095 | -0.0374 | -0.04043 | -0.05409 | 0.007002 | 0.008198 |
| Height | rs1907067 | rs1262551 | T | C | 6 | MIR588 | Male only | True | 0.4936 | 0.4935 | 0.4966 | 0.494 | 34577 | 25698 | 193785 | 166603 | -0.005143 | -0.03738 | -0.04042 | -0.0541 | 0.007002 | 0.008198 |
| Height | rs1907067 | rs1262552 | T | C | 6 | MIR588 | Male only | True | 0.4935 | 0.4934 | 0.4966 | 0.4939 | 34577 | 25698 | 193785 | 166603 | -0.005278 | -0.03762 | -0.04037 | -0.0541 | 0.007002 | 0.008198 |
| Height | rs1907067 | rs1262553 | C | T | 6 | MIR588 | Male only | True | 0.4948 | 0.495 | 0.4962 | 0.4937 | 34577 | 25698 | 193785 | 166603 | -0.005725 | -0.03845 | -0.04039 | -0.05426 | 0.007002 | 0.008197 |
| Height | rs1907067 | rs1015446 | A | G | 6 | MIR588 | Male only | True | 0.4942 | 0.4942 | 0.4965 | 0.4938 | 34577 | 25698 | 193785 | 166603 | -0.005014 | -0.03741 | -0.04016 | -0.05383 | 0.007003 | 0.008195 |
| Height | rs62466068 | rs62466068 | C | T | 7 | CDK6 | Female only | True | 0.1654 | 0.1647 | 0.1704 | 0.1693 | 34577 | 25698 | 193785 | 166603 | -0.05399 | -0.01821 | -0.05223 | -0.04789 | 0.00942 | 0.01101 |
| Height | rs62466068 | rs3731321 | C | T | 7 | CDK6 | Female only | True | 0.1656 | 0.1648 | 0.171 | 0.1701 | 34577 | 25698 | 193785 | 166603 | -0.05185 | -0.01605 | -0.05156 | -0.04811 | 0.009485 | 0.01109 |
| Height | rs16869 | rs11854876 | C | T | 15 | PML | Female only | True | 0.1705 | 0.1733 | 0.1783 | 0.1787 | 34577 | 25698 | 193785 | 166603 | -0.04757 | -0.005916 | -0.03883 | -0.03526 | 0.009418 | 0.01077 |
| Height | rs16869 | rs10152126 | T | C | 15 | PML | Female only | True | 0.1705 | 0.1733 | 0.1782 | 0.1786 | 34577 | 25698 | 193785 | 166603 | -0.04733 | -0.005987 | -0.03882 | -0.03532 | 0.009418 | 0.01077 |
| Height | rs16869 | rs11630778 | T | C | 15 | PML | Female only | True | 0.1707 | 0.1734 | 0.1783 | 0.1787 | 34577 | 25698 | 193785 | 166603 | -0.04704 | -0.006176 | -0.03898 | -0.0352 | 0.00937 | 0.01071 |
| Height | rs16869 | rs12050565 | G | A | 15 | PML | Female only | True | 0.171 | 0.1737 | 0.1785 | 0.1789 | 34577 | 25698 | 193785 | 166603 | -0.04795 | -0.00696 | -0.03881 | -0.03558 | 0.009362 | 0.01071 |
| Height | rs16869 | rs10152898 | T | G | 15 | PML | Female only | True | 0.1716 | 0.174 | 0.1781 | 0.1784 | 34577 | 25698 | 193785 | 166603 | -0.04706 | -0.006052 | -0.03941 | -0.03497 | 0.009316 | 0.01067 |
| Height | rs16869 | rs10152975 | C | T | 15 | PML | Female only | True | 0.1721 | 0.1747 | 0.1797 | 0.1797 | 34577 | 25698 | 193785 | 166603 | -0.04807 | -0.006567 | -0.03868 | -0.03535 | 0.009333 | 0.01068 |
| Height | rs16869 | rs11072456 | G | T | 15 | PML | Female only | True | 0.1722 | 0.1747 | 0.1797 | 0.1798 | 34577 | 25698 | 193785 | 166603 | -0.04802 | -0.006791 | -0.03896 | -0.03537 | 0.009336 | 0.01068 |
| Height | rs16869 | rs11072457 | C | T | 15 | PML | Female only | True | 0.1732 | 0.1755 | 0.18 | 0.1801 | 34577 | 25698 | 193785 | 166603 | -0.0489 | -0.00703 | -0.03892 | -0.0353 | 0.009307 | 0.01066 |
| Height | rs16869 | rs11636435 | T | G | 15 | PML | Female only | True | 0.1737 | 0.1763 | 0.1814 | 0.1814 | 34577 | 25698 | 193785 | 166603 | -0.04886 | -0.009041 | -0.03878 | -0.03448 | 0.009303 | 0.01064 |
| Height | rs16869 | rs34080912 | T | C | 15 | PML | Female only | True | 0.1732 | 0.176 | 0.1811 | 0.1811 | 34577 | 25698 | 193785 | 166603 | -0.04866 | -0.008469 | -0.03869 | -0.03455 | 0.009315 | 0.01065 |
| Height | rs16869 | rs28405144 | T | G | 15 | PML | Female only | True | 0.1735 | 0.1761 | 0.1814 | 0.1815 | 34577 | 25698 | 193785 | 166603 | -0.04782 | -0.00878 | -0.03891 | -0.03472 | 0.009308 | 0.01065 |
| Height | rs16869 | rs12898704 | T | C | 15 | PML | Female only | True | 0.1731 | 0.1759 | 0.1812 | 0.1812 | 34577 | 25698 | 193785 | 166603 | -0.04769 | -0.00869 | -0.03896 | -0.03474 | 0.009316 | 0.01065 |
| Height | rs16869 | rs7182319 | A | G | 15 | PML | Female only | True | 0.2407 | 0.2425 | 0.2439 | 0.2439 | 34577 | 25698 | 193785 | 166603 | -0.04308 | -0.0104 | -0.02947 | -0.02625 | 0.008207 | 0.009434 |
| Height | rs16869 | rs12907194 | T | C | 15 | PML | Female only | True | 0.1705 | 0.1734 | 0.1785 | 0.1786 | 34577 | 25698 | 193785 | 166603 | -0.049 | -0.007805 | -0.03984 | -0.03522 | 0.00938 | 0.01071 |
| Height | rs16869 | rs12907609 | A | G | 15 | PML | Female only | True | 0.1705 | 0.1734 | 0.1785 | 0.1786 | 34577 | 25698 | 193785 | 166603 | -0.049 | -0.007805 | -0.03984 | -0.03522 | 0.00938 | 0.01071 |
| Height | rs16869 | rs8039184 | A | G | 15 | PML | Female only | True | 0.2676 | 0.2682 | 0.2683 | 0.2685 | 34577 | 25698 | 193785 | 166603 | -0.04174 | -0.01269 | -0.02758 | -0.02171 | 0.007927 | 0.009124 |
| Height | rs16869 | rs7174985 | G | T | 15 | PML | Female only | True | 0.3306 | 0.3317 | 0.3317 | 0.3316 | 34577 | 25698 | 193785 | 166603 | -0.03843 | -0.01622 | -0.02675 | -0.02203 | 0.007451 | 0.008588 |

| trait | lead_snp | ID | ALLELE1 | ALLELE0 | CHR | Gene_Symbol | independent_loci_in_sex_P1e2 | in_embl_any_or_gene | A1FREQ_VUMC_female | A1FREQ_VUMC_male | A1FREQ_UKB_female | A1FREQ_UKB_male | N_VUMC_female | N_VUMC_male | N_UKB_female | N_UKB_male | BETA_VUMC_female | BETA_VUMC_male | BETA_UKB_female | BETA_UKB_male | SE_VUMC_female | SE_VUMC_male |
|---|---|---|---|---|---|---|---|---|---|---|---|---|---|---|---|---|---|---|---|---|---|---|
| Height | rs16869 | rs11636934 | G | A | 15 | PML | Female only | True | 0.2699 | 0.271 | 0.2708 | 0.2712 | 34577 | 25698 | 193785 | 166603 | -0.04258 | -0.01412 | -0.02842 | -0.02227 | 0.007889 | 0.009089 |
| Height | rs16869 | rs2507 | G | A | 15 | PML | Female only | True | 0.2709 | 0.272 | 0.2707 | 0.2711 | 34577 | 25698 | 193785 | 166603 | -0.04204 | -0.01431 | -0.02849 | -0.0224 | 0.007854 | 0.009049 |
| Height | rs16869 | rs9630436 | C | T | 15 | PML | Female only | True | 0.3014 | 0.3035 | 0.3041 | 0.3039 | 34577 | 25698 | 193785 | 166603 | -0.04023 | -0.01403 | -0.02855 | -0.02547 | 0.007644 | 0.008794 |
| Height | rs16869 | rs9630437 | C | T | 15 | PML | Female only | True | 0.1708 | 0.1739 | 0.1785 | 0.1787 | 34577 | 25698 | 193785 | 166603 | -0.0495 | -0.01088 | -0.04053 | -0.03569 | 0.009367 | 0.01069 |
| Height | rs16869 | rs11072460 | G | T | 15 | PML | Female only | True | 0.3011 | 0.3031 | 0.3038 | 0.3034 | 34577 | 25698 | 193785 | 166603 | -0.04043 | -0.01496 | -0.02847 | -0.02551 | 0.007637 | 0.008791 |
| Height | rs16869 | rs4993000 | T | C | 15 | PML | Female only | True | 0.3295 | 0.3304 | 0.3304 | 0.3301 | 34577 | 25698 | 193785 | 166603 | -0.03849 | -0.01683 | -0.02684 | -0.02186 | 0.007446 | 0.008581 |
| Height | rs16869 | rs3816197 | T | C | 15 | PML | Female only | True | 0.3016 | 0.3037 | 0.3035 | 0.3031 | 34577 | 25698 | 193785 | 166603 | -0.03931 | -0.01452 | -0.0284 | -0.02559 | 0.007611 | 0.008758 |
| Height | rs16869 | rs11072468 | C | T | 15 | PML | Female only | True | 0.243 | 0.244 | 0.2462 | 0.2464 | 34577 | 25698 | 193785 | 166603 | -0.04687 | -0.01619 | -0.02989 | -0.02667 | 0.008201 | 0.009465 |
| Height | rs16869 | rs11637420 | C | A | 15 | PML | Female only | True | 0.1761 | 0.1788 | 0.184 | 0.1842 | 34577 | 25698 | 193785 | 166603 | -0.05171 | -0.01554 | -0.03962 | -0.03458 | 0.009283 | 0.01061 |
| Height | rs16869 | rs16869 | C | T | 15 | PML | Female only | True | 0.243 | 0.244 | 0.2463 | 0.2465 | 34577 | 25698 | 193785 | 166603 | -0.04696 | -0.01635 | -0.03007 | -0.02667 | 0.008207 | 0.009471 |
| Height | rs16869 | rs12901885 | T | C | 15 | PML | Female only | True | 0.1762 | 0.1788 | 0.184 | 0.1842 | 34577 | 25698 | 193785 | 166603 | -0.05158 | -0.01579 | -0.03961 | -0.0346 | 0.009279 | 0.01061 |
| Height | rs16869 | rs11858669 | A | G | 15 | PML | Female only | True | 0.1762 | 0.1789 | 0.1842 | 0.1845 | 34577 | 25698 | 193785 | 166603 | -0.05162 | -0.01592 | -0.03971 | -0.03464 | 0.009278 | 0.01061 |
| Height | rs12148139 | rs12148209 | T | C | 15 | ADAMTSL3 | Female only | True | 0.07073 | 0.07112 | 0.07818 | 0.07802 | 34577 | 25698 | 193785 | 166603 | 0.06605 | 0.003184 | 0.01026 | 0.02076 | 0.01365 | 0.01594 |
| Height | rs78015020 | rs78015020 | C | T | 15 | ADAMTSL3 | Female only | True | 0.08636 | 0.08349 | 0.08535 | 0.08573 | 34577 | 25698 | 193785 | 166603 | -0.07559 | -0.01593 | -0.03282 | -0.03893 | 0.01263 | 0.01498 |
| Height | rs12148139 | rs55726315 | G | A | 15 | ADAMTSL3 | Female only | True | 0.1206 | 0.1197 | 0.1298 | 0.1286 | 34577 | 25698 | 193785 | 166603 | 0.05892 | 0.007371 | 0.01081 | 0.02307 | 0.01067 | 0.01254 |
| Height | rs12148139 | rs7164317 | C | T | 15 | ADAMTSL3 | Female only | True | 0.1223 | 0.1215 | 0.1308 | 0.1296 | 34577 | 25698 | 193785 | 166603 | 0.05692 | 0.006488 | 0.01096 | 0.02263 | 0.01061 | 0.01246 |
| Height | rs12148139 | rs59144084 | T | G | 15 | ADAMTSL3 | Female only | True | 0.107 | 0.1067 | 0.1158 | 0.1148 | 34577 | 25698 | 193785 | 166603 | 0.05817 | 0.01439 | 0.008223 | 0.02289 | 0.01126 | 0.0132 |
| Height | rs12148139 | rs12591049 | G | T | 15 | ADAMTSL3 | Female only | True | 0.1231 | 0.1227 | 0.1315 | 0.1305 | 34577 | 25698 | 193785 | 166603 | 0.05759 | 0.006732 | 0.01115 | 0.0223 | 0.01057 | 0.0124 |
| Height | rs12148139 | rs12593448 | T | C | 15 | ADAMTSL3 | Female only | True | 0.1221 | 0.1215 | 0.1306 | 0.1295 | 34577 | 25698 | 193785 | 166603 | 0.05596 | 0.006675 | 0.01107 | 0.0218 | 0.0106 | 0.01244 |
| Height | rs12148139 | rs72761818 | A | G | 15 | ADAMTSL3 | Female only | True | 0.1199 | 0.119 | 0.1297 | 0.1286 | 34577 | 25698 | 193785 | 166603 | 0.05718 | 0.007604 | 0.0111 | 0.02285 | 0.01068 | 0.01256 |
| Height | rs12148139 | rs72761820 | T | C | 15 | ADAMTSL3 | Female only | True | 0.1203 | 0.1194 | 0.1298 | 0.1288 | 34577 | 25698 | 193785 | 166603 | 0.05775 | 0.007282 | 0.01075 | 0.02284 | 0.01067 | 0.01255 |
| Height | rs12148139 | rs72761824 | A | G | 15 | ADAMTSL3 | Female only | True | 0.1196 | 0.1186 | 0.1293 | 0.1282 | 34577 | 25698 | 193785 | 166603 | 0.0577 | 0.008092 | 0.01111 | 0.0228 | 0.0107 | 0.01258 |
| Height | rs12148139 | rs60619082 | A | C | 15 | ADAMTSL3 | Female only | True | 0.1225 | 0.1219 | 0.131 | 0.1299 | 34577 | 25698 | 193785 | 166603 | 0.05729 | 0.007024 | 0.01119 | 0.02267 | 0.01059 | 0.01242 |
| Height | rs12148139 | rs10152418 | A | G | 15 | ADAMTSL3 | Female only | True | 0.1221 | 0.1215 | 0.1307 | 0.1296 | 34577 | 25698 | 193785 | 166603 | 0.05641 | 0.006104 | 0.01123 | 0.02296 | 0.01059 | 0.01242 |
| Height | rs12148139 | rs8026130 | G | A | 15 | ADAMTSL3 | Female only | True | 0.123 | 0.1225 | 0.1312 | 0.1301 | 34577 | 25698 | 193785 | 166603 | 0.0576 | 0.007234 | 0.01116 | 0.02312 | 0.01057 | 0.0124 |
| trait | lead_snp | ID | ALLELE1 | ALLELE0 | CHR | Gene_Symbol | independent_loci_in_sex_P1e2 | in_embl_any_or_gene | A1FREQ_VUMC_female | A1FREQ_VUMC_male | A1FREQ_UKB_female | A1FREQ_UKB_male | N_VUMC_female | N_VUMC_male | N_UKB_female | N_UKB_male | BETA_VUMC_female | BETA_VUMC_male | BETA_UKB_female | BETA_UKB_male | SE_VUMC_female | SE_VUMC_male |
| Height | rs12148139 | rs12595343 | G | A | 15 | ADAMTSL3 | Female only | True | 0.1199 | 0.1191 | 0.1294 | 0.1283 | 34577 | 25698 | 193785 | 166603 | 0.05788 | 0.008582 | 0.01112 | 0.023 | 0.01069 | 0.01255 |
| Height | rs12148139 | rs72761831 | G | T | 15 | ADAMTSL3 | Female only | True | 0.1044 | 0.1045 | 0.1137 | 0.1128 | 34577 | 25698 | 193785 | 166603 | 0.0622 | 0.01553 | 0.007876 | 0.02252 | 0.01137 | 0.01331 |

| trait | lead_snp | ID | ALLELE1 | ALLELE0 | CHR | Gene_Symbol | independent_loci_in_sex_P1e2 | in_embl_any_or_gene | A1FREQ_VUMC_female | A1FREQ_VUMC_male | A1FREQ_UKB_female | A1FREQ_UKB_male | N_VUMC_female | N_VUMC_male | N_UKB_female | N_UKB_male | BETA_VUMC_female | BETA_VUMC_male | BETA_UKB_female | BETA_UKB_male | SE_VUMC_female | SE_VUMC_male |
|---|---|---|---|---|---|---|---|---|---|---|---|---|---|---|---|---|---|---|---|---|---|---|
| Height | rs12148139 | rs1834524 | G | A | 15 | ADAMTSL3 | Female only | True | 0.1203 | 0.1197 | 0.1298 | 0.1288 | 34577 | 25698 | 193785 | 166603 | 0.05868 | 0.007909 | 0.01096 | 0.02337 | 0.01067 | 0.01254 |
| Height | rs12148139 | rs17158691 | T | C | 15 | ADAMTSL3 | Female only | True | 0.1196 | 0.1186 | 0.1294 | 0.1284 | 34577 | 25698 | 193785 | 166603 | 0.05718 | 0.007383 | 0.0112 | 0.02283 | 0.01072 | 0.0126 |
| Height | rs12148139 | rs17158688 | A | G | 15 | ADAMTSL3 | Female only | True | 0.1202 | 0.1194 | 0.1298 | 0.1287 | 34577 | 25698 | 193785 | 166603 | 0.05833 | 0.008487 | 0.01294 | 0.02287 | 0.01071 | 0.01258 |
| Height | rs12148139 | rs28548261 | A | G | 15 | ADAMTSL3 | Female only | True | 0.1225 | 0.1221 | 0.1312 | 0.1301 | 34577 | 25698 | 193785 | 166603 | 0.05757 | 0.006714 | 0.01161 | 0.02339 | 0.01062 | 0.01245 |
| Height | rs12148139 | rs28478148 | G | A | 15 | ADAMTSL3 | Female only | True | 0.1222 | 0.1219 | 0.1311 | 0.1301 | 34577 | 25698 | 193785 | 166603 | 0.05787 | 0.006716 | 0.01165 | 0.02331 | 0.01063 | 0.01246 |
| Height | rs12148139 | rs9788762 | C | T | 15 | ADAMTSL3 | Female only | True | 0.1095 | 0.108 | 0.1172 | 0.1161 | 34577 | 25698 | 193785 | 166603 | 0.06568 | 0.006125 | 0.01291 | 0.02637 | 0.01117 | 0.01313 |
| Height | rs12148139 | rs7168677 | A | G | 15 | ADAMTSL3 | Female only | True | 0.1093 | 0.1077 | 0.1169 | 0.1158 | 34577 | 25698 | 193785 | 166603 | 0.06587 | 0.007038 | 0.01312 | 0.0264 | 0.01115 | 0.01311 |
| Height | rs12148139 | rs57372822 | C | T | 15 | ADAMTSL3 | Female only | True | 0.1157 | 0.115 | 0.1244 | 0.1233 | 34577 | 25698 | 193785 | 166603 | 0.06389 | 0.01156 | 0.01079 | 0.02444 | 0.01087 | 0.01277 |
| Height | rs12148139 | rs1896815 | C | A | 15 | ADAMTSL3 | Female only | True | 0.116 | 0.1153 | 0.124 | 0.1229 | 34577 | 25698 | 193785 | 166603 | 0.06397 | 0.01093 | 0.01074 | 0.02407 | 0.01086 | 0.01275 |
| Height | rs12148139 | rs1896814 | G | A | 15 | ADAMTSL3 | Female only | True | 0.1156 | 0.1148 | 0.124 | 0.1228 | 34577 | 25698 | 193785 | 166603 | 0.06392 | 0.01086 | 0.01064 | 0.02408 | 0.01088 | 0.01278 |
| Height | rs12148139 | rs61260043 | G | A | 15 | ADAMTSL3 | Female only | True | 0.1158 | 0.115 | 0.1244 | 0.1233 | 34577 | 25698 | 193785 | 166603 | 0.0636 | 0.01133 | 0.01093 | 0.02452 | 0.01087 | 0.01277 |
| Height | rs12148139 | rs72746903 | G | A | 15 | ADAMTSL3 | Female only | True | 0.1159 | 0.115 | 0.1242 | 0.1231 | 34577 | 25698 | 193785 | 166603 | 0.06313 | 0.01107 | 0.01102 | 0.0245 | 0.01087 | 0.01277 |
| Height | rs12148139 | rs17158659 | A | G | 15 | ADAMTSL3 | Female only | True | 0.1098 | 0.1078 | 0.1172 | 0.116 | 34577 | 25698 | 193785 | 166603 | 0.06436 | 0.006555 | 0.01313 | 0.02644 | 0.01112 | 0.0131 |
| Height | rs12148139 | rs59195990 | T | C | 15 | ADAMTSL3 | Female only | True | 0.1087 | 0.1069 | 0.1168 | 0.1156 | 34577 | 25698 | 193785 | 166603 | 0.06577 | 0.005607 | 0.01317 | 0.02659 | 0.01117 | 0.01315 |
| Height | rs12148139 | rs12148139 | C | A | 15 | ADAMTSL3 | Female only | True | 0.1037 | 0.1032 | 0.1119 | 0.111 | 34577 | 25698 | 193785 | 166603 | 0.06875 | 0.01594 | 0.007294 | 0.02359 | 0.01142 | 0.01341 |
| Height | rs12148139 | rs17158657 | A | G | 15 | ADAMTSL3 | Female only | True | 0.1102 | 0.1082 | 0.1172 | 0.116 | 34577 | 25698 | 193785 | 166603 | 0.06465 | 0.006559 | 0.01318 | 0.02636 | 0.0111 | 0.01308 |
| Height | rs12148139 | rs72746905 | A | G | 15 | ADAMTSL3 | Female only | True | 0.1088 | 0.107 | 0.1172 | 0.116 | 34577 | 25698 | 193785 | 166603 | 0.06554 | 0.00632 | 0.01335 | 0.02628 | 0.01117 | 0.01315 |
| Height | rs12148139 | rs12593547 | T | G | 15 | ADAMTSL3 | Female only | True | 0.1099 | 0.1079 | 0.1173 | 0.1161 | 34577 | 25698 | 193785 | 166603 | 0.06421 | 0.0061 | 0.01304 | 0.02649 | 0.01112 | 0.0131 |
| Height | rs12148139 | rs58956574 | A | G | 15 | ADAMTSL3 | Female only | True | 0.1087 | 0.1069 | 0.1168 | 0.1156 | 34577 | 25698 | 193785 | 166603 | 0.06568 | 0.005895 | 0.01304 | 0.02666 | 0.01117 | 0.01314 |
| Height | rs12148139 | rs57921785 | T | C | 15 | ADAMTSL3 | Female only | True | 0.1103 | 0.1084 | 0.1174 | 0.1162 | 34577 | 25698 | 193785 | 166603 | 0.06506 | 0.007283 | 0.01299 | 0.02623 | 0.01108 | 0.01306 |
| Height | rs12148139 | rs56091379 | T | C | 15 | ADAMTSL3 | Female only | True | 0.1103 | 0.1083 | 0.1175 | 0.1163 | 34577 | 25698 | 193785 | 166603 | 0.06437 | 0.007262 | 0.01312 | 0.02603 | 0.01109 | 0.01308 |
| Height | rs78015020 | rs113261930 | T | C | 15 | ADAMTSL3 | Female only | True | 0.08947 | 0.08591 | 0.08848 | 0.08921 | 34577 | 25698 | 193785 | 166603 | -0.06496 | -0.01394 | -0.03814 | -0.04188 | 0.01245 | 0.0148 |
| Height | rs12148139 | rs72746921 | G | T | 15 | ADAMTSL3 | Female only | True | 0.1154 | 0.1144 | 0.1241 | 0.123 | 34577 | 25698 | 193785 | 166603 | 0.06284 | 0.01012 | 0.01101 | 0.02453 | 0.01088 | 0.01279 |
| Height | rs12148139 | rs7495206 | T | G | 15 | ADAMTSL3 | Female only | True | 0.109 | 0.1073 | 0.117 | 0.1158 | 34577 | 25698 | 193785 | 166603 | 0.06611 | 0.006732 | 0.01308 | 0.02649 | 0.01115 | 0.01312 |
| Height | rs12148139 | rs72746925 | T | C | 15 | ADAMTSL3 | Female only | True | 0.1086 | 0.1068 | 0.1161 | 0.115 | 34577 | 25698 | 193785 | 166603 | 0.065 | 0.005377 | 0.01364 | 0.02658 | 0.01119 | 0.01316 |
| Height | rs78015020 | rs62025778 | A | G | 15 | ADAMTSL3 | Female only | True | 0.07757 | 0.07414 | 0.07614 | 0.07672 | 34577 | 25698 | 193785 | 166603 | -0.06852 | -0.01716 | -0.03756 | -0.03736 | 0.01302 | 0.01553 |
| Height | rs12148139 | rs56176147 | A | G | 15 | ADAMTSL3 | Female only | True | 0.109 | 0.1072 | 0.1167 | 0.1156 | 34577 | 25698 | 193785 | 166603 | 0.06555 | 0.006302 | 0.01381 | 0.02635 | 0.01117 | 0.01314 |
| Height | rs12148139 | rs55776473 | T | C | 15 | ADAMTSL3 | Female only | True | 0.1155 | 0.1144 | 0.1239 | 0.1228 | 34577 | 25698 | 193785 | 166603 | 0.06349 | 0.009128 | 0.01158 | 0.02458 | 0.01088 | 0.01279 |
| Height | rs12148139 | rs72746936 | G | A | 15 | ADAMTSL3 | Female only | True | 0.1088 | 0.107 | 0.1165 | 0.1153 | 34577 | 25698 | 193785 | 166603 | 0.0654 | 0.005271 | 0.01367 | 0.02666 | 0.01119 | 0.01316 |

| trait | lead_snp | ID | ALLELE1 | ALLELE0 | CHR | Gene_Symbol | independent_loci_in_sex_P1e2 | in_embl_any_or_gene | A1FREQ_VUMC_female | A1FREQ_VUMC_male | A1FREQ_UKB_female | A1FREQ_UKB_male | N_VUMC_female | N_VUMC_male | N_UKB_female | N_UKB_male | BETA_VUMC_female | BETA_VUMC_male | BETA_UKB_female | BETA_UKB_male | SE_VUMC_female | SE_VUMC_male |
|---|---|---|---|---|---|---|---|---|---|---|---|---|---|---|---|---|---|---|---|---|---|---|
| Height | rs12148139 | rs72746937 | T | C | 15 | ADAMTSL3 | Female only | True | 0.1088 | 0.107 | 0.1165 | 0.1153 | 34577 | 25698 | 193785 | 166603 | 0.06538 | 0.005286 | 0.01366 | 0.02665 | 0.01119 | 0.01316 |
| Height | rs12148139 | rs72746938 | G | T | 15 | ADAMTSL3 | Female only | True | 0.1154 | 0.1144 | 0.1238 | 0.1227 | 34577 | 25698 | 193785 | 166603 | 0.0628 | 0.009511 | 0.01141 | 0.02464 | 0.01089 | 0.0128 |
| Height | rs12148139 | rs58836704 | A | G | 15 | ADAMTSL3 | Female only | True | 0.1085 | 0.1068 | 0.1164 | 0.1152 | 34577 | 25698 | 193785 | 166603 | 0.06416 | 0.00529 | 0.01345 | 0.02673 | 0.0112 | 0.01317 |
| Height | rs12148139 | rs7162500 | A | G | 15 | ADAMTSL3 | Female only | True | 0.1087 | 0.1068 | 0.1163 | 0.1151 | 34577 | 25698 | 193785 | 166603 | 0.06363 | 0.004128 | 0.01365 | 0.02672 | 0.01122 | 0.01319 |
| Height | rs12148139 | rs72746948 | G | A | 15 | ADAMTSL3 | Female only | True | 0.1085 | 0.1066 | 0.1163 | 0.1151 | 34577 | 25698 | 193785 | 166603 | 0.06397 | 0.004472 | 0.01353 | 0.02683 | 0.01123 | 0.0132 |
| Height | rs12148139 | rs57092166 | A | G | 15 | ADAMTSL3 | Female only | True | 0.1084 | 0.1066 | 0.1162 | 0.115 | 34577 | 25698 | 193785 | 166603 | 0.06376 | 0.004586 | 0.01351 | 0.02666 | 0.01123 | 0.0132 |
| Height | rs12148139 | rs61036216 | T | C | 15 | ADAMTSL3 | Female only | True | 0.1085 | 0.1066 | 0.1163 | 0.1151 | 34577 | 25698 | 193785 | 166603 | 0.06392 | 0.00447 | 0.01354 | 0.02683 | 0.01123 | 0.0132 |
| Height | rs12148139 | rs12594012 | A | G | 15 | ADAMTSL3 | Female only | True | 0.1086 | 0.1067 | 0.1163 | 0.1151 | 34577 | 25698 | 193785 | 166603 | 0.0638 | 0.004445 | 0.0136 | 0.02693 | 0.01123 | 0.0132 |
| Height | rs12148139 | rs59199978 | G | A | 15 | ADAMTSL3 | Female only | True | 0.1734 | 0.1739 | 0.1818 | 0.1804 | 34577 | 25698 | 193785 | 166603 | 0.03028 | 0.001949 | 0.007662 | 0.01744 | 0.009211 | 0.01077 |
| Height | rs12148139 | rs72746959 | G | A | 15 | ADAMTSL3 | Female only | True | 0.1154 | 0.1142 | 0.1236 | 0.1226 | 34577 | 25698 | 193785 | 166603 | 0.06026 | 0.00854 | 0.01139 | 0.02523 | 0.01094 | 0.01284 |
| Height | rs12148139 | rs113817743 | A | G | 15 | ADAMTSL3 | Female only | True | 0.1088 | 0.1069 | 0.116 | 0.1149 | 34577 | 25698 | 193785 | 166603 | 0.06198 | 0.003884 | 0.01382 | 0.02709 | 0.01123 | 0.01319 |
| Height | rs4414460 | rs28578450 | A | C | 15 | ADAMTSL3 | Female only | True | 0.2088 | 0.2045 | 0.2094 | 0.2068 | 34577 | 25698 | 193785 | 166603 | 0.03163 | 0.02189 | 0.02736 | 0.02898 | 0.008584 | 0.01006 |
| Height | rs4414460 | rs2086441 | A | G | 15 | ADAMTSL3 | Female only | True | 0.2088 | 0.2046 | 0.2094 | 0.2068 | 34577 | 25698 | 193785 | 166603 | 0.03174 | 0.0223 | 0.02738 | 0.02851 | 0.008583 | 0.01006 |
| Height | rs4414460 | rs28712829 | T | C | 15 | ADAMTSL3 | Female only | True | 0.2084 | 0.2043 | 0.2093 | 0.2067 | 34577 | 25698 | 193785 | 166603 | 0.03224 | 0.02181 | 0.02723 | 0.02863 | 0.00859 | 0.01007 |
| Height | rs4414460 | rs899927 | T | C | 15 | ADAMTSL3 | Female only | True | 0.3208 | 0.3177 | 0.3235 | 0.321 | 34577 | 25698 | 193785 | 166603 | 0.04605 | 0.02038 | 0.03095 | 0.03235 | 0.007481 | 0.008718 |
| Height | rs4414460 | rs4414460 | G | A | 15 | ADAMTSL3 | Female only | True | 0.3211 | 0.3179 | 0.3234 | 0.321 | 34577 | 25698 | 193785 | 166603 | 0.04629 | 0.02028 | 0.03104 | 0.0324 | 0.00748 | 0.008721 |
| Height | rs17370572 | rs62027787 | T | C | 15 | ADAMTSL3 | Female only | True | 0.1578 | 0.1586 | 0.1571 | 0.1573 | 34577 | 25698 | 193785 | 166603 | -0.06343 | -0.02778 | -0.04746 | -0.04002 | 0.009603 | 0.01114 |
| Height | rs17370572 | rs17370572 | T | C | 15 | ADAMTSL3 | Female only | True | 0.1635 | 0.1644 | 0.163 | 0.1632 | 34577 | 25698 | 193785 | 166603 | -0.06342 | -0.02555 | -0.04744 | -0.03921 | 0.009444 | 0.01095 |
| Height | rs17370572 | rs56373916 | C | A | 15 | ADAMTSL3 | Female only | True | 0.267 | 0.269 | 0.2671 | 0.2672 | 34577 | 25698 | 193785 | 166603 | -0.02416 | -0.02146 | -0.03081 | -0.02287 | 0.007871 | 0.009185 |
| Height | rs17370572 | rs11855613 | T | C | 15 | ADAMTSL3 | Female only | True | 0.2667 | 0.2686 | 0.2669 | 0.2671 | 34577 | 25698 | 193785 | 166603 | -0.02431 | -0.02165 | -0.03088 | -0.02278 | 0.007874 | 0.009187 |
| Height | rs17370572 | rs112409013 | G | A | 15 | ADAMTSL3 | Female only | True | 0.2667 | 0.2686 | 0.2667 | 0.2669 | 34577 | 25698 | 193785 | 166603 | -0.02363 | -0.02059 | -0.03109 | -0.02251 | 0.007876 | 0.009186 |
| Height | rs17370572 | rs138532692 | G | A | 15 | ADAMTSL3 | Female only | True | 0.1657 | 0.167 | 0.1652 | 0.1653 | 34577 | 25698 | 193785 | 166603 | -0.05978 | -0.02455 | -0.04896 | -0.03766 | 0.009408 | 0.01089 |
| Height | rs17370572 | rs55781136 | G | A | 15 | ADAMTSL3 | Female only | True | 0.2667 | 0.2687 | 0.2667 | 0.2669 | 34577 | 25698 | 193785 | 166603 | -0.02351 | -0.02058 | -0.03107 | -0.02244 | 0.007876 | 0.009186 |
| Height | rs17370572 | rs12148239 | G | T | 15 | ADAMTSL3 | Female only | True | 0.2735 | 0.2753 | 0.274 | 0.2743 | 34577 | 25698 | 193785 | 166603 | -0.02081 | -0.01792 | -0.03125 | -0.01941 | 0.007811 | 0.009119 |
| RDWSD | rs4849439 | rs1480494 | G | A | 2 | DPP10 | Male only | False | 0.1193 | 0.1197 | 0.111 | 0.1111 | 31506 | 24073 | 188075 | 162398 | -0.01035 | -0.07474 | -0.009376 | -0.006324 | 0.01202 | 0.0138 |
| RDWSD | rs4849439 | rs2421508 | C | T | 2 | DPP10 | Male only | False | 0.1194 | 0.1198 | 0.111 | 0.1112 | 31506 | 24073 | 188075 | 162398 | -0.0102 | -0.07503 | -0.009337 | -0.006402 | 0.01201 | 0.0138 |
| RDWSD | rs4849439 | rs7355679 | A | G | 2 | DPP10 | Male only | False | 0.1146 | 0.115 | 0.1063 | 0.1064 | 31506 | 24073 | 188075 | 162398 | -0.009486 | -0.07026 | -0.008192 | -0.00596 | 0.01226 | 0.01411 |
| RDWSD | rs4849439 | rs13382538 | C | T | 2 | DPP10 | Male only | False | 0.126 | 0.1258 | 0.1166 | 0.1164 | 31506 | 24073 | 188075 | 162398 | -0.008187 | -0.07209 | -0.00775 | -0.007248 | 0.01172 | 0.01352 |
| RDWSD | rs4849439 | rs13425437 | G | T | 2 | DPP10 | Male only | False | 0.06849 | 0.06847 | 0.06019 | 0.06023 | 31506 | 24073 | 188075 | 162398 | 0.01435 | -0.08555 | -0.01553 | -0.002669 | 0.01532 | 0.01781 |

| trait | lead_snp | ID | ALLELE1 | ALLELE0 | CHR | Gene_Symbol | independent_loci_in_sex_P1e2 | in_embl_any_or_gene | A1FREQ_VUMC_female | A1FREQ_VUMC_male | A1FREQ_UKB_female | A1FREQ_UKB_male | N_VUMC_female | N_VUMC_male | N_UKB_female | N_UKB_male | BETA_VUMC_female | BETA_VUMC_male | BETA_UKB_female | BETA_UKB_male | SE_VUMC_female | SE_VUMC_male |
|---|---|---|---|---|---|---|---|---|---|---|---|---|---|---|---|---|---|---|---|---|---|---|
| RDWSD | rs4849439 | rs13431747 | C | T | 2 | DPP10 | Male only | False | 0.126 | 0.1258 | 0.1167 | 0.1165 | 31506 | 24073 | 188075 | 162398 | -0.008152 | -0.07247 | -0.007641 | -0.00737 | 0.01172 | 0.01352 |
| RDWSD | rs4849439 | rs11688638 | A | C | 2 | DPP10 | Male only | False | 0.1189 | 0.1193 | 0.1106 | 0.1107 | 31506 | 24073 | 188075 | 162398 | -0.009838 | -0.07407 | -0.009157 | -0.006486 | 0.01203 | 0.01383 |
| RDWSD | rs4849439 | rs10168227 | G | A | 2 | DPP10 | Male only | False | 0.1259 | 0.1257 | 0.1166 | 0.1164 | 31506 | 24073 | 188075 | 162398 | -0.008278 | -0.07212 | -0.007696 | -0.007325 | 0.01173 | 0.01353 |
| RDWSD | rs4849439 | rs13430949 | T | C | 2 | DPP10 | Male only | False | 0.119 | 0.1193 | 0.1106 | 0.1107 | 31506 | 24073 | 188075 | 162398 | -0.01001 | -0.07405 | -0.009206 | -0.006423 | 0.01203 | 0.01383 |
| RDWSD | rs4849439 | rs10181163 | G | A | 2 | DPP10 | Male only | False | 0.126 | 0.1258 | 0.1167 | 0.1165 | 31506 | 24073 | 188075 | 162398 | -0.008334 | -0.07237 | -0.007753 | -0.00737 | 0.01172 | 0.01352 |
| RDWSD | rs4849439 | rs57315279 | A | G | 2 | DPP10 | Male only | False | 0.1255 | 0.1253 | 0.1162 | 0.1159 | 31506 | 24073 | 188075 | 162398 | -0.008073 | -0.0719 | -0.007666 | -0.007205 | 0.01175 | 0.01354 |
| RDWSD | rs4849439 | rs7588105 | G | T | 2 | DPP10 | Male only | False | 0.126 | 0.1257 | 0.1167 | 0.1165 | 31506 | 24073 | 188075 | 162398 | -0.008239 | -0.07376 | -0.007888 | -0.007239 | 0.01172 | 0.01352 |
| RDWSD | rs4849439 | rs1480501 | C | T | 2 | DPP10 | Male only | False | 0.1246 | 0.1247 | 0.1154 | 0.1151 | 31506 | 24073 | 188075 | 162398 | -0.006432 | -0.07608 | -0.008807 | -0.00704 | 0.01176 | 0.01357 |
| RDWSD | rs4849439 | rs4849439 | C | T | 2 | DPP10 | Male only | False | 0.1235 | 0.1236 | 0.1152 | 0.1149 | 31506 | 24073 | 188075 | 162398 | -0.007944 | -0.07652 | -0.008627 | -0.00696 | 0.01182 | 0.01362 |
| RDWSD | rs4849439 | rs6542303 | C | A | 2 | DPP10 | Male only | False | 0.1251 | 0.125 | 0.1158 | 0.1155 | 31506 | 24073 | 188075 | 162398 | -0.007205 | -0.07476 | -0.008746 | -0.006928 | 0.01176 | 0.01356 |
| RDWSD | rs4849439 | rs2035325 | G | A | 2 | DPP10 | Male only | False | 0.1239 | 0.1238 | 0.1149 | 0.1145 | 31506 | 24073 | 188075 | 162398 | -0.006634 | -0.07396 | -0.008653 | -0.006953 | 0.0118 | 0.0136 |
| RDWSD | rs4849439 | rs55657426 | G | A | 2 | DPP10 | Male only | False | 0.1154 | 0.1155 | 0.1082 | 0.1083 | 31506 | 24073 | 188075 | 162398 | -0.005933 | -0.07639 | -0.009305 | -0.006807 | 0.01217 | 0.01401 |
| RDWSD | rs10901256 | rs8176635 | A | G | 9 | ABO | Female only | True | 0.326 | 0.3232 | 0.3297 | 0.3295 | 31506 | 24073 | 188075 | 162398 | -0.04074 | -0.008986 | -0.02651 | -0.02405 | 0.00828 | 0.009634 |
| RDWSD | rs10901256 | rs9411492 | C | T | 9 | ABO | Female only | True | 0.2524 | 0.2494 | 0.2547 | 0.2549 | 31506 | 24073 | 188075 | 162398 | -0.04928 | -0.01666 | -0.03402 | -0.03435 | 0.008959 | 0.01034 |
| RDWSD | rs10901256 | rs10901256 | C | T | 9 | ABO | Female only | True | 0.2541 | 0.2507 | 0.2558 | 0.2559 | 31506 | 24073 | 188075 | 162398 | -0.05068 | -0.01766 | -0.03441 | -0.03434 | 0.008942 | 0.01032 |
| RDWSD | rs10901256 | rs10901257 | A | G | 9 | ABO | Female only | True | 0.2542 | 0.2507 | 0.2559 | 0.2561 | 31506 | 24073 | 188075 | 162398 | -0.05054 | -0.01749 | -0.03443 | -0.03438 | 0.008941 | 0.01031 |
| RDWSD | rs10901256 | rs4962125 | T | C | 9 | ABO | Female only | True | 0.2544 | 0.2518 | 0.2569 | 0.2571 | 31506 | 24073 | 188075 | 162398 | -0.04857 | -0.01812 | -0.0351 | -0.03387 | 0.008954 | 0.01032 |
| RDWSD | rs10901256 | rs17150482 | C | T | 9 | ABO | Female only | True | 0.2755 | 0.2751 | 0.285 | 0.2854 | 31506 | 24073 | 188075 | 162398 | -0.04052 | -0.01838 | -0.03022 | -0.02961 | 0.008747 | 0.01006 |
| RDWSD | rs74979599 | rs11023877 | C | T | 11 | SOX6 | Female only | True | 0.07744 | 0.07843 | 0.0845 | 0.08292 | 31506 | 24073 | 188075 | 162398 | 0.09242 | 0.02362 | 0.09229 | 0.09148 | 0.01457 | 0.01678 |
| RDWSD | rs74979599 | rs11023878 | A | G | 11 | SOX6 | Female only | True | 0.07744 | 0.07845 | 0.0845 | 0.08292 | 31506 | 24073 | 188075 | 162398 | 0.09229 | 0.02383 | 0.0923 | 0.09148 | 0.01456 | 0.01678 |
| RDWSD | rs74979599 | rs11023883 | G | A | 11 | SOX6 | Female only | True | 0.07638 | 0.07776 | 0.08352 | 0.08179 | 31506 | 24073 | 188075 | 162398 | 0.09735 | 0.02552 | 0.1003 | 0.09689 | 0.01463 | 0.01688 |
| RDWSD | rs74979599 | rs17539593 | A | C | 11 | SOX6 | Female only | True | 0.07671 | 0.07816 | 0.08376 | 0.08209 | 31506 | 24073 | 188075 | 162398 | 0.09819 | 0.02453 | 0.1 | 0.09675 | 0.0146 | 0.01682 |
| RDWSD | rs74979599 | rs11023888 | G | T | 11 | SOX6 | Female only | True | 0.07641 | 0.07782 | 0.08354 | 0.08181 | 31506 | 24073 | 188075 | 162398 | 0.09741 | 0.02508 | 0.1003 | 0.09701 | 0.01462 | 0.01687 |
| RDWSD | rs74979599 | rs78243491 | C | T | 11 | SOX6 | Female only | True | 0.07723 | 0.07871 | 0.08378 | 0.08211 | 31506 | 24073 | 188075 | 162398 | 0.0972 | 0.02347 | 0.1001 | 0.09688 | 0.01449 | 0.01668 |
| RDWSD | rs74979599 | rs11512059 | T | C | 11 | SOX6 | Female only | True | 0.07672 | 0.07817 | 0.08378 | 0.08211 | 31506 | 24073 | 188075 | 162398 | 0.09812 | 0.02393 | 0.1001 | 0.09694 | 0.01459 | 0.0168 |
| RDWSD | rs74979599 | rs11023895 | A | G | 11 | SOX6 | Female only | True | 0.07663 | 0.0781 | 0.08365 | 0.08199 | 31506 | 24073 | 188075 | 162398 | 0.09811 | 0.02364 | 0.1005 | 0.09703 | 0.0146 | 0.0168 |
| RDWSD | rs74979599 | rs74375633 | T | C | 11 | SOX6 | Female only | True | 0.07681 | 0.0782 | 0.08384 | 0.08217 | 31506 | 24073 | 188075 | 162398 | 0.09768 | 0.02304 | 0.1002 | 0.09693 | 0.01459 | 0.0168 |
| RDWSD | rs74979599 | rs11023905 | C | T | 11 | SOX6 | Female only | True | 0.1087 | 0.1091 | 0.1142 | 0.112 | 31506 | 24073 | 188075 | 162398 | 0.06836 | 0.01643 | 0.07172 | 0.07312 | 0.01242 | 0.01438 |

| trait | lead_snp | ID | ALLELE1 | ALLELE0 | CHR | Gene_Symbol | independent_loci_in_sex_P1e2 | in_embl_any_or_gene | A1FREQ_VUMC_female | A1FREQ_VUMC_male | A1FREQ_UKB_female | A1FREQ_UKB_male | N_VUMC_female | N_VUMC_male | N_UKB_female | N_UKB_male | BETA_VUMC_female | BETA_VUMC_male | BETA_UKB_female | BETA_UKB_male | SE_VUMC_female | SE_VUMC_male |
|---|---|---|---|---|---|---|---|---|---|---|---|---|---|---|---|---|---|---|---|---|---|---|
| RDWSD | rs74979599 | rs78936881 | T | C | 11 | SOX6 | Female only | True | 0.07735 | 0.07874 | 0.08393 | 0.08223 | 31506 | 24073 | 188075 | 162398 | 0.09835 | 0.02717 | 0.09952 | 0.0968 | 0.01454 | 0.01674 |
| RDWSD | rs74979599 | rs16932896 | T | C | 11 | SOX6 | Female only | True | 0.1064 | 0.1068 | 0.1127 | 0.1106 | 31506 | 24073 | 188075 | 162398 | 0.06799 | 0.0171 | 0.07172 | 0.07364 | 0.01264 | 0.01461 |
| RDWSD | rs74979599 | rs11023911 | T | G | 11 | SOX6 | Female only | True | 0.07722 | 0.0786 | 0.08397 | 0.08227 | 31506 | 24073 | 188075 | 162398 | 0.09864 | 0.02748 | 0.09959 | 0.09679 | 0.01459 | 0.01679 |
| RDWSD | rs74979599 | rs12281323 | G | A | 11 | SOX6 | Female only | True | 0.1074 | 0.1079 | 0.1139 | 0.1118 | 31506 | 24073 | 188075 | 162398 | 0.06927 | 0.0166 | 0.07175 | 0.07382 | 0.0126 | 0.01456 |
| RDWSD | rs74979599 | rs75817865 | T | C | 11 | SOX6 | Female only | True | 0.07714 | 0.07854 | 0.08395 | 0.08226 | 31506 | 24073 | 188075 | 162398 | 0.099 | 0.02742 | 0.09952 | 0.09681 | 0.01464 | 0.01685 |
| RDWSD | rs74979599 | rs1393923 | G | A | 11 | SOX6 | Female only | True | 0.1078 | 0.1082 | 0.1142 | 0.112 | 31506 | 24073 | 188075 | 162398 | 0.06882 | 0.0177 | 0.07175 | 0.07311 | 0.01257 | 0.01454 |
| RDWSD | rs74979599 | rs11023914 | C | T | 11 | SOX6 | Female only | True | 0.1078 | 0.1082 | 0.1142 | 0.112 | 31506 | 24073 | 188075 | 162398 | 0.06873 | 0.01769 | 0.0718 | 0.07309 | 0.01257 | 0.01454 |
| RDWSD | rs74979599 | rs11023919 | G | A | 11 | SOX6 | Female only | True | 0.07698 | 0.07828 | 0.08393 | 0.08224 | 31506 | 24073 | 188075 | 162398 | 0.1004 | 0.02724 | 0.09956 | 0.09688 | 0.01478 | 0.017 |
| RDWSD | rs74979599 | rs11023922 | T | C | 11 | SOX6 | Female only | True | 0.07695 | 0.07826 | 0.08386 | 0.08225 | 31506 | 24073 | 188075 | 162398 | 0.1004 | 0.02724 | 0.09954 | 0.09749 | 0.01478 | 0.017 |
| RDWSD | rs74979599 | rs74979599 | G | A | 11 | SOX6 | Female only | True | 0.07692 | 0.07814 | 0.08391 | 0.08218 | 31506 | 24073 | 188075 | 162398 | 0.1011 | 0.02786 | 0.09998 | 0.09716 | 0.01482 | 0.01705 |
| RDWSD | rs9932423 | rs2932690 | G | A | 16 | PIEZO1 | Female only | True | 0.3136 | 0.3121 | 0.3224 | 0.3223 | 31506 | 24073 | 188075 | 162398 | -0.05382 | -0.009168 | -0.01868 | -0.003446 | 0.008443 | 0.009718 |
| RDWSD | rs9932423 | rs9933309 | T | C | 16 | PIEZO1 | Female only | True | 0.2927 | 0.2921 | 0.3042 | 0.3042 | 31506 | 24073 | 188075 | 162398 | -0.0558 | -0.01092 | -0.01862 | -0.005283 | 0.008551 | 0.009866 |
| RDWSD | rs9932423 | rs9932423 | A | C | 16 | PIEZO1 | Female only | True | 0.2967 | 0.2964 | 0.3075 | 0.3075 | 31506 | 24073 | 188075 | 162398 | -0.0579 | -0.009618 | -0.01832 | -0.005281 | 0.008521 | 0.009822 |
| SYSTOLIC | rs7136259 | rs1689040 | T | C | 12 | ATP2B1 | Female only | True | 0.4022 | 0.4062 | 0.4064 | 0.4063 | 35815 | 26956 | 182645 | 157514 | -0.04329 | -0.01783 | -0.02921 | -0.01987 | 0.006818 | 0.007708 |
| SYSTOLIC | rs7136259 | rs1401982 | G | A | 12 | ATP2B1 | Female only | True | 0.403 | 0.4071 | 0.4066 | 0.4065 | 35815 | 26956 | 182645 | 157514 | -0.04321 | -0.01849 | -0.02913 | -0.01997 | 0.006816 | 0.007704 |
| SYSTOLIC | rs7136259 | rs2070759 | G | T | 12 | ATP2B1 | Female only | True | 0.4678 | 0.4711 | 0.4704 | 0.4703 | 35815 | 26956 | 182645 | 157514 | -0.04054 | -0.01011 | -0.02706 | -0.01558 | 0.006702 | 0.007584 |
| SYSTOLIC | rs7136259 | rs7297206 | C | T | 12 | ATP2B1 | Female only | True | 0.4002 | 0.4039 | 0.4028 | 0.4027 | 35815 | 26956 | 182645 | 157514 | -0.04257 | -0.0187 | -0.02923 | -0.01977 | 0.006849 | 0.007758 |
| SYSTOLIC | rs7136259 | rs2681485 | G | A | 12 | ATP2B1 | Female only | True | 0.4031 | 0.4072 | 0.4065 | 0.4064 | 35815 | 26956 | 182645 | 157514 | -0.04358 | -0.01882 | -0.0292 | -0.0201 | 0.006816 | 0.007706 |
| SYSTOLIC | rs7136259 | rs10858914 | C | T | 12 | ATP2B1 | Female only | True | 0.4703 | 0.4732 | 0.4706 | 0.4706 | 35815 | 26956 | 182645 | 157514 | -0.04185 | -0.009986 | -0.02691 | -0.01549 | 0.006725 | 0.007615 |
| SYSTOLIC | rs7136259 | rs939329 | G | A | 12 | ATP2B1 | Female only | True | 0.4702 | 0.473 | 0.4708 | 0.4708 | 35815 | 26956 | 182645 | 157514 | -0.04157 | -0.01006 | -0.02714 | -0.0153 | 0.006721 | 0.007612 |
| SYSTOLIC | rs7136259 | rs10858915 | G | A | 12 | ATP2B1 | Female only | True | 0.4937 | 0.496 | 0.4922 | 0.4922 | 35815 | 26956 | 182645 | 157514 | -0.04132 | -0.0043 | -0.02649 | -0.01376 | 0.006726 | 0.007615 |
| SYSTOLIC | rs7136259 | rs7136259 | T | C | 12 | ATP2B1 | Female only | True | 0.4137 | 0.4182 | 0.4176 | 0.4182 | 35815 | 26956 | 182645 | 157514 | -0.04377 | -0.0176 | -0.02789 | -0.0198 | 0.006781 | 0.007686 |
| TBil | rs17869072 | rs6746331 | A | G | 2 | TRPM8 | Female only | True | 0.2128 | 0.2095 | 0.2024 | 0.2028 | 31936 | 24820 | 184244 | 158585 | 0.03266 | 0.01439 | 0.04187 | 0.0306 | 0.008981 | 0.01047 |
| TBil | rs17869072 | rs11563204 | A | G | 2 | TRPM8 | Female only | True | 0.2126 | 0.2097 | 0.2023 | 0.2026 | 31936 | 24820 | 184244 | 158585 | 0.0317 | 0.01329 | 0.04074 | 0.03015 | 0.008985 | 0.01046 |
| TBil | rs17869072 | rs73996666 | T | C | 2 | TRPM8 | Female only | True | 0.2226 | 0.2197 | 0.212 | 0.2127 | 31936 | 24820 | 184244 | 158585 | 0.02873 | 0.01496 | 0.0422 | 0.02943 | 0.008833 | 0.01028 |
| TBil | rs17869072 | rs28953469 | T | G | 2 | TRPM8 | Female only | True | 0.2219 | 0.219 | 0.2123 | 0.213 | 31936 | 24820 | 184244 | 158585 | 0.02828 | 0.01504 | 0.04193 | 0.02917 | 0.008865 | 0.01031 |
| TBil | rs17869072 | rs28953470 | A | G | 2 | TRPM8 | Female only | True | 0.2219 | 0.219 | 0.2123 | 0.213 | 31936 | 24820 | 184244 | 158585 | 0.02833 | 0.01506 | 0.04181 | 0.02883 | 0.008865 | 0.01031 |
| TBil | rs17869072 | rs17869072 | C | T | 2 | TRPM8 | Female only | True | 0.1532 | 0.1535 | 0.1484 | 0.149 | 31936 | 24820 | 184244 | 158585 | 0.05645 | 0.0265 | 0.0692 | 0.05397 | 0.01029 | 0.01183 |
| TBil | rs17869072 | rs17865680 | G | A | 2 | TRPM8 | Female only | True | 0.2218 | 0.2192 | 0.2124 | 0.213 | 31936 | 24820 | 184244 | 158585 | 0.02835 | 0.01476 | 0.04187 | 0.02888 | 0.008873 | 0.01032 |

| trait | lead_snp | ID | ALLELE1 | ALLELE0 | CHR | Gene_Symbol | independent_loci_in_sex_P1e2 | in_embl_any_or_gene | A1FREQ_VUMC_female | A1FREQ_VUMC_male | A1FREQ_UKB_female | A1FREQ_UKB_male | N_VUMC_female | N_VUMC_male | N_UKB_female | N_UKB_male | BETA_VUMC_female | BETA_VUMC_male | BETA_UKB_female | BETA_UKB_male | SE_VUMC_female | SE_VUMC_male |
|---|---|---|---|---|---|---|---|---|---|---|---|---|---|---|---|---|---|---|---|---|---|---|
| TBil | rs17869072 | rs75371085 | A | G | 2 | TRPM8 | Female only | True | 0.1597 | 0.1601 | 0.1531 | 0.1536 | 31936 | 24820 | 184244 | 158585 | 0.05374 | 0.02315 | 0.06439 | 0.04958 | 0.01008 | 0.01162 |
| TBil | rs17869072 | rs78170816 | T | C | 2 | TRPM8 | Female only | True | 0.2222 | 0.2196 | 0.2129 | 0.2134 | 31936 | 24820 | 184244 | 158585 | 0.02766 | 0.01279 | 0.04085 | 0.02785 | 0.008868 | 0.01031 |
| TBil | rs17869072 | rs113993121 | C | A | 2 | TRPM8 | Female only | True | 0.2214 | 0.2189 | 0.2122 | 0.2127 | 31936 | 24820 | 184244 | 158585 | 0.02879 | 0.0148 | 0.04175 | 0.02868 | 0.008883 | 0.01033 |
| trait | lead_snp | ID | ALLELE1 | ALLELE0 | CHR | Gene_Symbol | independent_loci_in_sex_P1e2 | in_embl_any_or_gene | A1FREQ_VUMC_female | A1FREQ_VUMC_male | A1FREQ_UKB_female | A1FREQ_UKB_male | N_VUMC_female | N_VUMC_male | N_UKB_female | N_UKB_male | BETA_VUMC_female | BETA_VUMC_male | BETA_UKB_female | BETA_UKB_male | SE_VUMC_female | SE_VUMC_male |
| TBil | rs17869072 | rs75090001 | T | C | 2 | TRPM8 | Female only | True | 0.2214 | 0.2189 | 0.2122 | 0.2127 | 31936 | 24820 | 184244 | 158585 | 0.02891 | 0.0148 | 0.04176 | 0.02869 | 0.008883 | 0.01033 |
| TBil | rs17869072 | rs17864779 | T | C | 2 | TRPM8 | Female only | True | 0.2213 | 0.2188 | 0.2118 | 0.2123 | 31936 | 24820 | 184244 | 158585 | 0.02877 | 0.01478 | 0.04079 | 0.0278 | 0.008886 | 0.01034 |
| TBil | rs17869072 | rs11563202 | C | T | 2 | TRPM8 | Female only | True | 0.1631 | 0.1638 | 0.1553 | 0.1555 | 31936 | 24820 | 184244 | 158585 | 0.05128 | 0.01589 | 0.06104 | 0.04565 | 0.009954 | 0.01148 |
| TBil | rs17869072 | rs10490019 | A | G | 2 | TRPM8 | Female only | True | 0.1621 | 0.1626 | 0.1547 | 0.1549 | 31936 | 24820 | 184244 | 158585 | 0.05253 | 0.01873 | 0.06242 | 0.04706 | 0.009998 | 0.01153 |
| TBil | rs17869072 | rs28948677 | A | G | 2 | TRPM8 | Female only | True | 0.1626 | 0.1633 | 0.1553 | 0.1555 | 31936 | 24820 | 184244 | 158585 | 0.05143 | 0.01587 | 0.06103 | 0.04564 | 0.009986 | 0.01151 |
| TBil | rs17869072 | rs11563201 | T | C | 2 | TRPM8 | Female only | True | 0.1626 | 0.1633 | 0.1553 | 0.1555 | 31936 | 24820 | 184244 | 158585 | 0.0514 | 0.01586 | 0.06095 | 0.04559 | 0.009986 | 0.01151 |
| TBil | rs17869072 | rs11563199 | T | C | 2 | TRPM8 | Female only | True | 0.1621 | 0.1626 | 0.1547 | 0.1549 | 31936 | 24820 | 184244 | 158585 | 0.05248 | 0.01875 | 0.0622 | 0.04688 | 0.009999 | 0.01153 |
| TBil | rs17869072 | rs17869077 | T | C | 2 | TRPM8 | Female only | True | 0.1621 | 0.1626 | 0.1547 | 0.1549 | 31936 | 24820 | 184244 | 158585 | 0.05248 | 0.01875 | 0.0622 | 0.04688 | 0.009999 | 0.01153 |
| TBil | rs17869072 | rs11901004 | T | G | 2 | TRPM8 | Female only | True | 0.1621 | 0.1627 | 0.1547 | 0.1549 | 31936 | 24820 | 184244 | 158585 | 0.05233 | 0.01878 | 0.06219 | 0.04687 | 0.009998 | 0.01153 |
| TBil | rs17869072 | rs11901051 | T | G | 2 | TRPM8 | Female only | True | 0.1621 | 0.1627 | 0.1547 | 0.1549 | 31936 | 24820 | 184244 | 158585 | 0.05227 | 0.01877 | 0.06217 | 0.04686 | 0.009998 | 0.01153 |
| TBil | rs17869072 | rs17865682 | G | A | 2 | TRPM8 | Female only | True | 0.1626 | 0.1633 | 0.1553 | 0.1555 | 31936 | 24820 | 184244 | 158585 | 0.05115 | 0.01586 | 0.06082 | 0.04544 | 0.009986 | 0.01151 |
| TBil | rs17869072 | rs17865683 | C | T | 2 | TRPM8 | Female only | True | 0.1621 | 0.1627 | 0.1547 | 0.1549 | 31936 | 24820 | 184244 | 158585 | 0.05227 | 0.01876 | 0.06221 | 0.04686 | 0.009998 | 0.01153 |
| TBil | rs17869072 | rs17864782 | A | C | 2 | TRPM8 | Female only | True | 0.1621 | 0.1627 | 0.1547 | 0.1549 | 31936 | 24820 | 184244 | 158585 | 0.05224 | 0.01877 | 0.0622 | 0.04686 | 0.009998 | 0.01153 |
| TBil | rs17869072 | rs17869079 | T | C | 2 | TRPM8 | Female only | True | 0.1621 | 0.1627 | 0.1547 | 0.1549 | 31936 | 24820 | 184244 | 158585 | 0.05223 | 0.01877 | 0.0622 | 0.04686 | 0.009998 | 0.01153 |
| TBil | rs17869072 | rs17864783 | G | T | 2 | TRPM8 | Female only | True | 0.1621 | 0.1627 | 0.1548 | 0.155 | 31936 | 24820 | 184244 | 158585 | 0.05215 | 0.01878 | 0.06225 | 0.04692 | 0.009998 | 0.01153 |
| TBil | rs17869072 | rs17866592 | C | T | 2 | TRPM8 | Female only | True | 0.1622 | 0.1628 | 0.1548 | 0.155 | 31936 | 24820 | 184244 | 158585 | 0.05221 | 0.01894 | 0.06206 | 0.04673 | 0.009997 | 0.01153 |
| TBil | rs17869072 | rs17864785 | C | T | 2 | TRPM8 | Female only | True | 0.1626 | 0.1633 | 0.1554 | 0.1555 | 31936 | 24820 | 184244 | 158585 | 0.05104 | 0.01591 | 0.06072 | 0.04539 | 0.009985 | 0.01151 |
| TBil | rs17869072 | rs17865685 | T | C | 2 | TRPM8 | Female only | True | 0.1636 | 0.1642 | 0.1557 | 0.1559 | 31936 | 24820 | 184244 | 158585 | 0.05157 | 0.01628 | 0.06086 | 0.0454 | 0.009944 | 0.01146 |
| TBil | rs17869072 | rs17865686 | A | G | 2 | TRPM8 | Female only | True | 0.166 | 0.1667 | 0.159 | 0.1593 | 31936 | 24820 | 184244 | 158585 | 0.04868 | 0.01742 | 0.05795 | 0.04247 | 0.009905 | 0.01142 |
| TBil | rs17869072 | rs17874959 | T | C | 2 | TRPM8 | Female only | True | 0.1661 | 0.1668 | 0.1591 | 0.1593 | 31936 | 24820 | 184244 | 158585 | 0.04923 | 0.0171 | 0.05823 | 0.0427 | 0.009905 | 0.01142 |
| Weight | rs11546878 | rs11546878 | T | C | 3 | EIF2B5 | Female only | True | 0.1758 | 0.1714 | 0.1742 | 0.1759 | 35804 | 26877 | 193627 | 166489 | -0.05931 | -0.01663 | -0.01921 | -0.02987 | 0.009482 | 0.01123 |
| Weight | rs11546878 | rs11545169 | T | G | 3 | EIF2B5 | Female only | True | 0.1587 | 0.1567 | 0.1607 | 0.1625 | 35804 | 26877 | 193627 | 166489 | -0.05085 | -0.02349 | -0.01319 | -0.02721 | 0.009988 | 0.01172 |
| Weight | rs11546878 | rs2293605 | T | C | 3 | EIF2B5 | Female only | True | 0.1227 | 0.1201 | 0.1198 | 0.1213 | 35804 | 26877 | 193627 | 166489 | -0.0557 | -0.008097 | -0.01723 | -0.02839 | 0.01101 | 0.013 |
| Weight | rs11242816 | rs11242816 | T | C | 6 | NQO2 | Male only | False | 0.06736 | 0.06519 | 0.06914 | 0.06897 | 35804 | 26877 | 193627 | 166489 | 0.01288 | 0.09482 | -0.001225 | 0.005167 | 0.01451 | 0.01732 |

| trait | lead_snp | ID | ALLELE1 | ALLELE0 | CHR | Gene_Symbol | independent_loci_in_sex_P1e2 | in_embl_any_or_gene | A1FREQ_VUMC_female | A1FREQ_VUMC_male | A1FREQ_UKB_female | A1FREQ_UKB_male | N_VUMC_female | N_VUMC_male | N_UKB_female | N_UKB_male | BETA_VUMC_female | BETA_VUMC_male | BETA_UKB_female | BETA_UKB_male | SE_VUMC_female | SE_VUMC_male |
|---|---|---|---|---|---|---|---|---|---|---|---|---|---|---|---|---|---|---|---|---|---|---|
| Weight | rs11242816 | rs28383589 | T | G | 6 | NQO2 | Male only | False | 0.06975 | 0.06756 | 0.07141 | 0.07111 | 35804 | 26877 | 193627 | 166489 | 0.01349 | 0.08991 | 8.26e-04 | 0.004257 | 0.01425 | 0.01697 |
| Weight | rs11242816 | rs3757080 | T | C | 6 | NQO2 | Male only | False | 0.05548 | 0.05558 | 0.05773 | 0.05728 | 35804 | 26877 | 193627 | 166489 | 0.009089 | 0.05742 | -3.39e-04 | 0.01464 | 0.01572 | 0.01848 |
| Weight | rs11242816 | rs28383634 | A | G | 6 | NQO2 | Male only | False | 0.06997 | 0.07015 | 0.07015 | 0.06944 | 35804 | 26877 | 193627 | 166489 | 6.57e-04 | 0.06063 | 0.003965 | 0.009436 | 0.01412 | 0.01663 |
| Weight | rs11242816 | rs3823093 | C | A | 6 | NQO2 | Male only | False | 0.06729 | 0.06751 | 0.06956 | 0.06882 | 35804 | 26877 | 193627 | 166489 | 0.003092 | 0.06301 | 0.00328 | 0.009251 | 0.01453 | 0.01711 |
| Weight | rs11242816 | rs3799204 | G | A | 6 | NQO2 | Male only | False | 0.06942 | 0.06952 | 0.06965 | 0.06895 | 35804 | 26877 | 193627 | 166489 | 0.00102 | 0.0613 | 0.003191 | 0.009586 | 0.01415 | 0.01668 |
| Weight | rs11242816 | rs4149365 | T | C | 6 | NQO2 | Male only | False | 0.05512 | 0.05541 | 0.05776 | 0.05729 | 35804 | 26877 | 193627 | 166489 | 0.009369 | 0.05745 | -6.21e-04 | 0.01465 | 0.01579 | 0.01856 |
| Weight | rs11242816 | rs4149366 | T | C | 6 | NQO2 | Male only | False | 0.06984 | 0.07005 | 0.07025 | 0.06955 | 35804 | 26877 | 193627 | 166489 | 8.54e-04 | 0.06164 | 0.004212 | 0.00946 | 0.01413 | 0.01663 |
| Weight | rs11242816 | rs4149367 | T | C | 6 | NQO2 | Male only | False | 0.07023 | 0.07042 | 0.07028 | 0.06958 | 35804 | 26877 | 193627 | 166489 | 0.001372 | 0.06133 | 0.004168 | 0.009559 | 0.01406 | 0.01655 |
| Weight | rs11242816 | rs10223372 | G | T | 6 | NQO2 | Male only | False | 0.05651 | 0.05707 | 0.05808 | 0.05758 | 35804 | 26877 | 193627 | 166489 | 0.008977 | 0.06022 | -0.001419 | 0.01525 | 0.01567 | 0.01834 |
| Weight | rs11242816 | rs57563768 | C | T | 6 | NQO2 | Male only | False | 0.05711 | 0.05756 | 0.05826 | 0.0577 | 35804 | 26877 | 193627 | 166489 | 0.007366 | 0.0606 | -0.001094 | 0.01586 | 0.01562 | 0.01831 |
| Weight | rs1231281 | rs516246 | C | T | 19 | RASIP1 | Female only | True | 0.4988 | 0.4972 | 0.4886 | 0.4906 | 35804 | 26877 | 193627 | 166489 | 0.02981 | 0.01368 | 0.007682 | 0.002387 | 0.007172 | 0.008483 |
| Weight | rs1231281 | rs492602 | A | G | 19 | RASIP1 | Female only | True | 0.499 | 0.4972 | 0.4887 | 0.4907 | 35804 | 26877 | 193627 | 166489 | 0.02978 | 0.01343 | 0.007708 | 0.002417 | 0.007171 | 0.008484 |
| Weight | rs1231281 | rs681343 | C | T | 19 | RASIP1 | Female only | True | 0.499 | 0.4973 | 0.4886 | 0.4906 | 35804 | 26877 | 193627 | 166489 | 0.02977 | 0.01344 | 0.007754 | 0.002207 | 0.007172 | 0.008484 |
| Weight | rs1231281 | rs601338 | G | A | 19 | RASIP1 | Female only | True | 0.4964 | 0.4981 | 0.4887 | 0.4906 | 35804 | 26877 | 193627 | 166489 | 0.02996 | 0.01357 | 0.007568 | 0.002191 | 0.007242 | 0.008566 |
| Weight | rs1231281 | rs602662 | G | A | 19 | RASIP1 | Female only | True | 0.4787 | 0.4777 | 0.4612 | 0.4634 | 35804 | 26877 | 193627 | 166489 | 0.03274 | 0.007831 | 0.007534 | 0.002817 | 0.007255 | 0.008559 |
| Weight | rs1231281 | rs485186 | A | G | 19 | RASIP1 | Female only | True | 0.4726 | 0.472 | 0.46 | 0.4623 | 35804 | 26877 | 193627 | 166489 | 0.03302 | 0.008212 | 0.007334 | 0.002901 | 0.007189 | 0.008485 |
| Weight | rs1231281 | rs485073 | A | G | 19 | RASIP1 | Female only | True | 0.479 | 0.4781 | 0.4619 | 0.4642 | 35804 | 26877 | 193627 | 166489 | 0.03339 | 0.008002 | 0.007465 | 0.003034 | 0.007268 | 0.008578 |
| Weight | rs1231281 | rs603985 | T | C | 19 | RASIP1 | Female only | True | 0.4791 | 0.4782 | 0.462 | 0.4643 | 35804 | 26877 | 193627 | 166489 | 0.03361 | 0.008079 | 0.007555 | 0.003019 | 0.007268 | 0.008579 |
| Weight | rs1231281 | rs571689 | C | T | 19 | RASIP1 | Female only | True | 0.4792 | 0.4789 | 0.4624 | 0.4645 | 35804 | 26877 | 193627 | 166489 | 0.03317 | 0.01033 | 0.007333 | 0.003063 | 0.007253 | 0.00856 |
| Weight | rs1231281 | rs570794 | T | C | 19 | RASIP1 | Female only | True | 0.4792 | 0.4789 | 0.4624 | 0.4645 | 35804 | 26877 | 193627 | 166489 | 0.03317 | 0.01034 | 0.007335 | 0.003077 | 0.007253 | 0.00856 |
| Weight | rs1231281 | 19:49207718 | T | C | 19 | RASIP1 | Female only | True | 0.4792 | 0.4789 | | | 35804 | 26877 | | | 0.03317 | 0.01034 | | | 0.007253 | 0.00856 |
| Weight | rs1231281 | rs2251034 | G | A | 19 | RASIP1 | Female only | True | 0.4758 | 0.4753 | 0.463 | 0.4651 | 35804 | 26877 | 193627 | 166489 | 0.03293 | 0.00897 | 0.007269 | 0.003 | 0.007185 | 0.008483 |
| Weight | rs1231281 | rs507855 | A | G | 19 | RASIP1 | Female only | True | 0.4813 | 0.4808 | 0.4637 | 0.4658 | 35804 | 26877 | 193627 | 166489 | 0.03307 | 0.00971 | 0.007354 | 0.002873 | 0.007265 | 0.008576 |
| Weight | rs1231281 | rs507766 | T | C | 19 | RASIP1 | Female only | True | 0.4813 | 0.4809 | 0.4637 | 0.4658 | 35804 | 26877 | 193627 | 166489 | 0.03303 | 0.009698 | 0.007389 | 0.002878 | 0.007266 | 0.008577 |
| Weight | rs1231281 | rs507711 | C | T | 19 | RASIP1 | Female only | True | 0.4813 | 0.4809 | 0.4637 | 0.4658 | 35804 | 26877 | 193627 | 166489 | 0.0331 | 0.0096 | 0.007375 | 0.002905 | 0.007266 | 0.008577 |
| Weight | rs1231281 | rs504963 | G | A | 19 | RASIP1 | Female only | True | 0.4801 | 0.4798 | 0.4628 | 0.4648 | 35804 | 26877 | 193627 | 166489 | 0.03286 | 0.009703 | 0.007364 | 0.003019 | 0.007252 | 0.008559 |
| Weight | rs1231281 | rs632111 | A | G | 19 | RASIP1 | Female only | True | 0.4767 | 0.476 | 0.4633 | 0.4655 | 35804 | 26877 | 193627 | 166489 | 0.03292 | 0.009214 | 0.00731 | 0.003071 | 0.007185 | 0.008481 |
| Weight | rs1231281 | rs503279 | T | C | 19 | RASIP1 | Female only | True | 0.4798 | 0.4795 | 0.4626 | 0.4646 | 35804 | 26877 | 193627 | 166489 | 0.03326 | 0.009671 | 0.00734 | 0.003057 | 0.007253 | 0.00856 |
| Weight | rs1231281 | rs633372 | G | A | 19 | RASIP1 | Female only | True | 0.4809 | 0.4803 | 0.4631 | 0.4652 | 35804 | 26877 | 193627 | 166489 | 0.03338 | 0.008802 | 0.007273 | 0.002968 | 0.007249 | 0.008558 |

| trait | lead_snp | ID | ALLELE1 | ALLELE0 | CHR | Gene_Symbol | independent_loci_in_sex_P1e2 | in_embl_any_or_gene | A1FREQ_VUMC_female | A1FREQ_VUMC_male | A1FREQ_UKB_female | A1FREQ_UKB_male | N_VUMC_female | N_VUMC_male | N_UKB_female | N_UKB_male | BETA_VUMC_female | BETA_VUMC_male | BETA_UKB_female | BETA_UKB_male | SE_VUMC_female | SE_VUMC_male |
|---|---|---|---|---|---|---|---|---|---|---|---|---|---|---|---|---|---|---|---|---|---|---|
| Weight | rs1231281 | rs2548459 | T | C | 19 | RASIP1 | Female only | True | 0.4813 | 0.4808 | 0.4633 | 0.4655 | 35804 | 26877 | 193627 | 166489 | 0.03305 | 0.008787 | 0.007177 | 0.00299 | 0.007253 | 0.008561 |
| Weight | rs1231281 | rs692854 | C | A | 19 | RASIP1 | Female only | True | 0.4811 | 0.4809 | 0.4642 | 0.4664 | 35804 | 26877 | 193627 | 166489 | 0.03369 | 0.009644 | 0.007255 | 0.00289 | 0.007253 | 0.008563 |
| Weight | rs1231281 | rs1704773 | A | G | 19 | RASIP1 | Female only | True | 0.4822 | 0.4817 | 0.4647 | 0.4669 | 35804 | 26877 | 193627 | 166489 | 0.03375 | 0.009194 | 0.007247 | 0.00281 | 0.00725 | 0.008563 |
| Weight | rs1231281 | rs646327 | A | G | 19 | RASIP1 | Female only | True | 0.4845 | 0.4843 | 0.4683 | 0.4705 | 35804 | 26877 | 193627 | 166489 | 0.0353 | 0.009928 | 0.007248 | 0.003111 | 0.007254 | 0.008569 |
| Weight | rs1231281 | rs676388 | T | C | 19 | RASIP1 | Female only | True | 0.4798 | 0.479 | 0.4632 | 0.4653 | 35804 | 26877 | 193627 | 166489 | 0.03651 | 0.009929 | 0.007875 | 0.003096 | 0.007263 | 0.008574 |
| Weight | rs1231281 | rs546308 | G | A | 19 | RASIP1 | Female only | True | 0.3998 | 0.3958 | 0.389 | 0.3914 | 35804 | 26877 | 193627 | 166489 | 0.02524 | -0.00194 | 0.005352 | 0.00616 | 0.007307 | 0.008705 |
| Weight | rs1231281 | rs3859439 | T | C | 19 | RASIP1 | Female only | True | 0.4034 | 0.3994 | 0.3882 | 0.3905 | 35804 | 26877 | 193627 | 166489 | 0.02474 | -0.001721 | 0.005157 | 0.006691 | 0.007364 | 0.008774 |

| trait | lead_snp | ID | ALLELE1 | ALLELE0 | CHR | Gene_Symbol | independent_loci_in_sex_P1e2 | in_embl_any_or_gene | A1FREQ_VUMC_female | A1FREQ_VUMC_male | A1FREQ_UKB_female | A1FREQ_UKB_male | N_VUMC_female | N_VUMC_male | N_UKB_female | N_UKB_male | BETA_VUMC_female | BETA_VUMC_male | BETA_UKB_female | BETA_UKB_male | SE_VUMC_female | SE_VUMC_male |
|---|---|---|---|---|---|---|---|---|---|---|---|---|---|---|---|---|---|---|---|---|---|---|
| Weight | rs1231281 | rs519757 | A | C | 19 | RASIP1 | Female only | True | 0.4032 | 0.3991 | 0.3879 | 0.3904 | 35804 | 26877 | 193627 | 166489 | 0.02456 | -0.001905 | 0.005211 | 0.006779 | 0.007367 | 0.008778 |
| Weight | rs1231281 | rs12979144 | A | G | 19 | RASIP1 | Female only | True | 0.4136 | 0.4103 | 0.4118 | 0.4145 | 35804 | 26877 | 193627 | 166489 | 0.03992 | 0.009721 | 0.009511 | 0.006429 | 0.007351 | 0.008723 |
| Weight | rs1231281 | rs12978499 | C | A | 19 | RASIP1 | Female only | True | 0.4136 | 0.4103 | 0.4118 | 0.4145 | 35804 | 26877 | 193627 | 166489 | 0.04001 | 0.009719 | 0.009525 | 0.006437 | 0.007351 | 0.008723 |
| Weight | rs1231281 | rs418464 | C | T | 19 | RASIP1 | Female only | True | 0.4909 | 0.4934 | 0.4975 | 0.5 | 35804 | 26877 | 193627 | 166489 | -0.03926 | -0.009164 | -0.01074 | -0.01004 | 0.007223 | 0.008528 |
| Weight | rs1231281 | 19:49239053 | C | T | 19 | RASIP1 | Female only | True | 0.4886 | 0.4914 | | | 35804 | 26877 | | | -0.03923 | -0.01053 | | | 0.007248 | 0.008566 |
| Weight | rs1231281 | rs1231281 | A | G | 19 | RASIP1 | Female only | True | 0.491 | 0.4935 | 0.4979 | 0.4995 | 35804 | 26877 | 193627 | 166489 | -0.04006 | -0.008644 | -0.01095 | -0.009859 | 0.007223 | 0.008528 |
| Weight | rs1231281 | rs380743 | A | G | 19 | RASIP1 | Female only | True | 0.4856 | 0.4884 | 0.4966 | 0.4938 | 35804 | 26877 | 193627 | 166489 | -0.04 | -0.009076 | -0.0113 | -0.01051 | 0.007271 | 0.008581 |
| Weight | rs1231281 | rs427248 | T | C | 19 | RASIP1 | Female only | True | 0.4856 | 0.4884 | 0.4965 | 0.4938 | 35804 | 26877 | 193627 | 166489 | -0.04005 | -0.008816 | -0.01119 | -0.01034 | 0.007271 | 0.008581 |
| Weight | rs1231281 | rs56066508 | T | C | 19 | RASIP1 | Female only | True | 0.4285 | 0.4249 | 0.427 | 0.4296 | 35804 | 26877 | 193627 | 166489 | 0.04022 | 0.01232 | 0.009028 | 0.007246 | 0.007313 | 0.008645 |
| Weight | rs1231281 | rs973579 | G | A | 19 | RASIP1 | Female only | True | 0.491 | 0.4934 | 0.4975 | 0.5 | 35804 | 26877 | 193627 | 166489 | -0.0395 | -0.009234 | -0.01073 | -0.01005 | 0.007221 | 0.008525 |
| Weight | rs1231281 | rs2307018 | A | C | 19 | RASIP1 | Female only | True | 0.4254 | 0.4217 | 0.4255 | 0.4279 | 35804 | 26877 | 193627 | 166489 | 0.03991 | 0.01186 | 0.008567 | 0.00741 | 0.007313 | 0.008652 |
| Weight | rs1231281 | rs2307019 | A | G | 19 | RASIP1 | Female only | True | 0.4288 | 0.425 | 0.4255 | 0.4279 | 35804 | 26877 | 193627 | 166489 | 0.0396 | 0.01156 | 0.008475 | 0.007423 | 0.007257 | 0.008586 |
| Weight | rs1231281 | rs838147 | A | G | 19 | RASIP1 | Female only | True | 0.4959 | 0.4979 | 0.4967 | 0.4993 | 35804 | 26877 | 193627 | 166489 | -0.03854 | -0.01035 | -0.01051 | -0.009531 | 0.007165 | 0.008471 |
| Weight | rs1231281 | rs12462324 | G | A | 19 | RASIP1 | Female only | True | 0.4247 | 0.4215 | 0.4251 | 0.4275 | 35804 | 26877 | 193627 | 166489 | 0.03933 | 0.01277 | 0.008401 | 0.007003 | 0.007311 | 0.008654 |
| Weight | rs1231281 | rs12460295 | G | T | 19 | RASIP1 | Female only | True | 0.4247 | 0.4215 | 0.4251 | 0.4275 | 35804 | 26877 | 193627 | 166489 | 0.03933 | 0.01274 | 0.008391 | 0.007027 | 0.007311 | 0.008654 |
| Weight | rs1231281 | rs11672046 | T | C | 19 | RASIP1 | Female only | True | 0.4247 | 0.4215 | 0.4252 | 0.4276 | 35804 | 26877 | 193627 | 166489 | 0.03916 | 0.01265 | 0.00839 | 0.006994 | 0.00731 | 0.008653 |
| Weight | rs1231281 | rs8108136 | T | C | 19 | RASIP1 | Female only | True | 0.4247 | 0.4215 | 0.4252 | 0.4276 | 35804 | 26877 | 193627 | 166489 | 0.03915 | 0.01265 | 0.008401 | 0.006968 | 0.00731 | 0.008653 |
| Weight | rs1231281 | rs8108468 | A | G | 19 | RASIP1 | Female only | True | 0.4281 | 0.4248 | 0.4252 | 0.4276 | 35804 | 26877 | 193627 | 166489 | 0.0389 | 0.01231 | 0.008404 | 0.006963 | 0.007255 | 0.008588 |
| Weight | rs1231281 | rs8111288 | A | G | 19 | RASIP1 | Female only | True | 0.4247 | 0.4215 | 0.4252 | 0.4276 | 35804 | 26877 | 193627 | 166489 | 0.03915 | 0.01264 | 0.008416 | 0.007012 | 0.00731 | 0.008653 |
| Weight | rs1231281 | rs8111399 | T | G | 19 | RASIP1 | Female only | True | 0.4281 | 0.4248 | 0.4252 | 0.4276 | 35804 | 26877 | 193627 | 166489 | 0.0389 | 0.01211 | 0.008408 | 0.007005 | 0.007255 | 0.008588 |

| trait | lead_snp | ID | ALLELE1 | ALLELE0 | CHR | Gene_Symbol | independent_loci_in_sex_P1e2 | in_embl_any_or_gene | A1FREQ_VUMC_female | A1FREQ_VUMC_male | A1FREQ_UKB_female | A1FREQ_UKB_male | N_VUMC_female | N_VUMC_male | N_UKB_female | N_UKB_male | BETA_VUMC_female | BETA_VUMC_male | BETA_UKB_female | BETA_UKB_male | SE_VUMC_female | SE_VUMC_male |
|---|---|---|---|---|---|---|---|---|---|---|---|---|---|---|---|---|---|---|---|---|---|---|
| Weight | rs1231281 | rs8111208 | C | A | 19 | RASIP1 | Female only | True | 0.4247 | 0.4215 | 0.4252 | 0.4276 | 35804 | 26877 | 193627 | 166489 | 0.03913 | 0.01247 | 0.008416 | 0.006995 | 0.007311 | 0.008653 |
| Weight | rs1231281 | rs11083948 | G | A | 19 | RASIP1 | Female only | True | 0.4247 | 0.4215 | 0.4251 | 0.4276 | 35804 | 26877 | 193627 | 166489 | 0.03914 | 0.01247 | 0.008436 | 0.00702 | 0.00731 | 0.008653 |
| Weight | rs1231281 | rs12611201 | A | G | 19 | RASIP1 | Female only | True | 0.4247 | 0.4215 | 0.4251 | 0.4276 | 35804 | 26877 | 193627 | 166489 | 0.03914 | 0.01247 | 0.008412 | 0.006984 | 0.00731 | 0.008653 |
| Weight | rs1231281 | rs12611203 | A | G | 19 | RASIP1 | Female only | True | 0.4249 | 0.4217 | 0.4252 | 0.4276 | 35804 | 26877 | 193627 | 166489 | 0.03913 | 0.01257 | 0.008409 | 0.006971 | 0.007312 | 0.008655 |
| Weight | rs1231281 | rs12611211 | A | G | 19 | RASIP1 | Female only | True | 0.4249 | 0.4216 | 0.4251 | 0.4276 | 35804 | 26877 | 193627 | 166489 | 0.03914 | 0.01258 | 0.008416 | 0.007012 | 0.007312 | 0.008656 |
| Weight | rs1231281 | rs12982115 | G | A | 19 | RASIP1 | Female only | True | 0.4247 | 0.4214 | 0.4251 | 0.4276 | 35804 | 26877 | 193627 | 166489 | 0.03898 | 0.01279 | 0.008442 | 0.006967 | 0.007312 | 0.008656 |
| Weight | rs1231281 | rs838145 | G | A | 19 | RASIP1 | Female only | True | 0.4495 | 0.4499 | 0.4596 | 0.4578 | 35804 | 26877 | 193627 | 166489 | -0.03536 | -0.01844 | -0.01108 | -0.007054 | 0.00722 | 0.008514 |
| Weight | rs1231281 | rs8102032 | T | C | 19 | RASIP1 | Female only | True | 0.425 | 0.4217 | 0.4254 | 0.4278 | 35804 | 26877 | 193627 | 166489 | 0.03927 | 0.01267 | 0.008392 | 0.007005 | 0.007312 | 0.008657 |
| Weight | rs1231281 | rs8104897 | A | C | 19 | RASIP1 | Female only | True | 0.425 | 0.4217 | 0.4254 | 0.4278 | 35804 | 26877 | 193627 | 166489 | 0.03929 | 0.01267 | 0.008392 | 0.007019 | 0.007312 | 0.008657 |
| Weight | rs1231281 | rs838133 | A | G | 19 | RASIP1 | Female only | True | 0.4449 | 0.4505 | 0.4531 | 0.4505 | 35804 | 26877 | 193627 | 166489 | -0.04003 | -0.01029 | -0.01111 | -0.01163 | 0.007248 | 0.008577 |
| QRS_DURATION | rs4685095 | rs11128689 | G | A | 3 | RP11-536I6.2 | Female only | False | 0.4427 | 0.4447 | 0.4445 | 0.4289 | 21446 | 19269 | 5542 | 5273 | 0.05381 | 0.02185 | 0.03269 | 0.05138 | 0.009679 | 0.01043 |
| QRS_DURATION | rs4685095 | rs1871856 | A | G | 3 | RP11-536I6.2 | Female only | False | 0.4405 | 0.4426 | 0.4442 | 0.4287 | 21446 | 19269 | 5542 | 5273 | 0.0539 | 0.02089 | 0.03341 | 0.05263 | 0.009647 | 0.0104 |
| QRS_DURATION | rs4685095 | rs1871855 | T | C | 3 | RP11-536I6.2 | Female only | False | 0.4425 | 0.4445 | 0.4442 | 0.4287 | 21446 | 19269 | 5542 | 5273 | 0.05419 | 0.02095 | 0.03341 | 0.05261 | 0.00968 | 0.01044 |
| QRS_DURATION | rs4685095 | rs13061705 | T | C | 3 | RP11-536I6.2 | Female only | False | 0.3243 | 0.3228 | 0.3245 | 0.3157 | 21446 | 19269 | 5542 | 5273 | 0.04583 | 0.01673 | 0.03466 | 0.05512 | 0.01024 | 0.01108 |
| QRS_DURATION | rs4685095 | rs4684190 | A | G | 3 | RP11-536I6.2 | Female only | False | 0.4424 | 0.4445 | 0.4443 | 0.4287 | 21446 | 19269 | 5542 | 5273 | 0.05411 | 0.02075 | 0.0334 | 0.05258 | 0.00968 | 0.01044 |
| QRS_DURATION | rs4685095 | rs4685095 | G | A | 3 | RP11-536I6.2 | Female only | False | 0.4425 | 0.4446 | 0.4442 | 0.4287 | 21446 | 19269 | 5542 | 5273 | 0.0542 | 0.02072 | 0.03347 | 0.05255 | 0.009679 | 0.01044 |
| QRS_DURATION | rs4685095 | rs4685096 | G | T | 3 | RP11-536I6.2 | Female only | False | 0.4426 | 0.4447 | 0.4443 | 0.4288 | 21446 | 19269 | 5542 | 5273 | 0.05413 | 0.02072 | 0.03377 | 0.05256 | 0.009679 | 0.01043 |
| QRS_DURATION | rs4685095 | rs9817692 | C | T | 3 | RP11-536I6.2 | Female only | False | 0.4426 | 0.4447 | 0.4445 | 0.4287 | 21446 | 19269 | 5542 | 5273 | 0.0541 | 0.02036 | 0.0338 | 0.05243 | 0.009683 | 0.01044 |
| QRS_DURATION | rs4685095 | rs9817854 | C | T | 3 | RP11-536I6.2 | Female only | False | 0.4426 | 0.4447 | 0.4443 | 0.4287 | 21446 | 19269 | 5542 | 5273 | 0.0541 | 0.02036 | 0.03337 | 0.05243 | 0.009683 | 0.01044 |
| QRS_DURATION | rs4685095 | rs9855606 | A | G | 3 | RP11-536I6.2 | Female only | False | 0.4425 | 0.4446 | 0.4443 | 0.4287 | 21446 | 19269 | 5542 | 5273 | 0.05403 | 0.02039 | 0.03339 | 0.05237 | 0.009684 | 0.01044 |
| QRS_DURATION | rs4685095 | rs4684191 | G | A | 3 | RP11-536I6.2 | Female only | False | 0.4425 | 0.4448 | 0.4443 | 0.4287 | 21446 | 19269 | 5542 | 5273 | 0.05402 | 0.02047 | 0.03371 | 0.05244 | 0.009684 | 0.01044 |
| QRS_DURATION | rs4685095 | rs34666135 | A | C | 3 | RP11-536I6.2 | Female only | False | 0.298 | 0.2971 | 0.2993 | 0.2895 | 21446 | 19269 | 5542 | 5273 | 0.04485 | 0.01806 | 0.02431 | 0.04705 | 0.01045 | 0.01131 |
| QRS_DURATION | rs4685095 | rs36039266 | C | T | 3 | RP11-536I6.2 | Female only | False | 0.2979 | 0.2971 | 0.2994 | 0.2898 | 21446 | 19269 | 5542 | 5273 | 0.04465 | 0.01761 | 0.02393 | 0.04794 | 0.01046 | 0.01131 |

| trait | lead_snp | ID | ALLELE1 | ALLELE0 | CHR | Gene_Symbol | independent_loci_in_sex_P1e2 | in_embl_any_or_gene | A1FREQ_VUMC_female | A1FREQ_VUMC_male | A1FREQ_UKB_female | A1FREQ_UKB_male | N_VUMC_female | N_VUMC_male | N_UKB_female | N_UKB_male | BETA_VUMC_female | BETA_VUMC_male | BETA_UKB_female | BETA_UKB_male | SE_VUMC_female | SE_VUMC_male |
|---|---|---|---|---|---|---|---|---|---|---|---|---|---|---|---|---|---|---|---|---|---|---|
| QRS_DURATION | rs4685095 | rs12714901 | G | T | 3 | RP11-536I6.2 | Female only | False | 0.2976 | 0.2968 | 0.2993 | 0.2894 | 21446 | 19269 | 5542 | 5273 | 0.04437 | 0.0166 | 0.02426 | 0.04735 | 0.01046 | 0.01132 |
| QRS_DURATION | rs9812912 | rs62241189 | A | G | 3 | SCN5A | Female only | True | 0.1664 | 0.1675 | 0.1675 | 0.1646 | 21446 | 19269 | 5542 | 5273 | -0.07539 | -0.03221 | -0.07498 | -0.09105 | 0.01274 | 0.01384 |
| QRS_DURATION | rs9812912 | rs9812912 | T | C | 3 | SCN5A | Female only | True | 0.155 | 0.1563 | 0.1574 | 0.1552 | 21446 | 19269 | 5542 | 5273 | -0.08527 | -0.03353 | -0.07733 | -0.08312 | 0.01318 | 0.01431 |
| QRS_DURATION | rs6764249 | rs6764249 | C | T | 3 | SCN10A | Female only | True | 0.1542 | 0.158 | 0.1468 | 0.1489 | 21446 | 19269 | 5542 | 5273 | -0.07665 | -0.02727 | -0.05496 | -0.03813 | 0.01312 | 0.01423 |
| QRS_DURATION | rs6764249 | rs62242856 | G | A | 3 | SCN10A | Female only | True | 0.1182 | 0.1218 | 0.116 | 0.1157 | 21446 | 19269 | 5542 | 5273 | -0.05675 | -0.01476 | -0.0354 | -0.002187 | 0.01478 | 0.01588 |
| QRS_DURATION | rs6764249 | rs62243860 | C | T | 3 | SCN10A | Female only | True | 0.1097 | 0.1125 | 0.1065 | 0.1072 | 21446 | 19269 | 5542 | 5273 | -0.05656 | -0.01373 | -0.04034 | -0.001344 | 0.01521 | 0.01633 |
| QRS_DURATION | rs6764249 | rs11129799 | T | C | 3 | SCN10A | Female only | True | 0.1992 | 0.1981 | 0.1947 | 0.1936 | 21446 | 19269 | 5542 | 5273 | -0.06045 | -0.03014 | -0.04368 | -0.008249 | 0.01203 | 0.01303 |
| QRS_DURATION | rs6764249 | rs7429946 | G | A | 3 | SCN10A | Female only | True | 0.1077 | 0.1108 | 0.1056 | 0.107 | 21446 | 19269 | 5542 | 5273 | -0.05925 | -0.01623 | -0.04438 | 0.004259 | 0.01533 | 0.01643 |
| QRS_DURATION | rs6764249 | rs6599244 | G | A | 3 | SCN10A | Female only | True | 0.1093 | 0.1122 | 0.1062 | 0.1068 | 21446 | 19269 | 5542 | 5273 | -0.05733 | -0.01448 | -0.04153 | -0.001357 | 0.01523 | 0.01635 |
| QRS_DURATION | rs6764249 | rs11715513 | A | G | 3 | SCN10A | Female only | True | 0.1074 | 0.1104 | 0.105 | 0.1055 | 21446 | 19269 | 5542 | 5273 | -0.0596 | -0.01454 | -0.04588 | 0.005411 | 0.01545 | 0.01655 |
| QRS_DURATION | rs7752142 | rs205915 | A | C | 6 | SLC35F1 | Female only | True | 0.3183 | 0.3183 | 0.3119 | 0.3127 | 21446 | 19269 | 5542 | 5273 | -0.04819 | -0.02209 | -0.01182 | -0.0594 | 0.0103 | 0.01124 |
| QRS_DURATION | rs7752142 | rs6936121 | A | G | 6 | SLC35F1 | Female only | True | 0.4034 | 0.4049 | 0.3977 | 0.4004 | 21446 | 19269 | 5542 | 5273 | -0.04635 | -0.02499 | -0.02544 | -0.06848 | 0.009649 | 0.01053 |
| QRS_DURATION | rs7752142 | rs12202829 | A | C | 6 | SLC35F1 | Female only | True | 0.313 | 0.313 | 0.312 | 0.3129 | 21446 | 19269 | 5542 | 5273 | -0.04869 | -0.02145 | -0.01079 | -0.05899 | 0.01022 | 0.01115 |
| QRS_DURATION | rs7752142 | rs9374723 | A | G | 6 | SLC35F1 | Female only | True | 0.312 | 0.312 | 0.312 | 0.3129 | 21446 | 19269 | 5542 | 5273 | -0.04841 | -0.02128 | -0.01093 | -0.05893 | 0.01026 | 0.0112 |
| QRS_DURATION | rs7752142 | rs9398485 | C | T | 6 | SLC35F1 | Female only | True | 0.3122 | 0.3121 | 0.3119 | 0.3129 | 21446 | 19269 | 5542 | 5273 | -0.04822 | -0.02102 | -0.01065 | -0.05854 | 0.01026 | 0.0112 |
| QRS_DURATION | rs7752142 | rs4259282 | T | C | 6 | SLC35F1 | Female only | True | 0.4553 | 0.4557 | 0.4443 | 0.4458 | 21446 | 19269 | 5542 | 5273 | -0.04813 | -0.01691 | -0.02672 | -0.07822 | 0.009571 | 0.01042 |
| QRS_DURATION | rs7752142 | rs2105020 | A | C | 6 | SLC35F1 | Female only | True | 0.3121 | 0.312 | 0.3119 | 0.3128 | 21446 | 19269 | 5542 | 5273 | -0.04815 | -0.02119 | -0.01066 | -0.05848 | 0.01026 | 0.0112 |
| trait | lead_snp | ID | ALLELE1 | ALLELE0 | CHR | Gene_Symbol | independent_loci_in_sex_P1e2 | in_embl_any_or_gene | A1FREQ_VUMC_female | A1FREQ_VUMC_male | A1FREQ_UKB_female | A1FREQ_UKB_male | N_VUMC_female | N_VUMC_male | N_UKB_female | N_UKB_male | BETA_VUMC_female | BETA_VUMC_male | BETA_UKB_female | BETA_UKB_male | SE_VUMC_female | SE_VUMC_male |
| QRS_DURATION | rs7752142 | rs2356077 | G | A | 6 | SLC35F1 | Female only | True | 0.3122 | 0.3118 | 0.312 | 0.3132 | 21446 | 19269 | 5542 | 5273 | -0.04894 | -0.02143 | -0.01172 | -0.05798 | 0.01027 | 0.0112 |
| QRS_DURATION | rs7752142 | rs7752142 | A | G | 6 | SLC35F1 | Female only | True | 0.4121 | 0.4113 | 0.4088 | 0.4081 | 21446 | 19269 | 5542 | 5273 | -0.05393 | -0.01561 | -0.02982 | -0.08403 | 0.009624 | 0.01051 |

| trait | lead_snp | ID | ALLELE1 | ALLELE0 | CHR | Gene_Symbol | independent_loci_in_sex_P1e2 | in_embl_any_or_gene | A1FREQ_VUMC_female | A1FREQ_VUMC_male | A1FREQ_UKB_female | A1FREQ_UKB_male | N_VUMC_female | N_VUMC_male | N_UKB_female | N_UKB_male | BETA_VUMC_female | BETA_VUMC_male | BETA_UKB_female | BETA_UKB_male | SE_VUMC_female | SE_VUMC_male |
|---|---|---|---|---|---|---|---|---|---|---|---|---|---|---|---|---|---|---|---|---|---|---|
| QRS_DURATION | rs7752142 | rs9387565 | A | G | 6 | SLC35F1 | Female only | True | 0.3122 | 0.3119 | 0.3115 | 0.3127 | 21446 | 19269 | 5542 | 5273 | -0.04741 | -0.02132 | -0.01064 | -0.05859 | 0.01027 | 0.0112 |
| QRS_DURATION | rs663651 | rs881433 | A | C | 18 | SETBP1 | Male only | True | 0.2992 | 0.3017 | 0.3059 | 0.306 | 21446 | 19269 | 5542 | 5273 | -0.02034 | -0.05857 | -0.03975 | -0.03225 | 0.01035 | 0.01131 |
| QRS_DURATION | rs663651 | rs881434 | T | C | 18 | SETBP1 | Male only | True | 0.2987 | 0.3015 | 0.3064 | 0.3063 | 21446 | 19269 | 5542 | 5273 | -0.02043 | -0.06003 | -0.04072 | -0.03216 | 0.01039 | 0.01134 |
| QRS_DURATION | rs663651 | rs34535142 | A | G | 18 | SETBP1 | Male only | True | 0.3055 | 0.3093 | 0.3117 | 0.3125 | 21446 | 19269 | 5542 | 5273 | -0.02189 | -0.06339 | -0.03649 | -0.03325 | 0.01035 | 0.01128 |
| QRS_DURATION | rs663651 | rs8088989 | A | G | 18 | SETBP1 | Male only | True | 0.3055 | 0.3093 | 0.3117 | 0.3125 | 21446 | 19269 | 5542 | 5273 | -0.02189 | -0.06341 | -0.03649 | -0.03325 | 0.01035 | 0.01128 |
| QRS_DURATION | rs663651 | rs663651 | G | A | 18 | SETBP1 | Male only | True | 0.4381 | 0.44 | 0.4345 | 0.4358 | 21446 | 19269 | 5542 | 5273 | -0.01458 | -0.06615 | -0.03264 | -0.02748 | 0.009587 | 0.01048 |
| QRS_DURATION | rs663651 | rs618472 | T | G | 18 | SETBP1 | Male only | True | 0.4441 | 0.4459 | 0.4414 | 0.4435 | 21446 | 19269 | 5542 | 5273 | -0.01339 | -0.06513 | -0.02977 | -0.03655 | 0.009579 | 0.01047 |
| QRS_DURATION | rs663651 | rs684656 | C | T | 18 | SETBP1 | Male only | True | 0.4466 | 0.4484 | 0.4416 | 0.4436 | 21446 | 19269 | 5542 | 5273 | -0.01324 | -0.06556 | -0.02971 | -0.03583 | 0.009618 | 0.01051 |
| QRS_DURATION | rs663651 | rs2361050 | T | G | 18 | SETBP1 | Male only | True | 0.3038 | 0.308 | 0.3094 | 0.3104 | 21446 | 19269 | 5542 | 5273 | -0.02275 | -0.06523 | -0.02258 | -0.03668 | 0.01036 | 0.01132 |
| QRS_DURATION | rs663651 | rs638807 | T | G | 18 | SETBP1 | Male only | True | 0.4465 | 0.4483 | 0.4415 | 0.4436 | 21446 | 19269 | 5542 | 5273 | -0.01298 | -0.0657 | -0.02971 | -0.03584 | 0.009617 | 0.01051 |
| QRS_DURATION | rs663651 | rs685553 | T | C | 18 | SETBP1 | Male only | True | 0.4465 | 0.4483 | 0.4415 | 0.4436 | 21446 | 19269 | 5542 | 5273 | -0.01312 | -0.06561 | -0.02972 | -0.03584 | 0.009617 | 0.01051 |
| QRS_DURATION | rs663651 | rs9807721 | A | G | 18 | SETBP1 | Male only | True | 0.3028 | 0.3071 | 0.3085 | 0.3098 | 21446 | 19269 | 5542 | 5273 | -0.02266 | -0.06572 | -0.02386 | -0.03701 | 0.01038 | 0.01133 |
| QRS_DURATION | rs663651 | rs628221 | T | C | 18 | SETBP1 | Male only | True | 0.4453 | 0.4471 | 0.4403 | 0.4428 | 21446 | 19269 | 5542 | 5273 | -0.01351 | -0.06595 | -0.0291 | -0.03658 | 0.009626 | 0.01052 |
| QRS_DURATION | rs663651 | rs687781 | A | G | 18 | SETBP1 | Male only | True | 0.4657 | 0.4627 | 0.4688 | 0.4637 | 21446 | 19269 | 5542 | 5273 | -0.005743 | 0.04152 | 0.03525 | 0.03655 | 0.009585 | 0.01046 |
| Chol | rs2927472 | rs6509173 | A | G | 19 | PVRL2 | Female only | True | 0.2048 | 0.2006 | 0.207 | 0.2074 | 19280 | 15405 | 184998 | 159280 | -0.08684 | -0.02842 | -0.09454 | -0.06688 | 0.01204 | 0.0132 |
| Chol | rs2927472 | rs4803760 | T | C | 19 | PVRL2 | Female only | True | 0.2017 | 0.1965 | 0.2043 | 0.2046 | 19280 | 15405 | 184998 | 159280 | -0.08882 | -0.03037 | -0.09741 | -0.06972 | 0.01196 | 0.01313 |
| Chol | rs2927472 | rs2927472 | T | C | 19 | PVRL2 | Female only | True | 0.1599 | 0.1598 | 0.164 | 0.1641 | 19280 | 15405 | 184998 | 159280 | -0.1012 | -0.02761 | -0.1227 | -0.08258 | 0.01307 | 0.01423 |
| Chol | rs2927472 | rs404935 | A | G | 19 | PVRL2 | Female only | True | 0.1629 | 0.1622 | 0.1689 | 0.1686 | 19280 | 15405 | 184998 | 159280 | -0.09588 | -0.02149 | -0.1162 | -0.08136 | 0.01295 | 0.01418 |
| Chol | rs2927472 | rs395908 | A | G | 19 | PVRL2 | Female only | True | 0.1632 | 0.1626 | 0.1691 | 0.1689 | 19280 | 15405 | 184998 | 159280 | -0.09658 | -0.02163 | -0.116 | -0.08142 | 0.01292 | 0.01415 |
| HDL.C | rs972283 | rs4731702 | T | C | 7 | RP11-36B6.1 | Female only | True | 0.486 | 0.4873 | 0.494 | 0.4965 | 18580 | 14828 | 168089 | 147044 | 0.0538 | 0.007558 | 0.04709 | 0.01964 | 0.01008 | 0.01087 |
| HDL.C | rs972283 | rs11979110 | T | C | 7 | RP11-36B6.1 | Female only | True | 0.4783 | 0.4794 | 0.4941 | 0.4964 | 18580 | 14828 | 168089 | 147044 | 0.05467 | 0.007443 | 0.04723 | 0.01953 | 0.01023 | 0.01104 |
| HDL.C | rs972283 | rs13230111 | G | A | 7 | RP11-36B6.1 | Female only | True | 0.4779 | 0.4791 | 0.4937 | 0.4959 | 18580 | 14828 | 168089 | 147044 | 0.05411 | 0.007625 | 0.04745 | 0.01946 | 0.01023 | 0.01104 |

| trait | lead_snp | ID | ALLELE1 | ALLELE0 | CHR | Gene_Symbol | independent_loci_in_sex_P1e2 | in_embl_any_or_gene | A1FREQ_VUMC_female | A1FREQ_VUMC_male | A1FREQ_UKB_female | A1FREQ_UKB_male | N_VUMC_female | N_VUMC_male | N_UKB_female | N_UKB_male | BETA_VUMC_female | BETA_VUMC_male | BETA_UKB_female | BETA_UKB_male | SE_VUMC_female | SE_VUMC_male |
|---|---|---|---|---|---|---|---|---|---|---|---|---|---|---|---|---|---|---|---|---|---|---|
| HDL.C | rs972283 | rs13233731 | A | G | 7 | RP11-36B6.1 | Female only | True | 0.4779 | 0.4791 | 0.4937 | 0.4959 | 18580 | 14828 | 168089 | 147044 | 0.0541 | 0.007625 | 0.04746 | 0.01946 | 0.01023 | 0.01104 |
| HDL.C | rs972283 | rs13234407 | A | G | 7 | RP11-36B6.1 | Female only | True | 0.4851 | 0.4866 | 0.4937 | 0.4959 | 18580 | 14828 | 168089 | 147044 | 0.05335 | 0.007423 | 0.04746 | 0.01945 | 0.01008 | 0.01087 |
| HDL.C | rs972283 | rs13228906 | A | G | 7 | RP11-36B6.1 | Female only | True | 0.4777 | 0.479 | 0.4935 | 0.4958 | 18580 | 14828 | 168089 | 147044 | 0.05435 | 0.00741 | 0.04752 | 0.01948 | 0.01023 | 0.01104 |
| HDL.C | rs972283 | rs17185445 | T | C | 7 | RP11-36B6.1 | Female only | True | 0.4777 | 0.4789 | 0.4934 | 0.4957 | 18580 | 14828 | 168089 | 147044 | 0.05402 | 0.007106 | 0.0475 | 0.01951 | 0.01023 | 0.01104 |
| HDL.C | rs972283 | rs3996352 | G | A | 7 | RP11-36B6.1 | Female only | True | 0.4776 | 0.4789 | 0.4933 | 0.4955 | 18580 | 14828 | 168089 | 147044 | 0.05406 | 0.006975 | 0.04754 | 0.0195 | 0.01023 | 0.01104 |
| HDL.C | rs972283 | rs34748838 | T | C | 7 | RP11-36B6.1 | Female only | True | 0.4736 | 0.4751 | 0.4897 | 0.4921 | 18580 | 14828 | 168089 | 147044 | 0.05594 | 0.004778 | 0.04683 | 0.019 | 0.01024 | 0.01106 |
| HDL.C | rs972283 | rs972283 | A | G | 7 | RP11-36B6.1 | Female only | True | 0.4807 | 0.4828 | 0.4895 | 0.492 | 18580 | 14828 | 168089 | 147044 | 0.05593 | 0.004255 | 0.0468 | 0.01861 | 0.0101 | 0.0109 |
| HDL.C | rs28623088 | rs1388943 | G | A | 8 | CSGALNACT1 | Male only | True | 0.4742 | 0.4738 | 0.4751 | 0.4722 | 18580 | 14828 | 168089 | 147044 | -0.01963 | -0.04477 | -0.02052 | -0.02189 | 0.01009 | 0.01104 |
| HDL.C | rs28623088 | rs17091113 | T | C | 8 | CSGALNACT1 | Male only | True | 0.369 | 0.3689 | 0.3638 | 0.366 | 18580 | 14828 | 168089 | 147044 | 0.02456 | 0.06463 | 0.02605 | 0.02893 | 0.01043 | 0.01138 |
| HDL.C | rs28623088 | rs28623088 | C | A | 8 | CSGALNACT1 | Male only | True | 0.3625 | 0.3628 | 0.3591 | 0.3616 | 18580 | 14828 | 168089 | 147044 | 0.02631 | 0.06688 | 0.02692 | 0.02957 | 0.0105 | 0.01147 |
| HDL.C | rs28623088 | rs17091133 | G | A | 8 | CSGALNACT1 | Male only | True | 0.3622 | 0.3622 | 0.359 | 0.3615 | 18580 | 14828 | 168089 | 147044 | 0.02602 | 0.0663 | 0.02656 | 0.02958 | 0.0105 | 0.01148 |
| HEIGHT | rs6743685 | rs12620853 | A | G | 2 | LTBP1 | Female only | True | 0.5014 | 0.5019 | 0.4955 | 0.4937 | 19052 | 12621 | 193785 | 166603 | -0.05536 | -0.02624 | -0.01807 | -0.02662 | 0.009545 | 0.01198 |
| HEIGHT | rs6743685 | rs6739323 | A | G | 2 | LTBP1 | Female only | True | 0.445 | 0.4474 | 0.4435 | 0.4425 | 19052 | 12621 | 193785 | 166603 | -0.04696 | -0.01588 | -0.01483 | -0.02221 | 0.009596 | 0.01204 |
| HEIGHT | rs6743685 | rs60692621 | T | C | 2 | LTBP1 | Female only | True | 0.4907 | 0.4897 | 0.4947 | 0.4966 | 19052 | 12621 | 193785 | 166603 | 0.05427 | 0.02399 | 0.01792 | 0.02611 | 0.009554 | 0.01199 |
| HEIGHT | rs6743685 | rs6743685 | A | C | 2 | LTBP1 | Female only | True | 0.4821 | 0.4853 | 0.4762 | 0.4747 | 19052 | 12621 | 193785 | 166603 | -0.05585 | -0.02644 | -0.01892 | -0.02654 | 0.009531 | 0.01197 |
| HEIGHT | rs6743685 | rs1947444 | A | G | 2 | LTBP1 | Female only | True | 0.4915 | 0.49 | 0.4937 | 0.4958 | 19052 | 12621 | 193785 | 166603 | 0.05432 | 0.02295 | 0.01828 | 0.02629 | 0.009521 | 0.01195 |
| HEIGHT | rs6743685 | rs17640469 | A | G | 2 | LTBP1 | Female only | True | 0.4459 | 0.4484 | 0.445 | 0.444 | 19052 | 12621 | 193785 | 166603 | -0.0475 | -0.01436 | -0.01511 | -0.02236 | 0.009583 | 0.01202 |
| HEIGHT | rs6743685 | rs6728094 | G | A | 2 | LTBP1 | Female only | True | 0.4663 | 0.4676 | 0.47 | 0.4683 | 19052 | 12621 | 193785 | 166603 | -0.04625 | -0.01284 | -0.01358 | -0.0218 | 0.009553 | 0.01195 |
| HEIGHT | rs6743685 | rs55911139 | C | A | 2 | LTBP1 | Female only | True | 0.4902 | 0.4918 | 0.4846 | 0.4832 | 19052 | 12621 | 193785 | 166603 | -0.05542 | -0.02319 | -0.01797 | -0.02677 | 0.009534 | 0.01195 |
| HEIGHT | rs6743685 | rs4670184 | C | T | 2 | LTBP1 | Female only | True | 0.4514 | 0.4534 | 0.4503 | 0.4493 | 19052 | 12621 | 193785 | 166603 | -0.04651 | -0.0148 | -0.01363 | -0.0223 | 0.009588 | 0.012 |
| HEIGHT | rs6743685 | rs4670186 | G | A | 2 | LTBP1 | Female only | True | 0.4902 | 0.4918 | 0.4846 | 0.4832 | 19052 | 12621 | 193785 | 166603 | -0.0554 | -0.02306 | -0.01801 | -0.02699 | 0.009533 | 0.01194 |
| HEIGHT | rs6743685 | rs75429464 | C | T | 2 | LTBP1 | Female only | True | 0.4828 | 0.4831 | | | 19052 | 12621 | | | -0.05546 | -0.02711 | | | 0.009535 | 0.01195 |
| HEIGHT | rs6743685 | rs34577456 | C | T | 2 | LTBP1 | Female only | True | 0.4768 | 0.4778 | 0.4732 | 0.4719 | 19052 | 12621 | 193785 | 166603 | -0.05514 | -0.02648 | -0.01731 | -0.02729 | 0.009551 | 0.01196 |
| HEIGHT | rs6743685 | rs62146677 | T | C | 2 | LTBP1 | Female only | True | 0.4952 | 0.4946 | 0.4991 | 0.499 | 19052 | 12621 | 193785 | 166603 | 0.05538 | 0.02449 | 0.01631 | 0.02711 | 0.00956 | 0.01198 |
| HEIGHT | rs6743685 | rs10495783 | C | T | 2 | LTBP1 | Female only | True | 0.4434 | 0.4446 | 0.4433 | 0.4422 | 19052 | 12621 | 193785 | 166603 | -0.04763 | -0.0178 | -0.01351 | -0.02243 | 0.009606 | 0.01204 |
| HEIGHT | rs6743685 | rs62146678 | T | C | 2 | LTBP1 | Female only | True | 0.4952 | 0.4945 | 0.4991 | 0.4991 | 19052 | 12621 | 193785 | 166603 | 0.05538 | 0.0245 | 0.0163 | 0.02709 | 0.00956 | 0.01198 |
| HEIGHT | rs6743685 | rs4670778 | T | C | 2 | LTBP1 | Female only | True | 0.495 | 0.4945 | 0.499 | 0.4991 | 19052 | 12621 | 193785 | 166603 | 0.05506 | 0.0245 | 0.01643 | 0.02701 | 0.009562 | 0.01198 |
| HEIGHT | rs6743685 | rs4670225 | A | G | 2 | LTBP1 | Female only | True | 0.4435 | 0.4446 | 0.4431 | 0.4421 | 19052 | 12621 | 193785 | 166603 | -0.04772 | -0.01812 | -0.0137 | -0.02233 | 0.009607 | 0.01204 |
| HEIGHT | rs6743685 | rs7595240 | T | C | 2 | LTBP1 | Female only | True | 0.495 | 0.4946 | 0.4991 | 0.4991 | 19052 | 12621 | 193785 | 166603 | 0.05518 | 0.02463 | 0.01646 | 0.027 | 0.009562 | 0.01197 |

| trait | lead_snp | ID | ALLELE1 | ALLELE0 | CHR | Gene_Symbol | independent_loci_in_sex_P1e2 | in_embl_any_or_gene | A1FREQ_VUMC_female | A1FREQ_VUMC_male | A1FREQ_UKB_female | A1FREQ_UKB_male | N_VUMC_female | N_VUMC_male | N_UKB_female | N_UKB_male | BETA_VUMC_female | BETA_VUMC_male | BETA_UKB_female | BETA_UKB_male | SE_VUMC_female | SE_VUMC_male |
|---|---|---|---|---|---|---|---|---|---|---|---|---|---|---|---|---|---|---|---|---|---|---|
| HEIGHT | rs6743685 | rs6543698 | C | T | 2 | LTBP1 | Female only | True | 0.4771 | 0.478 | 0.4731 | 0.4718 | 19052 | 12621 | 193785 | 166603 | -0.05526 | -0.02596 | -0.01735 | -0.02708 | 0.009556 | 0.01197 |
| HEIGHT | rs6743685 | rs6543699 | A | G | 2 | LTBP1 | Female only | True | 0.4951 | 0.4945 | 0.4992 | 0.499 | 19052 | 12621 | 193785 | 166603 | 0.05499 | 0.02456 | 0.01637 | 0.02691 | 0.009564 | 0.01198 |
| HEIGHT | rs6743685 | rs6543700 | T | C | 2 | LTBP1 | Female only | True | 0.4777 | 0.4782 | 0.4731 | 0.4718 | 19052 | 12621 | 193785 | 166603 | -0.05554 | -0.02488 | -0.01748 | -0.02717 | 0.009554 | 0.01196 |
| HEIGHT | rs6743685 | rs12611549 | A | G | 2 | LTBP1 | Female only | True | 0.4949 | 0.4944 | 0.4992 | 0.499 | 19052 | 12621 | 193785 | 166603 | 0.0546 | 0.02518 | 0.01646 | 0.02657 | 0.009558 | 0.01197 |
| HEIGHT | rs6743685 | rs12617948 | G | A | 2 | LTBP1 | Female only | True | 0.495 | 0.4945 | 0.4993 | 0.4989 | 19052 | 12621 | 193785 | 166603 | 0.05455 | 0.02526 | 0.0165 | 0.02657 | 0.009557 | 0.01197 |
| HEIGHT | rs6743685 | rs12617970 | C | A | 2 | LTBP1 | Female only | True | 0.495 | 0.4944 | 0.4993 | 0.4989 | 19052 | 12621 | 193785 | 166603 | 0.05455 | 0.02533 | 0.0165 | 0.02657 | 0.009557 | 0.01197 |
| HEIGHT | rs6743685 | rs726429 | G | A | 2 | LTBP1 | Female only | True | 0.4952 | 0.4946 | 0.4994 | 0.4988 | 19052 | 12621 | 193785 | 166603 | 0.05428 | 0.02551 | 0.01648 | 0.02665 | 0.00955 | 0.01196 |

| trait | lead_snp | ID | ALLELE1 | ALLELE0 | CHR | Gene_Symbol | independent_loci_in_sex_P1e2 | in_embl_any_or_gene | A1FREQ_VUMC_female | A1FREQ_VUMC_male | A1FREQ_UKB_female | A1FREQ_UKB_male | N_VUMC_female | N_VUMC_male | N_UKB_female | N_UKB_male | BETA_VUMC_female | BETA_VUMC_male | BETA_UKB_female | BETA_UKB_male | SE_VUMC_female | SE_VUMC_male |
|---|---|---|---|---|---|---|---|---|---|---|---|---|---|---|---|---|---|---|---|---|---|---|
| HEIGHT | rs6743685 | rs6751657 | T | C | 2 | LTBP1 | Female only | True | 0.4753 | 0.4763 | 0.4728 | 0.4715 | 19052 | 12621 | 193785 | 166603 | -0.05444 | -0.02698 | -0.01759 | -0.02683 | 0.009522 | 0.01193 |
| HEIGHT | rs6743685 | rs6707904 | G | A | 2 | LTBP1 | Female only | True | 0.4952 | 0.4945 | 0.4993 | 0.4988 | 19052 | 12621 | 193785 | 166603 | 0.05422 | 0.02533 | 0.01647 | 0.02666 | 0.00955 | 0.01196 |
| HEIGHT | rs6743685 | rs6736908 | C | T | 2 | LTBP1 | Female only | True | 0.4763 | 0.4775 | 0.4728 | 0.4715 | 19052 | 12621 | 193785 | 166603 | -0.05487 | -0.02692 | -0.01758 | -0.02682 | 0.009541 | 0.01195 |
| HEIGHT | rs6743685 | rs5006125 | G | A | 2 | LTBP1 | Female only | True | 0.4764 | 0.4774 | 0.4728 | 0.4715 | 19052 | 12621 | 193785 | 166603 | -0.05457 | -0.02692 | -0.01755 | -0.02682 | 0.009542 | 0.01195 |
| HEIGHT | rs6743685 | rs5006120 | T | C | 2 | LTBP1 | Female only | True | 0.4764 | 0.4775 | 0.4729 | 0.4716 | 19052 | 12621 | 193785 | 166603 | -0.05456 | -0.02692 | -0.01757 | -0.02684 | 0.009542 | 0.01195 |
| HEIGHT | rs6743685 | rs4670822 | T | C | 2 | LTBP1 | Female only | True | 0.4764 | 0.4775 | 0.4728 | 0.4715 | 19052 | 12621 | 193785 | 166603 | -0.05464 | -0.02692 | -0.01753 | -0.02687 | 0.009542 | 0.01195 |
| HEIGHT | rs6743685 | rs4670828 | A | G | 2 | LTBP1 | Female only | True | 0.4953 | 0.4945 | 0.4994 | 0.4988 | 19052 | 12621 | 193785 | 166603 | 0.05453 | 0.02531 | 0.0165 | 0.02668 | 0.00955 | 0.01196 |
| HEIGHT | rs6743685 | rs4670833 | G | A | 2 | LTBP1 | Female only | True | 0.4763 | 0.4776 | 0.4729 | 0.4715 | 19052 | 12621 | 193785 | 166603 | -0.05486 | -0.02712 | -0.01753 | -0.02689 | 0.009543 | 0.01195 |
| HEIGHT | rs6743685 | rs4670836 | G | A | 2 | LTBP1 | Female only | True | 0.4953 | 0.4945 | 0.4994 | 0.4988 | 19052 | 12621 | 193785 | 166603 | 0.05453 | 0.02531 | 0.0165 | 0.02673 | 0.00955 | 0.01196 |
| HEIGHT | rs6743685 | rs4670849 | A | G | 2 | LTBP1 | Female only | True | 0.4953 | 0.4945 | 0.4994 | 0.4988 | 19052 | 12621 | 193785 | 166603 | 0.05453 | 0.02532 | 0.01651 | 0.02673 | 0.00955 | 0.01196 |
| HEIGHT | rs6743685 | rs4670857 | A | G | 2 | LTBP1 | Female only | True | 0.4763 | 0.4774 | 0.4728 | 0.4714 | 19052 | 12621 | 193785 | 166603 | -0.05421 | -0.02675 | -0.01746 | -0.02685 | 0.009544 | 0.01195 |
| HEIGHT | rs6743685 | rs4670860 | G | A | 2 | LTBP1 | Female only | True | 0.4765 | 0.4775 | 0.4729 | 0.4715 | 19052 | 12621 | 193785 | 166603 | -0.05451 | -0.02694 | -0.01754 | -0.02688 | 0.009541 | 0.01195 |
| HEIGHT | rs6743685 | rs4670251 | G | A | 2 | LTBP1 | Female only | True | 0.4952 | 0.4945 | 0.4993 | 0.4988 | 19052 | 12621 | 193785 | 166603 | 0.05421 | 0.02532 | 0.01645 | 0.0267 | 0.00955 | 0.01196 |
| HEIGHT | rs6743685 | rs4670252 | A | G | 2 | LTBP1 | Female only | True | 0.4765 | 0.4775 | 0.4729 | 0.4715 | 19052 | 12621 | 193785 | 166603 | -0.05451 | -0.02694 | -0.01752 | -0.02685 | 0.009541 | 0.01195 |
| HEIGHT | rs6743685 | rs4670863 | C | A | 2 | LTBP1 | Female only | True | 0.4765 | 0.4775 | 0.4729 | 0.4716 | 19052 | 12621 | 193785 | 166603 | -0.05451 | -0.02693 | -0.01754 | -0.02688 | 0.009541 | 0.01195 |
| HEIGHT | rs6743685 | rs6759943 | T | C | 2 | LTBP1 | Female only | True | 0.4765 | 0.4775 | 0.4729 | 0.4715 | 19052 | 12621 | 193785 | 166603 | -0.05451 | -0.02694 | -0.01755 | -0.02689 | 0.009541 | 0.01195 |
| HEIGHT | rs6743685 | rs2367619 | G | A | 2 | LTBP1 | Female only | True | 0.4765 | 0.4775 | 0.4729 | 0.4715 | 19052 | 12621 | 193785 | 166603 | -0.05451 | -0.02701 | -0.01754 | -0.02688 | 0.009541 | 0.01195 |
| HEIGHT | rs6743685 | rs6720059 | A | G | 2 | LTBP1 | Female only | True | 0.4763 | 0.4769 | 0.4723 | 0.471 | 19052 | 12621 | 193785 | 166603 | -0.05441 | -0.02679 | -0.01716 | -0.02686 | 0.009542 | 0.01195 |
| HEIGHT | rs6743685 | rs6748806 | G | A | 2 | LTBP1 | Female only | True | 0.4763 | 0.4769 | 0.4723 | 0.471 | 19052 | 12621 | 193785 | 166603 | -0.05441 | -0.0268 | -0.01716 | -0.02689 | 0.009542 | 0.01195 |
| HEIGHT | rs6743685 | rs6720185 | A | C | 2 | LTBP1 | Female only | True | 0.4763 | 0.4769 | 0.4723 | 0.471 | 19052 | 12621 | 193785 | 166603 | -0.0544 | -0.02679 | -0.01715 | -0.02686 | 0.009542 | 0.01195 |

| trait | lead_snp | ID | ALLELE1 | ALLELE0 | CHR | Gene_Symbol | independent_loci_in_sex_P1e2 | in_embl_any_or_gene | A1FREQ_VUMC_female | A1FREQ_VUMC_male | A1FREQ_UKB_female | A1FREQ_UKB_male | N_VUMC_female | N_VUMC_male | N_UKB_female | N_UKB_male | BETA_VUMC_female | BETA_VUMC_male | BETA_UKB_female | BETA_UKB_male | SE_VUMC_female | SE_VUMC_male |
|---|---|---|---|---|---|---|---|---|---|---|---|---|---|---|---|---|---|---|---|---|---|---|
| HEIGHT | rs6743685 | rs6720405 | C | A | 2 | LTBP1 | Female only | True | 0.4954 | 0.4951 | 0.4999 | 0.4982 | 19052 | 12621 | 193785 | 166603 | 0.05409 | 0.02516 | 0.01605 | 0.02667 | 0.009551 | 0.01196 |
| HEIGHT | rs6743685 | rs10211514 | G | A | 2 | LTBP1 | Female only | True | 0.4762 | 0.477 | 0.4723 | 0.471 | 19052 | 12621 | 193785 | 166603 | -0.05433 | -0.02675 | -0.01718 | -0.02688 | 0.009541 | 0.01195 |
| HEIGHT | rs6743685 | rs17566780 | A | G | 2 | LTBP1 | Female only | True | 0.4954 | 0.4948 | 0.4997 | 0.4984 | 19052 | 12621 | 193785 | 166603 | 0.05381 | 0.02508 | 0.01599 | 0.02655 | 0.00955 | 0.01196 |
| HEIGHT | rs6743685 | rs1065324 | A | G | 2 | LTBP1 | Female only | True | 0.4965 | 0.496 | 0.4997 | 0.4984 | 19052 | 12621 | 193785 | 166603 | 0.0539 | 0.02513 | 0.01616 | 0.02666 | 0.009529 | 0.01193 |
| HEIGHT | rs6743685 | rs6707911 | G | A | 2 | LTBP1 | Female only | True | 0.4428 | 0.4437 | 0.4419 | 0.441 | 19052 | 12621 | 193785 | 166603 | -0.04768 | -0.01937 | -0.0134 | -0.02181 | 0.009604 | 0.01203 |
| HEIGHT | rs6743685 | rs6707962 | C | A | 2 | LTBP1 | Female only | True | 0.4751 | 0.476 | 0.4724 | 0.4711 | 19052 | 12621 | 193785 | 166603 | -0.05442 | -0.02693 | -0.01718 | -0.0267 | 0.009524 | 0.01192 |
| HEIGHT | rs6743685 | rs6708836 | G | A | 2 | LTBP1 | Female only | True | 0.4428 | 0.4436 | 0.4416 | 0.4407 | 19052 | 12621 | 193785 | 166603 | -0.04721 | -0.0182 | -0.01362 | -0.02176 | 0.009604 | 0.01203 |
| HEIGHT | rs6743685 | rs56282338 | A | G | 2 | LTBP1 | Female only | True | 0.4955 | 0.495 | 0.4998 | 0.4984 | 19052 | 12621 | 193785 | 166603 | 0.05422 | 0.0254 | 0.01607 | 0.02658 | 0.009552 | 0.01196 |
| HEIGHT | rs6743685 | rs6712473 | G | T | 2 | LTBP1 | Female only | True | 0.4761 | 0.477 | 0.4724 | 0.4711 | 19052 | 12621 | 193785 | 166603 | -0.05459 | -0.02726 | -0.01725 | -0.02678 | 0.009543 | 0.01195 |
| HEIGHT | rs6743685 | rs4542826 | G | A | 2 | LTBP1 | Female only | True | 0.4425 | 0.4433 | 0.4415 | 0.4407 | 19052 | 12621 | 193785 | 166603 | -0.04785 | -0.01887 | -0.01352 | -0.02173 | 0.009608 | 0.01203 |
| HEIGHT | rs6743685 | rs17641062 | G | A | 2 | LTBP1 | Female only | True | 0.4955 | 0.495 | 0.4998 | 0.4983 | 19052 | 12621 | 193785 | 166603 | 0.05422 | 0.02531 | 0.01613 | 0.02658 | 0.009552 | 0.01196 |
| HEIGHT | rs6743685 | rs62146704 | G | A | 2 | LTBP1 | Female only | True | 0.4425 | 0.4433 | 0.4415 | 0.4407 | 19052 | 12621 | 193785 | 166603 | -0.04786 | -0.01886 | -0.01358 | -0.02174 | 0.009608 | 0.01203 |
| HEIGHT | rs6743685 | rs17641115 | C | T | 2 | LTBP1 | Female only | True | 0.4425 | 0.4432 | 0.4414 | 0.4406 | 19052 | 12621 | 193785 | 166603 | -0.04762 | -0.01869 | -0.01356 | -0.02172 | 0.009618 | 0.01204 |
| HEIGHT | rs6743685 | rs4670928 | C | T | 2 | LTBP1 | Female only | True | 0.4998 | 0.499 | 0.4956 | 0.4938 | 19052 | 12621 | 193785 | 166603 | 0.05558 | 0.02318 | 0.01615 | 0.02627 | 0.009568 | 0.01195 |
| HEIGHT | rs6743685 | rs55957962 | A | G | 2 | LTBP1 | Female only | True | 0.4035 | 0.4044 | 0.3971 | 0.3962 | 19052 | 12621 | 193785 | 166603 | -0.0476 | -0.005768 | -0.004431 | -0.01218 | 0.009771 | 0.01223 |
| HEIGHT | rs6743685 | rs7584473 | G | T | 2 | LTBP1 | Female only | True | 0.4851 | 0.4845 | 0.4885 | 0.4894 | 19052 | 12621 | 193785 | 166603 | 0.05076 | 0.02055 | 0.01621 | 0.02587 | 0.009585 | 0.01199 |
| HEIGHT | rs6743685 | rs17641180 | G | A | 2 | LTBP1 | Female only | True | 0.492 | 0.4916 | 0.4953 | 0.4965 | 19052 | 12621 | 193785 | 166603 | 0.05437 | 0.02118 | 0.01579 | 0.02509 | 0.009577 | 0.01199 |
| HEIGHT | rs6743685 | rs9308931 | G | T | 2 | LTBP1 | Female only | True | 0.4466 | 0.4476 | 0.4468 | 0.4465 | 19052 | 12621 | 193785 | 166603 | -0.04718 | -0.01493 | -0.01302 | -0.02023 | 0.009639 | 0.01207 |
| HEIGHT | rs6743685 | rs60000782 | G | A | 2 | LTBP1 | Female only | True | 0.4917 | 0.4914 | 0.4952 | 0.4964 | 19052 | 12621 | 193785 | 166603 | 0.05404 | 0.02055 | 0.01586 | 0.02515 | 0.009576 | 0.01199 |
| HEIGHT | rs6743685 | rs7423390 | T | C | 2 | LTBP1 | Female only | True | 0.4541 | 0.4547 | 0.454 | 0.454 | 19052 | 12621 | 193785 | 166603 | -0.04326 | -0.01373 | -0.01395 | -0.0211 | 0.00963 | 0.01205 |
| HEIGHT | rs6743685 | rs964703 | C | A | 2 | LTBP1 | Female only | True | 0.4877 | 0.4884 | 0.485 | 0.4845 | 19052 | 12621 | 193785 | 166603 | -0.05035 | -0.02195 | -0.01767 | -0.02629 | 0.009561 | 0.01197 |
| HEIGHT | rs6743685 | rs964702 | T | C | 2 | LTBP1 | Female only | True | 0.4468 | 0.4473 | 0.4466 | 0.4464 | 19052 | 12621 | 193785 | 166603 | -0.04766 | -0.01472 | -0.01333 | -0.02045 | 0.009639 | 0.01207 |
| HEIGHT | rs6743685 | rs2367623 | A | C | 2 | LTBP1 | Female only | True | 0.4716 | 0.4706 | 0.4733 | 0.4743 | 19052 | 12621 | 193785 | 166603 | 0.05012 | 0.01485 | 0.01684 | 0.02593 | 0.009588 | 0.01197 |
| HEIGHT | rs6743685 | rs996103 | T | C | 2 | LTBP1 | Female only | True | 0.4716 | 0.4706 | 0.4733 | 0.4743 | 19052 | 12621 | 193785 | 166603 | 0.05005 | 0.01486 | 0.01688 | 0.02584 | 0.009588 | 0.01197 |
| HEIGHT | rs6743685 | rs7580348 | T | C | 2 | LTBP1 | Female only | True | 0.4787 | 0.4777 | 0.4806 | 0.4818 | 19052 | 12621 | 193785 | 166603 | 0.05385 | 0.01487 | 0.01627 | 0.02506 | 0.009583 | 0.01197 |
| HEIGHT | rs6743685 | rs6705772 | C | A | 2 | LTBP1 | Female only | True | 0.4918 | 0.4915 | 0.4949 | 0.4961 | 19052 | 12621 | 193785 | 166603 | 0.05443 | 0.02119 | 0.01582 | 0.02515 | 0.009577 | 0.01199 |
| HEIGHT | rs6743685 | rs58774073 | T | C | 2 | LTBP1 | Female only | True | 0.4869 | 0.4869 | 0.4896 | 0.4909 | 19052 | 12621 | 193785 | 166603 | 0.05379 | 0.02437 | 0.01589 | 0.0255 | 0.009577 | 0.01201 |
| HEIGHT | rs6743685 | rs58920936 | C | T | 2 | LTBP1 | Female only | True | 0.4802 | 0.4804 | 0.4805 | 0.4794 | 19052 | 12621 | 193785 | 166603 | -0.04568 | -0.01551 | -0.01196 | -0.02027 | 0.009588 | 0.01203 |
| HEIGHT | rs6743685 | rs4671010 | C | A | 2 | LTBP1 | Female only | True | 0.491 | 0.4907 | 0.494 | 0.4953 | 19052 | 12621 | 193785 | 166603 | 0.05486 | 0.02159 | 0.01597 | 0.02528 | 0.009577 | 0.01199 |

| trait | lead_snp | ID | ALLELE1 | ALLELE0 | CHR | Gene_Symbol | independent_loci_in_sex_P1e2 | in_embl_any_or_gene | A1FREQ_VUMC_female | A1FREQ_VUMC_male | A1FREQ_UKB_female | A1FREQ_UKB_male | N_VUMC_female | N_VUMC_male | N_UKB_female | N_UKB_male | BETA_VUMC_female | BETA_VUMC_male | BETA_UKB_female | BETA_UKB_male | SE_VUMC_female | SE_VUMC_male |
|---|---|---|---|---|---|---|---|---|---|---|---|---|---|---|---|---|---|---|---|---|---|---|
| HEIGHT | rs6743685 | rs2367624 | T | C | 2 | LTBP1 | Female only | True | 0.4479 | 0.4487 | 0.4482 | 0.4477 | 19052 | 12621 | 193785 | 166603 | -0.04759 | -0.01464 | -0.01324 | -0.02051 | 0.009637 | 0.01207 |
| HEIGHT | rs6743685 | rs1377947 | T | C | 2 | LTBP1 | Female only | True | 0.448 | 0.4488 | 0.4483 | 0.4478 | 19052 | 12621 | 193785 | 166603 | -0.04751 | -0.01483 | -0.0132 | -0.0205 | 0.009636 | 0.01207 |
| HEIGHT | rs6743685 | rs2061027 | G | A | 2 | LTBP1 | Female only | True | 0.491 | 0.4907 | 0.494 | 0.4953 | 19052 | 12621 | 193785 | 166603 | 0.0549 | 0.02156 | 0.01592 | 0.02531 | 0.009578 | 0.012 |
| HEIGHT | rs6743685 | rs2061026 | A | G | 2 | LTBP1 | Female only | True | 0.4931 | 0.4945 | 0.4914 | 0.4907 | 19052 | 12621 | 193785 | 166603 | -0.05412 | -0.01761 | -0.01698 | -0.02521 | 0.009567 | 0.01195 |
| HEIGHT | rs6743685 | rs1377946 | A | G | 2 | LTBP1 | Female only | True | 0.4529 | 0.4546 | 0.4525 | 0.4529 | 19052 | 12621 | 193785 | 166603 | -0.04319 | -0.003732 | -0.01176 | -0.0184 | 0.009633 | 0.01199 |
| HEIGHT | rs6743685 | rs4671016 | T | C | 2 | LTBP1 | Female only | True | 0.4472 | 0.4493 | 0.4465 | 0.4471 | 19052 | 12621 | 193785 | 166603 | -0.04443 | -0.001428 | -0.01143 | -0.01742 | 0.009651 | 0.01202 |
| HEIGHT | rs6743685 | rs1454379 | G | A | 2 | LTBP1 | Female only | True | 0.4821 | 0.4843 | 0.4791 | 0.4794 | 19052 | 12621 | 193785 | 166603 | -0.05263 | -0.009763 | -0.01481 | -0.02194 | 0.009564 | 0.01192 |
| HEIGHT | rs6743685 | rs1454378 | G | A | 2 | LTBP1 | Female only | True | 0.4894 | 0.4878 | 0.4931 | 0.4934 | 19052 | 12621 | 193785 | 166603 | 0.05202 | 0.00683 | 0.01381 | 0.02185 | 0.00957 | 0.01192 |
| HEIGHT | rs6743685 | rs17641906 | G | A | 2 | LTBP1 | Female only | True | 0.4887 | 0.4873 | 0.4927 | 0.493 | 19052 | 12621 | 193785 | 166603 | 0.05155 | 0.005814 | 0.01405 | 0.02208 | 0.009568 | 0.01192 |
| HEIGHT | rs6743685 | rs2290449 | G | A | 2 | LTBP1 | Female only | True | 0.4497 | 0.4521 | 0.45 | 0.4506 | 19052 | 12621 | 193785 | 166603 | -0.04383 | 2.52e-04 | -0.0109 | -0.01689 | 0.009631 | 0.01201 |
| HEIGHT | rs6743685 | rs11124310 | T | C | 2 | LTBP1 | Female only | True | 0.4784 | 0.4771 | 0.4814 | 0.4816 | 19052 | 12621 | 193785 | 166603 | 0.04544 | 0.008167 | 0.0146 | 0.02309 | 0.009573 | 0.01194 |
| trait | lead_snp | ID | ALLELE1 | ALLELE0 | CHR | Gene_Symbol | independent_loci_in_sex_P1e2 | in_embl_any_or_gene | A1FREQ_VUMC_female | A1FREQ_VUMC_male | A1FREQ_UKB_female | A1FREQ_UKB_male | N_VUMC_female | N_VUMC_male | N_UKB_female | N_UKB_male | BETA_VUMC_female | BETA_VUMC_male | BETA_UKB_female | BETA_UKB_male | SE_VUMC_female | SE_VUMC_male |
| HEIGHT | rs6743685 | rs17568051 | A | G | 2 | LTBP1 | Female only | True | 0.4854 | 0.4842 | 0.4888 | 0.4892 | 19052 | 12621 | 193785 | 166603 | 0.04927 | 0.008171 | 0.01398 | 0.02234 | 0.009567 | 0.01194 |
| HEIGHT | rs6743685 | rs4671024 | A | G | 2 | LTBP1 | Female only | True | 0.4824 | 0.4808 | 0.4856 | 0.4857 | 19052 | 12621 | 193785 | 166603 | 0.04626 | 0.005671 | 0.01473 | 0.02256 | 0.009566 | 0.01191 |
| HEIGHT | rs6743685 | rs7608222 | T | C | 2 | LTBP1 | Female only | True | 0.4825 | 0.481 | 0.4858 | 0.4859 | 19052 | 12621 | 193785 | 166603 | 0.04674 | 0.00624 | 0.01463 | 0.02251 | 0.009564 | 0.01191 |
| HEIGHT | rs6743685 | rs7608535 | A | G | 2 | LTBP1 | Female only | True | 0.4898 | 0.4883 | 0.4932 | 0.4935 | 19052 | 12621 | 193785 | 166603 | 0.05062 | 0.006278 | 0.01396 | 0.02163 | 0.009561 | 0.01191 |
| HEIGHT | rs6743685 | rs2290448 | T | C | 2 | LTBP1 | Female only | True | 0.4836 | 0.482 | 0.4858 | 0.4859 | 19052 | 12621 | 193785 | 166603 | 0.04642 | 0.006222 | 0.01466 | 0.02249 | 0.009545 | 0.01189 |
| HEIGHT | rs6743685 | rs17642074 | G | A | 2 | LTBP1 | Female only | True | 0.4833 | 0.4817 | 0.4862 | 0.4864 | 19052 | 12621 | 193785 | 166603 | 0.04685 | 0.006577 | 0.01492 | 0.02241 | 0.009563 | 0.01191 |
| HEIGHT | rs6743685 | rs56163907 | G | A | 2 | LTBP1 | Female only | True | 0.4905 | 0.489 | 0.4938 | 0.4941 | 19052 | 12621 | 193785 | 166603 | 0.051 | 0.007153 | 0.01418 | 0.02164 | 0.00956 | 0.01191 |
| HEIGHT | rs6743685 | rs1454376 | G | A | 2 | LTBP1 | Female only | True | 0.4802 | 0.4826 | 0.4786 | 0.4788 | 19052 | 12621 | 193785 | 166603 | -0.05116 | -0.009186 | -0.01522 | -0.02174 | 0.009535 | 0.01189 |
| HEIGHT | rs6743685 | rs62146732 | A | G | 2 | LTBP1 | Female only | True | 0.4829 | 0.4813 | 0.486 | 0.4861 | 19052 | 12621 | 193785 | 166603 | 0.04671 | 0.006395 | 0.01475 | 0.0226 | 0.009564 | 0.01191 |
| HEIGHT | rs6743685 | rs13021881 | T | C | 2 | LTBP1 | Female only | True | 0.4807 | 0.4823 | 0.4786 | 0.4789 | 19052 | 12621 | 193785 | 166603 | -0.05047 | -0.009784 | -0.01501 | -0.02204 | 0.009563 | 0.01192 |
| HEIGHT | rs6743685 | rs2367508 | A | G | 2 | LTBP1 | Female only | True | 0.4835 | 0.4865 | 0.4821 | 0.4821 | 19052 | 12621 | 193785 | 166603 | -0.04811 | -0.009277 | -0.01462 | -0.02127 | 0.009574 | 0.01191 |
| HEIGHT | rs6743685 | rs7590965 | A | C | 2 | LTBP1 | Female only | True | 0.4715 | 0.4721 | 0.4653 | 0.4646 | 19052 | 12621 | 193785 | 166603 | -0.04559 | 0.008446 | -0.003276 | -0.01071 | 0.009576 | 0.01192 |
| HEIGHT | rs6743685 | rs2367509 | A | G | 2 | LTBP1 | Female only | True | 0.4449 | 0.4471 | 0.4382 | 0.4382 | 19052 | 12621 | 193785 | 166603 | -0.04482 | 0.004518 | -0.004338 | -0.01094 | 0.00957 | 0.01196 |
| HEIGHT | rs6743685 | rs62146734 | A | G | 2 | LTBP1 | Female only | True | 0.4825 | 0.4805 | 0.4836 | 0.4839 | 19052 | 12621 | 193785 | 166603 | 0.04278 | 0.007402 | 0.01385 | 0.02111 | 0.009572 | 0.0119 |
| HEIGHT | rs6743685 | rs6750739 | A | G | 2 | LTBP1 | Female only | True | 0.4184 | 0.4209 | 0.4136 | 0.414 | 19052 | 12621 | 193785 | 166603 | -0.03863 | 0.007678 | -1.45e-04 | -0.007438 | 0.009675 | 0.01208 |
| HEIGHT | rs6743685 | rs12151817 | T | C | 2 | LTBP1 | Female only | True | 0.4202 | 0.4229 | 0.414 | 0.4144 | 19052 | 12621 | 193785 | 166603 | -0.03817 | 0.007278 | 1.05e-04 | -0.00766 | 0.009704 | 0.0121 |

| trait | lead_snp | ID | ALLELE1 | ALLELE0 | CHR | Gene_Symbol | independent_loci_in_sex_P1e2 | in_embl_any_or_gene | A1FREQ_VUMC_female | A1FREQ_VUMC_male | A1FREQ_UKB_female | A1FREQ_UKB_male | N_VUMC_female | N_VUMC_male | N_UKB_female | N_UKB_male | BETA_VUMC_female | BETA_VUMC_male | BETA_UKB_female | BETA_UKB_male | SE_VUMC_female | SE_VUMC_male |
|---|---|---|---|---|---|---|---|---|---|---|---|---|---|---|---|---|---|---|---|---|---|---|
| HEIGHT | rs11125609 | rs11125609 | C | T | 2 | EFEMP1 | Male only | True | 0.2911 | 0.2887 | 0.2857 | 0.2852 | 19052 | 12621 | 193785 | 166603 | -0.008428 | -0.07237 | -0.03518 | -0.03168 | 0.01056 | 0.01314 |
| HEIGHT | rs11125609 | rs1346791 | C | T | 2 | EFEMP1 | Male only | True | 0.3875 | 0.385 | 0.3751 | 0.3759 | 19052 | 12621 | 193785 | 166603 | -0.01703 | -0.0608 | -0.03395 | -0.03168 | 0.009829 | 0.01225 |
| HEIGHT | rs11125609 | rs3791666 | C | T | 2 | EFEMP1 | Male only | True | 0.3072 | 0.3066 | 0.2998 | 0.3 | 19052 | 12621 | 193785 | 166603 | -0.005332 | -0.05845 | -0.02943 | -0.02528 | 0.01038 | 0.01289 |
| HEIGHT | rs11049356 | rs61920200 | A | G | 12 | CCDC91 | Female only | True | 0.338 | 0.3381 | 0.341 | 0.3416 | 19052 | 12621 | 193785 | 166603 | -0.03604 | 0.01111 | -0.02757 | -0.03141 | 0.01011 | 0.01265 |
| HEIGHT | rs11049356 | 12:28239325 | A | G | 12 | CCDC91 | Female only | True | 0.3373 | 0.3376 | | | 19052 | 12621 | | | -0.03565 | 0.01106 | | | 0.01015 | 0.0127 |
| HEIGHT | rs11049356 | rs10771405 | T | C | 12 | CCDC91 | Female only | True | 0.3437 | 0.3442 | 0.3507 | 0.3512 | 19052 | 12621 | 193785 | 166603 | -0.03793 | 0.01121 | -0.02867 | -0.03116 | 0.01011 | 0.01268 |
| HEIGHT | rs11049356 | rs10843095 | G | A | 12 | CCDC91 | Female only | True | 0.3427 | 0.3438 | 0.35 | 0.3508 | 19052 | 12621 | 193785 | 166603 | -0.04339 | 0.01099 | -0.03285 | -0.03256 | 0.01012 | 0.01268 |
| HEIGHT | rs11049356 | rs28380759 | T | G | 12 | CCDC91 | Female only | True | 0.3414 | 0.3429 | 0.3493 | 0.3501 | 19052 | 12621 | 193785 | 166603 | -0.04348 | 0.01107 | -0.0329 | -0.0326 | 0.01014 | 0.0127 |
| HEIGHT | rs11049356 | rs11049352 | A | G | 12 | CCDC91 | Female only | True | 0.2959 | 0.2987 | 0.3034 | 0.3035 | 19052 | 12621 | 193785 | 166603 | -0.0559 | 0.01232 | -0.04008 | -0.03728 | 0.01053 | 0.01312 |
| HEIGHT | rs11049356 | rs10843097 | T | C | 12 | CCDC91 | Female only | True | 0.2959 | 0.2987 | 0.3034 | 0.3035 | 19052 | 12621 | 193785 | 166603 | -0.05592 | 0.01235 | -0.04008 | -0.0373 | 0.01053 | 0.01312 |
| HEIGHT | rs11049356 | rs10843099 | G | A | 12 | CCDC91 | Female only | True | 0.2964 | 0.2996 | 0.3041 | 0.3042 | 19052 | 12621 | 193785 | 166603 | -0.05529 | 0.01415 | -0.03997 | -0.03752 | 0.01051 | 0.0131 |
| HEIGHT | rs11049356 | rs10843100 | C | T | 12 | CCDC91 | Female only | True | 0.2967 | 0.3001 | 0.3044 | 0.3044 | 19052 | 12621 | 193785 | 166603 | -0.05465 | 0.01337 | -0.04004 | -0.03759 | 0.01051 | 0.01309 |
| HEIGHT | rs11049356 | rs11049356 | T | C | 12 | CCDC91 | Female only | True | 0.3204 | 0.3237 | 0.3251 | 0.3248 | 19052 | 12621 | 193785 | 166603 | -0.05957 | 0.01204 | -0.03859 | -0.03681 | 0.0103 | 0.01286 |
| HEIGHT | rs11049356 | rs899505 | A | C | 12 | CCDC91 | Female only | True | 0.3149 | 0.3184 | 0.3194 | 0.3193 | 19052 | 12621 | 193785 | 166603 | -0.0582 | 0.011 | -0.03875 | -0.03621 | 0.01034 | 0.01288 |
| HEIGHT | rs11049356 | rs10843102 | A | G | 12 | CCDC91 | Female only | True | 0.2964 | 0.2994 | 0.3043 | 0.3044 | 19052 | 12621 | 193785 | 166603 | -0.05483 | 0.0137 | -0.04019 | -0.03742 | 0.01053 | 0.01311 |
| HEIGHT | rs11049356 | rs10843104 | C | T | 12 | CCDC91 | Female only | True | 0.305 | 0.3094 | 0.3076 | 0.3072 | 19052 | 12621 | 193785 | 166603 | -0.05087 | 0.007763 | -0.0398 | -0.03906 | 0.01039 | 0.01293 |
| HEIGHT | rs11049356 | rs11049359 | C | T | 12 | CCDC91 | Female only | True | 0.3 | 0.3038 | 0.3051 | 0.3047 | 19052 | 12621 | 193785 | 166603 | -0.05482 | 0.01229 | -0.03995 | -0.03872 | 0.01046 | 0.01304 |
| HEIGHT | rs11049356 | rs11049360 | T | C | 12 | CCDC91 | Female only | True | 0.3006 | 0.3041 | 0.3057 | 0.3053 | 19052 | 12621 | 193785 | 166603 | -0.05285 | 0.01046 | -0.03998 | -0.0387 | 0.01045 | 0.01303 |
| HEIGHT | rs11049356 | rs7305286 | G | A | 12 | CCDC91 | Female only | True | 0.3006 | 0.3041 | 0.3057 | 0.3053 | 19052 | 12621 | 193785 | 166603 | -0.05277 | 0.01045 | -0.03997 | -0.03865 | 0.01045 | 0.01303 |
| HEIGHT | rs11049356 | rs10219470 | T | C | 12 | CCDC91 | Female only | True | 0.3178 | 0.3211 | 0.3181 | 0.3177 | 19052 | 12621 | 193785 | 166603 | -0.05687 | 0.01036 | -0.03997 | -0.03755 | 0.01029 | 0.01283 |
| HEIGHT | rs11049356 | rs10843110 | G | T | 12 | CCDC91 | Female only | True | 0.3176 | 0.3209 | 0.318 | 0.3176 | 19052 | 12621 | 193785 | 166603 | -0.05723 | 0.01039 | -0.03995 | -0.03774 | 0.0103 | 0.01284 |
| HEIGHT | rs11049356 | rs11049363 | A | G | 12 | CCDC91 | Female only | True | 0.2993 | 0.3024 | 0.3037 | 0.3034 | 19052 | 12621 | 193785 | 166603 | -0.05327 | 0.00937 | -0.04033 | -0.03924 | 0.01046 | 0.01305 |
| HEIGHT | rs11049356 | rs10843111 | T | C | 12 | CCDC91 | Female only | True | 0.3134 | 0.316 | 0.3162 | 0.3162 | 19052 | 12621 | 193785 | 166603 | -0.05597 | 0.008647 | -0.0401 | -0.03882 | 0.01035 | 0.0129 |
| HEIGHT | rs11049356 | rs10843112 | G | T | 12 | CCDC91 | Female only | True | 0.3154 | 0.3182 | 0.319 | 0.3189 | 19052 | 12621 | 193785 | 166603 | -0.05429 | 0.00822 | -0.03991 | -0.03861 | 0.01033 | 0.01288 |
| HEIGHT | rs11049356 | rs11049369 | G | A | 12 | CCDC91 | Female only | True | 0.3103 | 0.3137 | 0.3144 | 0.3142 | 19052 | 12621 | 193785 | 166603 | -0.05467 | 0.006917 | -0.04098 | -0.03979 | 0.01038 | 0.01295 |
| HEIGHT | rs11049356 | rs11049370 | T | C | 12 | CCDC91 | Female only | True | 0.3103 | 0.3137 | 0.3143 | 0.3141 | 19052 | 12621 | 193785 | 166603 | -0.05438 | 0.007312 | -0.04101 | -0.03988 | 0.01038 | 0.01294 |
| HEIGHT | rs11049356 | rs10771408 | T | C | 12 | CCDC91 | Female only | True | 0.3303 | 0.333 | 0.3299 | 0.3297 | 19052 | 12621 | 193785 | 166603 | -0.05795 | 0.01188 | -0.04069 | -0.03781 | 0.01021 | 0.01273 |
| HEIGHT | rs11049356 | rs7980151 | A | G | 12 | CCDC91 | Female only | True | 0.3311 | 0.334 | 0.3287 | 0.3286 | 19052 | 12621 | 193785 | 166603 | -0.05446 | 0.007858 | -0.04114 | -0.03825 | 0.01019 | 0.01272 |
| HEIGHT | rs11049356 | rs10843114 | T | C | 12 | CCDC91 | Female only | True | 0.3305 | 0.3335 | 0.328 | 0.3278 | 19052 | 12621 | 193785 | 166603 | -0.05571 | 0.008494 | -0.04108 | -0.03817 | 0.01019 | 0.01272 |

| trait | lead_snp | ID | ALLELE1 | ALLELE0 | CHR | Gene_Symbol | independent_loci_in_sex_P1e2 | in_embl_any_or_gene | A1FREQ_VUMC_female | A1FREQ_VUMC_male | A1FREQ_UKB_female | A1FREQ_UKB_male | N_VUMC_female | N_VUMC_male | N_UKB_female | N_UKB_male | BETA_VUMC_female | BETA_VUMC_male | BETA_UKB_female | BETA_UKB_male | SE_VUMC_female | SE_VUMC_male |
|---|---|---|---|---|---|---|---|---|---|---|---|---|---|---|---|---|---|---|---|---|---|---|
| HEIGHT | rs11049356 | rs11049376 | A | C | 12 | CCDC91 | Female only | True | 0.3066 | 0.3084 | 0.3113 | 0.3112 | 19052 | 12621 | 193785 | 166603 | -0.05515 | 0.005699 | -0.04165 | -0.04047 | 0.0104 | 0.01299 |
| HEIGHT | rs11049356 | rs10771409 | G | A | 12 | CCDC91 | Female only | True | 0.3326 | 0.3358 | 0.3308 | 0.3307 | 19052 | 12621 | 193785 | 166603 | -0.05425 | 0.007942 | -0.04072 | -0.0378 | 0.01018 | 0.01269 |
| HEIGHT | rs11049356 | rs10771410 | G | T | 12 | CCDC91 | Female only | True | 0.3334 | 0.3366 | 0.3308 | 0.3307 | 19052 | 12621 | 193785 | 166603 | -0.05415 | 0.007971 | -0.04072 | -0.0378 | 0.01015 | 0.01266 |
| HEIGHT | rs11049356 | rs1479492 | C | T | 12 | CCDC91 | Female only | True | 0.3326 | 0.3358 | 0.3308 | 0.3306 | 19052 | 12621 | 193785 | 166603 | -0.05432 | 0.008057 | -0.04072 | -0.0378 | 0.01018 | 0.01269 |
| HEIGHT | rs11049356 | rs7955094 | T | C | 12 | CCDC91 | Female only | True | 0.3106 | 0.314 | 0.3126 | 0.3124 | 19052 | 12621 | 193785 | 166603 | -0.05332 | 0.004154 | -0.04132 | -0.04013 | 0.01036 | 0.01292 |
| HEIGHT | rs11049356 | rs7955118 | T | C | 12 | CCDC91 | Female only | True | 0.3066 | 0.3084 | 0.3113 | 0.3112 | 19052 | 12621 | 193785 | 166603 | -0.05542 | 0.005999 | -0.04163 | -0.04039 | 0.0104 | 0.01299 |
| HEIGHT | rs11049356 | rs7955237 | T | C | 12 | CCDC91 | Female only | True | 0.3066 | 0.3083 | 0.3113 | 0.3112 | 19052 | 12621 | 193785 | 166603 | -0.0555 | 0.006044 | -0.04163 | -0.04039 | 0.0104 | 0.01299 |
| HEIGHT | rs11049356 | rs7958401 | A | G | 12 | CCDC91 | Female only | True | 0.3088 | 0.3108 | 0.3142 | 0.314 | 19052 | 12621 | 193785 | 166603 | -0.05382 | 0.005711 | -0.04129 | -0.03995 | 0.01037 | 0.01297 |
| HEIGHT | rs11049356 | rs17801436 | C | T | 12 | CCDC91 | Female only | True | 0.3087 | 0.3107 | 0.3142 | 0.314 | 19052 | 12621 | 193785 | 166603 | -0.05389 | 0.005334 | -0.04122 | -0.03997 | 0.01038 | 0.01297 |
| HEIGHT | rs11049356 | rs17801442 | A | G | 12 | CCDC91 | Female only | True | 0.3125 | 0.3161 | 0.3155 | 0.3153 | 19052 | 12621 | 193785 | 166603 | -0.05169 | 0.004345 | -0.04095 | -0.03979 | 0.01034 | 0.01291 |
| HEIGHT | rs11049356 | rs11049384 | A | G | 12 | CCDC91 | Female only | True | 0.3125 | 0.3161 | 0.3155 | 0.3153 | 19052 | 12621 | 193785 | 166603 | -0.05169 | 0.004344 | -0.04095 | -0.03979 | 0.01034 | 0.01291 |
| HEIGHT | rs11049356 | rs11049385 | G | A | 12 | CCDC91 | Female only | True | 0.3075 | 0.3095 | 0.3128 | 0.3127 | 19052 | 12621 | 193785 | 166603 | -0.05422 | 0.006546 | -0.04161 | -0.03999 | 0.0104 | 0.013 |
| HEIGHT | rs11049356 | rs11049387 | T | C | 12 | CCDC91 | Female only | True | 0.3076 | 0.3097 | 0.3128 | 0.3127 | 19052 | 12621 | 193785 | 166603 | -0.05421 | 0.006319 | -0.04161 | -0.03999 | 0.0104 | 0.01301 |
| HEIGHT | rs11049356 | rs12371462 | C | A | 12 | CCDC91 | Female only | True | 0.3086 | 0.3108 | 0.3141 | 0.3139 | 19052 | 12621 | 193785 | 166603 | -0.05407 | 0.005087 | -0.04113 | -0.03975 | 0.01038 | 0.01298 |
| HEIGHT | rs11049356 | rs11049389 | A | G | 12 | CCDC91 | Female only | True | 0.3124 | 0.3161 | 0.3153 | 0.3151 | 19052 | 12621 | 193785 | 166603 | -0.05174 | 0.004343 | -0.04084 | -0.03961 | 0.01034 | 0.01292 |
| HEIGHT | rs11049356 | rs12368652 | T | C | 12 | CCDC91 | Female only | True | 0.3125 | 0.3163 | 0.3155 | 0.3153 | 19052 | 12621 | 193785 | 166603 | -0.05173 | 0.003948 | -0.04099 | -0.0397 | 0.01034 | 0.01291 |
| HEIGHT | rs11049356 | rs11049392 | C | T | 12 | CCDC91 | Female only | True | 0.3125 | 0.3163 | 0.3154 | 0.3152 | 19052 | 12621 | 193785 | 166603 | -0.05174 | 0.003941 | -0.04102 | -0.03968 | 0.01034 | 0.01291 |
| HEIGHT | rs11049356 | rs11049393 | A | G | 12 | CCDC91 | Female only | True | 0.3088 | 0.3109 | 0.3142 | 0.314 | 19052 | 12621 | 193785 | 166603 | -0.05392 | 0.005109 | -0.04128 | -0.03986 | 0.01037 | 0.01297 |
| HEIGHT | rs11049356 | rs11049394 | A | G | 12 | CCDC91 | Female only | True | 0.3129 | 0.3168 | 0.3156 | 0.3153 | 19052 | 12621 | 193785 | 166603 | -0.05126 | 0.004693 | -0.04094 | -0.03975 | 0.01033 | 0.01291 |
| HEIGHT | rs11049356 | rs11049395 | G | A | 12 | CCDC91 | Female only | True | 0.313 | 0.3169 | 0.3155 | 0.3153 | 19052 | 12621 | 193785 | 166603 | -0.05134 | 0.004741 | -0.04091 | -0.03977 | 0.01033 | 0.01291 |
| HEIGHT | rs11049356 | rs11049397 | G | A | 12 | CCDC91 | Female only | True | 0.3129 | 0.3168 | 0.3156 | 0.3153 | 19052 | 12621 | 193785 | 166603 | -0.05126 | 0.004693 | -0.04094 | -0.03976 | 0.01033 | 0.01291 |
| HEIGHT | rs11049356 | rs10492369 | A | G | 12 | CCDC91 | Female only | True | 0.3091 | 0.3114 | 0.3143 | 0.3141 | 19052 | 12621 | 193785 | 166603 | -0.05342 | 0.005866 | -0.04119 | -0.03993 | 0.01037 | 0.01296 |
| HEIGHT | rs11049356 | rs11049398 | G | A | 12 | CCDC91 | Female only | True | 0.3129 | 0.3168 | 0.3155 | 0.3153 | 19052 | 12621 | 193785 | 166603 | -0.05126 | 0.004693 | -0.04092 | -0.03976 | 0.01033 | 0.01291 |
| HEIGHT | rs11049356 | rs11049399 | T | C | 12 | CCDC91 | Female only | True | 0.3123 | 0.3163 | 0.3154 | 0.3151 | 19052 | 12621 | 193785 | 166603 | -0.05071 | 0.005119 | -0.04096 | -0.0397 | 0.01034 | 0.01291 |
| HEIGHT | rs11049356 | rs61920227 | T | C | 12 | CCDC91 | Female only | True | 0.3123 | 0.3163 | 0.3154 | 0.3151 | 19052 | 12621 | 193785 | 166603 | -0.05071 | 0.005119 | -0.04096 | -0.0397 | 0.01034 | 0.01291 |
| HEIGHT | rs11049356 | rs11049400 | A | G | 12 | CCDC91 | Female only | True | 0.3123 | 0.3163 | 0.3154 | 0.3151 | 19052 | 12621 | 193785 | 166603 | -0.05071 | 0.005119 | -0.04096 | -0.03969 | 0.01034 | 0.01291 |
| HEIGHT | rs11049356 | rs11049401 | T | C | 12 | CCDC91 | Female only | True | 0.3123 | 0.3163 | 0.3154 | 0.3151 | 19052 | 12621 | 193785 | 166603 | -0.05075 | 0.00512 | -0.04096 | -0.03969 | 0.01034 | 0.01291 |

| trait | lead_snp | ID | ALLELE1 | ALLELE0 | CHR | Gene_Symbol | independent_loci_in_sex_P1e2 | in_embl_any_or_gene | A1FREQ_VUMC_female | A1FREQ_VUMC_male | A1FREQ_UKB_female | A1FREQ_UKB_male | N_VUMC_female | N_VUMC_male | N_UKB_female | N_UKB_male | BETA_VUMC_female | BETA_VUMC_male | BETA_UKB_female | BETA_UKB_male | SE_VUMC_female | SE_VUMC_male |
|---|---|---|---|---|---|---|---|---|---|---|---|---|---|---|---|---|---|---|---|---|---|---|
| HEIGHT | rs11049356 | rs11049402 | G | T | 12 | CCDC91 | Female only | True | 0.3125 | 0.3165 | 0.3155 | 0.3151 | 19052 | 12621 | 193785 | 166603 | -0.0507 | 0.003947 | -0.04099 | -0.03971 | 0.01034 | 0.01291 |
| HEIGHT | rs11049356 | rs11049403 | G | A | 12 | CCDC91 | Female only | True | 0.3085 | 0.3109 | 0.3141 | 0.3138 | 19052 | 12621 | 193785 | 166603 | -0.05272 | 0.006309 | -0.04121 | -0.03989 | 0.01038 | 0.01297 |
| HEIGHT | rs11049356 | rs1551987 | A | C | 12 | CCDC91 | Female only | True | 0.3124 | 0.3164 | 0.3154 | 0.3151 | 19052 | 12621 | 193785 | 166603 | -0.05067 | 0.004318 | -0.04098 | -0.03972 | 0.01034 | 0.01291 |
| HEIGHT | rs11049356 | rs1551986 | T | C | 12 | CCDC91 | Female only | True | 0.3124 | 0.3164 | 0.3154 | 0.3151 | 19052 | 12621 | 193785 | 166603 | -0.05065 | 0.00434 | -0.04098 | -0.03971 | 0.01034 | 0.01291 |
| HEIGHT | rs11049356 | rs11049405 | C | A | 12 | CCDC91 | Female only | True | 0.3124 | 0.3164 | 0.3154 | 0.3151 | 19052 | 12621 | 193785 | 166603 | -0.05069 | 0.004286 | -0.04097 | -0.03975 | 0.01034 | 0.01291 |
| HEIGHT | rs11049356 | rs11049407 | G | A | 12 | CCDC91 | Female only | True | 0.3124 | 0.3165 | 0.3154 | 0.3151 | 19052 | 12621 | 193785 | 166603 | -0.05071 | 0.004287 | -0.04098 | -0.03971 | 0.01034 | 0.01291 |
| HEIGHT | rs11049356 | rs11049409 | T | C | 12 | CCDC91 | Female only | True | 0.3124 | 0.3165 | 0.3154 | 0.3151 | 19052 | 12621 | 193785 | 166603 | -0.05074 | 0.004289 | -0.04098 | -0.0397 | 0.01034 | 0.01291 |
| HEIGHT | rs11049356 | rs11049410 | G | A | 12 | CCDC91 | Female only | True | 0.3084 | 0.3109 | 0.3141 | 0.3138 | 19052 | 12621 | 193785 | 166603 | -0.05271 | 0.006358 | -0.04122 | -0.03984 | 0.01038 | 0.01297 |
| HEIGHT | rs11049356 | rs61920228 | T | C | 12 | CCDC91 | Female only | True | 0.3124 | 0.3165 | 0.3154 | 0.3151 | 19052 | 12621 | 193785 | 166603 | -0.05071 | 0.004287 | -0.04099 | -0.0397 | 0.01034 | 0.01291 |
| HEIGHT | rs11049356 | rs7306838 | T | C | 12 | CCDC91 | Female only | True | 0.3132 | 0.3173 | 0.3154 | 0.3151 | 19052 | 12621 | 193785 | 166603 | -0.05064 | 0.004308 | -0.04098 | -0.0397 | 0.01031 | 0.01287 |
| HEIGHT | rs11049356 | rs11049412 | C | T | 12 | CCDC91 | Female only | True | 0.3123 | 0.3164 | 0.3154 | 0.3151 | 19052 | 12621 | 193785 | 166603 | -0.05062 | 0.004297 | -0.04099 | -0.03967 | 0.01034 | 0.01291 |
| HEIGHT | rs11049356 | rs11049413 | C | T | 12 | CCDC91 | Female only | True | 0.3123 | 0.3164 | 0.3154 | 0.3151 | 19052 | 12621 | 193785 | 166603 | -0.05063 | 0.004303 | -0.04099 | -0.03967 | 0.01034 | 0.01291 |
| HEIGHT | rs11049356 | rs11049414 | G | A | 12 | CCDC91 | Female only | True | 0.3123 | 0.3164 | 0.3154 | 0.3151 | 19052 | 12621 | 193785 | 166603 | -0.05063 | 0.004303 | -0.04099 | -0.03967 | 0.01034 | 0.01291 |
| HEIGHT | rs11049356 | rs2061758 | T | C | 12 | CCDC91 | Female only | True | 0.3123 | 0.3164 | 0.3154 | 0.3151 | 19052 | 12621 | 193785 | 166603 | -0.05063 | 0.004301 | -0.04099 | -0.03969 | 0.01034 | 0.01291 |
| HEIGHT | rs11049356 | rs1478336 | T | C | 12 | CCDC91 | Female only | True | 0.3123 | 0.3165 | 0.3154 | 0.3151 | 19052 | 12621 | 193785 | 166603 | -0.05092 | 0.0041 | -0.04101 | -0.03967 | 0.01034 | 0.01291 |
| HEIGHT | rs11049356 | rs1478335 | T | G | 12 | CCDC91 | Female only | True | 0.3123 | 0.3165 | 0.3154 | 0.3151 | 19052 | 12621 | 193785 | 166603 | -0.05089 | 0.004099 | -0.04101 | -0.03967 | 0.01034 | 0.01291 |
| HEIGHT | rs11049356 | rs1478334 | C | A | 12 | CCDC91 | Female only | True | 0.3123 | 0.3165 | 0.3154 | 0.3151 | 19052 | 12621 | 193785 | 166603 | -0.05089 | 0.004095 | -0.04098 | -0.03968 | 0.01034 | 0.01291 |
| HEIGHT | rs11049356 | rs11049415 | T | C | 12 | CCDC91 | Female only | True | 0.3128 | 0.3171 | 0.3159 | 0.3154 | 19052 | 12621 | 193785 | 166603 | -0.05055 | 0.004448 | -0.04065 | -0.03942 | 0.01034 | 0.0129 |
| HEIGHT | rs11049356 | rs12372059 | A | G | 12 | CCDC91 | Female only | True | 0.3123 | 0.3169 | 0.3155 | 0.3152 | 19052 | 12621 | 193785 | 166603 | -0.05164 | 0.003876 | -0.04068 | -0.03964 | 0.01034 | 0.0129 |
| HEIGHT | rs11049356 | rs12372073 | A | G | 12 | CCDC91 | Female only | True | 0.3123 | 0.3169 | 0.3155 | 0.3152 | 19052 | 12621 | 193785 | 166603 | -0.05164 | 0.003876 | -0.04068 | -0.03964 | 0.01034 | 0.0129 |
| HEIGHT | rs11049356 | rs56318627 | T | C | 12 | CCDC91 | Female only | True | 0.3123 | 0.3169 | 0.3155 | 0.3152 | 19052 | 12621 | 193785 | 166603 | -0.05155 | 0.003962 | -0.04069 | -0.03966 | 0.01034 | 0.0129 |
| HEIGHT | rs11049356 | rs11049419 | A | G | 12 | CCDC91 | Female only | True | 0.3123 | 0.3169 | 0.3155 | 0.3152 | 19052 | 12621 | 193785 | 166603 | -0.05155 | 0.003962 | -0.04069 | -0.03966 | 0.01034 | 0.0129 |
| HEIGHT | rs11049356 | rs11049420 | A | G | 12 | CCDC91 | Female only | True | 0.3123 | 0.3169 | 0.3155 | 0.3152 | 19052 | 12621 | 193785 | 166603 | -0.05155 | 0.003962 | -0.04069 | -0.03966 | 0.01034 | 0.0129 |
| HEIGHT | rs11049356 | rs11049421 | C | T | 12 | CCDC91 | Female only | True | 0.3123 | 0.3169 | 0.3155 | 0.3152 | 19052 | 12621 | 193785 | 166603 | -0.05155 | 0.003962 | -0.04069 | -0.03966 | 0.01034 | 0.0129 |
| HEIGHT | rs11049356 | rs11049422 | C | T | 12 | CCDC91 | Female only | True | 0.3123 | 0.3169 | 0.3154 | 0.3152 | 19052 | 12621 | 193785 | 166603 | -0.05154 | 0.003962 | -0.04068 | -0.03966 | 0.01034 | 0.0129 |
| HEIGHT | rs11049356 | rs11049423 | T | C | 12 | CCDC91 | Female only | True | 0.3123 | 0.3169 | 0.3155 | 0.3152 | 19052 | 12621 | 193785 | 166603 | -0.05155 | 0.003962 | -0.04069 | -0.03965 | 0.01034 | 0.0129 |
| HEIGHT | rs11049356 | rs11049424 | A | G | 12 | CCDC91 | Female only | True | 0.3088 | 0.3131 | 0.3119 | 0.3115 | 19052 | 12621 | 193785 | 166603 | -0.05123 | 0.003615 | -0.04084 | -0.0397 | 0.01042 | 0.01299 |
| HEIGHT | rs11049356 | rs11049425 | G | A | 12 | CCDC91 | Female only | True | 0.3083 | 0.3128 | 0.3113 | 0.3109 | 19052 | 12621 | 193785 | 166603 | -0.05118 | 0.003182 | -0.04091 | -0.03971 | 0.01043 | 0.013 |
| HEIGHT | rs11049356 | rs11049427 | T | C | 12 | CCDC91 | Female only | True | 0.3085 | 0.313 | 0.3115 | 0.3111 | 19052 | 12621 | 193785 | 166603 | -0.05133 | 0.00327 | -0.04079 | -0.03965 | 0.01043 | 0.01299 |

| trait | lead_snp | ID | ALLELE1 | ALLELE0 | CHR | Gene_Symbol | independent_loci_in_sex_P1e2 | in_embl_any_or_gene | A1FREQ_VUMC_female | A1FREQ_VUMC_male | A1FREQ_UKB_female | A1FREQ_UKB_male | N_VUMC_female | N_VUMC_male | N_UKB_female | N_UKB_male | BETA_VUMC_female | BETA_VUMC_male | BETA_UKB_female | BETA_UKB_male | SE_VUMC_female | SE_VUMC_male |
|---|---|---|---|---|---|---|---|---|---|---|---|---|---|---|---|---|---|---|---|---|---|---|
| HEIGHT | rs11049356 | rs11049430 | A | G | 12 | CCDC91 | Female only | True | 0.3121 | 0.3166 | 0.3154 | 0.3151 | 19052 | 12621 | 193785 | 166603 | -0.05171 | 0.003863 | -0.04071 | -0.03963 | 0.01034 | 0.0129 |
| HEIGHT | rs11049356 | rs4554927 | T | C | 12 | CCDC91 | Female only | True | 0.312 | 0.3166 | 0.3154 | 0.3151 | 19052 | 12621 | 193785 | 166603 | -0.0515 | 0.004726 | -0.04067 | -0.03961 | 0.01034 | 0.01291 |
| HEIGHT | rs11049356 | rs61920243 | C | T | 12 | CCDC91 | Female only | True | 0.312 | 0.3167 | 0.3154 | 0.3151 | 19052 | 12621 | 193785 | 166603 | -0.05147 | 0.004778 | -0.04069 | -0.03959 | 0.01034 | 0.0129 |
| HEIGHT | rs11049356 | rs7974882 | A | G | 12 | CCDC91 | Female only | True | 0.312 | 0.3167 | 0.3147 | 0.3144 | 19052 | 12621 | 193785 | 166603 | -0.05152 | 0.004638 | -0.04067 | -0.03962 | 0.01034 | 0.0129 |
| HEIGHT | rs11049356 | rs7961395 | C | T | 12 | CCDC91 | Female only | True | 0.312 | 0.3167 | 0.3147 | 0.3144 | 19052 | 12621 | 193785 | 166603 | -0.05152 | 0.004638 | -0.04069 | -0.03963 | 0.01034 | 0.0129 |
| HEIGHT | rs11049356 | rs7961407 | C | T | 12 | CCDC91 | Female only | True | 0.3122 | 0.3167 | 0.3091 | 0.3089 | 19052 | 12621 | 193785 | 166603 | -0.05145 | 0.003997 | -0.03997 | -0.03999 | 0.01032 | 0.01289 |
| HEIGHT | rs11049356 | rs7974904 | A | G | 12 | CCDC91 | Female only | True | 0.3109 | 0.3155 | 0.3088 | 0.3085 | 19052 | 12621 | 193785 | 166603 | -0.05115 | 0.004292 | -0.04005 | -0.03998 | 0.01035 | 0.01293 |
| HEIGHT | rs11049356 | rs7974701 | G | A | 12 | CCDC91 | Female only | True | 0.312 | 0.3168 | 0.3154 | 0.3152 | 19052 | 12621 | 193785 | 166603 | -0.05147 | 0.004562 | -0.04068 | -0.03958 | 0.01034 | 0.0129 |
| HEIGHT | rs11049356 | rs7961769 | C | T | 12 | CCDC91 | Female only | True | 0.3107 | 0.3152 | 0.3144 | 0.3143 | 19052 | 12621 | 193785 | 166603 | -0.05126 | 0.003851 | -0.04067 | -0.03958 | 0.01036 | 0.01294 |
| HEIGHT | rs11049356 | rs61920244 | C | A | 12 | CCDC91 | Female only | True | 0.3121 | 0.3169 | 0.3154 | 0.3152 | 19052 | 12621 | 193785 | 166603 | -0.05143 | 0.004466 | -0.04068 | -0.03957 | 0.01034 | 0.0129 |
| HEIGHT | rs11049356 | rs61920245 | C | T | 12 | CCDC91 | Female only | True | 0.3121 | 0.3169 | 0.3154 | 0.3152 | 19052 | 12621 | 193785 | 166603 | -0.05143 | 0.004503 | -0.04069 | -0.03953 | 0.01034 | 0.01291 |
| HEIGHT | rs11049356 | rs34378873 | T | C | 12 | CCDC91 | Female only | True | 0.3121 | 0.3169 | 0.3154 | 0.3152 | 19052 | 12621 | 193785 | 166603 | -0.05143 | 0.004466 | -0.04068 | -0.03957 | 0.01034 | 0.0129 |
| HEIGHT | rs11049356 | rs11049437 | A | G | 12 | CCDC91 | Female only | True | 0.312 | 0.3168 | 0.3154 | 0.3152 | 19052 | 12621 | 193785 | 166603 | -0.05149 | 0.004627 | -0.0407 | -0.03954 | 0.01034 | 0.0129 |
| HEIGHT | rs11049356 | rs11049438 | C | T | 12 | CCDC91 | Female only | True | 0.312 | 0.3167 | 0.3154 | 0.3152 | 19052 | 12621 | 193785 | 166603 | -0.05149 | 0.0046 | -0.04069 | -0.03954 | 0.01034 | 0.0129 |
| HEIGHT | rs11049356 | rs11049439 | G | T | 12 | CCDC91 | Female only | True | 0.3128 | 0.3176 | 0.3154 | 0.3152 | 19052 | 12621 | 193785 | 166603 | -0.0514 | 0.004603 | -0.04071 | -0.03955 | 0.01031 | 0.01287 |
| HEIGHT | rs11049356 | rs11049440 | C | T | 12 | CCDC91 | Female only | True | 0.3121 | 0.3169 | 0.3154 | 0.3152 | 19052 | 12621 | 193785 | 166603 | -0.05143 | 0.004466 | -0.04069 | -0.03953 | 0.01034 | 0.0129 |
| HEIGHT | rs11049356 | rs11049441 | A | G | 12 | CCDC91 | Female only | True | 0.3121 | 0.3169 | 0.3154 | 0.3152 | 19052 | 12621 | 193785 | 166603 | -0.05143 | 0.004466 | -0.04069 | -0.03953 | 0.01034 | 0.0129 |
| HEIGHT | rs11049356 | rs11049442 | A | C | 12 | CCDC91 | Female only | True | 0.312 | 0.3167 | 0.3154 | 0.3152 | 19052 | 12621 | 193785 | 166603 | -0.0515 | 0.004603 | -0.0407 | -0.03954 | 0.01034 | 0.0129 |
| HEIGHT | rs11049356 | rs11049443 | G | A | 12 | CCDC91 | Female only | True | 0.312 | 0.3167 | 0.3154 | 0.3152 | 19052 | 12621 | 193785 | 166603 | -0.0515 | 0.004603 | -0.0407 | -0.03954 | 0.01034 | 0.0129 |
| trait | lead_snp | ID | ALLELE1 | ALLELE0 | CHR | Gene_Symbol | independent_loci_in_sex_P1e2 | in_embl_any_or_gene | A1FREQ_VUMC_female | A1FREQ_VUMC_male | A1FREQ_UKB_female | A1FREQ_UKB_male | N_VUMC_female | N_VUMC_male | N_UKB_female | N_UKB_male | BETA_VUMC_female | BETA_VUMC_male | BETA_UKB_female | BETA_UKB_male | SE_VUMC_female | SE_VUMC_male |
| HEIGHT | rs11049356 | rs11049444 | A | G | 12 | CCDC91 | Female only | True | 0.312 | 0.3167 | 0.3154 | 0.3152 | 19052 | 12621 | 193785 | 166603 | -0.0515 | 0.004603 | -0.0407 | -0.03954 | 0.01034 | 0.0129 |
| HEIGHT | rs11049356 | rs60193273 | C | T | 12 | CCDC91 | Female only | True | 0.312 | 0.3167 | 0.3154 | 0.3152 | 19052 | 12621 | 193785 | 166603 | -0.0515 | 0.004603 | -0.0407 | -0.03954 | 0.01034 | 0.0129 |
| HEIGHT | rs11049356 | rs11049446 | C | T | 12 | CCDC91 | Female only | True | 0.312 | 0.3167 | 0.3154 | 0.3152 | 19052 | 12621 | 193785 | 166603 | -0.0515 | 0.004603 | -0.0407 | -0.03954 | 0.01034 | 0.0129 |
| HEIGHT | rs11049356 | rs11049447 | G | A | 12 | CCDC91 | Female only | True | 0.312 | 0.3167 | 0.3154 | 0.3152 | 19052 | 12621 | 193785 | 166603 | -0.0515 | 0.004603 | -0.0407 | -0.03954 | 0.01034 | 0.0129 |
| HEIGHT | rs11049356 | rs10843123 | G | A | 12 | CCDC91 | Female only | True | 0.312 | 0.3167 | 0.3154 | 0.3152 | 19052 | 12621 | 193785 | 166603 | -0.0515 | 0.004603 | -0.0407 | -0.03954 | 0.01034 | 0.0129 |
| HEIGHT | rs11049356 | rs10843124 | G | A | 12 | CCDC91 | Female only | True | 0.312 | 0.3168 | 0.3154 | 0.3152 | 19052 | 12621 | 193785 | 166603 | -0.05143 | 0.004521 | -0.04066 | -0.03954 | 0.01034 | 0.0129 |
| HEIGHT | rs11049356 | rs10843125 | T | C | 12 | CCDC91 | Female only | True | 0.312 | 0.3167 | 0.3154 | 0.3152 | 19052 | 12621 | 193785 | 166603 | -0.0515 | 0.004603 | -0.0407 | -0.03954 | 0.01034 | 0.0129 |
| HEIGHT | rs11049356 | rs12146881 | T | C | 12 | CCDC91 | Female only | True | 0.312 | 0.3167 | 0.3154 | 0.3152 | 19052 | 12621 | 193785 | 166603 | -0.0515 | 0.004603 | -0.0407 | -0.03954 | 0.01034 | 0.0129 |

| trait | lead_snp | ID | ALLELE1 | ALLELE0 | CHR | Gene_Symbol | independent_loci_in_sex_P1e2 | in_embl_any_or_gene | A1FREQ_VUMC_female | A1FREQ_VUMC_male | A1FREQ_UKB_female | A1FREQ_UKB_male | N_VUMC_female | N_VUMC_male | N_UKB_female | N_UKB_male | BETA_VUMC_female | BETA_VUMC_male | BETA_UKB_female | BETA_UKB_male | SE_VUMC_female | SE_VUMC_male |
|---|---|---|---|---|---|---|---|---|---|---|---|---|---|---|---|---|---|---|---|---|---|---|
| HEIGHT | rs11049356 | rs11049448 | A | G | 12 | CCDC91 | Female only | True | 0.312 | 0.3168 | 0.3154 | 0.3152 | 19052 | 12621 | 193785 | 166603 | -0.0515 | 0.004597 | -0.04075 | -0.03958 | 0.01034 | 0.0129 |
| HEIGHT | rs11049356 | rs10843127 | G | A | 12 | CCDC91 | Female only | True | 0.3119 | 0.3167 | 0.3154 | 0.3152 | 19052 | 12621 | 193785 | 166603 | -0.05147 | 0.004464 | -0.0407 | -0.03954 | 0.01034 | 0.01291 |
| HEIGHT | rs11049356 | rs10843128 | C | T | 12 | CCDC91 | Female only | True | 0.3119 | 0.3167 | 0.3154 | 0.3152 | 19052 | 12621 | 193785 | 166603 | -0.05147 | 0.004464 | -0.0407 | -0.03954 | 0.01034 | 0.01291 |
| HEIGHT | rs11049356 | rs10843129 | A | G | 12 | CCDC91 | Female only | True | 0.3119 | 0.3167 | 0.3154 | 0.3152 | 19052 | 12621 | 193785 | 166603 | -0.05147 | 0.004464 | -0.0407 | -0.03954 | 0.01034 | 0.01291 |
| HEIGHT | rs11049356 | rs10843130 | A | G | 12 | CCDC91 | Female only | True | 0.3119 | 0.3167 | 0.3154 | 0.3152 | 19052 | 12621 | 193785 | 166603 | -0.05147 | 0.004464 | -0.0407 | -0.03954 | 0.01034 | 0.01291 |
| HEIGHT | rs11049356 | rs61920253 | A | G | 12 | CCDC91 | Female only | True | 0.3119 | 0.3167 | 0.3154 | 0.3152 | 19052 | 12621 | 193785 | 166603 | -0.05147 | 0.004461 | -0.0407 | -0.03954 | 0.01034 | 0.01291 |
| HEIGHT | rs11049356 | rs11612143 | C | T | 12 | CCDC91 | Female only | True | 0.3111 | 0.316 | 0.3144 | 0.3142 | 19052 | 12621 | 193785 | 166603 | -0.05145 | 0.004514 | -0.04107 | -0.03895 | 0.01036 | 0.01293 |
| HEIGHT | rs11049356 | rs11609999 | T | C | 12 | CCDC91 | Female only | True | 0.3113 | 0.3162 | 0.3145 | 0.3143 | 19052 | 12621 | 193785 | 166603 | -0.05132 | 0.004459 | -0.04105 | -0.03899 | 0.01036 | 0.01293 |
| HEIGHT | rs11049356 | rs11610011 | T | C | 12 | CCDC91 | Female only | True | 0.3119 | 0.3167 | 0.3155 | 0.3152 | 19052 | 12621 | 193785 | 166603 | -0.05147 | 0.004464 | -0.04066 | -0.0396 | 0.01034 | 0.01291 |
| HEIGHT | rs11049356 | rs11049453 | A | G | 12 | CCDC91 | Female only | True | 0.3118 | 0.3166 | 0.3154 | 0.3152 | 19052 | 12621 | 193785 | 166603 | -0.0515 | 0.004548 | -0.04069 | -0.03954 | 0.01034 | 0.01292 |
| HEIGHT | rs11049356 | rs7957056 | G | A | 12 | CCDC91 | Female only | True | 0.3118 | 0.3166 | 0.3154 | 0.3152 | 19052 | 12621 | 193785 | 166603 | -0.0515 | 0.004584 | -0.04069 | -0.03954 | 0.01034 | 0.01292 |
| HEIGHT | rs11049356 | rs7957059 | G | A | 12 | CCDC91 | Female only | True | 0.3118 | 0.3166 | 0.3154 | 0.3152 | 19052 | 12621 | 193785 | 166603 | -0.0515 | 0.004549 | -0.04069 | -0.03954 | 0.01034 | 0.01292 |
| HEIGHT | rs11049356 | rs7957382 | C | A | 12 | CCDC91 | Female only | True | 0.3118 | 0.3166 | 0.3154 | 0.3152 | 19052 | 12621 | 193785 | 166603 | -0.0515 | 0.004549 | -0.04069 | -0.03954 | 0.01034 | 0.01292 |
| HEIGHT | rs11049356 | rs7957503 | G | A | 12 | CCDC91 | Female only | True | 0.3118 | 0.3166 | 0.3154 | 0.3152 | 19052 | 12621 | 193785 | 166603 | -0.05153 | 0.004425 | -0.04069 | -0.03954 | 0.01035 | 0.01292 |
| HEIGHT | rs11049356 | rs73261702 | G | A | 12 | CCDC91 | Female only | True | 0.3118 | 0.3166 | 0.3154 | 0.3152 | 19052 | 12621 | 193785 | 166603 | -0.05153 | 0.004425 | -0.04069 | -0.03954 | 0.01035 | 0.01292 |
| HEIGHT | rs11049356 | rs11049454 | G | A | 12 | CCDC91 | Female only | True | 0.3081 | 0.3108 | 0.3141 | 0.3139 | 19052 | 12621 | 193785 | 166603 | -0.05347 | 0.005298 | -0.04096 | -0.03976 | 0.01038 | 0.01299 |
| HEIGHT | rs11049356 | rs11049455 | A | G | 12 | CCDC91 | Female only | True | 0.3119 | 0.3166 | 0.3154 | 0.3152 | 19052 | 12621 | 193785 | 166603 | -0.0515 | 0.004341 | -0.0407 | -0.03955 | 0.01034 | 0.01292 |
| HEIGHT | rs11049356 | rs11049457 | C | T | 12 | CCDC91 | Female only | True | 0.3119 | 0.3166 | 0.3154 | 0.3152 | 19052 | 12621 | 193785 | 166603 | -0.0515 | 0.004341 | -0.0407 | -0.03955 | 0.01034 | 0.01292 |
| HEIGHT | rs11049356 | rs10843131 | T | G | 12 | CCDC91 | Female only | True | 0.3119 | 0.3166 | 0.3154 | 0.3152 | 19052 | 12621 | 193785 | 166603 | -0.05149 | 0.004347 | -0.0407 | -0.03955 | 0.01034 | 0.01292 |
| HEIGHT | rs11049356 | rs10843132 | G | A | 12 | CCDC91 | Female only | True | 0.3119 | 0.3166 | 0.3154 | 0.3152 | 19052 | 12621 | 193785 | 166603 | -0.0515 | 0.00434 | -0.0407 | -0.03954 | 0.01034 | 0.01292 |
| HEIGHT | rs11049356 | rs10843134 | G | A | 12 | CCDC91 | Female only | True | 0.3081 | 0.3108 | 0.3141 | 0.3139 | 19052 | 12621 | 193785 | 166603 | -0.05349 | 0.005202 | -0.04096 | -0.03976 | 0.01038 | 0.01299 |
| HEIGHT | rs11049356 | rs10843135 | A | G | 12 | CCDC91 | Female only | True | 0.3119 | 0.3166 | 0.3154 | 0.3152 | 19052 | 12621 | 193785 | 166603 | -0.05145 | 0.004341 | -0.04065 | -0.03951 | 0.01034 | 0.01292 |
| HEIGHT | rs11049356 | rs11049459 | C | T | 12 | CCDC91 | Female only | True | 0.3119 | 0.3166 | 0.3154 | 0.3152 | 19052 | 12621 | 193785 | 166603 | -0.0515 | 0.00434 | -0.0407 | -0.03954 | 0.01034 | 0.01292 |
| HEIGHT | rs11049356 | rs11049460 | A | G | 12 | CCDC91 | Female only | True | 0.3119 | 0.3166 | 0.3154 | 0.3152 | 19052 | 12621 | 193785 | 166603 | -0.0515 | 0.00434 | -0.0407 | -0.03954 | 0.01034 | 0.01292 |
| HEIGHT | rs11049356 | rs1824768 | T | C | 12 | CCDC91 | Female only | True | 0.3081 | 0.3108 | 0.3141 | 0.3139 | 19052 | 12621 | 193785 | 166603 | -0.05344 | 0.00524 | -0.04094 | -0.03973 | 0.01038 | 0.01299 |
| HEIGHT | rs11049356 | rs1824767 | G | A | 12 | CCDC91 | Female only | True | 0.3119 | 0.3166 | 0.3154 | 0.3152 | 19052 | 12621 | 193785 | 166603 | -0.0515 | 0.00434 | -0.0407 | -0.03953 | 0.01034 | 0.01292 |
| HEIGHT | rs11049356 | rs1824766 | G | A | 12 | CCDC91 | Female only | True | 0.3119 | 0.3166 | 0.3154 | 0.3152 | 19052 | 12621 | 193785 | 166603 | -0.0515 | 0.00434 | -0.0407 | -0.03953 | 0.01034 | 0.01292 |
| HEIGHT | rs11049356 | rs1585682 | G | A | 12 | CCDC91 | Female only | True | 0.3119 | 0.3166 | 0.3154 | 0.3152 | 19052 | 12621 | 193785 | 166603 | -0.0515 | 0.00434 | -0.0407 | -0.03953 | 0.01034 | 0.01292 |
| HEIGHT | rs11049356 | rs9645745 | A | G | 12 | CCDC91 | Female only | True | 0.3119 | 0.3166 | 0.3154 | 0.3152 | 19052 | 12621 | 193785 | 166603 | -0.0515 | 0.00434 | -0.0407 | -0.03953 | 0.01034 | 0.01292 |

| trait | lead_snp | ID | ALLELE1 | ALLELE0 | CHR | Gene_Symbol | independent_loci_in_sex_P1e2 | in_embl_any_or_gene | A1FREQ_VUMC_female | A1FREQ_VUMC_male | A1FREQ_UKB_female | A1FREQ_UKB_male | N_VUMC_female | N_VUMC_male | N_UKB_female | N_UKB_male | BETA_VUMC_female | BETA_VUMC_male | BETA_UKB_female | BETA_UKB_male | SE_VUMC_female | SE_VUMC_male |
|---|---|---|---|---|---|---|---|---|---|---|---|---|---|---|---|---|---|---|---|---|---|---|
| HEIGHT | rs11049356 | rs7973241 | G | A | 12 | CCDC91 | Female only | True | 0.3119 | 0.3166 | 0.3154 | 0.3152 | 19052 | 12621 | 193785 | 166603 | -0.0515 | 0.00434 | -0.0407 | -0.03953 | 0.01034 | 0.01292 |
| HEIGHT | rs11049356 | rs7971033 | C | T | 12 | CCDC91 | Female only | True | 0.3119 | 0.3166 | 0.3154 | 0.3152 | 19052 | 12621 | 193785 | 166603 | -0.05149 | 0.004338 | -0.0407 | -0.03953 | 0.01034 | 0.01292 |
| HEIGHT | rs11049356 | rs7956418 | A | G | 12 | CCDC91 | Female only | True | 0.3119 | 0.3166 | 0.3154 | 0.3152 | 19052 | 12621 | 193785 | 166603 | -0.05149 | 0.004338 | -0.0407 | -0.03953 | 0.01034 | 0.01292 |
| HEIGHT | rs11049356 | rs7956671 | T | C | 12 | CCDC91 | Female only | True | 0.3119 | 0.3166 | 0.3153 | 0.315 | 19052 | 12621 | 193785 | 166603 | -0.05155 | 0.004338 | -0.04078 | -0.03949 | 0.01034 | 0.01292 |
| HEIGHT | rs11049356 | rs11494779 | T | C | 12 | CCDC91 | Female only | True | 0.3119 | 0.3167 | 0.3153 | 0.315 | 19052 | 12621 | 193785 | 166603 | -0.05155 | 0.004347 | -0.04078 | -0.03949 | 0.01034 | 0.01292 |
| HEIGHT | rs11049356 | rs11519094 | G | A | 12 | CCDC91 | Female only | True | 0.3119 | 0.3166 | 0.3153 | 0.315 | 19052 | 12621 | 193785 | 166603 | -0.05155 | 0.004338 | -0.04078 | -0.03949 | 0.01034 | 0.01292 |
| HEIGHT | rs11049356 | rs11513465 | T | C | 12 | CCDC91 | Female only | True | 0.3122 | 0.317 | 0.3156 | 0.3153 | 19052 | 12621 | 193785 | 166603 | -0.05128 | 0.004224 | -0.04083 | -0.03944 | 0.01035 | 0.01292 |
| HEIGHT | rs11049356 | rs11513250 | T | C | 12 | CCDC91 | Female only | True | 0.3121 | 0.3168 | 0.3156 | 0.3153 | 19052 | 12621 | 193785 | 166603 | -0.0515 | 0.004356 | -0.04087 | -0.03951 | 0.01034 | 0.01292 |
| HEIGHT | rs11049356 | rs11519330 | C | T | 12 | CCDC91 | Female only | True | 0.3119 | 0.3166 | 0.3153 | 0.315 | 19052 | 12621 | 193785 | 166603 | -0.05156 | 0.004341 | -0.04078 | -0.03949 | 0.01034 | 0.01292 |
| HEIGHT | rs11049356 | rs11519331 | A | G | 12 | CCDC91 | Female only | True | 0.3119 | 0.3166 | 0.3153 | 0.315 | 19052 | 12621 | 193785 | 166603 | -0.05155 | 0.004337 | -0.04077 | -0.03949 | 0.01034 | 0.01292 |
| HEIGHT | rs11049356 | rs11513466 | G | A | 12 | CCDC91 | Female only | True | 0.3119 | 0.3166 | 0.3153 | 0.315 | 19052 | 12621 | 193785 | 166603 | -0.05156 | 0.004341 | -0.04078 | -0.03949 | 0.01034 | 0.01292 |
| HEIGHT | rs11049356 | rs11513467 | C | T | 12 | CCDC91 | Female only | True | 0.3119 | 0.3166 | 0.3153 | 0.315 | 19052 | 12621 | 193785 | 166603 | -0.05155 | 0.004338 | -0.04078 | -0.03949 | 0.01034 | 0.01292 |
| HEIGHT | rs11049356 | rs11519332 | A | G | 12 | CCDC91 | Female only | True | 0.3119 | 0.3166 | 0.3153 | 0.315 | 19052 | 12621 | 193785 | 166603 | -0.05155 | 0.004338 | -0.04078 | -0.03949 | 0.01034 | 0.01292 |
| HEIGHT | rs11049356 | rs6487669 | A | C | 12 | CCDC91 | Female only | True | 0.3119 | 0.3166 | 0.3153 | 0.315 | 19052 | 12621 | 193785 | 166603 | -0.05155 | 0.004338 | -0.04078 | -0.03949 | 0.01034 | 0.01292 |
| HEIGHT | rs11049356 | rs6487671 | A | G | 12 | CCDC91 | Female only | True | 0.3119 | 0.3166 | 0.3152 | 0.315 | 19052 | 12621 | 193785 | 166603 | -0.05156 | 0.004334 | -0.04078 | -0.0395 | 0.01034 | 0.01292 |
| HEIGHT | rs11049356 | rs11049465 | T | C | 12 | CCDC91 | Female only | True | 0.308 | 0.3108 | 0.314 | 0.3137 | 19052 | 12621 | 193785 | 166603 | -0.05362 | 0.005292 | -0.04102 | -0.0397 | 0.01038 | 0.01299 |
| HEIGHT | rs11049356 | rs7980441 | T | C | 12 | CCDC91 | Female only | True | 0.3078 | 0.3105 | 0.3139 | 0.3136 | 19052 | 12621 | 193785 | 166603 | -0.05371 | 0.004624 | -0.04109 | -0.03967 | 0.0104 | 0.01301 |
| HEIGHT | rs11049356 | rs11049467 | A | G | 12 | CCDC91 | Female only | True | 0.3078 | 0.3105 | 0.3139 | 0.3137 | 19052 | 12621 | 193785 | 166603 | -0.0536 | 0.004617 | -0.04109 | -0.0397 | 0.01039 | 0.01301 |
| HEIGHT | rs11049356 | rs11049468 | T | C | 12 | CCDC91 | Female only | True | 0.3079 | 0.3105 | 0.3139 | 0.3136 | 19052 | 12621 | 193785 | 166603 | -0.05361 | 0.004666 | -0.04111 | -0.03972 | 0.01039 | 0.01301 |
| HEIGHT | rs11049356 | rs11049469 | A | C | 12 | CCDC91 | Female only | True | 0.3078 | 0.3105 | 0.3139 | 0.3136 | 19052 | 12621 | 193785 | 166603 | -0.05366 | 0.004594 | -0.04112 | -0.03972 | 0.0104 | 0.01301 |
| HEIGHT | rs11049356 | rs10843137 | A | C | 12 | CCDC91 | Female only | True | 0.3117 | 0.3165 | 0.3153 | 0.315 | 19052 | 12621 | 193785 | 166603 | -0.05156 | 0.003665 | -0.04079 | -0.03956 | 0.01035 | 0.01295 |
| HEIGHT | rs11049356 | rs10843138 | A | G | 12 | CCDC91 | Female only | True | 0.3117 | 0.3164 | 0.3153 | 0.315 | 19052 | 12621 | 193785 | 166603 | -0.05157 | 0.003751 | -0.0408 | -0.03953 | 0.01035 | 0.01294 |
| HEIGHT | rs11049356 | rs1841962 | C | T | 12 | CCDC91 | Female only | True | 0.3171 | 0.3225 | 0.3162 | 0.316 | 19052 | 12621 | 193785 | 166603 | -0.04948 | 0.00242 | -0.04046 | -0.03982 | 0.01034 | 0.0129 |
| trait | lead_snp | ID | ALLELE1 | ALLELE0 | CHR | Gene_Symbol | independent_loci_in_sex_P1e2 | in_embl_any_or_gene | A1FREQ_VUMC_female | A1FREQ_VUMC_male | A1FREQ_UKB_female | A1FREQ_UKB_male | N_VUMC_female | N_VUMC_male | N_UKB_female | N_UKB_male | BETA_VUMC_female | BETA_VUMC_male | BETA_UKB_female | BETA_UKB_male | SE_VUMC_female | SE_VUMC_male |
| HEIGHT | rs11049356 | rs1871152 | G | A | 12 | CCDC91 | Female only | True | 0.3186 | 0.3243 | 0.3172 | 0.3169 | 19052 | 12621 | 193785 | 166603 | -0.0485 | 0.001036 | -0.04022 | -0.03964 | 0.01033 | 0.01289 |
| HEIGHT | rs11049356 | rs10843140 | C | T | 12 | CCDC91 | Female only | True | 0.3079 | 0.3105 | 0.314 | 0.3137 | 19052 | 12621 | 193785 | 166603 | -0.0536 | 0.004667 | -0.04106 | -0.03973 | 0.01039 | 0.01301 |
| HEIGHT | rs11049356 | rs11049471 | T | C | 12 | CCDC91 | Female only | True | 0.3117 | 0.3164 | 0.3153 | 0.315 | 19052 | 12621 | 193785 | 166603 | -0.0516 | 0.003732 | -0.04081 | -0.0395 | 0.01035 | 0.01294 |
| HEIGHT | rs11049356 | rs11049472 | C | T | 12 | CCDC91 | Female only | True | 0.3079 | 0.3105 | 0.314 | 0.3137 | 19052 | 12621 | 193785 | 166603 | -0.05358 | 0.004481 | -0.04106 | -0.03973 | 0.01039 | 0.01301 |

| trait | lead_snp | ID | ALLELE1 | ALLELE0 | CHR | Gene_Symbol | independent_loci_in_sex_P1e2 | in_embl_any_or_gene | A1FREQ_VUMC_female | A1FREQ_VUMC_male | A1FREQ_UKB_female | A1FREQ_UKB_male | N_VUMC_female | N_VUMC_male | N_UKB_female | N_UKB_male | BETA_VUMC_female | BETA_VUMC_male | BETA_UKB_female | BETA_UKB_male | SE_VUMC_female | SE_VUMC_male |
|---|---|---|---|---|---|---|---|---|---|---|---|---|---|---|---|---|---|---|---|---|---|---|
| HEIGHT | rs11049356 | rs11049474 | G | A | 12 | CCDC91 | Female only | True | 0.3079 | 0.3105 | 0.314 | 0.3137 | 19052 | 12621 | 193785 | 166603 | -0.05362 | 0.004518 | -0.04107 | -0.03974 | 0.01039 | 0.01301 |
| HEIGHT | rs11049356 | rs7969582 | G | A | 12 | CCDC91 | Female only | True | 0.3171 | 0.3225 | 0.3161 | 0.3159 | 19052 | 12621 | 193785 | 166603 | -0.04959 | 0.002129 | -0.04051 | -0.03971 | 0.01034 | 0.01291 |
| HEIGHT | rs11049356 | rs11049475 | A | G | 12 | CCDC91 | Female only | True | 0.3116 | 0.3163 | 0.3153 | 0.315 | 19052 | 12621 | 193785 | 166603 | -0.05147 | 0.00358 | -0.04074 | -0.03955 | 0.01036 | 0.01295 |
| HEIGHT | rs11049356 | rs7303747 | A | G | 12 | CCDC91 | Female only | True | 0.3079 | 0.3107 | 0.314 | 0.3137 | 19052 | 12621 | 193785 | 166603 | -0.05355 | 0.004281 | -0.04125 | -0.03976 | 0.0104 | 0.01302 |
| HEIGHT | rs11049356 | rs11049476 | G | A | 12 | CCDC91 | Female only | True | 0.3115 | 0.3159 | 0.3153 | 0.315 | 19052 | 12621 | 193785 | 166603 | -0.05195 | 0.00279 | -0.04112 | -0.03963 | 0.01035 | 0.01294 |
| HEIGHT | rs11049356 | rs11049478 | A | G | 12 | CCDC91 | Female only | True | 0.308 | 0.3105 | 0.314 | 0.3137 | 19052 | 12621 | 193785 | 166603 | -0.05325 | 0.003927 | -0.04144 | -0.03974 | 0.01039 | 0.01301 |
| HEIGHT | rs11049356 | rs10843141 | T | G | 12 | CCDC91 | Female only | True | 0.3081 | 0.3105 | 0.314 | 0.3138 | 19052 | 12621 | 193785 | 166603 | -0.0534 | 0.004006 | -0.0415 | -0.03968 | 0.01038 | 0.01301 |
| HEIGHT | rs11049356 | rs11049480 | G | A | 12 | CCDC91 | Female only | True | 0.3114 | 0.3159 | 0.3152 | 0.315 | 19052 | 12621 | 193785 | 166603 | -0.05209 | 0.001985 | -0.0413 | -0.03954 | 0.01035 | 0.01295 |
| HEIGHT | rs11049356 | rs11049481 | T | C | 12 | CCDC91 | Female only | True | 0.2973 | 0.299 | 0.3031 | 0.3033 | 19052 | 12621 | 193785 | 166603 | -0.05081 | -0.001565 | -0.04286 | -0.03979 | 0.01051 | 0.01318 |
| HEIGHT | rs11049356 | rs11049484 | G | T | 12 | CCDC91 | Female only | True | 0.3002 | 0.3018 | 0.306 | 0.3061 | 19052 | 12621 | 193785 | 166603 | -0.05071 | -8.02e-04 | -0.04239 | -0.03966 | 0.01044 | 0.0131 |
| HEIGHT | rs11049356 | rs11049488 | A | G | 12 | CCDC91 | Female only | True | 0.301 | 0.3029 | 0.3059 | 0.306 | 19052 | 12621 | 193785 | 166603 | -0.05089 | -0.001446 | -0.04238 | -0.03955 | 0.01039 | 0.01305 |
| HEIGHT | rs11049356 | rs57618147 | A | C | 12 | CCDC91 | Female only | True | 0.2999 | 0.3018 | 0.3059 | 0.306 | 19052 | 12621 | 193785 | 166603 | -0.051 | -0.001435 | -0.04238 | -0.03955 | 0.01043 | 0.01309 |
| HEIGHT | rs11049356 | rs716687 | A | G | 12 | CCDC91 | Female only | True | 0.2999 | 0.3018 | 0.3059 | 0.3059 | 19052 | 12621 | 193785 | 166603 | -0.05102 | -0.001412 | -0.04238 | -0.03955 | 0.01043 | 0.01309 |
| HEIGHT | rs11049356 | rs11049493 | G | A | 12 | CCDC91 | Female only | True | 0.3 | 0.3019 | 0.3059 | 0.306 | 19052 | 12621 | 193785 | 166603 | -0.05082 | -0.001183 | -0.04235 | -0.03959 | 0.01043 | 0.0131 |
| HEIGHT | rs11049356 | rs11049495 | A | G | 12 | CCDC91 | Female only | True | 0.3 | 0.3019 | 0.3059 | 0.306 | 19052 | 12621 | 193785 | 166603 | -0.05082 | -0.001179 | -0.04234 | -0.03956 | 0.01043 | 0.0131 |
| HEIGHT | rs11049356 | rs11049496 | G | A | 12 | CCDC91 | Female only | True | 0.3 | 0.3019 | 0.3059 | 0.306 | 19052 | 12621 | 193785 | 166603 | -0.05082 | -0.001177 | -0.04235 | -0.03956 | 0.01043 | 0.0131 |
| HEIGHT | rs11049356 | rs10843146 | C | T | 12 | CCDC91 | Female only | True | 0.3 | 0.3018 | 0.3059 | 0.306 | 19052 | 12621 | 193785 | 166603 | -0.05059 | -0.001331 | -0.04235 | -0.03958 | 0.01043 | 0.0131 |
| HEIGHT | rs11049356 | rs2045887 | A | G | 12 | CCDC91 | Female only | True | 0.2999 | 0.3018 | 0.3058 | 0.3059 | 19052 | 12621 | 193785 | 166603 | -0.05088 | -0.00154 | -0.04226 | -0.03947 | 0.01044 | 0.01311 |
| HEIGHT | rs11049356 | rs2172299 | A | G | 12 | CCDC91 | Female only | True | 0.2995 | 0.3009 | 0.3055 | 0.3055 | 19052 | 12621 | 193785 | 166603 | -0.05055 | -0.001857 | -0.04223 | -0.0393 | 0.01045 | 0.01313 |
| HEIGHT | rs11049356 | rs11049503 | A | G | 12 | CCDC91 | Female only | True | 0.2995 | 0.3009 | 0.3055 | 0.3056 | 19052 | 12621 | 193785 | 166603 | -0.0506 | -0.001808 | -0.04228 | -0.03934 | 0.01045 | 0.01313 |
| HEIGHT | rs11049356 | rs61922974 | A | G | 12 | CCDC91 | Female only | True | 0.2995 | 0.3009 | 0.3055 | 0.3056 | 19052 | 12621 | 193785 | 166603 | -0.05061 | -0.001875 | -0.04228 | -0.03936 | 0.01045 | 0.01313 |
| HEIGHT | rs11049356 | rs61922975 | A | G | 12 | CCDC91 | Female only | True | 0.2995 | 0.3009 | 0.3055 | 0.3055 | 19052 | 12621 | 193785 | 166603 | -0.05061 | -0.001857 | -0.04226 | -0.03929 | 0.01045 | 0.01313 |
| HEIGHT | rs11049356 | rs61922976 | G | A | 12 | CCDC91 | Female only | True | 0.2995 | 0.3009 | 0.3055 | 0.3056 | 19052 | 12621 | 193785 | 166603 | -0.05062 | -0.001862 | -0.04226 | -0.03933 | 0.01045 | 0.01313 |
| HEIGHT | rs11049356 | rs61922977 | G | T | 12 | CCDC91 | Female only | True | 0.2993 | 0.3007 | 0.3055 | 0.3055 | 19052 | 12621 | 193785 | 166603 | -0.05069 | -0.002159 | -0.04233 | -0.0393 | 0.01045 | 0.01313 |
| HEIGHT | rs11049356 | rs11049505 | C | T | 12 | CCDC91 | Female only | True | 0.2993 | 0.3007 | 0.3054 | 0.3055 | 19052 | 12621 | 193785 | 166603 | -0.05071 | -0.002175 | -0.0423 | -0.03926 | 0.01045 | 0.01313 |
| HEIGHT | rs11049356 | rs11049509 | A | G | 12 | CCDC91 | Female only | True | 0.2993 | 0.3008 | 0.3055 | 0.3055 | 19052 | 12621 | 193785 | 166603 | -0.05076 | -0.002141 | -0.04236 | -0.03929 | 0.01045 | 0.01313 |
| HEIGHT | rs11049356 | rs11049510 | A | G | 12 | CCDC91 | Female only | True | 0.2993 | 0.3008 | 0.3055 | 0.3055 | 19052 | 12621 | 193785 | 166603 | -0.05074 | -0.002145 | -0.04232 | -0.03928 | 0.01045 | 0.01313 |
| HEIGHT | rs11049356 | rs4357727 | C | T | 12 | CCDC91 | Female only | True | 0.3651 | 0.3675 | 0.3583 | 0.3582 | 19052 | 12621 | 193785 | 166603 | -0.04368 | -0.002464 | -0.03371 | -0.03082 | 0.009958 | 0.01248 |
| HEIGHT | rs11049356 | rs17432330 | G | T | 12 | CCDC91 | Female only | True | 0.2986 | 0.2998 | 0.3048 | 0.3049 | 19052 | 12621 | 193785 | 166603 | -0.05075 | -0.00104 | -0.04238 | -0.03934 | 0.01046 | 0.01315 |

| trait | lead_snp | ID | ALLELE1 | ALLELE0 | CHR | Gene_Symbol | independent_loci_in_sex_P1e2 | in_embl_any_or_gene | A1FREQ_VUMC_female | A1FREQ_VUMC_male | A1FREQ_UKB_female | A1FREQ_UKB_male | N_VUMC_female | N_VUMC_male | N_UKB_female | N_UKB_male | BETA_VUMC_female | BETA_VUMC_male | BETA_UKB_female | BETA_UKB_male | SE_VUMC_female | SE_VUMC_male |
|---|---|---|---|---|---|---|---|---|---|---|---|---|---|---|---|---|---|---|---|---|---|---|
| HEIGHT | rs11049356 | rs12372448 | T | C | 12 | CCDC91 | Female only | True | 0.2994 | 0.3009 | 0.3055 | 0.3055 | 19052 | 12621 | 193785 | 166603 | -0.05058 | -0.001784 | -0.04232 | -0.03932 | 0.01045 | 0.01313 |
| HEIGHT | rs11049356 | rs11049515 | A | G | 12 | CCDC91 | Female only | True | 0.2996 | 0.3009 | 0.3055 | 0.3056 | 19052 | 12621 | 193785 | 166603 | -0.05061 | -0.002281 | -0.04234 | -0.03924 | 0.01045 | 0.01313 |
| HEIGHT | rs11049356 | rs61922979 | C | T | 12 | CCDC91 | Female only | True | 0.2996 | 0.3009 | 0.3055 | 0.3055 | 19052 | 12621 | 193785 | 166603 | -0.05061 | -0.002296 | -0.04235 | -0.03923 | 0.01045 | 0.01313 |
| HEIGHT | rs11049356 | rs11049518 | C | A | 12 | CCDC91 | Female only | True | 0.2995 | 0.3009 | 0.3055 | 0.3055 | 19052 | 12621 | 193785 | 166603 | -0.05033 | -0.001999 | -0.04235 | -0.03921 | 0.01045 | 0.01313 |
| HEIGHT | rs11049356 | rs12368409 | T | C | 12 | CCDC91 | Female only | True | 0.2996 | 0.301 | 0.3055 | 0.3056 | 19052 | 12621 | 193785 | 166603 | -0.05059 | -0.001913 | -0.04236 | -0.03921 | 0.01045 | 0.01313 |
| HEIGHT | rs11049356 | rs10843150 | G | A | 12 | CCDC91 | Female only | True | 0.2996 | 0.3009 | 0.3055 | 0.3056 | 19052 | 12621 | 193785 | 166603 | -0.05047 | -0.001971 | -0.04231 | -0.0392 | 0.01045 | 0.01313 |
| HEIGHT | rs11049356 | rs3825246 | G | A | 12 | CCDC91 | Female only | True | 0.2995 | 0.3007 | 0.3054 | 0.3055 | 19052 | 12621 | 193785 | 166603 | -0.05037 | -0.002039 | -0.0424 | -0.03919 | 0.01046 | 0.01314 |
| HEIGHT | rs11049356 | rs3782517 | G | A | 12 | CCDC91 | Female only | True | 0.2995 | 0.3007 | 0.3054 | 0.3055 | 19052 | 12621 | 193785 | 166603 | -0.05037 | -0.001976 | -0.0424 | -0.03919 | 0.01046 | 0.01314 |
| HEIGHT | rs11049356 | rs11049520 | A | G | 12 | CCDC91 | Female only | True | 0.2996 | 0.3008 | 0.3054 | 0.3055 | 19052 | 12621 | 193785 | 166603 | -0.05037 | -0.002031 | -0.0424 | -0.03918 | 0.01046 | 0.01314 |
| HEIGHT | rs11049356 | rs11049524 | T | C | 12 | CCDC91 | Female only | True | 0.2995 | 0.3008 | 0.3054 | 0.3055 | 19052 | 12621 | 193785 | 166603 | -0.05037 | -0.001966 | -0.04244 | -0.03917 | 0.01046 | 0.01314 |
| HEIGHT | rs11049356 | rs11049526 | C | T | 12 | CCDC91 | Female only | True | 0.2996 | 0.3008 | 0.3055 | 0.3055 | 19052 | 12621 | 193785 | 166603 | -0.05042 | -0.001983 | -0.04241 | -0.0392 | 0.01046 | 0.01313 |
| HEIGHT | rs11049356 | rs9645730 | C | T | 12 | CCDC91 | Female only | True | 0.2996 | 0.3008 | 0.3051 | 0.3051 | 19052 | 12621 | 193785 | 166603 | -0.05014 | -0.002301 | -0.0424 | -0.03932 | 0.01046 | 0.01313 |
| HEIGHT | rs11049356 | rs11049527 | C | T | 12 | CCDC91 | Female only | True | 0.2996 | 0.3009 | 0.3055 | 0.3056 | 19052 | 12621 | 193785 | 166603 | -0.05034 | -0.002185 | -0.04242 | -0.0392 | 0.01045 | 0.01313 |
| HEIGHT | rs11049356 | rs10843151 | A | G | 12 | CCDC91 | Female only | True | 0.2996 | 0.3009 | 0.3055 | 0.3056 | 19052 | 12621 | 193785 | 166603 | -0.05038 | -0.002072 | -0.0424 | -0.0392 | 0.01046 | 0.01313 |
| HEIGHT | rs11049356 | rs10843152 | C | T | 12 | CCDC91 | Female only | True | 0.2996 | 0.3009 | 0.3055 | 0.3055 | 19052 | 12621 | 193785 | 166603 | -0.05034 | -0.002178 | -0.04241 | -0.0392 | 0.01045 | 0.01313 |
| HEIGHT | rs11049356 | rs10843153 | A | G | 12 | CCDC91 | Female only | True | 0.2997 | 0.3011 | 0.3055 | 0.3056 | 19052 | 12621 | 193785 | 166603 | -0.05035 | -0.002074 | -0.04241 | -0.03921 | 0.01046 | 0.01313 |
| HEIGHT | rs11049356 | rs10506029 | C | T | 12 | CCDC91 | Female only | True | 0.2997 | 0.3009 | 0.3055 | 0.3056 | 19052 | 12621 | 193785 | 166603 | -0.05035 | -0.002169 | -0.0424 | -0.03921 | 0.01045 | 0.01313 |
| HEIGHT | rs11049356 | rs11049531 | C | T | 12 | CCDC91 | Female only | True | 0.2995 | 0.3008 | 0.3055 | 0.3055 | 19052 | 12621 | 193785 | 166603 | -0.05034 | -0.002095 | -0.04244 | -0.03925 | 0.01046 | 0.01314 |
| HEIGHT | rs11049356 | rs11049532 | A | G | 12 | CCDC91 | Female only | True | 0.2995 | 0.3007 | 0.3055 | 0.3056 | 19052 | 12621 | 193785 | 166603 | -0.05034 | -0.002102 | -0.04248 | -0.03928 | 0.01046 | 0.01314 |
| HEIGHT | rs11049356 | rs11049537 | A | C | 12 | CCDC91 | Female only | True | 0.3014 | 0.303 | 0.3086 | 0.3086 | 19052 | 12621 | 193785 | 166603 | -0.04879 | -0.00201 | -0.0415 | -0.03998 | 0.01043 | 0.01311 |
| HEIGHT | rs11049356 | rs10843155 | G | A | 12 | CCDC91 | Female only | True | 0.3018 | 0.3035 | 0.3089 | 0.3089 | 19052 | 12621 | 193785 | 166603 | -0.04912 | -0.002335 | -0.04155 | -0.03994 | 0.01043 | 0.0131 |
| HEIGHT | rs11049356 | rs11049538 | C | T | 12 | CCDC91 | Female only | True | 0.3018 | 0.3035 | 0.3088 | 0.3088 | 19052 | 12621 | 193785 | 166603 | -0.04913 | -0.002368 | -0.04151 | -0.04 | 0.01042 | 0.0131 |
| HEIGHT | rs11049356 | rs11049541 | T | C | 12 | CCDC91 | Female only | True | 0.3023 | 0.3039 | 0.3091 | 0.309 | 19052 | 12621 | 193785 | 166603 | -0.04852 | -0.001803 | -0.04144 | -0.04001 | 0.01041 | 0.01309 |
| HEIGHT | rs11049356 | rs11049543 | C | A | 12 | CCDC91 | Female only | True | 0.3024 | 0.304 | 0.3092 | 0.3091 | 19052 | 12621 | 193785 | 166603 | -0.04869 | -0.001689 | -0.04143 | -0.04001 | 0.01041 | 0.01308 |
| HEIGHT | rs11049356 | rs11049544 | T | C | 12 | CCDC91 | Female only | True | 0.3024 | 0.3039 | 0.3092 | 0.3091 | 19052 | 12621 | 193785 | 166603 | -0.04867 | -0.001707 | -0.04139 | -0.04002 | 0.01041 | 0.01308 |
| HEIGHT | rs11049356 | rs11049545 | A | C | 12 | CCDC91 | Female only | True | 0.3016 | 0.3034 | 0.3083 | 0.3083 | 19052 | 12621 | 193785 | 166603 | -0.04847 | -0.003522 | -0.04137 | -0.03993 | 0.01041 | 0.01308 |
| HEIGHT | rs11049356 | rs10843157 | C | T | 12 | CCDC91 | Female only | True | 0.3021 | 0.3037 | 0.309 | 0.3089 | 19052 | 12621 | 193785 | 166603 | -0.04874 | -0.00255 | -0.04148 | -0.03995 | 0.01041 | 0.01308 |

| trait | lead_snp | ID | ALLELE1 | ALLELE0 | CHR | Gene_Symbol | independent_loci_in_sex_P1e2 | in_embl_any_or_gene | A1FREQ_VUMC_female | A1FREQ_VUMC_male | A1FREQ_UKB_female | A1FREQ_UKB_male | N_VUMC_female | N_VUMC_male | N_UKB_female | N_UKB_male | BETA_VUMC_female | BETA_VUMC_male | BETA_UKB_female | BETA_UKB_male | SE_VUMC_female | SE_VUMC_male |
|---|---|---|---|---|---|---|---|---|---|---|---|---|---|---|---|---|---|---|---|---|---|---|
| HEIGHT | rs11049356 | rs11049547 | A | G | 12 | CCDC91 | Female only | True | 0.302 | 0.3035 | 0.3087 | 0.3086 | 19052 | 12621 | 193785 | 166603 | -0.04867 | -0.002585 | -0.04145 | -0.03989 | 0.01041 | 0.01308 |
| HEIGHT | rs11049356 | rs11049548 | C | T | 12 | CCDC91 | Female only | True | 0.3036 | 0.3051 | 0.309 | 0.3089 | 19052 | 12621 | 193785 | 166603 | -0.04859 | -0.002478 | -0.04149 | -0.03994 | 0.01036 | 0.01302 |
| HEIGHT | rs11049356 | rs10843158 | T | C | 12 | CCDC91 | Female only | True | 0.302 | 0.3034 | 0.3088 | 0.3088 | 19052 | 12621 | 193785 | 166603 | -0.0487 | -0.002261 | -0.0415 | -0.03986 | 0.01041 | 0.01308 |
| HEIGHT | rs11049356 | rs10843159 | T | C | 12 | CCDC91 | Female only | True | 0.3019 | 0.3032 | 0.3088 | 0.3088 | 19052 | 12621 | 193785 | 166603 | -0.04857 | -0.00165 | -0.04155 | -0.03997 | 0.01041 | 0.01309 |
| HEIGHT | rs11049356 | rs61920564 | T | G | 12 | CCDC91 | Female only | True | 0.3018 | 0.3032 | 0.3087 | 0.3086 | 19052 | 12621 | 193785 | 166603 | -0.04852 | -0.002365 | -0.04151 | -0.04007 | 0.01042 | 0.01309 |
| HEIGHT | rs11049356 | rs11049550 | G | A | 12 | CCDC91 | Female only | True | 0.302 | 0.3035 | 0.3089 | 0.3088 | 19052 | 12621 | 193785 | 166603 | -0.04856 | -0.001969 | -0.04161 | -0.04005 | 0.01041 | 0.01309 |
| HEIGHT | rs11049356 | rs11049551 | T | C | 12 | CCDC91 | Female only | True | 0.3031 | 0.3045 | 0.3097 | 0.3096 | 19052 | 12621 | 193785 | 166603 | -0.04858 | -0.002118 | -0.04151 | -0.0401 | 0.01041 | 0.01309 |
| HEIGHT | rs11049356 | rs11049552 | T | C | 12 | CCDC91 | Female only | True | 0.302 | 0.3034 | 0.3089 | 0.3088 | 19052 | 12621 | 193785 | 166603 | -0.0486 | -0.002089 | -0.04158 | -0.04008 | 0.01041 | 0.01309 |
| HEIGHT | rs11049356 | rs11049553 | C | T | 12 | CCDC91 | Female only | True | 0.3021 | 0.3036 | 0.309 | 0.3089 | 19052 | 12621 | 193785 | 166603 | -0.04872 | -0.002612 | -0.04154 | -0.04005 | 0.01041 | 0.01308 |
| HEIGHT | rs11049356 | rs11049554 | T | G | 12 | CCDC91 | Female only | True | 0.3018 | 0.3034 | 0.3089 | 0.3089 | 19052 | 12621 | 193785 | 166603 | -0.04876 | -0.002602 | -0.04154 | -0.04008 | 0.01041 | 0.01309 |
| HEIGHT | rs11049356 | rs12371222 | G | A | 12 | CCDC91 | Female only | True | 0.3018 | 0.3034 | 0.3089 | 0.3088 | 19052 | 12621 | 193785 | 166603 | -0.04877 | -0.002525 | -0.04149 | -0.04015 | 0.01041 | 0.01309 |
| HEIGHT | rs11049356 | rs11049556 | A | C | 12 | CCDC91 | Female only | True | 0.3016 | 0.303 | 0.3087 | 0.3087 | 19052 | 12621 | 193785 | 166603 | -0.04863 | -0.001942 | -0.04159 | -0.04004 | 0.01042 | 0.0131 |
| HEIGHT | rs2280470 | rs2293087 | T | G | 15 | ACAN | Female only | True | 0.4247 | 0.4251 | 0.4146 | 0.4129 | 19052 | 12621 | 193785 | 166603 | 0.04255 | 0.01145 | 0.01851 | 0.01941 | 0.009649 | 0.01204 |
| HEIGHT | rs2280470 | rs12913174 | T | C | 15 | ACAN | Female only | True | 0.4196 | 0.4196 | 0.4114 | 0.4097 | 19052 | 12621 | 193785 | 166603 | 0.04235 | 0.01167 | 0.01848 | 0.01892 | 0.009614 | 0.01199 |
| HEIGHT | rs2280470 | rs11630187 | A | G | 15 | ACAN | Female only | True | 0.2208 | 0.2202 | 0.2129 | 0.2117 | 19052 | 12621 | 193785 | 166603 | 0.04933 | 0.02003 | 0.03319 | 0.03377 | 0.01144 | 0.01435 |
| HEIGHT | rs2280470 | rs12901266 | A | C | 15 | ACAN | Female only | True | 0.2216 | 0.221 | 0.2139 | 0.2127 | 19052 | 12621 | 193785 | 166603 | 0.05243 | 0.0205 | 0.03313 | 0.03421 | 0.01142 | 0.01433 |
| HEIGHT | rs2280470 | 15:89390434 | G | A | 15 | ACAN | Female only | True | 0.3848 | 0.3882 | | | 19052 | 12621 | | | 0.03526 | 0.005622 | | | 0.009804 | 0.01215 |
| HEIGHT | rs2280470 | rs12904298 | A | G | 15 | ACAN | Female only | True | 0.2235 | 0.2222 | 0.2167 | 0.2155 | 19052 | 12621 | 193785 | 166603 | 0.04947 | 0.02063 | 0.0328 | 0.03415 | 0.01139 | 0.01433 |
| HEIGHT | rs2280470 | rs2280470 | A | G | 15 | ACAN | Female only | True | 0.3302 | 0.33 | 0.3218 | 0.321 | 19052 | 12621 | 193785 | 166603 | 0.05688 | 0.02337 | 0.0428 | 0.03771 | 0.01009 | 0.01257 |
| HEIGHT | rs2280470 | rs11635309 | A | G | 15 | ACAN | Female only | True | 0.2215 | 0.2204 | 0.2144 | 0.2134 | 19052 | 12621 | 193785 | 166603 | 0.04888 | 0.02124 | 0.03336 | 0.03412 | 0.01141 | 0.01437 |
| HEIGHT | rs2280470 | rs2351491 | C | T | 15 | ACAN | Female only | True | 0.3644 | 0.3637 | 0.3525 | 0.352 | 19052 | 12621 | 193785 | 166603 | 0.03652 | 0.01241 | 0.01884 | 0.01776 | 0.009888 | 0.01231 |
| HEIGHT | rs2280470 | rs938608 | G | T | 15 | ACAN | Female only | True | 0.3624 | 0.3624 | 0.3523 | 0.3517 | 19052 | 12621 | 193785 | 166603 | 0.03663 | 0.01246 | 0.01906 | 0.01832 | 0.009897 | 0.01233 |
| HEIGHT | rs2280470 | rs2882676 | A | C | 15 | ACAN | Female only | True | 0.361 | 0.3608 | 0.3517 | 0.351 | 19052 | 12621 | 193785 | 166603 | 0.03608 | 0.0123 | 0.01881 | 0.01838 | 0.009865 | 0.01229 |
| HEIGHT | rs2280470 | rs698621 | T | G | 15 | ACAN | Female only | True | 0.3664 | 0.3657 | 0.3579 | 0.3575 | 19052 | 12621 | 193785 | 166603 | 0.03364 | 0.01001 | 0.01809 | 0.01684 | 0.00982 | 0.01225 |
| HEIGHT | rs3764419 | rs8068504 | A | C | 17 | ATAD5 | Female only | True | 0.2965 | 0.3001 | 0.3 | 0.2985 | 19052 | 12621 | 193785 | 166603 | -0.03162 | -0.007342 | -0.03134 | -0.02784 | 0.0105 | 0.013 |
| HEIGHT | rs3764419 | rs3764419 | A | C | 17 | ATAD5 | Female only | True | 0.3837 | 0.3861 | 0.3826 | 0.3811 | 19052 | 12621 | 193785 | 166603 | -0.06 | -0.01776 | -0.04541 | -0.04505 | 0.009771 | 0.01216 |
| HEIGHT | rs3764419 | rs62070643 | G | A | 17 | ATAD5 | Female only | True | 0.269 | 0.2654 | 0.27 | 0.2684 | 19052 | 12621 | 193785 | 166603 | -0.05641 | -0.02895 | -0.05678 | -0.05794 | 0.01069 | 0.0135 |
| HEIGHT | rs3764419 | rs62070644 | T | G | 17 | ATAD5 | Female only | True | 0.2664 | 0.2629 | 0.2701 | 0.2684 | 19052 | 12621 | 193785 | 166603 | -0.05716 | -0.02875 | -0.05688 | -0.05792 | 0.0108 | 0.01363 |
| HEIGHT | rs3764419 | rs62070645 | A | C | 17 | ATAD5 | Female only | True | 0.2683 | 0.2653 | 0.2699 | 0.2682 | 19052 | 12621 | 193785 | 166603 | -0.05791 | -0.02724 | -0.05688 | -0.05828 | 0.0107 | 0.0135 |

| trait | lead_snp | ID | ALLELE1 | ALLELE0 | CHR | Gene_Symbol | independent_loci_in_sex_P1e2 | in_embl_any_or_gene | A1FREQ_VUMC_female | A1FREQ_VUMC_male | A1FREQ_UKB_female | A1FREQ_UKB_male | N_VUMC_female | N_VUMC_male | N_UKB_female | N_UKB_male | BETA_VUMC_female | BETA_VUMC_male | BETA_UKB_female | BETA_UKB_male | SE_VUMC_female | SE_VUMC_male |
|---|---|---|---|---|---|---|---|---|---|---|---|---|---|---|---|---|---|---|---|---|---|---|
| HEIGHT | rs3764419 | rs7342976 | G | A | 17 | ATAD5 | Female only | True | 0.3841 | 0.3867 | 0.3829 | 0.3814 | 19052 | 12621 | 193785 | 166603 | -0.05952 | -0.01761 | -0.04549 | -0.04506 | 0.009771 | 0.01216 |
| HEIGHT | rs3764419 | rs62070648 | A | G | 17 | ATAD5 | Female only | True | 0.2659 | 0.263 | 0.27 | 0.2683 | 19052 | 12621 | 193785 | 166603 | -0.05859 | -0.027 | -0.05686 | -0.05818 | 0.0108 | 0.01362 |
| HEIGHT | rs3764419 | rs7223535 | A | G | 17 | ATAD5 | Female only | True | 0.2679 | 0.2645 | 0.2713 | 0.2696 | 19052 | 12621 | 193785 | 166603 | -0.05743 | -0.02833 | -0.0573 | -0.05801 | 0.01078 | 0.0136 |
| HEIGHT | rs3764419 | rs113934718 | A | C | 17 | ATAD5 | Female only | True | 0.2649 | 0.2617 | 0.2689 | 0.2671 | 19052 | 12621 | 193785 | 166603 | -0.05638 | -0.02686 | -0.05673 | -0.05739 | 0.01084 | 0.01368 |
| HEIGHT | rs3764419 | rs62070652 | T | C | 17 | ATAD5 | Female only | True | 0.2662 | 0.2629 | 0.2705 | 0.2688 | 19052 | 12621 | 193785 | 166603 | -0.05696 | -0.02745 | -0.05678 | -0.058 | 0.0108 | 0.01362 |
| HEIGHT | rs3764419 | rs9897628 | G | A | 17 | ATAD5 | Female only | True | 0.3805 | 0.3833 | 0.3824 | 0.3809 | 19052 | 12621 | 193785 | 166603 | -0.06033 | -0.01767 | -0.04549 | -0.04471 | 0.009852 | 0.01225 |
| HEIGHT | rs3764419 | rs35958868 | A | G | 17 | ATAD5 | Female only | True | 0.2667 | 0.2629 | 0.268 | 0.2666 | 19052 | 12621 | 193785 | 166603 | -0.05586 | -0.02442 | -0.05649 | -0.05737 | 0.01073 | 0.01354 |
| HEIGHT | rs3764419 | rs3760318 | A | G | 17 | ATAD5 | Female only | True | 0.3743 | 0.3762 | 0.3758 | 0.3746 | 19052 | 12621 | 193785 | 166603 | -0.05817 | -0.01739 | -0.04631 | -0.0458 | 0.00982 | 0.01224 |
| HEIGHT | rs3764419 | rs11867227 | A | G | 17 | ATAD5 | Female only | True | 0.2648 | 0.261 | 0.2685 | 0.2671 | 19052 | 12621 | 193785 | 166603 | -0.05734 | -0.02619 | -0.05662 | -0.05719 | 0.01082 | 0.01366 |
| HEIGHT | rs3764419 | rs35840638 | A | G | 17 | ATAD5 | Female only | True | 0.2636 | 0.2601 | 0.268 | 0.2666 | 19052 | 12621 | 193785 | 166603 | -0.05844 | -0.02589 | -0.05658 | -0.05751 | 0.01084 | 0.01368 |
| Monocy | rs143792309 | rs78517377 | C | A | 1 | EPHX4 | Female only | False | 0.07146 | 0.07013 | 0.07303 | 0.07324 | 15947 | 11630 | 187760 | 162101 | 0.0927 | 0.0167 | 0.07137 | 0.06309 | 0.01958 | 0.02353 |
| Monocy | rs143792309 | rs145328508 | T | C | 1 | EPHX4 | Female only | False | 0.0701 | 0.06897 | 0.06946 | 0.06969 | 15947 | 11630 | 187760 | 162101 | 0.09885 | 0.01745 | 0.08042 | 0.06793 | 0.01985 | 0.02379 |
| Monocy | rs143792309 | rs143792309 | T | C | 1 | EPHX4 | Female only | False | 0.06397 | 0.06403 | 0.06395 | 0.06411 | 15947 | 11630 | 187760 | 162101 | 0.116 | 0.04155 | 0.09492 | 0.08728 | 0.02076 | 0.02464 |
| Monocy | rs74767605 | rs78539283 | C | T | 1 | RPAP2 | Female only | True | 0.06857 | 0.06861 | 0.06783 | 0.06773 | 15947 | 11630 | 187760 | 162101 | 0.1105 | 0.0344 | 0.09223 | 0.09058 | 0.02004 | 0.02359 |
| Monocy | rs74767605 | rs80105455 | T | C | 1 | RPAP2 | Female only | True | 0.1018 | 0.1017 | 0.1027 | 0.1023 | 15947 | 11630 | 187760 | 162101 | 0.09316 | 0.03315 | 0.07472 | 0.06201 | 0.0166 | 0.0195 |
| Monocy | rs74767605 | rs74767605 | G | A | 1 | RPAP2 | Female only | True | 0.08847 | 0.08934 | 0.09034 | 0.09002 | 15947 | 11630 | 187760 | 162101 | 0.1069 | 0.02387 | 0.08293 | 0.07355 | 0.01781 | 0.02075 |
| Monocy | rs74767605 | rs6658232 | A | C | 1 | RPAP2 | Female only | True | 0.08969 | 0.09101 | 0.09125 | 0.09095 | 15947 | 11630 | 187760 | 162101 | 0.09292 | 0.02052 | 0.06962 | 0.0602 | 0.01735 | 0.02025 |
| Monocy | 1:93262946 | 1:93035861 | T | C | 1 | EVI5 | Female only | True | 0.1 | 0.09961 | | | 15947 | 11630 | | | 0.08619 | 0.02967 | | | 0.01658 | 0.01952 |
| Monocy | 1:93262946 | rs11164778 | T | C | 1 | EVI5 | Female only | True | 0.1047 | 0.1043 | 0.1011 | 0.1006 | 15947 | 11630 | 187760 | 162101 | 0.08781 | 0.03029 | 0.06741 | 0.0549 | 0.01669 | 0.01965 |
| Monocy | 1:93262946 | rs4847280 | G | A | 1 | EVI5 | Female only | True | 0.1047 | 0.1043 | 0.1011 | 0.1006 | 15947 | 11630 | 187760 | 162101 | 0.08781 | 0.03029 | 0.06746 | 0.05452 | 0.01669 | 0.01965 |
| Monocy | 1:93262946 | rs914966 | A | G | 1 | EVI5 | Female only | True | 0.1047 | 0.1043 | 0.1011 | 0.1006 | 15947 | 11630 | 187760 | 162101 | 0.08781 | 0.03029 | 0.06746 | 0.05452 | 0.01669 | 0.01965 |
| Monocy | 1:93262946 | rs9651256 | C | T | 1 | EVI5 | Female only | True | 0.1047 | 0.1043 | 0.1011 | 0.1006 | 15947 | 11630 | 187760 | 162101 | 0.08783 | 0.03029 | 0.06747 | 0.05454 | 0.0167 | 0.01965 |
| Monocy | 1:93262946 | rs2893217 | T | G | 1 | EVI5 | Female only | True | 0.1047 | 0.1043 | 0.1011 | 0.1006 | 15947 | 11630 | 187760 | 162101 | 0.08781 | 0.03029 | 0.06746 | 0.05452 | 0.01669 | 0.01965 |
| Monocy | 1:93262946 | rs2391164 | T | C | 1 | EVI5 | Female only | True | 0.1047 | 0.1043 | 0.1011 | 0.1006 | 15947 | 11630 | 187760 | 162101 | 0.08781 | 0.03029 | 0.06746 | 0.05452 | 0.01669 | 0.01965 |
| Monocy | 1:93262946 | rs12046739 | T | C | 1 | EVI5 | Female only | True | 0.1047 | 0.1043 | 0.1011 | 0.1006 | 15947 | 11630 | 187760 | 162101 | 0.08781 | 0.03029 | 0.06746 | 0.05452 | 0.01669 | 0.01965 |
| Monocy | 1:93262946 | rs10874715 | T | C | 1 | EVI5 | Female only | True | 0.1047 | 0.1043 | 0.1011 | 0.1006 | 15947 | 11630 | 187760 | 162101 | 0.08781 | 0.03029 | 0.06746 | 0.05452 | 0.01669 | 0.01965 |
| Monocy | 1:93262946 | rs10874716 | G | A | 1 | EVI5 | Female only | True | 0.1047 | 0.1043 | 0.1011 | 0.1006 | 15947 | 11630 | 187760 | 162101 | 0.08781 | 0.03029 | 0.06746 | 0.05452 | 0.01669 | 0.01965 |
| Monocy | 1:93262946 | rs4847305 | C | T | 1 | EVI5 | Female only | True | 0.1049 | 0.1045 | 0.1012 | 0.1008 | 15947 | 11630 | 187760 | 162101 | 0.08691 | 0.03041 | 0.06762 | 0.05468 | 0.01669 | 0.01963 |
| Monocy | 1:93262946 | rs7516537 | A | G | 1 | EVI5 | Female only | True | 0.1048 | 0.1045 | 0.1011 | 0.1006 | 15947 | 11630 | 187760 | 162101 | 0.08793 | 0.02977 | 0.06745 | 0.05456 | 0.01669 | 0.01964 |

| trait | lead_snp | ID | ALLELE1 | ALLELE0 | CHR | Gene_Symbol | independent_loci_in_sex_P1e2 | in_embl_any_or_gene | A1FREQ_VUMC_female | A1FREQ_VUMC_male | A1FREQ_UKB_female | A1FREQ_UKB_male | N_VUMC_female | N_VUMC_male | N_UKB_female | N_UKB_male | BETA_VUMC_female | BETA_VUMC_male | BETA_UKB_female | BETA_UKB_male | SE_VUMC_female | SE_VUMC_male |
|---|---|---|---|---|---|---|---|---|---|---|---|---|---|---|---|---|---|---|---|---|---|---|
| Monocy | 1:93262946 | rs7553367 | C | T | 1 | EVI5 | Female only | True | 0.1047 | 0.1043 | 0.1011 | 0.1006 | 15947 | 11630 | 187760 | 162101 | 0.08781 | 0.03029 | 0.06746 | 0.05452 | 0.01669 | 0.01965 |
| Monocy | 1:93262946 | rs3928821 | C | A | 1 | EVI5 | Female only | True | 0.1047 | 0.1043 | 0.1011 | 0.1006 | 15947 | 11630 | 187760 | 162101 | 0.08781 | 0.03029 | 0.06746 | 0.05452 | 0.01669 | 0.01965 |
| trait | lead_snp | ID | ALLELE1 | ALLELE0 | CHR | Gene_Symbol | independent_loci_in_sex_P1e2 | in_embl_any_or_gene | A1FREQ_VUMC_female | A1FREQ_VUMC_male | A1FREQ_UKB_female | A1FREQ_UKB_male | N_VUMC_female | N_VUMC_male | N_UKB_female | N_UKB_male | BETA_VUMC_female | BETA_VUMC_male | BETA_UKB_female | BETA_UKB_male | SE_VUMC_female | SE_VUMC_male |
| Monocy | 1:93262946 | rs6671114 | T | C | 1 | EVI5 | Female only | True | 0.1046 | 0.1042 | 0.1011 | 0.1006 | 15947 | 11630 | 187760 | 162101 | 0.08822 | 0.03041 | 0.06746 | 0.05452 | 0.0167 | 0.01965 |
| Monocy | 1:93262946 | rs10874717 | A | G | 1 | EVI5 | Female only | True | 0.1046 | 0.1042 | 0.1011 | 0.1006 | 15947 | 11630 | 187760 | 162101 | 0.08822 | 0.03041 | 0.06746 | 0.05452 | 0.0167 | 0.01965 |
| Monocy | 1:93262946 | rs6603993 | T | C | 1 | EVI5 | Female only | True | 0.1046 | 0.1042 | 0.1011 | 0.1006 | 15947 | 11630 | 187760 | 162101 | 0.08822 | 0.03041 | 0.06746 | 0.05452 | 0.0167 | 0.01965 |
| Monocy | 1:93262946 | rs4847327 | A | G | 1 | EVI5 | Female only | True | 0.1046 | 0.1042 | 0.1013 | 0.1008 | 15947 | 11630 | 187760 | 162101 | 0.08822 | 0.03041 | 0.06734 | 0.05381 | 0.0167 | 0.01965 |
| Monocy | 1:93262946 | rs6603994 | G | A | 1 | EVI5 | Female only | True | 0.1046 | 0.1042 | 0.1011 | 0.1006 | 15947 | 11630 | 187760 | 162101 | 0.08822 | 0.03041 | 0.06746 | 0.05452 | 0.0167 | 0.01965 |
| Monocy | 1:93262946 | rs6603995 | T | C | 1 | EVI5 | Female only | True | 0.1046 | 0.1042 | 0.101 | 0.1006 | 15947 | 11630 | 187760 | 162101 | 0.08841 | 0.03029 | 0.06744 | 0.05451 | 0.0167 | 0.01965 |
| Monocy | 1:93262946 | rs6422515 | T | C | 1 | EVI5 | Female only | True | 0.1046 | 0.1042 | 0.101 | 0.1006 | 15947 | 11630 | 187760 | 162101 | 0.0883 | 0.03041 | 0.06745 | 0.05456 | 0.0167 | 0.01965 |
| Monocy | 1:93262946 | rs3898499 | C | T | 1 | EVI5 | Female only | True | 0.1046 | 0.1042 | 0.101 | 0.1006 | 15947 | 11630 | 187760 | 162101 | 0.0883 | 0.03041 | 0.06745 | 0.05456 | 0.0167 | 0.01965 |
| Monocy | 1:93262946 | rs3898498 | T | C | 1 | EVI5 | Female only | True | 0.1046 | 0.1042 | 0.101 | 0.1006 | 15947 | 11630 | 187760 | 162101 | 0.0883 | 0.03041 | 0.06745 | 0.05456 | 0.0167 | 0.01965 |
| Monocy | 1:93262946 | rs963044 | T | C | 1 | EVI5 | Female only | True | 0.1042 | 0.1042 | 0.101 | 0.1005 | 15947 | 11630 | 187760 | 162101 | 0.08857 | 0.03022 | 0.06768 | 0.05479 | 0.01672 | 0.01965 |
| Monocy | 1:93262946 | rs963043 | C | T | 1 | EVI5 | Female only | True | 0.1046 | 0.1042 | 0.101 | 0.1006 | 15947 | 11630 | 187760 | 162101 | 0.0883 | 0.03041 | 0.06745 | 0.05456 | 0.0167 | 0.01965 |
| Monocy | 1:93262946 | rs6691038 | T | C | 1 | EVI5 | Female only | True | 0.1135 | 0.1124 | 0.1103 | 0.1096 | 15947 | 11630 | 187760 | 162101 | 0.08132 | 0.02939 | 0.05319 | 0.04093 | 0.01619 | 0.01908 |
| Monocy | 1:93262946 | rs6702031 | G | A | 1 | EVI5 | Female only | True | 0.1046 | 0.1042 | 0.1011 | 0.1006 | 15947 | 11630 | 187760 | 162101 | 0.08827 | 0.03038 | 0.06748 | 0.05458 | 0.0167 | 0.01965 |
| Monocy | 1:93262946 | rs6678864 | C | A | 1 | EVI5 | Female only | True | 0.1046 | 0.1042 | 0.101 | 0.1006 | 15947 | 11630 | 187760 | 162101 | 0.0883 | 0.03041 | 0.06745 | 0.05456 | 0.0167 | 0.01965 |
| Monocy | 1:93262946 | rs7418569 | T | C | 1 | EVI5 | Female only | True | 0.1046 | 0.1042 | 0.101 | 0.1006 | 15947 | 11630 | 187760 | 162101 | 0.0883 | 0.03041 | 0.06738 | 0.05459 | 0.0167 | 0.01965 |
| Monocy | 1:93262946 | rs9659957 | A | G | 1 | EVI5 | Female only | True | 0.1046 | 0.1042 | 0.101 | 0.1006 | 15947 | 11630 | 187760 | 162101 | 0.0883 | 0.03041 | 0.06738 | 0.05459 | 0.0167 | 0.01965 |
| Monocy | 1:93262946 | rs78123931 | G | A | 1 | EVI5 | Female only | True | 0.1046 | 0.1042 | | | 15947 | 11630 | | | 0.0883 | 0.03041 | | | 0.0167 | 0.01965 |
| Monocy | 1:93262946 | rs10747445 | G | A | 1 | EVI5 | Female only | True | 0.1047 | 0.1044 | 0.101 | 0.1006 | 15947 | 11630 | 187760 | 162101 | 0.08861 | 0.02972 | 0.06738 | 0.05461 | 0.0167 | 0.01965 |
| Monocy | 1:93262946 | rs2027060 | A | C | 1 | EVI5 | Female only | True | 0.1046 | 0.1043 | 0.1009 | 0.1004 | 15947 | 11630 | 187760 | 162101 | 0.08873 | 0.0311 | 0.06762 | 0.05496 | 0.01671 | 0.01965 |
| Monocy | 1:93262946 | rs2027059 | T | C | 1 | EVI5 | Female only | True | 0.1046 | 0.1043 | 0.1009 | 0.1004 | 15947 | 11630 | 187760 | 162101 | 0.08873 | 0.0311 | 0.06763 | 0.05496 | 0.01671 | 0.01965 |
| Monocy | 1:93262946 | rs2039452 | T | C | 1 | EVI5 | Female only | True | 0.1046 | 0.1043 | 0.1009 | 0.1004 | 15947 | 11630 | 187760 | 162101 | 0.08873 | 0.0311 | 0.06763 | 0.05496 | 0.01671 | 0.01965 |
| Monocy | 1:93262946 | rs2039451 | G | T | 1 | EVI5 | Female only | True | 0.1046 | 0.1043 | 0.1009 | 0.1004 | 15947 | 11630 | 187760 | 162101 | 0.08873 | 0.0311 | 0.06763 | 0.05496 | 0.01671 | 0.01965 |
| Monocy | 1:93262946 | rs1537498 | C | A | 1 | EVI5 | Female only | True | 0.1046 | 0.1043 | 0.1009 | 0.1004 | 15947 | 11630 | 187760 | 162101 | 0.08873 | 0.0311 | 0.06756 | 0.05486 | 0.01671 | 0.01965 |
| Monocy | 1:93262946 | rs7515478 | C | A | 1 | EVI5 | Female only | True | 0.1046 | 0.1043 | 0.1009 | 0.1004 | 15947 | 11630 | 187760 | 162101 | 0.08872 | 0.03111 | 0.06756 | 0.05486 | 0.01671 | 0.01965 |
| Monocy | 1:93262946 | rs7515866 | C | T | 1 | EVI5 | Female only | True | 0.1046 | 0.1043 | 0.1008 | 0.1004 | 15947 | 11630 | 187760 | 162101 | 0.08873 | 0.0311 | 0.06761 | 0.05497 | 0.01671 | 0.01965 |

| trait | lead_snp | ID | ALLELE1 | ALLELE0 | CHR | Gene_Symbol | independent_loci_in_sex_P1e2 | in_embl_any_or_gene | A1FREQ_VUMC_female | A1FREQ_VUMC_male | A1FREQ_UKB_female | A1FREQ_UKB_male | N_VUMC_female | N_VUMC_male | N_UKB_female | N_UKB_male | BETA_VUMC_female | BETA_VUMC_male | BETA_UKB_female | BETA_UKB_male | SE_VUMC_female | SE_VUMC_male |
|---|---|---|---|---|---|---|---|---|---|---|---|---|---|---|---|---|---|---|---|---|---|---|
| Monocy | 1:93262946 | rs2050597 | G | T | 1 | EVI5 | Female only | True | 0.1046 | 0.1043 | 0.1008 | 0.1004 | 15947 | 11630 | 187760 | 162101 | 0.08879 | 0.0311 | 0.06761 | 0.05497 | 0.01671 | 0.01965 |
| Monocy | 1:93262946 | rs2039453 | A | C | 1 | EVI5 | Female only | True | 0.1046 | 0.1043 | 0.1008 | 0.1004 | 15947 | 11630 | 187760 | 162101 | 0.08879 | 0.0311 | 0.06761 | 0.05497 | 0.01671 | 0.01965 |
| Monocy | 1:93262946 | rs6668283 | A | G | 1 | EVI5 | Female only | True | 0.1046 | 0.1043 | 0.1008 | 0.1004 | 15947 | 11630 | 187760 | 162101 | 0.08881 | 0.0311 | 0.06762 | 0.055 | 0.01671 | 0.01965 |
| Monocy | 1:93262946 | rs6668381 | A | G | 1 | EVI5 | Female only | True | 0.1046 | 0.1043 | 0.1009 | 0.1004 | 15947 | 11630 | 187760 | 162101 | 0.08882 | 0.03099 | 0.06763 | 0.05489 | 0.01671 | 0.01965 |
| Monocy | 1:93262946 | rs6603998 | A | G | 1 | EVI5 | Female only | True | 0.1046 | 0.1043 | 0.1008 | 0.1004 | 15947 | 11630 | 187760 | 162101 | 0.08879 | 0.03116 | 0.06761 | 0.05497 | 0.01671 | 0.01965 |
| Monocy | 1:93262946 | rs6603999 | C | T | 1 | EVI5 | Female only | True | 0.1046 | 0.1043 | 0.1009 | 0.1004 | 15947 | 11630 | 187760 | 162101 | 0.08871 | 0.03118 | 0.06767 | 0.05497 | 0.01671 | 0.01965 |
| Monocy | 1:93262946 | rs7534997 | T | C | 1 | EVI5 | Female only | True | 0.1046 | 0.1043 | 0.1009 | 0.1004 | 15947 | 11630 | 187760 | 162101 | 0.08879 | 0.03117 | 0.06763 | 0.05502 | 0.01671 | 0.01965 |
| Monocy | 1:93262946 | rs4847232 | C | T | 1 | EVI5 | Female only | True | 0.1046 | 0.1044 | 0.1009 | 0.1004 | 15947 | 11630 | 187760 | 162101 | 0.08872 | 0.03118 | 0.06763 | 0.05504 | 0.01672 | 0.01965 |
| Monocy | 1:93262946 | rs4847234 | T | C | 1 | EVI5 | Female only | True | 0.1046 | 0.1044 | 0.1009 | 0.1004 | 15947 | 11630 | 187760 | 162101 | 0.08872 | 0.03118 | 0.06763 | 0.05505 | 0.01672 | 0.01965 |
| Monocy | 1:93262946 | rs6668584 | C | T | 1 | EVI5 | Female only | True | 0.1046 | 0.1044 | 0.1009 | 0.1004 | 15947 | 11630 | 187760 | 162101 | 0.08872 | 0.03118 | 0.06763 | 0.05505 | 0.01672 | 0.01965 |
| Monocy | 1:93262946 | rs2095268 | A | G | 1 | EVI5 | Female only | True | 0.1042 | 0.1043 | 0.1008 | 0.1003 | 15947 | 11630 | 187760 | 162101 | 0.089 | 0.03099 | 0.06786 | 0.05528 | 0.01674 | 0.01966 |
| Monocy | 1:93262946 | rs2095267 | T | C | 1 | EVI5 | Female only | True | 0.1046 | 0.1044 | 0.1009 | 0.1004 | 15947 | 11630 | 187760 | 162101 | 0.08872 | 0.03118 | 0.06763 | 0.05505 | 0.01672 | 0.01965 |
| Monocy | 1:93262946 | rs2095266 | A | G | 1 | EVI5 | Female only | True | 0.1046 | 0.1044 | 0.1009 | 0.1005 | 15947 | 11630 | 187760 | 162101 | 0.08872 | 0.03117 | 0.06757 | 0.05474 | 0.01672 | 0.01965 |
| Monocy | 1:93262946 | rs10874721 | A | G | 1 | EVI5 | Female only | True | 0.1047 | 0.1044 | 0.1009 | 0.1004 | 15947 | 11630 | 187760 | 162101 | 0.08887 | 0.03112 | 0.06763 | 0.05505 | 0.01672 | 0.01965 |
| Monocy | 1:93262946 | rs9633481 | C | A | 1 | EVI5 | Female only | True | 0.106 | 0.1054 | 0.1017 | 0.1014 | 15947 | 11630 | 187760 | 162101 | 0.08928 | 0.03271 | 0.06812 | 0.05549 | 0.0167 | 0.01965 |
| Monocy | 1:93262946 | rs10874723 | T | C | 1 | EVI5 | Female only | True | 0.1047 | 0.1044 | 0.1009 | 0.1004 | 15947 | 11630 | 187760 | 162101 | 0.08887 | 0.03112 | 0.06763 | 0.05505 | 0.01672 | 0.01965 |
| Monocy | 1:93262946 | rs7514716 | T | C | 1 | EVI5 | Female only | True | 0.09949 | 0.09925 | 0.1009 | 0.1004 | 15947 | 11630 | 187760 | 162101 | 0.08842 | 0.03128 | 0.06763 | 0.05505 | 0.01662 | 0.01955 |
| Monocy | 1:93262946 | rs7538520 | C | A | 1 | EVI5 | Female only | True | 0.1047 | 0.1045 | 0.1009 | 0.1004 | 15947 | 11630 | 187760 | 162101 | 0.08889 | 0.03103 | 0.06763 | 0.05505 | 0.01672 | 0.01965 |
| Monocy | 1:93262946 | 1:93093234 | C | T | 1 | EVI5 | Female only | True | 0.1047 | 0.1044 | | | 15947 | 11630 | | | 0.0889 | 0.03109 | | | 0.01672 | 0.01965 |
| Monocy | 1:93262946 | rs1556564 | A | C | 1 | EVI5 | Female only | True | 0.1047 | 0.1044 | 0.1009 | 0.1004 | 15947 | 11630 | 187760 | 162101 | 0.0889 | 0.03109 | 0.06763 | 0.05505 | 0.01672 | 0.01965 |
| Monocy | 1:93262946 | rs1556565 | T | C | 1 | EVI5 | Female only | True | 0.1047 | 0.1044 | 0.1009 | 0.1004 | 15947 | 11630 | 187760 | 162101 | 0.0889 | 0.03109 | 0.06763 | 0.05505 | 0.01672 | 0.01965 |
| Monocy | 1:93262946 | rs10874728 | C | T | 1 | EVI5 | Female only | True | 0.1048 | 0.1045 | 0.1009 | 0.1004 | 15947 | 11630 | 187760 | 162101 | 0.08886 | 0.03096 | 0.06761 | 0.05505 | 0.01672 | 0.01966 |
| Monocy | 1:93262946 | rs4526624 | A | G | 1 | EVI5 | Female only | True | 0.1049 | 0.1046 | 0.1009 | 0.1004 | 15947 | 11630 | 187760 | 162101 | 0.08875 | 0.03089 | 0.06763 | 0.05505 | 0.01672 | 0.01966 |
| Monocy | 1:93262946 | rs4417075 | C | T | 1 | EVI5 | Female only | True | 0.1049 | 0.1046 | 0.1009 | 0.1004 | 15947 | 11630 | 187760 | 162101 | 0.08875 | 0.03089 | 0.06763 | 0.05505 | 0.01672 | 0.01966 |
| Monocy | 1:93262946 | rs3936946 | A | G | 1 | EVI5 | Female only | True | 0.1049 | 0.1046 | 0.1009 | 0.1004 | 15947 | 11630 | 187760 | 162101 | 0.08875 | 0.03089 | 0.06763 | 0.05505 | 0.01672 | 0.01966 |
| Monocy | 1:93262946 | rs10782938 | C | T | 1 | EVI5 | Female only | True | 0.1049 | 0.1046 | 0.1009 | 0.1004 | 15947 | 11630 | 187760 | 162101 | 0.08875 | 0.03089 | 0.06762 | 0.05521 | 0.01672 | 0.01966 |
| Monocy | 1:93262946 | rs10874729 | G | A | 1 | EVI5 | Female only | True | 0.1049 | 0.1046 | 0.1009 | 0.1005 | 15947 | 11630 | 187760 | 162101 | 0.08875 | 0.03089 | 0.06752 | 0.05559 | 0.01672 | 0.01966 |
| Monocy | 1:93262946 | rs10782940 | G | T | 1 | EVI5 | Female only | True | 0.1049 | 0.1046 | 0.1009 | 0.1005 | 15947 | 11630 | 187760 | 162101 | 0.08875 | 0.03089 | 0.06752 | 0.05559 | 0.01672 | 0.01966 |
| Monocy | 1:93262946 | rs7529332 | G | A | 1 | EVI5 | Female only | True | 0.1049 | 0.1046 | 0.1009 | 0.1005 | 15947 | 11630 | 187760 | 162101 | 0.08874 | 0.03082 | 0.06751 | 0.05562 | 0.01672 | 0.01966 |

| trait | lead_snp | ID | ALLELE1 | ALLELE0 | CHR | Gene_Symbol | independent_loci_in_sex_P1e2 | in_embl_any_or_gene | A1FREQ_VUMC_female | A1FREQ_VUMC_male | A1FREQ_UKB_female | A1FREQ_UKB_male | N_VUMC_female | N_VUMC_male | N_UKB_female | N_UKB_male | BETA_VUMC_female | BETA_VUMC_male | BETA_UKB_female | BETA_UKB_male | SE_VUMC_female | SE_VUMC_male |
|---|---|---|---|---|---|---|---|---|---|---|---|---|---|---|---|---|---|---|---|---|---|---|
| Monocy | 1:93262946 | rs7529630 | G | A | 1 | EVI5 | Female only | True | 0.1049 | 0.1046 | 0.1009 | 0.1005 | 15947 | 11630 | 187760 | 162101 | 0.08875 | 0.03089 | 0.06751 | 0.05563 | 0.01672 | 0.01966 |
| Monocy | 1:93262946 | rs10874730 | T | C | 1 | EVI5 | Female only | True | 0.1051 | 0.1046 | 0.1011 | 0.1006 | 15947 | 11630 | 187760 | 162101 | 0.08834 | 0.03206 | 0.06696 | 0.05565 | 0.0167 | 0.01965 |
| Monocy | 1:93262946 | rs9439917 | T | C | 1 | EVI5 | Female only | True | 0.09893 | 0.099 | 0.1008 | 0.1004 | 15947 | 11630 | 187760 | 162101 | 0.08853 | 0.03127 | 0.06769 | 0.05593 | 0.01668 | 0.01959 |
| Monocy | 1:93262946 | rs11164790 | A | G | 1 | EVI5 | Female only | True | 0.105 | 0.1046 | 0.1009 | 0.1005 | 15947 | 11630 | 187760 | 162101 | 0.08862 | 0.03074 | 0.06746 | 0.05569 | 0.01672 | 0.01966 |
| Monocy | 1:93262946 | rs4120650 | C | T | 1 | EVI5 | Female only | True | 0.105 | 0.1046 | 0.1009 | 0.1005 | 15947 | 11630 | 187760 | 162101 | 0.08862 | 0.03074 | 0.06746 | 0.05577 | 0.01672 | 0.01966 |
| Monocy | 1:93262946 | rs10747447 | G | A | 1 | EVI5 | Female only | True | 0.105 | 0.1046 | 0.1009 | 0.1005 | 15947 | 11630 | 187760 | 162101 | 0.08864 | 0.03074 | 0.06737 | 0.0558 | 0.01672 | 0.01966 |
| Monocy | 1:93262946 | rs10747448 | T | C | 1 | EVI5 | Female only | True | 0.105 | 0.1046 | 0.1009 | 0.1005 | 15947 | 11630 | 187760 | 162101 | 0.08864 | 0.03074 | 0.06737 | 0.0558 | 0.01672 | 0.01966 |
| Monocy | 1:93262946 | rs10874731 | G | A | 1 | EVI5 | Female only | True | 0.105 | 0.1046 | 0.1009 | 0.1005 | 15947 | 11630 | 187760 | 162101 | 0.08864 | 0.03074 | 0.06735 | 0.05582 | 0.01672 | 0.01966 |
| Monocy | 1:93262946 | rs4847247 | G | A | 1 | EVI5 | Female only | True | 0.105 | 0.1046 | 0.1009 | 0.1005 | 15947 | 11630 | 187760 | 162101 | 0.08864 | 0.03074 | 0.06737 | 0.0558 | 0.01672 | 0.01966 |
| Monocy | 1:93262946 | rs10747449 | C | A | 1 | EVI5 | Female only | True | 0.105 | 0.1047 | 0.1009 | 0.1005 | 15947 | 11630 | 187760 | 162101 | 0.08851 | 0.03027 | 0.06742 | 0.05582 | 0.01671 | 0.01966 |
| Monocy | 1:93262946 | rs10747450 | A | G | 1 | EVI5 | Female only | True | 0.105 | 0.1047 | 0.1009 | 0.1005 | 15947 | 11630 | 187760 | 162101 | 0.08851 | 0.03027 | 0.06741 | 0.05582 | 0.01671 | 0.01966 |
| Monocy | 1:93262946 | rs75758615 | T | C | 1 | EVI5 | Female only | True | 0.08953 | 0.09107 | 0.0916 | 0.09133 | 15947 | 11630 | 187760 | 162101 | 0.09357 | 0.02053 | 0.06978 | 0.06027 | 0.01736 | 0.02024 |
| Monocy | 1:93262946 | rs78886094 | G | A | 1 | EVI5 | Female only | True | 0.09798 | 0.09796 | 0.1001 | 0.0997 | 15947 | 11630 | 187760 | 162101 | 0.08872 | 0.0317 | 0.06799 | 0.05667 | 0.01681 | 0.01973 |
| Monocy | 1:93262946 | rs10747451 | A | G | 1 | EVI5 | Female only | True | 0.1041 | 0.1036 | 0.1001 | 0.09978 | 15947 | 11630 | 187760 | 162101 | 0.08882 | 0.03114 | 0.06776 | 0.0564 | 0.01684 | 0.0198 |
| Monocy | 1:93262946 | rs1999819 | C | T | 1 | EVI5 | Female only | True | 0.09965 | 0.09927 | 0.1006 | 0.1003 | 15947 | 11630 | 187760 | 162101 | 0.08775 | 0.03088 | 0.06762 | 0.05614 | 0.01661 | 0.01954 |
| Monocy | 1:93262946 | rs6679839 | G | A | 1 | EVI5 | Female only | True | 0.105 | 0.1047 | 0.1006 | 0.1003 | 15947 | 11630 | 187760 | 162101 | 0.0881 | 0.03044 | 0.06762 | 0.05621 | 0.0167 | 0.01965 |
| Monocy | 1:93262946 | rs2152765 | A | G | 1 | EVI5 | Female only | True | 0.105 | 0.1046 | 0.1006 | 0.1003 | 15947 | 11630 | 187760 | 162101 | 0.08784 | 0.03046 | 0.06758 | 0.05623 | 0.01671 | 0.01966 |
| Monocy | 1:93262946 | rs1415295 | C | A | 1 | EVI5 | Female only | True | 0.105 | 0.1046 | 0.1008 | 0.1004 | 15947 | 11630 | 187760 | 162101 | 0.08814 | 0.03043 | 0.06739 | 0.05599 | 0.0167 | 0.01966 |
| Monocy | 1:93262946 | rs945724 | C | T | 1 | EVI5 | Female only | True | 0.1049 | 0.1046 | 0.1003 | 0.09979 | 15947 | 11630 | 187760 | 162101 | 0.08829 | 0.03 | 0.06756 | 0.05707 | 0.01673 | 0.01969 |
| Monocy | 1:93262946 | rs1932966 | G | A | 1 | EVI5 | Female only | True | 0.1048 | 0.1046 | 0.1003 | 0.0998 | 15947 | 11630 | 187760 | 162101 | 0.08798 | 0.03003 | 0.06749 | 0.05704 | 0.01673 | 0.01969 |
| Monocy | 1:93262946 | rs6604003 | T | C | 1 | EVI5 | Female only | True | 0.1048 | 0.1046 | 0.1003 | 0.09982 | 15947 | 11630 | 187760 | 162101 | 0.08807 | 0.03005 | 0.06741 | 0.057 | 0.01673 | 0.01969 |
| Monocy | 1:93262946 | rs7416169 | T | C | 1 | EVI5 | Female only | True | 0.1044 | 0.1045 | 0.1002 | 0.09974 | 15947 | 11630 | 187760 | 162101 | 0.08852 | 0.02982 | 0.06777 | 0.05731 | 0.01675 | 0.01969 |
| Monocy | 1:93262946 | rs2391201 | G | A | 1 | EVI5 | Female only | True | 0.1049 | 0.1046 | 0.1003 | 0.09983 | 15947 | 11630 | 187760 | 162101 | 0.08827 | 0.03 | 0.06754 | 0.05699 | 0.01673 | 0.01969 |
| Monocy | 1:93262946 | rs6604004 | G | A | 1 | EVI5 | Female only | True | 0.1049 | 0.1046 | 0.1003 | 0.09984 | 15947 | 11630 | 187760 | 162101 | 0.08827 | 0.03 | 0.06751 | 0.057 | 0.01673 | 0.01969 |
| Monocy | 1:93262946 | 1:93151766 | T | C | 1 | EVI5 | Female only | True | 0.1045 | 0.1046 | | | 15947 | 11630 | | | 0.08931 | 0.02973 | | | 0.01675 | 0.01969 |
| Monocy | 1:93262946 | rs7540407 | A | G | 1 | EVI5 | Female only | True | 0.1052 | 0.1049 | 0.1003 | 0.09983 | 15947 | 11630 | 187760 | 162101 | 0.08826 | 0.03014 | 0.06751 | 0.05699 | 0.01673 | 0.0197 |
| Monocy | 1:93262946 | rs4847266 | G | A | 1 | EVI5 | Female only | True | 0.1052 | 0.1049 | 0.1003 | 0.09983 | 15947 | 11630 | 187760 | 162101 | 0.08826 | 0.03014 | 0.06754 | 0.05699 | 0.01673 | 0.0197 |

| trait | lead_snp | ID | ALLELE1 | ALLELE0 | CHR | Gene_Symbol | independent_loci_in_sex_P1e2 | in_embl_any_or_gene | A1FREQ_VUMC_female | A1FREQ_VUMC_male | A1FREQ_UKB_female | A1FREQ_UKB_male | N_VUMC_female | N_VUMC_male | N_UKB_female | N_UKB_male | BETA_VUMC_female | BETA_VUMC_male | BETA_UKB_female | BETA_UKB_male | SE_VUMC_female | SE_VUMC_male |
|---|---|---|---|---|---|---|---|---|---|---|---|---|---|---|---|---|---|---|---|---|---|---|
| Monocy | 1:93262946 | rs2097838 | T | C | 1 | EVI5 | Female only | True | 0.1052 | 0.1049 | 0.1003 | 0.09984 | 15947 | 11630 | 187760 | 162101 | 0.08826 | 0.03014 | 0.06753 | 0.05701 | 0.01673 | 0.0197 |
| Monocy | 1:93262946 | rs2391200 | C | T | 1 | EVI5 | Female only | True | 0.1049 | 0.1049 | 0.1004 | 0.09993 | 15947 | 11630 | 187760 | 162101 | 0.08879 | 0.02976 | 0.06757 | 0.05715 | 0.01675 | 0.0197 |
| Monocy | 1:93262946 | rs2065916 | T | C | 1 | EVI5 | Female only | True | 0.1052 | 0.1049 | 0.1003 | 0.09981 | 15947 | 11630 | 187760 | 162101 | 0.08825 | 0.03012 | 0.06749 | 0.05707 | 0.01673 | 0.0197 |
| Monocy | 1:93262946 | rs2065917 | T | C | 1 | EVI5 | Female only | True | 0.1052 | 0.1049 | 0.1003 | 0.0998 | 15947 | 11630 | 187760 | 162101 | 0.08826 | 0.03014 | 0.06758 | 0.05708 | 0.01673 | 0.0197 |
| Monocy | 1:93262946 | rs2391199 | T | C | 1 | EVI5 | Female only | True | 0.0993 | 0.09893 | 0.1002 | 0.09977 | 15947 | 11630 | 187760 | 162101 | 0.08787 | 0.03075 | 0.06763 | 0.05718 | 0.01663 | 0.01957 |
| Monocy | 1:93262946 | rs914064 | C | T | 1 | EVI5 | Female only | True | 0.1058 | 0.1054 | 0.101 | 0.1005 | 15947 | 11630 | 187760 | 162101 | 0.08796 | 0.02867 | 0.06733 | 0.05714 | 0.01673 | 0.01969 |
| Monocy | 1:93262946 | rs914065 | T | C | 1 | EVI5 | Female only | True | 0.1053 | 0.1049 | 0.1003 | 0.0998 | 15947 | 11630 | 187760 | 162101 | 0.08824 | 0.03019 | 0.06759 | 0.05709 | 0.01673 | 0.0197 |
| Monocy | 1:93262946 | rs2005720 | C | T | 1 | EVI5 | Female only | True | 0.1053 | 0.1049 | 0.1002 | 0.09969 | 15947 | 11630 | 187760 | 162101 | 0.0883 | 0.03019 | 0.06776 | 0.05724 | 0.01673 | 0.0197 |
| Monocy | 1:93262946 | rs1017008 | C | T | 1 | EVI5 | Female only | True | 0.1053 | 0.1049 | 0.1001 | 0.09958 | 15947 | 11630 | 187760 | 162101 | 0.0883 | 0.03023 | 0.06802 | 0.05738 | 0.01673 | 0.0197 |
| Monocy | 1:93262946 | rs927339 | A | G | 1 | EVI5 | Female only | True | 0.1053 | 0.1049 | 0.1003 | 0.09981 | 15947 | 11630 | 187760 | 162101 | 0.0883 | 0.03023 | 0.06753 | 0.05704 | 0.01673 | 0.0197 |
| Monocy | 1:93262946 | rs10218567 | C | T | 1 | EVI5 | Female only | True | 0.1053 | 0.1049 | 0.1003 | 0.0998 | 15947 | 11630 | 187760 | 162101 | 0.08831 | 0.03023 | 0.06753 | 0.05705 | 0.01673 | 0.0197 |
| Monocy | 1:93262946 | rs10218670 | A | G | 1 | EVI5 | Female only | True | 0.1053 | 0.1049 | 0.1003 | 0.0998 | 15947 | 11630 | 187760 | 162101 | 0.0883 | 0.03023 | 0.06755 | 0.05704 | 0.01673 | 0.0197 |
| Monocy | 1:93262946 | rs6671421 | A | G | 1 | EVI5 | Female only | True | 0.1056 | 0.1052 | 0.1003 | 0.09981 | 15947 | 11630 | 187760 | 162101 | 0.08833 | 0.03029 | 0.06754 | 0.05704 | 0.01674 | 0.01971 |
| Monocy | 1:93262946 | rs7521345 | T | C | 1 | EVI5 | Female only | True | 0.1056 | 0.1052 | 0.1003 | 0.09981 | 15947 | 11630 | 187760 | 162101 | 0.08825 | 0.0303 | 0.06752 | 0.05709 | 0.01674 | 0.01971 |
| Monocy | 1:93262946 | 1:93172758 | C | A | 1 | EVI5 | Female only | True | 0.1024 | 0.1025 | | | 15947 | 11630 | | | 0.08611 | 0.03128 | | | 0.01649 | 0.01938 |
| Monocy | 1:93262946 | rs723909 | G | T | 1 | EVI5 | Female only | True | 0.1056 | 0.1052 | 0.1003 | 0.0998 | 15947 | 11630 | 187760 | 162101 | 0.08825 | 0.03029 | 0.06752 | 0.05711 | 0.01674 | 0.01971 |
| Monocy | 1:93262946 | rs2077529 | C | T | 1 | EVI5 | Female only | True | 0.1056 | 0.1052 | 0.1003 | 0.09981 | 15947 | 11630 | 187760 | 162101 | 0.08825 | 0.03029 | 0.0675 | 0.05712 | 0.01674 | 0.01971 |
| Monocy | 1:93262946 | rs6665579 | C | A | 1 | EVI5 | Female only | True | 0.1056 | 0.1052 | 0.1003 | 0.09981 | 15947 | 11630 | 187760 | 162101 | 0.08826 | 0.03029 | 0.0675 | 0.05709 | 0.01674 | 0.01971 |
| Monocy | 1:93262946 | rs1337109 | C | A | 1 | EVI5 | Female only | True | 0.1059 | 0.1055 | 0.1003 | 0.09982 | 15947 | 11630 | 187760 | 162101 | 0.08826 | 0.03029 | 0.06753 | 0.05703 | 0.01674 | 0.01971 |
| Monocy | 1:93262946 | rs4847285 | A | C | 1 | EVI5 | Female only | True | 0.1062 | 0.1057 | 0.1005 | 0.1 | 15947 | 11630 | 187760 | 162101 | 0.08768 | 0.03124 | 0.06748 | 0.05638 | 0.01672 | 0.0197 |
| Monocy | 1:93262946 | rs11164799 | A | G | 1 | EVI5 | Female only | True | 0.1054 | 0.1054 | 0.1003 | 0.09976 | 15947 | 11630 | 187760 | 162101 | 0.08861 | 0.03009 | 0.06777 | 0.05723 | 0.01677 | 0.01971 |
| Monocy | 1:93262946 | rs6673043 | A | G | 1 | EVI5 | Female only | True | 0.0958 | 0.09755 | 0.09081 | 0.09039 | 15947 | 11630 | 187760 | 162101 | 0.09372 | 0.01901 | 0.07053 | 0.06094 | 0.01749 | 0.0204 |
| Monocy | 1:93262946 | rs1182636 | T | C | 1 | EVI5 | Female only | True | 0.08901 | 0.09077 | 0.09067 | 0.09028 | 15947 | 11630 | 187760 | 162101 | 0.09352 | 0.02034 | 0.07081 | 0.06117 | 0.01742 | 0.0203 |
| Monocy | 1:93262946 | rs13138867 | C | T | 1 | EVI5 | Female only | True | 0.1062 | 0.1057 | | | 15947 | 11630 | | | 0.08773 | 0.03126 | | | 0.01672 | 0.0197 |
| Monocy | 1:93262946 | rs6604010 | G | A | 1 | EVI5 | Female only | True | 0.1067 | 0.1062 | 0.1003 | 0.09988 | 15947 | 11630 | 187760 | 162101 | 0.08766 | 0.03136 | 0.06774 | 0.05648 | 0.01673 | 0.01973 |
| Monocy | 1:93262946 | rs6604011 | T | C | 1 | EVI5 | Female only | True | 0.1067 | 0.1062 | 0.1003 | 0.09986 | 15947 | 11630 | 187760 | 162101 | 0.08777 | 0.03136 | 0.06778 | 0.05654 | 0.01673 | 0.01973 |
| Monocy | 1:93262946 | rs6604012 | T | C | 1 | EVI5 | Female only | True | 0.1066 | 0.106 | 0.1003 | 0.09978 | 15947 | 11630 | 187760 | 162101 | 0.08843 | 0.03131 | 0.06785 | 0.0567 | 0.01674 | 0.01975 |
| Monocy | 1:93262946 | rs757469 | T | C | 1 | EVI5 | Female only | True | 0.1064 | 0.1059 | 0.1003 | 0.09976 | 15947 | 11630 | 187760 | 162101 | 0.09026 | 0.03044 | 0.06774 | 0.0571 | 0.01676 | 0.01975 |
| Monocy | 1:93262946 | rs2008797 | A | G | 1 | EVI5 | Female only | True | 0.1063 | 0.1058 | 0.1003 | 0.09974 | 15947 | 11630 | 187760 | 162101 | 0.08903 | 0.03033 | 0.06766 | 0.05711 | 0.01677 | 0.01976 |

| trait | lead_snp | ID | ALLELE1 | ALLELE0 | CHR | Gene_Symbol | independent_loci_in_sex_P1e2 | in_embl_any_or_gene | A1FREQ_VUMC_female | A1FREQ_VUMC_male | A1FREQ_UKB_female | A1FREQ_UKB_male | N_VUMC_female | N_VUMC_male | N_UKB_female | N_UKB_male | BETA_VUMC_female | BETA_VUMC_male | BETA_UKB_female | BETA_UKB_male | SE_VUMC_female | SE_VUMC_male |
|---|---|---|---|---|---|---|---|---|---|---|---|---|---|---|---|---|---|---|---|---|---|---|
| Monocy | 1:93262946 | rs6703467 | G | A | 1 | EVI5 | Female only | True | 0.1065 | 0.106 | 0.1004 | 0.09985 | 15947 | 11630 | 187760 | 162101 | 0.08813 | 0.03136 | 0.06762 | 0.05657 | 0.01675 | 0.01975 |
| Monocy | 1:93262946 | rs2107520 | C | T | 1 | EVI5 | Female only | True | 0.1065 | 0.106 | 0.1004 | 0.09985 | 15947 | 11630 | 187760 | 162101 | 0.08813 | 0.03135 | 0.06762 | 0.05655 | 0.01675 | 0.01975 |
| Monocy | 1:93262946 | rs1932967 | C | T | 1 | EVI5 | Female only | True | 0.1066 | 0.1061 | 0.1003 | 0.09984 | 15947 | 11630 | 187760 | 162101 | 0.08812 | 0.03127 | 0.06759 | 0.05656 | 0.01676 | 0.01976 |
| Monocy | 1:93262946 | rs2391196 | C | A | 1 | EVI5 | Female only | True | 0.1066 | 0.1061 | 0.1003 | 0.09984 | 15947 | 11630 | 187760 | 162101 | 0.08812 | 0.03127 | 0.06759 | 0.05656 | 0.01676 | 0.01976 |
| Monocy | 1:93262946 | rs10735783 | T | C | 1 | EVI5 | Female only | True | 0.1066 | 0.1061 | 0.1003 | 0.09985 | 15947 | 11630 | 187760 | 162101 | 0.0881 | 0.03124 | 0.0676 | 0.0565 | 0.01676 | 0.01976 |
| Monocy | 1:93262946 | rs7516141 | A | G | 1 | EVI5 | Female only | True | 0.1066 | 0.1061 | 0.1003 | 0.09984 | 15947 | 11630 | 187760 | 162101 | 0.08812 | 0.03127 | 0.06759 | 0.05656 | 0.01676 | 0.01976 |
| Monocy | 1:93262946 | rs7549844 | C | A | 1 | EVI5 | Female only | True | 0.1066 | 0.1061 | 0.1003 | 0.09984 | 15947 | 11630 | 187760 | 162101 | 0.08812 | 0.03127 | 0.06759 | 0.05656 | 0.01676 | 0.01976 |
| Monocy | 1:93262946 | rs6661600 | A | C | 1 | EVI5 | Female only | True | 0.1063 | 0.1059 | 0.1002 | 0.09969 | 15947 | 11630 | 187760 | 162101 | 0.0887 | 0.03029 | 0.06764 | 0.05717 | 0.01678 | 0.01977 |
| Monocy | 1:93262946 | rs2107521 | C | T | 1 | EVI5 | Female only | True | 0.1066 | 0.1061 | 0.1003 | 0.09984 | 15947 | 11630 | 187760 | 162101 | 0.08837 | 0.03123 | 0.06764 | 0.05646 | 0.01675 | 0.01975 |
| Monocy | 1:93262946 | rs7526582 | A | C | 1 | EVI5 | Female only | True | 0.1063 | 0.1059 | 0.1002 | 0.09969 | 15947 | 11630 | 187760 | 162101 | 0.0887 | 0.03029 | 0.06761 | 0.05711 | 0.01678 | 0.01977 |
| trait | lead_snp | ID | ALLELE1 | ALLELE0 | CHR | Gene_Symbol | independent_loci_in_sex_P1e2 | in_embl_any_or_gene | A1FREQ_VUMC_female | A1FREQ_VUMC_male | A1FREQ_UKB_female | A1FREQ_UKB_male | N_VUMC_female | N_VUMC_male | N_UKB_female | N_UKB_male | BETA_VUMC_female | BETA_VUMC_male | BETA_UKB_female | BETA_UKB_male | SE_VUMC_female | SE_VUMC_male |
| Monocy | 1:93262946 | rs2180977 | C | A | 1 | EVI5 | Female only | True | 0.1066 | 0.1061 | 0.1003 | 0.09985 | 15947 | 11630 | 187760 | 162101 | 0.08841 | 0.03122 | 0.0676 | 0.05648 | 0.01675 | 0.01975 |
| Monocy | 1:93262946 | rs976208 | C | T | 1 | EVI5 | Female only | True | 0.1066 | 0.1062 | 0.1002 | 0.09968 | 15947 | 11630 | 187760 | 162101 | 0.08868 | 0.0303 | 0.06762 | 0.05713 | 0.01679 | 0.01978 |
| Monocy | 1:93262946 | rs976209 | A | G | 1 | EVI5 | Female only | True | 0.1066 | 0.1062 | 0.1002 | 0.09969 | 15947 | 11630 | 187760 | 162101 | 0.08868 | 0.0303 | 0.06761 | 0.05711 | 0.01679 | 0.01978 |
| Monocy | 1:93262946 | rs4264007 | G | A | 1 | EVI5 | Female only | True | 0.107 | 0.1065 | 0.1004 | 0.09994 | 15947 | 11630 | 187760 | 162101 | 0.08751 | 0.03137 | 0.06746 | 0.05629 | 0.01675 | 0.01975 |
| Monocy | 1:93262946 | rs10782942 | A | G | 1 | EVI5 | Female only | True | 0.1068 | 0.1063 | 0.1003 | 0.0998 | 15947 | 11630 | 187760 | 162101 | 0.08804 | 0.03037 | 0.06751 | 0.05694 | 0.01677 | 0.01976 |
| Monocy | 1:93262946 | rs4638137 | A | G | 1 | EVI5 | Female only | True | 0.1068 | 0.1063 | 0.1003 | 0.09979 | 15947 | 11630 | 187760 | 162101 | 0.08804 | 0.03036 | 0.06752 | 0.05696 | 0.01677 | 0.01976 |
| Monocy | 1:93262946 | rs1182639 | A | G | 1 | EVI5 | Female only | True | 0.08908 | 0.09095 | 0.09085 | 0.09046 | 15947 | 11630 | 187760 | 162101 | 0.09281 | 0.01998 | 0.07034 | 0.06071 | 0.01742 | 0.0203 |
| Monocy | 1:93262946 | rs2893224 | T | C | 1 | EVI5 | Female only | True | 0.1071 | 0.1065 | 0.1004 | 0.09995 | 15947 | 11630 | 187760 | 162101 | 0.08752 | 0.03137 | 0.06745 | 0.05637 | 0.01675 | 0.01975 |
| Monocy | 1:93262946 | rs6674324 | G | T | 1 | EVI5 | Female only | True | 0.107 | 0.1065 | 0.1004 | 0.09994 | 15947 | 11630 | 187760 | 162101 | 0.08747 | 0.03136 | 0.06746 | 0.05635 | 0.01675 | 0.01975 |
| Monocy | 1:93262946 | rs980938 | C | T | 1 | EVI5 | Female only | True | 0.107 | 0.1065 | 0.1004 | 0.09994 | 15947 | 11630 | 187760 | 162101 | 0.08747 | 0.03136 | 0.06746 | 0.05635 | 0.01675 | 0.01975 |
| Monocy | 1:93262946 | rs980940 | C | T | 1 | EVI5 | Female only | True | 0.107 | 0.1065 | 0.1004 | 0.09994 | 15947 | 11630 | 187760 | 162101 | 0.08748 | 0.03136 | 0.06745 | 0.05634 | 0.01675 | 0.01975 |
| Monocy | 1:93262946 | rs2158446 | T | G | 1 | EVI5 | Female only | True | 0.1074 | 0.1069 | 0.1004 | 0.09994 | 15947 | 11630 | 187760 | 162101 | 0.0875 | 0.0314 | 0.06746 | 0.05634 | 0.01676 | 0.01977 |
| Monocy | 1:93262946 | rs4847304 | C | T | 1 | EVI5 | Female only | True | 0.107 | 0.1065 | 0.1002 | 0.09969 | 15947 | 11630 | 187760 | 162101 | 0.08862 | 0.03032 | 0.0676 | 0.05716 | 0.01679 | 0.01979 |
| Monocy | 1:93262946 | rs2025101 | G | A | 1 | EVI5 | Female only | True | 0.1074 | 0.1069 | 0.1004 | 0.09994 | 15947 | 11630 | 187760 | 162101 | 0.08752 | 0.0314 | 0.06745 | 0.05634 | 0.01676 | 0.01977 |
| Monocy | 1:93262946 | rs6660296 | C | T | 1 | EVI5 | Female only | True | 0.1076 | 0.1072 | 0.1009 | 0.1003 | 15947 | 11630 | 187760 | 162101 | 0.08859 | 0.03031 | 0.06769 | 0.05703 | 0.01679 | 0.0198 |
| Monocy | 1:93262946 | rs1182640 | T | C | 1 | EVI5 | Female only | True | 0.09833 | 0.09828 | 0.1002 | 0.09965 | 15947 | 11630 | 187760 | 162101 | 0.08871 | 0.03072 | 0.06776 | 0.05735 | 0.01672 | 0.01967 |
| Monocy | 1:93262946 | rs1337108 | T | C | 1 | EVI5 | Female only | True | 0.1071 | 0.1066 | 0.1002 | 0.09973 | 15947 | 11630 | 187760 | 162101 | 0.08892 | 0.0303 | 0.06762 | 0.05713 | 0.01678 | 0.01979 |

| trait | lead_snp | ID | ALLELE1 | ALLELE0 | CHR | Gene_Symbol | independent_loci_in_sex_P1e2 | in_embl_any_or_gene | A1FREQ_VUMC_female | A1FREQ_VUMC_male | A1FREQ_UKB_female | A1FREQ_UKB_male | N_VUMC_female | N_VUMC_male | N_UKB_female | N_UKB_male | BETA_VUMC_female | BETA_VUMC_male | BETA_UKB_female | BETA_UKB_male | SE_VUMC_female | SE_VUMC_male |
|---|---|---|---|---|---|---|---|---|---|---|---|---|---|---|---|---|---|---|---|---|---|---|
| Monocy | 1:93262946 | rs6604015 | C | T | 1 | EVI5 | Female only | True | 0.1071 | 0.1065 | 0.1002 | 0.09974 | 15947 | 11630 | 187760 | 162101 | 0.08879 | 0.03027 | 0.0676 | 0.05709 | 0.01678 | 0.01979 |
| Monocy | 1:93262946 | rs2391194 | G | A | 1 | EVI5 | Female only | True | 0.1071 | 0.1065 | 0.1002 | 0.09974 | 15947 | 11630 | 187760 | 162101 | 0.08879 | 0.03027 | 0.0676 | 0.05709 | 0.01678 | 0.01979 |
| Monocy | 1:93262946 | rs2298911 | A | G | 1 | EVI5 | Female only | True | 0.1071 | 0.1065 | 0.1002 | 0.09974 | 15947 | 11630 | 187760 | 162101 | 0.08879 | 0.03027 | 0.0676 | 0.05709 | 0.01678 | 0.01979 |
| Monocy | 1:93262946 | rs2298912 | T | C | 1 | EVI5 | Female only | True | 0.1071 | 0.1065 | 0.1002 | 0.09973 | 15947 | 11630 | 187760 | 162101 | 0.08879 | 0.03027 | 0.0676 | 0.05711 | 0.01678 | 0.01979 |
| Monocy | 1:93262946 | rs4847309 | T | C | 1 | EVI5 | Female only | True | 0.09905 | 0.0985 | 0.1003 | 0.09973 | 15947 | 11630 | 187760 | 162101 | 0.0884 | 0.03049 | 0.06762 | 0.0571 | 0.01665 | 0.01962 |
| Monocy | 1:93262946 | rs4847310 | G | A | 1 | EVI5 | Female only | True | 0.1069 | 0.1068 | 0.1002 | 0.09965 | 15947 | 11630 | 187760 | 162101 | 0.08903 | 0.02999 | 0.06782 | 0.05743 | 0.01681 | 0.01979 |
| Monocy | 1:93262946 | rs6604018 | G | A | 1 | EVI5 | Female only | True | 0.1073 | 0.1068 | 0.1002 | 0.09972 | 15947 | 11630 | 187760 | 162101 | 0.08874 | 0.03019 | 0.06765 | 0.05713 | 0.01679 | 0.01979 |
| Monocy | 1:93262946 | rs6604020 | T | C | 1 | EVI5 | Female only | True | 0.1077 | 0.1071 | 0.1003 | 0.09978 | 15947 | 11630 | 187760 | 162101 | 0.0887 | 0.03033 | 0.06756 | 0.05714 | 0.01679 | 0.0198 |
| Monocy | 1:93262946 | rs6604022 | C | T | 1 | EVI5 | Female only | True | 0.1077 | 0.1071 | 0.1003 | 0.09978 | 15947 | 11630 | 187760 | 162101 | 0.0887 | 0.03033 | 0.06756 | 0.05715 | 0.01679 | 0.0198 |
| Monocy | 1:93262946 | rs6696636 | A | C | 1 | EVI5 | Female only | True | 0.107 | 0.1063 | 0.09975 | 0.09911 | 15947 | 11630 | 187760 | 162101 | 0.08957 | 0.03043 | 0.06793 | 0.05809 | 0.01685 | 0.01987 |
| Monocy | 1:93262946 | rs3790445 | C | T | 1 | EVI5 | Female only | True | 0.1077 | 0.1071 | 0.1003 | 0.09979 | 15947 | 11630 | 187760 | 162101 | 0.0887 | 0.03033 | 0.06756 | 0.05704 | 0.01679 | 0.0198 |
| Monocy | 1:93262946 | rs11164805 | C | T | 1 | EVI5 | Female only | True | 0.1081 | 0.1076 | 0.1002 | 0.09971 | 15947 | 11630 | 187760 | 162101 | 0.08861 | 0.02986 | 0.06767 | 0.0571 | 0.0168 | 0.01981 |
| Monocy | 1:93262946 | rs11164806 | T | C | 1 | EVI5 | Female only | True | 0.1081 | 0.1076 | 0.1002 | 0.09971 | 15947 | 11630 | 187760 | 162101 | 0.08861 | 0.02986 | 0.06767 | 0.05707 | 0.0168 | 0.01981 |
| Monocy | 1:93262946 | rs1182632 | A | G | 1 | EVI5 | Female only | True | 0.08911 | 0.09087 | 0.09088 | 0.09045 | 15947 | 11630 | 187760 | 162101 | 0.09329 | 0.01969 | 0.0707 | 0.06098 | 0.01741 | 0.02031 |
| Monocy | 1:93262946 | rs1182633 | G | A | 1 | EVI5 | Female only | True | 0.08903 | 0.09082 | 0.09081 | 0.09038 | 15947 | 11630 | 187760 | 162101 | 0.09368 | 0.01977 | 0.07045 | 0.06086 | 0.01742 | 0.02031 |
| Monocy | 1:93262946 | rs4847335 | A | C | 1 | EVI5 | Female only | True | 0.1081 | 0.1076 | 0.1003 | 0.09972 | 15947 | 11630 | 187760 | 162101 | 0.08861 | 0.02986 | 0.06766 | 0.05708 | 0.0168 | 0.01981 |
| Monocy | 1:93262946 | rs7554183 | A | G | 1 | EVI5 | Female only | True | 0.1081 | 0.1076 | 0.1003 | 0.09973 | 15947 | 11630 | 187760 | 162101 | 0.08861 | 0.02986 | 0.06765 | 0.05702 | 0.0168 | 0.01981 |
| Monocy | 1:93262946 | rs7543327 | C | T | 1 | EVI5 | Female only | True | 0.1084 | 0.1078 | 0.1004 | 0.09987 | 15947 | 11630 | 187760 | 162101 | 0.08816 | 0.03084 | 0.0676 | 0.05646 | 0.01677 | 0.01979 |
| Monocy | 1:93262946 | rs7554868 | A | G | 1 | EVI5 | Female only | True | 0.1081 | 0.1077 | 0.1004 | 0.09981 | 15947 | 11630 | 187760 | 162101 | 0.08861 | 0.02981 | 0.06776 | 0.05742 | 0.0168 | 0.0198 |
| Monocy | 1:93262946 | rs11164808 | A | C | 1 | EVI5 | Female only | True | 0.09905 | 0.09851 | 0.1003 | 0.09972 | 15947 | 11630 | 187760 | 162101 | 0.0884 | 0.03038 | 0.06766 | 0.05707 | 0.01665 | 0.01962 |
| Monocy | 1:93262946 | rs11164809 | A | G | 1 | EVI5 | Female only | True | 0.1077 | 0.1076 | 0.1002 | 0.09965 | 15947 | 11630 | 187760 | 162101 | 0.0889 | 0.02966 | 0.06787 | 0.05733 | 0.01682 | 0.01981 |
| Monocy | 1:93262946 | rs7552085 | C | T | 1 | EVI5 | Female only | True | 0.1081 | 0.1077 | 0.1003 | 0.0998 | 15947 | 11630 | 187760 | 162101 | 0.08849 | 0.03008 | 0.06781 | 0.05699 | 0.01681 | 0.01981 |
| Monocy | 1:93262946 | rs35054144 | C | T | 1 | EVI5 | Female only | True | 0.09817 | 0.09826 | 0.1002 | 0.09967 | 15947 | 11630 | 187760 | 162101 | 0.08943 | 0.03116 | 0.06784 | 0.05723 | 0.01674 | 0.01968 |
| Monocy | 1:93262946 | rs10874740 | A | G | 1 | EVI5 | Female only | True | 0.1078 | 0.1079 | 0.1001 | 0.09961 | 15947 | 11630 | 187760 | 162101 | 0.08945 | 0.03022 | 0.06794 | 0.05743 | 0.01684 | 0.01982 |
| Monocy | 1:93262946 | rs7513815 | T | G | 1 | EVI5 | Female only | True | 0.1082 | 0.1079 | 0.1002 | 0.09964 | 15947 | 11630 | 187760 | 162101 | 0.08919 | 0.03041 | 0.06772 | 0.05722 | 0.01682 | 0.01982 |
| Monocy | 1:93262946 | rs6665121 | T | C | 1 | EVI5 | Female only | True | 0.1084 | 0.1079 | 0.1003 | 0.09975 | 15947 | 11630 | 187760 | 162101 | 0.08896 | 0.0303 | 0.06763 | 0.05707 | 0.0168 | 0.01981 |
| Monocy | 1:93262946 | rs12045019 | A | C | 1 | EVI5 | Female only | True | 0.1084 | 0.1079 | 0.1003 | 0.09973 | 15947 | 11630 | 187760 | 162101 | 0.08893 | 0.03029 | 0.06761 | 0.05708 | 0.01681 | 0.01981 |
| Monocy | 1:93262946 | rs6685705 | T | C | 1 | EVI5 | Female only | True | 0.1089 | 0.1087 | 0.1002 | 0.0997 | 15947 | 11630 | 187760 | 162101 | 0.08997 | 0.03123 | 0.06752 | 0.05712 | 0.01682 | 0.01981 |
| Monocy | 1:93262946 | rs6694690 | G | A | 1 | EVI5 | Female only | True | 0.1092 | 0.1088 | 0.1003 | 0.09979 | 15947 | 11630 | 187760 | 162101 | 0.08954 | 0.03221 | 0.06743 | 0.05677 | 0.01679 | 0.0198 |

| trait | lead_snp | ID | ALLELE1 | ALLELE0 | CHR | Gene_Symbol | independent_loci_in_sex_P1e2 | in_embl_any_or_gene | A1FREQ_VUMC_female | A1FREQ_VUMC_male | A1FREQ_UKB_female | A1FREQ_UKB_male | N_VUMC_female | N_VUMC_male | N_UKB_female | N_UKB_male | BETA_VUMC_female | BETA_VUMC_male | BETA_UKB_female | BETA_UKB_male | SE_VUMC_female | SE_VUMC_male |
|---|---|---|---|---|---|---|---|---|---|---|---|---|---|---|---|---|---|---|---|---|---|---|
| Monocy | 1:93262946 | rs4847217 | T | C | 1 | EVI5 | Female only | True | 0.1085 | 0.1086 | 0.1002 | 0.09963 | 15947 | 11630 | 187760 | 162101 | 0.09021 | 0.03103 | 0.06764 | 0.05734 | 0.01684 | 0.01981 |
| Monocy | 1:93262946 | rs6693371 | G | T | 1 | EVI5 | Female only | True | 0.109 | 0.1085 | 0.1 | 0.09943 | 15947 | 11630 | 187760 | 162101 | 0.09011 | 0.03353 | 0.06731 | 0.05766 | 0.01682 | 0.01983 |
| Monocy | 1:93262946 | rs10782943 | G | A | 1 | EVI5 | Female only | True | 0.109 | 0.1085 | 0.09998 | 0.09942 | 15947 | 11630 | 187760 | 162101 | 0.09011 | 0.03353 | 0.06731 | 0.05767 | 0.01682 | 0.01983 |
| Monocy | 1:93262946 | rs11164812 | A | G | 1 | EVI5 | Female only | True | 0.109 | 0.1085 | 0.1002 | 0.09966 | 15947 | 11630 | 187760 | 162101 | 0.09014 | 0.03407 | 0.06724 | 0.05748 | 0.01682 | 0.01983 |
| Monocy | 1:93262946 | rs11164813 | A | G | 1 | EVI5 | Female only | True | 0.109 | 0.1085 | 0.1001 | 0.0995 | 15947 | 11630 | 187760 | 162101 | 0.09013 | 0.03407 | 0.06706 | 0.05746 | 0.01682 | 0.01983 |
| Monocy | 1:93262946 | rs7551880 | A | G | 1 | EVI5 | Female only | True | 0.1089 | 0.1085 | 0.1 | 0.09947 | 15947 | 11630 | 187760 | 162101 | 0.09003 | 0.03374 | 0.0671 | 0.05748 | 0.01682 | 0.01983 |
| Monocy | 1:93262946 | rs4847370 | C | T | 1 | EVI5 | Female only | True | 0.1089 | 0.1085 | 0.1 | 0.09947 | 15947 | 11630 | 187760 | 162101 | 0.09004 | 0.03374 | 0.06711 | 0.05749 | 0.01682 | 0.01983 |
| Monocy | 1:93262946 | rs7551340 | G | T | 1 | EVI5 | Female only | True | 0.109 | 0.1085 | 0.1001 | 0.09953 | 15947 | 11630 | 187760 | 162101 | 0.08952 | 0.03373 | 0.06721 | 0.05716 | 0.01681 | 0.01983 |
| Monocy | 1:93262946 | rs7543964 | T | C | 1 | EVI5 | Female only | True | 0.1077 | 0.1072 | 0.1001 | 0.09953 | 15947 | 11630 | 187760 | 162101 | 0.08948 | 0.03517 | 0.06721 | 0.05716 | 0.01679 | 0.0198 |
| Monocy | 1:93262946 | rs9439924 | C | T | 1 | EVI5 | Female only | True | 0.107 | 0.107 | 0.09995 | 0.09937 | 15947 | 11630 | 187760 | 162101 | 0.09007 | 0.03491 | 0.06741 | 0.05782 | 0.01684 | 0.01981 |
| Monocy | 1:93262946 | 1:93262946 | G | T | 1 | EVI5 | Female only | True | 0.102 | 0.102 | | | 15947 | 11630 | | | 0.092 | 0.02861 | | | 0.01653 | 0.0194 |
| Monocy | 1:93262946 | rs7513624 | G | T | 1 | EVI5 | Female only | True | 0.1116 | 0.1113 | 0.1091 | 0.1088 | 15947 | 11630 | 187760 | 162101 | 0.08408 | 0.04006 | 0.06344 | 0.05501 | 0.0158 | 0.0186 |
| Monocy | 1:93262946 | rs10735785 | A | G | 1 | EVI5 | Female only | True | 0.1018 | 0.1021 | 0.09309 | 0.09271 | 15947 | 11630 | 187760 | 162101 | 0.09559 | 0.02695 | 0.06928 | 0.05964 | 0.01727 | 0.02031 |
| Monocy | 1:93262946 | rs7515098 | T | C | 1 | EVI5 | Female only | True | 0.107 | 0.1071 | 0.0999 | 0.09932 | 15947 | 11630 | 187760 | 162101 | 0.0904 | 0.03472 | 0.06706 | 0.0578 | 0.01684 | 0.01981 |
| trait | lead_snp | ID | ALLELE1 | ALLELE0 | CHR | Gene_Symbol | independent_loci_in_sex_P1e2 | in_embl_any_or_gene | A1FREQ_VUMC_female | A1FREQ_VUMC_male | A1FREQ_UKB_female | A1FREQ_UKB_male | N_VUMC_female | N_VUMC_male | N_UKB_female | N_UKB_male | BETA_VUMC_female | BETA_VUMC_male | BETA_UKB_female | BETA_UKB_male | SE_VUMC_female | SE_VUMC_male |
| Monocy | 1:93262946 | rs12029026 | T | C | 1 | EVI5 | Female only | True | 0.1077 | 0.1072 | 0.1 | 0.09944 | 15947 | 11630 | 187760 | 162101 | 0.08948 | 0.03518 | 0.06679 | 0.05717 | 0.01679 | 0.0198 |
| Monocy | 1:93262946 | rs1977224 | G | A | 1 | EVI5 | Female only | True | 0.1076 | 0.1072 | 0.09993 | 0.0994 | 15947 | 11630 | 187760 | 162101 | 0.08663 | 0.03342 | 0.06683 | 0.05708 | 0.01682 | 0.01979 |
| Monocy | 1:93262946 | rs4847221 | T | C | 1 | EVI5 | Female only | True | 0.1103 | 0.1103 | 0.1084 | 0.1081 | 15947 | 11630 | 187760 | 162101 | 0.08263 | 0.03375 | 0.06245 | 0.05344 | 0.01591 | 0.01868 |
| Monocy | 1:93262946 | rs1977225 | C | T | 1 | EVI5 | Female only | True | 0.1189 | 0.1188 | 0.1083 | 0.108 | 15947 | 11630 | 187760 | 162101 | 0.08245 | 0.03514 | 0.06277 | 0.05368 | 0.01603 | 0.01885 |
| Monocy | 1:93262946 | rs1977226 | T | C | 1 | EVI5 | Female only | True | 0.119 | 0.1189 | 0.1085 | 0.1082 | 15947 | 11630 | 187760 | 162101 | 0.08221 | 0.03526 | 0.06272 | 0.0537 | 0.01602 | 0.01885 |
| Monocy | 1:93262946 | rs1977227 | A | G | 1 | EVI5 | Female only | True | 0.1175 | 0.1172 | 0.1073 | 0.107 | 15947 | 11630 | 187760 | 162101 | 0.08583 | 0.03588 | 0.06366 | 0.05366 | 0.01614 | 0.01902 |
| Monocy | 1:93262946 | rs1467559 | G | T | 1 | EVI5 | Female only | True | 0.1189 | 0.1188 | 0.1083 | 0.108 | 15947 | 11630 | 187760 | 162101 | 0.08251 | 0.03515 | 0.06276 | 0.05368 | 0.01603 | 0.01885 |
| Monocy | 1:93262946 | rs1977228 | T | G | 1 | EVI5 | Female only | True | 0.119 | 0.1189 | 0.1083 | 0.108 | 15947 | 11630 | 187760 | 162101 | 0.08248 | 0.03524 | 0.0627 | 0.05363 | 0.01603 | 0.01885 |
| Monocy | 1:93262946 | rs1977229 | T | C | 1 | EVI5 | Female only | True | 0.1186 | 0.1184 | 0.1075 | 0.1073 | 15947 | 11630 | 187760 | 162101 | 0.08137 | 0.03444 | 0.0632 | 0.05385 | 0.01606 | 0.01888 |
| Monocy | 1:93262946 | rs2391192 | C | T | 1 | EVI5 | Female only | True | 0.1187 | 0.1185 | 0.1078 | 0.1074 | 15947 | 11630 | 187760 | 162101 | 0.08275 | 0.03392 | 0.06268 | 0.0539 | 0.01605 | 0.01887 |
| Monocy | 1:93262946 | rs2025100 | G | A | 1 | EVI5 | Female only | True | 0.1186 | 0.1184 | 0.1077 | 0.1073 | 15947 | 11630 | 187760 | 162101 | 0.08264 | 0.03349 | 0.06277 | 0.05411 | 0.01605 | 0.01887 |
| Monocy | 1:93262946 | rs6687994 | G | T | 1 | EVI5 | Female only | True | 0.1184 | 0.1182 | 0.1076 | 0.1071 | 15947 | 11630 | 187760 | 162101 | 0.08329 | 0.03216 | 0.06317 | 0.05426 | 0.01606 | 0.0189 |
| Monocy | 1:93262946 | rs9662515 | A | C | 1 | EVI5 | Female only | True | 0.1184 | 0.1182 | 0.1076 | 0.1071 | 15947 | 11630 | 187760 | 162101 | 0.08329 | 0.03216 | 0.06313 | 0.05435 | 0.01606 | 0.0189 |

| trait | lead_snp | ID | ALLELE1 | ALLELE0 | CHR | Gene_Symbol | independent_loci_in_sex_P1e2 | in_embl_any_or_gene | A1FREQ_VUMC_female | A1FREQ_VUMC_male | A1FREQ_UKB_female | A1FREQ_UKB_male | N_VUMC_female | N_VUMC_male | N_UKB_female | N_UKB_male | BETA_VUMC_female | BETA_VUMC_male | BETA_UKB_female | BETA_UKB_male | SE_VUMC_female | SE_VUMC_male |
|---|---|---|---|---|---|---|---|---|---|---|---|---|---|---|---|---|---|---|---|---|---|---|
| Monocy | 1:93262946 | rs7543112 | T | C | 1 | EVI5 | Female only | True | 0.1175 | 0.1174 | 0.1076 | 0.1071 | 15947 | 11630 | 187760 | 162101 | 0.08313 | 0.03248 | 0.06308 | 0.05433 | 0.01606 | 0.01888 |
| Monocy | 1:93262946 | rs11164821 | T | C | 1 | EVI5 | Female only | True | 0.1175 | 0.1174 | 0.1076 | 0.1072 | 15947 | 11630 | 187760 | 162101 | 0.08313 | 0.03248 | 0.06307 | 0.05433 | 0.01606 | 0.01888 |
| Monocy | 1:93262946 | rs7532731 | A | G | 1 | EVI5 | Female only | True | 0.1081 | 0.1076 | 0.1004 | 0.09974 | 15947 | 11630 | 187760 | 162101 | 0.08953 | 0.03072 | 0.06509 | 0.05558 | 0.01671 | 0.01965 |
| Monocy | 1:93262946 | rs7543077 | G | A | 1 | EVI5 | Female only | True | 0.1175 | 0.1174 | 0.1076 | 0.1072 | 15947 | 11630 | 187760 | 162101 | 0.08308 | 0.03248 | 0.06306 | 0.05432 | 0.01606 | 0.01888 |
| Monocy | 1:93262946 | rs10782944 | C | A | 1 | EVI5 | Female only | True | 0.1174 | 0.1174 | 0.1076 | 0.1071 | 15947 | 11630 | 187760 | 162101 | 0.08298 | 0.03124 | 0.06308 | 0.05449 | 0.01607 | 0.01889 |
| Monocy | 1:93262946 | rs7527356 | C | T | 1 | EVI5 | Female only | True | 0.1173 | 0.1173 | 0.1076 | 0.1071 | 15947 | 11630 | 187760 | 162101 | 0.0829 | 0.03133 | 0.06308 | 0.05441 | 0.01607 | 0.01889 |
| Monocy | 1:93262946 | rs1929138 | A | G | 1 | EVI5 | Female only | True | 0.1173 | 0.1173 | 0.1076 | 0.1071 | 15947 | 11630 | 187760 | 162101 | 0.0829 | 0.03132 | 0.06308 | 0.05441 | 0.01607 | 0.01889 |
| Monocy | 1:93262946 | rs7541829 | T | C | 1 | EVI5 | Female only | True | 0.1173 | 0.1173 | 0.1076 | 0.1071 | 15947 | 11630 | 187760 | 162101 | 0.0829 | 0.03132 | 0.06307 | 0.05449 | 0.01607 | 0.01889 |
| Monocy | 1:93262946 | rs6604025 | C | A | 1 | EVI5 | Female only | True | 0.1173 | 0.1173 | 0.1076 | 0.1071 | 15947 | 11630 | 187760 | 162101 | 0.0829 | 0.03132 | 0.06308 | 0.0544 | 0.01607 | 0.01889 |
| Monocy | 1:93262946 | rs10874742 | T | C | 1 | EVI5 | Female only | True | 0.1098 | 0.1097 | 0.1077 | 0.1072 | 15947 | 11630 | 187760 | 162101 | 0.08222 | 0.03053 | 0.06272 | 0.05429 | 0.01593 | 0.01873 |
| Monocy | 1:93262946 | rs6659942 | G | T | 1 | EVI5 | Female only | True | 0.1164 | 0.1165 | 0.1068 | 0.1063 | 15947 | 11630 | 187760 | 162101 | 0.08427 | 0.03337 | 0.06337 | 0.0538 | 0.01608 | 0.01889 |
| Monocy | 1:93262946 | rs4847222 | A | G | 1 | EVI5 | Female only | True | 0.1163 | 0.1164 | 0.1068 | 0.1063 | 15947 | 11630 | 187760 | 162101 | 0.08421 | 0.03302 | 0.06335 | 0.05381 | 0.01609 | 0.0189 |
| Monocy | 1:93262946 | rs2244588 | G | A | 1 | EVI5 | Female only | True | 0.0956 | 0.09803 | 0.08905 | 0.08846 | 15947 | 11630 | 187760 | 162101 | 0.09564 | 0.01851 | 0.0689 | 0.05935 | 0.01762 | 0.02045 |
| Monocy | 1:93262946 | rs2244496 | C | A | 1 | EVI5 | Female only | True | 0.098 | 0.09846 | 0.09846 | 0.09771 | 15947 | 11630 | 187760 | 162101 | 0.09049 | 0.02884 | 0.06607 | 0.05577 | 0.01675 | 0.01962 |
| Monocy | 1:93262946 | rs2774940 | T | G | 1 | EVI5 | Female only | True | 0.1076 | 0.1075 | 0.09985 | 0.09918 | 15947 | 11630 | 187760 | 162101 | 0.08817 | 0.02929 | 0.06496 | 0.05492 | 0.0167 | 0.01958 |
| Monocy | 1:93262946 | rs2391179 | T | C | 1 | EVI5 | Female only | True | 0.1167 | 0.1171 | 0.1069 | 0.1064 | 15947 | 11630 | 187760 | 162101 | 0.08494 | 0.03408 | 0.06356 | 0.05385 | 0.01606 | 0.01883 |
| Monocy | 1:93262946 | rs2989014 | G | A | 1 | EVI5 | Female only | True | 0.1165 | 0.1171 | 0.1069 | 0.1064 | 15947 | 11630 | 187760 | 162101 | 0.08515 | 0.03421 | 0.06353 | 0.05388 | 0.01607 | 0.01883 |
| Monocy | 1:93262946 | rs2774941 | A | C | 1 | EVI5 | Female only | True | 0.105 | 0.1056 | 0.09839 | 0.09763 | 15947 | 11630 | 187760 | 162101 | 0.09057 | 0.03022 | 0.06609 | 0.05589 | 0.01687 | 0.01976 |
| Monocy | 1:93262946 | rs2774943 | T | G | 1 | EVI5 | Female only | True | 0.1158 | 0.1161 | 0.1067 | 0.1061 | 15947 | 11630 | 187760 | 162101 | 0.08612 | 0.03415 | 0.06394 | 0.05337 | 0.01612 | 0.01891 |
| Monocy | 1:93262946 | rs1411706 | A | G | 1 | EVI5 | Female only | True | 0.1069 | 0.1067 | 0.09959 | 0.09886 | 15947 | 11630 | 187760 | 162101 | 0.0917 | 0.03057 | 0.06525 | 0.05446 | 0.01675 | 0.01966 |
| Monocy | 1:93262946 | rs2811602 | C | T | 1 | EVI5 | Female only | True | 0.1067 | 0.1066 | 0.09939 | 0.09876 | 15947 | 11630 | 187760 | 162101 | 0.09107 | 0.02782 | 0.06314 | 0.05283 | 0.01676 | 0.01963 |
| Monocy | 1:93262946 | rs2811599 | A | G | 1 | EVI5 | Female only | True | 0.1044 | 0.1049 | 0.09788 | 0.0972 | 15947 | 11630 | 187760 | 162101 | 0.08973 | 0.02687 | 0.06436 | 0.0546 | 0.0169 | 0.01978 |
| Monocy | 1:93262946 | rs2774947 | C | T | 1 | EVI5 | Female only | True | 0.09456 | 0.09714 | 0.08838 | 0.08792 | 15947 | 11630 | 187760 | 162101 | 0.09404 | 0.01566 | 0.06715 | 0.05786 | 0.01768 | 0.02047 |
| Monocy | 1:93262946 | rs145469268 | A | G | 1 | EVI5 | Female only | True | 0.09779 | 0.09841 | 0.09799 | 0.09741 | 15947 | 11630 | 187760 | 162101 | 0.09018 | 0.02561 | 0.0657 | 0.05571 | 0.01694 | 0.01983 |
| Monocy | 1:93262946 | rs78663632 | A | G | 1 | EVI5 | Female only | True | 0.06647 | 0.06786 | 0.06644 | 0.06582 | 15947 | 11630 | 187760 | 162101 | 0.1039 | 0.03543 | 0.08542 | 0.07674 | 0.02032 | 0.02378 |
| Monocy | rs1449264 | rs1449264 | C | T | 2 | ITGA4 | Female only | True | 0.3382 | 0.333 | 0.3442 | 0.3432 | 15947 | 11630 | 187760 | 162101 | 0.07124 | 0.01442 | 0.06336 | 0.06256 | 0.01053 | 0.01252 |
| Monocy | rs1449264 | rs17224872 | T | C | 2 | ITGA4 | Female only | True | 0.4205 | 0.4188 | 0.4258 | 0.426 | 15947 | 11630 | 187760 | 162101 | 0.04427 | 0.008133 | 0.04757 | 0.0451 | 0.01006 | 0.01185 |
| LYMPRE | rs3011200 | rs1925381 | C | T | 1 | RP11-334L9.1 | Female only | False | 0.2902 | 0.2916 | 0.297 | 0.2973 | 25619 | 19940 | 187760 | 162101 | 0.04052 | 0.002013 | 2.12e-05 | 0.007228 | 0.009001 | 0.009647 |
| LYMPRE | rs3011200 | rs3011200 | G | A | 1 | RP11-334L9.1 | Female only | False | 0.3805 | 0.3833 | 0.3847 | 0.3844 | 25619 | 19940 | 187760 | 162101 | 0.04893 | 0.01178 | 1.00e-05 | 0.006143 | 0.008383 | 0.008981 |

| trait | lead_snp | ID | ALLELE1 | ALLELE0 | CHR | Gene_Symbol | independent_loci_in_sex_P1e2 | in_embl_any_or_gene | A1FREQ_VUMC_female | A1FREQ_VUMC_male | A1FREQ_UKB_female | A1FREQ_UKB_male | N_VUMC_female | N_VUMC_male | N_UKB_female | N_UKB_male | BETA_VUMC_female | BETA_VUMC_male | BETA_UKB_female | BETA_UKB_male | SE_VUMC_female | SE_VUMC_male |
|---|---|---|---|---|---|---|---|---|---|---|---|---|---|---|---|---|---|---|---|---|---|---|
| LYMPRE | rs3011200 | rs4970606 | G | A | 1 | RP11-334L9.1 | Female only | False | 0.2778 | 0.2786 | 0.2827 | 0.2825 | 25619 | 19940 | 187760 | 162101 | 0.04417 | 0.001858 | 8.78e-04 | 0.007391 | 0.009119 | 0.0098 |
| LYMPRE | rs3011200 | rs12137575 | C | A | 1 | RP11-334L9.1 | Female only | False | 0.2791 | 0.2803 | 0.2831 | 0.2829 | 25619 | 19940 | 187760 | 162101 | 0.04268 | 0.002086 | 0.001036 | 0.007347 | 0.009108 | 0.009782 |
| LYMPRE | rs3011200 | rs4970609 | G | A | 1 | RP11-334L9.1 | Female only | False | 0.3161 | 0.3196 | 0.3213 | 0.3205 | 25619 | 19940 | 187760 | 162101 | 0.03816 | 0.01338 | -6.88e-04 | 0.003929 | 0.00874 | 0.009398 |
| LYMPRE | rs3011200 | rs4970610 | T | C | 1 | RP11-334L9.1 | Female only | False | 0.3151 | 0.3185 | 0.3213 | 0.3204 | 25619 | 19940 | 187760 | 162101 | 0.03835 | 0.01334 | -6.55e-04 | 0.003942 | 0.008764 | 0.009425 |
| LYMPRE | rs3011200 | rs627552 | A | C | 1 | RP11-334L9.1 | Female only | False | 0.3162 | 0.3196 | 0.3215 | 0.3207 | 25619 | 19940 | 187760 | 162101 | 0.03849 | 0.01302 | -5.92e-04 | 0.003766 | 0.008755 | 0.009421 |
| LYMPRE | rs3011200 | rs1759398 | C | T | 1 | RP11-334L9.1 | Female only | False | 0.3162 | 0.3195 | 0.3216 | 0.3207 | 25619 | 19940 | 187760 | 162101 | 0.03851 | 0.01296 | -6.01e-04 | 0.003782 | 0.008756 | 0.009422 |
| Baso | rs56373544 | rs9866019 | C | T | 3 | RAB7A | Female only | False | 0.4258 | 0.4215 | 0.4113 | 0.4127 | 15240 | 11083 | 187760 | 162101 | -0.057 | -0.01304 | -0.0149 | -0.01118 | 0.01052 | 0.01223 |
| Baso | rs56373544 | rs7372263 | C | A | 3 | RAB7A | Female only | False | 0.3773 | 0.3713 | 0.3634 | 0.3646 | 15240 | 11083 | 187760 | 162101 | -0.06686 | -0.02128 | -0.01762 | -0.01378 | 0.01068 | 0.01236 |
| Baso | rs56373544 | rs12497923 | G | A | 3 | RAB7A | Female only | False | 0.4254 | 0.4209 | 0.4112 | 0.4126 | 15240 | 11083 | 187760 | 162101 | -0.05775 | -0.01295 | -0.01499 | -0.01121 | 0.01053 | 0.01224 |
| Baso | rs56373544 | rs56373544 | G | A | 3 | RAB7A | Female only | False | 0.3802 | 0.3742 | 0.3635 | 0.3646 | 15240 | 11083 | 187760 | 162101 | -0.06716 | -0.0224 | -0.01765 | -0.01375 | 0.01072 | 0.0124 |
| Baso | rs56373544 | rs6791451 | C | T | 3 | RAB7A | Female only | False | 0.4333 | 0.4283 | 0.4226 | 0.4249 | 15240 | 11083 | 187760 | 162101 | -0.05298 | -0.02801 | -0.0131 | -0.01191 | 0.01051 | 0.0121 |
| Baso | rs56373544 | rs1683791 | T | C | 3 | RAB7A | Female only | False | 0.4363 | 0.4312 | 0.4252 | 0.4276 | 15240 | 11083 | 187760 | 162101 | -0.05308 | -0.02778 | -0.0126 | -0.01187 | 0.01052 | 0.01211 |
| Baso | rs56373544 | rs1683804 | T | C | 3 | RAB7A | Female only | False | 0.4328 | 0.4277 | 0.4226 | 0.4249 | 15240 | 11083 | 187760 | 162101 | -0.05311 | -0.02766 | -0.01306 | -0.01175 | 0.01051 | 0.0121 |
| Baso | rs56373544 | rs1680789 | T | G | 3 | RAB7A | Female only | False | 0.4347 | 0.4291 | 0.4242 | 0.4266 | 15240 | 11083 | 187760 | 162101 | -0.05181 | -0.02776 | -0.01315 | -0.01149 | 0.0105 | 0.01209 |
| Baso | rs56373544 | rs6767396 | C | T | 3 | RAB7A | Female only | False | 0.4373 | 0.4316 | 0.4262 | 0.4283 | 15240 | 11083 | 187760 | 162101 | -0.05048 | -0.02693 | -0.01361 | -0.01181 | 0.01048 | 0.01206 |
| Baso | rs56373544 | rs7642004 | C | A | 3 | RAB7A | Female only | False | 0.438 | 0.4323 | 0.4263 | 0.4284 | 15240 | 11083 | 187760 | 162101 | -0.04997 | -0.02692 | -0.01352 | -0.01158 | 0.0105 | 0.01207 |
| Baso | rs56373544 | rs11917573 | T | G | 3 | RAB7A | Female only | False | 0.4374 | 0.4317 | 0.4263 | 0.4284 | 15240 | 11083 | 187760 | 162101 | -0.05024 | -0.02646 | -0.01352 | -0.0115 | 0.0105 | 0.01208 |
| Baso | rs56373544 | rs2128490 | G | A | 3 | RAB7A | Female only | False | 0.4374 | 0.4317 | 0.4264 | 0.4284 | 15240 | 11083 | 187760 | 162101 | -0.05027 | -0.02646 | -0.01351 | -0.01146 | 0.0105 | 0.01208 |
| Baso | rs56373544 | rs1683813 | G | T | 3 | RAB7A | Female only | False | 0.4589 | 0.4535 | 0.4522 | 0.4543 | 15240 | 11083 | 187760 | 162101 | -0.04604 | -0.02805 | -0.01303 | -0.01046 | 0.01042 | 0.01191 |
| Baso | rs10875486 | rs10875486 | T | G | 20 | C20orf112 | Female only | True | 0.2413 | 0.2384 | 0.2388 | 0.2373 | 15240 | 11083 | 187760 | 162101 | -0.0675 | -0.03497 | -0.01774 | -0.009631 | 0.01218 | 0.01408 |
| Baso | rs10875486 | rs6057602 | G | T | 20 | C20orf112 | Female only | True | 0.2945 | 0.2954 | 0.3023 | 0.3009 | 15240 | 11083 | 187760 | 162101 | -0.05859 | -0.02454 | -0.01428 | -0.006798 | 0.01142 | 0.01311 |
| Baso | rs10875486 | rs6058753 | G | A | 20 | C20orf112 | Female only | True | 0.2896 | 0.2901 | 0.2962 | 0.2946 | 15240 | 11083 | 187760 | 162101 | -0.05393 | -0.02431 | -0.01432 | -0.006638 | 0.01148 | 0.0132 |
| Baso | rs6141766 | rs1407256 | G | A | 20 | C20orf203 | Female only | True | 0.1487 | 0.1493 | 0.1493 | 0.1496 | 15240 | 11083 | 187760 | 162101 | -0.09418 | -0.03389 | -0.01928 | -0.01036 | 0.01456 | 0.01672 |
| Baso | rs6141766 | rs79384779 | T | C | 20 | C20orf203 | Female only | True | 0.1464 | 0.1475 | 0.1466 | 0.1467 | 15240 | 11083 | 187760 | 162101 | -0.095 | -0.03506 | -0.02087 | -0.01102 | 0.01466 | 0.0168 |
| Baso | rs6141766 | rs12481717 | A | G | 20 | C20orf203 | Female only | True | 0.1467 | 0.1474 | 0.1469 | 0.1471 | 15240 | 11083 | 187760 | 162101 | -0.09376 | -0.036 | -0.021 | -0.01088 | 0.01465 | 0.01681 |
| Baso | rs6141766 | rs6141766 | G | A | 20 | C20orf203 | Female only | True | 0.1527 | 0.1536 | 0.1505 | 0.1506 | 15240 | 11083 | 187760 | 162101 | -0.09803 | -0.02567 | -0.01864 | -0.009944 | 0.01443 | 0.01659 |
| Baso | rs6141766 | rs12624715 | G | A | 20 | C20orf203 | Female only | True | 0.152 | 0.1535 | 0.1504 | 0.1504 | 15240 | 11083 | 187760 | 162101 | -0.09689 | -0.02589 | -0.01884 | -0.01005 | 0.01444 | 0.01656 |

| trait | lead_snp | ID | ALLELE1 | ALLELE0 | CHR | Gene_Symbol | independent_loci_in_sex_P1e2 | in_embl_any_or_gene | A1FREQ_VUMC_female | A1FREQ_VUMC_male | A1FREQ_UKB_female | A1FREQ_UKB_male | N_VUMC_female | N_VUMC_male | N_UKB_female | N_UKB_male | BETA_VUMC_female | BETA_VUMC_male | BETA_UKB_female | BETA_UKB_male | SE_VUMC_female | SE_VUMC_male |
|---|---|---|---|---|---|---|---|---|---|---|---|---|---|---|---|---|---|---|---|---|---|---|
| Baso | rs6141766 | rs6141328 | A | G | 20 | C20orf203 | Female only | True | 0.1531 | 0.1541 | 0.1508 | 0.1508 | 15240 | 11083 | 187760 | 162101 | -0.09735 | -0.02476 | -0.01901 | -0.009963 | 0.0144 | 0.01654 |
| Baso | rs6141766 | rs73107865 | T | C | 20 | C20orf203 | Female only | True | 0.1538 | 0.1565 | 0.1525 | 0.1525 | 15240 | 11083 | 187760 | 162101 | -0.09701 | -0.02485 | -0.02093 | -0.007796 | 0.01436 | 0.0164 |
| Baso | rs6141766 | rs6141768 | A | G | 20 | C20orf203 | Female only | True | 0.1555 | 0.1579 | 0.153 | 0.153 | 15240 | 11083 | 187760 | 162101 | -0.09674 | -0.0232 | -0.02086 | -0.007601 | 0.01428 | 0.01631 |
| Baso | rs6141766 | rs6087964 | A | G | 20 | C20orf203 | Female only | True | 0.232 | 0.2344 | 0.2259 | 0.2257 | 15240 | 11083 | 187760 | 162101 | -0.08205 | -0.02568 | -0.02101 | -0.008429 | 0.01241 | 0.0141 |
| Baso | rs6141766 | rs6058799 | A | G | 20 | C20orf203 | Female only | True | 0.2322 | 0.2347 | 0.2261 | 0.226 | 15240 | 11083 | 187760 | 162101 | -0.08097 | -0.02521 | -0.02112 | -0.008434 | 0.01239 | 0.01407 |
| Baso | rs6141766 | rs6579029 | T | C | 20 | C20orf203 | Female only | True | 0.232 | 0.2345 | 0.2261 | 0.2259 | 15240 | 11083 | 187760 | 162101 | -0.08131 | -0.02572 | -0.02112 | -0.008413 | 0.0124 | 0.01408 |
| Baso | rs6141766 | rs6057617 | A | G | 20 | C20orf203 | Female only | True | 0.2327 | 0.2352 | 0.2261 | 0.226 | 15240 | 11083 | 187760 | 162101 | -0.08093 | -0.02415 | -0.02118 | -0.008452 | 0.01238 | 0.01405 |
| Baso | rs6141766 | rs12480157 | A | G | 20 | C20orf203 | Female only | True | 0.2295 | 0.2321 | 0.2262 | 0.2262 | 15240 | 11083 | 187760 | 162101 | -0.08061 | -0.0243 | -0.02103 | -0.008188 | 0.01233 | 0.014 |
| Baso | rs6141766 | rs6119932 | G | T | 20 | C20orf203 | Female only | True | 0.2336 | 0.2358 | 0.2264 | 0.2264 | 15240 | 11083 | 187760 | 162101 | -0.08159 | -0.02444 | -0.02139 | -0.00838 | 0.01237 | 0.01405 |
| Baso | rs6141766 | rs6057618 | A | G | 20 | C20orf203 | Female only | True | 0.2316 | 0.2334 | 0.2251 | 0.2252 | 15240 | 11083 | 187760 | 162101 | -0.08094 | -0.02544 | -0.02148 | -0.008578 | 0.0124 | 0.01408 |
| Baso | rs6141766 | rs6057619 | T | C | 20 | C20orf203 | Female only | True | 0.2279 | 0.2312 | 0.2245 | 0.2245 | 15240 | 11083 | 187760 | 162101 | -0.08219 | -0.02389 | -0.02152 | -0.008995 | 0.01236 | 0.01402 |
| Baso | rs6141766 | rs6058807 | T | C | 20 | C20orf203 | Female only | True | 0.232 | 0.2344 | 0.2253 | 0.2252 | 15240 | 11083 | 187760 | 162101 | -0.08331 | -0.02295 | -0.02106 | -0.009222 | 0.0124 | 0.01407 |
| Baso | rs6141766 | rs6057620 | A | G | 20 | C20orf203 | Female only | True | 0.2321 | 0.2344 | 0.2253 | 0.2253 | 15240 | 11083 | 187760 | 162101 | -0.08316 | -0.02378 | -0.02116 | -0.009144 | 0.01239 | 0.01408 |
| Baso | rs6141766 | rs6057621 | A | G | 20 | C20orf203 | Female only | True | 0.2312 | 0.2328 | 0.2246 | 0.2246 | 15240 | 11083 | 187760 | 162101 | -0.08263 | -0.02415 | -0.02153 | -0.009192 | 0.01241 | 0.01409 |
| Baso | rs6141766 | rs6058809 | T | C | 20 | C20orf203 | Female only | True | 0.2303 | 0.2331 | 0.224 | 0.2241 | 15240 | 11083 | 187760 | 162101 | -0.08272 | -0.02326 | -0.02125 | -0.008623 | 0.01244 | 0.01411 |
| Baso | rs6141766 | rs6119935 | C | A | 20 | C20orf203 | Female only | True | 0.231 | 0.2345 | 0.2243 | 0.2243 | 15240 | 11083 | 187760 | 162101 | -0.08278 | -0.02177 | -0.02084 | -0.008428 | 0.01242 | 0.01408 |
| Baso | rs6141766 | rs6058812 | A | G | 20 | C20orf203 | Female only | True | 0.2308 | 0.2339 | 0.2242 | 0.2242 | 15240 | 11083 | 187760 | 162101 | -0.0826 | -0.02358 | -0.02096 | -0.008525 | 0.01245 | 0.01412 |
| Baso | rs6141766 | rs6058813 | C | T | 20 | C20orf203 | Female only | True | 0.2312 | 0.2344 | 0.2243 | 0.2243 | 15240 | 11083 | 187760 | 162101 | -0.08234 | -0.02337 | -0.02084 | -0.008489 | 0.01244 | 0.01411 |
| Baso | rs6141766 | rs11905501 | C | A | 20 | C20orf203 | Female only | True | 0.2312 | 0.2344 | 0.2244 | 0.2243 | 15240 | 11083 | 187760 | 162101 | -0.08234 | -0.02338 | -0.02078 | -0.008497 | 0.01244 | 0.01412 |
| Baso | rs6141766 | rs28456308 | C | T | 20 | C20orf203 | Female only | True | 0.2361 | 0.2389 | 0.2295 | 0.2298 | 15240 | 11083 | 187760 | 162101 | -0.07802 | -0.01989 | -0.01994 | -0.01019 | 0.01235 | 0.01399 |
| Baso | rs6141766 | rs28402080 | A | G | 20 | C20orf203 | Female only | True | 0.2364 | 0.2391 | 0.2296 | 0.2298 | 15240 | 11083 | 187760 | 162101 | -0.07758 | -0.02041 | -0.02002 | -0.01022 | 0.01235 | 0.01399 |
| Baso | rs6141766 | rs28583637 | G | A | 20 | C20orf203 | Female only | True | 0.2364 | 0.2391 | 0.2296 | 0.2299 | 15240 | 11083 | 187760 | 162101 | -0.07738 | -0.02039 | -0.02015 | -0.01015 | 0.01235 | 0.01399 |
| Baso | rs28510728 | rs28698192 | T | C | 20 | COMMD7 | Female only | True | 0.2369 | 0.2398 | 0.2296 | 0.2299 | 15240 | 11083 | 187760 | 162101 | -0.07653 | -0.0223 | -0.02012 | -0.01024 | 0.01233 | 0.01398 |
| Baso | rs28510728 | rs28605309 | A | G | 20 | COMMD7 | Female only | True | 0.2369 | 0.2398 | 0.2296 | 0.23 | 15240 | 11083 | 187760 | 162101 | -0.07653 | -0.02231 | -0.02011 | -0.01022 | 0.01233 | 0.01398 |
| Baso | rs28510728 | rs28565102 | T | G | 20 | COMMD7 | Female only | True | 0.2369 | 0.2398 | 0.2296 | 0.2299 | 15240 | 11083 | 187760 | 162101 | -0.07653 | -0.02231 | -0.02009 | -0.01023 | 0.01233 | 0.01398 |
| Baso | rs28510728 | rs28510728 | G | A | 20 | COMMD7 | Female only | True | 0.2373 | 0.2402 | 0.2297 | 0.23 | 15240 | 11083 | 187760 | 162101 | -0.07665 | -0.02267 | -0.02027 | -0.01045 | 0.01234 | 0.01399 |
| Baso | rs28510728 | rs28707468 | T | C | 20 | COMMD7 | Female only | True | 0.2375 | 0.2404 | 0.2297 | 0.2301 | 15240 | 11083 | 187760 | 162101 | -0.07636 | -0.02215 | -0.02024 | -0.01046 | 0.01234 | 0.01398 |
| Baso | rs28510728 | rs28369623 | T | C | 20 | COMMD7 | Female only | True | 0.2375 | 0.2404 | 0.2297 | 0.2301 | 15240 | 11083 | 187760 | 162101 | -0.07636 | -0.02215 | -0.02027 | -0.01045 | 0.01234 | 0.01398 |
| Baso | rs28510728 | rs9421 | C | T | 20 | COMMD7 | Female only | True | 0.2373 | 0.24 | 0.229 | 0.2293 | 15240 | 11083 | 187760 | 162101 | -0.07603 | -0.02094 | -0.02045 | -0.01024 | 0.01234 | 0.01399 |

| trait | lead_snp | ID | ALLELE1 | ALLELE0 | CHR | Gene_Symbol | independent_loci_in_sex_P1e2 | in_embl_any_or_gene | A1FREQ_VUMC_female | A1FREQ_VUMC_male | A1FREQ_UKB_female | A1FREQ_UKB_male | N_VUMC_female | N_VUMC_male | N_UKB_female | N_UKB_male | BETA_VUMC_female | BETA_VUMC_male | BETA_UKB_female | BETA_UKB_male | SE_VUMC_female | SE_VUMC_male |
|---|---|---|---|---|---|---|---|---|---|---|---|---|---|---|---|---|---|---|---|---|---|---|
| Baso | rs28510728 | rs28548540 | A | G | 20 | COMMD7 | Female only | True | 0.2375 | 0.2402 | 0.2289 | 0.2292 | 15240 | 11083 | 187760 | 162101 | -0.07514 | -0.01975 | -0.02054 | -0.01015 | 0.01234 | 0.01399 |
| Baso | rs28510728 | rs28730896 | A | G | 20 | COMMD7 | Female only | True | 0.2371 | 0.2397 | 0.2289 | 0.2292 | 15240 | 11083 | 187760 | 162101 | -0.07607 | -0.02098 | -0.02056 | -0.01016 | 0.01235 | 0.014 |
| Baso | rs28510728 | rs28612490 | G | A | 20 | COMMD7 | Female only | True | 0.2369 | 0.2395 | 0.2288 | 0.2291 | 15240 | 11083 | 187760 | 162101 | -0.07578 | -0.02128 | -0.02051 | -0.01017 | 0.01235 | 0.014 |
| Baso | rs28510728 | rs28695612 | G | T | 20 | COMMD7 | Female only | True | 0.237 | 0.2396 | 0.2288 | 0.2291 | 15240 | 11083 | 187760 | 162101 | -0.07579 | -0.02124 | -0.02051 | -0.01017 | 0.01235 | 0.014 |
| Baso | rs28510728 | rs28421806 | A | G | 20 | COMMD7 | Female only | True | 0.237 | 0.2396 | 0.2288 | 0.2291 | 15240 | 11083 | 187760 | 162101 | -0.07579 | -0.02124 | -0.02051 | -0.01017 | 0.01235 | 0.014 |
| Baso | rs28510728 | rs28456361 | T | C | 20 | COMMD7 | Female only | True | 0.237 | 0.2396 | 0.2288 | 0.2291 | 15240 | 11083 | 187760 | 162101 | -0.07579 | -0.02124 | -0.02049 | -0.01018 | 0.01235 | 0.014 |
| Baso | rs28510728 | rs28439340 | G | A | 20 | COMMD7 | Female only | True | 0.237 | 0.2396 | 0.2288 | 0.2291 | 15240 | 11083 | 187760 | 162101 | -0.07579 | -0.02124 | -0.02049 | -0.01018 | 0.01235 | 0.014 |
| Baso | rs28510728 | rs28450123 | C | T | 20 | COMMD7 | Female only | True | 0.237 | 0.2396 | 0.2288 | 0.2291 | 15240 | 11083 | 187760 | 162101 | -0.07579 | -0.02124 | -0.02049 | -0.01018 | 0.01235 | 0.014 |
| Baso | rs28510728 | rs28437232 | C | T | 20 | COMMD7 | Female only | True | 0.237 | 0.2396 | 0.2288 | 0.2291 | 15240 | 11083 | 187760 | 162101 | -0.07578 | -0.02125 | -0.02048 | -0.01019 | 0.01235 | 0.014 |
| Baso | rs28510728 | rs28523897 | C | T | 20 | COMMD7 | Female only | True | 0.237 | 0.2396 | 0.2288 | 0.2291 | 15240 | 11083 | 187760 | 162101 | -0.07579 | -0.02124 | -0.02049 | -0.01019 | 0.01235 | 0.014 |
| Baso | rs28510728 | rs28542091 | T | C | 20 | COMMD7 | Female only | True | 0.2371 | 0.2396 | 0.2288 | 0.2291 | 15240 | 11083 | 187760 | 162101 | -0.07582 | -0.02131 | -0.02052 | -0.01019 | 0.01235 | 0.014 |
| Baso | rs28510728 | rs28548407 | T | C | 20 | COMMD7 | Female only | True | 0.2375 | 0.2402 | 0.2288 | 0.2291 | 15240 | 11083 | 187760 | 162101 | -0.07464 | -0.02115 | -0.02049 | -0.01018 | 0.01233 | 0.01398 |
| Baso | rs28510728 | rs28481782 | T | C | 20 | COMMD7 | Female only | True | 0.2375 | 0.2402 | 0.2288 | 0.2291 | 15240 | 11083 | 187760 | 162101 | -0.07464 | -0.02115 | -0.02048 | -0.01018 | 0.01233 | 0.01398 |
| Baso | rs28510728 | rs28536730 | A | C | 20 | COMMD7 | Female only | True | 0.2375 | 0.2402 | 0.2288 | 0.2291 | 15240 | 11083 | 187760 | 162101 | -0.07464 | -0.02115 | -0.02049 | -0.01016 | 0.01233 | 0.01398 |
| Baso | rs28510728 | rs6058819 | T | C | 20 | COMMD7 | Female only | True | 0.2376 | 0.2403 | 0.2288 | 0.2291 | 15240 | 11083 | 187760 | 162101 | -0.07472 | -0.02131 | -0.02043 | -0.01016 | 0.01233 | 0.01397 |
| Baso | rs28510728 | rs6058820 | T | C | 20 | COMMD7 | Female only | True | 0.2376 | 0.2402 | 0.2288 | 0.2291 | 15240 | 11083 | 187760 | 162101 | -0.07473 | -0.021 | -0.02046 | -0.01015 | 0.01233 | 0.01398 |
| Baso | rs28510728 | rs6058821 | C | T | 20 | COMMD7 | Female only | True | 0.2375 | 0.24 | 0.2285 | 0.2288 | 15240 | 11083 | 187760 | 162101 | -0.07472 | -0.02071 | -0.02034 | -0.01024 | 0.01233 | 0.01398 |
| Baso | rs28510728 | rs6057623 | G | T | 20 | COMMD7 | Female only | True | 0.2375 | 0.2399 | 0.2285 | 0.2288 | 15240 | 11083 | 187760 | 162101 | -0.07492 | -0.02084 | -0.02031 | -0.01022 | 0.01233 | 0.01398 |
| Baso | rs28510728 | rs6057625 | G | A | 20 | COMMD7 | Female only | True | 0.2375 | 0.24 | 0.2285 | 0.2288 | 15240 | 11083 | 187760 | 162101 | -0.07481 | -0.02062 | -0.02031 | -0.01023 | 0.01233 | 0.01398 |
| Baso | rs28510728 | rs6057626 | T | C | 20 | COMMD7 | Female only | True | 0.2375 | 0.24 | 0.2285 | 0.2288 | 15240 | 11083 | 187760 | 162101 | -0.07483 | -0.02061 | -0.02031 | -0.01019 | 0.01233 | 0.01398 |
| Baso | rs28510728 | rs6058825 | G | A | 20 | COMMD7 | Female only | True | 0.238 | 0.2404 | 0.2288 | 0.2292 | 15240 | 11083 | 187760 | 162101 | -0.07427 | -0.01993 | -0.02025 | -0.01017 | 0.01234 | 0.01398 |
| Baso | rs28510728 | rs6058828 | A | G | 20 | COMMD7 | Female only | True | 0.2375 | 0.2399 | 0.2285 | 0.2288 | 15240 | 11083 | 187760 | 162101 | -0.07491 | -0.02084 | -0.02031 | -0.01019 | 0.01233 | 0.01398 |
| trait | lead_snp | ID | ALLELE1 | ALLELE0 | CHR | Gene_Symbol | independent_loci_in_sex_P1e2 | in_embl_any_or_gene | A1FREQ_VUMC_female | A1FREQ_VUMC_male | A1FREQ_UKB_female | A1FREQ_UKB_male | N_VUMC_female | N_VUMC_male | N_UKB_female | N_UKB_male | BETA_VUMC_female | BETA_VUMC_male | BETA_UKB_female | BETA_UKB_male | SE_VUMC_female | SE_VUMC_male |
| Baso | rs28510728 | rs6057628 | C | T | 20 | COMMD7 | Female only | True | 0.2375 | 0.2399 | 0.2285 | 0.2288 | 15240 | 11083 | 187760 | 162101 | -0.0749 | -0.02085 | -0.0203 | -0.01023 | 0.01233 | 0.01398 |
| Baso | rs28510728 | rs2377665 | T | C | 20 | COMMD7 | Female only | True | 0.2398 | 0.2421 | 0.2305 | 0.2309 | 15240 | 11083 | 187760 | 162101 | -0.07446 | -0.01809 | -0.01949 | -0.009997 | 0.01231 | 0.01396 |
| Baso | rs28510728 | rs6058829 | G | A | 20 | COMMD7 | Female only | True | 0.2375 | 0.24 | 0.2285 | 0.2288 | 15240 | 11083 | 187760 | 162101 | -0.07481 | -0.02061 | -0.02031 | -0.01023 | 0.01233 | 0.01398 |
| Baso | rs28510728 | rs4425162 | A | G | 20 | COMMD7 | Female only | True | 0.2375 | 0.24 | 0.2285 | 0.2288 | 15240 | 11083 | 187760 | 162101 | -0.07482 | -0.02061 | -0.02032 | -0.01023 | 0.01233 | 0.01398 |
| Baso | rs28510728 | rs6058832 | A | G | 20 | COMMD7 | Female only | True | 0.2373 | 0.2397 | 0.2283 | 0.2287 | 15240 | 11083 | 187760 | 162101 | -0.07516 | -0.02154 | -0.02036 | -0.01032 | 0.01234 | 0.01397 |

| trait | lead_snp | ID | ALLELE1 | ALLELE0 | CHR | Gene_Symbol | independent_loci_in_sex_P1e2 | in_embl_any_or_gene | A1FREQ_VUMC_female | A1FREQ_VUMC_male | A1FREQ_UKB_female | A1FREQ_UKB_male | N_VUMC_female | N_VUMC_male | N_UKB_female | N_UKB_male | BETA_VUMC_female | BETA_VUMC_male | BETA_UKB_female | BETA_UKB_male | SE_VUMC_female | SE_VUMC_male |
|---|---|---|---|---|---|---|---|---|---|---|---|---|---|---|---|---|---|---|---|---|---|---|
| Baso | rs28510728 | rs2064933 | A | G | 20 | COMMD7 | Female only | True | 0.2326 | 0.2351 | 0.2283 | 0.2287 | 15240 | 11083 | 187760 | 162101 | -0.07505 | -0.02234 | -0.02031 | -0.01032 | 0.01227 | 0.0139 |
| Baso | rs28510728 | rs2064935 | G | A | 20 | COMMD7 | Female only | True | 0.2372 | 0.2396 | 0.2283 | 0.2287 | 15240 | 11083 | 187760 | 162101 | -0.07529 | -0.02178 | -0.02036 | -0.01032 | 0.01234 | 0.01398 |
| Baso | rs28510728 | rs6119939 | T | C | 20 | COMMD7 | Female only | True | 0.2373 | 0.2397 | 0.2283 | 0.2287 | 15240 | 11083 | 187760 | 162101 | -0.07523 | -0.02156 | -0.02036 | -0.01033 | 0.01234 | 0.01397 |
| Baso | rs28510728 | rs7360824 | A | G | 20 | COMMD7 | Female only | True | 0.2375 | 0.2399 | 0.2284 | 0.2288 | 15240 | 11083 | 187760 | 162101 | -0.07537 | -0.0212 | -0.02037 | -0.0104 | 0.01234 | 0.01398 |
| Baso | rs28510728 | rs6087976 | G | A | 20 | COMMD7 | Female only | True | 0.2372 | 0.2396 | 0.2283 | 0.2287 | 15240 | 11083 | 187760 | 162101 | -0.07529 | -0.02178 | -0.02036 | -0.01033 | 0.01234 | 0.01398 |
| QRS.DURATION | rs17226667 | rs9401060 | A | G | 6 | RP11-57K17.1 | Female only | True | 0.5005 | 0.4954 | 0.4927 | 0.4957 | 7224 | 6316 | 5542 | 5273 | 0.08012 | 0.004666 | 0.03852 | 0.04617 | 0.017 | 0.01891 |
| QRS.DURATION | rs17226667 | rs9489327 | C | T | 6 | RP11-57K17.1 | Female only | True | 0.488 | 0.4835 | 0.4972 | 0.4949 | 7224 | 6316 | 5542 | 5273 | 0.07992 | 5.51e-04 | 0.04009 | 0.05157 | 0.01695 | 0.0189 |
| QRS.DURATION | rs17226667 | rs6569009 | A | G | 6 | RP11-57K17.1 | Female only | True | 0.4659 | 0.4608 | 0.4763 | 0.474 | 7224 | 6316 | 5542 | 5273 | 0.08802 | -0.005428 | 0.04481 | 0.05773 | 0.01702 | 0.01884 |
| QRS.DURATION | rs17226667 | rs4946333 | A | G | 6 | RP11-57K17.1 | Female only | True | 0.4966 | 0.4918 | 0.4994 | 0.4987 | 7224 | 6316 | 5542 | 5273 | 0.08398 | 0.002057 | 0.03942 | 0.05194 | 0.01702 | 0.0189 |
| QRS.DURATION | rs17226667 | rs6912208 | A | G | 6 | RP11-57K17.1 | Female only | True | 0.4952 | 0.4907 | 0.4995 | 0.4984 | 7224 | 6316 | 5542 | 5273 | 0.08424 | 7.97e-04 | 0.03976 | 0.05145 | 0.01701 | 0.0189 |
| QRS.DURATION | rs17226667 | rs283077 | G | T | 6 | RP11-57K17.1 | Female only | True | 0.4952 | 0.491 | 0.4951 | 0.4983 | 7224 | 6316 | 5542 | 5273 | 0.08173 | 4.28e-04 | 0.03959 | 0.04896 | 0.01699 | 0.01887 |
| QRS.DURATION | rs17226667 | rs281868 | A | G | 6 | RP11-57K17.1 | Female only | True | 0.4938 | 0.4892 | 0.4996 | 0.4982 | 7224 | 6316 | 5542 | 5273 | 0.0844 | 1.28e-04 | 0.03884 | 0.05075 | 0.01695 | 0.01883 |
| QRS.DURATION | rs17226667 | rs17079898 | A | G | 6 | RP11-57K17.1 | Female only | True | 0.4634 | 0.4594 | 0.4739 | 0.4712 | 7224 | 6316 | 5542 | 5273 | 0.08944 | -0.007922 | 0.04497 | 0.05953 | 0.01702 | 0.01882 |
| QRS.DURATION | rs17226667 | rs89107 | A | G | 6 | RP11-57K17.1 | Female only | True | 0.4937 | 0.4892 | 0.4996 | 0.4982 | 7224 | 6316 | 5542 | 5273 | 0.08431 | 7.41e-05 | 0.03879 | 0.05075 | 0.01696 | 0.01883 |
| QRS.DURATION | rs17226667 | rs731695 | C | T | 6 | RP11-57K17.1 | Female only | True | 0.4659 | 0.4619 | 0.4779 | 0.4736 | 7224 | 6316 | 5542 | 5273 | 0.08532 | -0.00774 | 0.04497 | 0.05841 | 0.01698 | 0.0188 |
| QRS.DURATION | rs17226667 | rs927348 | A | G | 6 | RP11-57K17.1 | Female only | True | 0.4632 | 0.4587 | 0.4735 | 0.4704 | 7224 | 6316 | 5542 | 5273 | 0.08967 | -0.007546 | 0.04515 | 0.0608 | 0.01703 | 0.01883 |
| QRS.DURATION | rs17226667 | rs72965660 | G | A | 6 | RP11-57K17.1 | Female only | True | 0.4654 | 0.4612 | 0.4778 | 0.4735 | 7224 | 6316 | 5542 | 5273 | 0.08653 | -0.006986 | 0.04511 | 0.05887 | 0.01702 | 0.01883 |
| QRS.DURATION | rs17226667 | rs55697000 | T | G | 6 | RP11-57K17.1 | Female only | True | 0.463 | 0.4585 | 0.4733 | 0.4703 | 7224 | 6316 | 5542 | 5273 | 0.08954 | -0.007471 | 0.04527 | 0.0612 | 0.01703 | 0.01884 |
| QRS.DURATION | rs17226667 | rs7452622 | C | A | 6 | RP11-57K17.1 | Female only | True | 0.4631 | 0.4593 | 0.4763 | 0.4716 | 7224 | 6316 | 5542 | 5273 | 0.08711 | -0.007147 | 0.04568 | 0.05702 | 0.01703 | 0.01883 |
| QRS.DURATION | rs17226667 | rs11758375 | C | T | 6 | RP11-57K17.1 | Female only | True | 0.463 | 0.4594 | 0.4767 | 0.4714 | 7224 | 6316 | 5542 | 5273 | 0.08726 | -0.006696 | 0.0462 | 0.05647 | 0.01703 | 0.01882 |
| QRS.DURATION | rs17226667 | rs4294041 | G | A | 6 | RP11-57K17.1 | Female only | True | 0.4232 | 0.4221 | 0.4383 | 0.4359 | 7224 | 6316 | 5542 | 5273 | 0.08698 | -0.0063 | 0.05209 | 0.07162 | 0.01714 | 0.01891 |

| trait | lead_snp | ID | ALLELE1 | ALLELE0 | CHR | Gene_Symbol | independent_loci_in_sex_P1e2 | in_embl_any_or_gene | A1FREQ_VUMC_female | A1FREQ_VUMC_male | A1FREQ_UKB_female | A1FREQ_UKB_male | N_VUMC_female | N_VUMC_male | N_UKB_female | N_UKB_male | BETA_VUMC_female | BETA_VUMC_male | BETA_UKB_female | BETA_UKB_male | SE_VUMC_female | SE_VUMC_male |
|---|---|---|---|---|---|---|---|---|---|---|---|---|---|---|---|---|---|---|---|---|---|---|
| QRS.DURATION | rs17226667 | rs11758402 | G | A | 6 | RP11-57K17.1 | Female only | True | 0.4207 | 0.4193 | 0.4337 | 0.4324 | 7224 | 6316 | 5542 | 5273 | 0.09034 | -0.006507 | 0.05206 | 0.07376 | 0.01715 | 0.01893 |
| QRS.DURATION | rs17226667 | rs6940985 | G | T | 6 | RP11-57K17.1 | Female only | True | 0.4675 | 0.4641 | 0.4864 | 0.4822 | 7224 | 6316 | 5542 | 5273 | 0.08185 | -4.80e-04 | 0.04879 | 0.06565 | 0.01698 | 0.01867 |
| QRS.DURATION | rs17226667 | rs11968293 | A | C | 6 | RP11-57K17.1 | Female only | True | 0.5008 | 0.5028 | 0.486 | 0.4855 | 7224 | 6316 | 5542 | 5273 | -0.08577 | 0.00198 | -0.05128 | -0.06549 | 0.0169 | 0.01855 |
| QRS.DURATION | rs17226667 | rs7772845 | T | C | 6 | RP11-57K17.1 | Female only | True | 0.4353 | 0.4372 | 0.4436 | 0.4496 | 7224 | 6316 | 5542 | 5273 | 0.08345 | -0.005916 | 0.04571 | 0.06925 | 0.01707 | 0.01884 |
| QRS.DURATION | rs17226667 | rs2093369 | A | G | 6 | RP11-57K17.1 | Female only | True | 0.4372 | 0.4386 | 0.4457 | 0.4516 | 7224 | 6316 | 5542 | 5273 | 0.07994 | -0.004171 | 0.04741 | 0.06678 | 0.01709 | 0.01885 |
| QRS.DURATION | rs17226667 | rs6906332 | T | C | 6 | RP11-57K17.1 | Female only | True | 0.4358 | 0.4367 | 0.4441 | 0.4501 | 7224 | 6316 | 5542 | 5273 | 0.081 | -0.003119 | 0.04569 | 0.06934 | 0.01708 | 0.01885 |
| QRS.DURATION | rs17226667 | rs6911339 | A | G | 6 | RP11-57K17.1 | Female only | True | 0.4358 | 0.4367 | 0.4441 | 0.4501 | 7224 | 6316 | 5542 | 5273 | 0.081 | -0.00312 | 0.0457 | 0.06934 | 0.01708 | 0.01885 |
| QRS.DURATION | rs17226667 | rs72967533 | C | T | 6 | RP11-57K17.1 | Female only | True | 0.4752 | 0.4742 | 0.4883 | 0.4886 | 7224 | 6316 | 5542 | 5273 | 0.09176 | 9.51e-04 | 0.05001 | 0.07642 | 0.01692 | 0.01859 |
| QRS.DURATION | rs17226667 | rs11153730 | C | T | 6 | RP11-57K17.1 | Female only | True | 0.4836 | 0.4836 | 0.4983 | 0.4969 | 7224 | 6316 | 5542 | 5273 | 0.08995 | 0.007994 | 0.05323 | 0.07681 | 0.01688 | 0.01859 |
| QRS.DURATION | rs17226667 | rs1743241 | G | A | 6 | RP11-57K17.1 | Female only | True | 0.4481 | 0.4422 | 0.4292 | 0.4318 | 7224 | 6316 | 5542 | 5273 | -0.06686 | 0.00193 | -0.03675 | -0.06096 | 0.01674 | 0.01877 |
| QRS.DURATION | rs17226667 | rs11970286 | T | C | 6 | RP11-57K17.1 | Female only | True | 0.4437 | 0.4455 | 0.4636 | 0.4589 | 7224 | 6316 | 5542 | 5273 | 0.08778 | 0.01861 | 0.05519 | 0.07867 | 0.01698 | 0.01881 |
| QRS.DURATION | rs17226667 | rs6569014 | G | T | 6 | RP11-57K17.1 | Female only | True | 0.4609 | 0.4553 | 0.4417 | 0.4439 | 7224 | 6316 | 5542 | 5273 | -0.06863 | -7.99e-04 | -0.03999 | -0.06251 | 0.01688 | 0.01886 |
| QRS.DURATION | rs17226667 | rs12665409 | G | A | 6 | RP11-57K17.1 | Female only | True | 0.4636 | 0.4588 | 0.444 | 0.4469 | 7224 | 6316 | 5542 | 5273 | -0.07162 | -0.002287 | -0.04398 | -0.05902 | 0.01688 | 0.01886 |
| QRS.DURATION | rs17226667 | rs9489355 | A | G | 6 | RP11-57K17.1 | Female only | True | 0.4372 | 0.433 | 0.4222 | 0.4243 | 7224 | 6316 | 5542 | 5273 | -0.06564 | -0.007658 | -0.03624 | -0.05825 | 0.01698 | 0.01898 |
| QRS.DURATION | rs17226667 | rs6569015 | C | T | 6 | RP11-57K17.1 | Female only | True | 0.4636 | 0.4588 | 0.4442 | 0.4469 | 7224 | 6316 | 5542 | 5273 | -0.07165 | -0.002294 | -0.04438 | -0.05896 | 0.01688 | 0.01886 |
| QRS.DURATION | rs17226667 | rs2077162 | G | A | 6 | RP11-57K17.1 | Female only | True | 0.4636 | 0.4588 | 0.4441 | 0.4469 | 7224 | 6316 | 5542 | 5273 | -0.07164 | -0.002294 | -0.04391 | -0.05896 | 0.01688 | 0.01886 |
| QRS.DURATION | rs17226667 | rs28655926 | A | G | 6 | RP11-57K17.1 | Female only | True | 0.4636 | 0.4588 | 0.4441 | 0.4469 | 7224 | 6316 | 5542 | 5273 | -0.07166 | -0.002299 | -0.04392 | -0.05893 | 0.01688 | 0.01886 |
| QRS.DURATION | rs17226667 | rs9320648 | A | C | 6 | RP11-57K17.1 | Female only | True | 0.4379 | 0.4338 | 0.4222 | 0.4243 | 7224 | 6316 | 5542 | 5273 | -0.066 | -0.007826 | -0.03623 | -0.05823 | 0.01699 | 0.019 |
| QRS.DURATION | rs17226667 | rs7764272 | A | C | 6 | RP11-57K17.1 | Female only | True | 0.4371 | 0.433 | 0.4222 | 0.4244 | 7224 | 6316 | 5542 | 5273 | -0.06567 | -0.007654 | -0.03651 | -0.05829 | 0.01698 | 0.01898 |
| QRS.DURATION | rs17226667 | rs7764280 | A | G | 6 | RP11-57K17.1 | Female only | True | 0.437 | 0.433 | 0.4222 | 0.4244 | 7224 | 6316 | 5542 | 5273 | -0.06584 | -0.007659 | -0.03637 | -0.05823 | 0.01698 | 0.01898 |

| trait | lead_snp | ID | ALLELE1 | ALLELE0 | CHR | Gene_Symbol | independent_loci_in_sex_P1e2 | in_embl_any_or_gene | A1FREQ_VUMC_female | A1FREQ_VUMC_male | A1FREQ_UKB_female | A1FREQ_UKB_male | N_VUMC_female | N_VUMC_male | N_UKB_female | N_UKB_male | BETA_VUMC_female | BETA_VUMC_male | BETA_UKB_female | BETA_UKB_male | SE_VUMC_female | SE_VUMC_male |
|---|---|---|---|---|---|---|---|---|---|---|---|---|---|---|---|---|---|---|---|---|---|---|
| QRS.DURATION | rs17226667 | rs9320649 | G | A | 6 | RP11-57K17.1 | Female only | True | 0.4372 | 0.4331 | 0.4222 | 0.4242 | 7224 | 6316 | 5542 | 5273 | -0.06623 | -0.007862 | -0.03636 | -0.05824 | 0.01698 | 0.01898 |
| QRS.DURATION | rs17226667 | rs6920640 | T | G | 6 | RP11-57K17.1 | Female only | True | 0.4632 | 0.4584 | 0.444 | 0.4466 | 7224 | 6316 | 5542 | 5273 | -0.07187 | -0.002538 | -0.04421 | -0.05891 | 0.01689 | 0.01886 |
| QRS.DURATION | rs17226667 | rs6903459 | G | T | 6 | RP11-57K17.1 | Female only | True | 0.4978 | 0.495 | 0.4763 | 0.4819 | 7224 | 6316 | 5542 | 5273 | -0.06595 | -0.01453 | -0.04238 | -0.05906 | 0.01695 | 0.0189 |
| QRS.DURATION | rs17226667 | rs7767864 | T | C | 6 | RP11-57K17.1 | Female only | True | 0.4987 | 0.4966 | 0.4775 | 0.4835 | 7224 | 6316 | 5542 | 5273 | -0.06598 | -0.0133 | -0.04256 | -0.05736 | 0.01693 | 0.01892 |
| QRS.DURATION | rs17226667 | rs57912492 | T | C | 6 | RP11-57K17.1 | Female only | True | 0.4133 | 0.4128 | 0.4346 | 0.4252 | 7224 | 6316 | 5542 | 5273 | 0.08843 | 0.03075 | 0.06109 | 0.07245 | 0.01714 | 0.0191 |
| QRS.DURATION | rs17226667 | rs56403768 | C | T | 6 | RP11-57K17.1 | Female only | True | 0.4133 | 0.4124 | 0.4341 | 0.425 | 7224 | 6316 | 5542 | 5273 | 0.08793 | 0.03101 | 0.06119 | 0.0715 | 0.01714 | 0.0191 |
| QRS.DURATION | rs17226667 | rs11153732 | C | T | 6 | RP11-57K17.1 | Female only | True | 0.4995 | 0.4974 | 0.4798 | 0.485 | 7224 | 6316 | 5542 | 5273 | -0.06702 | -0.01318 | -0.04314 | -0.05512 | 0.01691 | 0.01886 |
| QRS.DURATION | rs17226667 | rs2078079 | G | A | 6 | RP11-57K17.1 | Female only | True | 0.4743 | 0.472 | 0.457 | 0.4622 | 7224 | 6316 | 5542 | 5273 | -0.06224 | -0.01716 | -0.03647 | -0.05379 | 0.01694 | 0.01888 |
| QRS.DURATION | rs17226667 | rs3890198 | T | C | 6 | RP11-57K17.1 | Female only | True | 0.4726 | 0.4682 | 0.4528 | 0.4559 | 7224 | 6316 | 5542 | 5273 | -0.07323 | -0.002899 | -0.04518 | -0.06012 | 0.01687 | 0.01885 |
| QRS.DURATION | rs17226667 | rs2078383 | T | C | 6 | RP11-57K17.1 | Female only | True | 0.4394 | 0.4413 | 0.4616 | 0.4538 | 7224 | 6316 | 5542 | 5273 | 0.09382 | 0.02022 | 0.06041 | 0.07655 | 0.01699 | 0.01886 |
| QRS.DURATION | rs17226667 | rs35730412 | T | C | 6 | RP11-57K17.1 | Female only | True | 0.4718 | 0.4675 | 0.452 | 0.4553 | 7224 | 6316 | 5542 | 5273 | -0.07246 | -0.003529 | -0.04556 | -0.05996 | 0.01688 | 0.01887 |
| QRS.DURATION | rs17226667 | rs12660455 | C | T | 6 | RP11-57K17.1 | Female only | True | 0.4729 | 0.4675 | 0.4526 | 0.4554 | 7224 | 6316 | 5542 | 5273 | -0.07299 | -0.005497 | -0.04419 | -0.05953 | 0.01688 | 0.01888 |
| QRS.DURATION | rs17226667 | rs6936178 | C | T | 6 | RP11-57K17.1 | Female only | True | 0.4729 | 0.4675 | 0.4526 | 0.4554 | 7224 | 6316 | 5542 | 5273 | -0.07299 | -0.005497 | -0.04419 | -0.05954 | 0.01688 | 0.01888 |
| QRS.DURATION | rs17226667 | rs17226667 | G | A | 6 | RP11-57K17.1 | Female only | True | 0.4435 | 0.4471 | 0.4655 | 0.4602 | 7224 | 6316 | 5542 | 5273 | 0.09463 | 0.02146 | 0.05986 | 0.07647 | 0.017 | 0.01884 |
| QRS.DURATION | rs17226667 | rs3951042 | C | T | 6 | RP11-57K17.1 | Female only | True | 0.473 | 0.4675 | 0.4526 | 0.4552 | 7224 | 6316 | 5542 | 5273 | -0.07308 | -0.005504 | -0.04427 | -0.05948 | 0.01688 | 0.01888 |
| QRS.DURATION | rs17226667 | rs7746778 | C | A | 6 | RP11-57K17.1 | Female only | True | 0.4729 | 0.4677 | 0.4525 | 0.4553 | 7224 | 6316 | 5542 | 5273 | -0.07308 | -0.006125 | -0.0442 | -0.05945 | 0.01688 | 0.01889 |
| QRS.DURATION | rs17226667 | rs3951043 | T | C | 6 | RP11-57K17.1 | Female only | True | 0.4734 | 0.4684 | 0.4526 | 0.4552 | 7224 | 6316 | 5542 | 5273 | -0.07434 | -0.005401 | -0.04394 | -0.05848 | 0.0169 | 0.01892 |
| QRS.DURATION | rs17226667 | rs10457331 | C | A | 6 | RP11-57K17.1 | Female only | True | 0.4733 | 0.4684 | 0.4529 | 0.4557 | 7224 | 6316 | 5542 | 5273 | -0.07405 | -0.005304 | -0.04385 | -0.06033 | 0.0169 | 0.01892 |
| QRS.DURATION | rs17226667 | rs17227124 | C | T | 6 | RP11-57K17.1 | Female only | True | 0.4437 | 0.4461 | 0.4649 | 0.4596 | 7224 | 6316 | 5542 | 5273 | 0.0937 | 0.02105 | 0.05785 | 0.07612 | 0.01699 | 0.01886 |

| trait | lead_snp | ID | ALLELE1 | ALLELE0 | CHR | Gene_Symbol | independent_loci_in_sex_P1e2 | in_embl_any_or_gene | A1FREQ_VUMC_female | A1FREQ_VUMC_male | A1FREQ_UKB_female | A1FREQ_UKB_male | N_VUMC_female | N_VUMC_male | N_UKB_female | N_UKB_male | BETA_VUMC_female | BETA_VUMC_male | BETA_UKB_female | BETA_UKB_male | SE_VUMC_female | SE_VUMC_male |
|---|---|---|---|---|---|---|---|---|---|---|---|---|---|---|---|---|---|---|---|---|---|---|
| QRS.DURATION | rs55892056 | rs2356176 | G | A | 6 | CEP85L | Female only | True | 0.4732 | 0.4684 | 0.4529 | 0.4558 | 7224 | 6316 | 5542 | 5273 | -0.07372 | -0.005351 | -0.04385 | -0.05994 | 0.0169 | 0.01892 |
| QRS.DURATION | rs55892056 | rs144175702 | G | T | 6 | CEP85L | Female only | True | 0.4731 | 0.4684 | 0.4529 | 0.4558 | 7224 | 6316 | 5542 | 5273 | -0.07378 | -0.005342 | -0.0439 | -0.05996 | 0.0169 | 0.01892 |
| QRS.DURATION | rs55892056 | rs6922637 | C | T | 6 | CEP85L | Female only | True | 0.4734 | 0.4687 | 0.4529 | 0.4558 | 7224 | 6316 | 5542 | 5273 | -0.07384 | -0.005252 | -0.04384 | -0.05997 | 0.0169 | 0.01893 |
| QRS.DURATION | rs55892056 | rs56399949 | G | A | 6 | CEP85L | Female only | True | 0.4426 | 0.4449 | 0.4649 | 0.4595 | 7224 | 6316 | 5542 | 5273 | 0.09345 | 0.02117 | 0.05779 | 0.076 | 0.01702 | 0.01889 |
| QRS.DURATION | rs55892056 | rs57813349 | C | A | 6 | CEP85L | Female only | True | 0.4717 | 0.4678 | 0.4528 | 0.4555 | 7224 | 6316 | 5542 | 5273 | -0.07382 | -0.00535 | -0.04375 | -0.06081 | 0.0169 | 0.01893 |
| QRS.DURATION | rs55892056 | rs2356183 | A | G | 6 | CEP85L | Female only | True | 0.4725 | 0.4679 | 0.4529 | 0.4558 | 7224 | 6316 | 5542 | 5273 | -0.07392 | -0.005105 | -0.04382 | -0.05997 | 0.01688 | 0.0189 |
| QRS.DURATION | rs55892056 | rs17825393 | C | T | 6 | CEP85L | Female only | True | 0.4433 | 0.4457 | 0.4652 | 0.46 | 7224 | 6316 | 5542 | 5273 | 0.09356 | 0.02069 | 0.05745 | 0.07424 | 0.01699 | 0.01885 |
| QRS.DURATION | rs55892056 | rs11153748 | A | G | 6 | CEP85L | Female only | True | 0.4725 | 0.4679 | 0.4526 | 0.4553 | 7224 | 6316 | 5542 | 5273 | -0.07392 | -0.005105 | -0.04388 | -0.05811 | 0.01688 | 0.0189 |
| QRS.DURATION | rs55892056 | rs10456917 | A | C | 6 | CEP85L | Female only | True | 0.4711 | 0.4661 | 0.4516 | 0.4538 | 7224 | 6316 | 5542 | 5273 | -0.07437 | -0.003863 | -0.04423 | -0.05668 | 0.01687 | 0.01892 |
| QRS.DURATION | rs55892056 | rs10457335 | G | A | 6 | CEP85L | Female only | True | 0.4734 | 0.4687 | 0.4526 | 0.4553 | 7224 | 6316 | 5542 | 5273 | -0.07384 | -0.005262 | -0.04392 | -0.0581 | 0.0169 | 0.01893 |
| QRS.DURATION | rs55892056 | 6:118781359 | A | G | 6 | CEP85L | Female only | True | 0.472 | 0.467 | | | 7224 | 6316 | | | -0.07428 | -0.004014 | | | 0.0169 | 0.01894 |
| QRS.DURATION | rs55892056 | rs10457338 | T | C | 6 | CEP85L | Female only | True | 0.472 | 0.467 | 0.4519 | 0.4542 | 7224 | 6316 | 5542 | 5273 | -0.07425 | -0.004026 | -0.04427 | -0.05914 | 0.0169 | 0.01894 |
| QRS.DURATION | rs55892056 | rs3798420 | T | C | 6 | CEP85L | Female only | True | 0.472 | 0.467 | 0.4516 | 0.4537 | 7224 | 6316 | 5542 | 5273 | -0.07425 | -0.004026 | -0.04423 | -0.05728 | 0.0169 | 0.01894 |
| QRS.DURATION | rs55892056 | rs11752626 | T | G | 6 | CEP85L | Female only | True | 0.444 | 0.4466 | 0.4661 | 0.4616 | 7224 | 6316 | 5542 | 5273 | 0.09372 | 0.02 | 0.05789 | 0.07326 | 0.01701 | 0.0189 |
| QRS.DURATION | rs55892056 | rs17825652 | A | G | 6 | CEP85L | Female only | True | 0.4426 | 0.4449 | 0.4651 | 0.4601 | 7224 | 6316 | 5542 | 5273 | 0.09329 | 0.02123 | 0.05768 | 0.07481 | 0.01703 | 0.01889 |
| QRS.DURATION | rs55892056 | rs78757409 | T | C | 6 | CEP85L | Female only | True | 0.4439 | 0.4466 | 0.4661 | 0.4616 | 7224 | 6316 | 5542 | 5273 | 0.09369 | 0.02001 | 0.05791 | 0.07328 | 0.01701 | 0.01889 |
| QRS.DURATION | rs55892056 | rs11965985 | C | T | 6 | CEP85L | Female only | True | 0.4439 | 0.4466 | 0.4662 | 0.4617 | 7224 | 6316 | 5542 | 5273 | 0.09369 | 0.02007 | 0.05771 | 0.07319 | 0.01701 | 0.01889 |
| QRS.DURATION | rs55892056 | rs12661338 | A | C | 6 | CEP85L | Female only | True | 0.4412 | 0.4433 | 0.4639 | 0.4589 | 7224 | 6316 | 5542 | 5273 | 0.0922 | 0.02048 | 0.05912 | 0.07447 | 0.01705 | 0.01892 |
| QRS.DURATION | rs55892056 | rs6569020 | C | T | 6 | CEP85L | Female only | True | 0.473 | 0.4686 | 0.4525 | 0.4551 | 7224 | 6316 | 5542 | 5273 | -0.0725 | -0.006065 | -0.04366 | -0.05904 | 0.01692 | 0.01894 |
| QRS.DURATION | rs55892056 | rs2213857 | A | G | 6 | CEP85L | Female only | True | 0.4733 | 0.4688 | 0.4525 | 0.4552 | 7224 | 6316 | 5542 | 5273 | -0.07475 | -0.004967 | -0.0439 | -0.05791 | 0.01691 | 0.01893 |

| trait | lead_snp | ID | ALLELE1 | ALLELE0 | CHR | Gene_Symbol | independent_loci_in_sex_P1e2 | in_embl_any_or_gene | A1FREQ_VUMC_female | A1FREQ_VUMC_male | A1FREQ_UKB_female | A1FREQ_UKB_male | N_VUMC_female | N_VUMC_male | N_UKB_female | N_UKB_male | BETA_VUMC_female | BETA_VUMC_male | BETA_UKB_female | BETA_UKB_male | SE_VUMC_female | SE_VUMC_male |
|---|---|---|---|---|---|---|---|---|---|---|---|---|---|---|---|---|---|---|---|---|---|---|
| QRS.DURATION | rs55892056 | rs2213856 | C | T | 6 | CEP85L | Female only | True | 0.4722 | 0.4677 | 0.4525 | 0.4552 | 7224 | 6316 | 5542 | 5273 | -0.07484 | -0.004825 | -0.04398 | -0.0578 | 0.01688 | 0.0189 |
| QRS.DURATION | rs55892056 | rs4307206 | A | C | 6 | CEP85L | Female only | True | 0.4435 | 0.4462 | 0.4662 | 0.4616 | 7224 | 6316 | 5542 | 5273 | 0.09335 | 0.02055 | 0.0578 | 0.0729 | 0.01701 | 0.0189 |
| QRS.DURATION | rs55892056 | rs13219555 | G | A | 6 | CEP85L | Female only | True | 0.4732 | 0.4688 | 0.4528 | 0.4554 | 7224 | 6316 | 5542 | 5273 | -0.07312 | -0.006656 | -0.04398 | -0.05852 | 0.01693 | 0.01894 |
| QRS.DURATION | rs55892056 | rs12194555 | C | A | 6 | CEP85L | Female only | True | 0.4695 | 0.4653 | 0.4516 | 0.4536 | 7224 | 6316 | 5542 | 5273 | -0.07372 | -0.004838 | -0.04406 | -0.0584 | 0.01687 | 0.01893 |
| QRS.DURATION | rs55892056 | rs6925641 | C | T | 6 | CEP85L | Female only | True | 0.4717 | 0.4669 | 0.4517 | 0.4538 | 7224 | 6316 | 5542 | 5273 | -0.0729 | -0.005718 | -0.04411 | -0.05779 | 0.01692 | 0.01895 |
| QRS.DURATION | rs55892056 | rs11967375 | G | A | 6 | CEP85L | Female only | True | 0.4424 | 0.4449 | 0.465 | 0.46 | 7224 | 6316 | 5542 | 5273 | 0.09363 | 0.02166 | 0.0577 | 0.07504 | 0.01703 | 0.0189 |
| QRS.DURATION | rs55892056 | rs12199463 | G | A | 6 | CEP85L | Female only | True | 0.4724 | 0.4675 | 0.452 | 0.4539 | 7224 | 6316 | 5542 | 5273 | -0.07427 | -0.004524 | -0.04458 | -0.0575 | 0.01691 | 0.01896 |
| QRS.DURATION | rs55892056 | rs67832971 | C | T | 6 | CEP85L | Female only | True | 0.4724 | 0.4675 | 0.452 | 0.4542 | 7224 | 6316 | 5542 | 5273 | -0.07416 | -0.004492 | -0.04469 | -0.05767 | 0.01691 | 0.01895 |
| QRS.DURATION | rs55892056 | rs11967858 | T | C | 6 | CEP85L | Female only | True | 0.442 | 0.4445 | 0.465 | 0.46 | 7224 | 6316 | 5542 | 5273 | 0.09319 | 0.02159 | 0.05773 | 0.07482 | 0.01704 | 0.0189 |
| QRS.DURATION | rs55892056 | rs11153753 | T | C | 6 | CEP85L | Female only | True | 0.4722 | 0.4674 | 0.452 | 0.4541 | 7224 | 6316 | 5542 | 5273 | -0.07447 | -0.00403 | -0.04469 | -0.05752 | 0.01691 | 0.01896 |
| QRS.DURATION | rs55892056 | rs113127380 | C | T | 6 | CEP85L | Female only | True | 0.4436 | 0.4463 | 0.4659 | 0.4613 | 7224 | 6316 | 5542 | 5273 | 0.09357 | 0.01973 | 0.05773 | 0.07375 | 0.01702 | 0.0189 |
| QRS.DURATION | rs55892056 | rs74680048 | C | T | 6 | CEP85L | Female only | True | 0.4722 | 0.4672 | | | 7224 | 6316 | | | -0.07423 | -0.003996 | | | 0.01691 | 0.01896 |
| QRS.DURATION | rs55892056 | rs113989395 | T | C | 6 | CEP85L | Female only | True | 0.444 | 0.4467 | | | 7224 | 6316 | | | 0.09372 | 0.02009 | | | 0.01701 | 0.01891 |
| QRS.DURATION | rs55892056 | rs4946347 | G | A | 6 | CEP85L | Female only | True | 0.4787 | 0.474 | 0.4579 | 0.4603 | 7224 | 6316 | 5542 | 5273 | -0.07113 | -0.00509 | -0.04382 | -0.05843 | 0.01692 | 0.01891 |
| QRS.DURATION | rs55892056 | rs13217710 | A | G | 6 | CEP85L | Female only | True | 0.4773 | 0.4725 | 0.4569 | 0.4589 | 7224 | 6316 | 5542 | 5273 | -0.07171 | -0.003817 | -0.04403 | -0.05768 | 0.01691 | 0.01892 |
| QRS.DURATION | rs55892056 | rs7757892 | G | A | 6 | CEP85L | Female only | True | 0.4773 | 0.4725 | 0.4569 | 0.4589 | 7224 | 6316 | 5542 | 5273 | -0.0715 | -0.003828 | -0.04409 | -0.05741 | 0.01691 | 0.01892 |
| QRS.DURATION | rs55892056 | rs11758454 | C | A | 6 | CEP85L | Female only | True | 0.4385 | 0.4412 | 0.4611 | 0.4566 | 7224 | 6316 | 5542 | 5273 | 0.09081 | 0.0198 | 0.05732 | 0.07382 | 0.01705 | 0.01892 |
| QRS.DURATION | rs55892056 | rs12207209 | T | G | 6 | CEP85L | Female only | True | 0.4775 | 0.4726 | 0.4568 | 0.4588 | 7224 | 6316 | 5542 | 5273 | -0.07109 | -0.004226 | -0.04402 | -0.05743 | 0.01691 | 0.01892 |
| QRS.DURATION | rs55892056 | rs973944 | T | G | 6 | CEP85L | Female only | True | 0.4721 | 0.4672 | 0.452 | 0.4541 | 7224 | 6316 | 5542 | 5273 | -0.07407 | -0.003783 | -0.04469 | -0.05754 | 0.01691 | 0.01896 |
| QRS.DURATION | rs55892056 | rs6938041 | C | A | 6 | CEP85L | Female only | True | 0.4774 | 0.4725 | 0.4571 | 0.4594 | 7224 | 6316 | 5542 | 5273 | -0.07043 | -0.004577 | -0.04414 | -0.05955 | 0.01691 | 0.01892 |

| trait | lead_snp | ID | ALLELE1 | ALLELE0 | CHR | Gene_Symbol | independent_loci_in_sex_P1e2 | in_embl_any_or_gene | A1FREQ_VUMC_female | A1FREQ_VUMC_male | A1FREQ_UKB_female | A1FREQ_UKB_male | N_VUMC_female | N_VUMC_male | N_UKB_female | N_UKB_male | BETA_VUMC_female | BETA_VUMC_male | BETA_UKB_female | BETA_UKB_male | SE_VUMC_female | SE_VUMC_male |
|---|---|---|---|---|---|---|---|---|---|---|---|---|---|---|---|---|---|---|---|---|---|---|
| QRS.DURATION | rs55892056 | rs6938226 | C | A | 6 | CEP85L | Female only | True | 0.4774 | 0.4725 | 0.4572 | 0.4595 | 7224 | 6316 | 5542 | 5273 | -0.07042 | -0.004579 | -0.04399 | -0.05948 | 0.01691 | 0.01892 |
| QRS.DURATION | rs55892056 | rs11753128 | C | T | 6 | CEP85L | Female only | True | 0.4774 | 0.4725 | 0.4572 | 0.4596 | 7224 | 6316 | 5542 | 5273 | -0.07043 | -0.004577 | -0.044 | -0.05978 | 0.01691 | 0.01892 |
| QRS.DURATION | rs55892056 | rs12192682 | A | G | 6 | CEP85L | Female only | True | 0.4708 | 0.4659 | 0.4524 | 0.4549 | 7224 | 6316 | 5542 | 5273 | -0.07324 | -0.003799 | -0.04426 | -0.06007 | 0.01687 | 0.01892 |
| QRS.DURATION | rs55892056 | rs12206329 | G | T | 6 | CEP85L | Female only | True | 0.4708 | 0.4659 | 0.4524 | 0.4548 | 7224 | 6316 | 5542 | 5273 | -0.07324 | -0.003799 | -0.0444 | -0.05983 | 0.01687 | 0.01892 |
| QRS.DURATION | rs55892056 | rs3752581 | G | A | 6 | CEP85L | Female only | True | 0.4711 | 0.4661 | 0.4524 | 0.4548 | 7224 | 6316 | 5542 | 5273 | -0.07257 | -0.004364 | -0.04446 | -0.05938 | 0.01687 | 0.01891 |
| QRS.DURATION | rs55892056 | rs12211576 | G | A | 6 | CEP85L | Female only | True | 0.4711 | 0.466 | 0.4524 | 0.4547 | 7224 | 6316 | 5542 | 5273 | -0.07259 | -0.004268 | -0.04447 | -0.05937 | 0.01687 | 0.01891 |
| QRS.DURATION | rs55892056 | rs13192574 | C | T | 6 | CEP85L | Female only | True | 0.4721 | 0.467 | 0.4525 | 0.4549 | 7224 | 6316 | 5542 | 5273 | -0.07278 | -0.003962 | -0.04434 | -0.0596 | 0.0169 | 0.01895 |
| QRS.DURATION | rs55892056 | rs2356497 | T | C | 6 | CEP85L | Female only | True | 0.4442 | 0.447 | 0.4655 | 0.4607 | 7224 | 6316 | 5542 | 5273 | 0.09201 | 0.02043 | 0.05762 | 0.07542 | 0.01701 | 0.01891 |
| QRS.DURATION | rs17226667 | rs11753886 | A | G | 6 | RP11-57K17.1 | Female only | True | 0.3135 | 0.3153 | 0.3251 | 0.3238 | 7224 | 6316 | 5542 | 5273 | 0.07156 | 0.01933 | 0.05979 | 0.07821 | 0.0182 | 0.02039 |
| QRS.DURATION | rs55892056 | rs12198461 | T | G | 6 | CEP85L | Female only | True | 0.472 | 0.4669 | 0.4524 | 0.4547 | 7224 | 6316 | 5542 | 5273 | -0.07276 | -0.004384 | -0.04453 | -0.0593 | 0.01691 | 0.01896 |
| QRS.DURATION | rs55892056 | rs72952771 | T | C | 6 | CEP85L | Female only | True | 0.4443 | 0.4474 | 0.4652 | 0.4607 | 7224 | 6316 | 5542 | 5273 | 0.09137 | 0.02118 | 0.05668 | 0.07559 | 0.01698 | 0.01887 |
| QRS.DURATION | rs55892056 | rs12197337 | C | T | 6 | CEP85L | Female only | True | 0.4712 | 0.466 | 0.4525 | 0.4547 | 7224 | 6316 | 5542 | 5273 | -0.07263 | -0.004267 | -0.04387 | -0.06051 | 0.01687 | 0.01891 |
| QRS.DURATION | rs55892056 | rs763254 | C | T | 6 | CEP85L | Female only | True | 0.4452 | 0.4481 | 0.4653 | 0.4607 | 7224 | 6316 | 5542 | 5273 | 0.09198 | 0.02028 | 0.05678 | 0.07547 | 0.01698 | 0.01886 |
| QRS.DURATION | rs55892056 | rs3734381 | T | C | 6 | CEP85L | Female only | True | 0.4692 | 0.4651 | 0.4525 | 0.4545 | 7224 | 6316 | 5542 | 5273 | -0.07352 | -0.003201 | -0.04458 | -0.06021 | 0.01688 | 0.01893 |
| QRS.DURATION | rs55892056 | rs7758614 | T | C | 6 | CEP85L | Female only | True | 0.4688 | 0.4638 | 0.4504 | 0.4522 | 7224 | 6316 | 5542 | 5273 | -0.07517 | -0.005044 | -0.04718 | -0.06042 | 0.01687 | 0.01893 |
| trait | lead_snp | ID | ALLELE1 | ALLELE0 | CHR | Gene_Symbol | independent_loci_in_sex_P1e2 | in_embl_any_or_gene | A1FREQ_VUMC_female | A1FREQ_VUMC_male | A1FREQ_UKB_female | A1FREQ_UKB_male | N_VUMC_female | N_VUMC_male | N_UKB_female | N_UKB_male | BETA_VUMC_female | BETA_VUMC_male | BETA_UKB_female | BETA_UKB_male | SE_VUMC_female | SE_VUMC_male |
| QRS.DURATION | rs55892056 | rs35029749 | C | A | 6 | CEP85L | Female only | True | 0.4457 | 0.4486 | 0.4675 | 0.4632 | 7224 | 6316 | 5542 | 5273 | 0.09341 | 0.02138 | 0.0592 | 0.07639 | 0.01702 | 0.01889 |
| QRS.DURATION | rs55892056 | rs72952795 | T | G | 6 | CEP85L | Female only | True | 0.4468 | 0.4497 | 0.4674 | 0.4632 | 7224 | 6316 | 5542 | 5273 | 0.09323 | 0.02115 | 0.05925 | 0.07642 | 0.01697 | 0.01885 |
| QRS.DURATION | rs55892056 | rs7763596 | G | A | 6 | CEP85L | Female only | True | 0.4687 | 0.4642 | 0.4503 | 0.452 | 7224 | 6316 | 5542 | 5273 | -0.07398 | -0.005712 | -0.04722 | -0.06141 | 0.0169 | 0.01897 |
| QRS.DURATION | rs55892056 | rs11968176 | A | C | 6 | CEP85L | Female only | True | 0.4466 | 0.4497 | 0.4674 | 0.4632 | 7224 | 6316 | 5542 | 5273 | 0.09279 | 0.02193 | 0.05927 | 0.07629 | 0.01697 | 0.01885 |

| trait | lead_snp | ID | ALLELE1 | ALLELE0 | CHR | Gene_Symbol | independent_loci_in_sex_P1e2 | in_embl_any_or_gene | A1FREQ_VUMC_female | A1FREQ_VUMC_male | A1FREQ_UKB_female | A1FREQ_UKB_male | N_VUMC_female | N_VUMC_male | N_UKB_female | N_UKB_male | BETA_VUMC_female | BETA_VUMC_male | BETA_UKB_female | BETA_UKB_male | SE_VUMC_female | SE_VUMC_male |
|---|---|---|---|---|---|---|---|---|---|---|---|---|---|---|---|---|---|---|---|---|---|---|
| QRS.DURATION | rs55892056 | rs11968351 | G | A | 6 | CEP85L | Female only | True | 0.469 | 0.4637 | 0.4503 | 0.4522 | 7224 | 6316 | 5542 | 5273 | -0.074 | -0.005923 | -0.04723 | -0.06041 | 0.01687 | 0.01892 |
| QRS.DURATION | rs55892056 | rs13220480 | G | A | 6 | CEP85L | Female only | True | 0.4691 | 0.464 | 0.4503 | 0.4522 | 7224 | 6316 | 5542 | 5273 | -0.07428 | -0.005503 | -0.04728 | -0.06035 | 0.01687 | 0.01893 |
| QRS.DURATION | rs55892056 | rs12203006 | C | T | 6 | CEP85L | Female only | True | 0.4691 | 0.464 | 0.4503 | 0.4522 | 7224 | 6316 | 5542 | 5273 | -0.07427 | -0.005503 | -0.04729 | -0.06037 | 0.01687 | 0.01893 |
| QRS.DURATION | rs55892056 | rs7766375 | G | A | 6 | CEP85L | Female only | True | 0.4683 | 0.4633 | 0.4503 | 0.4521 | 7224 | 6316 | 5542 | 5273 | -0.0737 | -0.005956 | -0.04729 | -0.06058 | 0.01688 | 0.01892 |
| QRS.DURATION | rs55892056 | rs35962656 | G | T | 6 | CEP85L | Female only | True | 0.4689 | 0.4638 | 0.4503 | 0.4522 | 7224 | 6316 | 5542 | 5273 | -0.07458 | -0.005044 | -0.04729 | -0.06034 | 0.01687 | 0.01893 |
| QRS.DURATION | rs55892056 | rs7769001 | A | G | 6 | CEP85L | Female only | True | 0.4689 | 0.4639 | 0.4503 | 0.4523 | 7224 | 6316 | 5542 | 5273 | -0.07458 | -0.00506 | -0.04728 | -0.06013 | 0.01687 | 0.01893 |
| QRS.DURATION | rs55892056 | rs2184402 | T | C | 6 | CEP85L | Female only | True | 0.4689 | 0.4639 | 0.4503 | 0.4523 | 7224 | 6316 | 5542 | 5273 | -0.07456 | -0.00506 | -0.04727 | -0.06011 | 0.01687 | 0.01893 |
| QRS.DURATION | rs55892056 | rs6902812 | A | G | 6 | CEP85L | Female only | True | 0.4689 | 0.4639 | 0.4503 | 0.4523 | 7224 | 6316 | 5542 | 5273 | -0.07456 | -0.00506 | -0.04727 | -0.06012 | 0.01687 | 0.01893 |
| QRS.DURATION | rs55892056 | rs2883956 | T | C | 6 | CEP85L | Female only | True | 0.4702 | 0.4653 | 0.4503 | 0.4523 | 7224 | 6316 | 5542 | 5273 | -0.07449 | -0.005219 | -0.04727 | -0.06011 | 0.01691 | 0.01897 |
| QRS.DURATION | rs55892056 | rs11153763 | G | A | 6 | CEP85L | Female only | True | 0.4683 | 0.4634 | 0.4503 | 0.4522 | 7224 | 6316 | 5542 | 5273 | -0.0737 | -0.005973 | -0.04727 | -0.06032 | 0.01688 | 0.01892 |
| QRS.DURATION | rs55892056 | rs13191610 | G | A | 6 | CEP85L | Female only | True | 0.4684 | 0.4634 | 0.4507 | 0.4527 | 7224 | 6316 | 5542 | 5273 | -0.07351 | -0.005973 | -0.04764 | -0.06062 | 0.01688 | 0.01892 |
| QRS.DURATION | rs55892056 | rs4946350 | A | C | 6 | CEP85L | Female only | True | 0.4698 | 0.4648 | 0.4506 | 0.4527 | 7224 | 6316 | 5542 | 5273 | -0.07345 | -0.006115 | -0.04759 | -0.0606 | 0.01692 | 0.01896 |
| QRS.DURATION | rs55892056 | rs10457342 | A | G | 6 | CEP85L | Female only | True | 0.4689 | 0.4639 | 0.4504 | 0.4521 | 7224 | 6316 | 5542 | 5273 | -0.07458 | -0.00506 | -0.04704 | -0.05931 | 0.01687 | 0.01893 |
| QRS.DURATION | rs55892056 | rs6569024 | A | G | 6 | CEP85L | Female only | True | 0.4454 | 0.4484 | 0.4673 | 0.463 | 7224 | 6316 | 5542 | 5273 | 0.09344 | 0.02168 | 0.05882 | 0.07573 | 0.01703 | 0.0189 |
| QRS.DURATION | rs55892056 | rs11153767 | C | T | 6 | CEP85L | Female only | True | 0.4689 | 0.4638 | 0.4504 | 0.4523 | 7224 | 6316 | 5542 | 5273 | -0.07458 | -0.005044 | -0.04728 | -0.06002 | 0.01687 | 0.01893 |
| QRS.DURATION | rs55892056 | rs6906287 | C | T | 6 | CEP85L | Female only | True | 0.4471 | 0.4501 | 0.4676 | 0.4631 | 7224 | 6316 | 5542 | 5273 | 0.09376 | 0.02148 | 0.05962 | 0.0757 | 0.01697 | 0.01885 |
| QRS.DURATION | rs55892056 | rs6913012 | G | A | 6 | CEP85L | Female only | True | 0.4679 | 0.4638 | 0.4504 | 0.451 | 7224 | 6316 | 5542 | 5273 | -0.07403 | -0.009856 | -0.0456 | -0.0592 | 0.0169 | 0.01892 |
| QRS.DURATION | rs55892056 | rs9320663 | A | G | 6 | CEP85L | Female only | True | 0.4682 | 0.4635 | 0.4513 | 0.4524 | 7224 | 6316 | 5542 | 5273 | -0.07237 | -0.005517 | -0.04649 | -0.05741 | 0.01689 | 0.01887 |
| QRS.DURATION | rs55892056 | rs17427116 | A | G | 6 | CEP85L | Female only | True | 0.4477 | 0.4505 | 0.4671 | 0.463 | 7224 | 6316 | 5542 | 5273 | 0.09177 | 0.02158 | 0.05876 | 0.07214 | 0.01698 | 0.0188 |
| QRS.DURATION | rs55892056 | rs9320665 | G | A | 6 | CEP85L | Female only | True | 0.4682 | 0.4635 | 0.4513 | 0.4523 | 7224 | 6316 | 5542 | 5273 | -0.07237 | -0.005517 | -0.04652 | -0.05742 | 0.01689 | 0.01887 |

| trait | lead_snp | ID | ALLELE1 | ALLELE0 | CHR | Gene_Symbol | independent_loci_in_sex_P1e2 | in_embl_any_or_gene | A1FREQ_VUMC_female | A1FREQ_VUMC_male | A1FREQ_UKB_female | A1FREQ_UKB_male | N_VUMC_female | N_VUMC_male | N_UKB_female | N_UKB_male | BETA_VUMC_female | BETA_VUMC_male | BETA_UKB_female | BETA_UKB_male | SE_VUMC_female | SE_VUMC_male |
|---|---|---|---|---|---|---|---|---|---|---|---|---|---|---|---|---|---|---|---|---|---|---|
| QRS.DURATION | rs55892056 | rs11153768 | T | C | 6 | CEP85L | Female only | True | 0.4465 | 0.4492 | 0.467 | 0.4629 | 7224 | 6316 | 5542 | 5273 | 0.09188 | 0.02162 | 0.05876 | 0.07221 | 0.01702 | 0.01885 |
| QRS.DURATION | rs55892056 | rs10872167 | G | A | 6 | CEP85L | Female only | True | 0.4477 | 0.4505 | 0.467 | 0.463 | 7224 | 6316 | 5542 | 5273 | 0.09142 | 0.02158 | 0.05874 | 0.07211 | 0.01698 | 0.0188 |
| QRS.DURATION | rs55892056 | rs2798321 | G | T | 6 | CEP85L | Female only | True | 0.4696 | 0.4641 | 0.4525 | 0.4532 | 7224 | 6316 | 5542 | 5273 | -0.06946 | -0.005623 | -0.04715 | -0.05663 | 0.01697 | 0.01894 |
| QRS.DURATION | rs55892056 | rs11756438 | A | C | 6 | CEP85L | Female only | True | 0.4648 | 0.4641 | 0.4786 | 0.4794 | 7224 | 6316 | 5542 | 5273 | 0.09277 | 0.009338 | 0.05782 | 0.06729 | 0.01687 | 0.01875 |
| QRS.DURATION | rs55892056 | rs11756440 | A | C | 6 | CEP85L | Female only | True | 0.4634 | 0.4624 | 0.4785 | 0.4793 | 7224 | 6316 | 5542 | 5273 | 0.09269 | 0.009478 | 0.05762 | 0.06721 | 0.01691 | 0.01879 |
| QRS.DURATION | rs55892056 | rs55892056 | T | C | 6 | CEP85L | Female only | True | 0.4406 | 0.442 | 0.46 | 0.4579 | 7224 | 6316 | 5542 | 5273 | 0.09507 | 0.01344 | 0.05726 | 0.07088 | 0.01697 | 0.0188 |
| QRS.DURATION | rs55892056 | rs2638551 | C | T | 6 | CEP85L | Female only | True | 0.4268 | 0.431 | 0.4417 | 0.4438 | 7224 | 6316 | 5542 | 5273 | 0.07252 | -0.009539 | 0.06147 | 0.0517 | 0.01695 | 0.01889 |
| QRS.DURATION | rs55892056 | rs2638552 | T | C | 6 | CEP85L | Female only | True | 0.4258 | 0.4299 | 0.4417 | 0.4438 | 7224 | 6316 | 5542 | 5273 | 0.07278 | -0.009448 | 0.06164 | 0.0517 | 0.01699 | 0.01893 |
| QRS.DURATION | rs55892056 | rs9489497 | C | T | 6 | CEP85L | Female only | True | 0.4258 | 0.4299 | 0.4417 | 0.4439 | 7224 | 6316 | 5542 | 5273 | 0.07278 | -0.009448 | 0.06147 | 0.05155 | 0.01699 | 0.01893 |
| QRS.DURATION | rs55892056 | rs902779 | G | T | 6 | CEP85L | Female only | True | 0.4258 | 0.4299 | 0.4417 | 0.4438 | 7224 | 6316 | 5542 | 5273 | 0.07278 | -0.009448 | 0.06147 | 0.05169 | 0.01699 | 0.01893 |
| QRS.DURATION | rs55892056 | rs1343287 | C | T | 6 | CEP85L | Female only | True | 0.4259 | 0.4301 | 0.4417 | 0.4438 | 7224 | 6316 | 5542 | 5273 | 0.07271 | -0.009266 | 0.0614 | 0.05169 | 0.01698 | 0.01892 |
| QRS.DURATION | rs55892056 | rs2638544 | A | G | 6 | CEP85L | Female only | True | 0.4259 | 0.4301 | 0.4417 | 0.4438 | 7224 | 6316 | 5542 | 5273 | 0.07272 | -0.009268 | 0.0614 | 0.0517 | 0.01698 | 0.01892 |
| QRS.DURATION | rs55892056 | rs1844964 | T | C | 6 | CEP85L | Female only | True | 0.4259 | 0.4301 | 0.4418 | 0.4438 | 7224 | 6316 | 5542 | 5273 | 0.07271 | -0.009266 | 0.06157 | 0.0517 | 0.01698 | 0.01892 |
| QRS.DURATION | rs55892056 | rs17349329 | G | T | 6 | CEP85L | Female only | True | 0.3902 | 0.3937 | 0.4046 | 0.407 | 7224 | 6316 | 5542 | 5273 | 0.07921 | -0.009122 | 0.06718 | 0.05961 | 0.01714 | 0.01911 |
| QRS.DURATION | rs55892056 | rs2798326 | T | C | 6 | CEP85L | Female only | True | 0.4259 | 0.4302 | 0.4417 | 0.4438 | 7224 | 6316 | 5542 | 5273 | 0.07316 | -0.008999 | 0.06141 | 0.05169 | 0.01699 | 0.01893 |
| QRS.DURATION | rs55892056 | rs2798327 | T | C | 6 | CEP85L | Female only | True | 0.4259 | 0.4303 | 0.4418 | 0.4438 | 7224 | 6316 | 5542 | 5273 | 0.07309 | -0.00899 | 0.06141 | 0.05169 | 0.01699 | 0.01893 |
| QRS.DURATION | rs55892056 | rs2798328 | T | C | 6 | CEP85L | Female only | True | 0.4259 | 0.4303 | 0.4418 | 0.4438 | 7224 | 6316 | 5542 | 5273 | 0.0731 | -0.008998 | 0.06141 | 0.05169 | 0.01699 | 0.01893 |
| QRS.DURATION | rs55892056 | rs11754023 | C | T | 6 | CEP85L | Female only | True | 0.3902 | 0.3938 | 0.4046 | 0.4069 | 7224 | 6316 | 5542 | 5273 | 0.07966 | -0.008955 | 0.06727 | 0.05962 | 0.01714 | 0.01911 |
| QRS.DURATION | rs55892056 | rs2638529 | C | T | 6 | CEP85L | Female only | True | 0.4259 | 0.4304 | 0.4419 | 0.444 | 7224 | 6316 | 5542 | 5273 | 0.073 | -0.009559 | 0.06158 | 0.05185 | 0.01699 | 0.01893 |
| QRS.DURATION | rs55892056 | rs6937176 | G | A | 6 | CEP85L | Female only | True | 0.3902 | 0.3939 | 0.4046 | 0.407 | 7224 | 6316 | 5542 | 5273 | 0.07877 | -0.009573 | 0.06742 | 0.0596 | 0.01715 | 0.0191 |

| trait | lead_snp | ID | ALLELE1 | ALLELE0 | CHR | Gene_Symbol | independent_loci_in_sex_P1e2 | in_embl_any_or_gene | A1FREQ_VUMC_female | A1FREQ_VUMC_male | A1FREQ_UKB_female | A1FREQ_UKB_male | N_VUMC_female | N_VUMC_male | N_UKB_female | N_UKB_male | BETA_VUMC_female | BETA_VUMC_male | BETA_UKB_female | BETA_UKB_male | SE_VUMC_female | SE_VUMC_male |
|---|---|---|---|---|---|---|---|---|---|---|---|---|---|---|---|---|---|---|---|---|---|---|
| QRS.DURATION | rs55892056 | rs72962906 | T | C | 6 | CEP85L | Female only | True | 0.3902 | 0.3939 | 0.4046 | 0.407 | 7224 | 6316 | 5542 | 5273 | 0.07877 | -0.009571 | 0.06754 | 0.0596 | 0.01715 | 0.0191 |
| QRS.DURATION | rs55892056 | rs60497831 | A | G | 6 | CEP85L | Female only | True | 0.3892 | 0.3929 | 0.4046 | 0.407 | 7224 | 6316 | 5542 | 5273 | 0.07909 | -0.009472 | 0.06738 | 0.05956 | 0.01719 | 0.01914 |
| QRS.DURATION | rs55892056 | rs56344535 | C | T | 6 | CEP85L | Female only | True | 0.3657 | 0.3711 | 0.3852 | 0.3848 | 7224 | 6316 | 5542 | 5273 | 0.08389 | -0.003491 | 0.06798 | 0.06318 | 0.0174 | 0.01932 |
| QRS.DURATION | rs55892056 | rs72962938 | C | T | 6 | CEP85L | Female only | True | 0.365 | 0.3711 | 0.3852 | 0.3848 | 7224 | 6316 | 5542 | 5273 | 0.08349 | -0.002819 | 0.06789 | 0.06255 | 0.01741 | 0.01931 |
| QRS.DURATION | rs55892056 | rs4945624 | T | G | 6 | CEP85L | Female only | True | 0.364 | 0.37 | 0.385 | 0.3847 | 7224 | 6316 | 5542 | 5273 | 0.08441 | -0.002825 | 0.0673 | 0.06237 | 0.01746 | 0.01937 |
| QRS.DURATION | rs55892056 | rs4027595 | C | T | 6 | CEP85L | Female only | True | 0.3901 | 0.3944 | 0.4049 | 0.4072 | 7224 | 6316 | 5542 | 5273 | 0.07743 | -0.007323 | 0.06706 | 0.05965 | 0.01715 | 0.01906 |
| QRS.DURATION | rs55892056 | 6:119017844 | G | T | 6 | CEP85L | Female only | True | 0.427 | 0.4317 | | | 7224 | 6316 | | | 0.07154 | -0.00824 | | | 0.01695 | 0.01887 |
| QRS.DURATION | rs55892056 | rs57237379 | A | G | 6 | CEP85L | Female only | True | 0.3897 | 0.394 | 0.4046 | 0.4072 | 7224 | 6316 | 5542 | 5273 | 0.07803 | -0.007784 | 0.06736 | 0.05994 | 0.01714 | 0.01907 |
| QRS.DURATION | rs55892056 | rs1771756 | T | C | 6 | CEP85L | Female only | True | 0.4254 | 0.4304 | 0.442 | 0.444 | 7224 | 6316 | 5542 | 5273 | 0.07283 | -0.008432 | 0.06049 | 0.05244 | 0.01697 | 0.0189 |
| QRS.DURATION | rs55892056 | rs4946356 | A | G | 6 | CEP85L | Female only | True | 0.4119 | 0.4172 | 0.4253 | 0.4286 | 7224 | 6316 | 5542 | 5273 | 0.07356 | -0.007734 | 0.06202 | 0.05822 | 0.01703 | 0.0189 |
| QRS.DURATION | rs55892056 | rs2046951 | C | T | 6 | CEP85L | Female only | True | 0.4119 | 0.4171 | 0.4252 | 0.4282 | 7224 | 6316 | 5542 | 5273 | 0.07397 | -0.008136 | 0.06234 | 0.05794 | 0.01705 | 0.0189 |
| QRS.DURATION | rs55892056 | rs9481850 | A | C | 6 | CEP85L | Female only | True | 0.4269 | 0.4321 | 0.442 | 0.4442 | 7224 | 6316 | 5542 | 5273 | 0.0722 | -0.008966 | 0.06036 | 0.05119 | 0.01701 | 0.01891 |
| QRS.DURATION | rs55892056 | rs17825844 | A | G | 6 | CEP85L | Female only | True | 0.3647 | 0.3711 | 0.3846 | 0.3839 | 7224 | 6316 | 5542 | 5273 | 0.08408 | -0.002107 | 0.06697 | 0.05985 | 0.01745 | 0.01932 |
| QRS.DURATION | rs55892056 | rs7746210 | G | A | 6 | CEP85L | Female only | True | 0.3642 | 0.3704 | 0.3847 | 0.3843 | 7224 | 6316 | 5542 | 5273 | 0.08377 | -0.003091 | 0.06683 | 0.05918 | 0.01751 | 0.01938 |
| UricA | rs1048252 | rs10939515 | A | G | 4 | SLC2A9 | Female only | True | 0.1623 | 0.151 | 0.1618 | 0.1621 | 6711 | 6411 | 184755 | 159081 | -0.1188 | -0.007205 | -0.1529 | -0.0681 | 0.02301 | 0.02258 |
| UricA | rs1048252 | rs1401438 | A | G | 4 | SLC2A9 | Female only | True | 0.1682 | 0.1594 | 0.1674 | 0.1677 | 6711 | 6411 | 184755 | 159081 | -0.09776 | -0.004525 | -0.1586 | -0.06041 | 0.02244 | 0.0219 |
| trait | lead_snp | ID | ALLELE1 | ALLELE0 | CHR | Gene_Symbol | independent_loci_in_sex_P1e2 | in_embl_any_or_gene | A1FREQ_VUMC_female | A1FREQ_VUMC_male | A1FREQ_UKB_female | A1FREQ_UKB_male | N_VUMC_female | N_VUMC_male | N_UKB_female | N_UKB_male | BETA_VUMC_female | BETA_VUMC_male | BETA_UKB_female | BETA_UKB_male | SE_VUMC_female | SE_VUMC_male |
| UricA | rs1048252 | rs11735883 | A | G | 4 | SLC2A9 | Female only | True | 0.1679 | 0.1594 | 0.1673 | 0.1676 | 6711 | 6411 | 184755 | 159081 | -0.09882 | -0.004266 | -0.1589 | -0.06059 | 0.02245 | 0.0219 |
| UricA | rs1048252 | rs11722614 | T | C | 4 | SLC2A9 | Female only | True | 0.1665 | 0.158 | 0.1663 | 0.1667 | 6711 | 6411 | 184755 | 159081 | -0.09849 | -0.00138 | -0.1609 | -0.06128 | 0.02258 | 0.02204 |
| UricA | rs7698858 | rs4697892 | A | C | 4 | SLC2A9 | Female only | True | 0.2182 | 0.2082 | 0.2163 | 0.2154 | 6711 | 6411 | 184755 | 159081 | -0.1319 | -0.02826 | -0.2015 | -0.0891 | 0.02066 | 0.01995 |
| UricA | rs7698858 | rs1980220 | A | G | 4 | SLC2A9 | Female only | True | 0.2181 | 0.208 | 0.2159 | 0.2149 | 6711 | 6411 | 184755 | 159081 | -0.1307 | -0.02764 | -0.2017 | -0.08906 | 0.02066 | 0.01994 |
| UricA | rs7698858 | rs57731161 | T | C | 4 | SLC2A9 | Female only | True | 0.2184 | 0.208 | 0.2162 | 0.2152 | 6711 | 6411 | 184755 | 159081 | -0.1302 | -0.02891 | -0.2015 | -0.08895 | 0.02066 | 0.01995 |

| trait | lead_snp | ID | ALLELE1 | ALLELE0 | CHR | Gene_Symbol | independent_loci_in_sex_P1e2 | in_embl_any_or_gene | A1FREQ_VUMC_female | A1FREQ_VUMC_male | A1FREQ_UKB_female | A1FREQ_UKB_male | N_VUMC_female | N_VUMC_male | N_UKB_female | N_UKB_male | BETA_VUMC_female | BETA_VUMC_male | BETA_UKB_female | BETA_UKB_male | SE_VUMC_female | SE_VUMC_male |
|---|---|---|---|---|---|---|---|---|---|---|---|---|---|---|---|---|---|---|---|---|---|---|
| UricA | rs7698858 | rs4621429 | G | A | 4 | SLC2A9 | Female only | True | 0.218 | 0.2078 | 0.2159 | 0.2149 | 6711 | 6411 | 184755 | 159081 | -0.1315 | -0.02822 | -0.2018 | -0.08892 | 0.02067 | 0.01994 |
| UricA | rs7698858 | rs4697895 | C | T | 4 | SLC2A9 | Female only | True | 0.218 | 0.2081 | 0.216 | 0.215 | 6711 | 6411 | 184755 | 159081 | -0.1314 | -0.02824 | -0.2016 | -0.08883 | 0.02065 | 0.01992 |
| UricA | rs7698858 | rs16889260 | T | C | 4 | SLC2A9 | Female only | True | 0.1916 | 0.1807 | 0.1882 | 0.187 | 6711 | 6411 | 184755 | 159081 | -0.146 | -0.03697 | -0.2138 | -0.1006 | 0.02164 | 0.02097 |
| UricA | rs7698858 | rs10939552 | A | C | 4 | SLC2A9 | Female only | True | 0.2187 | 0.2084 | 0.2155 | 0.2145 | 6711 | 6411 | 184755 | 159081 | -0.1275 | -0.02912 | -0.2008 | -0.08877 | 0.02059 | 0.01988 |
| UricA | rs7698858 | rs1976792 | A | G | 4 | SLC2A9 | Female only | True | 0.1663 | 0.1555 | 0.1622 | 0.1617 | 6711 | 6411 | 184755 | 159081 | -0.143 | -0.02819 | -0.2176 | -0.09646 | 0.02271 | 0.02224 |
| UricA | rs7698858 | rs11721988 | G | A | 4 | SLC2A9 | Female only | True | 0.2185 | 0.2081 | 0.2157 | 0.2148 | 6711 | 6411 | 184755 | 159081 | -0.1297 | -0.0283 | -0.2025 | -0.08891 | 0.02058 | 0.01987 |
| UricA | rs7698858 | rs2176644 | T | C | 4 | SLC2A9 | Female only | True | 0.2193 | 0.2089 | 0.2158 | 0.2149 | 6711 | 6411 | 184755 | 159081 | -0.1292 | -0.02826 | -0.2023 | -0.08867 | 0.0205 | 0.0198 |
| UricA | rs7698858 | rs4697898 | T | C | 4 | SLC2A9 | Female only | True | 0.2185 | 0.2081 | 0.2157 | 0.2148 | 6711 | 6411 | 184755 | 159081 | -0.1297 | -0.02828 | -0.2026 | -0.08894 | 0.02058 | 0.01987 |
| UricA | rs7698858 | rs4697899 | A | C | 4 | SLC2A9 | Female only | True | 0.2185 | 0.2081 | 0.2157 | 0.2148 | 6711 | 6411 | 184755 | 159081 | -0.1297 | -0.02828 | -0.2026 | -0.08894 | 0.02058 | 0.01987 |
| UricA | rs7698858 | rs10939567 | A | G | 4 | SLC2A9 | Female only | True | 0.2197 | 0.2086 | 0.216 | 0.215 | 6711 | 6411 | 184755 | 159081 | -0.1299 | -0.02892 | -0.2026 | -0.089 | 0.02058 | 0.01986 |
| UricA | rs7698858 | rs6826806 | G | A | 4 | SLC2A9 | Female only | True | 0.2301 | 0.2175 | 0.223 | 0.2222 | 6711 | 6411 | 184755 | 159081 | -0.1277 | -0.03742 | -0.1987 | -0.08678 | 0.02035 | 0.0196 |
| UricA | rs7698858 | rs4507358 | T | G | 4 | SLC2A9 | Female only | True | 0.23 | 0.2174 | 0.2231 | 0.2224 | 6711 | 6411 | 184755 | 159081 | -0.1275 | -0.03746 | -0.1989 | -0.08682 | 0.02036 | 0.0196 |
| UricA | rs7698858 | rs2001412 | C | T | 4 | SLC2A9 | Female only | True | 0.2201 | 0.209 | 0.2162 | 0.2154 | 6711 | 6411 | 184755 | 159081 | -0.1297 | -0.02643 | -0.2031 | -0.0892 | 0.02056 | 0.01983 |
| UricA | rs7698858 | rs1106058 | A | G | 4 | SLC2A9 | Female only | True | 0.22 | 0.209 | 0.2162 | 0.2154 | 6711 | 6411 | 184755 | 159081 | -0.1305 | -0.02662 | -0.2031 | -0.08918 | 0.02057 | 0.01983 |
| UricA | rs7698858 | rs4697903 | A | G | 4 | SLC2A9 | Female only | True | 0.2291 | 0.2165 | 0.2216 | 0.221 | 6711 | 6411 | 184755 | 159081 | -0.1312 | -0.03736 | -0.1979 | -0.08648 | 0.02034 | 0.01958 |
| UricA | rs7698858 | rs2139243 | G | A | 4 | SLC2A9 | Female only | True | 0.2202 | 0.2092 | 0.2161 | 0.2153 | 6711 | 6411 | 184755 | 159081 | -0.1293 | -0.0282 | -0.203 | -0.08918 | 0.02057 | 0.01983 |
| UricA | rs7698858 | rs57235613 | T | C | 4 | SLC2A9 | Female only | True | 0.1669 | 0.1559 | 0.1624 | 0.1619 | 6711 | 6411 | 184755 | 159081 | -0.1432 | -0.02883 | -0.2177 | -0.09661 | 0.02265 | 0.0222 |
| UricA | rs7698858 | rs883041 | T | C | 4 | SLC2A9 | Female only | True | 0.2301 | 0.2177 | 0.2231 | 0.2225 | 6711 | 6411 | 184755 | 159081 | -0.1303 | -0.03775 | -0.1991 | -0.08689 | 0.02036 | 0.01958 |
| UricA | rs1048252 | rs1048252 | T | C | 4 | SLC2A9 | Female only | True | 0.1727 | 0.1616 | 0.168 | 0.1675 | 6711 | 6411 | 184755 | 159081 | -0.1518 | -0.02796 | -0.2087 | -0.094 | 0.02241 | 0.02186 |
| UricA | rs7698858 | rs1568318 | C | T | 4 | SLC2A9 | Female only | True | 0.1992 | 0.1871 | 0.1943 | 0.193 | 6711 | 6411 | 184755 | 159081 | -0.1536 | -0.03269 | -0.2081 | -0.09897 | 0.02135 | 0.02063 |
| UricA | rs7698858 | rs2867394 | A | G | 4 | SLC2A9 | Female only | True | 0.1931 | 0.1816 | 0.1884 | 0.1874 | 6711 | 6411 | 184755 | 159081 | -0.1438 | -0.03559 | -0.2152 | -0.1011 | 0.02152 | 0.02086 |
| UricA | rs7698858 | rs6840999 | A | G | 4 | SLC2A9 | Female only | True | 0.1933 | 0.1815 | 0.1886 | 0.1876 | 6711 | 6411 | 184755 | 159081 | -0.146 | -0.03287 | -0.2157 | -0.1014 | 0.02156 | 0.02091 |
| UricA | rs7698858 | rs6839302 | T | C | 4 | SLC2A9 | Female only | True | 0.2096 | 0.1966 | 0.2013 | 0.2004 | 6711 | 6411 | 184755 | 159081 | -0.1512 | -0.04311 | -0.2036 | -0.09613 | 0.02107 | 0.02032 |
| UricA | rs7698858 | rs9998156 | G | A | 4 | SLC2A9 | Female only | True | 0.2096 | 0.1963 | 0.2013 | 0.2004 | 6711 | 6411 | 184755 | 159081 | -0.1528 | -0.04438 | -0.2041 | -0.09633 | 0.02111 | 0.02036 |
| UricA | rs7698858 | rs10939599 | A | C | 4 | SLC2A9 | Female only | True | 0.1993 | 0.1873 | 0.1942 | 0.1931 | 6711 | 6411 | 184755 | 159081 | -0.1544 | -0.03237 | -0.209 | -0.09922 | 0.02137 | 0.02066 |
| UricA | rs7698858 | rs6818572 | A | G | 4 | SLC2A9 | Female only | True | 0.1993 | 0.1868 | 0.194 | 0.1929 | 6711 | 6411 | 184755 | 159081 | -0.1542 | -0.03314 | -0.2103 | -0.09987 | 0.02141 | 0.02071 |
| UricA | rs7698858 | rs1107710 | G | A | 4 | SLC2A9 | Female only | True | 0.1996 | 0.187 | 0.1942 | 0.1931 | 6711 | 6411 | 184755 | 159081 | -0.1542 | -0.033 | -0.211 | -0.1 | 0.0214 | 0.02071 |
| UricA | rs7698858 | rs938562 | T | C | 4 | SLC2A9 | Female only | True | 0.1995 | 0.1869 | 0.1941 | 0.193 | 6711 | 6411 | 184755 | 159081 | -0.155 | -0.03309 | -0.211 | -0.1001 | 0.02141 | 0.02072 |
| UricA | rs7698858 | rs12644047 | A | G | 4 | SLC2A9 | Female only | True | 0.1942 | 0.1814 | 0.1886 | 0.1877 | 6711 | 6411 | 184755 | 159081 | -0.1487 | -0.04015 | -0.219 | -0.1031 | 0.02163 | 0.02101 |

| trait | lead_snp | ID | ALLELE1 | ALLELE0 | CHR | Gene_Symbol | independent_loci_in_sex_P1e2 | in_embl_any_or_gene | A1FREQ_VUMC_female | A1FREQ_VUMC_male | A1FREQ_UKB_female | A1FREQ_UKB_male | N_VUMC_female | N_VUMC_male | N_UKB_female | N_UKB_male | BETA_VUMC_female | BETA_VUMC_male | BETA_UKB_female | BETA_UKB_male | SE_VUMC_female | SE_VUMC_male |
|---|---|---|---|---|---|---|---|---|---|---|---|---|---|---|---|---|---|---|---|---|---|---|
| UricA | rs7698858 | rs7698858 | C | T | 4 | SLC2A9 | Female only | True | 0.2491 | 0.2384 | 0.2492 | 0.2479 | 6711 | 6411 | 184755 | 159081 | -0.1517 | -0.03951 | -0.1958 | -0.08814 | 0.01963 | 0.01892 |
| LYMPM | rs112247182 | rs146359295 | T | | 23 | RP11-585K14.2 | Female only | False | 0.05201 | 0.05319 | 0.05462 | 0.05478 | 5362 | 5485 | 187760 | 162101 | -0.1262 | 0.02975 | -0.004585 | 9.62e-04 | 0.04174 | 0.02848 |
| LYMPM | rs112247182 | rs16982266 | T | | 23 | RP11-585K14.2 | Female only | False | 0.05247 | 0.05317 | 0.05461 | 0.05474 | 5362 | 5485 | 187760 | 162101 | -0.1092 | 0.03031 | -0.004156 | 4.54e-04 | 0.04152 | 0.02846 |
| LYMPM | rs112247182 | rs5961593 | A | | 23 | RP11-585K14.2 | Female only | False | 0.05247 | 0.05317 | 0.0547 | 0.05473 | 5362 | 5485 | 187760 | 162101 | -0.1092 | 0.03034 | -0.00409 | 4.95e-04 | 0.04152 | 0.02845 |
| LYMPM | rs112247182 | rs5961594 | T | | 23 | RP11-585K14.2 | Female only | False | 0.05217 | 0.05314 | 0.05461 | 0.05464 | 5362 | 5485 | 187760 | 162101 | -0.1092 | 0.03032 | -0.003442 | 8.41e-04 | 0.04164 | 0.02845 |
| LYMPM | rs112247182 | rs149288444 | C | | 23 | RP11-585K14.2 | Female only | False | 0.0522 | 0.05315 | 0.05457 | 0.05468 | 5362 | 5485 | 187760 | 162101 | -0.1099 | 0.03027 | -0.004142 | 7.35e-04 | 0.04163 | 0.02845 |
| LYMPM | rs112247182 | rs5961595 | T | | 23 | RP11-585K14.2 | Female only | False | 0.05238 | 0.05316 | 0.05473 | 0.05467 | 5362 | 5485 | 187760 | 162101 | -0.1101 | 0.03 | -0.003372 | 8.10e-04 | 0.0416 | 0.02842 |
| LYMPM | rs112247182 | rs116548335 | T | | 23 | RP11-585K14.2 | Female only | False | 0.05599 | 0.05773 | 0.05852 | 0.05856 | 5362 | 5485 | 187760 | 162101 | -0.1434 | 0.01571 | -0.003104 | 2.88e-04 | 0.04028 | 0.02735 |
| LYMPM | rs112247182 | rs17304601 | C | | 23 | RP11-585K14.2 | Female only | False | 0.05588 | 0.05771 | 0.05851 | 0.05853 | 5362 | 5485 | 187760 | 162101 | -0.1449 | 0.01582 | -0.002213 | 3.01e-04 | 0.04033 | 0.02736 |
| LYMPM | rs112247182 | rs144757126 | G | | 23 | RP11-585K14.2 | Female only | False | 0.05561 | 0.05787 | 0.05851 | 0.05849 | 5362 | 5485 | 187760 | 162101 | -0.1428 | 0.01576 | -0.002281 | 6.08e-04 | 0.04042 | 0.02732 |
| LYMPM | rs112247182 | rs138842878 | C | | 23 | RP11-585K14.2 | Female only | False | 0.05561 | 0.05787 | 0.05851 | 0.05849 | 5362 | 5485 | 187760 | 162101 | -0.1428 | 0.01576 | -0.002281 | 6.09e-04 | 0.04042 | 0.02732 |
| LYMPM | rs112247182 | rs115647281 | G | | 23 | RP11-585K14.2 | Female only | False | 0.05563 | 0.0579 | 0.05843 | 0.05859 | 5362 | 5485 | 187760 | 162101 | -0.1427 | 0.01568 | -0.00327 | 5.95e-04 | 0.0404 | 0.02731 |
| LYMPM | rs112247182 | rs17304615 | A | | 23 | RP11-585K14.2 | Female only | False | 0.05563 | 0.0579 | 0.05843 | 0.0586 | 5362 | 5485 | 187760 | 162101 | -0.1427 | 0.01568 | -0.003253 | 6.11e-04 | 0.0404 | 0.0273 |
| LYMPM | rs112247182 | rs17219546 | A | | 23 | RP11-585K14.2 | Female only | False | 0.05563 | 0.0579 | 0.05849 | 0.05859 | 5362 | 5485 | 187760 | 162101 | -0.1427 | 0.01573 | -0.003197 | 6.01e-04 | 0.0404 | 0.02731 |
| LYMPM | rs112247182 | rs55884691 | T | | 23 | RP11-585K14.2 | Female only | False | 0.05563 | 0.05789 | 0.05847 | 0.05859 | 5362 | 5485 | 187760 | 162101 | -0.1427 | 0.01572 | -0.00328 | 6.30e-04 | 0.0404 | 0.02731 |
| LYMPM | rs112247182 | rs58056218 | T | | 23 | RP11-585K14.2 | Female only | False | 0.05562 | 0.05802 | 0.05843 | 0.05861 | 5362 | 5485 | 187760 | 162101 | -0.1401 | 0.0157 | -0.003092 | 6.67e-04 | 0.04033 | 0.02726 |
| LYMPM | rs112247182 | rs55782540 | T | | 23 | RP11-585K14.2 | Female only | False | 0.05563 | 0.05802 | 0.05847 | 0.05864 | 5362 | 5485 | 187760 | 162101 | -0.1404 | 0.0157 | -0.003026 | 5.90e-04 | 0.04032 | 0.02726 |
| LYMPM | rs112247182 | rs17219553 | G | | 23 | RP11-585K14.2 | Female only | False | 0.0559 | 0.05801 | 0.05855 | 0.05866 | 5362 | 5485 | 187760 | 162101 | -0.1406 | 0.01569 | -0.002889 | 4.58e-04 | 0.04025 | 0.02726 |
| LYMPM | rs112247182 | rs142422482 | G | | 23 | RP11-585K14.2 | Female only | False | 0.0559 | 0.05802 | 0.05849 | 0.05866 | 5362 | 5485 | 187760 | 162101 | -0.1406 | 0.01567 | -0.002929 | 4.54e-04 | 0.04024 | 0.02726 |
| LYMPM | rs112247182 | rs144330635 | G | | 23 | RP11-585K14.2 | Female only | False | 0.0559 | 0.05802 | 0.05849 | 0.05866 | 5362 | 5485 | 187760 | 162101 | -0.1406 | 0.01567 | -0.002932 | 4.36e-04 | 0.04024 | 0.02726 |
| LYMPM | rs112247182 | rs116089980 | A | | 23 | RP11-585K14.2 | Female only | False | 0.0559 | 0.05802 | 0.0585 | 0.05866 | 5362 | 5485 | 187760 | 162101 | -0.1405 | 0.01567 | -0.002966 | 4.41e-04 | 0.04024 | 0.02726 |
| LYMPM | rs112247182 | rs5961597 | A | | 23 | RP11-585K14.2 | Female only | False | 0.05707 | 0.05838 | 0.05875 | 0.05883 | 5362 | 5485 | 187760 | 162101 | -0.1219 | 0.01616 | -0.002807 | 1.27e-04 | 0.03986 | 0.02718 |
| LYMPM | rs112247182 | rs56114849 | G | | 23 | RP11-585K14.2 | Female only | False | 0.05586 | 0.058 | 0.05847 | 0.05862 | 5362 | 5485 | 187760 | 162101 | -0.1401 | 0.01568 | -0.00293 | 5.53e-04 | 0.04027 | 0.02727 |
| LYMPM | rs112247182 | rs55670546 | A | | 23 | RP11-585K14.2 | Female only | False | 0.05605 | 0.05819 | 0.05862 | 0.05877 | 5362 | 5485 | 187760 | 162101 | -0.1403 | 0.01589 | -0.002465 | 2.73e-04 | 0.04024 | 0.02724 |
| LYMPM | rs112247182 | rs58371007 | A | | 23 | RP11-585K14.2 | Female only | False | 0.05588 | 0.05799 | 0.05848 | 0.05863 | 5362 | 5485 | 187760 | 162101 | -0.1406 | 0.01564 | -0.002941 | 4.13e-04 | 0.04026 | 0.02727 |
| LYMPM | rs112247182 | rs115472624 | A | | 23 | RP11-585K14.2 | Female only | False | 0.0559 | 0.05802 | 0.05868 | 0.05866 | 5362 | 5485 | 187760 | 162101 | -0.1407 | 0.01568 | -0.00205 | 4.42e-04 | 0.04024 | 0.02726 |
| trait | lead_snp | ID | ALLELE1 | ALLELE0 | CHR | Gene_Symbol | independent_loci_in_sex_P1e2 | in_embl_any_or_gene | A1FREQ_VUMC_female | A1FREQ_VUMC_male | A1FREQ_UKB_female | A1FREQ_UKB_male | N_VUMC_female | N_VUMC_male | N_UKB_female | N_UKB_male | BETA_VUMC_female | BETA_VUMC_male | BETA_UKB_female | BETA_UKB_male | SE_VUMC_female | SE_VUMC_male |
| LYMPM | rs112247182 | rs114290791 | C | | 23 | RP11-585K14.2 | Female only | False | 0.0559 | 0.05802 | 0.0586 | 0.05867 | 5362 | 5485 | 187760 | 162101 | -0.1407 | 0.01568 | -0.002806 | 4.57e-04 | 0.04024 | 0.02726 |

| trait | lead_snp | ID | ALLELE1 | ALLELE0 | CHR | Gene_Symbol | independent_loci_in_sex_P1e2 | in_embl_any_or_gene | A1FREQ_VUMC_female | A1FREQ_VUMC_male | A1FREQ_UKB_female | A1FREQ_UKB_male | N_VUMC_female | N_VUMC_male | N_UKB_female | N_UKB_male | BETA_VUMC_female | BETA_VUMC_male | BETA_UKB_female | BETA_UKB_male | SE_VUMC_female | SE_VUMC_male |
|---|---|---|---|---|---|---|---|---|---|---|---|---|---|---|---|---|---|---|---|---|---|---|
| LYMPM | rs112247182 | rs116805972 | A | | 23 | RP11-585K14.2 | Female only | False | 0.05591 | 0.05803 | 0.05869 | 0.05867 | 5362 | 5485 | 187760 | 162101 | -0.1408 | 0.01567 | -0.002112 | 4.55e-04 | 0.04024 | 0.02726 |
| LYMPM | rs112247182 | rs116567335 | T | | 23 | RP11-585K14.2 | Female only | False | 0.05591 | 0.05802 | 0.05868 | 0.05867 | 5362 | 5485 | 187760 | 162101 | -0.1406 | 0.01569 | -0.002116 | 4.60e-04 | 0.04023 | 0.02726 |
| LYMPM | rs112247182 | rs1562605 | C | | 23 | RP11-585K14.2 | Female only | False | 0.0559 | 0.05802 | 0.05858 | 0.05865 | 5362 | 5485 | 187760 | 162101 | -0.1407 | 0.01571 | -0.002928 | 4.47e-04 | 0.04024 | 0.02726 |
| LYMPM | rs112247182 | rs7058294 | G | | 23 | RP11-585K14.2 | Female only | False | 0.0559 | 0.05802 | 0.05861 | 0.05866 | 5362 | 5485 | 187760 | 162101 | -0.1406 | 0.01568 | -0.002649 | 4.38e-04 | 0.04024 | 0.02726 |
| LYMPM | rs112247182 | rs56955060 | T | | 23 | RP11-585K14.2 | Female only | False | 0.0559 | 0.05802 | 0.05853 | 0.05866 | 5362 | 5485 | 187760 | 162101 | -0.1407 | 0.01567 | -0.00276 | 4.49e-04 | 0.04024 | 0.02726 |
| LYMPM | rs112247182 | rs17304657 | G | | 23 | RP11-585K14.2 | Female only | False | 0.05587 | 0.05802 | 0.05873 | 0.05865 | 5362 | 5485 | 187760 | 162101 | -0.1401 | 0.0157 | -0.001864 | 5.47e-04 | 0.04026 | 0.02726 |
| LYMPM | rs112247182 | rs148775317 | A | | 23 | RP11-585K14.2 | Female only | False | 0.05586 | 0.05801 | 0.05874 | 0.05864 | 5362 | 5485 | 187760 | 162101 | -0.1402 | 0.01569 | -0.001908 | 5.09e-04 | 0.04027 | 0.02726 |
| LYMPM | rs112247182 | rs116162447 | G | | 23 | RP11-585K14.2 | Female only | False | 0.05589 | 0.05802 | 0.05876 | 0.05865 | 5362 | 5485 | 187760 | 162101 | -0.1409 | 0.01571 | -0.001847 | 5.51e-04 | 0.04024 | 0.02726 |
| LYMPM | rs112247182 | rs115944192 | G | | 23 | RP11-585K14.2 | Female only | False | 0.05589 | 0.05803 | 0.05978 | 0.05875 | 5362 | 5485 | 187760 | 162101 | -0.1409 | 0.01571 | -0.001248 | 3.61e-04 | 0.04024 | 0.02726 |
| LYMPM | rs112247182 | rs141666026 | A | | 23 | RP11-585K14.2 | Female only | False | 0.0559 | 0.05803 | 0.05878 | 0.05867 | 5362 | 5485 | 187760 | 162101 | -0.1408 | 0.01571 | -0.001891 | 4.86e-04 | 0.04024 | 0.02726 |
| LYMPM | rs112247182 | rs7066705 | G | | 23 | RP11-585K14.2 | Female only | False | 0.05589 | 0.05802 | 0.05878 | 0.05866 | 5362 | 5485 | 187760 | 162101 | -0.1408 | 0.01571 | -0.001896 | 4.74e-04 | 0.04024 | 0.02726 |
| LYMPM | rs112247182 | rs7067127 | T | | 23 | RP11-585K14.2 | Female only | False | 0.05589 | 0.05802 | 0.05875 | 0.05864 | 5362 | 5485 | 187760 | 162101 | -0.1409 | 0.0157 | -0.001898 | 5.60e-04 | 0.04024 | 0.02726 |
| LYMPM | rs112247182 | rs116259316 | C | | 23 | RP11-585K14.2 | Female only | False | 0.05589 | 0.05802 | 0.05874 | 0.05864 | 5362 | 5485 | 187760 | 162101 | -0.1409 | 0.01568 | -0.00198 | 5.72e-04 | 0.04025 | 0.02726 |
| LYMPM | rs112247182 | rs115552759 | T | | 23 | RP11-585K14.2 | Female only | False | 0.05594 | 0.05803 | 0.05876 | 0.05866 | 5362 | 5485 | 187760 | 162101 | -0.1433 | 0.01572 | -0.001907 | 6.16e-04 | 0.04024 | 0.02726 |
| LYMPM | rs112247182 | rs1106873 | C | | 23 | RP11-585K14.2 | Female only | False | 0.05604 | 0.05816 | 0.05877 | 0.05867 | 5362 | 5485 | 187760 | 162101 | -0.1404 | 0.01562 | -0.001904 | 5.43e-04 | 0.04014 | 0.0272 |
| LYMPM | rs112247182 | rs56404144 | G | | 23 | RP11-585K14.2 | Female only | False | 0.05461 | 0.0562 | 0.05896 | 0.05902 | 5362 | 5485 | 187760 | 162101 | -0.1657 | 0.02055 | -0.001993 | 9.23e-04 | 0.04068 | 0.02769 |
| LYMPM | rs112247182 | rs10521587 | T | | 23 | RP11-585K14.2 | Female only | False | 0.05469 | 0.05618 | 0.05899 | 0.05901 | 5362 | 5485 | 187760 | 162101 | -0.1662 | 0.02058 | -0.0018 | 0.001249 | 0.04064 | 0.0277 |
| LYMPM | rs112247182 | rs55649140 | C | | 23 | RP11-585K14.2 | Female only | False | 0.05357 | 0.05469 | 0.05808 | 0.05825 | 5362 | 5485 | 187760 | 162101 | -0.166 | 0.02855 | -0.001637 | 0.002353 | 0.04112 | 0.02808 |
| LYMPM | rs112247182 | rs56221421 | G | | 23 | RP11-585K14.2 | Female only | False | 0.05357 | 0.05469 | 0.0581 | 0.05826 | 5362 | 5485 | 187760 | 162101 | -0.166 | 0.02856 | -0.001643 | 0.002321 | 0.04112 | 0.02807 |
| LYMPM | rs112247182 | rs137977889 | G | | 23 | RP11-585K14.2 | Female only | False | 0.05382 | 0.0549 | 0.05809 | 0.05828 | 5362 | 5485 | 187760 | 162101 | -0.1663 | 0.02665 | -0.00152 | 0.002445 | 0.04102 | 0.02802 |
| LYMPM | rs112247182 | rs17304720 | C | | 23 | RP11-585K14.2 | Female only | False | 0.05304 | 0.05354 | 0.05736 | 0.05759 | 5362 | 5485 | 187760 | 162101 | -0.1574 | 0.0296 | -0.001962 | 0.003005 | 0.04137 | 0.02847 |
| LYMPM | rs112247182 | rs7063372 | A | | 23 | RP11-585K14.2 | Female only | False | 0.07068 | 0.0714 | 0.0744 | 0.0748 | 5362 | 5485 | 187760 | 162101 | -0.1966 | 0.001185 | 0.002029 | 9.21e-04 | 0.03628 | 0.02485 |
| LYMPM | rs112247182 | rs7061873 | A | | 23 | RP11-585K14.2 | Female only | False | 0.07082 | 0.07147 | 0.0745 | 0.07488 | 5362 | 5485 | 187760 | 162101 | -0.1972 | 9.10e-04 | 0.002255 | 0.001333 | 0.03623 | 0.02483 |
| LYMPM | rs112247182 | rs73436536 | T | | 23 | RP11-585K14.2 | Female only | False | 0.07091 | 0.07148 | 0.07455 | 0.07495 | 5362 | 5485 | 187760 | 162101 | -0.1978 | 1.78e-04 | 0.002404 | 0.001054 | 0.03607 | 0.02477 |
| LYMPM | rs112247182 | rs73436601 | T | | 23 | RP11-585K14.2 | Female only | False | 0.07382 | 0.07481 | 0.0768 | 0.07725 | 5362 | 5485 | 187760 | 162101 | -0.1996 | -4.78e-05 | 0.001045 | 4.57e-04 | 0.03546 | 0.02425 |
| LYMPM | rs112247182 | rs996058 | T | | 23 | RP11-585K14.2 | Female only | False | 0.0742 | 0.07487 | 0.07677 | 0.07726 | 5362 | 5485 | 187760 | 162101 | -0.1982 | -2.51e-04 | 0.001543 | 2.46e-04 | 0.03541 | 0.02424 |
| LYMPM | rs112247182 | rs113020723 | C | | 23 | RP11-585K14.2 | Female only | False | 0.07402 | 0.07487 | 0.07674 | 0.07721 | 5362 | 5485 | 187760 | 162101 | -0.1968 | -2.53e-04 | 0.001381 | 3.23e-04 | 0.03545 | 0.02424 |
| LYMPM | rs112247182 | rs7052521 | A | | 23 | RP11-585K14.2 | Female only | False | 0.07414 | 0.07521 | 0.07684 | 0.07742 | 5362 | 5485 | 187760 | 162101 | -0.1953 | 6.14e-04 | 9.11e-04 | 4.12e-04 | 0.03542 | 0.02419 |
| LYMPM | rs112247182 | rs55798514 | A | | 23 | RP11-585K14.2 | Female only | False | 0.07429 | 0.07542 | 0.07687 | 0.0775 | 5362 | 5485 | 187760 | 162101 | -0.1957 | 0.001437 | 0.001259 | 3.96e-04 | 0.03538 | 0.02415 |

| trait | lead_snp | ID | ALLELE1 | ALLELE0 | CHR | Gene_Symbol | independent_loci_in_sex_P1e2 | in_embl_any_or_gene | A1FREQ_VUMC_female | A1FREQ_VUMC_male | A1FREQ_UKB_female | A1FREQ_UKB_male | N_VUMC_female | N_VUMC_male | N_UKB_female | N_UKB_male | BETA_VUMC_female | BETA_VUMC_male | BETA_UKB_female | BETA_UKB_male | SE_VUMC_female | SE_VUMC_male |
|---|---|---|---|---|---|---|---|---|---|---|---|---|---|---|---|---|---|---|---|---|---|---|
| LYMPM | rs112247182 | rs16997018 | C | | 23 | RP11-585K14.2 | Female only | False | 0.07432 | 0.07501 | 0.07657 | 0.07724 | 5362 | 5485 | 187760 | 162101 | -0.1926 | -0.002482 | 4.82e-04 | 0.001123 | 0.03528 | 0.02418 |
| LYMPM | rs112247182 | rs73448818 | C | | 23 | RP11-585K14.2 | Female only | False | 0.0742 | 0.07499 | 0.07662 | 0.07732 | 5362 | 5485 | 187760 | 162101 | -0.195 | -0.001533 | 7.16e-04 | 0.001096 | 0.03537 | 0.02422 |
| LYMPM | rs112247182 | rs7878812 | C | | 23 | RP11-585K14.2 | Female only | False | 0.0741 | 0.07495 | 0.07672 | 0.07742 | 5362 | 5485 | 187760 | 162101 | -0.1949 | -0.00139 | 0.001082 | 9.95e-04 | 0.03541 | 0.02422 |
| LYMPM | rs112247182 | rs7065808 | T | | 23 | RP11-585K14.2 | Female only | False | 0.07406 | 0.07509 | 0.07708 | 0.07726 | 5362 | 5485 | 187760 | 162101 | -0.1979 | -0.001681 | 0.002289 | 0.001402 | 0.0354 | 0.0242 |
| LYMPM | rs112247182 | rs112977121 | A | | 23 | RP11-585K14.2 | Female only | False | 0.07406 | 0.07509 | 0.07663 | 0.07723 | 5362 | 5485 | 187760 | 162101 | -0.1979 | -0.001681 | 0.001552 | 0.00122 | 0.0354 | 0.0242 |
| LYMPM | rs112247182 | rs1462648 | A | | 23 | RP11-585K14.2 | Female only | False | 0.07441 | 0.07548 | 0.07669 | 0.07726 | 5362 | 5485 | 187760 | 162101 | -0.1964 | -0.001493 | 0.001622 | 0.001216 | 0.03527 | 0.0241 |
| LYMPM | rs112247182 | rs73432891 | C | | 23 | RP11-585K14.2 | Female only | False | 0.0742 | 0.07531 | 0.0767 | 0.07727 | 5362 | 5485 | 187760 | 162101 | -0.1972 | -0.001643 | 0.001629 | 0.001181 | 0.03537 | 0.02417 |
| LYMPM | rs112247182 | rs111847052 | G | | 23 | RP11-585K14.2 | Female only | False | 0.0742 | 0.07531 | 0.07668 | 0.07726 | 5362 | 5485 | 187760 | 162101 | -0.1972 | -0.001643 | 0.001624 | 0.001218 | 0.03537 | 0.02417 |
| LYMPM | rs112247182 | rs73434716 | T | | 23 | RP11-585K14.2 | Female only | False | 0.07426 | 0.07548 | 0.0767 | 0.07728 | 5362 | 5485 | 187760 | 162101 | -0.1966 | -0.00209 | 0.001586 | 0.001131 | 0.03547 | 0.02423 |
| LYMPM | rs112247182 | rs35327795 | T | | 23 | RP11-585K14.2 | Female only | False | 0.07424 | 0.07547 | 0.07668 | 0.07726 | 5362 | 5485 | 187760 | 162101 | -0.1964 | -0.002098 | 0.00153 | 0.001185 | 0.03548 | 0.02423 |
| LYMPM | rs112247182 | rs112999455 | G | | 23 | RP11-585K14.2 | Female only | False | 0.07426 | 0.07548 | 0.07669 | 0.07727 | 5362 | 5485 | 187760 | 162101 | -0.1966 | -0.00209 | 0.001608 | 0.001161 | 0.03548 | 0.02423 |
| LYMPM | rs112247182 | rs73434740 | A | | 23 | RP11-585K14.2 | Female only | False | 0.06785 | 0.07004 | 0.07109 | 0.07165 | 5362 | 5485 | 187760 | 162101 | -0.2134 | -0.006436 | -0.001078 | 0.001481 | 0.03732 | 0.02525 |
| LYMPM | rs112247182 | rs73434798 | C | | 23 | RP11-585K14.2 | Female only | False | 0.06778 | 0.07003 | 0.0711 | 0.07165 | 5362 | 5485 | 187760 | 162101 | -0.2133 | -0.006452 | -0.001101 | 0.0014 | 0.03735 | 0.02525 |
| LYMPM | rs112247182 | rs113216425 | C | | 23 | RP11-585K14.2 | Female only | False | 0.06778 | 0.07003 | 0.07109 | 0.07165 | 5362 | 5485 | 187760 | 162101 | -0.2133 | -0.006452 | -0.001067 | 0.001438 | 0.03735 | 0.02525 |
| LYMPM | rs112247182 | rs112247182 | A | | 23 | RP11-585K14.2 | Female only | False | 0.0678 | 0.06993 | 0.07108 | 0.07159 | 5362 | 5485 | 187760 | 162101 | -0.2138 | -0.006839 | -0.001111 | 0.001522 | 0.03732 | 0.02527 |
| NEUTM | rs73432891 | rs116548335 | T | | 23 | MTND6P12 | Female only | False | 0.05658 | 0.05804 | 0.05852 | 0.05856 | 5147 | 5352 | 187760 | 162101 | 0.1231 | -0.01804 | 3.53e-04 | -0.001416 | 0.04028 | 0.02733 |
| NEUTM | rs73432891 | rs17304601 | C | | 23 | MTND6P12 | Female only | False | 0.05646 | 0.05803 | 0.05851 | 0.05853 | 5147 | 5352 | 187760 | 162101 | 0.1244 | -0.01815 | -5.32e-04 | -0.001412 | 0.04033 | 0.02734 |
| NEUTM | rs73432891 | rs144757126 | G | | 23 | MTND6P12 | Female only | False | 0.05619 | 0.05819 | 0.05851 | 0.05849 | 5147 | 5352 | 187760 | 162101 | 0.1225 | -0.01829 | -1.72e-04 | -0.001767 | 0.04041 | 0.0273 |
| NEUTM | rs73432891 | rs138842878 | C | | 23 | MTND6P12 | Female only | False | 0.05619 | 0.05819 | 0.05851 | 0.05849 | 5147 | 5352 | 187760 | 162101 | 0.1225 | -0.01829 | -1.71e-04 | -0.001767 | 0.04041 | 0.0273 |
| NEUTM | rs73432891 | rs115647281 | G | | 23 | MTND6P12 | Female only | False | 0.05621 | 0.05822 | 0.05843 | 0.05859 | 5147 | 5352 | 187760 | 162101 | 0.1226 | -0.01818 | 7.93e-04 | -0.001722 | 0.0404 | 0.02729 |
| NEUTM | rs73432891 | rs17304615 | A | | 23 | MTND6P12 | Female only | False | 0.05621 | 0.05822 | 0.05843 | 0.0586 | 5147 | 5352 | 187760 | 162101 | 0.1226 | -0.01819 | 7.94e-04 | -0.001744 | 0.0404 | 0.02728 |
| NEUTM | rs73432891 | rs17219546 | A | | 23 | MTND6P12 | Female only | False | 0.05621 | 0.05822 | 0.05849 | 0.05859 | 5147 | 5352 | 187760 | 162101 | 0.1226 | -0.01821 | 7.34e-04 | -0.00173 | 0.0404 | 0.02729 |
| NEUTM | rs73432891 | rs55884691 | T | | 23 | MTND6P12 | Female only | False | 0.05621 | 0.05821 | 0.05847 | 0.05859 | 5147 | 5352 | 187760 | 162101 | 0.1226 | -0.01821 | 7.82e-04 | -0.001765 | 0.0404 | 0.02729 |
| NEUTM | rs73432891 | rs58056218 | T | | 23 | MTND6P12 | Female only | False | 0.0562 | 0.05834 | 0.05843 | 0.05861 | 5147 | 5352 | 187760 | 162101 | 0.1213 | -0.01837 | 6.84e-04 | -0.001825 | 0.04033 | 0.02724 |
| NEUTM | rs73432891 | rs55782540 | T | | 23 | MTND6P12 | Female only | False | 0.05621 | 0.05834 | 0.05847 | 0.05864 | 5147 | 5352 | 187760 | 162101 | 0.1216 | -0.01837 | 6.31e-04 | -0.001757 | 0.04032 | 0.02724 |
| NEUTM | rs73432891 | rs17219553 | G | | 23 | MTND6P12 | Female only | False | 0.05649 | 0.05833 | 0.05855 | 0.05866 | 5147 | 5352 | 187760 | 162101 | 0.1231 | -0.01836 | 3.54e-04 | -0.001605 | 0.04024 | 0.02724 |
| NEUTM | rs73432891 | rs142422482 | G | | 23 | MTND6P12 | Female only | False | 0.05649 | 0.05834 | 0.05849 | 0.05866 | 5147 | 5352 | 187760 | 162101 | 0.1232 | -0.01835 | 4.10e-04 | -0.001596 | 0.04023 | 0.02724 |
| NEUTM | rs73432891 | rs144330635 | G | | 23 | MTND6P12 | Female only | False | 0.05649 | 0.05834 | 0.05849 | 0.05866 | 5147 | 5352 | 187760 | 162101 | 0.1232 | -0.01835 | 4.10e-04 | -0.001572 | 0.04023 | 0.02724 |
| NEUTM | rs73432891 | rs116089980 | A | | 23 | MTND6P12 | Female only | False | 0.05649 | 0.05834 | 0.0585 | 0.05866 | 5147 | 5352 | 187760 | 162101 | 0.1231 | -0.01836 | 4.49e-04 | -0.001576 | 0.04023 | 0.02724 |

| trait | lead_snp | ID | ALLELE1 | ALLELE0 | CHR | Gene_Symbol | independent_loci_in_sex_P1e2 | in_embl_any_or_gene | A1FREQ_VUMC_female | A1FREQ_VUMC_male | A1FREQ_UKB_female | A1FREQ_UKB_male | N_VUMC_female | N_VUMC_male | N_UKB_female | N_UKB_male | BETA_VUMC_female | BETA_VUMC_male | BETA_UKB_female | BETA_UKB_male | SE_VUMC_female | SE_VUMC_male |
|---|---|---|---|---|---|---|---|---|---|---|---|---|---|---|---|---|---|---|---|---|---|---|
| NEUTM | rs73432891 | rs5961597 | A | | 23 | MTND6P12 | Female only | False | 0.05761 | 0.05871 | 0.05875 | 0.05883 | 5147 | 5352 | 187760 | 162101 | 0.1122 | -0.01796 | 2.41e-04 | -0.001234 | 0.03987 | 0.02716 |
| trait | lead_snp | ID | ALLELE1 | ALLELE0 | CHR | Gene_Symbol | independent_loci_in_sex_P1e2 | in_embl_any_or_gene | A1FREQ_VUMC_female | A1FREQ_VUMC_male | A1FREQ_UKB_female | A1FREQ_UKB_male | N_VUMC_female | N_VUMC_male | N_UKB_female | N_UKB_male | BETA_VUMC_female | BETA_VUMC_male | BETA_UKB_female | BETA_UKB_male | SE_VUMC_female | SE_VUMC_male |
| NEUTM | rs73432891 | rs56114849 | G | | 23 | MTND6P12 | Female only | False | 0.05644 | 0.05832 | 0.05847 | 0.05862 | 5147 | 5352 | 187760 | 162101 | 0.1229 | -0.01836 | 4.37e-04 | -0.001661 | 0.04026 | 0.02725 |
| NEUTM | rs73432891 | rs55670546 | A | | 23 | MTND6P12 | Female only | False | 0.05659 | 0.05852 | 0.05862 | 0.05877 | 5147 | 5352 | 187760 | 162101 | 0.1233 | -0.01753 | 3.27e-05 | -0.001527 | 0.04024 | 0.02722 |
| NEUTM | rs73432891 | rs58371007 | A | | 23 | MTND6P12 | Female only | False | 0.05647 | 0.05831 | 0.05848 | 0.05863 | 5147 | 5352 | 187760 | 162101 | 0.1232 | -0.01827 | 3.72e-04 | -0.001538 | 0.04025 | 0.02725 |
| NEUTM | rs73432891 | rs115472624 | A | | 23 | MTND6P12 | Female only | False | 0.05649 | 0.05834 | 0.05868 | 0.05866 | 5147 | 5352 | 187760 | 162101 | 0.1232 | -0.01835 | -4.89e-04 | -0.00156 | 0.04023 | 0.02724 |
| NEUTM | rs73432891 | rs114290791 | C | | 23 | MTND6P12 | Female only | False | 0.05649 | 0.05834 | 0.0586 | 0.05867 | 5147 | 5352 | 187760 | 162101 | 0.1232 | -0.01835 | 2.19e-04 | -0.001584 | 0.04023 | 0.02724 |
| NEUTM | rs73432891 | rs116805972 | A | | 23 | MTND6P12 | Female only | False | 0.0565 | 0.05835 | 0.05869 | 0.05867 | 5147 | 5352 | 187760 | 162101 | 0.1233 | -0.01832 | -3.37e-04 | -0.001576 | 0.04023 | 0.02724 |
| NEUTM | rs73432891 | rs116567335 | T | | 23 | MTND6P12 | Female only | False | 0.05649 | 0.05835 | 0.05868 | 0.05867 | 5147 | 5352 | 187760 | 162101 | 0.1232 | -0.01837 | -3.39e-04 | -0.001584 | 0.04023 | 0.02724 |
| NEUTM | rs73432891 | rs1562605 | C | | 23 | MTND6P12 | Female only | False | 0.05649 | 0.05834 | 0.05858 | 0.05865 | 5147 | 5352 | 187760 | 162101 | 0.1232 | -0.01836 | 4.36e-04 | -0.001559 | 0.04023 | 0.02724 |
| NEUTM | rs73432891 | rs7058294 | G | | 23 | MTND6P12 | Female only | False | 0.05649 | 0.05834 | 0.05861 | 0.05866 | 5147 | 5352 | 187760 | 162101 | 0.1232 | -0.01835 | 1.13e-04 | -0.001548 | 0.04023 | 0.02724 |
| NEUTM | rs73432891 | rs56955060 | T | | 23 | MTND6P12 | Female only | False | 0.05648 | 0.05834 | 0.05853 | 0.05866 | 5147 | 5352 | 187760 | 162101 | 0.1233 | -0.01834 | 2.81e-04 | -0.001558 | 0.04024 | 0.02724 |
| NEUTM | rs73432891 | rs17304657 | G | | 23 | MTND6P12 | Female only | False | 0.05645 | 0.05834 | 0.05873 | 0.05865 | 5147 | 5352 | 187760 | 162101 | 0.1228 | -0.01837 | -6.91e-04 | -0.001615 | 0.04025 | 0.02724 |
| NEUTM | rs73432891 | rs148775317 | A | | 23 | MTND6P12 | Female only | False | 0.05644 | 0.05833 | 0.05874 | 0.05864 | 5147 | 5352 | 187760 | 162101 | 0.1229 | -0.01838 | -6.11e-04 | -0.001587 | 0.04026 | 0.02724 |
| NEUTM | rs73432891 | rs116162447 | G | | 23 | MTND6P12 | Female only | False | 0.05648 | 0.05835 | 0.05876 | 0.05865 | 5147 | 5352 | 187760 | 162101 | 0.1234 | -0.01837 | -6.76e-04 | -0.001614 | 0.04024 | 0.02724 |
| NEUTM | rs73432891 | rs115944192 | G | | 23 | MTND6P12 | Female only | False | 0.05648 | 0.05835 | 0.05978 | 0.05875 | 5147 | 5352 | 187760 | 162101 | 0.1234 | -0.01837 | -0.001181 | -0.001438 | 0.04024 | 0.02724 |
| NEUTM | rs73432891 | rs141666026 | A | | 23 | MTND6P12 | Female only | False | 0.05649 | 0.05835 | 0.05878 | 0.05867 | 5147 | 5352 | 187760 | 162101 | 0.1233 | -0.01838 | -6.55e-04 | -0.001529 | 0.04023 | 0.02723 |
| NEUTM | rs73432891 | rs7066705 | G | | 23 | MTND6P12 | Female only | False | 0.05648 | 0.05834 | 0.05878 | 0.05866 | 5147 | 5352 | 187760 | 162101 | 0.1233 | -0.01838 | -6.71e-04 | -0.00154 | 0.04023 | 0.02724 |
| NEUTM | rs73432891 | rs7067127 | T | | 23 | MTND6P12 | Female only | False | 0.05647 | 0.05834 | 0.05875 | 0.05864 | 5147 | 5352 | 187760 | 162101 | 0.1234 | -0.01836 | -6.16e-04 | -0.00164 | 0.04024 | 0.02724 |
| NEUTM | rs73432891 | rs116259316 | C | | 23 | MTND6P12 | Female only | False | 0.05647 | 0.05834 | 0.05874 | 0.05864 | 5147 | 5352 | 187760 | 162101 | 0.1234 | -0.01835 | -5.37e-04 | -0.001626 | 0.04024 | 0.02724 |
| NEUTM | rs73432891 | rs115552759 | T | | 23 | MTND6P12 | Female only | False | 0.05653 | 0.05835 | 0.05876 | 0.05866 | 5147 | 5352 | 187760 | 162101 | 0.1252 | -0.01839 | -6.22e-04 | -0.001665 | 0.04023 | 0.02724 |
| NEUTM | rs73432891 | rs1106873 | C | | 23 | MTND6P12 | Female only | False | 0.05664 | 0.05848 | 0.05877 | 0.05867 | 5147 | 5352 | 187760 | 162101 | 0.1226 | -0.01839 | -5.63e-04 | -0.001595 | 0.04013 | 0.02717 |
| NEUTM | rs73432891 | rs56404144 | G | | 23 | MTND6P12 | Female only | False | 0.05504 | 0.05647 | 0.05896 | 0.05902 | 5147 | 5352 | 187760 | 162101 | 0.139 | -0.01977 | -0.001213 | -0.00166 | 0.04072 | 0.02768 |
| NEUTM | rs73432891 | rs10521587 | T | | 23 | MTND6P12 | Female only | False | 0.05513 | 0.05645 | 0.05899 | 0.05901 | 5147 | 5352 | 187760 | 162101 | 0.1387 | -0.01977 | -0.001373 | -0.002022 | 0.04067 | 0.02769 |
| NEUTM | rs73432891 | rs55649140 | C | | 23 | MTND6P12 | Female only | False | 0.05406 | 0.05493 | 0.05808 | 0.05825 | 5147 | 5352 | 187760 | 162101 | 0.1377 | -0.02503 | -0.001655 | -0.003034 | 0.04114 | 0.02807 |
| NEUTM | rs73432891 | rs56221421 | G | | 23 | MTND6P12 | Female only | False | 0.05407 | 0.05493 | 0.0581 | 0.05826 | 5147 | 5352 | 187760 | 162101 | 0.1376 | -0.02503 | -0.001735 | -0.002972 | 0.04114 | 0.02807 |
| NEUTM | rs73432891 | rs137977889 | G | | 23 | MTND6P12 | Female only | False | 0.05432 | 0.05514 | 0.05809 | 0.05828 | 5147 | 5352 | 187760 | 162101 | 0.1379 | -0.02413 | -0.001844 | -0.003095 | 0.04104 | 0.02802 |
| NEUTM | rs73432891 | rs17304720 | C | | 23 | MTND6P12 | Female only | False | 0.0536 | 0.05356 | 0.05736 | 0.05759 | 5147 | 5352 | 187760 | 162101 | 0.137 | -0.02758 | -0.00161 | -0.003542 | 0.04137 | 0.02853 |

| trait | lead_snp | ID | ALLELE1 | ALLELE0 | CHR | Gene_Symbol | independent_loci_in_sex_P1e2 | in_embl_any_or_gene | A1FREQ_VUMC_female | A1FREQ_VUMC_male | A1FREQ_UKB_female | A1FREQ_UKB_male | N_VUMC_female | N_VUMC_male | N_UKB_female | N_UKB_male | BETA_VUMC_female | BETA_VUMC_male | BETA_UKB_female | BETA_UKB_male | SE_VUMC_female | SE_VUMC_male |
|---|---|---|---|---|---|---|---|---|---|---|---|---|---|---|---|---|---|---|---|---|---|---|
| NEUTM | rs73432891 | rs7063372 | A | | 23 | MTND6P12 | Female only | False | 0.07132 | 0.07187 | 0.0744 | 0.0748 | 5147 | 5352 | 187760 | 162101 | 0.1947 | -0.004618 | -0.007622 | -2.98e-04 | 0.03635 | 0.02482 |
| NEUTM | rs73432891 | rs7061873 | A | | 23 | MTND6P12 | Female only | False | 0.07147 | 0.07194 | 0.0745 | 0.07488 | 5147 | 5352 | 187760 | 162101 | 0.1959 | -0.004366 | -0.007742 | -5.58e-04 | 0.0363 | 0.0248 |
| NEUTM | rs73432891 | rs73436536 | T | | 23 | MTND6P12 | Female only | False | 0.07154 | 0.07196 | 0.07455 | 0.07495 | 5147 | 5352 | 187760 | 162101 | 0.1981 | -0.004761 | -0.007819 | -2.70e-04 | 0.03614 | 0.02474 |
| NEUTM | rs73432891 | rs73436601 | T | | 23 | MTND6P12 | Female only | False | 0.07438 | 0.07518 | 0.0768 | 0.07725 | 5147 | 5352 | 187760 | 162101 | 0.206 | -0.002004 | -0.006395 | 5.88e-04 | 0.03555 | 0.02424 |
| NEUTM | rs73432891 | rs996058 | T | | 23 | MTND6P12 | Female only | False | 0.07471 | 0.07524 | 0.07677 | 0.07726 | 5147 | 5352 | 187760 | 162101 | 0.2058 | -0.001992 | -0.00684 | 8.02e-04 | 0.0355 | 0.02423 |
| NEUTM | rs73432891 | rs113020723 | C | | 23 | MTND6P12 | Female only | False | 0.07451 | 0.07524 | 0.07674 | 0.07721 | 5147 | 5352 | 187760 | 162101 | 0.2041 | -0.001991 | -0.006732 | 6.88e-04 | 0.03554 | 0.02423 |
| NEUTM | rs73432891 | rs7052521 | A | | 23 | MTND6P12 | Female only | False | 0.07464 | 0.07559 | 0.07684 | 0.07742 | 5147 | 5352 | 187760 | 162101 | 0.2029 | -0.004561 | -0.006374 | 5.15e-04 | 0.03551 | 0.02418 |
| NEUTM | rs73432891 | rs55798514 | A | | 23 | MTND6P12 | Female only | False | 0.07478 | 0.0758 | 0.07687 | 0.0775 | 5147 | 5352 | 187760 | 162101 | 0.2037 | -0.005203 | -0.006643 | 4.59e-04 | 0.03548 | 0.02414 |
| NEUTM | rs73432891 | rs16997018 | C | | 23 | MTND6P12 | Female only | False | 0.0748 | 0.07538 | 0.07657 | 0.07724 | 5147 | 5352 | 187760 | 162101 | 0.2013 | -0.001386 | -0.006024 | -2.95e-04 | 0.03538 | 0.02417 |
| NEUTM | rs73432891 | rs73448818 | C | | 23 | MTND6P12 | Female only | False | 0.07468 | 0.07536 | 0.07662 | 0.07732 | 5147 | 5352 | 187760 | 162101 | 0.2038 | -0.00211 | -0.006216 | -2.99e-04 | 0.03547 | 0.0242 |
| NEUTM | rs73432891 | rs7878812 | C | | 23 | MTND6P12 | Female only | False | 0.07458 | 0.07532 | 0.07672 | 0.07742 | 5147 | 5352 | 187760 | 162101 | 0.2041 | -0.002464 | -0.006478 | -1.34e-04 | 0.03551 | 0.02421 |
| NEUTM | rs73432891 | rs7065808 | T | | 23 | MTND6P12 | Female only | False | 0.07454 | 0.07546 | 0.07708 | 0.07726 | 5147 | 5352 | 187760 | 162101 | 0.2063 | -0.001341 | -0.007757 | -7.58e-04 | 0.0355 | 0.02419 |
| NEUTM | rs73432891 | rs112977121 | A | | 23 | MTND6P12 | Female only | False | 0.07454 | 0.07546 | 0.07663 | 0.07723 | 5147 | 5352 | 187760 | 162101 | 0.2063 | -0.001341 | -0.006748 | -5.33e-04 | 0.0355 | 0.02419 |
| NEUTM | rs73432891 | rs1462648 | A | | 23 | MTND6P12 | Female only | False | 0.0749 | 0.07587 | 0.07669 | 0.07726 | 5147 | 5352 | 187760 | 162101 | 0.2059 | -8.87e-04 | -0.00673 | -5.22e-04 | 0.03537 | 0.02409 |
| NEUTM | rs73432891 | rs73432891 | C | | 23 | MTND6P12 | Female only | False | 0.07468 | 0.07569 | 0.0767 | 0.07727 | 5147 | 5352 | 187760 | 162101 | 0.2068 | -7.90e-04 | -0.006736 | -5.00e-04 | 0.03547 | 0.02415 |
| NEUTM | rs73432891 | rs111847052 | G | | 23 | MTND6P12 | Female only | False | 0.07468 | 0.07569 | 0.07668 | 0.07726 | 5147 | 5352 | 187760 | 162101 | 0.2068 | -7.90e-04 | -0.006733 | -5.24e-04 | 0.03547 | 0.02415 |
| NEUTM | rs73432891 | rs73434716 | T | | 23 | MTND6P12 | Female only | False | 0.07469 | 0.07587 | 0.0767 | 0.07728 | 5147 | 5352 | 187760 | 162101 | 0.2072 | -4.47e-04 | -0.006671 | -4.50e-04 | 0.03558 | 0.02421 |
| NEUTM | rs73432891 | rs35327795 | T | | 23 | MTND6P12 | Female only | False | 0.07467 | 0.07585 | 0.07668 | 0.07726 | 5147 | 5352 | 187760 | 162101 | 0.207 | -4.42e-04 | -0.006622 | -5.12e-04 | 0.03559 | 0.02422 |
| NEUTM | rs73432891 | rs112999455 | G | | 23 | MTND6P12 | Female only | False | 0.07469 | 0.07587 | 0.07669 | 0.07727 | 5147 | 5352 | 187760 | 162101 | 0.2071 | -4.47e-04 | -0.006693 | -4.75e-04 | 0.03559 | 0.02421 |
| NEUTM | rs73432891 | rs73434740 | A | | 23 | MTND6P12 | Female only | False | 0.06852 | 0.07028 | 0.07109 | 0.07165 | 5147 | 5352 | 187760 | 162101 | 0.2156 | 0.00461 | -0.003872 | -3.45e-04 | 0.03733 | 0.02524 |
| NEUTM | rs73432891 | rs73434798 | C | | 23 | MTND6P12 | Female only | False | 0.06846 | 0.07027 | 0.0711 | 0.07165 | 5147 | 5352 | 187760 | 162101 | 0.2155 | 0.004609 | -0.003848 | -2.70e-04 | 0.03736 | 0.02524 |
| NEUTM | rs73432891 | rs113216425 | C | | 23 | MTND6P12 | Female only | False | 0.06846 | 0.07027 | 0.07109 | 0.07165 | 5147 | 5352 | 187760 | 162101 | 0.2155 | 0.004609 | -0.003836 | -3.12e-04 | 0.03736 | 0.02524 |
| NEUTM | rs73432891 | rs112247182 | A | | 23 | MTND6P12 | Female only | False | 0.06848 | 0.07017 | 0.07108 | 0.07159 | 5147 | 5352 | 187760 | 162101 | 0.216 | 0.005456 | -0.00391 | -3.66e-04 | 0.03733 | 0.02526 |
| VD25 | rs1993116 | rs7119073 | G | A | 11 | CYP2R1 | Female only | False | 0.3977 | 0.4135 | 0.4184 | 0.4193 | 2437 | 949 | 174597 | 154650 | 0.1031 | 0.04513 | 0.03724 | 0.03944 | 0.02629 | 0.03825 |
| VD25 | rs1993116 | rs10741653 | C | A | 11 | CYP2R1 | Female only | False | 0.3976 | 0.4134 | 0.4184 | 0.4193 | 2437 | 949 | 174597 | 154650 | 0.1027 | 0.04577 | 0.03726 | 0.03941 | 0.02629 | 0.03821 |
| VD25 | rs1993116 | rs11023310 | T | G | 11 | CYP2R1 | Female only | False | 0.3975 | 0.4132 | 0.4185 | 0.4193 | 2437 | 949 | 174597 | 154650 | 0.1025 | 0.04668 | 0.03724 | 0.0394 | 0.02627 | 0.03816 |
| VD25 | rs1993116 | rs10741654 | C | A | 11 | CYP2R1 | Female only | False | 0.3913 | 0.4046 | 0.4171 | 0.4181 | 2437 | 949 | 174597 | 154650 | 0.1052 | 0.04021 | 0.03702 | 0.03875 | 0.02611 | 0.03793 |
| VD25 | rs1993116 | rs10832295 | T | C | 11 | CYP2R1 | Female only | False | 0.3957 | 0.409 | 0.4171 | 0.4181 | 2437 | 949 | 174597 | 154650 | 0.1053 | 0.04032 | 0.037 | 0.03881 | 0.02629 | 0.03821 |
| VD25 | rs1993116 | rs76376118 | T | C | 11 | CYP2R1 | Female only | False | 0.3958 | 0.4091 | | | 2437 | 949 | | | 0.1053 | 0.04003 | | | 0.02629 | 0.03822 |

| trait | lead_snp | ID | ALLELE1 | ALLELE0 | CHR | Gene_Symbol | independent_loci_in_sex_P1e2 | in_embl_any_or_gene | A1FREQ_VUMC_female | A1FREQ_VUMC_male | A1FREQ_UKB_female | A1FREQ_UKB_male | N_VUMC_female | N_VUMC_male | N_UKB_female | N_UKB_male | BETA_VUMC_female | BETA_VUMC_male | BETA_UKB_female | BETA_UKB_male | SE_VUMC_female | SE_VUMC_male |
|---|---|---|---|---|---|---|---|---|---|---|---|---|---|---|---|---|---|---|---|---|---|---|
| VD25 | rs1993116 | rs9971395 | G | A | 11 | CYP2R1 | Female only | False | 0.3958 | 0.4092 | 0.4171 | 0.4181 | 2437 | 949 | 174597 | 154650 | 0.1054 | 0.0404 | 0.03703 | 0.03878 | 0.0263 | 0.03822 |
| VD25 | rs1993116 | rs10734227 | C | T | 11 | CYP2R1 | Female only | False | 0.3959 | 0.4082 | 0.4173 | 0.4182 | 2437 | 949 | 174597 | 154650 | 0.1047 | 0.03661 | 0.03708 | 0.03877 | 0.02632 | 0.03819 |
| VD25 | rs1993116 | rs6486202 | A | C | 11 | CYP2R1 | Female only | False | 0.3958 | 0.4082 | 0.4172 | 0.4181 | 2437 | 949 | 174597 | 154650 | 0.1046 | 0.0366 | 0.0371 | 0.03873 | 0.02632 | 0.03819 |
| VD25 | rs1993116 | rs1542291 | T | G | 11 | CYP2R1 | Female only | False | 0.3958 | 0.4085 | 0.4169 | 0.4179 | 2437 | 949 | 174597 | 154650 | 0.1058 | 0.03855 | 0.03703 | 0.03884 | 0.02634 | 0.0382 |
| VD25 | rs1993116 | rs4757268 | A | G | 11 | CYP2R1 | Female only | False | 0.3909 | 0.4052 | 0.4171 | 0.4181 | 2437 | 949 | 174597 | 154650 | 0.1057 | 0.04278 | 0.03718 | 0.03892 | 0.02612 | 0.03789 |
| VD25 | rs1993116 | rs10766193 | C | T | 11 | CYP2R1 | Female only | False | 0.3959 | 0.4092 | 0.4169 | 0.4179 | 2437 | 949 | 174597 | 154650 | 0.1056 | 0.04092 | 0.03706 | 0.0388 | 0.02633 | 0.03815 |
| VD25 | rs1993116 | rs2349793 | A | G | 11 | CYP2R1 | Female only | False | 0.396 | 0.4105 | 0.4171 | 0.4181 | 2437 | 949 | 174597 | 154650 | 0.105 | 0.04256 | 0.03715 | 0.03888 | 0.02633 | 0.03822 |
| VD25 | rs1993116 | rs10741656 | G | A | 11 | CYP2R1 | Female only | False | 0.396 | 0.4105 | 0.4171 | 0.4181 | 2437 | 949 | 174597 | 154650 | 0.105 | 0.04256 | 0.03712 | 0.03884 | 0.02633 | 0.03822 |
| VD25 | rs1993116 | rs3740823 | A | G | 11 | CYP2R1 | Female only | False | 0.396 | 0.4105 | 0.4171 | 0.4181 | 2437 | 949 | 174597 | 154650 | 0.105 | 0.04256 | 0.03712 | 0.03884 | 0.02633 | 0.03822 |
| VD25 | rs1993116 | rs1839834 | A | G | 11 | CYP2R1 | Female only | False | 0.396 | 0.4094 | 0.4169 | 0.4179 | 2437 | 949 | 174597 | 154650 | 0.1051 | 0.03983 | 0.03704 | 0.03878 | 0.02633 | 0.03817 |
| VD25 | rs1993116 | rs1597689 | G | A | 11 | CYP2R1 | Female only | False | 0.3966 | 0.4122 | 0.4177 | 0.4186 | 2437 | 949 | 174597 | 154650 | 0.1026 | 0.03663 | 0.03731 | 0.03874 | 0.02631 | 0.03816 |
| VD25 | rs1993116 | rs1037378 | G | A | 11 | CYP2R1 | Female only | False | 0.4423 | 0.4622 | 0.4615 | 0.4615 | 2437 | 949 | 174597 | 154650 | 0.1376 | 0.05862 | 0.1004 | 0.1008 | 0.02637 | 0.0376 |
| VD25 | rs1993116 | rs1037379 | A | G | 11 | CYP2R1 | Female only | False | 0.3559 | 0.3718 | 0.3729 | 0.3746 | 2437 | 949 | 174597 | 154650 | 0.15 | 0.06144 | 0.0847 | 0.08559 | 0.02712 | 0.03881 |
| VD25 | rs1993116 | rs11023364 | G | A | 11 | CYP2R1 | Female only | False | 0.4399 | 0.4292 | 0.4196 | 0.4185 | 2437 | 949 | 174597 | 154650 | -0.1241 | -0.02512 | -0.08386 | -0.08959 | 0.02667 | 0.03879 |
| VD25 | rs1993116 | rs7125781 | T | C | 11 | CYP2R1 | Female only | False | 0.3563 | 0.3718 | 0.3732 | 0.3748 | 2437 | 949 | 174597 | 154650 | 0.1519 | 0.05829 | 0.08483 | 0.08573 | 0.02715 | 0.039 |
| VD25 | rs1993116 | rs6486205 | T | G | 11 | CYP2R1 | Female only | False | 0.3564 | 0.3716 | 0.3732 | 0.3748 | 2437 | 949 | 174597 | 154650 | 0.1519 | 0.05769 | 0.08479 | 0.08573 | 0.02716 | 0.03902 |
| VD25 | rs1993116 | rs7116978 | T | C | 11 | CYP2R1 | Female only | False | 0.3564 | 0.3716 | 0.3732 | 0.3748 | 2437 | 949 | 174597 | 154650 | 0.1519 | 0.05769 | 0.08483 | 0.08574 | 0.02716 | 0.03902 |
| VD25 | rs1993116 | rs10832311 | G | T | 11 | CYP2R1 | Female only | False | 0.4398 | 0.4292 | 0.4195 | 0.4184 | 2437 | 949 | 174597 | 154650 | -0.1239 | -0.02517 | -0.08385 | -0.08965 | 0.02668 | 0.03879 |
| VD25 | rs1993116 | rs1868997 | T | C | 11 | CYP2R1 | Female only | False | 0.3565 | 0.3714 | 0.3732 | 0.3749 | 2437 | 949 | 174597 | 154650 | 0.1519 | 0.05728 | 0.08488 | 0.08559 | 0.02717 | 0.03904 |
| VD25 | rs1993116 | rs1993116 | A | G | 11 | CYP2R1 | Female only | False | 0.3885 | 0.3957 | 0.4124 | 0.4147 | 2437 | 949 | 174597 | 154650 | 0.1584 | 0.05171 | 0.09255 | 0.09441 | 0.02638 | 0.03836 |
| VD25 | rs1993116 | rs10500804 | G | T | 11 | CYP2R1 | Female only | False | 0.4415 | 0.4315 | 0.4212 | 0.4202 | 2437 | 949 | 174597 | 154650 | -0.1297 | -0.02364 | -0.08458 | -0.08998 | 0.02672 | 0.03867 |
| VD25 | rs1993116 | rs10766196 | G | A | 11 | CYP2R1 | Female only | False | 0.4407 | 0.4316 | 0.4216 | 0.4207 | 2437 | 949 | 174597 | 154650 | -0.1319 | -0.02358 | -0.08467 | -0.08994 | 0.02673 | 0.03865 |
| VD25 | rs1993116 | rs12794714 | A | G | 11 | CYP2R1 | Female only | False | 0.4452 | 0.4341 | 0.4215 | 0.4205 | 2437 | 949 | 174597 | 154650 | -0.1303 | -0.02352 | -0.08465 | -0.09005 | 0.02643 | 0.0382 |
| VD25 | rs1993116 | rs10741657 | A | G | 11 | CYP2R1 | Female only | False | 0.3826 | 0.3878 | 0.4043 | 0.4065 | 2437 | 949 | 174597 | 154650 | 0.1575 | 0.05087 | 0.08926 | 0.09129 | 0.02644 | 0.03819 |
| VD25 | rs1993116 | rs2060793 | A | G | 11 | CYP2R1 | Female only | False | 0.3881 | 0.3937 | 0.4051 | 0.4071 | 2437 | 949 | 174597 | 154650 | 0.1592 | 0.05364 | 0.08937 | 0.09112 | 0.02665 | 0.03857 |
| VD25 | rs1993116 | rs1562902 | C | T | 11 | CYP2R1 | Female only | False | 0.431 | 0.4328 | 0.4532 | 0.455 | 2437 | 949 | 174597 | 154650 | 0.09541 | -7.48e-04 | 0.0576 | 0.06396 | 0.02599 | 0.03798 |
| VD25 | rs1993116 | rs1553006 | A | G | 11 | CYP2R1 | Female only | False | 0.4365 | 0.4375 | 0.4524 | 0.4541 | 2437 | 949 | 174597 | 154650 | 0.09611 | 0.001246 | 0.05746 | 0.06357 | 0.02626 | 0.03846 |

| trait | lead_snp | ID | ALLELE1 | ALLELE0 | CHR | Gene_Symbol | independent_loci_in_sex_P1e2 | in_embl_any_or_gene | A1FREQ_VUMC_female | A1FREQ_VUMC_male | A1FREQ_UKB_female | A1FREQ_UKB_male | N_VUMC_female | N_VUMC_male | N_UKB_female | N_UKB_male | BETA_VUMC_female | BETA_VUMC_male | BETA_UKB_female | BETA_UKB_male | SE_VUMC_female | SE_VUMC_male |
|---|---|---|---|---|---|---|---|---|---|---|---|---|---|---|---|---|---|---|---|---|---|---|
| WBCrt | rs13329952 | rs71384446 | C | A | 16 | UMOD | Male only | True | 0.2335 | 0.2373 | 0.2358 | 0.2351 | 4458 | 4826 | 184912 | 159192 | -0.03099 | -0.1269 | -0.05905 | -0.05718 | 0.02437 | 0.02399 |
| WBCrt | rs13329952 | rs9928936 | A | G | 16 | UMOD | Male only | True | 0.177 | 0.185 | 0.1819 | 0.1809 | 4458 | 4826 | 184912 | 159192 | -0.04847 | -0.1408 | -0.07652 | -0.0714 | 0.02696 | 0.02654 |
| WBCrt | rs13329952 | rs60136849 | C | T | 16 | UMOD | Male only | True | 0.1764 | 0.1846 | 0.1815 | 0.1805 | 4458 | 4826 | 184912 | 159192 | -0.04796 | -0.1423 | -0.07698 | -0.07147 | 0.02698 | 0.02657 |
| WBCrt | rs13329952 | rs6497475 | T | C | 16 | UMOD | Male only | True | 0.2354 | 0.2443 | 0.2318 | 0.2305 | 4458 | 4826 | 184912 | 159192 | -0.0335 | -0.1321 | -0.05952 | -0.05223 | 0.0248 | 0.02415 |
| WBCrt | rs13329952 | rs34262842 | G | A | 16 | UMOD | Male only | True | 0.159 | 0.1675 | 0.1671 | 0.1664 | 4458 | 4826 | 184912 | 159192 | -0.04653 | -0.1657 | -0.07956 | -0.07492 | 0.02814 | 0.02756 |
| WBCrt | rs13329952 | rs11647727 | A | G | 16 | UMOD | Male only | True | 0.2396 | 0.2512 | 0.2408 | 0.2397 | 4458 | 4826 | 184912 | 159192 | -0.02663 | -0.1406 | -0.05779 | -0.05278 | 0.02424 | 0.02355 |
| WBCrt | rs13329952 | rs34356953 | T | G | 16 | UMOD | Male only | True | 0.1763 | 0.1847 | 0.1817 | 0.1808 | 4458 | 4826 | 184912 | 159192 | -0.04782 | -0.1422 | -0.0769 | -0.07175 | 0.027 | 0.02654 |
| WBCrt | rs13329952 | rs12934320 | T | C | 16 | UMOD | Male only | True | 0.1763 | 0.1847 | 0.1817 | 0.1808 | 4458 | 4826 | 184912 | 159192 | -0.04782 | -0.1422 | -0.07699 | -0.07171 | 0.027 | 0.02654 |
| WBCrt | rs13329952 | rs12934455 | T | C | 16 | UMOD | Male only | True | 0.1596 | 0.1681 | 0.1671 | 0.1665 | 4458 | 4826 | 184912 | 159192 | -0.04584 | -0.1651 | -0.07953 | -0.07517 | 0.02803 | 0.02746 |
| WBCrt | rs13329952 | rs9928003 | C | A | 16 | UMOD | Male only | True | 0.1752 | 0.1833 | 0.1807 | 0.1798 | 4458 | 4826 | 184912 | 159192 | -0.051 | -0.146 | -0.07708 | -0.07265 | 0.02703 | 0.02662 |
| WBCrt | rs13329952 | rs28640218 | T | G | 16 | UMOD | Male only | True | 0.1883 | 0.1996 | 0.192 | 0.1912 | 4458 | 4826 | 184912 | 159192 | -0.03606 | -0.1588 | -0.07353 | -0.07179 | 0.02633 | 0.02571 |
| WBCrt | rs13329952 | rs28544423 | T | C | 16 | UMOD | Male only | True | 0.1772 | 0.1858 | 0.1829 | 0.1819 | 4458 | 4826 | 184912 | 159192 | -0.04741 | -0.1429 | -0.07691 | -0.07289 | 0.02694 | 0.02648 |
| WBCrt | rs13329952 | rs13335818 | T | C | 16 | UMOD | Male only | True | 0.1762 | 0.1845 | 0.1819 | 0.181 | 4458 | 4826 | 184912 | 159192 | -0.04999 | -0.1473 | -0.07689 | -0.07373 | 0.02696 | 0.02655 |
| WBCrt | rs13329952 | rs7193058 | G | A | 16 | UMOD | Male only | True | 0.2479 | 0.2596 | 0.2434 | 0.2422 | 4458 | 4826 | 184912 | 159192 | -0.02524 | -0.1453 | -0.05844 | -0.05399 | 0.02435 | 0.02371 |
| WBCrt | rs13329952 | rs34882080 | G | A | 16 | UMOD | Male only | True | 0.1774 | 0.1858 | 0.1829 | 0.182 | 4458 | 4826 | 184912 | 159192 | -0.04798 | -0.1437 | -0.07664 | -0.07291 | 0.02689 | 0.0265 |
| WBCrt | rs13329952 | rs35650857 | T | C | 16 | UMOD | Male only | True | 0.1774 | 0.1858 | 0.1829 | 0.182 | 4458 | 4826 | 184912 | 159192 | -0.04798 | -0.1437 | -0.07664 | -0.07291 | 0.02689 | 0.0265 |
| WBCrt | rs13329952 | rs36060036 | T | C | 16 | UMOD | Male only | True | 0.1598 | 0.1681 | 0.1682 | 0.1676 | 4458 | 4826 | 184912 | 159192 | -0.04661 | -0.1669 | -0.07926 | -0.0757 | 0.02807 | 0.02751 |
| WBCrt | rs13329952 | rs4293393 | G | A | 16 | UMOD | Male only | True | 0.1785 | 0.1864 | 0.1834 | 0.1824 | 4458 | 4826 | 184912 | 159192 | -0.04438 | -0.1511 | -0.07685 | -0.07283 | 0.02689 | 0.02648 |
| WBCrt | rs13329952 | rs28362063 | C | T | 16 | UMOD | Male only | True | 0.1771 | 0.1847 | 0.1825 | 0.1816 | 4458 | 4826 | 184912 | 159192 | -0.04735 | -0.1468 | -0.07707 | -0.07405 | 0.0269 | 0.02649 |
| WBCrt | rs13329952 | rs13333226 | G | A | 16 | UMOD | Male only | True | 0.1805 | 0.1883 | 0.1841 | 0.1831 | 4458 | 4826 | 184912 | 159192 | -0.04758 | -0.1473 | -0.07704 | -0.07295 | 0.02674 | 0.02629 |
| WBCrt | rs13329952 | rs13329952 | C | T | 16 | UMOD | Male only | True | 0.1891 | 0.1998 | 0.1919 | 0.1912 | 4458 | 4826 | 184912 | 159192 | -0.04086 | -0.1619 | -0.07397 | -0.07325 | 0.02625 | 0.02568 |
| WBCrt | rs13329952 | rs9933330 | T | C | 16 | UMOD | Male only | True | 0.1765 | 0.1841 | 0.1825 | 0.1817 | 4458 | 4826 | 184912 | 159192 | -0.04793 | -0.1475 | -0.07722 | -0.07399 | 0.02699 | 0.02658 |
| WBCrt | rs13329952 | rs7204775 | C | T | 16 | UMOD | Male only | True | 0.1899 | 0.2007 | 0.1929 | 0.1921 | 4458 | 4826 | 184912 | 159192 | -0.03934 | -0.158 | -0.07383 | -0.07221 | 0.02628 | 0.0257 |
| WBCrt | rs13329952 | rs7203642 | G | A | 16 | UMOD | Male only | True | 0.1903 | 0.201 | 0.1925 | 0.1918 | 4458 | 4826 | 184912 | 159192 | -0.03922 | -0.1554 | -0.07373 | -0.07256 | 0.02621 | 0.02564 |
| WBCrt | rs13329952 | rs28688991 | T | C | 16 | UMOD | Male only | True | 0.1905 | 0.2014 | 0.1929 | 0.1921 | 4458 | 4826 | 184912 | 159192 | -0.03831 | -0.1574 | -0.07388 | -0.07214 | 0.0262 | 0.02561 |
| WBCrt | rs13329952 | rs12922822 | T | C | 16 | UMOD | Male only | True | 0.1771 | 0.1847 | 0.1824 | 0.1815 | 4458 | 4826 | 184912 | 159192 | -0.0469 | -0.147 | -0.07719 | -0.07379 | 0.0269 | 0.02649 |
| WBCrt | rs13329952 | rs12917707 | T | G | 16 | UMOD | Male only | True | 0.1764 | 0.184 | 0.182 | 0.1811 | 4458 | 4826 | 184912 | 159192 | -0.04938 | -0.1464 | -0.07691 | -0.07347 | 0.02694 | 0.02652 |
| BASOPHILS | rs5911858 | rs2884138 | C | | 23 | TENM1 | Male only | False | 0.1777 | 0.1806 | 0.1784 | 0.1772 | 885 | 592 | 187758 | 162098 | 0.01872 | -0.2512 | -2.91e-04 | -0.002714 | 0.0562 | 0.05302 |
| BASOPHILS | rs5911858 | rs5911855 | C | | 23 | TENM1 | Male only | False | 0.1803 | 0.1786 | 0.1817 | 0.1802 | 885 | 592 | 187758 | 162098 | 0.02865 | -0.255 | -7.40e-04 | -0.003879 | 0.05557 | 0.05287 |

| trait | lead_snp | ID | ALLELE1 | ALLELE0 | CHR | Gene_Symbol | independent_loci_in_sex_P1e2 | in_embl_any_or_gene | A1FREQ_VUMC_female | A1FREQ_VUMC_male | A1FREQ_UKB_female | A1FREQ_UKB_male | N_VUMC_female | N_VUMC_male | N_UKB_female | N_UKB_male | BETA_VUMC_female | BETA_VUMC_male | BETA_UKB_female | BETA_UKB_male | SE_VUMC_female | SE_VUMC_male |
|---|---|---|---|---|---|---|---|---|---|---|---|---|---|---|---|---|---|---|---|---|---|---|
| BASOPHILS | rs5911858 | rs5911857 | G | | 23 | TENM1 | Male only | False | 0.2938 | 0.2713 | 0.3005 | 0.2996 | 885 | 592 | 187758 | 162098 | 0.03733 | -0.1819 | -4.94e-04 | -8.51e-04 | 0.04799 | 0.04667 |
| BASOPHILS | rs5911858 | rs5911858 | G | | 23 | TENM1 | Male only | False | 0.2226 | 0.2069 | 0.2266 | 0.2247 | 885 | 592 | 187758 | 162098 | 0.05954 | -0.2861 | 0.002035 | 7.44e-05 | 0.05125 | 0.0505 |

| SE_UKB_female | SE_UKB_male | LOG10P_VUMC_female | LOG10P_VUMC_male | LOG10P_UKB_female | LOG10P_UKB_male | female_effect_direction_replication | male_effect_direction_replication | sex_effect_comparison_replication | pvalue_effect_comparison_replication | replication_type |
|---|---|---|---|---|---|---|---|---|---|---|
| 0.003588 | 0.003882 | 5.3 | 0.1943 | 30.04 | 22.31 | True | True | True | True | Sex-specific 1e-2 VUMC, sex-differential UKB |
| 0.003585 | 0.003878 | 5.091 | 0.1608 | 29.98 | 22.28 | True | True | True | True | Sex-specific 1e-2 VUMC, sex-differential UKB |
| 0.003605 | 0.003896 | 4.727 | 0.1588 | 29.51 | 22.27 | True | True | True | True | Sex-specific 1e-2 VUMC, sex-differential UKB |
| 0.003586 | 0.003879 | 5.214 | 0.164 | 30.19 | 22.2 | True | True | True | True | Sex-specific 1e-2 VUMC, sex-differential UKB |
| 0.003586 | 0.003879 | 5.222 | 0.1686 | 30.19 | 21.61 | True | True | True | True | Sex-specific 1e-2 VUMC, sex-differential UKB |
| 0.003627 | 0.003918 | 5.47 | 0.04075 | 28.89 | 21.98 | True | False | True | True | Sex-specific 1e-2 VUMC, sex-differential UKB |
| 0.003288 | 0.003559 | 9.008 | 1.878 | 82.49 | 59.62 | True | True | True | True | Sex-specific 1e-2 VUMC, sex-differential UKB |
| 0.003634 | 0.003927 | 5.246 | 0.09556 | 28.78 | 22.34 | True | False | True | True | Sex-specific 1e-2 VUMC, sex-differential UKB |
| 0.003586 | 0.00388 | 5.28 | 0.1597 | 30.07 | 22.17 | True | True | True | True | Sex-specific 1e-2 VUMC, sex-differential UKB |
| 0.003592 | 0.003882 | 5.378 | 0.1086 | 30.39 | 20.26 | True | False | True | True | Sex-specific 1e-2 VUMC, sex-differential UKB |
| 0.003596 | 0.003886 | 5.526 | 0.1618 | 30.35 | 20.23 | True | False | True | True | Sex-specific 1e-2 VUMC, sex-differential UKB |
| 0.003571 | 0.003862 | 5.763 | 0.1057 | 30.69 | 20.11 | True | False | True | True | Sex-specific 1e-2 VUMC, sex-differential UKB |
| 0.003571 | 0.003863 | 5.695 | 0.1125 | 30.66 | 20.1 | True | False | True | True | Sex-specific 1e-2 VUMC, sex-differential UKB |
| 0.003595 | 0.003886 | 5.762 | 0.1593 | 30.62 | 20.3 | True | False | True | True | Sex-specific 1e-2 VUMC, sex-differential UKB |
| 0.003268 | 0.003538 | 8.865 | 1.779 | 86.41 | 58.6 | True | True | True | True | Sex-specific 1e-2 VUMC, sex-differential UKB |
| 0.003555 | 0.003846 | 5.342 | 0.01038 | 32.04 | 20.51 | True | False | True | True | Sex-specific 1e-2 VUMC, sex-differential UKB |
| 0.003238 | 0.003511 | 7.297 | 1.476 | 59.45 | 41.15 | True | True | True | True | Sex-specific 1e-2 VUMC, sex-differential UKB |
| 0.003395 | 0.003681 | 5.796 | 1.235 | 51.13 | 39.5 | True | True | True | True | Sex-specific 1e-2 VUMC, sex-differential UKB |
| 0.003272 | 0.00354 | 9.03 | 1.493 | 70.08 | 60.68 | True | True | False | True | |
| 0.003309 | 0.003584 | 6.941 | 1.194 | 53.57 | 49.33 | True | True | False | True | |
| 0.003309 | 0.003583 | 7.036 | 1.137 | 53.33 | 49.21 | True | True | False | True | |
| 0.003308 | 0.003582 | 6.988 | 1.078 | 53.25 | 49.05 | True | True | False | True | |
| 0.003307 | 0.003581 | 6.957 | 0.9989 | 52.91 | 48.96 | True | True | False | True | |
| 0.003271 | 0.003538 | 8.866 | 1.323 | 69.18 | 60.27 | True | True | False | True | |
| 0.003309 | 0.003583 | 6.949 | 1.024 | 52.9 | 48.75 | True | True | False | True | |

| SE_UKB_female | SE_UKB_male | LOG10P_VUMC_female | LOG10P_VUMC_male | LOG10P_UKB_female | LOG10P_UKB_male | female_effect_direction_replication | male_effect_direction_replication | sex_effect_comparison_replication | pvalue_effect_comparison_replication | replication_type |
|---|---|---|---|---|---|---|---|---|---|---|
| 0.003272 | 0.003539 | 8.687 | 1.27 | 69.03 | 60.13 | True | True | False | True | |
| 0.003272 | 0.003539 | 8.705 | 1.335 | 69.11 | 60.21 | True | True | False | True | |
| 0.003272 | 0.003539 | 8.707 | 1.343 | 69.02 | 60.25 | True | True | False | True | |
| 0.003302 | 0.003576 | 7.256 | 1.321 | 53.99 | 50.16 | True | True | False | True | |
| 0.003272 | 0.003539 | 8.695 | 1.215 | 68.76 | 59.41 | True | True | False | True | |
| 0.003273 | 0.00354 | 8.717 | 1.235 | 68.84 | 59.68 | True | True | False | True | |
| 0.003272 | 0.003539 | 8.714 | 1.221 | 68.97 | 59.21 | True | True | False | True | |
| 0.003272 | 0.003539 | 8.714 | 1.22 | 68.92 | 59.19 | True | True | False | True | |
| 0.003272 | 0.003539 | 9.006 | 1.293 | 68.95 | 59.13 | True | True | False | True | |
| 0.003276 | 0.003545 | 8.071 | 1.069 | 62.78 | 56.2 | True | True | False | True | |
| 0.003272 | 0.003539 | 8.717 | 1.22 | 68.99 | 59.23 | True | True | False | True | |
| 0.003273 | 0.003539 | 8.714 | 1.22 | 68.88 | 59.36 | True | True | False | True | |
| 0.003273 | 0.00354 | 8.687 | 1.234 | 68.88 | 59.42 | True | True | False | True | |
| 0.003273 | 0.00354 | 8.681 | 1.23 | 68.87 | 59.41 | True | True | False | True | |
| 0.003273 | 0.00354 | 8.705 | 1.221 | 68.97 | 59.22 | True | True | False | True | |
| 0.003305 | 0.003578 | 7.535 | 1.362 | 54.57 | 49.32 | True | True | False | True | |
| 0.003275 | 0.003542 | 8.758 | 1.267 | 68.5 | 58.59 | True | True | True | True | Sex-specific 1e-2 VUMC, sex-differential UKB |
| 0.003305 | 0.003578 | 7.29 | 1.262 | 54.44 | 49.54 | True | True | False | True | |
| 0.003273 | 0.003541 | 8.845 | 1.278 | 67.8 | 57.96 | True | True | True | True | Sex-specific 1e-2 VUMC, sex-differential UKB |
| 0.003273 | 0.003541 | 8.852 | 1.276 | 67.47 | 58.08 | True | True | False | True | |
| 0.003462 | 0.003746 | 6.904 | 0.8412 | 43.26 | 41.6 | True | True | False | True | |
| | | 9.363 | 1.149 | | | False | False | False | False | |
| | | 9.274 | 1.142 | | | False | False | False | False | |
| 0.003273 | 0.003508 | 1.649 | 7.447 | 14.77 | 22.55 | True | True | True | True | Sex-specific 1e-2 VUMC, sex-differential UKB |
| 0.003271 | 0.003504 | 1.437 | 7.67 | 14.57 | 23.08 | True | True | True | True | Sex-specific 1e-2 VUMC, sex-differential UKB |
| 0.003258 | 0.003492 | 1.46 | 7.656 | 16.99 | 23.58 | True | True | True | True | Sex-specific 1e-2 VUMC, sex-differential UKB |
| 0.003266 | 0.003501 | 0.9328 | 7.953 | 19.14 | 25.15 | True | True | True | True | Sex-specific 1e-2 VUMC, sex-differential UKB |
| 0.005166 | 0.005536 | 5.058 | 0.6755 | 2.389 | 4.162 | True | True | False | False | |

| SE_UKB_female | SE_UKB_male | LOG10P_VUMC_female | LOG10P_VUMC_male | LOG10P_UKB_female | LOG10P_UKB_male | female_effect_direction_replication | male_effect_direction_replication | sex_effect_comparison_replication | pvalue_effect_comparison_replication | replication_type |
|---|---|---|---|---|---|---|---|---|---|---|
| 0.005165 | 0.005535 | 5.041 | 0.6668 | 2.364 | 4.128 | True | True | False | False | |
| 0.005179 | 0.005551 | 5.51 | 0.6861 | 2.342 | 4.214 | True | True | False | False | |
| 0.005159 | 0.005531 | 5.577 | 0.6512 | 2.426 | 4.209 | True | True | False | False | |
| 0.005061 | 0.005431 | 5.114 | 1.05 | 2.471 | 3.083 | True | True | False | False | |
| 0.005159 | 0.005531 | 5.423 | 0.6683 | 2.426 | 4.232 | True | True | False | False | |
| 0.005063 | 0.005432 | 4.985 | 1.067 | 2.441 | 3.053 | True | True | False | False | |
| 0.00516 | 0.005532 | 5.435 | 0.6615 | 2.428 | 4.263 | True | True | False | False | |
| 0.005163 | 0.005533 | 5.423 | 0.6553 | 2.398 | 4.316 | True | True | False | False | |
| 0.005062 | 0.005431 | 4.981 | 1.063 | 2.449 | 3.042 | True | True | False | False | |
| 0.004933 | 0.005299 | 4.61 | 0.9885 | 2.843 | 2.921 | True | True | False | False | |
| 0.004934 | 0.005299 | 4.552 | 0.97 | 2.806 | 2.958 | True | True | False | False | |
| 0.004959 | 0.005312 | 6.338 | 1.18 | 3.155 | 6.038 | True | True | False | False | |
| 0.005133 | 0.0055 | 5.592 | 0.9297 | 3.113 | 4.952 | True | True | False | False | |
| 0.005098 | 0.005465 | 5.642 | 1.014 | 2.783 | 5.706 | True | True | False | False | |
| 0.00509 | 0.005457 | 5.539 | 0.9915 | 2.728 | 5.611 | True | True | False | False | |
| 0.00509 | 0.005457 | 5.539 | 0.9946 | 2.751 | 5.624 | True | True | False | False | |
| 0.004833 | 0.005182 | 4.956 | 1.317 | 2.849 | 5.381 | True | True | False | False | |
| 0.004848 | 0.005202 | 4.275 | 1.541 | 2.892 | 5.153 | True | True | False | False | |
| 0.00485 | 0.005202 | 4.939 | 1.519 | 2.919 | 5.186 | True | True | False | False | |
| 0.00509 | 0.005457 | 5.682 | 0.9708 | 2.696 | 5.582 | True | True | False | False | |
| 0.004853 | 0.005205 | 4.923 | 1.498 | 2.929 | 5.044 | True | True | False | False | |
| 0.005094 | 0.005461 | 5.508 | 0.9884 | 2.744 | 5.415 | True | True | False | False | |
| 0.005087 | 0.005454 | 5.378 | 0.943 | 2.82 | 5.435 | True | True | False | False | |
| 0.005217 | 0.005596 | 6.376 | 0.645 | 3.009 | 6.492 | True | True | False | False | |
| 0.004552 | 0.004881 | 2.801 | 0.05681 | 0.2145 | 0.1886 | False | True | False | True | |

| SE_UKB_female | SE_UKB_male | LOG10P_VUMC_female | LOG10P_VUMC_male | LOG10P_UKB_female | LOG10P_UKB_male | female_effect_direction_replication | male_effect_direction_replication | sex_effect_comparison_replication | pvalue_effect_comparison_replication | replication_type |
|---|---|---|---|---|---|---|---|---|---|---|
| 0.005256 | 0.005639 | 6.451 | 0.7699 | 2.83 | 6.189 | True | True | False | False | |
| 0.004581 | 0.004911 | 2.933 | 0.09776 | 0.4742 | 0.2641 | False | True | False | True | |
| 0.00458 | 0.004909 | 2.958 | 0.08014 | 0.4892 | 0.2526 | False | True | False | True | |
| 0.00458 | 0.00491 | 2.883 | 0.05718 | 0.4502 | 0.2586 | False | True | False | True | |
| 0.00458 | 0.00491 | 2.887 | 0.05679 | 0.4537 | 0.2596 | False | True | False | True | |
| 0.004593 | 0.004924 | 2.884 | 0.06739 | 0.4412 | 0.2536 | False | True | False | True | |
| 0.00458 | 0.00491 | 2.887 | 0.05681 | 0.4508 | 0.259 | False | True | False | True | |
| 0.00458 | 0.00491 | 2.886 | 0.05692 | 0.4601 | 0.2578 | False | True | False | True | |
| 0.00458 | 0.00491 | 2.866 | 0.04642 | 0.4508 | 0.259 | False | True | False | True | |
| 0.004582 | 0.004911 | 2.873 | 0.0464 | 0.4383 | 0.2414 | False | True | False | True | |
| 0.00458 | 0.004911 | 2.873 | 0.0464 | 0.4576 | 0.2689 | False | True | False | True | |
| 0.00458 | 0.004911 | 2.873 | 0.0464 | 0.458 | 0.2688 | False | True | False | True | |
| 0.004581 | 0.00491 | 2.87 | 0.0467 | 0.4745 | 0.2906 | False | True | False | True | |
| 0.004575 | 0.004907 | 2.87 | 0.0467 | 0.431 | 0.2658 | False | True | False | True | |
| 0.004581 | 0.004911 | 2.867 | 0.04436 | 0.4525 | 0.2626 | False | True | False | True | |
| 0.004581 | 0.004911 | 2.867 | 0.04436 | 0.4579 | 0.2619 | False | True | False | True | |
| 0.005202 | 0.005577 | 6.058 | 0.7205 | 2.769 | 6.485 | True | True | False | False | |
| 0.004582 | 0.004911 | 2.89 | 0.03049 | 0.486 | 0.2551 | False | True | False | True | |
| 0.005242 | 0.005619 | 5.705 | 0.4596 | 3.008 | 6.354 | True | True | False | False | |
| 0.004582 | 0.004911 | 2.889 | 0.0318 | 0.4698 | 0.2559 | False | True | False | True | |
| 0.004582 | 0.004911 | 2.889 | 0.0318 | 0.4698 | 0.2559 | False | True | False | True | |
| 0.004582 | 0.004911 | 2.85 | 0.02938 | 0.4723 | 0.2581 | False | True | False | True | |
| 0.004587 | 0.004917 | 2.944 | 0.03942 | 0.4129 | 0.1692 | False | True | False | True | |
| 0.004587 | 0.004916 | 2.944 | 0.03939 | 0.4056 | 0.1734 | False | True | False | True | |
| 0.004587 | 0.004917 | 2.956 | 0.03738 | 0.4069 | 0.1663 | False | True | False | True | |
| 0.004587 | 0.004916 | 2.96 | 0.03736 | 0.4056 | 0.1734 | False | True | False | True | |
| 0.004587 | 0.004916 | 2.959 | 0.03742 | 0.4033 | 0.1723 | False | True | False | True | |
| 0.004587 | 0.004917 | 2.96 | 0.03736 | 0.4072 | 0.1769 | False | True | False | True | |

| SE_UKB_female | SE_UKB_male | LOG10P_VUMC_female | LOG10P_VUMC_male | LOG10P_UKB_female | LOG10P_UKB_male | female_effect_direction_replication | male_effect_direction_replication | sex_effect_comparison_replication | pvalue_effect_comparison_replication | replication_type |
|---|---|---|---|---|---|---|---|---|---|---|
| 0.004587 | 0.004916 | 2.96 | 0.03736 | 0.4052 | 0.1735 | False | True | False | True | |
| 0.004587 | 0.004916 | 2.962 | 0.03704 | 0.4056 | 0.1734 | False | True | False | True | |
| 0.004586 | 0.004916 | 2.985 | 0.01632 | 0.3837 | 0.1727 | False | True | False | True | |
| 0.004588 | 0.004917 | 2.96 | 0.0376 | 0.4035 | 0.1717 | False | True | False | True | |
| 0.005228 | 0.005604 | 6.554 | 0.713 | 3.105 | 7.225 | True | True | False | False | |
| 0.004587 | 0.004916 | 2.941 | 0.04159 | 0.4051 | 0.1741 | False | True | False | True | |
| 0.00458 | 0.004909 | 3.012 | 0.03621 | 0.3943 | 0.1349 | False | True | False | True | |
| 0.004586 | 0.004916 | 3.012 | 0.03621 | 0.4094 | 0.1658 | False | True | False | True | |
| 0.004586 | 0.004916 | 3.012 | 0.03621 | 0.4094 | 0.1658 | False | True | False | True | |
| 0.004586 | 0.004916 | 3.012 | 0.03621 | 0.4094 | 0.1658 | False | True | False | True | |
| 0.004588 | 0.004917 | 3.08 | 0.02525 | 0.4217 | 0.1619 | False | True | False | True | |
| 0.0046 | 0.004931 | 3.062 | 0.05682 | 0.51 | 0.2629 | False | True | False | True | |
| 0.0046 | 0.004931 | 3.062 | 0.05683 | 0.5104 | 0.2638 | False | True | False | True | |
| 0.005234 | 0.005614 | 6.557 | 0.6238 | 2.942 | 6.947 | True | True | False | False | |

| SE_UKB_female | SE_UKB_male | LOG10P_VUMC_female | LOG10P_VUMC_male | LOG10P_UKB_female | LOG10P_UKB_male | female_effect_direction_replication | male_effect_direction_replication | sex_effect_comparison_replication | pvalue_effect_comparison_replication | replication_type |
|---|---|---|---|---|---|---|---|---|---|---|
| 0.0046 | 0.004931 | 3.062 | 0.05683 | 0.5115 | 0.2638 | False | True | False | True | |
| 0.0046 | 0.004931 | 3.062 | 0.05683 | 0.5104 | 0.2638 | False | True | False | True | |
| 0.0046 | 0.004931 | 3.062 | 0.05683 | 0.511 | 0.2711 | False | True | False | True | |
| 0.0046 | 0.004931 | 3.062 | 0.05683 | 0.5104 | 0.2638 | False | True | False | True | |
| 0.004599 | 0.004932 | 3.059 | 0.0569 | 0.5615 | 0.2753 | False | True | False | True | |
| 0.0046 | 0.004931 | 3.062 | 0.05683 | 0.5104 | 0.2638 | False | True | False | True | |
| 0.0046 | 0.004931 | 3.062 | 0.05709 | 0.5112 | 0.2649 | False | True | False | True | |
| 0.0046 | 0.004931 | 3.067 | 0.05691 | 0.51 | 0.261 | False | True | False | True | |
| 0.0046 | 0.004931 | 3.062 | 0.05709 | 0.5104 | 0.2638 | False | True | False | True | |
| 0.0046 | 0.004931 | 3.062 | 0.05683 | 0.5094 | 0.2638 | False | True | False | True | |
| 0.005248 | 0.005628 | 6.739 | 0.689 | 2.836 | 6.797 | True | True | False | False | |

| SE_UKB_female | SE_UKB_male | LOG10P_VUMC_female | LOG10P_VUMC_male | LOG10P_UKB_female | LOG10P_UKB_male | female_effect_direction_replication | male_effect_direction_replication | sex_effect_comparison_replication | pvalue_effect_comparison_replication | replication_type |
|---|---|---|---|---|---|---|---|---|---|---|
| 0.0046 | 0.004931 | 3.063 | 0.06228 | 0.5104 | 0.2638 | False | True | False | True | |
| 0.0046 | 0.004931 | 3.063 | 0.06201 | 0.5104 | 0.2638 | False | True | False | True | |
| 0.0046 | 0.004931 | 3.063 | 0.06201 | 0.5094 | 0.2638 | False | True | False | True | |
| 0.0046 | 0.004931 | 3.063 | 0.06228 | 0.5094 | 0.2638 | False | True | False | True | |
| 0.005248 | 0.005628 | 6.737 | 0.6871 | 2.836 | 6.797 | True | True | False | False | |
| 0.0046 | 0.004931 | 3.063 | 0.06228 | 0.5094 | 0.2638 | False | True | False | True | |
| 0.0046 | 0.004931 | 3.063 | 0.06228 | 0.5094 | 0.2638 | False | True | False | True | |
| 0.0046 | 0.004931 | 3.059 | 0.06251 | 0.5144 | 0.2637 | False | True | False | True | |
| | | 3.063 | 0.06228 | | | False | False | False | False | |
| 0.0046 | 0.004931 | 3.064 | 0.06229 | 0.5149 | 0.2626 | False | True | False | True | |
| 0.0046 | 0.004932 | 3.085 | 0.05268 | 0.5211 | 0.2727 | False | True | False | True | |
| 0.004594 | 0.004925 | 3.126 | 0.0485 | 0.4915 | 0.3142 | False | True | False | True | |
| 0.004594 | 0.004924 | 3.127 | 0.04955 | 0.4805 | 0.2973 | False | True | False | True | |
| 0.004594 | 0.004924 | 3.126 | 0.04956 | 0.4803 | 0.3036 | False | True | False | True | |
| 0.004594 | 0.004924 | 3.241 | 0.03598 | 0.5252 | 0.3018 | False | True | False | True | |
| 0.004594 | 0.004924 | 3.239 | 0.03592 | 0.4935 | 0.2848 | False | True | False | True | |
| 0.004584 | 0.004913 | 3.14 | 0.04735 | 0.2916 | 0.2341 | False | True | False | True | |
| 0.004594 | 0.004924 | 3.15 | 0.05185 | 0.4805 | 0.2973 | False | True | False | True | |
| 0.004594 | 0.004924 | 3.093 | 0.05341 | 0.4818 | 0.2952 | False | True | False | True | |
| 0.005242 | 0.00562 | 6.748 | 0.695 | 2.901 | 6.676 | True | True | False | False | |
| 0.005242 | 0.00562 | 6.748 | 0.6963 | 2.901 | 6.676 | True | True | False | False | |
| 0.004594 | 0.004924 | 3.15 | 0.05185 | 0.4844 | 0.2974 | False | True | False | True | |
| 0.004595 | 0.004924 | 3.25 | 0.04017 | 0.4941 | 0.2906 | False | True | False | True | |
| 0.004593 | 0.004924 | 3.15 | 0.05185 | 0.4731 | 0.2999 | False | True | False | True | |
| 0.004594 | 0.004924 | 3.15 | 0.05185 | 0.4845 | 0.2973 | False | True | False | True | |
| 0.005243 | 0.005621 | 6.747 | 0.6963 | 2.805 | 6.787 | True | True | False | False | |
| 0.004594 | 0.004924 | 3.149 | 0.05186 | 0.4827 | 0.3013 | False | True | False | True | |

| SE_UKB_female | SE_UKB_male | LOG10P_VUMC_female | LOG10P_VUMC_male | LOG10P_UKB_female | LOG10P_UKB_male | female_effect_direction_replication | male_effect_direction_replication | sex_effect_comparison_replication | pvalue_effect_comparison_replication | replication_type |
|---|---|---|---|---|---|---|---|---|---|---|
| 0.004559 | 0.004885 | 3.15 | 0.05187 | 0.4251 | 0.2567 | False | True | False | True | |
| 0.004592 | 0.004922 | 3.126 | 0.04616 | 0.5228 | 0.3107 | False | True | False | True | |
| 0.005236 | 0.005613 | 6.714 | 0.6333 | 2.727 | 6.557 | True | True | False | False | |
| 0.005241 | 0.005619 | 6.705 | 0.6337 | 2.912 | 6.652 | True | True | False | False | |
| 0.004595 | 0.004925 | 3.099 | 0.07983 | 0.483 | 0.2885 | False | True | False | True | |
| 0.004594 | 0.004925 | 3.099 | 0.07979 | 0.4862 | 0.2853 | False | True | False | True | |
| 0.005298 | 0.005678 | 6.421 | 0.5364 | 3.008 | 6.762 | True | True | False | False | |
| 0.004594 | 0.004925 | 3.099 | 0.07979 | 0.4862 | 0.2853 | False | True | False | True | |
| 0.004595 | 0.004926 | 3.132 | 0.08134 | 0.5033 | 0.2945 | False | True | False | True | |
| 0.004595 | 0.004924 | 3.147 | 0.04741 | 0.4782 | 0.2506 | False | True | False | True | |
| 0.004822 | 0.005174 | 3.317 | 0.06756 | 0.9772 | 0.5515 | False | True | False | True | |
| 0.004775 | 0.005121 | 3.803 | 0.1348 | 0.4405 | 0.2526 | False | True | False | True | |
| 0.004793 | 0.005139 | 3.478 | 0.1074 | 0.5612 | 0.265 | False | True | False | True | |
| 0.004346 | 0.004654 | 4.377 | 0.3891 | 0.05606 | 0.2232 | True | True | False | False | |
| 0.004773 | 0.005118 | 3.841 | 0.1204 | 0.4366 | 0.2637 | False | True | False | True | |
| 0.004773 | 0.005118 | 3.841 | 0.1155 | 0.4323 | 0.2629 | False | True | False | True | |
| 0.004772 | 0.005118 | 3.821 | 0.1058 | 0.4424 | 0.2658 | False | True | False | True | |
| 0.004772 | 0.005118 | 3.828 | 0.1096 | 0.4396 | 0.268 | False | True | False | True | |
| 0.006683 | 0.007174 | 3.682 | 0.601 | 1.12 | 1.98 | True | True | False | False | |
| 0.004669 | 0.005011 | 3.553 | 0.05564 | 0.3479 | 0.5486 | False | True | True | False | |
| 0.004794 | 0.00514 | 3.44 | 0.09083 | 0.5197 | 0.1892 | False | True | False | True | |
| 0.005417 | 0.00581 | 7.032 | 0.6474 | 2.457 | 5.794 | True | True | False | False | |

| SE_UKB_female | SE_UKB_male | LOG10P_VUMC_female | LOG10P_VUMC_male | LOG10P_UKB_female | LOG10P_UKB_male | female_effect_direction_replication | male_effect_direction_replication | sex_effect_comparison_replication | pvalue_effect_comparison_replication | replication_type |
|---|---|---|---|---|---|---|---|---|---|---|
| 0.005418 | 0.005812 | 7.043 | 0.6469 | 2.448 | 5.825 | True | True | False | False | |
| 0.005447 | 0.005842 | 6.826 | 0.6922 | 2.519 | 5.919 | True | True | False | False | |
| 0.005412 | 0.005806 | 7.107 | 0.6293 | 2.585 | 5.799 | True | True | False | False | |
| | | 6.82 | 0.6968 | | | False | False | False | False | |

| SE_UKB_female | SE_UKB_male | LOG10P_VUMC_female | LOG10P_VUMC_male | LOG10P_UKB_female | LOG10P_UKB_male | female_effect_direction_replication | male_effect_direction_replication | sex_effect_comparison_replication | pvalue_effect_comparison_replication | replication_type |
|---|---|---|---|---|---|---|---|---|---|---|
| 0.005415 | 0.005806 | 7.027 | 0.6381 | 2.456 | 5.82 | True | True | False | False | |
| 0.005421 | 0.005813 | 7.093 | 0.651 | 2.52 | 5.961 | True | True | False | False | |
| 0.006692 | 0.00718 | 3.989 | 0.5254 | 0.9844 | 2.14 | True | True | False | False | |
| 0.005412 | 0.005802 | 7.302 | 0.71 | 2.615 | 6.058 | True | True | False | False | |
| 0.005414 | 0.005804 | 7.367 | 0.7039 | 2.658 | 6.075 | True | True | False | False | |
| 0.004224 | 0.004513 | 5.983 | 0.2275 | 1.312 | 1.984 | True | True | False | False | |
| 0.004238 | 0.004528 | 6.222 | 0.2511 | 1.565 | 2.184 | True | True | False | False | |
| 0.005414 | 0.005803 | 7.055 | 0.6861 | 2.593 | 6.174 | True | True | False | False | |
| 0.005416 | 0.005807 | 6.925 | 0.6897 | 2.626 | 6.006 | True | True | False | False | |
| 0.005416 | 0.005807 | 6.925 | 0.6905 | 2.64 | 6.073 | True | True | False | False | |
| 0.005416 | 0.005805 | 6.821 | 0.6649 | 2.614 | 5.917 | True | True | False | False | |
| 0.005417 | 0.005807 | 6.886 | 0.6658 | 2.614 | 5.977 | True | True | False | False | |
| 0.005421 | 0.005813 | 6.881 | 0.6365 | 2.628 | 5.816 | True | True | False | False | |
| 0.003392 | 0.00364 | 5.786 | 1.297 | 10.69 | 22.3 | True | True | False | False | |
| 0.003392 | 0.00364 | 5.74 | 1.293 | 10.67 | 22.27 | True | True | False | False | |
| 0.003392 | 0.00364 | 5.786 | 1.297 | 10.7 | 22.31 | True | True | False | False | |
| 0.003392 | 0.003638 | 5.755 | 1.298 | 10.63 | 22.17 | True | True | False | False | |
| 0.003392 | 0.003638 | 5.898 | 1.434 | 10.72 | 22.41 | True | True | False | False | |
| 0.003392 | 0.003639 | 5.854 | 1.429 | 10.73 | 22.65 | True | True | False | False | |
| 0.003391 | 0.003638 | 5.822 | 1.429 | 10.7 | 22.17 | True | True | False | False | |
| 0.003392 | 0.003638 | 5.784 | 1.414 | 10.74 | 22.32 | True | True | False | False | |
| 0.003391 | 0.003638 | 5.853 | 1.426 | 10.69 | 22.28 | True | True | False | False | |
| 0.003391 | 0.003639 | 5.533 | 1.352 | 11.02 | 22.82 | True | True | False | False | |
| 0.003392 | 0.003638 | 5.73 | 1.247 | 10.73 | 22.38 | True | True | False | False | |
| 0.003392 | 0.003638 | 5.73 | 1.247 | 10.75 | 22.42 | True | True | False | False | |
| 0.003392 | 0.003638 | 5.73 | 1.243 | 10.76 | 22.42 | True | True | False | False | |
| 0.003393 | 0.003639 | 5.659 | 1.243 | 10.79 | 22.39 | True | True | False | False | |
| 0.003392 | 0.003638 | 5.695 | 1.246 | 10.79 | 22.43 | True | True | False | False | |

| SE_UKB_female | SE_UKB_male | LOG10P_VUMC_female | LOG10P_VUMC_male | LOG10P_UKB_female | LOG10P_UKB_male | female_effect_direction_replication | male_effect_direction_replication | sex_effect_comparison_replication | pvalue_effect_comparison_replication | replication_type |
|---|---|---|---|---|---|---|---|---|---|---|
| 0.003392 | 0.003638 | 5.688 | 1.247 | 10.82 | 22.47 | True | True | False | False | |
| 0.003392 | 0.003638 | 5.681 | 1.243 | 10.8 | 22.41 | True | True | False | False | |
| 0.003392 | 0.003638 | 5.679 | 1.244 | 10.79 | 22.41 | True | True | False | False | |
| 0.003412 | 0.003663 | 5.526 | 0.97 | 10.46 | 19.12 | True | True | False | False | |
| 0.003392 | 0.003639 | 5.578 | 1.188 | 10.79 | 22.77 | True | True | False | False | |
| 0.003395 | 0.003642 | 5.601 | 1.192 | 10.89 | 22.92 | True | True | False | False | |
| 0.003393 | 0.003639 | 5.59 | 1.19 | 10.78 | 22.74 | True | True | False | False | |
| 0.003393 | 0.003639 | 5.59 | 1.19 | 10.78 | 22.75 | True | True | False | False | |
| 0.003393 | 0.003639 | 5.588 | 1.19 | 10.76 | 22.83 | True | True | False | False | |
| 0.003393 | 0.003639 | 5.587 | 1.189 | 10.8 | 22.76 | True | True | False | False | |
| 0.003393 | 0.003639 | 5.598 | 1.189 | 10.77 | 22.76 | True | True | False | False | |
| 0.003397 | 0.003644 | 5.594 | 1.189 | 10.78 | 23.49 | True | True | False | False | |
| 0.003413 | 0.003663 | 5.442 | 0.9276 | 10.45 | 19.43 | True | True | False | False | |
| 0.003398 | 0.003644 | 5.975 | 1.175 | 9.873 | 21.75 | True | True | False | False | |
| 0.0034 | 0.003645 | 5.949 | 1.179 | 9.832 | 21.74 | True | True | False | False | |
| | | 7.523 | 1.821 | | | False | False | False | False | |
| | | 7.663 | 1.556 | | | False | False | False | False | |
| | | 6.045 | 1.468 | | | False | False | False | False | |
| | | 7.684 | 1.543 | | | False | False | False | False | |
| 0.003779 | 0.004042 | 7.827 | 1.243 | 9.572 | 24.67 | True | True | False | False | |
| 0.004643 | 0.004955 | 3.902 | 0.8253 | 0.3016 | 4.931 | True | True | False | False | |
| 0.003759 | 0.004022 | 7.891 | 1.472 | 10.02 | 25.25 | True | True | False | False | |
| 0.003759 | 0.004022 | 7.875 | 1.47 | 9.884 | 24.71 | True | True | False | False | |
| 0.003764 | 0.004026 | 7.891 | 1.46 | 10 | 25.22 | True | True | False | False | |
| 0.004058 | 0.004357 | 7.563 | 0.8039 | 34.18 | 36.57 | True | True | False | False | |
| 0.004109 | 0.004411 | 6.913 | 0.6596 | 36.27 | 36.19 | True | True | False | True | |
| 0.004058 | 0.004358 | 7.599 | 0.8098 | 34.1 | 36.15 | True | True | False | False | |
| 0.004272 | 0.004587 | 6.915 | 0.6492 | 38.75 | 41.74 | True | True | False | False | |

| SE_UKB_female | SE_UKB_male | LOG10P_VUMC_female | LOG10P_VUMC_male | LOG10P_UKB_female | LOG10P_UKB_male | female_effect_direction_replication | male_effect_direction_replication | sex_effect_comparison_replication | pvalue_effect_comparison_replication | replication_type |
|---|---|---|---|---|---|---|---|---|---|---|
| 0.004273 | 0.004589 | 7.009 | 0.6468 | 38.73 | 41.5 | True | True | False | False | |
| 0.004272 | 0.004587 | 6.912 | 0.6553 | 38.59 | 41.47 | True | True | False | False | |
| 0.004863 | 0.005223 | 5.571 | 0.6323 | 25.32 | 24.04 | True | True | False | True | |
| 0.004858 | 0.005218 | 5.737 | 0.6522 | 25.31 | 24.11 | True | True | False | True | |
| 0.004858 | 0.005218 | 5.673 | 0.6389 | 25.35 | 24.1 | True | True | False | True | |
| 0.00331 | 0.003548 | 8.763 | 0.9475 | 3.473 | 8.086 | True | True | False | False | |
| 0.003311 | 0.003548 | 8.795 | 0.9498 | 3.482 | 8.036 | True | True | False | False | |
| 0.003311 | 0.003548 | 8.77 | 0.9458 | 3.484 | 8.042 | True | True | False | False | |
| 0.003264 | 0.003498 | 7.863 | 0.5762 | 1.446 | 4.016 | True | True | False | False | |
| 0.003265 | 0.003498 | 7.848 | 0.5722 | 1.442 | 4.075 | True | True | False | False | |
| 0.003387 | 0.003629 | 11.5 | 1.079 | 5.041 | 7.615 | True | True | False | False | |
| 0.003303 | 0.003541 | 8.824 | 1.056 | 3.073 | 7.081 | True | True | False | False | |
| 0.003265 | 0.003498 | 7.835 | 0.5742 | 1.435 | 4.061 | True | True | False | False | |
| | | 11.3 | 1.021 | | | False | False | False | False | |
| 0.003311 | 0.003548 | 8.733 | 0.936 | 3.468 | 8.03 | True | True | False | False | |
| 0.003387 | 0.003629 | 11.37 | 1.018 | 5.173 | 7.425 | True | True | False | False | |
| 0.003311 | 0.003548 | 8.826 | 0.945 | 3.411 | 7.994 | True | True | False | False | |
| 0.003265 | 0.003499 | 8.001 | 0.4857 | 1.308 | 3.943 | True | True | False | False | |
| 0.003313 | 0.00355 | 8.901 | 0.9768 | 3.687 | 8.107 | True | True | False | False | |
| 0.003265 | 0.003499 | 7.879 | 0.5817 | 1.405 | 4.031 | True | True | False | False | |
| 0.003446 | 0.003704 | 7.738 | 0.4341 | 48 | 37.37 | True | True | True | True | Sex-specific 1e-2 VUMC, sex-differential UKB |
| 0.003445 | 0.003703 | 7.809 | 0.4197 | 47.8 | 37.32 | True | True | True | True | Sex-specific 1e-2 VUMC, sex-differential UKB |
| 0.003518 | 0.003783 | 6.617 | 0.1298 | 40.15 | 36.37 | True | False | False | True | |
| 0.003438 | 0.003697 | 7.49 | 0.4355 | 48.83 | 37.99 | True | True | True | True | Sex-specific 1e-2 VUMC, sex-differential UKB |
| 0.003438 | 0.003696 | 7.498 | 0.425 | 48.77 | 37.96 | True | True | True | True | Sex-specific 1e-2 VUMC, sex-differential UKB |
| 0.003512 | 0.003778 | 6.436 | 0.1109 | 40.92 | 36.81 | True | False | False | True | |
| 0.003438 | 0.003696 | 7.489 | 0.4314 | 48.79 | 38.12 | True | True | True | True | Sex-specific 1e-2 VUMC, sex-differential UKB |

| SE_UKB_female | SE_UKB_male | LOG10P_VUMC_female | LOG10P_VUMC_male | LOG10P_UKB_female | LOG10P_UKB_male | female_effect_direction_replication | male_effect_direction_replication | sex_effect_comparison_replication | pvalue_effect_comparison_replication | replication_type |
|---|---|---|---|---|---|---|---|---|---|---|
| 0.003438 | 0.003696 | 7.498 | 0.4247 | 48.96 | 38.08 | True | True | True | True | Sex-specific 1e-2 VUMC, sex-differential UKB |
| 0.003438 | 0.003696 | 7.506 | 0.4295 | 48.92 | 38.13 | True | True | True | True | Sex-specific 1e-2 VUMC, sex-differential UKB |
| 0.003438 | 0.003696 | 7.5 | 0.4243 | 48.91 | 38.13 | True | True | True | True | Sex-specific 1e-2 VUMC, sex-differential UKB |
| 0.00344 | 0.003697 | 7.738 | 0.3945 | 48.36 | 37.92 | True | True | True | True | Sex-specific 1e-2 VUMC, sex-differential UKB |
| 0.00344 | 0.003697 | 7.701 | 0.3836 | 48.3 | 37.66 | True | True | True | True | Sex-specific 1e-2 VUMC, sex-differential UKB |
| 0.003487 | 0.00375 | 5.749 | 0.1665 | 42.26 | 31.64 | True | True | True | True | Sex-specific 1e-2 VUMC, sex-differential UKB |
| 0.003481 | 0.003744 | 5.074 | 0.1665 | 43.05 | 31.32 | True | True | True | True | Sex-specific 1e-2 VUMC, sex-differential UKB |
| 0.00358 | 0.00385 | 4.579 | 0.2914 | 35 | 30.74 | True | False | False | True | |
| 0.003569 | 0.00384 | 4.476 | 0.3906 | 35.79 | 30.43 | True | False | True | True | Sex-specific 1e-2 VUMC, sex-differential UKB |
| 0.003448 | 0.003708 | 7.262 | 0.3935 | 49.9 | 37.54 | True | True | True | True | Sex-specific 1e-2 VUMC, sex-differential UKB |
| 0.003492 | 0.003756 | 4.941 | 0.2256 | 43.19 | 31.41 | True | True | True | True | Sex-specific 1e-2 VUMC, sex-differential UKB |
| 0.003492 | 0.003755 | 5.087 | 0.1728 | 43.22 | 31.67 | True | True | True | True | Sex-specific 1e-2 VUMC, sex-differential UKB |
| 0.003605 | 0.003875 | 4.791 | 0.4026 | 36.2 | 32.35 | True | False | False | True | |
| 0.003602 | 0.00387 | 5.166 | 0.4698 | 36.21 | 31.99 | True | False | False | True | |
| 0.003787 | 0.004066 | 7.338 | 0.27 | 47.03 | 39.02 | True | False | True | True | Sex-specific 1e-2 VUMC, sex-differential UKB |
| 0.003593 | 0.003857 | 10.42 | 0.6598 | 64.09 | 45.42 | True | True | True | True | Sex-specific 1e-2 VUMC, sex-differential UKB |
| 0.003311 | 0.003565 | 5.351 | 0.03345 | 27.3 | 12.47 | True | True | True | True | Sex-specific 1e-2 VUMC, sex-differential UKB |
| 0.003592 | 0.003855 | 10.42 | 0.4275 | 63.71 | 45.21 | True | True | True | True | Sex-specific 1e-2 VUMC, sex-differential UKB |
| 0.003337 | 0.003576 | 7.517 | 1.595 | 14.54 | 11.15 | True | True | True | True | Sex-specific 1e-2 VUMC, sex-differential UKB |
| 0.003336 | 0.003575 | 7.55 | 1.629 | 14.75 | 11.12 | True | True | True | True | Sex-specific 1e-2 VUMC, sex-differential UKB |
| 0.003355 | 0.003597 | 7.186 | 1.382 | 15.27 | 11.14 | True | True | True | True | Sex-specific 1e-2 VUMC, sex-differential UKB |
| 0.003292 | 0.003527 | 6.935 | 1.484 | 10.81 | 9.003 | True | True | True | True | Sex-specific 1e-2 VUMC, sex-differential UKB |
| 0.003351 | 0.00359 | 6.867 | 1.637 | 13.98 | 10.68 | True | True | True | True | Sex-specific 1e-2 VUMC, sex-differential UKB |
| 0.003346 | 0.003585 | 6.934 | 1.594 | 13.87 | 10.73 | True | True | True | True | Sex-specific 1e-2 VUMC, sex-differential UKB |
| 0.003361 | 0.003604 | 6.483 | 1.397 | 14.67 | 10.2 | True | True | True | True | Sex-specific 1e-2 VUMC, sex-differential UKB |
| 0.00336 | 0.003601 | 6.311 | 1.325 | 14.59 | 10.06 | True | True | True | True | Sex-specific 1e-2 VUMC, sex-differential UKB |
| 0.003353 | 0.003591 | 7.245 | 1.127 | 14.54 | 8.919 | True | True | True | True | Sex-specific 1e-2 VUMC, sex-differential UKB |

| SE_UKB_female | SE_UKB_male | LOG10P_VUMC_female | LOG10P_VUMC_male | LOG10P_UKB_female | LOG10P_UKB_male | female_effect_direction_replication | male_effect_direction_replication | sex_effect_comparison_replication | pvalue_effect_comparison_replication | replication_type |
|---|---|---|---|---|---|---|---|---|---|---|
| 0.003374 | 0.003616 | 6.835 | 0.9484 | 15.12 | 8.801 | True | True | True | True | Sex-specific 1e-2 VUMC, sex-differential UKB |
| 0.003352 | 0.003589 | 7.17 | 1.137 | 14.61 | 8.883 | True | True | True | True | Sex-specific 1e-2 VUMC, sex-differential UKB |
| 0.003374 | 0.003616 | 6.865 | 1.002 | 15.09 | 8.761 | True | True | True | True | Sex-specific 1e-2 VUMC, sex-differential UKB |
| 0.003375 | 0.003616 | 6.798 | 0.9434 | 15.21 | 8.535 | True | True | True | True | Sex-specific 1e-2 VUMC, sex-differential UKB |
| 0.003375 | 0.003617 | 6.805 | 0.9646 | 15.09 | 8.336 | True | True | True | True | Sex-specific 1e-2 VUMC, sex-differential UKB |
| 0.003352 | 0.003589 | 7.13 | 1.13 | 14.72 | 8.622 | True | True | True | True | Sex-specific 1e-2 VUMC, sex-differential UKB |
| SE_UKB_female | SE_UKB_male | LOG10P_VUMC_female | LOG10P_VUMC_male | LOG10P_UKB_female | LOG10P_UKB_male | female_effect_direction_replication | male_effect_direction_replication | sex_effect_comparison_replication | pvalue_effect_comparison_replication | replication_type |
| 0.003304 | 0.003537 | 7.114 | 1.22 | 11.14 | 7.131 | True | True | True | True | Sex-specific 1e-2 VUMC, sex-specific 5e-8 UKB |
| 0.003351 | 0.00359 | 7.085 | 1.154 | 14.78 | 8.481 | True | True | True | True | Sex-specific 1e-2 VUMC, sex-differential UKB |
| 0.003375 | 0.003617 | 6.843 | 1.035 | 14.72 | 8.398 | True | True | True | True | Sex-specific 1e-2 VUMC, sex-differential UKB |
| 0.003375 | 0.003617 | 6.745 | 1.048 | 14.75 | 8.446 | True | True | True | True | Sex-specific 1e-2 VUMC, sex-differential UKB |
| 0.003375 | 0.003616 | 6.91 | 1.011 | 14.68 | 8.56 | True | True | True | True | Sex-specific 1e-2 VUMC, sex-differential UKB |
| 0.003375 | 0.003617 | 6.629 | 1.023 | 14.67 | 8.524 | True | True | True | True | Sex-specific 1e-2 VUMC, sex-differential UKB |
| 0.003375 | 0.003616 | 6.651 | 1.039 | 14.67 | 8.573 | True | True | True | True | Sex-specific 1e-2 VUMC, sex-differential UKB |
| 0.003375 | 0.003616 | 6.647 | 1.036 | 14.62 | 8.582 | True | True | True | True | Sex-specific 1e-2 VUMC, sex-differential UKB |
| 0.003375 | 0.003616 | 6.643 | 1.038 | 14.63 | 8.579 | True | True | True | True | Sex-specific 1e-2 VUMC, sex-differential UKB |
| 0.003375 | 0.003617 | 6.577 | 1.051 | 14.65 | 8.629 | True | True | True | True | Sex-specific 1e-2 VUMC, sex-differential UKB |
| 0.003375 | 0.003616 | 6.582 | 1.052 | 14.65 | 8.645 | True | True | True | True | Sex-specific 1e-2 VUMC, sex-differential UKB |
| 0.003374 | 0.003617 | 6.625 | 1.032 | 14.55 | 8.421 | True | True | True | True | Sex-specific 1e-2 VUMC, sex-differential UKB |
| 0.003375 | 0.003617 | 6.549 | 1.041 | 14.58 | 8.455 | True | True | True | True | Sex-specific 1e-2 VUMC, sex-differential UKB |
| 0.003304 | 0.003538 | 6.895 | 1.329 | 10.6 | 7.332 | True | True | True | True | Sex-specific 1e-2 VUMC, sex-differential UKB |
| 0.003305 | 0.003538 | 6.879 | 1.39 | 10.57 | 7.297 | True | True | True | True | Sex-specific 1e-2 VUMC, sex-specific 5e-8 UKB |
| 0.003375 | 0.003616 | 6.468 | 1.052 | 14.55 | 8.671 | True | True | True | True | Sex-specific 1e-2 VUMC, sex-differential UKB |
| 0.003374 | 0.003616 | 6.499 | 1.061 | 14.44 | 8.626 | True | True | True | True | Sex-specific 1e-2 VUMC, sex-differential UKB |
| 0.003374 | 0.003616 | 6.442 | 0.9719 | 14.48 | 8.468 | True | True | True | True | Sex-specific 1e-2 VUMC, sex-differential UKB |
| 0.003372 | 0.003614 | 6.27 | 0.9761 | 14.45 | 8.42 | True | True | True | True | Sex-specific 1e-2 VUMC, sex-differential UKB |

| SE_UKB_female | SE_UKB_male | LOG10P_VUMC_female | LOG10P_VUMC_male | LOG10P_UKB_female | LOG10P_UKB_male | female_effect_direction_replication | male_effect_direction_replication | sex_effect_comparison_replication | pvalue_effect_comparison_replication | replication_type |
|---|---|---|---|---|---|---|---|---|---|---|
| 0.00331 | 0.003544 | 6.557 | 1.438 | 10.85 | 7.487 | True | True | True | True | Sex-specific 1e-2 VUMC, sex-differential UKB |
| 0.003379 | 0.003622 | 5.916 | 1.083 | 14.86 | 8.436 | True | True | True | True | Sex-specific 1e-2 VUMC, sex-differential UKB |
| 0.003309 | 0.003542 | 6.543 | 1.415 | 10.39 | 7.028 | True | True | True | True | Sex-specific 1e-2 VUMC, sex-specific 5e-8 UKB |
| 0.003324 | 0.003561 | 6.225 | 1.178 | 10.67 | 6.68 | True | True | True | True | Sex-specific 1e-2 VUMC, sex-specific 5e-8 UKB |
| 0.003336 | 0.003583 | 6.457 | 0.6402 | 0.7417 | 1.294 | True | True | False | False | |
| 0.003336 | 0.003583 | 6.482 | 0.6658 | 0.7444 | 1.299 | True | True | False | False | |
| 0.003327 | 0.003574 | 6.745 | 0.742 | 0.8181 | 1.263 | True | True | False | False | |
| 0.003332 | 0.003579 | 6.541 | 0.6716 | 0.754 | 1.268 | True | True | False | False | |
| 0.003333 | 0.00358 | 6.501 | 0.7494 | 0.759 | 1.282 | True | True | False | False | |
| | | 6.464 | 0.6643 | | | False | False | False | False | |
| 0.003297 | 0.003543 | 6.495 | 0.8644 | 0.6951 | 1.507 | True | True | False | False | |
| 0.003297 | 0.003543 | 6.48 | 0.8635 | 0.6988 | 1.529 | True | True | False | False | |
| 0.003297 | 0.003543 | 6.481 | 0.8628 | 0.7087 | 1.533 | True | True | False | False | |
| 0.003297 | 0.003543 | 6.483 | 0.8629 | 0.7126 | 1.538 | True | True | False | False | |
| 0.003298 | 0.003543 | 6.438 | 0.9725 | 0.693 | 1.554 | True | True | False | False | |
| 0.00329 | 0.003532 | 4.296 | 0.9277 | 0.2922 | 1.109 | True | True | False | False | |
| 0.003299 | 0.003544 | 6.669 | 0.8557 | 0.7114 | 1.527 | True | True | False | False | |
| 0.003301 | 0.003542 | 4.178 | 0.8882 | 0.3149 | 1.117 | True | True | False | False | |
| 0.003302 | 0.003548 | 7.168 | 0.9993 | 0.5233 | 1.327 | True | True | False | False | |
| 0.003301 | 0.003547 | 7.361 | 1.002 | 0.5288 | 1.31 | True | True | False | False | |
| 0.003301 | 0.003547 | 7.361 | 1.002 | 0.5293 | 1.307 | True | True | False | False | |
| 0.003301 | 0.003547 | 7.361 | 1.002 | 0.5295 | 1.306 | True | True | False | False | |
| 0.0033 | 0.003547 | 7.378 | 0.9229 | 0.5411 | 1.323 | True | True | False | False | |
| 0.0033 | 0.003547 | 7.378 | 0.9229 | 0.5424 | 1.319 | True | True | False | False | |
| 0.0033 | 0.003547 | 7.378 | 0.923 | 0.5424 | 1.318 | True | True | False | False | |
| 0.0033 | 0.003547 | 7.378 | 0.9229 | 0.5424 | 1.318 | True | True | False | False | |
| 0.0033 | 0.003547 | 7.378 | 0.9229 | 0.5469 | 1.318 | True | True | False | False | |

| SE_UKB_female | SE_UKB_male | LOG10P_VUMC_female | LOG10P_VUMC_male | LOG10P_UKB_female | LOG10P_UKB_male | female_effect_direction_replication | male_effect_direction_replication | sex_effect_comparison_replication | pvalue_effect_comparison_replication | replication_type |
|---|---|---|---|---|---|---|---|---|---|---|
| 0.0033 | 0.003547 | 7.383 | 0.9137 | 0.5423 | 1.319 | True | True | False | False | |
| 0.0033 | 0.003547 | 7.378 | 0.923 | 0.5424 | 1.318 | True | True | False | False | |
| 0.0033 | 0.003547 | 7.378 | 0.9233 | 0.5428 | 1.318 | True | True | False | False | |
| 0.0033 | 0.003547 | 7.381 | 0.914 | 0.5444 | 1.322 | True | True | False | False | |
| 0.0033 | 0.003547 | 7.377 | 0.9233 | 0.5423 | 1.318 | True | True | False | False | |
| 0.0033 | 0.003547 | 7.377 | 0.9233 | 0.5423 | 1.318 | True | True | False | False | |
| 0.0033 | 0.003547 | 7.377 | 0.9233 | 0.5421 | 1.318 | True | True | False | False | |
| 0.003302 | 0.003549 | 7.483 | 0.923 | 0.5575 | 1.314 | True | True | False | False | |
| 0.003302 | 0.003548 | 7.482 | 0.923 | 0.5471 | 1.302 | True | True | False | False | |
| 0.0033 | 0.003547 | 7.218 | 0.9085 | 0.563 | 1.302 | True | True | False | False | |
| 0.0033 | 0.003547 | 7.374 | 0.9255 | 0.5482 | 1.31 | True | True | False | False | |
| 0.0033 | 0.003547 | 7.376 | 0.9235 | 0.5436 | 1.316 | True | True | False | False | |
| 0.0033 | 0.003547 | 7.377 | 0.9239 | 0.5434 | 1.316 | True | True | False | False | |
| 0.003301 | 0.003547 | 7.351 | 0.8706 | 0.5266 | 1.299 | True | True | False | False | |
| 0.003301 | 0.003547 | 7.394 | 0.8726 | 0.5344 | 1.262 | True | True | False | False | |
| 0.0033 | 0.003547 | 7.369 | 0.9213 | 0.5509 | 1.288 | True | True | False | False | |
| 0.0033 | 0.003546 | 7.369 | 0.9234 | 0.5498 | 1.299 | True | True | False | False | |
| 0.0033 | 0.003547 | 7.379 | 0.9194 | 0.5507 | 1.292 | True | True | False | False | |
| 0.0033 | 0.003546 | 7.373 | 0.9194 | 0.5721 | 1.3 | True | True | False | False | |
| 0.0033 | 0.003546 | 7.332 | 0.9087 | 0.5721 | 1.3 | True | True | False | False | |
| 0.003297 | 0.003543 | 6.487 | 0.8965 | 0.5072 | 1.298 | True | True | False | False | |
| 0.003299 | 0.003545 | 7.369 | 0.9306 | 0.5687 | 1.379 | True | True | False | False | |
| 0.0033 | 0.003546 | 7.378 | 0.9315 | 0.5423 | 1.304 | True | True | False | False | |
| 0.0033 | 0.003546 | 7.363 | 0.9283 | 0.5421 | 1.307 | True | True | False | False | |
| 0.0033 | 0.003547 | 7.384 | 0.9232 | 0.517 | 1.287 | True | True | False | False | |

| SE_UKB_female | SE_UKB_male | LOG10P_VUMC_female | LOG10P_VUMC_male | LOG10P_UKB_female | LOG10P_UKB_male | female_effect_direction_replication | male_effect_direction_replication | sex_effect_comparison_replication | pvalue_effect_comparison_replication | replication_type |
|---|---|---|---|---|---|---|---|---|---|---|
| 0.0033 | 0.003547 | 7.384 | 0.9232 | 0.517 | 1.287 | True | True | False | False | |
| 0.0033 | 0.003547 | 7.384 | 0.9232 | 0.517 | 1.287 | True | True | False | False | |
| 0.0033 | 0.003547 | 7.384 | 0.9232 | 0.517 | 1.287 | True | True | False | False | |
| 0.0033 | 0.003547 | 7.263 | 0.9142 | 0.5394 | 1.294 | True | True | False | False | |
| 0.0033 | 0.003547 | 7.263 | 0.9142 | 0.5394 | 1.294 | True | True | False | False | |
| 0.0033 | 0.003547 | 7.264 | 0.9142 | 0.5415 | 1.292 | True | True | False | False | |
| 0.0033 | 0.003547 | 7.264 | 0.9143 | 0.5437 | 1.287 | True | True | False | False | |
| 0.0033 | 0.003547 | 7.264 | 0.9143 | 0.5414 | 1.294 | True | True | False | False | |
| 0.0033 | 0.003547 | 7.283 | 0.9191 | 0.556 | 1.283 | True | True | False | False | |
| 0.0033 | 0.003547 | 7.268 | 0.9145 | 0.5558 | 1.282 | True | True | False | False | |
| 0.0033 | 0.003547 | 7.276 | 0.9211 | 0.5548 | 1.283 | True | True | False | False | |
| 0.0033 | 0.003547 | 7.283 | 0.9176 | 0.5545 | 1.283 | True | True | False | False | |
| 0.003308 | 0.003554 | 7.303 | 0.93 | 0.5953 | 1.291 | True | True | False | False | |
| 0.003261 | 0.003506 | 5.711 | 1.369 | 1.94 | 2.497 | True | True | False | False | |
| 0.00326 | 0.003507 | 5.033 | 1.305 | 1.838 | 2.498 | True | True | False | False | |
| 0.00326 | 0.003506 | 5.806 | 1.321 | 1.905 | 2.601 | True | True | False | False | |
| 0.003261 | 0.003506 | 5.63 | 1.417 | 1.961 | 2.474 | True | True | False | False | |
| 0.003261 | 0.003506 | 5.643 | 1.421 | 1.964 | 2.474 | True | True | False | False | |
| 0.003263 | 0.003508 | 5.659 | 1.436 | 1.974 | 2.39 | True | True | False | False | |
| 0.003261 | 0.003506 | 5.875 | 1.313 | 1.951 | 2.531 | True | True | False | False | |
| 0.003261 | 0.003506 | 5.775 | 1.352 | 1.988 | 2.427 | True | True | False | False | |
| 0.00326 | 0.003507 | 5.25 | 1.264 | 1.847 | 2.449 | True | True | False | False | |
| 0.00326 | 0.003507 | 5.281 | 1.256 | 1.875 | 2.466 | True | True | False | False | |
| 0.003261 | 0.003506 | 5.794 | 1.334 | 2.037 | 2.421 | True | True | False | False | |
| 0.003261 | 0.003506 | 5.794 | 1.334 | 2.046 | 2.408 | True | True | False | False | |
| 0.003261 | 0.003506 | 5.734 | 1.404 | 2.06 | 2.389 | True | True | False | False | |

| SE_UKB_female | SE_UKB_male | LOG10P_VUMC_female | LOG10P_VUMC_male | LOG10P_UKB_female | LOG10P_UKB_male | female_effect_direction_replication | male_effect_direction_replication | sex_effect_comparison_replication | pvalue_effect_comparison_replication | replication_type |
|---|---|---|---|---|---|---|---|---|---|---|
| 0.003261 | 0.003506 | 5.728 | 1.403 | 2.058 | 2.385 | True | True | False | False | |
| 0.003261 | 0.003506 | 5.787 | 1.334 | 2.046 | 2.408 | True | True | False | False | |
| 0.003261 | 0.003506 | 5.786 | 1.334 | 1.96 | 2.404 | True | True | False | False | |
| 0.003261 | 0.003506 | 5.791 | 1.336 | 1.96 | 2.417 | True | True | False | False | |
| 0.003261 | 0.003506 | 5.816 | 1.327 | 2.029 | 2.417 | True | True | False | False | |
| 0.003261 | 0.003506 | 5.816 | 1.327 | 2.029 | 2.417 | True | True | False | False | |
| 0.003261 | 0.003506 | 5.816 | 1.327 | 2.029 | 2.417 | True | True | False | False | |
| 0.003261 | 0.003506 | 5.816 | 1.327 | 2.032 | 2.409 | True | True | False | False | |
| 0.003261 | 0.003506 | 5.809 | 1.363 | 2.026 | 2.379 | True | True | False | False | |
| 0.003261 | 0.003507 | 5.786 | 1.346 | 2.009 | 2.428 | True | True | False | False | |
| 0.003261 | 0.003507 | 5.789 | 1.344 | 2.008 | 2.429 | True | True | False | False | |
| 0.00326 | 0.003507 | 5.714 | 1.25 | 1.953 | 2.347 | True | True | False | False | |
| 0.00326 | 0.003508 | 5.012 | 1.159 | 1.833 | 2.349 | True | True | False | False | |
| 0.00326 | 0.003507 | 5.701 | 1.255 | 1.956 | 2.33 | True | True | False | False | |
| 0.00326 | 0.003507 | 5.701 | 1.255 | 1.956 | 2.33 | True | True | False | False | |
| 0.00326 | 0.003507 | 5.701 | 1.255 | 1.956 | 2.33 | True | True | False | False | |
| 0.00326 | 0.003507 | 5.701 | 1.255 | 1.956 | 2.33 | True | True | False | False | |
| 0.00326 | 0.003507 | 5.7 | 1.255 | 1.957 | 2.33 | True | True | False | False | |
| 0.003261 | 0.003507 | 5.716 | 1.255 | 1.967 | 2.327 | True | True | False | False | |
| 0.003261 | 0.003507 | 5.718 | 1.255 | 1.962 | 2.312 | True | True | False | False | |
| 0.003262 | 0.003509 | 5.729 | 1.249 | 1.92 | 2.349 | True | True | False | False | |
| 0.00326 | 0.003508 | 5.78 | 1.195 | 1.926 | 2.378 | True | True | False | False | |
| 0.00326 | 0.003507 | 5.887 | 1.112 | 2.025 | 2.446 | True | True | False | False | |
| SE_UKB_female | SE_UKB_male | LOG10P_VUMC_female | LOG10P_VUMC_male | LOG10P_UKB_female | LOG10P_UKB_male | female_effect_direction_replication | male_effect_direction_replication | sex_effect_comparison_replication | pvalue_effect_comparison_replication | replication_type |
| 0.00326 | 0.003508 | 6.015 | 1.13 | 2.057 | 2.445 | True | True | False | False | |
| 0.00326 | 0.003507 | 5.888 | 1.162 | 2.1 | 2.452 | True | True | False | False | |

| SE_UKB_female | SE_UKB_male | LOG10P_VUMC_female | LOG10P_VUMC_male | LOG10P_UKB_female | LOG10P_UKB_male | female_effect_direction_replication | male_effect_direction_replication | sex_effect_comparison_replication | pvalue_effect_comparison_replication | replication_type |
|---|---|---|---|---|---|---|---|---|---|---|
| 0.00326 | 0.003508 | 5.894 | 1.107 | 2.036 | 2.486 | True | True | False | False | |
| 0.00326 | 0.003506 | 5.936 | 1.107 | 2.124 | 2.509 | True | True | False | False | |
| 0.00326 | 0.003507 | 5.895 | 1.107 | 2.073 | 2.515 | True | True | False | False | |
| 0.003447 | 0.003686 | 6.114 | 0.2316 | 27.16 | 33.27 | True | True | False | False | |
| 0.003415 | 0.00365 | 6.268 | 0.2307 | 28.17 | 33.89 | True | True | False | False | |
| 0.003286 | 0.003516 | 10.5 | 1.728 | 63.63 | 63.94 | True | True | False | False | |
| 0.004347 | 0.004645 | 6.865 | 0.904 | 6.582 | 1.089 | True | True | True | True | Sex-specific 1e-2 VUMC, sex-differential UKB |
| 0.004333 | 0.004628 | 6.602 | 0.9516 | 6.355 | 0.9665 | True | True | True | True | Sex-specific 1e-2 VUMC, sex-differential UKB |
| 0.004618 | 0.00494 | 4.515 | 0.2646 | 2.627 | 1.62 | True | True | True | True | Sex-specific 1e-2 VUMC, sex-differential UKB |
| 0.004617 | 0.004939 | 4.493 | 0.2744 | 2.677 | 1.638 | True | True | True | True | Sex-specific 1e-2 VUMC, sex-differential UKB |
| 0.004619 | 0.00494 | 4.466 | 0.2975 | 2.652 | 1.645 | True | True | True | True | Sex-specific 1e-2 VUMC, sex-differential UKB |
| 0.004619 | 0.00494 | 4.538 | 0.3116 | 2.617 | 1.622 | True | True | True | True | Sex-specific 1e-2 VUMC, sex-differential UKB |
| 0.004618 | 0.004939 | 4.46 | 0.3163 | 2.824 | 1.56 | True | True | True | True | Sex-specific 1e-2 VUMC, sex-differential UKB |
| 0.003602 | 0.003852 | 2.628 | 1.3 | 1.765 | 0.1389 | True | True | True | True | Sex-specific 1e-2 VUMC, sex-differential UKB |
| 0.004133 | 0.004424 | 7.61 | 1.416 | 7.904 | 1.399 | True | True | True | True | Sex-specific 1e-2 VUMC & UKB |
| 0.004878 | 0.005227 | 4.537 | 0.6588 | 6.83 | 0.723 | True | True | True | True | Sex-specific 1e-2 VUMC, sex-differential UKB |
| 0.004448 | 0.004769 | 4.871 | 0.8744 | 5.393 | 0.8177 | True | True | True | True | Sex-specific 1e-2 VUMC, sex-differential UKB |
| 0.003788 | 0.004073 | 2.007 | 0.2046 | 1.258 | 0.7116 | True | True | True | True | Sex-specific 1e-2 VUMC, sex-differential UKB |
| 0.004383 | 0.004704 | 7.365 | 0.05118 | 6.971 | 13.4 | True | True | False | False | |
| 0.004398 | 0.004722 | 7.126 | 0.03705 | 6.545 | 13.3 | True | True | False | False | |
| 0.004383 | 0.004704 | 7.345 | 0.05461 | 6.988 | 13.38 | True | True | False | False | |
| 0.004412 | 0.004734 | 6.938 | 0.08775 | 6.509 | 13.07 | True | True | False | False | |
| 0.004382 | 0.004704 | 7.327 | 0.05586 | 6.962 | 13.32 | True | True | False | False | |
| 0.004381 | 0.004703 | 7.323 | 0.04963 | 6.87 | 13.22 | True | True | False | False | |
| 0.004411 | 0.004733 | 6.935 | 0.08097 | 6.426 | 12.97 | True | True | False | False | |
| 0.004383 | 0.004705 | 7.234 | 0.04577 | 6.989 | 13.16 | True | True | False | False | |

| SE_UKB_female | SE_UKB_male | LOG10P_VUMC_female | LOG10P_VUMC_male | LOG10P_UKB_female | LOG10P_UKB_male | female_effect_direction_replication | male_effect_direction_replication | sex_effect_comparison_replication | pvalue_effect_comparison_replication | replication_type |
|---|---|---|---|---|---|---|---|---|---|---|
| 0.004402 | 0.004727 | 6.741 | 0.05822 | 6.923 | 13.46 | True | True | False | False | |
| 0.004421 | 0.004745 | 6.533 | 0.08671 | 6.707 | 12.84 | True | True | False | False | |
| 0.004405 | 0.004732 | 6.717 | 0.05296 | 7.176 | 13.38 | True | True | False | False | |
| 0.00442 | 0.004743 | 6.567 | 0.0879 | 6.844 | 12.92 | True | True | False | False | |
| 0.004406 | 0.004732 | 6.714 | 0.05282 | 7.125 | 13.1 | True | True | False | False | |
| 0.00442 | 0.004743 | 6.569 | 0.08798 | 6.852 | 12.91 | True | True | False | False | |
| 0.00446 | 0.00479 | 6.342 | 0.1534 | 6.696 | 13.5 | True | True | False | False | |
| 0.004461 | 0.004792 | 6.311 | 0.1485 | 6.774 | 13.43 | True | True | False | False | |
| 0.004405 | 0.004731 | 6.717 | 0.04954 | 7.066 | 13.33 | True | True | False | False | |
| 0.004406 | 0.004731 | 6.721 | 0.04741 | 7.028 | 13.3 | True | True | False | False | |
| 0.004406 | 0.004731 | 6.718 | 0.04737 | 7.055 | 13.32 | True | True | False | False | |
| 0.004412 | 0.004738 | 6.715 | 0.03759 | 6.968 | 12.95 | True | True | False | False | |
| 0.004422 | 0.004746 | 6.547 | 0.06964 | 6.686 | 12.88 | True | True | False | False | |
| 0.003513 | 0.00373 | 2.794 | 0.3824 | 5.318 | 4.647 | True | True | True | True | Sex-specific 1e-2 VUMC, sex-differential UKB |
| 0.003513 | 0.00373 | 2.838 | 0.3585 | 5.348 | 4.675 | True | True | True | True | Sex-specific 1e-2 VUMC, sex-differential UKB |
| 0.003512 | 0.003729 | 2.854 | 0.3534 | 5.454 | 4.645 | True | True | True | True | Sex-specific 1e-2 VUMC, sex-differential UKB |
| 0.003289 | 0.003491 | 8.095 | 1.961 | 12.41 | 14.7 | True | True | False | False | |
| | | 4.097 | 1.225 | | | False | False | False | False | |
| | | 4.594 | 1.319 | | | False | False | False | False | |
| 0.00351 | 0.003729 | 2.318 | 0.6202 | 5.043 | 4.958 | True | True | False | True | |
| 0.003509 | 0.003727 | 2.292 | 0.5715 | 5.075 | 5.007 | True | True | False | True | |
| 0.003298 | 0.003498 | 4.115 | 1.302 | 9.119 | 9.133 | True | True | False | False | |
| 0.003301 | 0.003502 | 3.864 | 1.449 | 9.628 | 8.878 | True | True | False | True | |
| | | 3.874 | 0.6541 | | | False | False | False | False | |
| 0.003338 | 0.003552 | 4.079 | 0.6037 | 18.93 | 22.13 | True | True | False | False | |
| 0.003452 | 0.003669 | 3.135 | 0.5952 | 17.57 | 13.88 | True | True | True | True | Sex-specific 1e-2 VUMC, sex-differential UKB |
| | | 3.053 | 0.5317 | | | False | False | False | False | |
| | | 3.11 | 0.5972 | | | False | False | False | False | |
| | | 3.162 | 0.6116 | | | False | False | False | False | |

| SE_UKB_female | SE_UKB_male | LOG10P_VUMC_female | LOG10P_VUMC_male | LOG10P_UKB_female | LOG10P_UKB_male | female_effect_direction_replication | male_effect_direction_replication | sex_effect_comparison_replication | pvalue_effect_comparison_replication | replication_type |
|---|---|---|---|---|---|---|---|---|---|---|
| | | 3.163 | 0.6124 | | | False | False | False | False | |
| | | 3.119 | 0.596 | | | False | False | False | False | |
| | | 3.119 | 0.5959 | | | False | False | False | False | |
| SE_UKB_female | SE_UKB_male | LOG10P_VUMC_female | LOG10P_VUMC_male | LOG10P_UKB_female | LOG10P_UKB_male | female_effect_direction_replication | male_effect_direction_replication | sex_effect_comparison_replication | pvalue_effect_comparison_replication | replication_type |
| | | 3.119 | 0.596 | | | False | False | False | False | |
| | | 3.11 | 0.5972 | | | False | False | False | False | |
| | | 3.119 | 0.5959 | | | False | False | False | False | |
| | | 3.11 | 0.5972 | | | False | False | False | False | |
| | | 3.11 | 0.5972 | | | False | False | False | False | |
| | | 3.12 | 0.5953 | | | False | False | False | False | |
| | | 3.12 | 0.5952 | | | False | False | False | False | |
| | | 3.12 | 0.5952 | | | False | False | False | False | |
| | | 3.11 | 0.5972 | | | False | False | False | False | |
| 0.003451 | 0.003669 | 3.11 | 0.5972 | 17.5 | 13.79 | True | True | True | True | Sex-specific 1e-2 VUMC, sex-differential UKB |
| 0.003309 | 0.00352 | 4.732 | 0.5591 | 25.92 | 27.05 | True | True | False | False | |
| 0.00352 | 0.003746 | 3.03 | 0.6481 | 16.31 | 12.68 | True | True | True | True | Sex-specific 1e-2 VUMC, sex-differential UKB |
| 0.003509 | 0.003729 | 3.122 | 0.5917 | 16.49 | 12.89 | True | True | True | True | Sex-specific 1e-2 VUMC, sex-differential UKB |
| 0.003452 | 0.003669 | 3.119 | 0.5966 | 17.48 | 13.76 | True | True | True | True | Sex-specific 1e-2 VUMC, sex-differential UKB |
| 0.003461 | 0.003679 | 3.143 | 0.5539 | 16.71 | 13.19 | True | True | True | True | Sex-specific 1e-2 VUMC, sex-differential UKB |
| 0.003452 | 0.003669 | 3.119 | 0.596 | 17.51 | 13.77 | True | True | True | True | Sex-specific 1e-2 VUMC, sex-differential UKB |
| 0.003451 | 0.003669 | 3.121 | 0.5959 | 17.46 | 13.77 | True | True | True | True | Sex-specific 1e-2 VUMC, sex-differential UKB |
| 0.003451 | 0.003669 | 3.121 | 0.5959 | 17.46 | 13.77 | True | True | True | True | Sex-specific 1e-2 VUMC, sex-differential UKB |
| 0.003451 | 0.003669 | 3.121 | 0.5959 | 17.46 | 13.77 | True | True | True | True | Sex-specific 1e-2 VUMC, sex-differential UKB |
| 0.003451 | 0.003669 | 3.121 | 0.5952 | 17.46 | 13.77 | True | True | True | True | Sex-specific 1e-2 VUMC, sex-differential UKB |
| 0.003452 | 0.003669 | 3.115 | 0.5979 | 17.46 | 13.77 | True | True | True | True | Sex-specific 1e-2 VUMC, sex-differential UKB |
| 0.003452 | 0.003669 | 3.115 | 0.5979 | 17.46 | 13.77 | True | True | True | True | Sex-specific 1e-2 VUMC, sex-differential UKB |
| 0.003451 | 0.003669 | 3.12 | 0.5955 | 17.46 | 13.77 | True | True | True | True | Sex-specific 1e-2 VUMC, sex-differential UKB |
| 0.003451 | 0.003669 | 3.12 | 0.5955 | 17.46 | 13.77 | True | True | True | True | Sex-specific 1e-2 VUMC, sex-differential UKB |
| 0.003451 | 0.003669 | 3.121 | 0.5955 | 17.46 | 13.77 | True | True | True | True | Sex-specific 1e-2 VUMC, sex-differential UKB |
| 0.003483 | 0.003702 | 3.15 | 0.5752 | 17.95 | 13.91 | True | True | True | True | Sex-specific 1e-2 VUMC, sex-differential UKB |
| 0.003452 | 0.00367 | 3.115 | 0.5952 | 17.43 | 13.79 | True | True | True | True | Sex-specific 1e-2 VUMC, sex-differential UKB |
| 0.003452 | 0.00367 | 3.115 | 0.5952 | 17.44 | 13.78 | True | True | True | True | Sex-specific 1e-2 VUMC, sex-differential UKB |

| SE_UKB_female | SE_UKB_male | LOG10P_VUMC_female | LOG10P_VUMC_male | LOG10P_UKB_female | LOG10P_UKB_male | female_effect_direction_replication | male_effect_direction_replication | sex_effect_comparison_replication | pvalue_effect_comparison_replication | replication_type |
|---|---|---|---|---|---|---|---|---|---|---|
| 0.003452 | 0.003669 | 3.114 | 0.5952 | 17.44 | 13.79 | True | True | True | True | Sex-specific 1e-2 VUMC, sex-differential UKB |
| 0.003452 | 0.003669 | 3.114 | 0.5952 | 17.43 | 13.8 | True | True | True | True | Sex-specific 1e-2 VUMC, sex-differential UKB |
| 0.003452 | 0.003669 | 3.114 | 0.5952 | 17.46 | 13.78 | True | True | True | True | Sex-specific 1e-2 VUMC, sex-differential UKB |
| 0.003451 | 0.003669 | 3.119 | 0.5952 | 17.48 | 13.78 | True | True | True | True | Sex-specific 1e-2 VUMC, sex-differential UKB |
| 0.003453 | 0.00367 | 3.12 | 0.5949 | 17.46 | 13.81 | True | True | True | True | Sex-specific 1e-2 VUMC, sex-differential UKB |
| 0.003453 | 0.00367 | 3.119 | 0.5956 | 17.47 | 13.81 | True | True | True | True | Sex-specific 1e-2 VUMC, sex-differential UKB |
| 0.003453 | 0.003671 | 3.12 | 0.5956 | 17.46 | 13.8 | True | True | True | True | Sex-specific 1e-2 VUMC, sex-differential UKB |
| 0.003452 | 0.003669 | 3.119 | 0.596 | 17.47 | 13.76 | True | True | True | True | Sex-specific 1e-2 VUMC, sex-differential UKB |
| 0.003451 | 0.003669 | 3.11 | 0.5972 | 17.46 | 13.79 | True | True | True | True | Sex-specific 1e-2 VUMC, sex-differential UKB |
| 0.003451 | 0.003669 | 3.119 | 0.596 | 17.46 | 13.77 | True | True | True | True | Sex-specific 1e-2 VUMC, sex-differential UKB |
| 0.003451 | 0.003669 | 3.119 | 0.596 | 17.48 | 13.78 | True | True | True | True | Sex-specific 1e-2 VUMC, sex-differential UKB |
| 0.003451 | 0.003669 | 3.119 | 0.596 | 17.48 | 13.77 | True | True | True | True | Sex-specific 1e-2 VUMC, sex-differential UKB |
| 0.003451 | 0.003669 | 3.119 | 0.5957 | 17.47 | 13.77 | True | True | True | True | Sex-specific 1e-2 VUMC, sex-differential UKB |
| 0.003451 | 0.003669 | 3.119 | 0.5957 | 17.47 | 13.77 | True | True | True | True | Sex-specific 1e-2 VUMC, sex-differential UKB |
| 0.003451 | 0.003669 | 3.119 | 0.5957 | 17.44 | 13.79 | True | True | True | True | Sex-specific 1e-2 VUMC, sex-differential UKB |
| 0.003451 | 0.003669 | 3.119 | 0.596 | 17.47 | 13.77 | True | True | True | True | Sex-specific 1e-2 VUMC, sex-differential UKB |
| 0.003451 | 0.003669 | 3.119 | 0.5962 | 17.43 | 13.77 | True | True | True | True | Sex-specific 1e-2 VUMC, sex-differential UKB |
| 0.003451 | 0.003669 | 3.119 | 0.596 | 17.48 | 13.77 | True | True | True | True | Sex-specific 1e-2 VUMC, sex-differential UKB |
| 0.003451 | 0.003669 | 3.111 | 0.5976 | 17.44 | 13.86 | True | True | True | True | Sex-specific 1e-2 VUMC, sex-differential UKB |
| 0.003452 | 0.003669 | 3.122 | 0.5968 | 17.47 | 13.78 | True | True | True | True | Sex-specific 1e-2 VUMC, sex-differential UKB |
| 0.003452 | 0.003669 | 3.122 | 0.5968 | 17.47 | 13.78 | True | True | True | True | Sex-specific 1e-2 VUMC, sex-differential UKB |
| 0.003452 | 0.003669 | 3.119 | 0.596 | 17.47 | 13.78 | True | True | True | True | Sex-specific 1e-2 VUMC, sex-differential UKB |
| 0.003451 | 0.003669 | 3.12 | 0.5955 | 17.46 | 13.78 | True | True | True | True | Sex-specific 1e-2 VUMC, sex-differential UKB |
| 0.003451 | 0.003669 | 3.128 | 0.5958 | 17.43 | 13.77 | True | True | True | True | Sex-specific 1e-2 VUMC, sex-differential UKB |
| 0.003452 | 0.003669 | 3.12 | 0.5943 | 17.46 | 13.78 | True | True | True | True | Sex-specific 1e-2 VUMC, sex-differential UKB |
| 0.003452 | 0.003669 | 3.156 | 0.5837 | 17.46 | 13.79 | True | True | True | True | Sex-specific 1e-2 VUMC, sex-differential UKB |

| SE_UKB_female | SE_UKB_male | LOG10P_VUMC_female | LOG10P_VUMC_male | LOG10P_UKB_female | LOG10P_UKB_male | female_effect_direction_replication | male_effect_direction_replication | sex_effect_comparison_replication | pvalue_effect_comparison_replication | replication_type |
|---|---|---|---|---|---|---|---|---|---|---|
| 0.003454 | 0.003672 | 3.145 | 0.5804 | 17.47 | 13.86 | True | True | True | True | Sex-specific 1e-2 VUMC, sex-differential UKB |
| 0.003451 | 0.003669 | 3.145 | 0.5836 | 17.46 | 13.76 | True | True | True | True | Sex-specific 1e-2 VUMC, sex-differential UKB |
| 0.003452 | 0.003669 | 3.146 | 0.5834 | 17.46 | 13.77 | True | True | True | True | Sex-specific 1e-2 VUMC, sex-differential UKB |
| 0.003452 | 0.003669 | 3.146 | 0.5834 | 17.47 | 13.77 | True | True | True | True | Sex-specific 1e-2 VUMC, sex-differential UKB |
| 0.003452 | 0.003669 | 3.122 | 0.5948 | 17.45 | 13.78 | True | True | True | True | Sex-specific 1e-2 VUMC, sex-differential UKB |
| 0.003451 | 0.003669 | 3.122 | 0.5949 | 17.44 | 13.78 | True | True | True | True | Sex-specific 1e-2 VUMC, sex-differential UKB |
| SE_UKB_female | SE_UKB_male | LOG10P_VUMC_female | LOG10P_VUMC_male | LOG10P_UKB_female | LOG10P_UKB_male | female_effect_direction_replication | male_effect_direction_replication | sex_effect_comparison_replication | pvalue_effect_comparison_replication | replication_type |
| 0.003276 | 0.003489 | 7.621 | 1.043 | 34.64 | 32.15 | True | True | False | True | |
| 0.003451 | 0.003669 | 3.122 | 0.5949 | 17.46 | 13.77 | True | True | True | True | Sex-specific 1e-2 VUMC, sex-differential UKB |
| 0.003451 | 0.003669 | 3.122 | 0.5949 | 17.46 | 13.77 | True | True | True | True | Sex-specific 1e-2 VUMC, sex-differential UKB |
| 0.003452 | 0.003669 | 3.122 | 0.5949 | 17.38 | 13.8 | True | True | True | True | Sex-specific 1e-2 VUMC, sex-differential UKB |
| 0.003469 | 0.003688 | 3.109 | 0.6023 | 17.24 | 13.57 | True | True | True | True | Sex-specific 1e-2 VUMC, sex-differential UKB |
| 0.00345 | 0.003668 | 3.138 | 0.5951 | 17.22 | 13.87 | True | True | True | True | Sex-specific 1e-2 VUMC, sex-differential UKB |
| 0.00345 | 0.003667 | 3.125 | 0.5942 | 17.22 | 13.86 | True | True | True | True | Sex-specific 1e-2 VUMC, sex-differential UKB |
| | | 3.169 | 0.6159 | | | False | False | False | False | |
| 0.003365 | 0.003573 | 5.699 | 0.3812 | 30.13 | 30.12 | True | True | False | True | |
| 0.005859 | 0.006276 | 7.627 | 0.9898 | 55.15 | 47.38 | True | True | True | True | Sex-specific 1e-2 VUMC, sex-differential UKB |
| 0.005859 | 0.006276 | 7.611 | 1.005 | 55.16 | 47.37 | True | True | True | True | Sex-specific 1e-2 VUMC, sex-differential UKB |
| 0.005903 | 0.006327 | 8.335 | 1.093 | 63.95 | 52.17 | True | True | True | True | Sex-specific 1e-2 VUMC, sex-differential UKB |
| 0.005892 | 0.006314 | 8.498 | 1.109 | 63.89 | 52.24 | True | True | True | True | Sex-specific 1e-2 VUMC, sex-differential UKB |
| 0.005902 | 0.006325 | 8.366 | 1.053 | 64 | 52.32 | True | True | True | True | Sex-specific 1e-2 VUMC, sex-differential UKB |
| 0.005891 | 0.006312 | 8.465 | 1.022 | 63.96 | 52.4 | True | True | True | True | Sex-specific 1e-2 VUMC, sex-differential UKB |
| 0.005892 | 0.006313 | 8.486 | 1.04 | 63.97 | 52.44 | True | True | True | True | Sex-specific 1e-2 VUMC, sex-differential UKB |
| 0.005899 | 0.006321 | 8.447 | 1.03 | 64.29 | 52.42 | True | True | True | True | Sex-specific 1e-2 VUMC, sex-differential UKB |
| 0.005891 | 0.006313 | 8.519 | 1.017 | 64.08 | 52.44 | True | True | True | True | Sex-specific 1e-2 VUMC, sex-differential UKB |
| 0.005123 | 0.005487 | 6.824 | 0.8147 | 43.78 | 39.76 | True | True | False | True | |

| SE_UKB_female | SE_UKB_male | LOG10P_VUMC_female | LOG10P_VUMC_male | LOG10P_UKB_female | LOG10P_UKB_male | female_effect_direction_replication | male_effect_direction_replication | sex_effect_comparison_replication | pvalue_effect_comparison_replication | replication_type |
|---|---|---|---|---|---|---|---|---|---|---|
| 0.005874 | 0.006296 | 8.492 | 1.168 | 63.61 | 52.59 | True | True | True | True | Sex-specific 1e-2 VUMC, sex-differential UKB |
| 0.005164 | 0.005529 | 6.719 | 0.915 | 43.1 | 39.72 | True | True | False | True | |
| 0.005874 | 0.006295 | 8.48 | 1.189 | 63.7 | 52.58 | True | True | True | True | Sex-specific 1e-2 VUMC, sex-differential UKB |
| 0.005129 | 0.005491 | 6.732 | 0.8095 | 43.73 | 40.45 | True | True | False | True | |
| 0.005874 | 0.006295 | 8.491 | 1.188 | 63.62 | 52.6 | True | True | True | True | Sex-specific 1e-2 VUMC, sex-differential UKB |
| 0.005123 | 0.005487 | 6.843 | 0.863 | 43.82 | 39.75 | True | True | False | True | |
| 0.005123 | 0.005487 | 6.815 | 0.8624 | 43.88 | 39.74 | True | True | False | True | |
| 0.005873 | 0.006295 | 8.545 | 1.156 | 63.67 | 52.69 | True | True | True | True | Sex-specific 1e-2 VUMC, sex-differential UKB |
| 0.005873 | 0.006291 | 8.591 | 1.156 | 63.66 | 53.4 | True | True | True | True | Sex-specific 1e-2 VUMC, sex-differential UKB |
| 0.005879 | 0.006302 | 8.664 | 1.165 | 64.09 | 52.86 | True | True | True | True | Sex-specific 1e-2 VUMC, sex-differential UKB |
| 0.00336 | 0.003573 | 5.814 | 0.1661 | 7.707 | 0.3746 | True | True | True | True | Sex-specific 1e-2 VUMC & UKB |
| 0.003367 | 0.003578 | 5.251 | 0.1405 | 8.69 | 0.4146 | True | True | True | True | Sex-specific 1e-2 VUMC & UKB |
| 0.003354 | 0.003569 | 5.328 | 0.1993 | 8.179 | 0.8662 | True | True | True | True | Sex-specific 1e-2 VUMC & UKB |
| 0.003487 | 0.003701 | 2.512 | 0.2989 | 6.649 | 0.2036 | True | False | True | True | Sex-specific 1e-2 VUMC, sex-differential UKB |
| 0.003511 | 0.003729 | 2.344 | 0.293 | 8.016 | 0.5564 | True | False | True | True | Sex-specific 1e-2 VUMC & UKB |
| 0.003347 | 0.003565 | 5.77 | 0.1472 | 9.22 | 0.7679 | True | True | True | True | Sex-specific 1e-2 VUMC & UKB |
| 0.003277 | 0.003478 | 8.633 | 0.7887 | 13.68 | 0.3548 | True | True | True | True | Sex-specific 1e-2 VUMC & UKB |
| 0.003354 | 0.003565 | 7.448 | 0.2715 | 12.13 | 0.3326 | True | True | True | True | Sex-specific 1e-2 VUMC & UKB |
| 0.003361 | 0.00357 | 6.932 | 0.2719 | 13.14 | 0.4372 | True | True | True | True | Sex-specific 1e-2 VUMC & UKB |
| 0.005299 | 0.005696 | 0.9814 | 4.81 | 21.58 | 20.92 | True | True | True | False | |
| 0.005315 | 0.005713 | 0.9256 | 5.204 | 22.08 | 21.2 | True | True | True | False | |
| 0.00527 | 0.005661 | 0.9332 | 5.427 | 24.06 | 22.65 | True | True | True | False | |
| 0.005216 | 0.005606 | 0.8734 | 5.31 | 24.02 | 22.46 | True | True | True | False | |
| 0.005266 | 0.005658 | 0.9768 | 5.346 | 24.12 | 22.97 | True | True | True | False | |
| 0.005216 | 0.005605 | 0.856 | 5.343 | 24.16 | 22.59 | True | True | True | False | |
| 0.005269 | 0.00566 | 1.013 | 5.475 | 24.08 | 23.35 | True | True | True | False | |
| 0.005264 | 0.005655 | 1.063 | 4.973 | 23.89 | 22.16 | True | True | True | False | |

| SE_UKB_female | SE_UKB_male | LOG10P_VUMC_female | LOG10P_VUMC_male | LOG10P_UKB_female | LOG10P_UKB_male | female_effect_direction_replication | male_effect_direction_replication | sex_effect_comparison_replication | pvalue_effect_comparison_replication | replication_type |
|---|---|---|---|---|---|---|---|---|---|---|
| 0.005257 | 0.005649 | 1.094 | 4.855 | 23.98 | 21.82 | True | True | True | False | |
| 0.004766 | 0.005111 | 1.025 | 6.342 | 26.71 | 25.56 | True | True | True | False | |
| 0.004766 | 0.005111 | 1.019 | 6.305 | 26.72 | 25.54 | True | True | True | False | |
| 0.004759 | 0.005104 | 1.029 | 6.271 | 27.16 | 25.47 | True | True | True | False | |
| 0.004765 | 0.005109 | 1.03 | 6.363 | 26.71 | 25.49 | True | True | True | False | |
| 0.003566 | 0.003829 | 0.1914 | 5.175 | 8.371 | 7.85 | False | True | True | False | |
| 0.003566 | 0.003829 | 0.1936 | 5.175 | 8.257 | 7.67 | False | True | True | False | |
| 0.004765 | 0.005109 | 1.029 | 6.272 | 26.71 | 25.52 | True | True | True | False | |
| 0.003565 | 0.003828 | 0.1922 | 5.179 | 8.401 | 7.823 | False | True | True | False | |
| 0.004765 | 0.005109 | 1.029 | 6.272 | 26.7 | 25.52 | True | True | True | False | |
| 0.004767 | 0.005111 | 1.018 | 6.253 | 26.76 | 25.53 | True | True | True | False | |
| 0.004767 | 0.005111 | 1.021 | 6.232 | 26.76 | 25.53 | True | True | True | False | |
| 0.004767 | 0.005112 | 1.031 | 6.346 | 26.61 | 25.48 | True | True | True | False | |
| 0.004767 | 0.005112 | 1.031 | 6.346 | 26.61 | 25.48 | True | True | True | False | |
| 0.004767 | 0.005112 | 1.031 | 6.346 | 26.63 | 25.47 | True | True | True | False | |
| 0.004767 | 0.005112 | 1.036 | 6.33 | 26.52 | 25.46 | True | True | True | False | |
| 0.003713 | 0.003989 | 0.3203 | 6.06 | 11.73 | 12.39 | True | True | True | True | Sex-specific 1e-2 VUMC, sex-differential UKB |
| 0.004771 | 0.005118 | 1.038 | 6.325 | 26.15 | 25.28 | True | True | True | False | |
| 0.003569 | 0.003834 | 0.143 | 5.105 | 8.292 | 7.951 | False | True | True | False | |
| 0.004767 | 0.005112 | 1.038 | 6.326 | 26.52 | 25.45 | True | True | True | False | |
| 0.004767 | 0.005112 | 1.037 | 6.326 | 26.52 | 25.45 | True | True | True | False | |
| 0.004767 | 0.005112 | 1.032 | 6.326 | 26.52 | 25.45 | True | True | True | False | |
| 0.004768 | 0.005112 | 1.031 | 6.328 | 26.48 | 25.44 | True | True | True | False | |
| 0.004767 | 0.005112 | 1.032 | 6.326 | 26.52 | 25.45 | True | True | True | False | |
| 0.004767 | 0.005112 | 1.032 | 6.326 | 26.52 | 25.45 | True | True | True | False | |

| SE_UKB_female | SE_UKB_male | LOG10P_VUMC_female | LOG10P_VUMC_male | LOG10P_UKB_female | LOG10P_UKB_male | female_effect_direction_replication | male_effect_direction_replication | sex_effect_comparison_replication | pvalue_effect_comparison_replication | replication_type |
|---|---|---|---|---|---|---|---|---|---|---|
| 0.003569 | 0.003834 | 0.1416 | 5.103 | 8.371 | 7.937 | False | True | True | False | |
| 0.004767 | 0.005112 | 0.9872 | 6.479 | 26.55 | 25.38 | True | True | True | False | |
| 0.004765 | 0.005109 | 0.9872 | 6.479 | 26.49 | 25.23 | True | True | True | False | |
| 0.004768 | 0.005112 | 0.9768 | 6.498 | 26.36 | 25.24 | True | True | True | False | |
| 0.004665 | 0.005005 | 0.5921 | 5.444 | 22.4 | 23.27 | True | True | True | True | Sex-specific 1e-2 VUMC, sex-differential UKB |
| 0.004768 | 0.005112 | 0.9769 | 6.498 | 26.36 | 25.23 | True | True | True | False | |
| 0.004768 | 0.005112 | 0.9769 | 6.498 | 26.36 | 25.23 | True | True | True | False | |
| 0.004767 | 0.005111 | 0.9751 | 6.456 | 26.29 | 25.21 | True | True | True | False | |
| 0.004766 | 0.005111 | 0.9752 | 6.456 | 26.28 | 25.1 | True | True | True | False | |
| 0.004766 | 0.005111 | 0.9752 | 6.456 | 26.29 | 25.22 | True | True | True | False | |
| 0.004765 | 0.00511 | 0.9752 | 6.456 | 26.02 | 24.99 | True | True | True | False | |
| 0.004767 | 0.005112 | 0.9994 | 6.472 | 26.23 | 25.23 | True | True | True | False | |
| 0.004767 | 0.005112 | 0.9994 | 6.472 | 26.24 | 25.23 | True | True | True | False | |
| 0.004767 | 0.005112 | 1.001 | 6.457 | 26.23 | 25.23 | True | True | True | False | |
| 0.004767 | 0.005112 | 1.001 | 6.461 | 26.24 | 25.23 | True | True | True | False | |
| 0.004767 | 0.005112 | 1.001 | 6.457 | 26.23 | 25.23 | True | True | True | False | |
| 0.004774 | 0.005118 | 0.9511 | 6.342 | 26.13 | 25.04 | True | True | True | False | |
| 0.003713 | 0.003989 | 0.3317 | 6.021 | 11.64 | 12.26 | True | True | True | True | Sex-specific 1e-2 VUMC, sex-differential UKB |
| 0.004767 | 0.005112 | 1.001 | 6.452 | 26.27 | 25.23 | True | True | True | False | |
| 0.004781 | 0.005127 | 0.9184 | 6.206 | 26.7 | 25.16 | True | True | True | False | |
| 0.004781 | 0.005127 | 0.9184 | 6.206 | 26.7 | 25.16 | True | True | True | False | |
| 0.004778 | 0.005124 | 0.9229 | 6.162 | 26.62 | 25.03 | True | True | True | False | |
| 0.004781 | 0.005128 | 0.9208 | 6.163 | 26.69 | 25.11 | True | True | True | False | |
| 0.00478 | 0.005127 | 0.9033 | 6.146 | 26.54 | 25.12 | True | True | True | False | |
| 0.004781 | 0.005129 | 0.9249 | 6.097 | 26.78 | 25.22 | True | True | True | False | |
| 0.004781 | 0.005127 | 0.9223 | 6.163 | 26.67 | 25.16 | True | True | True | False | |

| SE_UKB_female | SE_UKB_male | LOG10P_VUMC_female | LOG10P_VUMC_male | LOG10P_UKB_female | LOG10P_UKB_male | female_effect_direction_replication | male_effect_direction_replication | sex_effect_comparison_replication | pvalue_effect_comparison_replication | replication_type |
|---|---|---|---|---|---|---|---|---|---|---|
| 0.004781 | 0.005127 | 0.9223 | 6.163 | 26.68 | 25.16 | True | True | True | False | |
| 0.004776 | 0.005124 | 0.9227 | 6.162 | 26.75 | 24.88 | True | True | True | False | |
| 0.00478 | 0.005127 | 0.9227 | 6.162 | 26.59 | 24.91 | True | True | True | False | |
| 0.00358 | 0.003845 | 0.14 | 5.285 | 9.482 | 8.154 | False | True | False | False | |
| 0.003573 | 0.003838 | 0.1968 | 4.934 | 8.402 | 7.859 | False | True | True | False | |
| 0.004781 | 0.005127 | 0.9232 | 6.163 | 26.72 | 25.18 | True | True | True | False | |
| 0.004781 | 0.005127 | 0.9233 | 6.162 | 26.73 | 25.17 | True | True | True | False | |
| 0.004788 | 0.005136 | 0.9256 | 6.16 | 26.7 | 25.16 | True | True | True | False | |
| 0.004793 | 0.00514 | 0.9217 | 6.107 | 26.73 | 25.13 | True | True | True | False | |
| 0.004865 | 0.005218 | 0.9539 | 6.315 | 26.16 | 24.57 | True | True | True | False | |
| 0.004862 | 0.005214 | 0.9536 | 6.355 | 26.12 | 24.61 | True | True | True | False | |
| 0.00478 | 0.005127 | 0.9001 | 6.176 | 26.65 | 25.15 | True | True | True | False | |
| 0.00478 | 0.005127 | 0.9001 | 6.176 | 26.69 | 25.17 | True | True | True | False | |
| 0.00478 | 0.005127 | 0.9001 | 6.176 | 26.7 | 25.19 | True | True | True | False | |
| 0.003538 | 0.003801 | 0.1405 | 4.821 | 7.468 | 6.618 | False | True | True | False | |
| 0.004068 | 0.004361 | 0.9515 | 3.527 | 17.48 | 17.16 | True | True | True | False | |
| 0.004791 | 0.005139 | 0.9445 | 6.07 | 26.47 | 25.19 | True | True | True | False | |
| 0.004748 | 0.005092 | 0.9275 | 6.134 | 26.03 | 25.24 | True | True | True | False | |
| 0.004779 | 0.005126 | 0.9391 | 6.135 | 26.72 | 25.08 | True | True | True | False | |
| 0.004807 | 0.005157 | 0.8767 | 6.109 | 26.18 | 25.12 | True | True | True | False | |
| 0.004805 | 0.005155 | 0.8767 | 6.113 | 26.29 | 24.95 | True | True | True | False | |
| 0.004748 | 0.005095 | 1.027 | 7.765 | 25.69 | 25.92 | True | True | True | True | Sex-specific 1e-2 VUMC, sex-differential UKB |
| 0.00478 | 0.005134 | 1.136 | 7.276 | 25.14 | 26.02 | True | True | True | True | Sex-specific 1e-2 VUMC, sex-differential UKB |
| SE_UKB_female | SE_UKB_male | LOG10P_VUMC_female | LOG10P_VUMC_male | LOG10P_UKB_female | LOG10P_UKB_male | female_effect_direction_replication | male_effect_direction_replication | sex_effect_comparison_replication | pvalue_effect_comparison_replication | replication_type |
| 0.003812 | 0.004106 | 0.2126 | 8.373 | 12.2 | 14.24 | True | True | True | True | Sex-specific 1e-2 VUMC, sex-differential UKB |
| 0.004783 | 0.005134 | 1.077 | 7.351 | 25.35 | 25.71 | True | True | True | True | Sex-specific 1e-2 VUMC, sex-differential UKB |

| SE_UKB_female | SE_UKB_male | LOG10P_VUMC_female | LOG10P_VUMC_male | LOG10P_UKB_female | LOG10P_UKB_male | female_effect_direction_replication | male_effect_direction_replication | sex_effect_comparison_replication | pvalue_effect_comparison_replication | replication_type |
|---|---|---|---|---|---|---|---|---|---|---|
| 0.005183 | 0.005569 | 1.453 | 6.027 | 26.69 | 26.68 | True | True | True | False | |
| 0.004775 | 0.005127 | 1.129 | 7.109 | 26.06 | 25.53 | True | True | True | False | |
| 0.004775 | 0.005127 | 1.185 | 6.945 | 26.04 | 25.51 | True | True | True | False | |
| 0.004772 | 0.005125 | 1.194 | 7.159 | 26.07 | 25.82 | True | True | True | False | |
| 0.004772 | 0.005124 | 1.203 | 7.151 | 26.15 | 25.89 | True | True | True | False | |
| 0.005195 | 0.005583 | 1.463 | 5.678 | 27.6 | 26.82 | True | True | True | False | |
| 0.004773 | 0.005125 | 1.192 | 7.143 | 26.01 | 25.89 | True | True | True | False | |
| 0.004956 | 0.005327 | 1.289 | 6.067 | 28.46 | 26.63 | True | True | True | False | |
| 0.004946 | 0.005318 | 1.399 | 5.573 | 28.56 | 26.66 | True | True | True | False | |
| 0.004956 | 0.005327 | 1.288 | 6.063 | 28.39 | 26.68 | True | True | True | False | |
| 0.004958 | 0.005329 | 1.26 | 6.09 | 28.38 | 26.75 | True | True | True | False | |
| 0.004956 | 0.005327 | 1.316 | 6.028 | 28.18 | 26.56 | True | True | True | False | |
| 0.005195 | 0.005585 | 1.593 | 5.886 | 27.89 | 26.97 | True | True | True | False | |
| 0.005164 | 0.005551 | 1.591 | 5.886 | 27.69 | 26.68 | True | True | True | False | |
| 0.004954 | 0.005326 | 1.316 | 6.028 | 28.16 | 26.59 | True | True | True | False | |
| 0.003511 | 0.003775 | 0.00939 | 4.665 | 4.881 | 5.875 | True | True | True | True | Sex-specific 1e-2 VUMC, sex-differential UKB |
| 0.004959 | 0.005329 | 1.336 | 5.72 | 28.06 | 26.8 | True | True | True | False | |
| 0.004126 | 0.004431 | 0.54 | 3.881 | 12.37 | 11.31 | True | True | True | False | |
| 0.004127 | 0.004432 | 0.5311 | 3.813 | 12.41 | 11.31 | True | True | True | False | |
| 0.004126 | 0.004432 | 0.5305 | 3.813 | 12.43 | 11.29 | True | True | True | False | |
| 0.004126 | 0.004432 | 0.5408 | 3.886 | 12.39 | 11.32 | True | True | True | False | |
| 0.004126 | 0.004432 | 0.5385 | 3.878 | 12.38 | 11.32 | True | True | True | False | |
| 0.004126 | 0.004431 | 0.5385 | 3.878 | 12.36 | 11.3 | True | True | True | False | |
| 0.004126 | 0.004432 | 0.5404 | 3.882 | 12.36 | 11.4 | True | True | True | False | |
| 0.004961 | 0.00533 | 1.351 | 5.694 | 27.95 | 26.94 | True | True | True | False | |
| 0.004126 | 0.004432 | 0.5379 | 3.812 | 12.38 | 11.37 | True | True | True | False | |

| SE_UKB_female | SE_UKB_male | LOG10P_VUMC_female | LOG10P_VUMC_male | LOG10P_UKB_female | LOG10P_UKB_male | female_effect_direction_replication | male_effect_direction_replication | sex_effect_comparison_replication | pvalue_effect_comparison_replication | replication_type |
|---|---|---|---|---|---|---|---|---|---|---|
| 0.004127 | 0.004432 | 0.5397 | 3.819 | 12.31 | 11.28 | True | True | True | False | |
| 0.004755 | 0.005105 | 1.463 | 6.202 | 26.01 | 26.89 | True | True | True | True | Sex-specific 1e-2 VUMC, sex-differential UKB |
| 0.004126 | 0.004431 | 0.5562 | 3.839 | 12.28 | 11.35 | True | True | True | False | |
| 0.004126 | 0.004432 | 0.5581 | 3.846 | 12.28 | 11.36 | True | True | True | False | |
| 0.004126 | 0.004432 | 0.558 | 3.846 | 12.28 | 11.35 | True | True | True | False | |
| 0.004789 | 0.005143 | 1.751 | 6.273 | 26.02 | 27.11 | True | True | True | True | Sex-specific 1e-2 VUMC, sex-differential UKB |
| 0.004789 | 0.005143 | 1.803 | 6.147 | 26.19 | 27.08 | True | True | True | True | Sex-specific 1e-2 VUMC, sex-differential UKB |
| 0.004789 | 0.005143 | 1.75 | 6.273 | 26.22 | 27.11 | True | True | True | True | Sex-specific 1e-2 VUMC, sex-differential UKB |
| 0.004146 | 0.004453 | 0.6781 | 3.791 | 12.28 | 11.28 | True | True | True | False | |
| 0.004148 | 0.004454 | 0.6686 | 3.787 | 12.2 | 11.25 | True | True | True | False | |
| 0.004126 | 0.004431 | 0.5516 | 3.81 | 12.49 | 11.23 | True | True | True | False | |
| 0.004133 | 0.004439 | 0.5758 | 3.788 | 12.43 | 11.23 | True | True | True | False | |
| 0.005169 | 0.005555 | 1.765 | 5.828 | 27.86 | 27.66 | True | True | True | False | |
| 0.00496 | 0.00533 | 1.431 | 5.665 | 28.24 | 27.04 | True | True | True | False | |
| 0.004687 | 0.005031 | 1.284 | 5.997 | 25.97 | 25.89 | True | True | True | False | |
| 0.00496 | 0.005329 | 1.417 | 5.586 | 28.39 | 27 | True | True | True | False | |
| 0.004689 | 0.005032 | 1.258 | 5.926 | 25.62 | 25.65 | True | True | True | True | Sex-specific 1e-2 VUMC, sex-differential UKB |
| 0.004687 | 0.005032 | 1.257 | 5.927 | 26.04 | 26.04 | True | True | True | False | |
| 0.004687 | 0.005032 | 1.243 | 5.995 | 26.03 | 26.05 | True | True | True | True | Sex-specific 1e-2 VUMC, sex-differential UKB |
| 0.004955 | 0.005324 | 1.414 | 5.585 | 28.15 | 26.73 | True | True | True | False | |
| 0.004955 | 0.005324 | 1.414 | 5.606 | 28.15 | 26.73 | True | True | True | False | |
| 0.004687 | 0.005032 | 1.271 | 5.959 | 26.04 | 26.05 | True | True | True | True | Sex-specific 1e-2 VUMC, sex-differential UKB |
| 0.004707 | 0.005054 | 1.289 | 6.079 | 25.86 | 26.76 | True | True | True | True | Sex-specific 1e-2 VUMC, sex-differential UKB |
| 0.004958 | 0.005325 | 1.413 | 5.597 | 28.26 | 26.81 | True | True | True | False | |
| 0.004958 | 0.005326 | 1.413 | 5.597 | 28.28 | 26.84 | True | True | True | False | |
| 0.004689 | 0.005032 | 1.275 | 6.072 | 26.38 | 26.29 | True | True | True | False | |

| SE_UKB_female | SE_UKB_male | LOG10P_VUMC_female | LOG10P_VUMC_male | LOG10P_UKB_female | LOG10P_UKB_male | female_effect_direction_replication | male_effect_direction_replication | sex_effect_comparison_replication | pvalue_effect_comparison_replication | replication_type |
|---|---|---|---|---|---|---|---|---|---|---|
| 0.004958 | 0.005326 | 1.421 | 5.563 | 28.31 | 27.01 | True | True | True | False | |
| 0.004958 | 0.005326 | 1.422 | 5.563 | 28.3 | 26.98 | True | True | True | False | |
| 0.004955 | 0.005322 | 1.411 | 5.828 | 28.23 | 27.01 | True | True | True | False | |
| 0.004628 | 0.004965 | 0.907 | 4.883 | 26.77 | 25.97 | True | True | True | False | |
| 0.005161 | 0.005547 | 1.705 | 5.822 | 27.14 | 26.84 | True | True | True | False | |
| 0.004643 | 0.004982 | 0.881 | 4.914 | 26.79 | 26.93 | True | True | True | True | Sex-specific 1e-2 VUMC, sex-differential UKB |
| SE_UKB_female | SE_UKB_male | LOG10P_VUMC_female | LOG10P_VUMC_male | LOG10P_UKB_female | LOG10P_UKB_male | female_effect_direction_replication | male_effect_direction_replication | sex_effect_comparison_replication | pvalue_effect_comparison_replication | replication_type |
| 0.004965 | 0.005335 | 1.488 | 5.588 | 29.75 | 29.05 | True | True | True | False | |
| 0.004771 | 0.005119 | 1.166 | 6.071 | 27.29 | 28.24 | True | True | True | True | Sex-specific 1e-2 VUMC, sex-differential UKB |
| 0.004962 | 0.005332 | 1.499 | 5.53 | 29.67 | 28.84 | True | True | True | False | |
| 0.005168 | 0.005557 | 1.804 | 5.779 | 28.73 | 29.09 | True | True | True | True | Sex-specific 1e-2 VUMC, sex-differential UKB |
| 0.004767 | 0.005116 | 1.196 | 5.878 | 27.22 | 27.96 | True | True | True | True | Sex-specific 1e-2 VUMC, sex-differential UKB |
| 0.005169 | 0.005558 | 1.88 | 5.624 | 28.77 | 29.06 | True | True | True | True | Sex-specific 1e-2 VUMC, sex-differential UKB |
| 0.004766 | 0.005113 | 1.196 | 5.888 | 27.18 | 27.69 | True | True | True | True | Sex-specific 1e-2 VUMC, sex-differential UKB |
| 0.004766 | 0.005114 | 1.107 | 5.853 | 27.16 | 28.02 | True | True | True | True | Sex-specific 1e-2 VUMC, sex-differential UKB |
| 0.004768 | 0.005116 | 1.196 | 5.89 | 27.27 | 28.03 | True | True | True | True | Sex-specific 1e-2 VUMC, sex-differential UKB |
| | | 1.197 | 5.892 | | | False | False | False | False | |
| 0.004768 | 0.005116 | 1.198 | 5.889 | 27.24 | 27.95 | True | True | True | True | Sex-specific 1e-2 VUMC, sex-differential UKB |
| 0.004962 | 0.005331 | 1.514 | 5.453 | 29.7 | 28.72 | True | True | True | False | |
| 0.004768 | 0.005116 | 1.196 | 5.899 | 27.24 | 27.98 | True | True | True | True | Sex-specific 1e-2 VUMC, sex-differential UKB |
| 0.00517 | 0.005558 | 1.9 | 5.686 | 28.8 | 29 | True | True | True | True | Sex-specific 1e-2 VUMC, sex-differential UKB |
| 0.003262 | 0.003509 | 2.08 | 0.6194 | 14.44 | 10.68 | True | True | True | True | Sex-specific 1e-2 VUMC, sex-differential UKB |
| 0.003327 | 0.003578 | 7.746 | 1.527 | 25.81 | 22.66 | True | True | False | True | |
| 0.003327 | 0.003578 | 7.746 | 1.527 | 25.76 | 22.66 | True | True | False | True | |
| 0.003263 | 0.003509 | 2.002 | 0.9216 | 16.77 | 11.83 | True | True | True | True | Sex-specific 1e-2 VUMC, sex-differential UKB |
| 0.003263 | 0.003509 | 2.015 | 0.9357 | 16.79 | 11.78 | True | True | True | True | Sex-specific 1e-2 VUMC, sex-differential UKB |

| SE_UKB_female | SE_UKB_male | LOG10P_VUMC_female | LOG10P_VUMC_male | LOG10P_UKB_female | LOG10P_UKB_male | female_effect_direction_replication | male_effect_direction_replication | sex_effect_comparison_replication | pvalue_effect_comparison_replication | replication_type |
|---|---|---|---|---|---|---|---|---|---|---|
| 0.003288 | 0.003539 | 7.319 | 1.655 | 22.23 | 17.92 | True | True | True | True | Sex-specific 1e-2 VUMC, sex-differential UKB |
| 0.003256 | 0.003502 | 3.094 | 1.306 | 18.17 | 13.3 | True | True | True | True | Sex-specific 1e-2 VUMC, sex-differential UKB |
| 0.003269 | 0.003517 | 5.309 | 1.635 | 21.5 | 15.88 | True | True | True | True | Sex-specific 1e-2 VUMC, sex-differential UKB |
| 0.003269 | 0.003517 | 5.309 | 1.636 | 21.55 | 15.87 | True | True | True | True | Sex-specific 1e-2 VUMC, sex-differential UKB |
| 0.003254 | 0.003496 | 4.193 | 1.849 | 24.11 | 16.22 | True | True | True | True | Sex-specific 1e-2 VUMC, sex-differential UKB |
| 0.003272 | 0.003521 | 5.464 | 1.651 | 21.73 | 16.04 | True | True | True | True | Sex-specific 1e-2 VUMC, sex-differential UKB |
| 0.003271 | 0.003519 | 5.327 | 1.678 | 21.55 | 15.89 | True | True | True | True | Sex-specific 1e-2 VUMC, sex-differential UKB |
| 0.003271 | 0.003519 | 5.342 | 1.637 | 21.7 | 15.75 | True | True | True | True | Sex-specific 1e-2 VUMC, sex-differential UKB |
| 0.00327 | 0.003517 | 5.358 | 1.63 | 21.71 | 15.7 | True | True | True | True | Sex-specific 1e-2 VUMC, sex-differential UKB |
| 0.00327 | 0.003518 | 5.358 | 1.63 | 21.69 | 15.69 | True | True | True | True | Sex-specific 1e-2 VUMC, sex-differential UKB |
| 0.003269 | 0.003517 | 5.359 | 1.632 | 21.61 | 15.67 | True | True | True | True | Sex-specific 1e-2 VUMC, sex-differential UKB |
| 0.003269 | 0.003517 | 5.335 | 1.631 | 21.64 | 15.68 | True | True | True | True | Sex-specific 1e-2 VUMC, sex-differential UKB |
| 0.003269 | 0.003517 | 5.331 | 1.63 | 21.61 | 15.64 | True | True | True | True | Sex-specific 1e-2 VUMC, sex-differential UKB |
| 0.003277 | 0.003525 | 5.329 | 1.632 | 21.41 | 15.47 | True | True | True | True | Sex-specific 1e-2 VUMC, sex-differential UKB |
| 0.003269 | 0.003517 | 5.344 | 1.631 | 21.62 | 15.65 | True | True | True | True | Sex-specific 1e-2 VUMC, sex-differential UKB |
| 0.003269 | 0.003517 | 5.352 | 1.63 | 21.61 | 15.65 | True | True | True | True | Sex-specific 1e-2 VUMC, sex-differential UKB |
| 0.003255 | 0.003512 | 3.247 | 1.275 | 17.93 | 13.18 | True | True | True | True | Sex-specific 1e-2 VUMC, sex-differential UKB |
| 0.003255 | 0.003512 | 3.258 | 1.272 | 17.92 | 13.19 | True | True | True | True | Sex-specific 1e-2 VUMC, sex-differential UKB |
| 0.003255 | 0.003512 | 3.258 | 1.271 | 17.92 | 13.19 | True | True | True | True | Sex-specific 1e-2 VUMC, sex-differential UKB |
| 0.003255 | 0.003512 | 3.204 | 1.25 | 17.88 | 13.33 | True | True | True | True | Sex-specific 1e-2 VUMC, sex-differential UKB |
| 0.003256 | 0.003513 | 3.217 | 1.184 | 17.79 | 13.15 | True | True | True | True | Sex-specific 1e-2 VUMC, sex-differential UKB |
| 0.003256 | 0.003514 | 3.216 | 1.191 | 17.76 | 13.18 | True | True | True | True | Sex-specific 1e-2 VUMC, sex-differential UKB |
| 0.003256 | 0.003514 | 3.216 | 1.191 | 17.76 | 13.18 | True | True | True | True | Sex-specific 1e-2 VUMC, sex-differential UKB |
| 0.003255 | 0.003513 | 3.225 | 1.152 | 17.63 | 13.19 | True | True | True | True | Sex-specific 1e-2 VUMC, sex-differential UKB |
| 0.003254 | 0.003511 | 3.184 | 1.142 | 17.55 | 12.98 | True | True | True | True | Sex-specific 1e-2 VUMC, sex-differential UKB |
| 0.003254 | 0.003511 | 3.217 | 1.152 | 17.53 | 13.03 | True | True | True | True | Sex-specific 1e-2 VUMC, sex-differential UKB |

| SE_UKB_female | SE_UKB_male | LOG10P_VUMC_female | LOG10P_VUMC_male | LOG10P_UKB_female | LOG10P_UKB_male | female_effect_direction_replication | male_effect_direction_replication | sex_effect_comparison_replication | pvalue_effect_comparison_replication | replication_type |
|---|---|---|---|---|---|---|---|---|---|---|
| 0.003255 | 0.003512 | 3.231 | 1.154 | 17.47 | 12.95 | True | True | True | True | Sex-specific 1e-2 VUMC, sex-differential UKB |
| 0.003254 | 0.003511 | 3.231 | 1.154 | 17.62 | 13.04 | True | True | True | True | Sex-specific 1e-2 VUMC, sex-differential UKB |
| 0.003251 | 0.0035 | 2.363 | 1.316 | 19.7 | 13.4 | True | True | True | True | Sex-specific 1e-2 VUMC, sex-differential UKB |
| 0.003751 | 0.004023 | 2.23 | 0.1907 | 12.07 | 11.52 | True | True | False | True | |
| 0.00345 | 0.003706 | 7.368 | 0.02334 | 14.02 | 13.05 | True | True | False | True | |
| 0.00345 | 0.003707 | 7.181 | 0.04836 | 14.01 | 13.01 | True | True | False | True | |
| 0.003689 | 0.003957 | 2.897 | 0.02118 | 11.55 | 11.12 | True | True | False | True | |
| 0.003453 | 0.00371 | 6.894 | 0.03587 | 14.05 | 13.04 | True | True | False | True | |
| 0.003693 | 0.003961 | 2.802 | 0.028 | 11.55 | 11.2 | True | True | False | True | |
| 0.003449 | 0.003706 | 6.608 | 0.02558 | 14.85 | 14.12 | True | False | False | True | |
| 0.003377 | 0.003632 | 5.149 | 0.07843 | 25.67 | 21.41 | True | True | True | True | Sex-specific 1e-2 VUMC, sex-differential UKB |
| 0.003548 | 0.003809 | 5.855 | 1.096 | 29.31 | 22.84 | True | True | True | True | Sex-specific 1e-2 VUMC, sex-differential UKB |
| 0.003548 | 0.003809 | 5.982 | 1.064 | 29.32 | 22.82 | True | True | True | True | Sex-specific 1e-2 VUMC, sex-differential UKB |
| 0.003506 | 0.003766 | 7.577 | 0.8386 | 28.11 | 22.73 | True | True | True | True | Sex-specific 1e-2 VUMC, sex-differential UKB |
| 0.003552 | 0.003814 | 6.298 | 1.088 | 29.14 | 23.36 | True | True | True | True | Sex-specific 1e-2 VUMC, sex-differential UKB |
| 0.003552 | 0.003815 | 6.278 | 1.091 | 29.13 | 23.36 | True | True | True | True | Sex-specific 1e-2 VUMC, sex-differential UKB |
| 0.003507 | 0.003767 | 7.62 | 0.8462 | 28.09 | 22.73 | True | True | True | True | Sex-specific 1e-2 VUMC, sex-differential UKB |
| 0.003552 | 0.003815 | 6.299 | 1.088 | 29.18 | 23.44 | True | True | True | True | Sex-specific 1e-2 VUMC, sex-differential UKB |
| 0.003509 | 0.003769 | 7.378 | 0.8558 | 28.63 | 22.88 | True | True | True | True | Sex-specific 1e-2 VUMC, sex-differential UKB |
| 0.003508 | 0.003767 | 7.404 | 0.8348 | 28.38 | 22.85 | True | True | True | True | Sex-specific 1e-2 VUMC, sex-differential UKB |
| 0.003509 | 0.003769 | 7.377 | 0.8557 | 28.62 | 22.89 | True | True | True | True | Sex-specific 1e-2 VUMC, sex-differential UKB |
| 0.003509 | 0.003769 | 7.376 | 0.8558 | 28.62 | 22.89 | True | True | True | True | Sex-specific 1e-2 VUMC, sex-differential UKB |
| 0.003465 | 0.00372 | 6.77 | 0.5954 | 26.72 | 20 | True | True | True | True | Sex-specific 1e-2 VUMC, sex-differential UKB |
| 0.003465 | 0.00372 | 6.77 | 0.5954 | 26.72 | 20 | True | True | True | True | Sex-specific 1e-2 VUMC, sex-differential UKB |
| 0.003465 | 0.00372 | 6.769 | 0.6117 | 26.59 | 19.89 | True | True | True | True | Sex-specific 1e-2 VUMC, sex-differential UKB |

| SE_UKB_female | SE_UKB_male | LOG10P_VUMC_female | LOG10P_VUMC_male | LOG10P_UKB_female | LOG10P_UKB_male | female_effect_direction_replication | male_effect_direction_replication | sex_effect_comparison_replication | pvalue_effect_comparison_replication | replication_type |
|---|---|---|---|---|---|---|---|---|---|---|
| 0.003465 | 0.003721 | 6.779 | 0.6124 | 26.59 | 19.82 | True | True | True | True | Sex-specific 1e-2 VUMC, sex-differential UKB |
| 0.003465 | 0.00372 | 6.798 | 0.605 | 26.62 | 20.05 | True | True | True | True | Sex-specific 1e-2 VUMC, sex-differential UKB |
| 0.00346 | 0.003717 | 6.972 | 0.5138 | 26.43 | 20.13 | True | True | True | True | Sex-specific 1e-2 VUMC, sex-differential UKB |
| 0.003465 | 0.00372 | 6.794 | 0.613 | 26.6 | 19.83 | True | True | True | True | Sex-specific 1e-2 VUMC, sex-differential UKB |
| 0.003463 | 0.003718 | 7.073 | 0.6011 | 26.15 | 19.68 | True | True | True | True | Sex-specific 1e-2 VUMC, sex-differential UKB |
| 0.003465 | 0.00372 | 6.796 | 0.6127 | 26.58 | 19.8 | True | True | True | True | Sex-specific 1e-2 VUMC, sex-differential UKB |
| 0.003465 | 0.00372 | 6.796 | 0.6127 | 26.58 | 19.8 | True | True | True | True | Sex-specific 1e-2 VUMC, sex-differential UKB |
| 0.003465 | 0.00372 | 6.78 | 0.6083 | 26.62 | 19.81 | True | True | True | True | Sex-specific 1e-2 VUMC, sex-differential UKB |
| 0.003465 | 0.00372 | 6.808 | 0.6157 | 26.59 | 19.82 | True | True | True | True | Sex-specific 1e-2 VUMC, sex-differential UKB |
| 0.003464 | 0.00372 | 7.046 | 0.6132 | 26.4 | 19.73 | True | True | True | True | Sex-specific 1e-2 VUMC, sex-differential UKB |
| 0.003464 | 0.00372 | 7.047 | 0.6131 | 26.39 | 19.73 | True | True | True | True | Sex-specific 1e-2 VUMC, sex-differential UKB |
| 0.003463 | 0.003718 | 7.072 | 0.5988 | 26.17 | 19.7 | True | True | True | True | Sex-specific 1e-2 VUMC, sex-differential UKB |
| 0.003554 | 0.003817 | 6.016 | 1.074 | 29.32 | 22.98 | True | True | True | True | Sex-specific 1e-2 VUMC, sex-differential UKB |
| 0.003463 | 0.003718 | 7.078 | 0.6023 | 26.21 | 19.65 | True | True | True | True | Sex-specific 1e-2 VUMC, sex-differential UKB |
| 0.003463 | 0.003718 | 7.036 | 0.615 | 26.2 | 19.63 | True | True | True | True | Sex-specific 1e-2 VUMC, sex-differential UKB |
| 0.003503 | 0.003761 | 5.638 | 0.8404 | 27.33 | 20.14 | True | True | True | True | Sex-specific 1e-2 VUMC, sex-differential UKB |
| 0.003474 | 0.003729 | 4.335 | 0.8762 | 24.14 | 15.95 | True | True | True | True | Sex-specific 1e-2 VUMC, sex-differential UKB |
| 0.003437 | 0.00369 | 5.606 | 0.6411 | 23.21 | 15.54 | True | True | True | True | Sex-specific 1e-2 VUMC, sex-differential UKB |
| 0.003474 | 0.003729 | 4.369 | 0.8821 | 24.14 | 15.96 | True | True | True | True | Sex-specific 1e-2 VUMC, sex-differential UKB |
| 0.003436 | 0.003689 | 5.611 | 0.6414 | 23.21 | 15.54 | True | True | True | True | Sex-specific 1e-2 VUMC, sex-differential UKB |
| 0.003524 | 0.003785 | 4.628 | 1.135 | 26.9 | 19.47 | True | True | True | True | Sex-specific 1e-2 VUMC, sex-differential UKB |
| 0.003482 | 0.00374 | 5.686 | 0.8788 | 26.34 | 19.19 | True | True | True | True | Sex-specific 1e-2 VUMC, sex-differential UKB |
| 0.003522 | 0.003782 | 4.956 | 1.132 | 26.77 | 19.86 | True | True | True | True | Sex-specific 1e-2 VUMC, sex-differential UKB |
| 0.003524 | 0.003785 | 4.624 | 1.14 | 26.9 | 19.5 | True | True | True | True | Sex-specific 1e-2 VUMC, sex-differential UKB |
| 0.003526 | 0.003788 | 4.749 | 1.143 | 27.16 | 19.58 | True | True | True | True | Sex-specific 1e-2 VUMC, sex-differential UKB |
| 0.004309 | 0.004887 | 8.211 | 0.4392 | 34.57 | 37.19 | True | True | False | False | |

| SE_UKB_female | SE_UKB_male | LOG10P_VUMC_female | LOG10P_VUMC_male | LOG10P_UKB_female | LOG10P_UKB_male | female_effect_direction_replication | male_effect_direction_replication | sex_effect_comparison_replication | pvalue_effect_comparison_replication | replication_type |
|---|---|---|---|---|---|---|---|---|---|---|
| 0.003985 | 0.004539 | 7.405 | 0.3403 | 38 | 35.98 | True | True | False | True | |
| 0.003986 | 0.004539 | 7.192 | 0.3211 | 37.97 | 35.98 | True | True | False | True | |
| 0.00431 | 0.004886 | 7.823 | 0.4136 | 34.44 | 37.36 | True | True | False | False | |
| 0.004374 | 0.00496 | 7.657 | 0.439 | 32.8 | 35.28 | True | True | False | False | |
| 0.004355 | 0.004939 | 7.497 | 0.4218 | 32.66 | 35.47 | True | True | False | False | |
| 0.004241 | 0.004808 | 8.767 | 0.5169 | 35.94 | 38.92 | True | True | False | False | |
| 0.004239 | 0.004807 | 8.859 | 0.525 | 35.88 | 38.87 | True | True | False | False | |
| 0.00424 | 0.004808 | 8.872 | 0.4909 | 36.01 | 38.92 | True | True | False | False | |
| 0.004107 | 0.004658 | 4.153 | 0.04682 | 13.32 | 10.95 | True | False | False | True | |
| 0.004241 | 0.004809 | 8.878 | 0.4923 | 36.13 | 39 | True | True | False | False | |
| 0.004241 | 0.00481 | 8.879 | 0.4923 | 36.12 | 39.01 | True | True | False | False | |
| 0.00424 | 0.004807 | 8.951 | 0.5096 | 36.03 | 38.85 | True | True | False | False | |
| 0.004241 | 0.004809 | 8.958 | 0.5042 | 36.12 | 38.83 | True | True | False | False | |
| 0.004241 | 0.004809 | 8.957 | 0.5032 | 36.11 | 38.82 | True | True | False | False | |
| 0.004688 | 0.005328 | 9.108 | 0.291 | 27.83 | 30.63 | True | True | False | False | |
| 0.004734 | 0.005376 | 8.848 | 0.3018 | 29.67 | 30.67 | True | True | False | False | |
| 0.004737 | 0.005379 | 8.747 | 0.2947 | 29.73 | 30.86 | True | True | False | False | |
| 0.004284 | 0.004885 | 9.832 | 0.06512 | 33.18 | 33.24 | True | True | False | False | |
| 0.004751 | 0.005394 | 8.753 | 0.2737 | 28.78 | 30.94 | True | True | False | False | |
| 0.006448 | 0.007339 | 4.543 | 0.1511 | 13.11 | 14.47 | True | False | False | False | |
| 0.006446 | 0.007337 | 4.608 | 0.1575 | 13.22 | 14.52 | True | False | False | False | |
| 0.004744 | 0.005387 | 8.875 | 0.358 | 29.71 | 32.07 | True | True | False | False | |
| 0.006477 | 0.007379 | 4.997 | 0.1724 | 12.18 | 13.86 | True | False | False | False | |
| 0.006532 | 0.007435 | 4.408 | 0.007412 | 10.81 | 14.25 | True | False | False | False | |
| SE_UKB_female | SE_UKB_male | LOG10P_VUMC_female | LOG10P_VUMC_male | LOG10P_UKB_female | LOG10P_UKB_male | female_effect_direction_replication | male_effect_direction_replication | sex_effect_comparison_replication | pvalue_effect_comparison_replication | replication_type |
| 0.006622 | 0.007551 | 4.504 | 0.1051 | 10.66 | 12.84 | True | False | False | False | |
| 0.005388 | 0.006164 | 7.444 | 0.3554 | 26.85 | 26.65 | True | True | False | True | |

| SE_UKB_female | SE_UKB_male | LOG10P_VUMC_female | LOG10P_VUMC_male | LOG10P_UKB_female | LOG10P_UKB_male | female_effect_direction_replication | male_effect_direction_replication | sex_effect_comparison_replication | pvalue_effect_comparison_replication | replication_type |
|---|---|---|---|---|---|---|---|---|---|---|
| 0.006682 | 0.007624 | 3.83 | 0.2467 | 10.52 | 12.55 | True | False | False | False | |
| 0.006563 | 0.007482 | 4.144 | 0.09531 | 9.968 | 12.52 | True | False | False | False | |
| 0.006502 | 0.007418 | 3.998 | 0.03779 | 10.69 | 12.52 | True | False | False | False | |
| 0.006502 | 0.007418 | 3.998 | 0.03779 | 10.7 | 12.52 | True | False | False | False | |
| 0.006498 | 0.007413 | 3.996 | 0.03633 | 10.62 | 12.52 | True | False | False | False | |
| 0.005611 | 0.006431 | 5.073 | 0.2234 | 21.3 | 23.3 | True | True | False | False | |
| 0.006496 | 0.007411 | 3.997 | 0.04126 | 10.46 | 12.69 | True | False | False | False | |
| 0.006491 | 0.007405 | 3.993 | 0.04022 | 10.59 | 12.43 | True | False | False | False | |
| 0.006498 | 0.00741 | 4.034 | 0.05802 | 10.5 | 12.47 | True | False | False | False | |
| 0.006538 | 0.007457 | 4.013 | 0.05334 | 10.14 | 12.23 | True | False | False | False | |
| 0.005608 | 0.006431 | 5.001 | 0.2207 | 21.24 | 23.47 | True | True | False | False | |
| 0.005608 | 0.006431 | 4.966 | 0.2158 | 21.23 | 23.47 | True | True | False | False | |
| 0.005606 | 0.006431 | 4.88 | 0.2194 | 21.23 | 23.54 | True | True | False | False | |
| 0.005607 | 0.006431 | 4.985 | 0.2178 | 21.24 | 23.55 | True | True | False | False | |
| 0.005602 | 0.006425 | 4.599 | 0.1824 | 20.67 | 23.13 | True | True | False | False | |
| 0.005607 | 0.00643 | 5.087 | 0.2019 | 20.95 | 23.54 | True | True | False | False | |
| 0.006627 | 0.007559 | 3.653 | 0.3049 | 10.05 | 12.12 | True | False | False | False | |
| 0.006503 | 0.00742 | 3.737 | 0.08661 | 9.924 | 12.19 | True | False | False | False | |
| 0.006505 | 0.007421 | 3.773 | 0.06803 | 9.9 | 12.17 | True | False | False | False | |
| 0.006564 | 0.007482 | 3.892 | 0.1613 | 9.183 | 12.16 | True | False | False | False | |
| 0.006564 | 0.007482 | 3.889 | 0.1619 | 9.183 | 12.12 | True | False | False | False | |
| 0.006565 | 0.007482 | 3.897 | 0.1634 | 9.182 | 12.12 | True | False | False | False | |
| 0.006582 | 0.007504 | 3.4 | 0.1522 | 9.203 | 11.97 | True | False | False | False | |
| 0.007179 | 0.008177 | 3.582 | 0.3276 | 7.26 | 10.88 | True | False | False | False | |
| 0.003345 | 0.003806 | 7.068 | 0.8955 | 6.24 | 4.377 | True | True | True | True | Sex-specific 1e-2 VUMC, sex-differential UKB |
| 0.003344 | 0.003806 | 6.958 | 0.7926 | 6.12 | 4.496 | True | True | True | True | Sex-specific 1e-2 VUMC, sex-differential UKB |
| 0.003159 | 0.00359 | 8.711 | 1.037 | 22.38 | 16.73 | True | True | True | True | Sex-specific 1e-2 VUMC, sex-differential UKB |

| SE_UKB_female | SE_UKB_male | LOG10P_VUMC_female | LOG10P_VUMC_male | LOG10P_UKB_female | LOG10P_UKB_male | female_effect_direction_replication | male_effect_direction_replication | sex_effect_comparison_replication | pvalue_effect_comparison_replication | replication_type |
|---|---|---|---|---|---|---|---|---|---|---|
| 0.003112 | 0.003544 | 4.954 | 1.327 | 16.63 | 15.93 | True | True | False | True | |
| 0.00311 | 0.003543 | 5.031 | 1.25 | 15.9 | 15.46 | True | True | False | True | |
| 0.003106 | 0.003539 | 5.071 | 1.296 | 15.86 | 15.37 | True | True | False | True | |
| 0.003846 | 0.004285 | 5.113 | 0.09944 | 1.717 | 2.99 | True | False | False | False | |
| 0.003854 | 0.004298 | 4.99 | 0.1184 | 1.849 | 3.147 | True | False | False | False | |
| 0.003813 | 0.004255 | 6.172 | 0.2595 | 3.57 | 4.047 | True | False | False | False | |
| 0.004158 | 0.004646 | 6.296 | 0.1826 | 3.296 | 6.25 | True | False | False | False | |
| 0.004281 | 0.004781 | 4.947 | 0.0523 | 6.907 | 6.327 | True | False | False | True | |
| 0.004073 | 0.004552 | 5.069 | 0.1022 | 4.943 | 5.788 | True | True | False | False | |
| 0.004338 | 0.004848 | 5.085 | 0.0895 | 6.827 | 5.703 | True | False | False | True | |
| 0.003727 | 0.004165 | 7.439 | 0.2415 | 3.449 | 5.043 | True | False | False | False | |
| 0.003729 | 0.004166 | 7.444 | 0.2276 | 3.511 | 5.21 | True | False | False | False | |
| 0.003729 | 0.004167 | 7.297 | 0.2293 | 3.568 | 5.2 | True | False | False | False | |
| 0.003815 | 0.004274 | 6.996 | 0.2045 | 4.504 | 3.767 | True | False | False | True | |
| 0.003988 | 0.004471 | 5.521 | 0.07692 | 4.984 | 5.259 | True | True | False | False | |
| 0.003815 | 0.004274 | 6.995 | 0.2047 | 4.503 | 3.755 | True | False | False | True | |
| 0.003815 | 0.004274 | 7.046 | 0.2146 | 4.479 | 3.843 | True | False | False | True | |
| 0.003987 | 0.004469 | 5.433 | 0.07814 | 4.984 | 5.423 | True | True | False | False | |
| 0.004441 | 0.004727 | 1.379 | 2.211 | 13.01 | 12.88 | True | True | True | False | |
| 0.003511 | 0.003744 | 0.09181 | 4.871 | 13.37 | 16.37 | True | True | True | True | Sex-specific 1e-2 VUMC, sex-differential UKB |
| 0.003512 | 0.003744 | 0.09238 | 4.85 | 13.4 | 16.43 | True | True | True | True | Sex-specific 1e-2 VUMC, sex-differential UKB |
| 0.003529 | 0.003758 | 0.04311 | 4.192 | 12.57 | 15.83 | False | True | True | True | Sex-specific 1e-2 VUMC, sex-differential UKB |
| 0.004409 | 0.004705 | 1.174 | 3.257 | 11.91 | 12.76 | True | True | True | True | Sex-specific 1e-2 VUMC, sex-differential UKB |
| | | 0.04617 | 3.53 | | | False | False | False | False | |
| 0.00354 | 0.003775 | 0.2283 | 5.126 | 13.6 | 19.18 | True | True | True | True | Sex-specific 1e-2 VUMC, sex-differential UKB |
| 0.003541 | 0.003776 | 0.2472 | 5.116 | 13.7 | 19.15 | True | True | True | True | Sex-specific 1e-2 VUMC, sex-differential UKB |

| SE_UKB_female | SE_UKB_male | LOG10P_VUMC_female | LOG10P_VUMC_male | LOG10P_UKB_female | LOG10P_UKB_male | female_effect_direction_replication | male_effect_direction_replication | sex_effect_comparison_replication | pvalue_effect_comparison_replication | replication_type |
|---|---|---|---|---|---|---|---|---|---|---|
| 0.003538 | 0.003773 | 0.2288 | 5.143 | 13.74 | 19.18 | True | True | True | True | Sex-specific 1e-2 VUMC, sex-differential UKB |
| 0.003694 | 0.003937 | 0.04703 | 3.392 | 4.251 | 14.12 | True | True | True | True | Sex-specific 1e-2 VUMC, sex-specific 5e-8 UKB |
| 0.003694 | 0.003937 | 0.04703 | 3.392 | 4.258 | 14.12 | True | True | True | True | Sex-specific 1e-2 VUMC, sex-specific 5e-8 UKB |
| 0.003695 | 0.003937 | 0.04711 | 3.391 | 4.264 | 14.11 | True | True | True | True | Sex-specific 1e-2 VUMC, sex-specific 5e-8 UKB |
| 0.003695 | 0.003937 | 0.04713 | 3.391 | 4.267 | 14.1 | True | True | True | True | Sex-specific 1e-2 VUMC, sex-specific 5e-8 UKB |
| SE_UKB_female | SE_UKB_male | LOG10P_VUMC_female | LOG10P_VUMC_male | LOG10P_UKB_female | LOG10P_UKB_male | female_effect_direction_replication | male_effect_direction_replication | sex_effect_comparison_replication | pvalue_effect_comparison_replication | replication_type |
| 0.003695 | 0.003937 | 0.02913 | 3.338 | 4.248 | 14.16 | True | True | True | True | Sex-specific 1e-2 VUMC, sex-specific 5e-8 UKB |
| 0.003538 | 0.003773 | 0.2455 | 5.31 | 13.75 | 19.1 | True | True | True | True | Sex-specific 1e-2 VUMC, sex-differential UKB |
| 0.0037 | 0.00394 | 0.1135 | 3.302 | 3.91 | 14.07 | True | True | True | True | Sex-specific 1e-2 VUMC, sex-specific 5e-8 UKB |
| 0.00369 | 0.003934 | 0.07312 | 3.756 | 4.342 | 14.87 | True | True | True | True | Sex-specific 1e-2 VUMC, sex-specific 5e-8 UKB |
| 0.003542 | 0.003777 | 0.246 | 5.928 | 14.44 | 19.81 | True | True | True | True | Sex-specific 1e-2 VUMC, sex-differential UKB |
| 0.003542 | 0.003778 | 0.2458 | 5.887 | 14.45 | 19.81 | True | True | True | True | Sex-specific 1e-2 VUMC, sex-differential UKB |
| 0.003542 | 0.003778 | 0.2414 | 5.887 | 14.47 | 19.88 | True | True | True | True | Sex-specific 1e-2 VUMC, sex-differential UKB |
| 0.003693 | 0.003937 | 0.06531 | 3.835 | 4.333 | 14.83 | True | True | True | True | Sex-specific 1e-2 VUMC, sex-specific 5e-8 UKB |
| 0.00367 | 0.00391 | 0.4427 | 6.092 | 19.47 | 21.64 | True | True | True | True | Sex-specific 1e-2 VUMC, sex-differential UKB |
| 0.003707 | 0.00395 | 0.00244 | 4.148 | 4.244 | 15.22 | True | True | True | True | Sex-specific 1e-2 VUMC, sex-specific 5e-8 UKB |
| 0.003832 | 0.004086 | 0.7746 | 5.124 | 14.67 | 16.34 | True | True | True | True | Sex-specific 1e-2 VUMC, sex-differential UKB |
| 0.003725 | 0.003967 | 0.0354 | 4.024 | 4.303 | 15.17 | True | True | True | True | Sex-specific 1e-2 VUMC, sex-specific 5e-8 UKB |
| 0.003686 | 0.003927 | 0.03159 | 4.453 | 3.68 | 14.35 | True | True | True | True | Sex-specific 1e-2 VUMC, sex-specific 5e-8 UKB |
| 0.003598 | 0.003836 | 0.5873 | 7.586 | 20.56 | 24.57 | True | True | True | True | Sex-specific 1e-2 VUMC, sex-differential UKB |
| 0.003828 | 0.004078 | 0.678 | 7.558 | 22.7 | 26.82 | True | True | True | True | Sex-specific 1e-2 VUMC, sex-differential UKB |
| 0.003687 | 0.003927 | 0.04726 | 4.479 | 3.821 | 14.38 | True | True | True | True | Sex-specific 1e-2 VUMC, sex-specific 5e-8 UKB |
| 0.003827 | 0.004077 | 0.6724 | 7.602 | 22.72 | 26.91 | True | True | True | True | Sex-specific 1e-2 VUMC, sex-differential UKB |
| 0.003455 | 0.003686 | 0.3835 | 10.44 | 13.65 | 24.94 | True | True | True | True | Sex-specific 1e-2 VUMC, sex-differential UKB |
| 0.003684 | 0.003924 | 0.03525 | 4.398 | 3.821 | 14.28 | True | True | True | True | Sex-specific 1e-2 VUMC, sex-specific 5e-8 UKB |
| 0.003455 | 0.003686 | 0.3917 | 10.49 | 13.66 | 24.86 | True | True | True | True | Sex-specific 1e-2 VUMC, sex-differential UKB |

| SE_UKB_female | SE_UKB_male | LOG10P_VUMC_female | LOG10P_VUMC_male | LOG10P_UKB_female | LOG10P_UKB_male | female_effect_direction_replication | male_effect_direction_replication | sex_effect_comparison_replication | pvalue_effect_comparison_replication | replication_type |
|---|---|---|---|---|---|---|---|---|---|---|
| 0.003465 | 0.003691 | 0.6739 | 8.983 | 15.04 | 27.43 | True | True | True | True | Sex-specific 1e-2 VUMC, sex-differential UKB |
| 0.003826 | 0.004081 | 0.6555 | 4.007 | 4.294 | 11.32 | True | True | True | True | Sex-specific 1e-2 VUMC, sex-specific 5e-8 UKB |
| 0.003482 | 0.003709 | 1.181 | 7.312 | 19.73 | 35.59 | True | True | True | True | Sex-specific 1e-2 VUMC, sex-differential UKB |
| 0.003828 | 0.004083 | 0.6555 | 3.92 | 4.219 | 11.29 | True | True | True | True | Sex-specific 1e-2 VUMC, sex-specific 5e-8 UKB |
| 0.003828 | 0.004083 | 0.6603 | 3.837 | 4.174 | 11.31 | True | True | True | True | Sex-specific 1e-2 VUMC, sex-specific 5e-8 UKB |
| 0.003827 | 0.004082 | 0.7129 | 3.852 | 4.165 | 11.54 | True | True | True | True | Sex-specific 1e-2 VUMC, sex-specific 5e-8 UKB |
| 0.003686 | 0.003931 | 0.1217 | 4.091 | 7.933 | 16.79 | True | True | True | True | Sex-specific 1e-2 VUMC, sex-differential UKB |
| 0.003493 | 0.003703 | 1.717 | 5.696 | 19.94 | 33.26 | True | True | True | True | Sex-specific 1e-2 VUMC, sex-differential UKB |
| 0.003526 | 0.003754 | 0.8821 | 6.864 | 24.23 | 44.34 | True | True | True | True | Sex-specific 1e-2 VUMC, sex-differential UKB |
| 0.003526 | 0.003753 | 0.934 | 7.035 | 24.18 | 44.38 | True | True | True | True | Sex-specific 1e-2 VUMC, sex-differential UKB |
| 0.003472 | 0.00369 | 1.131 | 6.884 | 20.56 | 39.69 | True | True | True | True | Sex-specific 1e-2 VUMC, sex-differential UKB |
| 0.003523 | 0.003751 | 0.8558 | 6.918 | 24.03 | 44.33 | True | True | True | True | Sex-specific 1e-2 VUMC, sex-differential UKB |
| 0.003472 | 0.00369 | 1.041 | 6.748 | 20.56 | 39.68 | True | True | True | True | Sex-specific 1e-2 VUMC, sex-differential UKB |
| 0.003472 | 0.00369 | 1.046 | 6.749 | 20.61 | 39.71 | True | True | True | True | Sex-specific 1e-2 VUMC, sex-differential UKB |
| 0.003497 | 0.003725 | 0.8765 | 7.997 | 23.86 | 47.75 | True | True | True | True | Sex-specific 1e-2 VUMC, sex-differential UKB |
| 0.003472 | 0.00369 | 1.046 | 6.749 | 20.62 | 39.73 | True | True | True | True | Sex-specific 1e-2 VUMC, sex-differential UKB |
| 0.003472 | 0.00369 | 1.044 | 6.732 | 20.6 | 39.55 | True | True | True | True | Sex-specific 1e-2 VUMC, sex-differential UKB |
| 0.003472 | 0.00369 | 1.05 | 6.767 | 20.56 | 39.76 | True | True | True | True | Sex-specific 1e-2 VUMC, sex-differential UKB |
| 0.003472 | 0.00369 | 1.05 | 6.745 | 20.57 | 39.76 | True | True | True | True | Sex-specific 1e-2 VUMC, sex-differential UKB |
| 0.002827 | 0.003052 | 9.619 | 1.896 | 0.5201 | 2.699 | True | True | False | False | |
| 0.0028 | 0.003025 | 10.42 | 1.875 | 0.613 | 2.94 | True | True | False | False | |
| 0.002835 | 0.00306 | 10.25 | 1.982 | 0.3452 | 3.379 | True | True | False | False | |
| 0.0028 | 0.003024 | 10.35 | 1.912 | 0.617 | 2.95 | True | True | False | False | |
| 0.002827 | 0.003052 | 9.624 | 1.887 | 0.5299 | 2.709 | True | True | False | False | |
| 0.002801 | 0.003025 | 10.6 | 1.81 | 0.6314 | 3.073 | True | True | False | False | |
| 0.002801 | 0.003025 | 10.54 | 1.779 | 0.6159 | 3.059 | True | True | False | False | |

| SE_UKB_female | SE_UKB_male | LOG10P_VUMC_female | LOG10P_VUMC_male | LOG10P_UKB_female | LOG10P_UKB_male | female_effect_direction_replication | male_effect_direction_replication | sex_effect_comparison_replication | pvalue_effect_comparison_replication | replication_type |
|---|---|---|---|---|---|---|---|---|---|---|
| 0.002807 | 0.003032 | 10.98 | 1.946 | 0.4354 | 3.655 | True | True | False | False | |
| 0.002827 | 0.003052 | 9.612 | 1.866 | 0.5255 | 2.714 | True | True | False | False | |
| 0.002827 | 0.003052 | 9.573 | 1.852 | 0.5288 | 2.71 | True | True | False | False | |
| 0.0028 | 0.003025 | 10.32 | 1.858 | 0.6298 | 2.956 | True | True | False | False | |
| 0.002801 | 0.003025 | 10.32 | 1.9 | 0.6422 | 2.965 | True | True | False | False | |
| 0.002801 | 0.003025 | 10.47 | 1.764 | 0.6198 | 3.079 | True | True | False | False | |
| 0.002828 | 0.003052 | 9.799 | 1.77 | 0.5679 | 2.789 | True | True | False | False | |
| 0.002801 | 0.003025 | 10.49 | 1.775 | 0.6073 | 2.962 | True | True | False | False | |
| 0.0028 | 0.003024 | 10.56 | 1.714 | 0.6371 | 3.121 | True | True | False | False | |
| 0.002799 | 0.003023 | 10.59 | 1.757 | 0.6343 | 3.029 | True | True | False | False | |
| 0.00279 | 0.003013 | 10.78 | 1.577 | 0.8185 | 3.117 | True | True | False | False | |
| 0.002808 | 0.003033 | 10.87 | 1.88 | 0.3891 | 3.796 | True | True | False | False | |
| 0.002808 | 0.003033 | 10.58 | 1.907 | 0.3457 | 3.756 | True | True | False | False | |
| 0.002807 | 0.003032 | 10.94 | 1.879 | 0.3968 | 3.9 | True | True | False | False | |

| SE_UKB_female | SE_UKB_male | LOG10P_VUMC_female | LOG10P_VUMC_male | LOG10P_UKB_female | LOG10P_UKB_male | female_effect_direction_replication | male_effect_direction_replication | sex_effect_comparison_replication | pvalue_effect_comparison_replication | replication_type |
|---|---|---|---|---|---|---|---|---|---|---|
| 0.002807 | 0.003032 | 10.94 | 1.879 | 0.3969 | 3.901 | True | True | False | False | |
| 0.002807 | 0.003032 | 10.91 | 1.855 | 0.3938 | 3.904 | True | True | False | False | |
| 0.002807 | 0.003032 | 10.91 | 1.853 | 0.4039 | 3.891 | True | True | False | False | |
| 0.002807 | 0.003032 | 10.83 | 1.899 | 0.3554 | 3.844 | True | True | False | False | |
| 0.002939 | 0.003173 | 8.75 | 0.9498 | 0.4486 | 2.567 | True | True | False | False | |
| 0.002941 | 0.003175 | 8.71 | 0.9129 | 0.4308 | 2.719 | True | True | False | False | |
| 0.002947 | 0.003181 | 8.819 | 0.9618 | 0.3769 | 2.7 | True | True | False | False | |
| 0.002947 | 0.003181 | 8.821 | 0.9505 | 0.3723 | 2.693 | True | True | False | False | |
| 0.002941 | 0.003175 | 8.712 | 0.9164 | 0.4359 | 2.691 | True | True | False | False | |
| 0.002941 | 0.003175 | 8.716 | 0.9169 | 0.4347 | 2.697 | True | True | False | False | |
| 0.002943 | 0.003176 | 8.536 | 0.933 | 0.4373 | 2.713 | True | True | False | False | |

| SE_UKB_female | SE_UKB_male | LOG10P_VUMC_female | LOG10P_VUMC_male | LOG10P_UKB_female | LOG10P_UKB_male | female_effect_direction_replication | male_effect_direction_replication | sex_effect_comparison_replication | pvalue_effect_comparison_replication | replication_type |
|---|---|---|---|---|---|---|---|---|---|---|
| 0.002943 | 0.003176 | 8.532 | 0.9336 | 0.4349 | 2.724 | True | True | False | False | |
| 0.002943 | 0.003176 | 8.649 | 0.9117 | 0.4395 | 2.665 | True | True | False | False | |
| 0.002946 | 0.00318 | 8.619 | 0.8974 | 0.4438 | 2.587 | True | True | False | False | |
| 0.002813 | 0.003037 | 8.604 | 0.4854 | 0.228 | 2.363 | True | True | False | False | |
| 0.002814 | 0.003038 | 8.665 | 0.4752 | 0.2254 | 2.4 | True | True | False | False | |
| 0.003408 | 0.003684 | 6.66 | 0.4239 | 0.05649 | 1.207 | True | True | False | False | |
| 0.003114 | 0.003364 | 7.54 | 0.905 | 0.05315 | 1.247 | False | True | False | False | |
| 0.00351 | 0.003789 | 4.631 | 0.8134 | 0.3124 | 1.121 | True | False | False | False | |
| 0.003501 | 0.003777 | 4.909 | 0.5068 | 0.2948 | 1.15 | True | False | False | False | |
| 0.003507 | 0.003786 | 5.082 | 0.6357 | 0.2898 | 1.187 | True | False | False | False | |
| 0.003498 | 0.003774 | 5.069 | 0.4876 | 0.2686 | 1.132 | True | False | False | False | |
| 0.003507 | 0.003785 | 4.888 | 0.724 | 0.3002 | 1.073 | True | False | False | False | |
| 0.003504 | 0.003783 | 4.87 | 0.7319 | 0.2943 | 1.064 | True | False | False | False | |
| 0.003476 | 0.003751 | 5.055 | 0.7029 | 0.3212 | 1.008 | True | False | False | False | |
| 0.003406 | 0.003677 | 3.764 | 1.022 | 0.3835 | 0.5994 | True | False | False | False | |
| 0.00347 | 0.003744 | 4.72 | 0.616 | 0.4392 | 0.798 | True | False | False | False | |
| 0.00347 | 0.003744 | 4.64 | 0.5381 | 0.4409 | 0.7683 | True | False | False | False | |
| 0.00347 | 0.003747 | 5.133 | 0.3951 | 0.3332 | 0.5674 | True | False | False | False | |
| 0.00347 | 0.003748 | 5.359 | 0.4015 | 0.3029 | 0.5201 | True | False | False | False | |
| 0.003469 | 0.003746 | 5.311 | 0.3953 | 0.3272 | 0.5025 | True | False | False | False | |
| 0.003468 | 0.003746 | 5.222 | 0.3659 | 0.351 | 0.5178 | True | False | False | False | |
| 0.003469 | 0.003747 | 5.22 | 0.366 | 0.3468 | 0.5164 | True | False | False | False | |
| 0.003472 | 0.00375 | 5.309 | 0.3808 | 0.332 | 0.5217 | True | False | False | False | |
| 0.003472 | 0.00375 | 5.384 | 0.4007 | 0.3464 | 0.5198 | True | False | False | False | |
| 0.003469 | 0.003747 | 5.254 | 0.3854 | 0.3423 | 0.5191 | True | False | False | False | |
| 0.00347 | 0.003748 | 5.314 | 0.4014 | 0.3503 | 0.5233 | True | False | False | False | |
| 0.00347 | 0.003748 | 5.193 | 0.3876 | 0.3515 | 0.5082 | True | False | False | False | |

| SE_UKB_female | SE_UKB_male | LOG10P_VUMC_female | LOG10P_VUMC_male | LOG10P_UKB_female | LOG10P_UKB_male | female_effect_direction_replication | male_effect_direction_replication | sex_effect_comparison_replication | pvalue_effect_comparison_replication | replication_type |
|---|---|---|---|---|---|---|---|---|---|---|
| 0.003469 | 0.003747 | 5.359 | 0.4008 | 0.316 | 0.5306 | True | False | False | False | |
| 0.00347 | 0.003748 | 5.344 | 0.4077 | 0.3635 | 0.5038 | True | False | False | False | |
| 0.003479 | 0.003758 | 4.977 | 0.3519 | 0.3549 | 0.5033 | True | False | False | False | |
| 0.003476 | 0.003755 | 5.201 | 0.436 | 0.3067 | 0.5253 | True | False | False | False | |
| 0.003473 | 0.003752 | 5.228 | 0.3863 | 0.347 | 0.5149 | True | False | False | False | |
| 0.003474 | 0.003752 | 5.25 | 0.3908 | 0.3463 | 0.5401 | True | False | False | False | |
| 0.00345 | 0.003729 | 4.795 | 0.31 | 0.411 | 0.6717 | True | False | False | False | |
| 0.003444 | 0.003723 | 4.914 | 0.313 | 0.372 | 0.5472 | True | False | False | False | |
| 0.003445 | 0.003723 | 4.981 | 0.2764 | 0.3738 | 0.5437 | True | False | False | False | |
| 0.003446 | 0.003725 | 5.084 | 0.2705 | 0.3919 | 0.565 | True | False | False | False | |
| 0.003447 | 0.003726 | 4.992 | 0.2859 | 0.3934 | 0.5744 | True | False | False | False | |
| 0.003372 | 0.003638 | 5.531 | 0.6969 | 1.223 | 0.5296 | True | True | True | True | Sex-specific 1e-2 VUMC, sex-differential UKB |
| 0.003446 | 0.003725 | 5.089 | 0.2652 | 0.3882 | 0.554 | True | False | False | False | |
| 0.003446 | 0.003725 | 5.089 | 0.2652 | 0.3881 | 0.554 | True | False | False | False | |
| 0.003478 | 0.003759 | 5.607 | 0.3578 | 0.3802 | 0.5896 | True | False | False | False | |
| 0.003446 | 0.003725 | 5.129 | 0.2827 | 0.3886 | 0.5527 | True | False | False | False | |
| 0.003372 | 0.003638 | 5.51 | 0.6777 | 1.217 | 0.5062 | True | True | True | True | Sex-specific 1e-2 VUMC, sex-differential UKB |
| 0.003372 | 0.003638 | 5.508 | 0.6802 | 1.222 | 0.5083 | True | True | True | True | Sex-specific 1e-2 VUMC, sex-differential UKB |
| 0.003478 | 0.003759 | 5.566 | 0.3569 | 0.3574 | 0.5845 | True | False | False | False | |
| 0.003372 | 0.003638 | 5.543 | 0.6652 | 1.184 | 0.4885 | True | True | True | True | Sex-specific 1e-2 VUMC, sex-differential UKB |
| 0.003372 | 0.003639 | 5.573 | 0.6691 | 1.222 | 0.4839 | True | True | True | True | Sex-specific 1e-2 VUMC, sex-differential UKB |
| 0.003447 | 0.003726 | 5.174 | 0.2907 | 0.3922 | 0.5313 | True | False | False | False | |

| SE_UKB_female | SE_UKB_male | LOG10P_VUMC_female | LOG10P_VUMC_male | LOG10P_UKB_female | LOG10P_UKB_male | female_effect_direction_replication | male_effect_direction_replication | sex_effect_comparison_replication | pvalue_effect_comparison_replication | replication_type |
|---|---|---|---|---|---|---|---|---|---|---|
| 0.003374 | 0.00364 | 5.703 | 0.6422 | 1.193 | 0.453 | True | True | True | True | Sex-specific 1e-2 VUMC, sex-differential UKB |
| 0.003449 | 0.003728 | 5.309 | 0.3088 | 0.3902 | 0.5387 | True | False | False | False | |
| 0.003374 | 0.00364 | 5.814 | 0.6418 | 1.184 | 0.4519 | True | True | True | True | Sex-specific 1e-2 VUMC, sex-differential UKB |

| SE_UKB_female | SE_UKB_male | LOG10P_VUMC_female | LOG10P_VUMC_male | LOG10P_UKB_female | LOG10P_UKB_male | female_effect_direction_replication | male_effect_direction_replication | sex_effect_comparison_replication | pvalue_effect_comparison_replication | replication_type |
|---|---|---|---|---|---|---|---|---|---|---|
| 0.003401 | 0.003667 | 5.735 | 0.4717 | 1.671 | 0.8457 | True | True | True | True | Sex-specific 1e-2 VUMC, sex-differential UKB |
| 0.003408 | 0.003679 | 6.192 | 0.1416 | 0.8856 | 0.7945 | True | False | False | True | |
| 0.0034 | 0.003665 | 5.745 | 0.4736 | 1.637 | 0.8343 | True | True | True | True | Sex-specific 1e-2 VUMC, sex-differential UKB |
| 0.003367 | 0.00363 | 9.321 | 0.1225 | 0.202 | 1.287 | True | True | False | False | |
| 0.003367 | 0.003629 | 9.313 | 0.1259 | 0.2057 | 1.289 | True | True | False | False | |
| 0.003398 | 0.003663 | 5.88 | 0.4778 | 1.643 | 0.844 | True | True | True | True | Sex-specific 1e-2 VUMC, sex-differential UKB |
| 0.003371 | 0.003635 | 9.339 | 0.01255 | 0.2121 | 1.198 | True | False | False | False | |
| 0.003365 | 0.003627 | 9.472 | 0.07916 | 0.2095 | 1.268 | True | True | False | False | |
| 0.00337 | 0.003633 | 9.121 | 0.04748 | 0.1878 | 1.17 | True | False | False | False | |
| 0.003364 | 0.003626 | 9.37 | 0.08742 | 0.2016 | 1.289 | True | True | False | False | |
| 0.00337 | 0.003634 | 8.991 | 0.01707 | 0.2218 | 1.202 | True | False | False | False | |
| 0.003363 | 0.003624 | 9.671 | 0.1197 | 0.2041 | 1.183 | True | True | False | False | |
| 0.003369 | 0.003632 | 9.382 | 0.007927 | 0.193 | 1.105 | True | False | False | False | |
| 0.00337 | 0.003633 | 9.343 | 0.006212 | 0.1856 | 1.089 | True | False | False | False | |
| 0.003365 | 0.003626 | 9.6 | 0.09347 | 0.1742 | 1.171 | True | True | False | False | |
| 0.003372 | 0.003635 | 9.29 | 0.007654 | 0.1952 | 1.123 | True | False | False | False | |
| 0.003402 | 0.00367 | 5.661 | 0.5297 | 1.123 | 0.5974 | True | True | True | True | Sex-specific 1e-2 VUMC, sex-differential UKB |
| 0.003318 | 0.003581 | 7.027 | 0.8173 | 1.289 | 1.238 | True | True | False | True | |
| 0.003267 | 0.003519 | 6.766 | 0.3098 | 1.789 | 1.225 | True | True | True | True | Sex-specific 1e-2 VUMC, sex-differential UKB |
| 0.003277 | 0.003537 | 6.675 | 0.3004 | 1.126 | 1.041 | True | True | False | True | |
| 0.003317 | 0.00358 | 7.043 | 0.8049 | 1.291 | 1.242 | True | True | False | True | |
| 0.003317 | 0.00358 | 7.081 | 0.7861 | 1.289 | 1.242 | True | True | False | True | |
| 0.003308 | 0.003573 | 6.6 | 0.5626 | 1.298 | 1.081 | True | True | True | True | Sex-specific 1e-2 VUMC, sex-differential UKB |
| 0.003308 | 0.003573 | 6.567 | 0.5741 | 1.252 | 1.063 | True | True | True | True | Sex-specific 1e-2 VUMC, sex-differential UKB |
| 0.003401 | 0.003669 | 5.803 | 0.5149 | 1.117 | 0.6284 | True | True | True | True | Sex-specific 1e-2 VUMC, sex-differential UKB |
| 0.003277 | 0.003537 | 6.653 | 0.3049 | 1.066 | 1.009 | True | True | False | True | |
| 0.003308 | 0.003573 | 6.585 | 0.574 | 1.251 | 1.061 | True | True | True | True | Sex-specific 1e-2 VUMC, sex-differential UKB |

| SE_UKB_female | SE_UKB_male | LOG10P_VUMC_female | LOG10P_VUMC_male | LOG10P_UKB_female | LOG10P_UKB_male | female_effect_direction_replication | male_effect_direction_replication | sex_effect_comparison_replication | pvalue_effect_comparison_replication | replication_type |
|---|---|---|---|---|---|---|---|---|---|---|
| 0.003308 | 0.003573 | 6.584 | 0.5739 | 1.247 | 1.07 | True | True | True | True | Sex-specific 1e-2 VUMC, sex-differential UKB |
| 0.003308 | 0.003572 | 6.389 | 0.7602 | 1.228 | 1.166 | True | True | False | True | |
| 0.003308 | 0.003573 | 6.57 | 0.5738 | 1.248 | 1.073 | True | True | True | True | Sex-specific 1e-2 VUMC, sex-differential UKB |
| 0.003309 | 0.003573 | 6.564 | 0.5018 | 1.284 | 1.076 | True | True | True | True | Sex-specific 1e-2 VUMC, sex-differential UKB |
| 0.003278 | 0.003537 | 6.666 | 0.251 | 1.125 | 1.027 | True | True | False | True | |
| 0.003309 | 0.003573 | 6.524 | 0.5048 | 1.294 | 1.057 | True | True | True | True | Sex-specific 1e-2 VUMC, sex-differential UKB |
| 0.003309 | 0.003573 | 6.526 | 0.499 | 1.29 | 1.058 | True | True | True | True | Sex-specific 1e-2 VUMC, sex-differential UKB |
| 0.003307 | 0.003571 | 6.467 | 0.4863 | 1.342 | 1.121 | True | True | True | True | Sex-specific 1e-2 VUMC, sex-differential UKB |
| 0.003314 | 0.003579 | 6.665 | 0.4857 | 1.413 | 1.177 | True | True | True | True | Sex-specific 1e-2 VUMC, sex-differential UKB |
| 0.003314 | 0.00358 | 6.656 | 0.4189 | 1.473 | 1.141 | True | True | True | True | Sex-specific 1e-2 VUMC, sex-differential UKB |
| 0.003292 | 0.003558 | 7.655 | 0.1402 | 1.606 | 0.8221 | True | True | True | True | Sex-specific 1e-2 VUMC, sex-differential UKB |
| 0.003427 | 0.003696 | 6.686 | 0.375 | 2.565 | 0.9504 | True | True | True | True | Sex-specific 1e-2 VUMC, sex-differential UKB |
| 0.003271 | 0.00353 | 8.217 | 0.02615 | 1.5 | 0.9691 | True | True | True | True | Sex-specific 1e-2 VUMC, sex-differential UKB |
| 0.003361 | 0.003623 | 6.914 | 0.1598 | 2.269 | 0.822 | True | True | True | True | Sex-specific 1e-2 VUMC, sex-differential UKB |
| 0.003424 | 0.003694 | 6.734 | 0.3603 | 2.607 | 0.9132 | True | True | True | True | Sex-specific 1e-2 VUMC, sex-differential UKB |
| 0.003427 | 0.003697 | 6.796 | 0.3663 | 2.626 | 0.8327 | True | True | True | True | Sex-specific 1e-2 VUMC, sex-differential UKB |
| 0.003428 | 0.003698 | 6.911 | 0.3472 | 2.703 | 0.8639 | True | True | True | True | Sex-specific 1e-2 VUMC, sex-differential UKB |
| 0.003432 | 0.003701 | 6.884 | 0.3468 | 2.657 | 0.8411 | True | True | True | True | Sex-specific 1e-2 VUMC, sex-differential UKB |
| 0.003362 | 0.003624 | 6.93 | 0.1586 | 2.379 | 0.8654 | True | True | True | True | Sex-specific 1e-2 VUMC, sex-differential UKB |
| 0.003271 | 0.00353 | 8.135 | 0.03045 | 1.483 | 1.011 | True | True | True | True | Sex-specific 1e-2 VUMC, sex-differential UKB |
| 0.003366 | 0.003629 | 6.381 | 0.08441 | 2.044 | 0.5698 | True | True | True | True | Sex-specific 1e-2 VUMC, sex-differential UKB |
| 0.003277 | 0.003537 | 8.274 | 0.02904 | 1.546 | 0.9511 | True | True | True | True | Sex-specific 1e-2 VUMC, sex-differential UKB |
| 0.003277 | 0.003536 | 8.006 | 0.02037 | 1.548 | 0.9902 | True | True | True | True | Sex-specific 1e-2 VUMC, sex-differential UKB |
| 0.003284 | 0.003533 | 6.761 | 0.8296 | 2.212 | 1.626 | True | True | True | True | Sex-specific 1e-2 VUMC, sex-differential UKB |
| 0.003288 | 0.003537 | 6.901 | 0.7933 | 2.194 | 1.629 | True | True | True | True | Sex-specific 1e-2 VUMC, sex-differential UKB |
| 0.00409 | 0.004379 | 6.3 | 1.563 | 46.78 | 26.47 | True | True | True | True | Sex-specific 1e-2 VUMC, sex-differential UKB |

| SE_UKB_female | SE_UKB_male | LOG10P_VUMC_female | LOG10P_VUMC_male | LOG10P_UKB_female | LOG10P_UKB_male | female_effect_direction_replication | male_effect_direction_replication | sex_effect_comparison_replication | pvalue_effect_comparison_replication | replication_type |
|---|---|---|---|---|---|---|---|---|---|---|
| 0.00409 | 0.004379 | 6.302 | 1.564 | 46.79 | 26.47 | True | True | True | True | Sex-specific 1e-2 VUMC, sex-differential UKB |
| 0.00408 | 0.004367 | 6.141 | 1.487 | 46.55 | 27.41 | True | True | True | True | Sex-specific 1e-2 VUMC, sex-differential UKB |
| 0.004081 | 0.004368 | 6.162 | 1.478 | 46.49 | 27.42 | True | True | True | True | Sex-specific 1e-2 VUMC, sex-differential UKB |
| 0.004077 | 0.004364 | 6.162 | 1.475 | 46.67 | 27.59 | True | True | True | True | Sex-specific 1e-2 VUMC, sex-differential UKB |
| SE_UKB_female | SE_UKB_male | LOG10P_VUMC_female | LOG10P_VUMC_male | LOG10P_UKB_female | LOG10P_UKB_male | female_effect_direction_replication | male_effect_direction_replication | sex_effect_comparison_replication | pvalue_effect_comparison_replication | replication_type |
| 0.004077 | 0.004363 | 6.156 | 1.432 | 46.69 | 27.54 | True | True | True | True | Sex-specific 1e-2 VUMC, sex-differential UKB |
| 0.004077 | 0.004363 | 6.162 | 1.475 | 46.68 | 27.59 | True | True | True | True | Sex-specific 1e-2 VUMC, sex-differential UKB |
| 0.004077 | 0.004363 | 6.162 | 1.475 | 46.69 | 27.59 | True | True | True | True | Sex-specific 1e-2 VUMC, sex-differential UKB |
| 0.004077 | 0.004363 | 6.055 | 1.417 | 46.81 | 27.47 | True | True | True | True | Sex-specific 1e-2 VUMC, sex-differential UKB |
| 0.004076 | 0.004363 | 6.162 | 1.456 | 46.69 | 27.59 | True | True | True | True | Sex-specific 1e-2 VUMC, sex-differential UKB |
| 0.004076 | 0.004363 | 6.162 | 1.455 | 46.69 | 27.59 | True | True | True | True | Sex-specific 1e-2 VUMC, sex-differential UKB |
| 0.004076 | 0.004363 | 6.162 | 1.455 | 46.71 | 27.58 | True | True | True | True | Sex-specific 1e-2 VUMC, sex-differential UKB |
| 0.004077 | 0.004363 | 6.164 | 1.467 | 46.69 | 27.61 | True | True | True | True | Sex-specific 1e-2 VUMC, sex-differential UKB |
| 0.004076 | 0.004362 | 6.236 | 1.454 | 46.72 | 27.59 | True | True | True | True | Sex-specific 1e-2 VUMC, sex-differential UKB |
| 0.004076 | 0.004362 | 6.222 | 1.441 | 46.75 | 27.59 | True | True | True | True | Sex-specific 1e-2 VUMC, sex-differential UKB |
| 0.004076 | 0.004362 | 6.137 | 1.443 | 46.52 | 27.7 | True | True | True | True | Sex-specific 1e-2 VUMC, sex-differential UKB |
| 0.004076 | 0.004363 | 6.127 | 1.436 | 46.83 | 27.62 | True | True | True | True | Sex-specific 1e-2 VUMC, sex-differential UKB |
| 0.004076 | 0.004363 | 6.131 | 1.448 | 46.81 | 27.59 | True | True | True | True | Sex-specific 1e-2 VUMC, sex-differential UKB |
| 0.004076 | 0.004363 | 6.145 | 1.432 | 46.87 | 27.67 | True | True | True | True | Sex-specific 1e-2 VUMC, sex-differential UKB |
| 0.004076 | 0.004363 | 6.145 | 1.44 | 46.92 | 27.7 | True | True | True | True | Sex-specific 1e-2 VUMC, sex-differential UKB |
| 0.004076 | 0.004362 | 6.15 | 1.438 | 46.95 | 27.67 | True | True | True | True | Sex-specific 1e-2 VUMC, sex-differential UKB |
| 0.004076 | 0.004362 | 6.148 | 1.438 | 46.88 | 27.59 | True | True | True | True | Sex-specific 1e-2 VUMC, sex-differential UKB |
| 0.004076 | 0.004363 | 6.119 | 1.439 | 46.93 | 27.44 | True | True | True | True | Sex-specific 1e-2 VUMC, sex-differential UKB |
| 0.004076 | 0.004363 | 6.127 | 1.44 | 47.01 | 27.48 | True | True | True | True | Sex-specific 1e-2 VUMC, sex-differential UKB |
| 0.004077 | 0.004364 | 6.144 | 1.532 | 47.01 | 27.51 | True | True | True | True | Sex-specific 1e-2 VUMC, sex-differential UKB |
| 0.004079 | 0.004366 | 6.175 | 1.524 | 47.28 | 27.58 | True | True | True | True | Sex-specific 1e-2 VUMC, sex-differential UKB |

| SE_UKB_female | SE_UKB_male | LOG10P_VUMC_female | LOG10P_VUMC_male | LOG10P_UKB_female | LOG10P_UKB_male | female_effect_direction_replication | male_effect_direction_replication | sex_effect_comparison_replication | pvalue_effect_comparison_replication | replication_type |
|---|---|---|---|---|---|---|---|---|---|---|
| 0.004087 | 0.004374 | 5.99 | 1.484 | 46.6 | 26.9 | True | True | True | True | Sex-specific 1e-2 VUMC, sex-differential UKB |
| 0.004081 | 0.004366 | 6.344 | 1.491 | 48.5 | 27.83 | True | True | True | True | Sex-specific 1e-2 VUMC, sex-differential UKB |
| | | 6.388 | 1.378 | | | False | False | False | False | |
| 0.004071 | 0.004358 | 6.446 | 1.481 | 46.8 | 26.38 | True | True | True | True | Sex-specific 1e-2 VUMC, sex-differential UKB |
| 0.004071 | 0.004357 | 6.443 | 1.479 | 46.67 | 26.42 | True | True | True | True | Sex-specific 1e-2 VUMC, sex-differential UKB |
| 0.004085 | 0.004372 | 6.316 | 1.606 | 47.91 | 27.96 | True | True | True | True | Sex-specific 1e-2 VUMC, sex-differential UKB |
| 0.004032 | 0.004315 | 7.244 | 1.502 | 49.08 | 31.2 | True | True | True | True | Sex-specific 1e-2 VUMC, sex-differential UKB |
| 0.004031 | 0.004315 | 7.236 | 1.522 | 49.1 | 31.31 | True | True | True | True | Sex-specific 1e-2 VUMC, sex-differential UKB |
| 0.004032 | 0.004315 | 7.371 | 1.467 | 49.31 | 31.38 | True | True | True | True | Sex-specific 1e-2 VUMC, sex-differential UKB |
| 0.00403 | 0.004314 | 7.26 | 1.497 | 49.07 | 31.31 | True | True | True | True | Sex-specific 1e-2 VUMC, sex-differential UKB |
| 0.00403 | 0.004313 | 7.349 | 1.497 | 49.11 | 31.35 | True | True | True | True | Sex-specific 1e-2 VUMC, sex-differential UKB |
| 0.004026 | 0.004308 | 7.218 | 1.475 | 49.68 | 31.52 | True | True | True | True | Sex-specific 1e-2 VUMC, sex-differential UKB |
| 0.004025 | 0.004307 | 7.07 | 1.529 | 49.29 | 31.35 | True | True | True | True | Sex-specific 1e-2 VUMC, sex-differential UKB |
| 0.004024 | 0.004305 | 7.015 | 1.511 | 49.49 | 31.25 | True | True | True | True | Sex-specific 1e-2 VUMC, sex-differential UKB |
| 0.004024 | 0.004306 | 7.085 | 1.48 | 49.81 | 31.36 | True | True | True | True | Sex-specific 1e-2 VUMC, sex-differential UKB |
| 0.004017 | 0.004298 | 6.937 | 1.528 | 49.55 | 31.35 | True | True | True | True | Sex-specific 1e-2 VUMC, sex-differential UKB |
| 0.004017 | 0.004298 | 6.984 | 1.523 | 49.65 | 31.33 | True | True | True | True | Sex-specific 1e-2 VUMC, sex-differential UKB |
| 0.004017 | 0.004298 | 6.934 | 1.561 | 49.56 | 31.35 | True | True | True | True | Sex-specific 1e-2 VUMC, sex-differential UKB |
| 0.004017 | 0.004298 | 6.932 | 1.564 | 49.57 | 31.34 | True | True | True | True | Sex-specific 1e-2 VUMC, sex-differential UKB |
| 0.004017 | 0.004298 | 6.99 | 1.574 | 49.79 | 31.04 | True | True | True | True | Sex-specific 1e-2 VUMC, sex-differential UKB |
| 0.004018 | 0.004298 | 7.034 | 1.529 | 49.89 | 31.46 | True | True | True | True | Sex-specific 1e-2 VUMC, sex-differential UKB |
| 0.004017 | 0.004298 | 6.934 | 1.561 | 49.57 | 31.34 | True | True | True | True | Sex-specific 1e-2 VUMC, sex-differential UKB |
| 0.004014 | 0.004294 | 6.99 | 1.544 | 48.86 | 31.04 | True | True | True | True | Sex-specific 1e-2 VUMC, sex-differential UKB |
| 0.003998 | 0.004274 | 6.991 | 1.544 | 48.52 | 31.06 | True | True | True | True | Sex-specific 1e-2 VUMC, sex-differential UKB |
| 0.003998 | 0.004274 | 6.992 | 1.544 | 48.52 | 31.06 | True | True | True | True | Sex-specific 1e-2 VUMC, sex-differential UKB |
| 0.003998 | 0.004274 | 6.991 | 1.544 | 48.52 | 31.06 | True | True | True | True | Sex-specific 1e-2 VUMC, sex-differential UKB |

| SE_UKB_female | SE_UKB_male | LOG10P_VUMC_female | LOG10P_VUMC_male | LOG10P_UKB_female | LOG10P_UKB_male | female_effect_direction_replication | male_effect_direction_replication | sex_effect_comparison_replication | pvalue_effect_comparison_replication | replication_type |
|---|---|---|---|---|---|---|---|---|---|---|
| 0.004 | 0.004278 | 6.905 | 1.562 | 49.21 | 31.36 | True | True | True | True | Sex-specific 1e-2 VUMC, sex-differential UKB |
| 0.003999 | 0.004277 | 6.864 | 1.514 | 49.15 | 30.93 | True | True | True | True | Sex-specific 1e-2 VUMC, sex-differential UKB |
| 0.003999 | 0.004276 | 6.884 | 1.585 | 49.02 | 31.32 | True | True | True | True | Sex-specific 1e-2 VUMC, sex-differential UKB |
| 0.003999 | 0.004276 | 6.883 | 1.585 | 49.02 | 31.31 | True | True | True | True | Sex-specific 1e-2 VUMC, sex-differential UKB |
| 0.003998 | 0.004276 | 6.884 | 1.585 | 49.02 | 31.32 | True | True | True | True | Sex-specific 1e-2 VUMC, sex-differential UKB |
| 0.003999 | 0.004276 | 6.884 | 1.584 | 49 | 31.31 | True | True | True | True | Sex-specific 1e-2 VUMC, sex-differential UKB |
| 0.003998 | 0.004276 | 6.884 | 1.585 | 49.01 | 31.3 | True | True | True | True | Sex-specific 1e-2 VUMC, sex-differential UKB |
| 0.003998 | 0.004276 | 6.884 | 1.585 | 49.01 | 31.3 | True | True | True | True | Sex-specific 1e-2 VUMC, sex-differential UKB |
| 0.003998 | 0.004276 | 6.884 | 1.585 | 49.01 | 31.3 | True | True | True | True | Sex-specific 1e-2 VUMC, sex-differential UKB |
| 0.003998 | 0.004276 | 6.884 | 1.585 | 49.01 | 31.3 | True | True | True | True | Sex-specific 1e-2 VUMC, sex-differential UKB |
| 0.003998 | 0.004276 | 6.885 | 1.585 | 49.01 | 31.3 | True | True | True | True | Sex-specific 1e-2 VUMC, sex-differential UKB |
| 0.003998 | 0.004276 | 6.884 | 1.585 | 49.01 | 31.3 | True | True | True | True | Sex-specific 1e-2 VUMC, sex-differential UKB |
| 0.003998 | 0.004276 | 6.885 | 1.585 | 49.01 | 31.3 | True | True | True | True | Sex-specific 1e-2 VUMC, sex-differential UKB |
| SE_UKB_female | SE_UKB_male | LOG10P_VUMC_female | LOG10P_VUMC_male | LOG10P_UKB_female | LOG10P_UKB_male | female_effect_direction_replication | male_effect_direction_replication | sex_effect_comparison_replication | pvalue_effect_comparison_replication | replication_type |
| 0.003998 | 0.004276 | 6.894 | 1.586 | 49.01 | 31.3 | True | True | True | True | Sex-specific 1e-2 VUMC, sex-differential UKB |
| 0.003998 | 0.004276 | 6.874 | 1.603 | 49.01 | 31.3 | True | True | True | True | Sex-specific 1e-2 VUMC, sex-differential UKB |
| | | 6.874 | 1.603 | | | False | False | False | False | |
| 0.003998 | 0.004276 | 6.874 | 1.603 | 49.01 | 31.3 | True | True | True | True | Sex-specific 1e-2 VUMC, sex-differential UKB |
| 0.003998 | 0.004276 | 6.873 | 1.603 | 49.01 | 31.29 | True | True | True | True | Sex-specific 1e-2 VUMC, sex-differential UKB |
| 0.003998 | 0.004276 | 6.873 | 1.603 | 49.01 | 31.3 | True | True | True | True | Sex-specific 1e-2 VUMC, sex-differential UKB |
| 0.003999 | 0.004276 | 6.964 | 1.569 | 49.34 | 31.42 | True | True | True | True | Sex-specific 1e-2 VUMC, sex-differential UKB |
| 0.003998 | 0.004276 | 6.873 | 1.603 | 49.01 | 31.3 | True | True | True | True | Sex-specific 1e-2 VUMC, sex-differential UKB |
| 0.003998 | 0.004276 | 6.873 | 1.603 | 49.01 | 31.3 | True | True | True | True | Sex-specific 1e-2 VUMC, sex-differential UKB |
| 0.003998 | 0.004276 | 6.875 | 1.604 | 48.99 | 31.28 | True | True | True | True | Sex-specific 1e-2 VUMC, sex-differential UKB |
| 0.003998 | 0.004276 | 6.891 | 1.613 | 49.01 | 31.3 | True | True | True | True | Sex-specific 1e-2 VUMC, sex-differential UKB |
| 0.003998 | 0.004276 | 6.871 | 1.6 | 49.01 | 31.3 | True | True | True | True | Sex-specific 1e-2 VUMC, sex-differential UKB |

| SE_UKB_female | SE_UKB_male | LOG10P_VUMC_female | LOG10P_VUMC_male | LOG10P_UKB_female | LOG10P_UKB_male | female_effect_direction_replication | male_effect_direction_replication | sex_effect_comparison_replication | pvalue_effect_comparison_replication | replication_type |
|---|---|---|---|---|---|---|---|---|---|---|
| 0.003999 | 0.004276 | 6.946 | 1.583 | 49.34 | 31.41 | True | True | True | True | Sex-specific 1e-2 VUMC, sex-differential UKB |
| 0.003998 | 0.004276 | 6.871 | 1.6 | 49.01 | 31.3 | True | True | True | True | Sex-specific 1e-2 VUMC, sex-differential UKB |
| | | 6.961 | 1.567 | | | False | False | False | False | |
| 0.003998 | 0.004276 | 6.871 | 1.6 | 49 | 31.32 | True | True | True | True | Sex-specific 1e-2 VUMC, sex-differential UKB |
| 0.003998 | 0.004276 | 6.795 | 1.583 | 49 | 31.33 | True | True | True | True | Sex-specific 1e-2 VUMC, sex-differential UKB |
| 0.003998 | 0.004276 | 6.795 | 1.583 | 49 | 31.32 | True | True | True | True | Sex-specific 1e-2 VUMC, sex-differential UKB |
| 0.003998 | 0.004276 | 6.795 | 1.583 | 49 | 31.33 | True | True | True | True | Sex-specific 1e-2 VUMC, sex-differential UKB |
| 0.003998 | 0.004276 | 6.795 | 1.583 | 49 | 31.33 | True | True | True | True | Sex-specific 1e-2 VUMC, sex-differential UKB |
| 0.003998 | 0.004276 | 6.795 | 1.583 | 49 | 31.33 | True | True | True | True | Sex-specific 1e-2 VUMC, sex-differential UKB |
| | | 6.161 | 1.53 | | | False | False | False | False | |
| 0.003998 | 0.004276 | 6.795 | 1.583 | 48.96 | 31.31 | True | True | True | True | Sex-specific 1e-2 VUMC, sex-differential UKB |
| 0.003998 | 0.004276 | 6.795 | 1.583 | 48.96 | 31.31 | True | True | True | True | Sex-specific 1e-2 VUMC, sex-differential UKB |
| 0.003996 | 0.004273 | 6.795 | 1.583 | 48.93 | 31.31 | True | True | True | True | Sex-specific 1e-2 VUMC, sex-differential UKB |
| 0.003995 | 0.004271 | 6.795 | 1.583 | 48.9 | 31.3 | True | True | True | True | Sex-specific 1e-2 VUMC, sex-differential UKB |
| | | 6.825 | 1.591 | | | False | False | False | False | |
| | | 6.823 | 1.591 | | | False | False | False | False | |
| | | 6.755 | 1.6 | | | False | False | False | False | |
| | | 6.755 | 1.596 | | | False | False | False | False | |
| | | 6.755 | 1.596 | | | False | False | False | False | |
| | | 6.756 | 1.598 | | | False | False | False | False | |
| | | 6.753 | 1.598 | | | False | False | False | False | |
| | | 6.798 | 1.597 | | | False | False | False | False | |
| 0.003643 | 0.003889 | 7.936 | 1.04 | 19.98 | 25.98 | True | True | False | False | |
| 0.003643 | 0.003889 | 7.953 | 1.035 | 20.49 | 26.16 | True | True | False | False | |
| 0.003642 | 0.003888 | 7.95 | 1.048 | 20.46 | 26.21 | True | True | False | False | |
| 0.003643 | 0.003888 | 7.981 | 1.024 | 20.5 | 26.18 | True | True | False | False | |
| 0.003643 | 0.003888 | 7.979 | 1.024 | 20.49 | 26.18 | True | True | False | False | |
| 0.003643 | 0.003889 | 7.901 | 1.015 | 20.5 | 26.16 | True | True | False | False | |
| 0.003635 | 0.003881 | 7.887 | 0.9817 | 19.17 | 25.64 | True | True | False | False | |
| 0.003644 | 0.00389 | 7.899 | 0.9647 | 20.25 | 26.24 | True | True | False | False | |
| 0.003644 | 0.00389 | 7.901 | 0.9649 | 20.19 | 26.25 | True | True | False | False | |

| SE_UKB_female | SE_UKB_male | LOG10P_VUMC_female | LOG10P_VUMC_male | LOG10P_UKB_female | LOG10P_UKB_male | female_effect_direction_replication | male_effect_direction_replication | sex_effect_comparison_replication | pvalue_effect_comparison_replication | replication_type |
|---|---|---|---|---|---|---|---|---|---|---|
| 0.003635 | 0.003881 | 7.887 | 0.9823 | 19.18 | 25.66 | True | True | False | False | |
| 0.003644 | 0.00389 | 7.968 | 1.004 | 19.77 | 26.43 | True | True | False | False | |
| 0.003644 | 0.00389 | 7.968 | 1.004 | 19.75 | 26.43 | True | True | False | False | |
| 0.003644 | 0.00389 | 7.744 | 1.049 | 19.96 | 26.77 | True | True | False | False | |
| 0.003645 | 0.003891 | 7.768 | 1.063 | 20.01 | 26.66 | True | True | False | False | |
| 0.003645 | 0.00389 | 7.768 | 1.064 | 20.02 | 26.67 | True | True | False | False | |
| 0.003641 | 0.003886 | 7.797 | 1.25 | 20.69 | 28.17 | True | True | False | False | |
| 0.003632 | 0.003876 | 8.414 | 1.365 | 21.56 | 29.51 | True | True | False | False | |
| 0.003631 | 0.003874 | 8.426 | 1.393 | 21.82 | 29.19 | True | True | False | False | |
| 0.003578 | 0.003815 | 8.099 | 1.416 | 23.43 | 30.4 | True | True | False | False | |
| 0.003657 | 0.0039 | 8.14 | 1.572 | 22.96 | 30.51 | True | True | False | False | |
| 0.003767 | 0.004033 | 9.236 | 1.37 | 30.42 | 34.02 | True | True | False | False | |
| 0.003702 | 0.003966 | 9.29 | 1.235 | 28.73 | 31.89 | True | True | False | False | |
| 0.003702 | 0.003965 | 9.336 | 1.296 | 28.67 | 31.87 | True | True | False | False | |
| 0.003701 | 0.003965 | 9.294 | 1.259 | 28.14 | 32.07 | True | True | False | False | |
| 0.004284 | 0.004592 | 6.773 | 1.424 | 14.18 | 18.91 | True | True | False | False | |
| 0.003701 | 0.003966 | 9.288 | 1.256 | 28.08 | 32.05 | True | True | False | False | |
| 0.0037 | 0.003964 | 9.318 | 1.294 | 28.06 | 31.83 | True | True | False | False | |
| 0.005077 | 0.005432 | 1.187 | 7.407 | 26.1 | 23.06 | True | True | True | False | |
| 0.005067 | 0.005423 | 1.188 | 7.424 | 26.15 | 22.76 | True | True | False | False | |
| 0.004863 | 0.005206 | 1.511 | 7.315 | 26.47 | 20.44 | True | True | False | False | |
| 0.005033 | 0.005381 | 1.326 | 7.738 | 27.91 | 22.48 | True | True | False | False | |
| 0.005108 | 0.005462 | 0.9118 | 7.589 | 29.06 | 23.2 | True | True | False | False | |
| 0.005032 | 0.005379 | 1.349 | 7.651 | 27.93 | 22.46 | True | True | False | False | |
| 0.005032 | 0.005379 | 1.349 | 7.651 | 27.93 | 22.46 | True | True | False | False | |
| 0.005032 | 0.005379 | 1.349 | 7.651 | 27.93 | 22.47 | True | True | False | False | |
| 0.00503 | 0.005376 | 1.378 | 7.689 | 27.86 | 22.45 | True | True | False | False | |

| SE_UKB_female | SE_UKB_male | LOG10P_VUMC_female | LOG10P_VUMC_male | LOG10P_UKB_female | LOG10P_UKB_male | female_effect_direction_replication | male_effect_direction_replication | sex_effect_comparison_replication | pvalue_effect_comparison_replication | replication_type |
|---|---|---|---|---|---|---|---|---|---|---|
| 0.005033 | 0.005381 | 1.381 | 7.827 | 27.85 | 22.81 | True | True | False | False | |
| 0.005035 | 0.005383 | 1.378 | 7.828 | 27.76 | 23.03 | True | True | False | False | |
| | | 1.414 | 7.764 | | | False | False | False | False | |
| 0.005178 | 0.005535 | 0.9659 | 7.628 | 29.18 | 23.84 | True | True | False | False | |
| 0.005033 | 0.005381 | 1.416 | 7.75 | 27.77 | 23.09 | True | True | False | False | |
| 0.005033 | 0.005381 | 1.429 | 7.648 | 27.76 | 23.08 | True | True | False | False | |
| 0.005094 | 0.005446 | 1.452 | 7.635 | 27.29 | 23.09 | True | True | False | False | |
| 0.005033 | 0.005381 | 1.432 | 7.626 | 27.78 | 23.12 | True | True | False | False | |
| 0.005033 | 0.005381 | 1.447 | 7.627 | 27.76 | 23.16 | True | True | False | False | |
| 0.005033 | 0.005381 | 1.442 | 7.609 | 27.77 | 23.16 | True | True | False | False | |
| 0.005032 | 0.00538 | 1.444 | 7.607 | 27.88 | 23.13 | True | True | False | False | |
| 0.005032 | 0.00538 | 1.444 | 7.607 | 27.87 | 23.13 | True | True | False | False | |
| 0.005253 | 0.005619 | 0.5758 | 7.713 | 28.18 | 24.12 | True | True | False | False | |
| 0.00503 | 0.005378 | 1.454 | 7.526 | 27.99 | 23.19 | True | True | False | False | |
| 0.005031 | 0.005379 | 1.459 | 7.529 | 28.03 | 22.98 | True | True | False | False | |
| 0.005031 | 0.00538 | 1.417 | 7.475 | 28.27 | 23.13 | True | True | False | False | |
| 0.005031 | 0.00538 | 1.411 | 7.391 | 28.28 | 23.13 | True | True | False | False | |
| 0.00503 | 0.005379 | 1.414 | 7.387 | 28.41 | 23.11 | True | True | False | False | |
| 0.00503 | 0.005379 | 1.42 | 7.471 | 28.43 | 23.11 | True | True | False | False | |
| 0.004235 | 0.004522 | 0.1779 | 4.907 | 17.74 | 12.07 | True | True | False | False | |
| 0.005153 | 0.005513 | 0.9414 | 7.366 | 29.54 | 23.66 | True | True | False | False | |
| 0.005028 | 0.005378 | 1.424 | 7.326 | 28.55 | 23.21 | True | True | False | False | |
| 0.005028 | 0.005378 | 1.427 | 7.321 | 28.57 | 23.21 | True | True | False | False | |
| 0.005028 | 0.005378 | 1.424 | 7.326 | 28.58 | 23.21 | True | True | False | False | |
| 0.005028 | 0.005378 | 1.43 | 7.409 | 28.58 | 23.2 | True | True | False | False | |
| 0.004231 | 0.004517 | 0.1415 | 4.48 | 17.92 | 11.54 | True | True | False | False | |
| 0.004917 | 0.005265 | 1.085 | 8.013 | 28.64 | 22.41 | True | True | False | False | |
| 0.005028 | 0.005377 | 1.462 | 7.37 | 28.66 | 23.35 | True | True | False | False | |

| SE_UKB_female | SE_UKB_male | LOG10P_VUMC_female | LOG10P_VUMC_male | LOG10P_UKB_female | LOG10P_UKB_male | female_effect_direction_replication | male_effect_direction_replication | sex_effect_comparison_replication | pvalue_effect_comparison_replication | replication_type |
|---|---|---|---|---|---|---|---|---|---|---|
| 0.005028 | 0.005377 | 1.461 | 7.372 | 28.65 | 23.35 | True | True | False | False | |
| 0.005028 | 0.005377 | 1.477 | 7.263 | 28.83 | 23.3 | True | True | False | False | |
| 0.004712 | 0.005044 | 0.7191 | 6.526 | 25.01 | 20.68 | True | True | False | False | |
| 0.004715 | 0.005047 | 0.6887 | 6.643 | 25.14 | 20.73 | True | True | False | False | |
| 0.005027 | 0.005376 | 1.494 | 7.264 | 28.92 | 23.23 | True | True | False | False | |
| 0.004715 | 0.005047 | 0.7273 | 6.707 | 25.11 | 20.72 | True | True | False | False | |
| 0.004711 | 0.005043 | 0.7103 | 6.524 | 25.37 | 20.9 | True | True | False | False | |
| 0.004937 | 0.005285 | 0.7267 | 6.692 | 24.68 | 20.5 | True | True | False | False | |
| 0.004714 | 0.005046 | 0.7177 | 6.684 | 25.22 | 20.82 | True | True | False | False | |
| 0.005027 | 0.005376 | 1.478 | 7.266 | 29.04 | 23.33 | True | True | False | False | |
| 0.004712 | 0.005044 | 0.7106 | 6.654 | 25.47 | 20.54 | True | True | False | False | |
| 0.004716 | 0.005047 | 0.6839 | 6.427 | 25.26 | 20.88 | True | True | False | False | |
| 0.004714 | 0.005045 | 0.6522 | 6.405 | 25.17 | 20.78 | True | True | False | False | |
| | | 1.489 | 7.288 | | | False | False | False | False | |
| 0.004716 | 0.005047 | 0.6757 | 6.57 | 25.29 | 20.87 | True | True | False | False | |
| 0.005026 | 0.005374 | 1.456 | 7.239 | 29.02 | 23.21 | True | True | False | False | |
| 0.005027 | 0.005376 | 1.445 | 7.239 | 28.99 | 23.43 | True | True | False | False | |
| 0.00471 | 0.00504 | 0.6504 | 6.245 | 25.52 | 20.67 | True | True | False | False | |
| 0.005027 | 0.005376 | 1.447 | 7.243 | 28.98 | 23.45 | True | True | False | False | |
| 0.004715 | 0.005046 | 0.6786 | 6.473 | 25.19 | 20.92 | True | True | False | False | |
| 0.004716 | 0.005047 | 0.6466 | 6.367 | 25.27 | 21.03 | True | True | False | False | |
| 0.004715 | 0.005045 | 0.6172 | 6.55 | 25.47 | 21.17 | True | True | False | False | |
| SE_UKB_female | SE_UKB_male | LOG10P_VUMC_female | LOG10P_VUMC_male | LOG10P_UKB_female | LOG10P_UKB_male | female_effect_direction_replication | male_effect_direction_replication | sex_effect_comparison_replication | pvalue_effect_comparison_replication | replication_type |
| 0.004996 | 0.005346 | 0.6083 | 6.301 | 24.76 | 21.08 | True | True | False | False | |
| 0.004286 | 0.004584 | 0.1585 | 5.818 | 22.21 | 17.85 | True | True | False | False | |
| 0.005028 | 0.005377 | 1.463 | 7.335 | 29.07 | 23.83 | True | True | False | False | |
| 0.004714 | 0.005046 | 0.6302 | 6.373 | 25.47 | 21.23 | True | True | False | False | |

| SE_UKB_female | SE_UKB_male | LOG10P_VUMC_female | LOG10P_VUMC_male | LOG10P_UKB_female | LOG10P_UKB_male | female_effect_direction_replication | male_effect_direction_replication | sex_effect_comparison_replication | pvalue_effect_comparison_replication | replication_type |
|---|---|---|---|---|---|---|---|---|---|---|
| 0.004716 | 0.005048 | 0.6544 | 6.532 | 25.39 | 21.2 | True | True | False | False | |
| 0.005028 | 0.005377 | 1.462 | 7.339 | 28.98 | 23.72 | True | True | False | False | |
| 0.004717 | 0.005049 | 0.663 | 6.467 | 25.28 | 21.23 | True | True | False | False | |
| 0.004715 | 0.005046 | 0.6122 | 6.367 | 25.37 | 21.22 | True | True | False | False | |
| 0.005029 | 0.005377 | 1.434 | 7.291 | 28.99 | 23.98 | True | True | False | False | |
| 0.004999 | 0.005347 | 1.433 | 7.309 | 28.59 | 23.84 | True | True | False | False | |
| 0.004717 | 0.005049 | 0.6358 | 6.528 | 25.3 | 21.24 | True | True | False | False | |
| 0.004633 | 0.004958 | 0.434 | 6.032 | 23.03 | 19.86 | True | True | False | False | |
| 0.004717 | 0.005049 | 0.6351 | 6.534 | 25.3 | 21.24 | True | True | False | False | |
| 0.004717 | 0.005049 | 0.6351 | 6.534 | 25.3 | 21.24 | True | True | False | False | |
| 0.005028 | 0.005378 | 1.432 | 7.331 | 28.93 | 23.86 | True | True | False | False | |
| 0.004716 | 0.005047 | 0.6114 | 6.372 | 25.33 | 21.21 | True | True | False | False | |
| 0.004237 | 0.004531 | 0.04756 | 5.751 | 20.4 | 16.71 | True | True | False | False | |
| 0.005103 | 0.005461 | 0.9145 | 7.218 | 29.7 | 24.23 | True | True | False | False | |
| 0.004778 | 0.005115 | 0.3055 | 6.393 | 26.07 | 21.61 | True | True | False | False | |
| 0.004777 | 0.005115 | 0.3581 | 6.454 | 26.16 | 21.58 | True | True | False | False | |
| 0.005083 | 0.005436 | 1.006 | 7.179 | 30.67 | 24.47 | True | True | False | False | |
| 0.005072 | 0.005426 | 1.031 | 7.314 | 27.47 | 22.82 | True | True | False | False | |
| 0.005058 | 0.005412 | 1.03 | 7.312 | 29.59 | 24.59 | True | True | False | False | |
| 0.00507 | 0.005421 | 0.9995 | 7.109 | 30.46 | 24.86 | True | True | False | False | |
| 0.00507 | 0.005422 | 0.9785 | 7.015 | 30.69 | 25.04 | True | True | False | False | |
| 0.005071 | 0.005422 | 0.9791 | 6.996 | 30.72 | 24.96 | True | True | False | False | |
| 0.005071 | 0.005422 | 0.9797 | 6.994 | 30.69 | 24.86 | True | True | False | False | |
| 0.005071 | 0.005422 | 0.9816 | 7.004 | 30.75 | 24.85 | True | True | False | False | |
| 0.005071 | 0.005422 | 0.9868 | 6.992 | 30.73 | 24.86 | True | True | False | False | |
| 0.005071 | 0.005422 | 0.9865 | 6.991 | 30.72 | 24.88 | True | True | False | False | |

| SE_UKB_female | SE_UKB_male | LOG10P_VUMC_female | LOG10P_VUMC_male | LOG10P_UKB_female | LOG10P_UKB_male | female_effect_direction_replication | male_effect_direction_replication | sex_effect_comparison_replication | pvalue_effect_comparison_replication | replication_type |
|---|---|---|---|---|---|---|---|---|---|---|
| 0.004573 | 0.004896 | 0.6244 | 6.292 | 20.22 | 16.54 | True | True | False | False | |
| 0.00515 | 0.005504 | 1.008 | 7.022 | 30.65 | 24.6 | True | True | False | False | |
| 0.005062 | 0.005415 | 1.023 | 7.193 | 30.06 | 24.78 | True | True | False | False | |
| 0.004575 | 0.004898 | 0.6307 | 6.442 | 20.3 | 15.38 | True | True | False | False | |
| 0.004561 | 0.004885 | 0.6307 | 6.442 | 20.54 | 16.52 | True | True | False | False | |
| 0.004552 | 0.004873 | 0.6107 | 6.595 | 21.19 | 16.79 | True | True | False | False | |
| 0.004551 | 0.004872 | 0.6405 | 6.679 | 21.12 | 16.69 | True | True | False | False | |
| 0.00507 | 0.005422 | 1.034 | 6.939 | 31 | 24.86 | True | True | False | False | |
| 0.004561 | 0.004885 | 0.5848 | 6.252 | 21.01 | 16.82 | True | True | False | False | |
| 0.005067 | 0.005418 | 0.9982 | 6.633 | 32.1 | 25.51 | True | True | False | False | |
| 0.003815 | 0.004073 | 0.02676 | 4.134 | 12.87 | 11.37 | True | True | False | False | |
| 0.004516 | 0.004825 | 0.3481 | 6.446 | 23.42 | 17.97 | True | True | False | False | |
| 0.004516 | 0.004825 | 0.3455 | 6.476 | 23.5 | 17.93 | True | True | False | False | |
| 0.005067 | 0.005418 | 0.9845 | 6.6 | 31.95 | 25.37 | True | True | False | False | |
| 0.005067 | 0.005418 | 0.9838 | 6.599 | 32.16 | 25.48 | True | True | False | False | |
| 0.003946 | 0.004217 | 0.1993 | 4.775 | 13.95 | 13.13 | True | True | True | False | |
| 0.005067 | 0.005417 | 0.9836 | 6.599 | 32.18 | 25.55 | True | True | False | False | |
| 0.005067 | 0.005417 | 0.9836 | 6.599 | 32.18 | 25.56 | True | True | False | False | |
| 0.004515 | 0.004822 | 0.3511 | 6.583 | 23.05 | 18.43 | True | True | False | False | |
| 0.005067 | 0.005417 | 0.9836 | 6.599 | 32.21 | 25.58 | True | True | False | False | |
| 0.005067 | 0.005417 | 0.9836 | 6.599 | 32.2 | 25.59 | True | True | False | False | |
| 0.005067 | 0.005417 | 0.9836 | 6.599 | 32.21 | 25.58 | True | True | False | False | |
| 0.004461 | 0.004764 | 0.7247 | 6.731 | 22.83 | 17.71 | True | True | False | False | |
| 0.005067 | 0.005417 | 0.9835 | 6.596 | 32.16 | 25.58 | True | True | False | False | |
| | | 0.9835 | 6.596 | | | False | False | False | False | |
| 0.005067 | 0.005417 | 0.9836 | 6.595 | 32.16 | 25.6 | True | True | False | False | |

| SE_UKB_female | SE_UKB_male | LOG10P_VUMC_female | LOG10P_VUMC_male | LOG10P_UKB_female | LOG10P_UKB_male | female_effect_direction_replication | male_effect_direction_replication | sex_effect_comparison_replication | pvalue_effect_comparison_replication | replication_type |
|---|---|---|---|---|---|---|---|---|---|---|
| 0.004461 | 0.004764 | 0.7251 | 6.732 | 22.71 | 17.72 | True | True | False | False | |
| 0.00446 | 0.004764 | 0.7225 | 6.738 | 22.89 | 17.6 | True | True | False | False | |
| 0.00446 | 0.004764 | 0.729 | 6.725 | 22.8 | 17.68 | True | True | False | False | |
| 0.004455 | 0.004758 | 0.755 | 6.74 | 22.03 | 17.74 | True | True | False | False | |
| SE_UKB_female | SE_UKB_male | LOG10P_VUMC_female | LOG10P_VUMC_male | LOG10P_UKB_female | LOG10P_UKB_male | female_effect_direction_replication | male_effect_direction_replication | sex_effect_comparison_replication | pvalue_effect_comparison_replication | replication_type |
| 0.003799 | 0.004056 | 0.07563 | 4.054 | 12.92 | 10.67 | True | True | False | False | |
| 0.0038 | 0.004057 | 0.07269 | 4.073 | 12.82 | 10.67 | True | True | False | False | |
| 0.0038 | 0.004057 | 0.07272 | 4.085 | 12.82 | 10.8 | True | True | False | False | |
| 0.003801 | 0.004057 | 0.07239 | 4.087 | 12.8 | 10.8 | True | True | False | False | |
| 0.003801 | 0.004058 | 0.07283 | 4.086 | 12.77 | 10.85 | True | True | False | False | |
| 0.003801 | 0.004057 | 0.07392 | 4.085 | 12.79 | 10.81 | True | True | False | False | |
| 0.003801 | 0.004057 | 0.07392 | 4.085 | 12.79 | 10.81 | True | True | False | False | |
| 0.0038 | 0.004057 | 0.07392 | 4.085 | 12.72 | 10.75 | True | True | False | False | |
| 0.005067 | 0.005417 | 0.9848 | 6.599 | 32.12 | 25.63 | True | True | False | False | |
| 0.003801 | 0.004058 | 0.07399 | 4.087 | 12.86 | 10.83 | True | True | False | False | |
| 0.003798 | 0.004055 | 0.07075 | 4.086 | 12.23 | 10.23 | True | True | False | False | |
| 0.003801 | 0.004058 | 0.0741 | 4.084 | 12.85 | 10.79 | True | True | False | False | |
| 0.003801 | 0.004057 | 0.0741 | 4.084 | 12.85 | 10.78 | True | True | False | False | |
| 0.005067 | 0.005417 | 0.9835 | 6.599 | 32.14 | 25.64 | True | True | False | False | |
| 0.003801 | 0.004057 | 0.07248 | 4.084 | 12.86 | 10.77 | True | True | False | False | |
| 0.0038 | 0.004057 | 0.07408 | 4.084 | 12.85 | 10.77 | True | True | False | False | |
| 0.00507 | 0.005422 | 0.9839 | 6.598 | 32.54 | 24.74 | True | True | False | False | |
| 0.0038 | 0.004057 | 0.07415 | 4.083 | 12.85 | 10.78 | True | True | False | False | |
| 0.0038 | 0.004057 | 0.07415 | 4.083 | 12.87 | 10.78 | True | True | False | False | |
| 0.003801 | 0.004057 | 0.07415 | 4.083 | 12.86 | 10.8 | True | True | False | False | |
| 0.004461 | 0.004765 | 0.7547 | 6.834 | 22.66 | 17.85 | True | True | False | False | |

| SE_UKB_female | SE_UKB_male | LOG10P_VUMC_female | LOG10P_VUMC_male | LOG10P_UKB_female | LOG10P_UKB_male | female_effect_direction_replication | male_effect_direction_replication | sex_effect_comparison_replication | pvalue_effect_comparison_replication | replication_type |
|---|---|---|---|---|---|---|---|---|---|---|
| 0.0038 | 0.004057 | 0.0773 | 4.059 | 12.93 | 10.81 | True | True | False | False | |
| 0.003801 | 0.004057 | 0.0734 | 4.082 | 12.85 | 10.78 | True | True | False | False | |
| 0.003801 | 0.004058 | 0.06327 | 4.099 | 13.04 | 10.79 | True | True | False | False | |
| 0.003802 | 0.00406 | 0.05604 | 4.167 | 13.1 | 10.85 | True | True | False | False | |
| 0.005067 | 0.005417 | 0.9845 | 6.601 | 32.07 | 25.78 | True | True | False | False | |
| 0.005118 | 0.005471 | 0.9884 | 6.621 | 31.95 | 25.63 | True | True | False | False | |
| 0.003801 | 0.004058 | 0.07028 | 4.03 | 12.81 | 10.78 | True | True | False | False | |
| 0.003801 | 0.004058 | 0.07437 | 4.082 | 12.87 | 10.81 | True | True | False | False | |
| 0.005067 | 0.005417 | 0.9846 | 6.602 | 32.08 | 25.78 | True | True | False | False | |
| 0.0038 | 0.004057 | 0.07752 | 4.058 | 12.96 | 10.83 | True | True | False | False | |
| 0.005067 | 0.005417 | 0.9847 | 6.602 | 32.08 | 25.78 | True | True | False | False | |
| 0.003801 | 0.004058 | 0.07456 | 4.081 | 12.89 | 10.83 | True | True | False | False | |
| 0.003801 | 0.004058 | 0.07456 | 4.081 | 12.9 | 10.84 | True | True | False | False | |
| 0.0038 | 0.004058 | 0.07467 | 4.082 | 12.9 | 10.92 | True | True | False | False | |
| 0.003801 | 0.004058 | 0.07451 | 4.079 | 12.9 | 10.85 | True | True | False | False | |
| 0.003801 | 0.004058 | 0.07451 | 4.079 | 12.88 | 10.84 | True | True | False | False | |
| 0.003801 | 0.004058 | 0.07451 | 4.079 | 12.9 | 10.85 | True | True | False | False | |
| 0.003801 | 0.004058 | 0.0744 | 4.079 | 12.9 | 10.85 | True | True | False | False | |
| 0.005067 | 0.005417 | 0.9854 | 6.605 | 32.08 | 25.8 | True | True | False | False | |
| 0.003801 | 0.004058 | 0.07447 | 4.077 | 12.91 | 10.84 | True | True | False | False | |
| 0.003801 | 0.004058 | 0.07444 | 4.079 | 12.92 | 10.84 | True | True | False | False | |
| 0.003801 | 0.004058 | 0.07444 | 4.079 | 12.93 | 10.84 | True | True | False | False | |
| 0.003801 | 0.004058 | 0.07444 | 4.079 | 12.99 | 10.75 | True | True | False | False | |
| 0.003801 | 0.004057 | 0.07444 | 4.079 | 12.95 | 10.73 | True | True | False | False | |
| 0.003801 | 0.004058 | 0.0745 | 4.078 | 12.92 | 10.82 | True | True | False | False | |
| 0.003801 | 0.004058 | 0.07434 | 4.079 | 12.93 | 10.83 | True | True | False | False | |
| | | 0.9883 | 6.609 | | | False | False | False | False | |

| SE_UKB_female | SE_UKB_male | LOG10P_VUMC_female | LOG10P_VUMC_male | LOG10P_UKB_female | LOG10P_UKB_male | female_effect_direction_replication | male_effect_direction_replication | sex_effect_comparison_replication | pvalue_effect_comparison_replication | replication_type |
|---|---|---|---|---|---|---|---|---|---|---|
| 0.00446 | 0.004763 | 0.7758 | 6.856 | 22.61 | 17.8 | True | True | False | False | |
| 0.003801 | 0.004058 | 0.07474 | 4.078 | 12.9 | 10.83 | True | True | False | False | |
| 0.003801 | 0.004058 | 0.07332 | 4.084 | 12.9 | 10.84 | True | True | False | False | |
| 0.004461 | 0.004765 | 0.755 | 6.838 | 22.6 | 17.91 | True | True | False | False | |
| 0.003801 | 0.004058 | 0.07341 | 4.083 | 12.9 | 10.85 | True | True | False | False | |
| 0.005067 | 0.005418 | 0.9869 | 6.609 | 32.07 | 25.86 | True | True | False | False | |
| 0.0038 | 0.004057 | 0.0705 | 4.136 | 12.96 | 10.69 | True | True | False | False | |
| 0.005074 | 0.005425 | 0.976 | 6.698 | 32.47 | 26.06 | True | True | False | False | |
| 0.003782 | 0.004039 | 0.1602 | 4.488 | 13.25 | 10.69 | True | True | False | False | |
| 0.003782 | 0.004039 | 0.163 | 4.54 | 13.23 | 10.69 | True | True | False | False | |
| 0.003845 | 0.00411 | 0.1078 | 4.876 | 12.87 | 11.15 | True | True | False | False | |
| 0.00378 | 0.004036 | 0.1301 | 4.722 | 13.16 | 10.6 | True | True | False | False | |
| 0.003779 | 0.004036 | 0.1306 | 4.722 | 13.15 | 10.59 | True | True | False | False | |
| 0.003779 | 0.004035 | 0.13 | 4.722 | 13.26 | 10.54 | True | True | False | False | |
| 0.003779 | 0.004035 | 0.13 | 4.722 | 13.14 | 10.59 | True | True | False | False | |
| 0.003781 | 0.004036 | 0.13 | 4.722 | 12.92 | 10.64 | True | True | False | False | |
| 0.004516 | 0.004826 | 0.3648 | 6.866 | 23.39 | 18.3 | True | True | False | False | |
| 0.005066 | 0.005416 | 0.9722 | 6.757 | 32.05 | 25.85 | True | True | False | False | |
| 0.004434 | 0.004739 | 1.207 | 8.409 | 22.47 | 16.78 | True | True | False | False | |
| 0.003779 | 0.004035 | 0.1268 | 4.722 | 13.11 | 10.61 | True | True | False | False | |
| 0.003779 | 0.004035 | 0.1305 | 4.723 | 13.11 | 10.6 | True | True | False | False | |
| 0.003779 | 0.004035 | 0.1297 | 4.722 | 13.05 | 10.63 | True | True | False | False | |
| 0.003783 | 0.004039 | 0.1264 | 4.761 | 13.13 | 10.66 | True | True | False | False | |
| 0.003781 | 0.004039 | 0.1198 | 5.346 | 13.41 | 10.51 | True | True | False | False | |
| 0.005084 | 0.005435 | 0.962 | 6.768 | 32.08 | 26.01 | True | True | False | False | |

| SE_UKB_female | SE_UKB_male | LOG10P_VUMC_female | LOG10P_VUMC_male | LOG10P_UKB_female | LOG10P_UKB_male | female_effect_direction_replication | male_effect_direction_replication | sex_effect_comparison_replication | pvalue_effect_comparison_replication | replication_type |
|---|---|---|---|---|---|---|---|---|---|---|
| 0.005074 | 0.005424 | 1.035 | 6.301 | 32.25 | 26.76 | True | True | False | False | |
| 0.004528 | 0.004837 | 0.2834 | 6.503 | 23.39 | 18.01 | True | True | False | False | |
| 0.005062 | 0.005412 | 1.031 | 6.328 | 32.26 | 26.79 | True | True | False | False | |
| 0.005062 | 0.005412 | 1.031 | 6.328 | 32.29 | 26.79 | True | True | False | False | |
| 0.004113 | 0.00439 | 0.1589 | 4.307 | 18.57 | 12.7 | True | True | False | False | |
| 0.004108 | 0.004385 | 0.1615 | 4.442 | 18.17 | 12.6 | True | True | False | False | |
| 0.004109 | 0.004387 | 0.1624 | 4.444 | 18.14 | 12.59 | True | True | False | False | |
| 0.004109 | 0.004386 | 0.1814 | 4.558 | 18.27 | 12.57 | True | True | False | False | |
| 0.005061 | 0.005412 | 1.074 | 6.344 | 32.45 | 26.71 | True | True | False | False | |
| 0.005062 | 0.005412 | 1.075 | 6.342 | 32.47 | 26.7 | True | True | False | False | |
| 0.004502 | 0.004812 | 0.331 | 6.564 | 23.19 | 17.59 | True | True | False | False | |
| 0.00411 | 0.004385 | 0.1606 | 4.454 | 18.48 | 12.37 | True | True | False | False | |
| 0.004109 | 0.004387 | 0.1606 | 4.454 | 18.15 | 12.58 | True | True | False | False | |
| 0.004109 | 0.004386 | 0.1596 | 4.453 | 18.19 | 12.63 | True | True | False | False | |
| 0.004112 | 0.004388 | 0.1685 | 4.428 | 18.47 | 12.44 | True | True | False | False | |
| 0.004743 | 0.005065 | 1.21 | 6.539 | 29.98 | 24.19 | True | True | False | False | |
| 0.004405 | 0.004707 | 0.7699 | 6.611 | 22.74 | 16.72 | True | True | False | False | |
| 0.004013 | 0.004282 | 0.2852 | 4.695 | 16.31 | 11.19 | True | True | False | False | |
| 0.004405 | 0.004706 | 0.77 | 6.612 | 22.74 | 16.71 | True | True | False | False | |
| 0.004393 | 0.004695 | 0.716 | 6.961 | 22.57 | 16.37 | True | True | False | False | |
| 0.004407 | 0.004711 | 0.7958 | 7.55 | 22.09 | 16.57 | True | True | False | False | |
| 0.004391 | 0.004692 | 0.7049 | 7.038 | 22.73 | 16.44 | True | True | False | False | |
| 0.004392 | 0.004693 | 0.7009 | 7.034 | 22.79 | 16.31 | True | True | False | False | |
| 0.004391 | 0.004692 | 0.7023 | 7.012 | 22.86 | 16.26 | True | True | False | False | |
| 0.004386 | 0.004687 | 0.7143 | 7.026 | 22.63 | 16.22 | True | True | False | False | |
| 0.00439 | 0.004691 | 0.7146 | 7.027 | 22.59 | 16.41 | True | True | False | False | |

| SE_UKB_female | SE_UKB_male | LOG10P_VUMC_female | LOG10P_VUMC_male | LOG10P_UKB_female | LOG10P_UKB_male | female_effect_direction_replication | male_effect_direction_replication | sex_effect_comparison_replication | pvalue_effect_comparison_replication | replication_type |
|---|---|---|---|---|---|---|---|---|---|---|
| 0.005034 | 0.005382 | 0.9858 | 5.882 | 33.31 | 26.53 | True | True | False | False | |
| 0.004389 | 0.004691 | 0.717 | 7.026 | 22.58 | 16.45 | True | True | False | False | |
| 0.004391 | 0.004692 | 0.7144 | 7.027 | 22.71 | 16.42 | True | True | False | False | |
| 0.004373 | 0.004673 | 0.6986 | 6.99 | 23.52 | 16.56 | True | True | False | False | |
| 0.005034 | 0.005383 | 0.9875 | 5.927 | 33.33 | 26.69 | True | True | False | False | |
| 0.005036 | 0.005385 | 0.9955 | 5.943 | 33.25 | 26.66 | True | True | False | False | |
| 0.004391 | 0.004694 | 0.7365 | 7.052 | 22.97 | 16.71 | True | True | False | False | |
| 0.005032 | 0.005381 | 0.9705 | 6.01 | 32.97 | 26.83 | True | True | False | False | |
| 0.005034 | 0.005383 | 0.9487 | 6.012 | 33.42 | 26.75 | True | True | False | False | |
| 0.004394 | 0.0047 | 0.6164 | 6.379 | 19.95 | 15.02 | True | True | False | False | |
| 0.005057 | 0.005408 | 1.06 | 6.737 | 32.81 | 26.9 | True | True | False | False | |
| 0.005042 | 0.005393 | 1.011 | 6.713 | 32.63 | 26.31 | True | True | False | False | |
| 0.005056 | 0.005408 | 1.043 | 6.746 | 32.82 | 26.77 | True | True | False | False | |
| 0.005056 | 0.005408 | 1.043 | 6.746 | 32.78 | 26.81 | True | True | False | False | |
| 0.005031 | 0.005381 | 0.9091 | 6.071 | 33.4 | 26.63 | True | True | False | False | |
| 0.005056 | 0.005408 | 1.044 | 6.746 | 32.78 | 26.81 | True | True | False | False | |
| 0.005031 | 0.005381 | 0.9091 | 6.071 | 33.4 | 26.63 | True | True | False | False | |
| 0.005031 | 0.005381 | 0.9087 | 6.073 | 33.38 | 26.68 | True | True | False | False | |
| 0.004383 | 0.004689 | 0.6275 | 6.124 | 19.93 | 14.8 | True | True | False | False | |
| 0.005056 | 0.005408 | 1.048 | 6.746 | 32.72 | 26.88 | True | True | False | False | |
| 0.005056 | 0.005408 | 1.048 | 6.746 | 32.72 | 26.89 | True | True | False | False | |
| SE_UKB_female | SE_UKB_male | LOG10P_VUMC_female | LOG10P_VUMC_male | LOG10P_UKB_female | LOG10P_UKB_male | female_effect_direction_replication | male_effect_direction_replication | sex_effect_comparison_replication | pvalue_effect_comparison_replication | replication_type |
| 0.004384 | 0.004691 | 0.6281 | 6.169 | 20.17 | 14.82 | True | True | False | False | |
| 0.004384 | 0.004691 | 0.6294 | 6.172 | 20.22 | 14.87 | True | True | False | False | |
| 0.004385 | 0.004691 | 0.6289 | 6.169 | 20.24 | 14.89 | True | True | False | False | |
| 0.004379 | 0.004684 | 0.6435 | 6.041 | 20.59 | 14.75 | True | True | False | False | |

| SE_UKB_female | SE_UKB_male | LOG10P_VUMC_female | LOG10P_VUMC_male | LOG10P_UKB_female | LOG10P_UKB_male | female_effect_direction_replication | male_effect_direction_replication | sex_effect_comparison_replication | pvalue_effect_comparison_replication | replication_type |
|---|---|---|---|---|---|---|---|---|---|---|
| 0.004864 | 0.005206 | 0.7075 | 5.896 | 28.06 | 23.32 | True | True | False | False | |
| 0.005028 | 0.005376 | 0.9966 | 6.236 | 33.36 | 27.05 | True | True | False | False | |
| 0.00505 | 0.005399 | 1.127 | 6.607 | 32.78 | 27.33 | True | True | False | False | |
| 0.00505 | 0.005399 | 1.127 | 6.607 | 32.81 | 27.33 | True | True | False | False | |
| 0.005072 | 0.005422 | 1.134 | 6.499 | 32.79 | 27.81 | True | True | False | False | |
| 0.005073 | 0.005421 | 1.124 | 6.521 | 32.84 | 27.9 | True | True | False | False | |
| 0.005064 | 0.005411 | 1.071 | 6.695 | 32.75 | 29.02 | True | True | True | False | |
| 0.005063 | 0.00541 | 1.071 | 6.695 | 32.74 | 29.18 | True | True | True | False | |
| 0.005063 | 0.005409 | 1.042 | 6.793 | 32.51 | 29.3 | True | True | True | False | |
| 0.005058 | 0.005401 | 1.007 | 6.569 | 32.19 | 29.29 | True | True | True | False | |
| 0.005061 | 0.005406 | 1.061 | 6.785 | 32.42 | 29.37 | True | True | True | False | |
| 0.005058 | 0.005401 | 1.009 | 6.578 | 32.17 | 29.3 | True | True | True | False | |
| 0.005058 | 0.005401 | 1.009 | 6.578 | 32.15 | 29.31 | True | True | True | False | |
| 0.005061 | 0.005406 | 1.064 | 6.781 | 32.21 | 29.47 | True | True | True | False | |
| 0.005061 | 0.005406 | 1.064 | 6.781 | 32.21 | 29.47 | True | True | True | False | |
| 0.005057 | 0.0054 | 1.011 | 6.574 | 31.97 | 29.38 | True | True | True | False | |
| 0.005058 | 0.005402 | 1.055 | 6.771 | 32.03 | 29.76 | True | True | True | False | |
| 0.003268 | 0.003493 | 1.047 | 7.123 | 28.31 | 21.17 | True | True | False | False | |
| 0.003256 | 0.003476 | 1.611 | 5.554 | 25.69 | 19.49 | True | True | False | False | |
| 0.003256 | 0.003476 | 1.628 | 5.551 | 25.71 | 19.57 | True | True | False | False | |
| 0.003349 | 0.003585 | 1.004 | 5.789 | 14.24 | 11.24 | True | True | False | False | |
| 0.003264 | 0.003489 | 1.189 | 7.525 | 28.51 | 20.67 | True | True | False | False | |
| 0.003354 | 0.003591 | 1.08 | 5.373 | 13.84 | 11.51 | True | True | False | False | |
| 0.003277 | 0.003507 | 1.207 | 7.321 | 26.75 | 17.26 | True | True | False | False | |
| 0.003386 | 0.003633 | 1.933 | 5.772 | 16.64 | 13.2 | True | True | False | False | |
| 0.003287 | 0.003516 | 1.581 | 8.447 | 32.76 | 21.85 | True | True | False | False | |
| 0.003621 | 0.00391 | 0.5832 | 4.997 | 6.518 | 2.213 | True | True | False | False | |

| SE_UKB_female | SE_UKB_male | LOG10P_VUMC_female | LOG10P_VUMC_male | LOG10P_UKB_female | LOG10P_UKB_male | female_effect_direction_replication | male_effect_direction_replication | sex_effect_comparison_replication | pvalue_effect_comparison_replication | replication_type |
|---|---|---|---|---|---|---|---|---|---|---|
| 0.003621 | 0.003911 | 0.5878 | 5.014 | 6.594 | 2.24 | True | True | False | False | |
| 0.003619 | 0.00391 | 0.5884 | 5.024 | 6.615 | 2.23 | True | True | False | False | |
| 0.003619 | 0.003909 | 0.5885 | 5.023 | 6.608 | 2.239 | True | True | False | False | |
| 0.003555 | 0.00384 | 0.6404 | 4.895 | 6.001 | 2.653 | True | True | False | False | |
| 0.003629 | 0.003919 | 0.5949 | 5.161 | 6.526 | 2.312 | True | True | False | False | |
| 0.003307 | 0.003564 | 0.9047 | 7.342 | 4.751 | 1.294 | True | True | False | False | |
| 0.003546 | 0.00383 | 0.6325 | 4.862 | 6.124 | 2.616 | True | True | False | False | |
| 0.003619 | 0.003909 | 0.5881 | 5.122 | 6.674 | 2.281 | True | True | False | False | |
| 0.003619 | 0.003909 | 0.5896 | 5.137 | 6.666 | 2.215 | True | True | False | False | |
| 0.003619 | 0.003909 | 0.5848 | 5.122 | 6.674 | 2.28 | True | True | False | False | |
| 0.003619 | 0.003909 | 0.5872 | 5.123 | 6.709 | 2.273 | True | True | False | False | |
| 0.003546 | 0.00383 | 0.633 | 4.859 | 6.165 | 2.613 | True | True | False | False | |
| 0.003546 | 0.00383 | 0.6333 | 4.859 | 6.165 | 2.61 | True | True | False | False | |
| 0.003619 | 0.003909 | 0.5874 | 5.135 | 6.716 | 2.298 | True | True | False | False | |
| 0.003619 | 0.003909 | 0.5869 | 5.124 | 6.709 | 2.274 | True | True | False | False | |
| 0.003619 | 0.003909 | 0.5941 | 5.126 | 6.688 | 2.275 | True | True | False | False | |
| 0.003549 | 0.003832 | 0.6409 | 4.869 | 6.115 | 2.595 | True | True | False | False | |
| 0.003572 | 0.003859 | 0.5484 | 5.075 | 6.791 | 2.763 | True | True | False | False | |
| 0.003572 | 0.003859 | 0.5485 | 5.075 | 6.793 | 2.763 | True | True | False | False | |
| 0.003411 | 0.003687 | 1.127 | 6.432 | 6.788 | 1.954 | True | True | False | False | |
| 0.003411 | 0.003687 | 1.123 | 6.364 | 6.792 | 1.956 | True | True | False | False | |
| 0.003572 | 0.003859 | 0.5422 | 5.061 | 6.795 | 2.761 | True | True | False | False | |
| 0.003411 | 0.003687 | 1.128 | 6.358 | 6.798 | 1.952 | True | True | False | False | |
| 0.003411 | 0.003687 | 1.121 | 6.356 | 6.796 | 1.952 | True | True | False | False | |
| 0.003411 | 0.003687 | 1.124 | 6.367 | 6.776 | 1.943 | True | True | False | False | |
| 0.003572 | 0.003858 | 0.4666 | 5.214 | 6.773 | 2.748 | True | True | False | False | |
| 0.003571 | 0.003858 | 0.4631 | 5.207 | 6.776 | 2.747 | True | True | False | False | |
| 0.003568 | 0.003855 | 0.4459 | 5.147 | 6.699 | 2.693 | True | True | False | False | |

| SE_UKB_female | SE_UKB_male | LOG10P_VUMC_female | LOG10P_VUMC_male | LOG10P_UKB_female | LOG10P_UKB_male | female_effect_direction_replication | male_effect_direction_replication | sex_effect_comparison_replication | pvalue_effect_comparison_replication | replication_type |
|---|---|---|---|---|---|---|---|---|---|---|
| 0.003564 | 0.00385 | 0.4025 | 5.398 | 6.86 | 2.723 | True | True | False | False | |
| SE_UKB_female | SE_UKB_male | LOG10P_VUMC_female | LOG10P_VUMC_male | LOG10P_UKB_female | LOG10P_UKB_male | female_effect_direction_replication | male_effect_direction_replication | sex_effect_comparison_replication | pvalue_effect_comparison_replication | replication_type |
| 0.003564 | 0.003851 | 0.441 | 5.373 | 6.897 | 2.715 | True | True | False | False | |
| 0.003564 | 0.00385 | 0.4566 | 5.346 | 6.917 | 2.728 | True | True | False | False | |
| 0.003565 | 0.003852 | 0.4599 | 5.235 | 6.881 | 2.799 | True | True | False | False | |
| 0.003564 | 0.00385 | 0.4566 | 5.394 | 6.912 | 2.727 | True | True | False | False | |
| 0.003565 | 0.003852 | 0.4592 | 5.234 | 6.887 | 2.791 | True | True | False | False | |
| 0.003564 | 0.003851 | 0.4568 | 5.403 | 6.92 | 2.721 | True | True | False | False | |
| 0.003566 | 0.003853 | 0.4528 | 5.287 | 6.879 | 2.814 | True | True | False | False | |
| 0.003575 | 0.003862 | 0.5009 | 5.311 | 6.828 | 2.76 | True | True | False | False | |
| 0.003576 | 0.003863 | 0.498 | 5.29 | 6.858 | 2.748 | True | True | False | False | |
| 0.003565 | 0.003852 | 0.4563 | 5.24 | 6.913 | 2.803 | True | True | False | False | |
| 0.003564 | 0.003851 | 0.4559 | 5.402 | 6.926 | 2.721 | True | True | False | False | |
| 0.003567 | 0.003853 | 0.4546 | 5.311 | 6.947 | 2.85 | True | True | False | False | |
| 0.003566 | 0.003853 | 0.4616 | 5.315 | 6.923 | 2.807 | True | True | False | False | |
| 0.003567 | 0.003854 | 0.462 | 5.315 | 6.967 | 2.862 | True | True | False | False | |
| 0.003567 | 0.003854 | 0.5345 | 5.689 | 7.003 | 2.861 | True | True | False | False | |
| 0.003566 | 0.003853 | 0.5396 | 5.625 | 6.911 | 2.788 | True | True | False | False | |
| 0.003575 | 0.003862 | 0.54 | 5.577 | 6.983 | 2.854 | True | True | False | False | |
| 0.003579 | 0.003868 | 0.5486 | 5.572 | 6.991 | 2.932 | True | True | False | False | |
| 0.003574 | 0.003863 | 0.5051 | 5.997 | 7.353 | 2.838 | True | True | False | False | |
| 0.003589 | 0.003879 | 0.6127 | 5.395 | 7.522 | 3.304 | True | True | False | False | |
| 0.003271 | 0.003532 | 0.5486 | 4.6 | 0.06677 | 0.1211 | True | True | True | True | Sex-specific 1e-2 VUMC, sex-differential UKB |
| 0.003277 | 0.003538 | 0.495 | 4.665 | 0.08295 | 0.1172 | True | True | True | True | Sex-specific 1e-2 VUMC, sex-differential UKB |
| 0.003279 | 0.00354 | 0.4228 | 4.731 | 0.081 | 0.1289 | True | True | True | True | Sex-specific 1e-2 VUMC, sex-differential UKB |
| 0.003285 | 0.003545 | 0.7685 | 5.32 | 0.2244 | 0.09823 | False | True | False | False | |
| 0.003351 | 0.00362 | 0.4849 | 4.492 | 0.3099 | 0.1777 | False | True | False | False | |
| 0.003351 | 0.003619 | 0.4961 | 4.493 | 0.2911 | 0.1716 | False | True | False | False | |

| SE_UKB_female | SE_UKB_male | LOG10P_VUMC_female | LOG10P_VUMC_male | LOG10P_UKB_female | LOG10P_UKB_male | female_effect_direction_replication | male_effect_direction_replication | sex_effect_comparison_replication | pvalue_effect_comparison_replication | replication_type |
|---|---|---|---|---|---|---|---|---|---|---|
| 0.003293 | 0.003562 | 0.1413 | 5.857 | 0.5046 | 0.09576 | False | True | False | False | |
| 0.003267 | 0.003527 | 0.5891 | 6.325 | 0.3989 | 0.007928 | False | True | False | False | |
| 0.003293 | 0.00356 | 0.1357 | 6.182 | 0.4945 | 0.1142 | False | True | False | False | |
| 0.00327 | 0.003533 | 0.5256 | 7.717 | 0.2561 | 0.002238 | False | True | False | False | |
| 0.004073 | 0.004384 | 6.736 | 0.4751 | 33.9 | 24.14 | True | True | True | True | Sex-specific 1e-2 VUMC, sex-differential UKB |
| 0.004875 | 0.005241 | 6.793 | 1.041 | 37.17 | 25.05 | True | True | True | True | Sex-specific 1e-2 VUMC, sex-differential UKB |
| 0.004902 | 0.005274 | 7.777 | 1.092 | 40.3 | 28.43 | True | True | True | True | Sex-specific 1e-2 VUMC, sex-differential UKB |
| 0.00486 | 0.005235 | 7.928 | 1.088 | 47.57 | 29.41 | True | True | True | True | Sex-specific 1e-2 VUMC, sex-differential UKB |
| 0.005207 | 0.005607 | 7.156 | 1.038 | 54.68 | 25.45 | True | True | True | True | Sex-specific 1e-2 VUMC, sex-differential UKB |
| 0.005205 | 0.005603 | 7.162 | 0.973 | 54.79 | 25.55 | True | True | True | True | Sex-specific 1e-2 VUMC, sex-differential UKB |
| 0.005211 | 0.005611 | 7.222 | 1.034 | 55.31 | 25.74 | True | True | True | True | Sex-specific 1e-2 VUMC, sex-differential UKB |
| 0.00521 | 0.005608 | 7.449 | 1.032 | 54.37 | 25.86 | True | True | True | True | Sex-specific 1e-2 VUMC, sex-differential UKB |
| 0.00517 | 0.005562 | 7.181 | 1.411 | 54.09 | 26.58 | True | True | True | True | Sex-specific 1e-2 VUMC, sex-differential UKB |
| 0.00521 | 0.005608 | 7.449 | 1.032 | 54.36 | 25.86 | True | True | True | True | Sex-specific 1e-2 VUMC, sex-differential UKB |
| 0.005208 | 0.005605 | 7.39 | 1.044 | 53.88 | 25.89 | True | True | True | True | Sex-specific 1e-2 VUMC, sex-differential UKB |
| 0.00521 | 0.005606 | 7.418 | 1.035 | 54.17 | 26.01 | True | True | True | True | Sex-specific 1e-2 VUMC, sex-differential UKB |
| 0.005169 | 0.00556 | 7.09 | 1.419 | 53.87 | 26.75 | True | True | True | True | Sex-specific 1e-2 VUMC, sex-differential UKB |
| 0.005165 | 0.005559 | 6.643 | 1.339 | 54.8 | 26.4 | True | True | True | True | Sex-specific 1e-2 VUMC, sex-differential UKB |
| 0.0052 | 0.005601 | 6.656 | 0.9864 | 54.41 | 25.63 | True | True | True | True | Sex-specific 1e-2 VUMC, sex-differential UKB |
| 0.005208 | 0.00561 | 6.886 | 1.048 | 53.91 | 26.19 | True | True | True | True | Sex-specific 1e-2 VUMC, sex-differential UKB |
| 0.005234 | 0.005634 | 6.694 | 1.319 | 52.86 | 26.16 | True | True | True | True | Sex-specific 1e-2 VUMC, sex-differential UKB |
| 0.005233 | 0.005634 | 6.735 | 1.317 | 52.85 | 26.16 | True | True | True | True | Sex-specific 1e-2 VUMC, sex-differential UKB |
| 0.005205 | 0.005607 | 7.049 | 1.235 | 53.99 | 25.73 | True | True | True | True | Sex-specific 1e-2 VUMC, sex-differential UKB |
| 0.005205 | 0.005606 | 6.987 | 1.188 | 53.47 | 26.1 | True | True | True | True | Sex-specific 1e-2 VUMC, sex-differential UKB |
| 0.005204 | 0.005605 | 7.047 | 1.239 | 53.73 | 26.02 | True | True | True | True | Sex-specific 1e-2 VUMC, sex-differential UKB |
| 0.005194 | 0.005596 | 7.138 | 1.258 | 53.84 | 25.99 | True | True | True | True | Sex-specific 1e-2 VUMC, sex-differential UKB |

| SE_UKB_female | SE_UKB_male | LOG10P_VUMC_female | LOG10P_VUMC_male | LOG10P_UKB_female | LOG10P_UKB_male | female_effect_direction_replication | male_effect_direction_replication | sex_effect_comparison_replication | pvalue_effect_comparison_replication | replication_type |
|---|---|---|---|---|---|---|---|---|---|---|
| 0.005259 | 0.005664 | 6.824 | 1.241 | 53.12 | 26.08 | True | True | True | True | Sex-specific 1e-2 VUMC, sex-differential UKB |
| 0.005263 | 0.005668 | 7.147 | 1.313 | 53.27 | 25.92 | True | True | True | True | Sex-specific 1e-2 VUMC, sex-differential UKB |
| 0.005263 | 0.005668 | 7.246 | 1.355 | 53.1 | 25.75 | True | True | True | True | Sex-specific 1e-2 VUMC, sex-differential UKB |
| 0.005258 | 0.005662 | 7.039 | 1.216 | 53.04 | 25.67 | True | True | True | True | Sex-specific 1e-2 VUMC, sex-differential UKB |
| 0.006646 | 0.007156 | 10.05 | 0.7218 | 44.18 | 22.62 | True | True | True | True | Sex-specific 1e-2 VUMC, sex-differential UKB |
| 0.003556 | 0.003834 | 7.297 | 1.821 | 18.17 | 7.886 | True | True | True | True | Sex-specific 1e-2 VUMC, sex-differential UKB |
| 0.003543 | 0.003822 | 7.543 | 1.436 | 18.46 | 7.849 | True | True | True | True | Sex-specific 1e-2 VUMC, sex-differential UKB |
| 0.003543 | 0.003822 | 7.543 | 1.439 | 18.45 | 7.843 | True | True | True | True | Sex-specific 1e-2 VUMC, sex-differential UKB |
| SE_UKB_female | SE_UKB_male | LOG10P_VUMC_female | LOG10P_VUMC_male | LOG10P_UKB_female | LOG10P_UKB_male | female_effect_direction_replication | male_effect_direction_replication | sex_effect_comparison_replication | pvalue_effect_comparison_replication | replication_type |
| 0.003543 | 0.003822 | 7.559 | 1.443 | 18.44 | 7.839 | True | True | True | True | Sex-specific 1e-2 VUMC, sex-differential UKB |
| 0.003543 | 0.003822 | 7.555 | 1.443 | 18.45 | 7.846 | True | True | True | True | Sex-specific 1e-2 VUMC, sex-differential UKB |
| 0.003544 | 0.003823 | 7.557 | 1.45 | 18.45 | 7.833 | True | True | True | True | Sex-specific 1e-2 VUMC, sex-differential UKB |
| 0.003556 | 0.003834 | 7.294 | 1.817 | 18.15 | 7.857 | True | True | True | True | Sex-specific 1e-2 VUMC, sex-differential UKB |
| 0.003543 | 0.003822 | 7.554 | 1.422 | 18.4 | 7.796 | True | True | True | True | Sex-specific 1e-2 VUMC, sex-differential UKB |
| 0.003556 | 0.003835 | 7.21 | 1.907 | 17.96 | 7.849 | True | True | True | True | Sex-specific 1e-2 VUMC, sex-differential UKB |
| 0.003543 | 0.003823 | 7.54 | 1.435 | 18.47 | 7.837 | True | True | True | True | Sex-specific 1e-2 VUMC, sex-differential UKB |
| 0.003543 | 0.003823 | 7.597 | 1.426 | 18.49 | 7.825 | True | True | True | True | Sex-specific 1e-2 VUMC, sex-differential UKB |
| 0.003543 | 0.003822 | 7.582 | 1.429 | 18.48 | 7.78 | True | True | True | True | Sex-specific 1e-2 VUMC, sex-differential UKB |
| 0.003543 | 0.003822 | 7.542 | 1.437 | 18.42 | 7.788 | True | True | True | True | Sex-specific 1e-2 VUMC, sex-differential UKB |
| 0.003555 | 0.003833 | 7.234 | 1.777 | 18.2 | 7.776 | True | True | True | True | Sex-specific 1e-2 VUMC, sex-differential UKB |
| 0.003556 | 0.003834 | 7.218 | 1.908 | 18.02 | 7.726 | True | True | True | True | Sex-specific 1e-2 VUMC, sex-differential UKB |
| 0.003556 | 0.003834 | 7.12 | 1.915 | 18 | 7.738 | True | True | True | True | Sex-specific 1e-2 VUMC, sex-differential UKB |
| 0.003543 | 0.003822 | 7.446 | 1.449 | 18.54 | 7.705 | True | True | True | True | Sex-specific 1e-2 VUMC, sex-differential UKB |
| 0.003543 | 0.003822 | 7.446 | 1.449 | 18.54 | 7.705 | True | True | True | True | Sex-specific 1e-2 VUMC, sex-differential UKB |
| 0.003556 | 0.003833 | 7.192 | 1.836 | 18.3 | 7.748 | True | True | True | True | Sex-specific 1e-2 VUMC, sex-differential UKB |
| 0.003543 | 0.003822 | 7.446 | 1.46 | 18.51 | 7.716 | True | True | True | True | Sex-specific 1e-2 VUMC, sex-differential UKB |

| SE_UKB_female | SE_UKB_male | LOG10P_VUMC_female | LOG10P_VUMC_male | LOG10P_UKB_female | LOG10P_UKB_male | female_effect_direction_replication | male_effect_direction_replication | sex_effect_comparison_replication | pvalue_effect_comparison_replication | replication_type |
|---|---|---|---|---|---|---|---|---|---|---|
| 0.003555 | 0.003833 | 7.199 | 1.838 | 18.23 | 7.762 | True | True | True | True | Sex-specific 1e-2 VUMC, sex-differential UKB |
| 0.003543 | 0.003822 | 7.445 | 1.46 | 18.5 | 7.746 | True | True | True | True | Sex-specific 1e-2 VUMC, sex-differential UKB |
| 0.003548 | 0.003827 | 7.371 | 1.48 | 18.67 | 7.772 | True | True | True | True | Sex-specific 1e-2 VUMC, sex-differential UKB |
| 0.003518 | 0.003796 | 6.782 | 1.313 | 17.63 | 7.309 | True | True | True | True | Sex-specific 1e-2 VUMC, sex-differential UKB |
| 0.003562 | 0.003841 | 7.069 | 1.938 | 18.31 | 7.774 | True | True | True | True | Sex-specific 1e-2 VUMC, sex-differential UKB |
| 0.003518 | 0.003795 | 6.811 | 1.323 | 17.82 | 7.137 | True | True | True | True | Sex-specific 1e-2 VUMC, sex-specific 5e-8 UKB |
| 0.003549 | 0.003828 | 7.392 | 1.468 | 18.87 | 7.755 | True | True | True | True | Sex-specific 1e-2 VUMC, sex-differential UKB |
| 0.003562 | 0.003841 | 6.973 | 1.92 | 18.3 | 7.762 | True | True | True | True | Sex-specific 1e-2 VUMC, sex-differential UKB |
| 0.003549 | 0.003828 | 7.377 | 1.459 | 18.83 | 7.696 | True | True | True | True | Sex-specific 1e-2 VUMC, sex-differential UKB |
| 0.003549 | 0.003828 | 7.343 | 1.478 | 18.82 | 7.673 | True | True | True | True | Sex-specific 1e-2 VUMC, sex-differential UKB |
| 0.00358 | 0.003862 | 7.564 | 0.6659 | 14.28 | 5.972 | True | True | True | True | Sex-specific 1e-2 VUMC, sex-specific 5e-8 UKB |
| 0.003595 | 0.003876 | 7.684 | 0.8279 | 14.6 | 5.738 | True | True | True | True | Sex-specific 1e-2 VUMC, sex-specific 5e-8 UKB |
| 0.003533 | 0.00381 | 4.902 | 0.8836 | 14.01 | 10.03 | True | True | True | True | Sex-specific 1e-2 VUMC, sex-differential UKB |
| 0.003297 | 0.003553 | 7.56 | 0.5045 | 16.81 | 6.352 | True | True | True | True | Sex-specific 1e-2 VUMC, sex-specific 5e-8 UKB |
| 0.003297 | 0.003553 | 7.591 | 0.5299 | 16.94 | 6.316 | True | True | True | True | Sex-specific 1e-2 VUMC, sex-specific 5e-8 UKB |
| 0.004827 | 0.005154 | 0.4557 | 6.184 | 0.1086 | 1.855 | True | True | True | True | Sex-specific 1e-2 VUMC, sex-differential UKB |
| 0.004585 | 0.004902 | 0.5111 | 6.728 | 0.2882 | 1.823 | True | True | True | True | Sex-specific 1e-2 VUMC, sex-differential UKB |
| 0.004583 | 0.004899 | 0.546 | 6.711 | 0.2992 | 1.872 | True | True | True | True | Sex-specific 1e-2 VUMC, sex-differential UKB |
| 0.004632 | 0.004951 | 0.5161 | 6.848 | 0.4076 | 1.648 | True | True | True | True | Sex-specific 1e-2 VUMC, sex-differential UKB |
| 0.004581 | 0.004897 | 0.5248 | 6.816 | 0.294 | 1.921 | True | True | True | True | Sex-specific 1e-2 VUMC, sex-differential UKB |
| 0.004581 | 0.004897 | 0.5314 | 6.836 | 0.2885 | 1.922 | True | True | True | True | Sex-specific 1e-2 VUMC, sex-differential UKB |
| 0.004579 | 0.004896 | 0.5332 | 6.837 | 0.2882 | 1.929 | True | True | True | True | Sex-specific 1e-2 VUMC, sex-differential UKB |
| 0.004579 | 0.004896 | 0.5261 | 6.817 | 0.2837 | 1.907 | True | True | True | True | Sex-specific 1e-2 VUMC, sex-differential UKB |
| 0.004586 | 0.004907 | 0.607 | 7.505 | 0.3427 | 2.342 | True | True | True | True | Sex-specific 1e-2 VUMC, sex-differential UKB |
| 0.004583 | 0.004903 | 0.522 | 7.53 | 0.4202 | 2.698 | True | True | True | True | Sex-specific 1e-2 VUMC, sex-differential UKB |
| 0.004582 | 0.004903 | 0.5703 | 7.414 | 0.4279 | 2.688 | True | True | True | True | Sex-specific 1e-2 VUMC, sex-differential UKB |

| SE_UKB_female | SE_UKB_male | LOG10P_VUMC_female | LOG10P_VUMC_male | LOG10P_UKB_female | LOG10P_UKB_male | female_effect_direction_replication | male_effect_direction_replication | sex_effect_comparison_replication | pvalue_effect_comparison_replication | replication_type |
|---|---|---|---|---|---|---|---|---|---|---|
| 0.004584 | 0.004906 | 0.5682 | 7.421 | 0.4334 | 2.697 | True | True | True | True | Sex-specific 1e-2 VUMC, sex-differential UKB |
| 0.004587 | 0.004908 | 0.5276 | 7.557 | 0.447 | 2.724 | True | True | True | True | Sex-specific 1e-2 VUMC, sex-differential UKB |
| 0.004588 | 0.004909 | 0.5999 | 7.62 | 0.4255 | 2.768 | True | True | True | True | Sex-specific 1e-2 VUMC, sex-differential UKB |
| 0.004588 | 0.004909 | 0.5979 | 7.616 | 0.4259 | 2.76 | True | True | True | True | Sex-specific 1e-2 VUMC, sex-differential UKB |
| 0.004589 | 0.00491 | 0.5854 | 7.754 | 0.4415 | 2.765 | True | True | True | True | Sex-specific 1e-2 VUMC, sex-differential UKB |
| 0.004663 | 0.004986 | 0.5472 | 8.278 | 0.2036 | 2.966 | True | True | True | True | Sex-specific 1e-2 VUMC, sex-differential UKB |
| 0.004617 | 0.004939 | 0.5638 | 7.806 | 0.4004 | 2.708 | True | True | True | True | Sex-specific 1e-2 VUMC, sex-differential UKB |
| 0.004618 | 0.00494 | 0.578 | 7.772 | 0.4179 | 2.793 | True | True | True | True | Sex-specific 1e-2 VUMC, sex-differential UKB |
| 0.00462 | 0.004942 | 0.5478 | 7.872 | 0.4134 | 2.876 | True | True | True | True | Sex-specific 1e-2 VUMC, sex-differential UKB |
| 0.004639 | 0.004962 | 0.3765 | 7.611 | 0.3722 | 2.789 | True | True | True | True | Sex-specific 1e-2 VUMC, sex-differential UKB |
| 0.004637 | 0.00496 | 0.3075 | 7.572 | 0.3129 | 2.775 | True | True | True | True | Sex-specific 1e-2 VUMC, sex-differential UKB |
| 0.004636 | 0.004959 | 0.2747 | 7.555 | 0.2981 | 2.734 | True | True | True | True | Sex-specific 1e-2 VUMC, sex-differential UKB |
| 0.004638 | 0.00496 | 0.2667 | 7.62 | 0.324 | 2.73 | True | True | True | True | Sex-specific 1e-2 VUMC, sex-differential UKB |
| 0.004636 | 0.004958 | 0.2715 | 7.616 | 0.3178 | 2.722 | True | True | True | True | Sex-specific 1e-2 VUMC, sex-differential UKB |
| 0.004637 | 0.004959 | 0.2464 | 7.601 | 0.3162 | 2.863 | True | True | True | True | Sex-specific 1e-2 VUMC, sex-differential UKB |
| 0.004524 | 0.00484 | 0.7243 | 6.378 | 0.3584 | 2.199 | True | True | True | True | Sex-specific 1e-2 VUMC, sex-differential UKB |
| 0.003848 | 0.004151 | 8.623 | 0.9808 | 1.672 | 0.4081 | True | False | True | True | Sex-specific 1e-2 VUMC, sex-differential UKB |
| SE_UKB_female | SE_UKB_male | LOG10P_VUMC_female | LOG10P_VUMC_male | LOG10P_UKB_female | LOG10P_UKB_male | female_effect_direction_replication | male_effect_direction_replication | sex_effect_comparison_replication | pvalue_effect_comparison_replication | replication_type |
| 0.003848 | 0.00415 | 7.876 | 0.8433 | 1.747 | 0.3611 | True | False | True | True | Sex-specific 1e-2 VUMC, sex-differential UKB |
| 0.003846 | 0.004148 | 8.565 | 0.8158 | 1.794 | 0.3947 | True | False | True | True | Sex-specific 1e-2 VUMC, sex-differential UKB |
| 0.003821 | 0.004122 | 8.303 | 0.6951 | 2.347 | 0.5384 | True | False | True | True | Sex-specific 1e-2 VUMC, sex-differential UKB |
| 0.003817 | 0.004118 | 8.943 | 0.6548 | 2.313 | 0.5733 | True | False | True | True | Sex-specific 1e-2 VUMC, sex-differential UKB |
| 0.003298 | 0.00355 | 9.097 | 1.204 | 1.716 | 0.2373 | True | False | True | True | Sex-specific 1e-2 VUMC, sex-differential UKB |
| 0.003296 | 0.003547 | 8.92 | 1.163 | 1.837 | 0.2513 | True | False | True | True | Sex-specific 1e-2 VUMC, sex-differential UKB |
| 0.003296 | 0.003547 | 8.93 | 1.205 | 1.844 | 0.2454 | True | False | True | True | Sex-specific 1e-2 VUMC, sex-differential UKB |
| 0.003296 | 0.003547 | 8.836 | 1.082 | 1.862 | 0.2519 | True | False | True | True | Sex-specific 1e-2 VUMC, sex-differential UKB |

| SE_UKB_female | SE_UKB_male | LOG10P_VUMC_female | LOG10P_VUMC_male | LOG10P_UKB_female | LOG10P_UKB_male | female_effect_direction_replication | male_effect_direction_replication | sex_effect_comparison_replication | pvalue_effect_comparison_replication | replication_type |
|---|---|---|---|---|---|---|---|---|---|---|
| 0.003295 | 0.003546 | 8.804 | 1.099 | 1.815 | 0.2656 | True | False | True | True | Sex-specific 1e-2 VUMC, sex-differential UKB |
| 0.003294 | 0.003545 | 8.853 | 1.097 | 1.935 | 0.2261 | True | False | True | True | Sex-specific 1e-2 VUMC, sex-differential UKB |
| 0.003301 | 0.003551 | 8.929 | 1.187 | 1.959 | 0.2125 | True | False | True | True | Sex-specific 1e-2 VUMC, sex-differential UKB |
| 0.00331 | 0.003562 | 9.253 | 1.037 | 1.928 | 0.1776 | True | False | True | True | Sex-specific 1e-2 VUMC, sex-differential UKB |
| 0.003299 | 0.003549 | 9.3 | 0.9793 | 1.894 | 0.1717 | True | False | True | True | Sex-specific 1e-2 VUMC, sex-differential UKB |
| 0.003297 | 0.003547 | 9.29 | 1.038 | 2.009 | 0.1711 | True | False | True | True | Sex-specific 1e-2 VUMC, sex-differential UKB |
| 0.003301 | 0.003551 | 9.121 | 1.048 | 2.157 | 0.1534 | True | False | True | True | Sex-specific 1e-2 VUMC, sex-differential UKB |
| 0.003302 | 0.003554 | 8.725 | 1.052 | 2.312 | 0.1161 | True | False | True | True | Sex-specific 1e-2 VUMC, sex-differential UKB |
| 0.003299 | 0.003551 | 8.937 | 1.008 | 2.183 | 0.1244 | True | False | True | True | Sex-specific 1e-2 VUMC, sex-differential UKB |
| 0.003299 | 0.003552 | 8.752 | 1.065 | 2.204 | 0.118 | True | False | True | True | Sex-specific 1e-2 VUMC, sex-differential UKB |
| 0.003299 | 0.003551 | 8.87 | 1.07 | 2.192 | 0.1187 | True | False | True | True | Sex-specific 1e-2 VUMC, sex-differential UKB |
| 0.003298 | 0.003551 | 8.872 | 1.07 | 2.176 | 0.1174 | True | False | True | True | Sex-specific 1e-2 VUMC, sex-differential UKB |
| 0.00327 | 0.003519 | 8.086 | 0.759 | 1.826 | 0.0894 | True | False | True | True | Sex-specific 1e-2 VUMC, sex-differential UKB |
| 0.003268 | 0.003517 | 7.97 | 0.7671 | 1.839 | 0.07947 | True | False | True | True | Sex-specific 1e-2 VUMC, sex-differential UKB |
| 0.003268 | 0.003517 | 7.945 | 0.7756 | 1.846 | 0.08914 | True | False | True | True | Sex-specific 1e-2 VUMC, sex-differential UKB |
| 0.003267 | 0.003515 | 7.746 | 0.8053 | 1.873 | 0.08593 | True | False | True | True | Sex-specific 1e-2 VUMC, sex-differential UKB |
| 0.003266 | 0.003515 | 7.746 | 0.8057 | 1.873 | 0.08588 | True | False | True | True | Sex-specific 1e-2 VUMC, sex-differential UKB |
| 0.003266 | 0.003515 | 7.746 | 0.8054 | 1.857 | 0.08181 | True | False | True | True | Sex-specific 1e-2 VUMC, sex-differential UKB |
| 0.003265 | 0.003514 | 7.804 | 0.8161 | 1.872 | 0.05616 | True | False | True | True | Sex-specific 1e-2 VUMC, sex-differential UKB |
| 0.003265 | 0.003514 | 7.767 | 0.7819 | 1.862 | 0.0595 | True | False | True | True | Sex-specific 1e-2 VUMC, sex-differential UKB |
| 0.003265 | 0.003513 | 7.767 | 0.7819 | 1.871 | 0.06045 | True | False | True | True | Sex-specific 1e-2 VUMC, sex-differential UKB |
| 0.003267 | 0.003517 | 7.694 | 0.7576 | 1.91 | 0.1018 | True | False | True | True | Sex-specific 1e-2 VUMC, sex-differential UKB |
| 0.003269 | 0.003518 | 7.674 | 0.7111 | 1.852 | 0.09347 | True | False | True | True | Sex-specific 1e-2 VUMC, sex-differential UKB |
| 0.003269 | 0.003518 | 7.833 | 0.7213 | 1.847 | 0.09419 | True | False | True | True | Sex-specific 1e-2 VUMC, sex-differential UKB |
| 0.003269 | 0.003518 | 7.833 | 0.7414 | 1.838 | 0.09852 | True | False | True | True | Sex-specific 1e-2 VUMC, sex-differential UKB |
| 0.003269 | 0.003518 | 7.804 | 0.7359 | 1.837 | 0.09558 | True | False | True | True | Sex-specific 1e-2 VUMC, sex-differential UKB |

| SE_UKB_female | SE_UKB_male | LOG10P_VUMC_female | LOG10P_VUMC_male | LOG10P_UKB_female | LOG10P_UKB_male | female_effect_direction_replication | male_effect_direction_replication | sex_effect_comparison_replication | pvalue_effect_comparison_replication | replication_type |
|---|---|---|---|---|---|---|---|---|---|---|
| 0.003269 | 0.003518 | 7.804 | 0.7359 | 1.835 | 0.096 | True | False | True | True | Sex-specific 1e-2 VUMC, sex-differential UKB |
| 0.003269 | 0.003518 | 7.804 | 0.7359 | 1.83 | 0.09633 | True | False | True | True | Sex-specific 1e-2 VUMC, sex-differential UKB |
| 0.003269 | 0.003518 | 7.803 | 0.7359 | 1.83 | 0.09658 | True | False | True | True | Sex-specific 1e-2 VUMC, sex-differential UKB |
| 0.003269 | 0.003518 | 7.803 | 0.7359 | 1.83 | 0.09658 | True | False | True | True | Sex-specific 1e-2 VUMC, sex-differential UKB |
| 0.003269 | 0.003518 | 7.806 | 0.7376 | 1.831 | 0.09693 | True | False | True | True | Sex-specific 1e-2 VUMC, sex-differential UKB |
| 0.003269 | 0.003518 | 7.806 | 0.7375 | 1.831 | 0.09753 | True | False | True | True | Sex-specific 1e-2 VUMC, sex-differential UKB |
| 0.003269 | 0.003518 | 7.778 | 0.7393 | 1.834 | 0.09539 | True | False | True | True | Sex-specific 1e-2 VUMC, sex-differential UKB |
| 0.003269 | 0.003518 | 7.709 | 0.6876 | 1.83 | 0.09776 | True | False | True | True | Sex-specific 1e-2 VUMC, sex-differential UKB |
| 0.003269 | 0.003518 | 7.709 | 0.6876 | 1.829 | 0.09782 | True | False | True | True | Sex-specific 1e-2 VUMC, sex-differential UKB |
| 0.003269 | 0.003518 | 7.709 | 0.6876 | 1.832 | 0.1002 | True | False | True | True | Sex-specific 1e-2 VUMC, sex-differential UKB |
| 0.003269 | 0.003518 | 7.694 | 0.7059 | 1.83 | 0.1025 | True | False | True | True | Sex-specific 1e-2 VUMC, sex-differential UKB |
| 0.003269 | 0.003518 | 7.662 | 0.6965 | 1.821 | 0.1037 | True | False | True | True | Sex-specific 1e-2 VUMC, sex-differential UKB |
| 0.003271 | 0.003519 | 7.703 | 0.687 | 1.771 | 0.09036 | True | False | True | True | Sex-specific 1e-2 VUMC, sex-differential UKB |
| 0.003271 | 0.003519 | 7.688 | 0.6923 | 1.785 | 0.09089 | True | False | True | True | Sex-specific 1e-2 VUMC, sex-differential UKB |
| 0.003271 | 0.003519 | 7.662 | 0.6917 | 1.785 | 0.09054 | True | False | True | True | Sex-specific 1e-2 VUMC, sex-differential UKB |
| 0.003271 | 0.003519 | 7.66 | 0.692 | 1.781 | 0.09436 | True | False | True | True | Sex-specific 1e-2 VUMC, sex-differential UKB |
| 0.003271 | 0.003519 | 7.591 | 0.6474 | 1.739 | 0.09182 | True | False | True | True | Sex-specific 1e-2 VUMC, sex-differential UKB |
| 0.003271 | 0.003519 | 7.688 | 0.6923 | 1.777 | 0.09574 | True | False | True | True | Sex-specific 1e-2 VUMC, sex-differential UKB |
| 0.003271 | 0.003519 | 7.69 | 0.6919 | 1.775 | 0.09273 | True | False | True | True | Sex-specific 1e-2 VUMC, sex-differential UKB |
| 0.003264 | 0.003512 | 7.814 | 0.5988 | 1.725 | 0.1278 | True | False | True | True | Sex-specific 1e-2 VUMC, sex-differential UKB |
| 0.003271 | 0.003519 | 7.662 | 0.6916 | 1.775 | 0.09306 | True | False | True | True | Sex-specific 1e-2 VUMC, sex-differential UKB |
| 0.003271 | 0.00352 | 7.662 | 0.6917 | 1.776 | 0.09458 | True | False | True | True | Sex-specific 1e-2 VUMC, sex-differential UKB |
| 0.003272 | 0.00352 | 7.762 | 0.7393 | 1.776 | 0.07766 | True | False | True | True | Sex-specific 1e-2 VUMC, sex-differential UKB |
| 0.003272 | 0.00352 | 7.772 | 0.7233 | 1.781 | 0.07155 | True | False | True | True | Sex-specific 1e-2 VUMC, sex-differential UKB |
| 0.003272 | 0.00352 | 7.806 | 0.7396 | 1.777 | 0.07767 | True | False | True | True | Sex-specific 1e-2 VUMC, sex-differential UKB |
| 0.003272 | 0.00352 | 7.783 | 0.7408 | 1.778 | 0.07952 | True | False | True | True | Sex-specific 1e-2 VUMC, sex-differential UKB |

| SE_UKB_female | SE_UKB_male | LOG10P_VUMC_female | LOG10P_VUMC_male | LOG10P_UKB_female | LOG10P_UKB_male | female_effect_direction_replication | male_effect_direction_replication | sex_effect_comparison_replication | pvalue_effect_comparison_replication | replication_type |
|---|---|---|---|---|---|---|---|---|---|---|
| 0.003272 | 0.00352 | 7.748 | 0.7379 | 1.816 | 0.0675 | True | False | True | True | Sex-specific 1e-2 VUMC, sex-differential UKB |
| 0.003272 | 0.00352 | 7.806 | 0.7395 | 1.778 | 0.07761 | True | False | True | True | Sex-specific 1e-2 VUMC, sex-differential UKB |
| 0.003265 | 0.003507 | 0.04022 | 7.248 | 1.998 | 3.015 | False | True | True | True | Sex-specific 1e-2 VUMC, sex-differential UKB |
| 0.003264 | 0.003506 | 0.05752 | 7.391 | 2.123 | 3.059 | False | True | True | True | Sex-specific 1e-2 VUMC, sex-differential UKB |
| 0.003264 | 0.003506 | 0.05354 | 7.369 | 2.071 | 3.016 | False | True | True | True | Sex-specific 1e-2 VUMC, sex-differential UKB |
| 0.003266 | 0.00351 | 0.1083 | 3.04 | 1.454 | 1.309 | False | True | True | False | |
| 0.003259 | 0.003504 | 0.03024 | 3.403 | 2.221 | 2.399 | False | True | True | True | Sex-specific 1e-2 VUMC, sex-differential UKB |
| 0.003259 | 0.003503 | 0.06029 | 3.607 | 2.263 | 2.431 | False | True | True | True | Sex-specific 1e-2 VUMC, sex-differential UKB |
| 0.003266 | 0.00351 | 0.07488 | 2.716 | 1.403 | 1.339 | False | True | True | False | |
| 0.003259 | 0.003504 | 0.03669 | 3.411 | 2.286 | 2.385 | False | True | True | True | Sex-specific 1e-2 VUMC, sex-differential UKB |
| 0.003259 | 0.003503 | 0.01775 | 3.285 | 2.253 | 2.449 | False | True | True | True | Sex-specific 1e-2 VUMC, sex-differential UKB |
| 0.003266 | 0.00351 | 0.1037 | 2.915 | 1.453 | 1.263 | False | True | False | False | |
| 0.003261 | 0.003505 | 0.01558 | 3.274 | 2.248 | 2.505 | False | True | True | True | Sex-specific 1e-2 VUMC, sex-differential UKB |
| 0.003259 | 0.003504 | 0.02243 | 3.308 | 2.227 | 2.431 | False | True | True | True | Sex-specific 1e-2 VUMC, sex-differential UKB |
| 0.003266 | 0.00351 | 0.08158 | 2.721 | 1.492 | 1.313 | False | True | False | False | |
| 0.003259 | 0.003503 | 0.04384 | 3.387 | 2.303 | 2.456 | False | True | True | True | Sex-specific 1e-2 VUMC, sex-differential UKB |
| 0.003265 | 0.00351 | 0.1229 | 2.922 | 1.409 | 1.337 | False | True | True | False | |
| 0.003266 | 0.003509 | 0.1212 | 2.906 | 1.452 | 1.323 | False | True | True | False | |
| 0.003259 | 0.003503 | 0.06828 | 3.404 | 2.17 | 2.451 | False | True | True | True | Sex-specific 1e-2 VUMC, sex-differential UKB |
| 0.003259 | 0.003502 | 0.04354 | 3.19 | 2.222 | 2.516 | False | True | True | True | Sex-specific 1e-2 VUMC, sex-differential UKB |
| 0.003259 | 0.003502 | 0.04966 | 3.342 | 2.256 | 2.464 | False | True | True | True | Sex-specific 1e-2 VUMC, sex-differential UKB |
| 0.003266 | 0.00351 | 0.1187 | 2.853 | 1.421 | 1.342 | False | True | True | False | |
| 0.003266 | 0.00351 | 0.1228 | 2.854 | 1.455 | 1.329 | False | True | True | False | |
| 0.00326 | 0.003503 | 0.05367 | 3.348 | 2.28 | 2.48 | False | True | True | True | Sex-specific 1e-2 VUMC, sex-differential UKB |
| 0.00326 | 0.003504 | 0.04675 | 3.336 | 2.274 | 2.459 | False | True | True | True | Sex-specific 1e-2 VUMC, sex-differential UKB |
| 0.003262 | 0.003506 | 0.0412 | 3.284 | 2.339 | 2.501 | False | True | True | True | Sex-specific 1e-2 VUMC, sex-differential UKB |
| 0.003261 | 0.003504 | 0.05465 | 3.353 | 2.263 | 2.447 | False | True | True | True | Sex-specific 1e-2 VUMC, sex-differential UKB |
| 0.003262 | 0.003505 | 0.06362 | 3.294 | 2.41 | 2.53 | False | True | True | True | Sex-specific 1e-2 VUMC, sex-differential UKB |
| 0.003269 | 0.003513 | 0.05352 | 3.233 | 2.206 | 2.417 | False | True | True | True | Sex-specific 1e-2 VUMC, sex-differential UKB |

| SE_UKB_female | SE_UKB_male | LOG10P_VUMC_female | LOG10P_VUMC_male | LOG10P_UKB_female | LOG10P_UKB_male | female_effect_direction_replication | male_effect_direction_replication | sex_effect_comparison_replication | pvalue_effect_comparison_replication | replication_type |
|---|---|---|---|---|---|---|---|---|---|---|
| 0.00378 | 0.004091 | 7.231 | 0.3424 | 6.611 | 1.124 | True | True | True | True | Sex-specific 1e-2 VUMC, sex-differential UKB |
| 0.003774 | 0.004085 | 7.332 | 0.3201 | 6.603 | 1.13 | True | True | True | True | Sex-specific 1e-2 VUMC, sex-differential UKB |
| 0.003774 | 0.004085 | 7.261 | 0.3202 | 6.6 | 1.115 | True | True | True | True | Sex-specific 1e-2 VUMC, sex-differential UKB |
| 0.003774 | 0.004085 | 7.269 | 0.3201 | 6.578 | 1.119 | True | True | True | True | Sex-specific 1e-2 VUMC, sex-differential UKB |
| 0.003774 | 0.004085 | 7.27 | 0.3202 | 6.578 | 1.112 | True | True | True | True | Sex-specific 1e-2 VUMC, sex-differential UKB |
| 0.003774 | 0.004085 | 7.269 | 0.3202 | 6.577 | 1.113 | True | True | True | True | Sex-specific 1e-2 VUMC, sex-differential UKB |
| 0.003774 | 0.004085 | 7.269 | 0.3197 | 6.573 | 1.114 | True | True | True | True | Sex-specific 1e-2 VUMC, sex-differential UKB |
| 0.003774 | 0.004085 | 7.269 | 0.3202 | 6.572 | 1.113 | True | True | True | True | Sex-specific 1e-2 VUMC, sex-differential UKB |
| 0.003774 | 0.004085 | 7.159 | 0.3694 | 6.572 | 1.118 | True | True | True | True | Sex-specific 1e-2 VUMC, sex-differential UKB |
| 0.003774 | 0.004085 | 7.159 | 0.3694 | 6.567 | 1.113 | True | True | True | True | Sex-specific 1e-2 VUMC, sex-differential UKB |
| 0.003774 | 0.004085 | 7.159 | 0.3694 | 6.567 | 1.113 | True | True | True | True | Sex-specific 1e-2 VUMC, sex-differential UKB |
| 0.003774 | 0.004085 | 7.159 | 0.3694 | 6.566 | 1.113 | True | True | True | True | Sex-specific 1e-2 VUMC, sex-differential UKB |
| 0.00377 | 0.004082 | 7.263 | 0.3818 | 6.849 | 1.127 | True | True | True | True | Sex-specific 1e-2 VUMC, sex-differential UKB |
| 0.003774 | 0.004085 | 7.159 | 0.3694 | 6.564 | 1.113 | True | True | True | True | Sex-specific 1e-2 VUMC, sex-differential UKB |
| 0.003774 | 0.004085 | 7.159 | 0.3694 | 6.565 | 1.118 | True | True | True | True | Sex-specific 1e-2 VUMC, sex-differential UKB |
| 0.003774 | 0.004085 | 7.16 | 0.3693 | 6.565 | 1.118 | True | True | True | True | Sex-specific 1e-2 VUMC, sex-differential UKB |
| 0.003774 | 0.004085 | 7.16 | 0.3693 | 6.559 | 1.116 | True | True | True | True | Sex-specific 1e-2 VUMC, sex-differential UKB |
| 0.003774 | 0.004086 | 7.126 | 0.3543 | 6.559 | 1.112 | True | True | True | True | Sex-specific 1e-2 VUMC, sex-differential UKB |
| 0.00377 | 0.004082 | 7.282 | 0.3769 | 6.865 | 1.067 | True | True | True | True | Sex-specific 1e-2 VUMC, sex-differential UKB |
| 0.003464 | 0.003737 | 3.225 | 0.6787 | 7.076 | 0.4371 | True | True | True | True | Sex-specific 1e-2 VUMC, sex-differential UKB |
| 0.003773 | 0.004083 | 7.057 | 0.3809 | 6.304 | 1.054 | True | True | True | True | Sex-specific 1e-2 VUMC, sex-differential UKB |
| 0.003387 | 0.003656 | 4.037 | 1.202 | 2.269 | 2.169 | True | True | False | True | |
| 0.003773 | 0.004084 | 7.206 | 0.3601 | 6.315 | 1.068 | True | True | True | True | Sex-specific 1e-2 VUMC, sex-differential UKB |
| 0.003776 | 0.004087 | 7.195 | 0.3652 | 6.608 | 1.061 | True | True | True | True | Sex-specific 1e-2 VUMC, sex-differential UKB |
| 0.003796 | 0.004109 | 7.225 | 0.3528 | 6.692 | 1.084 | True | True | True | True | Sex-specific 1e-2 VUMC, sex-differential UKB |
| 0.003775 | 0.004086 | 7.174 | 0.3555 | 6.544 | 1.068 | True | True | True | True | Sex-specific 1e-2 VUMC, sex-differential UKB |

| SE_UKB_female | SE_UKB_male | LOG10P_VUMC_female | LOG10P_VUMC_male | LOG10P_UKB_female | LOG10P_UKB_male | female_effect_direction_replication | male_effect_direction_replication | sex_effect_comparison_replication | pvalue_effect_comparison_replication | replication_type |
|---|---|---|---|---|---|---|---|---|---|---|
| 0.003463 | 0.003737 | 3.281 | 0.6745 | 7.639 | 0.4121 | True | True | True | True | Sex-specific 1e-2 VUMC & UKB |
| 0.003463 | 0.003737 | 3.238 | 0.6482 | 7.614 | 0.4178 | True | True | True | True | Sex-specific 1e-2 VUMC & UKB |
| 0.003463 | 0.003737 | 3.231 | 0.6481 | 7.648 | 0.4074 | True | True | True | True | Sex-specific 1e-2 VUMC & UKB |
| 0.003463 | 0.003737 | 3.254 | 0.6594 | 7.642 | 0.3967 | True | True | True | True | Sex-specific 1e-2 VUMC & UKB |
| 0.003777 | 0.004089 | 7.31 | 0.3687 | 6.638 | 1.062 | True | True | True | True | Sex-specific 1e-2 VUMC, sex-differential UKB |
| SE_UKB_female | SE_UKB_male | LOG10P_VUMC_female | LOG10P_VUMC_male | LOG10P_UKB_female | LOG10P_UKB_male | female_effect_direction_replication | male_effect_direction_replication | sex_effect_comparison_replication | pvalue_effect_comparison_replication | replication_type |
| 0.003462 | 0.003737 | 3.269 | 0.6701 | 7.761 | 0.3623 | True | True | True | True | Sex-specific 1e-2 VUMC & UKB |
| 0.003776 | 0.004088 | 7.142 | 0.3802 | 6.66 | 1.059 | True | True | True | True | Sex-specific 1e-2 VUMC, sex-differential UKB |
| 0.003462 | 0.003736 | 3.294 | 0.6608 | 7.78 | 0.359 | True | True | True | True | Sex-specific 1e-2 VUMC & UKB |
| 0.003776 | 0.004088 | 7.143 | 0.3857 | 6.717 | 1.04 | True | True | True | True | Sex-specific 1e-2 VUMC, sex-differential UKB |
| 0.003464 | 0.003738 | 3.294 | 0.6904 | 7.852 | 0.3269 | True | True | True | True | Sex-specific 1e-2 VUMC & UKB |
| 0.003464 | 0.003738 | 3.293 | 0.6903 | 7.858 | 0.3266 | True | True | True | True | Sex-specific 1e-2 VUMC & UKB |
| 0.003463 | 0.003737 | 3.271 | 0.6524 | 7.687 | 0.3809 | True | True | True | True | Sex-specific 1e-2 VUMC & UKB |
| 0.003463 | 0.003737 | 3.27 | 0.6523 | 7.711 | 0.3823 | True | True | True | True | Sex-specific 1e-2 VUMC & UKB |
| 0.003463 | 0.003737 | 3.27 | 0.6523 | 7.681 | 0.3812 | True | True | True | True | Sex-specific 1e-2 VUMC & UKB |
| 0.003463 | 0.003738 | 3.315 | 0.6485 | 7.756 | 0.3809 | True | True | True | True | Sex-specific 1e-2 VUMC & UKB |
| 0.003464 | 0.003739 | 3.247 | 0.6484 | 7.638 | 0.3773 | True | True | True | True | Sex-specific 1e-2 VUMC & UKB |
| 0.003384 | 0.003652 | 4.269 | 1.226 | 1.902 | 2.167 | True | True | False | False | |
| 0.003777 | 0.004089 | 7.43 | 0.3471 | 6.558 | 1.037 | True | True | True | True | Sex-specific 1e-2 VUMC, sex-differential UKB |
| 0.003384 | 0.003652 | 4.271 | 1.22 | 1.91 | 2.168 | True | True | False | False | |
| 0.003373 | 0.003643 | 4.162 | 1.097 | 1.931 | 2.311 | True | True | False | False | |
| 0.003373 | 0.003642 | 4.059 | 1.137 | 1.946 | 2.259 | True | True | False | False | |
| 0.003373 | 0.003642 | 4.112 | 1.147 | 1.935 | 2.259 | True | True | False | False | |
| 0.003373 | 0.003642 | 4.134 | 1.117 | 1.931 | 2.26 | True | True | False | False | |
| 0.003373 | 0.003642 | 4.02 | 1.081 | 1.955 | 2.272 | True | True | False | False | |
| 0.003778 | 0.00409 | 7.459 | 0.3603 | 6.57 | 1.047 | True | True | True | True | Sex-specific 1e-2 VUMC, sex-differential UKB |

| SE_UKB_female | SE_UKB_male | LOG10P_VUMC_female | LOG10P_VUMC_male | LOG10P_UKB_female | LOG10P_UKB_male | female_effect_direction_replication | male_effect_direction_replication | sex_effect_comparison_replication | pvalue_effect_comparison_replication | replication_type |
|---|---|---|---|---|---|---|---|---|---|---|
| 0.003373 | 0.003642 | 4.065 | 1.087 | 1.941 | 2.318 | True | True | False | False | |
| 0.003373 | 0.003642 | 4.046 | 1.142 | 1.928 | 2.329 | True | True | False | False | |
| 0.003777 | 0.004088 | 7.266 | 0.3899 | 6.561 | 1.042 | True | True | True | True | Sex-specific 1e-2 VUMC, sex-differential UKB |
| 0.003374 | 0.003643 | 3.867 | 1.133 | 1.949 | 2.366 | True | True | False | False | |
| 0.003373 | 0.003642 | 3.823 | 1.174 | 1.953 | 2.33 | True | True | False | False | |
| 0.003371 | 0.00364 | 3.964 | 1.093 | 2.045 | 2.319 | True | True | False | False | |
| 0.003369 | 0.003639 | 4.061 | 1.082 | 2.185 | 2.33 | True | True | False | False | |
| 0.003371 | 0.00364 | 3.852 | 1.125 | 2.029 | 2.363 | True | True | False | False | |
| 0.003369 | 0.003639 | 4 | 1.073 | 2.182 | 2.319 | True | True | False | False | |
| 0.003773 | 0.004084 | 7.404 | 0.3834 | 6.415 | 1.01 | True | True | True | True | Sex-specific 1e-2 VUMC, sex-differential UKB |
| 0.003371 | 0.00364 | 4.068 | 1.137 | 1.94 | 2.329 | True | True | False | False | |
| 0.003371 | 0.003639 | 4.15 | 1.102 | 1.947 | 2.3 | True | True | False | False | |
| 0.003369 | 0.003639 | 4.129 | 1.089 | 2.095 | 2.336 | True | True | False | False | |
| 0.003371 | 0.00364 | 4.083 | 1.135 | 1.944 | 2.318 | True | True | False | False | |
| 0.00337 | 0.003639 | 5.02 | 0.01107 | 8.649 | 0.3525 | True | False | True | True | Sex-specific 1e-2 VUMC & UKB |
| 0.003372 | 0.003641 | 5.054 | 0.004801 | 8.625 | 0.3347 | True | False | True | True | Sex-specific 1e-2 VUMC & UKB |
| 0.003371 | 0.003641 | 5.042 | 0.003944 | 8.656 | 0.3656 | True | False | True | True | Sex-specific 1e-2 VUMC & UKB |
| 0.003618 | 0.003894 | 8.076 | 1.326 | 40.3 | 15.96 | True | True | True | True | Sex-specific 1e-2 VUMC, sex-differential UKB |
| 0.003725 | 0.004013 | 8.729 | 1.559 | 40.25 | 16.87 | True | True | True | True | Sex-specific 1e-2 VUMC, sex-differential UKB |
| 0.003725 | 0.004012 | 8.747 | 1.557 | 40.35 | 16.9 | True | True | True | True | Sex-specific 1e-2 VUMC, sex-differential UKB |
| 0.003718 | 0.004005 | 8.961 | 1.599 | 40.41 | 17.12 | True | True | True | True | Sex-specific 1e-2 VUMC, sex-differential UKB |
| 0.003712 | 0.004 | 8.858 | 1.413 | 39.98 | 17.47 | True | True | True | True | Sex-specific 1e-2 VUMC, sex-differential UKB |
| 0.003702 | 0.003989 | 9.136 | 1.451 | 39.82 | 17.79 | True | True | True | True | Sex-specific 1e-2 VUMC, sex-differential UKB |
| 0.003703 | 0.003989 | 9 | 1.511 | 40.18 | 17.8 | True | True | True | True | Sex-specific 1e-2 VUMC, sex-differential UKB |
| 0.003678 | 0.003963 | 8.549 | 1.382 | 37.66 | 16.42 | True | True | True | True | Sex-specific 1e-2 VUMC, sex-differential UKB |
| 0.003702 | 0.003988 | 9.128 | 1.435 | 39.87 | 17.79 | True | True | True | True | Sex-specific 1e-2 VUMC, sex-differential UKB |
| 0.003704 | 0.003991 | 9.072 | 1.573 | 41.01 | 17.81 | True | True | True | True | Sex-specific 1e-2 VUMC, sex-differential UKB |
| 0.003695 | 0.003982 | 8.922 | 1.396 | 40.07 | 17.59 | True | True | True | True | Sex-specific 1e-2 VUMC, sex-differential UKB |

| SE_UKB_female | SE_UKB_male | LOG10P_VUMC_female | LOG10P_VUMC_male | LOG10P_UKB_female | LOG10P_UKB_male | female_effect_direction_replication | male_effect_direction_replication | sex_effect_comparison_replication | pvalue_effect_comparison_replication | replication_type |
|---|---|---|---|---|---|---|---|---|---|---|
| 0.003691 | 0.003977 | 9.092 | 1.438 | 40.3 | 17.66 | True | True | True | True | Sex-specific 1e-2 VUMC, sex-differential UKB |
| 0.003696 | 0.003983 | 9.389 | 1.419 | 40.64 | 18.14 | True | True | True | True | Sex-specific 1e-2 VUMC, sex-differential UKB |
| 0.003697 | 0.003983 | 9.394 | 1.42 | 40.44 | 18.17 | True | True | True | True | Sex-specific 1e-2 VUMC, sex-differential UKB |
| 0.003716 | 0.004002 | 9.311 | 1.45 | 40.52 | 18.33 | True | True | True | True | Sex-specific 1e-2 VUMC, sex-differential UKB |
| 0.003707 | 0.003993 | 8.92 | 1.338 | 40.51 | 17.54 | True | True | True | True | Sex-specific 1e-2 VUMC, sex-differential UKB |
| 0.003696 | 0.003982 | 9.315 | 1.318 | 40.16 | 18.26 | True | True | True | True | Sex-specific 1e-2 VUMC, sex-differential UKB |
| 0.003696 | 0.003981 | 9.344 | 1.37 | 40.11 | 17.93 | True | True | True | True | Sex-specific 1e-2 VUMC, sex-differential UKB |
| 0.003696 | 0.003981 | 9.277 | 1.386 | 40.25 | 18.26 | True | True | True | True | Sex-specific 1e-2 VUMC, sex-differential UKB |
| 0.003701 | 0.003985 | 9.366 | 1.378 | 40.49 | 17.87 | True | True | True | True | Sex-specific 1e-2 VUMC, sex-differential UKB |
| 0.003719 | 0.004005 | 9.191 | 1.364 | 41.04 | 17.47 | True | True | True | True | Sex-specific 1e-2 VUMC, sex-differential UKB |
| 0.003696 | 0.003981 | 9.331 | 1.374 | 40.22 | 18.17 | True | True | True | True | Sex-specific 1e-2 VUMC, sex-differential UKB |
| 0.003731 | 0.004019 | 8.846 | 1.218 | 39.8 | 17.16 | True | True | True | True | Sex-specific 1e-2 VUMC, sex-differential UKB |
| SE_UKB_female | SE_UKB_male | LOG10P_VUMC_female | LOG10P_VUMC_male | LOG10P_UKB_female | LOG10P_UKB_male | female_effect_direction_replication | male_effect_direction_replication | sex_effect_comparison_replication | pvalue_effect_comparison_replication | replication_type |
| 0.003708 | 0.003993 | 8.98 | 1.363 | 40.47 | 17.97 | True | True | True | True | Sex-specific 1e-2 VUMC, sex-differential UKB |
| 0.003708 | 0.003992 | 9.125 | 1.362 | 40.13 | 17.89 | True | True | True | True | Sex-specific 1e-2 VUMC, sex-differential UKB |
| 0.003731 | 0.004017 | 8.267 | 1.233 | 38.7 | 15.73 | True | True | True | True | Sex-specific 1e-2 VUMC, sex-differential UKB |
| 0.003616 | 0.003888 | 10.28 | 1.683 | 37.55 | 18.55 | True | True | True | True | Sex-specific 1e-2 VUMC, sex-differential UKB |
| 0.003443 | 0.003708 | 6.557 | 1.364 | 19.85 | 9.48 | True | True | True | True | Sex-specific 1e-2 VUMC, sex-differential UKB |
| 0.004257 | 0.004568 | 5.211 | 0.271 | 14.9 | 7.039 | True | True | True | True | Sex-specific 1e-2 VUMC, sex-specific 5e-8 UKB |
| 0.004201 | 0.004511 | 5.612 | 0.3218 | 16.24 | 7.772 | True | True | True | True | Sex-specific 1e-2 VUMC, sex-differential UKB |
| 0.004201 | 0.004511 | 5.639 | 0.3168 | 16.2 | 7.772 | True | True | True | True | Sex-specific 1e-2 VUMC, sex-differential UKB |
| 0.004204 | 0.004513 | 5.596 | 0.3213 | 16.1 | 7.896 | True | True | True | True | Sex-specific 1e-2 VUMC, sex-differential UKB |
| 0.004219 | 0.004528 | 5.499 | 0.3353 | 16.17 | 7.901 | True | True | True | True | Sex-specific 1e-2 VUMC, sex-differential UKB |
| 0.004219 | 0.004528 | 5.546 | 0.3302 | 16.11 | 7.878 | True | True | True | True | Sex-specific 1e-2 VUMC, sex-differential UKB |
| 0.003968 | 0.004267 | 6.421 | 0.7336 | 24.64 | 15.08 | True | True | True | True | Sex-specific 1e-2 VUMC, sex-differential UKB |
| 0.004258 | 0.004569 | 5.17 | 0.2618 | 14.94 | 7.222 | True | True | True | True | Sex-specific 1e-2 VUMC, sex-specific 5e-8 UKB |

| SE_UKB_female | SE_UKB_male | LOG10P_VUMC_female | LOG10P_VUMC_male | LOG10P_UKB_female | LOG10P_UKB_male | female_effect_direction_replication | male_effect_direction_replication | sex_effect_comparison_replication | pvalue_effect_comparison_replication | replication_type |
|---|---|---|---|---|---|---|---|---|---|---|
| 0.003968 | 0.004266 | 6.42 | 0.741 | 24.64 | 15.13 | True | True | True | True | Sex-specific 1e-2 VUMC, sex-differential UKB |
| 0.004299 | 0.004612 | 4.463 | 0.1737 | 14.14 | 6.408 | True | True | True | True | Sex-specific 1e-2 VUMC, sex-specific 5e-8 UKB |
| 0.004302 | 0.004615 | 4.563 | 0.1508 | 14.16 | 6.517 | True | True | True | True | Sex-specific 1e-2 VUMC, sex-specific 5e-8 UKB |
| 0.004671 | 0.005017 | 8.932 | 1.233 | 37.58 | 23.16 | True | True | True | True | Sex-specific 1e-2 VUMC, sex-differential UKB |
| 0.004668 | 0.005013 | 8.831 | 1.306 | 37.73 | 23.23 | True | True | True | True | Sex-specific 1e-2 VUMC, sex-differential UKB |
| 0.004667 | 0.005011 | 8.881 | 1.284 | 37.65 | 22.8 | True | True | True | True | Sex-specific 1e-2 VUMC, sex-differential UKB |
| 0.004667 | 0.005011 | 8.881 | 1.284 | 37.67 | 22.79 | True | True | True | True | Sex-specific 1e-2 VUMC, sex-differential UKB |
| 0.004667 | 0.005011 | 8.828 | 1.323 | 37.65 | 22.79 | True | True | True | True | Sex-specific 1e-2 VUMC, sex-differential UKB |
| 0.004668 | 0.005013 | 8.792 | 1.317 | 37.54 | 22.68 | True | True | True | True | Sex-specific 1e-2 VUMC, sex-differential UKB |
| 0.004659 | 0.005 | 8.957 | 1.416 | 37.25 | 22.18 | True | True | True | True | Sex-specific 1e-2 VUMC, sex-differential UKB |
| 0.004668 | 0.005012 | 9.014 | 1.325 | 37.52 | 22.64 | True | True | True | True | Sex-specific 1e-2 VUMC, sex-differential UKB |
| 0.004668 | 0.005012 | 9.043 | 1.309 | 37.55 | 22.6 | True | True | True | True | Sex-specific 1e-2 VUMC, sex-differential UKB |
| 0.004667 | 0.005011 | 9.101 | 1.289 | 37.44 | 22.69 | True | True | True | True | Sex-specific 1e-2 VUMC, sex-differential UKB |
| 0.004668 | 0.005012 | 8.994 | 1.321 | 37.56 | 22.48 | True | True | True | True | Sex-specific 1e-2 VUMC, sex-differential UKB |
| 0.004666 | 0.00501 | 8.918 | 1.349 | 37.8 | 22.45 | True | True | True | True | Sex-specific 1e-2 VUMC, sex-differential UKB |
| 0.004667 | 0.005011 | 9.115 | 1.393 | 37.72 | 22.52 | True | True | True | True | Sex-specific 1e-2 VUMC, sex-differential UKB |
| 0.004667 | 0.005011 | 9.123 | 1.409 | 37.69 | 22.49 | True | True | True | True | Sex-specific 1e-2 VUMC, sex-differential UKB |
| 0.004667 | 0.005011 | 8.948 | 1.373 | 37.83 | 22.47 | True | True | True | True | Sex-specific 1e-2 VUMC, sex-differential UKB |
| 0.004667 | 0.005011 | 8.954 | 1.379 | 37.8 | 22.46 | True | True | True | True | Sex-specific 1e-2 VUMC, sex-differential UKB |
| 0.004666 | 0.00501 | 8.99 | 1.391 | 37.66 | 22.55 | True | True | True | True | Sex-specific 1e-2 VUMC, sex-differential UKB |
| 0.004666 | 0.005011 | 8.975 | 1.386 | 37.83 | 22.44 | True | True | True | True | Sex-specific 1e-2 VUMC, sex-differential UKB |
| 0.004666 | 0.005011 | 9.021 | 1.388 | 37.87 | 22.46 | True | True | True | True | Sex-specific 1e-2 VUMC, sex-differential UKB |
| 0.004666 | 0.005011 | 9.119 | 1.41 | 37.63 | 22.48 | True | True | True | True | Sex-specific 1e-2 VUMC, sex-differential UKB |
| 0.004666 | 0.00501 | 9.005 | 1.377 | 37.73 | 22.56 | True | True | True | True | Sex-specific 1e-2 VUMC, sex-differential UKB |
| 0.004666 | 0.00501 | 8.999 | 1.378 | 37.75 | 22.62 | True | True | True | True | Sex-specific 1e-2 VUMC, sex-differential UKB |
| 0.004666 | 0.00501 | 8.99 | 1.379 | 37.71 | 22.55 | True | True | True | True | Sex-specific 1e-2 VUMC, sex-differential UKB |

| SE_UKB_female | SE_UKB_male | LOG10P_VUMC_female | LOG10P_VUMC_male | LOG10P_UKB_female | LOG10P_UKB_male | female_effect_direction_replication | male_effect_direction_replication | sex_effect_comparison_replication | pvalue_effect_comparison_replication | replication_type |
|---|---|---|---|---|---|---|---|---|---|---|
| 0.004666 | 0.00501 | 8.984 | 1.384 | 37.69 | 22.6 | True | True | True | True | Sex-specific 1e-2 VUMC, sex-differential UKB |
| 0.004665 | 0.00501 | 8.993 | 1.392 | 37.79 | 22.53 | True | True | True | True | Sex-specific 1e-2 VUMC, sex-differential UKB |
| 0.004666 | 0.00501 | 8.996 | 1.381 | 37.63 | 22.57 | True | True | True | True | Sex-specific 1e-2 VUMC, sex-differential UKB |
| 0.004666 | 0.00501 | 8.955 | 1.379 | 37.64 | 22.52 | True | True | True | True | Sex-specific 1e-2 VUMC, sex-differential UKB |
| 0.0046 | 0.004938 | 9.381 | 1.65 | 39.44 | 24.71 | True | True | True | True | Sex-specific 1e-2 VUMC, sex-differential UKB |
| 0.004666 | 0.00501 | 8.932 | 1.356 | 37.74 | 22.6 | True | True | True | True | Sex-specific 1e-2 VUMC, sex-differential UKB |
| 0.004666 | 0.00501 | 9.025 | 1.38 | 37.61 | 22.55 | True | True | True | True | Sex-specific 1e-2 VUMC, sex-differential UKB |
| 0.004666 | 0.00501 | 8.84 | 1.361 | 37.67 | 22.6 | True | True | True | True | Sex-specific 1e-2 VUMC, sex-differential UKB |
| 0.004666 | 0.00501 | 8.812 | 1.36 | 37.46 | 22.53 | True | True | True | True | Sex-specific 1e-2 VUMC, sex-differential UKB |
| 0.004666 | 0.00501 | 8.811 | 1.363 | 37.45 | 22.57 | True | True | True | True | Sex-specific 1e-2 VUMC, sex-differential UKB |
| 0.004666 | 0.00501 | 8.819 | 1.354 | 37.31 | 22.52 | True | True | True | True | Sex-specific 1e-2 VUMC, sex-differential UKB |
| 0.004666 | 0.00501 | 8.83 | 1.354 | 37.52 | 22.53 | True | True | True | True | Sex-specific 1e-2 VUMC, sex-differential UKB |
| 0.004666 | 0.00501 | 8.868 | 1.365 | 37.51 | 22.53 | True | True | True | True | Sex-specific 1e-2 VUMC, sex-differential UKB |
| 0.004666 | 0.00501 | 8.844 | 1.353 | 37.42 | 22.53 | True | True | True | True | Sex-specific 1e-2 VUMC, sex-differential UKB |
| 0.004666 | 0.00501 | 8.785 | 1.343 | 37.4 | 22.53 | True | True | True | True | Sex-specific 1e-2 VUMC, sex-differential UKB |
| 0.004666 | 0.00501 | 8.744 | 1.334 | 37.48 | 22.53 | True | True | True | True | Sex-specific 1e-2 VUMC, sex-differential UKB |
| 0.004666 | 0.00501 | 8.743 | 1.334 | 37.48 | 22.53 | True | True | True | True | Sex-specific 1e-2 VUMC, sex-differential UKB |
| 0.004666 | 0.00501 | 8.747 | 1.322 | 37.48 | 22.53 | True | True | True | True | Sex-specific 1e-2 VUMC, sex-differential UKB |
| 0.004661 | 0.005005 | 8.732 | 1.325 | 37.22 | 22.52 | True | True | True | True | Sex-specific 1e-2 VUMC, sex-differential UKB |
| 0.005064 | 0.005465 | 4.73 | 1.963 | 39.87 | 15.64 | True | True | True | True | Sex-specific 1e-2 VUMC, sex-differential UKB |
| 0.005065 | 0.005466 | 4.681 | 1.998 | 39.9 | 15.66 | True | True | True | True | Sex-specific 1e-2 VUMC, sex-differential UKB |
| SE_UKB_female | SE_UKB_male | LOG10P_VUMC_female | LOG10P_VUMC_male | LOG10P_UKB_female | LOG10P_UKB_male | female_effect_direction_replication | male_effect_direction_replication | sex_effect_comparison_replication | pvalue_effect_comparison_replication | replication_type |
| 0.004424 | 0.00477 | 8.525 | 1.657 | 64.52 | 28.32 | True | True | True | True | Sex-specific 1e-2 VUMC, sex-differential UKB |
| 0.004433 | 0.004781 | 8.757 | 1.642 | 64.17 | 27.69 | True | True | True | True | Sex-specific 1e-2 VUMC, sex-differential UKB |
| 0.004832 | 0.005228 | 9.753 | 1.208 | 59.28 | 35.69 | True | True | True | True | Sex-specific 1e-2 VUMC, sex-differential UKB |
| 0.004829 | 0.005223 | 9.606 | 1.204 | 59.16 | 35.92 | True | True | True | True | Sex-specific 1e-2 VUMC, sex-differential UKB |

| SE_UKB_female | SE_UKB_male | LOG10P_VUMC_female | LOG10P_VUMC_male | LOG10P_UKB_female | LOG10P_UKB_male | female_effect_direction_replication | male_effect_direction_replication | sex_effect_comparison_replication | pvalue_effect_comparison_replication | replication_type |
|---|---|---|---|---|---|---|---|---|---|---|
| 0.00483 | 0.005224 | 9.549 | 1.16 | 59.24 | 35.93 | True | True | True | True | Sex-specific 1e-2 VUMC, sex-differential UKB |
| 0.004831 | 0.005225 | 9.639 | 1.202 | 59.06 | 35.83 | True | True | True | True | Sex-specific 1e-2 VUMC, sex-differential UKB |
| 0.00483 | 0.005223 | 9.644 | 1.232 | 59.01 | 35.85 | True | True | True | True | Sex-specific 1e-2 VUMC, sex-differential UKB |
| 0.00483 | 0.005223 | 9.644 | 1.232 | 59.01 | 35.85 | True | True | True | True | Sex-specific 1e-2 VUMC, sex-differential UKB |
| 0.004902 | 0.005304 | 9.238 | 1.169 | 58.17 | 36.1 | True | True | True | True | Sex-specific 1e-2 VUMC, sex-differential UKB |
| 0.004901 | 0.005303 | 9.076 | 1.18 | 57.98 | 36.22 | True | True | True | True | Sex-specific 1e-2 VUMC, sex-differential UKB |
| 0.0049 | 0.005302 | 9.257 | 1.143 | 57.94 | 35.94 | True | True | True | True | Sex-specific 1e-2 VUMC, sex-differential UKB |
| 0.004839 | 0.005233 | 9.514 | 1.343 | 58.02 | 35.5 | True | True | True | True | Sex-specific 1e-2 VUMC, sex-differential UKB |
| 0.004928 | 0.005329 | 8.535 | 1.005 | 55.68 | 36.07 | True | True | True | True | Sex-specific 1e-2 VUMC, sex-differential UKB |
| 0.004929 | 0.00533 | 8.565 | 0.9862 | 56 | 36.47 | True | True | True | True | Sex-specific 1e-2 VUMC, sex-differential UKB |
| 0.004928 | 0.005328 | 8.417 | 0.9525 | 55.73 | 36.29 | True | True | True | True | Sex-specific 1e-2 VUMC, sex-differential UKB |
| 0.004927 | 0.005327 | 8.537 | 1.005 | 55.85 | 36.4 | True | True | True | True | Sex-specific 1e-2 VUMC, sex-differential UKB |
| 0.004927 | 0.005327 | 8.433 | 0.9983 | 55.82 | 36.22 | True | True | True | True | Sex-specific 1e-2 VUMC, sex-differential UKB |
| 0.004927 | 0.005327 | 8.449 | 1.001 | 55.74 | 36.36 | True | True | True | True | Sex-specific 1e-2 VUMC, sex-differential UKB |
| 0.00493 | 0.005331 | 8.325 | 1.018 | 55.75 | 36.55 | True | True | True | True | Sex-specific 1e-2 VUMC, sex-differential UKB |
| 0.004929 | 0.005328 | 8.296 | 0.9364 | 55.99 | 36.46 | True | True | True | True | Sex-specific 1e-2 VUMC, sex-differential UKB |
| 0.004941 | 0.005339 | 8.45 | 1.138 | 56 | 36.04 | True | True | True | True | Sex-specific 1e-2 VUMC, sex-differential UKB |
| 0.004941 | 0.005339 | 8.45 | 1.138 | 56 | 36.05 | True | True | True | True | Sex-specific 1e-2 VUMC, sex-differential UKB |
| 0.004941 | 0.005339 | 8.449 | 1.138 | 56 | 36.05 | True | True | True | True | Sex-specific 1e-2 VUMC, sex-differential UKB |
| 0.004945 | 0.005344 | 8.492 | 1.16 | 55.91 | 35.98 | True | True | True | True | Sex-specific 1e-2 VUMC, sex-differential UKB |
| 0.004926 | 0.005323 | 8.911 | 1.196 | 56.44 | 36.24 | True | True | True | True | Sex-specific 1e-2 VUMC, sex-differential UKB |
| 0.004928 | 0.005324 | 9.017 | 1.206 | 56.33 | 36.05 | True | True | True | True | Sex-specific 1e-2 VUMC, sex-differential UKB |
| 0.004927 | 0.005324 | 9.016 | 1.206 | 56.27 | 36.01 | True | True | True | True | Sex-specific 1e-2 VUMC, sex-differential UKB |
| 0.004938 | 0.005335 | 9.2 | 1.249 | 55.58 | 36.47 | True | True | True | True | Sex-specific 1e-2 VUMC, sex-differential UKB |
| 0.00494 | 0.005337 | 9.244 | 1.249 | 55.69 | 36.37 | True | True | True | True | Sex-specific 1e-2 VUMC, sex-differential UKB |
| 0.00504 | 0.005448 | 9.271 | 1.355 | 54.24 | 34.59 | True | True | True | True | Sex-specific 1e-2 VUMC, sex-differential UKB |

| SE_UKB_female | SE_UKB_male | LOG10P_VUMC_female | LOG10P_VUMC_male | LOG10P_UKB_female | LOG10P_UKB_male | female_effect_direction_replication | male_effect_direction_replication | sex_effect_comparison_replication | pvalue_effect_comparison_replication | replication_type |
|---|---|---|---|---|---|---|---|---|---|---|
| 0.004643 | 0.004995 | 5.727 | 1.585 | 34.5 | 24.82 | True | True | True | True | Sex-specific 1e-2 VUMC, sex-differential UKB |
| 0.004581 | 0.004928 | 5.163 | 1.53 | 30.9 | 21.96 | True | True | True | True | Sex-specific 1e-2 VUMC, sex-differential UKB |
| 0.004235 | 0.004551 | 4.402 | 0.6259 | 33.23 | 25.25 | True | True | True | True | Sex-specific 1e-2 VUMC, sex-differential UKB |
| 0.004235 | 0.00455 | 4.393 | 0.6541 | 33.28 | 25.41 | True | True | True | True | Sex-specific 1e-2 VUMC, sex-differential UKB |
| 0.004233 | 0.004549 | 4.3 | 0.6151 | 33.16 | 25.07 | True | True | True | True | Sex-specific 1e-2 VUMC, sex-differential UKB |
| 0.004234 | 0.004549 | 4.425 | 0.7162 | 33.56 | 25.48 | True | True | True | True | Sex-specific 1e-2 VUMC, sex-differential UKB |
| 0.004231 | 0.004546 | 4.347 | 0.5866 | 33.38 | 25.44 | True | True | True | True | Sex-specific 1e-2 VUMC, sex-differential UKB |
| 0.004231 | 0.004545 | 4.376 | 0.6053 | 33.34 | 25.33 | True | True | True | True | Sex-specific 1e-2 VUMC, sex-differential UKB |
| 0.004639 | 0.004988 | 5.847 | 1.454 | 35.13 | 25.73 | True | True | True | True | Sex-specific 1e-2 VUMC, sex-differential UKB |
| 0.004556 | 0.004901 | 5.744 | 1.173 | 28.98 | 22.42 | True | True | True | True | Sex-specific 1e-2 VUMC, sex-differential UKB |
| 0.004539 | 0.004878 | 5.618 | 1.047 | 29.52 | 23.04 | True | True | True | True | Sex-specific 1e-2 VUMC, sex-differential UKB |
| 0.004551 | 0.004885 | 7.183 | 1.318 | 34.21 | 26.02 | True | True | True | True | Sex-specific 1e-2 VUMC, sex-differential UKB |
| 0.004554 | 0.00489 | 7.265 | 1.316 | 33.33 | 25.78 | True | True | True | True | Sex-specific 1e-2 VUMC, sex-differential UKB |
| 0.004555 | 0.00489 | 7.333 | 1.327 | 33.38 | 25.54 | True | True | True | True | Sex-specific 1e-2 VUMC, sex-differential UKB |
| 0.004581 | 0.00492 | 7.303 | 1.391 | 33.03 | 26.02 | True | True | True | True | Sex-specific 1e-2 VUMC, sex-differential UKB |
| 0.004553 | 0.004889 | 7.488 | 1.453 | 34.91 | 27.25 | True | True | True | True | Sex-specific 1e-2 VUMC, sex-differential UKB |
| 0.003681 | 0.003959 | 9.407 | 1.463 | 35.49 | 20.9 | True | True | True | True | Sex-specific 1e-2 VUMC, sex-differential UKB |
| 0.003507 | 0.003777 | 7.353 | 1.415 | 40.4 | 22.61 | True | True | True | True | Sex-specific 1e-2 VUMC, sex-differential UKB |
| 0.003481 | 0.003746 | 7.636 | 1.281 | 40.99 | 22.98 | True | True | True | True | Sex-specific 1e-2 VUMC, sex-differential UKB |
| 0.00339 | 0.00365 | 5.481 | 1.058 | 37.85 | 16.92 | True | True | True | True | Sex-specific 1e-2 VUMC, sex-differential UKB |
| 0.003685 | 0.003964 | 9.279 | 1.626 | 36.1 | 21.33 | True | True | True | True | Sex-specific 1e-2 VUMC, sex-differential UKB |
| 0.003478 | 0.003743 | 7.476 | 1.248 | 40.08 | 22.49 | True | True | True | True | Sex-specific 1e-2 VUMC, sex-differential UKB |
| 0.003505 | 0.003776 | 7.371 | 1.413 | 40.5 | 22.59 | True | True | True | True | Sex-specific 1e-2 VUMC, sex-differential UKB |
| 0.003685 | 0.003964 | 9.18 | 1.756 | 36.12 | 21.66 | True | True | True | True | Sex-specific 1e-2 VUMC, sex-differential UKB |
| 0.00368 | 0.00396 | 8.556 | 1.85 | 36.62 | 22.16 | True | True | True | True | Sex-specific 1e-2 VUMC, sex-differential UKB |
| 0.003508 | 0.003773 | 6.34 | 1.504 | 33.82 | 16.11 | True | True | True | True | Sex-specific 1e-2 VUMC, sex-differential UKB |

| SE_UKB_female | SE_UKB_male | LOG10P_VUMC_female | LOG10P_VUMC_male | LOG10P_UKB_female | LOG10P_UKB_male | female_effect_direction_replication | male_effect_direction_replication | sex_effect_comparison_replication | pvalue_effect_comparison_replication | replication_type |
|---|---|---|---|---|---|---|---|---|---|---|
| 0.00368 | 0.00396 | 8.549 | 1.853 | 36.68 | 22.15 | True | True | True | True | Sex-specific 1e-2 VUMC, sex-differential UKB |
| 0.003386 | 0.003646 | 5.151 | 1.327 | 38.7 | 17.79 | True | True | True | True | Sex-specific 1e-2 VUMC, sex-differential UKB |
| 0.003385 | 0.003645 | 5.005 | 1.372 | 38.98 | 17.77 | True | True | True | True | Sex-specific 1e-2 VUMC, sex-differential UKB |
| 0.003385 | 0.003645 | 5.304 | 1.31 | 39.86 | 18.39 | True | True | True | True | Sex-specific 1e-2 VUMC, sex-differential UKB |
| SE_UKB_female | SE_UKB_male | LOG10P_VUMC_female | LOG10P_VUMC_male | LOG10P_UKB_female | LOG10P_UKB_male | female_effect_direction_replication | male_effect_direction_replication | sex_effect_comparison_replication | pvalue_effect_comparison_replication | replication_type |
| 0.003675 | 0.003955 | 8.48 | 1.75 | 35.94 | 22.12 | True | True | True | True | Sex-specific 1e-2 VUMC, sex-differential UKB |
| 0.003675 | 0.003955 | 8.472 | 1.75 | 35.93 | 22.14 | True | True | True | True | Sex-specific 1e-2 VUMC, sex-differential UKB |
| 0.003694 | 0.003976 | 8.296 | 1.753 | 35.29 | 21.8 | True | True | True | True | Sex-specific 1e-2 VUMC, sex-differential UKB |
| 0.003696 | 0.003979 | 8.255 | 1.761 | 35.21 | 21.77 | True | True | True | True | Sex-specific 1e-2 VUMC, sex-differential UKB |
| 0.003503 | 0.003774 | 6.755 | 1.659 | 41.52 | 23.67 | True | True | True | True | Sex-specific 1e-2 VUMC, sex-differential UKB |
| 0.00368 | 0.003959 | 8.685 | 1.83 | 37.04 | 22.37 | True | True | True | True | Sex-specific 1e-2 VUMC, sex-differential UKB |
| 0.003386 | 0.003646 | 5.325 | 1.289 | 39.93 | 18.34 | True | True | True | True | Sex-specific 1e-2 VUMC, sex-differential UKB |
| 0.00365 | 0.003927 | 8.194 | 1.654 | 34.84 | 20.28 | True | True | True | True | Sex-specific 1e-2 VUMC, sex-differential UKB |
| 0.003649 | 0.003926 | 8.197 | 1.654 | 34.93 | 20.07 | True | True | True | True | Sex-specific 1e-2 VUMC, sex-differential UKB |
| 0.003662 | 0.003939 | 7.86 | 1.606 | 34.84 | 19.99 | True | True | True | True | Sex-specific 1e-2 VUMC, sex-differential UKB |
| 0.003628 | 0.003898 | 8.024 | 1.494 | 35.09 | 20.13 | True | True | True | True | Sex-specific 1e-2 VUMC, sex-differential UKB |
| 0.003657 | 0.003934 | 8.091 | 1.732 | 35.87 | 20.5 | True | True | True | True | Sex-specific 1e-2 VUMC, sex-differential UKB |
| 0.00366 | 0.003938 | 7.912 | 1.725 | 35.94 | 20.55 | True | True | True | True | Sex-specific 1e-2 VUMC, sex-differential UKB |
| 0.003661 | 0.003939 | 7.922 | 1.717 | 35.93 | 20.56 | True | True | True | True | Sex-specific 1e-2 VUMC, sex-differential UKB |
| 0.003405 | 0.003672 | 8.51 | 1.183 | 31.59 | 18.92 | True | True | True | True | Sex-specific 1e-2 VUMC, sex-differential UKB |
| 0.003412 | 0.003679 | 8.538 | 0.8564 | 27.84 | 17.72 | True | True | True | True | Sex-specific 1e-2 VUMC, sex-differential UKB |
| 0.003637 | 0.003929 | 7.34 | 1.091 | 11.93 | 4.025 | True | True | True | True | Sex-specific 1e-2 VUMC, sex-specific 5e-8 UKB |
| 0.003636 | 0.003929 | 7.603 | 1.063 | 12.16 | 3.959 | True | True | True | True | Sex-specific 1e-2 VUMC, sex-specific 5e-8 UKB |
| 0.003636 | 0.003929 | 7.777 | 1.011 | 12.08 | 3.861 | True | True | True | True | Sex-specific 1e-2 VUMC, sex-specific 5e-8 UKB |
| 0.004141 | 0.004467 | 8.067 | 0.6971 | 16.59 | 9.177 | True | True | True | True | Sex-specific 1e-2 VUMC, sex-differential UKB |
| 0.003636 | 0.003929 | 7.7 | 1.094 | 12.23 | 3.915 | True | True | True | True | Sex-specific 1e-2 VUMC, sex-specific 5e-8 UKB |

| SE_UKB_female | SE_UKB_male | LOG10P_VUMC_female | LOG10P_VUMC_male | LOG10P_UKB_female | LOG10P_UKB_male | female_effect_direction_replication | male_effect_direction_replication | sex_effect_comparison_replication | pvalue_effect_comparison_replication | replication_type |
|---|---|---|---|---|---|---|---|---|---|---|
| 0.004143 | 0.004469 | 8.046 | 0.7019 | 16.55 | 9.147 | True | True | True | True | Sex-specific 1e-2 VUMC, sex-differential UKB |
| 0.01787 | 0.01988 | 1.012 | 4.962 | 0.267 | 0.2219 | False | True | False | False | |
| 0.01785 | 0.01985 | 1.043 | 4.839 | 0.3248 | 0.1916 | False | True | False | False | |
| 0.01776 | 0.01979 | 1.238 | 5.443 | 0.2771 | 0.2709 | False | True | True | False | |
| 0.01782 | 0.01987 | 1.04 | 4.834 | 0.2473 | 0.2259 | False | True | True | False | |
| 0.01774 | 0.01978 | 1.252 | 5.236 | 0.2964 | 0.262 | False | True | True | False | |
| 0.023 | 0.02561 | 0.2537 | 7.207 | 0.2775 | 0.8326 | False | True | True | True | Sex-specific 1e-2 VUMC, sex-differential UKB |
| 0.02299 | 0.02558 | 0.1881 | 6.936 | 0.301 | 0.7729 | False | True | True | True | Sex-specific 1e-2 VUMC, sex-differential UKB |
| 0.023 | 0.02557 | 0.2385 | 7.252 | 0.2854 | 0.8288 | False | True | True | True | Sex-specific 1e-2 VUMC, sex-differential UKB |
| 0.02422 | 0.02707 | 0.1849 | 7.457 | 0.2207 | 0.6422 | False | True | True | True | Sex-specific 1e-2 VUMC, sex-differential UKB |
| 0.02302 | 0.02555 | 0.2446 | 7.366 | 0.2791 | 0.9276 | False | True | True | True | Sex-specific 1e-2 VUMC, sex-differential UKB |
| 0.02302 | 0.02555 | 0.2446 | 7.366 | 0.2794 | 0.9283 | False | True | True | True | Sex-specific 1e-2 VUMC, sex-differential UKB |
| 0.02306 | 0.0256 | 0.2465 | 7.428 | 0.2724 | 0.9627 | False | True | True | True | Sex-specific 1e-2 VUMC, sex-differential UKB |
| 0.02304 | 0.02558 | 0.2584 | 7.435 | 0.2784 | 0.9573 | False | True | True | True | Sex-specific 1e-2 VUMC, sex-differential UKB |
| 0.02302 | 0.02555 | 0.2576 | 7.424 | 0.2805 | 0.9217 | False | True | True | True | Sex-specific 1e-2 VUMC, sex-differential UKB |
| 0.02302 | 0.02555 | 0.2576 | 7.424 | 0.2801 | 0.9235 | False | True | True | True | Sex-specific 1e-2 VUMC, sex-differential UKB |
| 0.02302 | 0.02555 | 0.2576 | 7.424 | 0.2801 | 0.9235 | False | True | True | True | Sex-specific 1e-2 VUMC, sex-differential UKB |
| 0.02303 | 0.02554 | 0.258 | 7.423 | 0.2905 | 0.9433 | False | True | True | True | Sex-specific 1e-2 VUMC, sex-differential UKB |
| 0.02304 | 0.02555 | 0.2593 | 7.57 | 0.2877 | 0.9603 | False | True | True | True | Sex-specific 1e-2 VUMC, sex-differential UKB |
| 0.02304 | 0.02555 | 0.2664 | 7.595 | 0.2915 | 0.9584 | False | True | True | True | Sex-specific 1e-2 VUMC, sex-differential UKB |
| 0.02304 | 0.02555 | 0.2631 | 7.565 | 0.2967 | 0.9612 | False | True | True | True | Sex-specific 1e-2 VUMC, sex-differential UKB |
| 0.02304 | 0.02555 | 0.2664 | 7.595 | 0.2972 | 0.9654 | False | True | True | True | Sex-specific 1e-2 VUMC, sex-differential UKB |
| 0.02305 | 0.02555 | 0.2667 | 7.596 | 0.3081 | 0.9765 | False | True | True | True | Sex-specific 1e-2 VUMC, sex-differential UKB |
| 0.02305 | 0.02555 | 0.2667 | 7.24 | 0.3007 | 0.9805 | False | True | True | True | Sex-specific 1e-2 VUMC, sex-differential UKB |
| 0.02322 | 0.0257 | 0.1484 | 6.54 | 0.3078 | 0.8847 | False | True | True | True | Sex-specific 1e-2 VUMC, sex-differential UKB |
| 0.02306 | 0.02555 | 0.2665 | 7.213 | 0.3131 | 0.9951 | False | True | True | True | Sex-specific 1e-2 VUMC, sex-differential UKB |
| 0.02306 | 0.02555 | 0.2665 | 7.213 | 0.3132 | 0.9972 | False | True | True | True | Sex-specific 1e-2 VUMC, sex-differential UKB |
| 0.02306 | 0.02555 | 0.2665 | 7.213 | 0.3134 | 0.9968 | False | True | True | True | Sex-specific 1e-2 VUMC, sex-differential UKB |

| SE_UKB_female | SE_UKB_male | LOG10P_VUMC_female | LOG10P_VUMC_male | LOG10P_UKB_female | LOG10P_UKB_male | female_effect_direction_replication | male_effect_direction_replication | sex_effect_comparison_replication | pvalue_effect_comparison_replication | replication_type |
|---|---|---|---|---|---|---|---|---|---|---|
| 0.02321 | 0.02568 | 0.1641 | 6.525 | 0.2893 | 0.9265 | False | True | True | True | Sex-specific 1e-2 VUMC, sex-differential UKB |
| 0.02321 | 0.02569 | 0.153 | 6.5 | 0.2995 | 0.9229 | False | True | True | True | Sex-specific 1e-2 VUMC, sex-differential UKB |
| 0.02305 | 0.02555 | 0.2645 | 7.18 | 0.3098 | 0.9938 | False | True | True | True | Sex-specific 1e-2 VUMC, sex-differential UKB |
| 0.02307 | 0.02554 | 0.2633 | 7.172 | 0.3196 | 0.9982 | False | True | True | True | Sex-specific 1e-2 VUMC, sex-differential UKB |
| 0.02306 | 0.02553 | 0.2428 | 7.164 | 0.3135 | 0.9898 | False | True | True | True | Sex-specific 1e-2 VUMC, sex-differential UKB |
| 0.02309 | 0.02557 | 0.1692 | 6.85 | 0.3151 | 0.8961 | False | True | True | True | Sex-specific 1e-2 VUMC, sex-differential UKB |
| 0.02305 | 0.02552 | 0.2427 | 7.164 | 0.3108 | 0.9893 | False | True | True | True | Sex-specific 1e-2 VUMC, sex-differential UKB |
| 0.003401 | 0.003675 | 8.235 | 1.642 | 24.68 | 20.85 | True | True | True | True | Sex-specific 1e-2 VUMC, sex-differential UKB |
| 0.003401 | 0.003676 | 8.564 | 1.849 | 25.04 | 20.93 | True | True | True | True | Sex-specific 1e-2 VUMC, sex-differential UKB |
| 0.003223 | 0.003485 | 5.81 | 1.062 | 14.95 | 12.85 | True | True | True | True | Sex-specific 1e-2 VUMC, sex-differential UKB |
| 0.003221 | 0.003484 | 5.784 | 0.996 | 14.86 | 12.32 | True | True | True | True | Sex-specific 1e-2 VUMC, sex-differential UKB |

| SE_UKB_female | SE_UKB_male | LOG10P_VUMC_female | LOG10P_VUMC_male | LOG10P_UKB_female | LOG10P_UKB_male | female_effect_direction_replication | male_effect_direction_replication | sex_effect_comparison_replication | pvalue_effect_comparison_replication | replication_type |
|---|---|---|---|---|---|---|---|---|---|---|
| 0.003221 | 0.003484 | 5.754 | 0.9932 | 14.81 | 12.35 | True | True | True | True | Sex-specific 1e-2 VUMC, sex-differential UKB |
| 0.003221 | 0.003483 | 5.827 | 0.9939 | 14.95 | 12.56 | True | True | True | True | Sex-specific 1e-2 VUMC, sex-differential UKB |
| 0.003221 | 0.003483 | 5.782 | 0.998 | 14.88 | 12.34 | True | True | True | True | Sex-specific 1e-2 VUMC, sex-differential UKB |
| 0.00322 | 0.003483 | 5.77 | 1.009 | 14.84 | 12.32 | True | True | True | True | Sex-specific 1e-2 VUMC, sex-differential UKB |
| 0.00322 | 0.003482 | 5.763 | 0.9953 | 14.88 | 12.25 | True | True | True | True | Sex-specific 1e-2 VUMC, sex-differential UKB |
| 0.003394 | 0.00367 | 8.564 | 1.759 | 24.95 | 20.48 | True | True | True | True | Sex-specific 1e-2 VUMC, sex-differential UKB |
| 0.00322 | 0.003483 | 5.7 | 0.9737 | 14.94 | 12.2 | True | True | True | True | Sex-specific 1e-2 VUMC, sex-differential UKB |
| 0.003395 | 0.003671 | 8.628 | 1.697 | 24.84 | 20.52 | True | True | True | True | Sex-specific 1e-2 VUMC, sex-differential UKB |
| 0.003396 | 0.003672 | 8.587 | 1.709 | 24.91 | 20.55 | True | True | True | True | Sex-specific 1e-2 VUMC, sex-differential UKB |
| 0.003395 | 0.003672 | 8.443 | 1.628 | 24.45 | 19.59 | True | True | True | True | Sex-specific 1e-2 VUMC, sex-differential UKB |
| 0.003227 | 0.003489 | 5.18 | 0.7698 | 15.06 | 11.37 | True | True | True | True | Sex-specific 1e-2 VUMC, sex-differential UKB |
| 0.003672 | 0.003958 | 0.8581 | 7.561 | 0.2407 | 6.958 | True | True | True | True | Sex-specific 1e-2 VUMC, sex-differential UKB |
| 0.003673 | 0.003958 | 0.848 | 7.533 | 0.2368 | 6.971 | True | True | True | True | Sex-specific 1e-2 VUMC, sex-differential UKB |
| 0.003748 | 0.004041 | 0.74 | 6.046 | 0.03048 | 5.677 | False | True | True | True | Sex-specific 1e-2 VUMC, sex-differential UKB |

| SE_UKB_female | SE_UKB_male | LOG10P_VUMC_female | LOG10P_VUMC_male | LOG10P_UKB_female | LOG10P_UKB_male | female_effect_direction_replication | male_effect_direction_replication | sex_effect_comparison_replication | pvalue_effect_comparison_replication | replication_type |
|---|---|---|---|---|---|---|---|---|---|---|
| 0.003748 | 0.004041 | 0.7448 | 6.046 | 0.02943 | 5.694 | False | True | True | True | Sex-specific 1e-2 VUMC, sex-differential UKB |
| 0.003749 | 0.004043 | 0.7475 | 5.952 | 0.04404 | 5.791 | False | True | True | True | Sex-specific 1e-2 VUMC, sex-differential UKB |
| 0.003673 | 0.003958 | 0.869 | 7.774 | 0.2472 | 6.862 | True | True | True | True | Sex-specific 1e-2 VUMC, sex-differential UKB |
| 0.003749 | 0.004042 | 0.7194 | 6.064 | 0.03637 | 5.656 | False | True | True | True | Sex-specific 1e-2 VUMC, sex-differential UKB |
| 0.003749 | 0.004043 | 0.7216 | 6.065 | 0.03632 | 5.657 | False | True | True | True | Sex-specific 1e-2 VUMC, sex-differential UKB |
| 0.00375 | 0.004044 | 0.7649 | 5.854 | 0.04632 | 5.875 | False | True | True | True | Sex-specific 1e-2 VUMC, sex-differential UKB |
| 0.003751 | 0.004045 | 0.7405 | 5.834 | 0.05236 | 5.822 | False | True | True | True | Sex-specific 1e-2 VUMC, sex-differential UKB |
| 0.003674 | 0.00396 | 0.8179 | 7.895 | 0.2357 | 7.018 | True | True | True | True | Sex-specific 1e-2 VUMC, sex-differential UKB |
| 0.003755 | 0.00405 | 0.7794 | 5.656 | 0.07225 | 5.771 | False | True | True | True | Sex-specific 1e-2 VUMC, sex-differential UKB |
| 0.003753 | 0.004047 | 0.7246 | 5.835 | 0.05319 | 5.668 | False | True | True | True | Sex-specific 1e-2 VUMC, sex-differential UKB |
| 0.003752 | 0.004046 | 0.7236 | 5.907 | 0.03842 | 5.741 | False | True | True | True | Sex-specific 1e-2 VUMC, sex-differential UKB |
| 0.003753 | 0.004048 | 0.7667 | 5.587 | 0.05628 | 5.831 | False | True | True | True | Sex-specific 1e-2 VUMC, sex-differential UKB |
| 0.003753 | 0.004048 | 0.7326 | 5.576 | 0.04769 | 5.82 | False | True | True | True | Sex-specific 1e-2 VUMC, sex-differential UKB |
| 0.003752 | 0.004046 | 0.7527 | 5.757 | 0.04317 | 5.646 | False | True | True | True | Sex-specific 1e-2 VUMC, sex-differential UKB |
| 0.003753 | 0.004047 | 0.758 | 5.74 | 0.04562 | 5.781 | False | True | True | True | Sex-specific 1e-2 VUMC, sex-differential UKB |
| 0.003486 | 0.003769 | 6.651 | 1.425 | 8.815 | 12.94 | True | True | False | False | |
| 0.0037 | 0.004013 | 7.497 | 1.158 | 8.049 | 17.47 | True | True | False | False | |
| 0.003691 | 0.004003 | 7.531 | 1.177 | 8.28 | 17.4 | True | True | False | False | |
| 0.003694 | 0.004005 | 7.449 | 1.115 | 8.802 | 17.3 | True | True | False | False | |
| 0.003693 | 0.004004 | 7.312 | 1.168 | 8.535 | 17.66 | True | True | False | False | |
| 0.003696 | 0.004007 | 7.242 | 1.117 | 8.816 | 17.18 | True | True | False | False | |
| 0.003691 | 0.004003 | 7.571 | 1.153 | 8.44 | 17.77 | True | True | False | False | |
| 0.003692 | 0.004003 | 7.615 | 1.174 | 8.449 | 17.88 | True | True | False | False | |
| 0.003692 | 0.004004 | 7.414 | 1.122 | 8.352 | 17.85 | True | True | False | False | |
| 0.003692 | 0.004004 | 7.378 | 1.12 | 8.347 | 17.85 | True | True | False | False | |
| 0.003694 | 0.004006 | 7.337 | 1.119 | 8.496 | 17.83 | True | True | False | False | |

| SE_UKB_female | SE_UKB_male | LOG10P_VUMC_female | LOG10P_VUMC_male | LOG10P_UKB_female | LOG10P_UKB_male | female_effect_direction_replication | male_effect_direction_replication | sex_effect_comparison_replication | pvalue_effect_comparison_replication | replication_type |
|---|---|---|---|---|---|---|---|---|---|---|
| 0.003695 | 0.004006 | 7.397 | 1.165 | 8.255 | 17.81 | True | True | False | False | |
| 0.003695 | 0.004008 | 7.479 | 1.174 | 8.29 | 17.96 | True | True | False | False | |
| 0.003416 | 0.003687 | 5.853 | 0.4441 | 7.734 | 15.1 | True | True | False | False | |
| 0.003415 | 0.003685 | 5.8 | 0.4904 | 8.115 | 15.53 | True | True | False | False | |
| 0.003427 | 0.003714 | 9.356 | 1.012 | 12.49 | 23.26 | True | True | False | False | |
| 0.003664 | 0.003976 | 7.035 | 1.139 | 8.278 | 17.64 | True | True | False | False | |
| 0.003662 | 0.003974 | 7.322 | 1.029 | 8.072 | 17.73 | True | True | False | False | |
| 0.00366 | 0.003972 | 7.297 | 0.9978 | 8.233 | 17.38 | True | True | False | False | |
| 0.003425 | 0.003711 | 9.763 | 0.921 | 12.32 | 23.22 | True | True | False | False | |
| 0.003427 | 0.003714 | 9.482 | 0.9946 | 12.32 | 23.47 | True | True | False | False | |
| 0.003663 | 0.003975 | 7.266 | 0.9713 | 8.261 | 17.42 | True | True | False | False | |
| 0.003663 | 0.003975 | 7.339 | 0.9798 | 8.252 | 17.43 | True | True | False | False | |
| 0.003662 | 0.003973 | 7.345 | 0.9939 | 8.235 | 17.46 | True | True | False | False | |
| 0.003662 | 0.003973 | 7.331 | 0.9939 | 8.238 | 17.46 | True | True | False | False | |
| 0.00366 | 0.003971 | 7.333 | 1.092 | 8.037 | 17.74 | True | True | False | False | |
| 0.003658 | 0.003969 | 7.557 | 1.012 | 8.035 | 17.69 | True | True | False | False | |
| 0.003658 | 0.003969 | 7.499 | 1.083 | 8.052 | 17.73 | True | True | False | False | |
| 0.003658 | 0.003969 | 7.384 | 1.08 | 8.031 | 17.67 | True | True | False | False | |
| 0.003661 | 0.003972 | 7.474 | 1.074 | 8.095 | 17.52 | True | True | False | False | |
| 0.003665 | 0.003977 | 7.096 | 1.131 | 7.86 | 17.31 | True | True | False | False | |
| 0.003661 | 0.003973 | 7.289 | 1.049 | 8.025 | 17.42 | True | True | False | False | |
| 0.003401 | 0.003676 | 5.767 | 0.6222 | 6.967 | 16.8 | True | True | False | False | |
| 0.003664 | 0.003976 | 7.12 | 1.111 | 7.965 | 17.29 | True | True | False | False | |
| 0.003666 | 0.003978 | 6.889 | 1.155 | 7.713 | 17.45 | True | True | False | False | |
| 0.003398 | 0.003673 | 5.711 | 0.6295 | 6.984 | 16.4 | True | True | False | False | |

| SE_UKB_female | SE_UKB_male | LOG10P_VUMC_female | LOG10P_VUMC_male | LOG10P_UKB_female | LOG10P_UKB_male | female_effect_direction_replication | male_effect_direction_replication | sex_effect_comparison_replication | pvalue_effect_comparison_replication | replication_type |
|---|---|---|---|---|---|---|---|---|---|---|
| 0.003398 | 0.003673 | 5.626 | 0.6448 | 7.057 | 16.43 | True | True | False | False | |
| 0.003403 | 0.00368 | 5.958 | 0.6749 | 6.363 | 16.09 | True | True | False | False | |
| 0.003399 | 0.003674 | 5.603 | 0.7037 | 7.218 | 16.31 | True | True | False | False | |
| 0.003396 | 0.003669 | 5.946 | 0.6705 | 6.912 | 17.28 | True | True | False | False | |
| 0.003396 | 0.00367 | 5.994 | 0.6556 | 6.856 | 17.22 | True | True | False | False | |
| 0.003396 | 0.00367 | 5.828 | 0.6376 | 7.117 | 16.86 | True | True | False | False | |
| 0.003399 | 0.003673 | 5.758 | 0.6531 | 7.153 | 16.83 | True | True | False | False | |
| 0.003403 | 0.003678 | 5.183 | 0.7272 | 7.238 | 16.11 | True | True | False | False | |
| 0.003404 | 0.003683 | 5.265 | 0.5354 | 6.406 | 15.95 | True | True | False | False | |
| 0.003662 | 0.003972 | 7.786 | 0.7553 | 7.757 | 15.84 | True | True | False | False | |
| 0.003405 | 0.003684 | 5.564 | 0.5664 | 6.409 | 16.57 | True | True | False | False | |
| 0.003405 | 0.003684 | 5.47 | 0.513 | 6.328 | 16.61 | True | True | False | False | |
| 0.003414 | 0.003695 | 5.453 | 0.5121 | 6.183 | 16.44 | True | True | False | False | |
| 0.003415 | 0.003696 | 5.414 | 0.4815 | 6.242 | 16.33 | True | True | False | False | |
| 0.003419 | 0.0037 | 5.75 | 0.4896 | 6.147 | 16.61 | True | True | False | False | |
| 0.003426 | 0.003707 | 5.711 | 0.4353 | 6.097 | 16.81 | True | True | False | False | |
| 0.003422 | 0.003703 | 5.58 | 0.4103 | 6.172 | 16.53 | True | True | False | False | |
| 0.003422 | 0.003703 | 5.591 | 0.3662 | 6.106 | 16.8 | True | True | False | False | |
| 0.003431 | 0.003697 | 3.884 | 0.1362 | 2.359 | 0.7891 | True | True | True | True | Sex-specific 1e-2 VUMC, sex-differential UKB |
| 0.003397 | 0.00367 | 3.461 | 0.01258 | 0.8382 | 0.04453 | True | False | True | True | Sex-specific 1e-2 VUMC, sex-differential UKB |
| 0.0034 | 0.003671 | 3.075 | 0.05958 | 0.6976 | 0.007292 | True | False | True | True | Sex-specific 1e-2 VUMC, sex-differential UKB |
| 0.003397 | 0.00367 | 3.52 | 0.07836 | 0.7254 | 0.01954 | True | False | True | True | Sex-specific 1e-2 VUMC, sex-differential UKB |
| 0.003396 | 0.003669 | 3.544 | 0.1218 | 0.8754 | 0.01043 | True | True | True | True | Sex-specific 1e-2 VUMC, sex-differential UKB |
| 0.003396 | 0.003669 | 3.549 | 0.1166 | 0.8678 | 0.009098 | True | True | True | True | Sex-specific 1e-2 VUMC, sex-differential UKB |
| 0.003393 | 0.003667 | 3.624 | 0.1301 | 1.328 | 0.02636 | True | False | True | True | Sex-specific 1e-2 VUMC, sex-differential UKB |
| 0.003393 | 0.003667 | 3.608 | 0.1291 | 1.321 | 0.02433 | True | False | True | True | Sex-specific 1e-2 VUMC, sex-differential UKB |
| 0.003466 | 0.003739 | 4.658 | 0.9871 | 3.362 | 0.1616 | True | False | True | True | Sex-specific 1e-2 VUMC, sex-differential UKB |
| 0.003437 | 0.003707 | 7.331 | 0.1585 | 4.207 | 0.9014 | True | False | True | True | Sex-specific 1e-2 VUMC, sex-differential UKB |

| SE_UKB_female | SE_UKB_male | LOG10P_VUMC_female | LOG10P_VUMC_male | LOG10P_UKB_female | LOG10P_UKB_male | female_effect_direction_replication | male_effect_direction_replication | sex_effect_comparison_replication | pvalue_effect_comparison_replication | replication_type |
|---|---|---|---|---|---|---|---|---|---|---|
| 0.003548 | 0.003812 | 5.038 | 0.05379 | 7.814 | 7.907 | True | True | False | False | |
| 0.003585 | 0.003853 | 5.023 | 0.1233 | 9.862 | 8.174 | True | True | True | True | Sex-specific 1e-2 VUMC, sex-differential UKB |
| 0.003556 | 0.00382 | 4.947 | 0.09176 | 8.056 | 7.716 | True | True | False | True | |
| 0.003587 | 0.003855 | 5.05 | 0.1495 | 9.793 | 8.105 | True | True | True | True | Sex-specific 1e-2 VUMC, sex-differential UKB |
| 0.003831 | 0.004117 | 6.473 | 0.002796 | 13.23 | 8.494 | True | True | True | True | Sex-specific 1e-2 VUMC, sex-differential UKB |
| 0.003561 | 0.003835 | 7.466 | 0.232 | 13.5 | 7.74 | True | False | True | True | Sex-specific 1e-2 VUMC, sex-differential UKB |
| 0.003561 | 0.003836 | 7.23 | 0.352 | 13.46 | 7.675 | True | False | True | True | Sex-specific 1e-2 VUMC, sex-differential UKB |
| 0.003567 | 0.003841 | 7.328 | 0.2567 | 13.36 | 7.513 | True | False | True | True | Sex-specific 1e-2 VUMC, sex-differential UKB |
| 0.003565 | 0.00384 | 7.342 | 0.3269 | 13.39 | 7.339 | True | False | True | True | Sex-specific 1e-2 VUMC, sex-differential UKB |
| 0.003564 | 0.00384 | 7.308 | 0.3013 | 13.33 | 7.382 | True | False | True | True | Sex-specific 1e-2 VUMC, sex-differential UKB |
| 0.003324 | 0.003578 | 5.632 | 0.07009 | 10.7 | 5.816 | True | False | True | True | Sex-specific 1e-2 VUMC, sex-specific 5e-8 UKB |
| 0.003564 | 0.003839 | 7.287 | 0.298 | 13.36 | 7.392 | True | False | True | True | Sex-specific 1e-2 VUMC, sex-differential UKB |
| 0.003564 | 0.003839 | 7.376 | 0.2575 | 13.29 | 7.337 | True | False | True | True | Sex-specific 1e-2 VUMC, sex-differential UKB |
| 0.003324 | 0.003578 | 5.706 | 0.06739 | 10.68 | 5.77 | True | False | True | True | Sex-specific 1e-2 VUMC, sex-specific 5e-8 UKB |
| 0.003313 | 0.003565 | 5.052 | 0.04987 | 10.68 | 5.481 | True | False | True | True | Sex-specific 1e-2 VUMC, sex-specific 5e-8 UKB |
| 0.003312 | 0.003564 | 5.117 | 0.06115 | 10.65 | 5.469 | True | False | True | True | Sex-specific 1e-2 VUMC, sex-specific 5e-8 UKB |
| 0.003312 | 0.003565 | 4.952 | 0.05783 | 10.83 | 5.434 | True | False | True | True | Sex-specific 1e-2 VUMC, sex-specific 5e-8 UKB |
| 0.003312 | 0.003565 | 5.07 | 0.06387 | 10.59 | 5.463 | True | False | True | True | Sex-specific 1e-2 VUMC, sex-specific 5e-8 UKB |
| 0.003312 | 0.003565 | 5.068 | 0.06603 | 10.61 | 5.467 | True | False | True | True | Sex-specific 1e-2 VUMC, sex-specific 5e-8 UKB |
| 0.003312 | 0.003565 | 5.071 | 0.06904 | 10.65 | 5.479 | True | False | True | True | Sex-specific 1e-2 VUMC, sex-specific 5e-8 UKB |
| 0.003491 | 0.003758 | 6.458 | 0.3563 | 10.66 | 6.117 | True | False | True | True | Sex-specific 1e-2 VUMC, sex-specific 5e-8 UKB |
| 0.003314 | 0.003566 | 4.793 | 0.05853 | 10.52 | 5.496 | True | False | True | True | Sex-specific 1e-2 VUMC, sex-specific 5e-8 UKB |
| 0.003537 | 0.003807 | 6.432 | 0.1996 | 13.19 | 6.9 | True | False | True | True | Sex-specific 1e-2 VUMC, sex-specific 5e-8 UKB |
| 0.003491 | 0.003758 | 6.29 | 0.3584 | 10.7 | 6.021 | True | False | True | True | Sex-specific 1e-2 VUMC, sex-specific 5e-8 UKB |
| 0.003314 | 0.003567 | 4.798 | 0.05857 | 10.53 | 5.506 | True | False | True | True | Sex-specific 1e-2 VUMC, sex-specific 5e-8 UKB |
| 0.00349 | 0.003757 | 6.29 | 0.355 | 10.71 | 6.037 | True | False | True | True | Sex-specific 1e-2 VUMC, sex-specific 5e-8 UKB |

| SE_UKB_female | SE_UKB_male | LOG10P_VUMC_female | LOG10P_VUMC_male | LOG10P_UKB_female | LOG10P_UKB_male | female_effect_direction_replication | male_effect_direction_replication | sex_effect_comparison_replication | pvalue_effect_comparison_replication | replication_type |
|---|---|---|---|---|---|---|---|---|---|---|
| 0.00349 | 0.003757 | 6.302 | 0.3568 | 10.74 | 6.019 | True | False | True | True | Sex-specific 1e-2 VUMC, sex-specific 5e-8 UKB |
| 0.003314 | 0.003567 | 4.8 | 0.06556 | 10.58 | 5.511 | True | False | True | True | Sex-specific 1e-2 VUMC, sex-specific 5e-8 UKB |
| 0.00349 | 0.003757 | 6.285 | 0.3604 | 10.74 | 6.03 | True | False | True | True | Sex-specific 1e-2 VUMC, sex-specific 5e-8 UKB |
| 0.00349 | 0.003757 | 6.285 | 0.3604 | 10.74 | 6.03 | True | False | True | True | Sex-specific 1e-2 VUMC, sex-specific 5e-8 UKB |
| 0.003314 | 0.003567 | 4.8 | 0.05633 | 10.59 | 5.511 | True | False | True | True | Sex-specific 1e-2 VUMC, sex-specific 5e-8 UKB |
| 0.00349 | 0.003757 | 6.296 | 0.3657 | 10.76 | 6.028 | True | False | True | True | Sex-specific 1e-2 VUMC, sex-specific 5e-8 UKB |
| 0.00349 | 0.003757 | 6.302 | 0.3565 | 10.74 | 6.038 | True | False | True | True | Sex-specific 1e-2 VUMC, sex-specific 5e-8 UKB |
| 0.003534 | 0.003805 | 6.355 | 0.2569 | 13.23 | 6.886 | True | False | True | True | Sex-specific 1e-2 VUMC, sex-specific 5e-8 UKB |
| 0.00349 | 0.003757 | 6.321 | 0.3615 | 10.76 | 6.035 | True | False | True | True | Sex-specific 1e-2 VUMC, sex-specific 5e-8 UKB |
| 0.00349 | 0.003757 | 6.321 | 0.3613 | 10.76 | 6.036 | True | False | True | True | Sex-specific 1e-2 VUMC, sex-specific 5e-8 UKB |
| 0.00349 | 0.003757 | 6.321 | 0.3615 | 10.76 | 6.035 | True | False | True | True | Sex-specific 1e-2 VUMC, sex-specific 5e-8 UKB |
| 0.00349 | 0.003757 | 6.321 | 0.3615 | 10.76 | 6.037 | True | False | True | True | Sex-specific 1e-2 VUMC, sex-specific 5e-8 UKB |
| 0.003491 | 0.003757 | 6.315 | 0.3649 | 10.79 | 6.076 | True | False | True | True | Sex-specific 1e-2 VUMC, sex-specific 5e-8 UKB |
| 0.00349 | 0.003757 | 6.328 | 0.368 | 10.74 | 6.07 | True | False | True | True | Sex-specific 1e-2 VUMC, sex-specific 5e-8 UKB |
| 0.003314 | 0.003567 | 4.877 | 0.0534 | 10.51 | 5.413 | True | False | True | True | Sex-specific 1e-2 VUMC, sex-specific 5e-8 UKB |
| 0.003534 | 0.003806 | 6.439 | 0.2349 | 13.18 | 6.72 | True | False | True | True | Sex-specific 1e-2 VUMC, sex-specific 5e-8 UKB |
| 0.003491 | 0.003758 | 6.421 | 0.3173 | 10.69 | 5.865 | True | False | True | True | Sex-specific 1e-2 VUMC, sex-specific 5e-8 UKB |
| 0.00374 | 0.004005 | 5.467 | 0.1338 | 10.11 | 4.221 | True | True | True | True | Sex-specific 1e-2 VUMC, sex-specific 5e-8 UKB |
| 0.003739 | 0.004003 | 5.373 | 0.1474 | 10.08 | 4.08 | True | True | True | True | Sex-specific 1e-2 VUMC, sex-specific 5e-8 UKB |
| 0.003737 | 0.004001 | 5.298 | 0.1692 | 10.21 | 4.278 | True | True | True | True | Sex-specific 1e-2 VUMC, sex-specific 5e-8 UKB |
| 0.003736 | 0.004 | 5.288 | 0.1694 | 10.19 | 4.267 | True | True | True | True | Sex-specific 1e-2 VUMC, sex-specific 5e-8 UKB |
| 0.003735 | 0.003999 | 5.203 | 0.1493 | 10.14 | 4.149 | True | True | True | True | Sex-specific 1e-2 VUMC, sex-specific 5e-8 UKB |
| 0.003735 | 0.003999 | 5.214 | 0.1852 | 10.18 | 4.256 | True | True | True | True | Sex-specific 1e-2 VUMC, sex-specific 5e-8 UKB |
| 0.003735 | 0.003999 | 5.204 | 0.1371 | 10.1 | 4.221 | True | True | True | True | Sex-specific 1e-2 VUMC, sex-specific 5e-8 UKB |
| 0.003511 | 0.003771 | 5.596 | 0.2704 | 7.772 | 3.086 | True | False | True | True | Sex-specific 1e-2 VUMC, sex-specific 5e-8 UKB |

| SE_UKB_female | SE_UKB_male | LOG10P_VUMC_female | LOG10P_VUMC_male | LOG10P_UKB_female | LOG10P_UKB_male | female_effect_direction_replication | male_effect_direction_replication | sex_effect_comparison_replication | pvalue_effect_comparison_replication | replication_type |
|---|---|---|---|---|---|---|---|---|---|---|
| 0.003735 | 0.003999 | 5.192 | 0.1177 | 10.15 | 4.092 | True | True | True | True | Sex-specific 1e-2 VUMC, sex-specific 5e-8 UKB |
| 0.003735 | 0.003999 | 5.18 | 0.1361 | 10.13 | 4.115 | True | True | True | True | Sex-specific 1e-2 VUMC, sex-specific 5e-8 UKB |
| 0.003735 | 0.003999 | 5.167 | 0.1343 | 10.12 | 4.122 | True | True | True | True | Sex-specific 1e-2 VUMC, sex-specific 5e-8 UKB |
| 0.003734 | 0.003999 | 5.197 | 0.1338 | 10 | 4.167 | True | True | True | True | Sex-specific 1e-2 VUMC, sex-specific 5e-8 UKB |
| 0.003735 | 0.003999 | 5.219 | 0.1355 | 10.03 | 4.16 | True | True | True | True | Sex-specific 1e-2 VUMC, sex-specific 5e-8 UKB |
| 0.003735 | 0.003999 | 5.209 | 0.1322 | 10 | 4.164 | True | True | True | True | Sex-specific 1e-2 VUMC, sex-specific 5e-8 UKB |
| 0.00352 | 0.003781 | 5.504 | 0.2799 | 7.625 | 3.219 | True | False | True | True | Sex-specific 1e-2 VUMC, sex-specific 5e-8 UKB |
| 0.003521 | 0.003781 | 5.478 | 0.2775 | 7.622 | 3.223 | True | False | True | True | Sex-specific 1e-2 VUMC, sex-specific 5e-8 UKB |
| 0.003546 | 0.003808 | 4.799 | 0.1288 | 6.297 | 1.857 | True | False | True | True | Sex-specific 1e-2 VUMC, sex-differential UKB |
| 0.003598 | 0.003863 | 4.352 | 0.03988 | 8.079 | 2.081 | True | False | True | True | Sex-specific 1e-2 VUMC, sex-specific 5e-8 UKB |
| 0.003597 | 0.003863 | 4.308 | 0.0412 | 8.089 | 2.051 | True | False | True | True | Sex-specific 1e-2 VUMC, sex-specific 5e-8 UKB |
| 0.003597 | 0.003863 | 4.331 | 0.05749 | 8.062 | 2.039 | True | False | True | True | Sex-specific 1e-2 VUMC, sex-specific 5e-8 UKB |
| 0.003302 | 0.003575 | 3.958 | 0.1067 | 10.14 | 8.539 | True | False | True | True | Sex-specific 1e-2 VUMC, sex-differential UKB |
| 0.003302 | 0.003575 | 3.932 | 0.09305 | 10.19 | 8.648 | True | False | True | True | Sex-specific 1e-2 VUMC, sex-differential UKB |
| 0.003302 | 0.003575 | 3.984 | 0.1087 | 10.13 | 8.572 | True | False | True | True | Sex-specific 1e-2 VUMC, sex-differential UKB |
| 0.003302 | 0.003575 | 3.889 | 0.09799 | 10.07 | 8.452 | True | False | True | True | Sex-specific 1e-2 VUMC, sex-differential UKB |
| 0.003301 | 0.003575 | 3.938 | 0.1154 | 10.12 | 8.498 | True | False | True | True | Sex-specific 1e-2 VUMC, sex-differential UKB |
| 0.0033 | 0.003574 | 3.962 | 0.09953 | 10.09 | 8.502 | True | False | True | True | Sex-specific 1e-2 VUMC, sex-differential UKB |
| 0.0033 | 0.003573 | 4.01 | 0.1239 | 10.1 | 8.447 | True | False | True | True | Sex-specific 1e-2 VUMC, sex-differential UKB |
| 0.0033 | 0.003573 | 4.002 | 0.1273 | 10.02 | 8.458 | True | False | True | True | Sex-specific 1e-2 VUMC, sex-differential UKB |
| 0.0033 | 0.003573 | 3.977 | 0.1048 | 10.11 | 8.458 | True | False | True | True | Sex-specific 1e-2 VUMC, sex-differential UKB |
| 0.003305 | 0.003578 | 4.156 | 0.1873 | 10.72 | 7.93 | True | False | True | True | Sex-specific 1e-2 VUMC, sex-differential UKB |
| 0.003302 | 0.003575 | 4.121 | 0.1901 | 10.76 | 7.933 | True | False | True | True | Sex-specific 1e-2 VUMC, sex-differential UKB |
| 0.003302 | 0.003576 | 4.151 | 0.1918 | 10.81 | 7.969 | True | False | True | True | Sex-specific 1e-2 VUMC, sex-differential UKB |
| 0.003305 | 0.003579 | 4.171 | 0.1721 | 10.68 | 7.849 | True | False | True | True | Sex-specific 1e-2 VUMC, sex-differential UKB |
| 0.003305 | 0.003579 | 4.172 | 0.1736 | 10.64 | 7.848 | True | False | True | True | Sex-specific 1e-2 VUMC, sex-differential UKB |

| SE_UKB_female | SE_UKB_male | LOG10P_VUMC_female | LOG10P_VUMC_male | LOG10P_UKB_female | LOG10P_UKB_male | female_effect_direction_replication | male_effect_direction_replication | sex_effect_comparison_replication | pvalue_effect_comparison_replication | replication_type |
|---|---|---|---|---|---|---|---|---|---|---|
| 0.003306 | 0.003581 | 4.108 | 0.1957 | 10.63 | 7.794 | True | False | True | True | Sex-specific 1e-2 VUMC, sex-differential UKB |
| 0.003307 | 0.003581 | 4.124 | 0.2061 | 10.7 | 7.784 | True | False | True | True | Sex-specific 1e-2 VUMC, sex-differential UKB |
| 0.003257 | 0.003527 | 3.332 | 0.6495 | 7.519 | 5.977 | True | False | True | True | Sex-specific 1e-2 VUMC, sex-specific 5e-8 UKB |
| 0.003325 | 0.003601 | 3.972 | 0.1182 | 11.35 | 8.5 | True | False | True | True | Sex-specific 1e-2 VUMC, sex-differential UKB |
| 0.0033 | 0.003574 | 4.19 | 0.205 | 11.84 | 7.642 | True | False | True | True | Sex-specific 1e-2 VUMC, sex-differential UKB |
| 0.003302 | 0.003576 | 4.166 | 0.2143 | 11.52 | 7.698 | True | False | True | True | Sex-specific 1e-2 VUMC, sex-differential UKB |
| 0.003301 | 0.003576 | 4.2 | 0.215 | 11.76 | 7.529 | True | False | True | True | Sex-specific 1e-2 VUMC, sex-differential UKB |
| 0.003301 | 0.003576 | 4.137 | 0.2138 | 11.43 | 7.721 | True | False | True | True | Sex-specific 1e-2 VUMC, sex-differential UKB |
| 0.0033 | 0.003574 | 3.98 | 0.2661 | 11.79 | 7.775 | True | False | True | True | Sex-specific 1e-2 VUMC, sex-differential UKB |
| 0.0033 | 0.003575 | 4.029 | 0.309 | 11.61 | 7.836 | True | False | True | True | Sex-specific 1e-2 VUMC, sex-differential UKB |

| SE_UKB_female | SE_UKB_male | LOG10P_VUMC_female | LOG10P_VUMC_male | LOG10P_UKB_female | LOG10P_UKB_male | female_effect_direction_replication | male_effect_direction_replication | sex_effect_comparison_replication | pvalue_effect_comparison_replication | replication_type |
|---|---|---|---|---|---|---|---|---|---|---|
| 0.003142 | 0.003402 | 7.367 | 0.3588 | 28.93 | 19.94 | True | True | True | True | Sex-specific 1e-2 VUMC, sex-differential UKB |
| 0.003144 | 0.003404 | 7.302 | 0.3157 | 28.99 | 19.68 | True | True | True | True | Sex-specific 1e-2 VUMC, sex-differential UKB |
| 0.003143 | 0.003404 | 7.062 | 0.3125 | 28.62 | 18.88 | True | True | True | True | Sex-specific 1e-2 VUMC, sex-differential UKB |
| 0.003307 | 0.003584 | 4.503 | 0.2899 | 11.33 | 7.807 | True | False | True | True | Sex-specific 1e-2 VUMC, sex-differential UKB |
| 0.003144 | 0.003406 | 7.81 | 0.2908 | 27.35 | 18.34 | True | True | True | True | Sex-specific 1e-2 VUMC, sex-differential UKB |
| 0.003306 | 0.003583 | 4.592 | 0.2672 | 11.31 | 7.709 | True | False | True | True | Sex-specific 1e-2 VUMC, sex-differential UKB |
| 0.003142 | 0.003402 | 7.48 | 0.3378 | 27.96 | 19.01 | True | True | True | True | Sex-specific 1e-2 VUMC, sex-differential UKB |
| 0.003303 | 0.00358 | 4.369 | 0.2668 | 11.93 | 8.157 | True | False | True | True | Sex-specific 1e-2 VUMC, sex-differential UKB |
| 0.003147 | 0.003407 | 7.594 | 0.3331 | 26.91 | 18.51 | True | True | True | True | Sex-specific 1e-2 VUMC, sex-differential UKB |
| 0.003142 | 0.003403 | 7.291 | 0.2797 | 28.47 | 18.54 | True | True | True | True | Sex-specific 1e-2 VUMC, sex-differential UKB |
| 0.003306 | 0.003583 | 4.167 | 0.3023 | 11.31 | 7.841 | True | False | True | True | Sex-specific 1e-2 VUMC, sex-differential UKB |
| 0.003141 | 0.003401 | 7.147 | 0.3347 | 28.03 | 18.95 | True | True | True | True | Sex-specific 1e-2 VUMC, sex-differential UKB |
| 0.003298 | 0.003575 | 4.127 | 0.3526 | 12.04 | 7.95 | True | False | True | True | Sex-specific 1e-2 VUMC, sex-differential UKB |
| 0.003305 | 0.003582 | 4.468 | 0.284 | 11.36 | 7.91 | True | False | True | True | Sex-specific 1e-2 VUMC, sex-differential UKB |
| 0.00314 | 0.003401 | 7.139 | 0.3423 | 28.52 | 18.41 | True | True | True | True | Sex-specific 1e-2 VUMC, sex-differential UKB |

| SE_UKB_female | SE_UKB_male | LOG10P_VUMC_female | LOG10P_VUMC_male | LOG10P_UKB_female | LOG10P_UKB_male | female_effect_direction_replication | male_effect_direction_replication | sex_effect_comparison_replication | pvalue_effect_comparison_replication | replication_type |
|---|---|---|---|---|---|---|---|---|---|---|
| 0.003302 | 0.003579 | 4.121 | 0.3646 | 11.36 | 7.466 | True | False | True | True | Sex-specific 1e-2 VUMC, sex-differential UKB |
| 0.003299 | 0.003576 | 4.107 | 0.3424 | 12.19 | 7.839 | True | False | True | True | Sex-specific 1e-2 VUMC, sex-differential UKB |
| 0.00314 | 0.0034 | 7.116 | 0.3408 | 28.43 | 18.76 | True | True | True | True | Sex-specific 1e-2 VUMC, sex-differential UKB |
| 0.00314 | 0.0034 | 7.087 | 0.3362 | 28.3 | 18.94 | True | True | True | True | Sex-specific 1e-2 VUMC, sex-differential UKB |
| 0.00314 | 0.0034 | 7.075 | 0.3454 | 28.34 | 18.7 | True | True | True | True | Sex-specific 1e-2 VUMC, sex-differential UKB |
| 0.003306 | 0.003583 | 4.37 | 0.3087 | 11.33 | 7.954 | True | False | True | True | Sex-specific 1e-2 VUMC, sex-differential UKB |
| 0.00314 | 0.003401 | 7.075 | 0.3454 | 28.37 | 18.9 | True | True | True | True | Sex-specific 1e-2 VUMC, sex-differential UKB |
| 0.003141 | 0.003403 | 7.31 | 0.2813 | 28.71 | 18.32 | True | True | True | True | Sex-specific 1e-2 VUMC, sex-differential UKB |
| 0.003141 | 0.003401 | 7.186 | 0.346 | 28.33 | 18.74 | True | True | True | True | Sex-specific 1e-2 VUMC, sex-differential UKB |
| 0.003141 | 0.003401 | 7.163 | 0.3451 | 28.36 | 18.69 | True | True | True | True | Sex-specific 1e-2 VUMC, sex-differential UKB |
| 0.003141 | 0.003401 | 7.164 | 0.3453 | 28.3 | 18.7 | True | True | True | True | Sex-specific 1e-2 VUMC, sex-differential UKB |
| 0.003303 | 0.00358 | 4.26 | 0.2431 | 12.12 | 8.184 | True | False | True | True | Sex-specific 1e-2 VUMC, sex-differential UKB |
| 0.003141 | 0.003401 | 7.2 | 0.3344 | 28.25 | 18.93 | True | True | True | True | Sex-specific 1e-2 VUMC, sex-differential UKB |
| 0.003141 | 0.003401 | 7.213 | 0.3424 | 28.32 | 18.75 | True | True | True | True | Sex-specific 1e-2 VUMC, sex-differential UKB |
| 0.003299 | 0.003576 | 4.159 | 0.3488 | 12 | 8.072 | True | False | True | True | Sex-specific 1e-2 VUMC, sex-differential UKB |
| 0.003142 | 0.003404 | 7.4 | 0.263 | 28.6 | 18.56 | True | True | True | True | Sex-specific 1e-2 VUMC, sex-differential UKB |
| 0.003142 | 0.003402 | 7.275 | 0.3262 | 28.26 | 18.97 | True | True | True | True | Sex-specific 1e-2 VUMC, sex-differential UKB |
| 0.003142 | 0.003402 | 7.2 | 0.3311 | 28.2 | 18.98 | True | True | True | True | Sex-specific 1e-2 VUMC, sex-differential UKB |
| 0.003142 | 0.003403 | 7.126 | 0.334 | 28.23 | 19.14 | True | True | True | True | Sex-specific 1e-2 VUMC, sex-differential UKB |
| 0.003309 | 0.003586 | 4.369 | 0.2957 | 11.32 | 7.995 | True | False | True | True | Sex-specific 1e-2 VUMC, sex-differential UKB |
| 0.003309 | 0.003586 | 4.318 | 0.2972 | 11.31 | 7.946 | True | False | True | True | Sex-specific 1e-2 VUMC, sex-differential UKB |
| 0.00331 | 0.003587 | 4.346 | 0.3129 | 11.33 | 7.964 | True | False | True | True | Sex-specific 1e-2 VUMC, sex-differential UKB |
| 0.00331 | 0.003587 | 4.313 | 0.2992 | 11.3 | 7.95 | True | False | True | True | Sex-specific 1e-2 VUMC, sex-differential UKB |
| 0.003144 | 0.003405 | 7.115 | 0.3464 | 28.32 | 19.15 | True | True | True | True | Sex-specific 1e-2 VUMC, sex-differential UKB |
| 0.003144 | 0.003405 | 7.153 | 0.2635 | 28.41 | 19.12 | True | True | True | True | Sex-specific 1e-2 VUMC, sex-differential UKB |
| 0.003144 | 0.003405 | 7.149 | 0.2597 | 28.35 | 19.08 | True | True | True | True | Sex-specific 1e-2 VUMC, sex-differential UKB |

| SE_UKB_female | SE_UKB_male | LOG10P_VUMC_female | LOG10P_VUMC_male | LOG10P_UKB_female | LOG10P_UKB_male | female_effect_direction_replication | male_effect_direction_replication | sex_effect_comparison_replication | pvalue_effect_comparison_replication | replication_type |
|---|---|---|---|---|---|---|---|---|---|---|
| 0.003313 | 0.003591 | 4.459 | 0.4217 | 11.34 | 7.902 | True | False | True | True | Sex-specific 1e-2 VUMC, sex-differential UKB |
| 0.003176 | 0.003439 | 7.081 | 0.1926 | 30.5 | 18.27 | True | True | True | True | Sex-specific 1e-2 VUMC, sex-differential UKB |
| 0.003177 | 0.003439 | 7.131 | 0.2158 | 30.42 | 18.22 | True | True | True | True | Sex-specific 1e-2 VUMC, sex-differential UKB |
| 0.003177 | 0.00344 | 6.929 | 0.2015 | 29.89 | 18.24 | True | True | True | True | Sex-specific 1e-2 VUMC, sex-differential UKB |
| 0.003178 | 0.00344 | 7 | 0.2099 | 30.09 | 18.24 | True | True | True | True | Sex-specific 1e-2 VUMC, sex-differential UKB |
| 0.003177 | 0.003438 | 6.945 | 0.2815 | 29.67 | 18.69 | True | True | True | True | Sex-specific 1e-2 VUMC, sex-differential UKB |
| 0.003177 | 0.003437 | 6.914 | 0.2821 | 29.87 | 18.73 | True | True | True | True | Sex-specific 1e-2 VUMC, sex-differential UKB |
| 0.003412 | 0.003694 | 5.025 | 0.2507 | 14.1 | 9.314 | True | False | True | True | Sex-specific 1e-2 VUMC, sex-differential UKB |
| 0.003949 | 0.00428 | 3.72 | 0.327 | 1.89 | 1.408 | True | True | True | True | Sex-specific 1e-2 VUMC, sex-differential UKB |
| 0.003612 | 0.003917 | 7.055 | 0.07382 | 13.04 | 4.13 | True | True | True | True | Sex-specific 1e-2 VUMC, sex-specific 5e-8 UKB |
| 0.003612 | 0.003917 | 7.221 | 0.0565 | 12.99 | 4.237 | True | True | True | True | Sex-specific 1e-2 VUMC, sex-specific 5e-8 UKB |
| 0.004318 | 0.004674 | 6.53 | 0.07606 | 3.599 | 1.275 | True | True | True | True | Sex-specific 1e-2 VUMC, sex-differential UKB |
| 0.004346 | 0.004707 | 6.826 | 0.109 | 3.279 | 1.105 | True | True | True | True | Sex-specific 1e-2 VUMC, sex-differential UKB |
| 0.003661 | 0.003968 | 7.148 | 0.1269 | 13.54 | 3.854 | True | True | True | True | Sex-specific 1e-2 VUMC, sex-specific 5e-8 UKB |
| 0.004353 | 0.004717 | 6.765 | 0.1128 | 3.126 | 0.9587 | True | True | True | True | Sex-specific 1e-2 VUMC, sex-differential UKB |
| 0.003643 | 0.003949 | 6.594 | 0.1526 | 13.79 | 4.322 | True | True | True | True | Sex-specific 1e-2 VUMC, sex-specific 5e-8 UKB |
| 0.00364 | 0.003945 | 6.594 | 0.1255 | 13.51 | 4.284 | True | True | True | True | Sex-specific 1e-2 VUMC, sex-specific 5e-8 UKB |
| 0.003914 | 0.004242 | 6.217 | 0.211 | 10.28 | 3.521 | True | True | True | True | Sex-specific 1e-2 VUMC, sex-specific 5e-8 UKB |
| 0.00364 | 0.003944 | 6.499 | 0.1596 | 13.44 | 4.187 | True | True | True | True | Sex-specific 1e-2 VUMC, sex-specific 5e-8 UKB |
| SE_UKB_female | SE_UKB_male | LOG10P_VUMC_female | LOG10P_VUMC_male | LOG10P_UKB_female | LOG10P_UKB_male | female_effect_direction_replication | male_effect_direction_replication | sex_effect_comparison_replication | pvalue_effect_comparison_replication | replication_type |
| 0.00364 | 0.003944 | 6.589 | 0.1409 | 13.46 | 4.189 | True | True | True | True | Sex-specific 1e-2 VUMC, sex-specific 5e-8 UKB |
| 0.003912 | 0.00424 | 6.192 | 0.2136 | 10.19 | 3.418 | True | True | True | True | Sex-specific 1e-2 VUMC, sex-specific 5e-8 UKB |
| 0.003565 | 0.003866 | 5.567 | 0.09418 | 12.79 | 5.27 | True | True | True | True | Sex-specific 1e-2 VUMC, sex-specific 5e-8 UKB |
| 0.00363 | 0.003933 | 6.2 | 0.1253 | 13.61 | 4.475 | True | True | True | True | Sex-specific 1e-2 VUMC, sex-specific 5e-8 UKB |
| 0.003629 | 0.003932 | 6.128 | 0.1183 | 13.6 | 4.482 | True | True | True | True | Sex-specific 1e-2 VUMC, sex-specific 5e-8 UKB |
| 0.003556 | 0.003856 | 5.2 | 0.06954 | 12.97 | 5.598 | True | True | True | True | Sex-specific 1e-2 VUMC, sex-specific 5e-8 UKB |

| SE_UKB_female | SE_UKB_male | LOG10P_VUMC_female | LOG10P_VUMC_male | LOG10P_UKB_female | LOG10P_UKB_male | female_effect_direction_replication | male_effect_direction_replication | sex_effect_comparison_replication | pvalue_effect_comparison_replication | replication_type |
|---|---|---|---|---|---|---|---|---|---|---|
| 0.00363 | 0.003933 | 6.049 | 0.1195 | 13.54 | 4.459 | True | True | True | True | Sex-specific 1e-2 VUMC, sex-specific 5e-8 UKB |
| 0.003554 | 0.003853 | 5.257 | 0.07571 | 13.25 | 5.744 | True | True | True | True | Sex-specific 1e-2 VUMC, sex-specific 5e-8 UKB |
| 0.003895 | 0.004223 | 5.84 | 0.2558 | 10.48 | 3.782 | True | True | True | True | Sex-specific 1e-2 VUMC, sex-specific 5e-8 UKB |
| 0.003896 | 0.004223 | 5.861 | 0.2552 | 10.48 | 3.832 | True | True | True | True | Sex-specific 1e-2 VUMC, sex-specific 5e-8 UKB |
| 0.003896 | 0.004224 | 5.853 | 0.2576 | 10.53 | 3.884 | True | True | True | True | Sex-specific 1e-2 VUMC, sex-specific 5e-8 UKB |
| 0.003896 | 0.004223 | 5.836 | 0.2552 | 10.5 | 3.858 | True | True | True | True | Sex-specific 1e-2 VUMC, sex-specific 5e-8 UKB |
| 0.003814 | 0.004136 | 5.353 | 0.2946 | 11.55 | 4.246 | True | True | True | True | Sex-specific 1e-2 VUMC, sex-specific 5e-8 UKB |
| 0.003896 | 0.004224 | 5.848 | 0.2559 | 10.4 | 3.805 | True | True | True | True | Sex-specific 1e-2 VUMC, sex-specific 5e-8 UKB |
| 0.003945 | 0.004276 | 5.826 | 0.1988 | 10.53 | 3.619 | True | True | True | True | Sex-specific 1e-2 VUMC, sex-specific 5e-8 UKB |
| 0.003896 | 0.004224 | 5.818 | 0.2746 | 10.6 | 3.832 | True | True | True | True | Sex-specific 1e-2 VUMC, sex-specific 5e-8 UKB |
| 0.003897 | 0.004224 | 5.82 | 0.2748 | 10.6 | 3.863 | True | True | True | True | Sex-specific 1e-2 VUMC, sex-specific 5e-8 UKB |
| 0.003815 | 0.004137 | 5.289 | 0.3337 | 11.61 | 4.235 | True | True | True | True | Sex-specific 1e-2 VUMC, sex-specific 5e-8 UKB |
| 0.003895 | 0.004221 | 5.835 | 0.3526 | 10.77 | 3.934 | True | True | True | True | Sex-specific 1e-2 VUMC, sex-specific 5e-8 UKB |
| 0.003895 | 0.004221 | 5.831 | 0.3498 | 10.76 | 3.923 | True | True | True | True | Sex-specific 1e-2 VUMC, sex-specific 5e-8 UKB |
| 0.003745 | 0.004058 | 7.444 | 1.205 | 17.35 | 6.309 | True | True | True | True | Sex-specific 1e-2 VUMC, sex-specific 5e-8 UKB |
| 0.003745 | 0.004058 | 7.495 | 1.248 | 17.33 | 6.31 | True | True | True | True | Sex-specific 1e-2 VUMC, sex-specific 5e-8 UKB |
| 0.003745 | 0.004058 | 7.482 | 1.243 | 17.44 | 6.372 | True | True | True | True | Sex-specific 1e-2 VUMC, sex-specific 5e-8 UKB |
| 0.003889 | 0.004206 | 7.856 | 0.7503 | 14.91 | 4.928 | True | True | True | True | Sex-specific 1e-2 VUMC, sex-specific 5e-8 UKB |
| 0.003887 | 0.004204 | 7.818 | 0.7422 | 14.83 | 4.974 | True | True | True | True | Sex-specific 1e-2 VUMC, sex-specific 5e-8 UKB |
| 0.003975 | 0.004296 | 8.725 | 0.6613 | 13.94 | 4.893 | True | True | True | True | Sex-specific 1e-2 VUMC, sex-specific 5e-8 UKB |
| 0.003976 | 0.004297 | 8.714 | 0.6644 | 13.91 | 4.751 | True | True | True | True | Sex-specific 1e-2 VUMC, sex-specific 5e-8 UKB |
| 0.003744 | 0.004057 | 7.523 | 1.206 | 17.54 | 6.44 | True | True | True | True | Sex-specific 1e-2 VUMC, sex-specific 5e-8 UKB |
| 0.003744 | 0.004057 | 7.377 | 1.219 | 17.61 | 6.2 | True | True | True | True | Sex-specific 1e-2 VUMC, sex-specific 5e-8 UKB |
| 0.003745 | 0.004057 | 7.459 | 1.259 | 17.58 | 6.201 | True | True | True | True | Sex-specific 1e-2 VUMC, sex-specific 5e-8 UKB |
| 0.003745 | 0.004057 | 7.47 | 1.26 | 17.57 | 6.217 | True | True | True | True | Sex-specific 1e-2 VUMC, sex-specific 5e-8 UKB |
| 0.003745 | 0.004057 | 7.471 | 1.255 | 17.55 | 6.222 | True | True | True | True | Sex-specific 1e-2 VUMC, sex-specific 5e-8 UKB |

| SE_UKB_female | SE_UKB_male | LOG10P_VUMC_female | LOG10P_VUMC_male | LOG10P_UKB_female | LOG10P_UKB_male | female_effect_direction_replication | male_effect_direction_replication | sex_effect_comparison_replication | pvalue_effect_comparison_replication | replication_type |
|---|---|---|---|---|---|---|---|---|---|---|
| 0.003884 | 0.004201 | 7.933 | 0.7528 | 15.08 | 4.852 | True | True | True | True | Sex-specific 1e-2 VUMC, sex-specific 5e-8 UKB |
| 0.003975 | 0.004296 | 8.603 | 0.6541 | 14.06 | 4.636 | True | True | True | True | Sex-specific 1e-2 VUMC, sex-specific 5e-8 UKB |
| 0.004544 | 0.004911 | 8.808 | 0.6538 | 4.324 | 2.011 | True | True | True | True | Sex-specific 1e-2 VUMC, sex-differential UKB |
| 0.003984 | 0.004306 | 8.097 | 0.5561 | 14.8 | 4.841 | True | True | True | True | Sex-specific 1e-2 VUMC, sex-specific 5e-8 UKB |
| 0.003521 | 0.003817 | 7.577 | 0.9231 | 22.52 | 6.542 | True | True | True | True | Sex-specific 1e-2 VUMC, sex-specific 5e-8 UKB |
| 0.003989 | 0.004311 | 8.298 | 0.5591 | 15.01 | 4.834 | True | True | True | True | Sex-specific 1e-2 VUMC, sex-specific 5e-8 UKB |
| 0.003988 | 0.004311 | 8.339 | 0.5758 | 15 | 4.863 | True | True | True | True | Sex-specific 1e-2 VUMC, sex-specific 5e-8 UKB |
| 0.00352 | 0.003815 | 7.122 | 0.7663 | 22.48 | 6.345 | True | True | True | True | Sex-specific 1e-2 VUMC, sex-specific 5e-8 UKB |
| 0.003896 | 0.004207 | 7.043 | 0.3033 | 16.7 | 5.145 | True | True | True | True | Sex-specific 1e-2 VUMC, sex-specific 5e-8 UKB |
| 0.003671 | 0.003982 | 6.161 | 1.066 | 19.93 | 6.134 | True | True | True | True | Sex-specific 1e-2 VUMC, sex-specific 5e-8 UKB |
| 0.004173 | 0.00451 | 6.542 | 0.535 | 14.57 | 4.869 | True | True | True | True | Sex-specific 1e-2 VUMC, sex-specific 5e-8 UKB |
| 0.004169 | 0.004505 | 6.243 | 0.5006 | 14.35 | 4.825 | True | True | True | True | Sex-specific 1e-2 VUMC, sex-specific 5e-8 UKB |
| 0.004227 | 0.00457 | 6.011 | 0.5077 | 14.15 | 5.214 | True | True | True | True | Sex-specific 1e-2 VUMC, sex-specific 5e-8 UKB |
| 0.004174 | 0.00451 | 6.493 | 0.5337 | 14.48 | 4.908 | True | True | True | True | Sex-specific 1e-2 VUMC, sex-specific 5e-8 UKB |
| 0.004169 | 0.004505 | 6.237 | 0.4569 | 14.32 | 4.854 | True | True | True | True | Sex-specific 1e-2 VUMC, sex-specific 5e-8 UKB |
| 0.004171 | 0.004507 | 6.497 | 0.5169 | 14.59 | 4.87 | True | True | True | True | Sex-specific 1e-2 VUMC, sex-specific 5e-8 UKB |
| 0.004172 | 0.004508 | 6.365 | 0.4834 | 14.65 | 4.878 | True | True | True | True | Sex-specific 1e-2 VUMC, sex-specific 5e-8 UKB |
| 0.004171 | 0.004507 | 6.406 | 0.4969 | 14.62 | 4.855 | True | True | True | True | Sex-specific 1e-2 VUMC, sex-specific 5e-8 UKB |
| 0.004171 | 0.004507 | 6.406 | 0.4968 | 14.64 | 4.859 | True | True | True | True | Sex-specific 1e-2 VUMC, sex-specific 5e-8 UKB |
| 0.004816 | 0.005206 | 7.103 | 0.5444 | 4.171 | 1.783 | True | True | True | True | Sex-specific 1e-2 VUMC, sex-differential UKB |
| | | 6.64 | 0.4574 | | | False | False | False | False | |
| 0.004816 | 0.005206 | 7.042 | 0.5262 | 4.177 | 1.785 | True | True | True | True | Sex-specific 1e-2 VUMC, sex-differential UKB |
| 0.004826 | 0.005218 | 7.288 | 0.5362 | 4.05 | 1.845 | True | True | True | True | Sex-specific 1e-2 VUMC, sex-differential UKB |
| 0.003876 | 0.004205 | 5.879 | 1.02 | 18.41 | 6.286 | True | True | True | True | Sex-specific 1e-2 VUMC, sex-specific 5e-8 UKB |
| 0.004826 | 0.005218 | 7.199 | 0.533 | 4.061 | 1.848 | True | True | True | True | Sex-specific 1e-2 VUMC, sex-differential UKB |
| 0.004817 | 0.005207 | 6.93 | 0.5198 | 4.172 | 1.842 | True | True | True | True | Sex-specific 1e-2 VUMC, sex-differential UKB |

| SE_UKB_female | SE_UKB_male | LOG10P_VUMC_female | LOG10P_VUMC_male | LOG10P_UKB_female | LOG10P_UKB_male | female_effect_direction_replication | male_effect_direction_replication | sex_effect_comparison_replication | pvalue_effect_comparison_replication | replication_type |
|---|---|---|---|---|---|---|---|---|---|---|
| 0.004822 | 0.005213 | 6.945 | 0.5344 | 4.177 | 1.785 | True | True | True | True | Sex-specific 1e-2 VUMC, sex-differential UKB |
| 0.004823 | 0.005213 | 6.979 | 0.5469 | 4.193 | 1.765 | True | True | True | True | Sex-specific 1e-2 VUMC, sex-differential UKB |
| SE_UKB_female | SE_UKB_male | LOG10P_VUMC_female | LOG10P_VUMC_male | LOG10P_UKB_female | LOG10P_UKB_male | female_effect_direction_replication | male_effect_direction_replication | sex_effect_comparison_replication | pvalue_effect_comparison_replication | replication_type |
| 0.004859 | 0.005254 | 6.727 | 0.5874 | 4.012 | 1.832 | True | True | True | True | Sex-specific 1e-2 VUMC, sex-differential UKB |
| 0.004832 | 0.005224 | 7.142 | 0.5565 | 4.119 | 1.774 | True | True | True | True | Sex-specific 1e-2 VUMC, sex-differential UKB |
| 0.004832 | 0.005224 | 7.198 | 0.558 | 4.12 | 1.774 | True | True | True | True | Sex-specific 1e-2 VUMC, sex-differential UKB |
| 0.003871 | 0.004199 | 5.333 | 0.963 | 18.25 | 5.911 | True | True | True | True | Sex-specific 1e-2 VUMC, sex-specific 5e-8 UKB |
| 0.004822 | 0.005214 | 6.863 | 0.5148 | 4.246 | 1.752 | True | True | True | True | Sex-specific 1e-2 VUMC, sex-differential UKB |
| 0.004822 | 0.005214 | 6.867 | 0.5339 | 4.231 | 1.751 | True | True | True | True | Sex-specific 1e-2 VUMC, sex-differential UKB |
| 0.004822 | 0.005214 | 6.876 | 0.5497 | 4.22 | 1.743 | True | True | True | True | Sex-specific 1e-2 VUMC, sex-differential UKB |
| 0.004822 | 0.005214 | 6.859 | 0.5208 | 4.174 | 1.766 | True | True | True | True | Sex-specific 1e-2 VUMC, sex-differential UKB |
| 0.004822 | 0.005214 | 6.859 | 0.5203 | 4.204 | 1.746 | True | True | True | True | Sex-specific 1e-2 VUMC, sex-differential UKB |
| 0.004822 | 0.005214 | 6.864 | 0.5189 | 4.163 | 1.766 | True | True | True | True | Sex-specific 1e-2 VUMC, sex-differential UKB |
| 0.004821 | 0.005213 | 6.79 | 0.5401 | 4.258 | 1.722 | True | True | True | True | Sex-specific 1e-2 VUMC, sex-differential UKB |
| 0.004822 | 0.005214 | 6.905 | 0.5392 | 4.169 | 1.752 | True | True | True | True | Sex-specific 1e-2 VUMC, sex-differential UKB |
| 0.003868 | 0.004198 | 5.263 | 0.9601 | 18.24 | 5.978 | True | True | True | True | Sex-specific 1e-2 VUMC, sex-specific 5e-8 UKB |
| 0.004822 | 0.005214 | 6.898 | 0.5534 | 4.205 | 1.742 | True | True | True | True | Sex-specific 1e-2 VUMC, sex-differential UKB |
| 0.003869 | 0.004198 | 5.21 | 0.9753 | 18.2 | 6.033 | True | True | True | True | Sex-specific 1e-2 VUMC, sex-specific 5e-8 UKB |
| 0.004831 | 0.005224 | 7.078 | 0.5842 | 4.168 | 1.889 | True | True | True | True | Sex-specific 1e-2 VUMC, sex-differential UKB |
| 0.004831 | 0.005224 | 7.126 | 0.5848 | 4.166 | 1.9 | True | True | True | True | Sex-specific 1e-2 VUMC, sex-differential UKB |
| 0.004831 | 0.005224 | 7.121 | 0.586 | 4.169 | 1.888 | True | True | True | True | Sex-specific 1e-2 VUMC, sex-differential UKB |
| 0.004831 | 0.005224 | 7.131 | 0.585 | 4.167 | 1.888 | True | True | True | True | Sex-specific 1e-2 VUMC, sex-differential UKB |
| 0.004874 | 0.005265 | 6.88 | 0.5098 | 4.053 | 1.888 | True | True | True | True | Sex-specific 1e-2 VUMC, sex-differential UKB |
| 0.004867 | 0.005258 | 7.05 | 0.5095 | 4.022 | 1.842 | True | True | True | True | Sex-specific 1e-2 VUMC, sex-differential UKB |
| 0.004866 | 0.005258 | 7.152 | 0.521 | 4.034 | 1.854 | True | True | True | True | Sex-specific 1e-2 VUMC, sex-differential UKB |
| 0.004866 | 0.005258 | 7.177 | 0.5387 | 4.01 | 1.859 | True | True | True | True | Sex-specific 1e-2 VUMC, sex-differential UKB |

| SE_UKB_female | SE_UKB_male | LOG10P_VUMC_female | LOG10P_VUMC_male | LOG10P_UKB_female | LOG10P_UKB_male | female_effect_direction_replication | male_effect_direction_replication | sex_effect_comparison_replication | pvalue_effect_comparison_replication | replication_type |
|---|---|---|---|---|---|---|---|---|---|---|
| 0.004876 | 0.005268 | 7.43 | 0.5284 | 3.908 | 1.903 | True | True | True | True | Sex-specific 1e-2 VUMC, sex-differential UKB |
| 0.004872 | 0.005265 | 7.214 | 0.4769 | 3.825 | 1.923 | True | True | True | True | Sex-specific 1e-2 VUMC, sex-differential UKB |
| 0.004872 | 0.005265 | 7.22 | 0.4779 | 3.811 | 1.926 | True | True | True | True | Sex-specific 1e-2 VUMC, sex-differential UKB |
| 0.004872 | 0.005266 | 7.266 | 0.4612 | 3.811 | 1.928 | True | True | True | True | Sex-specific 1e-2 VUMC, sex-differential UKB |
| 0.004873 | 0.005267 | 7.246 | 0.4929 | 3.839 | 1.935 | True | True | True | True | Sex-specific 1e-2 VUMC, sex-differential UKB |
| 0.004872 | 0.005266 | 7.121 | 0.5033 | 3.837 | 1.925 | True | True | True | True | Sex-specific 1e-2 VUMC, sex-differential UKB |
| 0.004873 | 0.005267 | 7.231 | 0.4653 | 3.854 | 1.934 | True | True | True | True | Sex-specific 1e-2 VUMC, sex-differential UKB |
| 0.004873 | 0.005267 | 7.2 | 0.4693 | 3.858 | 1.914 | True | True | True | True | Sex-specific 1e-2 VUMC, sex-differential UKB |
| 0.004874 | 0.005268 | 7.159 | 0.4876 | 3.87 | 1.922 | True | True | True | True | Sex-specific 1e-2 VUMC, sex-differential UKB |
| 0.004873 | 0.005267 | 7.186 | 0.4636 | 3.851 | 1.915 | True | True | True | True | Sex-specific 1e-2 VUMC, sex-differential UKB |
| 0.004872 | 0.005266 | 7.165 | 0.4524 | 3.863 | 1.912 | True | True | True | True | Sex-specific 1e-2 VUMC, sex-differential UKB |
| 0.004872 | 0.005266 | 7.166 | 0.4518 | 3.833 | 1.904 | True | True | True | True | Sex-specific 1e-2 VUMC, sex-differential UKB |
| 0.004877 | 0.005273 | 7.136 | 0.4506 | 3.758 | 1.869 | True | True | True | True | Sex-specific 1e-2 VUMC, sex-differential UKB |
| 0.004872 | 0.005266 | 7.187 | 0.4619 | 3.829 | 1.895 | True | True | True | True | Sex-specific 1e-2 VUMC, sex-differential UKB |
| 0.004871 | 0.005266 | 7.18 | 0.4547 | 3.839 | 1.89 | True | True | True | True | Sex-specific 1e-2 VUMC, sex-differential UKB |
| 0.004872 | 0.005266 | 7.125 | 0.4579 | 3.859 | 1.898 | True | True | True | True | Sex-specific 1e-2 VUMC, sex-differential UKB |
| 0.004872 | 0.005266 | 7.124 | 0.4584 | 3.865 | 1.896 | True | True | True | True | Sex-specific 1e-2 VUMC, sex-differential UKB |
| 0.00487 | 0.005265 | 7.095 | 0.3549 | 3.794 | 1.895 | True | True | True | True | Sex-specific 1e-2 VUMC, sex-differential UKB |
| 0.00487 | 0.005265 | 7.147 | 0.3914 | 3.824 | 1.886 | True | True | True | True | Sex-specific 1e-2 VUMC, sex-differential UKB |
| 0.00487 | 0.005265 | 7.148 | 0.3914 | 3.825 | 1.886 | True | True | True | True | Sex-specific 1e-2 VUMC, sex-differential UKB |
| 0.00487 | 0.005265 | 7.148 | 0.3914 | 3.825 | 1.886 | True | True | True | True | Sex-specific 1e-2 VUMC, sex-differential UKB |
| 0.004893 | 0.005291 | 7.073 | 0.4057 | 3.814 | 1.938 | True | True | True | True | Sex-specific 1e-2 VUMC, sex-differential UKB |
| 0.004871 | 0.005266 | 7.113 | 0.376 | 3.817 | 1.889 | True | True | True | True | Sex-specific 1e-2 VUMC, sex-differential UKB |
| 0.004871 | 0.005266 | 7.113 | 0.376 | 3.817 | 1.889 | True | True | True | True | Sex-specific 1e-2 VUMC, sex-differential UKB |
| 0.004545 | 0.004911 | 8.958 | 0.4787 | 4.145 | 1.762 | True | True | True | True | Sex-specific 1e-2 VUMC, sex-differential UKB |
| 0.00487 | 0.005265 | 7.14 | 0.3897 | 3.841 | 1.889 | True | True | True | True | Sex-specific 1e-2 VUMC, sex-differential UKB |

| SE_UKB_female | SE_UKB_male | LOG10P_VUMC_female | LOG10P_VUMC_male | LOG10P_UKB_female | LOG10P_UKB_male | female_effect_direction_replication | male_effect_direction_replication | sex_effect_comparison_replication | pvalue_effect_comparison_replication | replication_type |
|---|---|---|---|---|---|---|---|---|---|---|
| 0.00487 | 0.005265 | 7.116 | 0.3729 | 3.823 | 1.889 | True | True | True | True | Sex-specific 1e-2 VUMC, sex-differential UKB |
| 0.00487 | 0.005265 | 7.116 | 0.3729 | 3.823 | 1.889 | True | True | True | True | Sex-specific 1e-2 VUMC, sex-differential UKB |
| 0.00487 | 0.005265 | 7.111 | 0.3758 | 3.823 | 1.889 | True | True | True | True | Sex-specific 1e-2 VUMC, sex-differential UKB |
| 0.00487 | 0.005265 | 7.154 | 0.4436 | 3.868 | 1.89 | True | True | True | True | Sex-specific 1e-2 VUMC, sex-differential UKB |
| 0.00487 | 0.005265 | 7.139 | 0.4482 | 3.856 | 1.885 | True | True | True | True | Sex-specific 1e-2 VUMC, sex-differential UKB |
| 0.00487 | 0.005265 | 7.139 | 0.4482 | 3.856 | 1.885 | True | True | True | True | Sex-specific 1e-2 VUMC, sex-differential UKB |
| 0.004871 | 0.005266 | 7.016 | 0.4605 | 3.84 | 1.898 | True | True | True | True | Sex-specific 1e-2 VUMC, sex-differential UKB |
| 0.00487 | 0.005265 | 7.14 | 0.4498 | 3.846 | 1.891 | True | True | True | True | Sex-specific 1e-2 VUMC, sex-differential UKB |
| 0.004869 | 0.005264 | 7.122 | 0.4119 | 3.815 | 1.908 | True | True | True | True | Sex-specific 1e-2 VUMC, sex-differential UKB |
| 0.004543 | 0.00491 | 8.992 | 0.5419 | 4.186 | 1.812 | True | True | True | True | Sex-specific 1e-2 VUMC, sex-differential UKB |
| 0.004867 | 0.00526 | 7.112 | 0.4364 | 3.845 | 1.937 | True | True | True | True | Sex-specific 1e-2 VUMC, sex-differential UKB |
| SE_UKB_female | SE_UKB_male | LOG10P_VUMC_female | LOG10P_VUMC_male | LOG10P_UKB_female | LOG10P_UKB_male | female_effect_direction_replication | male_effect_direction_replication | sex_effect_comparison_replication | pvalue_effect_comparison_replication | replication_type |
| 0.00487 | 0.005265 | 7.112 | 0.4382 | 3.816 | 1.913 | True | True | True | True | Sex-specific 1e-2 VUMC, sex-differential UKB |
| 0.00487 | 0.005264 | 7.132 | 0.452 | 3.845 | 1.895 | True | True | True | True | Sex-specific 1e-2 VUMC, sex-differential UKB |
| 0.004871 | 0.005266 | 7.101 | 0.4723 | 3.865 | 1.89 | True | True | True | True | Sex-specific 1e-2 VUMC, sex-differential UKB |
| 0.004552 | 0.004919 | 8.664 | 0.5426 | 4.221 | 1.75 | True | True | True | True | Sex-specific 1e-2 VUMC, sex-differential UKB |
| 0.004551 | 0.004918 | 8.657 | 0.5464 | 4.248 | 1.769 | True | True | True | True | Sex-specific 1e-2 VUMC, sex-differential UKB |
| 0.00488 | 0.005276 | 6.916 | 0.4415 | 3.913 | 1.845 | True | True | True | True | Sex-specific 1e-2 VUMC, sex-differential UKB |
| 0.004552 | 0.004919 | 8.668 | 0.5277 | 4.232 | 1.76 | True | True | True | True | Sex-specific 1e-2 VUMC, sex-differential UKB |
| 0.00455 | 0.004918 | 8.582 | 0.6031 | 4.122 | 1.762 | True | True | True | True | Sex-specific 1e-2 VUMC, sex-differential UKB |
| 0.00488 | 0.005277 | 6.783 | 0.516 | 3.81 | 1.833 | True | True | True | True | Sex-specific 1e-2 VUMC, sex-differential UKB |
| 0.00488 | 0.005276 | 6.868 | 0.4788 | 3.969 | 1.879 | True | True | True | True | Sex-specific 1e-2 VUMC, sex-differential UKB |
| 0.00488 | 0.005277 | 6.777 | 0.5524 | 3.846 | 1.834 | True | True | True | True | Sex-specific 1e-2 VUMC, sex-differential UKB |
| 0.004879 | 0.005276 | 6.792 | 0.5384 | 3.853 | 1.828 | True | True | True | True | Sex-specific 1e-2 VUMC, sex-differential UKB |
| 0.004879 | 0.005275 | 6.775 | 0.5188 | 3.854 | 1.851 | True | True | True | True | Sex-specific 1e-2 VUMC, sex-differential UKB |
| 0.00488 | 0.005277 | 6.796 | 0.5423 | 3.884 | 1.807 | True | True | True | True | Sex-specific 1e-2 VUMC, sex-differential UKB |

| SE_UKB_female | SE_UKB_male | LOG10P_VUMC_female | LOG10P_VUMC_male | LOG10P_UKB_female | LOG10P_UKB_male | female_effect_direction_replication | male_effect_direction_replication | sex_effect_comparison_replication | pvalue_effect_comparison_replication | replication_type |
|---|---|---|---|---|---|---|---|---|---|---|
| 0.004879 | 0.005276 | 6.717 | 0.509 | 3.844 | 1.816 | True | True | True | True | Sex-specific 1e-2 VUMC, sex-differential UKB |
| 0.004879 | 0.005276 | 6.795 | 0.4974 | 3.855 | 1.787 | True | True | True | True | Sex-specific 1e-2 VUMC, sex-differential UKB |
| 0.004551 | 0.004919 | 8.594 | 0.6014 | 4.198 | 1.681 | True | True | True | True | Sex-specific 1e-2 VUMC, sex-differential UKB |
| 0.00488 | 0.005277 | 6.681 | 0.5621 | 3.843 | 1.774 | True | True | True | True | Sex-specific 1e-2 VUMC, sex-differential UKB |
| 0.004879 | 0.005276 | 6.803 | 0.5252 | 3.861 | 1.771 | True | True | True | True | Sex-specific 1e-2 VUMC, sex-differential UKB |
| 0.004558 | 0.004926 | 8.466 | 0.6134 | 4.115 | 1.662 | True | True | True | True | Sex-specific 1e-2 VUMC, sex-differential UKB |
| 0.004883 | 0.005279 | 6.72 | 0.5388 | 3.839 | 1.771 | True | True | True | True | Sex-specific 1e-2 VUMC, sex-differential UKB |
| 0.004884 | 0.005281 | 6.774 | 0.5215 | 3.853 | 1.73 | True | True | True | True | Sex-specific 1e-2 VUMC, sex-differential UKB |
| 0.00488 | 0.005277 | 6.737 | 0.5479 | 3.869 | 1.763 | True | True | True | True | Sex-specific 1e-2 VUMC, sex-differential UKB |
| 0.004878 | 0.005275 | 6.612 | 0.5298 | 3.864 | 1.793 | True | True | True | True | Sex-specific 1e-2 VUMC, sex-differential UKB |
| 0.004551 | 0.00492 | 8.409 | 0.6377 | 4.204 | 1.62 | True | True | True | True | Sex-specific 1e-2 VUMC, sex-differential UKB |
| 0.004879 | 0.005276 | 6.751 | 0.5294 | 3.81 | 1.781 | True | True | True | True | Sex-specific 1e-2 VUMC, sex-differential UKB |
| 0.004551 | 0.004919 | 8.425 | 0.6276 | 4.187 | 1.668 | True | True | True | True | Sex-specific 1e-2 VUMC, sex-differential UKB |
| 0.004879 | 0.005276 | 6.714 | 0.5191 | 3.85 | 1.736 | True | True | True | True | Sex-specific 1e-2 VUMC, sex-differential UKB |
| 0.004879 | 0.005276 | 6.759 | 0.5248 | 3.864 | 1.747 | True | True | True | True | Sex-specific 1e-2 VUMC, sex-differential UKB |
| 0.004879 | 0.005276 | 6.684 | 0.5244 | 3.839 | 1.757 | True | True | True | True | Sex-specific 1e-2 VUMC, sex-differential UKB |
| 0.004552 | 0.00492 | 8.557 | 0.6201 | 4.176 | 1.616 | True | True | True | True | Sex-specific 1e-2 VUMC, sex-differential UKB |
| 0.004551 | 0.004919 | 8.559 | 0.6211 | 4.157 | 1.633 | True | True | True | True | Sex-specific 1e-2 VUMC, sex-differential UKB |
| 0.004551 | 0.004919 | 8.559 | 0.6211 | 4.156 | 1.633 | True | True | True | True | Sex-specific 1e-2 VUMC, sex-differential UKB |
| 0.00488 | 0.005276 | 6.825 | 0.5291 | 3.864 | 1.748 | True | True | True | True | Sex-specific 1e-2 VUMC, sex-differential UKB |
| 0.004551 | 0.004919 | 8.676 | 0.614 | 4.158 | 1.629 | True | True | True | True | Sex-specific 1e-2 VUMC, sex-differential UKB |
| 0.004879 | 0.005276 | 6.827 | 0.5204 | 3.857 | 1.754 | True | True | True | True | Sex-specific 1e-2 VUMC, sex-differential UKB |
| 0.00488 | 0.005276 | 6.853 | 0.5207 | 3.865 | 1.747 | True | True | True | True | Sex-specific 1e-2 VUMC, sex-differential UKB |
| 0.004879 | 0.005276 | 6.807 | 0.5305 | 3.873 | 1.778 | True | True | True | True | Sex-specific 1e-2 VUMC, sex-differential UKB |
| 0.004879 | 0.005276 | 6.807 | 0.5289 | 3.87 | 1.776 | True | True | True | True | Sex-specific 1e-2 VUMC, sex-differential UKB |
| 0.004552 | 0.00492 | 8.561 | 0.617 | 4.15 | 1.634 | True | True | True | True | Sex-specific 1e-2 VUMC, sex-differential UKB |

| SE_UKB_female | SE_UKB_male | LOG10P_VUMC_female | LOG10P_VUMC_male | LOG10P_UKB_female | LOG10P_UKB_male | female_effect_direction_replication | male_effect_direction_replication | sex_effect_comparison_replication | pvalue_effect_comparison_replication | replication_type |
|---|---|---|---|---|---|---|---|---|---|---|
| 0.004552 | 0.00492 | 8.562 | 0.6209 | 4.139 | 1.641 | True | True | True | True | Sex-specific 1e-2 VUMC, sex-differential UKB |
| 0.00488 | 0.005277 | 6.822 | 0.5376 | 3.859 | 1.748 | True | True | True | True | Sex-specific 1e-2 VUMC, sex-differential UKB |
| 0.004879 | 0.005276 | 6.76 | 0.5419 | 3.862 | 1.781 | True | True | True | True | Sex-specific 1e-2 VUMC, sex-differential UKB |
| 0.004879 | 0.005276 | 6.755 | 0.5433 | 3.861 | 1.781 | True | True | True | True | Sex-specific 1e-2 VUMC, sex-differential UKB |
| 0.00462 | 0.004992 | 8.58 | 0.5305 | 4.078 | 1.609 | True | True | True | True | Sex-specific 1e-2 VUMC, sex-differential UKB |
| 0.004879 | 0.005276 | 6.752 | 0.543 | 3.865 | 1.787 | True | True | True | True | Sex-specific 1e-2 VUMC, sex-differential UKB |
| 0.004881 | 0.005277 | 6.778 | 0.5767 | 3.904 | 1.776 | True | True | True | True | Sex-specific 1e-2 VUMC, sex-differential UKB |
| 0.004551 | 0.004919 | 8.516 | 0.6247 | 4.156 | 1.657 | True | True | True | True | Sex-specific 1e-2 VUMC, sex-differential UKB |
| 0.004879 | 0.005275 | 6.762 | 0.5349 | 3.868 | 1.767 | True | True | True | True | Sex-specific 1e-2 VUMC, sex-differential UKB |
| 0.004551 | 0.004918 | 8.494 | 0.6156 | 4.154 | 1.651 | True | True | True | True | Sex-specific 1e-2 VUMC, sex-differential UKB |
| 0.00455 | 0.004917 | 8.501 | 0.6169 | 4.095 | 1.679 | True | True | True | True | Sex-specific 1e-2 VUMC, sex-differential UKB |
| 0.00455 | 0.004918 | 8.501 | 0.6127 | 4.079 | 1.647 | True | True | True | True | Sex-specific 1e-2 VUMC, sex-differential UKB |
| 0.00455 | 0.004918 | 8.493 | 0.6333 | 4.072 | 1.646 | True | True | True | True | Sex-specific 1e-2 VUMC, sex-differential UKB |
| 0.00455 | 0.004917 | 8.538 | 0.6403 | 4.086 | 1.674 | True | True | True | True | Sex-specific 1e-2 VUMC, sex-differential UKB |
| 0.004551 | 0.004917 | 8.518 | 0.6411 | 4.078 | 1.666 | True | True | True | True | Sex-specific 1e-2 VUMC, sex-differential UKB |
| 0.00455 | 0.004917 | 8.493 | 0.6333 | 4.07 | 1.642 | True | True | True | True | Sex-specific 1e-2 VUMC, sex-differential UKB |
| 0.00455 | 0.004917 | 8.493 | 0.6333 | 4.07 | 1.643 | True | True | True | True | Sex-specific 1e-2 VUMC, sex-differential UKB |
| 0.004137 | 0.004474 | 6.127 | 0.7569 | 2.81 | 1.666 | True | True | True | True | Sex-specific 1e-2 VUMC, sex-differential UKB |
| 0.004138 | 0.004475 | 6.225 | 0.7919 | 2.827 | 1.658 | True | True | True | True | Sex-specific 1e-2 VUMC, sex-differential UKB |
| 0.00455 | 0.004918 | 8.461 | 0.6367 | 4.141 | 1.643 | True | True | True | True | Sex-specific 1e-2 VUMC, sex-differential UKB |
| SE_UKB_female | SE_UKB_male | LOG10P_VUMC_female | LOG10P_VUMC_male | LOG10P_UKB_female | LOG10P_UKB_male | female_effect_direction_replication | male_effect_direction_replication | sex_effect_comparison_replication | pvalue_effect_comparison_replication | replication_type |
| 0.00455 | 0.004917 | 8.468 | 0.649 | 4.154 | 1.609 | True | True | True | True | Sex-specific 1e-2 VUMC, sex-differential UKB |
| 0.00455 | 0.004917 | 8.464 | 0.6495 | 4.16 | 1.646 | True | True | True | True | Sex-specific 1e-2 VUMC, sex-differential UKB |
| 0.004516 | 0.004884 | 8.436 | 0.5124 | 4.023 | 1.646 | True | True | True | True | Sex-specific 1e-2 VUMC, sex-differential UKB |
| 0.00455 | 0.004917 | 8.467 | 0.6465 | 4.158 | 1.642 | True | True | True | True | Sex-specific 1e-2 VUMC, sex-differential UKB |
| 0.00455 | 0.004917 | 8.392 | 0.6287 | 4.096 | 1.609 | True | True | True | True | Sex-specific 1e-2 VUMC, sex-differential UKB |

| SE_UKB_female | SE_UKB_male | LOG10P_VUMC_female | LOG10P_VUMC_male | LOG10P_UKB_female | LOG10P_UKB_male | female_effect_direction_replication | male_effect_direction_replication | sex_effect_comparison_replication | pvalue_effect_comparison_replication | replication_type |
|---|---|---|---|---|---|---|---|---|---|---|
| 0.004516 | 0.004884 | 8.36 | 0.5141 | 4.002 | 1.651 | True | True | True | True | Sex-specific 1e-2 VUMC, sex-differential UKB |
| 0.00455 | 0.004917 | 8.408 | 0.6477 | 4.08 | 1.641 | True | True | True | True | Sex-specific 1e-2 VUMC, sex-differential UKB |
| 0.004878 | 0.005274 | 6.715 | 0.5736 | 3.862 | 1.722 | True | True | True | True | Sex-specific 1e-2 VUMC, sex-differential UKB |
| 0.00455 | 0.004917 | 8.439 | 0.6467 | 4.146 | 1.614 | True | True | True | True | Sex-specific 1e-2 VUMC, sex-differential UKB |
| 0.00455 | 0.004916 | 8.437 | 0.6475 | 4.073 | 1.645 | True | True | True | True | Sex-specific 1e-2 VUMC, sex-differential UKB |
| 0.00455 | 0.004917 | 8.39 | 0.6507 | 4.124 | 1.65 | True | True | True | True | Sex-specific 1e-2 VUMC, sex-differential UKB |
| 0.00455 | 0.004916 | 8.428 | 0.6521 | 4.145 | 1.641 | True | True | True | True | Sex-specific 1e-2 VUMC, sex-differential UKB |
| 0.004878 | 0.005274 | 6.706 | 0.5649 | 3.856 | 1.722 | True | True | True | True | Sex-specific 1e-2 VUMC, sex-differential UKB |
| 0.004516 | 0.004884 | 8.399 | 0.5195 | 3.995 | 1.614 | True | True | True | True | Sex-specific 1e-2 VUMC, sex-differential UKB |
| 0.004516 | 0.004884 | 8.4 | 0.5196 | 3.995 | 1.618 | True | True | True | True | Sex-specific 1e-2 VUMC, sex-differential UKB |
| 0.004876 | 0.005272 | 6.662 | 0.572 | 3.841 | 1.745 | True | True | True | True | Sex-specific 1e-2 VUMC, sex-differential UKB |
| 0.004515 | 0.004882 | 8.439 | 0.5106 | 3.988 | 1.624 | True | True | True | True | Sex-specific 1e-2 VUMC, sex-differential UKB |
| 0.004537 | 0.004902 | 8.25 | 0.6726 | 4.155 | 1.625 | True | True | True | True | Sex-specific 1e-2 VUMC, sex-differential UKB |
| 0.004833 | 0.00523 | 6.61 | 0.4602 | 3.705 | 1.699 | True | True | True | True | Sex-specific 1e-2 VUMC, sex-differential UKB |
| 0.004514 | 0.004882 | 8.263 | 0.4909 | 4.021 | 1.632 | True | True | True | True | Sex-specific 1e-2 VUMC, sex-differential UKB |
| 0.004831 | 0.005228 | 6.579 | 0.4327 | 3.675 | 1.725 | True | True | True | True | Sex-specific 1e-2 VUMC, sex-differential UKB |
| 0.004832 | 0.005229 | 6.626 | 0.4442 | 3.53 | 1.629 | True | True | True | True | Sex-specific 1e-2 VUMC, sex-differential UKB |
| 0.004551 | 0.004916 | 8.373 | 0.6355 | 4.037 | 1.561 | True | True | True | True | Sex-specific 1e-2 VUMC, sex-differential UKB |
| 0.00483 | 0.005226 | 6.591 | 0.4728 | 3.6 | 1.642 | True | True | True | True | Sex-specific 1e-2 VUMC, sex-differential UKB |
| 0.004512 | 0.004879 | 8.306 | 0.499 | 3.87 | 1.538 | True | True | True | True | Sex-specific 1e-2 VUMC, sex-differential UKB |
| 0.004828 | 0.005225 | 6.532 | 0.4385 | 3.584 | 1.673 | True | True | True | True | Sex-specific 1e-2 VUMC, sex-differential UKB |
| 0.004827 | 0.005224 | 6.818 | 0.4033 | 3.612 | 1.642 | True | True | True | True | Sex-specific 1e-2 VUMC, sex-differential UKB |
| 0.004875 | 0.005269 | 6.628 | 0.5584 | 3.815 | 1.699 | True | True | True | True | Sex-specific 1e-2 VUMC, sex-differential UKB |
| 0.004548 | 0.004914 | 8.34 | 0.6158 | 4.07 | 1.557 | True | True | True | True | Sex-specific 1e-2 VUMC, sex-differential UKB |
| 0.004828 | 0.005225 | 6.563 | 0.4455 | 3.568 | 1.627 | True | True | True | True | Sex-specific 1e-2 VUMC, sex-differential UKB |
| 0.004827 | 0.005223 | 6.5 | 0.4503 | 3.568 | 1.661 | True | True | True | True | Sex-specific 1e-2 VUMC, sex-differential UKB |

| SE_UKB_female | SE_UKB_male | LOG10P_VUMC_female | LOG10P_VUMC_male | LOG10P_UKB_female | LOG10P_UKB_male | female_effect_direction_replication | male_effect_direction_replication | sex_effect_comparison_replication | pvalue_effect_comparison_replication | replication_type |
|---|---|---|---|---|---|---|---|---|---|---|
| 0.004511 | 0.004877 | 8.24 | 0.5059 | 3.873 | 1.524 | True | True | True | True | Sex-specific 1e-2 VUMC, sex-differential UKB |
| 0.004827 | 0.005223 | 6.491 | 0.4538 | 3.58 | 1.633 | True | True | True | True | Sex-specific 1e-2 VUMC, sex-differential UKB |
| 0.004827 | 0.005223 | 6.544 | 0.5015 | 3.691 | 1.639 | True | True | True | True | Sex-specific 1e-2 VUMC, sex-differential UKB |
| 0.004814 | 0.005208 | 6.779 | 0.4594 | 3.678 | 1.648 | True | True | True | True | Sex-specific 1e-2 VUMC, sex-differential UKB |
| 0.004802 | 0.005198 | 6.456 | 0.5384 | 4.024 | 1.724 | True | True | True | True | Sex-specific 1e-2 VUMC, sex-differential UKB |
| 0.004836 | 0.005232 | 6.851 | 0.4535 | 3.603 | 1.649 | True | True | True | True | Sex-specific 1e-2 VUMC, sex-differential UKB |
| 0.00484 | 0.005237 | 6.84 | 0.4453 | 3.622 | 1.689 | True | True | True | True | Sex-specific 1e-2 VUMC, sex-differential UKB |
| 0.004841 | 0.005238 | 6.609 | 0.4838 | 3.606 | 1.83 | True | True | True | True | Sex-specific 1e-2 VUMC, sex-differential UKB |
| 0.003144 | 0.003397 | 0.4734 | 6.797 | 38.39 | 68.2 | True | True | True | True | Sex-specific 1e-2 VUMC, sex-differential UKB |
| 0.003144 | 0.003396 | 0.4261 | 6.755 | 38.4 | 67.58 | True | True | True | True | Sex-specific 1e-2 VUMC, sex-differential UKB |
| 0.003142 | 0.003394 | 0.4478 | 6.822 | 38.64 | 67.83 | True | True | True | True | Sex-specific 1e-2 VUMC, sex-differential UKB |
| 0.003142 | 0.003394 | 0.4882 | 6.943 | 38.47 | 67.91 | True | True | True | True | Sex-specific 1e-2 VUMC, sex-differential UKB |
| 0.003141 | 0.003393 | 0.4437 | 6.779 | 38.61 | 67.93 | True | True | True | True | Sex-specific 1e-2 VUMC, sex-differential UKB |
| 0.003141 | 0.003393 | 0.4495 | 6.713 | 38.74 | 67.92 | True | True | True | True | Sex-specific 1e-2 VUMC, sex-differential UKB |
| | | 0.4178 | 6.803 | | | False | False | False | False | |
| 0.00314 | 0.003392 | 0.4043 | 6.429 | 38.77 | 67.8 | True | True | True | True | Sex-specific 1e-2 VUMC, sex-differential UKB |
| 0.003134 | 0.003385 | 0.4479 | 6.808 | 35.29 | 61.37 | True | True | True | True | Sex-specific 1e-2 VUMC, sex-differential UKB |
| 0.00314 | 0.003391 | 0.4625 | 6.72 | 38.74 | 67.9 | True | True | True | True | Sex-specific 1e-2 VUMC, sex-differential UKB |
| 0.00314 | 0.003392 | 0.4527 | 6.697 | 39.01 | 67.79 | True | True | True | True | Sex-specific 1e-2 VUMC, sex-differential UKB |
| 0.003155 | 0.003409 | 0.3983 | 6.746 | 41.7 | 68.11 | True | True | True | True | Sex-specific 1e-2 VUMC, sex-differential UKB |
| 0.003136 | 0.003389 | 0.2687 | 6.5 | 29.59 | 51.4 | True | True | True | True | Sex-specific 1e-2 VUMC, sex-differential UKB |
| 0.003138 | 0.003391 | 0.3789 | 6.759 | 39.3 | 67.88 | True | True | True | True | Sex-specific 1e-2 VUMC, sex-differential UKB |
| 0.00314 | 0.003392 | 0.3877 | 6.739 | 39.41 | 68.14 | True | True | True | True | Sex-specific 1e-2 VUMC, sex-differential UKB |
| 0.003139 | 0.003392 | 0.3365 | 6.968 | 39.53 | 68.42 | True | True | True | True | Sex-specific 1e-2 VUMC, sex-differential UKB |
| 0.003139 | 0.003392 | 0.3361 | 6.828 | 39.49 | 68.34 | True | True | True | True | Sex-specific 1e-2 VUMC, sex-differential UKB |
| 0.003139 | 0.003392 | 0.3569 | 7.125 | 39.49 | 68.24 | True | True | True | True | Sex-specific 1e-2 VUMC, sex-differential UKB |

| SE_UKB_female | SE_UKB_male | LOG10P_VUMC_female | LOG10P_VUMC_male | LOG10P_UKB_female | LOG10P_UKB_male | female_effect_direction_replication | male_effect_direction_replication | sex_effect_comparison_replication | pvalue_effect_comparison_replication | replication_type |
|---|---|---|---|---|---|---|---|---|---|---|
| 0.003139 | 0.003392 | 0.3485 | 7.156 | 39.35 | 68.24 | True | True | True | True | Sex-specific 1e-2 VUMC, sex-differential UKB |
| 0.003139 | 0.003391 | 0.3606 | 7.126 | 39.5 | 68.33 | True | True | True | True | Sex-specific 1e-2 VUMC, sex-differential UKB |
| 0.003139 | 0.003391 | 0.3606 | 7.126 | 39.5 | 68.33 | True | True | True | True | Sex-specific 1e-2 VUMC, sex-differential UKB |
| SE_UKB_female | SE_UKB_male | LOG10P_VUMC_female | LOG10P_VUMC_male | LOG10P_UKB_female | LOG10P_UKB_male | female_effect_direction_replication | male_effect_direction_replication | sex_effect_comparison_replication | pvalue_effect_comparison_replication | replication_type |
| 0.003139 | 0.003391 | 0.3493 | 7.011 | 39.59 | 68.14 | True | True | True | True | Sex-specific 1e-2 VUMC, sex-differential UKB |
| 0.003139 | 0.003392 | 0.3593 | 7.209 | 39.51 | 68.34 | True | True | True | True | Sex-specific 1e-2 VUMC, sex-differential UKB |
| 0.003139 | 0.003392 | 0.3448 | 7.1 | 39.58 | 68.31 | True | True | True | True | Sex-specific 1e-2 VUMC, sex-differential UKB |
| 0.003137 | 0.00339 | 0.3444 | 7.134 | 39.64 | 68.24 | True | True | True | True | Sex-specific 1e-2 VUMC, sex-differential UKB |
| 0.003137 | 0.00339 | 0.3535 | 7.145 | 39.56 | 68.35 | True | True | True | True | Sex-specific 1e-2 VUMC, sex-differential UKB |
| 0.003137 | 0.00339 | 0.3414 | 7.139 | 39.7 | 68.19 | True | True | True | True | Sex-specific 1e-2 VUMC, sex-differential UKB |
| 0.003141 | 0.003393 | 0.3843 | 7.021 | 39.34 | 67.58 | True | True | True | True | Sex-specific 1e-2 VUMC, sex-differential UKB |
| 0.003138 | 0.00339 | 0.3526 | 7.148 | 39.65 | 68.19 | True | True | True | True | Sex-specific 1e-2 VUMC, sex-differential UKB |
| 0.003137 | 0.00339 | 0.3697 | 7.208 | 39.67 | 68.29 | True | True | True | True | Sex-specific 1e-2 VUMC, sex-differential UKB |
| 0.003138 | 0.003391 | 0.3365 | 7.051 | 39.36 | 68.05 | True | True | True | True | Sex-specific 1e-2 VUMC, sex-differential UKB |
| 0.003137 | 0.00339 | 0.3336 | 6.959 | 39.85 | 68.24 | True | True | True | True | Sex-specific 1e-2 VUMC, sex-differential UKB |
| 0.003138 | 0.003391 | 0.2746 | 6.694 | 39.81 | 67.7 | True | True | True | True | Sex-specific 1e-2 VUMC, sex-differential UKB |
| 0.003138 | 0.003391 | 0.292 | 6.693 | 39.64 | 67.87 | True | True | True | True | Sex-specific 1e-2 VUMC, sex-differential UKB |
| 0.003138 | 0.003392 | 0.324 | 7.062 | 39.72 | 68.2 | True | True | True | True | Sex-specific 1e-2 VUMC, sex-differential UKB |
| 0.003138 | 0.003392 | 0.3198 | 6.928 | 39.77 | 68.08 | True | True | True | True | Sex-specific 1e-2 VUMC, sex-differential UKB |
| 0.003138 | 0.003391 | 0.3134 | 6.962 | 39.61 | 68.28 | True | True | True | True | Sex-specific 1e-2 VUMC, sex-differential UKB |
| 0.003138 | 0.003392 | 0.3277 | 6.926 | 39.54 | 68.18 | True | True | True | True | Sex-specific 1e-2 VUMC, sex-differential UKB |
| 0.003138 | 0.003391 | 0.3252 | 7.029 | 39.63 | 68.2 | True | True | True | True | Sex-specific 1e-2 VUMC, sex-differential UKB |
| 0.003138 | 0.003392 | 0.3136 | 6.801 | 39.68 | 68.28 | True | True | True | True | Sex-specific 1e-2 VUMC, sex-differential UKB |
| 0.003138 | 0.003392 | 0.36 | 7.027 | 39.7 | 68.2 | True | True | True | True | Sex-specific 1e-2 VUMC, sex-differential UKB |
| 0.003138 | 0.003392 | 0.3361 | 6.851 | 39.62 | 67.75 | True | True | True | True | Sex-specific 1e-2 VUMC, sex-differential UKB |
| 0.003138 | 0.003392 | 0.3468 | 7.12 | 39.69 | 68.22 | True | True | True | True | Sex-specific 1e-2 VUMC, sex-differential UKB |

| SE_UKB_female | SE_UKB_male | LOG10P_VUMC_female | LOG10P_VUMC_male | LOG10P_UKB_female | LOG10P_UKB_male | female_effect_direction_replication | male_effect_direction_replication | sex_effect_comparison_replication | pvalue_effect_comparison_replication | replication_type |
|---|---|---|---|---|---|---|---|---|---|---|
| 0.003138 | 0.003392 | 0.3425 | 7.131 | 39.68 | 68.22 | True | True | True | True | Sex-specific 1e-2 VUMC, sex-differential UKB |
| 0.003138 | 0.003392 | 0.3716 | 7.182 | 39.73 | 68.18 | True | True | True | True | Sex-specific 1e-2 VUMC, sex-differential UKB |
| 0.003141 | 0.003396 | 0.3011 | 6.96 | 39.21 | 67.69 | True | True | True | True | Sex-specific 1e-2 VUMC, sex-differential UKB |
| 0.003138 | 0.003392 | 0.3433 | 6.959 | 39.75 | 67.69 | True | True | True | True | Sex-specific 1e-2 VUMC, sex-differential UKB |
| 0.003138 | 0.003392 | 0.3608 | 7.155 | 39.53 | 68.26 | True | True | True | True | Sex-specific 1e-2 VUMC, sex-differential UKB |
| 0.003138 | 0.003392 | 0.3536 | 7.188 | 39.69 | 68.03 | True | True | True | True | Sex-specific 1e-2 VUMC, sex-differential UKB |
| 0.003138 | 0.003392 | 0.3583 | 7.23 | 39.44 | 68.23 | True | True | True | True | Sex-specific 1e-2 VUMC, sex-differential UKB |
| 0.003138 | 0.003392 | 0.3409 | 7.172 | 39.69 | 68.31 | True | True | True | True | Sex-specific 1e-2 VUMC, sex-differential UKB |
| 0.003138 | 0.003391 | 0.3407 | 7.04 | 39.36 | 68.36 | True | True | True | True | Sex-specific 1e-2 VUMC, sex-differential UKB |
| 0.003137 | 0.003392 | 0.3519 | 7.285 | 39.22 | 68.4 | True | True | True | True | Sex-specific 1e-2 VUMC, sex-differential UKB |
| 0.003137 | 0.003391 | 0.3709 | 7.242 | 39.19 | 68.17 | True | True | True | True | Sex-specific 1e-2 VUMC, sex-differential UKB |
| 0.003137 | 0.003391 | 0.3781 | 7.302 | 39.18 | 68.16 | True | True | True | True | Sex-specific 1e-2 VUMC, sex-differential UKB |
| 0.003137 | 0.003391 | 0.3525 | 7.281 | 39.07 | 68.24 | True | True | True | True | Sex-specific 1e-2 VUMC, sex-differential UKB |
| 0.003137 | 0.003391 | 0.3612 | 7.311 | 39.03 | 68.27 | True | True | True | True | Sex-specific 1e-2 VUMC, sex-differential UKB |
| 0.003137 | 0.003391 | 0.3498 | 7.231 | 38.99 | 68.31 | True | True | True | True | Sex-specific 1e-2 VUMC, sex-differential UKB |
| 0.003137 | 0.003391 | 0.3437 | 7.234 | 39.03 | 68.3 | True | True | True | True | Sex-specific 1e-2 VUMC, sex-differential UKB |
| 0.003137 | 0.003391 | 0.3371 | 7.22 | 39 | 68.07 | True | True | True | True | Sex-specific 1e-2 VUMC, sex-differential UKB |
| 0.003138 | 0.003392 | 0.368 | 7.324 | 38.95 | 68.16 | True | True | True | True | Sex-specific 1e-2 VUMC, sex-differential UKB |
|  |  | 0.3227 | 7.053 |  |  | False | False | False | False |  |
| 0.003125 | 0.00338 | 0.6425 | 7.313 | 44.79 | 65.17 | True | True | True | True | Sex-specific 1e-2 VUMC, sex-differential UKB |
| 0.003128 | 0.003384 | 0.7718 | 6.249 | 38.58 | 61.04 | True | True | True | True | Sex-specific 1e-2 VUMC, sex-differential UKB |
| 0.003124 | 0.003381 | 0.507 | 7.246 | 41.81 | 62.71 | True | True | True | True | Sex-specific 1e-2 VUMC, sex-differential UKB |
| 0.003125 | 0.003381 | 0.541 | 6.947 | 41.96 | 62.85 | True | True | True | True | Sex-specific 1e-2 VUMC, sex-differential UKB |
| 0.003125 | 0.003381 | 0.5417 | 7.293 | 42 | 62.59 | True | True | True | True | Sex-specific 1e-2 VUMC, sex-differential UKB |
| 0.003125 | 0.003381 | 0.5365 | 6.842 | 42.12 | 62.28 | True | True | True | True | Sex-specific 1e-2 VUMC, sex-differential UKB |
| 0.003124 | 0.003381 | 0.5397 | 7.035 | 41.89 | 62.45 | True | True | True | True | Sex-specific 1e-2 VUMC, sex-differential UKB |

| SE_UKB_female | SE_UKB_male | LOG10P_VUMC_female | LOG10P_VUMC_male | LOG10P_UKB_female | LOG10P_UKB_male | female_effect_direction_replication | male_effect_direction_replication | sex_effect_comparison_replication | pvalue_effect_comparison_replication | replication_type |
|---|---|---|---|---|---|---|---|---|---|---|
| 0.003125 | 0.003381 | 0.4934 | 7.183 | 42 | 62.22 | True | True | True | True | Sex-specific 1e-2 VUMC, sex-differential UKB |
| 0.003125 | 0.003381 | 0.4965 | 7.187 | 42.01 | 62.27 | True | True | True | True | Sex-specific 1e-2 VUMC, sex-differential UKB |
| 0.003125 | 0.003381 | 0.5234 | 7.15 | 41.69 | 62.2 | True | True | True | True | Sex-specific 1e-2 VUMC, sex-differential UKB |
| 0.003124 | 0.003379 | 0.471 | 6.846 | 41.47 | 61.7 | True | True | True | True | Sex-specific 1e-2 VUMC, sex-differential UKB |
| 0.003124 | 0.003379 | 0.4806 | 6.873 | 41.36 | 61.61 | True | True | True | True | Sex-specific 1e-2 VUMC, sex-differential UKB |
| 0.003124 | 0.003379 | 0.4806 | 6.873 | 41.36 | 61.62 | True | True | True | True | Sex-specific 1e-2 VUMC, sex-differential UKB |
| 0.003124 | 0.003379 | 0.513 | 6.946 | 41.38 | 61.61 | True | True | True | True | Sex-specific 1e-2 VUMC, sex-differential UKB |
| 0.003124 | 0.003379 | 0.5403 | 7.286 | 41.46 | 61.95 | True | True | True | True | Sex-specific 1e-2 VUMC, sex-differential UKB |
| 0.003124 | 0.003379 | 0.5617 | 7.12 | 41.42 | 62.09 | True | True | True | True | Sex-specific 1e-2 VUMC, sex-differential UKB |
| 0.003124 | 0.003379 | 0.5617 | 7.12 | 41.42 | 62.09 | True | True | True | True | Sex-specific 1e-2 VUMC, sex-differential UKB |
| 0.003124 | 0.003379 | 0.5157 | 7.077 | 41.49 | 61.71 | True | True | True | True | Sex-specific 1e-2 VUMC, sex-differential UKB |
| 0.003124 | 0.003379 | 0.5227 | 6.804 | 41.45 | 61.89 | True | True | True | True | Sex-specific 1e-2 VUMC, sex-differential UKB |
| SE_UKB_female | SE_UKB_male | LOG10P_VUMC_female | LOG10P_VUMC_male | LOG10P_UKB_female | LOG10P_UKB_male | female_effect_direction_replication | male_effect_direction_replication | sex_effect_comparison_replication | pvalue_effect_comparison_replication | replication_type |
| 0.003124 | 0.003379 | 0.5426 | 6.803 | 41.35 | 61.72 | True | True | True | True | Sex-specific 1e-2 VUMC, sex-differential UKB |
| 0.003124 | 0.003379 | 0.4824 | 6.796 | 41.33 | 61.66 | True | True | True | True | Sex-specific 1e-2 VUMC, sex-differential UKB |
| 0.003124 | 0.003379 | 0.5547 | 7.157 | 41.27 | 62.01 | True | True | True | True | Sex-specific 1e-2 VUMC, sex-differential UKB |
| 0.003124 | 0.003379 | 0.623 | 6.647 | 40.5 | 61.51 | True | True | True | True | Sex-specific 1e-2 VUMC, sex-differential UKB |
| 0.003124 | 0.003379 | 0.6034 | 6.663 | 40.38 | 61.69 | True | True | True | True | Sex-specific 1e-2 VUMC, sex-differential UKB |
| 0.003125 | 0.00338 | 0.5619 | 6.455 | 40.38 | 61.25 | True | True | True | True | Sex-specific 1e-2 VUMC, sex-differential UKB |
| 0.003124 | 0.003379 | 0.5753 | 6.887 | 40.46 | 61.56 | True | True | True | True | Sex-specific 1e-2 VUMC, sex-differential UKB |
| 0.003124 | 0.003379 | 0.5804 | 6.676 | 40.43 | 61.64 | True | True | True | True | Sex-specific 1e-2 VUMC, sex-differential UKB |
| 0.003125 | 0.003381 | 0.5847 | 6.761 | 40.35 | 61.33 | True | True | True | True | Sex-specific 1e-2 VUMC, sex-differential UKB |
| 0.003124 | 0.003379 | 0.5519 | 6.823 | 40.56 | 61.38 | True | True | True | True | Sex-specific 1e-2 VUMC, sex-differential UKB |
| 0.003124 | 0.003379 | 0.5477 | 6.838 | 40.56 | 61.49 | True | True | True | True | Sex-specific 1e-2 VUMC, sex-differential UKB |
| 0.003124 | 0.003379 | 0.5454 | 6.764 | 40.55 | 61.5 | True | True | True | True | Sex-specific 1e-2 VUMC, sex-differential UKB |
| 0.003124 | 0.003379 | 0.5504 | 6.445 | 40.42 | 61.42 | True | True | True | True | Sex-specific 1e-2 VUMC, sex-differential UKB |

| SE_UKB_female | SE_UKB_male | LOG10P_VUMC_female | LOG10P_VUMC_male | LOG10P_UKB_female | LOG10P_UKB_male | female_effect_direction_replication | male_effect_direction_replication | sex_effect_comparison_replication | pvalue_effect_comparison_replication | replication_type |
|---|---|---|---|---|---|---|---|---|---|---|
| 0.003124 | 0.003379 | 0.6107 | 6.696 | 40.5 | 61.84 | True | True | True | True | Sex-specific 1e-2 VUMC, sex-differential UKB |
| 0.003124 | 0.003379 | 0.6183 | 6.689 | 40.4 | 61.74 | True | True | True | True | Sex-specific 1e-2 VUMC, sex-differential UKB |
| 0.003128 | 0.003383 | 0.6635 | 6.633 | 40.98 | 61.89 | True | True | True | True | Sex-specific 1e-2 VUMC, sex-differential UKB |
| 0.003124 | 0.003379 | 0.5528 | 6.76 | 40.92 | 61.45 | True | True | True | True | Sex-specific 1e-2 VUMC, sex-differential UKB |
| 0.003124 | 0.003379 | 0.523 | 6.79 | 40.8 | 61.32 | True | True | True | True | Sex-specific 1e-2 VUMC, sex-differential UKB |
| 0.003124 | 0.003379 | 0.6233 | 6.655 | 41.05 | 61.47 | True | True | True | True | Sex-specific 1e-2 VUMC, sex-differential UKB |
| 0.003124 | 0.003379 | 0.6481 | 6.656 | 41.02 | 61.66 | True | True | True | True | Sex-specific 1e-2 VUMC, sex-differential UKB |
| 0.003124 | 0.003379 | 0.6334 | 6.833 | 41.17 | 61.68 | True | True | True | True | Sex-specific 1e-2 VUMC, sex-differential UKB |
| 0.003124 | 0.003379 | 0.6203 | 6.919 | 41.17 | 61.55 | True | True | True | True | Sex-specific 1e-2 VUMC, sex-differential UKB |
| 0.003126 | 0.003381 | 0.595 | 6.613 | 41.23 | 60.25 | True | True | True | True | Sex-specific 1e-2 VUMC, sex-differential UKB |
| 0.003124 | 0.003379 | 0.6442 | 6.638 | 41.36 | 61.38 | True | True | True | True | Sex-specific 1e-2 VUMC, sex-differential UKB |
| 0.003124 | 0.003379 | 0.6128 | 6.643 | 41.36 | 61.34 | True | True | True | True | Sex-specific 1e-2 VUMC, sex-differential UKB |
| 0.003125 | 0.00338 | 0.616 | 6.46 | 41.42 | 61.37 | True | True | True | True | Sex-specific 1e-2 VUMC, sex-differential UKB |
| 0.003124 | 0.003379 | 0.6327 | 6.672 | 41.35 | 61.43 | True | True | True | True | Sex-specific 1e-2 VUMC, sex-differential UKB |
| 0.003124 | 0.003379 | 0.6194 | 6.447 | 41.27 | 61.25 | True | True | True | True | Sex-specific 1e-2 VUMC, sex-differential UKB |
| 0.003125 | 0.00338 | 0.6321 | 6.682 | 41.12 | 61.74 | True | True | True | True | Sex-specific 1e-2 VUMC, sex-differential UKB |
| 0.003125 | 0.00338 | 0.5581 | 6.433 | 40.94 | 61.23 | True | True | True | True | Sex-specific 1e-2 VUMC, sex-differential UKB |
| 0.003136 | 0.003394 | 0.5167 | 5.021 | 37.72 | 57.08 | True | True | True | True | Sex-specific 1e-2 VUMC, sex-differential UKB |
| 0.003125 | 0.00338 | 0.6279 | 6.474 | 40.99 | 61.49 | True | True | True | True | Sex-specific 1e-2 VUMC, sex-differential UKB |
| 0.003125 | 0.00338 | 0.602 | 6.501 | 40.93 | 61.43 | True | True | True | True | Sex-specific 1e-2 VUMC, sex-differential UKB |
| | | 0.6022 | 6.484 | | | False | False | False | False | |
| | | 0.6132 | 6.741 | | | False | False | False | False | |
| 0.003125 | 0.00338 | 0.5204 | 6.339 | 40.85 | 61.16 | True | True | True | True | Sex-specific 1e-2 VUMC, sex-differential UKB |
| 0.003125 | 0.00338 | 0.5659 | 6.474 | 40.94 | 61.28 | True | True | True | True | Sex-specific 1e-2 VUMC, sex-differential UKB |
| 0.003125 | 0.00338 | 0.5588 | 6.372 | 40.72 | 61 | True | True | True | True | Sex-specific 1e-2 VUMC, sex-differential UKB |
| 0.003125 | 0.00338 | 0.5945 | 6.529 | 40.88 | 61.4 | True | True | True | True | Sex-specific 1e-2 VUMC, sex-differential UKB |
| 0.003125 | 0.00338 | 0.593 | 6.571 | 40.85 | 61.26 | True | True | True | True | Sex-specific 1e-2 VUMC, sex-differential UKB |

| SE_UKB_female | SE_UKB_male | LOG10P_VUMC_female | LOG10P_VUMC_male | LOG10P_UKB_female | LOG10P_UKB_male | female_effect_direction_replication | male_effect_direction_replication | sex_effect_comparison_replication | pvalue_effect_comparison_replication | replication_type |
|---|---|---|---|---|---|---|---|---|---|---|
| 0.003125 | 0.00338 | 0.5885 | 6.472 | 40.89 | 61.41 | True | True | True | True | Sex-specific 1e-2 VUMC, sex-differential UKB |
| 0.003125 | 0.00338 | 0.5288 | 6.2 | 40.85 | 61.04 | True | True | True | True | Sex-specific 1e-2 VUMC, sex-differential UKB |
| 0.003125 | 0.00338 | 0.5883 | 6.494 | 40.9 | 61.39 | True | True | True | True | Sex-specific 1e-2 VUMC, sex-differential UKB |
| 0.003125 | 0.00338 | 0.5644 | 6.442 | 40.94 | 61.26 | True | True | True | True | Sex-specific 1e-2 VUMC, sex-differential UKB |
| 0.003127 | 0.003382 | 0.5894 | 6.513 | 40.83 | 61.38 | True | True | True | True | Sex-specific 1e-2 VUMC, sex-differential UKB |
| 0.003138 | 0.003398 | 0.341 | 4.607 | 32.74 | 52.02 | True | True | True | True | Sex-specific 1e-2 VUMC, sex-differential UKB |
| 0.003129 | 0.003386 | 0.3379 | 5.662 | 37.65 | 57.13 | True | True | True | True | Sex-specific 1e-2 VUMC, sex-differential UKB |
| 0.003129 | 0.003386 | 0.2945 | 5.356 | 37.61 | 56.77 | True | True | True | True | Sex-specific 1e-2 VUMC, sex-differential UKB |
| 0.003129 | 0.003386 | 0.3387 | 5.622 | 37.65 | 57.14 | True | True | True | True | Sex-specific 1e-2 VUMC, sex-differential UKB |
| 0.003135 | 0.003392 | 0.3264 | 5.205 | 33.36 | 50.1 | True | True | True | True | Sex-specific 1e-2 VUMC, sex-differential UKB |
| 0.003136 | 0.003393 | 0.3229 | 5.213 | 33.34 | 50.07 | True | True | True | True | Sex-specific 1e-2 VUMC, sex-differential UKB |
| 0.003129 | 0.003386 | 0.3057 | 5.369 | 37.6 | 56.79 | True | True | True | True | Sex-specific 1e-2 VUMC, sex-differential UKB |
| 0.003129 | 0.003386 | 0.3502 | 5.545 | 37.61 | 57.1 | True | True | True | True | Sex-specific 1e-2 VUMC, sex-differential UKB |
| 0.003129 | 0.003386 | 0.3557 | 5.523 | 37.64 | 57.16 | True | True | True | True | Sex-specific 1e-2 VUMC, sex-differential UKB |
| 0.003129 | 0.003386 | 0.3226 | 5.318 | 37.57 | 56.82 | True | True | True | True | Sex-specific 1e-2 VUMC, sex-differential UKB |
| 0.003136 | 0.003393 | 0.3246 | 5.107 | 33.18 | 50.22 | True | True | True | True | Sex-specific 1e-2 VUMC, sex-differential UKB |
| 0.003129 | 0.003386 | 0.3453 | 5.506 | 37.63 | 57.15 | True | True | True | True | Sex-specific 1e-2 VUMC, sex-differential UKB |
| 0.003136 | 0.003393 | 0.3401 | 5.079 | 33.22 | 50.07 | True | True | True | True | Sex-specific 1e-2 VUMC, sex-differential UKB |
| 0.00313 | 0.003387 | 0.3713 | 5.525 | 37.56 | 57.11 | True | True | True | True | Sex-specific 1e-2 VUMC, sex-differential UKB |
| 0.00313 | 0.003387 | 0.3329 | 5.249 | 37.46 | 56.76 | True | True | True | True | Sex-specific 1e-2 VUMC, sex-differential UKB |


| SE_UKB_female | SE_UKB_male | LOG10P_VUMC_female | LOG10P_VUMC_male | LOG10P_UKB_female | LOG10P_UKB_male | female_effect_direction_replication | male_effect_direction_replication | sex_effect_comparison_replication | pvalue_effect_comparison_replication | replication_type |
|---|---|---|---|---|---|---|---|---|---|---|

| 0.00313 | 0.003387 | 0.3496 | 5.3 | 37.34 | 56.7 | True | True | True | True | Sex-specific 1e-2 VUMC, sex-differential UKB |
| 0.00313 | 0.003387 | 0.3855 | 5.552 | 37.38 | 57.03 | True | True | True | True | Sex-specific 1e-2 VUMC, sex-differential UKB |
| 0.00313 | 0.003387 | 0.3426 | 5.287 | 37.33 | 56.7 | True | True | True | True | Sex-specific 1e-2 VUMC, sex-differential UKB |
| 0.00313 | 0.003387 | 0.3819 | 5.55 | 37.39 | 57.05 | True | True | True | True | Sex-specific 1e-2 VUMC, sex-differential UKB |
| 0.00313 | 0.003387 | 0.3341 | 5.265 | 37.32 | 56.67 | True | True | True | True | Sex-specific 1e-2 VUMC, sex-differential UKB |

| SE_UKB_female | SE_UKB_male | LOG10P_VUMC_female | LOG10P_VUMC_male | LOG10P_UKB_female | LOG10P_UKB_male | female_effect_direction_replication | male_effect_direction_replication | sex_effect_comparison_replication | pvalue_effect_comparison_replication | replication_type |
|---|---|---|---|---|---|---|---|---|---|---|
| 0.00313 | 0.003387 | 0.3754 | 5.534 | 37.34 | 57.01 | True | True | True | True | Sex-specific 1e-2 VUMC, sex-differential UKB |
| 0.00313 | 0.003387 | 0.3744 | 5.537 | 37.34 | 57.02 | True | True | True | True | Sex-specific 1e-2 VUMC, sex-differential UKB |
| 0.00313 | 0.003387 | 0.3127 | 5.202 | 37.31 | 56.66 | True | True | True | True | Sex-specific 1e-2 VUMC, sex-differential UKB |
| 0.00313 | 0.003387 | 0.3779 | 5.54 | 37.35 | 57.02 | True | True | True | True | Sex-specific 1e-2 VUMC, sex-differential UKB |
| 0.00313 | 0.003387 | 0.374 | 5.557 | 37.36 | 57 | True | True | True | True | Sex-specific 1e-2 VUMC, sex-differential UKB |
| 0.00313 | 0.003387 | 0.3294 | 5.295 | 37.34 | 56.66 | True | True | True | True | Sex-specific 1e-2 VUMC, sex-differential UKB |
| 0.00313 | 0.003387 | 0.3309 | 5.296 | 37.42 | 56.66 | True | True | True | True | Sex-specific 1e-2 VUMC, sex-differential UKB |
| 0.00313 | 0.003387 | 0.3347 | 5.29 | 37.41 | 56.67 | True | True | True | True | Sex-specific 1e-2 VUMC, sex-differential UKB |
| 0.00313 | 0.003387 | 0.3459 | 5.352 | 37.32 | 56.65 | True | True | True | True | Sex-specific 1e-2 VUMC, sex-differential UKB |
| 0.00313 | 0.003387 | 0.3835 | 5.564 | 37.36 | 56.99 | True | True | True | True | Sex-specific 1e-2 VUMC, sex-differential UKB |
| 0.003137 | 0.003395 | 0.3242 | 5.301 | 36.79 | 55.86 | True | True | True | True | Sex-specific 1e-2 VUMC, sex-differential UKB |
| 0.004152 | 0.004507 | 8.002 | 1.008 | 35.55 | 25.64 | True | True | True | True | Sex-specific 1e-2 VUMC, sex-differential UKB |
| 0.004174 | 0.004529 | 7.339 | 0.8295 | 34.32 | 25.62 | True | True | True | True | Sex-specific 1e-2 VUMC, sex-differential UKB |
| 0.004086 | 0.004435 | 6.356 | 0.2345 | 20.69 | 14.73 | True | True | True | True | Sex-specific 1e-2 VUMC, sex-differential UKB |
| 0.004085 | 0.004434 | 6.3 | 0.2379 | 20.69 | 14.78 | True | True | True | True | Sex-specific 1e-2 VUMC, sex-differential UKB |
| 0.004084 | 0.004432 | 6.288 | 0.2485 | 20.86 | 14.7 | True | True | True | True | Sex-specific 1e-2 VUMC, sex-differential UKB |
| 0.004081 | 0.00443 | 6.519 | 0.2876 | 20.72 | 15.02 | True | True | True | True | Sex-specific 1e-2 VUMC, sex-differential UKB |
| 0.004084 | 0.004433 | 6.359 | 0.2438 | 21.31 | 14.52 | True | True | True | True | Sex-specific 1e-2 VUMC, sex-differential UKB |
| 0.004071 | 0.004421 | 6.586 | 0.2687 | 20.68 | 14.89 | True | True | True | True | Sex-specific 1e-2 VUMC, sex-differential UKB |
| 0.004072 | 0.004422 | 6.571 | 0.2798 | 20.95 | 14.9 | True | True | True | True | Sex-specific 1e-2 VUMC, sex-differential UKB |
| 0.004068 | 0.004417 | 6.828 | 0.2927 | 20.96 | 14.87 | True | True | True | True | Sex-specific 1e-2 VUMC, sex-differential UKB |
| 0.004057 | 0.004406 | 6.823 | 0.4027 | 20.92 | 14.3 | True | True | True | True | Sex-specific 1e-2 VUMC, sex-differential UKB |
| 0.004059 | 0.004409 | 6.757 | 0.37 | 20.8 | 14.33 | True | True | True | True | Sex-specific 1e-2 VUMC, sex-differential UKB |
| 0.004056 | 0.004406 | 6.555 | 0.3876 | 21.06 | 14.48 | True | True | True | True | Sex-specific 1e-2 VUMC, sex-differential UKB |
| 0.004059 | 0.004408 | 6.512 | 0.3823 | 21.09 | 14.49 | True | True | True | True | Sex-specific 1e-2 VUMC, sex-differential UKB |
| 0.003642 | 0.003948 | 6.815 | 0.5683 | 15.22 | 10.53 | True | True | True | True | Sex-specific 1e-2 VUMC, sex-differential UKB |

| SE_UKB_female | SE_UKB_male | LOG10P_VUMC_female | LOG10P_VUMC_male | LOG10P_UKB_female | LOG10P_UKB_male | female_effect_direction_replication | male_effect_direction_replication | sex_effect_comparison_replication | pvalue_effect_comparison_replication | replication_type |
|---|---|---|---|---|---|---|---|---|---|---|
| 0.004084 | 0.004434 | 6.756 | 0.3315 | 21.75 | 14.7 | True | True | True | True | Sex-specific 1e-2 VUMC, sex-differential UKB |
| 0.004084 | 0.004434 | 6.756 | 0.3315 | 21.75 | 14.7 | True | True | True | True | Sex-specific 1e-2 VUMC, sex-differential UKB |
| 0.003532 | 0.003829 | 6.855 | 0.7847 | 14.23 | 7.847 | True | True | True | True | Sex-specific 1e-2 VUMC, sex-differential UKB |
| 0.003331 | 0.003607 | 6.601 | 1.229 | 15.01 | 8.998 | True | True | True | True | Sex-specific 1e-2 VUMC, sex-differential UKB |
| 0.003522 | 0.003817 | 7.171 | 0.9193 | 15.15 | 8.266 | True | True | True | True | Sex-specific 1e-2 VUMC, sex-differential UKB |
| 0.003521 | 0.003817 | 7.062 | 0.9443 | 15.23 | 8.354 | True | True | True | True | Sex-specific 1e-2 VUMC, sex-differential UKB |
| 0.003405 | 0.003688 | 6.848 | 0.9566 | 16.28 | 11.3 | True | True | True | True | Sex-specific 1e-2 VUMC, sex-differential UKB |
| 0.004084 | 0.004434 | 6.899 | 0.5104 | 22.48 | 15.08 | True | True | True | True | Sex-specific 1e-2 VUMC, sex-differential UKB |
| 0.003406 | 0.003688 | 6.922 | 1.051 | 16.2 | 11.34 | True | True | True | True | Sex-specific 1e-2 VUMC, sex-differential UKB |
| 0.003331 | 0.003606 | 6.629 | 1.302 | 15.11 | 8.873 | True | True | True | True | Sex-specific 1e-2 VUMC, sex-differential UKB |
| 0.003406 | 0.003688 | 6.619 | 1.012 | 16.12 | 11.4 | True | True | True | True | Sex-specific 1e-2 VUMC, sex-differential UKB |
| 0.003634 | 0.003935 | 7.959 | 1.059 | 15.71 | 10.92 | True | True | True | True | Sex-specific 1e-2 VUMC, sex-differential UKB |
| 0.004041 | 0.004382 | 7.595 | 0.8448 | 21.96 | 14.52 | True | True | True | True | Sex-specific 1e-2 VUMC, sex-differential UKB |
| 0.003635 | 0.003935 | 7.978 | 1.074 | 15.88 | 10.91 | True | True | True | True | Sex-specific 1e-2 VUMC, sex-differential UKB |
| 0.004041 | 0.004382 | 7.567 | 0.8647 | 21.95 | 14.54 | True | True | True | True | Sex-specific 1e-2 VUMC, sex-differential UKB |
| 0.004038 | 0.004377 | 7.577 | 0.8745 | 22.09 | 14.6 | True | True | True | True | Sex-specific 1e-2 VUMC, sex-differential UKB |
| 0.006153 | 0.006654 | 5.887 | 0.07485 | 1.021 | 2.744 | True | True | False | False | |
| 0.005728 | 0.006191 | 8.662 | 0.5415 | 7.996 | 9.49 | True | True | False | False | |
| 0.004899 | 0.005318 | 7.475 | 0.2543 | 1.563 | 4.843 | True | True | False | False | |
| 0.004882 | 0.005298 | 7.092 | 0.2201 | 1.606 | 4.711 | True | True | False | False | |
| 0.005156 | 0.005595 | 6.619 | 0.5595 | 0.9557 | 4.365 | True | True | False | False | |
| 0.004869 | 0.005283 | 7.292 | 0.2313 | 1.657 | 4.613 | True | True | False | False | |
| 0.004882 | 0.005299 | 6.882 | 0.228 | 1.632 | 4.41 | True | True | False | False | |
| 0.004894 | 0.005313 | 7.06 | 0.2637 | 1.632 | 4.77 | True | True | False | False | |
| 0.004895 | 0.005311 | 7.2 | 0.2505 | 1.551 | 4.767 | True | True | False | False | |
| 0.004903 | 0.005323 | 7.16 | 0.284 | 1.63 | 4.736 | True | True | False | False | |

| SE_UKB_female | SE_UKB_male | LOG10P_VUMC_female | LOG10P_VUMC_male | LOG10P_UKB_female | LOG10P_UKB_male | female_effect_direction_replication | male_effect_direction_replication | sex_effect_comparison_replication | pvalue_effect_comparison_replication | replication_type |
|---|---|---|---|---|---|---|---|---|---|---|
| 0.004877 | 0.005292 | 7.201 | 0.2428 | 1.662 | 4.736 | True | True | False | False | |
| 0.004882 | 0.005297 | 6.996 | 0.2055 | 1.67 | 4.836 | True | True | False | False | |
| 0.004875 | 0.005289 | 7.293 | 0.2521 | 1.656 | 4.908 | True | True | False | False | |
| SE_UKB_female | SE_UKB_male | LOG10P_VUMC_female | LOG10P_VUMC_male | LOG10P_UKB_female | LOG10P_UKB_male | female_effect_direction_replication | male_effect_direction_replication | sex_effect_comparison_replication | pvalue_effect_comparison_replication | replication_type |
| 0.004904 | 0.005323 | 7.216 | 0.3061 | 1.631 | 4.808 | True | True | False | False | |
| 0.005196 | 0.005638 | 7.344 | 0.614 | 0.8876 | 4.188 | True | True | False | False | |
| 0.004898 | 0.005315 | 7.417 | 0.2773 | 1.599 | 4.958 | True | True | False | False | |
| 0.004906 | 0.005323 | 7.017 | 0.2535 | 1.649 | 4.748 | True | True | False | False | |
| 0.004903 | 0.00532 | 7.288 | 0.301 | 2.081 | 4.767 | True | True | False | False | |
| 0.004883 | 0.005296 | 7.229 | 0.2295 | 1.759 | 4.999 | True | True | False | False | |
| 0.004884 | 0.005298 | 7.285 | 0.2293 | 1.768 | 4.965 | True | True | False | False | |
| 0.005072 | 0.005512 | 8.382 | 0.1932 | 1.961 | 5.767 | True | True | False | False | |
| 0.00507 | 0.00551 | 8.454 | 0.2281 | 2.016 | 5.779 | True | True | False | False | |
| 0.004979 | 0.005408 | 8.375 | 0.4373 | 1.52 | 5.206 | True | True | False | False | |
| 0.004981 | 0.00541 | 8.414 | 0.4075 | 1.507 | 5.064 | True | True | False | False | |
| 0.004982 | 0.005411 | 8.376 | 0.403 | 1.486 | 5.068 | True | True | False | False | |
| 0.004973 | 0.005401 | 8.315 | 0.4262 | 1.554 | 5.249 | True | True | False | False | |
| 0.004975 | 0.005404 | 8.198 | 0.4134 | 1.573 | 5.237 | True | True | False | False | |
| 0.005056 | 0.005494 | 8.149 | 0.2099 | 2.026 | 5.827 | True | True | False | False | |
| 0.005063 | 0.005504 | 8.405 | 0.1741 | 2.032 | 5.866 | True | True | False | False | |
| 0.005212 | 0.005659 | 8.759 | 0.6297 | 0.7913 | 4.514 | True | True | False | False | |
| 0.005056 | 0.005494 | 8.242 | 0.2105 | 2.04 | 5.796 | True | True | False | False | |
| 0.005049 | 0.005491 | 8.356 | 0.2002 | 2.086 | 5.767 | True | True | False | False | |
| 0.005056 | 0.005494 | 8.116 | 0.1928 | 2.005 | 5.846 | True | True | False | False | |
| 0.005064 | 0.005504 | 8.388 | 0.1846 | 2 | 5.892 | True | True | False | False | |
| 0.005053 | 0.005491 | 8.371 | 0.2389 | 1.993 | 5.751 | True | True | False | False | |
| 0.00505 | 0.005489 | 8.192 | 0.2376 | 2.027 | 5.675 | True | True | False | False | |

| SE_UKB_female | SE_UKB_male | LOG10P_VUMC_female | LOG10P_VUMC_male | LOG10P_UKB_female | LOG10P_UKB_male | female_effect_direction_replication | male_effect_direction_replication | sex_effect_comparison_replication | pvalue_effect_comparison_replication | replication_type |
|---|---|---|---|---|---|---|---|---|---|---|
| 0.005676 | 0.006125 | 6.742 | 0.4605 | 10.74 | 11.1 | True | True | False | False | |
| 0.004978 | 0.005407 | 8.11 | 0.3675 | 1.568 | 5.244 | True | True | False | False | |
| 0.005064 | 0.005504 | 8.511 | 0.2161 | 2.009 | 5.829 | True | True | False | False | |
| 0.005077 | 0.005518 | 8.201 | 0.1656 | 2.143 | 5.836 | True | True | False | False | |
| 0.005983 | 0.006461 | 6.846 | 0.5699 | 9.462 | 8.13 | True | True | True | True | Sex-specific 1e-2 VUMC, sex-differential UKB |
| 0.005073 | 0.005513 | 8.356 | 0.1997 | 2.188 | 5.756 | True | True | False | False | |
| 0.004982 | 0.00541 | 8.272 | 0.323 | 1.696 | 5.255 | True | True | False | False | |
| 0.005074 | 0.005515 | 8.299 | 0.162 | 2.151 | 5.872 | True | True | False | False | |
| 0.005074 | 0.005515 | 8.295 | 0.1625 | 2.147 | 5.869 | True | True | False | False | |
| 0.004985 | 0.005413 | 8.088 | 0.3397 | 1.657 | 5.272 | True | True | False | False | |
| 0.005077 | 0.005518 | 7.991 | 0.1624 | 2.092 | 5.894 | True | True | False | False | |
| 0.005078 | 0.005519 | 7.847 | 0.1225 | 2.143 | 5.891 | True | True | False | False | |
| 0.005078 | 0.005519 | 7.918 | 0.1339 | 2.111 | 5.934 | True | True | False | False | |
| 0.00508 | 0.005522 | 7.863 | 0.1377 | 2.106 | 5.859 | True | True | False | False | |
| 0.005079 | 0.00552 | 7.905 | 0.1338 | 2.116 | 5.934 | True | True | False | False | |
| 0.005079 | 0.00552 | 7.878 | 0.133 | 2.13 | 5.97 | True | True | False | False | |
| 0.004278 | 0.004649 | 2.996 | 0.06735 | 1.135 | 3.755 | True | True | False | False | |
| 0.004997 | 0.005425 | 7.444 | 0.2959 | 1.645 | 5.479 | True | True | False | False | |
| 0.0051 | 0.005541 | 7.464 | 0.1144 | 2.172 | 5.993 | True | True | False | False | |
| 0.003928 | 0.004279 | 3.64 | 1.528 | 11.48 | 10.9 | True | True | False | True | |
| 0.003928 | 0.004279 | 3.663 | 1.573 | 11.5 | 10.57 | True | True | False | True | |
| 0.003929 | 0.00428 | 3.757 | 1.519 | 11.38 | 10.65 | True | True | False | True | |
| 0.003496 | 0.003796 | 9.126 | 1.712 | 18.06 | 16.81 | True | True | False | True | |
| 0.003497 | 0.003795 | 9.214 | 1.698 | 18.15 | 16.86 | True | True | False | True | |
| 0.004411 | 0.004779 | 10.4 | 1.898 | 26.26 | 16.25 | True | True | True | True | Sex-specific 1e-2 VUMC, sex-differential UKB |
| 0.00434 | 0.004703 | 10.73 | 1.708 | 27.08 | 16.12 | True | True | True | True | Sex-specific 1e-2 VUMC, sex-differential UKB |
| 0.003543 | 0.003836 | 2.668 | 1.711 | 17.47 | 8.605 | True | True | True | True | Sex-specific 1e-2 VUMC, sex-differential UKB |
| 0.003543 | 0.003836 | 2.694 | 1.734 | 17.53 | 8.539 | True | True | True | True | Sex-specific 1e-2 VUMC, sex-differential UKB |

| SE_UKB_female | SE_UKB_male | LOG10P_VUMC_female | LOG10P_VUMC_male | LOG10P_UKB_female | LOG10P_UKB_male | female_effect_direction_replication | male_effect_direction_replication | sex_effect_comparison_replication | pvalue_effect_comparison_replication | replication_type |
|---|---|---|---|---|---|---|---|---|---|---|
| 0.003542 | 0.003834 | 2.569 | 1.602 | 17.78 | 8.363 | True | True | True | True | Sex-specific 1e-2 VUMC, sex-differential UKB |
| 0.004316 | 0.004678 | 9.681 | 1.618 | 29.09 | 15.08 | True | True | True | True | Sex-specific 1e-2 VUMC, sex-differential UKB |
| 0.003541 | 0.003834 | 2.548 | 1.601 | 17.75 | 8.314 | True | True | True | True | Sex-specific 1e-2 VUMC, sex-differential UKB |
| 0.003512 | 0.003801 | 2.113 | 1.306 | 18.25 | 6.485 | True | True | True | True | Sex-specific 1e-2 VUMC, sex-specific 5e-8 UKB |
| 0.0052 | 0.005515 | 0.4099 | 7.211 | 1.147 | 0.5994 | True | True | False | False | |
| 0.005198 | 0.005513 | 0.4027 | 7.265 | 1.14 | 0.6098 | True | True | False | False | |
| 0.005313 | 0.00564 | 0.3574 | 6.199 | 0.9096 | 0.5366 | True | True | False | False | |
| 0.005092 | 0.005408 | 0.3143 | 7.016 | 0.8927 | 0.7443 | True | True | False | False | |
| 0.006872 | 0.007278 | 0.4572 | 5.808 | 1.624 | 0.1464 | False | True | False | False | |

| SE_UKB_female | SE_UKB_male | LOG10P_VUMC_female | LOG10P_VUMC_male | LOG10P_UKB_female | LOG10P_UKB_male | female_effect_direction_replication | male_effect_direction_replication | sex_effect_comparison_replication | pvalue_effect_comparison_replication | replication_type |
|---|---|---|---|---|---|---|---|---|---|---|
| 0.005089 | 0.005405 | 0.3127 | 7.082 | 0.8753 | 0.7626 | True | True | False | False | |
| 0.005204 | 0.005521 | 0.3837 | 7.073 | 1.105 | 0.6197 | True | True | False | False | |
| 0.005089 | 0.005405 | 0.3185 | 7.012 | 0.8846 | 0.7562 | True | True | False | False | |
| 0.005202 | 0.005519 | 0.3921 | 7.069 | 1.115 | 0.6117 | True | True | False | False | |
| 0.005084 | 0.0054 | 0.3213 | 7.062 | 0.8953 | 0.7636 | True | True | False | False | |
| 0.005096 | 0.005413 | 0.3081 | 6.956 | 0.8776 | 0.7371 | True | True | False | False | |
| 0.005087 | 0.005402 | 0.3169 | 7.312 | 0.9173 | 0.7443 | True | True | False | False | |
| 0.005113 | 0.005429 | 0.2332 | 7.69 | 1.071 | 0.7104 | True | True | False | False | |
| 0.005116 | 0.005432 | 0.2997 | 7.714 | 1.037 | 0.6987 | True | True | False | False | |
| 0.005109 | 0.005426 | 0.2676 | 7.455 | 1.061 | 0.6953 | True | True | False | False | |
| 0.005125 | 0.005444 | 0.2412 | 7.269 | 1.039 | 0.6957 | True | True | False | False | |
| 0.005254 | 0.005573 | 0.2036 | 7.3 | 1.116 | 0.6538 | True | True | False | False | |
| 0.003469 | 0.003683 | 6.063 | 0.4547 | 13.67 | 10.19 | True | True | True | True | Sex-specific 1e-2 VUMC, sex-differential UKB |
| 0.003759 | 0.003994 | 7.423 | 0.9701 | 18.85 | 17.09 | True | True | False | True | |
| 0.003753 | 0.003989 | 7.838 | 1.061 | 19.32 | 17.12 | True | True | True | True | Sex-specific 1e-2 VUMC, sex-differential UKB |
| 0.003753 | 0.003989 | 7.802 | 1.046 | 19.34 | 17.16 | True | True | True | True | Sex-specific 1e-2 VUMC, sex-differential UKB |
| 0.003751 | 0.003985 | 7.237 | 1.102 | 20.09 | 16.72 | True | True | True | True | Sex-specific 1e-2 VUMC, sex-differential UKB |

| SE_UKB_female | SE_UKB_male | LOG10P_VUMC_female | LOG10P_VUMC_male | LOG10P_UKB_female | LOG10P_UKB_male | female_effect_direction_replication | male_effect_direction_replication | sex_effect_comparison_replication | pvalue_effect_comparison_replication | replication_type |
|---|---|---|---|---|---|---|---|---|---|---|
| 0.003618 | 0.003839 | 5.442 | 1.17 | 16.17 | 13.91 | True | True | True | True | Sex-specific 1e-2 VUMC, sex-differential UKB |
| 0.005859 | 0.006276 | 9.653 | 0.7979 | 55.15 | 47.38 | True | True | True | True | Sex-specific 1e-2 VUMC, sex-differential UKB |
| 0.005859 | 0.006276 | 9.629 | 0.8081 | 55.16 | 47.37 | True | True | True | True | Sex-specific 1e-2 VUMC, sex-differential UKB |
| 0.005903 | 0.006327 | 10.54 | 0.8843 | 63.95 | 52.17 | True | True | True | True | Sex-specific 1e-2 VUMC, sex-differential UKB |
| 0.005892 | 0.006314 | 10.76 | 0.8396 | 63.89 | 52.24 | True | True | True | True | Sex-specific 1e-2 VUMC, sex-differential UKB |
| 0.005902 | 0.006325 | 10.57 | 0.8634 | 64 | 52.32 | True | True | True | True | Sex-specific 1e-2 VUMC, sex-differential UKB |
| 0.005891 | 0.006312 | 10.71 | 0.7971 | 63.96 | 52.4 | True | True | True | True | Sex-specific 1e-2 VUMC, sex-differential UKB |
| 0.005892 | 0.006313 | 10.76 | 0.8118 | 63.97 | 52.44 | True | True | True | True | Sex-specific 1e-2 VUMC, sex-differential UKB |
| 0.005899 | 0.006321 | 10.74 | 0.797 | 64.29 | 52.42 | True | True | True | True | Sex-specific 1e-2 VUMC, sex-differential UKB |
| 0.005891 | 0.006313 | 10.67 | 0.7691 | 64.08 | 52.44 | True | True | True | True | Sex-specific 1e-2 VUMC, sex-differential UKB |
| 0.005123 | 0.005487 | 7.427 | 0.5966 | 43.78 | 39.76 | True | True | False | True | |
| 0.005874 | 0.006296 | 10.88 | 0.9809 | 63.61 | 52.59 | True | True | True | True | Sex-specific 1e-2 VUMC, sex-differential UKB |
| 0.005164 | 0.005529 | 7.123 | 0.6164 | 43.1 | 39.72 | True | True | False | True | |
| 0.005874 | 0.006295 | 10.86 | 0.9925 | 63.7 | 52.58 | True | True | True | True | Sex-specific 1e-2 VUMC, sex-differential UKB |
| 0.005129 | 0.005491 | 7.418 | 0.5951 | 43.73 | 40.45 | True | True | False | True | |
| 0.005874 | 0.006295 | 10.87 | 0.9843 | 63.62 | 52.6 | True | True | True | True | Sex-specific 1e-2 VUMC, sex-differential UKB |
| 0.005123 | 0.005487 | 7.353 | 0.6505 | 43.82 | 39.75 | True | True | False | True | |
| 0.005123 | 0.005487 | 7.337 | 0.6499 | 43.88 | 39.74 | True | True | False | True | |
| 0.005873 | 0.006295 | 10.96 | 0.9623 | 63.67 | 52.69 | True | True | True | True | Sex-specific 1e-2 VUMC, sex-differential UKB |
| 0.005873 | 0.006291 | 10.96 | 0.9623 | 63.66 | 53.4 | True | True | True | True | Sex-specific 1e-2 VUMC, sex-differential UKB |
| 0.005879 | 0.006302 | 11.05 | 0.9906 | 64.09 | 52.86 | True | True | True | True | Sex-specific 1e-2 VUMC, sex-differential UKB |
| 0.003494 | 0.003713 | 9.735 | 0.4616 | 7.047 | 0.4518 | True | True | True | True | Sex-specific 1e-2 VUMC, sex-differential UKB |
| 0.003537 | 0.003761 | 10.17 | 0.5709 | 6.849 | 0.7956 | True | True | True | True | Sex-specific 1e-2 VUMC, sex-differential UKB |
| 0.003527 | 0.003749 | 10.97 | 0.4849 | 6.686 | 0.7987 | True | True | True | True | Sex-specific 1e-2 VUMC, sex-differential UKB |
| 0.003118 | 0.003515 | 9.664 | 1.684 | 20.14 | 7.801 | True | True | True | True | Sex-specific 1e-2 VUMC, sex-differential UKB |
| 0.003117 | 0.003514 | 9.636 | 1.785 | 20.03 | 7.88 | True | True | True | True | Sex-specific 1e-2 VUMC, sex-differential UKB |

| SE_UKB_female | SE_UKB_male | LOG10P_VUMC_female | LOG10P_VUMC_male | LOG10P_UKB_female | LOG10P_UKB_male | female_effect_direction_replication | male_effect_direction_replication | sex_effect_comparison_replication | pvalue_effect_comparison_replication | replication_type |
|---|---|---|---|---|---|---|---|---|---|---|
| 0.003064 | 0.003458 | 8.836 | 0.7383 | 17.98 | 5.179 | True | True | True | True | Sex-specific 1e-2 VUMC, sex-specific 5e-8 UKB |
| 0.003123 | 0.003521 | 9.29 | 1.799 | 20.09 | 7.703 | True | True | True | True | Sex-specific 1e-2 VUMC, sex-differential UKB |
| 0.003117 | 0.003514 | 9.79 | 1.836 | 20.12 | 7.973 | True | True | True | True | Sex-specific 1e-2 VUMC, sex-differential UKB |
| 0.003064 | 0.003458 | 9.313 | 0.7219 | 17.79 | 5.127 | True | True | True | True | Sex-specific 1e-2 VUMC, sex-specific 5e-8 UKB |
| 0.003064 | 0.003457 | 9.207 | 0.7296 | 18.08 | 5.019 | True | True | True | True | Sex-specific 1e-2 VUMC, sex-specific 5e-8 UKB |
| 0.003059 | 0.00345 | 9.092 | 0.2424 | 17.32 | 4.174 | True | True | True | True | Sex-specific 1e-2 VUMC, sex-specific 5e-8 UKB |
| 0.003101 | 0.003497 | 9.963 | 1.657 | 18.63 | 7.826 | True | True | True | True | Sex-specific 1e-2 VUMC, sex-differential UKB |
| 0.004107 | 0.004436 | 3.559 | 0.7714 | 23.67 | 11.27 | True | True | True | True | Sex-specific 1e-2 VUMC, sex-differential UKB |
| 0.004106 | 0.004435 | 3.378 | 0.6902 | 22.48 | 10.97 | True | True | True | True | Sex-specific 1e-2 VUMC, sex-differential UKB |
| 0.004026 | 0.004346 | 2.941 | 0.8373 | 24.97 | 10.9 | True | True | True | True | Sex-specific 1e-2 VUMC, sex-differential UKB |
| 0.004023 | 0.004343 | 2.847 | 0.8391 | 24.7 | 10.73 | True | True | True | True | Sex-specific 1e-2 VUMC, sex-differential UKB |
| 0.004024 | 0.004344 | 2.855 | 0.8406 | 24.56 | 10.49 | True | True | True | True | Sex-specific 1e-2 VUMC, sex-differential UKB |
| 0.004637 | 0.005009 | 7.38 | 1.6 | 49.6 | 26.34 | True | True | True | True | Sex-specific 1e-2 VUMC, sex-differential UKB |
| 0.004025 | 0.004345 | 2.855 | 0.816 | 24.61 | 10.52 | True | True | True | True | Sex-specific 1e-2 VUMC, sex-differential UKB |
| 0.004577 | 0.004943 | 7.011 | 1.334 | 44.21 | 22.94 | True | True | True | True | Sex-specific 1e-2 VUMC, sex-differential UKB |
| 0.00402 | 0.004341 | 2.742 | 0.668 | 23.52 | 9.853 | True | True | True | True | Sex-specific 1e-2 VUMC, sex-differential UKB |
| 0.004026 | 0.004347 | 2.925 | 0.8183 | 24.46 | 10.38 | True | True | True | True | Sex-specific 1e-2 VUMC, sex-differential UKB |
| SE_UKB_female | SE_UKB_male | LOG10P_VUMC_female | LOG10P_VUMC_male | LOG10P_UKB_female | LOG10P_UKB_male | female_effect_direction_replication | male_effect_direction_replication | sex_effect_comparison_replication | pvalue_effect_comparison_replication | replication_type |
| 0.004026 | 0.004347 | 2.945 | 0.8183 | 24.47 | 10.38 | True | True | True | True | Sex-specific 1e-2 VUMC, sex-differential UKB |
| 0.00403 | 0.004352 | 2.919 | 0.8161 | 23.35 | 9.775 | True | True | True | True | Sex-specific 1e-2 VUMC, sex-differential UKB |
| 0.004543 | 0.004912 | 6.589 | 0.7797 | 40.41 | 19.82 | True | True | True | True | Sex-specific 1e-2 VUMC, sex-differential UKB |
| 0.00455 | 0.00492 | 6.826 | 0.9817 | 42.08 | 20.94 | True | True | True | True | Sex-specific 1e-2 VUMC, sex-differential UKB |
| 0.004542 | 0.004912 | 6.585 | 0.7745 | 40.4 | 19.81 | True | True | True | True | Sex-specific 1e-2 VUMC, sex-differential UKB |
| 0.004542 | 0.004911 | 6.578 | 0.7736 | 40.32 | 19.78 | True | True | True | True | Sex-specific 1e-2 VUMC, sex-differential UKB |
| 0.004548 | 0.004917 | 6.814 | 0.9833 | 41.83 | 20.81 | True | True | True | True | Sex-specific 1e-2 VUMC, sex-differential UKB |
| 0.004548 | 0.004917 | 6.815 | 0.9833 | 41.83 | 20.81 | True | True | True | True | Sex-specific 1e-2 VUMC, sex-differential UKB |

| SE_UKB_female | SE_UKB_male | LOG10P_VUMC_female | LOG10P_VUMC_male | LOG10P_UKB_female | LOG10P_UKB_male | female_effect_direction_replication | male_effect_direction_replication | sex_effect_comparison_replication | pvalue_effect_comparison_replication | replication_type |
|---|---|---|---|---|---|---|---|---|---|---|
| 0.004548 | 0.004917 | 6.781 | 0.9857 | 41.82 | 20.8 | True | True | True | True | Sex-specific 1e-2 VUMC, sex-differential UKB |
| 0.004548 | 0.004917 | 6.765 | 0.9855 | 41.8 | 20.8 | True | True | True | True | Sex-specific 1e-2 VUMC, sex-differential UKB |
| 0.00454 | 0.00491 | 6.521 | 0.774 | 40.17 | 19.66 | True | True | True | True | Sex-specific 1e-2 VUMC, sex-differential UKB |
| 0.004548 | 0.004917 | 6.765 | 0.9844 | 41.85 | 20.8 | True | True | True | True | Sex-specific 1e-2 VUMC, sex-differential UKB |
| 0.004548 | 0.004917 | 6.758 | 0.9853 | 41.84 | 20.79 | True | True | True | True | Sex-specific 1e-2 VUMC, sex-differential UKB |
| 0.004548 | 0.004917 | 6.756 | 0.9853 | 41.84 | 20.79 | True | True | True | True | Sex-specific 1e-2 VUMC, sex-differential UKB |
| 0.004547 | 0.004916 | 6.739 | 0.9857 | 41.92 | 20.86 | True | True | True | True | Sex-specific 1e-2 VUMC, sex-differential UKB |
| 0.004547 | 0.004916 | 6.753 | 0.9982 | 41.66 | 20.69 | True | True | True | True | Sex-specific 1e-2 VUMC, sex-differential UKB |
| 0.004541 | 0.00491 | 6.497 | 0.7775 | 40.04 | 19.62 | True | True | True | True | Sex-specific 1e-2 VUMC, sex-differential UKB |
| 0.004537 | 0.004907 | 6.669 | 0.8086 | 40.28 | 19.66 | True | True | True | True | Sex-specific 1e-2 VUMC, sex-differential UKB |
| 0.004498 | 0.004864 | 6.05 | 0.8956 | 37.23 | 17.6 | True | True | True | True | Sex-specific 1e-2 VUMC, sex-differential UKB |
| 0.004498 | 0.004864 | 6.176 | 0.8718 | 37.58 | 17.78 | True | True | True | True | Sex-specific 1e-2 VUMC, sex-differential UKB |
| 0.004247 | 0.004548 | 9.401 | 0.8578 | 5.216 | 10.29 | True | True | False | False | |
| 0.004381 | 0.004691 | 6.449 | 1.347 | 2.585 | 8.179 | True | True | False | False | |
| 0.004958 | 0.005302 | 6.37 | 0.273 | 3.291 | 7.065 | True | True | False | False | |
| 0.006356 | 0.006836 | 0.4261 | 7.358 | 0.07201 | 0.3471 | False | True | True | True | Sex-specific 1e-2 VUMC, sex-differential UKB |
| 0.006236 | 0.006717 | 0.4635 | 6.929 | 0.04833 | 0.2788 | True | True | True | True | Sex-specific 1e-2 VUMC, sex-differential UKB |
| 0.006909 | 0.007459 | 0.2494 | 2.723 | 0.01735 | 1.303 | False | True | True | True | Sex-specific 1e-2 VUMC, sex-differential UKB |
| 0.006292 | 0.006797 | 0.01644 | 3.576 | 0.2769 | 0.7823 | True | True | True | True | Sex-specific 1e-2 VUMC, sex-differential UKB |
| 0.006319 | 0.006831 | 0.08013 | 3.638 | 0.2192 | 0.7553 | True | True | True | True | Sex-specific 1e-2 VUMC, sex-differential UKB |
| 0.006319 | 0.006829 | 0.0257 | 3.624 | 0.2121 | 0.7948 | True | True | True | True | Sex-specific 1e-2 VUMC, sex-differential UKB |
| 0.006889 | 0.007436 | 0.2572 | 2.706 | 0.03239 | 1.312 | False | True | True | True | Sex-specific 1e-2 VUMC, sex-differential UKB |
| 0.006292 | 0.006798 | 0.02146 | 3.677 | 0.2982 | 0.785 | True | True | True | True | Sex-specific 1e-2 VUMC, sex-differential UKB |
| 0.006291 | 0.006797 | 0.03516 | 3.677 | 0.2944 | 0.7969 | True | True | True | True | Sex-specific 1e-2 VUMC, sex-differential UKB |
| 0.006875 | 0.007422 | 0.2466 | 2.989 | 0.07757 | 1.399 | False | True | True | True | Sex-specific 1e-2 VUMC, sex-differential UKB |
| 0.006869 | 0.007419 | 0.1957 | 3.029 | 0.05874 | 1.487 | False | True | True | True | Sex-specific 1e-2 VUMC, sex-differential UKB |

| SE_UKB_female | SE_UKB_male | LOG10P_VUMC_female | LOG10P_VUMC_male | LOG10P_UKB_female | LOG10P_UKB_male | female_effect_direction_replication | male_effect_direction_replication | sex_effect_comparison_replication | pvalue_effect_comparison_replication | replication_type |
|---|---|---|---|---|---|---|---|---|---|---|
| 0.003225 | 0.003472 | 4.491 | 0.9712 | 1.764 | 0.3083 | True | True | True | True | Sex-specific 1e-2 VUMC, sex-differential UKB |
| 0.003225 | 0.003472 | 4.485 | 0.9457 | 1.774 | 0.3131 | True | True | True | True | Sex-specific 1e-2 VUMC, sex-differential UKB |
| 0.003225 | 0.003471 | 4.479 | 0.946 | 1.79 | 0.2798 | True | True | True | True | Sex-specific 1e-2 VUMC, sex-differential UKB |
| 0.003225 | 0.003472 | 4.455 | 0.9465 | 1.722 | 0.2774 | True | True | True | True | Sex-specific 1e-2 VUMC, sex-differential UKB |
| 0.003235 | 0.003482 | 5.193 | 0.4434 | 1.702 | 0.3783 | True | True | True | True | Sex-specific 1e-2 VUMC, sex-differential UKB |
| 0.003236 | 0.003483 | 5.358 | 0.4774 | 1.63 | 0.3927 | True | True | True | True | Sex-specific 1e-2 VUMC, sex-differential UKB |
| 0.003242 | 0.003489 | 5.36 | 0.4548 | 1.672 | 0.415 | True | True | True | True | Sex-specific 1e-2 VUMC, sex-differential UKB |
| 0.003242 | 0.003489 | 5.426 | 0.4605 | 1.704 | 0.4124 | True | True | True | True | Sex-specific 1e-2 VUMC, sex-differential UKB |
| 0.003236 | 0.003481 | 5.32 | 0.6432 | 1.63 | 0.4214 | True | True | True | True | Sex-specific 1e-2 VUMC, sex-differential UKB |
| 0.003236 | 0.003481 | 5.32 | 0.644 | 1.631 | 0.424 | True | True | True | True | Sex-specific 1e-2 VUMC, sex-differential UKB |
| | | 5.32 | 0.644 | | | False | False | False | False | |
| 0.003236 | 0.003481 | 5.34 | 0.5371 | 1.608 | 0.4102 | True | True | True | True | Sex-specific 1e-2 VUMC, sex-differential UKB |
| 0.00324 | 0.003485 | 5.274 | 0.5891 | 1.634 | 0.3875 | True | True | True | True | Sex-specific 1e-2 VUMC, sex-differential UKB |
| 0.00324 | 0.003485 | 5.261 | 0.5881 | 1.646 | 0.3883 | True | True | True | True | Sex-specific 1e-2 VUMC, sex-differential UKB |
| 0.00324 | 0.003485 | 5.282 | 0.5799 | 1.642 | 0.3931 | True | True | True | True | Sex-specific 1e-2 VUMC, sex-differential UKB |
| 0.003235 | 0.003481 | 5.231 | 0.5902 | 1.641 | 0.4136 | True | True | True | True | Sex-specific 1e-2 VUMC, sex-differential UKB |
| 0.003235 | 0.003481 | 5.337 | 0.5571 | 1.622 | 0.4229 | True | True | True | True | Sex-specific 1e-2 VUMC, sex-differential UKB |
| 0.003235 | 0.003481 | 5.344 | 0.5874 | 1.633 | 0.4205 | True | True | True | True | Sex-specific 1e-2 VUMC, sex-differential UKB |
| 0.003236 | 0.003481 | 5.384 | 0.5175 | 1.609 | 0.4047 | True | True | True | True | Sex-specific 1e-2 VUMC, sex-differential UKB |
| 0.003237 | 0.003482 | 5.284 | 0.5162 | 1.575 | 0.4083 | True | True | True | True | Sex-specific 1e-2 VUMC, sex-differential UKB |
| 0.003235 | 0.00348 | 5.469 | 0.5849 | 1.603 | 0.3911 | True | True | True | True | Sex-specific 1e-2 VUMC, sex-differential UKB |
| 0.003235 | 0.003481 | 5.489 | 0.5483 | 1.6 | 0.3773 | True | True | True | True | Sex-specific 1e-2 VUMC, sex-differential UKB |
| 0.003235 | 0.00348 | 5.944 | 0.608 | 1.601 | 0.4302 | True | True | True | True | Sex-specific 1e-2 VUMC, sex-differential UKB |
| 0.003234 | 0.003479 | 6.302 | 0.6076 | 1.827 | 0.4276 | True | True | True | True | Sex-specific 1e-2 VUMC, sex-differential UKB |
| 0.003303 | 0.003554 | 3.257 | 0.08427 | 0.978 | 1.08 | True | False | False | False | |
| 0.003305 | 0.003556 | 3.107 | 0.0734 | 0.9257 | 1.223 | True | False | False | False | |

| SE_UKB_female | SE_UKB_male | LOG10P_VUMC_female | LOG10P_VUMC_male | LOG10P_UKB_female | LOG10P_UKB_male | female_effect_direction_replication | male_effect_direction_replication | sex_effect_comparison_replication | pvalue_effect_comparison_replication | replication_type |
|---|---|---|---|---|---|---|---|---|---|---|
| 0.003305 | 0.003556 | 3.067 | 0.08185 | 0.9398 | 1.247 | True | False | False | False | |
| 0.003271 | 0.003513 | 7.25 | 0.5766 | 2.439 | 1.172 | True | True | True | True | Sex-specific 1e-2 VUMC, sex-differential UKB |
| 0.003271 | 0.003513 | 7.281 | 0.5764 | 2.445 | 1.174 | True | True | True | True | Sex-specific 1e-2 VUMC, sex-differential UKB |
| 0.003218 | 0.003465 | 7.26 | 0.5489 | 3.073 | 2.423 | True | True | True | True | Sex-specific 1e-2 VUMC, sex-differential UKB |
| | | 7.207 | 0.6594 | | | False | False | False | False | |
| 0.003217 | 0.003465 | 7.536 | 0.5075 | 3.175 | 2.353 | True | True | True | True | Sex-specific 1e-2 VUMC, sex-differential UKB |
| 0.003234 | 0.003484 | 7.424 | 0.5373 | 3.324 | 2.592 | True | True | True | True | Sex-specific 1e-2 VUMC, sex-differential UKB |
| 0.003234 | 0.003484 | 7.439 | 0.5168 | 3.268 | 2.523 | True | True | True | True | Sex-specific 1e-2 VUMC, sex-differential UKB |
| 0.003251 | 0.003493 | 7.42 | 0.8123 | 2.261 | 1.42 | True | True | True | True | Sex-specific 1e-2 VUMC, sex-differential UKB |
| 0.003216 | 0.003464 | 7.347 | 0.5548 | 3.071 | 2.428 | True | True | True | True | Sex-specific 1e-2 VUMC, sex-differential UKB |
| 0.003251 | 0.003492 | 7.315 | 0.7686 | 2.075 | 1.471 | True | True | True | True | Sex-specific 1e-2 VUMC, sex-differential UKB |
| 0.003251 | 0.003492 | 7.315 | 0.7489 | 2.039 | 1.475 | True | True | True | True | Sex-specific 1e-2 VUMC, sex-differential UKB |
| 0.003218 | 0.003466 | 7.126 | 0.6536 | 2.964 | 2.224 | True | True | True | True | Sex-specific 1e-2 VUMC, sex-differential UKB |
| 0.003255 | 0.003497 | 7.128 | 0.8535 | 2.006 | 1.345 | True | True | True | True | Sex-specific 1e-2 VUMC, sex-differential UKB |
| 0.003255 | 0.003497 | 7.127 | 0.851 | 2.002 | 1.352 | True | True | True | True | Sex-specific 1e-2 VUMC, sex-differential UKB |
| 0.003256 | 0.003497 | 7.073 | 0.8427 | 2.002 | 1.342 | True | True | True | True | Sex-specific 1e-2 VUMC, sex-differential UKB |
| 0.003256 | 0.003498 | 7.068 | 0.8419 | 2.006 | 1.334 | True | True | True | True | Sex-specific 1e-2 VUMC, sex-differential UKB |
| 0.003256 | 0.003498 | 7.085 | 0.8188 | 2.007 | 1.332 | True | True | True | True | Sex-specific 1e-2 VUMC, sex-differential UKB |
| 0.003256 | 0.003498 | 7.068 | 0.8418 | 2.011 | 1.347 | True | True | True | True | Sex-specific 1e-2 VUMC, sex-differential UKB |
| 0.003256 | 0.003498 | 7.085 | 0.8004 | 2.008 | 1.344 | True | True | True | True | Sex-specific 1e-2 VUMC, sex-differential UKB |
| 0.003257 | 0.003498 | 7.063 | 0.8248 | 2.011 | 1.342 | True | True | True | True | Sex-specific 1e-2 VUMC, sex-differential UKB |
| 0.003257 | 0.003499 | 7.067 | 0.8251 | 2.018 | 1.349 | True | True | True | True | Sex-specific 1e-2 VUMC, sex-differential UKB |
| 0.003257 | 0.003499 | 7.065 | 0.8248 | 2.009 | 1.338 | True | True | True | True | Sex-specific 1e-2 VUMC, sex-differential UKB |
| 0.003257 | 0.003499 | 7.059 | 0.8348 | 2.007 | 1.334 | True | True | True | True | Sex-specific 1e-2 VUMC, sex-differential UKB |
| 0.003257 | 0.003499 | 7.063 | 0.8349 | 2.01 | 1.346 | True | True | True | True | Sex-specific 1e-2 VUMC, sex-differential UKB |
| 0.003258 | 0.0035 | 7.009 | 0.8557 | 2.019 | 1.332 | True | True | True | True | Sex-specific 1e-2 VUMC, sex-differential UKB |

| SE_UKB_female | SE_UKB_male | LOG10P_VUMC_female | LOG10P_VUMC_male | LOG10P_UKB_female | LOG10P_UKB_male | female_effect_direction_replication | male_effect_direction_replication | sex_effect_comparison_replication | pvalue_effect_comparison_replication | replication_type |
|---|---|---|---|---|---|---|---|---|---|---|
| 0.003251 | 0.003498 | 6.013 | 1.518 | 3.187 | 1.359 | True | True | True | True | Sex-specific 1e-2 VUMC, sex-differential UKB |
| 0.003261 | 0.003503 | 7.107 | 0.8437 | 1.997 | 1.341 | True | True | True | True | Sex-specific 1e-2 VUMC, sex-differential UKB |
| 0.003261 | 0.003503 | 7.111 | 0.8434 | 1.997 | 1.346 | True | True | True | True | Sex-specific 1e-2 VUMC, sex-differential UKB |
| 0.003323 | 0.003583 | 7.476 | 0.6376 | 3.083 | 2.932 | True | True | False | True | |
| 0.01941 | 0.01978 | 7.566 | 1.44 | 1.036 | 2.027 | True | True | False | False | |
| 0.01941 | 0.01978 | 7.637 | 1.351 | 1.069 | 2.106 | True | True | False | False | |
| 0.01941 | 0.01978 | 7.665 | 1.35 | 1.069 | 2.105 | True | True | False | False | |
| 0.02035 | 0.02117 | 5.121 | 0.8824 | 1.053 | 2.034 | True | True | False | False | |
| 0.01941 | 0.01979 | 7.644 | 1.33 | 1.069 | 2.102 | True | True | False | False | |
| 0.01941 | 0.01979 | 7.668 | 1.327 | 1.072 | 2.099 | True | True | False | False | |
| 0.01941 | 0.01979 | 7.65 | 1.327 | 1.087 | 2.1 | True | True | False | False | |
| 0.01942 | 0.01979 | 7.638 | 1.292 | 1.087 | 2.092 | True | True | False | False | |
| 0.01941 | 0.01979 | 7.638 | 1.291 | 1.067 | 2.092 | True | True | False | False | |
| 0.01941 | 0.01979 | 7.617 | 1.294 | 1.068 | 2.088 | True | True | False | False | |
| 0.01942 | 0.0198 | 7.613 | 1.302 | 1.083 | 2.092 | True | True | False | False | |
| 0.02101 | 0.02151 | 4.752 | 0.958 | 0.6066 | 1.541 | True | True | False | False | |
| 0.02101 | 0.02151 | 4.71 | 0.9227 | 0.5939 | 1.587 | True | True | False | False | |
| 0.02102 | 0.02152 | 4.655 | 0.8465 | 0.6045 | 1.555 | True | True | False | False | |
| 0.02595 | 0.02709 | 8.488 | 1.699 | 2.412 | 3.107 | True | True | False | False | |
| 0.02653 | 0.02783 | 10.01 | 1.718 | 2.446 | 2.548 | True | True | False | False | |
| 0.02755 | 0.02781 | 8.286 | 1.257 | 1.336 | 0.7687 | True | True | True | True | Sex-specific 1e-2 VUMC, sex-differential UKB |
| 0.03021 | 0.03086 | 3.907 | 0.4523 | 0.6173 | 0.02525 | True | True | True | True | Sex-specific 1e-2 VUMC, sex-differential UKB |
| 0.03115 | 0.03163 | 3.697 | 0.3973 | 0.709 | 0.01497 | True | True | True | True | Sex-specific 1e-2 VUMC, sex-differential UKB |
| 0.02375 | 0.02443 | 6.297 | 1.684 | 1.18 | 0.1334 | True | True | True | True | Sex-specific 1e-2 VUMC, sex-differential UKB |
| 0.03123 | 0.03168 | 3.954 | 0.4906 | 0.8084 | 0.04912 | True | False | True | True | Sex-specific 1e-2 VUMC, sex-differential UKB |
| 0.03116 | 0.03171 | 3.775 | 0.4253 | 0.7383 | 0.01509 | True | True | True | True | Sex-specific 1e-2 VUMC, sex-differential UKB |
| 0.03154 | 0.03205 | 3.941 | 0.4206 | 0.8361 | 0.0625 | True | False | True | True | Sex-specific 1e-2 VUMC, sex-differential UKB |
| 0.02034 | 0.02094 | 5.535 | 1.307 | 0.2509 | 2.339 | True | True | False | False | |
| 0.01942 | 0.01976 | 5.807 | 1.754 | 0.7207 | 3.272 | True | True | False | False | |
| 0.02032 | 0.02092 | 5.719 | 1.264 | 0.2252 | 2.316 | True | True | False | False | |
| 0.02031 | 0.02092 | 5.619 | 1.24 | 0.2287 | 2.313 | True | True | False | False | |
| 0.0203 | 0.02092 | 5.58 | 1.217 | 0.2219 | 2.288 | True | True | False | False | |
| 0.01917 | 0.01956 | 6.307 | 0.9797 | 0.7868 | 4.192 | True | True | False | False | |
| 0.0203 | 0.02091 | 5.566 | 1.233 | 0.2222 | 2.285 | True | True | False | False | |
| SE_UKB_female | SE_UKB_male | LOG10P_VUMC_female | LOG10P_VUMC_male | LOG10P_UKB_female | LOG10P_UKB_male | female_effect_direction_replication | male_effect_direction_replication | sex_effect_comparison_replication | pvalue_effect_comparison_replication | replication_type |
| 0.0203 | 0.02091 | 5.728 | 1.253 | 0.2489 | 2.254 | True | True | False | False | |
| 0.01942 | 0.01974 | 7.679 | 0.8611 | 0.9043 | 4.674 | True | True | False | False | |

| SE_UKB_female | SE_UKB_male | LOG10P_VUMC_female | LOG10P_VUMC_male | LOG10P_UKB_female | LOG10P_UKB_male | female_effect_direction_replication | male_effect_direction_replication | sex_effect_comparison_replication | pvalue_effect_comparison_replication | replication_type |
|---|---|---|---|---|---|---|---|---|---|---|
| 0.02028 | 0.02089 | 5.412 | 1.243 | 0.2219 | 2.297 | True | True | False | False | |
| 0.02046 | 0.02103 | 1.306 | 6.649 | 1.283 | 0.9023 | True | True | False | False | |
| 0.02044 | 0.02101 | 1.307 | 6.918 | 1.333 | 0.8995 | True | True | False | False | |
| 0.02035 | 0.02083 | 1.463 | 7.718 | 1.137 | 0.9562 | True | True | False | False | |
| 0.02035 | 0.02083 | 1.463 | 7.721 | 1.137 | 0.9566 | True | True | False | False | |
| 0.01908 | 0.01968 | 0.8921 | 9.567 | 1.06 | 0.7888 | True | True | False | False | |
| 0.01908 | 0.0197 | 0.7905 | 9.304 | 0.9255 | 1.196 | True | True | True | True | Sex-specific 1e-2 VUMC, sex-differential UKB |
| 0.01907 | 0.0197 | 0.7732 | 9.354 | 0.9234 | 1.161 | True | True | True | True | Sex-specific 1e-2 VUMC, sex-differential UKB |
| 0.02049 | 0.02107 | 1.55 | 8.086 | 0.5678 | 1.088 | True | True | True | True | Sex-specific 1e-2 VUMC, sex-differential UKB |
| 0.01907 | 0.0197 | 0.7518 | 9.388 | 0.9235 | 1.161 | True | True | True | True | Sex-specific 1e-2 VUMC, sex-differential UKB |
| 0.01907 | 0.0197 | 0.7635 | 9.362 | 0.9237 | 1.161 | True | True | True | True | Sex-specific 1e-2 VUMC, sex-differential UKB |
| 0.02052 | 0.02109 | 1.537 | 8.179 | 0.6108 | 1.1 | True | True | True | True | Sex-specific 1e-2 VUMC, sex-differential UKB |
| 0.0191 | 0.01974 | 0.7945 | 9.441 | 0.8938 | 1.195 | True | True | True | True | Sex-specific 1e-2 VUMC, sex-differential UKB |
| 0.01888 | 0.01976 | 0.2604 | 4.141 | 1.208 | 1.192 | False | True | True | False | |
| 0.003949 | 0.004342 | 12.26 | 1.504 | 125.76 | 52.78 | True | True | True | True | Sex-specific 1e-2 VUMC, sex-differential UKB |
| 0.003966 | 0.004357 | 12.95 | 1.683 | 132.29 | 56.88 | True | True | True | True | Sex-specific 1e-2 VUMC, sex-differential UKB |
| 0.004309 | 0.00474 | 14 | 1.281 | 177.26 | 67.2 | True | True | True | True | Sex-specific 1e-2 VUMC, sex-differential UKB |
| 0.004243 | 0.004677 | 12.88 | 0.8874 | 163.96 | 66.99 | True | True | True | True | Sex-specific 1e-2 VUMC, sex-differential UKB |
| 0.00424 | 0.004673 | 13.12 | 0.899 | 163.78 | 67.2 | True | True | True | True | Sex-specific 1e-2 VUMC, sex-differential UKB |
| 0.00343 | 0.003689 | 7.022 | 0.3127 | 42.13 | 6.995 | True | True | True | True | Sex-specific 1e-2 VUMC, sex-specific 5e-8 UKB |
| 0.003433 | 0.003692 | 7.044 | 0.3009 | 42.31 | 6.913 | True | True | True | True | Sex-specific 1e-2 VUMC, sex-specific 5e-8 UKB |
| 0.003434 | 0.003693 | 6.916 | 0.3101 | 42.68 | 6.865 | True | True | True | True | Sex-specific 1e-2 VUMC, sex-specific 5e-8 UKB |
| 0.003434 | 0.003693 | 6.914 | 0.3101 | 42.69 | 6.862 | True | True | True | True | Sex-specific 1e-2 VUMC, sex-specific 5e-8 UKB |
| 0.003434 | 0.003693 | 6.923 | 0.3056 | 42.68 | 6.859 | True | True | True | True | Sex-specific 1e-2 VUMC, sex-specific 5e-8 UKB |
| 0.003436 | 0.003694 | 6.967 | 0.2991 | 42.75 | 6.872 | True | True | True | True | Sex-specific 1e-2 VUMC, sex-specific 5e-8 UKB |
| 0.003436 | 0.003695 | 6.892 | 0.2841 | 42.71 | 6.891 | True | True | True | True | Sex-specific 1e-2 VUMC, sex-specific 5e-8 UKB |
| 0.003437 | 0.003696 | 6.902 | 0.2777 | 42.77 | 6.879 | True | True | True | True | Sex-specific 1e-2 VUMC, sex-specific 5e-8 UKB |
| 0.003436 | 0.003693 | 7.331 | 0.1766 | 41.54 | 6.575 | True | True | True | True | Sex-specific 1e-2 VUMC, sex-specific 5e-8 UKB |
| 0.003432 | 0.003688 | 7.513 | 0.1572 | 41.59 | 6.347 | True | True | True | True | Sex-specific 1e-2 VUMC, sex-specific 5e-8 UKB |

| SE_UKB_female | SE_UKB_male | LOG10P_VUMC_female | LOG10P_VUMC_male | LOG10P_UKB_female | LOG10P_UKB_male | female_effect_direction_replication | male_effect_direction_replication | sex_effect_comparison_replication | pvalue_effect_comparison_replication | replication_type |
|---|---|---|---|---|---|---|---|---|---|---|
| 0.003454 | 0.003708 | 1.285 | 4.304 | 8.547 | 8.449 | True | True | True | False | |
| 0.003576 | 0.003838 | 1.732 | 7.863 | 12.49 | 13.32 | True | True | True | True | Sex-specific 1e-2 VUMC, sex-differential UKB |
| 0.003583 | 0.003845 | 1.913 | 8.256 | 13.24 | 13.83 | True | True | True | True | Sex-specific 1e-2 VUMC, sex-differential UKB |
| 0.003582 | 0.003843 | 1.879 | 8.115 | 12.92 | 13.85 | True | True | True | True | Sex-specific 1e-2 VUMC, sex-differential UKB |
| 0.003125 | 0.003395 | 8.179 | 1.546 | 8.137 | 14.35 | True | True | False | False | |
| 0.003144 | 0.003414 | 6.005 | 0.7281 | 5.62 | 10.11 | True | True | False | False | |
| 0.003125 | 0.003395 | 7.871 | 1.343 | 8.005 | 13.83 | True | True | False | False | |
| 0.003127 | 0.003398 | 8.335 | 1.565 | 8.838 | 14.25 | True | True | False | False | |
| 0.003126 | 0.003395 | 7.936 | 1.261 | 8.302 | 14.02 | True | True | False | False | |
| 0.003144 | 0.003413 | 6.145 | 0.6345 | 5.809 | 10.24 | True | True | False | False | |
| 0.003132 | 0.003401 | 5.89 | 0.5488 | 4.839 | 9.835 | True | True | False | False | |
| 0.003125 | 0.003395 | 8.212 | 1.282 | 8.049 | 14.5 | True | True | False | False | |
| 0.003139 | 0.003409 | 5.911 | 0.663 | 4.849 | 10.22 | True | True | False | False | |
| 0.003125 | 0.003395 | 8.207 | 1.272 | 8.084 | 14.73 | True | True | False | False | |
| | | 8.221 | 1.633 | | | False | False | False | False | |
| 0.003124 | 0.003395 | 8.109 | 1.57 | 7.525 | 15.04 | True | True | False | False | |
| 0.003122 | 0.003392 | 8.159 | 1.388 | 6.759 | 14.88 | True | True | False | False | |
| 0.00314 | 0.003409 | 6.148 | 0.8562 | 4.77 | 10.32 | True | True | False | False | |
| 0.003122 | 0.003392 | 8.16 | 1.389 | 6.754 | 14.86 | True | True | False | False | |
| 0.003122 | 0.003393 | 8.071 | 1.39 | 6.847 | 14.77 | True | True | False | False | |
| 0.003141 | 0.003412 | 6.167 | 0.8785 | 4.89 | 10.23 | True | True | False | False | |
| 0.003122 | 0.003393 | 8.103 | 1.401 | 6.871 | 14.75 | True | True | False | False | |
| 0.003126 | 0.003398 | 8.135 | 1.521 | 7.543 | 14.79 | True | True | False | False | |
| 0.003123 | 0.003394 | 8.048 | 1.395 | 6.801 | 14.65 | True | True | False | False | |
| 0.003126 | 0.003398 | 8.212 | 1.425 | 7.648 | 14.89 | True | True | False | False | |
| 0.003123 | 0.003394 | 7.954 | 1.451 | 6.866 | 14.3 | True | True | False | False | |

| SE_UKB_female | SE_UKB_male | LOG10P_VUMC_female | LOG10P_VUMC_male | LOG10P_UKB_female | LOG10P_UKB_male | female_effect_direction_replication | male_effect_direction_replication | sex_effect_comparison_replication | pvalue_effect_comparison_replication | replication_type |
|---|---|---|---|---|---|---|---|---|---|---|
| 0.003123 | 0.003394 | 7.941 | 1.458 | 6.894 | 14.31 | True | True | False | False | |
| 0.003123 | 0.003394 | 7.941 | 1.465 | 6.896 | 14.31 | True | True | False | False | |
| 0.003123 | 0.003394 | 7.879 | 1.482 | 6.88 | 14.39 | True | True | False | False | |

| SE_UKB_female | SE_UKB_male | LOG10P_VUMC_female | LOG10P_VUMC_male | LOG10P_UKB_female | LOG10P_UKB_male | female_effect_direction_replication | male_effect_direction_replication | sex_effect_comparison_replication | pvalue_effect_comparison_replication | replication_type |
|---|---|---|---|---|---|---|---|---|---|---|
| 0.003126 | 0.003398 | 7.966 | 1.625 | 7.734 | 14.54 | True | True | False | False | |
| 0.003123 | 0.003394 | 7.863 | 1.466 | 6.874 | 14.4 | True | True | False | False | |
| 0.003126 | 0.003398 | 8.051 | 1.614 | 7.724 | 14.53 | True | True | False | False | |
| 0.003126 | 0.003398 | 7.971 | 1.614 | 7.705 | 14.53 | True | True | False | False | |
| 0.003126 | 0.003398 | 7.966 | 1.614 | 7.716 | 14.55 | True | True | False | False | |
| 0.003126 | 0.003398 | 7.989 | 1.614 | 7.689 | 14.58 | True | True | False | False | |
| 0.003123 | 0.003394 | 7.947 | 1.464 | 6.896 | 14.42 | True | True | False | False | |
| 0.003126 | 0.003398 | 8.047 | 1.633 | 7.685 | 14.6 | True | True | False | False | |
| 0.003123 | 0.003394 | 7.947 | 1.464 | 6.896 | 14.46 | True | True | False | False | |
| 0.003123 | 0.003394 | 7.946 | 1.464 | 6.905 | 14.46 | True | True | False | False | |
| 0.003126 | 0.003398 | 7.871 | 1.598 | 7.628 | 14.56 | True | True | False | False | |
| 0.003126 | 0.003398 | 7.956 | 1.617 | 7.693 | 14.6 | True | True | False | False | |
| 0.003123 | 0.003394 | 7.861 | 1.465 | 6.861 | 14.43 | True | True | False | False | |
| 0.003126 | 0.003398 | 7.956 | 1.617 | 7.681 | 14.56 | True | True | False | False | |
| 0.003126 | 0.003398 | 7.955 | 1.616 | 7.696 | 14.59 | True | True | False | False | |
| 0.003126 | 0.003398 | 7.956 | 1.616 | 7.702 | 14.61 | True | True | False | False | |
| 0.003126 | 0.003398 | 7.956 | 1.623 | 7.693 | 14.59 | True | True | False | False | |
| 0.003127 | 0.003399 | 7.927 | 1.603 | 7.389 | 14.56 | True | True | False | False | |
| 0.003127 | 0.003399 | 7.926 | 1.603 | 7.392 | 14.59 | True | True | False | False | |
| 0.003127 | 0.003399 | 7.923 | 1.603 | 7.382 | 14.57 | True | True | False | False | |
| 0.003123 | 0.003394 | 7.829 | 1.452 | 6.556 | 14.4 | True | True | False | False | |
| 0.003127 | 0.003398 | 7.906 | 1.599 | 7.409 | 14.59 | True | True | False | False | |

| SE_UKB_female | SE_UKB_male | LOG10P_VUMC_female | LOG10P_VUMC_male | LOG10P_UKB_female | LOG10P_UKB_male | female_effect_direction_replication | male_effect_direction_replication | sex_effect_comparison_replication | pvalue_effect_comparison_replication | replication_type |
|---|---|---|---|---|---|---|---|---|---|---|
| 0.003124 | 0.003395 | 7.756 | 1.444 | 6.516 | 14.28 | True | True | False | False | |
| 0.003124 | 0.003395 | 7.811 | 1.454 | 6.638 | 14.39 | True | True | False | False | |
| 0.003144 | 0.003415 | 6.161 | 0.969 | 4.691 | 9.767 | True | True | False | False | |
| 0.003127 | 0.003399 | 7.957 | 1.622 | 7.404 | 14.39 | True | True | False | False | |
| 0.003144 | 0.003415 | 6.053 | 0.8852 | 4.828 | 9.721 | True | True | False | False | |
| 0.003124 | 0.003395 | 7.863 | 1.473 | 6.571 | 14.3 | True | True | False | False | |
| 0.003127 | 0.003399 | 7.974 | 1.648 | 7.462 | 14.48 | True | True | False | False | |
| 0.003144 | 0.003416 | 6.197 | 0.9331 | 4.768 | 9.699 | True | True | False | False | |
| 0.003124 | 0.003395 | 7.861 | 1.465 | 6.612 | 14.31 | True | True | False | False | |
| 0.003144 | 0.003416 | 6.199 | 0.9322 | 4.806 | 9.711 | True | True | False | False | |
| 0.003144 | 0.003416 | 6.133 | 0.9183 | 4.789 | 9.684 | True | True | False | False | |
| 0.003126 | 0.003397 | 8.202 | 1.28 | 6.625 | 13.98 | True | True | False | False | |
| 0.003195 | 0.003474 | 5.956 | 0.1957 | 0.7812 | 3.345 | True | True | False | False | |
| 0.003132 | 0.003403 | 6.927 | 1.063 | 6.645 | 13.53 | True | True | False | False | |
| 0.003127 | 0.003399 | 7.865 | 1.112 | 6.351 | 12.8 | True | True | False | False | |
| 0.003144 | 0.003417 | 6.006 | 0.6651 | 4.463 | 8.496 | True | True | False | False | |
| 0.003126 | 0.003399 | 7.778 | 1.062 | 6.405 | 12.86 | True | True | False | False | |
| 0.00314 | 0.00341 | 5.152 | 0.5945 | 5.049 | 9.216 | True | True | False | False | |
| 0.003129 | 0.0034 | 6.858 | 1.176 | 7.784 | 13.98 | True | True | False | False | |
| 0.003142 | 0.003415 | 6.118 | 0.6522 | 4.654 | 8.675 | True | True | False | False | |
| 0.003134 | 0.003403 | 6.765 | 0.6678 | 7.117 | 13.59 | True | True | False | False | |
| 0.003134 | 0.003403 | 6.749 | 0.6686 | 7.142 | 13.51 | True | True | False | False | |
| 0.003129 | 0.0034 | 7.717 | 0.6695 | 6.704 | 12.77 | True | True | False | False | |
| 0.003126 | 0.003398 | 7.878 | 1.112 | 6.383 | 12.86 | True | True | False | False | |
| 0.003125 | 0.003398 | 7.711 | 1.372 | 6.43 | 13.21 | True | True | False | False | |
| 0.003128 | 0.003398 | 5.722 | 0.7048 | 3.883 | 8.611 | True | True | False | False | |
| 0.003126 | 0.003398 | 7.993 | 1.144 | 6.489 | 13 | True | True | False | False | |

| SE_UKB_female | SE_UKB_male | LOG10P_VUMC_female | LOG10P_VUMC_male | LOG10P_UKB_female | LOG10P_UKB_male | female_effect_direction_replication | male_effect_direction_replication | sex_effect_comparison_replication | pvalue_effect_comparison_replication | replication_type |
|---|---|---|---|---|---|---|---|---|---|---|
| 0.003142 | 0.003414 | 6.103 | 0.6476 | 4.598 | 8.721 | True | True | False | False | |
| 0.003142 | 0.003414 | 6.086 | 0.6593 | 4.574 | 8.716 | True | True | False | False | |
| 0.003126 | 0.003399 | 8.003 | 1.141 | 6.45 | 13.01 | True | True | False | False | |
| 0.00313 | 0.0034 | 7.812 | 0.8517 | 7.24 | 12.91 | True | True | False | False | |
| 0.003152 | 0.003419 | 5.135 | 0.1217 | 3.72 | 7.134 | True | True | False | False | |
| 0.003158 | 0.003425 | 5.383 | 0.04314 | 3.53 | 6.433 | True | True | False | False | |
| 0.003142 | 0.00341 | 7.426 | 0.3844 | 5.611 | 9.908 | True | True | False | False | |
| 0.00314 | 0.003408 | 7.262 | 0.2466 | 4.961 | 9.837 | True | True | False | False | |
| 0.00314 | 0.003408 | 7.147 | 0.2036 | 5.116 | 10.03 | True | True | False | False | |
| 0.003155 | 0.003423 | 5.272 | 0.007321 | 3.259 | 6.094 | True | False | False | False | |
| 0.003147 | 0.003414 | 5.685 | 0.3063 | 5.458 | 10.86 | True | True | False | False | |

| SE_UKB_female | SE_UKB_male | LOG10P_VUMC_female | LOG10P_VUMC_male | LOG10P_UKB_female | LOG10P_UKB_male | female_effect_direction_replication | male_effect_direction_replication | sex_effect_comparison_replication | pvalue_effect_comparison_replication | replication_type |
|---|---|---|---|---|---|---|---|---|---|---|
| 0.003143 | 0.003411 | 6.583 | 0.3064 | 5.061 | 10.24 | True | True | False | False | |
| 0.003147 | 0.003414 | 5.877 | 0.1979 | 5.54 | 10.41 | True | True | False | False | |
| 0.003147 | 0.003414 | 5.99 | 0.2216 | 5.476 | 10.36 | True | True | False | False | |
| 0.003143 | 0.003411 | 6.922 | 0.2232 | 5.049 | 9.641 | True | True | False | False | |
| 0.003147 | 0.003414 | 5.937 | 0.2213 | 5.499 | 10.35 | True | True | False | False | |
| 0.003147 | 0.003413 | 6.016 | 0.236 | 5.67 | 10.28 | True | True | False | False | |
| 0.003143 | 0.003411 | 7.02 | 0.2611 | 5.193 | 9.651 | True | True | False | False | |
| 0.003145 | 0.003413 | 7.091 | 0.3569 | 5.885 | 9.724 | True | True | False | False | |
| 0.003147 | 0.003414 | 5.983 | 0.2282 | 5.557 | 10.44 | True | True | False | False | |
| 0.003147 | 0.003414 | 6.883 | 0.3854 | 5.735 | 9.967 | True | True | False | False | |
| 0.003149 | 0.003418 | 6.3 | 0.3606 | 5.462 | 9.311 | True | True | False | False | |
| 0.003154 | 0.003423 | 5.714 | 0.3199 | 0.5243 | 2.753 | True | False | False | False | |
| 0.003172 | 0.00344 | 5.549 | 0.1515 | 0.766 | 2.831 | True | False | False | False | |
| 0.003148 | 0.003415 | 5.105 | 0.2725 | 4.966 | 9.196 | True | True | False | False | |
| 0.003193 | 0.003467 | 4.184 | 0.2798 | 0.016 | 1.496 | True | False | False | False | |

| SE_UKB_female | SE_UKB_male | LOG10P_VUMC_female | LOG10P_VUMC_male | LOG10P_UKB_female | LOG10P_UKB_male | female_effect_direction_replication | male_effect_direction_replication | sex_effect_comparison_replication | pvalue_effect_comparison_replication | replication_type |
|---|---|---|---|---|---|---|---|---|---|---|
| 0.003193 | 0.003467 | 4.077 | 0.2615 | 0.01151 | 1.566 | False | False | False | False | |
| 0.003502 | 0.003797 | 0.3716 | 7.439 | 23.02 | 16.14 | True | True | False | False | |
| 0.003232 | 0.003499 | 1.08 | 6.157 | 25.07 | 18.85 | True | True | False | False | |
| 0.003419 | 0.003703 | 0.2165 | 5.238 | 17.12 | 11.06 | True | True | False | False | |
| 0.003312 | 0.003599 | 3.437 | 0.4206 | 16.07 | 17.57 | True | False | False | False | |
| | | 3.35 | 0.4157 | | | False | False | False | False | |
| 0.003292 | 0.003576 | 3.753 | 0.4239 | 17.51 | 17.52 | True | False | False | False | |
| 0.003286 | 0.003563 | 4.74 | 0.4131 | 22.8 | 19.19 | True | False | True | True | Sex-specific 1e-2 VUMC, sex-differential UKB |
| 0.00329 | 0.003567 | 4.741 | 0.4162 | 22.81 | 19.19 | True | False | True | True | Sex-specific 1e-2 VUMC, sex-differential UKB |
| 0.003404 | 0.003686 | 6.96 | 0.4589 | 31.28 | 23.3 | True | False | True | True | Sex-specific 1e-2 VUMC, sex-differential UKB |
| 0.003403 | 0.003686 | 6.963 | 0.46 | 31.29 | 23.34 | True | False | True | True | Sex-specific 1e-2 VUMC, sex-differential UKB |
| 0.0034 | 0.003683 | 6.838 | 0.5523 | 31.16 | 23.65 | True | False | True | True | Sex-specific 1e-2 VUMC, sex-differential UKB |
| 0.0034 | 0.003682 | 6.695 | 0.5127 | 31.28 | 23.74 | True | False | True | True | Sex-specific 1e-2 VUMC, sex-differential UKB |
| 0.003345 | 0.00362 | 8.136 | 0.457 | 30.06 | 23.55 | True | False | True | True | Sex-specific 1e-2 VUMC, sex-differential UKB |
| 0.003357 | 0.003633 | 7.739 | 0.4057 | 30.08 | 22.67 | True | False | True | True | Sex-specific 1e-2 VUMC, sex-differential UKB |
| 0.003402 | 0.003685 | 6.722 | 0.5283 | 31.47 | 23.49 | True | False | True | True | Sex-specific 1e-2 VUMC, sex-differential UKB |
| 0.003395 | 0.003675 | 6.016 | 0.261 | 31 | 25.65 | True | False | True | True | Sex-specific 1e-2 VUMC, sex-differential UKB |
| 0.003395 | 0.003677 | 6.794 | 0.4611 | 31.22 | 25.19 | True | False | True | True | Sex-specific 1e-2 VUMC, sex-differential UKB |
| 0.003393 | 0.003675 | 6.376 | 0.3745 | 31.32 | 25.2 | True | False | True | True | Sex-specific 1e-2 VUMC, sex-differential UKB |
| 0.003393 | 0.003674 | 6.36 | 0.3744 | 31.3 | 25.15 | True | False | True | True | Sex-specific 1e-2 VUMC, sex-differential UKB |
| 0.003357 | 0.003633 | 7.482 | 0.3777 | 31.95 | 24.3 | True | False | True | True | Sex-specific 1e-2 VUMC, sex-differential UKB |
| 0.003356 | 0.003633 | 7.561 | 0.3783 | 31.93 | 24.54 | True | False | True | True | Sex-specific 1e-2 VUMC, sex-differential UKB |
| 0.003399 | 0.00368 | 6.451 | 0.3254 | 31.73 | 25.8 | True | False | True | True | Sex-specific 1e-2 VUMC, sex-differential UKB |
| 0.003363 | 0.003639 | 7.195 | 0.2988 | 32.04 | 25.83 | True | False | True | True | Sex-specific 1e-2 VUMC, sex-differential UKB |
| 0.003355 | 0.003632 | 6.83 | 0.2812 | 31.9 | 25.66 | True | False | True | True | Sex-specific 1e-2 VUMC, sex-differential UKB |
| 0.003373 | 0.00365 | 6.852 | 0.2268 | 33.22 | 26.94 | True | False | True | True | Sex-specific 1e-2 VUMC, sex-differential UKB |

| SE_UKB_female | SE_UKB_male | LOG10P_VUMC_female | LOG10P_VUMC_male | LOG10P_UKB_female | LOG10P_UKB_male | female_effect_direction_replication | male_effect_direction_replication | sex_effect_comparison_replication | pvalue_effect_comparison_replication | replication_type |
|---|---|---|---|---|---|---|---|---|---|---|
| 0.003373 | 0.00365 | 6.788 | 0.2425 | 33.27 | 27.05 | True | False | True | True | Sex-specific 1e-2 VUMC, sex-differential UKB |
| 0.003327 | 0.0036 | 7.856 | 0.4553 | 33.66 | 25.07 | True | False | True | True | Sex-specific 1e-2 VUMC, sex-differential UKB |
| 0.003332 | 0.003605 | 7.047 | 0.2703 | 34.28 | 25.57 | True | False | True | True | Sex-specific 1e-2 VUMC, sex-differential UKB |
| 0.003334 | 0.003606 | 7.334 | 0.2975 | 34.15 | 25.45 | True | False | True | True | Sex-specific 1e-2 VUMC, sex-differential UKB |
| 0.003382 | 0.003658 | 6.945 | 0.1799 | 34.11 | 27.72 | True | False | True | True | Sex-specific 1e-2 VUMC, sex-differential UKB |
| 0.003324 | 0.003597 | 7.01 | 0.2745 | 33.76 | 25.1 | True | False | True | True | Sex-specific 1e-2 VUMC, sex-differential UKB |
| 0.003324 | 0.003597 | 7.017 | 0.2765 | 33.77 | 25.1 | True | False | True | True | Sex-specific 1e-2 VUMC, sex-differential UKB |
| 0.003324 | 0.003597 | 7.025 | 0.2793 | 33.76 | 25.1 | True | False | True | True | Sex-specific 1e-2 VUMC, sex-differential UKB |
| 0.003378 | 0.003654 | 6.577 | 0.1262 | 33.66 | 27.32 | True | False | True | True | Sex-specific 1e-2 VUMC, sex-differential UKB |
| 0.003382 | 0.003658 | 7.004 | 0.191 | 34.08 | 27.61 | True | False | True | True | Sex-specific 1e-2 VUMC, sex-differential UKB |
| 0.003382 | 0.003658 | 7.024 | 0.1927 | 34.08 | 27.6 | True | False | True | True | Sex-specific 1e-2 VUMC, sex-differential UKB |
| 0.00337 | 0.003647 | 6.671 | 0.1807 | 33.78 | 27.18 | True | False | True | True | Sex-specific 1e-2 VUMC, sex-differential UKB |
| 0.00337 | 0.003647 | 6.686 | 0.167 | 33.66 | 27.21 | True | False | True | True | Sex-specific 1e-2 VUMC, sex-differential UKB |
| 0.003367 | 0.003643 | 6.242 | 0.1329 | 33.29 | 27.03 | True | False | True | True | Sex-specific 1e-2 VUMC, sex-differential UKB |
| 0.003367 | 0.003643 | 6.242 | 0.1328 | 33.29 | 27.03 | True | False | True | True | Sex-specific 1e-2 VUMC, sex-differential UKB |
| 0.00338 | 0.003657 | 6.728 | 0.2113 | 34.09 | 27.09 | True | False | True | True | Sex-specific 1e-2 VUMC, sex-differential UKB |
| 0.00338 | 0.003657 | 6.729 | 0.2027 | 34.09 | 27.09 | True | False | True | True | Sex-specific 1e-2 VUMC, sex-differential UKB |
| 0.003371 | 0.003648 | 6.721 | 0.158 | 33.51 | 26.91 | True | False | True | True | Sex-specific 1e-2 VUMC, sex-differential UKB |
| 0.003367 | 0.003644 | 6.251 | 0.1327 | 33.12 | 26.78 | True | False | True | True | Sex-specific 1e-2 VUMC, sex-differential UKB |
| SE_UKB_female | SE_UKB_male | LOG10P_VUMC_female | LOG10P_VUMC_male | LOG10P_UKB_female | LOG10P_UKB_male | female_effect_direction_replication | male_effect_direction_replication | sex_effect_comparison_replication | pvalue_effect_comparison_replication | replication_type |
| 0.003365 | 0.003641 | 6.254 | 0.1193 | 33.39 | 26.95 | True | False | True | True | Sex-specific 1e-2 VUMC, sex-differential UKB |
| 0.003365 | 0.003641 | 6.255 | 0.1191 | 33.45 | 26.92 | True | False | True | True | Sex-specific 1e-2 VUMC, sex-differential UKB |
| 0.003368 | 0.003645 | 6.694 | 0.1588 | 33.79 | 27.1 | True | False | True | True | Sex-specific 1e-2 VUMC, sex-differential UKB |
| 0.003364 | 0.00364 | 6.153 | 0.145 | 33.32 | 27.02 | True | False | True | True | Sex-specific 1e-2 VUMC, sex-differential UKB |
| 0.003364 | 0.00364 | 6.169 | 0.1467 | 33.28 | 27.04 | True | False | True | True | Sex-specific 1e-2 VUMC, sex-differential UKB |
| 0.003364 | 0.00364 | 6.153 | 0.145 | 33.32 | 27.03 | True | False | True | True | Sex-specific 1e-2 VUMC, sex-differential UKB |

| SE_UKB_female | SE_UKB_male | LOG10P_VUMC_female | LOG10P_VUMC_male | LOG10P_UKB_female | LOG10P_UKB_male | female_effect_direction_replication | male_effect_direction_replication | sex_effect_comparison_replication | pvalue_effect_comparison_replication | replication_type |
|---|---|---|---|---|---|---|---|---|---|---|
| 0.003368 | 0.003644 | 6.587 | 0.1865 | 33.66 | 27.2 | True | False | True | True | Sex-specific 1e-2 VUMC, sex-differential UKB |
| 0.003364 | 0.00364 | 6.153 | 0.145 | 33.29 | 27.03 | True | False | True | True | Sex-specific 1e-2 VUMC, sex-differential UKB |
| 0.003365 | 0.003641 | 6.028 | 0.16 | 33.35 | 26.95 | True | False | True | True | Sex-specific 1e-2 VUMC, sex-differential UKB |
| 0.003365 | 0.003641 | 6.028 | 0.16 | 33.35 | 26.95 | True | False | True | True | Sex-specific 1e-2 VUMC, sex-differential UKB |
| 0.003365 | 0.003641 | 6.028 | 0.16 | 33.35 | 26.94 | True | False | True | True | Sex-specific 1e-2 VUMC, sex-differential UKB |
| 0.003365 | 0.003641 | 6.035 | 0.1601 | 33.35 | 26.94 | True | False | True | True | Sex-specific 1e-2 VUMC, sex-differential UKB |
| 0.003364 | 0.003641 | 6.028 | 0.1193 | 33.41 | 26.97 | True | False | True | True | Sex-specific 1e-2 VUMC, sex-differential UKB |
| 0.003368 | 0.003645 | 6.425 | 0.203 | 33.68 | 27.15 | True | False | True | True | Sex-specific 1e-2 VUMC, sex-differential UKB |
| 0.003365 | 0.003641 | 6.019 | 0.132 | 33.38 | 26.98 | True | False | True | True | Sex-specific 1e-2 VUMC, sex-differential UKB |
| 0.003365 | 0.003641 | 6.016 | 0.1327 | 33.38 | 26.97 | True | False | True | True | Sex-specific 1e-2 VUMC, sex-differential UKB |
| 0.003364 | 0.00364 | 6.026 | 0.1309 | 33.38 | 27.02 | True | False | True | True | Sex-specific 1e-2 VUMC, sex-differential UKB |
| 0.003364 | 0.00364 | 6.03 | 0.1309 | 33.4 | 26.97 | True | False | True | True | Sex-specific 1e-2 VUMC, sex-differential UKB |
| 0.003363 | 0.003639 | 6.037 | 0.131 | 33.41 | 26.97 | True | False | True | True | Sex-specific 1e-2 VUMC, sex-differential UKB |
| 0.003367 | 0.003643 | 6.424 | 0.2049 | 33.71 | 27.1 | True | False | True | True | Sex-specific 1e-2 VUMC, sex-differential UKB |
| 0.003363 | 0.003639 | 6.03 | 0.1309 | 33.42 | 26.97 | True | False | True | True | Sex-specific 1e-2 VUMC, sex-differential UKB |
| 0.003363 | 0.003639 | 6.043 | 0.132 | 33.41 | 26.97 | True | False | True | True | Sex-specific 1e-2 VUMC, sex-differential UKB |
| 0.003363 | 0.00364 | 6.01 | 0.1312 | 33.42 | 26.93 | True | False | True | True | Sex-specific 1e-2 VUMC, sex-differential UKB |
| 0.003363 | 0.00364 | 6.012 | 0.1315 | 33.42 | 26.92 | True | False | True | True | Sex-specific 1e-2 VUMC, sex-differential UKB |
| 0.003363 | 0.00364 | 6.012 | 0.1315 | 33.42 | 26.92 | True | False | True | True | Sex-specific 1e-2 VUMC, sex-differential UKB |
| 0.003363 | 0.00364 | 6.012 | 0.1314 | 33.42 | 26.96 | True | False | True | True | Sex-specific 1e-2 VUMC, sex-differential UKB |
| 0.003363 | 0.003639 | 6.074 | 0.1245 | 33.46 | 26.94 | True | False | True | True | Sex-specific 1e-2 VUMC, sex-differential UKB |
| 0.003363 | 0.003639 | 6.067 | 0.1245 | 33.46 | 26.93 | True | False | True | True | Sex-specific 1e-2 VUMC, sex-differential UKB |
| 0.003363 | 0.003639 | 6.067 | 0.1243 | 33.42 | 26.95 | True | False | True | True | Sex-specific 1e-2 VUMC, sex-differential UKB |
| 0.003362 | 0.003638 | 5.994 | 0.1365 | 32.92 | 26.62 | True | False | True | True | Sex-specific 1e-2 VUMC, sex-differential UKB |
| 0.003363 | 0.00364 | 6.231 | 0.117 | 32.94 | 26.89 | True | False | True | True | Sex-specific 1e-2 VUMC, sex-differential UKB |
| 0.003363 | 0.00364 | 6.231 | 0.117 | 32.94 | 26.89 | True | False | True | True | Sex-specific 1e-2 VUMC, sex-differential UKB |

| SE_UKB_female | SE_UKB_male | LOG10P_VUMC_female | LOG10P_VUMC_male | LOG10P_UKB_female | LOG10P_UKB_male | female_effect_direction_replication | male_effect_direction_replication | sex_effect_comparison_replication | pvalue_effect_comparison_replication | replication_type |
|---|---|---|---|---|---|---|---|---|---|---|
| 0.003363 | 0.003639 | 6.212 | 0.1199 | 32.95 | 26.92 | True | False | True | True | Sex-specific 1e-2 VUMC, sex-differential UKB |
| 0.003363 | 0.003639 | 6.212 | 0.1199 | 32.96 | 26.92 | True | False | True | True | Sex-specific 1e-2 VUMC, sex-differential UKB |
| 0.003363 | 0.003639 | 6.212 | 0.1199 | 32.96 | 26.92 | True | False | True | True | Sex-specific 1e-2 VUMC, sex-differential UKB |
| 0.003363 | 0.003639 | 6.212 | 0.1199 | 32.96 | 26.92 | True | False | True | True | Sex-specific 1e-2 VUMC, sex-differential UKB |
| 0.003363 | 0.003639 | 6.208 | 0.1199 | 32.94 | 26.91 | True | False | True | True | Sex-specific 1e-2 VUMC, sex-differential UKB |
| 0.003363 | 0.003639 | 6.211 | 0.1199 | 32.96 | 26.91 | True | False | True | True | Sex-specific 1e-2 VUMC, sex-differential UKB |
| 0.003389 | 0.003668 | 6.05 | 0.1075 | 32.71 | 26.57 | True | False | True | True | Sex-specific 1e-2 VUMC, sex-differential UKB |
| 0.003391 | 0.00367 | 6.03 | 0.09334 | 32.78 | 26.54 | True | False | True | True | Sex-specific 1e-2 VUMC, sex-differential UKB |
| 0.00339 | 0.003669 | 6.066 | 0.09623 | 32.61 | 26.49 | True | False | True | True | Sex-specific 1e-2 VUMC, sex-differential UKB |
| 0.003363 | 0.003639 | 6.246 | 0.1165 | 32.99 | 26.88 | True | False | True | True | Sex-specific 1e-2 VUMC, sex-differential UKB |
| 0.003363 | 0.003639 | 6.202 | 0.1462 | 32.92 | 26.86 | True | False | True | True | Sex-specific 1e-2 VUMC, sex-differential UKB |
| 0.003363 | 0.003639 | 6.197 | 0.1481 | 32.96 | 26.82 | True | False | True | True | Sex-specific 1e-2 VUMC, sex-differential UKB |
| 0.003368 | 0.003644 | 6.208 | 0.1431 | 32.84 | 26.8 | True | False | True | True | Sex-specific 1e-2 VUMC, sex-differential UKB |
| 0.003368 | 0.003644 | 6.208 | 0.1431 | 32.86 | 26.81 | True | False | True | True | Sex-specific 1e-2 VUMC, sex-differential UKB |
| 0.003409 | 0.003688 | 6.213 | 0.1212 | 31.01 | 26.65 | True | False | False | True | |
| 0.003411 | 0.003691 | 6.113 | 0.1308 | 31.11 | 26.61 | True | False | True | True | Sex-specific 1e-2 VUMC, sex-differential UKB |
| 0.003363 | 0.003638 | 6.197 | 0.1405 | 32.95 | 26.83 | True | False | True | True | Sex-specific 1e-2 VUMC, sex-differential UKB |
| 0.003367 | 0.003643 | 6.126 | 0.1158 | 32.86 | 26.75 | True | False | True | True | Sex-specific 1e-2 VUMC, sex-differential UKB |
| 0.003363 | 0.003638 | 6.187 | 0.1371 | 32.95 | 26.82 | True | False | True | True | Sex-specific 1e-2 VUMC, sex-differential UKB |
| 0.003363 | 0.003638 | 6.187 | 0.1384 | 32.97 | 26.76 | True | False | True | True | Sex-specific 1e-2 VUMC, sex-differential UKB |
| 0.003363 | 0.003638 | 6.187 | 0.1371 | 32.95 | 26.82 | True | False | True | True | Sex-specific 1e-2 VUMC, sex-differential UKB |
| 0.003363 | 0.003638 | 6.201 | 0.1427 | 32.99 | 26.77 | True | False | True | True | Sex-specific 1e-2 VUMC, sex-differential UKB |
| 0.003363 | 0.003638 | 6.201 | 0.1418 | 32.97 | 26.78 | True | False | True | True | Sex-specific 1e-2 VUMC, sex-differential UKB |
| 0.003363 | 0.003638 | 6.211 | 0.1423 | 32.99 | 26.78 | True | False | True | True | Sex-specific 1e-2 VUMC, sex-differential UKB |
| 0.003363 | 0.003638 | 6.187 | 0.1371 | 32.96 | 26.76 | True | False | True | True | Sex-specific 1e-2 VUMC, sex-differential UKB |
| 0.003363 | 0.003638 | 6.187 | 0.1371 | 32.96 | 26.76 | True | False | True | True | Sex-specific 1e-2 VUMC, sex-differential UKB |

| SE_UKB_female | SE_UKB_male | LOG10P_VUMC_female | LOG10P_VUMC_male | LOG10P_UKB_female | LOG10P_UKB_male | female_effect_direction_replication | male_effect_direction_replication | sex_effect_comparison_replication | pvalue_effect_comparison_replication | replication_type |
|---|---|---|---|---|---|---|---|---|---|---|
| 0.003363 | 0.003638 | 6.202 | 0.1419 | 32.98 | 26.78 | True | False | True | True | Sex-specific 1e-2 VUMC, sex-differential UKB |
| 0.003363 | 0.003638 | 6.202 | 0.1419 | 32.98 | 26.78 | True | False | True | True | Sex-specific 1e-2 VUMC, sex-differential UKB |

| SE_UKB_female | SE_UKB_male | LOG10P_VUMC_female | LOG10P_VUMC_male | LOG10P_UKB_female | LOG10P_UKB_male | female_effect_direction_replication | male_effect_direction_replication | sex_effect_comparison_replication | pvalue_effect_comparison_replication | replication_type |
|---|---|---|---|---|---|---|---|---|---|---|
| 0.003363 | 0.003638 | 6.202 | 0.1419 | 32.98 | 26.78 | True | False | True | True | Sex-specific 1e-2 VUMC, sex-differential UKB |
| 0.003363 | 0.003638 | 6.202 | 0.1419 | 32.98 | 26.78 | True | False | True | True | Sex-specific 1e-2 VUMC, sex-differential UKB |
| 0.003363 | 0.003638 | 6.202 | 0.1419 | 32.98 | 26.78 | True | False | True | True | Sex-specific 1e-2 VUMC, sex-differential UKB |
| 0.003363 | 0.003638 | 6.202 | 0.1419 | 32.98 | 26.78 | True | False | True | True | Sex-specific 1e-2 VUMC, sex-differential UKB |
| 0.003363 | 0.003638 | 6.202 | 0.1419 | 32.98 | 26.78 | True | False | True | True | Sex-specific 1e-2 VUMC, sex-differential UKB |
| 0.003363 | 0.003638 | 6.186 | 0.139 | 32.91 | 26.78 | True | False | True | True | Sex-specific 1e-2 VUMC, sex-differential UKB |
| 0.003363 | 0.003638 | 6.202 | 0.1419 | 32.98 | 26.78 | True | False | True | True | Sex-specific 1e-2 VUMC, sex-differential UKB |
| 0.003363 | 0.003638 | 6.202 | 0.1419 | 32.98 | 26.78 | True | False | True | True | Sex-specific 1e-2 VUMC, sex-differential UKB |
| 0.003363 | 0.003638 | 6.202 | 0.1417 | 33.07 | 26.82 | True | False | True | True | Sex-specific 1e-2 VUMC, sex-differential UKB |
| 0.003363 | 0.003638 | 6.192 | 0.1369 | 32.98 | 26.78 | True | False | True | True | Sex-specific 1e-2 VUMC, sex-differential UKB |
| 0.003363 | 0.003638 | 6.192 | 0.1369 | 32.98 | 26.78 | True | False | True | True | Sex-specific 1e-2 VUMC, sex-differential UKB |
| 0.003363 | 0.003638 | 6.192 | 0.1369 | 32.98 | 26.78 | True | False | True | True | Sex-specific 1e-2 VUMC, sex-differential UKB |
| 0.003363 | 0.003638 | 6.192 | 0.1369 | 32.98 | 26.78 | True | False | True | True | Sex-specific 1e-2 VUMC, sex-differential UKB |
| 0.003363 | 0.003638 | 6.191 | 0.1368 | 32.98 | 26.78 | True | False | True | True | Sex-specific 1e-2 VUMC, sex-differential UKB |
| 0.003369 | 0.003646 | 6.168 | 0.1384 | 33.45 | 25.91 | True | False | True | True | Sex-specific 1e-2 VUMC, sex-differential UKB |
| 0.003368 | 0.003645 | 6.142 | 0.1366 | 33.43 | 25.98 | True | False | True | True | Sex-specific 1e-2 VUMC, sex-differential UKB |
| 0.003362 | 0.003638 | 6.192 | 0.1369 | 32.93 | 26.86 | True | False | True | True | Sex-specific 1e-2 VUMC, sex-differential UKB |
| 0.003363 | 0.003638 | 6.196 | 0.1398 | 32.97 | 26.77 | True | False | True | True | Sex-specific 1e-2 VUMC, sex-differential UKB |
| 0.003363 | 0.003638 | 6.195 | 0.141 | 32.97 | 26.78 | True | False | True | True | Sex-specific 1e-2 VUMC, sex-differential UKB |
| 0.003363 | 0.003638 | 6.195 | 0.1399 | 32.97 | 26.78 | True | False | True | True | Sex-specific 1e-2 VUMC, sex-differential UKB |
| 0.003363 | 0.003638 | 6.195 | 0.1398 | 32.97 | 26.78 | True | False | True | True | Sex-specific 1e-2 VUMC, sex-differential UKB |
| 0.003363 | 0.003638 | 6.198 | 0.1355 | 32.97 | 26.78 | True | False | True | True | Sex-specific 1e-2 VUMC, sex-differential UKB |
| 0.003363 | 0.003638 | 6.198 | 0.1355 | 32.97 | 26.78 | True | False | True | True | Sex-specific 1e-2 VUMC, sex-differential UKB |

| SE_UKB_female | SE_UKB_male | LOG10P_VUMC_female | LOG10P_VUMC_male | LOG10P_UKB_female | LOG10P_UKB_male | female_effect_direction_replication | male_effect_direction_replication | sex_effect_comparison_replication | pvalue_effect_comparison_replication | replication_type |
|---|---|---|---|---|---|---|---|---|---|---|
| 0.003366 | 0.003642 | 6.587 | 0.1654 | 33.32 | 27.01 | True | False | True | True | Sex-specific 1e-2 VUMC, sex-differential UKB |
| 0.003363 | 0.003638 | 6.194 | 0.1326 | 32.99 | 26.78 | True | False | True | True | Sex-specific 1e-2 VUMC, sex-differential UKB |
| 0.003363 | 0.003638 | 6.194 | 0.1326 | 32.99 | 26.78 | True | False | True | True | Sex-specific 1e-2 VUMC, sex-differential UKB |
| 0.003363 | 0.003638 | 6.193 | 0.1328 | 32.98 | 26.79 | True | False | True | True | Sex-specific 1e-2 VUMC, sex-differential UKB |
| 0.003363 | 0.003638 | 6.194 | 0.1326 | 32.99 | 26.78 | True | False | True | True | Sex-specific 1e-2 VUMC, sex-differential UKB |
| 0.003366 | 0.003642 | 6.59 | 0.1619 | 33.32 | 27.01 | True | False | True | True | Sex-specific 1e-2 VUMC, sex-differential UKB |
| 0.003363 | 0.003638 | 6.183 | 0.1326 | 32.91 | 26.74 | True | False | True | True | Sex-specific 1e-2 VUMC, sex-differential UKB |
| 0.003363 | 0.003638 | 6.194 | 0.1326 | 32.99 | 26.78 | True | False | True | True | Sex-specific 1e-2 VUMC, sex-differential UKB |
| 0.003363 | 0.003638 | 6.194 | 0.1326 | 32.99 | 26.78 | True | False | True | True | Sex-specific 1e-2 VUMC, sex-differential UKB |
| 0.003367 | 0.003643 | 6.578 | 0.1633 | 33.28 | 26.96 | True | False | True | True | Sex-specific 1e-2 VUMC, sex-differential UKB |
| 0.003363 | 0.003638 | 6.194 | 0.1326 | 32.99 | 26.76 | True | False | True | True | Sex-specific 1e-2 VUMC, sex-differential UKB |
| 0.003363 | 0.003638 | 6.194 | 0.1326 | 32.99 | 26.76 | True | False | True | True | Sex-specific 1e-2 VUMC, sex-differential UKB |
| 0.003363 | 0.003638 | 6.194 | 0.1326 | 32.99 | 26.76 | True | False | True | True | Sex-specific 1e-2 VUMC, sex-differential UKB |
| 0.003363 | 0.003638 | 6.194 | 0.1326 | 32.99 | 26.76 | True | False | True | True | Sex-specific 1e-2 VUMC, sex-differential UKB |
| 0.003363 | 0.003638 | 6.194 | 0.1326 | 32.99 | 26.76 | True | False | True | True | Sex-specific 1e-2 VUMC, sex-differential UKB |
| 0.003363 | 0.003638 | 6.193 | 0.1325 | 32.98 | 26.76 | True | False | True | True | Sex-specific 1e-2 VUMC, sex-differential UKB |
| 0.003363 | 0.003638 | 6.193 | 0.1325 | 32.98 | 26.76 | True | False | True | True | Sex-specific 1e-2 VUMC, sex-differential UKB |
| 0.003363 | 0.003639 | 6.204 | 0.1325 | 33.09 | 26.7 | True | False | True | True | Sex-specific 1e-2 VUMC, sex-differential UKB |
| 0.003363 | 0.003639 | 6.204 | 0.1328 | 33.1 | 26.7 | True | False | True | True | Sex-specific 1e-2 VUMC, sex-differential UKB |
| 0.003363 | 0.003639 | 6.204 | 0.1325 | 33.1 | 26.7 | True | False | True | True | Sex-specific 1e-2 VUMC, sex-differential UKB |
| 0.003364 | 0.00364 | 6.144 | 0.1285 | 33.16 | 26.62 | True | False | True | True | Sex-specific 1e-2 VUMC, sex-differential UKB |
| 0.003364 | 0.00364 | 6.194 | 0.1331 | 33.23 | 26.71 | True | False | True | True | Sex-specific 1e-2 VUMC, sex-differential UKB |
| 0.003363 | 0.003639 | 6.206 | 0.1326 | 33.1 | 26.7 | True | False | True | True | Sex-specific 1e-2 VUMC, sex-differential UKB |
| 0.003363 | 0.003639 | 6.204 | 0.1325 | 33.09 | 26.7 | True | False | True | True | Sex-specific 1e-2 VUMC, sex-differential UKB |
| 0.003363 | 0.003639 | 6.206 | 0.1326 | 33.1 | 26.7 | True | False | True | True | Sex-specific 1e-2 VUMC, sex-differential UKB |
| 0.003363 | 0.003639 | 6.204 | 0.1325 | 33.09 | 26.7 | True | False | True | True | Sex-specific 1e-2 VUMC, sex-differential UKB |

| SE_UKB_female | SE_UKB_male | LOG10P_VUMC_female | LOG10P_VUMC_male | LOG10P_UKB_female | LOG10P_UKB_male | female_effect_direction_replication | male_effect_direction_replication | sex_effect_comparison_replication | pvalue_effect_comparison_replication | replication_type |
|---|---|---|---|---|---|---|---|---|---|---|
| 0.003363 | 0.003639 | 6.204 | 0.1325 | 33.09 | 26.7 | True | False | True | True | Sex-specific 1e-2 VUMC, sex-differential UKB |
| 0.003363 | 0.003639 | 6.204 | 0.1325 | 33.09 | 26.7 | True | False | True | True | Sex-specific 1e-2 VUMC, sex-differential UKB |
| 0.003364 | 0.00364 | 6.207 | 0.1323 | 33.09 | 26.7 | True | False | True | True | Sex-specific 1e-2 VUMC, sex-differential UKB |
| 0.003367 | 0.003643 | 6.617 | 0.1652 | 33.41 | 26.92 | True | False | True | True | Sex-specific 1e-2 VUMC, sex-differential UKB |
| 0.003368 | 0.003644 | 6.622 | 0.1413 | 33.51 | 26.87 | True | False | True | True | Sex-specific 1e-2 VUMC, sex-differential UKB |
| 0.003368 | 0.003644 | 6.599 | 0.141 | 33.5 | 26.9 | True | False | True | True | Sex-specific 1e-2 VUMC, sex-differential UKB |
| 0.003368 | 0.003644 | 6.602 | 0.1427 | 33.53 | 26.92 | True | False | True | True | Sex-specific 1e-2 VUMC, sex-differential UKB |
| 0.003368 | 0.003644 | 6.612 | 0.1402 | 33.54 | 26.92 | True | False | True | True | Sex-specific 1e-2 VUMC, sex-differential UKB |
| 0.003363 | 0.003639 | 6.195 | 0.1095 | 33.11 | 26.78 | True | False | True | True | Sex-specific 1e-2 VUMC, sex-differential UKB |
| 0.003363 | 0.003639 | 6.198 | 0.1124 | 33.12 | 26.75 | True | False | True | True | Sex-specific 1e-2 VUMC, sex-differential UKB |
| 0.003361 | 0.003636 | 5.764 | 0.06995 | 32.63 | 27.16 | True | False | True | True | Sex-specific 1e-2 VUMC, sex-differential UKB |
| SE_UKB_female | SE_UKB_male | LOG10P_VUMC_female | LOG10P_VUMC_male | LOG10P_UKB_female | LOG10P_UKB_male | female_effect_direction_replication | male_effect_direction_replication | sex_effect_comparison_replication | pvalue_effect_comparison_replication | replication_type |
| 0.003358 | 0.003633 | 5.572 | 0.02876 | 32.31 | 26.98 | True | False | True | True | Sex-specific 1e-2 VUMC, sex-differential UKB |
| 0.003367 | 0.003644 | 6.603 | 0.1428 | 33.46 | 26.94 | True | False | True | True | Sex-specific 1e-2 VUMC, sex-differential UKB |
| 0.003363 | 0.003639 | 6.204 | 0.1117 | 33.13 | 26.71 | True | False | True | True | Sex-specific 1e-2 VUMC, sex-differential UKB |
| 0.003367 | 0.003644 | 6.597 | 0.1363 | 33.47 | 26.95 | True | False | True | True | Sex-specific 1e-2 VUMC, sex-differential UKB |
| 0.003367 | 0.003644 | 6.606 | 0.1376 | 33.48 | 26.96 | True | False | True | True | Sex-specific 1e-2 VUMC, sex-differential UKB |
| 0.003361 | 0.003637 | 5.786 | 0.06099 | 32.7 | 27.01 | True | False | True | True | Sex-specific 1e-2 VUMC, sex-differential UKB |
| 0.003364 | 0.00364 | 6.171 | 0.1067 | 33.02 | 26.76 | True | False | True | True | Sex-specific 1e-2 VUMC, sex-differential UKB |
| 0.00337 | 0.003647 | 6.588 | 0.1295 | 33.72 | 26.94 | True | False | True | True | Sex-specific 1e-2 VUMC, sex-differential UKB |
| 0.003367 | 0.003644 | 6.283 | 0.08127 | 33.56 | 26.82 | True | False | True | True | Sex-specific 1e-2 VUMC, sex-differential UKB |
| 0.00337 | 0.003648 | 6.528 | 0.1176 | 34 | 26.9 | True | False | True | True | Sex-specific 1e-2 VUMC, sex-differential UKB |
| 0.003371 | 0.003648 | 6.567 | 0.1202 | 34.1 | 26.81 | True | False | True | True | Sex-specific 1e-2 VUMC, sex-differential UKB |
| 0.003367 | 0.003644 | 6.312 | 0.0564 | 33.85 | 26.7 | True | False | True | True | Sex-specific 1e-2 VUMC, sex-differential UKB |
| 0.003416 | 0.003697 | 5.879 | 0.04314 | 35.37 | 26.29 | True | True | True | True | Sex-specific 1e-2 VUMC, sex-differential UKB |
| 0.003392 | 0.003671 | 5.93 | 0.02173 | 35.1 | 26.47 | True | True | True | True | Sex-specific 1e-2 VUMC, sex-differential UKB |

| SE_UKB_female | SE_UKB_male | LOG10P_VUMC_female | LOG10P_VUMC_male | LOG10P_UKB_female | LOG10P_UKB_male | female_effect_direction_replication | male_effect_direction_replication | sex_effect_comparison_replication | pvalue_effect_comparison_replication | replication_type |
|---|---|---|---|---|---|---|---|---|---|---|
| 0.003391 | 0.00367 | 6.008 | 0.04012 | 35.09 | 26.35 | True | True | True | True | Sex-specific 1e-2 VUMC, sex-differential UKB |
| 0.003391 | 0.00367 | 5.994 | 0.03966 | 35.09 | 26.34 | True | True | True | True | Sex-specific 1e-2 VUMC, sex-differential UKB |
| 0.003392 | 0.00367 | 5.997 | 0.03899 | 35.09 | 26.34 | True | True | True | True | Sex-specific 1e-2 VUMC, sex-differential UKB |
| 0.003391 | 0.00367 | 5.954 | 0.03244 | 35.05 | 26.39 | True | True | True | True | Sex-specific 1e-2 VUMC, sex-differential UKB |
| 0.003391 | 0.00367 | 5.954 | 0.0323 | 35.04 | 26.36 | True | True | True | True | Sex-specific 1e-2 VUMC, sex-differential UKB |
| 0.003391 | 0.00367 | 5.953 | 0.03227 | 35.04 | 26.36 | True | True | True | True | Sex-specific 1e-2 VUMC, sex-differential UKB |
| 0.003391 | 0.00367 | 5.905 | 0.03666 | 35.05 | 26.39 | True | True | True | True | Sex-specific 1e-2 VUMC, sex-differential UKB |
| 0.003391 | 0.00367 | 5.964 | 0.04263 | 34.9 | 26.24 | True | True | True | True | Sex-specific 1e-2 VUMC, sex-differential UKB |
| 0.003392 | 0.003671 | 5.88 | 0.05181 | 34.84 | 26.01 | True | True | True | True | Sex-specific 1e-2 VUMC, sex-differential UKB |
| 0.003392 | 0.00367 | 5.89 | 0.05036 | 34.93 | 26.06 | True | True | True | True | Sex-specific 1e-2 VUMC, sex-differential UKB |
| 0.003392 | 0.00367 | 5.891 | 0.05234 | 34.93 | 26.1 | True | True | True | True | Sex-specific 1e-2 VUMC, sex-differential UKB |
| 0.003392 | 0.003671 | 5.891 | 0.05182 | 34.9 | 26.01 | True | True | True | True | Sex-specific 1e-2 VUMC, sex-differential UKB |
| 0.003391 | 0.00367 | 5.893 | 0.05196 | 34.9 | 26.06 | True | True | True | True | Sex-specific 1e-2 VUMC, sex-differential UKB |
| 0.003391 | 0.00367 | 5.907 | 0.06078 | 35.02 | 26.03 | True | True | True | True | Sex-specific 1e-2 VUMC, sex-differential UKB |
| 0.003391 | 0.00367 | 5.912 | 0.06125 | 34.96 | 25.97 | True | True | True | True | Sex-specific 1e-2 VUMC, sex-differential UKB |
| 0.003391 | 0.00367 | 5.922 | 0.06024 | 35.07 | 26 | True | True | True | True | Sex-specific 1e-2 VUMC, sex-differential UKB |
| 0.003391 | 0.00367 | 5.919 | 0.06039 | 35.01 | 26 | True | True | True | True | Sex-specific 1e-2 VUMC, sex-differential UKB |
| 0.003261 | 0.003521 | 4.939 | 0.07395 | 24.32 | 17.67 | True | True | True | True | Sex-specific 1e-2 VUMC, sex-differential UKB |
| 0.003395 | 0.003674 | 5.91 | 0.02827 | 35.02 | 26.02 | True | True | True | True | Sex-specific 1e-2 VUMC, sex-differential UKB |
| 0.003391 | 0.00367 | 5.885 | 0.04967 | 35.01 | 26.05 | True | True | True | True | Sex-specific 1e-2 VUMC, sex-differential UKB |
| 0.003391 | 0.003669 | 5.889 | 0.06446 | 35.04 | 25.95 | True | True | True | True | Sex-specific 1e-2 VUMC, sex-differential UKB |
| 0.003391 | 0.003669 | 5.889 | 0.06489 | 35.06 | 25.95 | True | True | True | True | Sex-specific 1e-2 VUMC, sex-differential UKB |
| 0.00339 | 0.003669 | 5.831 | 0.05602 | 35.07 | 25.92 | True | True | True | True | Sex-specific 1e-2 VUMC, sex-differential UKB |
| 0.00339 | 0.003669 | 5.885 | 0.05346 | 35.09 | 25.92 | True | True | True | True | Sex-specific 1e-2 VUMC, sex-differential UKB |
| 0.00339 | 0.003669 | 5.861 | 0.05518 | 35 | 25.9 | True | True | True | True | Sex-specific 1e-2 VUMC, sex-differential UKB |
| 0.00339 | 0.003669 | 5.836 | 0.05718 | 35.16 | 25.89 | True | True | True | True | Sex-specific 1e-2 VUMC, sex-differential UKB |

| SE_UKB_female | SE_UKB_male | LOG10P_VUMC_female | LOG10P_VUMC_male | LOG10P_UKB_female | LOG10P_UKB_male | female_effect_direction_replication | male_effect_direction_replication | sex_effect_comparison_replication | pvalue_effect_comparison_replication | replication_type |
|---|---|---|---|---|---|---|---|---|---|---|
| 0.00339 | 0.003669 | 5.836 | 0.0553 | 35.16 | 25.89 | True | True | True | True | Sex-specific 1e-2 VUMC, sex-differential UKB |
| 0.00339 | 0.003669 | 5.837 | 0.05692 | 35.16 | 25.88 | True | True | True | True | Sex-specific 1e-2 VUMC, sex-differential UKB |
| 0.00339 | 0.003669 | 5.836 | 0.055 | 35.21 | 25.88 | True | True | True | True | Sex-specific 1e-2 VUMC, sex-differential UKB |
| 0.00339 | 0.003669 | 5.849 | 0.05553 | 35.17 | 25.91 | True | True | True | True | Sex-specific 1e-2 VUMC, sex-differential UKB |
| 0.003392 | 0.003671 | 5.789 | 0.06502 | 35.12 | 26.03 | True | True | True | True | Sex-specific 1e-2 VUMC, sex-differential UKB |
| 0.00339 | 0.003669 | 5.833 | 0.06156 | 35.18 | 25.91 | True | True | True | True | Sex-specific 1e-2 VUMC, sex-differential UKB |
| 0.00339 | 0.003669 | 5.838 | 0.05819 | 35.14 | 25.9 | True | True | True | True | Sex-specific 1e-2 VUMC, sex-differential UKB |
| 0.00339 | 0.003669 | 5.833 | 0.06134 | 35.17 | 25.91 | True | True | True | True | Sex-specific 1e-2 VUMC, sex-differential UKB |
| 0.00339 | 0.003669 | 5.834 | 0.05822 | 35.17 | 25.92 | True | True | True | True | Sex-specific 1e-2 VUMC, sex-differential UKB |
| 0.00339 | 0.003669 | 5.834 | 0.06107 | 35.15 | 25.92 | True | True | True | True | Sex-specific 1e-2 VUMC, sex-differential UKB |
| 0.00339 | 0.003669 | 5.831 | 0.05884 | 35.21 | 25.97 | True | True | True | True | Sex-specific 1e-2 VUMC, sex-differential UKB |
| 0.003391 | 0.00367 | 5.831 | 0.05905 | 35.27 | 26.01 | True | True | True | True | Sex-specific 1e-2 VUMC, sex-differential UKB |
| 0.00338 | 0.003659 | 5.541 | 0.05647 | 33.9 | 27.05 | True | True | True | True | Sex-specific 1e-2 VUMC, sex-differential UKB |
| 0.00338 | 0.003659 | 5.61 | 0.06624 | 33.98 | 26.99 | True | True | True | True | Sex-specific 1e-2 VUMC, sex-differential UKB |
| 0.00338 | 0.003659 | 5.611 | 0.06726 | 33.92 | 27.08 | True | True | True | True | Sex-specific 1e-2 VUMC, sex-differential UKB |
| 0.003381 | 0.00366 | 5.499 | 0.05041 | 33.8 | 27.07 | True | True | True | True | Sex-specific 1e-2 VUMC, sex-differential UKB |
| 0.003379 | 0.003659 | 5.536 | 0.04709 | 33.81 | 27.1 | True | True | True | True | Sex-specific 1e-2 VUMC, sex-differential UKB |
| 0.003379 | 0.003659 | 5.532 | 0.04762 | 33.75 | 27.12 | True | True | True | True | Sex-specific 1e-2 VUMC, sex-differential UKB |
| 0.003383 | 0.003662 | 5.488 | 0.1036 | 33.65 | 26.95 | True | True | True | True | Sex-specific 1e-2 VUMC, sex-differential UKB |
| 0.00338 | 0.003659 | 5.548 | 0.07293 | 33.88 | 27.02 | True | True | True | True | Sex-specific 1e-2 VUMC, sex-differential UKB |
| SE_UKB_female | SE_UKB_male | LOG10P_VUMC_female | LOG10P_VUMC_male | LOG10P_UKB_female | LOG10P_UKB_male | female_effect_direction_replication | male_effect_direction_replication | sex_effect_comparison_replication | pvalue_effect_comparison_replication | replication_type |
| 0.003382 | 0.003663 | 5.53 | 0.074 | 33.78 | 26.89 | True | True | True | True | Sex-specific 1e-2 VUMC, sex-differential UKB |
| 0.00338 | 0.003659 | 5.564 | 0.07109 | 33.9 | 27.01 | True | True | True | True | Sex-specific 1e-2 VUMC, sex-differential UKB |
| 0.00338 | 0.003659 | 5.536 | 0.06411 | 33.91 | 26.91 | True | True | True | True | Sex-specific 1e-2 VUMC, sex-differential UKB |
| 0.00338 | 0.003659 | 5.51 | 0.04592 | 33.99 | 27.04 | True | True | True | True | Sex-specific 1e-2 VUMC, sex-differential UKB |
| 0.003381 | 0.00366 | 5.497 | 0.06721 | 33.91 | 27.16 | True | True | True | True | Sex-specific 1e-2 VUMC, sex-differential UKB |

| SE_UKB_female | SE_UKB_male | LOG10P_VUMC_female | LOG10P_VUMC_male | LOG10P_UKB_female | LOG10P_UKB_male | female_effect_direction_replication | male_effect_direction_replication | sex_effect_comparison_replication | pvalue_effect_comparison_replication | replication_type |
|---|---|---|---|---|---|---|---|---|---|---|
| 0.00338 | 0.003659 | 5.508 | 0.05533 | 34.09 | 27.15 | True | True | True | True | Sex-specific 1e-2 VUMC, sex-differential UKB |
| 0.00338 | 0.003659 | 5.512 | 0.05974 | 33.93 | 27.21 | True | True | True | True | Sex-specific 1e-2 VUMC, sex-differential UKB |
| 0.00338 | 0.003659 | 5.516 | 0.05889 | 34.05 | 27.19 | True | True | True | True | Sex-specific 1e-2 VUMC, sex-differential UKB |
| 0.003379 | 0.003658 | 5.542 | 0.07481 | 33.98 | 27.15 | True | True | True | True | Sex-specific 1e-2 VUMC, sex-differential UKB |
| 0.00338 | 0.003659 | 5.548 | 0.07449 | 33.98 | 27.2 | True | True | True | True | Sex-specific 1e-2 VUMC, sex-differential UKB |
| 0.00338 | 0.003659 | 5.549 | 0.0721 | 33.9 | 27.28 | True | True | True | True | Sex-specific 1e-2 VUMC, sex-differential UKB |
| 0.003381 | 0.00366 | 5.517 | 0.05449 | 34.04 | 27.12 | True | True | True | True | Sex-specific 1e-2 VUMC, sex-differential UKB |
| 0.003186 | 0.003449 | 4.984 | 0.4666 | 8.205 | 7.738 | True | True | False | True | |
| 0.00318 | 0.003442 | 4.976 | 0.481 | 8.209 | 7.412 | True | True | False | True | |
| 0.003819 | 0.00415 | 4.791 | 0.7882 | 17.44 | 15.39 | True | True | False | True | |
| 0.003811 | 0.004142 | 5.36 | 0.8167 | 17.45 | 15.83 | True | True | False | True | |
| | | 3.491 | 0.1913 | | | False | False | False | False | |
| 0.003789 | 0.004118 | 4.856 | 0.8244 | 17.31 | 15.95 | True | True | False | True | |
| 0.003338 | 0.003624 | 7.759 | 1.2 | 36.9 | 24.63 | True | True | True | True | Sex-specific 1e-2 VUMC, sex-differential UKB |
| 0.003798 | 0.004129 | 4.734 | 0.8557 | 17.8 | 15.85 | True | True | False | True | |
| 0.003269 | 0.003542 | 3.654 | 0.5038 | 8.078 | 6.272 | True | True | True | True | Sex-specific 1e-2 VUMC, sex-specific 5e-8 UKB |
| 0.003272 | 0.003545 | 3.668 | 0.5056 | 8.239 | 6.629 | True | True | True | True | Sex-specific 1e-2 VUMC, sex-specific 5e-8 UKB |
| 0.003272 | 0.003545 | 3.594 | 0.499 | 8.044 | 6.664 | True | True | True | True | Sex-specific 1e-2 VUMC, sex-specific 5e-8 UKB |
| 0.003284 | 0.003553 | 3.213 | 0.3833 | 7.445 | 5.671 | True | True | True | True | Sex-specific 1e-2 VUMC, sex-specific 5e-8 UKB |
| 0.003412 | 0.00371 | 2.583 | 0.2424 | 19.39 | 13.21 | True | True | True | True | Sex-specific 1e-2 VUMC, sex-differential UKB |
| 0.003214 | 0.003487 | 9.086 | 0.8415 | 44.56 | 37.43 | True | True | True | True | Sex-specific 1e-2 VUMC, sex-differential UKB |
| 0.003516 | 0.003815 | 6.882 | 1.495 | 57.92 | 51.34 | True | True | False | True | |
| 0.003515 | 0.003815 | 6.923 | 1.458 | 58.12 | 51.32 | True | True | False | True | |
| 0.003517 | 0.003816 | 7.207 | 1.361 | 58.07 | 51.91 | True | True | False | True | |
| 0.003215 | 0.003488 | 8.952 | 0.8314 | 44.71 | 37.44 | True | True | True | True | Sex-specific 1e-2 VUMC, sex-differential UKB |
| 0.003516 | 0.003815 | 7.233 | 1.323 | 58.08 | 51.75 | True | True | False | True | |
| 0.003511 | 0.003811 | 7.003 | 1.429 | 59.1 | 51.55 | True | True | False | True | |

| SE_UKB_female | SE_UKB_male | LOG10P_VUMC_female | LOG10P_VUMC_male | LOG10P_UKB_female | LOG10P_UKB_male | female_effect_direction_replication | male_effect_direction_replication | sex_effect_comparison_replication | pvalue_effect_comparison_replication | replication_type |
|---|---|---|---|---|---|---|---|---|---|---|
| 0.003527 | 0.003828 | 6.707 | 1.304 | 57.46 | 50.05 | True | True | False | True | |
| 0.003522 | 0.003821 | 6.876 | 1.357 | 57.69 | 51.29 | True | True | False | True | |
| 0.003215 | 0.003487 | 9.037 | 0.8262 | 44.71 | 36.89 | True | True | True | True | Sex-specific 1e-2 VUMC, sex-differential UKB |
| 0.003523 | 0.003822 | 6.719 | 1.147 | 57.1 | 50.18 | True | True | False | True | |
| 0.003224 | 0.003494 | 8.5 | 0.8081 | 46.05 | 38.52 | True | True | True | True | Sex-specific 1e-2 VUMC, sex-differential UKB |
| 0.003521 | 0.003818 | 6.935 | 1.259 | 57.42 | 49.96 | True | True | False | True | |
| 0.003524 | 0.003821 | 7.157 | 1.234 | 57.26 | 50.43 | True | True | False | True | |
| 0.006293 | 0.006771 | 5.657 | 0.3208 | 29.07 | 19.92 | True | True | True | True | Sex-specific 1e-2 VUMC, sex-differential UKB |
| 0.006518 | 0.007013 | 6.197 | 0.3341 | 34.24 | 21.46 | True | True | True | True | Sex-specific 1e-2 VUMC, sex-differential UKB |
| 0.006738 | 0.007249 | 7.642 | 1.038 | 44.32 | 32.65 | True | True | True | True | Sex-specific 1e-2 VUMC, sex-differential UKB |
| 0.006547 | 0.007054 | 7.45 | 0.8393 | 44.32 | 37 | True | True | True | True | Sex-specific 1e-2 VUMC, sex-differential UKB |
| 0.005379 | 0.005799 | 7.702 | 1.05 | 43.13 | 25.96 | True | True | True | True | Sex-specific 1e-2 VUMC, sex-differential UKB |
| 0.005748 | 0.006189 | 8.704 | 0.6019 | 46.44 | 31.83 | True | True | True | True | Sex-specific 1e-2 VUMC, sex-differential UKB |
| 0.005657 | 0.00609 | 7.066 | 0.5075 | 34.07 | 22.31 | True | True | True | True | Sex-specific 1e-2 VUMC, sex-differential UKB |
| | | 6.697 | 0.8911 | | | False | False | False | False | |
| 0.005388 | 0.005812 | 6.842 | 0.9093 | 35.18 | 20.45 | True | True | True | True | Sex-specific 1e-2 VUMC, sex-differential UKB |
| 0.005387 | 0.00581 | 6.842 | 0.9093 | 35.24 | 20.2 | True | True | True | True | Sex-specific 1e-2 VUMC, sex-differential UKB |
| 0.005387 | 0.00581 | 6.842 | 0.9093 | 35.24 | 20.2 | True | True | True | True | Sex-specific 1e-2 VUMC, sex-differential UKB |
| 0.005387 | 0.00581 | 6.844 | 0.9092 | 35.24 | 20.2 | True | True | True | True | Sex-specific 1e-2 VUMC, sex-differential UKB |
| 0.005387 | 0.00581 | 6.842 | 0.9093 | 35.24 | 20.2 | True | True | True | True | Sex-specific 1e-2 VUMC, sex-differential UKB |
| 0.005387 | 0.00581 | 6.842 | 0.9093 | 35.24 | 20.2 | True | True | True | True | Sex-specific 1e-2 VUMC, sex-differential UKB |
| 0.005387 | 0.00581 | 6.842 | 0.9093 | 35.24 | 20.2 | True | True | True | True | Sex-specific 1e-2 VUMC, sex-differential UKB |
| 0.005387 | 0.00581 | 6.842 | 0.9093 | 35.24 | 20.2 | True | True | True | True | Sex-specific 1e-2 VUMC, sex-differential UKB |
| 0.005387 | 0.00581 | 6.842 | 0.9093 | 35.24 | 20.2 | True | True | True | True | Sex-specific 1e-2 VUMC, sex-differential UKB |
| 0.005384 | 0.005807 | 6.719 | 0.916 | 35.43 | 20.32 | True | True | True | True | Sex-specific 1e-2 VUMC, sex-differential UKB |
| 0.005386 | 0.005809 | 6.86 | 0.8873 | 35.24 | 20.23 | True | True | True | True | Sex-specific 1e-2 VUMC, sex-differential UKB |

| SE_UKB_female | SE_UKB_male | LOG10P_VUMC_female | LOG10P_VUMC_male | LOG10P_UKB_female | LOG10P_UKB_male | female_effect_direction_replication | male_effect_direction_replication | sex_effect_comparison_replication | pvalue_effect_comparison_replication | replication_type |
|---|---|---|---|---|---|---|---|---|---|---|
| 0.005387 | 0.00581 | 6.842 | 0.9093 | 35.24 | 20.2 | True | True | True | True | Sex-specific 1e-2 VUMC, sex-differential UKB |
| 0.005387 | 0.00581 | 6.842 | 0.9093 | 35.24 | 20.2 | True | True | True | True | Sex-specific 1e-2 VUMC, sex-differential UKB |

| SE_UKB_female | SE_UKB_male | LOG10P_VUMC_female | LOG10P_VUMC_male | LOG10P_UKB_female | LOG10P_UKB_male | female_effect_direction_replication | male_effect_direction_replication | sex_effect_comparison_replication | pvalue_effect_comparison_replication | replication_type |
|---|---|---|---|---|---|---|---|---|---|---|
| 0.005387 | 0.00581 | 6.898 | 0.9148 | 35.24 | 20.2 | True | True | True | True | Sex-specific 1e-2 VUMC, sex-differential UKB |
| 0.005387 | 0.00581 | 6.898 | 0.9148 | 35.24 | 20.2 | True | True | True | True | Sex-specific 1e-2 VUMC, sex-differential UKB |
| 0.005387 | 0.00581 | 6.898 | 0.9148 | 35.24 | 20.2 | True | True | True | True | Sex-specific 1e-2 VUMC, sex-differential UKB |
| 0.005383 | 0.005805 | 6.898 | 0.9148 | 35.16 | 19.72 | True | True | True | True | Sex-specific 1e-2 VUMC, sex-differential UKB |
| 0.005387 | 0.00581 | 6.898 | 0.9148 | 35.24 | 20.2 | True | True | True | True | Sex-specific 1e-2 VUMC, sex-differential UKB |
| 0.005388 | 0.00581 | 6.921 | 0.9094 | 35.21 | 20.19 | True | True | True | True | Sex-specific 1e-2 VUMC, sex-differential UKB |
| 0.005387 | 0.00581 | 6.909 | 0.9148 | 35.22 | 20.21 | True | True | True | True | Sex-specific 1e-2 VUMC, sex-differential UKB |
| 0.005387 | 0.00581 | 6.909 | 0.9148 | 35.22 | 20.21 | True | True | True | True | Sex-specific 1e-2 VUMC, sex-differential UKB |
| 0.005387 | 0.00581 | 6.909 | 0.9148 | 35.22 | 20.21 | True | True | True | True | Sex-specific 1e-2 VUMC, sex-differential UKB |
| 0.005389 | 0.005812 | 6.929 | 0.9062 | 35.43 | 20.37 | True | True | True | True | Sex-specific 1e-2 VUMC, sex-differential UKB |
| 0.005387 | 0.00581 | 6.909 | 0.9148 | 35.22 | 20.21 | True | True | True | True | Sex-specific 1e-2 VUMC, sex-differential UKB |
| 0.005203 | 0.005623 | 6.291 | 0.9083 | 23.8 | 12.47 | True | True | True | True | Sex-specific 1e-2 VUMC, sex-differential UKB |
| 0.005387 | 0.00581 | 6.905 | 0.9134 | 35.25 | 20.23 | True | True | True | True | Sex-specific 1e-2 VUMC, sex-differential UKB |
| 0.005387 | 0.00581 | 6.909 | 0.9148 | 35.22 | 20.21 | True | True | True | True | Sex-specific 1e-2 VUMC, sex-differential UKB |
| 0.005388 | 0.005811 | 6.909 | 0.9148 | 35.15 | 20.24 | True | True | True | True | Sex-specific 1e-2 VUMC, sex-differential UKB |
| 0.005388 | 0.005811 | 6.909 | 0.9148 | 35.15 | 20.24 | True | True | True | True | Sex-specific 1e-2 VUMC, sex-differential UKB |
| | | 6.909 | 0.9148 | | | False | False | False | False | |
| 0.005388 | 0.005811 | 6.948 | 0.8847 | 35.14 | 20.25 | True | True | True | True | Sex-specific 1e-2 VUMC, sex-differential UKB |
| 0.005393 | 0.005816 | 6.957 | 0.9447 | 35.33 | 20.46 | True | True | True | True | Sex-specific 1e-2 VUMC, sex-differential UKB |
| 0.005393 | 0.005816 | 6.957 | 0.9447 | 35.33 | 20.46 | True | True | True | True | Sex-specific 1e-2 VUMC, sex-differential UKB |
| 0.005393 | 0.005816 | 6.957 | 0.9447 | 35.33 | 20.46 | True | True | True | True | Sex-specific 1e-2 VUMC, sex-differential UKB |
| 0.005393 | 0.005816 | 6.957 | 0.9447 | 35.33 | 20.46 | True | True | True | True | Sex-specific 1e-2 VUMC, sex-differential UKB |
| 0.005392 | 0.005815 | 6.957 | 0.9447 | 35.27 | 20.4 | True | True | True | True | Sex-specific 1e-2 VUMC, sex-differential UKB |

| SE_UKB_female | SE_UKB_male | LOG10P_VUMC_female | LOG10P_VUMC_male | LOG10P_UKB_female | LOG10P_UKB_male | female_effect_direction_replication | male_effect_direction_replication | sex_effect_comparison_replication | pvalue_effect_comparison_replication | replication_type |
|---|---|---|---|---|---|---|---|---|---|---|
| 0.005392 | 0.005815 | 6.956 | 0.945 | 35.27 | 20.4 | True | True | True | True | Sex-specific 1e-2 VUMC, sex-differential UKB |
| 0.005393 | 0.005816 | 6.957 | 0.9447 | 35.32 | 20.47 | True | True | True | True | Sex-specific 1e-2 VUMC, sex-differential UKB |
| 0.005393 | 0.005816 | 6.966 | 0.9448 | 35.32 | 20.47 | True | True | True | True | Sex-specific 1e-2 VUMC, sex-differential UKB |
| 0.005393 | 0.005816 | 6.966 | 0.9448 | 35.32 | 20.47 | True | True | True | True | Sex-specific 1e-2 VUMC, sex-differential UKB |
| 0.005393 | 0.005816 | 6.967 | 0.9449 | 35.33 | 20.49 | True | True | True | True | Sex-specific 1e-2 VUMC, sex-differential UKB |
| 0.005393 | 0.005816 | 6.969 | 0.9402 | 35.34 | 20.42 | True | True | True | True | Sex-specific 1e-2 VUMC, sex-differential UKB |
| 0.005393 | 0.005816 | 6.965 | 0.9476 | 35.32 | 20.47 | True | True | True | True | Sex-specific 1e-2 VUMC, sex-differential UKB |
| 0.005393 | 0.005816 | 6.955 | 0.9485 | 35.37 | 20.47 | True | True | True | True | Sex-specific 1e-2 VUMC, sex-differential UKB |
| 0.005393 | 0.005816 | 6.966 | 0.9478 | 35.33 | 20.51 | True | True | True | True | Sex-specific 1e-2 VUMC, sex-differential UKB |
| 0.005393 | 0.005815 | 6.954 | 0.9484 | 35.34 | 20.52 | True | True | True | True | Sex-specific 1e-2 VUMC, sex-differential UKB |
| 0.005393 | 0.005815 | 6.954 | 0.9484 | 35.34 | 20.53 | True | True | True | True | Sex-specific 1e-2 VUMC, sex-differential UKB |
| 0.005393 | 0.005815 | 6.954 | 0.9484 | 35.34 | 20.53 | True | True | True | True | Sex-specific 1e-2 VUMC, sex-differential UKB |
| 0.005395 | 0.005817 | 6.976 | 0.9397 | 35.54 | 20.69 | True | True | True | True | Sex-specific 1e-2 VUMC, sex-differential UKB |
| 0.005393 | 0.005815 | 6.954 | 0.9484 | 35.34 | 20.53 | True | True | True | True | Sex-specific 1e-2 VUMC, sex-differential UKB |
| 0.005392 | 0.005814 | 6.954 | 0.9481 | 35.29 | 20.32 | True | True | True | True | Sex-specific 1e-2 VUMC, sex-differential UKB |
| 0.005393 | 0.005815 | 6.976 | 0.9456 | 35.34 | 20.53 | True | True | True | True | Sex-specific 1e-2 VUMC, sex-differential UKB |
| 0.005387 | 0.005809 | 7.049 | 1.017 | 35.91 | 20.89 | True | True | True | True | Sex-specific 1e-2 VUMC, sex-differential UKB |
| 0.005393 | 0.005815 | 6.976 | 0.9456 | 35.34 | 20.53 | True | True | True | True | Sex-specific 1e-2 VUMC, sex-differential UKB |
| 0.005393 | 0.005815 | 6.98 | 0.9606 | 35.34 | 20.53 | True | True | True | True | Sex-specific 1e-2 VUMC, sex-differential UKB |
| 0.005393 | 0.005815 | 6.978 | 0.9416 | 35.34 | 20.53 | True | True | True | True | Sex-specific 1e-2 VUMC, sex-differential UKB |
| | | 6.98 | 0.9444 | | | False | False | False | False | |
| 0.005393 | 0.005815 | 6.98 | 0.9444 | 35.34 | 20.53 | True | True | True | True | Sex-specific 1e-2 VUMC, sex-differential UKB |
| 0.005393 | 0.005815 | 6.98 | 0.9444 | 35.34 | 20.53 | True | True | True | True | Sex-specific 1e-2 VUMC, sex-differential UKB |
| 0.005393 | 0.005815 | 6.973 | 0.9381 | 35.32 | 20.53 | True | True | True | True | Sex-specific 1e-2 VUMC, sex-differential UKB |
| 0.005393 | 0.005815 | 6.956 | 0.9349 | 35.34 | 20.53 | True | True | True | True | Sex-specific 1e-2 VUMC, sex-differential UKB |
| 0.005393 | 0.005815 | 6.957 | 0.935 | 35.34 | 20.53 | True | True | True | True | Sex-specific 1e-2 VUMC, sex-differential UKB |

| SE_UKB_female | SE_UKB_male | LOG10P_VUMC_female | LOG10P_VUMC_male | LOG10P_UKB_female | LOG10P_UKB_male | female_effect_direction_replication | male_effect_direction_replication | sex_effect_comparison_replication | pvalue_effect_comparison_replication | replication_type |
|---|---|---|---|---|---|---|---|---|---|---|
| 0.005393 | 0.005815 | 6.956 | 0.9349 | 35.34 | 20.53 | True | True | True | True | Sex-specific 1e-2 VUMC, sex-differential UKB |
| 0.005394 | 0.005816 | 6.956 | 0.9349 | 35.31 | 20.64 | True | True | True | True | Sex-specific 1e-2 VUMC, sex-differential UKB |
| 0.005395 | 0.005817 | 6.956 | 0.9349 | 35.2 | 20.91 | True | True | True | True | Sex-specific 1e-2 VUMC, sex-differential UKB |
| 0.005395 | 0.005817 | 6.956 | 0.935 | 35.2 | 20.91 | True | True | True | True | Sex-specific 1e-2 VUMC, sex-differential UKB |
| 0.005395 | 0.005817 | 6.955 | 0.9321 | 35.19 | 20.93 | True | True | True | True | Sex-specific 1e-2 VUMC, sex-differential UKB |
| 0.005395 | 0.005817 | 6.956 | 0.9349 | 35.19 | 20.94 | True | True | True | True | Sex-specific 1e-2 VUMC, sex-differential UKB |
| 0.005392 | 0.005814 | 6.916 | 0.9882 | 34.67 | 20.98 | True | True | True | True | Sex-specific 1e-2 VUMC, sex-differential UKB |
| 0.005397 | 0.005818 | 6.955 | 0.957 | 35.34 | 21.15 | True | True | True | True | Sex-specific 1e-2 VUMC, sex-differential UKB |
| 0.005395 | 0.005816 | 6.94 | 0.9283 | 35.14 | 20.99 | True | True | True | True | Sex-specific 1e-2 VUMC, sex-differential UKB |
| 0.005394 | 0.005815 | 6.939 | 0.9285 | 35.15 | 21.05 | True | True | True | True | Sex-specific 1e-2 VUMC, sex-differential UKB |
| 0.005394 | 0.005815 | 6.943 | 0.9283 | 35.06 | 21.08 | True | True | True | True | Sex-specific 1e-2 VUMC, sex-differential UKB |

| SE_UKB_female | SE_UKB_male | LOG10P_VUMC_female | LOG10P_VUMC_male | LOG10P_UKB_female | LOG10P_UKB_male | female_effect_direction_replication | male_effect_direction_replication | sex_effect_comparison_replication | pvalue_effect_comparison_replication | replication_type |
|---|---|---|---|---|---|---|---|---|---|---|
| 0.005394 | 0.005815 | 6.943 | 0.9284 | 35.06 | 21.07 | True | True | True | True | Sex-specific 1e-2 VUMC, sex-differential UKB |
| 0.005394 | 0.005815 | 6.943 | 0.9284 | 35.04 | 21.09 | True | True | True | True | Sex-specific 1e-2 VUMC, sex-differential UKB |
| 0.005394 | 0.005815 | 6.943 | 0.9284 | 35.06 | 21.07 | True | True | True | True | Sex-specific 1e-2 VUMC, sex-differential UKB |
| 0.005394 | 0.005815 | 6.926 | 0.908 | 35.11 | 21.09 | True | True | True | True | Sex-specific 1e-2 VUMC, sex-differential UKB |
| 0.005394 | 0.005815 | 6.926 | 0.9081 | 35.11 | 21.09 | True | True | True | True | Sex-specific 1e-2 VUMC, sex-differential UKB |
| 0.005644 | 0.006074 | 7.15 | 0.5081 | 34.37 | 22.47 | True | True | True | True | Sex-specific 1e-2 VUMC, sex-differential UKB |
| 0.005424 | 0.005845 | 6.884 | 0.9662 | 35.3 | 21.49 | True | True | True | True | Sex-specific 1e-2 VUMC, sex-differential UKB |
| 0.005422 | 0.005844 | 6.873 | 0.9361 | 35.1 | 21.31 | True | True | True | True | Sex-specific 1e-2 VUMC, sex-differential UKB |
| 0.005404 | 0.005824 | 6.896 | 0.9429 | 35.18 | 21.26 | True | True | True | True | Sex-specific 1e-2 VUMC, sex-differential UKB |
| 0.005404 | 0.005824 | 6.875 | 0.9156 | 35.18 | 21.31 | True | True | True | True | Sex-specific 1e-2 VUMC, sex-differential UKB |
| 0.005404 | 0.005824 | 6.837 | 0.9162 | 35.14 | 21.32 | True | True | True | True | Sex-specific 1e-2 VUMC, sex-differential UKB |
| 0.005398 | 0.005819 | 6.882 | 0.9151 | 35.02 | 21.19 | True | True | True | True | Sex-specific 1e-2 VUMC, sex-differential UKB |
| 0.005409 | 0.005833 | 6.885 | 0.8941 | 35.06 | 21.87 | True | True | True | True | Sex-specific 1e-2 VUMC, sex-differential UKB |
| 0.005409 | 0.005833 | 6.84 | 0.8953 | 34.99 | 21.85 | True | True | True | True | Sex-specific 1e-2 VUMC, sex-differential UKB |

| SE_UKB_female | SE_UKB_male | LOG10P_VUMC_female | LOG10P_VUMC_male | LOG10P_UKB_female | LOG10P_UKB_male | female_effect_direction_replication | male_effect_direction_replication | sex_effect_comparison_replication | pvalue_effect_comparison_replication | replication_type |
|---|---|---|---|---|---|---|---|---|---|---|
| 0.005409 | 0.005833 | 6.852 | 0.8964 | 34.91 | 21.82 | True | True | True | True | Sex-specific 1e-2 VUMC, sex-differential UKB |
| 0.00541 | 0.005834 | 6.897 | 0.8861 | 35.25 | 22.04 | True | True | True | True | Sex-specific 1e-2 VUMC, sex-differential UKB |
| 0.005407 | 0.005832 | 6.883 | 0.8941 | 35.06 | 21.83 | True | True | True | True | Sex-specific 1e-2 VUMC, sex-differential UKB |
| 0.005407 | 0.005831 | 6.883 | 0.8941 | 35.03 | 21.84 | True | True | True | True | Sex-specific 1e-2 VUMC, sex-differential UKB |
| | | 7.015 | 0.8824 | | | False | False | False | False | |
| 0.005407 | 0.005832 | 6.878 | 0.8997 | 35.04 | 21.82 | True | True | True | True | Sex-specific 1e-2 VUMC, sex-differential UKB |
| 0.005407 | 0.005832 | 6.878 | 0.8997 | 35.06 | 21.83 | True | True | True | True | Sex-specific 1e-2 VUMC, sex-differential UKB |
| 0.005407 | 0.005831 | 6.878 | 0.8997 | 35.06 | 21.84 | True | True | True | True | Sex-specific 1e-2 VUMC, sex-differential UKB |
| 0.005408 | 0.005831 | 6.941 | 0.8835 | 35.09 | 21.94 | True | True | True | True | Sex-specific 1e-2 VUMC, sex-differential UKB |
| 0.005407 | 0.005832 | 6.877 | 0.8989 | 35.01 | 21.87 | True | True | True | True | Sex-specific 1e-2 VUMC, sex-differential UKB |
| 0.005408 | 0.005833 | 6.878 | 0.8997 | 35.09 | 21.89 | True | True | True | True | Sex-specific 1e-2 VUMC, sex-differential UKB |
| 0.005414 | 0.005837 | 6.898 | 0.9347 | 35.07 | 21.92 | True | True | True | True | Sex-specific 1e-2 VUMC, sex-differential UKB |
| 0.005402 | 0.005828 | 6.839 | 0.8375 | 34.91 | 21.96 | True | True | True | True | Sex-specific 1e-2 VUMC, sex-differential UKB |
| 0.005408 | 0.005833 | 6.875 | 0.9015 | 35.1 | 21.89 | True | True | True | True | Sex-specific 1e-2 VUMC, sex-differential UKB |
| 0.005414 | 0.005839 | 6.882 | 0.9015 | 35.2 | 21.96 | True | True | True | True | Sex-specific 1e-2 VUMC, sex-differential UKB |
| 0.005419 | 0.005845 | 6.882 | 0.9033 | 35.4 | 22.02 | True | True | True | True | Sex-specific 1e-2 VUMC, sex-differential UKB |
| 0.005408 | 0.005833 | 6.882 | 0.9032 | 35.04 | 21.85 | True | True | True | True | Sex-specific 1e-2 VUMC, sex-differential UKB |
| 0.005408 | 0.005833 | 6.883 | 0.9033 | 35.04 | 21.86 | True | True | True | True | Sex-specific 1e-2 VUMC, sex-differential UKB |
| 0.005408 | 0.005833 | 6.882 | 0.9032 | 35.06 | 21.85 | True | True | True | True | Sex-specific 1e-2 VUMC, sex-differential UKB |
| 0.005408 | 0.005833 | 6.884 | 0.9058 | 35.05 | 21.85 | True | True | True | True | Sex-specific 1e-2 VUMC, sex-differential UKB |
| 0.005409 | 0.005834 | 6.873 | 0.906 | 35.02 | 21.89 | True | True | True | True | Sex-specific 1e-2 VUMC, sex-differential UKB |
| | | 6.755 | 0.973 | | | False | False | False | False | |
| 0.005408 | 0.005833 | 6.874 | 0.9058 | 35.03 | 21.9 | True | True | True | True | Sex-specific 1e-2 VUMC, sex-differential UKB |
| 0.005408 | 0.005833 | 6.874 | 0.9058 | 35.02 | 21.91 | True | True | True | True | Sex-specific 1e-2 VUMC, sex-differential UKB |
| 0.005408 | 0.005833 | 6.874 | 0.9057 | 35.01 | 21.89 | True | True | True | True | Sex-specific 1e-2 VUMC, sex-differential UKB |
| 0.005408 | 0.005833 | 6.871 | 0.9051 | 35.04 | 21.85 | True | True | True | True | Sex-specific 1e-2 VUMC, sex-differential UKB |
| 0.005404 | 0.005827 | 6.807 | 0.9476 | 35.05 | 21.42 | True | True | True | True | Sex-specific 1e-2 VUMC, sex-differential UKB |

| SE_UKB_female | SE_UKB_male | LOG10P_VUMC_female | LOG10P_VUMC_male | LOG10P_UKB_female | LOG10P_UKB_male | female_effect_direction_replication | male_effect_direction_replication | sex_effect_comparison_replication | pvalue_effect_comparison_replication | replication_type |
|---|---|---|---|---|---|---|---|---|---|---|
| 0.00541 | 0.005834 | 6.902 | 0.8964 | 35.26 | 21.98 | True | True | True | True | Sex-specific 1e-2 VUMC, sex-differential UKB |
| 0.005668 | 0.006104 | 7.073 | 0.4541 | 34.81 | 22.74 | True | True | True | True | Sex-specific 1e-2 VUMC, sex-differential UKB |
| 0.005675 | 0.00611 | 7.102 | 0.4997 | 34.99 | 22.86 | True | True | True | True | Sex-specific 1e-2 VUMC, sex-differential UKB |
| | | 6.814 | 0.9488 | | | False | False | False | False | |
| 0.005412 | 0.005834 | 6.793 | 0.9515 | 35.21 | 21.44 | True | True | True | True | Sex-specific 1e-2 VUMC, sex-differential UKB |
| 0.005412 | 0.005834 | 6.809 | 0.9514 | 35.24 | 21.47 | True | True | True | True | Sex-specific 1e-2 VUMC, sex-differential UKB |
| 0.005414 | 0.005837 | 6.893 | 0.9475 | 35.29 | 21.58 | True | True | True | True | Sex-specific 1e-2 VUMC, sex-differential UKB |
| 0.00541 | 0.005835 | 7.144 | 0.9087 | 35.23 | 21.88 | True | True | True | True | Sex-specific 1e-2 VUMC, sex-differential UKB |
| 0.00541 | 0.005835 | 6.96 | 0.9036 | 35.15 | 21.89 | True | True | True | True | Sex-specific 1e-2 VUMC, sex-differential UKB |
| 0.005409 | 0.005833 | 6.844 | 0.9491 | 35.12 | 21.51 | True | True | True | True | Sex-specific 1e-2 VUMC, sex-differential UKB |
| 0.005409 | 0.005833 | 6.843 | 0.9489 | 35.12 | 21.5 | True | True | True | True | Sex-specific 1e-2 VUMC, sex-differential UKB |
| 0.00541 | 0.005833 | 6.84 | 0.9452 | 35.08 | 21.5 | True | True | True | True | Sex-specific 1e-2 VUMC, sex-differential UKB |
| 0.00541 | 0.005833 | 6.84 | 0.9452 | 35.08 | 21.5 | True | True | True | True | Sex-specific 1e-2 VUMC, sex-differential UKB |
| 0.00541 | 0.005833 | 6.836 | 0.9438 | 35.09 | 21.46 | True | True | True | True | Sex-specific 1e-2 VUMC, sex-differential UKB |
| 0.00541 | 0.005833 | 6.84 | 0.9452 | 35.08 | 21.5 | True | True | True | True | Sex-specific 1e-2 VUMC, sex-differential UKB |
| 0.00541 | 0.005833 | 6.84 | 0.9452 | 35.08 | 21.5 | True | True | True | True | Sex-specific 1e-2 VUMC, sex-differential UKB |
| 0.005412 | 0.005837 | 6.905 | 0.9015 | 35.09 | 21.91 | True | True | True | True | Sex-specific 1e-2 VUMC, sex-differential UKB |
| 0.005409 | 0.005833 | 6.878 | 0.9436 | 35.13 | 21.43 | True | True | True | True | Sex-specific 1e-2 VUMC, sex-differential UKB |
| 0.005412 | 0.005837 | 6.904 | 0.9016 | 35.07 | 21.88 | True | True | True | True | Sex-specific 1e-2 VUMC, sex-differential UKB |
| SE_UKB_female | SE_UKB_male | LOG10P_VUMC_female | LOG10P_VUMC_male | LOG10P_UKB_female | LOG10P_UKB_male | female_effect_direction_replication | male_effect_direction_replication | sex_effect_comparison_replication | pvalue_effect_comparison_replication | replication_type |
| 0.005409 | 0.005832 | 6.884 | 0.9432 | 35.1 | 21.45 | True | True | True | True | Sex-specific 1e-2 VUMC, sex-differential UKB |
| 0.005413 | 0.005838 | 6.896 | 0.9011 | 35.07 | 21.89 | True | True | True | True | Sex-specific 1e-2 VUMC, sex-differential UKB |
| 0.005412 | 0.005837 | 6.896 | 0.9011 | 35.07 | 21.87 | True | True | True | True | Sex-specific 1e-2 VUMC, sex-differential UKB |
| 0.005408 | 0.00583 | 6.757 | 0.9496 | 34.97 | 21.32 | True | True | True | True | Sex-specific 1e-2 VUMC, sex-differential UKB |
| 0.00541 | 0.005834 | 6.819 | 0.9053 | 35 | 21.77 | True | True | True | True | Sex-specific 1e-2 VUMC, sex-differential UKB |
| 0.00541 | 0.005834 | 6.818 | 0.9051 | 35 | 21.78 | True | True | True | True | Sex-specific 1e-2 VUMC, sex-differential UKB |

| SE_UKB_female | SE_UKB_male | LOG10P_VUMC_female | LOG10P_VUMC_male | LOG10P_UKB_female | LOG10P_UKB_male | female_effect_direction_replication | male_effect_direction_replication | sex_effect_comparison_replication | pvalue_effect_comparison_replication | replication_type |
|---|---|---|---|---|---|---|---|---|---|---|
| 0.005667 | 0.006102 | 7.002 | 0.4878 | 34.63 | 22.58 | True | True | True | True | Sex-specific 1e-2 VUMC, sex-differential UKB |
| 0.005407 | 0.00583 | 6.762 | 0.9501 | 34.97 | 21.39 | True | True | True | True | Sex-specific 1e-2 VUMC, sex-differential UKB |
| 0.005408 | 0.00583 | 6.751 | 0.9494 | 34.98 | 21.37 | True | True | True | True | Sex-specific 1e-2 VUMC, sex-differential UKB |
| 0.005408 | 0.00583 | 6.751 | 0.9494 | 34.98 | 21.37 | True | True | True | True | Sex-specific 1e-2 VUMC, sex-differential UKB |
| 0.005408 | 0.00583 | 6.752 | 0.9495 | 34.97 | 21.36 | True | True | True | True | Sex-specific 1e-2 VUMC, sex-differential UKB |
| 0.005408 | 0.00583 | 6.75 | 0.9503 | 34.98 | 21.36 | True | True | True | True | Sex-specific 1e-2 VUMC, sex-differential UKB |
| 0.005412 | 0.005837 | 6.884 | 0.9012 | 35.06 | 21.91 | True | True | True | True | Sex-specific 1e-2 VUMC, sex-differential UKB |
| 0.005408 | 0.00583 | 6.751 | 0.9503 | 34.97 | 21.36 | True | True | True | True | Sex-specific 1e-2 VUMC, sex-differential UKB |
| 0.005409 | 0.005835 | 6.88 | 0.9006 | 35.18 | 21.83 | True | True | True | True | Sex-specific 1e-2 VUMC, sex-differential UKB |
| 0.005414 | 0.005838 | 6.95 | 0.927 | 35.2 | 22.04 | True | True | True | True | Sex-specific 1e-2 VUMC, sex-differential UKB |
| 0.005411 | 0.005836 | 6.933 | 0.9006 | 35.1 | 21.9 | True | True | True | True | Sex-specific 1e-2 VUMC, sex-differential UKB |
| 0.005411 | 0.005836 | 6.915 | 0.8992 | 35.08 | 21.87 | True | True | True | True | Sex-specific 1e-2 VUMC, sex-differential UKB |
| 0.005411 | 0.005835 | 6.915 | 0.8993 | 35.08 | 21.87 | True | True | True | True | Sex-specific 1e-2 VUMC, sex-differential UKB |
| 0.005411 | 0.005836 | 6.914 | 0.8992 | 35.08 | 21.87 | True | True | True | True | Sex-specific 1e-2 VUMC, sex-differential UKB |
| 0.005411 | 0.005836 | 6.915 | 0.8992 | 35.08 | 21.88 | True | True | True | True | Sex-specific 1e-2 VUMC, sex-differential UKB |
| 0.00541 | 0.005835 | 6.959 | 0.92 | 35.1 | 21.88 | True | True | True | True | Sex-specific 1e-2 VUMC, sex-differential UKB |
| 0.005413 | 0.005838 | 6.927 | 0.887 | 35.27 | 22.1 | True | True | True | True | Sex-specific 1e-2 VUMC, sex-differential UKB |
| 0.005411 | 0.005836 | 6.905 | 0.8955 | 35.12 | 21.9 | True | True | True | True | Sex-specific 1e-2 VUMC, sex-differential UKB |
| 0.005411 | 0.005836 | 6.896 | 0.9012 | 35.03 | 21.9 | True | True | True | True | Sex-specific 1e-2 VUMC, sex-differential UKB |
| 0.005411 | 0.005837 | 6.896 | 0.9012 | 35.04 | 21.91 | True | True | True | True | Sex-specific 1e-2 VUMC, sex-differential UKB |
| 0.005428 | 0.005859 | 6.977 | 0.9006 | 35.19 | 22.44 | True | True | True | True | Sex-specific 1e-2 VUMC, sex-differential UKB |
| 0.005411 | 0.005836 | 6.896 | 0.9012 | 35.04 | 21.83 | True | True | True | True | Sex-specific 1e-2 VUMC, sex-differential UKB |
| 0.005411 | 0.005836 | 6.879 | 0.8805 | 35.15 | 21.87 | True | True | True | True | Sex-specific 1e-2 VUMC, sex-differential UKB |
| 0.005411 | 0.005836 | 6.879 | 0.8805 | 35.15 | 21.85 | True | True | True | True | Sex-specific 1e-2 VUMC, sex-differential UKB |
| 0.005667 | 0.006103 | 7.073 | 0.4785 | 34.98 | 22.77 | True | True | True | True | Sex-specific 1e-2 VUMC, sex-differential UKB |
| 0.005668 | 0.006104 | 7.119 | 0.4811 | 34.74 | 22.68 | True | True | True | True | Sex-specific 1e-2 VUMC, sex-differential UKB |

| SE_UKB_female | SE_UKB_male | LOG10P_VUMC_female | LOG10P_VUMC_male | LOG10P_UKB_female | LOG10P_UKB_male | female_effect_direction_replication | male_effect_direction_replication | sex_effect_comparison_replication | pvalue_effect_comparison_replication | replication_type |
|---|---|---|---|---|---|---|---|---|---|---|
| 0.00541 | 0.005835 | 6.879 | 0.8805 | 35.15 | 21.87 | True | True | True | True | Sex-specific 1e-2 VUMC, sex-differential UKB |
| 0.00541 | 0.005835 | 6.879 | 0.8804 | 35.14 | 21.82 | True | True | True | True | Sex-specific 1e-2 VUMC, sex-differential UKB |
| 0.005408 | 0.005831 | 6.832 | 0.9237 | 35.12 | 21.44 | True | True | True | True | Sex-specific 1e-2 VUMC, sex-differential UKB |
| 0.005408 | 0.005833 | 6.879 | 0.8787 | 35.28 | 22.13 | True | True | True | True | Sex-specific 1e-2 VUMC, sex-differential UKB |
| 0.00541 | 0.005835 | 6.959 | 0.9153 | 35.15 | 21.86 | True | True | True | True | Sex-specific 1e-2 VUMC, sex-differential UKB |
| 0.005412 | 0.005837 | 6.901 | 0.8721 | 35.34 | 22.04 | True | True | True | True | Sex-specific 1e-2 VUMC, sex-differential UKB |
| 0.005414 | 0.005838 | 6.853 | 0.8898 | 35.25 | 21.78 | True | True | True | True | Sex-specific 1e-2 VUMC, sex-differential UKB |
| 0.005415 | 0.005838 | 7.04 | 0.9457 | 35.28 | 21.95 | True | True | True | True | Sex-specific 1e-2 VUMC, sex-differential UKB |
| 0.005415 | 0.005839 | 6.962 | 0.8949 | 35.38 | 22.09 | True | True | True | True | Sex-specific 1e-2 VUMC, sex-differential UKB |
| 0.005414 | 0.005839 | 6.942 | 0.9033 | 35.16 | 21.94 | True | True | True | True | Sex-specific 1e-2 VUMC, sex-differential UKB |
| 0.00541 | 0.005835 | 6.921 | 0.8988 | 35.12 | 21.86 | True | True | True | True | Sex-specific 1e-2 VUMC, sex-differential UKB |
| 0.005411 | 0.005836 | 6.917 | 0.8986 | 35.08 | 21.86 | True | True | True | True | Sex-specific 1e-2 VUMC, sex-differential UKB |
| 0.005412 | 0.005837 | 7.055 | 0.9398 | 34.99 | 21.88 | True | True | True | True | Sex-specific 1e-2 VUMC, sex-differential UKB |
| 0.00541 | 0.005834 | 7.012 | 0.984 | 34.92 | 21.65 | True | True | True | True | Sex-specific 1e-2 VUMC, sex-differential UKB |
| 0.005412 | 0.005836 | 7.071 | 0.9308 | 35.1 | 22.05 | True | True | True | True | Sex-specific 1e-2 VUMC, sex-differential UKB |
| 0.005415 | 0.005841 | 7.077 | 1.041 | 34.74 | 22.25 | True | True | True | True | Sex-specific 1e-2 VUMC, sex-differential UKB |
| 0.005416 | 0.005842 | 7.077 | 1.041 | 34.72 | 22.25 | True | True | True | True | Sex-specific 1e-2 VUMC, sex-differential UKB |
| 0.00541 | 0.005835 | 7.081 | 1.067 | 34.73 | 22.16 | True | True | True | True | Sex-specific 1e-2 VUMC, sex-differential UKB |
| 0.005413 | 0.005838 | 7.08 | 1.067 | 34.51 | 22.12 | True | True | True | True | Sex-specific 1e-2 VUMC, sex-differential UKB |
| 0.005415 | 0.00584 | 7.063 | 1.051 | 34.52 | 22.12 | True | True | True | True | Sex-specific 1e-2 VUMC, sex-differential UKB |
| 0.005416 | 0.005841 | 7.065 | 1.051 | 34.52 | 22.13 | True | True | True | True | Sex-specific 1e-2 VUMC, sex-differential UKB |
| 0.005415 | 0.005839 | 6.996 | 1.051 | 34.63 | 21.89 | True | True | True | True | Sex-specific 1e-2 VUMC, sex-differential UKB |
| 0.005415 | 0.00584 | 7.005 | 1.121 | 34.63 | 21.89 | True | True | True | True | Sex-specific 1e-2 VUMC, sex-differential UKB |
| 0.005424 | 0.005848 | 7.051 | 1.107 | 34.73 | 22.32 | True | True | True | True | Sex-specific 1e-2 VUMC, sex-differential UKB |
| | | 7.583 | 0.8531 | | | False | False | False | False | |
| 0.005228 | 0.005625 | 6.989 | 1.505 | 33.14 | 21.85 | True | True | True | True | Sex-specific 1e-2 VUMC, sex-differential UKB |

| SE_UKB_female | SE_UKB_male | LOG10P_VUMC_female | LOG10P_VUMC_male | LOG10P_UKB_female | LOG10P_UKB_male | female_effect_direction_replication | male_effect_direction_replication | sex_effect_comparison_replication | pvalue_effect_comparison_replication | replication_type |
|---|---|---|---|---|---|---|---|---|---|---|
| 0.005621 | 0.006053 | 7.51 | 0.7339 | 34.16 | 22.17 | True | True | True | True | Sex-specific 1e-2 VUMC, sex-differential UKB |
| 0.005428 | 0.005853 | 7.099 | 1.099 | 34.31 | 22.27 | True | True | True | True | Sex-specific 1e-2 VUMC, sex-differential UKB |
| SE_UKB_female | SE_UKB_male | LOG10P_VUMC_female | LOG10P_VUMC_male | LOG10P_UKB_female | LOG10P_UKB_male | female_effect_direction_replication | male_effect_direction_replication | sex_effect_comparison_replication | pvalue_effect_comparison_replication | replication_type |
| 0.005424 | 0.005849 | 7.005 | 1.122 | 34.1 | 21.83 | True | True | True | True | Sex-specific 1e-2 VUMC, sex-differential UKB |
| 0.005425 | 0.005849 | 6.585 | 1.04 | 34.13 | 21.76 | True | True | True | True | Sex-specific 1e-2 VUMC, sex-differential UKB |
| 0.005233 | 0.005629 | 6.689 | 1.149 | 32.09 | 20.65 | True | True | True | True | Sex-specific 1e-2 VUMC, sex-differential UKB |
| 0.005237 | 0.005634 | 6.569 | 1.206 | 32.36 | 20.79 | True | True | True | True | Sex-specific 1e-2 VUMC, sex-differential UKB |
| 0.005236 | 0.005633 | 6.541 | 1.212 | 32.32 | 20.81 | True | True | True | True | Sex-specific 1e-2 VUMC, sex-differential UKB |
| 0.005269 | 0.005668 | 6.983 | 1.227 | 32.88 | 20.54 | True | True | True | True | Sex-specific 1e-2 VUMC, sex-differential UKB |
| 0.005237 | 0.005634 | 6.578 | 1.206 | 32.35 | 20.79 | True | True | True | True | Sex-specific 1e-2 VUMC, sex-differential UKB |
| 0.005238 | 0.005635 | 6.573 | 1.211 | 32.28 | 20.74 | True | True | True | True | Sex-specific 1e-2 VUMC, sex-differential UKB |
| 0.00526 | 0.005656 | 6.394 | 1.167 | 32.51 | 20.76 | True | True | True | True | Sex-specific 1e-2 VUMC, sex-differential UKB |
| 0.005252 | 0.005652 | 6.6 | 1.142 | 32.1 | 20.82 | True | True | True | True | Sex-specific 1e-2 VUMC, sex-differential UKB |
| 0.005254 | 0.005654 | 6.581 | 1.119 | 32.16 | 20.97 | True | True | True | True | Sex-specific 1e-2 VUMC, sex-differential UKB |
| 0.005256 | 0.005658 | 6.668 | 1.051 | 32.53 | 21.05 | True | True | True | True | Sex-specific 1e-2 VUMC, sex-differential UKB |
| 0.005257 | 0.005658 | 6.668 | 1.051 | 32.49 | 21.11 | True | True | True | True | Sex-specific 1e-2 VUMC, sex-differential UKB |
| 0.005258 | 0.00566 | 6.643 | 1.068 | 32.43 | 21.09 | True | True | True | True | Sex-specific 1e-2 VUMC, sex-differential UKB |
| 0.005258 | 0.00566 | 6.643 | 1.068 | 32.41 | 21.08 | True | True | True | True | Sex-specific 1e-2 VUMC, sex-differential UKB |
| 0.00542 | 0.005843 | 7.074 | 0.9278 | 32.49 | 20.73 | True | True | True | True | Sex-specific 1e-2 VUMC, sex-differential UKB |
| 0.005258 | 0.005661 | 6.636 | 1.068 | 32.39 | 21.08 | True | True | True | True | Sex-specific 1e-2 VUMC, sex-differential UKB |
| 0.005261 | 0.005664 | 6.616 | 1.008 | 32.39 | 21.18 | True | True | True | True | Sex-specific 1e-2 VUMC, sex-differential UKB |
| 0.005261 | 0.005664 | 6.605 | 1.012 | 32.39 | 21.12 | True | True | True | True | Sex-specific 1e-2 VUMC, sex-differential UKB |
| 0.005261 | 0.005664 | 6.605 | 1.012 | 32.39 | 21.12 | True | True | True | True | Sex-specific 1e-2 VUMC, sex-differential UKB |
| 0.005261 | 0.005664 | 6.605 | 1.012 | 32.38 | 21.18 | True | True | True | True | Sex-specific 1e-2 VUMC, sex-differential UKB |
| 0.005261 | 0.005664 | 6.605 | 1.012 | 32.38 | 21.11 | True | True | True | True | Sex-specific 1e-2 VUMC, sex-differential UKB |
| 0.005258 | 0.005661 | 6.608 | 0.9871 | 32.06 | 21.04 | True | True | True | True | Sex-specific 1e-2 VUMC, sex-differential UKB |

| SE_UKB_female | SE_UKB_male | LOG10P_VUMC_female | LOG10P_VUMC_male | LOG10P_UKB_female | LOG10P_UKB_male | female_effect_direction_replication | male_effect_direction_replication | sex_effect_comparison_replication | pvalue_effect_comparison_replication | replication_type |
|---|---|---|---|---|---|---|---|---|---|---|
| 0.005278 | 0.005681 | 6.793 | 1.112 | 32.46 | 20.55 | True | True | True | True | Sex-specific 1e-2 VUMC, sex-differential UKB |
| 0.005279 | 0.005681 | 6.781 | 1.094 | 32.45 | 20.56 | True | True | True | True | Sex-specific 1e-2 VUMC, sex-differential UKB |
| 0.005731 | 0.006171 | 7.241 | 0.4373 | 32.56 | 21.16 | True | True | True | True | Sex-specific 1e-2 VUMC, sex-differential UKB |
| 0.005467 | 0.005894 | 7.179 | 0.849 | 32.89 | 20.51 | True | True | True | True | Sex-specific 1e-2 VUMC, sex-differential UKB |
| 0.005436 | 0.005858 | 6.888 | 0.8704 | 32.18 | 20.16 | True | True | True | True | Sex-specific 1e-2 VUMC, sex-differential UKB |
| 0.005279 | 0.00568 | 6.911 | 1.153 | 32.65 | 20.59 | True | True | True | True | Sex-specific 1e-2 VUMC, sex-differential UKB |
| 0.005279 | 0.00568 | 6.936 | 1.159 | 32.62 | 20.61 | True | True | True | True | Sex-specific 1e-2 VUMC, sex-differential UKB |
| 0.00547 | 0.005899 | 7.1 | 0.8989 | 32.87 | 20.57 | True | True | True | True | Sex-specific 1e-2 VUMC, sex-differential UKB |
| 0.005285 | 0.005686 | 7.039 | 1.15 | 32.95 | 20.2 | True | True | True | True | Sex-specific 1e-2 VUMC, sex-differential UKB |
| 0.005446 | 0.005869 | 7.354 | 0.921 | 32.33 | 19.77 | True | True | True | True | Sex-specific 1e-2 VUMC, sex-differential UKB |
| 0.005438 | 0.005858 | 7.261 | 0.8056 | 30.44 | 18.72 | True | True | True | True | Sex-specific 1e-2 VUMC, sex-differential UKB |
| 0.005475 | 0.005902 | 6.957 | 0.7589 | 31.17 | 19.65 | True | True | True | True | Sex-specific 1e-2 VUMC, sex-differential UKB |
| 0.005745 | 0.006183 | 6.979 | 0.3523 | 30.82 | 20.09 | True | True | True | True | Sex-specific 1e-2 VUMC, sex-differential UKB |
| 0.005514 | 0.005939 | 6.995 | 0.7067 | 32 | 20.18 | True | True | True | True | Sex-specific 1e-2 VUMC, sex-differential UKB |
| 0.006624 | 0.007154 | 6.5 | 0.8655 | 37.31 | 26.12 | True | True | True | True | Sex-specific 1e-2 VUMC, sex-differential UKB |
| 0.003419 | 0.003678 | 10.87 | 0.603 | 75.85 | 64.12 | True | True | True | True | Sex-specific 1e-2 VUMC, sex-differential UKB |
| 0.003298 | 0.003543 | 4.964 | 0.3076 | 46.41 | 36.38 | True | True | True | True | Sex-specific 1e-2 VUMC, sex-differential UKB |
| 0.003575 | 0.003847 | 5.171 | 0.07846 | 0.00206 | 1.22 | True | True | False | False | |
| 0.003355 | 0.003617 | 8.275 | 0.7217 | 0.001035 | 1.048 | True | True | False | False | |
| 0.003629 | 0.003909 | 5.893 | 0.07079 | 0.09219 | 1.231 | True | True | False | False | |
| 0.003627 | 0.003908 | 5.554 | 0.08034 | 0.1106 | 1.221 | True | True | False | False | |
| 0.003468 | 0.003733 | 4.899 | 0.8113 | 0.07435 | 0.5338 | False | True | False | False | |
| 0.003468 | 0.003733 | 4.918 | 0.8044 | 0.07051 | 0.5362 | False | True | False | False | |
| 0.003468 | 0.003733 | 4.958 | 0.7773 | 0.06327 | 0.5044 | False | True | False | False | |
| 0.003469 | 0.003734 | 4.962 | 0.7723 | 0.06425 | 0.5072 | False | True | False | False | |
| 0.003312 | 0.003565 | 7.216 | 0.5431 | 5.166 | 2.767 | True | True | True | True | Sex-specific 1e-2 VUMC, sex-differential UKB |

| SE_UKB_female | SE_UKB_male | LOG10P_VUMC_female | LOG10P_VUMC_male | LOG10P_UKB_female | LOG10P_UKB_male | female_effect_direction_replication | male_effect_direction_replication | sex_effect_comparison_replication | pvalue_effect_comparison_replication | replication_type |
|---|---|---|---|---|---|---|---|---|---|---|
| 0.003387 | 0.003647 | 9.419 | 1.07 | 6.706 | 3.803 | True | True | True | True | Sex-specific 1e-2 VUMC, sex-differential UKB |
| 0.003312 | 0.003565 | 7.383 | 0.5373 | 5.218 | 2.78 | True | True | True | True | Sex-specific 1e-2 VUMC, sex-differential UKB |
| 0.003387 | 0.003648 | 9.427 | 1.15 | 6.728 | 3.789 | True | True | True | True | Sex-specific 1e-2 VUMC, sex-differential UKB |
| 0.003306 | 0.003561 | 6.336 | 1.686 | 4.127 | 3.082 | True | True | True | True | Sex-specific 1e-2 VUMC, sex-differential UKB |
| 0.0033 | 0.003554 | 6.349 | 1.662 | 3.869 | 3.075 | True | True | True | True | Sex-specific 1e-2 VUMC, sex-differential UKB |
| 0.003305 | 0.00356 | 6.366 | 1.652 | 4.11 | 3.014 | True | True | True | True | Sex-specific 1e-2 VUMC, sex-differential UKB |
| 0.003301 | 0.003557 | 6.097 | 1.664 | 4.167 | 2.906 | True | True | True | True | Sex-specific 1e-2 VUMC, sex-differential UKB |
| 0.0033 | 0.003557 | 5.833 | 1.593 | 4.429 | 3.047 | True | True | True | True | Sex-specific 1e-2 VUMC, sex-differential UKB |
| 0.003305 | 0.003561 | 5.712 | 1.589 | 4.369 | 2.941 | True | True | True | True | Sex-specific 1e-2 VUMC, sex-differential UKB |
| 0.003306 | 0.003563 | 5.769 | 1.546 | 4.363 | 2.906 | True | True | True | True | Sex-specific 1e-2 VUMC, sex-differential UKB |
| 0.003308 | 0.003565 | 5.774 | 1.546 | 4.355 | 2.885 | True | True | True | True | Sex-specific 1e-2 VUMC, sex-differential UKB |
| SE_UKB_female | SE_UKB_male | LOG10P_VUMC_female | LOG10P_VUMC_male | LOG10P_UKB_female | LOG10P_UKB_male | female_effect_direction_replication | male_effect_direction_replication | sex_effect_comparison_replication | pvalue_effect_comparison_replication | replication_type |
| 0.003302 | 0.003563 | 5.004 | 1.733 | 4.101 | 2.48 | True | True | True | True | Sex-specific 1e-2 VUMC, sex-differential UKB |
| 0.003837 | 0.004147 | 7.521 | 1.885 | 5.422 | 1.695 | True | True | True | True | Sex-specific 1e-2 VUMC, sex-differential UKB |
| 0.003554 | 0.00384 | 6.534 | 1.214 | 4.232 | 1.115 | True | True | True | True | Sex-specific 1e-2 VUMC, sex-differential UKB |
| 0.003595 | 0.003885 | 5.581 | 1.183 | 4.168 | 1.058 | True | True | True | True | Sex-specific 1e-2 VUMC, sex-differential UKB |
| 0.004632 | 0.00499 | 9.998 | 1.369 | 4.502 | 1.422 | True | True | True | True | Sex-specific 1e-2 VUMC, sex-differential UKB |
| 0.004663 | 0.005026 | 10.04 | 1.433 | 5.119 | 1.547 | True | True | True | True | Sex-specific 1e-2 VUMC, sex-differential UKB |
| 0.004659 | 0.005023 | 9.805 | 1.492 | 5.183 | 1.518 | True | True | True | True | Sex-specific 1e-2 VUMC, sex-differential UKB |
| 0.004574 | 0.004934 | 10.96 | 0.9143 | 4.335 | 1.358 | True | True | True | True | Sex-specific 1e-2 VUMC, sex-differential UKB |
| 0.004575 | 0.004933 | 10.72 | 0.9277 | 4.417 | 1.382 | True | True | True | True | Sex-specific 1e-2 VUMC, sex-differential UKB |
| 0.004572 | 0.004931 | 10.86 | 0.872 | 4.493 | 1.363 | True | True | True | True | Sex-specific 1e-2 VUMC, sex-differential UKB |
| 0.004543 | 0.004899 | 10.84 | 0.8871 | 5.39 | 0.9527 | True | True | True | True | Sex-specific 1e-2 VUMC, sex-differential UKB |
| 0.004538 | 0.004894 | 10.91 | 0.8101 | 5.368 | 0.9195 | True | True | True | True | Sex-specific 1e-2 VUMC, sex-differential UKB |
| 0.003921 | 0.004219 | 10.42 | 1.165 | 7.072 | 1.34 | True | True | True | True | Sex-specific 1e-2 VUMC, sex-differential UKB |
| 0.003919 | 0.004216 | 10.19 | 1.135 | 7.148 | 1.342 | True | True | True | True | Sex-specific 1e-2 VUMC, sex-differential UKB |

| SE_UKB_female | SE_UKB_male | LOG10P_VUMC_female | LOG10P_VUMC_male | LOG10P_UKB_female | LOG10P_UKB_male | female_effect_direction_replication | male_effect_direction_replication | sex_effect_comparison_replication | pvalue_effect_comparison_replication | replication_type |
|---|---|---|---|---|---|---|---|---|---|---|
| 0.003919 | 0.004216 | 10.27 | 1.169 | 7.151 | 1.337 | True | True | True | True | Sex-specific 1e-2 VUMC, sex-differential UKB |
| 0.003918 | 0.004216 | 10.2 | 1.067 | 7.189 | 1.347 | True | True | True | True | Sex-specific 1e-2 VUMC, sex-differential UKB |
| 0.003917 | 0.004215 | 10.2 | 1.083 | 7.1 | 1.284 | True | True | True | True | Sex-specific 1e-2 VUMC, sex-differential UKB |
| 0.003916 | 0.004213 | 10.38 | 1.086 | 7.324 | 1.331 | True | True | True | True | Sex-specific 1e-2 VUMC & UKB |
| 0.003924 | 0.004221 | 10.18 | 1.15 | 7.356 | 1.375 | True | True | True | True | Sex-specific 1e-2 VUMC & UKB |
| 0.003935 | 0.004233 | 10.53 | 1.054 | 7.341 | 1.473 | True | True | True | True | Sex-specific 1e-2 VUMC & UKB |
| 0.003922 | 0.004219 | 10.74 | 0.9871 | 7.103 | 1.54 | True | True | True | True | Sex-specific 1e-2 VUMC, sex-differential UKB |
| 0.00392 | 0.004216 | 10.71 | 1.04 | 7.17 | 1.522 | True | True | True | True | Sex-specific 1e-2 VUMC, sex-differential UKB |
| 0.003924 | 0.004221 | 10.56 | 1.063 | 7.386 | 1.531 | True | True | True | True | Sex-specific 1e-2 VUMC & UKB |
| 0.003926 | 0.004224 | 10.53 | 1.004 | 7.209 | 1.385 | True | True | True | True | Sex-specific 1e-2 VUMC, sex-differential UKB |
| 0.003921 | 0.004221 | 10.57 | 0.9129 | 6.973 | 1.339 | True | True | True | True | Sex-specific 1e-2 VUMC, sex-differential UKB |
| 0.003922 | 0.004221 | 10.49 | 1.023 | 7.04 | 1.362 | True | True | True | True | Sex-specific 1e-2 VUMC, sex-differential UKB |
| 0.003921 | 0.004221 | 10.44 | 1.01 | 6.968 | 1.354 | True | True | True | True | Sex-specific 1e-2 VUMC, sex-differential UKB |
| 0.003921 | 0.004221 | 10.44 | 1.01 | 6.934 | 1.356 | True | True | True | True | Sex-specific 1e-2 VUMC, sex-differential UKB |
| 0.003887 | 0.004182 | 9.579 | 0.8096 | 6.538 | 1.827 | True | True | True | True | Sex-specific 1e-2 VUMC, sex-differential UKB |
| 0.003886 | 0.004181 | 9.479 | 0.84 | 6.588 | 1.84 | True | True | True | True | Sex-specific 1e-2 VUMC, sex-differential UKB |
| 0.003885 | 0.00418 | 9.438 | 0.8385 | 6.669 | 1.818 | True | True | True | True | Sex-specific 1e-2 VUMC, sex-differential UKB |
| 0.003884 | 0.004178 | 9.264 | 0.956 | 6.656 | 1.845 | True | True | True | True | Sex-specific 1e-2 VUMC, sex-differential UKB |
| 0.003883 | 0.004178 | 9.264 | 0.9562 | 6.651 | 1.839 | True | True | True | True | Sex-specific 1e-2 VUMC, sex-differential UKB |
| 0.003883 | 0.004178 | 9.265 | 0.9562 | 6.643 | 1.845 | True | True | True | True | Sex-specific 1e-2 VUMC, sex-differential UKB |
| 0.003882 | 0.004176 | 9.28 | 0.9781 | 6.748 | 1.908 | True | True | True | True | Sex-specific 1e-2 VUMC, sex-differential UKB |
| 0.003882 | 0.004176 | 9.218 | 0.9463 | 6.733 | 1.913 | True | True | True | True | Sex-specific 1e-2 VUMC, sex-differential UKB |
| 0.003881 | 0.004176 | 9.219 | 0.9463 | 6.752 | 1.908 | True | True | True | True | Sex-specific 1e-2 VUMC, sex-differential UKB |
| 0.003884 | 0.00418 | 9.135 | 0.8711 | 6.849 | 1.846 | True | True | True | True | Sex-specific 1e-2 VUMC, sex-differential UKB |
| 0.003886 | 0.004181 | 8.943 | 0.8019 | 6.905 | 1.817 | True | True | True | True | Sex-specific 1e-2 VUMC, sex-differential UKB |
| 0.003886 | 0.004182 | 9.14 | 0.8728 | 6.915 | 1.821 | True | True | True | True | Sex-specific 1e-2 VUMC, sex-differential UKB |

| SE_UKB_female | SE_UKB_male | LOG10P_VUMC_female | LOG10P_VUMC_male | LOG10P_UKB_female | LOG10P_UKB_male | female_effect_direction_replication | male_effect_direction_replication | sex_effect_comparison_replication | pvalue_effect_comparison_replication | replication_type |
|---|---|---|---|---|---|---|---|---|---|---|
| 0.003887 | 0.004182 | 9.067 | 0.891 | 6.88 | 1.823 | True | True | True | True | Sex-specific 1e-2 VUMC, sex-differential UKB |
| 0.003886 | 0.004181 | 9.079 | 0.8885 | 6.884 | 1.824 | True | True | True | True | Sex-specific 1e-2 VUMC, sex-differential UKB |
| 0.003886 | 0.004181 | 9.079 | 0.8885 | 6.881 | 1.824 | True | True | True | True | Sex-specific 1e-2 VUMC, sex-differential UKB |
| 0.003886 | 0.004182 | 9.079 | 0.8885 | 6.87 | 1.826 | True | True | True | True | Sex-specific 1e-2 VUMC, sex-differential UKB |
| 0.003886 | 0.004182 | 9.079 | 0.8886 | 6.87 | 1.826 | True | True | True | True | Sex-specific 1e-2 VUMC, sex-differential UKB |
| 0.003886 | 0.004182 | 9.079 | 0.8886 | 6.87 | 1.826 | True | True | True | True | Sex-specific 1e-2 VUMC, sex-differential UKB |
| 0.003886 | 0.004182 | 9.078 | 0.8891 | 6.865 | 1.831 | True | True | True | True | Sex-specific 1e-2 VUMC, sex-differential UKB |
| 0.003886 | 0.004182 | 9.078 | 0.8888 | 6.869 | 1.829 | True | True | True | True | Sex-specific 1e-2 VUMC, sex-differential UKB |
| 0.003887 | 0.004182 | 9.086 | 0.8928 | 6.888 | 1.828 | True | True | True | True | Sex-specific 1e-2 VUMC, sex-differential UKB |
| 0.003886 | 0.004182 | 8.851 | 0.8855 | 6.867 | 1.827 | True | True | True | True | Sex-specific 1e-2 VUMC, sex-differential UKB |
| 0.003886 | 0.004182 | 8.851 | 0.8855 | 6.866 | 1.827 | True | True | True | True | Sex-specific 1e-2 VUMC, sex-differential UKB |
| 0.003886 | 0.004182 | 8.851 | 0.8856 | 6.869 | 1.822 | True | True | True | True | Sex-specific 1e-2 VUMC, sex-differential UKB |
| 0.003886 | 0.004181 | 8.868 | 0.8955 | 6.834 | 1.822 | True | True | True | True | Sex-specific 1e-2 VUMC, sex-differential UKB |
| 0.003886 | 0.004181 | 8.87 | 0.8766 | 6.85 | 1.819 | True | True | True | True | Sex-specific 1e-2 VUMC, sex-differential UKB |
| 0.003889 | 0.004183 | 8.865 | 0.8587 | 6.772 | 1.844 | True | True | True | True | Sex-specific 1e-2 VUMC, sex-differential UKB |
| 0.003889 | 0.004183 | 8.904 | 0.8664 | 6.751 | 1.836 | True | True | True | True | Sex-specific 1e-2 VUMC, sex-differential UKB |
| 0.003889 | 0.004183 | 8.882 | 0.8532 | 6.753 | 1.839 | True | True | True | True | Sex-specific 1e-2 VUMC, sex-differential UKB |
| 0.003889 | 0.004183 | 8.885 | 0.8525 | 6.757 | 1.829 | True | True | True | True | Sex-specific 1e-2 VUMC, sex-differential UKB |
| 0.003889 | 0.004183 | 8.759 | 0.8127 | 6.718 | 1.824 | True | True | True | True | Sex-specific 1e-2 VUMC, sex-differential UKB |
| 0.003889 | 0.004183 | 8.904 | 0.8664 | 6.752 | 1.83 | True | True | True | True | Sex-specific 1e-2 VUMC, sex-differential UKB |
| SE_UKB_female | SE_UKB_male | LOG10P_VUMC_female | LOG10P_VUMC_male | LOG10P_UKB_female | LOG10P_UKB_male | female_effect_direction_replication | male_effect_direction_replication | sex_effect_comparison_replication | pvalue_effect_comparison_replication | replication_type |
| 0.003889 | 0.004183 | 8.901 | 0.867 | 6.749 | 1.841 | True | True | True | True | Sex-specific 1e-2 VUMC, sex-differential UKB |
| 0.00388 | 0.004174 | 8.839 | 0.7101 | 6.295 | 1.78 | True | True | True | True | Sex-specific 1e-2 VUMC, sex-differential UKB |
| 0.003889 | 0.004183 | 8.882 | 0.8529 | 6.752 | 1.84 | True | True | True | True | Sex-specific 1e-2 VUMC, sex-differential UKB |
| 0.003889 | 0.004183 | 8.882 | 0.853 | 6.758 | 1.84 | True | True | True | True | Sex-specific 1e-2 VUMC, sex-differential UKB |
| 0.00389 | 0.004184 | 8.953 | 0.9095 | 6.778 | 1.865 | True | True | True | True | Sex-specific 1e-2 VUMC, sex-differential UKB |

| SE_UKB_female | SE_UKB_male | LOG10P_VUMC_female | LOG10P_VUMC_male | LOG10P_UKB_female | LOG10P_UKB_male | female_effect_direction_replication | male_effect_direction_replication | sex_effect_comparison_replication | pvalue_effect_comparison_replication | replication_type |
|---|---|---|---|---|---|---|---|---|---|---|
| 0.00389 | 0.004184 | 9.024 | 0.9666 | 6.752 | 1.865 | True | True | True | True | Sex-specific 1e-2 VUMC, sex-differential UKB |
| 0.00389 | 0.004184 | 8.985 | 0.924 | 6.778 | 1.866 | True | True | True | True | Sex-specific 1e-2 VUMC, sex-differential UKB |
| 0.00389 | 0.004184 | 8.971 | 0.9102 | 6.78 | 1.868 | True | True | True | True | Sex-specific 1e-2 VUMC, sex-differential UKB |
| 0.00389 | 0.004184 | 9.001 | 0.8885 | 6.789 | 1.888 | True | True | True | True | Sex-specific 1e-2 VUMC, sex-differential UKB |
| 0.00389 | 0.004184 | 8.985 | 0.924 | 6.782 | 1.867 | True | True | True | True | Sex-specific 1e-2 VUMC, sex-differential UKB |
| 0.01887 | 0.01949 | 5.611 | 0.09418 | 1.384 | 1.747 | True | True | False | False | |
| 0.01881 | 0.01952 | 5.616 | 0.01022 | 1.48 | 2.083 | True | True | False | False | |
| 0.0188 | 0.01948 | 6.632 | 0.1117 | 1.764 | 2.515 | True | False | False | False | |
| 0.01886 | 0.01953 | 6.093 | 0.03936 | 1.436 | 2.105 | True | True | False | False | |
| 0.01886 | 0.01953 | 6.136 | 0.01486 | 1.455 | 2.072 | True | True | False | False | |
| 0.01885 | 0.0195 | 5.82 | 0.007933 | 1.446 | 1.918 | True | True | False | False | |
| 0.01884 | 0.01951 | 6.192 | 0.002355 | 1.405 | 2.031 | True | True | False | False | |
| 0.01879 | 0.01947 | 6.828 | 0.1715 | 1.776 | 2.648 | True | False | False | False | |
| 0.01885 | 0.01951 | 6.179 | 0.001365 | 1.401 | 2.031 | True | True | False | False | |
| 0.01878 | 0.01942 | 6.297 | 0.1671 | 1.777 | 2.577 | True | False | False | False | |
| 0.01879 | 0.01947 | 6.855 | 0.162 | 1.787 | 2.744 | True | False | False | False | |
| 0.0188 | 0.01944 | 6.431 | 0.1484 | 1.784 | 2.606 | True | False | False | False | |
| 0.01881 | 0.01949 | 6.833 | 0.1601 | 1.792 | 2.769 | True | False | False | False | |
| 0.0188 | 0.01944 | 6.505 | 0.1523 | 1.82 | 2.472 | True | False | False | False | |
| 0.01881 | 0.01945 | 6.524 | 0.1414 | 1.851 | 2.432 | True | False | False | False | |
| 0.01908 | 0.01957 | 6.413 | 0.1314 | 2.197 | 3.594 | True | False | False | False | |
| 0.0191 | 0.01963 | 6.86 | 0.136 | 2.192 | 3.762 | True | False | False | False | |
| 0.01908 | 0.01942 | 5.844 | 0.008994 | 1.975 | 3.137 | True | False | False | False | |
| 0.01916 | 0.01935 | 6.41 | 0.03858 | 2.128 | 3.143 | True | False | False | False | |
| 0.01919 | 0.01946 | 5.992 | 0.1229 | 1.763 | 3.425 | True | False | False | False | |
| 0.01916 | 0.01946 | 5.534 | 0.08359 | 1.874 | 3.22 | True | False | False | False | |
| 0.01916 | 0.01945 | 5.674 | 0.0612 | 1.766 | 3.437 | True | False | False | False | |
| 0.01916 | 0.01945 | 5.674 | 0.06121 | 1.766 | 3.437 | True | False | False | False | |
| 0.01909 | 0.01934 | 7.234 | 0.01808 | 2.054 | 4.103 | True | True | False | False | |
| 0.01911 | 0.01925 | 7.005 | 0.1758 | 2.27 | 4.176 | True | True | False | False | |
| 0.0194 | 0.01952 | 4.187 | 0.03711 | 1.234 | 2.744 | True | False | False | False | |
| 0.01913 | 0.01931 | 6.63 | 0.4915 | 2.405 | 4.328 | True | True | False | False | |
| 0.01925 | 0.01943 | 4.321 | 0.01493 | 1.422 | 2.886 | True | True | False | False | |
| 0.01923 | 0.01945 | 4.657 | 0.04408 | 1.653 | 2.616 | True | True | False | False | |
| 0.01937 | 0.01971 | 3.958 | 0.1633 | 1.212 | 2.503 | True | True | False | False | |
| 0.01923 | 0.01945 | 4.66 | 0.04422 | 1.678 | 2.612 | True | True | False | False | |
| 0.01923 | 0.01945 | 4.659 | 0.04422 | 1.649 | 2.612 | True | True | False | False | |
| 0.01923 | 0.01945 | 4.662 | 0.04431 | 1.649 | 2.609 | True | True | False | False | |
| 0.01937 | 0.01971 | 3.988 | 0.1672 | 1.211 | 2.502 | True | True | False | False | |
| 0.01938 | 0.01971 | 3.959 | 0.1632 | 1.225 | 2.506 | True | True | False | False | |
| 0.01937 | 0.01971 | 3.978 | 0.1633 | 1.218 | 2.502 | True | True | False | False | |
| 0.01938 | 0.01972 | 4.017 | 0.1683 | 1.217 | 2.5 | True | True | False | False | |
| 0.01926 | 0.01948 | 4.682 | 0.04917 | 1.662 | 2.601 | True | True | False | False | |

| SE_UKB_female | SE_UKB_male | LOG10P_VUMC_female | LOG10P_VUMC_male | LOG10P_UKB_female | LOG10P_UKB_male | female_effect_direction_replication | male_effect_direction_replication | sex_effect_comparison_replication | pvalue_effect_comparison_replication | replication_type |
|---|---|---|---|---|---|---|---|---|---|---|
| 0.01931 | 0.01963 | 4.002 | 0.3547 | 1.55 | 2.579 | True | True | False | False | |
| 0.01929 | 0.01962 | 4.012 | 0.317 | 1.562 | 2.459 | True | True | False | False | |
| 0.01932 | 0.01976 | 6.605 | 0.9687 | 2.803 | 3.606 | True | True | False | False | |
| 0.01933 | 0.01977 | 6.541 | 0.9809 | 2.807 | 3.521 | True | True | False | False | |
| 0.01929 | 0.0196 | 4.132 | 0.3145 | 1.596 | 2.306 | True | True | False | False | |
| 0.01933 | 0.01977 | 3.622 | 0.4398 | 1.227 | 2.185 | True | True | False | False | |
| 0.01923 | 0.01946 | 4.846 | 0.05663 | 1.725 | 2.694 | True | True | False | False | |
| 0.01917 | 0.01945 | 7.475 | 0.5475 | 2.787 | 4.075 | True | True | False | False | |
| 0.01922 | 0.01947 | 4.753 | 0.06974 | 1.749 | 2.681 | True | True | False | False | |
| 0.01923 | 0.01945 | 4.815 | 0.113 | 1.666 | 2.653 | True | True | False | False | |
| 0.01923 | 0.01945 | 4.815 | 0.113 | 1.666 | 2.653 | True | True | False | False | |
| 0.01917 | 0.01939 | 7.583 | 0.5941 | 2.746 | 4.091 | True | True | False | False | |
| SE_UKB_female | SE_UKB_male | LOG10P_VUMC_female | LOG10P_VUMC_male | LOG10P_UKB_female | LOG10P_UKB_male | female_effect_direction_replication | male_effect_direction_replication | sex_effect_comparison_replication | pvalue_effect_comparison_replication | replication_type |
| 0.01923 | 0.01945 | 4.825 | 0.1132 | 1.671 | 2.65 | True | True | False | False | |
| 0.01923 | 0.01945 | 4.824 | 0.1274 | 1.666 | 2.648 | True | True | False | False | |
| 0.01923 | 0.01944 | 4.963 | 0.1105 | 1.65 | 2.578 | True | True | False | False | |
| 0.01923 | 0.01944 | 4.929 | 0.1083 | 1.645 | 2.716 | True | True | False | False | |
| 0.01916 | 0.01937 | 7.458 | 0.5777 | 2.595 | 4.066 | True | True | False | False | |
| 0.01923 | 0.01944 | 4.892 | 0.1094 | 1.645 | 2.686 | True | True | False | False | |
| 0.01923 | 0.01944 | 4.899 | 0.1092 | 1.648 | 2.687 | True | True | False | False | |
| 0.01923 | 0.01944 | 4.903 | 0.1071 | 1.645 | 2.689 | True | True | False | False | |
| 0.01916 | 0.01936 | 7.396 | 0.5813 | 2.591 | 4.056 | True | True | False | False | |
| 0.01923 | 0.01944 | 4.901 | 0.1093 | 1.639 | 2.752 | True | True | False | False | |
| 0.01923 | 0.01944 | 4.925 | 0.104 | 1.644 | 2.689 | True | True | False | False | |
| 0.01917 | 0.01938 | 7.434 | 0.5645 | 2.563 | 3.889 | True | True | False | False | |
| 0.01923 | 0.01945 | 4.925 | 0.104 | 1.647 | 2.55 | True | True | False | False | |
| 0.01924 | 0.01946 | 4.982 | 0.07665 | 1.667 | 2.444 | True | True | False | False | |
| 0.01923 | 0.01945 | 4.902 | 0.1073 | 1.649 | 2.549 | True | True | False | False | |
| | | 4.958 | 0.07978 | | | False | False | False | False | |
| 0.01923 | 0.01945 | 4.954 | 0.08003 | 1.67 | 2.626 | True | True | False | False | |
| 0.01924 | 0.01945 | 4.954 | 0.08004 | 1.667 | 2.489 | True | True | False | False | |
| 0.01915 | 0.01936 | 7.446 | 0.5378 | 2.599 | 3.806 | True | True | False | False | |
| 0.01916 | 0.01937 | 7.367 | 0.5832 | 2.581 | 3.944 | True | True | False | False | |
| 0.01915 | 0.01936 | 7.441 | 0.5381 | 2.6 | 3.808 | True | True | False | False | |
| 0.01916 | 0.01937 | 7.442 | 0.5403 | 2.585 | 3.799 | True | True | False | False | |
| 0.01918 | 0.01939 | 7.199 | 0.5541 | 2.686 | 3.906 | True | True | False | False | |
| 0.01923 | 0.01944 | 4.738 | 0.1256 | 1.634 | 2.619 | True | True | False | False | |
| 0.01924 | 0.01944 | 5.006 | 0.1007 | 1.648 | 2.536 | True | True | False | False | |
| 0.01923 | 0.01944 | 5.034 | 0.09773 | 1.652 | 2.528 | True | True | False | False | |
| 0.01916 | 0.01937 | 7.387 | 0.5577 | 2.591 | 3.77 | True | True | False | False | |
| 0.01923 | 0.01944 | 4.806 | 0.1395 | 1.653 | 2.58 | True | True | False | False | |
| 0.01924 | 0.01945 | 4.904 | 0.09786 | 1.657 | 2.57 | True | True | False | False | |
| 0.01924 | 0.01945 | 4.784 | 0.1176 | 1.66 | 2.526 | True | True | False | False | |
| 0.01917 | 0.01937 | 7.412 | 0.599 | 2.582 | 3.965 | True | True | False | False | |
| 0.01924 | 0.01945 | 4.949 | 0.09078 | 1.687 | 2.504 | True | True | False | False | |
| 0.01924 | 0.01945 | 4.935 | 0.09008 | 1.694 | 2.516 | True | True | False | False | |

| SE_UKB_female | SE_UKB_male | LOG10P_VUMC_female | LOG10P_VUMC_male | LOG10P_UKB_female | LOG10P_UKB_male | female_effect_direction_replication | male_effect_direction_replication | sex_effect_comparison_replication | pvalue_effect_comparison_replication | replication_type |
|---|---|---|---|---|---|---|---|---|---|---|
| 0.01917 | 0.01938 | 7.346 | 0.5965 | 2.585 | 3.943 | True | True | False | False | |
| 0.01924 | 0.01945 | 4.973 | 0.08007 | 1.694 | 2.506 | True | True | False | False | |
| 0.01915 | 0.01937 | 7.415 | 0.5278 | 2.587 | 3.849 | True | True | False | False | |
| | | 4.945 | 0.07931 | | | False | False | False | False | |
| | | 7.441 | 0.5405 | | | False | False | False | False | |
| 0.01921 | 0.01939 | 4.582 | 0.1036 | 1.647 | 2.586 | True | True | False | False | |
| 0.0192 | 0.01939 | 4.653 | 0.07565 | 1.66 | 2.531 | True | True | False | False | |
| 0.0192 | 0.01939 | 4.63 | 0.0759 | 1.663 | 2.511 | True | True | False | False | |
| 0.01917 | 0.01934 | 6.999 | 0.5297 | 2.554 | 3.865 | True | True | False | False | |
| 0.0192 | 0.01939 | 4.583 | 0.08448 | 1.659 | 2.513 | True | True | False | False | |
| 0.01924 | 0.01945 | 4.925 | 0.07478 | 1.694 | 2.507 | True | True | False | False | |
| 0.01919 | 0.01938 | 4.508 | 0.09214 | 1.668 | 2.672 | True | True | False | False | |
| 0.01919 | 0.01938 | 4.508 | 0.09219 | 1.659 | 2.665 | True | True | False | False | |
| 0.01919 | 0.01939 | 4.508 | 0.09214 | 1.66 | 2.688 | True | True | False | False | |
| 0.01923 | 0.01945 | 4.848 | 0.07529 | 1.67 | 2.693 | True | True | False | False | |
| 0.01923 | 0.01945 | 4.848 | 0.07529 | 1.678 | 2.677 | True | True | False | False | |
| 0.01923 | 0.01945 | 4.77 | 0.0875 | 1.682 | 2.644 | True | True | False | False | |
| 0.01923 | 0.01944 | 4.773 | 0.0854 | 1.683 | 2.643 | True | True | False | False | |
| 0.01923 | 0.01945 | 4.777 | 0.07863 | 1.674 | 2.658 | True | True | False | False | |
| 0.01915 | 0.01935 | 7.196 | 0.5532 | 2.58 | 4.008 | True | True | False | False | |
| 0.02049 | 0.02057 | 4.072 | 0.4644 | 2.451 | 3.837 | True | True | False | False | |
| 0.01923 | 0.01945 | 4.774 | 0.0877 | 1.686 | 2.636 | True | True | False | False | |
| 0.01915 | 0.01935 | 7.127 | 0.5823 | 2.509 | 4.023 | True | True | False | False | |
| 0.01922 | 0.01945 | 4.777 | 0.08537 | 1.648 | 2.728 | True | True | False | False | |
| 0.01914 | 0.01935 | 7.22 | 0.5494 | 2.519 | 4.012 | True | True | False | False | |
| 0.01922 | 0.01944 | 4.878 | 0.06264 | 1.69 | 2.707 | True | True | False | False | |
| 0.01922 | 0.01944 | 5.075 | 0.1024 | 1.849 | 2.722 | True | True | False | False | |
| SE_UKB_female | SE_UKB_male | LOG10P_VUMC_female | LOG10P_VUMC_male | LOG10P_UKB_female | LOG10P_UKB_male | female_effect_direction_replication | male_effect_direction_replication | sex_effect_comparison_replication | pvalue_effect_comparison_replication | replication_type |
| 0.01912 | 0.01933 | 7.394 | 0.589 | 2.705 | 4.105 | True | True | False | False | |
| 0.01912 | 0.01933 | 7.404 | 0.5822 | 2.709 | 4.109 | True | True | False | False | |
| 0.01922 | 0.01944 | 4.918 | 0.1173 | 1.852 | 2.797 | True | True | False | False | |
| 0.01912 | 0.01933 | 7.342 | 0.6114 | 2.711 | 4.096 | True | True | False | False | |
| 0.01922 | 0.01945 | 4.936 | 0.1225 | 1.853 | 2.721 | True | True | False | False | |
| 0.01922 | 0.01945 | 4.97 | 0.1128 | 1.856 | 2.716 | True | True | False | False | |
| 0.01922 | 0.01945 | 4.97 | 0.1128 | 1.857 | 2.717 | True | True | False | False | |
| 0.01922 | 0.01944 | 4.899 | 0.1232 | 1.857 | 2.734 | True | True | False | False | |
| 0.01922 | 0.01945 | 5.007 | 0.1024 | 1.857 | 2.715 | True | True | False | False | |
| 0.01923 | 0.01946 | 5.007 | 0.1028 | 1.855 | 2.695 | True | True | False | False | |
| 0.01923 | 0.01946 | 5.004 | 0.1028 | 1.854 | 2.694 | True | True | False | False | |
| 0.01923 | 0.01946 | 5.004 | 0.1028 | 1.854 | 2.695 | True | True | False | False | |
| 0.01923 | 0.01946 | 4.975 | 0.1061 | 1.854 | 2.694 | True | True | False | False | |
| 0.01923 | 0.01946 | 4.899 | 0.1236 | 1.854 | 2.71 | True | True | False | False | |
| 0.01924 | 0.01946 | 4.877 | 0.1236 | 1.875 | 2.733 | True | True | False | False | |
| 0.01924 | 0.01946 | 4.85 | 0.1266 | 1.872 | 2.732 | True | True | False | False | |
| 0.01924 | 0.01946 | 5.007 | 0.1028 | 1.838 | 2.634 | True | True | False | False | |
| 0.01914 | 0.01936 | 7.391 | 0.5998 | 2.672 | 4.033 | True | True | False | False | |

| SE_UKB_female | SE_UKB_male | LOG10P_VUMC_female | LOG10P_VUMC_male | LOG10P_UKB_female | LOG10P_UKB_male | female_effect_direction_replication | male_effect_direction_replication | sex_effect_comparison_replication | pvalue_effect_comparison_replication | replication_type |
|---|---|---|---|---|---|---|---|---|---|---|
| 0.01924 | 0.01948 | 5.007 | 0.1024 | 1.854 | 2.684 | True | True | False | False | |
| 0.01914 | 0.01936 | 7.479 | 0.5942 | 2.732 | 4.029 | True | True | False | False | |
| 0.01928 | 0.01955 | 4.926 | 0.2201 | 1.743 | 2.606 | True | True | False | False | |
| 0.01928 | 0.01953 | 4.74 | 0.1135 | 1.799 | 2.482 | True | True | False | False | |
| 0.01918 | 0.01938 | 7.189 | 0.6004 | 2.658 | 3.701 | True | True | False | False | |
| 0.01928 | 0.01953 | 4.74 | 0.1135 | 1.8 | 2.483 | True | True | False | False | |
| 0.01918 | 0.01938 | 7.169 | 0.5999 | 2.658 | 3.706 | True | True | False | False | |
| 0.01918 | 0.01938 | 7.138 | 0.6003 | 2.657 | 3.697 | True | True | False | False | |
| 0.01932 | 0.01957 | 4.369 | 0.1155 | 1.832 | 2.417 | True | True | False | False | |
| 0.01919 | 0.01942 | 7.417 | 0.2087 | 2.585 | 3.272 | True | True | False | False | |
| 0.01919 | 0.01942 | 7.373 | 0.2118 | 2.571 | 3.265 | True | True | False | False | |
| 0.01913 | 0.01945 | 7.673 | 0.3234 | 2.558 | 3.567 | True | True | False | False | |
| 0.01916 | 0.01947 | 4.726 | 0.2121 | 2.872 | 2.099 | True | False | True | True | Sex-specific 1e-2 VUMC, sex-differential UKB |
| 0.01916 | 0.01947 | 4.737 | 0.2092 | 2.886 | 2.099 | True | False | True | True | Sex-specific 1e-2 VUMC, sex-differential UKB |
| 0.01916 | 0.01947 | 4.737 | 0.2092 | 2.872 | 2.089 | True | False | True | True | Sex-specific 1e-2 VUMC, sex-differential UKB |
| 0.01916 | 0.01947 | 4.737 | 0.2092 | 2.873 | 2.099 | True | False | True | True | Sex-specific 1e-2 VUMC, sex-differential UKB |
| 0.01916 | 0.01947 | 4.73 | 0.2045 | 2.867 | 2.099 | True | False | True | True | Sex-specific 1e-2 VUMC, sex-differential UKB |
| 0.01916 | 0.01947 | 4.732 | 0.2046 | 2.867 | 2.099 | True | False | True | True | Sex-specific 1e-2 VUMC, sex-differential UKB |
| 0.01916 | 0.01947 | 4.73 | 0.2045 | 2.88 | 2.099 | True | False | True | True | Sex-specific 1e-2 VUMC, sex-differential UKB |
| 0.01941 | 0.01965 | 5.419 | 0.1986 | 3.267 | 2.614 | True | False | True | True | Sex-specific 1e-2 VUMC, sex-differential UKB |
| 0.01916 | 0.01947 | 4.781 | 0.1976 | 2.867 | 2.099 | True | False | True | True | Sex-specific 1e-2 VUMC, sex-differential UKB |
| 0.01916 | 0.01947 | 4.772 | 0.1974 | 2.867 | 2.099 | True | False | True | True | Sex-specific 1e-2 VUMC, sex-differential UKB |
| 0.01916 | 0.01947 | 4.773 | 0.1976 | 2.867 | 2.099 | True | False | True | True | Sex-specific 1e-2 VUMC, sex-differential UKB |
| 0.01941 | 0.01966 | 5.472 | 0.1943 | 3.273 | 2.613 | True | False | True | True | Sex-specific 1e-2 VUMC, sex-differential UKB |
| 0.01916 | 0.01949 | 4.76 | 0.2122 | 2.881 | 2.107 | True | False | True | True | Sex-specific 1e-2 VUMC, sex-differential UKB |
| 0.01941 | 0.01967 | 5.361 | 0.2103 | 3.287 | 2.61 | True | False | True | True | Sex-specific 1e-2 VUMC, sex-differential UKB |
| 0.01941 | 0.01967 | 5.361 | 0.2102 | 3.298 | 2.61 | True | False | True | True | Sex-specific 1e-2 VUMC, sex-differential UKB |
| 0.01941 | 0.01967 | 5.378 | 0.2071 | 3.284 | 2.608 | True | False | True | True | Sex-specific 1e-2 VUMC, sex-differential UKB |
| 0.01958 | 0.01986 | 5.845 | 0.06723 | 3.282 | 2.832 | True | False | True | True | Sex-specific 1e-2 VUMC, sex-differential UKB |
| 0.01959 | 0.01986 | 5.789 | 0.05358 | 3.272 | 2.785 | True | False | True | True | Sex-specific 1e-2 VUMC, sex-differential UKB |
| 0.0196 | 0.01986 | 5.878 | 0.05352 | 3.222 | 2.77 | True | False | True | True | Sex-specific 1e-2 VUMC, sex-differential UKB |

| SE_UKB_female | SE_UKB_male | LOG10P_VUMC_female | LOG10P_VUMC_male | LOG10P_UKB_female | LOG10P_UKB_male | female_effect_direction_replication | male_effect_direction_replication | sex_effect_comparison_replication | pvalue_effect_comparison_replication | replication_type |
|---|---|---|---|---|---|---|---|---|---|---|
| 0.01942 | 0.01968 | 5.201 | 0.1543 | 3.254 | 2.611 | True | False | True | True | Sex-specific 1e-2 VUMC, sex-differential UKB |
| | | 4.614 | 0.1789 | | | False | False | False | False | |
| 0.01944 | 0.01969 | 5.276 | 0.1655 | 3.273 | 2.63 | True | False | True | True | Sex-specific 1e-2 VUMC, sex-differential UKB |
| 0.01918 | 0.01951 | 4.749 | 0.1834 | 2.791 | 2.142 | True | False | True | True | Sex-specific 1e-2 VUMC, sex-differential UKB |
| 0.01937 | 0.01969 | 4.804 | 0.166 | 2.861 | 2.506 | True | False | True | True | Sex-specific 1e-2 VUMC, sex-differential UKB |
| 0.0194 | 0.01972 | 4.845 | 0.176 | 2.881 | 2.48 | True | False | True | True | Sex-specific 1e-2 VUMC, sex-differential UKB |
| 0.01927 | 0.01961 | 4.658 | 0.1969 | 2.759 | 2.043 | True | False | True | True | Sex-specific 1e-2 VUMC, sex-differential UKB |
| 0.01972 | 0.02002 | 5.842 | 0.03945 | 3.163 | 2.552 | True | False | True | True | Sex-specific 1e-2 VUMC, sex-differential UKB |
| 0.01973 | 0.02002 | 5.763 | 0.05883 | 3.149 | 2.505 | True | False | True | True | Sex-specific 1e-2 VUMC, sex-differential UKB |
| 0.004359 | 0.004807 | 6.61 | 0.1251 | 267.83 | 44.81 | True | True | True | True | Sex-specific 1e-2 VUMC, sex-differential UKB |
| 0.004307 | 0.004756 | 4.88 | 0.07764 | 294.92 | 36.22 | True | True | True | True | Sex-specific 1e-2 VUMC, sex-differential UKB |
| SE_UKB_female | SE_UKB_male | LOG10P_VUMC_female | LOG10P_VUMC_male | LOG10P_UKB_female | LOG10P_UKB_male | female_effect_direction_replication | male_effect_direction_replication | sex_effect_comparison_replication | pvalue_effect_comparison_replication | replication_type |
| 0.00431 | 0.00476 | 4.97 | 0.07286 | 295.76 | 36.38 | True | True | True | True | Sex-specific 1e-2 VUMC, sex-differential UKB |
| 0.004329 | 0.00478 | 4.892 | 0.02224 | 300.37 | 36.87 | True | True | True | True | Sex-specific 1e-2 VUMC, sex-differential UKB |
| 0.00391 | 0.004327 | 9.764 | 0.8055 | Inf | 93.35 | True | True | True | True | Sex-specific 1e-2 VUMC, sex-differential UKB |
| 0.00391 | 0.004328 | 9.595 | 0.7804 | Inf | 93.23 | True | True | True | True | Sex-specific 1e-2 VUMC, sex-differential UKB |
| 0.003909 | 0.004328 | 9.536 | 0.8313 | Inf | 93.03 | True | True | True | True | Sex-specific 1e-2 VUMC, sex-differential UKB |
| 0.00391 | 0.004328 | 9.705 | 0.8041 | Inf | 92.96 | True | True | True | True | Sex-specific 1e-2 VUMC, sex-differential UKB |
| 0.003909 | 0.004327 | 9.709 | 0.806 | Inf | 92.79 | True | True | True | True | Sex-specific 1e-2 VUMC, sex-differential UKB |
| 0.004105 | 0.004549 | 10.82 | 1.109 | Inf | 107.35 | True | True | True | True | Sex-specific 1e-2 VUMC, sex-differential UKB |
| 0.003908 | 0.004326 | 9.221 | 0.8452 | Inf | 92.73 | True | True | True | True | Sex-specific 1e-2 VUMC, sex-differential UKB |
| 0.004347 | 0.004816 | 9.517 | 0.6882 | Inf | 88.42 | True | True | True | True | Sex-specific 1e-2 VUMC, sex-differential UKB |
| 0.003895 | 0.004312 | 9.535 | 0.811 | Inf | 93.59 | True | True | True | True | Sex-specific 1e-2 VUMC, sex-differential UKB |
| 0.003895 | 0.004312 | 9.525 | 0.814 | Inf | 93.1 | True | True | True | True | Sex-specific 1e-2 VUMC, sex-differential UKB |
| 0.003895 | 0.004312 | 9.527 | 0.8104 | Inf | 93.66 | True | True | True | True | Sex-specific 1e-2 VUMC, sex-differential UKB |
| 0.003895 | 0.004312 | 9.527 | 0.8104 | Inf | 93.66 | True | True | True | True | Sex-specific 1e-2 VUMC, sex-differential UKB |

| SE_UKB_female | SE_UKB_male | LOG10P_VUMC_female | LOG10P_VUMC_male | LOG10P_UKB_female | LOG10P_UKB_male | female_effect_direction_replication | male_effect_direction_replication | sex_effect_comparison_replication | pvalue_effect_comparison_replication | replication_type |
|---|---|---|---|---|---|---|---|---|---|---|
| 0.003895 | 0.004313 | 9.566 | 0.8374 | Inf | 93.73 | True | True | True | True | Sex-specific 1e-2 VUMC, sex-differential UKB |
| 0.003852 | 0.004261 | 9.459 | 1.25 | Inf | 91.36 | True | True | True | True | Sex-specific 1e-2 VUMC, sex-differential UKB |
| 0.003852 | 0.004261 | 9.423 | 1.252 | Inf | 91.42 | True | True | True | True | Sex-specific 1e-2 VUMC, sex-differential UKB |
| 0.003896 | 0.004313 | 9.542 | 0.7389 | Inf | 94.19 | True | True | True | True | Sex-specific 1e-2 VUMC, sex-differential UKB |
| 0.003896 | 0.004314 | 9.65 | 0.7464 | Inf | 94.11 | True | True | True | True | Sex-specific 1e-2 VUMC, sex-differential UKB |
| 0.003866 | 0.004276 | 9.958 | 1.248 | Inf | 90.12 | True | True | True | True | Sex-specific 1e-2 VUMC, sex-differential UKB |
| 0.003896 | 0.004312 | 9.492 | 0.81 | Inf | 94.15 | True | True | True | True | Sex-specific 1e-2 VUMC, sex-differential UKB |
| 0.004342 | 0.004811 | 9.584 | 0.712 | Inf | 88.85 | True | True | True | True | Sex-specific 1e-2 VUMC, sex-differential UKB |
| 0.003853 | 0.004261 | 9.815 | 1.268 | Inf | 91.57 | True | True | True | True | Sex-specific 1e-2 VUMC, sex-differential UKB |
| 0.004286 | 0.004747 | 10.91 | 0.6969 | Inf | 86.45 | True | True | True | True | Sex-specific 1e-2 VUMC, sex-differential UKB |
| 0.004048 | 0.004484 | 12.2 | 0.9467 | Inf | 107.05 | True | True | True | True | Sex-specific 1e-2 VUMC, sex-differential UKB |
| 0.004096 | 0.004539 | 10.63 | 1.056 | Inf | 108.89 | True | True | True | True | Sex-specific 1e-2 VUMC, sex-differential UKB |
| 0.004091 | 0.004535 | 10.9 | 0.9355 | Inf | 109.96 | True | True | True | True | Sex-specific 1e-2 VUMC, sex-differential UKB |
| 0.003997 | 0.004424 | 12.14 | 1.47 | Inf | 103.83 | True | True | True | True | Sex-specific 1e-2 VUMC, sex-differential UKB |
| 0.003999 | 0.004427 | 12.33 | 1.533 | Inf | 104.11 | True | True | True | True | Sex-specific 1e-2 VUMC, sex-differential UKB |
| 0.004053 | 0.00449 | 12.3 | 0.9314 | Inf | 107.32 | True | True | True | True | Sex-specific 1e-2 VUMC, sex-differential UKB |
| 0.004057 | 0.004495 | 12.22 | 0.9605 | Inf | 108.50 | True | True | True | True | Sex-specific 1e-2 VUMC, sex-differential UKB |
| 0.004058 | 0.004495 | 12.24 | 0.9542 | Inf | 108.77 | True | True | True | True | Sex-specific 1e-2 VUMC, sex-differential UKB |
| 0.004059 | 0.004497 | 12.35 | 0.9578 | Inf | 108.75 | True | True | True | True | Sex-specific 1e-2 VUMC, sex-differential UKB |
| 0.004104 | 0.004547 | 11.2 | 1.252 | Inf | 112.95 | True | True | True | True | Sex-specific 1e-2 VUMC, sex-differential UKB |
| 0.003696 | 0.004101 | 13.96 | 1.435 | Inf | 101.56 | True | True | True | True | Sex-specific 1e-2 VUMC, sex-differential UKB |
| 0.007149 | 0.005436 | 2.602 | 0.5283 | 0.2829 | 0.06574 | True | True | True | True | Sex-specific 1e-2 VUMC, sex-differential UKB |
| 0.007147 | 0.005435 | 2.068 | 0.5424 | 0.2511 | 0.02991 | True | True | True | True | Sex-specific 1e-2 VUMC, sex-differential UKB |
| 0.00714 | 0.005436 | 2.068 | 0.5432 | 0.2466 | 0.03268 | True | True | True | True | Sex-specific 1e-2 VUMC, sex-differential UKB |
| 0.007143 | 0.005438 | 2.06 | 0.5428 | 0.2007 | 0.05697 | True | True | True | True | Sex-specific 1e-2 VUMC, sex-differential UKB |
| 0.007144 | 0.005434 | 2.082 | 0.5415 | 0.2502 | 0.04945 | True | True | True | True | Sex-specific 1e-2 VUMC, sex-differential UKB |

| SE_UKB_female | SE_UKB_male | LOG10P_VUMC_female | LOG10P_VUMC_male | LOG10P_UKB_female | LOG10P_UKB_male | female_effect_direction_replication | male_effect_direction_replication | sex_effect_comparison_replication | pvalue_effect_comparison_replication | replication_type |
|---|---|---|---|---|---|---|---|---|---|---|
| 0.00713 | 0.005433 | 2.088 | 0.536 | 0.1964 | 0.05475 | True | True | True | True | Sex-specific 1e-2 VUMC, sex-differential UKB |
| 0.006914 | 0.005262 | 3.431 | 0.2475 | 0.1848 | 0.01937 | True | True | True | True | Sex-specific 1e-2 VUMC, sex-differential UKB |
| 0.006914 | 0.005263 | 3.488 | 0.2495 | 0.1256 | 0.02028 | True | True | True | True | Sex-specific 1e-2 VUMC, sex-differential UKB |
| 0.00691 | 0.005262 | 3.388 | 0.2487 | 0.13 | 0.04193 | True | True | True | True | Sex-specific 1e-2 VUMC, sex-differential UKB |
| 0.00691 | 0.005262 | 3.388 | 0.2487 | 0.13 | 0.04198 | True | True | True | True | Sex-specific 1e-2 VUMC, sex-differential UKB |
| 0.006912 | 0.005254 | 3.385 | 0.2474 | 0.1965 | 0.04101 | True | True | True | True | Sex-specific 1e-2 VUMC, sex-differential UKB |
| 0.006911 | 0.005253 | 3.384 | 0.2473 | 0.1953 | 0.04222 | True | True | True | True | Sex-specific 1e-2 VUMC, sex-differential UKB |
| 0.006907 | 0.005253 | 3.386 | 0.2482 | 0.1914 | 0.04146 | True | True | True | True | Sex-specific 1e-2 VUMC, sex-differential UKB |
| 0.006907 | 0.005253 | 3.387 | 0.2481 | 0.1973 | 0.04354 | True | True | True | True | Sex-specific 1e-2 VUMC, sex-differential UKB |
| 0.006905 | 0.005248 | 3.29 | 0.2482 | 0.1843 | 0.04629 | True | True | True | True | Sex-specific 1e-2 VUMC, sex-differential UKB |
| 0.006904 | 0.005248 | 3.304 | 0.2482 | 0.1796 | 0.04073 | True | True | True | True | Sex-specific 1e-2 VUMC, sex-differential UKB |
| 0.006899 | 0.005247 | 3.321 | 0.248 | 0.1704 | 0.03133 | True | True | True | True | Sex-specific 1e-2 VUMC, sex-differential UKB |
| 0.006903 | 0.005246 | 3.322 | 0.2477 | 0.1731 | 0.03103 | True | True | True | True | Sex-specific 1e-2 VUMC, sex-differential UKB |
| 0.006902 | 0.005246 | 3.321 | 0.2476 | 0.1733 | 0.02976 | True | True | True | True | Sex-specific 1e-2 VUMC, sex-differential UKB |
| 0.006902 | 0.005246 | 3.321 | 0.2477 | 0.1756 | 0.03013 | True | True | True | True | Sex-specific 1e-2 VUMC, sex-differential UKB |
| 0.006888 | 0.005239 | 2.653 | 0.2579 | 0.1652 | 0.008459 | True | True | True | True | Sex-specific 1e-2 VUMC, sex-differential UKB |
| 0.006905 | 0.005249 | 3.299 | 0.2478 | 0.1731 | 0.03806 | True | True | True | True | Sex-specific 1e-2 VUMC, sex-differential UKB |
| 0.006901 | 0.005245 | 3.312 | 0.252 | 0.1421 | 0.01842 | True | True | True | True | Sex-specific 1e-2 VUMC, sex-differential UKB |
| 0.006905 | 0.005249 | 3.322 | 0.2469 | 0.1739 | 0.02815 | True | True | True | True | Sex-specific 1e-2 VUMC, sex-differential UKB |
| 0.006891 | 0.005246 | 3.326 | 0.2477 | 0.1157 | 0.03021 | True | True | True | True | Sex-specific 1e-2 VUMC, sex-differential UKB |
| SE_UKB_female | SE_UKB_male | LOG10P_VUMC_female | LOG10P_VUMC_male | LOG10P_UKB_female | LOG10P_UKB_male | female_effect_direction_replication | male_effect_direction_replication | sex_effect_comparison_replication | pvalue_effect_comparison_replication | replication_type |
| 0.006896 | 0.005245 | 3.326 | 0.2477 | 0.1649 | 0.03124 | True | True | True | True | Sex-specific 1e-2 VUMC, sex-differential UKB |
| 0.00689 | 0.005245 | 3.33 | 0.2477 | 0.1197 | 0.03113 | True | True | True | True | Sex-specific 1e-2 VUMC, sex-differential UKB |
| 0.00689 | 0.005245 | 3.325 | 0.248 | 0.1199 | 0.03146 | True | True | True | True | Sex-specific 1e-2 VUMC, sex-differential UKB |
| 0.006898 | 0.005246 | 3.327 | 0.2484 | 0.1731 | 0.03054 | True | True | True | True | Sex-specific 1e-2 VUMC, sex-differential UKB |
| 0.006895 | 0.005246 | 3.325 | 0.2479 | 0.1544 | 0.02992 | True | True | True | True | Sex-specific 1e-2 VUMC, sex-differential UKB |

| SE_UKB_female | SE_UKB_male | LOG10P_VUMC_female | LOG10P_VUMC_male | LOG10P_UKB_female | LOG10P_UKB_male | female_effect_direction_replication | male_effect_direction_replication | sex_effect_comparison_replication | pvalue_effect_comparison_replication | replication_type |
|---|---|---|---|---|---|---|---|---|---|---|
| 0.006901 | 0.005246 | 3.327 | 0.2477 | 0.1616 | 0.0307 | True | True | True | True | Sex-specific 1e-2 VUMC, sex-differential UKB |
| 0.006886 | 0.005245 | 3.3 | 0.2483 | 0.1042 | 0.03768 | True | True | True | True | Sex-specific 1e-2 VUMC, sex-differential UKB |
| 0.006883 | 0.005245 | 3.304 | 0.248 | 0.107 | 0.03493 | True | True | True | True | Sex-specific 1e-2 VUMC, sex-differential UKB |
| 0.006881 | 0.005243 | 3.335 | 0.2483 | 0.1033 | 0.03797 | True | True | True | True | Sex-specific 1e-2 VUMC, sex-differential UKB |
| 0.006827 | 0.005239 | 3.335 | 0.2483 | 0.06803 | 0.02451 | True | True | True | True | Sex-specific 1e-2 VUMC, sex-differential UKB |
| 0.006881 | 0.005243 | 3.332 | 0.2484 | 0.106 | 0.03333 | True | True | True | True | Sex-specific 1e-2 VUMC, sex-differential UKB |
| 0.006882 | 0.005245 | 3.331 | 0.2485 | 0.1063 | 0.03248 | True | True | True | True | Sex-specific 1e-2 VUMC, sex-differential UKB |
| 0.006884 | 0.005245 | 3.334 | 0.2482 | 0.1064 | 0.03861 | True | True | True | True | Sex-specific 1e-2 VUMC, sex-differential UKB |
| 0.006885 | 0.005246 | 3.335 | 0.2479 | 0.1114 | 0.03948 | True | True | True | True | Sex-specific 1e-2 VUMC, sex-differential UKB |
| 0.006884 | 0.005245 | 3.433 | 0.2486 | 0.1069 | 0.04264 | True | True | True | True | Sex-specific 1e-2 VUMC, sex-differential UKB |
| 0.006884 | 0.005245 | 3.328 | 0.2473 | 0.1068 | 0.03741 | True | True | True | True | Sex-specific 1e-2 VUMC, sex-differential UKB |
| 0.006888 | 0.005239 | 4.333 | 0.3391 | 0.1122 | 0.06541 | True | True | True | True | Sex-specific 1e-2 VUMC, sex-differential UKB |
| 0.006891 | 0.005244 | 4.364 | 0.3394 | 0.1002 | 0.0906 | True | True | True | True | Sex-specific 1e-2 VUMC, sex-differential UKB |
| 0.006937 | 0.005275 | 4.266 | 0.5097 | 0.08968 | 0.1834 | True | True | True | False | |
| 0.006936 | 0.005274 | 4.265 | 0.51 | 0.09007 | 0.1805 | True | True | True | False | |
| 0.006939 | 0.005275 | 4.299 | 0.4664 | 0.08268 | 0.1918 | True | True | True | False | |
| 0.006993 | 0.005313 | 3.846 | 0.525 | 0.1084 | 0.2428 | True | True | True | False | |
| 0.006194 | 0.004702 | 7.221 | 0.01685 | 0.1289 | 0.07332 | False | True | False | True | |
| 0.006185 | 0.004697 | 7.28 | 0.01288 | 0.1454 | 0.1099 | False | True | False | True | |
| 0.006182 | 0.004694 | 7.377 | 0.002492 | 0.1565 | 0.08498 | False | True | False | True | |
| 0.006096 | 0.004627 | 7.74 | 6.84e-04 | 0.06351 | 0.03555 | False | False | False | True | |
| 0.006097 | 0.004627 | 7.665 | 0.00361 | 0.09679 | 0.01879 | False | False | False | True | |
| 0.006098 | 0.004628 | 7.547 | 0.00363 | 0.08575 | 0.02487 | False | False | False | True | |
| 0.006093 | 0.004622 | 7.452 | 0.008889 | 0.05493 | 0.03203 | False | True | False | True | |
| 0.006092 | 0.00462 | 7.495 | 0.02111 | 0.07764 | 0.03071 | False | True | False | True | |
| 0.006089 | 0.004619 | 7.321 | 0.03705 | 0.02833 | 0.0926 | False | False | True | False | |

| SE_UKB_female | SE_UKB_male | LOG10P_VUMC_female | LOG10P_VUMC_male | LOG10P_UKB_female | LOG10P_UKB_male | female_effect_direction_replication | male_effect_direction_replication | sex_effect_comparison_replication | pvalue_effect_comparison_replication | replication_type |
|---|---|---|---|---|---|---|---|---|---|---|
| 0.006085 | 0.004615 | 7.451 | 0.02249 | 0.04271 | 0.09026 | False | False | True | False | |
| 0.006081 | 0.004612 | 7.43 | 0.02034 | 0.06612 | 0.08138 | False | False | False | False | |
| 0.006058 | 0.004614 | 7.647 | 0.02474 | 0.1515 | 0.1185 | False | False | False | True | |
| 0.00608 | 0.004616 | 7.647 | 0.02474 | 0.0977 | 0.1015 | False | False | False | False | |
| 0.006085 | 0.00462 | 7.59 | 0.02201 | 0.1025 | 0.1011 | False | False | False | True | |
| 0.006085 | 0.00462 | 7.606 | 0.02421 | 0.103 | 0.09784 | False | False | False | True | |
| 0.006085 | 0.00462 | 7.606 | 0.02421 | 0.1026 | 0.1012 | False | False | False | True | |
| 0.006085 | 0.00462 | 7.527 | 0.03092 | 0.09999 | 0.09331 | False | False | False | True | |
| 0.006086 | 0.004621 | 7.509 | 0.03104 | 0.09611 | 0.09826 | False | False | False | False | |
| 0.006086 | 0.004621 | 7.523 | 0.03092 | 0.1015 | 0.09603 | False | False | False | True | |
| 0.006322 | 0.0048 | 7.969 | 0.09757 | 0.06321 | 0.1206 | True | False | True | False | |
| 0.006322 | 0.0048 | 7.949 | 0.09782 | 0.06462 | 0.1132 | True | False | True | False | |
| 0.006323 | 0.0048 | 7.949 | 0.09782 | 0.06251 | 0.1166 | True | False | True | False | |
| 0.006324 | 0.004803 | 7.999 | 0.1042 | 0.06524 | 0.1242 | True | False | True | False | |
| 0.006914 | 0.005283 | 2.648 | 0.2931 | 0.01803 | 0.1031 | True | True | True | False | |
| 0.006914 | 0.005284 | 2.692 | 0.2951 | 0.02749 | 0.1027 | False | True | True | False | |
| 0.00691 | 0.005283 | 2.615 | 0.2986 | 0.008707 | 0.1319 | False | True | True | False | |
| 0.00691 | 0.005283 | 2.615 | 0.2986 | 0.008676 | 0.1319 | False | True | True | False | |
| 0.006912 | 0.005275 | 2.617 | 0.2965 | 0.04162 | 0.1283 | True | True | True | False | |
| 0.006911 | 0.005274 | 2.617 | 0.2967 | 0.04167 | 0.1303 | True | True | True | False | |
| 0.006907 | 0.005274 | 2.619 | 0.2972 | 0.03841 | 0.1291 | True | True | True | False | |
| 0.006907 | 0.005274 | 2.619 | 0.297 | 0.04105 | 0.132 | True | True | True | False | |
| 0.006905 | 0.005269 | 2.58 | 0.3009 | 0.03571 | 0.1372 | True | True | True | False | |
| 0.006904 | 0.005269 | 2.592 | 0.3009 | 0.03284 | 0.1315 | True | True | True | False | |
| 0.006899 | 0.005268 | 2.655 | 0.3006 | 0.01815 | 0.1189 | True | True | True | False | |
| 0.006903 | 0.005267 | 2.657 | 0.3005 | 0.02107 | 0.1181 | True | True | True | False | |
| 0.006902 | 0.005267 | 2.657 | 0.3006 | 0.02109 | 0.1161 | True | True | True | False | |
| 0.006902 | 0.005267 | 2.656 | 0.3007 | 0.02312 | 0.1165 | True | True | True | False | |
| 0.006888 | 0.00526 | 2.31 | 0.2938 | 0.01227 | 0.08909 | True | True | True | False | |

| SE_UKB_female | SE_UKB_male | LOG10P_VUMC_female | LOG10P_VUMC_male | LOG10P_UKB_female | LOG10P_UKB_male | female_effect_direction_replication | male_effect_direction_replication | sex_effect_comparison_replication | pvalue_effect_comparison_replication | replication_type |
|---|---|---|---|---|---|---|---|---|---|---|
| 0.006905 | 0.00527 | 2.643 | 0.3007 | 0.02248 | 0.1234 | True | True | True | False | |
| 0.006901 | 0.005266 | 2.66 | 0.2844 | 0.001647 | 0.1125 | True | True | True | False | |
| 0.006905 | 0.00527 | 2.658 | 0.2989 | 0.01906 | 0.1133 | True | True | True | False | |
| 0.006891 | 0.005267 | 2.658 | 0.3005 | 0.02527 | 0.1151 | False | True | True | False | |
| 0.006896 | 0.005266 | 2.658 | 0.3005 | 0.01112 | 0.1171 | True | True | True | False | |
| 0.00689 | 0.005266 | 2.662 | 0.3 | 0.0173 | 0.1165 | False | True | True | False | |
| 0.00689 | 0.005266 | 2.657 | 0.3009 | 0.01737 | 0.1172 | False | True | True | False | |
| 0.006898 | 0.005267 | 2.659 | 0.3008 | 0.02244 | 0.1151 | True | True | True | False | |
| 0.006895 | 0.005267 | 2.658 | 0.3007 | 0.00571 | 0.1142 | True | True | True | False | |
| 0.006901 | 0.005267 | 2.66 | 0.3004 | 0.01436 | 0.115 | True | True | True | False | |
| 0.006886 | 0.005266 | 2.641 | 0.3009 | 0.03616 | 0.1197 | False | True | True | False | |
| 0.006884 | 0.005265 | 2.645 | 0.301 | 0.03187 | 0.1174 | False | True | True | False | |
| 0.006881 | 0.005264 | 2.665 | 0.3009 | 0.03539 | 0.1197 | False | True | True | False | |
| 0.006827 | 0.005259 | 2.664 | 0.3011 | 0.06415 | 0.1054 | False | True | True | False | |
| 0.006881 | 0.005264 | 2.661 | 0.3013 | 0.03425 | 0.1127 | False | True | True | False | |
| 0.006882 | 0.005266 | 2.662 | 0.3012 | 0.03514 | 0.1136 | False | True | True | False | |
| 0.006884 | 0.005266 | 2.664 | 0.3008 | 0.03215 | 0.1217 | False | True | True | False | |
| 0.006885 | 0.005267 | 2.664 | 0.3005 | 0.0279 | 0.1206 | False | True | True | False | |
| 0.006884 | 0.005266 | 2.731 | 0.3014 | 0.03243 | 0.1239 | False | True | True | False | |
| 0.006884 | 0.005266 | 2.647 | 0.3023 | 0.02929 | 0.1181 | False | True | True | False | |
| 0.006888 | 0.00526 | 3.195 | 0.3233 | 0.06541 | 0.1236 | False | True | True | False | |
| 0.006891 | 0.005265 | 3.19 | 0.323 | 0.07467 | 0.1543 | False | True | True | False | |
| 0.006937 | 0.005296 | 3.086 | 0.4288 | 0.09077 | 0.2467 | False | True | True | False | |
| 0.006936 | 0.005296 | 3.086 | 0.4289 | 0.09558 | 0.2406 | False | True | True | False | |
| 0.006939 | 0.005296 | 3.11 | 0.41 | 0.1022 | 0.2527 | False | True | True | False | |
| 0.006993 | 0.005335 | 3.034 | 0.4767 | 0.0873 | 0.2953 | False | True | True | False | |
| 0.006194 | 0.004721 | 7.068 | 0.06935 | 0.6605 | 0.02244 | False | True | False | True | |

| SE_UKB_female | SE_UKB_male | LOG10P_VUMC_female | LOG10P_VUMC_male | LOG10P_UKB_female | LOG10P_UKB_male | female_effect_direction_replication | male_effect_direction_replication | sex_effect_comparison_replication | pvalue_effect_comparison_replication | replication_type |
|---|---|---|---|---|---|---|---|---|---|---|
| 0.006185 | 0.004715 | 7.17 | 0.06537 | 0.6764 | 0.04293 | False | True | False | True | |
| 0.006182 | 0.004713 | 7.371 | 0.07192 | 0.6862 | 0.0203 | False | True | False | True | |
| 0.006096 | 0.004646 | 8.164 | 0.02961 | 0.5314 | 0.04608 | False | False | False | True | |
| 0.006097 | 0.004646 | 8.168 | 0.02943 | 0.5818 | 0.06406 | False | False | False | True | |
| 0.006098 | 0.004647 | 8.031 | 0.02941 | 0.5693 | 0.05441 | False | False | False | True | |
| 0.006093 | 0.00464 | 7.953 | 0.0704 | 0.5294 | 0.04021 | False | False | False | True | |
| 0.006092 | 0.004638 | 8.023 | 0.08125 | 0.5599 | 0.03567 | False | False | False | True | |
| 0.006089 | 0.004637 | 7.895 | 0.02033 | 0.4915 | 0.02262 | False | True | False | True | |
| 0.006085 | 0.004634 | 8.036 | 0.03127 | 0.5128 | 0.02294 | False | True | False | True | |
| 0.006081 | 0.004631 | 8.042 | 0.03672 | 0.5424 | 0.01013 | False | True | False | True | |
| 0.006058 | 0.004632 | 8.206 | 0.01964 | 0.6981 | 0.06049 | False | True | False | True | |
| 0.00608 | 0.004634 | 8.206 | 0.01964 | 0.5734 | 0.04173 | False | True | False | True | |
| 0.006085 | 0.004639 | 8.233 | 0.01295 | 0.5706 | 0.04076 | False | True | False | True | |
| 0.006085 | 0.004639 | 8.256 | 0.01148 | 0.5714 | 0.03896 | False | True | False | True | |
| 0.006085 | 0.004639 | 8.256 | 0.01148 | 0.571 | 0.04097 | False | True | False | True | |
| 0.006085 | 0.004639 | 8.24 | 0.006446 | 0.5639 | 0.03497 | False | True | False | True | |
| 0.006086 | 0.004639 | 8.219 | 0.006364 | 0.5582 | 0.03992 | False | True | False | True | |
| 0.006086 | 0.00464 | 8.231 | 0.006446 | 0.5663 | 0.03693 | False | True | False | True | |
| 0.006322 | 0.004819 | 8.116 | 0.06799 | 0.2675 | 0.02549 | False | False | False | True | |
| 0.006322 | 0.004819 | 8.097 | 0.06797 | 0.2654 | 0.01988 | False | False | False | True | |
| 0.006323 | 0.00482 | 8.097 | 0.06797 | 0.2643 | 0.02298 | False | False | False | True | |
| 0.006324 | 0.004822 | 8.142 | 0.08143 | 0.2705 | 0.02709 | False | False | False | True | |
| 0.003418 | 0.003623 | 4.053 | 0.6233 | 26.91 | 26.85 | True | True | False | True | |
| 0.003418 | 0.003623 | 4.026 | 0.6364 | 26.93 | 26.81 | True | True | False | True | |
| 0.003418 | 0.003623 | 4.016 | 0.6552 | 26.9 | 26.81 | True | True | False | True | |
| 0.00342 | 0.003625 | 4.25 | 0.5389 | 26.56 | 25.94 | True | True | False | True | |
| 0.00342 | 0.003625 | 4.208 | 0.5357 | 26.54 | 26.02 | True | True | False | True | |
| | | 4.207 | 0.5304 | | | False | False | False | False | |

| SE_UKB_female | SE_UKB_male | LOG10P_VUMC_female | LOG10P_VUMC_male | LOG10P_UKB_female | LOG10P_UKB_male | female_effect_direction_replication | male_effect_direction_replication | sex_effect_comparison_replication | pvalue_effect_comparison_replication | replication_type |
|---|---|---|---|---|---|---|---|---|---|---|
| 0.00342 | 0.003625 | 4.213 | 0.5368 | 26.57 | 25.97 | True | True | False | True | |
| 0.003419 | 0.003624 | 4.159 | 0.4714 | 26.66 | 25.98 | True | True | False | True | |
| 0.003419 | 0.003623 | 4.148 | 0.4713 | 26.69 | 25.93 | True | True | False | True | |
| 0.00342 | 0.003624 | 4.227 | 0.5047 | 26.58 | 26.06 | True | True | False | True | |
| 0.00342 | 0.003624 | 4.283 | 0.587 | 26.79 | 26.17 | True | True | False | True | |

| SE_UKB_female | SE_UKB_male | LOG10P_VUMC_female | LOG10P_VUMC_male | LOG10P_UKB_female | LOG10P_UKB_male | female_effect_direction_replication | male_effect_direction_replication | sex_effect_comparison_replication | pvalue_effect_comparison_replication | replication_type |
|---|---|---|---|---|---|---|---|---|---|---|
| 0.00342 | 0.003624 | 4.22 | 0.5476 | 26.62 | 26.01 | True | True | False | True | |
| 0.00342 | 0.003624 | 4.179 | 0.5759 | 26.75 | 26.11 | True | True | False | True | |
| 0.00342 | 0.003624 | 4.179 | 0.5759 | 26.71 | 26.07 | True | True | False | True | |
| 0.00342 | 0.003624 | 4.179 | 0.5759 | 26.71 | 26.06 | True | True | False | True | |
| 0.00342 | 0.003624 | 4.182 | 0.5277 | 26.59 | 25.99 | True | True | False | True | |
| 0.003421 | 0.003626 | 4.013 | 0.4723 | 26.96 | 25.91 | True | True | False | True | |
| 0.003378 | 0.003567 | 6.739 | 0.9245 | 192.75 | 174.60 | True | True | False | True | |
| 0.003481 | 0.003684 | 7.501 | 0.9452 | 129.84 | 118.47 | True | True | False | True | |
| 0.003411 | 0.003611 | 5.488 | 0.2863 | 132.52 | 134.87 | True | True | False | False | |
| 0.003479 | 0.003683 | 7.657 | 0.8696 | 130.38 | 118.95 | True | True | False | True | |
| 0.003479 | 0.003683 | 7.651 | 0.856 | 130.27 | 118.94 | True | True | False | True | |
| 0.003479 | 0.003683 | 7.651 | 0.856 | 130.39 | 118.96 | True | True | False | True | |
| 0.003411 | 0.003611 | 5.468 | 0.287 | 132.52 | 135.06 | True | True | False | False | |
| 0.003479 | 0.003683 | 7.644 | 0.8466 | 130.50 | 118.56 | True | True | False | True | |
| 0.003405 | 0.003607 | 8.712 | 0.7503 | 161.65 | 149.95 | True | True | False | True | |
| 0.003409 | 0.003609 | 5.916 | 0.2668 | 134.89 | 136.26 | True | True | False | False | |
| 0.003408 | 0.003608 | 6.097 | 0.2662 | 135.25 | 136.15 | True | True | False | False | |
| 0.003408 | 0.003608 | 6.082 | 0.2691 | 135.22 | 136.54 | True | True | False | False | |
| 0.003417 | 0.003625 | 8.587 | 0.7379 | 149.41 | 138.95 | True | True | False | True | |
| 0.003419 | 0.003626 | 8.633 | 0.7843 | 149.65 | 138.32 | True | True | False | True | |

| SE_UKB_female | SE_UKB_male | LOG10P_VUMC_female | LOG10P_VUMC_male | LOG10P_UKB_female | LOG10P_UKB_male | female_effect_direction_replication | male_effect_direction_replication | sex_effect_comparison_replication | pvalue_effect_comparison_replication | replication_type |
|---|---|---|---|---|---|---|---|---|---|---|
| 0.003383 | 0.003585 | 3.618 | 0.006875 | 64.21 | 70.41 | True | False | False | False | |
| 0.003382 | 0.003582 | 3.598 | 0.01137 | 63.97 | 69.67 | True | True | False | False | |
| 0.003917 | 0.004225 | 0.6915 | 6.911 | 50.59 | 40.99 | True | True | False | False | |
| 0.004256 | 0.004602 | 1.141 | 6.949 | 71.48 | 53.53 | True | True | False | False | |
| 0.00426 | 0.004606 | 1.122 | 7.071 | 72.2 | 53.54 | True | True | False | False | |
| 0.003896 | 0.004209 | 0.7526 | 7.343 | 51.93 | 34.61 | True | True | False | False | |
| 0.004401 | 0.00476 | 1.008 | 8.736 | 72.24 | 55.05 | True | True | False | False | |
| 0.003845 | 0.004153 | 0.5655 | 8.628 | 50.3 | 36.26 | True | True | False | False | |
| 0.004255 | 0.004601 | 1.116 | 7.07 | 72.22 | 54.05 | True | True | False | False | |
| 0.004255 | 0.004601 | 1.116 | 7.07 | 72.39 | 54 | True | True | False | False | |
| 0.004401 | 0.004759 | 0.9914 | 8.736 | 72.22 | 55.43 | True | True | False | False | |
| 0.004262 | 0.004609 | 1.228 | 7.389 | 72.31 | 55.2 | True | True | False | False | |
| 0.004166 | 0.004501 | 0.7673 | 9.184 | 68.94 | 56.5 | True | True | False | False | |
| 0.004243 | 0.004587 | 1.106 | 7.166 | 72.63 | 56.08 | True | True | False | False | |
| 0.004251 | 0.004596 | 1.196 | 7.539 | 72.32 | 57.14 | True | True | False | False | |
| 0.003837 | 0.004142 | 0.5229 | 9.055 | 51.62 | 38.08 | True | True | False | False | |
| 0.004242 | 0.004585 | 1.129 | 7.236 | 72.18 | 56.16 | True | True | False | False | |
| 0.004242 | 0.004585 | 1.129 | 7.236 | 72.18 | 56.16 | True | True | False | False | |
| 0.004386 | 0.004742 | 1.014 | 8.882 | 72.19 | 56.61 | True | True | False | False | |
| 0.004232 | 0.004575 | 1.005 | 7.936 | 72.9 | 56.29 | True | True | False | False | |
| 0.00424 | 0.004583 | 1.106 | 7.525 | 73.05 | 57.95 | True | True | False | False | |
| 0.004226 | 0.004569 | 1.124 | 7.674 | 73.45 | 56.62 | True | True | False | False | |
| 0.004161 | 0.004495 | 0.9226 | 9.538 | 69.93 | 58.93 | True | True | False | False | |
| 0.004241 | 0.004585 | 1.121 | 7.543 | 73.27 | 57.82 | True | True | False | False | |
| 0.004151 | 0.004485 | 0.8714 | 9.109 | 69.98 | 57.55 | True | True | False | False | |
| 0.004156 | 0.00449 | 0.8712 | 8.866 | 69.61 | 57.97 | True | True | False | False | |
| 0.004151 | 0.004485 | 0.8426 | 9.099 | 70.06 | 57.44 | True | True | False | False | |

| SE_UKB_female | SE_UKB_male | LOG10P_VUMC_female | LOG10P_VUMC_male | LOG10P_UKB_female | LOG10P_UKB_male | female_effect_direction_replication | male_effect_direction_replication | sex_effect_comparison_replication | pvalue_effect_comparison_replication | replication_type |
|---|---|---|---|---|---|---|---|---|---|---|
| 0.004242 | 0.004585 | 1.09 | 7.546 | 73.2 | 57.51 | True | True | False | False | |
| 0.004244 | 0.004588 | 1.175 | 7.465 | 72.61 | 56.93 | True | True | False | False | |
| 0.003589 | 0.002742 | 0.1314 | 5.666 | 0.02904 | 0.4917 | False | True | True | True | Sex-specific 1e-2 VUMC, sex-differential UKB |
| 0.003554 | 0.002718 | 0.2174 | 5.85 | 0.07831 | 0.8138 | False | True | True | True | Sex-specific 1e-2 VUMC, sex-differential UKB |
| 0.003095 | 0.002356 | 0.3598 | 4.013 | 0.05891 | 0.1439 | False | True | True | True | Sex-specific 1e-2 VUMC, sex-differential UKB |
| 0.003375 | 0.002573 | 0.6103 | 7.834 | 0.2624 | 0.01013 | True | False | True | False | |

#### STable9: Unique loci identified as associated with per-person variances of clinical quantitative trait values using plink

116 rows × 18 columns

| pheno | CHR | Gene_Symbol | biotype | lead_snp | lead_BP | lead_P | TOTAL | NSIG | S05 | S01 | S001 | S0001 | independent_loci_in_sex_P1e6 | independent_loci_in_sex_P1e2 | independent_loci_in_sex_P5e8 | in_HLA | any_overlap |
|---|---|---|---|---|---|---|---|---|---|---|---|---|---|---|---|---|---|
| A.2Gg_ | 19 | OR7A5 | protein_coding | rs59889385 | 14943775 | 2.52e-08 | 15 | 8 | 0 | 0 | 0 | 7 | Female only | Female only | Female only | False | False |
| BASOAB | 19 | CTD-2540B15.12 | lincRNA | rs78744187 | 33754548 | 4.32e-36 | 5 | 0 | 0 | 0 | 0 | 5 | Both sexes | Both sexes | Both sexes | False | True |
| BASOAB | 3 | RPN1 | protein_coding | rs6782812 | 128317997 | 1.09e-19 | 53 | 0 | 0 | 1 | 0 | 52 | Both sexes | Both sexes | Both sexes | False | True |
| BASOAB | 19 | CTD-2540B15.12 | lincRNA | rs67228209 | 33751033 | 2.25e-10 | 3 | 0 | 0 | 0 | 0 | 3 | Female only | Both sexes | Female only | False | True |
| BASOAB | 3 | C3orf27 | protein_coding | rs6806253 | 128282697 | 8.12e-09 | 81 | 0 | 2 | 19 | 19 | 41 | Female only | Both sexes | Female only | False | True |
| BASOAB | 14 | CEBPE | protein_coding | rs8015478 | 23586018 | 1.11e-08 | 7 | 0 | 0 | 0 | 0 | 7 | Both sexes | Both sexes | Male only | False | True |
| BASOAB | 19 | CTD-2540B15.12 | lincRNA | rs10412166 | 33758201 | 2.49e-08 | 5 | 2 | 0 | 0 | 0 | 3 | Female only | Both sexes | Female only | False | True |
| BASORE | 19 | CTD-2540B15.12 | lincRNA | rs78744187 | 33754548 | 7.10e-47 | 5 | 0 | 0 | 0 | 0 | 5 | Both sexes | Both sexes | Both sexes | False | True |
| BASORE | 3 | RPN1 | protein_coding | rs6782812 | 128317997 | 1.00e-21 | 53 | 0 | 0 | 0 | 1 | 52 | Both sexes | Both sexes | Both sexes | False | True |
| BASORE | 19 | CTD-2540B15.12 | lincRNA | rs67228209 | 33751033 | 4.02e-11 | 3 | 0 | 0 | 0 | 0 | 3 | Both sexes | Both sexes | Both sexes | False | True |
| BASORE | 3 | C3orf27 | protein_coding | rs8179974 | 128281701 | 7.31e-10 | 123 | 0 | 0 | 15 | 26 | 82 | Female only | Both sexes | Female only | False | True |
| BASORE | 3 | CRBN | protein_coding | rs62228659 | 3217928 | 7.43e-10 | 41 | 5 | 16 | 0 | 0 | 20 | Female only | Female only | Female only | False | True |
| BASORE | 19 | CTD-2540B15.12 | lincRNA | rs12461296 | 33774591 | 9.40e-10 | 3 | 0 | 0 | 0 | 1 | 2 | Male only | Both sexes | Male only | False | True |
| BASORE | 14 | CEBPE | protein_coding | rs8015478 | 23586018 | 3.23e-09 | 7 | 0 | 0 | 0 | 0 | 7 | Both sexes | Both sexes | Male only | False | True |
| BASORE | 8 | CCDC26 | lincRNA | rs10100356 | 130626164 | 3.86e-08 | 31 | 16 | 0 | 0 | 0 | 15 | Female only | Female only | Female only | False | False |
| BAbs | 19 | CTD-2540B15.12 | lincRNA | rs78744187 | 33754548 | 7.51e-10 | 5 | 0 | 0 | 0 | 1 | 4 | Both sexes | Both sexes | Both sexes | False | True |
| BMI_CLEAN | 16 | FTO | protein_coding | rs55872725 | 53809123 | 5.25e-09 | 169 | 0 | 0 | 2 | 15 | 152 | Both sexes | Both sexes | Female only | False | True |
| Baso | 19 | CTD-2540B15.12 | lincRNA | rs78744187 | 33754548 | 7.99e-14 | 5 | 0 | 0 | 0 | 0 | 5 | Both sexes | Both sexes | Both sexes | False | True |
| Chol | 19 | LDLR | protein_coding | rs56289821 | 11188247 | 2.36e-08 | 75 | 0 | 0 | 0 | 38 | 37 | Male only | Both sexes | Male only | False | True |
| EOSIM | 5 | AC034220.3 | processed_transcript | rs4705938 | 131694077 | 4.80e-08 | 249 | 121 | 4 | 1 | 35 | 88 | Male only | Male only | Male only | False | False |
| EOSIRE | 11 | MAML2 | protein_coding | rs568477 | 95746897 | 1.04e-08 | 15 | 8 | 0 | 4 | 0 | 3 | Female only | Female only | Female only | False | False |
| EoAbs | 3 | RPN1 | protein_coding | rs6782812 | 128317997 | 2.25e-08 | 53 | 0 | 0 | 4 | 26 | 23 | Female only | Both sexes | Female only | False | True |
| EoAbs | 5 | C5orf56 | protein_coding | 5:131744482 | 131744482 | 3.19e-08 | 67 | 27 | 7 | 0 | 0 | 33 | Female only | Female only | Female only | False | True |
| Eos | 3 | C3orf27 | protein_coding | rs4328821 | 128316435 | 6.49e-10 | 53 | 0 | 0 | 4 | 20 | 29 | Female only | Both sexes | Female only | False | True |

| pheno | CHR | Gene_Symbol | biotype | lead_snp | lead_BP | lead_P | TOTAL | NSIG | S05 | S01 | S001 | S0001 | independent_loci_in_sex_P1e6 | independent_loci_in_sex_P1e2 | independent_loci_in_sex_P5e8 | in_HLA | any_overlap |
|---|---|---|---|---|---|---|---|---|---|---|---|---|---|---|---|---|---|
| Eos | 5 | C5orf56 | protein_coding | 5:131744482 | 131744482 | 6.54e-09 | 67 | 34 | 0 | 0 | 0 | 33 | Female only | Female only | Female only | False | False |
| FEV1_PPR | 8 | RP11-48B3.2 | pseudogene | rs73691933 | 81484389 | 1.47e-08 | 15 | 8 | 0 | 0 | 0 | 7 | Female only | Female only | Female only | False | False |
| FEV1_ZBD | 10 | LRRC20 | protein_coding | rs10762368 | 72109482 | 3.89e-08 | 25 | 13 | 0 | 0 | 2 | 10 | Male only | Male only | Male only | False | False |
| FRONTAL.AXIS.MEAN.QRS | 4 | SLC2A9 | protein_coding | rs4697693 | 9895860 | 2.98e-08 | 19 | 10 | 0 | 0 | 0 | 9 | Male only | Male only | Male only | False | False |
| GluBed | 10 | TCF7L2 | protein_coding | rs35198068 | 114754784 | 7.58e-13 | 47 | 0 | 0 | 0 | 0 | 47 | Both sexes | Both sexes | Male only | False | True |
| GluBed | 6 | HLA-DQA1 | protein_coding | rs9271364 | 32586787 | 2.89e-10 | 21 | 0 | 0 | 3 | 2 | 16 | Female only | Both sexes | Female only | True | False |
| Gluc | 10 | TCF7L2 | protein_coding | rs35198068 | 114754784 | 1.19e-16 | 47 | 0 | 0 | 0 | 0 | 47 | Both sexes | Both sexes | Both sexes | False | True |
| Gluc | 6 | HLA-DQB1 | protein_coding | rs9273364 | 32626302 | 1.33e-16 | 7 | 0 | 0 | 0 | 0 | 7 | Both sexes | Both sexes | Both sexes | True | True |
| Gluc | 6 | HLA-DQA1 | protein_coding | rs9272426 | 32605189 | 1.55e-10 | 55 | 4 | 0 | 24 | 1 | 26 | Male only | Both sexes | Male only | True | False |
| Gluc | 6 | HLA-DQA1 | protein_coding | rs9272454 | 32605525 | 2.75e-10 | 191 | 77 | 13 | 3 | 3 | 95 | Male only | Both sexes | Male only | True | False |
| Gluc | 6 | XXbac-BPG254F23.7 | lincRNA | rs3957148 | 32682137 | 3.18e-09 | 189 | 0 | 0 | 0 | 17 | 172 | Both sexes | Both sexes | Both sexes | True | False |
| Gluc | 6 | XXbac-BPG254F23.7 | lincRNA | rs9275618 | 32684387 | 7.78e-09 | 17 | 0 | 0 | 2 | 0 | 15 | Both sexes | Both sexes | Female only | True | False |
| Gluc | 6 | BTNL2 | protein_coding | rs4959098 | 32383256 | 2.17e-08 | 9 | 4 | 1 | 0 | 0 | 4 | Male only | Male only | Male only | True | False |
| Gluc | 6 | HLA-DQB1 | protein_coding | 6:32623223 | 32623223 | 4.96e-08 | 127 | 47 | 0 | 35 | 13 | 32 | Male only | Both sexes | Male only | True | False |
| HDL.C | 16 | AC012181.1 | miRNA | rs3764261 | 56993324 | 3.71e-16 | 41 | 0 | 0 | 0 | 0 | 41 | Both sexes | Both sexes | Both sexes | False | True |
| HDL.C | 16 | CETP | protein_coding | rs1864163 | 56997233 | 5.65e-10 | 11 | 0 | 0 | 0 | 0 | 11 | Female only | Both sexes | Female only | False | True |
| HEIGHT | 8 | ELP3 | protein_coding | rs7821358 | 28060690 | 1.13e-08 | 41 | 21 | 0 | 0 | 3 | 17 | Male only | Male only | Male only | False | False |
| HORIZONTAL.AXIS..TERMINAL.40.MS | 18 | MAPK4 | protein_coding | rs1370483 | 48228968 | 4.06e-08 | 5 | 3 | 0 | 0 | 0 | 2 | Female only | Female only | Female only | False | False |
| IGRE | 9 | LRRC8A | protein_coding | rs34305034 | 131654547 | 2.22e-11 | 3 | 0 | 0 | 0 | 0 | 3 | Both sexes | Both sexes | Male only | False | True |
| IPF | 3 | RP11-197K6.1 | lincRNA | rs1851684 | 191379840 | 3.74e-08 | 101 | 51 | 0 | 0 | 0 | 50 | Female only | Female only | Female only | False | False |
| LDL.C | 19 | APOE | protein_coding | rs7412 | 45412079 | 1.05e-08 | 9 | 0 | 0 | 0 | 4 | 5 | Both sexes | Both sexes | Both sexes | False | True |
| LDL.C | 1 | USP24 | protein_coding | rs1165226 | 55608005 | 1.65e-08 | 49 | 23 | 2 | 2 | 16 | 6 | Female only | Female only | Female only | False | False |
| MCHC | 1 | OR10X1 | protein_coding | rs857693 | 158556301 | 2.88e-08 | 109 | 39 | 16 | 0 | 20 | 34 | Male only | Male only | Male only | False | True |
| MPV | 17 | TAOK1 | protein_coding | rs9896579 | 27786460 | 3.70e-08 | 115 | 0 | 50 | 8 | 0 | 57 | Female only | Both sexes | Female only | False | True |
| MonAbs | 4 | CCSER1 | protein_coding | rs56047630 | 92495671 | 1.68e-08 | 21 | 3 | 7 | 1 | 0 | 10 | Male only | Both sexes | Male only | False | False |

| pheno | CHR | Gene_Symbol | biotype | lead_snp | lead_BP | lead_P | TOTAL | NSIG | S05 | S01 | S001 | S0001 | independent_loci_in_sex_P1e6 | independent_loci_in_sex_P1e2 | independent_loci_in_sex_P5e8 | in_HLA | any_overlap |
|---|---|---|---|---|---|---|---|---|---|---|---|---|---|---|---|---|---|
| Nt.PTH | 10 | RP11-95I16.2 | lincRNA | rs11815121 | 122719834 | 1.28e-08 | 11 | 6 | 0 | 0 | 0 | 5 | Male only | Male only | Male only | False | False |
| PR_INTERVAL | 10 | FUT11 | protein_coding | rs78790589 | 75539598 | 1.63e-08 | 125 | 0 | 0 | 1 | 16 | 108 | Male only | Both sexes | Male only | False | False |
| Prolac | 4 | CTD-2012I17.1 | lincRNA | rs76674008 | 135769888 | 1.61e-08 | 5 | 3 | 0 | 0 | 0 | 2 | Male only | Male only | Male only | False | False |
| QRS.DURATION | 23 | HNRNPCP10 | pseudogene | X:143731017:A:G | 143731017 | 5.36e-09 | 19 | 10 | 0 | 0 | 0 | 9 | Female only | Female only | Female only | False | False |
| R.IP.IDEAL.BODY.WEIGHT | 8 | ELP3 | protein_coding | rs7821358 | 28060690 | 3.58e-08 | 41 | 21 | 0 | 0 | 3 | 17 | Male only | Male only | Male only | False | False |
| R.RT..FEMALE.ADULT..IDEAL.BODY.WEIGHT | 8 | ELP3 | protein_coding | rs7821358 | 28060690 | 3.47e-08 | 41 | 21 | 0 | 0 | 4 | 16 | Male only | Male only | Male only | False | False |
| R.RT..MALE.ADULT..IDEAL.BODY.WEIGHT | 8 | ELP3 | protein_coding | rs7821358 | 28060690 | 3.47e-08 | 41 | 21 | 0 | 0 | 4 | 16 | Male only | Male only | Male only | False | False |
| SGOT | 8 | RP11-127H5.1 | protein_coding | rs4734807 | 105705585 | 3.19e-08 | 39 | 20 | 0 | 0 | 1 | 18 | Female only | Female only | Female only | False | False |
| T3FRE | 3 | RP11-340E6.1 | lincRNA | rs73226608 | 141388147 | 4.02e-08 | 9 | 5 | 0 | 0 | 0 | 4 | Female only | Female only | Female only | False | False |
| TBil | 2 | UGT1A8 | protein_coding | rs4148325 | 234673309 | 4.52e-172 | 219 | 0 | 0 | 0 | 0 | 219 | Both sexes | Both sexes | Both sexes | False | True |
| TBil | 2 | UGT1A8 | protein_coding | rs2012734 | 234622429 | 3.90e-83 | 15 | 0 | 0 | 0 | 0 | 15 | Both sexes | Both sexes | Both sexes | False | True |
| TBil | 2 | UGT1A8 | protein_coding | rs7592624 | 234602906 | 1.98e-51 | 35 | 0 | 0 | 0 | 0 | 35 | Both sexes | Both sexes | Both sexes | False | True |
| TBil | 2 | UGT1A8 | protein_coding | rs6751673 | 234604903 | 1.39e-37 | 31 | 0 | 0 | 0 | 12 | 19 | Both sexes | Both sexes | Both sexes | False | True |
| TBil | 2 | MROH2A | protein_coding | rs4663336 | 234692932 | 5.88e-20 | 5 | 0 | 0 | 0 | 0 | 5 | Both sexes | Both sexes | Both sexes | False | True |
| TBil | 2 | UGT1A8 | protein_coding | rs1112310 | 234567916 | 2.54e-19 | 25 | 0 | 0 | 0 | 0 | 25 | Both sexes | Both sexes | Both sexes | False | True |
| TBil | 2 | UGT1A8 | protein_coding | rs12479240 | 234570829 | 5.05e-18 | 45 | 0 | 0 | 8 | 14 | 23 | Female only | Both sexes | Female only | False | True |
| TBil | 2 | UGT1A8 | protein_coding | rs35203651 | 234679549 | 1.83e-17 | 25 | 0 | 0 | 7 | 3 | 15 | Female only | Both sexes | Female only | False | True |
| TBil | 2 | MROH2A | protein_coding | rs1587493 | 234689751 | 1.01e-13 | 21 | 0 | 0 | 0 | 0 | 21 | Both sexes | Both sexes | Both sexes | False | True |
| TBil | 2 | UGT1A8 | protein_coding | rs17863795 | 234651369 | 2.64e-13 | 91 | 0 | 0 | 0 | 0 | 91 | Both sexes | Both sexes | Both sexes | False | True |
| TBil | 2 | ATG16L1 | protein_coding | rs62192926 | 234148737 | 3.29e-12 | 5 | 0 | 1 | 0 | 1 | 3 | Female only | Both sexes | Female only | False | True |
| TBil | 2 | USP40 | protein_coding | rs12472244 | 234466843 | 3.83e-12 | 13 | 0 | 1 | 6 | 0 | 6 | Female only | Both sexes | Female only | False | True |
| TBil | 2 | DGKD | protein_coding | rs838718 | 234296650 | 3.02e-11 | 27 | 9 | 5 | 0 | 3 | 10 | Female only | Female only | Female only | False | True |
| TBil | 2 | USP40 | protein_coding | rs72982316 | 234416719 | 1.07e-09 | 87 | 0 | 0 | 13 | 7 | 67 | Female only | Both sexes | Female only | False | True |

| pheno | CHR | Gene_Symbol | biotype | lead_snp | lead_BP | lead_P | TOTAL | NSIG | S05 | S01 | S001 | S0001 | independent_loci_in_sex_P1e6 | independent_loci_in_sex_P1e2 | independent_loci_in_sex_P5e8 | in_HLA | any_overlap |
|---|---|---|---|---|---|---|---|---|---|---|---|---|---|---|---|---|---|
| TBil | 2 | DGKD | protein_coding | rs838709 | 234284744 | 5.94e-09 | 7 | 0 | 2 | 2 | 0 | 3 | Female only | Both sexes | Female only | False | True |
| TBil | 2 | MROH2A | protein_coding | rs6716168 | 234713429 | 8.89e-09 | 13 | 0 | 6 | 6 | 0 | 1 | Female only | Both sexes | Female only | False | True |
| TBil | 2 | MROH2A | protein_coding | rs2041651 | 234713812 | 4.54e-08 | 3 | 0 | 0 | 2 | 0 | 1 | Female only | Both sexes | Female only | False | True |
| TRPI | 21 | RNU2-55P | snRNA | rs2199168 | 24661858 | 6.80e-09 | 11 | 6 | 0 | 0 | 0 | 5 | Male only | Male only | Male only | False | False |
| TSH | 9 | RP11-23B15.1 | lincRNA | rs10739496 | 100552559 | 1.13e-19 | 43 | 0 | 0 | 0 | 3 | 40 | Female only | Both sexes | Female only | False | True |
| TSH | 6 | HLA-DQA1 | protein_coding | rs9272426 | 32605189 | 1.99e-15 | 55 | 0 | 3 | 3 | 20 | 29 | Female only | Both sexes | Female only | True | False |
| TSH | 6 | HLA-DQA1 | protein_coding | rs3129758 | 32584625 | 1.04e-14 | 203 | 0 | 78 | 18 | 5 | 102 | Female only | Both sexes | Female only | True | False |
| TSH | 9 | FOXE1 | protein_coding | rs10759960 | 100608745 | 2.53e-12 | 13 | 0 | 0 | 3 | 3 | 7 | Female only | Both sexes | Female only | False | True |
| TSH | 6 | HLA-DRB9 | pseudogene | rs9268848 | 32429303 | 6.34e-11 | 31 | 0 | 15 | 1 | 1 | 14 | Female only | Both sexes | Female only | True | False |
| TSH | 9 | C9orf156 | protein_coding | rs6478471 | 100664382 | 2.57e-10 | 97 | 0 | 0 | 49 | 0 | 48 | Female only | Both sexes | Female only | False | True |
| TSH | 6 | MTCO3P1 | pseudogene | rs3135002 | 32668439 | 8.60e-09 | 111 | 50 | 5 | 1 | 0 | 55 | Female only | Both sexes | Female only | True | False |
| TSH | 9 | C9orf156 | protein_coding | rs7028759 | 100677883 | 1.50e-08 | 3 | 0 | 2 | 0 | 1 | 0 | Female only | Female only | Female only | False | True |
| TSH | 5 | PDE8B | protein_coding | rs2046045 | 76535811 | 1.84e-08 | 29 | 0 | 1 | 0 | 0 | 28 | Male only | Both sexes | Male only | False | True |
| T_WAVE | 3 | AC027119.1 | lincRNA | rs13326481 | 6016016 | 4.88e-08 | 125 | 63 | 0 | 0 | 15 | 47 | Female only | Female only | Female only | False | False |
| Trigs | 11 | ZNF259 | protein_coding | rs17120029 | 116650118 | 2.30e-21 | 87 | 0 | 0 | 0 | 0 | 87 | Both sexes | Both sexes | Both sexes | False | True |
| Trigs | 11 | ZNF259 | protein_coding | rs3741298 | 116657561 | 9.32e-21 | 3 | 0 | 0 | 0 | 0 | 3 | Both sexes | Both sexes | Both sexes | False | True |
| Trigs | 8 | AC100802.3 | antisense | rs115849089 | 19912370 | 1.01e-17 | 155 | 0 | 0 | 0 | 0 | 155 | Both sexes | Both sexes | Both sexes | False | True |
| Trigs | 11 | AP000770.1 | lincRNA | 11:116572089 | 116572089 | 5.10e-17 | 7 | 0 | 0 | 0 | 0 | 7 | Both sexes | Both sexes | Both sexes | False | True |
| Trigs | 11 | ZNF259 | protein_coding | rs6589566 | 116652423 | 5.58e-15 | 33 | 0 | 0 | 0 | 0 | 33 | Both sexes | Both sexes | Both sexes | False | True |
| Trigs | 11 | AP006216.10 | antisense | rs7118999 | 116645275 | 1.32e-14 | 15 | 0 | 0 | 1 | 1 | 13 | Both sexes | Both sexes | Both sexes | False | True |
| Trigs | 8 | LPL | protein_coding | rs13702 | 19824492 | 1.57e-13 | 157 | 0 | 0 | 0 | 0 | 157 | Both sexes | Both sexes | Both sexes | False | True |
| Trigs | 11 | APOA1 | protein_coding | rs525028 | 116705516 | 2.46e-13 | 97 | 0 | 1 | 1 | 0 | 95 | Both sexes | Both sexes | Both sexes | False | True |
| Trigs | 11 | APOC3 | protein_coding | rs5141 | 116702123 | 1.39e-11 | 111 | 0 | 0 | 0 | 2 | 109 | Both sexes | Both sexes | Both sexes | False | True |
| Trigs | 11 | AP000936.5 | pseudogene | rs12366015 | 116990851 | 1.99e-11 | 9 | 0 | 0 | 0 | 0 | 9 | Both sexes | Both sexes | Both sexes | False | True |
| Trigs | 8 | RP11-136O12.2 | lincRNA | rs2954038 | 126507389 | 2.70e-11 | 43 | 0 | 0 | 0 | 4 | 39 | Male only | Both sexes | Male only | False | True |
| Trigs | 8 | RP11-136O12.2 | lincRNA | rs2954021 | 126482077 | 5.15e-11 | 41 | 0 | 0 | 0 | 0 | 41 | Both sexes | Both sexes | Male only | False | True |

| pheno | CHR | Gene_Symbol | biotype | lead_snp | lead_BP | lead_P | TOTAL | NSIG | S05 | S01 | S001 | S0001 | independent_loci_in_sex_P1e6 | independent_loci_in_sex_P1e2 | independent_loci_in_sex_P5e8 | in_HLA | any_overlap |
|---|---|---|---|---|---|---|---|---|---|---|---|---|---|---|---|---|---|
| Trigs | 7 | BAZ1B | protein_coding | rs1178977 | 72857049 | 1.46e-10 | 161 | 0 | 0 | 2 | 30 | 129 | Female only | Both sexes | Female only | False | True |
| Trigs | 11 | BUD13 | protein_coding | rs180327 | 116623659 | 6.60e-10 | 17 | 4 | 4 | 1 | 2 | 6 | Both sexes | Both sexes | Female only | False | True |
| Trigs | 2 | GCKR | protein_coding | rs1260326 | 27730940 | 8.00e-10 | 33 | 0 | 0 | 0 | 7 | 26 | Both sexes | Both sexes | Both sexes | False | True |
| Trigs | 8 | LPL | protein_coding | rs256 | 19811967 | 1.96e-09 | 23 | 0 | 1 | 6 | 7 | 9 | Male only | Both sexes | Male only | False | True |
| Trigs | 11 | BUD13 | protein_coding | rs11216105 | 116577441 | 2.21e-09 | 9 | 3 | 1 | 4 | 0 | 1 | Both sexes | Both sexes | Both sexes | False | True |
| Trigs | 11 | SIK3 | protein_coding | rs2289893 | 116720089 | 2.28e-09 | 3 | 0 | 0 | 0 | 0 | 3 | Both sexes | Both sexes | Female only | False | False |
| Trigs | 19 | APOC1 | protein_coding | rs5117 | 45418790 | 3.00e-09 | 11 | 1 | 0 | 1 | 4 | 5 | Male only | Both sexes | Male only | False | True |
| Trigs | 11 | PAFAH1B2 | protein_coding | rs7925256 | 117045335 | 6.33e-09 | 63 | 0 | 0 | 0 | 1 | 62 | Both sexes | Both sexes | Female only | False | True |
| Trigs | 11 | AP000770.1 | lincRNA | rs480823 | 116525730 | 6.40e-09 | 43 | 0 | 20 | 0 | 2 | 21 | Female only | Both sexes | Female only | False | True |
| Trigs | 11 | SIK3 | protein_coding | rs72645459 | 116762157 | 7.21e-09 | 9 | 0 | 1 | 0 | 2 | 6 | Female only | Both sexes | Female only | False | True |
| Trigs | 11 | AP000770.1 | lincRNA | rs12803249 | 116536418 | 1.06e-08 | 63 | 18 | 5 | 19 | 6 | 15 | Female only | Both sexes | Female only | False | True |
| Trigs | 19 | APOE | protein_coding | rs72654473 | 45414399 | 2.04e-08 | 7 | 0 | 0 | 0 | 3 | 4 | Male only | Both sexes | Male only | False | True |
| URBC | 2 | RP11-335E8.1 | pseudogene | rs10496203 | 76494081 | 3.34e-08 | 15 | 8 | 0 | 0 | 0 | 7 | Female only | Female only | Female only | False | False |
| WBC | 17 | PSMD3 | protein_coding | rs3816472 | 38142677 | 3.57e-11 | 117 | 0 | 9 | 29 | 17 | 62 | Female only | Both sexes | Female only | False | True |
| WBC | 17 | MED24 | protein_coding | rs8066846 | 38203652 | 9.32e-11 | 19 | 0 | 5 | 5 | 0 | 9 | Female only | Both sexes | Female only | False | True |
| WBC | 17 | PSMD3 | protein_coding | rs2305481 | 38138624 | 3.69e-09 | 85 | 0 | 1 | 6 | 26 | 52 | Female only | Both sexes | Female only | False | True |
| WBC | 17 | GSDMA | protein_coding | rs4239225 | 38127112 | 3.13e-08 | 15 | 0 | 0 | 0 | 8 | 7 | Female only | Both sexes | Female only | False | True |
| X_percent_SAT | 6 | SLC17A3 | protein_coding | rs79920061 | 25857920 | 2.27e-08 | 23 | 0 | 0 | 1 | 7 | 15 | Male only | Both sexes | Male only | False | True |

#### STable10: Sex interaction loci for per-person medians of clinical quantitative traits in BioVU identified from post-hoc t test and replication status

660 rows × 37 columns

STable10 — Columns 1–23 of 37

| lab | SNP | CHR | BP | A1 | tstat | P | adjP | Gene_Symbol | biotype | full_name | group | genome_wide | CHROM | GENPOS | ALLELE0 | ALLELE1 | male_A1FREQ | male_INFO | male_N | male_TEST | male_BETA | male_SE |
|---|---|---|---|---|---|---|---|---|---|---|---|---|---|---|---|---|---|---|---|---|---|---|
| Ca | rs4929816 | 10 | 125842025 | G | 5.65 | 8.03e-09 | 0.002952 | CHST15 | protein_coding | Calcium serum/plasma serum/plasma | metabolic | Yes | 10 | 125842025 | A | G | 0.3519 | 1.005 | 26934 | ADD | 0.03029 | 0.007585 |
| Ca | rs4929815 | 10 | 125843462 | A | 5.646 | 8.22e-09 | 0.00302 | CHST15 | protein_coding | Calcium serum/plasma serum/plasma | metabolic | Yes | 10 | 125843462 | G | A | 0.347 | 0.9818 | 26934 | ADD | 0.03037 | 0.007696 |
| Ca | rs28459345 | 10 | 125843997 | A | 5.611 | 1.01e-08 | 0.003706 | CHST15 | protein_coding | Calcium serum/plasma serum/plasma | metabolic | Yes | 10 | 125843997 | C | A | 0.3472 | 0.977 | 26934 | ADD | 0.03095 | 0.007715 |
| Ca | rs28423034 | 10 | 125846716 | T | 5.463 | 2.35e-08 | 0.008624 | CHST15 | protein_coding | Calcium serum/plasma serum/plasma | metabolic | Yes | 10 | 125846716 | C | T | 0.3496 | 0.9791 | 26934 | ADD | 0.03005 | 0.007694 |
| Ca | rs4929819 | 10 | 125832832 | C | 5.461 | 2.37e-08 | 0.008694 | CHST15 | protein_coding | Calcium serum/plasma serum/plasma | metabolic | Yes | 10 | 125832832 | A | C | 0.3471 | 0.9835 | 26934 | ADD | 0.03003 | 0.007689 |
| Ca | rs28565055 | 10 | 125850658 | A | 5.411 | 3.13e-08 | 0.0115 | CHST15 | protein_coding | Calcium serum/plasma serum/plasma | metabolic | Yes | 10 | 125850658 | G | A | 0.3412 | 0.9759 | 26934 | ADD | 0.03219 | 0.007752 |
| Ca | rs28517851 | 10 | 125854055 | T | 5.375 | 3.82e-08 | 0.01406 | CHST15 | protein_coding | Calcium serum/plasma serum/plasma | metabolic | Yes | 10 | 125854055 | C | T | 0.3408 | 0.9766 | 26934 | ADD | 0.03166 | 0.007751 |
| Ca | rs7089310 | 10 | 125857244 | C | 5.354 | 4.31e-08 | 0.01586 | CHST15 | protein_coding | Calcium serum/plasma serum/plasma | metabolic | Yes | 10 | 125857244 | T | C | 0.3424 | 0.977 | 26934 | ADD | 0.03204 | 0.007741 |
| Ca | rs28587839 | 10 | 125849315 | T | 5.338 | 4.70e-08 | 0.01728 | CHST15 | protein_coding | Calcium serum/plasma serum/plasma | metabolic | Yes | 10 | 125849315 | C | T | 0.341 | 0.976 | 26934 | ADD | 0.03137 | 0.007753 |
| Ca | rs28655605 | 10 | 125856417 | A | 5.333 | 4.83e-08 | 0.01775 | CHST15 | protein_coding | Calcium serum/plasma serum/plasma | metabolic | Yes | 10 | 125856417 | G | A | 0.3423 | 0.9772 | 26934 | ADD | 0.03178 | 0.007741 |
| Ca | rs11870198 | 17 | 30043800 | T | -5.249 | 7.66e-08 | 0.02816 | RNU6-1134P | snRNA | Calcium serum/plasma serum/plasma | metabolic | No | 17 | 30043800 | G | T | 0.1201 | 0.9857 | 26934 | ADD | -0.04378 | 0.01125 |
| Ca | rs4256916 | 10 | 125855817 | G | 5.246 | 7.77e-08 | 0.02856 | CHST15 | protein_coding | Calcium serum/plasma serum/plasma | metabolic | No | 10 | 125855817 | A | G | 0.3428 | 0.9769 | 26934 | ADD | 0.03139 | 0.007739 |
| Ca | rs11870268 | 17 | 30041829 | C | -5.164 | 1.21e-07 | 0.04451 | RNU6-1134P | snRNA | Calcium serum/plasma serum/plasma | metabolic | No | 17 | 30041829 | A | C | 0.1202 | 0.9939 | 26934 | ADD | -0.04212 | 0.01121 |
| Ca | rs11870296 | 17 | 30041922 | G | -5.164 | 1.21e-07 | 0.04451 | RNU6-1134P | snRNA | Calcium serum/plasma serum/plasma | metabolic | No | 17 | 30041922 | A | G | 0.1202 | 0.9939 | 26934 | ADD | -0.04212 | 0.01121 |
| Ca | rs11868149 | 17 | 30042215 | A | -5.152 | 1.29e-07 | 0.04747 | RNU6-1134P | snRNA | Calcium serum/plasma serum/plasma | metabolic | No | 17 | 30042215 | G | A | 0.1202 | 0.9937 | 26934 | ADD | -0.04196 | 0.01121 |
| Ca | rs59852342 | 17 | 30040615 | A | -5.146 | 1.33e-07 | 0.04882 | RNU6-1134P | snRNA | Calcium serum/plasma serum/plasma | metabolic | No | 17 | 30040615 | G | A | 0.1202 | 0.9939 | 26934 | ADD | -0.04191 | 0.0112 |
| Ca | rs7210416 | 17 | 30040353 | A | -5.145 | 1.34e-07 | 0.04914 | RNU6-1134P | snRNA | Calcium serum/plasma serum/plasma | metabolic | No | 17 | 30040353 | G | A | 0.1202 | 0.9937 | 26934 | ADD | -0.04192 | 0.01121 |
| Ca | rs7211769 | 17 | 30040155 | G | -5.145 | 1.34e-07 | 0.04914 | RNU6-1134P | snRNA | Calcium serum/plasma serum/plasma | metabolic | No | 17 | 30040155 | A | G | 0.1202 | 0.9937 | 26934 | ADD | -0.04192 | 0.01121 |
| Ca | rs9898609 | 17 | 30037705 | C | -5.142 | 1.36e-07 | 0.04998 | RP11-805L22.1 | pseudogene | Calcium serum/plasma serum/plasma | metabolic | No | 17 | 30037705 | T | C | 0.1202 | 0.9936 | 26934 | ADD | -0.04198 | 0.01121 |
| Cl | rs7541552 | 1 | 142692818 | A | 5.549 | 1.44e-08 | 0.0053 | RP11-417J8.3 | lincRNA | Chloride [Moles/volume] in Serum or Plasma | metabolic | Yes | 1 | 142692818 | G | A | 0.2883 | 0.9771 | 26963 | ADD | 0.02079 | 0.008762 |
| Creat | rs79622676 | 20 | 22416573 | T | -5.466 | 2.31e-08 | 0.008681 | RP5-1004I9.1 | lincRNA | Creatinine [Mass/volume] in Blood Creatinine [Mass/volume] in Blood | kidney | Yes | 20 | 22416573 | C | T | 0.1 | 0.9978 | 27025 | ADD | -0.06033 | 0.01495 |
| EGFR | rs79622676 | 20 | 22416573 | T | 5.313 | 5.39e-08 | 0.02015 | RP5-1004I9.1 | lincRNA | eGFR | kidney | No | 20 | 22416573 | C | T | 0.1001 | 0.9977 | 27030 | ADD | 0.0539 | 0.01407 |
| Gluc | rs12542862 | 8 | 129245387 | T | -5.571 | 1.26e-08 | 0.004884 | RN7SKP226 | misc_RNA | Glucose lab | metabolic | Yes | 8 | 129245387 | G | T | 0.22 | 0.9991 | 26960 | ADD | -0.0377 | 0.009464 |
| Gluc | rs10492294 | 8 | 129246415 | C | -5.553 | 1.40e-08 | 0.005409 | RN7SKP226 | misc_RNA | Glucose lab | metabolic | Yes | 8 | 129246415 | T | C | 0.2211 | 1.004 | 26960 | ADD | -0.03747 | 0.009423 |
| Gluc | rs12541389 | 8 | 129243389 | T | -5.55 | 1.43e-08 | 0.005503 | RN7SKP226 | misc_RNA | Glucose lab | metabolic | Yes | 8 | 129243389 | C | T | 0.22 | 0.9993 | 26960 | ADD | -0.03757 | 0.009464 |
| Gluc | rs16902734 | 8 | 129225868 | T | -5.542 | 1.50e-08 | 0.005785 | RN7SKP226 | misc_RNA | Glucose lab | metabolic | Yes | 8 | 129225868 | C | T | 0.2193 | 0.9984 | 26960 | ADD | -0.03731 | 0.009481 |

| lab | SNP | CHR | BP | A1 | tstat | P | adjP | Gene_Symbol | biotype | full_name | group | genome_wide | CHROM | GENPOS | ALLELE0 | ALLELE1 | male_A1FREQ | male_INFO | male_N | male_TEST | male_BETA | male_SE |
|---|---|---|---|---|---|---|---|---|---|---|---|---|---|---|---|---|---|---|---|---|---|---|
| Gluc | rs56406478 | 19 | 55637503 | T | 5.444 | 2.61e-08 | 0.01007 | CTD-2587H24.1 | pseudogene | Glucose lab | metabolic | Yes | 19 | 55637503 | C | T | 0.08754 | 0.9703 | 26960 | ADD | 0.06342 | 0.01407 |
| Gluc | rs17383047 | 8 | 129227763 | A | -5.405 | 3.25e-08 | 0.01255 | RN7SKP226 | misc_RNA | Glucose lab | metabolic | Yes | 8 | 129227763 | G | A | 0.2175 | 0.9976 | 26960 | ADD | -0.03614 | 0.009512 |
| Height | rs12923049 | 16 | 9068302 | C | -5.482 | 2.11e-08 | 0.01038 | RP11-77H9.8 | lincRNA | Height | vitals | Yes | 16 | 9068302 | T | C | 0.4755 | 1.009 | 25698 | ADD | -0.03509 | 0.008087 |
| Height | rs2312305 | 19 | 35924757 | T | -5.481 | 2.11e-08 | 0.0104 | AC002511.3 | lincRNA | Height | vitals | Yes | 19 | 35924757 | C | T | 0.2717 | 0.9838 | 25698 | ADD | -0.03123 | 0.009198 |
| Height | rs2040601 | 19 | 35924880 | C | -5.453 | 2.48e-08 | 0.01222 | AC002511.3 | lincRNA | Height | vitals | Yes | 19 | 35924880 | T | C | 0.2717 | 0.9837 | 25698 | ADD | -0.03113 | 0.009198 |
| Height | rs4143784 | 15 | 53618226 | G | 5.407 | 3.21e-08 | 0.01583 | WDR72 | protein_coding | Height | vitals | Yes | 15 | 53618226 | T | G | 0.4909 | 0.9957 | 25698 | ADD | 0.02806 | 0.008137 |
| Height | rs112459152 | 23 | 29663635 | G | -5.4 | 3.33e-08 | 0.01641 | IL1RAPL1 | protein_coding | Height | vitals | Yes | 23 | 29663635 | A | G | 0.1009 | 1.952 | 25698 | ADD | -0.01847 | 0.009643 |
| Height | rs4829120 | 23 | 29652995 | A | -5.389 | 3.55e-08 | 0.01749 | IL1RAPL1 | protein_coding | Height | vitals | Yes | 23 | 29652995 | G | A | 0.0882 | 1.98 | 25698 | ADD | -0.02187 | 0.01017 |
| Height | rs1486562 | 15 | 53615578 | A | 5.366 | 4.03e-08 | 0.01987 | WDR72 | protein_coding | Height | vitals | Yes | 15 | 53615578 | C | A | 0.4916 | 0.9936 | 25698 | ADD | 0.02801 | 0.008146 |
| Height | rs111729097 | 23 | 29663304 | A | -5.364 | 4.07e-08 | 0.02005 | IL1RAPL1 | protein_coding | Height | vitals | Yes | 23 | 29663304 | G | A | 0.08813 | 1.973 | 25698 | ADD | -0.02165 | 0.01019 |
| Height | rs1486558 | 15 | 53620073 | C | 5.362 | 4.13e-08 | 0.02032 | WDR72 | protein_coding | Height | vitals | Yes | 15 | 53620073 | T | C | 0.4887 | 0.9957 | 25698 | ADD | 0.02681 | 0.008138 |
| Height | rs7579631 | 2 | 212986818 | T | -5.349 | 4.43e-08 | 0.02184 | ERBB4 | protein_coding | Height | vitals | Yes | 2 | 212986818 | C | T | 0.4147 | 0.995 | 25698 | ADD | -0.02041 | 0.0083 |
| Height | rs7050277 | 23 | 29668249 | T | -5.341 | 4.63e-08 | 0.02279 | IL1RAPL1 | protein_coding | Height | vitals | Yes | 23 | 29668249 | C | T | 0.0882 | 1.974 | 25698 | ADD | -0.02089 | 0.01019 |
| Height | rs11884023 | 2 | 171139256 | T | 5.34 | 4.66e-08 | 0.02295 | MYO3B | protein_coding | Height | vitals | Yes | 2 | 171139256 | C | T | 0.3323 | 0.9999 | 25698 | ADD | 0.02836 | 0.008658 |
| Height | rs4908192 | 1 | 102300223 | T | -5.337 | 4.72e-08 | 0.02325 | OLFM3 | protein_coding | Height | vitals | Yes | 1 | 102300223 | C | T | 0.4152 | 0.9856 | 25698 | ADD | -0.024 | 0.008307 |
| Height | rs112618048 | 23 | 29669698 | A | -5.331 | 4.89e-08 | 0.02408 | IL1RAPL1 | protein_coding | Height | vitals | Yes | 23 | 29669698 | G | A | 0.08841 | 1.974 | 25698 | ADD | -0.02128 | 0.01018 |
| Height | rs10782854 | 1 | 102299894 | T | -5.315 | 5.32e-08 | 0.02623 | OLFM3 | protein_coding | Height | vitals | No | 1 | 102299894 | C | T | 0.4152 | 0.9857 | 25698 | ADD | -0.02398 | 0.008306 |
| Height | rs11902372 | 2 | 212980961 | T | -5.302 | 5.74e-08 | 0.02826 | ERBB4 | protein_coding | Height | vitals | No | 2 | 212980961 | C | T | 0.4169 | 0.9944 | 25698 | ADD | -0.01822 | 0.008297 |
| Height | rs12469791 | 2 | 212984819 | A | -5.298 | 5.87e-08 | 0.0289 | ERBB4 | protein_coding | Height | vitals | No | 2 | 212984819 | G | A | 0.4118 | 0.9938 | 25698 | ADD | -0.01953 | 0.008314 |
| Height | rs11887531 | 2 | 212979871 | T | -5.287 | 6.21e-08 | 0.03058 | ERBB4 | protein_coding | Height | vitals | No | 2 | 212979871 | C | T | 0.4164 | 0.994 | 25698 | ADD | -0.01867 | 0.0083 |
| Height | rs2136530 | 9 | 10047844 | T | -5.287 | 6.23e-08 | 0.03071 | PTPRD | protein_coding | Height | vitals | No | 9 | 10047844 | C | T | 0.1335 | 1.004 | 25698 | ADD | -0.03716 | 0.01191 |
| Height | rs61055972 | 23 | 29664772 | G | -5.282 | 6.38e-08 | 0.03144 | IL1RAPL1 | protein_coding | Height | vitals | No | 23 | 29664772 | A | G | 0.08815 | 1.974 | 25698 | ADD | -0.0208 | 0.01019 |
| Height | rs148533541 | 23 | 29663857 | T | -5.276 | 6.59e-08 | 0.03245 | IL1RAPL1 | protein_coding | Height | vitals | No | 23 | 29663857 | C | T | 0.08817 | 1.974 | 25698 | ADD | -0.02088 | 0.01019 |
| Height | 12:71074730 | 12 | 71074730 | T | -5.276 | 6.62e-08 | 0.03261 | PTPRR | protein_coding | Height | vitals | No | 12 | 71074730 | G | T | 0.139 | 0.9389 | 25698 | ADD | -0.05054 | 0.01214 |
| Height | rs66522864 | 2 | 212966947 | C | -5.269 | 6.87e-08 | 0.03384 | ERBB4 | protein_coding | Height | vitals | No | 2 | 212966947 | T | C | 0.4211 | 0.988 | 25698 | ADD | -0.02055 | 0.008329 |
| Height | rs7056388 | 23 | 29664027 | G | -5.267 | 6.94e-08 | 0.03419 | IL1RAPL1 | protein_coding | Height | vitals | No | 23 | 29664027 | A | G | 0.08816 | 1.974 | 25698 | ADD | -0.02089 | 0.01019 |
| Height | rs148127450 | 23 | 29665304 | T | -5.266 | 6.99e-08 | 0.03444 | IL1RAPL1 | protein_coding | Height | vitals | No | 23 | 29665304 | C | T | 0.0881 | 1.974 | 25698 | ADD | -0.02072 | 0.01019 |
| Height | rs140009106 | 23 | 29663441 | T | -5.265 | 7.00e-08 | 0.0345 | IL1RAPL1 | protein_coding | Height | vitals | No | 23 | 29663441 | C | T | 0.08816 | 1.974 | 25698 | ADD | -0.02089 | 0.01019 |
| Height | rs3136645 | 19 | 39398649 | C | 5.265 | 7.03e-08 | 0.03463 | NFKBIB | protein_coding | Height | vitals | No | 19 | 39398649 | T | C | 0.1767 | 0.9897 | 25698 | ADD | 0.02764 | 0.01071 |

| lab | SNP | CHR | BP | A1 | tstat | P | adjP | Gene_Symbol | biotype | full_name | group | genome_wide | CHROM | GENPOS | ALLELE0 | ALLELE1 | male_A1FREQ | male_INFO | male_N | male_TEST | male_BETA | male_SE |
|---|---|---|---|---|---|---|---|---|---|---|---|---|---|---|---|---|---|---|---|---|---|---|
| Height | rs186524101 | 23 | 29666326 | T | -5.242 | 7.96e-08 | 0.0392 | IL1RAPL1 | protein_coding | Height | vitals | No | 23 | 29666326 | C | T | 0.08837 | 1.974 | 25698 | ADD | -0.02129 | 0.01018 |
| Height | rs10204324 | 2 | 212984466 | C | -5.24 | 8.02e-08 | 0.03953 | ERBB4 | protein_coding | Height | vitals | No | 2 | 212984466 | T | C | 0.4205 | 0.9972 | 25698 | ADD | -0.01805 | 0.008275 |
| Height | rs147175681 | 23 | 29652410 | A | -5.238 | 8.11e-08 | 0.03997 | IL1RAPL1 | protein_coding | Height | vitals | No | 23 | 29652410 | G | A | 0.08673 | 1.983 | 25698 | ADD | -0.02059 | 0.01024 |
| Height | rs141855038 | 23 | 29642306 | T | -5.237 | 8.16e-08 | 0.04018 | IL1RAPL1 | protein_coding | Height | vitals | No | 23 | 29642306 | G | T | 0.08691 | 1.978 | 25698 | ADD | -0.02149 | 0.01024 |
| Height | rs7051233 | 23 | 29647078 | A | -5.236 | 8.20e-08 | 0.04041 | IL1RAPL1 | protein_coding | Height | vitals | No | 23 | 29647078 | G | A | 0.08646 | 1.997 | 25698 | ADD | -0.01965 | 0.01022 |
| lab | SNP | CHR | BP | A1 | tstat | P | adjP | Gene_Symbol | biotype | full_name | group | genome_wide | CHROM | GENPOS | ALLELE0 | ALLELE1 | male_A1FREQ | male_INFO | male_N | male_TEST | male_BETA | male_SE |
| Height | rs57912808 | 23 | 29660672 | C | -5.235 | 8.23e-08 | 0.04055 | IL1RAPL1 | protein_coding | Height | vitals | No | 23 | 29660672 | A | C | 0.1009 | 1.95 | 25698 | ADD | -0.01929 | 0.009648 |
| Height | rs17003955 | 23 | 108868153 | A | -5.235 | 8.26e-08 | 0.0407 | KCNE1L | protein_coding | Height | vitals | No | 23 | 108868153 | G | A | 0.1571 | 1.981 | 25698 | ADD | -0.02006 | 0.007918 |
| Height | rs12327821 | 19 | 35926220 | C | -5.233 | 8.35e-08 | 0.04113 | AC002511.3 | lincRNA | Height | vitals | No | 19 | 35926220 | T | C | 0.2192 | 0.9977 | 25698 | ADD | -0.02755 | 0.009818 |
| Height | rs138348474 | 23 | 29662975 | T | -5.227 | 8.60e-08 | 0.04234 | IL1RAPL1 | protein_coding | Height | vitals | No | 23 | 29662975 | C | T | 0.08797 | 1.974 | 25698 | ADD | -0.02065 | 0.0102 |
| Height | rs4571009 | 2 | 212983838 | G | -5.217 | 9.10e-08 | 0.04482 | ERBB4 | protein_coding | Height | vitals | No | 2 | 212983838 | T | G | 0.421 | 0.9942 | 25698 | ADD | -0.0183 | 0.008286 |
| Height | rs9973287 | 19 | 35924213 | G | -5.211 | 9.42e-08 | 0.04639 | AC002511.3 | lincRNA | Height | vitals | No | 19 | 35924213 | A | G | 0.2752 | 0.987 | 25698 | ADD | -0.02873 | 0.009147 |
| Height | rs56242159 | 2 | 212991972 | C | -5.21 | 9.47e-08 | 0.04664 | ERBB4 | protein_coding | Height | vitals | No | 2 | 212991972 | A | C | 0.4251 | 0.995 | 25698 | ADD | -0.01728 | 0.008271 |
| Height | rs6719517 | 2 | 212979399 | T | -5.21 | 9.47e-08 | 0.04665 | ERBB4 | protein_coding | Height | vitals | No | 2 | 212979399 | C | T | 0.4178 | 0.9939 | 25698 | ADD | -0.01811 | 0.008296 |
| Height | rs10879176 | 12 | 71043417 | A | -5.209 | 9.49e-08 | 0.04676 | PTPRR | protein_coding | Height | vitals | No | 12 | 71043417 | G | A | 0.1322 | 0.9952 | 25698 | ADD | -0.04855 | 0.01205 |
| Height | rs7576561 | 2 | 212986457 | T | -5.205 | 9.68e-08 | 0.04768 | ERBB4 | protein_coding | Height | vitals | No | 2 | 212986457 | C | T | 0.4218 | 0.9973 | 25698 | ADD | -0.01798 | 0.00827 |
| Hgb | rs5971686 | 23 | 33138772 | T | 5.183 | 1.09e-07 | 0.04088 | DMD | protein_coding | Hemoglobin | blood | No | 23 | 33138772 | C | T | 0.2841 | 1.948 | 26968 | ADD | 0.02367 | 0.006578 |
| K | rs7588970 | 2 | 176490599 | G | -5.676 | 6.89e-09 | 0.002504 | AC096649.3 | lincRNA | Potassium blood | metabolic | Yes | 2 | 176490599 | A | G | 0.4334 | 1.003 | 26975 | ADD | -0.02753 | 0.007536 |
| K | rs9287987 | 2 | 176487261 | C | -5.641 | 8.45e-09 | 0.003068 | AC096649.3 | lincRNA | Potassium blood | metabolic | Yes | 2 | 176487261 | T | C | 0.4347 | 1.007 | 26975 | ADD | -0.02704 | 0.007519 |
| K | rs9287986 | 2 | 176486968 | T | -5.625 | 9.29e-09 | 0.003373 | AC096649.3 | lincRNA | Potassium blood | metabolic | Yes | 2 | 176486968 | C | T | 0.4338 | 1.004 | 26975 | ADD | -0.02701 | 0.007534 |
| K | rs4371333 | 2 | 176492608 | A | -5.623 | 9.39e-09 | 0.003411 | AC096649.3 | lincRNA | Potassium blood | metabolic | Yes | 2 | 176492608 | G | A | 0.4338 | 1.003 | 26975 | ADD | -0.02706 | 0.007536 |
| K | rs12693069 | 2 | 176489738 | C | -5.613 | 9.97e-09 | 0.003622 | AC096649.3 | lincRNA | Potassium blood | metabolic | Yes | 2 | 176489738 | T | C | 0.4338 | 1.003 | 26975 | ADD | -0.02705 | 0.007535 |
| K | rs12469683 | 2 | 176488640 | G | -5.604 | 1.05e-08 | 0.003806 | AC096649.3 | lincRNA | Potassium blood | metabolic | Yes | 2 | 176488640 | A | G | 0.4338 | 1.003 | 26975 | ADD | -0.02697 | 0.007536 |
| K | rs6717474 | 2 | 176497023 | T | -5.555 | 1.39e-08 | 0.005037 | AC096649.3 | lincRNA | Potassium blood | metabolic | Yes | 2 | 176497023 | C | T | 0.4372 | 1.002 | 26975 | ADD | -0.02522 | 0.007532 |
| MCH | rs1111716 | 10 | 91673756 | G | 5.186 | 1.08e-07 | 0.04225 | RP11-478K7.2 | lincRNA | MCH [Entitic mass] by Automated count | blood | No | 10 | 91673756 | A | G | 0.3529 | 1.011 | 26968 | ADD | 0.03656 | 0.008499 |
| MCHC | rs3813851 | 7 | 55752684 | A | 5.269 | 6.85e-08 | 0.02696 | FKBP9L | pseudogene | MCHC [Mass/volume] by Automated count | blood | No | 7 | 55752684 | G | A | 0.09215 | 0.9821 | 26945 | ADD | 0.05955 | 0.01347 |
| MCHC | rs79597331 | 7 | 55753510 | A | 5.255 | 7.39e-08 | 0.02909 | FKBP9L | pseudogene | MCHC [Mass/volume] by Automated count | blood | No | 7 | 55753510 | G | A | 0.09214 | 0.9816 | 26945 | ADD | 0.05941 | 0.01348 |
| MCHC | rs4074684 | 7 | 55750966 | T | 5.23 | 8.46e-08 | 0.03329 | FKBP9L | pseudogene | MCHC [Mass/volume] by Automated count | blood | No | 7 | 55750966 | C | T | 0.093 | 0.9915 | 26945 | ADD | 0.0568 | 0.01336 |
| MCHC | rs4397354 | 7 | 55778416 | C | 5.222 | 8.83e-08 | 0.03476 | FKBP9L | pseudogene | MCHC [Mass/volume] by Automated count | blood | No | 7 | 55778416 | T | C | 0.08234 | 0.9918 | 26945 | ADD | 0.05056 | 0.01411 |

| lab | SNP | CHR | BP | A1 | tstat | P | adjP | Gene_Symbol | biotype | full_name | group | genome_wide | CHROM | GENPOS | ALLELE0 | ALLELE1 | male_A1FREQ | male_INFO | male_N | male_TEST | male_BETA | male_SE |
|---|---|---|---|---|---|---|---|---|---|---|---|---|---|---|---|---|---|---|---|---|---|---|
| MCHC | rs11764978 | 7 | 55771924 | A | 5.176 | 1.14e-07 | 0.04468 | FKBP9L | pseudogene | MCHC [Mass/volume] by Automated count | blood | No | 7 | 55771924 | G | A | 0.08817 | 0.9929 | 26945 | ADD | 0.05464 | 0.01367 |
| NRBC. | rs800372 | 12 | 70026161 | T | -5.548 | 1.45e-08 | 0.004882 | BEST3 | protein_coding | NRBC # | blood | Yes | 12 | 70026161 | C | T | 0.4984 | 0.9991 | 23718 | ADD | -0.038 | 0.008739 |
| NRBC. | rs776203 | 12 | 70028389 | G | -5.516 | 1.74e-08 | 0.005867 | BEST3 | protein_coding | NRBC # | blood | Yes | 12 | 70028389 | A | G | 0.4969 | 1.001 | 23718 | ADD | -0.03778 | 0.008732 |
| NRBC. | rs776207 | 12 | 70025422 | G | 5.361 | 4.15e-08 | 0.01402 | BEST3 | protein_coding | NRBC # | blood | Yes | 12 | 70025422 | A | G | 0.472 | 0.9939 | 23718 | ADD | 0.03712 | 0.008778 |
| NRBC. | rs776424 | 12 | 70023791 | C | -5.317 | 5.28e-08 | 0.01782 | BEST3 | protein_coding | NRBC # | blood | No | 12 | 70023791 | A | C | 0.4931 | 0.9969 | 23718 | ADD | -0.03651 | 0.008751 |
| NRBC. | rs800371 | 12 | 70026760 | A | 5.309 | 5.50e-08 | 0.01858 | BEST3 | protein_coding | NRBC # | blood | No | 12 | 70026760 | G | A | 0.473 | 0.9966 | 23718 | ADD | 0.03687 | 0.008766 |
| NRBC. | rs710743 | 12 | 70028053 | A | 5.309 | 5.51e-08 | 0.01861 | BEST3 | protein_coding | NRBC # | blood | No | 12 | 70028053 | G | A | 0.4739 | 0.9954 | 23718 | ADD | 0.03666 | 0.008769 |
| NRBC. | rs776420 | 12 | 70023370 | A | -5.3 | 5.80e-08 | 0.01961 | BEST3 | protein_coding | NRBC # | blood | No | 12 | 70023370 | C | A | 0.4918 | 0.995 | 23718 | ADD | -0.03617 | 0.008759 |
| NRBC. | rs776421 | 12 | 70023386 | A | -5.29 | 6.12e-08 | 0.02067 | BEST3 | protein_coding | NRBC # | blood | No | 12 | 70023386 | G | A | 0.4918 | 0.9952 | 23718 | ADD | -0.03605 | 0.008759 |
| NRBC. | rs776417 | 12 | 70022135 | T | -5.191 | 1.05e-07 | 0.0354 | BEST3 | protein_coding | NRBC # | blood | No | 12 | 70022135 | C | T | 0.4952 | 0.9978 | 23718 | ADD | -0.03617 | 0.008746 |
| NRBC. | rs776414 | 12 | 70021405 | G | -5.168 | 1.18e-07 | 0.03998 | BEST3 | protein_coding | NRBC # | blood | No | 12 | 70021405 | A | G | 0.4957 | 1.002 | 23718 | ADD | -0.03568 | 0.008727 |
| NRBC. | rs776418 | 12 | 70022403 | A | -5.162 | 1.22e-07 | 0.04138 | BEST3 | protein_coding | NRBC # | blood | No | 12 | 70022403 | G | A | 0.4951 | 0.9974 | 23718 | ADD | -0.03612 | 0.008748 |
| NRBC. | rs776209 | 12 | 70024459 | C | 5.161 | 1.23e-07 | 0.0416 | BEST3 | protein_coding | NRBC # | blood | No | 12 | 70024459 | T | C | 0.478 | 0.9926 | 23718 | ADD | 0.03549 | 0.008778 |
| O2SAT | rs6436688 | 2 | 228259302 | G | -5.388 | 3.56e-08 | 0.01271 | TM4SF20 | protein_coding | Oxygen saturation calculated from oxygen partial pressure in arterial blood | vitals | Yes | 2 | 228259302 | A | G | 0.4493 | 1 | 23729 | ADD | -0.02455 | 0.008806 |
| O2SAT | rs6734145 | 2 | 228258697 | G | -5.382 | 3.69e-08 | 0.01317 | TM4SF20 | protein_coding | Oxygen saturation calculated from oxygen partial pressure in arterial blood | vitals | Yes | 2 | 228258697 | A | G | 0.4506 | 1.001 | 23729 | ADD | -0.02472 | 0.008802 |
| O2SAT | rs7588185 | 2 | 228260311 | A | -5.286 | 6.26e-08 | 0.02234 | TM4SF20 | protein_coding | Oxygen saturation calculated from oxygen partial pressure in arterial blood | vitals | No | 2 | 228260311 | C | A | 0.4564 | 1.002 | 23729 | ADD | -0.02397 | 0.008794 |
| O2SAT | rs11695593 | 2 | 228260413 | G | -5.286 | 6.26e-08 | 0.02235 | TM4SF20 | protein_coding | Oxygen saturation calculated from oxygen partial pressure in arterial blood | vitals | No | 2 | 228260413 | A | G | 0.4564 | 1.002 | 23729 | ADD | -0.02397 | 0.008794 |
| O2SAT | rs6742775 | 2 | 228253937 | C | -5.137 | 1.39e-07 | 0.04973 | TM4SF20 | protein_coding | Oxygen saturation calculated from oxygen partial pressure in arterial blood | vitals | No | 2 | 228253937 | T | C | 0.4557 | 0.9992 | 23729 | ADD | -0.02234 | 0.008806 |
| PCV | rs7458505 | 7 | 155154992 | A | 5.18 | 1.11e-07 | 0.04072 | BLACE | protein_coding | Hematocrit [Volume Fraction] of Blood by Automated count | blood | No | 7 | 155154992 | C | A | 0.1819 | 1.002 | 27330 | ADD | 0.0375 | 0.01044 |
| PCV | rs7384797 | 7 | 155161787 | T | 5.145 | 1.34e-07 | 0.04894 | BLACE | protein_coding | Hematocrit [Volume Fraction] of Blood by Automated count | blood | No | 7 | 155161787 | C | T | 0.1825 | 1.001 | 27330 | ADD | 0.03866 | 0.01043 |
| Plt.Ct | rs12283565 | 11 | 76346193 | T | 5.457 | 2.42e-08 | 0.009397 | RP11-672A2.5 | lincRNA | Platelet Count | blood | Yes | 11 | 76346193 | C | T | 0.08052 | 1.004 | 26981 | ADD | 0.05022 | 0.01554 |
| Plt.Ct | rs12281800 | 11 | 76345510 | T | 5.417 | 3.03e-08 | 0.01174 | RP11-672A2.5 | lincRNA | Platelet Count | blood | Yes | 11 | 76345510 | C | T | 0.07994 | 1.004 | 26981 | ADD | 0.04967 | 0.01559 |
| Plt.Ct | rs13377416 | 11 | 76346302 | T | 5.395 | 3.43e-08 | 0.01331 | RP11-672A2.5 | lincRNA | Platelet Count | blood | Yes | 11 | 76346302 | C | T | 0.07994 | 1.004 | 26981 | ADD | 0.0494 | 0.01559 |
| Plt.Ct | rs7929272 | 11 | 76347944 | G | 5.38 | 3.73e-08 | 0.01445 | RP11-672A2.5 | lincRNA | Platelet Count | blood | Yes | 11 | 76347944 | A | G | 0.08003 | 1.003 | 26981 | ADD | 0.04904 | 0.01559 |
| Plt.Ct | rs61893488 | 11 | 76349107 | A | 5.319 | 5.22e-08 | 0.02025 | RP11-672A2.5 | lincRNA | Platelet Count | blood | No | 11 | 76349107 | G | A | 0.07943 | 1.003 | 26981 | ADD | 0.04822 | 0.01564 |
| Plt.Ct | rs12280865 | 11 | 29294512 | G | -5.277 | 6.58e-08 | 0.0255 | CTD-2063L20.1 | pseudogene | Platelet Count | blood | No | 11 | 29294512 | T | G | 0.05958 | 0.9907 | 26981 | ADD | -0.0736 | 0.01798 |
| Plt.Ct | rs12273525 | 11 | 29299867 | T | -5.264 | 7.06e-08 | 0.02739 | AC110056.1 | miRNA | Platelet Count | blood | No | 11 | 29299867 | C | T | 0.05959 | 0.9929 | 26981 | ADD | -0.07352 | 0.01796 |

| lab | SNP | CHR | BP | A1 | tstat | P | adjP | Gene_Symbol | biotype | full_name | group | genome_wide | CHROM | GENPOS | ALLELE0 | ALLELE1 | male_A1FREQ | male_INFO | male_N | male_TEST | male_BETA | male_SE |
|---|---|---|---|---|---|---|---|---|---|---|---|---|---|---|---|---|---|---|---|---|---|---|
| Plt.Ct | rs12280421 | 11 | 29299730 | C | -5.263 | 7.10e-08 | 0.02754 | AC110056.1 | miRNA | Platelet Count | blood | No | 11 | 29299730 | T | C | 0.05959 | 0.9929 | 26981 | ADD | -0.07352 | 0.01796 |
| Plt.Ct | rs12275758 | 11 | 76349629 | A | 5.247 | 7.73e-08 | 0.02996 | RP11-672A2.5 | lincRNA | Platelet Count | blood | No | 11 | 76349629 | G | A | 0.07977 | 1.002 | 26981 | ADD | 0.04775 | 0.01562 |
| Plt.Ct | rs12275480 | 11 | 29295106 | T | -5.21 | 9.43e-08 | 0.03655 | CTD-2063L20.1 | pseudogene | Platelet Count | blood | No | 11 | 29295106 | C | T | 0.05923 | 0.9893 | 26981 | ADD | -0.07295 | 0.01802 |
| Plt.Ct | rs12275107 | 11 | 29300424 | T | -5.182 | 1.10e-07 | 0.04246 | AC110056.1 | miRNA | Platelet Count | blood | No | 11 | 29300424 | C | T | 0.05928 | 0.9917 | 26981 | ADD | -0.07263 | 0.018 |
| Plt.Ct | rs12273418 | 11 | 29299705 | T | -5.178 | 1.12e-07 | 0.04359 | AC110056.1 | miRNA | Platelet Count | blood | No | 11 | 29299705 | C | T | 0.05925 | 0.9916 | 26981 | ADD | -0.07259 | 0.018 |
| Plt.Ct | rs11030657 | 11 | 29302414 | A | -5.169 | 1.18e-07 | 0.04567 | AC110056.1 | miRNA | Platelet Count | blood | No | 11 | 29302414 | G | A | 0.05926 | 0.9918 | 26981 | ADD | -0.07229 | 0.018 |
| RDW | rs28409799 | 15 | 58701806 | A | 5.186 | 1.07e-07 | 0.04178 | ALDH1A2 | protein_coding | Erythrocyte distribution width [Ratio] by Automated count Erythrocyte distribution width [Ratio] by Automated count | blood | No | 15 | 58701806 | C | A | 0.05287 | 0.9983 | 26955 | ADD | 0.05586 | 0.01872 |
| RespRt | rs17300961 | 23 | 14573869 | G | -5.665 | 7.35e-09 | 0.002495 | GLRA2 | protein_coding | Respiratory rate | vitals | Yes | 23 | 14573869 | T | G | 0.08409 | 1.806 | 23979 | ADD | -0.04216 | 0.01078 |
| WBC | rs12919171 | 16 | 7034920 | G | 5.478 | 2.16e-08 | 0.008445 | RBFOX1 | protein_coding | Leukocytes [#/volume] in Blood by Automated count | immune/inflammatory | Yes | 16 | 7034920 | A | G | 0.4228 | 0.9739 | 26968 | ADD | 0.03007 | 0.007856 |
| WBC | rs4289011 | 16 | 7034268 | C | 5.432 | 2.78e-08 | 0.0109 | RBFOX1 | protein_coding | Leukocytes [#/volume] in Blood by Automated count | immune/inflammatory | Yes | 16 | 7034268 | T | C | 0.4132 | 1.004 | 26968 | ADD | 0.02964 | 0.007761 |
| lab | SNP | CHR | BP | A1 | tstat | P | adjP | Gene_Symbol | biotype | full_name | group | genome_wide | CHROM | GENPOS | ALLELE0 | ALLELE1 | male_A1FREQ | male_INFO | male_N | male_TEST | male_BETA | male_SE |
| Weight | rs11233373 | 11 | 71083737 | T | 5.505 | 1.85e-08 | 0.007815 | AP002387.1 | pseudogene | Weight | vitals | Yes | 11 | 71083737 | C | T | 0.05051 | 1.001 | 26877 | ADD | 0.06258 | 0.0193 |
| Weight | rs11233358 | 11 | 71079402 | T | 5.496 | 1.94e-08 | 0.008231 | AP002387.1 | pseudogene | Weight | vitals | Yes | 11 | 71079402 | C | T | 0.05052 | 1.005 | 26877 | ADD | 0.06201 | 0.01926 |
| Weight | rs41515245 | 11 | 71100917 | A | 5.48 | 2.13e-08 | 0.009027 | AP002387.1 | pseudogene | Weight | vitals | Yes | 11 | 71100917 | G | A | 0.05045 | 1.005 | 26877 | ADD | 0.06142 | 0.01927 |
| Weight | rs144002293 | 11 | 71099165 | T | 5.433 | 2.77e-08 | 0.01175 | AP002387.1 | pseudogene | Weight | vitals | Yes | 11 | 71099165 | C | T | 0.05012 | 1.002 | 26877 | ADD | 0.06067 | 0.01936 |
| Weight | rs2421139 | 10 | 124796215 | A | 5.424 | 2.92e-08 | 0.01235 | ACADSB | protein_coding | Weight | vitals | Yes | 10 | 124796215 | G | A | 0.4491 | 1.004 | 26877 | ADD | 0.03601 | 0.008492 |
| Weight | rs9423323 | 10 | 124796626 | A | 5.404 | 3.25e-08 | 0.01377 | ACADSB | protein_coding | Weight | vitals | Yes | 10 | 124796626 | G | A | 0.45 | 1.001 | 26877 | ADD | 0.03585 | 0.008507 |
| Weight | rs9423255 | 10 | 124796483 | G | 5.387 | 3.57e-08 | 0.01513 | ACADSB | protein_coding | Weight | vitals | Yes | 10 | 124796483 | A | G | 0.4502 | 1.001 | 26877 | ADD | 0.03561 | 0.008507 |
| Weight | rs6599645 | 10 | 124799259 | C | 5.384 | 3.64e-08 | 0.01542 | ACADSB | protein_coding | Weight | vitals | Yes | 10 | 124799259 | T | C | 0.4476 | 0.9992 | 26877 | ADD | 0.03584 | 0.008517 |
| Weight | rs58298943 | 12 | 57391292 | T | 5.327 | 4.99e-08 | 0.02113 | HBCBP | protein_coding | Weight | vitals | Yes | 12 | 57391292 | C | T | 0.08428 | 1.002 | 26877 | ADD | 0.059 | 0.01522 |
| Weight | rs11073644 | 15 | 87095072 | T | 5.304 | 5.67e-08 | 0.02402 | AGBL1 | protein_coding | Weight | vitals | No | 15 | 87095072 | C | T | 0.4938 | 0.9884 | 26877 | ADD | 0.02765 | 0.008529 |
| Weight | rs3803018 | 12 | 57394123 | A | 5.292 | 6.06e-08 | 0.02566 | ZBTB39 | protein_coding | Weight | vitals | No | 12 | 57394123 | C | A | 0.08388 | 0.9991 | 26877 | ADD | 0.05943 | 0.01527 |
| Weight | rs28784669 | 15 | 87094519 | T | 5.276 | 6.59e-08 | 0.02792 | AGBL1 | protein_coding | Weight | vitals | No | 15 | 87094519 | C | T | 0.4936 | 0.988 | 26877 | ADD | 0.02741 | 0.008531 |
| Weight | rs2371717 | 12 | 57394654 | A | 5.248 | 7.69e-08 | 0.03255 | ZBTB39 | protein_coding | Weight | vitals | No | 12 | 57394654 | G | A | 0.08401 | 0.9995 | 26877 | ADD | 0.05879 | 0.01526 |
| Weight | rs4016338 | 12 | 57399015 | C | 5.243 | 7.92e-08 | 0.03353 | ZBTB39 | protein_coding | Weight | vitals | No | 12 | 57399015 | T | C | 0.08413 | 0.9996 | 26877 | ADD | 0.05891 | 0.01525 |
| Weight | rs35493121 | 12 | 57390038 | C | 5.222 | 8.85e-08 | 0.03748 | GPR182 | protein_coding | Weight | vitals | No | 12 | 57390038 | T | C | 0.08486 | 1.004 | 26877 | ADD | 0.05772 | 0.01515 |
| HR_INTERVAL | rs76628467 | 11 | 84889124 | G | -5.67 | 7.15e-09 | 0.002399 | DLG2 | protein_coding | Heart rate | vitals | Yes | 11 | 84889124 | A | G | 0.1251 | 0.965 | 19279 | ADD | -0.04653 | 0.01303 |

| lab | SNP | CHR | BP | A1 | tstat | P | adjP | Gene_Symbol | biotype | full_name | group | genome_wide | CHROM | GENPOS | ALLELE0 | ALLELE1 | male_A1FREQ | male_INFO | male_N | male_TEST | male_BETA | male_SE |
|---|---|---|---|---|---|---|---|---|---|---|---|---|---|---|---|---|---|---|---|---|---|---|
| HR_INTERVAL | rs28516854 | 15 | 56833144 | T | -5.56 | 1.35e-08 | 0.004522 | RP11-1129I3.1 | sense_overlapping | Heart rate | vitals | Yes | 15 | 56833144 | C | T | 0.1512 | 0.9875 | 19279 | ADD | -0.04153 | 0.0119 |
| HR_INTERVAL | rs6581776 | 12 | 68198021 | T | 5.307 | 5.57e-08 | 0.01867 | RP11-43N5.1 | lincRNA | Heart rate | vitals | No | 12 | 68198021 | C | T | 0.05657 | 1.011 | 19279 | ADD | 0.0776 | 0.01826 |
| HR_INTERVAL | rs7953226 | 12 | 68184633 | T | 5.256 | 7.38e-08 | 0.02475 | RP11-43N5.1 | lincRNA | Heart rate | vitals | No | 12 | 68184633 | G | T | 0.06238 | 0.9155 | 19279 | ADD | 0.07269 | 0.01832 |
| HR_INTERVAL | rs147278931 | 11 | 85306003 | A | -5.255 | 7.42e-08 | 0.0249 | DLG2 | protein_coding | Heart rate | vitals | No | 11 | 85306003 | G | A | 0.1098 | 0.9739 | 19279 | ADD | -0.05475 | 0.01373 |
| HR_INTERVAL | rs79636458 | 11 | 85162767 | G | -5.225 | 8.72e-08 | 0.02926 | DLG2 | protein_coding | Heart rate | vitals | No | 11 | 85162767 | A | G | 0.1133 | 0.9745 | 19279 | ADD | -0.05443 | 0.01353 |
| HR_INTERVAL | rs13329689 | 15 | 56840763 | A | -5.198 | 1.01e-07 | 0.03379 | RP11-1129I3.1 | sense_overlapping | Heart rate | vitals | No | 15 | 56840763 | C | A | 0.1468 | 0.9903 | 19279 | ADD | -0.0387 | 0.01203 |
| HR_INTERVAL | rs9325186 | 12 | 68202363 | A | 5.181 | 1.10e-07 | 0.03696 | RP11-43N5.1 | lincRNA | Heart rate | vitals | No | 12 | 68202363 | G | A | 0.05517 | 0.9449 | 19279 | ADD | 0.08598 | 0.0191 |
| HR_INTERVAL | rs3861888 | 12 | 68199770 | T | 5.177 | 1.13e-07 | 0.03775 | RP11-43N5.1 | lincRNA | Heart rate | vitals | No | 12 | 68199770 | C | T | 0.05517 | 0.9453 | 19279 | ADD | 0.08554 | 0.0191 |
| HR_INTERVAL | rs9920264 | 15 | 56841638 | T | -5.174 | 1.15e-07 | 0.03849 | RP11-1129I3.1 | sense_overlapping | Heart rate | vitals | No | 15 | 56841638 | G | T | 0.1513 | 0.9909 | 19279 | ADD | -0.03959 | 0.01188 |
| HR_INTERVAL | rs7955560 | 12 | 68199174 | G | 5.127 | 1.47e-07 | 0.04944 | RP11-43N5.1 | lincRNA | Heart rate | vitals | No | 12 | 68199174 | A | G | 0.05203 | 1.013 | 19279 | ADD | 0.08426 | 0.01896 |
| MEAN_QRS | rs2784755 | 10 | 36063725 | T | -5.324 | 5.09e-08 | 0.01866 | RPL7P37 | pseudogene | QRS axis | cardiovascular | No | 10 | 36063725 | C | T | 0.1577 | 0.9863 | 19258 | ADD | -0.08279 | 0.01457 |
| MEAN_QRS | rs2845920 | 10 | 36061143 | A | -5.289 | 6.16e-08 | 0.0226 | RPL7P37 | pseudogene | QRS axis | cardiovascular | No | 10 | 36061143 | G | A | 0.1636 | 0.9994 | 19258 | ADD | -0.08228 | 0.01426 |
| MEAN_QRS | rs2804817 | 10 | 36061337 | A | -5.284 | 6.32e-08 | 0.02318 | RPL7P37 | pseudogene | QRS axis | cardiovascular | No | 10 | 36061337 | C | A | 0.1627 | 0.992 | 19258 | ADD | -0.08307 | 0.01435 |
| MEAN_QRS | rs2804814 | 10 | 36071555 | A | -5.262 | 7.14e-08 | 0.02617 | RPL7P37 | pseudogene | QRS axis | cardiovascular | No | 10 | 36071555 | G | A | 0.158 | 0.9833 | 19258 | ADD | -0.08138 | 0.01459 |
| MEAN_QRS | rs12232965 | 2 | 33954560 | T | -5.179 | 1.12e-07 | 0.0409 | AC009499.1 | lincRNA | QRS axis | cardiovascular | No | 2 | 33954560 | C | T | 0.4764 | 1.001 | 19258 | ADD | -0.0322 | 0.01055 |
| PR_INTERVAL | rs12792987 | 11 | 15131899 | T | -5.403 | 3.28e-08 | 0.01183 | INSC | protein_coding | PR Interval mean | cardiovascular | Yes | 11 | 15131899 | C | T | 0.06719 | 0.98 | 19278 | ADD | -0.04833 | 0.02032 |
| PR_INTERVAL | rs11826529 | 11 | 15133198 | C | -5.175 | 1.14e-07 | 0.04127 | INSC | protein_coding | PR Interval mean | cardiovascular | No | 11 | 15133198 | T | C | 0.07226 | 0.9853 | 19278 | ADD | -0.04755 | 0.0196 |
| QT_INTERVAL_C | rs111823477 | 22 | 32659884 | T | 5.341 | 4.61e-08 | 0.01638 | RP1-90G24.10 | antisense | QT interval corrected | cardiovascular | Yes | 22 | 32659884 | C | T | 0.0863 | 0.9934 | 19274 | ADD | 0.0672 | 0.01695 |
| QT_INTERVAL_C | rs7285242 | 22 | 32663153 | C | 5.323 | 5.11e-08 | 0.01816 | RP1-90G24.10 | antisense | QT interval corrected | cardiovascular | No | 22 | 32663153 | T | C | 0.08661 | 0.9919 | 19274 | ADD | 0.06965 | 0.01697 |
| QT_INTERVAL_C | rs56240204 | 22 | 32662384 | C | 5.258 | 7.27e-08 | 0.02581 | RP1-90G24.10 | antisense | QT interval corrected | cardiovascular | No | 22 | 32662384 | A | C | 0.08695 | 0.9956 | 19274 | ADD | 0.06899 | 0.0169 |
| QT_INTERVAL_C | rs67817759 | 22 | 32660412 | A | 5.257 | 7.33e-08 | 0.02604 | RP1-90G24.10 | antisense | QT interval corrected | cardiovascular | No | 22 | 32660412 | G | A | 0.08579 | 0.994 | 19274 | ADD | 0.06689 | 0.017 |

| lab | SNP | CHR | BP | A1 | tstat | P | adjP | Gene_Symbol | biotype | full_name | group | genome_wide | CHROM | GENPOS | ALLELE0 | ALLELE1 | male_A1FREQ | male_INFO | male_N | male_TEST | male_BETA | male_SE |
|---|---|---|---|---|---|---|---|---|---|---|---|---|---|---|---|---|---|---|---|---|---|---|
| QT_INTERVAL_C | rs5998359 | 22 | 32658620 | G | 5.224 | 8.75e-08 | 0.03108 | RP1-90G24.10 | antisense | QT interval corrected | cardiovascular | No | 22 | 32658620 | A | G | 0.08761 | 0.9971 | 19274 | ADD | 0.06565 | 0.0168 |
| QT_INTERVAL_C | rs34754823 | 5 | 133318470 | G | 5.195 | 1.02e-07 | 0.03636 | VDAC1 | protein_coding | QT interval corrected | cardiovascular | No | 5 | 133318470 | A | G | 0.1198 | 0.9868 | 19274 | ADD | 0.05132 | 0.01469 |
| QT_INTERVAL_C | rs71587512 | 5 | 133320602 | A | 5.187 | 1.07e-07 | 0.03801 | VDAC1 | protein_coding | QT interval corrected | cardiovascular | No | 5 | 133320602 | G | A | 0.1197 | 0.9873 | 19274 | ADD | 0.05083 | 0.0147 |
| QT_INTERVAL_C | rs35644303 | 5 | 133331167 | C | 5.185 | 1.08e-07 | 0.03847 | VDAC1 | protein_coding | QT interval corrected | cardiovascular | No | 5 | 133331167 | T | C | 0.1203 | 0.9902 | 19274 | ADD | 0.04914 | 0.01464 |
| QT_INTERVAL_C | rs13162964 | 5 | 133325014 | T | 5.175 | 1.14e-07 | 0.04043 | VDAC1 | protein_coding | QT interval corrected | cardiovascular | No | 5 | 133325014 | C | T | 0.1197 | 0.9871 | 19274 | ADD | 0.04936 | 0.01469 |
| QT_INTERVAL_C | rs6893145 | 5 | 133328825 | C | 5.173 | 1.15e-07 | 0.04086 | VDAC1 | protein_coding | QT interval corrected | cardiovascular | No | 5 | 133328825 | T | C | 0.1202 | 0.9907 | 19274 | ADD | 0.04887 | 0.01464 |
| QT_INTERVAL_C | rs6897409 | 5 | 133328998 | C | 5.172 | 1.16e-07 | 0.04105 | VDAC1 | protein_coding | QT interval corrected | cardiovascular | No | 5 | 133328998 | T | C | 0.1202 | 0.9907 | 19274 | ADD | 0.04887 | 0.01464 |
| QT_INTERVAL_C | rs34133155 | 5 | 133338671 | G | 5.168 | 1.19e-07 | 0.04213 | VDAC1 | protein_coding | QT interval corrected | cardiovascular | No | 5 | 133338671 | T | G | 0.1207 | 0.9892 | 19274 | ADD | 0.05047 | 0.01462 |
| QT_INTERVAL_C | rs56326224 | 5 | 133337503 | G | 5.162 | 1.23e-07 | 0.04352 | VDAC1 | protein_coding | QT interval corrected | cardiovascular | No | 5 | 133337503 | A | G | 0.1207 | 0.9894 | 19274 | ADD | 0.05048 | 0.01462 |
| QT_INTERVAL_C | rs71587514 | 5 | 133338394 | A | 5.161 | 1.23e-07 | 0.04367 | VDAC1 | protein_coding | QT interval corrected | cardiovascular | No | 5 | 133338394 | G | A | 0.1207 | 0.9893 | 19274 | ADD | 0.05046 | 0.01462 |
| QT_INTERVAL_C | rs6880980 | 5 | 133326423 | G | 5.16 | 1.24e-07 | 0.04396 | VDAC1 | protein_coding | QT interval corrected | cardiovascular | No | 5 | 133326423 | T | G | 0.1201 | 0.9878 | 19274 | ADD | 0.04931 | 0.01467 |
| QT_INTERVAL_C | rs3733936 | 5 | 133340212 | T | 5.154 | 1.27e-07 | 0.04522 | VDAC1 | protein_coding | QT interval corrected | cardiovascular | No | 5 | 133340212 | C | T | 0.1196 | 0.9903 | 19274 | ADD | 0.04999 | 0.01468 |
| RATE_C | rs79346577 | 11 | 84989662 | T | -6.203 | 2.77e-10 | 9.79e-05 | DLG2 | protein_coding | Heart rate | vitals | Yes | 11 | 84989662 | C | T | 0.179 | 0.9785 | 19277 | ADD | -0.06863 | 0.01237 |
| RATE_C | rs75251022 | 11 | 85093218 | T | -6.085 | 5.83e-10 | 2.06e-04 | DLG2 | protein_coding | Heart rate | vitals | Yes | 11 | 85093218 | C | T | 0.154 | 0.9779 | 19277 | ADD | -0.06494 | 0.01314 |
| RATE_C | rs79636458 | 11 | 85162767 | G | -6.055 | 7.02e-10 | 2.48e-04 | DLG2 | protein_coding | Heart rate | vitals | Yes | 11 | 85162767 | A | G | 0.1133 | 0.9745 | 19277 | ADD | -0.07513 | 0.01499 |
| RATE_C | rs74851225 | 11 | 84655133 | A | -6.054 | 7.05e-10 | 2.49e-04 | DLG2 | protein_coding | Heart rate | vitals | Yes | 11 | 84655133 | G | A | 0.09954 | 0.9894 | 19277 | ADD | -0.08089 | 0.01575 |
| RATE_C | rs77106986 | 11 | 84655132 | C | -6.051 | 7.20e-10 | 2.55e-04 | DLG2 | protein_coding | Heart rate | vitals | Yes | 11 | 84655132 | T | C | 0.1024 | 0.9923 | 19277 | ADD | -0.07873 | 0.01553 |
| RATE_C | rs144961007 | 11 | 85316953 | T | -6.034 | 8.00e-10 | 2.83e-04 | DLG2 | protein_coding | Heart rate | vitals | Yes | 11 | 85316953 | C | T | 0.1059 | 0.973 | 19277 | ADD | -0.07745 | 0.01545 |
| RATE_C | rs147278931 | 11 | 85306003 | A | -5.984 | 1.09e-09 | 3.84e-04 | DLG2 | protein_coding | Heart rate | vitals | Yes | 11 | 85306003 | G | A | 0.1099 | 0.9739 | 19277 | ADD | -0.07542 | 0.0152 |
| RATE_C | rs78212243 | 11 | 84653467 | A | -5.963 | 1.24e-09 | 4.38e-04 | DLG2 | protein_coding | Heart rate | vitals | Yes | 11 | 84653467 | G | A | 0.1027 | 0.9924 | 19277 | ADD | -0.07764 | 0.0155 |
| RATE_C | rs77432810 | 11 | 84653997 | T | -5.956 | 1.29e-09 | 4.57e-04 | DLG2 | protein_coding | Heart rate | vitals | Yes | 11 | 84653997 | C | T | 0.1028 | 0.9924 | 19277 | ADD | -0.07806 | 0.0155 |
| RATE_C | rs74579964 | 11 | 84649222 | T | -5.953 | 1.31e-09 | 4.65e-04 | DLG2 | protein_coding | Heart rate | vitals | Yes | 11 | 84649222 | C | T | 0.1027 | 0.9923 | 19277 | ADD | -0.07697 | 0.01551 |
| RATE_C | rs77260179 | 11 | 84650401 | C | -5.938 | 1.44e-09 | 5.10e-04 | DLG2 | protein_coding | Heart rate | vitals | Yes | 11 | 84650401 | T | C | 0.1029 | 0.9919 | 19277 | ADD | -0.078 | 0.0155 |

| lab | SNP | CHR | BP | A1 | tstat | P | adjP | Gene_Symbol | biotype | full_name | group | genome_wide | CHROM | GENPOS | ALLELE0 | ALLELE1 | male_A1FREQ | male_INFO | male_N | male_TEST | male_BETA | male_SE |
|---|---|---|---|---|---|---|---|---|---|---|---|---|---|---|---|---|---|---|---|---|---|---|
| RATE_C | rs80063939 | 11 | 84654438 | G | -5.937 | 1.45e-09 | 5.13e-04 | DLG2 | protein_coding | Heart rate | vitals | Yes | 11 | 84654438 | A | G | 0.1029 | 0.9925 | 19277 | ADD | -0.07826 | 0.01549 |
| RATE_C | rs74958871 | 11 | 85144339 | A | -5.93 | 1.51e-09 | 5.35e-04 | DLG2 | protein_coding | Heart rate | vitals | Yes | 11 | 85144339 | G | A | 0.1119 | 0.9756 | 19277 | ADD | -0.07295 | 0.01506 |
| RATE_C | rs79772854 | 11 | 84652949 | C | -5.925 | 1.56e-09 | 5.51e-04 | DLG2 | protein_coding | Heart rate | vitals | Yes | 11 | 84652949 | T | C | 0.1029 | 0.992 | 19277 | ADD | -0.07723 | 0.0155 |
| RATE_C | rs118157676 | 11 | 85244366 | T | -5.925 | 1.56e-09 | 5.54e-04 | DLG2 | protein_coding | Heart rate | vitals | Yes | 11 | 85244366 | C | T | 0.11 | 0.9766 | 19277 | ADD | -0.07578 | 0.01517 |
| RATE_C | rs74730384 | 11 | 84654836 | C | -5.924 | 1.57e-09 | 5.55e-04 | DLG2 | protein_coding | Heart rate | vitals | Yes | 11 | 84654836 | T | C | 0.103 | 0.9924 | 19277 | ADD | -0.0782 | 0.01549 |
| RATE_C | rs56030411 | 11 | 84655283 | T | -5.914 | 1.67e-09 | 5.92e-04 | DLG2 | protein_coding | Heart rate | vitals | Yes | 11 | 84655283 | C | T | 0.103 | 0.9921 | 19277 | ADD | -0.07809 | 0.01549 |
| RATE_C | rs111675347 | 11 | 85128307 | C | -5.885 | 1.99e-09 | 7.04e-04 | DLG2 | protein_coding | Heart rate | vitals | Yes | 11 | 85128307 | T | C | 0.1675 | 0.9794 | 19277 | ADD | -0.06085 | 0.01269 |
| RATE_C | rs77719342 | 11 | 85237927 | C | -5.865 | 2.24e-09 | 7.93e-04 | DLG2 | protein_coding | Heart rate | vitals | Yes | 11 | 85237927 | A | C | 0.1068 | 0.9846 | 19277 | ADD | -0.07269 | 0.01532 |
| RATE_C | rs76628467 | 11 | 84889124 | G | -5.859 | 2.33e-09 | 8.26e-04 | DLG2 | protein_coding | Heart rate | vitals | Yes | 11 | 84889124 | A | G | 0.1251 | 0.965 | 19277 | ADD | -0.06005 | 0.01443 |
| RATE_C | rs146843948 | 11 | 84717140 | A | -5.819 | 2.97e-09 | 0.00105 | DLG2 | protein_coding | Heart rate | vitals | Yes | 11 | 84717140 | C | A | 0.1038 | 0.9893 | 19277 | ADD | -0.06976 | 0.01546 |
| RATE_C | rs80023956 | 11 | 85190497 | G | -5.698 | 6.06e-09 | 0.002143 | DLG2 | protein_coding | Heart rate | vitals | Yes | 11 | 85190497 | A | G | 0.113 | 0.9749 | 19277 | ADD | -0.07043 | 0.015 |
| RATE_C | rs61390712 | 11 | 84698622 | T | -5.679 | 6.77e-09 | 0.002396 | DLG2 | protein_coding | Heart rate | vitals | Yes | 11 | 84698622 | C | T | 0.1027 | 0.9919 | 19277 | ADD | -0.07066 | 0.01551 |
| RATE_C | rs60540551 | 11 | 84745255 | A | -5.665 | 7.35e-09 | 0.002603 | DLG2 | protein_coding | Heart rate | vitals | Yes | 11 | 84745255 | G | A | 0.1023 | 0.9883 | 19277 | ADD | -0.06872 | 0.01556 |
| RATE_C | rs150106533 | 11 | 85215621 | C | -5.655 | 7.81e-09 | 0.002765 | DLG2 | protein_coding | Heart rate | vitals | Yes | 11 | 85215621 | T | C | 0.1214 | 0.982 | 19277 | ADD | -0.06727 | 0.01449 |
| RATE_C | rs111729932 | 11 | 84651377 | C | -5.645 | 8.28e-09 | 0.002929 | DLG2 | protein_coding | Heart rate | vitals | Yes | 11 | 84651377 | A | C | 0.1067 | 0.989 | 19277 | ADD | -0.07127 | 0.01528 |
| RATE_C | rs17743946 | 11 | 85392873 | G | -5.622 | 9.46e-09 | 0.003348 | CREBZF | protein_coding | Heart rate | vitals | Yes | 11 | 85392873 | T | G | 0.1158 | 0.9835 | 19277 | ADD | -0.06241 | 0.01478 |
| RATE_C | rs56046810 | 11 | 84674001 | A | -5.568 | 1.29e-08 | 0.004563 | DLG2 | protein_coding | Heart rate | vitals | Yes | 11 | 84674001 | G | A | 0.1034 | 0.9919 | 19277 | ADD | -0.06994 | 0.01546 |
| RATE_C | rs76447323 | 11 | 85378422 | T | -5.554 | 1.39e-08 | 0.004935 | CREBZF | protein_coding | Heart rate | vitals | Yes | 11 | 85378422 | C | T | 0.09764 | 0.98 | 19277 | ADD | -0.06302 | 0.01598 |
| RATE_C | rs116867021 | 11 | 85035566 | C | -5.506 | 1.84e-08 | 0.006506 | DLG2 | protein_coding | Heart rate | vitals | Yes | 11 | 85035566 | T | C | 0.107 | 0.99 | 19277 | ADD | -0.06239 | 0.01527 |
| RATE_C | rs117080230 | 11 | 85112464 | C | -5.492 | 1.99e-08 | 0.007035 | DLG2 | protein_coding | Heart rate | vitals | Yes | 11 | 85112464 | A | C | 0.106 | 0.9794 | 19277 | ADD | -0.06311 | 0.01541 |
| RATE_C | rs140313369 | 11 | 84717195 | C | -5.473 | 2.22e-08 | 0.007848 | DLG2 | protein_coding | Heart rate | vitals | Yes | 11 | 84717195 | T | C | 0.08545 | 0.9813 | 19277 | ADD | -0.07057 | 0.01696 |
| RATE_C | rs79448310 | 11 | 85203364 | C | -5.472 | 2.22e-08 | 0.007862 | DLG2 | protein_coding | Heart rate | vitals | Yes | 11 | 85203364 | A | C | 0.1014 | 0.9906 | 19277 | ADD | -0.06712 | 0.01563 |
| RATE_C | rs74670688 | 11 | 84958425 | C | -5.471 | 2.24e-08 | 0.007928 | DLG2 | protein_coding | Heart rate | vitals | Yes | 11 | 84958425 | T | C | 0.1207 | 0.9882 | 19277 | ADD | -0.06173 | 0.0145 |
| RATE_C | rs138537852 | 11 | 85443325 | T | -5.45 | 2.51e-08 | 0.008892 | SYTL2 | protein_coding | Heart rate | vitals | Yes | 11 | 85443325 | C | T | 0.09697 | 0.9702 | 19277 | ADD | -0.06553 | 0.01611 |
| RATE_C | rs142781730 | 11 | 84689381 | A | -5.436 | 2.72e-08 | 0.009631 | DLG2 | protein_coding | Heart rate | vitals | Yes | 11 | 84689381 | G | A | 0.08578 | 0.9853 | 19277 | ADD | -0.07321 | 0.01689 |
| RATE_C | rs77386575 | 11 | 85463823 | A | -5.31 | 5.49e-08 | 0.01941 | SYTL2 | protein_coding | Heart rate | vitals | No | 11 | 85463823 | G | A | 0.09359 | 0.9621 | 19277 | ADD | -0.06418 | 0.01644 |
| RATE_C | rs118188129 | 11 | 84641700 | T | -5.174 | 1.14e-07 | 0.0405 | DLG2 | protein_coding | Heart rate | vitals | No | 11 | 84641700 | C | T | 0.09001 | 0.9934 | 19277 | ADD | -0.06288 | 0.01646 |
| SGOT | rs72675488 | 14 | 39875747 | A | 5.371 | 3.91e-08 | 0.01376 | FBXO33 | protein_coding | SGOT | liver | Yes | 14 | 39875747 | G | A | 0.06092 | 0.9874 | 15836 | ADD | 0.05469 | 0.02077 |
| SGOT | rs1955724 | 14 | 39902610 | A | 5.355 | 4.29e-08 | 0.01511 | FBXO33 | protein_coding | SGOT | liver | Yes | 14 | 39902610 | G | A | 0.06312 | 0.9794 | 15836 | ADD | 0.05776 | 0.02051 |

| lab | SNP | CHR | BP | A1 | tstat | P | adjP | Gene_Symbol | biotype | full_name | group | genome_wide | CHROM | GENPOS | ALLELE0 | ALLELE1 | male_A1FREQ | male_INFO | male_N | male_TEST | male_BETA | male_SE |
|---|---|---|---|---|---|---|---|---|---|---|---|---|---|---|---|---|---|---|---|---|---|---|
| SGOT | rs78665473 | 14 | 39979022 | G | 5.194 | 1.03e-07 | 0.03619 | RP11-111A21.1 | lincRNA | SGOT | liver | No | 14 | 39979022 | A | G | 0.06065 | 0.9741 | 15836 | ADD | 0.04971 | 0.02095 |
| EOSIM | rs13153393 | 5 | 167604213 | G | -5.513 | 1.76e-08 | 0.006243 | TENM2 | protein_coding | Eosinophils/100 leukocytes in Blood by Automated count | immune/inflammatory | Yes | 5 | 167604213 | A | G | 0.1121 | 0.9804 | 15117 | ADD | -0.0459 | 0.0179 |
| EOSIM | rs13180204 | 5 | 167622914 | C | -5.509 | 1.80e-08 | 0.006386 | TENM2 | protein_coding | Eosinophils/100 leukocytes in Blood by Automated count | immune/inflammatory | Yes | 5 | 167622914 | T | C | 0.1191 | 0.9951 | 15117 | ADD | -0.04397 | 0.0173 |
| EOSIM | rs34373271 | 5 | 167629483 | G | -5.469 | 2.27e-08 | 0.008037 | TENM2 | protein_coding | Eosinophils/100 leukocytes in Blood by Automated count | immune/inflammatory | Yes | 5 | 167629483 | A | G | 0.1193 | 0.994 | 15117 | ADD | -0.04499 | 0.0173 |
| Chol | 10:78918297 | 10 | 78918297 | A | 5.515 | 1.74e-08 | 0.006244 | KCNMA1 | protein_coding | Cholesterol [Mass/volume] in Serum or Plasma | metabolic | Yes | 10 | 78918297 | C | A | 0.08445 | 1.002 | 15405 | ADD | 0.08435 | 0.0187 |
| Chol | rs1319039 | 10 | 78931122 | C | 5.353 | 4.34e-08 | 0.01552 | KCNMA1 | protein_coding | Cholesterol [Mass/volume] in Serum or Plasma | metabolic | Yes | 10 | 78931122 | T | C | 0.08755 | 0.9889 | 15405 | ADD | 0.0821 | 0.01852 |
| Chol | rs76094981 | 2 | 156792830 | T | 5.293 | 6.03e-08 | 0.02159 | AC093375.1 | lincRNA | Cholesterol [Mass/volume] in Serum or Plasma | metabolic | No | 2 | 156792830 | C | T | 0.0555 | 1.005 | 15405 | ADD | 0.08567 | 0.02271 |
| Chol | rs10762749 | 10 | 78926083 | G | 5.271 | 6.78e-08 | 0.02427 | KCNMA1 | protein_coding | Cholesterol [Mass/volume] in Serum or Plasma | metabolic | No | 10 | 78926083 | A | G | 0.08793 | 0.9957 | 15405 | ADD | 0.08256 | 0.01842 |
| Chol | rs77477120 | 2 | 156795870 | T | 5.258 | 7.30e-08 | 0.02613 | AC093375.1 | lincRNA | Cholesterol [Mass/volume] in Serum or Plasma | metabolic | No | 2 | 156795870 | C | T | 0.0551 | 1.002 | 15405 | ADD | 0.08536 | 0.02282 |
| Chol | rs7412 | 19 | 45412079 | T | 5.247 | 7.73e-08 | 0.02765 | APOE | protein_coding | Cholesterol [Mass/volume] in Serum or Plasma | metabolic | No | 19 | 45412079 | C | T | 0.07758 | 1.01 | 15405 | ADD | -0.1611 | 0.01943 |
| Chol | rs11002051 | 10 | 78921392 | T | 5.237 | 8.18e-08 | 0.02928 | KCNMA1 | protein_coding | Cholesterol [Mass/volume] in Serum or Plasma | metabolic | No | 10 | 78921392 | C | T | 0.08739 | 0.9977 | 15405 | ADD | 0.08048 | 0.01846 |
| Chol | rs6434699 | 2 | 195511939 | G | 5.182 | 1.10e-07 | 0.03929 | AC006196.1 | lincRNA | Cholesterol [Mass/volume] in Serum or Plasma | metabolic | No | 2 | 195511939 | T | G | 0.2342 | 0.959 | 15405 | ADD | 0.04853 | 0.01258 |
| Chol | rs114758991 | 10 | 78914855 | T | 5.178 | 1.12e-07 | 0.0402 | KCNMA1 | protein_coding | Cholesterol [Mass/volume] in Serum or Plasma | metabolic | No | 10 | 78914855 | C | T | 0.07244 | 0.987 | 15405 | ADD | 0.08593 | 0.02023 |
| Chol | rs6727455 | 2 | 195514148 | G | 5.175 | 1.14e-07 | 0.04077 | AC006196.1 | lincRNA | Cholesterol [Mass/volume] in Serum or Plasma | metabolic | No | 2 | 195514148 | A | G | 0.2291 | 0.9845 | 15405 | ADD | 0.04838 | 0.01251 |
| Chol | rs78714935 | 2 | 195508862 | A | 5.139 | 1.38e-07 | 0.04943 | AC073973.1 | miRNA | Cholesterol [Mass/volume] in Serum or Plasma | metabolic | No | 2 | 195508862 | G | A | 0.2329 | 0.957 | 15405 | ADD | 0.04824 | 0.01261 |
| Chol | rs12471587 | 2 | 195493854 | C | 5.139 | 1.38e-07 | 0.04948 | AC073973.1 | miRNA | Cholesterol [Mass/volume] in Serum or Plasma | metabolic | No | 2 | 195493854 | T | C | 0.2279 | 0.9793 | 15405 | ADD | 0.04705 | 0.01256 |
| Trigs | rs5915786 | 23 | 4642016 | C | -5.378 | 3.76e-08 | 0.0142 | AC074035.1 | miRNA | Triglyceride [Mass/volume] in Serum or Plasma | metabolic | Yes | 23 | 4642016 | T | C | 0.374 | 1.994 | 15431 | ADD | -0.01966 | 0.007648 |
| Trigs | rs7057542 | 23 | 4643144 | G | -5.36 | 4.16e-08 | 0.01568 | AC074035.1 | miRNA | Triglyceride [Mass/volume] in Serum or Plasma | metabolic | Yes | 23 | 4643144 | A | G | 0.372 | 1.968 | 15431 | ADD | -0.01958 | 0.007708 |
| Trigs | rs7061816 | 23 | 4643018 | C | -5.36 | 4.16e-08 | 0.0157 | AC074035.1 | miRNA | Triglyceride [Mass/volume] in Serum or Plasma | metabolic | Yes | 23 | 4643018 | T | C | 0.372 | 1.968 | 15431 | ADD | -0.01959 | 0.007708 |
| Trigs | rs7057815 | 23 | 4643304 | G | -5.344 | 4.54e-08 | 0.01712 | AC074035.1 | miRNA | Triglyceride [Mass/volume] in Serum or Plasma | metabolic | Yes | 23 | 4643304 | A | G | 0.372 | 1.968 | 15431 | ADD | -0.01957 | 0.007708 |
| PTT.pt | rs9819879 | 3 | 166000429 | C | 5.529 | 1.61e-08 | 0.005435 | RP11-12N13.1 | pseudogene | aPTT in Platelet poor plasma | blood | Yes | 3 | 166000429 | A | C | 0.08292 | 1.004 | 16272 | ADD | 0.08125 | 0.01901 |
| PTT.pt | rs9852581 | 3 | 165999536 | T | 5.528 | 1.62e-08 | 0.005446 | RP11-12N13.1 | pseudogene | aPTT in Platelet poor plasma | blood | Yes | 3 | 165999536 | C | T | 0.08292 | 1.004 | 16272 | ADD | 0.08125 | 0.01901 |
| PTT.pt | rs11926722 | 3 | 165942772 | T | 5.505 | 1.85e-08 | 0.006221 | RP11-12N13.2 | pseudogene | aPTT in Platelet poor plasma | blood | Yes | 3 | 165942772 | C | T | 0.0821 | 0.9869 | 16272 | ADD | 0.08201 | 0.01926 |
| PTT.pt | rs9873795 | 3 | 166003323 | A | 5.353 | 4.33e-08 | 0.01459 | RP11-12N13.1 | pseudogene | aPTT in Platelet poor plasma | blood | Yes | 3 | 166003323 | G | A | 0.06742 | 1.012 | 16272 | ADD | 0.0728 | 0.02082 |
| PTT.pt | rs34218892 | 3 | 165964677 | C | 5.347 | 4.48e-08 | 0.01509 | RP11-12N13.1 | pseudogene | aPTT in Platelet poor plasma | blood | Yes | 3 | 165964677 | T | C | 0.06714 | 1.014 | 16272 | ADD | 0.071 | 0.02084 |
| PTT.pt | rs1371890 | 3 | 166008760 | G | 5.346 | 4.51e-08 | 0.01518 | RP11-12N13.1 | pseudogene | aPTT in Platelet poor plasma | blood | Yes | 3 | 166008760 | A | G | 0.06744 | 1.013 | 16272 | ADD | 0.07257 | 0.02081 |
| PTT.pt | rs11922089 | 3 | 166008835 | A | 5.346 | 4.51e-08 | 0.01518 | RP11-12N13.1 | pseudogene | aPTT in Platelet poor plasma | blood | Yes | 3 | 166008835 | G | A | 0.06744 | 1.013 | 16272 | ADD | 0.07257 | 0.02081 |
| PTT.pt | rs28504984 | 3 | 165968129 | A | 5.328 | 4.96e-08 | 0.01673 | RP11-12N13.1 | pseudogene | aPTT in Platelet poor plasma | blood | Yes | 3 | 165968129 | C | A | 0.06714 | 1.014 | 16272 | ADD | 0.07103 | 0.02084 |

| lab | SNP | CHR | BP | A1 | tstat | P | adjP | Gene_Symbol | biotype | full_name | group | genome_wide | CHROM | GENPOS | ALLELE0 | ALLELE1 | male_A1FREQ | male_INFO | male_N | male_TEST | male_BETA | male_SE |
|---|---|---|---|---|---|---|---|---|---|---|---|---|---|---|---|---|---|---|---|---|---|---|
| PTT.pt | rs34434720 | 3 | 165959008 | T | 5.328 | 4.97e-08 | 0.01675 | RP11-12N13.1 | pseudogene | aPTT in Platelet poor plasma | blood | Yes | 3 | 165959008 | C | T | 0.06718 | 1.014 | 16272 | ADD | 0.07095 | 0.02083 |
| PTT.pt | rs71302579 | 3 | 165988776 | C | 5.323 | 5.11e-08 | 0.01721 | RP11-12N13.1 | pseudogene | aPTT in Platelet poor plasma | blood | No | 3 | 165988776 | T | C | 0.06738 | 1.012 | 16272 | ADD | 0.0719 | 0.02083 |
| PTT.pt | rs13322959 | 3 | 165991203 | C | 5.318 | 5.24e-08 | 0.01767 | RP11-12N13.1 | pseudogene | aPTT in Platelet poor plasma | blood | No | 3 | 165991203 | T | C | 0.06753 | 1.012 | 16272 | ADD | 0.07249 | 0.0208 |
| lab | SNP | CHR | BP | A1 | tstat | P | adjP | Gene_Symbol | biotype | full_name | group | genome_wide | CHROM | GENPOS | ALLELE0 | ALLELE1 | male_A1FREQ | male_INFO | male_N | male_TEST | male_BETA | male_SE |
| PTT.pt | rs1440672 | 3 | 165995411 | G | 5.318 | 5.26e-08 | 0.01772 | RP11-12N13.1 | pseudogene | aPTT in Platelet poor plasma | blood | No | 3 | 165995411 | A | G | 0.06753 | 1.012 | 16272 | ADD | 0.07249 | 0.0208 |
| PTT.pt | rs9876567 | 3 | 165975608 | C | 5.314 | 5.35e-08 | 0.01803 | RP11-12N13.1 | pseudogene | aPTT in Platelet poor plasma | blood | No | 3 | 165975608 | T | C | 0.06715 | 1.014 | 16272 | ADD | 0.07103 | 0.02084 |
| PTT.pt | rs9810862 | 3 | 165986016 | T | 5.307 | 5.57e-08 | 0.01878 | RP11-12N13.1 | pseudogene | aPTT in Platelet poor plasma | blood | No | 3 | 165986016 | C | T | 0.06753 | 1.013 | 16272 | ADD | 0.07218 | 0.0208 |
| PTT.pt | rs11915213 | 3 | 165981577 | T | 5.301 | 5.76e-08 | 0.0194 | RP11-12N13.1 | pseudogene | aPTT in Platelet poor plasma | blood | No | 3 | 165981577 | C | T | 0.06753 | 1.013 | 16272 | ADD | 0.07201 | 0.02079 |
| PTT.pt | rs35688183 | 3 | 166115634 | G | 5.281 | 6.43e-08 | 0.02168 | RP11-12N13.1 | pseudogene | aPTT in Platelet poor plasma | blood | No | 3 | 166115634 | A | G | 0.05935 | 0.9923 | 16272 | ADD | 0.08114 | 0.02232 |
| PTT.pt | rs9848421 | 3 | 165953968 | C | 5.237 | 8.18e-08 | 0.02757 | RP11-12N13.2 | pseudogene | aPTT in Platelet poor plasma | blood | No | 3 | 165953968 | T | C | 0.06619 | 1.005 | 16272 | ADD | 0.07467 | 0.02107 |
| PTT.pt | rs34423841 | 3 | 165941325 | C | 5.231 | 8.43e-08 | 0.0284 | RP11-12N13.2 | pseudogene | aPTT in Platelet poor plasma | blood | No | 3 | 165941325 | T | C | 0.06691 | 0.9916 | 16272 | ADD | 0.07213 | 0.02111 |
| PTT.pt | rs35376432 | 3 | 165987121 | A | 5.221 | 8.92e-08 | 0.03006 | RP11-12N13.1 | pseudogene | aPTT in Platelet poor plasma | blood | No | 3 | 165987121 | G | A | 0.06657 | 1.01 | 16272 | ADD | 0.07488 | 0.02096 |
| PTT.pt | rs11927547 | 3 | 166003163 | T | 5.218 | 9.02e-08 | 0.0304 | RP11-12N13.1 | pseudogene | aPTT in Platelet poor plasma | blood | No | 3 | 166003163 | C | T | 0.06806 | 1.011 | 16272 | ADD | 0.07049 | 0.02074 |
| PTT.pt | rs28605977 | 3 | 166010530 | G | 5.218 | 9.03e-08 | 0.03042 | RP11-12N13.1 | pseudogene | aPTT in Platelet poor plasma | blood | No | 3 | 166010530 | T | G | 0.0682 | 1.013 | 16272 | ADD | 0.07089 | 0.0207 |
| PTT.pt | rs28446617 | 3 | 166010425 | C | 5.218 | 9.03e-08 | 0.03043 | RP11-12N13.1 | pseudogene | aPTT in Platelet poor plasma | blood | No | 3 | 166010425 | T | C | 0.0682 | 1.013 | 16272 | ADD | 0.07088 | 0.0207 |
| PTT.pt | rs9871920 | 3 | 165999127 | C | 5.218 | 9.04e-08 | 0.03046 | RP11-12N13.1 | pseudogene | aPTT in Platelet poor plasma | blood | No | 3 | 165999127 | T | C | 0.06806 | 1.011 | 16272 | ADD | 0.07049 | 0.02074 |
| PTT.pt | rs9872809 | 3 | 165999671 | C | 5.218 | 9.04e-08 | 0.03046 | RP11-12N13.1 | pseudogene | aPTT in Platelet poor plasma | blood | No | 3 | 165999671 | T | C | 0.06806 | 1.011 | 16272 | ADD | 0.07049 | 0.02074 |
| PTT.pt | rs28692775 | 3 | 166010854 | T | 5.218 | 9.05e-08 | 0.03049 | RP11-12N13.1 | pseudogene | aPTT in Platelet poor plasma | blood | No | 3 | 166010854 | C | T | 0.06821 | 1.013 | 16272 | ADD | 0.07078 | 0.0207 |
| PTT.pt | rs1440688 | 3 | 166009992 | C | 5.218 | 9.06e-08 | 0.03052 | RP11-12N13.1 | pseudogene | aPTT in Platelet poor plasma | blood | No | 3 | 166009992 | T | C | 0.06823 | 1.013 | 16272 | ADD | 0.07089 | 0.0207 |
| PTT.pt | rs13091733 | 3 | 166009894 | C | 5.217 | 9.10e-08 | 0.03066 | RP11-12N13.1 | pseudogene | aPTT in Platelet poor plasma | blood | No | 3 | 166009894 | T | C | 0.06822 | 1.013 | 16272 | ADD | 0.0709 | 0.0207 |
| PTT.pt | rs13062071 | 3 | 165986722 | G | 5.211 | 9.41e-08 | 0.03172 | RP11-12N13.1 | pseudogene | aPTT in Platelet poor plasma | blood | No | 3 | 165986722 | A | G | 0.06657 | 1.01 | 16272 | ADD | 0.07486 | 0.02096 |
| PTT.pt | rs10936481 | 3 | 166002471 | C | 5.206 | 9.64e-08 | 0.03248 | RP11-12N13.1 | pseudogene | aPTT in Platelet poor plasma | blood | No | 3 | 166002471 | T | C | 0.06823 | 1.012 | 16272 | ADD | 0.07087 | 0.02071 |
| PTT.pt | rs11927567 | 3 | 166003108 | A | 5.206 | 9.64e-08 | 0.03248 | RP11-12N13.1 | pseudogene | aPTT in Platelet poor plasma | blood | No | 3 | 166003108 | G | A | 0.06823 | 1.012 | 16272 | ADD | 0.07087 | 0.02071 |
| PTT.pt | rs9829942 | 3 | 166002035 | G | 5.206 | 9.66e-08 | 0.03254 | RP11-12N13.1 | pseudogene | aPTT in Platelet poor plasma | blood | No | 3 | 166002035 | A | G | 0.06823 | 1.012 | 16272 | ADD | 0.07087 | 0.02071 |
| PTT.pt | rs1440687 | 3 | 166009917 | T | 5.202 | 9.84e-08 | 0.03318 | RP11-12N13.1 | pseudogene | aPTT in Platelet poor plasma | blood | No | 3 | 166009917 | C | T | 0.06822 | 1.013 | 16272 | ADD | 0.07091 | 0.0207 |
| PTT.pt | rs11923918 | 3 | 166006629 | C | 5.202 | 9.88e-08 | 0.03331 | RP11-12N13.1 | pseudogene | aPTT in Platelet poor plasma | blood | No | 3 | 166006629 | T | C | 0.06824 | 1.012 | 16272 | ADD | 0.07073 | 0.0207 |
| PTT.pt | rs9845863 | 3 | 166008221 | T | 5.201 | 9.92e-08 | 0.03344 | RP11-12N13.1 | pseudogene | aPTT in Platelet poor plasma | blood | No | 3 | 166008221 | G | T | 0.06825 | 1.012 | 16272 | ADD | 0.07067 | 0.0207 |
| PTT.pt | rs1371891 | 3 | 166008590 | G | 5.199 | 1.00e-07 | 0.03371 | RP11-12N13.1 | pseudogene | aPTT in Platelet poor plasma | blood | No | 3 | 166008590 | A | G | 0.06825 | 1.012 | 16272 | ADD | 0.07065 | 0.0207 |
| PTT.pt | rs1371892 | 3 | 166008551 | T | 5.199 | 1.00e-07 | 0.03375 | RP11-12N13.1 | pseudogene | aPTT in Platelet poor plasma | blood | No | 3 | 166008551 | C | T | 0.06825 | 1.012 | 16272 | ADD | 0.07065 | 0.0207 |

| lab | SNP | CHR | BP | A1 | tstat | P | adjP | Gene_Symbol | biotype | full_name | group | genome_wide | CHROM | GENPOS | ALLELE0 | ALLELE1 | male_A1FREQ | male_INFO | male_N | male_TEST | male_BETA | male_SE |
|---|---|---|---|---|---|---|---|---|---|---|---|---|---|---|---|---|---|---|---|---|---|---|
| PTT.pt | rs2902332 | 3 | 166009142 | G | 5.199 | 1.00e-07 | 0.03375 | RP11-12N13.1 | pseudogene | aPTT in Platelet poor plasma | blood | No | 3 | 166009142 | A | G | 0.06825 | 1.012 | 16272 | ADD | 0.07065 | 0.0207 |
| PTT.pt | rs9829029 | 3 | 166004927 | A | 5.199 | 1.00e-07 | 0.03375 | RP11-12N13.1 | pseudogene | aPTT in Platelet poor plasma | blood | No | 3 | 166004927 | C | A | 0.06825 | 1.012 | 16272 | ADD | 0.07065 | 0.0207 |
| PTT.pt | rs9841424 | 3 | 166007749 | A | 5.199 | 1.00e-07 | 0.03375 | RP11-12N13.1 | pseudogene | aPTT in Platelet poor plasma | blood | No | 3 | 166007749 | G | A | 0.06825 | 1.012 | 16272 | ADD | 0.07065 | 0.0207 |
| PTT.pt | rs9859832 | 3 | 166006944 | C | 5.199 | 1.00e-07 | 0.03375 | RP11-12N13.1 | pseudogene | aPTT in Platelet poor plasma | blood | No | 3 | 166006944 | T | C | 0.06825 | 1.012 | 16272 | ADD | 0.07065 | 0.0207 |
| PTT.pt | rs9873210 | 3 | 166006696 | G | 5.199 | 1.00e-07 | 0.03377 | RP11-12N13.1 | pseudogene | aPTT in Platelet poor plasma | blood | No | 3 | 166006696 | A | G | 0.06824 | 1.012 | 16272 | ADD | 0.07065 | 0.0207 |
| PTT.pt | rs34772290 | 3 | 166216316 | G | 5.178 | 1.12e-07 | 0.03789 | RP11-450H5.1 | lincRNA | aPTT in Platelet poor plasma | blood | No | 3 | 166216316 | A | G | 0.06264 | 1.006 | 16272 | ADD | 0.07836 | 0.0216 |
| PTT.pt | rs35585812 | 3 | 166209806 | A | 5.173 | 1.15e-07 | 0.03883 | RP11-450H5.1 | lincRNA | aPTT in Platelet poor plasma | blood | No | 3 | 166209806 | G | A | 0.06272 | 1.007 | 16272 | ADD | 0.07734 | 0.02158 |
| PTT.pt | rs9880101 | 3 | 165944706 | A | 5.164 | 1.21e-07 | 0.04072 | RP11-12N13.2 | pseudogene | aPTT in Platelet poor plasma | blood | No | 3 | 165944706 | G | A | 0.06593 | 0.9972 | 16272 | ADD | 0.07508 | 0.02119 |
| PTT.pt | rs9879693 | 3 | 165944626 | T | 5.164 | 1.21e-07 | 0.04084 | RP11-12N13.2 | pseudogene | aPTT in Platelet poor plasma | blood | No | 3 | 165944626 | C | T | 0.06593 | 0.9972 | 16272 | ADD | 0.07507 | 0.02119 |
| PTT.pt | rs34635876 | 3 | 165944305 | C | 5.161 | 1.23e-07 | 0.04145 | RP11-12N13.2 | pseudogene | aPTT in Platelet poor plasma | blood | No | 3 | 165944305 | T | C | 0.06591 | 0.9963 | 16272 | ADD | 0.07518 | 0.0212 |
| PTT.pt | rs141519684 | 3 | 166219458 | T | 5.138 | 1.39e-07 | 0.04677 | RP11-450H5.1 | lincRNA | aPTT in Platelet poor plasma | blood | No | 3 | 166219458 | C | T | 0.06272 | 1.008 | 16272 | ADD | 0.07748 | 0.02158 |
| BLOOD.PRESSURE | rs2689177 | 1 | 238931099 | C | 5.32 | 5.19e-08 | 0.01833 | MIPEPP2 | pseudogene | Diastolic blood pressure | vitals | No | 1 | 238931099 | A | C | 0.4842 | 0.981 | 12921 | ADD | 0.05114 | 0.01132 |
| PULSE | rs9956549 | 18 | 4084772 | G | 5.335 | 4.79e-08 | 0.01721 | DLGAP1 | protein_coding | Heart rate | vitals | Yes | 18 | 4084772 | A | G | 0.1588 | 0.9952 | 12927 | ADD | 0.07574 | 0.01496 |
| WEIGHT_SCALE | rs4731998 | 7 | 80645119 | G | 5.305 | 5.63e-08 | 0.02137 | SEMA3C | protein_coding | Weight scale | vitals | No | 7 | 80645119 | A | G | 0.1797 | 0.999 | 12994 | ADD | 0.08104 | 0.01591 |
| WEIGHT_SCALE | rs2040978 | 7 | 80640930 | T | 5.271 | 6.80e-08 | 0.0258 | SEMA3C | protein_coding | Weight scale | vitals | No | 7 | 80640930 | G | T | 0.1799 | 1.005 | 12994 | ADD | 0.08039 | 0.01585 |
| WEIGHT_SCALE | rs11576565 | 1 | 72114787 | A | 5.156 | 1.26e-07 | 0.04792 | NEGR1 | protein_coding | Weight scale | vitals | No | 1 | 72114787 | G | A | 0.4859 | 0.9823 | 12994 | ADD | 0.02966 | 0.0123 |
| HDL.C | rs34277739 | 19 | 19910183 | G | 5.358 | 4.22e-08 | 0.01586 | ZNF506 | protein_coding | HDL Cholesterol | metabolic | Yes | 19 | 19910183 | A | G | 0.1826 | 0.9947 | 14828 | ADD | 0.04868 | 0.01416 |
| HDL.C | rs7252985 | 19 | 19908116 | T | 5.346 | 4.49e-08 | 0.01688 | ZNF506 | protein_coding | HDL Cholesterol | metabolic | Yes | 19 | 19908116 | G | T | 0.1826 | 0.9948 | 14828 | ADD | 0.04867 | 0.01416 |
| HDL.C | rs60041535 | 19 | 19922203 | G | 5.341 | 4.63e-08 | 0.01742 | ZNF506 | protein_coding | HDL Cholesterol | metabolic | Yes | 19 | 19922203 | A | G | 0.1823 | 0.9885 | 14828 | ADD | 0.04831 | 0.01421 |
| HDL.C | rs12975121 | 19 | 19905011 | T | 5.326 | 5.03e-08 | 0.0189 | ZNF506 | protein_coding | HDL Cholesterol | metabolic | No | 19 | 19905011 | C | T | 0.1827 | 0.9968 | 14828 | ADD | 0.04827 | 0.01414 |
| HDL.C | rs35018071 | 19 | 19914095 | G | 5.323 | 5.10e-08 | 0.01918 | ZNF506 | protein_coding | HDL Cholesterol | metabolic | No | 19 | 19914095 | T | G | 0.1826 | 0.9946 | 14828 | ADD | 0.04874 | 0.01416 |
| HDL.C | rs248955 | 19 | 19895707 | G | 5.223 | 8.81e-08 | 0.03313 | CTC-559E9.4 | lincRNA | HDL Cholesterol | metabolic | No | 19 | 19895707 | T | G | 0.1829 | 0.9982 | 14828 | ADD | 0.04744 | 0.01412 |
| HDL.C | rs23654 | 19 | 19897424 | C | 5.219 | 9.00e-08 | 0.03385 | ZNF506 | protein_coding | HDL Cholesterol | metabolic | No | 19 | 19897424 | T | C | 0.1839 | 1.005 | 14828 | ADD | 0.04714 | 0.01405 |
| LDL.C | rs7412 | 19 | 45412079 | T | 6.206 | 2.71e-10 | 9.67e-05 | APOE | protein_coding | LDL cholesterol | metabolic | Yes | 19 | 45412079 | C | T | 0.07767 | 1.006 | 14481 | ADD | -0.3048 | 0.01996 |
| LDL.C | rs1163962 | 12 | 17576764 | G | -5.391 | 3.50e-08 | 0.01246 | RP11-871F6.2 | pseudogene | LDL cholesterol | metabolic | Yes | 12 | 17576764 | A | G | 0.3289 | 0.9975 | 14481 | ADD | -0.05849 | 0.01136 |
| LDL.C | rs72654473 | 19 | 45414399 | A | 5.32 | 5.18e-08 | 0.01847 | APOE | protein_coding | LDL cholesterol | metabolic | No | 19 | 45414399 | C | A | 0.1057 | 0.9993 | 14481 | ADD | -0.1872 | 0.01744 |
| LDL.C | rs445925 | 19 | 45415640 | A | 5.25 | 7.61e-08 | 0.02711 | APOC1 | protein_coding | LDL cholesterol | metabolic | No | 19 | 45415640 | G | A | 0.1073 | 0.9986 | 14481 | ADD | -0.1856 | 0.01733 |

| lab | SNP | CHR | BP | A1 | tstat | P | adjP | Gene_Symbol | biotype | full_name | group | genome_wide | CHROM | GENPOS | ALLELE0 | ALLELE1 | male_A1FREQ | male_INFO | male_N | male_TEST | male_BETA | male_SE |
|---|---|---|---|---|---|---|---|---|---|---|---|---|---|---|---|---|---|---|---|---|---|---|
| HEIGHT | rs11750865 | 5 | 167471631 | G | 5.171 | 1.17e-07 | 0.0491 | TENM2 | protein_coding | Height | vitals | No | 5 | 167471631 | T | G | 0.07397 | 0.9678 | 12621 | ADD | 0.08764 | 0.0231 |
| NTAuto | rs6131114 | 20 | 10711916 | T | -5.241 | 7.99e-08 | 0.02973 | RP11-103J8.1 | antisense | Neutrophils | immune/inflammatory | No | 20 | 10711916 | C | T | 0.1058 | 0.9924 | 21662 | ADD | -0.0468 | 0.01336 |
| EOSIAB | rs944055 | 13 | 114843588 | A | -5.368 | 3.97e-08 | 0.01376 | RASA3 | protein_coding | Eosinophils [#/volume] in Blood by Automated count | immune/inflammatory | Yes | 13 | 114843588 | G | A | 0.4523 | 0.9931 | 12637 | ADD | -0.04753 | 0.01219 |
| EOSIAB | rs9525387 | 13 | 114848948 | C | -5.269 | 6.86e-08 | 0.02376 | RASA3 | protein_coding | Eosinophils [#/volume] in Blood by Automated count | immune/inflammatory | No | 13 | 114848948 | T | C | 0.4575 | 0.9918 | 12637 | ADD | -0.04743 | 0.01219 |
| EOSIAB | rs9525388 | 13 | 114848963 | G | -5.269 | 6.86e-08 | 0.02376 | RASA3 | protein_coding | Eosinophils [#/volume] in Blood by Automated count | immune/inflammatory | No | 13 | 114848963 | A | G | 0.4575 | 0.9918 | 12637 | ADD | -0.04743 | 0.01219 |
| EOSIAB | rs4883675 | 13 | 114842648 | T | -5.242 | 7.96e-08 | 0.02758 | RASA3 | protein_coding | Eosinophils [#/volume] in Blood by Automated count | immune/inflammatory | No | 13 | 114842648 | C | T | 0.4541 | 0.9974 | 12637 | ADD | -0.04588 | 0.01216 |
| EOSIAB | rs9525254 | 13 | 114853128 | A | -5.167 | 1.19e-07 | 0.0411 | RASA3 | protein_coding | Eosinophils [#/volume] in Blood by Automated count | immune/inflammatory | No | 13 | 114853128 | C | A | 0.4645 | 0.9882 | 12637 | ADD | -0.04552 | 0.0122 |
| R.APACHE.TEMPERATURE | rs78712959 | 12 | 75647582 | T | 5.197 | 1.01e-07 | 0.03401 | RP11-560G2.1 | antisense | Body temperature | vitals | No | 12 | 75647582 | C | T | 0.06216 | 0.964 | 11789 | ADD | 0.1291 | 0.02258 |
| lab | SNP | CHR | BP | A1 | tstat | P | adjP | Gene_Symbol | biotype | full_name | group | genome_wide | CHROM | GENPOS | ALLELE0 | ALLELE1 | male_A1FREQ | male_INFO | male_N | male_TEST | male_BETA | male_SE |
| RESPIRATIONS | rs1948089 | 2 | 77964983 | T | -5.306 | 5.59e-08 | 0.01916 | AC105399.2 | pseudogene | Respiratory rate | vitals | No | 2 | 77964983 | C | T | 0.403 | 0.9931 | 11399 | ADD | -0.0486 | 0.01319 |
| RESPIRATIONS | rs6732508 | 2 | 77966145 | A | -5.291 | 6.10e-08 | 0.02088 | AC105399.2 | pseudogene | Respiratory rate | vitals | No | 2 | 77966145 | C | A | 0.4032 | 0.9929 | 11399 | ADD | -0.04922 | 0.01319 |
| RESPIRATIONS | rs60909491 | 2 | 77964729 | A | -5.266 | 6.96e-08 | 0.02384 | AC105399.2 | pseudogene | Respiratory rate | vitals | No | 2 | 77964729 | G | A | 0.4029 | 0.993 | 11399 | ADD | -0.04814 | 0.01319 |
| RESPIRATIONS | rs6547157 | 2 | 77965060 | C | -5.265 | 7.01e-08 | 0.02401 | AC105399.2 | pseudogene | Respiratory rate | vitals | No | 2 | 77965060 | T | C | 0.4032 | 0.9935 | 11399 | ADD | -0.04828 | 0.01319 |
| RESPIRATIONS | rs6732620 | 2 | 77966169 | T | -5.26 | 7.21e-08 | 0.02469 | AC105399.2 | pseudogene | Respiratory rate | vitals | No | 2 | 77966169 | C | T | 0.4035 | 0.9931 | 11399 | ADD | -0.04888 | 0.01319 |
| RESPIRATIONS | rs6729714 | 2 | 77965996 | A | -5.259 | 7.26e-08 | 0.02486 | AC105399.2 | pseudogene | Respiratory rate | vitals | No | 2 | 77965996 | G | A | 0.4035 | 0.9931 | 11399 | ADD | -0.04889 | 0.01319 |
| RESPIRATIONS | rs6747628 | 2 | 77966286 | C | -5.258 | 7.28e-08 | 0.02494 | AC105399.2 | pseudogene | Respiratory rate | vitals | No | 2 | 77966286 | T | C | 0.4035 | 0.9931 | 11399 | ADD | -0.0489 | 0.01319 |
| RESPIRATIONS | rs6729146 | 2 | 77965750 | T | -5.258 | 7.30e-08 | 0.025 | AC105399.2 | pseudogene | Respiratory rate | vitals | No | 2 | 77965750 | C | T | 0.4035 | 0.9931 | 11399 | ADD | -0.04891 | 0.01319 |
| RESPIRATIONS | rs60025330 | 2 | 77964701 | A | -5.244 | 7.84e-08 | 0.02687 | AC105399.2 | pseudogene | Respiratory rate | vitals | No | 2 | 77964701 | G | A | 0.4033 | 0.993 | 11399 | ADD | -0.04787 | 0.01319 |
| RESPIRATIONS | rs6729102 | 2 | 77965521 | A | -5.244 | 7.85e-08 | 0.02689 | AC105399.2 | pseudogene | Respiratory rate | vitals | No | 2 | 77965521 | G | A | 0.4047 | 0.9934 | 11399 | ADD | -0.04832 | 0.01318 |
| RESPIRATIONS | rs59942226 | 2 | 77964399 | C | -5.237 | 8.17e-08 | 0.028 | AC105399.2 | pseudogene | Respiratory rate | vitals | No | 2 | 77964399 | T | C | 0.4033 | 0.9894 | 11399 | ADD | -0.04793 | 0.01322 |

| lab | SNP | CHR | BP | A1 | tstat | P | adjP | Gene_Symbol | biotype | full_name | group | genome_wide | CHROM | GENPOS | ALLELE0 | ALLELE1 | male_A1FREQ | male_INFO | male_N | male_TEST | male_BETA | male_SE |
|---|---|---|---|---|---|---|---|---|---|---|---|---|---|---|---|---|---|---|---|---|---|---|
| RESPIRATIONS | rs59322091 | 2 | 77964408 | T | -5.232 | 8.37e-08 | 0.02867 | AC105399.2 | pseudogene | Respiratory rate | vitals | No | 2 | 77964408 | G | T | 0.4033 | 0.9893 | 11399 | ADD | -0.04805 | 0.01322 |
| RESPIRATIONS | rs57528236 | 2 | 77964467 | G | -5.229 | 8.53e-08 | 0.02923 | AC105399.2 | pseudogene | Respiratory rate | vitals | No | 2 | 77964467 | A | G | 0.4035 | 0.9899 | 11399 | ADD | -0.04804 | 0.01321 |
| RESPIRATIONS | rs11887817 | 2 | 77966578 | G | -5.222 | 8.84e-08 | 0.03027 | AC105399.2 | pseudogene | Respiratory rate | vitals | No | 2 | 77966578 | A | G | 0.4027 | 0.9931 | 11399 | ADD | -0.04865 | 0.01319 |
| RESPIRATIONS | rs58134944 | 2 | 77964500 | T | -5.21 | 9.46e-08 | 0.0324 | AC105399.2 | pseudogene | Respiratory rate | vitals | No | 2 | 77964500 | C | T | 0.4029 | 0.9916 | 11399 | ADD | -0.04745 | 0.0132 |
| RESPIRATIONS | rs6547158 | 2 | 77965325 | C | -5.197 | 1.01e-07 | 0.03473 | AC105399.2 | pseudogene | Respiratory rate | vitals | No | 2 | 77965325 | T | C | 0.4055 | 0.9978 | 11399 | ADD | -0.04748 | 0.01315 |
| RESPIRATIONS | rs6743936 | 2 | 77965568 | C | -5.197 | 1.01e-07 | 0.03473 | AC105399.2 | pseudogene | Respiratory rate | vitals | No | 2 | 77965568 | T | C | 0.4044 | 0.9938 | 11399 | ADD | -0.0477 | 0.01318 |
| Eos | rs73596652 | 8 | 18889465 | C | 5.206 | 9.66e-08 | 0.03392 | PSD3 | protein_coding | Eos | immune/inflammatory | No | 8 | 18889465 | T | C | 0.1357 | 1.016 | 11491 | ADD | 0.07335 | 0.01776 |
| Eos | rs75213539 | 8 | 18875685 | G | 5.135 | 1.41e-07 | 0.0495 | PSD3 | protein_coding | Eos | immune/inflammatory | No | 8 | 18875685 | T | G | 0.1334 | 1.012 | 11491 | ADD | 0.07348 | 0.01792 |
| Eos | rs80038167 | 8 | 18874936 | A | 5.135 | 1.41e-07 | 0.04954 | PSD3 | protein_coding | Eos | immune/inflammatory | No | 8 | 18874936 | C | A | 0.1333 | 1.012 | 11491 | ADD | 0.0736 | 0.01793 |
| LYMPAB | rs586355 | 10 | 6558188 | T | -5.296 | 5.93e-08 | 0.02161 | PRKCQ | protein_coding | Lymphocytes [#/volume] in Blood by Automated count | immune/inflammatory | No | 10 | 6558188 | G | T | 0.1304 | 0.9927 | 19927 | ADD | -0.04844 | 0.01433 |
| LYMPAB | rs10763439 | 10 | 58702171 | C | -5.241 | 7.99e-08 | 0.02911 | MIR3924 | miRNA | Lymphocytes [#/volume] in Blood by Automated count | immune/inflammatory | No | 10 | 58702171 | T | C | 0.4835 | 0.9926 | 19927 | ADD | -0.03772 | 0.009654 |
| LYMPAB | rs12573519 | 10 | 58707149 | C | -5.219 | 9.01e-08 | 0.03282 | MIR3924 | miRNA | Lymphocytes [#/volume] in Blood by Automated count | immune/inflammatory | No | 10 | 58707149 | T | C | 0.477 | 0.9936 | 19927 | ADD | -0.03521 | 0.009652 |
| LYMPAB | rs627715 | 10 | 6555831 | A | -5.152 | 1.29e-07 | 0.04704 | PRKCQ | protein_coding | Lymphocytes [#/volume] in Blood by Automated count | immune/inflammatory | No | 10 | 6555831 | G | A | 0.1386 | 0.9998 | 19927 | ADD | -0.03956 | 0.01391 |
| MONORE | rs7260534 | 19 | 52074474 | T | 5.419 | 3.00e-08 | 0.01115 | ZNF175 | protein_coding | Monocytes/100 leukocytes in Blood by Automated count | immune/inflammatory | Yes | 19 | 52074474 | G | T | 0.3763 | 0.9968 | 19994 | ADD | 0.04324 | 0.009713 |
| MONORE | rs2864107 | 19 | 52069027 | A | 5.254 | 7.43e-08 | 0.02758 | SIGLEC29P | pseudogene | Monocytes/100 leukocytes in Blood by Automated count | immune/inflammatory | No | 19 | 52069027 | G | A | 0.3634 | 0.9861 | 19994 | ADD | 0.04096 | 0.009835 |
| MONORE | rs1457095 | 19 | 52072434 | G | 5.252 | 7.51e-08 | 0.02788 | ZNF175 | protein_coding | Monocytes/100 leukocytes in Blood by Automated count | immune/inflammatory | No | 19 | 52072434 | A | G | 0.3633 | 0.9863 | 19994 | ADD | 0.04105 | 0.009835 |
| MONORE | rs10500307 | 19 | 52071472 | A | 5.213 | 9.30e-08 | 0.0345 | ZNF175 | protein_coding | Monocytes/100 leukocytes in Blood by Automated count | immune/inflammatory | No | 19 | 52071472 | G | A | 0.3631 | 0.9852 | 19994 | ADD | 0.04059 | 0.009842 |
| BAbs | rs28759753 | 4 | 164238799 | A | 5.147 | 1.32e-07 | 0.04623 | NPY1R | protein_coding | Basophils [#/volume] in Blood by Automated count | immune/inflammatory | No | 4 | 164238799 | C | A | 0.07986 | 0.9968 | 10793 | ADD | 0.08197 | 0.02299 |
| EoAbs | rs2160871 | 3 | 10446826 | T | -5.418 | 3.01e-08 | 0.01071 | ATP2B2 | protein_coding | EoAbs | immune/inflammatory | Yes | 3 | 10446826 | C | T | 0.1884 | 0.9747 | 10829 | ADD | -0.06657 | 0.01606 |
| EoAbs | rs759692 | 3 | 10443000 | T | -5.351 | 4.37e-08 | 0.01553 | ATP2B2 | protein_coding | EoAbs | immune/inflammatory | Yes | 3 | 10443000 | C | T | 0.1945 | 0.9776 | 10829 | ADD | -0.06388 | 0.01584 |

| lab | SNP | CHR | BP | A1 | tstat | P | adjP | Gene_Symbol | biotype | full_name | group | genome_wide | CHROM | GENPOS | ALLELE0 | ALLELE1 | male_A1FREQ | male_INFO | male_N | male_TEST | male_BETA | male_SE |
|---|---|---|---|---|---|---|---|---|---|---|---|---|---|---|---|---|---|---|---|---|---|---|
| EoAbs | rs6786239 | 3 | 10448772 | T | -5.279 | 6.51e-08 | 0.02312 | ATP2B2 | protein_coding | EoAbs | immune/inflammatory | No | 3 | 10448772 | C | T | 0.1407 | 0.9783 | 10829 | ADD | -0.06896 | 0.01803 |
| LymAbs | rs12995157 | 2 | 238589737 | A | 5.174 | 1.15e-07 | 0.04035 | LRRFIP1 | protein_coding | Lymphocytes [#/volume] in Blood by Automated count | immune/inflammatory | No | 2 | 238589737 | G | A | 0.2409 | 0.9683 | 11025 | ADD | 0.05191 | 0.0145 |
| LymAbs | rs4142730 | 2 | 238591913 | A | 5.155 | 1.27e-07 | 0.04452 | LRRFIP1 | protein_coding | Lymphocytes [#/volume] in Blood by Automated count | immune/inflammatory | No | 2 | 238591913 | G | A | 0.2405 | 0.9681 | 11025 | ADD | 0.05182 | 0.01451 |
| HgbA1C | rs10507238 | 12 | 113749780 | A | -5.278 | 6.55e-08 | 0.02404 | SLC8B1 | protein_coding | Hemoglobin A1c (Glycated) Hemoglobin A1c (Glycated) | metabolic | No | 12 | 113749780 | G | A | 0.1048 | 0.9811 | 11801 | ADD | -0.06764 | 0.02015 |
| HgbA1C | rs56801977 | 12 | 113757109 | A | -5.198 | 1.01e-07 | 0.03698 | SLC8B1 | protein_coding | Hemoglobin A1c (Glycated) Hemoglobin A1c (Glycated) | metabolic | No | 12 | 113757109 | G | A | 0.1044 | 0.9842 | 11801 | ADD | -0.06598 | 0.02015 |
| HgbA1C | rs67763786 | 12 | 113770841 | C | -5.179 | 1.12e-07 | 0.04096 | SLC8B1 | protein_coding | Hemoglobin A1c (Glycated) Hemoglobin A1c (Glycated) | metabolic | No | 12 | 113770841 | T | C | 0.1058 | 0.9792 | 11801 | ADD | -0.06472 | 0.02008 |
| HgbA1C | rs67206313 | 12 | 113769569 | T | -5.167 | 1.19e-07 | 0.04372 | SLC8B1 | protein_coding | Hemoglobin A1c (Glycated) Hemoglobin A1c (Glycated) | metabolic | No | 12 | 113769569 | C | T | 0.1055 | 0.9804 | 11801 | ADD | -0.06369 | 0.02009 |
| HgbA1C | rs67521432 | 12 | 113769100 | G | -5.146 | 1.33e-07 | 0.04883 | SLC8B1 | protein_coding | Hemoglobin A1c (Glycated) Hemoglobin A1c (Glycated) | metabolic | No | 12 | 113769100 | T | G | 0.1055 | 0.9804 | 11801 | ADD | -0.06361 | 0.02009 |
| HgbA1C | rs66878077 | 12 | 113766589 | T | -5.144 | 1.34e-07 | 0.04937 | SLC8B1 | protein_coding | Hemoglobin A1c (Glycated) Hemoglobin A1c (Glycated) | metabolic | No | 12 | 113766589 | C | T | 0.1055 | 0.9804 | 11801 | ADD | -0.06362 | 0.02009 |
| HgbA1C | rs57296942 | 12 | 113766969 | T | -5.143 | 1.35e-07 | 0.04965 | SLC8B1 | protein_coding | Hemoglobin A1c (Glycated) Hemoglobin A1c (Glycated) | metabolic | No | 12 | 113766969 | C | T | 0.1055 | 0.9804 | 11801 | ADD | -0.06359 | 0.02009 |
| HgbA1C | rs59331454 | 12 | 113767219 | A | -5.143 | 1.35e-07 | 0.04965 | SLC8B1 | protein_coding | Hemoglobin A1c (Glycated) Hemoglobin A1c (Glycated) | metabolic | No | 12 | 113767219 | G | A | 0.1055 | 0.9804 | 11801 | ADD | -0.06359 | 0.02009 |
| HgbA1C | rs7969912 | 12 | 113767891 | G | -5.143 | 1.35e-07 | 0.04965 | SLC8B1 | protein_coding | Hemoglobin A1c (Glycated) Hemoglobin A1c (Glycated) | metabolic | No | 12 | 113767891 | A | G | 0.1055 | 0.9804 | 11801 | ADD | -0.06359 | 0.02009 |
| SedRat | rs9806363 | 15 | 79110302 | G | 5.249 | 7.65e-08 | 0.02745 | MORF4L1 | protein_coding | Erythrocyte sedimentation rate by Westergren method | blood | No | 15 | 79110302 | A | G | 0.4077 | 0.9729 | 8621 | ADD | 0.06566 | 0.01551 |
| SedRat | rs4380028 | 15 | 79111093 | T | 5.248 | 7.70e-08 | 0.02762 | MORF4L1 | protein_coding | Erythrocyte sedimentation rate by Westergren method | blood | No | 15 | 79111093 | C | T | 0.4077 | 0.9729 | 8621 | ADD | 0.06566 | 0.01551 |
| SedRat | rs12442476 | 15 | 79117856 | G | 5.245 | 7.80e-08 | 0.02797 | MORF4L1 | protein_coding | Erythrocyte sedimentation rate by Westergren method | blood | No | 15 | 79117856 | A | G | 0.4131 | 0.9734 | 8621 | ADD | 0.06507 | 0.01548 |
| SedRat | rs12915192 | 15 | 79115585 | G | 5.243 | 7.91e-08 | 0.02836 | MORF4L1 | protein_coding | Erythrocyte sedimentation rate by Westergren method | blood | No | 15 | 79115585 | A | G | 0.4133 | 0.9745 | 8621 | ADD | 0.06492 | 0.01547 |
| SedRat | rs9920493 | 15 | 79113846 | A | 5.242 | 7.96e-08 | 0.02854 | MORF4L1 | protein_coding | Erythrocyte sedimentation rate by Westergren method | blood | No | 15 | 79113846 | C | A | 0.4132 | 0.9745 | 8621 | ADD | 0.06491 | 0.01547 |
| SedRat | rs7165075 | 15 | 79115965 | G | 5.241 | 7.97e-08 | 0.02857 | MORF4L1 | protein_coding | Erythrocyte sedimentation rate by Westergren method | blood | No | 15 | 79115965 | A | G | 0.4132 | 0.9745 | 8621 | ADD | 0.0649 | 0.01547 |
| SedRat | rs6495335 | 15 | 79117133 | G | 5.241 | 7.97e-08 | 0.02857 | MORF4L1 | protein_coding | Erythrocyte sedimentation rate by Westergren method | blood | No | 15 | 79117133 | T | G | 0.4132 | 0.9745 | 8621 | ADD | 0.0649 | 0.01547 |
| SedRat | rs7165902 | 15 | 79115872 | T | 5.241 | 7.97e-08 | 0.02858 | MORF4L1 | protein_coding | Erythrocyte sedimentation rate by Westergren method | blood | No | 15 | 79115872 | G | T | 0.4132 | 0.9745 | 8621 | ADD | 0.0649 | 0.01547 |
| SedRat | rs9920968 | 15 | 79118747 | G | 5.241 | 7.97e-08 | 0.02858 | MORF4L1 | protein_coding | Erythrocyte sedimentation rate by Westergren method | blood | No | 15 | 79118747 | A | G | 0.4131 | 0.9735 | 8621 | ADD | 0.06512 | 0.01548 |
| SedRat | rs7167056 | 15 | 79113484 | G | 5.241 | 7.98e-08 | 0.0286 | MORF4L1 | protein_coding | Erythrocyte sedimentation rate by Westergren method | blood | No | 15 | 79113484 | T | G | 0.4133 | 0.9745 | 8621 | ADD | 0.06489 | 0.01547 |
| SedRat | rs7167242 | 15 | 79113541 | G | 5.241 | 8.01e-08 | 0.0287 | MORF4L1 | protein_coding | Erythrocyte sedimentation rate by Westergren method | blood | No | 15 | 79113541 | A | G | 0.4132 | 0.9744 | 8621 | ADD | 0.06488 | 0.01547 |
| SedRat | rs7166770 | 15 | 79113568 | A | 5.24 | 8.02e-08 | 0.02874 | MORF4L1 | protein_coding | Erythrocyte sedimentation rate by Westergren method | blood | No | 15 | 79113568 | G | A | 0.4132 | 0.9744 | 8621 | ADD | 0.0649 | 0.01547 |
| SedRat | rs8026598 | 15 | 79112938 | A | 5.239 | 8.08e-08 | 0.02898 | MORF4L1 | protein_coding | Erythrocyte sedimentation rate by Westergren method | blood | No | 15 | 79112938 | G | A | 0.4109 | 0.9845 | 8621 | ADD | 0.06458 | 0.0154 |
| SedRat | rs12898292 | 15 | 79112603 | A | 5.236 | 8.22e-08 | 0.02947 | MORF4L1 | protein_coding | Erythrocyte sedimentation rate by Westergren method | blood | No | 15 | 79112603 | G | A | 0.4137 | 0.9735 | 8621 | ADD | 0.06516 | 0.01548 |
| SedRat | rs12898392 | 15 | 79112432 | G | 5.235 | 8.26e-08 | 0.02961 | MORF4L1 | protein_coding | Erythrocyte sedimentation rate by Westergren method | blood | No | 15 | 79112432 | A | G | 0.4132 | 0.9745 | 8621 | ADD | 0.06514 | 0.01547 |
| SedRat | rs8041064 | 15 | 79115388 | A | 5.232 | 8.40e-08 | 0.03013 | MORF4L1 | protein_coding | Erythrocyte sedimentation rate by Westergren method | blood | No | 15 | 79115388 | G | A | 0.4132 | 0.9742 | 8621 | ADD | 0.06486 | 0.01547 |

| lab | SNP | CHR | BP | A1 | tstat | P | adjP | Gene_Symbol | biotype | full_name | group | genome_wide | CHROM | GENPOS | ALLELE0 | ALLELE1 | male_A1FREQ | male_INFO | male_N | male_TEST | male_BETA | male_SE |
|---|---|---|---|---|---|---|---|---|---|---|---|---|---|---|---|---|---|---|---|---|---|---|
| SedRat | rs7166340 | 15 | 79113293 | C | 5.229 | 8.50e-08 | 0.03049 | MORF4L1 | protein_coding | Erythrocyte sedimentation rate by Westergren method | blood | No | 15 | 79113293 | T | C | 0.4132 | 0.9745 | 8621 | ADD | 0.06492 | 0.01547 |
| lab | SNP | CHR | BP | A1 | tstat | P | adjP | Gene_Symbol | biotype | full_name | group | genome_wide | CHROM | GENPOS | ALLELE0 | ALLELE1 | male_A1FREQ | male_INFO | male_N | male_TEST | male_BETA | male_SE |
| SedRat | rs7167853 | 15 | 79113318 | T | 5.229 | 8.50e-08 | 0.03049 | MORF4L1 | protein_coding | Erythrocyte sedimentation rate by Westergren method | blood | No | 15 | 79113318 | C | T | 0.4132 | 0.9745 | 8621 | ADD | 0.06492 | 0.01547 |
| SedRat | rs8027833 | 15 | 79112837 | T | 5.229 | 8.51e-08 | 0.0305 | MORF4L1 | protein_coding | Erythrocyte sedimentation rate by Westergren method | blood | No | 15 | 79112837 | C | T | 0.4132 | 0.9745 | 8621 | ADD | 0.06492 | 0.01547 |
| SedRat | rs6495333 | 15 | 79113215 | G | 5.228 | 8.59e-08 | 0.03081 | MORF4L1 | protein_coding | Erythrocyte sedimentation rate by Westergren method | blood | No | 15 | 79113215 | A | G | 0.4132 | 0.9745 | 8621 | ADD | 0.06488 | 0.01547 |
| SedRat | rs7176187 | 15 | 79121373 | T | 5.225 | 8.73e-08 | 0.0313 | MORF4L1 | protein_coding | Erythrocyte sedimentation rate by Westergren method | blood | No | 15 | 79121373 | C | T | 0.4129 | 0.9731 | 8621 | ADD | 0.06493 | 0.01548 |
| SedRat | rs7403608 | 15 | 79115570 | G | 5.219 | 8.99e-08 | 0.03222 | MORF4L1 | protein_coding | Erythrocyte sedimentation rate by Westergren method | blood | No | 15 | 79115570 | A | G | 0.4083 | 0.9789 | 8621 | ADD | 0.06451 | 0.01546 |
| SedRat | rs7403728 | 15 | 79112016 | G | 5.217 | 9.09e-08 | 0.03259 | MORF4L1 | protein_coding | Erythrocyte sedimentation rate by Westergren method | blood | No | 15 | 79112016 | T | G | 0.4082 | 0.9789 | 8621 | ADD | 0.06463 | 0.01546 |
| SedRat | rs7403593 | 15 | 79111962 | G | 5.217 | 9.10e-08 | 0.03261 | MORF4L1 | protein_coding | Erythrocyte sedimentation rate by Westergren method | blood | No | 15 | 79111962 | A | G | 0.4082 | 0.9789 | 8621 | ADD | 0.06463 | 0.01546 |
| SedRat | rs34372090 | 15 | 79110735 | T | 5.214 | 9.25e-08 | 0.03315 | MORF4L1 | protein_coding | Erythrocyte sedimentation rate by Westergren method | blood | No | 15 | 79110735 | C | T | 0.4067 | 0.9733 | 8621 | ADD | 0.06548 | 0.01552 |
| SedRat | rs12898712 | 15 | 79112622 | G | 5.212 | 9.34e-08 | 0.03348 | MORF4L1 | protein_coding | Erythrocyte sedimentation rate by Westergren method | blood | No | 15 | 79112622 | T | G | 0.4137 | 0.9739 | 8621 | ADD | 0.0647 | 0.01547 |
| SedRat | rs12909317 | 15 | 79114419 | T | 5.211 | 9.41e-08 | 0.03374 | MORF4L1 | protein_coding | Erythrocyte sedimentation rate by Westergren method | blood | No | 15 | 79114419 | G | T | 0.4082 | 0.979 | 8621 | ADD | 0.06453 | 0.01546 |
| SedRat | rs7403708 | 15 | 79123885 | C | 5.209 | 9.50e-08 | 0.03405 | MORF4L1 | protein_coding | Erythrocyte sedimentation rate by Westergren method | blood | No | 15 | 79123885 | T | C | 0.4076 | 0.9777 | 8621 | ADD | 0.06485 | 0.01548 |
| SedRat | rs66925868 | 15 | 79115406 | G | 5.208 | 9.53e-08 | 0.03416 | MORF4L1 | protein_coding | Erythrocyte sedimentation rate by Westergren method | blood | No | 15 | 79115406 | A | G | 0.4057 | 0.9791 | 8621 | ADD | 0.0658 | 0.01549 |
| SedRat | rs7178051 | 15 | 79118296 | T | 5.205 | 9.70e-08 | 0.03479 | MORF4L1 | protein_coding | Erythrocyte sedimentation rate by Westergren method | blood | No | 15 | 79118296 | C | T | 0.4077 | 0.9777 | 8621 | ADD | 0.06477 | 0.01548 |
| SedRat | rs58021857 | 15 | 79119229 | A | 5.201 | 9.89e-08 | 0.03545 | MORF4L1 | protein_coding | Erythrocyte sedimentation rate by Westergren method | blood | No | 15 | 79119229 | C | A | 0.4077 | 0.9776 | 8621 | ADD | 0.06472 | 0.01548 |
| SedRat | rs12898452 | 15 | 79112628 | C | 5.199 | 1.00e-07 | 0.03585 | MORF4L1 | protein_coding | Erythrocyte sedimentation rate by Westergren method | blood | No | 15 | 79112628 | A | C | 0.4135 | 0.9739 | 8621 | ADD | 0.06453 | 0.01547 |
| SedRat | rs7402376 | 15 | 79113570 | G | 5.198 | 1.01e-07 | 0.03613 | MORF4L1 | protein_coding | Erythrocyte sedimentation rate by Westergren method | blood | No | 15 | 79113570 | A | G | 0.41 | 0.986 | 8621 | ADD | 0.064 | 0.0154 |
| SedRat | rs72749206 | 15 | 79122948 | A | 5.189 | 1.06e-07 | 0.03789 | MORF4L1 | protein_coding | Erythrocyte sedimentation rate by Westergren method | blood | No | 15 | 79122948 | G | A | 0.4051 | 0.9778 | 8621 | ADD | 0.06595 | 0.0155 |
| SedRat | rs56683682 | 15 | 79112524 | T | 5.187 | 1.07e-07 | 0.03826 | MORF4L1 | protein_coding | Erythrocyte sedimentation rate by Westergren method | blood | No | 15 | 79112524 | G | T | 0.4072 | 0.9794 | 8621 | ADD | 0.06444 | 0.01547 |
| SedRat | rs10083697 | 15 | 79124039 | A | 5.179 | 1.12e-07 | 0.04009 | MORF4L1 | protein_coding | Erythrocyte sedimentation rate by Westergren method | blood | No | 15 | 79124039 | G | A | 0.4075 | 0.9774 | 8621 | ADD | 0.06444 | 0.01548 |
| SedRat | rs10083696 | 15 | 79124016 | A | 5.167 | 1.19e-07 | 0.04266 | MORF4L1 | protein_coding | Erythrocyte sedimentation rate by Westergren method | blood | No | 15 | 79124016 | G | A | 0.4075 | 0.977 | 8621 | ADD | 0.0643 | 0.01548 |
| SedRat | rs4389112 | 15 | 79122099 | A | 5.159 | 1.24e-07 | 0.04444 | MORF4L1 | protein_coding | Erythrocyte sedimentation rate by Westergren method | blood | No | 15 | 79122099 | G | A | 0.407 | 0.9855 | 8621 | ADD | 0.06511 | 0.01543 |
| SedRat | rs7403714 | 15 | 79118650 | T | 5.157 | 1.26e-07 | 0.04501 | MORF4L1 | protein_coding | Erythrocyte sedimentation rate by Westergren method | blood | No | 15 | 79118650 | C | T | 0.4066 | 0.978 | 8621 | ADD | 0.06446 | 0.01548 |
| SedRat | rs8035093 | 15 | 79120670 | T | 5.148 | 1.32e-07 | 0.04719 | MORF4L1 | protein_coding | Erythrocyte sedimentation rate by Westergren method | blood | No | 15 | 79120670 | C | T | 0.4149 | 0.971 | 8621 | ADD | 0.0634 | 0.01549 |
| SedRat | rs62011002 | 15 | 79120578 | C | 5.142 | 1.36e-07 | 0.04879 | MORF4L1 | protein_coding | Erythrocyte sedimentation rate by Westergren method | blood | No | 15 | 79120578 | T | C | 0.4149 | 0.9711 | 8621 | ADD | 0.06332 | 0.01549 |
| SedRat | rs62011001 | 15 | 79120467 | G | 5.142 | 1.36e-07 | 0.0488 | MORF4L1 | protein_coding | Erythrocyte sedimentation rate by Westergren method | blood | No | 15 | 79120467 | A | G | 0.4149 | 0.9711 | 8621 | ADD | 0.06332 | 0.01549 |
| SedRat | rs4389113 | 15 | 79121739 | A | 5.139 | 1.38e-07 | 0.04951 | MORF4L1 | protein_coding | Erythrocyte sedimentation rate by Westergren method | blood | No | 15 | 79121739 | G | A | 0.4066 | 0.9779 | 8621 | ADD | 0.06425 | 0.01548 |

| lab | SNP | CHR | BP | A1 | tstat | P | adjP | Gene_Symbol | biotype | full_name | group | genome_wide | CHROM | GENPOS | ALLELE0 | ALLELE1 | male_A1FREQ | male_INFO | male_N | male_TEST | male_BETA | male_SE |
|---|---|---|---|---|---|---|---|---|---|---|---|---|---|---|---|---|---|---|---|---|---|---|
| MPV | rs55752763 | 8 | 96468350 | A | -5.47 | 2.24e-08 | 0.009201 | KB-1047C11.2 | lincRNA | Platelet mean volume [Entitic volume] in Blood by Automated count Platelet mean volume [Entitic volume] in Blood by Automated count | blood | Yes | 8 | 96468350 | G | A | 0.1999 | 0.9904 | 19784 | ADD | -0.04844 | 0.01172 |
| MPV | rs7017356 | 8 | 96461413 | A | -5.375 | 3.83e-08 | 0.01571 | KB-1047C11.2 | lincRNA | Platelet mean volume [Entitic volume] in Blood by Automated count Platelet mean volume [Entitic volume] in Blood by Automated count | blood | Yes | 8 | 96461413 | G | A | 0.1986 | 0.9922 | 19784 | ADD | -0.04664 | 0.01174 |
| MPV | rs6987245 | 8 | 96464039 | C | -5.369 | 3.95e-08 | 0.01621 | KB-1047C11.2 | lincRNA | Platelet mean volume [Entitic volume] in Blood by Automated count Platelet mean volume [Entitic volume] in Blood by Automated count | blood | Yes | 8 | 96464039 | A | C | 0.1985 | 0.9919 | 19784 | ADD | -0.04641 | 0.01174 |
| MPV | rs55876772 | 8 | 96463109 | G | -5.362 | 4.11e-08 | 0.01685 | KB-1047C11.2 | lincRNA | Platelet mean volume [Entitic volume] in Blood by Automated count Platelet mean volume [Entitic volume] in Blood by Automated count | blood | Yes | 8 | 96463109 | A | G | 0.1985 | 0.9924 | 19784 | ADD | -0.04645 | 0.01174 |
| MPV | rs55737626 | 8 | 96468384 | A | -5.358 | 4.20e-08 | 0.01722 | KB-1047C11.2 | lincRNA | Platelet mean volume [Entitic volume] in Blood by Automated count Platelet mean volume [Entitic volume] in Blood by Automated count | blood | Yes | 8 | 96468384 | G | A | 0.1985 | 0.9917 | 19784 | ADD | -0.0464 | 0.01175 |
| MPV | rs113411905 | 8 | 96465271 | C | -5.357 | 4.24e-08 | 0.01736 | KB-1047C11.2 | lincRNA | Platelet mean volume [Entitic volume] in Blood by Automated count Platelet mean volume [Entitic volume] in Blood by Automated count | blood | Yes | 8 | 96465271 | T | C | 0.1987 | 0.9913 | 19784 | ADD | -0.04617 | 0.01174 |
| MPV | rs7830718 | 8 | 96463289 | T | -5.312 | 5.43e-08 | 0.02227 | KB-1047C11.2 | lincRNA | Platelet mean volume [Entitic volume] in Blood by Automated count Platelet mean volume [Entitic volume] in Blood by Automated count | blood | No | 8 | 96463289 | C | T | 0.1987 | 0.9923 | 19784 | ADD | -0.04563 | 0.01174 |
| MPV | rs17220111 | 13 | 74627057 | G | 5.241 | 8.00e-08 | 0.03281 | KLF12 | protein_coding | Platelet mean volume [Entitic volume] in Blood by Automated count Platelet mean volume [Entitic volume] in Blood by Automated count | blood | No | 13 | 74627057 | T | G | 0.2819 | 1.001 | 19784 | ADD | 0.03665 | 0.01033 |
| MPV | rs4292657 | 8 | 96469866 | T | -5.213 | 9.27e-08 | 0.038 | KB-1047C11.2 | lincRNA | Platelet mean volume [Entitic volume] in Blood by Automated count Platelet mean volume [Entitic volume] in Blood by Automated count | blood | No | 8 | 96469866 | C | T | 0.2233 | 0.983 | 19784 | ADD | -0.0424 | 0.0113 |
| MPV | rs1466874 | 8 | 96469848 | T | -5.213 | 9.30e-08 | 0.03813 | KB-1047C11.2 | lincRNA | Platelet mean volume [Entitic volume] in Blood by Automated count Platelet mean volume [Entitic volume] in Blood by Automated count | blood | No | 8 | 96469848 | C | T | 0.2233 | 0.9829 | 19784 | ADD | -0.04239 | 0.0113 |
| MPV | rs1392789 | 8 | 96469004 | G | -5.204 | 9.77e-08 | 0.04005 | KB-1047C11.2 | lincRNA | Platelet mean volume [Entitic volume] in Blood by Automated count Platelet mean volume [Entitic volume] in Blood by Automated count | blood | No | 8 | 96469004 | A | G | 0.2232 | 0.9834 | 19784 | ADD | -0.04264 | 0.0113 |
| MPV | rs9530270 | 13 | 74622339 | A | 5.187 | 1.07e-07 | 0.04382 | KLF12 | protein_coding | Platelet mean volume [Entitic volume] in Blood by Automated count Platelet mean volume [Entitic volume] in Blood by Automated count | blood | No | 13 | 74622339 | C | A | 0.2904 | 0.9902 | 19784 | ADD | 0.03628 | 0.01029 |

| lab | SNP | CHR | BP | A1 | tstat | P | adjP | Gene_Symbol | biotype | full_name | group | genome_wide | CHROM | GENPOS | ALLELE0 | ALLELE1 | male_A1FREQ | male_INFO | male_N | male_TEST | male_BETA | male_SE |
|---|---|---|---|---|---|---|---|---|---|---|---|---|---|---|---|---|---|---|---|---|---|---|
| MPV | rs1466875 | 8 | 96469655 | C | -5.177 | 1.13e-07 | 0.04627 | KB-1047C11.2 | lincRNA | Platelet mean volume [Entitic volume] in Blood by Automated count Platelet mean volume [Entitic volume] in Blood by Automated count | blood | No | 8 | 96469655 | A | C | 0.2228 | 0.9801 | 19784 | ADD | -0.04242 | 0.01133 |
| MPV | rs1466878 | 8 | 96469578 | C | -5.176 | 1.14e-07 | 0.04658 | KB-1047C11.2 | lincRNA | Platelet mean volume [Entitic volume] in Blood by Automated count Platelet mean volume [Entitic volume] in Blood by Automated count | blood | No | 8 | 96469578 | A | C | 0.2233 | 0.9823 | 19784 | ADD | -0.04209 | 0.0113 |
| MPV | rs1466877 | 8 | 96469618 | C | -5.175 | 1.14e-07 | 0.04681 | KB-1047C11.2 | lincRNA | Platelet mean volume [Entitic volume] in Blood by Automated count Platelet mean volume [Entitic volume] in Blood by Automated count | blood | No | 8 | 96469618 | T | C | 0.2228 | 0.9801 | 19784 | ADD | -0.04245 | 0.01133 |
| MPV | rs112251874 | 8 | 96465415 | A | -5.162 | 1.22e-07 | 0.04997 | KB-1047C11.2 | lincRNA | Platelet mean volume [Entitic volume] in Blood by Automated count Platelet mean volume [Entitic volume] in Blood by Automated count | blood | No | 8 | 96465415 | G | A | 0.1813 | 0.9916 | 19784 | ADD | -0.04715 | 0.01216 |
| Iron | rs987280 | 5 | 85448604 | A | 5.6 | 1.08e-08 | 0.003601 | PTP4A1P4 | pseudogene | Iron [Mass/volume] in Serum or Plasma | blood | Yes | 5 | 85448604 | G | A | 0.4862 | 0.9958 | 7897 | ADD | 0.06138 | 0.01539 |
| Iron | rs1345624 | 5 | 85449508 | T | 5.594 | 1.11e-08 | 0.003712 | PTP4A1P4 | pseudogene | Iron [Mass/volume] in Serum or Plasma | blood | Yes | 5 | 85449508 | G | T | 0.4873 | 0.991 | 7897 | ADD | 0.06158 | 0.01543 |
| Iron | rs1422105 | 5 | 85448371 | A | 5.551 | 1.42e-08 | 0.004759 | PTP4A1P4 | pseudogene | Iron [Mass/volume] in Serum or Plasma | blood | Yes | 5 | 85448371 | C | A | 0.4872 | 0.9914 | 7897 | ADD | 0.06166 | 0.01543 |
| Iron | rs1422104 | 5 | 85447842 | T | 5.346 | 4.51e-08 | 0.0151 | PTP4A1P4 | pseudogene | Iron [Mass/volume] in Serum or Plasma | blood | Yes | 5 | 85447842 | G | T | 0.4983 | 0.9914 | 7897 | ADD | 0.05811 | 0.01542 |
| Iron | rs1363266 | 5 | 85451165 | G | 5.309 | 5.50e-08 | 0.01843 | PTP4A1P4 | pseudogene | Iron [Mass/volume] in Serum or Plasma | blood | No | 5 | 85451165 | A | G | 0.4983 | 0.9917 | 7897 | ADD | 0.05745 | 0.01542 |
| Iron | rs1345623 | 5 | 85446912 | T | 5.295 | 5.94e-08 | 0.01989 | PTP4A1P4 | pseudogene | Iron [Mass/volume] in Serum or Plasma | blood | No | 5 | 85446912 | C | T | 0.4992 | 0.9893 | 7897 | ADD | 0.05772 | 0.01544 |
| Iron | rs1345622 | 5 | 85446883 | A | -5.267 | 6.93e-08 | 0.02322 | PTP4A1P4 | pseudogene | Iron [Mass/volume] in Serum or Plasma | blood | No | 5 | 85446883 | G | A | 0.4828 | 0.9899 | 7897 | ADD | -0.06323 | 0.01544 |
| Iron | rs1363265 | 5 | 85450966 | G | -5.241 | 7.99e-08 | 0.02677 | PTP4A1P4 | pseudogene | Iron [Mass/volume] in Serum or Plasma | blood | No | 5 | 85450966 | A | G | 0.4828 | 0.9899 | 7897 | ADD | -0.06273 | 0.01544 |
| SUCrea | rs1901753 | 2 | 11142721 | G | -5.161 | 1.23e-07 | 0.04151 | KCNF1 | protein_coding | Creatinine [Mass/volume] in Urine | urinary | No | 2 | 11142721 | A | G | 0.4338 | 1.01 | 7600 | ADD | -0.03704 | 0.01507 |
| X_percent_SAT | rs1345624 | 5 | 85449508 | T | 5.314 | 5.37e-08 | 0.01809 | PTP4A1P4 | pseudogene | % saturation | other | No | 5 | 85449508 | G | T | 0.4851 | 1.001 | 6545 | ADD | 0.07663 | 0.01691 |
| X_percent_SAT | rs987280 | 5 | 85448604 | A | 5.31 | 5.48e-08 | 0.01846 | PTP4A1P4 | pseudogene | % saturation | other | No | 5 | 85448604 | G | A | 0.4839 | 1.006 | 6545 | ADD | 0.07625 | 0.01688 |
| X_percent_SAT | rs1422105 | 5 | 85448371 | A | 5.302 | 5.73e-08 | 0.0193 | PTP4A1P4 | pseudogene | % saturation | other | No | 5 | 85448371 | C | A | 0.4848 | 1.001 | 6545 | ADD | 0.07643 | 0.01691 |
| X_percent_SAT | rs1345623 | 5 | 85446912 | T | 5.135 | 1.41e-07 | 0.04752 | PTP4A1P4 | pseudogene | % saturation | other | No | 5 | 85446912 | C | T | 0.4968 | 1 | 6545 | ADD | 0.07308 | 0.01691 |
| X_percent_SAT | rs1422104 | 5 | 85447842 | T | 5.128 | 1.47e-07 | 0.04933 | PTP4A1P4 | pseudogene | % saturation | other | No | 5 | 85447842 | G | T | 0.4958 | 1.002 | 6545 | ADD | 0.07269 | 0.0169 |
| R.AN.HEART.RATE.MONITOR | rs2293105 | 7 | 45725133 | A | 5.473 | 2.21e-08 | 0.007275 | ADCY1 | protein_coding | Heart rate | vitals | Yes | 7 | 45725133 | G | A | 0.1939 | 0.9939 | 5668 | ADD | 0.09714 | 0.02208 |

| lab | SNP | CHR | BP | A1 | tstat | P | adjP | Gene_Symbol | biotype | full_name | group | genome_wide | CHROM | GENPOS | ALLELE0 | ALLELE1 | male_A1FREQ | male_INFO | male_N | male_TEST | male_BETA | male_SE |
|---|---|---|---|---|---|---|---|---|---|---|---|---|---|---|---|---|---|---|---|---|---|---|
| R.AN.HEART.RATE.MONITOR | rs2293104 | 7 | 45725485 | T | 5.465 | 2.31e-08 | 0.007597 | ADCY1 | protein_coding | Heart rate | vitals | Yes | 7 | 45725485 | G | T | 0.1938 | 0.9941 | 5668 | ADD | 0.09692 | 0.02208 |
| R.AN.HEART.RATE.MONITOR | rs78447894 | 2 | 234315394 | C | 5.274 | 6.67e-08 | 0.02192 | DGKD | protein_coding | Heart rate | vitals | No | 2 | 234315394 | A | C | 0.05264 | 0.9844 | 5668 | ADD | 0.1898 | 0.0393 |
| R.AN.PULSE.SPO2 | rs2293104 | 7 | 45725485 | T | 5.482 | 2.10e-08 | 0.006971 | ADCY1 | protein_coding | Heart rate by Pulse oximetry | vitals | Yes | 7 | 45725485 | G | T | 0.1939 | 0.9945 | 5626 | ADD | 0.08977 | 0.02176 |
| lab | SNP | CHR | BP | A1 | tstat | P | adjP | Gene_Symbol | biotype | full_name | group | genome_wide | CHROM | GENPOS | ALLELE0 | ALLELE1 | male_A1FREQ | male_INFO | male_N | male_TEST | male_BETA | male_SE |
| R.AN.PULSE.SPO2 | rs2293105 | 7 | 45725133 | A | 5.481 | 2.12e-08 | 0.00702 | ADCY1 | protein_coding | Heart rate by Pulse oximetry | vitals | Yes | 7 | 45725133 | G | A | 0.194 | 0.9943 | 5626 | ADD | 0.08973 | 0.02176 |
| R.AN.PULSE.SPO2 | rs11767059 | 7 | 45732186 | T | 5.129 | 1.46e-07 | 0.04843 | ADCY1 | protein_coding | Heart rate by Pulse oximetry | vitals | No | 7 | 45732186 | C | T | 0.1885 | 0.9987 | 5626 | ADD | 0.08138 | 0.02195 |
| TProt | rs12173947 | 6 | 416489 | T | -5.241 | 7.98e-08 | 0.02939 | IRF4 | protein_coding | Total protein | blood | No | 6 | 416489 | C | T | 0.1754 | 0.9909 | 19923 | ADD | -0.04104 | 0.01147 |
| TProt | rs12174418 | 6 | 416710 | C | -5.24 | 8.02e-08 | 0.02952 | IRF4 | protein_coding | Total protein | blood | No | 6 | 416710 | T | C | 0.1772 | 0.9899 | 19923 | ADD | -0.0416 | 0.01143 |
| TProt | rs34069958 | 6 | 413536 | A | -5.227 | 8.62e-08 | 0.03175 | IRF4 | protein_coding | Total protein | blood | No | 6 | 413536 | G | A | 0.1748 | 0.992 | 19923 | ADD | -0.04139 | 0.01148 |
| TProt | rs12176562 | 6 | 416359 | T | -5.184 | 1.09e-07 | 0.04 | IRF4 | protein_coding | Total protein | blood | No | 6 | 416359 | G | T | 0.1753 | 0.9911 | 19923 | ADD | -0.04051 | 0.01147 |
| FRONTAL.AXIS.P | rs6967128 | 7 | 12767684 | G | 5.381 | 3.70e-08 | 0.0125 | ARL4A | protein_coding | P wave axis | cardiovascular | Yes | 7 | 12767684 | A | G | 0.1607 | 1.005 | 6062 | ADD | 0.08578 | 0.02552 |
| FRONTAL.AXIS.P | rs6967288 | 7 | 12767770 | C | 5.364 | 4.08e-08 | 0.01378 | ARL4A | protein_coding | P wave axis | cardiovascular | Yes | 7 | 12767770 | A | C | 0.1604 | 1.001 | 6062 | ADD | 0.08782 | 0.0256 |
| FRONTAL.AXIS.TERMINAL.40.MS | rs938906 | 2 | 231555684 | T | -5.175 | 1.14e-07 | 0.03789 | AC098823.3 | lincRNA | Frontal axis terminal 40 ms | cardiovascular | No | 2 | 231555684 | C | T | 0.2081 | 0.9804 | 6296 | ADD | -0.07802 | 0.02281 |
| HORIZONTAL.AXIS..TERMINAL.40.MS | rs7921016 | 10 | 20301670 | G | 5.255 | 7.39e-08 | 0.02438 | PLXDC2 | protein_coding | QRS axis | cardiovascular | No | 10 | 20301670 | A | G | 0.4816 | 0.9969 | 6205 | ADD | 0.07217 | 0.01801 |
| HORIZONTAL.AXIS..TERMINAL.40.MS | rs7910531 | 10 | 20302141 | A | 5.128 | 1.46e-07 | 0.04832 | PLXDC2 | protein_coding | QRS axis | cardiovascular | No | 10 | 20302141 | G | A | 0.3399 | 0.9636 | 6205 | ADD | 0.08217 | 0.01932 |
| P.HORIZONTAL.AXIS | rs7995471 | 13 | 44888742 | T | 5.589 | 1.15e-08 | 0.003812 | SERP2 | protein_coding | P wave axis horizontal plane | cardiovascular | Yes | 13 | 44888742 | C | T | 0.06236 | 0.8965 | 5960 | ADD | 0.1409 | 0.04164 |

| lab | SNP | CHR | BP | A1 | tstat | P | adjP | Gene_Symbol | biotype | full_name | group | genome_wide | CHROM | GENPOS | ALLELE0 | ALLELE1 | male_A1FREQ | male_INFO | male_N | male_TEST | male_BETA | male_SE |
|---|---|---|---|---|---|---|---|---|---|---|---|---|---|---|---|---|---|---|---|---|---|---|
| PR.INTERVAL | rs6937361 | 6 | 123317677 | A | 5.27 | 6.82e-08 | 0.02257 | CLVS2 | protein_coding | PR Interval mean | cardiovascular | No | 6 | 123317677 | G | A | 0.4901 | 0.9715 | 6106 | ADD | 0.08476 | 0.01977 |
| QRS.DURATION | rs6581228 | 12 | 40015286 | C | -5.228 | 8.56e-08 | 0.02928 | ABCD2 | protein_coding | QRS duration | cardiovascular | No | 12 | 40015286 | T | C | 0.1592 | 0.9972 | 6316 | ADD | -0.1074 | 0.02571 |
| QRS.DURATION | rs11172794 | 12 | 39998833 | C | -5.164 | 1.21e-07 | 0.04148 | ABCD2 | protein_coding | QRS duration | cardiovascular | No | 12 | 39998833 | T | C | 0.1401 | 1.006 | 6316 | ADD | -0.1078 | 0.02699 |
| UricA | rs9994216 | 4 | 9984541 | G | 5.309 | 5.51e-08 | 0.01801 | SLC2A9 | protein_coding | Urate [Mass/volume] in Serum or Plasma | urinary | No | 4 | 9984541 | T | G | 0.235 | 1.005 | 6411 | ADD | -0.1192 | 0.01882 |
| UricA | rs1014290 | 4 | 10001861 | G | 5.282 | 6.39e-08 | 0.02088 | SLC2A9 | protein_coding | Urate [Mass/volume] in Serum or Plasma | urinary | No | 4 | 10001861 | A | G | 0.2284 | 0.9977 | 6411 | ADD | -0.116 | 0.01905 |
| UricA | rs3733588 | 4 | 9997303 | G | 5.272 | 6.76e-08 | 0.02211 | SLC2A9 | protein_coding | Urate [Mass/volume] in Serum or Plasma | urinary | No | 4 | 9997303 | A | G | 0.2286 | 0.9973 | 6411 | ADD | -0.1163 | 0.01905 |
| UricA | rs3775947 | 4 | 9995240 | C | 5.271 | 6.80e-08 | 0.02224 | SLC2A9 | protein_coding | Urate [Mass/volume] in Serum or Plasma | urinary | No | 4 | 9995240 | T | C | 0.231 | 0.9901 | 6411 | ADD | -0.114 | 0.01906 |
| UricA | rs13129697 | 4 | 9926967 | G | 5.247 | 7.74e-08 | 0.0253 | SLC2A9 | protein_coding | Urate [Mass/volume] in Serum or Plasma | urinary | No | 4 | 9926967 | T | G | 0.2573 | 0.9921 | 6411 | ADD | -0.114 | 0.01836 |
| UricA | rs7679724 | 4 | 9985376 | G | 5.238 | 8.14e-08 | 0.0266 | SLC2A9 | protein_coding | Urate [Mass/volume] in Serum or Plasma | urinary | No | 4 | 9985376 | T | G | 0.2411 | 1.008 | 6411 | ADD | -0.111 | 0.01863 |
| UricA | rs7660895 | 4 | 9985445 | A | 5.185 | 1.08e-07 | 0.03529 | SLC2A9 | protein_coding | Urate [Mass/volume] in Serum or Plasma | urinary | No | 4 | 9985445 | G | A | 0.2408 | 1.009 | 6411 | ADD | -0.1115 | 0.01863 |
| UricA | rs11723742 | 4 | 10111572 | G | 5.171 | 1.17e-07 | 0.03816 | WDR1 | protein_coding | Urate [Mass/volume] in Serum or Plasma | urinary | No | 4 | 10111572 | A | G | 0.2102 | 1.012 | 6411 | ADD | -0.1001 | 0.01948 |
| UricA | rs4529048 | 4 | 9997112 | C | 5.161 | 1.23e-07 | 0.04021 | SLC2A9 | protein_coding | Urate [Mass/volume] in Serum or Plasma | urinary | No | 4 | 9997112 | A | C | 0.2335 | 0.996 | 6411 | ADD | -0.114 | 0.01893 |
| UricA | rs62288518 | 4 | 10087464 | C | 5.13 | 1.45e-07 | 0.0473 | WDR1 | protein_coding | Urate [Mass/volume] in Serum or Plasma | urinary | No | 4 | 10087464 | T | C | 0.2008 | 1.017 | 6411 | ADD | -0.107 | 0.01976 |
| UricA | rs3775946 | 4 | 9995256 | A | 5.126 | 1.48e-07 | 0.04846 | SLC2A9 | protein_coding | Urate [Mass/volume] in Serum or Plasma | urinary | No | 4 | 9995256 | G | A | 0.2244 | 0.995 | 6411 | ADD | -0.1142 | 0.01921 |
| UricA | rs7663032 | 4 | 9993838 | C | 5.123 | 1.50e-07 | 0.04911 | SLC2A9 | protein_coding | Urate [Mass/volume] in Serum or Plasma | urinary | No | 4 | 9993838 | T | C | 0.2277 | 1.002 | 6411 | ADD | -0.1114 | 0.01905 |
| MONOA | rs3128734 | 10 | 43451430 | C | -5.351 | 4.38e-08 | 0.01446 | RP11-124O11.2 | lincRNA | Monocytes [#/volume] in Blood | immune/inflammatory | Yes | 10 | 43451430 | T | C | 0.2644 | 0.9978 | 5259 | ADD | -0.06116 | 0.0203 |
| R.VUMC.AN.RESPIRATION.RATE | rs55813256 | 17 | 74498386 | G | -5.163 | 1.22e-07 | 0.04066 | RHBDF2 | protein_coding | Respiratory rate | vitals | No | 17 | 74498386 | T | G | 0.06366 | 0.9706 | 4501 | ADD | -0.1183 | 0.04248 |
| R.VUMC.AN.RESPIRATION.RATE | rs4648329 | 17 | 74498342 | T | -5.152 | 1.29e-07 | 0.04313 | RHBDF2 | protein_coding | Respiratory rate | vitals | No | 17 | 74498342 | C | T | 0.06366 | 0.9705 | 4501 | ADD | -0.1182 | 0.04248 |
| X25DTot | rs731844 | 7 | 34150264 | C | 5.993 | 1.03e-09 | 3.45e-04 | BMPER | protein_coding | 25-Hydroxyvitamin D3+25-Hydroxyvitamin D2 [Mass/volume] in Serum or Plasma | metabolic | Yes | 7 | 34150264 | T | C | 0.4303 | 0.9796 | 2828 | ADD | 0.1269 | 0.02461 |
| X25DTot | rs9655014 | 7 | 34148632 | T | 5.974 | 1.16e-09 | 3.86e-04 | BMPER | protein_coding | 25-Hydroxyvitamin D3+25-Hydroxyvitamin D2 [Mass/volume] in Serum or Plasma | metabolic | Yes | 7 | 34148632 | C | T | 0.4206 | 0.9789 | 2828 | ADD | 0.1278 | 0.02471 |
| X25DTot | rs10228731 | 7 | 34150782 | G | 5.973 | 1.16e-09 | 3.89e-04 | BMPER | protein_coding | 25-Hydroxyvitamin D3+25-Hydroxyvitamin D2 [Mass/volume] in Serum or Plasma | metabolic | Yes | 7 | 34150782 | T | G | 0.4208 | 0.9776 | 2828 | ADD | 0.1278 | 0.02472 |
| X25DTot | rs10253869 | 7 | 34149922 | A | 5.971 | 1.18e-09 | 3.93e-04 | BMPER | protein_coding | 25-Hydroxyvitamin D3+25-Hydroxyvitamin D2 [Mass/volume] in Serum or Plasma | metabolic | Yes | 7 | 34149922 | G | A | 0.4204 | 0.9774 | 2828 | ADD | 0.1278 | 0.02473 |

| lab | SNP | CHR | BP | A1 | tstat | P | adjP | Gene_Symbol | biotype | full_name | group | genome_wide | CHROM | GENPOS | ALLELE0 | ALLELE1 | male_A1FREQ | male_INFO | male_N | male_TEST | male_BETA | male_SE |
|---|---|---|---|---|---|---|---|---|---|---|---|---|---|---|---|---|---|---|---|---|---|---|
| X25DTot | rs10224203 | 7 | 34149593 | C | 5.971 | 1.18e-09 | 3.94e-04 | BMPER | protein_coding | 25-Hydroxyvitamin D3+25-Hydroxyvitamin D2 [Mass/volume] in Serum or Plasma | metabolic | Yes | 7 | 34149593 | T | C | 0.4206 | 0.979 | 2828 | ADD | 0.1277 | 0.0247 |
| X25DTot | rs1362460 | 7 | 34147964 | C | 5.969 | 1.19e-09 | 3.98e-04 | BMPER | protein_coding | 25-Hydroxyvitamin D3+25-Hydroxyvitamin D2 [Mass/volume] in Serum or Plasma | metabolic | Yes | 7 | 34147964 | T | C | 0.4229 | 1.01 | 2828 | ADD | 0.1246 | 0.0243 |
| X25DTot | rs10229510 | 7 | 34151389 | G | 5.965 | 1.22e-09 | 4.09e-04 | BMPER | protein_coding | 25-Hydroxyvitamin D3+25-Hydroxyvitamin D2 [Mass/volume] in Serum or Plasma | metabolic | Yes | 7 | 34151389 | A | G | 0.4206 | 0.9779 | 2828 | ADD | 0.1276 | 0.02472 |
| X25DTot | rs4723351 | 7 | 34155813 | T | 5.941 | 1.42e-09 | 4.74e-04 | BMPER | protein_coding | 25-Hydroxyvitamin D3+25-Hydroxyvitamin D2 [Mass/volume] in Serum or Plasma | metabolic | Yes | 7 | 34155813 | G | T | 0.4199 | 0.9782 | 2828 | ADD | 0.1289 | 0.02472 |
| X25DTot | rs6462531 | 7 | 34157746 | G | 5.933 | 1.48e-09 | 4.96e-04 | BMPER | protein_coding | 25-Hydroxyvitamin D3+25-Hydroxyvitamin D2 [Mass/volume] in Serum or Plasma | metabolic | Yes | 7 | 34157746 | A | G | 0.4199 | 0.9773 | 2828 | ADD | 0.1283 | 0.02473 |
| X25DTot | rs918038 | 7 | 34153768 | A | 5.914 | 1.67e-09 | 5.57e-04 | BMPER | protein_coding | 25-Hydroxyvitamin D3+25-Hydroxyvitamin D2 [Mass/volume] in Serum or Plasma | metabolic | Yes | 7 | 34153768 | G | A | 0.4119 | 1.012 | 2828 | ADD | 0.1261 | 0.02438 |
| X25DTot | rs918037 | 7 | 34192011 | G | 5.49 | 2.01e-08 | 0.006698 | BMPER | protein_coding | 25-Hydroxyvitamin D3+25-Hydroxyvitamin D2 [Mass/volume] in Serum or Plasma | metabolic | Yes | 7 | 34192011 | A | G | 0.4232 | 1.006 | 2828 | ADD | 0.1117 | 0.02435 |
| IRF | rs62596093 | 23 | 57050631 | T | -5.317 | 5.26e-08 | 0.01756 | SPIN3 | protein_coding | Immature reticulocytes/Reticulocytes.total in Blood | blood | No | 23 | 57050631 | C | T | 0.07593 | 1.683 | 2822 | ADD | -0.126 | 0.03563 |
| IRF | rs137975667 | 23 | 57171004 | G | -5.275 | 6.63e-08 | 0.02211 | SPIN2A | protein_coding | Immature reticulocytes/Reticulocytes.total in Blood | blood | No | 23 | 57171004 | A | G | 0.07723 | 1.725 | 2822 | ADD | -0.1287 | 0.03494 |
| FEF25_75_PRD | rs11208 | 12 | 133161346 | C | -5.204 | 9.77e-08 | 0.03352 | FBRSL1 | protein_coding | FEF PRD | lung | No | 12 | 133161346 | T | C | 0.1776 | 1.035 | 2375 | ADD | -0.1123 | 0.03736 |
| UGluc | rs6969646 | 7 | 26533803 | T | 5.189 | 1.06e-07 | 0.03485 | AC004540.5 | lincRNA | Glucose urine dipstick | urinary | No | 7 | 26533803 | C | T | 0.15 | 0.9753 | 2730 | ADD | 0.1089 | 0.03373 |
| UGluc | rs7811559 | 7 | 26541157 | A | 5.173 | 1.15e-07 | 0.03782 | AC004540.5 | lincRNA | Glucose urine dipstick | urinary | No | 7 | 26541157 | G | A | 0.1505 | 0.9697 | 2730 | ADD | 0.1095 | 0.03377 |
| AlbCre | rs757066 | 23 | 15608952 | C | 5.148 | 1.32e-07 | 0.04272 | ACE2 | protein_coding | Microalbumin/Creatinine [Mass Ratio] in 24 hour Urine | liver | No | 23 | 15608952 | T | C | 0.3811 | 1.969 | 2218 | ADD | 0.07596 | 0.02209 |
| HBSA. | rs4668506 | 2 | 6908119 | T | 5.648 | 8.12e-09 | 0.002706 | LINC00487 | lincRNA | Hepatitis B virus surface Ab [Presence] in Serum | immune/inflammatory | Yes | 2 | 6908119 | C | T | 0.4845 | 1.013 | 2001 | ADD | 0.06917 | 0.02683 |
| HBSA. | rs11661287 | 18 | 71063187 | T | 5.129 | 1.46e-07 | 0.04852 | CTD-2354A18.1 | processed_transcript | Hepatitis B virus surface Ab [Presence] in Serum | immune/inflammatory | No | 18 | 71063187 | C | T | 0.1758 | 1.014 | 2001 | ADD | 0.137 | 0.03567 |
| C4Quan | rs12335173 | 8 | 142562065 | T | -5.424 | 2.92e-08 | 0.009869 | AC138647.1 | protein_coding | Complement C4 [Mass/volume] in Serum or Plasma | immune/inflammatory | Yes | 8 | 142562065 | C | T | 0.4694 | 1.021 | 1374 | ADD | -0.1186 | 0.03363 |
| C4Quan | rs11167070 | 8 | 142566709 | A | 5.196 | 1.02e-07 | 0.03436 | AC138647.1 | protein_coding | Complement C4 [Mass/volume] in Serum or Plasma | immune/inflammatory | No | 8 | 142566709 | C | A | 0.4936 | 1.008 | 1374 | ADD | 0.1211 | 0.03381 |
| C4Quan | rs10448124 | 8 | 142562270 | T | 5.182 | 1.10e-07 | 0.03713 | AC138647.1 | protein_coding | Complement C4 [Mass/volume] in Serum or Plasma | immune/inflammatory | No | 8 | 142562270 | C | T | 0.4935 | 1.01 | 1374 | ADD | 0.1202 | 0.03377 |
| C4Quan | rs7819351 | 8 | 142561839 | G | 5.182 | 1.10e-07 | 0.03714 | AC138647.1 | protein_coding | Complement C4 [Mass/volume] in Serum or Plasma | immune/inflammatory | No | 8 | 142561839 | A | G | 0.4935 | 1.01 | 1374 | ADD | 0.1202 | 0.03377 |
| C4Quan | rs2085839 | 8 | 142559685 | C | 5.182 | 1.10e-07 | 0.03716 | AC138647.1 | protein_coding | Complement C4 [Mass/volume] in Serum or Plasma | immune/inflammatory | No | 8 | 142559685 | T | C | 0.4935 | 1.01 | 1374 | ADD | 0.1201 | 0.03377 |
| C4Quan | rs7835035 | 8 | 142561809 | T | 5.182 | 1.10e-07 | 0.03716 | AC138647.1 | protein_coding | Complement C4 [Mass/volume] in Serum or Plasma | immune/inflammatory | No | 8 | 142561809 | G | T | 0.4935 | 1.01 | 1374 | ADD | 0.1201 | 0.03377 |

| lab | SNP | CHR | BP | A1 | tstat | P | adjP | Gene_Symbol | biotype | full_name | group | genome_wide | CHROM | GENPOS | ALLELE0 | ALLELE1 | male_A1FREQ | male_INFO | male_N | male_TEST | male_BETA | male_SE |
|---|---|---|---|---|---|---|---|---|---|---|---|---|---|---|---|---|---|---|---|---|---|---|
| DLCOOVA_PRD | rs4765821 | 12 | 1611846 | T | 5.584 | 1.18e-08 | 0.004023 | LINC00942 | lincRNA | DLCOOVA_PRD | lung | Yes | 12 | 1611846 | G | T | 0.3469 | 1.025 | 1954 | ADD | 0.06768 | 0.0295 |
| DLCOOVA_PRD | rs10773948 | 12 | 1612770 | C | 5.304 | 5.65e-08 | 0.0193 | LINC00942 | lincRNA | DLCOOVA_PRD | lung | No | 12 | 1612770 | T | C | 0.3344 | 1.015 | 1954 | ADD | 0.0574 | 0.02988 |
| DLCOOVA_PRD | rs4765822 | 12 | 1611894 | G | 5.242 | 7.96e-08 | 0.02719 | LINC00942 | lincRNA | DLCOOVA_PRD | lung | No | 12 | 1611894 | A | G | 0.331 | 1.03 | 1954 | ADD | 0.05626 | 0.02973 |
| DLCOOVA_PRD | rs7965881 | 12 | 1612090 | A | 5.216 | 9.16e-08 | 0.03127 | LINC00942 | lincRNA | DLCOOVA_PRD | lung | No | 12 | 1612090 | G | A | 0.3344 | 1.013 | 1954 | ADD | 0.05748 | 0.0299 |
| FNucCl | rs9635769 | 17 | 33288363 | C | -5.822 | 2.92e-09 | 9.67e-04 | CCT6B | protein_coding | Nucleated cells [#/volume] in Synovial fluid by Manual count | blood | Yes | 17 | 33288363 | T | C | 0.4258 | 0.9767 | 1952 | ADD | -0.1178 | 0.02997 |
| lab | SNP | CHR | BP | A1 | tstat | P | adjP | Gene_Symbol | biotype | full_name | group | genome_wide | CHROM | GENPOS | ALLELE0 | ALLELE1 | male_A1FREQ | male_INFO | male_N | male_TEST | male_BETA | male_SE |
| FNucCl | rs8073366 | 17 | 33260938 | G | -5.817 | 3.00e-09 | 9.94e-04 | CCT6B | protein_coding | Nucleated cells [#/volume] in Synovial fluid by Manual count | blood | Yes | 17 | 33260938 | A | G | 0.4252 | 0.9713 | 1952 | ADD | -0.1178 | 0.03006 |
| FNucCl | rs1482103 | 17 | 33287651 | A | -5.8 | 3.32e-09 | 0.001101 | CCT6B | protein_coding | Nucleated cells [#/volume] in Synovial fluid by Manual count | blood | Yes | 17 | 33287651 | G | A | 0.4232 | 0.9779 | 1952 | ADD | -0.1169 | 0.02998 |
| POCA1C | rs35819527 | 12 | 106361242 | C | 5.144 | 1.35e-07 | 0.04509 | ST13P3 | pseudogene | Hemoglobin A1c (Glycated) | blood | No | 12 | 106361242 | T | C | 0.4196 | 0.955 | 1866 | ADD | 0.1102 | 0.02881 |
| FEF25_75_PRL | rs1757387 | 13 | 104351911 | G | 5.168 | 1.19e-07 | 0.04038 | ATP6V1G1P7 | pseudogene | FEF PRL | lung | No | 13 | 104351911 | A | G | 0.266 | 0.9366 | 1581 | ADD | 0.1192 | 0.03777 |
| FEV1OFVC_PPR | rs45627133 | 20 | 48805769 | A | 5.142 | 1.36e-07 | 0.04644 | CEBPB | protein_coding | FEV1/FVC PPR | lung | No | 20 | 48805769 | G | A | 0.07243 | 0.9521 | 1604 | ADD | 0.2864 | 0.0663 |
| FEV1OFVC_PRD | rs28845518 | 3 | 88820652 | T | 5.174 | 1.15e-07 | 0.03792 | NDUFA5P5 | pseudogene | FEV1/FVC PRD | lung | No | 3 | 88820652 | C | T | 0.4505 | 0.9944 | 1604 | ADD | 0.1734 | 0.03485 |
| FEV1OFVC_PRD | rs11719154 | 3 | 88815878 | A | 5.174 | 1.15e-07 | 0.03795 | NDUFA5P5 | pseudogene | FEV1/FVC PRD | lung | No | 3 | 88815878 | G | A | 0.4528 | 0.9978 | 1604 | ADD | 0.1712 | 0.03476 |
| FEV1OFVC_PRD | rs1601106 | 3 | 88829421 | T | 5.159 | 1.24e-07 | 0.04106 | NARG2P2 | pseudogene | FEV1/FVC PRD | lung | No | 3 | 88829421 | C | T | 0.4503 | 0.9935 | 1604 | ADD | 0.1732 | 0.03487 |
| FEV1OFVC_PRD | rs1601108 | 3 | 88828983 | T | 5.159 | 1.24e-07 | 0.04106 | NARG2P2 | pseudogene | FEV1/FVC PRD | lung | No | 3 | 88828983 | C | T | 0.4503 | 0.9935 | 1604 | ADD | 0.1732 | 0.03487 |
| FEV1OFVC_PRD | rs9832849 | 3 | 88827372 | T | 5.146 | 1.33e-07 | 0.04392 | NARG2P2 | pseudogene | FEV1/FVC PRD | lung | No | 3 | 88827372 | C | T | 0.45 | 0.9944 | 1604 | ADD | 0.1726 | 0.03486 |
| FEV1OFVC_PRD | rs9810286 | 3 | 88812659 | A | 5.13 | 1.45e-07 | 0.04794 | NDUFA5P5 | pseudogene | FEV1/FVC PRD | lung | No | 3 | 88812659 | G | A | 0.4503 | 0.9938 | 1604 | ADD | 0.1713 | 0.03485 |
| FEV1OFVC_PRD | rs4254665 | 3 | 88803614 | A | 5.127 | 1.47e-07 | 0.04864 | NDUFA5P5 | pseudogene | FEV1/FVC PRD | lung | No | 3 | 88803614 | C | A | 0.4504 | 0.994 | 1604 | ADD | 0.1722 | 0.03484 |
| FEV1OFVC_PRD | rs1483473 | 3 | 88804168 | C | 5.124 | 1.49e-07 | 0.04933 | NDUFA5P5 | pseudogene | FEV1/FVC PRD | lung | No | 3 | 88804168 | A | C | 0.4506 | 0.9951 | 1604 | ADD | 0.1722 | 0.03482 |
| FEV1OFVC_PRL | rs2972332 | 5 | 76494268 | A | -5.365 | 4.06e-08 | 0.01319 | PDE8B | protein_coding | FEV1/FVC PRL | lung | Yes | 5 | 76494268 | C | A | 0.09411 | 1.019 | 1604 | ADD | -0.2725 | 0.06201 |
| FEV1OFVC_ZBD | rs45627133 | 20 | 48805769 | A | 5.183 | 1.09e-07 | 0.03702 | CEBPB | protein_coding | FEV1/FVC ZBD | lung | No | 20 | 48805769 | G | A | 0.07243 | 0.9521 | 1604 | ADD | 0.2859 | 0.06562 |
| Prolac | rs10003580 | 4 | 136536342 | A | -5.184 | 1.08e-07 | 0.03594 | TARS2P1 | pseudogene | Prolactin [Mass/volume] in Serum or Plasma | endocrine | No | 4 | 136536342 | G | A | 0.3131 | 1.033 | 964 | ADD | -0.1795 | 0.03915 |

| lab | SNP | CHR | BP | A1 | tstat | P | adjP | Gene_Symbol | biotype | full_name | group | genome_wide | CHROM | GENPOS | ALLELE0 | ALLELE1 | male_A1FREQ | male_INFO | male_N | male_TEST | male_BETA | male_SE |
|---|---|---|---|---|---|---|---|---|---|---|---|---|---|---|---|---|---|---|---|---|---|---|
| R.AN.ESOPHAGEAL.TEMPERATURE | rs35906209 | 5 | 38726470 | T | -5.132 | 1.44e-07 | 0.04726 | RP11-122C5.1 | lincRNA | Temperature - Distal airway | vitals | No | 5 | 38726470 | C | T | 0.1172 | 0.9695 | 1304 | ADD | -0.2137 | 0.05516 |
| AFPTum | rs1294941 | 7 | 45216646 | C | -5.189 | 1.06e-07 | 0.03518 | RAMP3 | protein_coding | Alpha-1-fetoprotein.tumor marker [Mass/volume] in Serum or Plasma | cancer | No | 7 | 45216646 | T | C | 0.4852 | 1.007 | 1721 | ADD | -0.06902 | 0.02929 |
| AFPTum | rs1294942 | 7 | 45216634 | T | -5.178 | 1.12e-07 | 0.0373 | RAMP3 | protein_coding | Alpha-1-fetoprotein.tumor marker [Mass/volume] in Serum or Plasma | cancer | No | 7 | 45216634 | C | T | 0.4814 | 0.9938 | 1721 | ADD | -0.06897 | 0.02949 |
| CRBC | rs708209 | 12 | 31554082 | G | 5.127 | 1.48e-07 | 0.04819 | DENND5B | protein_coding | Erythrocytes in Cerebral spinal fluid | other | No | 12 | 31554082 | A | G | 0.4783 | 0.9659 | 1072 | ADD | 0.1441 | 0.03682 |
| CRBC | rs793154 | 12 | 31565261 | A | 5.124 | 1.49e-07 | 0.04882 | DENND5B | protein_coding | Erythrocytes in Cerebral spinal fluid | other | No | 12 | 31565261 | G | A | 0.4786 | 0.9669 | 1072 | ADD | 0.1435 | 0.0368 |
| CRBC | rs793157 | 12 | 31567998 | A | 5.124 | 1.50e-07 | 0.04887 | DENND5B | protein_coding | Erythrocytes in Cerebral spinal fluid | other | No | 12 | 31567998 | G | A | 0.4786 | 0.9669 | 1072 | ADD | 0.1435 | 0.0368 |
| FEF25_75_PPR | rs57727667 | 4 | 77855208 | G | -5.398 | 3.37e-08 | 0.01119 | SEPT11 | protein_coding | FEF PPR | lung | Yes | 4 | 77855208 | A | G | 0.1268 | 0.9829 | 1210 | ADD | -0.235 | 0.05685 |
| FEF25_75_PPR | rs112359859 | 4 | 77858905 | T | -5.384 | 3.64e-08 | 0.0121 | SEPT11 | protein_coding | FEF PPR | lung | Yes | 4 | 77858905 | C | T | 0.1279 | 0.9814 | 1210 | ADD | -0.2366 | 0.05669 |
| FEV1_PRD | rs3105110 | 9 | 107859488 | A | -5.429 | 2.83e-08 | 0.009605 | RN7SKP191 | misc_RNA | FEV1 PRD | lung | Yes | 9 | 107859488 | G | A | 0.4218 | 0.9578 | 1214 | ADD | -0.1031 | 0.03559 |
| FEV1_PRD | rs2937368 | 9 | 107865861 | T | -5.338 | 4.70e-08 | 0.01591 | RN7SKP191 | misc_RNA | FEV1 PRD | lung | Yes | 9 | 107865861 | C | T | 0.3685 | 0.9466 | 1214 | ADD | -0.1161 | 0.03664 |
| FEV1_PRD | rs10991543 | 9 | 107863681 | A | -5.249 | 7.65e-08 | 0.02594 | RN7SKP191 | misc_RNA | FEV1 PRD | lung | No | 9 | 107863681 | G | A | 0.3678 | 0.9538 | 1214 | ADD | -0.1121 | 0.03651 |
| FVC_PRD | rs3105110 | 9 | 107859488 | A | -5.438 | 2.69e-08 | 0.009102 | RN7SKP191 | misc_RNA | FVC PRD | lung | Yes | 9 | 107859488 | G | A | 0.4218 | 0.9578 | 1214 | ADD | -0.1002 | 0.03501 |
| FVC_PRD | rs2937368 | 9 | 107865861 | T | -5.35 | 4.40e-08 | 0.01489 | RN7SKP191 | misc_RNA | FVC PRD | lung | Yes | 9 | 107865861 | C | T | 0.3685 | 0.9466 | 1214 | ADD | -0.114 | 0.03605 |
| FVC_PRD | rs10991543 | 9 | 107863681 | A | -5.262 | 7.11e-08 | 0.02407 | RN7SKP191 | misc_RNA | FVC PRD | lung | No | 9 | 107863681 | G | A | 0.3678 | 0.9538 | 1214 | ADD | -0.1101 | 0.03593 |
| Amyl | rs4595382 | 1 | 19720151 | G | 5.217 | 9.09e-08 | 0.03074 | CAPZB | protein_coding | Amylase [Enzymatic activity/volume] in Serum or Plasma | metabolic | No | 1 | 19720151 | T | G | 0.4454 | 0.9916 | 3776 | ADD | 0.06589 | 0.02258 |
| Meta | rs16997108 | 21 | 40357481 | A | 5.52 | 1.70e-08 | 0.005573 | AF064858.8 | lincRNA | Metamyelocytes/100 leukocytes in Blood by Manual count | blood | Yes | 21 | 40357481 | G | A | 0.0812 | 0.9946 | 3764 | ADD | 0.1996 | 0.03931 |
| Meta | rs4887309 | 15 | 88154744 | A | -5.482 | 2.10e-08 | 0.006897 | RP11-648K4.2 | lincRNA | Metamyelocytes/100 leukocytes in Blood by Manual count | blood | Yes | 15 | 88154744 | G | A | 0.3758 | 1.006 | 3764 | ADD | -0.07041 | 0.02207 |
| Meta | rs4887310 | 15 | 88154870 | A | -5.482 | 2.11e-08 | 0.006928 | RP11-648K4.2 | lincRNA | Metamyelocytes/100 leukocytes in Blood by Manual count | blood | Yes | 15 | 88154870 | G | A | 0.3759 | 1.006 | 3764 | ADD | -0.07038 | 0.02207 |
| Meta | rs4887308 | 15 | 88154706 | A | -5.479 | 2.14e-08 | 0.007023 | RP11-648K4.2 | lincRNA | Metamyelocytes/100 leukocytes in Blood by Manual count | blood | Yes | 15 | 88154706 | G | A | 0.376 | 1.004 | 3764 | ADD | -0.07019 | 0.02209 |
| Meta | rs6496427 | 15 | 88153004 | A | -5.288 | 6.18e-08 | 0.02031 | RP11-648K4.2 | lincRNA | Metamyelocytes/100 leukocytes in Blood by Manual count | blood | No | 15 | 88153004 | G | A | 0.3676 | 1.004 | 3764 | ADD | -0.0648 | 0.0222 |
| Meta | rs7350778 | 15 | 88152921 | A | -5.245 | 7.83e-08 | 0.02572 | RP11-648K4.2 | lincRNA | Metamyelocytes/100 leukocytes in Blood by Manual count | blood | No | 15 | 88152921 | C | A | 0.3697 | 1.005 | 3764 | ADD | -0.06463 | 0.02215 |
| Meta | rs4887190 | 15 | 88152928 | T | -5.244 | 7.88e-08 | 0.02587 | RP11-648K4.2 | lincRNA | Metamyelocytes/100 leukocytes in Blood by Manual count | blood | No | 15 | 88152928 | C | T | 0.3683 | 1.005 | 3764 | ADD | -0.06551 | 0.02217 |
| Meta | rs12905303 | 15 | 88153040 | C | -5.154 | 1.28e-07 | 0.04188 | RP11-648K4.2 | lincRNA | Metamyelocytes/100 leukocytes in Blood by Manual count | blood | No | 15 | 88153040 | T | C | 0.3814 | 1 | 3764 | ADD | -0.06615 | 0.02207 |
| Meta | rs7172845 | 15 | 88140936 | A | -5.146 | 1.33e-07 | 0.04362 | RP11-648K4.2 | lincRNA | Metamyelocytes/100 leukocytes in Blood by Manual count | blood | No | 15 | 88140936 | C | A | 0.3292 | 1.001 | 3764 | ADD | -0.06529 | 0.0228 |
| PCR | rs9856864 | 3 | 69762267 | A | 5.19 | 1.05e-07 | 0.03463 | MITF | protein_coding | Protein/Creatinine [Mass ratio] in Urine | kidney | No | 3 | 69762267 | G | A | 0.2233 | 1.01 | 2909 | ADD | 0.1076 | 0.02706 |

| lab | SNP | CHR | BP | A1 | tstat | P | adjP | Gene_Symbol | biotype | full_name | group | genome_wide | CHROM | GENPOS | ALLELE0 | ALLELE1 | male_A1FREQ | male_INFO | male_N | male_TEST | male_BETA | male_SE |
|---|---|---|---|---|---|---|---|---|---|---|---|---|---|---|---|---|---|---|---|---|---|---|
| LucAbs | rs28592167 | 7 | 100738224 | G | 5.158 | 1.25e-07 | 0.04118 | RP11-395B7.7 | sense_overlapping | Leukocytes [#/volume] in Blood by Automated count | blood | No | 7 | 100738224 | A | G | 0.09641 | 0.9939 | 2514 | ADD | 0.1948 | 0.04141 |
| LucAbs | rs10275377 | 7 | 100736712 | G | 5.15 | 1.30e-07 | 0.04285 | RP11-395B7.7 | sense_overlapping | Leukocytes [#/volume] in Blood by Automated count | blood | No | 7 | 100736712 | A | G | 0.09642 | 0.9911 | 2514 | ADD | 0.1938 | 0.04147 |
| LucAbs | rs10246242 | 7 | 100736928 | T | 5.15 | 1.30e-07 | 0.04289 | RP11-395B7.7 | sense_overlapping | Leukocytes [#/volume] in Blood by Automated count | blood | No | 7 | 100736928 | C | T | 0.09641 | 0.9911 | 2514 | ADD | 0.1938 | 0.04147 |
| ACAIgG | rs2287051 | 2 | 40547424 | C | 5.128 | 1.47e-07 | 0.04788 | SLC8A1 | protein_coding | Cardiolipin IgG Ab [Units/volume] in Serum by Immunoassay | immune/inflammatory | No | 2 | 40547424 | T | C | 0.2592 | 0.9877 | 675 | ADD | 0.2343 | 0.05542 |
| IC_PPR | rs34332103 | 1 | 238472852 | G | 5.208 | 9.56e-08 | 0.03167 | RNU6-725P | snRNA | IC PPR | lung | No | 1 | 238472852 | A | G | 0.4625 | 1.005 | 1168 | ADD | 0.1633 | 0.03739 |
| VC_PPR | rs72721845 | 4 | 150119475 | A | -5.246 | 7.77e-08 | 0.02538 | RP11-24I21.1 | lincRNA | VC PPR | lung | No | 4 | 150119475 | G | A | 0.07107 | 0.9059 | 1145 | ADD | -0.3395 | 0.07895 |
| VC_ZBD | rs13329905 | 16 | 88095095 | C | 5.244 | 7.87e-08 | 0.02619 | BANP | protein_coding | VC ZBD | lung | No | 16 | 88095095 | T | C | 0.1256 | 0.9956 | 1145 | ADD | 0.23 | 0.05376 |
| VC_ZBD | rs72818549 | 16 | 88085718 | C | 5.216 | 9.16e-08 | 0.03047 | BANP | protein_coding | VC ZBD | lung | No | 16 | 88085718 | T | C | 0.1249 | 0.9856 | 1145 | ADD | 0.2368 | 0.05417 |
| TUCrea | rs7685306 | 4 | 189663354 | C | -5.523 | 1.66e-08 | 0.005419 | RP11-756P10.3 | pseudogene | Creatinine [Mass/volume] in Urine | kidney | Yes | 4 | 189663354 | T | C | 0.2338 | 0.9661 | 1069 | ADD | -0.1582 | 0.04504 |
| TUCrea | rs10006229 | 4 | 189664412 | G | -5.51 | 1.79e-08 | 0.005826 | RP11-756P10.3 | pseudogene | Creatinine [Mass/volume] in Urine | kidney | Yes | 4 | 189664412 | A | G | 0.2335 | 0.964 | 1069 | ADD | -0.1581 | 0.04511 |
| CLymph | rs10254310 | 7 | 101063059 | G | -5.236 | 8.23e-08 | 0.02671 | COL26A1 | polymorphic_pseudogene | Lymphocytes/100 leukocytes in Cerebral spinal fluid by Manual count | immune/inflammatory | No | 7 | 101063059 | A | G | 0.1544 | 0.9826 | 881 | ADD | -0.2362 | 0.05757 |
| CLymph | rs7788624 | 7 | 101061317 | C | -5.174 | 1.14e-07 | 0.03714 | COL26A1 | polymorphic_pseudogene | Lymphocytes/100 leukocytes in Cerebral spinal fluid by Manual count | immune/inflammatory | No | 7 | 101061317 | A | C | 0.1605 | 0.9369 | 881 | ADD | -0.2362 | 0.05804 |
| R.ED.PRE.ARRIVAL.EMS.O2.SAT | rs11188661 | 10 | 97960993 | A | -5.388 | 3.57e-08 | 0.01149 | BLNK | protein_coding | Oxygen saturation in Arterial blood by Pulse oximetry | vitals | Yes | 10 | 97960993 | G | A | 0.3278 | 1 | 981 | ADD | -0.1734 | 0.04638 |
| R.ED.PRE.ARRIVAL.EMS.O2.SAT | rs10882743 | 10 | 97961748 | A | -5.379 | 3.76e-08 | 0.0121 | BLNK | protein_coding | Oxygen saturation in Arterial blood by Pulse oximetry | vitals | Yes | 10 | 97961748 | G | A | 0.3154 | 1.003 | 981 | ADD | -0.1706 | 0.04676 |
| ViARet | rs11786514 | 8 | 18787795 | G | -5.359 | 4.18e-08 | 0.01376 | PSD3 | protein_coding | Retinol [Mass/volume] in Serum or Plasma | metabolic | Yes | 8 | 18787795 | T | G | 0.2522 | 0.9761 | 795 | ADD | -0.1858 | 0.05117 |
| X.WBC | rs11650090 | 17 | 32776598 | T | -5.448 | 2.55e-08 | 0.008361 | AC011193.1 | lincRNA | # of WBC's | blood | Yes | 17 | 32776598 | C | T | 0.4487 | 0.9934 | 761 | ADD | -0.1681 | 0.04291 |
| TLC_PPR | rs13962 | 1 | 159175527 | A | -5.197 | 1.01e-07 | 0.03381 | DARC | protein_coding | TLC PPR | lung | No | 1 | 159175527 | G | A | 0.1744 | 1.068 | 869 | ADD | -0.1517 | 0.04997 |
| R.AN.MEAN.ARTERIAL.BLOOD.PRESSURE | rs1347823 | 2 | 148304924 | G | -5.208 | 9.53e-08 | 0.03172 | RNU6-715P | snRNA | Invasive Mean blood pressure | vitals | No | 2 | 148304924 | A | G | 0.2818 | 1.024 | 1168 | ADD | -0.1305 | 0.04081 |
| R.AN.MEAN.ARTERIAL.BLOOD.PRESSURE | rs17427741 | 4 | 38413462 | C | 5.165 | 1.20e-07 | 0.04 | RP11-83C7.2 | lincRNA | Invasive Mean blood pressure | vitals | No | 4 | 38413462 | T | C | 0.3354 | 0.9935 | 1168 | ADD | 0.1478 | 0.0396 |

| lab | SNP | CHR | BP | A1 | tstat | P | adjP | Gene_Symbol | biotype | full_name | group | genome_wide | CHROM | GENPOS | ALLELE0 | ALLELE1 | male_A1FREQ | male_INFO | male_N | male_TEST | male_BETA | male_SE |
|---|---|---|---|---|---|---|---|---|---|---|---|---|---|---|---|---|---|---|---|---|---|---|
| R.AN.MEAN.ARTERIAL.BLOOD.PRESSURE | rs4524073 | 2 | 148301416 | T | -5.144 | 1.34e-07 | 0.04474 | RNU6-715P | snRNA | Invasive Mean blood pressure | vitals | No | 2 | 148301416 | C | T | 0.289 | 1.002 | 1168 | ADD | -0.1282 | 0.04093 |
| R.AN.MEAN.ARTERIAL.BLOOD.PRESSURE | rs1344948 | 2 | 148311124 | T | -5.13 | 1.45e-07 | 0.04828 | RNU6-715P | snRNA | Invasive Mean blood pressure | vitals | No | 2 | 148311124 | G | T | 0.2854 | 1.009 | 1168 | ADD | -0.1259 | 0.04097 |
| R.AN.MEAN.ARTERIAL.BLOOD.PRESSURE | rs1487622 | 4 | 38414088 | T | 5.126 | 1.48e-07 | 0.04935 | RP11-83C7.2 | lincRNA | Invasive Mean blood pressure | vitals | No | 4 | 38414088 | C | T | 0.3354 | 0.9934 | 1168 | ADD | 0.1479 | 0.0396 |
| R.AN.MEAN.ARTERIAL.BLOOD.PRESSURE | rs901703 | 4 | 38413846 | T | 5.125 | 1.49e-07 | 0.04956 | RP11-83C7.2 | lincRNA | Invasive Mean blood pressure | vitals | No | 4 | 38413846 | C | T | 0.3329 | 1.004 | 1168 | ADD | 0.1473 | 0.03946 |
| TU.Pro | rs56043319 | 7 | 10247107 | A | 5.617 | 9.74e-09 | 0.003188 | AC006373.1 | lincRNA | 24 hour urinary protein | urinary | Yes | 7 | 10247107 | G | A | 0.2295 | 1.006 | 784 | ADD | 0.2343 | 0.04848 |
| TU.Pro | rs2351831 | 7 | 10251935 | C | 5.596 | 1.10e-08 | 0.003597 | AC006373.1 | lincRNA | 24 hour urinary protein | urinary | Yes | 7 | 10251935 | A | C | 0.23 | 1.009 | 784 | ADD | 0.2301 | 0.04837 |
| TU.Pro | rs2189951 | 7 | 10252234 | T | 5.588 | 1.15e-08 | 0.003752 | AC006373.1 | lincRNA | 24 hour urinary protein | urinary | Yes | 7 | 10252234 | C | T | 0.23 | 1.009 | 784 | ADD | 0.2301 | 0.04837 |
| TU.Pro | rs73056839 | 7 | 10256513 | G | 5.588 | 1.15e-08 | 0.003755 | HSPA8P8 | pseudogene | 24 hour urinary protein | urinary | Yes | 7 | 10256513 | A | G | 0.2295 | 1.01 | 784 | ADD | 0.2301 | 0.04838 |
| TU.Pro | rs73056835 | 7 | 10256268 | T | 5.584 | 1.18e-08 | 0.00385 | HSPA8P8 | pseudogene | 24 hour urinary protein | urinary | Yes | 7 | 10256268 | C | T | 0.2302 | 1.01 | 784 | ADD | 0.2299 | 0.04833 |
| TU.Pro | rs73056804 | 7 | 10251476 | T | 5.576 | 1.23e-08 | 0.004027 | AC006373.1 | lincRNA | 24 hour urinary protein | urinary | Yes | 7 | 10251476 | C | T | 0.2293 | 1.011 | 784 | ADD | 0.2305 | 0.04838 |
| TU.Pro | rs2189952 | 7 | 10252414 | T | 5.574 | 1.24e-08 | 0.00407 | AC006373.1 | lincRNA | 24 hour urinary protein | urinary | Yes | 7 | 10252414 | C | T | 0.2302 | 1.01 | 784 | ADD | 0.2299 | 0.04833 |
| TU.Pro | rs2189953 | 7 | 10252437 | T | 5.56 | 1.35e-08 | 0.004418 | AC006373.1 | lincRNA | 24 hour urinary protein | urinary | Yes | 7 | 10252437 | C | T | 0.2306 | 1.008 | 784 | ADD | 0.2288 | 0.04837 |
| TU.Pro | rs9692549 | 7 | 10245522 | A | 5.138 | 1.39e-07 | 0.04544 | AC006373.1 | lincRNA | 24 hour urinary protein | urinary | No | 7 | 10245522 | G | A | 0.1714 | 1.034 | 784 | ADD | 0.2259 | 0.05329 |
| TU.Pro | rs9692033 | 7 | 10245417 | G | 5.127 | 1.47e-07 | 0.04822 | AC006373.1 | lincRNA | 24 hour urinary protein | urinary | No | 7 | 10245417 | T | G | 0.1716 | 1.035 | 784 | ADD | 0.2258 | 0.05325 |
| FProt1 | rs4843400 | 16 | 86617795 | C | -5.533 | 1.57e-08 | 0.005098 | FOXL1 | protein_coding | Protein body fluid | metabolic | Yes | 16 | 86617795 | T | C | 0.4754 | 1.01 | 990 | ADD | -0.1139 | 0.03422 |
| FProt1 | rs34128649 | 1 | 106144478 | A | -5.212 | 9.34e-08 | 0.03024 | RP11-251P6.1 | lincRNA | Protein body fluid | metabolic | No | 1 | 106144478 | G | A | 0.3658 | 0.9699 | 990 | ADD | -0.1531 | 0.03645 |
| FProt1 | rs12049618 | 1 | 106147471 | G | -5.211 | 9.40e-08 | 0.03045 | RP11-251P6.1 | lincRNA | Protein body fluid | metabolic | No | 1 | 106147471 | T | G | 0.3658 | 0.9699 | 990 | ADD | -0.1531 | 0.03644 |
| FProt1 | rs35945222 | 1 | 106144184 | T | -5.211 | 9.41e-08 | 0.03046 | RP11-251P6.1 | lincRNA | Protein body fluid | metabolic | No | 1 | 106144184 | C | T | 0.3658 | 0.9699 | 990 | ADD | -0.1531 | 0.03644 |
| FProt1 | rs12029662 | 1 | 106140823 | T | -5.21 | 9.46e-08 | 0.03064 | RP11-251P6.1 | lincRNA | Protein body fluid | metabolic | No | 1 | 106140823 | C | T | 0.3659 | 0.97 | 990 | ADD | -0.153 | 0.03644 |
| FProt1 | rs1916821 | 1 | 106143126 | T | -5.21 | 9.46e-08 | 0.03064 | RP11-251P6.1 | lincRNA | Protein body fluid | metabolic | No | 1 | 106143126 | G | T | 0.3658 | 0.9701 | 990 | ADD | -0.153 | 0.03644 |

| lab | SNP | CHR | BP | A1 | tstat | P | adjP | Gene_Symbol | biotype | full_name | group | genome_wide | CHROM | GENPOS | ALLELE0 | ALLELE1 | male_A1FREQ | male_INFO | male_N | male_TEST | male_BETA | male_SE |
|---|---|---|---|---|---|---|---|---|---|---|---|---|---|---|---|---|---|---|---|---|---|---|
| FProt1 | rs6129996 | 20 | 40611969 | T | 5.203 | 9.80e-08 | 0.03174 | RP5-1121H13.4 | antisense | Protein body fluid | metabolic | No | 20 | 40611969 | C | T | 0.1944 | 1.002 | 990 | ADD | 0.1303 | 0.04386 |
| FProt1 | rs12730762 | 1 | 106153292 | T | -5.191 | 1.04e-07 | 0.03381 | RP11-251P6.1 | lincRNA | Protein body fluid | metabolic | No | 1 | 106153292 | C | T | 0.3664 | 0.9711 | 990 | ADD | -0.1528 | 0.03641 |
| FProt1 | rs11184648 | 1 | 106155888 | A | -5.191 | 1.05e-07 | 0.03393 | RP11-251P6.1 | lincRNA | Protein body fluid | metabolic | No | 1 | 106155888 | G | A | 0.3663 | 0.9709 | 990 | ADD | -0.1529 | 0.03642 |
| FProt1 | rs12024416 | 1 | 106156618 | G | -5.191 | 1.05e-07 | 0.03397 | RP11-251P6.1 | lincRNA | Protein body fluid | metabolic | No | 1 | 106156618 | A | G | 0.3663 | 0.9709 | 990 | ADD | -0.1529 | 0.03642 |
| FProt1 | rs9787052 | 1 | 106151776 | C | -5.19 | 1.05e-07 | 0.03405 | RP11-251P6.1 | lincRNA | Protein body fluid | metabolic | No | 1 | 106151776 | T | C | 0.3664 | 0.9708 | 990 | ADD | -0.1526 | 0.03642 |
| FProt1 | rs12044441 | 1 | 106134909 | T | -5.17 | 1.17e-07 | 0.03795 | RP11-251P6.1 | lincRNA | Protein body fluid | metabolic | No | 1 | 106134909 | G | T | 0.3648 | 0.9684 | 990 | ADD | -0.1506 | 0.0365 |
| FProt1 | rs11184643 | 1 | 106139540 | A | -5.17 | 1.17e-07 | 0.03802 | RP11-251P6.1 | lincRNA | Protein body fluid | metabolic | No | 1 | 106139540 | G | A | 0.3647 | 0.9686 | 990 | ADD | -0.1505 | 0.03649 |
| FProt1 | rs11184644 | 1 | 106139802 | A | -5.169 | 1.17e-07 | 0.03802 | RP11-251P6.1 | lincRNA | Protein body fluid | metabolic | No | 1 | 106139802 | G | A | 0.3647 | 0.9686 | 990 | ADD | -0.1505 | 0.03649 |
| FProt1 | rs10465777 | 1 | 106131731 | G | -5.158 | 1.25e-07 | 0.04036 | RP11-251P6.1 | lincRNA | Protein body fluid | metabolic | No | 1 | 106131731 | A | G | 0.3645 | 0.9645 | 990 | ADD | -0.1502 | 0.03658 |
| FProt1 | rs2037608 | 1 | 106136729 | G | -5.122 | 1.52e-07 | 0.04909 | RP11-251P6.1 | lincRNA | Protein body fluid | metabolic | No | 1 | 106136729 | A | G | 0.3647 | 0.9679 | 990 | ADD | -0.1505 | 0.03651 |
| RVOTLC_PRU | rs7976210 | 12 | 108502142 | G | 5.536 | 1.55e-08 | 0.005079 | WSCD2 | protein_coding | RVOTLC PRU | lung | Yes | 12 | 108502142 | A | G | 0.459 | 0.9699 | 926 | ADD | 0.1188 | 0.03956 |
| TIGIgG | rs197347 | 7 | 36422110 | T | -5.192 | 1.04e-07 | 0.03373 | KIAA0895 | protein_coding | Tissue transglutaminase IgG Ab [Units/volume] in Serum by Immunoassay | immune/inflammatory | No | 7 | 36422110 | G | T | 0.1659 | 0.9697 | 266 | ADD | -0.429 | 0.09759 |
| TIGIgG | rs60579881 | 6 | 23641124 | G | 5.154 | 1.28e-07 | 0.04139 | RP4-810F7.1 | pseudogene | Tissue transglutaminase IgG Ab [Units/volume] in Serum by Immunoassay | immune/inflammatory | No | 6 | 23641124 | A | G | 0.1049 | 0.9582 | 266 | ADD | 0.4241 | 0.115 |
| TIGIgG | rs75121997 | 6 | 23639696 | G | 5.135 | 1.41e-07 | 0.04586 | RP4-810F7.1 | pseudogene | Tissue transglutaminase IgG Ab [Units/volume] in Serum by Immunoassay | immune/inflammatory | No | 6 | 23639696 | A | G | 0.1053 | 0.9622 | 266 | ADD | 0.4229 | 0.1147 |
| Blasts | rs9637184 | 21 | 45991038 | G | 5.262 | 7.13e-08 | 0.0233 | TSPEAR | protein_coding | Blasts/100 leukocytes in Blood by Manual count | blood | No | 21 | 45991038 | A | G | 0.1959 | 1.017 | 867 | ADD | 0.1673 | 0.05198 |
| Blasts | rs11910575 | 21 | 45985047 | A | 5.204 | 9.74e-08 | 0.03179 | TSPEAR | protein_coding | Blasts/100 leukocytes in Blood by Manual count | blood | No | 21 | 45985047 | G | A | 0.1996 | 1.012 | 867 | ADD | 0.1711 | 0.05176 |
| Blasts | rs11911565 | 21 | 45984833 | C | 5.199 | 1.00e-07 | 0.03264 | TSPEAR | protein_coding | Blasts/100 leukocytes in Blood by Manual count | blood | No | 21 | 45984833 | T | C | 0.2006 | 1.021 | 867 | ADD | 0.1679 | 0.05141 |
| Blasts | rs9983521 | 21 | 45988906 | T | 5.158 | 1.25e-07 | 0.04079 | TSPEAR | protein_coding | Blasts/100 leukocytes in Blood by Manual count | blood | No | 21 | 45988906 | C | T | 0.197 | 1.017 | 867 | ADD | 0.1658 | 0.05187 |
| ABS_BASO_CT | rs1553938 | 11 | 106290444 | C | 5.676 | 6.90e-09 | 0.002274 | RP11-680E19.2 | pseudogene | Basophils [#/volume] in Blood by Automated count | blood | Yes | 11 | 106290444 | T | C | 0.1678 | 0.9479 | 638 | ADD | 0.3178 | 0.06525 |
| ABS_BASO_CT | rs975394 | 11 | 106292302 | A | 5.672 | 7.04e-09 | 0.002321 | RP11-680E19.2 | pseudogene | Basophils [#/volume] in Blood by Automated count | blood | Yes | 11 | 106292302 | G | A | 0.1703 | 0.9539 | 638 | ADD | 0.3106 | 0.06468 |
| ABS_BASO_CT | rs953816 | 11 | 106288554 | C | 5.651 | 7.99e-09 | 0.002636 | RP11-680E19.2 | pseudogene | Basophils [#/volume] in Blood by Automated count | blood | Yes | 11 | 106288554 | T | C | 0.1702 | 0.9607 | 638 | ADD | 0.3138 | 0.06446 |
| ABS_BASO_CT | rs2458326 | 11 | 106291464 | G | 5.534 | 1.56e-08 | 0.005154 | RP11-680E19.2 | pseudogene | Basophils [#/volume] in Blood by Automated count | blood | Yes | 11 | 106291464 | A | G | 0.1679 | 0.9483 | 638 | ADD | 0.3055 | 0.06526 |
| ABS_BASO_CT | rs975393 | 11 | 106292338 | C | 5.501 | 1.88e-08 | 0.006211 | RP11-680E19.2 | pseudogene | Basophils [#/volume] in Blood by Automated count | blood | Yes | 11 | 106292338 | A | C | 0.1805 | 0.963 | 638 | ADD | 0.2868 | 0.06278 |
| ABS_BASO_CT | rs2458343 | 11 | 106307233 | T | 5.45 | 2.53e-08 | 0.008326 | RP11-680E19.2 | pseudogene | Basophils [#/volume] in Blood by Automated count | blood | Yes | 11 | 106307233 | G | T | 0.1751 | 0.9505 | 638 | ADD | 0.2727 | 0.06391 |

| lab | SNP | CHR | BP | A1 | tstat | P | adjP | Gene_Symbol | biotype | full_name | group | genome_wide | CHROM | GENPOS | ALLELE0 | ALLELE1 | male_A1FREQ | male_INFO | male_N | male_TEST | male_BETA | male_SE |
|---|---|---|---|---|---|---|---|---|---|---|---|---|---|---|---|---|---|---|---|---|---|---|
| ABS_BASO_CT | rs2458348 | 11 | 106299661 | G | 5.444 | 2.61e-08 | 0.008597 | RP11-680E19.2 | pseudogene | Basophils [#/volume] in Blood by Automated count | blood | Yes | 11 | 106299661 | A | G | 0.1756 | 0.973 | 638 | ADD | 0.2679 | 0.06309 |
| ABS_BASO_CT | rs2513600 | 11 | 106299999 | C | 5.437 | 2.70e-08 | 0.008916 | RP11-680E19.2 | pseudogene | Basophils [#/volume] in Blood by Automated count | blood | Yes | 11 | 106299999 | T | C | 0.1734 | 0.9541 | 638 | ADD | 0.2723 | 0.06402 |
| ABS_BASO_CT | rs907648 | 11 | 106297486 | G | 5.413 | 3.11e-08 | 0.01024 | RP11-680E19.2 | pseudogene | Basophils [#/volume] in Blood by Automated count | blood | Yes | 11 | 106297486 | A | G | 0.1776 | 0.9683 | 638 | ADD | 0.2696 | 0.06298 |
| ABS_BASO_CT | rs975395 | 11 | 106292043 | A | 5.402 | 3.29e-08 | 0.01086 | RP11-680E19.2 | pseudogene | Basophils [#/volume] in Blood by Automated count | blood | Yes | 11 | 106292043 | G | A | 0.1786 | 0.9665 | 638 | ADD | 0.2766 | 0.06293 |
| ABS_BASO_CT | rs2942229 | 11 | 106293058 | C | 5.398 | 3.37e-08 | 0.0111 | RP11-680E19.2 | pseudogene | Basophils [#/volume] in Blood by Automated count | blood | Yes | 11 | 106293058 | A | C | 0.1774 | 0.9701 | 638 | ADD | 0.2693 | 0.06296 |
| ABS_BASO_CT | rs1512435 | 11 | 106307111 | T | 5.359 | 4.19e-08 | 0.01381 | RP11-680E19.2 | pseudogene | Basophils [#/volume] in Blood by Automated count | blood | Yes | 11 | 106307111 | G | T | 0.1736 | 0.9545 | 638 | ADD | 0.266 | 0.06397 |
| ABS_BASO_CT | rs2513614 | 11 | 106301973 | C | 5.352 | 4.36e-08 | 0.01436 | RP11-680E19.2 | pseudogene | Basophils [#/volume] in Blood by Automated count | blood | Yes | 11 | 106301973 | A | C | 0.1743 | 0.954 | 638 | ADD | 0.2652 | 0.06394 |
| ABS_BASO_CT | rs2513609 | 11 | 106315651 | A | 5.243 | 7.89e-08 | 0.02603 | RP11-680E19.2 | pseudogene | Basophils [#/volume] in Blood by Automated count | blood | No | 11 | 106315651 | G | A | 0.1732 | 0.954 | 638 | ADD | 0.2632 | 0.06404 |
| ABS_BASO_CT | rs4471339 | 10 | 90879234 | G | 5.152 | 1.29e-07 | 0.0426 | RP11-341B24.3 | pseudogene | Basophils [#/volume] in Blood by Automated count | blood | No | 10 | 90879234 | T | G | 0.05192 | 0.9688 | 638 | ADD | 0.4403 | 0.1082 |
| CNeut | rs141057315 | 23 | 124411728 | T | 5.147 | 1.32e-07 | 0.04307 | AL953885.1 | miRNA | Neutrophils/100 leukocytes in Cerebral spinal fluid by Manual count | immune/inflammatory | No | 23 | 124411728 | G | T | 0.2828 | 1.575 | 622 | ADD | 0.106 | 0.04122 |
| MmAcid | rs10510786 | 3 | 56491294 | G | 5.483 | 2.09e-08 | 0.006977 | ERC2 | protein_coding | Methylmalonic acid | metabolic | Yes | 3 | 56491294 | A | G | 0.3087 | 1.053 | 689 | ADD | 0.1783 | 0.05126 |
| MmAcid | rs11719646 | 3 | 56467691 | G | 5.482 | 2.11e-08 | 0.007041 | ERC2 | protein_coding | Methylmalonic acid | metabolic | Yes | 3 | 56467691 | A | G | 0.3462 | 1.048 | 689 | ADD | 0.1402 | 0.04983 |
| MmAcid | rs11708250 | 3 | 56481923 | C | 5.359 | 4.19e-08 | 0.01399 | ERC2 | protein_coding | Methylmalonic acid | metabolic | Yes | 3 | 56481923 | A | C | 0.3044 | 1.052 | 689 | ADD | 0.1662 | 0.05149 |
| MmAcid | rs6801539 | 3 | 56477803 | C | 5.327 | 4.99e-08 | 0.01667 | ERC2 | protein_coding | Methylmalonic acid | metabolic | Yes | 3 | 56477803 | T | C | 0.3045 | 1.052 | 689 | ADD | 0.1662 | 0.05148 |
| MmAcid | rs17825463 | 3 | 56484387 | A | 5.312 | 5.43e-08 | 0.01814 | ERC2 | protein_coding | Methylmalonic acid | metabolic | No | 3 | 56484387 | C | A | 0.3043 | 1.052 | 689 | ADD | 0.1664 | 0.0515 |
| MmAcid | rs11712094 | 3 | 56459364 | C | 5.294 | 5.98e-08 | 0.01997 | ERC2 | protein_coding | Methylmalonic acid | metabolic | No | 3 | 56459364 | T | C | 0.3052 | 1.048 | 689 | ADD | 0.1644 | 0.05155 |

| lab | SNP | CHR | BP | A1 | tstat | P | adjP | Gene_Symbol | biotype | full_name | group | genome_wide | CHROM | GENPOS | ALLELE0 | ALLELE1 | male_A1FREQ | male_INFO | male_N | male_TEST | male_BETA | male_SE |
|---|---|---|---|---|---|---|---|---|---|---|---|---|---|---|---|---|---|---|---|---|---|---|
| MmAcid | rs55833786 | 3 | 56466327 | C | 5.29 | 6.13e-08 | 0.02046 | ERC2 | protein_coding | Methylmalonic acid | metabolic | No | 3 | 56466327 | T | C | 0.3402 | 1.041 | 689 | ADD | 0.1295 | 0.05021 |
| MmAcid | rs11715404 | 3 | 56462386 | C | 5.248 | 7.67e-08 | 0.02562 | ERC2 | protein_coding | Methylmalonic acid | metabolic | No | 3 | 56462386 | T | C | 0.3394 | 1.042 | 689 | ADD | 0.1271 | 0.05021 |
| MmAcid | rs72875345 | 3 | 56478290 | T | 5.246 | 7.77e-08 | 0.02595 | ERC2 | protein_coding | Methylmalonic acid | metabolic | No | 3 | 56478290 | C | T | 0.3394 | 1.043 | 689 | ADD | 0.1251 | 0.0502 |
| MmAcid | rs11923602 | 3 | 56481177 | A | 5.235 | 8.25e-08 | 0.02755 | ERC2 | protein_coding | Methylmalonic acid | metabolic | No | 3 | 56481177 | C | A | 0.3393 | 1.042 | 689 | ADD | 0.1246 | 0.05021 |
| MmAcid | rs55783589 | 3 | 56497961 | G | 5.208 | 9.57e-08 | 0.03196 | ERC2 | protein_coding | Methylmalonic acid | metabolic | No | 3 | 56497961 | A | G | 0.3411 | 1.039 | 689 | ADD | 0.1282 | 0.0502 |
| MmAcid | rs57352922 | 3 | 56486642 | A | 5.168 | 1.19e-07 | 0.03962 | ERC2 | protein_coding | Methylmalonic acid | metabolic | No | 3 | 56486642 | G | A | 0.3392 | 1.042 | 689 | ADD | 0.1248 | 0.05022 |
| CREATININE | rs809474 | 20 | 19727276 | T | 5.185 | 1.08e-07 | 0.03497 | AL121761.2 | protein_coding | Creatinine | kidney | No | 20 | 19727276 | C | T | 0.2364 | 0.9838 | 755 | ADD | 0.2756 | 0.0521 |

| lab | SNP | CHR | BP | A1 | tstat | P | adjP | Gene_Symbol | biotype | full_name | group | genome_wide | CHROM | GENPOS | ALLELE0 | ALLELE1 | male_A1FREQ | male_INFO | male_N | male_TEST | male_BETA | male_SE |
|---|---|---|---|---|---|---|---|---|---|---|---|---|---|---|---|---|---|---|---|---|---|---|
| KapLCQ | rs10147215 | 14 | 94481458 | A | 5.469 | 2.26e-08 | 0.007433 | LINC00521 | processed_transcript | Kappa light chains.free [Mass/volume] in Serum | toxicology/pharmacology | Yes | 14 | 94481458 | C | A | 0.293 | 1.002 | 640 | ADD | 0.2057 | 0.04789 |
| KapLCQ | rs28552817 | 14 | 94476839 | C | 5.363 | 4.10e-08 | 0.01349 | LINC00521 | processed_transcript | Kappa light chains.free [Mass/volume] in Serum | toxicology/pharmacology | Yes | 14 | 94476839 | T | C | 0.3172 | 0.9875 | 640 | ADD | 0.2068 | 0.04707 |
| KapLCQ | rs112678733 | 15 | 84223085 | G | -5.299 | 5.83e-08 | 0.01918 | SH3GL3 | protein_coding | Kappa light chains.free [Mass/volume] in Serum | toxicology/pharmacology | No | 15 | 84223085 | A | G | 0.2538 | 0.975 | 640 | ADD | -0.1588 | 0.05055 |
| KapLCQ | rs7160644 | 14 | 94479804 | A | 5.265 | 7.01e-08 | 0.02308 | LINC00521 | processed_transcript | Kappa light chains.free [Mass/volume] in Serum | toxicology/pharmacology | No | 14 | 94479804 | G | A | 0.3109 | 0.9936 | 640 | ADD | 0.201 | 0.04723 |
| KapLCQ | rs4905142 | 14 | 94480348 | C | 5.263 | 7.10e-08 | 0.02338 | LINC00521 | processed_transcript | Kappa light chains.free [Mass/volume] in Serum | toxicology/pharmacology | No | 14 | 94480348 | T | C | 0.3102 | 0.9894 | 640 | ADD | 0.2014 | 0.04736 |
| KapLCQ | rs12592318 | 15 | 84219700 | A | -5.26 | 7.22e-08 | 0.02378 | SH3GL3 | protein_coding | Kappa light chains.free [Mass/volume] in Serum | toxicology/pharmacology | No | 15 | 84219700 | G | A | 0.252 | 0.9774 | 640 | ADD | -0.1616 | 0.05061 |
| KapLCQ | rs17585513 | 15 | 84240242 | T | -5.129 | 1.46e-07 | 0.04796 | SH3GL3 | protein_coding | Kappa light chains.free [Mass/volume] in Serum | toxicology/pharmacology | No | 15 | 84240242 | C | T | 0.2524 | 0.9804 | 640 | ADD | -0.1617 | 0.05049 |
| LamLCQ | rs61829349 | 1 | 197729197 | T | 5.934 | 1.48e-09 | 4.83e-04 | DENND1B | protein_coding | Lambda light chains.free [Mass/volume] in Serum or Plasma | immune/inflammatory | Yes | 1 | 197729197 | C | T | 0.08518 | 0.9828 | 639 | ADD | 0.3602 | 0.07346 |
| LamLCQ | rs55792017 | 1 | 197769890 | A | 5.912 | 1.69e-09 | 5.52e-04 | DENND1B | protein_coding | Lambda light chains.free [Mass/volume] in Serum or Plasma | immune/inflammatory | Yes | 1 | 197769890 | G | A | 0.08791 | 1.008 | 639 | ADD | 0.3467 | 0.07155 |
| LamLCQ | rs61829344 | 1 | 197683734 | C | 5.505 | 1.85e-08 | 0.006037 | DENND1B | protein_coding | Lambda light chains.free [Mass/volume] in Serum or Plasma | immune/inflammatory | Yes | 1 | 197683734 | A | C | 0.08786 | 0.9979 | 639 | ADD | 0.336 | 0.07202 |
| LamLCQ | rs61829440 | 1 | 197439044 | A | 5.44 | 2.67e-08 | 0.008713 | CRB1 | protein_coding | Lambda light chains.free [Mass/volume] in Serum or Plasma | immune/inflammatory | Yes | 1 | 197439044 | G | A | 0.08926 | 1.036 | 639 | ADD | 0.325 | 0.07026 |
| LamLCQ | rs3804511 | 6 | 52287195 | A | -5.287 | 6.20e-08 | 0.02026 | EFHC1 | protein_coding | Lambda light chains.free [Mass/volume] in Serum or Plasma | immune/inflammatory | No | 6 | 52287195 | G | A | 0.2823 | 0.9881 | 639 | ADD | -0.1558 | 0.04463 |
| LamLCQ | rs3804510 | 6 | 52287196 | A | -5.278 | 6.52e-08 | 0.0213 | EFHC1 | protein_coding | Lambda light chains.free [Mass/volume] in Serum or Plasma | immune/inflammatory | No | 6 | 52287196 | C | A | 0.2824 | 0.9883 | 639 | ADD | -0.1557 | 0.04462 |
| LamLCQ | rs2273120 | 6 | 52355206 | T | -5.2 | 9.98e-08 | 0.03258 | EFHC1 | protein_coding | Lambda light chains.free [Mass/volume] in Serum or Plasma | immune/inflammatory | No | 6 | 52355206 | C | T | 0.3206 | 0.9992 | 639 | ADD | -0.1425 | 0.04306 |
| eGFR | rs2099274 | 14 | 59217658 | T | -5.446 | 2.58e-08 | 0.008353 | AL121819.1 | miRNA | eGFR | kidney | Yes | 14 | 59217658 | C | T | 0.07937 | 1.012 | 680 | ADD | -0.3774 | 0.08491 |
| eGFR | rs11850622 | 14 | 59230660 | T | -5.379 | 3.75e-08 | 0.01217 | AL121819.1 | miRNA | eGFR | kidney | Yes | 14 | 59230660 | C | T | 0.07877 | 1.002 | 680 | ADD | -0.3666 | 0.08575 |
| eGFR | rs17095059 | 14 | 59224969 | A | -5.343 | 4.57e-08 | 0.01482 | AL121819.1 | miRNA | eGFR | kidney | Yes | 14 | 59224969 | C | A | 0.07856 | 1.014 | 680 | ADD | -0.367 | 0.0853 |
| eGFR | rs7664009 | 4 | 138864270 | G | -5.174 | 1.14e-07 | 0.03706 | LINC00616 | lincRNA | eGFR | kidney | No | 4 | 138864270 | T | G | 0.4193 | 0.9919 | 680 | ADD | -0.1486 | 0.04787 |
| eGFR | rs7695489 | 4 | 138864247 | T | -5.174 | 1.15e-07 | 0.03721 | LINC00616 | lincRNA | eGFR | kidney | No | 4 | 138864247 | C | T | 0.4193 | 0.9919 | 680 | ADD | -0.1486 | 0.04787 |
| eGFR | rs6815810 | 4 | 138850891 | C | -5.149 | 1.31e-07 | 0.0424 | LINC00616 | lincRNA | eGFR | kidney | No | 4 | 138850891 | T | C | 0.4198 | 0.9944 | 680 | ADD | -0.1479 | 0.04781 |
| eGFR | rs4640713 | 4 | 138853076 | G | -5.148 | 1.32e-07 | 0.04266 | LINC00616 | lincRNA | eGFR | kidney | No | 4 | 138853076 | T | G | 0.421 | 0.9993 | 680 | ADD | -0.1474 | 0.04767 |
| eGFR | rs7658015 | 4 | 138855571 | A | -5.143 | 1.35e-07 | 0.04377 | LINC00616 | lincRNA | eGFR | kidney | No | 4 | 138855571 | G | A | 0.4199 | 0.9951 | 680 | ADD | -0.1479 | 0.04779 |
| eGFR | rs17296269 | 4 | 138850236 | C | -5.143 | 1.35e-07 | 0.0438 | LINC00616 | lincRNA | eGFR | kidney | No | 4 | 138850236 | T | C | 0.4199 | 0.9951 | 680 | ADD | -0.1479 | 0.04779 |

| lab | SNP | CHR | BP | A1 | tstat | P | adjP | Gene_Symbol | biotype | full_name | group | genome_wide | CHROM | GENPOS | ALLELE0 | ALLELE1 | male_A1FREQ | male_INFO | male_N | male_TEST | male_BETA | male_SE |
|---|---|---|---|---|---|---|---|---|---|---|---|---|---|---|---|---|---|---|---|---|---|---|
| eGFR | rs7699753 | 4 | 138858226 | C | -5.143 | 1.35e-07 | 0.04384 | LINC00616 | lincRNA | eGFR | kidney | No | 4 | 138858226 | A | C | 0.4199 | 0.9951 | 680 | ADD | -0.1478 | 0.04779 |
| eGFR | rs7698174 | 4 | 138857345 | G | -5.142 | 1.36e-07 | 0.04398 | LINC00616 | lincRNA | eGFR | kidney | No | 4 | 138857345 | A | G | 0.4199 | 0.9948 | 680 | ADD | -0.1478 | 0.04779 |
| HomocB | rs35219779 | 23 | 4907860 | A | -5.186 | 1.07e-07 | 0.03514 | MTND6P12 | pseudogene | Homocysteine in Serum or Plasma | metabolic | No | 23 | 4907860 | G | A | 0.06367 | 1.937 | 538 | ADD | -0.2847 | 0.07406 |
| FET_AFD | rs1738899 | 14 | 97535923 | A | 5.603 | 1.05e-08 | 0.003406 | RP11-907D1.1 | lincRNA | FET AFD | lung | Yes | 14 | 97535923 | G | A | 0.0497 | 1.03 | 503 | ADD | 0.5969 | 0.1291 |
| FET_AFD | rs2629514 | 14 | 97536467 | G | 5.603 | 1.06e-08 | 0.003412 | RP11-907D1.1 | lincRNA | FET AFD | lung | Yes | 14 | 97536467 | A | G | 0.04967 | 1.029 | 503 | ADD | 0.5976 | 0.1292 |
| FEV1OFVC_AFD | rs144602263 | 23 | 150109020 | T | 5.15 | 1.31e-07 | 0.04369 | RNU6-383P | snRNA | FEV1/FVC AFD | lung | No | 23 | 150109020 | C | T | 0.06002 | 1.89 | 506 | ADD | 0.2418 | 0.08442 |
| CPepti | rs9400518 | 6 | 112388358 | G | -5.379 | 3.74e-08 | 0.0124 | WISP3 | protein_coding | C peptide [Mass/volume] in Serum or Plasma | endocrine | Yes | 6 | 112388358 | T | G | 0.1513 | 0.9315 | 476 | ADD | -0.3975 | 0.08392 |
| CPepti | rs9374307 | 6 | 112397878 | C | -5.357 | 4.24e-08 | 0.01405 | TUBE1 | protein_coding | C peptide [Mass/volume] in Serum or Plasma | endocrine | Yes | 6 | 112397878 | T | C | 0.1488 | 0.9382 | 476 | ADD | -0.3916 | 0.08405 |
| CPepti | rs9400519 | 6 | 112402709 | G | -5.352 | 4.35e-08 | 0.01442 | TUBE1 | protein_coding | C peptide [Mass/volume] in Serum or Plasma | endocrine | Yes | 6 | 112402709 | A | G | 0.1492 | 0.9405 | 476 | ADD | -0.3907 | 0.08387 |
| CPepti | rs9384817 | 6 | 112364361 | A | -5.273 | 6.73e-08 | 0.02229 | WISP3 | protein_coding | C peptide [Mass/volume] in Serum or Plasma | endocrine | No | 6 | 112364361 | G | A | 0.1478 | 0.9518 | 476 | ADD | -0.3747 | 0.0837 |
| CPepti | rs9398296 | 6 | 112363842 | T | -5.267 | 6.92e-08 | 0.02293 | WISP3 | protein_coding | C peptide [Mass/volume] in Serum or Plasma | endocrine | No | 6 | 112363842 | C | T | 0.1478 | 0.9518 | 476 | ADD | -0.3748 | 0.0837 |
| CPepti | rs9384816 | 6 | 112363749 | C | -5.256 | 7.37e-08 | 0.02442 | WISP3 | protein_coding | C peptide [Mass/volume] in Serum or Plasma | endocrine | No | 6 | 112363749 | T | C | 0.1488 | 0.9496 | 476 | ADD | -0.3734 | 0.08354 |
| CPepti | rs9487799 | 6 | 112363807 | A | -5.256 | 7.37e-08 | 0.02442 | WISP3 | protein_coding | C peptide [Mass/volume] in Serum or Plasma | endocrine | No | 6 | 112363807 | G | A | 0.1488 | 0.9496 | 476 | ADD | -0.3734 | 0.08354 |
| CPepti | rs9384821 | 6 | 112442147 | T | -5.173 | 1.15e-07 | 0.03824 | LAMA4 | protein_coding | C peptide [Mass/volume] in Serum or Plasma | endocrine | No | 6 | 112442147 | C | T | 0.1653 | 0.9585 | 476 | ADD | -0.3257 | 0.07949 |
| CPepti | rs9374305 | 6 | 112369643 | T | -5.127 | 1.47e-07 | 0.04875 | WISP3 | protein_coding | C peptide [Mass/volume] in Serum or Plasma | endocrine | No | 6 | 112369643 | C | T | 0.1472 | 0.9391 | 476 | ADD | -0.3761 | 0.0844 |
| UAlb | rs72882011 | 11 | 5647491 | C | -5.133 | 1.43e-07 | 0.0466 | HBG2 | protein_coding | Albumin [Presence] in Urine | urinary | No | 11 | 5647491 | T | C | 0.0731 | 1.067 | 602 | ADD | -0.3381 | 0.09518 |
| BBTAb | rs11794097 | 9 | 924014 | C | 5.453 | 2.47e-08 | 0.008224 | DMRT1 | protein_coding | Borrelia burgdorferi Ab [Units/volume] in Serum by Immunoassay | toxicology/pharmacology | Yes | 9 | 924014 | A | C | 0.0835 | 1.058 | 407 | ADD | 0.4831 | 0.1083 |
| BBTAb | rs73380437 | 9 | 927815 | A | 5.391 | 3.51e-08 | 0.01168 | DMRT1 | protein_coding | Borrelia burgdorferi Ab [Units/volume] in Serum by Immunoassay | toxicology/pharmacology | Yes | 9 | 927815 | C | A | 0.07964 | 1.075 | 407 | ADD | 0.4889 | 0.1099 |
| BBTAb | rs11789691 | 9 | 915218 | A | 5.362 | 4.12e-08 | 0.01372 | DMRT1 | protein_coding | Borrelia burgdorferi Ab [Units/volume] in Serum by Immunoassay | toxicology/pharmacology | Yes | 9 | 915218 | G | A | 0.08787 | 1.073 | 407 | ADD | 0.4735 | 0.105 |
| BBTAb | rs59989930 | 9 | 924396 | C | 5.321 | 5.16e-08 | 0.01718 | DMRT1 | protein_coding | Borrelia burgdorferi Ab [Units/volume] in Serum by Immunoassay | toxicology/pharmacology | No | 9 | 924396 | T | C | 0.08189 | 1.059 | 407 | ADD | 0.4764 | 0.1094 |
| BBTAb | 9:921385 | 9 | 921385 | A | 5.299 | 5.81e-08 | 0.01934 | DMRT1 | protein_coding | Borrelia burgdorferi Ab [Units/volume] in Serum by Immunoassay | toxicology/pharmacology | No | 9 | 921385 | G | A | 0.09281 | 1.064 | 407 | ADD | 0.4728 | 0.103 |
| BBTAb | rs58710178 | 9 | 921431 | A | 5.299 | 5.81e-08 | 0.01934 | DMRT1 | protein_coding | Borrelia burgdorferi Ab [Units/volume] in Serum by Immunoassay | toxicology/pharmacology | No | 9 | 921431 | G | A | 0.09281 | 1.064 | 407 | ADD | 0.4728 | 0.103 |
| BBTAb | rs11792914 | 9 | 923938 | T | 5.279 | 6.48e-08 | 0.02158 | DMRT1 | protein_coding | Borrelia burgdorferi Ab [Units/volume] in Serum by Immunoassay | toxicology/pharmacology | No | 9 | 923938 | C | T | 0.08428 | 1.059 | 407 | ADD | 0.479 | 0.1079 |
| BBTAb | rs73380407 | 9 | 922216 | G | 5.279 | 6.49e-08 | 0.02158 | DMRT1 | protein_coding | Borrelia burgdorferi Ab [Units/volume] in Serum by Immunoassay | toxicology/pharmacology | No | 9 | 922216 | A | G | 0.08428 | 1.059 | 407 | ADD | 0.479 | 0.1079 |
| BBTAb | rs73380405 | 9 | 922091 | T | 5.264 | 7.06e-08 | 0.02348 | DMRT1 | protein_coding | Borrelia burgdorferi Ab [Units/volume] in Serum by Immunoassay | toxicology/pharmacology | No | 9 | 922091 | G | T | 0.08421 | 1.058 | 407 | ADD | 0.4793 | 0.108 |

| lab | SNP | CHR | BP | A1 | tstat | P | adjP | Gene_Symbol | biotype | full_name | group | genome_wide | CHROM | GENPOS | ALLELE0 | ALLELE1 | male_A1FREQ | male_INFO | male_N | male_TEST | male_BETA | male_SE |
|---|---|---|---|---|---|---|---|---|---|---|---|---|---|---|---|---|---|---|---|---|---|---|
| BBTAb | rs10977534 | 9 | 924525 | T | 5.227 | 8.64e-08 | 0.02875 | DMRT1 | protein_coding | Borrelia burgdorferi Ab [Units/volume] in Serum by Immunoassay | toxicology/pharmacology | No | 9 | 924525 | G | T | 0.08967 | 1.082 | 407 | ADD | 0.4491 | 0.1037 |
| BBTAb | rs57482601 | 9 | 920519 | A | 5.221 | 8.89e-08 | 0.0296 | DMRT1 | protein_coding | Borrelia burgdorferi Ab [Units/volume] in Serum by Immunoassay | toxicology/pharmacology | No | 9 | 920519 | G | A | 0.09405 | 1.061 | 407 | ADD | 0.4622 | 0.1026 |
| BBTAb | rs10977499 | 9 | 919784 | A | 5.185 | 1.08e-07 | 0.03597 | DMRT1 | protein_coding | Borrelia burgdorferi Ab [Units/volume] in Serum by Immunoassay | toxicology/pharmacology | No | 9 | 919784 | G | A | 0.08569 | 1.058 | 407 | ADD | 0.4649 | 0.1071 |
| HSCRP | rs2113260 | 16 | 11455993 | T | -5.317 | 5.28e-08 | 0.01726 | CTD-3088G3.8 | protein_coding | C reactive protein (high sensitivity) | immune/inflammatory | No | 16 | 11455993 | C | T | 0.3389 | 1.079 | 585 | ADD | -0.2009 | 0.05497 |
| HSCRP | rs413024 | 16 | 11354091 | G | -5.233 | 8.32e-08 | 0.02721 | RMI2 | protein_coding | C reactive protein (high sensitivity) | immune/inflammatory | No | 16 | 11354091 | A | G | 0.2966 | 1.074 | 585 | ADD | -0.201 | 0.05707 |

| male_CHISQ | male_LOG10P | male_EXTRA | female_A1FREQ | female_INFO | female_N | female_TEST | female_BETA | female_SE | female_CHISQ | female_LOG10P | female_EXTRA | locus | nominal_replication |
|---|---|---|---|---|---|---|---|---|---|---|---|---|---|
| 15.95 | 4.188 | | 0.3496 | 0.9973 | 34341 | ADD | -0.02474 | 0.00701 | 12.46 | 3.381 | | loci_11 | Not replicated |
| 15.57 | 4.1 | | 0.3449 | 0.9764 | 34341 | ADD | -0.02541 | 0.007107 | 12.79 | 3.457 | | loci_11 | Not replicated |
| 16.1 | 4.22 | | 0.3446 | 0.9736 | 34341 | ADD | -0.0246 | 0.007118 | 11.94 | 3.261 | | loci_11 | Not replicated |
| 15.26 | 4.027 | | 0.3478 | 0.9729 | 34341 | ADD | -0.02391 | 0.007105 | 11.32 | 3.116 | | loci_11 | Not replicated |
| 15.26 | 4.027 | | 0.345 | 0.9741 | 34341 | ADD | -0.02393 | 0.007115 | 11.31 | 3.113 | | loci_11 | Not replicated |
| 17.24 | 4.483 | | 0.3397 | 0.9729 | 34341 | ADD | -0.02162 | 0.007145 | 9.156 | 2.606 | | loci_11 | Not replicated |
| 16.68 | 4.355 | | 0.3394 | 0.9741 | 34341 | ADD | -0.02178 | 0.007142 | 9.298 | 2.639 | | loci_11 | Not replicated |
| 17.13 | 4.456 | | 0.341 | 0.9753 | 34341 | ADD | -0.02111 | 0.007129 | 8.764 | 2.512 | | loci_11 | Not replicated |
| 16.38 | 4.285 | | 0.3396 | 0.9731 | 34341 | ADD | -0.02171 | 0.007145 | 9.232 | 2.624 | | loci_11 | Not replicated |
| 16.86 | 4.394 | | 0.341 | 0.9755 | 34341 | ADD | -0.02116 | 0.007129 | 8.806 | 2.522 | | loci_11 | Not replicated |
| 15.13 | 3.999 | | 0.1197 | 0.9831 | 34341 | ADD | 0.03205 | 0.01039 | 9.515 | 2.691 | | loci_41 | Not replicated |
| 16.45 | 4.302 | | 0.3416 | 0.9751 | 34341 | ADD | -0.02067 | 0.007127 | 8.41 | 2.428 | | loci_11 | Not replicated |
| 14.13 | 3.768 | | 0.1196 | 0.991 | 34341 | ADD | 0.03218 | 0.01035 | 9.665 | 2.726 | | loci_41 | Not replicated |
| 14.13 | 3.768 | | 0.1196 | 0.991 | 34341 | ADD | 0.03218 | 0.01035 | 9.665 | 2.726 | | loci_41 | Not replicated |
| 14.02 | 3.743 | | 0.1197 | 0.9907 | 34341 | ADD | 0.03217 | 0.01035 | 9.658 | 2.725 | | loci_41 | Not replicated |
| 13.99 | 3.736 | | 0.1197 | 0.991 | 34341 | ADD | 0.03213 | 0.01035 | 9.639 | 2.72 | | loci_41 | Not replicated |
| 13.99 | 3.736 | | 0.1196 | 0.991 | 34341 | ADD | 0.03211 | 0.01035 | 9.625 | 2.717 | | loci_41 | Not replicated |
| 13.99 | 3.736 | | 0.1196 | 0.991 | 34341 | ADD | 0.03211 | 0.01035 | 9.625 | 2.717 | | loci_41 | Not replicated |
| 14.03 | 3.745 | | 0.1195 | 0.9907 | 34341 | ADD | 0.03202 | 0.01035 | 9.563 | 2.702 | | loci_41 | Not replicated |
| 5.628 | 1.753 | | 0.2867 | 0.9711 | 34342 | ADD | -0.04243 | 0.007857 | 29.16 | 7.176 | | loci_singleton_20 | No replication available |
| 16.29 | 4.265 | | 0.09884 | 0.9928 | 34787 | ADD | 0.04386 | 0.01301 | 11.36 | 3.125 | | loci_singleton_21 | No replication available |
| 14.67 | 3.892 | | 0.09883 | 0.9928 | 34789 | ADD | -0.04133 | 0.01235 | 11.2 | 3.088 | | loci_singleton_22 | No replication available |
| 15.87 | 4.168 | | 0.2203 | 0.9943 | 34448 | ADD | 0.02808 | 0.008366 | 11.26 | 3.102 | | loci_87 | Not replicated |
| 15.81 | 4.155 | | 0.2214 | 0.9999 | 34448 | ADD | 0.02781 | 0.008328 | 11.15 | 3.075 | | loci_87 | Not replicated |
| 15.76 | 4.143 | | 0.2203 | 0.9942 | 34448 | ADD | 0.02796 | 0.008366 | 11.17 | 3.08 | | loci_87 | Not replicated |
| 15.48 | 4.08 | | 0.2198 | 0.9931 | 34448 | ADD | 0.02824 | 0.00838 | 11.36 | 3.124 | | loci_87 | Not replicated |
| 20.32 | 5.185 | | 0.08557 | 0.9616 | 34448 | ADD | -0.0327 | 0.01261 | 6.725 | 2.022 | | loci_singleton_27 | Not replicated |
| 14.43 | 3.838 | | 0.2177 | 0.9934 | 34448 | ADD | 0.02799 | 0.008407 | 11.09 | 3.061 | | loci_87 | Not replicated |
| 18.83 | 4.845 | | 0.4806 | 1.006 | 34577 | ADD | 0.01362 | 0.006913 | 3.882 | 1.312 | | loci_singleton_29 | Nominal replication |
| 11.53 | 3.164 | | 0.2699 | 0.9829 | 34577 | ADD | 0.02422 | 0.007879 | 9.447 | 2.675 | | loci_45 | Not replicated |
| 11.45 | 3.147 | | 0.27 | 0.9827 | 34577 | ADD | 0.02403 | 0.00788 | 9.297 | 2.639 | | loci_45 | Not replicated |
| 11.89 | 3.249 | | 0.4973 | 0.9884 | 34577 | ADD | -0.02037 | 0.006989 | 8.496 | 2.449 | | loci_32 | Not replicated |
| 3.668 | 1.256 | | 0.1004 | 0.9748 | 34577 | ADD | 0.04996 | 0.01169 | 18.27 | 4.718 | | loci_92 | Nominal replication |
| 4.622 | 1.501 | | 0.0878 | 0.996 | 34577 | ADD | 0.05 | 0.01228 | 16.57 | 4.328 | | loci_92 | Nominal replication |
| 11.82 | 3.232 | | 0.4979 | 0.9901 | 34577 | ADD | -0.02007 | 0.006982 | 8.263 | 2.393 | | loci_32 | Not replicated |
| 4.51 | 1.472 | | 0.08819 | 0.9912 | 34577 | ADD | 0.04997 | 0.01229 | 16.53 | 4.32 | | loci_92 | Nominal replication |
| 10.85 | 3.006 | | 0.495 | 0.9889 | 34577 | ADD | -0.02122 | 0.006987 | 9.22 | 2.621 | | loci_32 | Not replicated |
| 6.048 | 1.856 | | 0.4164 | 0.9859 | 34577 | ADD | 0.02843 | 0.007116 | 15.96 | 4.188 | | loci_53 | Not replicated |
| 4.207 | 1.395 | | 0.08817 | 0.992 | 34577 | ADD | 0.05038 | 0.01229 | 16.82 | 4.385 | | loci_92 | Nominal replication |
| 10.73 | 2.977 | | 0.3296 | 1.01 | 34577 | ADD | -0.02236 | 0.00737 | 9.204 | 2.617 | | loci_singleton_40 | Significant replication |
| 8.349 | 2.414 | | 0.4149 | 0.9911 | 34577 | ADD | 0.02471 | 0.0071 | 12.11 | 3.301 | | loci_1 | Not replicated |
| 4.375 | 1.438 | | 0.08838 | 0.9914 | 34577 | ADD | 0.0498 | 0.01228 | 16.45 | 4.302 | | loci_92 | Nominal replication |
| 8.335 | 2.41 | | 0.415 | 0.9912 | 34577 | ADD | 0.02453 | 0.0071 | 11.94 | 3.26 | | loci_1 | Not replicated |
| 4.822 | 1.551 | | 0.4193 | 0.9871 | 34577 | ADD | 0.03016 | 0.007109 | 18 | 4.655 | | loci_53 | Not replicated |
| 5.515 | 1.725 | | 0.4138 | 0.9844 | 34577 | ADD | 0.02893 | 0.007129 | 16.47 | 4.306 | | loci_53 | Not replicated |
| 5.059 | 1.611 | | 0.4185 | 0.9866 | 34577 | ADD | 0.0296 | 0.007112 | 17.32 | 4.5 | | loci_53 | Not replicated |

| male_CHISQ | male_LOG10P | male_EXTRA | female_A1FREQ | female_INFO | female_N | female_TEST | female_BETA | female_SE | female_CHISQ | female_LOG10P | female_EXTRA | locus | nominal_replication |
|---|---|---|---|---|---|---|---|---|---|---|---|---|---|
| 9.736 | 2.743 | | 0.1323 | 0.993 | 34577 | ADD | 0.03231 | 0.01029 | 9.862 | 2.773 | | loci_singleton_47 | Not replicated |
| 4.168 | 1.385 | | 0.08816 | 0.9914 | 34577 | ADD | 0.04971 | 0.01229 | 16.36 | 4.281 | | loci_92 | Nominal replication |
| 4.201 | 1.394 | | 0.08809 | 0.9915 | 34577 | ADD | 0.04956 | 0.01229 | 16.26 | 4.257 | | loci_92 | Nominal replication |
| 17.33 | 4.502 | | 0.1413 | 0.9356 | 34577 | ADD | 0.0197 | 0.01032 | 3.641 | 1.249 | | loci_24 | Not replicated |
| 6.088 | 1.866 | | 0.4225 | 0.9919 | 34577 | ADD | 0.02762 | 0.007103 | 15.13 | 3.998 | | loci_53 | Not replicated |
| 4.205 | 1.395 | | 0.0881 | 0.9916 | 34577 | ADD | 0.04942 | 0.01229 | 16.17 | 4.236 | | loci_92 | Nominal replication |
| 4.134 | 1.376 | | 0.08808 | 0.9913 | 34577 | ADD | 0.0496 | 0.0123 | 16.27 | 4.26 | | loci_92 | Nominal replication |
| 4.205 | 1.395 | | 0.08808 | 0.9917 | 34577 | ADD | 0.0494 | 0.01229 | 16.15 | 4.233 | | loci_92 | Nominal replication |
| 6.658 | 2.006 | | 0.1752 | 0.9904 | 34577 | ADD | -0.03437 | 0.009175 | 14.03 | 3.746 | | loci_singleton_55 | Not replicated |
| 4.378 | 1.439 | | 0.08827 | 0.9917 | 34577 | ADD | 0.04862 | 0.01228 | 15.67 | 4.123 | | loci_92 | Nominal replication |
| 4.758 | 1.535 | | 0.4225 | 0.9881 | 34577 | ADD | 0.02966 | 0.007097 | 17.46 | 4.533 | | loci_53 | Not replicated |
| 4.042 | 1.353 | | 0.08638 | 0.9966 | 34577 | ADD | 0.04976 | 0.01237 | 16.18 | 4.239 | | loci_92 | Nominal replication |
| 4.406 | 1.446 | | 0.08684 | 0.9929 | 34577 | ADD | 0.04881 | 0.01236 | 15.59 | 4.104 | | loci_92 | Nominal replication |
| 3.7 | 1.264 | | 0.08656 | 1.004 | 34577 | ADD | 0.0504 | 0.01231 | 16.75 | 4.371 | | loci_92 | Nominal replication |
| 3.999 | 1.342 | | 0.1004 | 0.974 | 34577 | ADD | 0.0471 | 0.0117 | 16.21 | 4.247 | | loci_92 | Nominal replication |
| 6.421 | 1.948 | | 0.1553 | 0.9879 | 34577 | ADD | 0.0345 | 0.009622 | 12.85 | 3.473 | | loci_singleton_62 | Not replicated |
| 7.875 | 2.3 | | 0.2183 | 0.9926 | 34577 | ADD | 0.02898 | 0.008423 | 11.84 | 3.236 | | loci_45 | Not replicated |
| 4.098 | 1.367 | | 0.08786 | 0.9918 | 34577 | ADD | 0.04922 | 0.01231 | 16 | 4.198 | | loci_92 | Nominal replication |
| 4.876 | 1.565 | | 0.423 | 0.9871 | 34577 | ADD | 0.02925 | 0.007101 | 16.97 | 4.42 | | loci_53 | Not replicated |
| 9.862 | 2.773 | | 0.2735 | 0.9869 | 34577 | ADD | 0.02368 | 0.007834 | 9.14 | 2.602 | | loci_45 | Not replicated |
| 4.363 | 1.435 | | 0.4265 | 0.986 | 34577 | ADD | 0.03013 | 0.007095 | 18.04 | 4.665 | | loci_53 | Not replicated |
| 4.765 | 1.537 | | 0.42 | 0.987 | 34577 | ADD | 0.02942 | 0.007107 | 17.14 | 4.458 | | loci_53 | Not replicated |
| 16.24 | 4.253 | | 0.1343 | 0.9917 | 34577 | ADD | 0.02027 | 0.01024 | 3.914 | 1.32 | | loci_24 | Not replicated |
| 4.727 | 1.527 | | 0.4233 | 0.9895 | 34577 | ADD | 0.02938 | 0.00709 | 17.17 | 4.466 | | loci_53 | Not replicated |
| 12.94 | 3.494 | | 0.275 | 0.9652 | 35678 | ADD | -0.02762 | 0.008019 | 11.87 | 3.243 | | loci_singleton_71 | Not replicated |
| 13.35 | 3.587 | | 0.4301 | 1.008 | 34362 | ADD | 0.02723 | 0.006645 | 16.8 | 4.381 | | loci_51 | Nominal replication |
| 12.94 | 3.492 | | 0.4315 | 1.012 | 34362 | ADD | 0.02726 | 0.006631 | 16.91 | 4.406 | | loci_51 | Nominal replication |
| 12.85 | 3.472 | | 0.4306 | 1.008 | 34362 | ADD | 0.02725 | 0.006644 | 16.82 | 4.387 | | loci_51 | Nominal replication |
| 12.89 | 3.481 | | 0.4305 | 1.007 | 34362 | ADD | 0.0272 | 0.006647 | 16.75 | 4.37 | | loci_51 | Nominal replication |
| 12.89 | 3.481 | | 0.4306 | 1.008 | 34362 | ADD | 0.02709 | 0.006644 | 16.63 | 4.342 | | loci_51 | Nominal replication |
| 12.81 | 3.463 | | 0.4306 | 1.008 | 34362 | ADD | 0.02709 | 0.006644 | 16.62 | 4.341 | | loci_51 | Nominal replication |
| 11.22 | 3.091 | | 0.434 | 1.006 | 34362 | ADD | 0.02835 | 0.006643 | 18.21 | 4.704 | | loci_51 | Nominal replication |
| 18.5 | 4.771 | | 0.3592 | 1.001 | 35668 | ADD | -0.01562 | 0.006983 | 5.002 | 1.597 | | loci_singleton_79 | Not replicated |
| 19.54 | 5.006 | | 0.09211 | 1.002 | 35644 | ADD | -0.02486 | 0.0109 | 5.2 | 1.646 | | loci_80 | Not replicated |
| 19.43 | 4.982 | | 0.09207 | 1.001 | 35644 | ADD | -0.0248 | 0.0109 | 5.174 | 1.64 | | loci_80 | Not replicated |
| 18.09 | 4.676 | | 0.09276 | 1.012 | 35644 | ADD | -0.02628 | 0.01081 | 5.906 | 1.821 | | loci_80 | Not replicated |
| 12.84 | 3.47 | | 0.08194 | 0.9984 | 35644 | ADD | -0.03733 | 0.01151 | 10.51 | 2.927 | | loci_80 | Not replicated |
| 15.98 | 4.193 | | 0.08819 | 1.001 | 35644 | ADD | -0.02968 | 0.01113 | 7.115 | 2.117 | | loci_80 | Not replicated |
| 18.91 | 4.862 | | 0.4982 | 1.001 | 31113 | ADD | 0.02465 | 0.007485 | 10.85 | 3.005 | | loci_23 | Not replicated |
| 18.72 | 4.82 | | 0.4961 | 0.9988 | 31113 | ADD | 0.02451 | 0.007495 | 10.7 | 2.97 | | loci_23 | Not replicated |
| 17.88 | 4.63 | | 0.4727 | 1.004 | 31113 | ADD | -0.02358 | 0.007487 | 9.916 | 2.786 | | loci_23 | Not replicated |
| 17.41 | 4.52 | | 0.4932 | 1.002 | 31113 | ADD | 0.02359 | 0.007486 | 9.926 | 2.788 | | loci_23 | Not replicated |
| 17.69 | 4.585 | | 0.4745 | 1.002 | 31113 | ADD | -0.02323 | 0.007494 | 9.608 | 2.713 | | loci_23 | Not replicated |
| 17.48 | 4.536 | | 0.4753 | 1.001 | 31113 | ADD | -0.02345 | 0.007495 | 9.79 | 2.756 | | loci_23 | Not replicated |
| 17.05 | 4.439 | | 0.4917 | 0.9999 | 31113 | ADD | 0.02379 | 0.007494 | 10.08 | 2.824 | | loci_23 | Not replicated |

| male_CHISQ | male_LOG10P | male_EXTRA | female_A1FREQ | female_INFO | female_N | female_TEST | female_BETA | female_SE | female_CHISQ | female_LOG10P | female_EXTRA | locus | nominal_replication |
|---|---|---|---|---|---|---|---|---|---|---|---|---|---|
| 16.94 | 4.414 | | 0.4917 | 1 | 31113 | ADD | 0.0238 | 0.007494 | 10.08 | 2.825 | | loci_23 | Not replicated |
| 17.11 | 4.451 | | 0.4944 | 1.003 | 31113 | ADD | 0.02246 | 0.007481 | 9.013 | 2.572 | | loci_23 | Not replicated |
| 16.72 | 4.363 | | 0.4949 | 1.007 | 31113 | ADD | 0.02257 | 0.007466 | 9.139 | 2.602 | | loci_23 | Not replicated |
| 17.05 | 4.438 | | 0.4943 | 1.003 | 31113 | ADD | 0.02219 | 0.007481 | 8.8 | 2.521 | | loci_23 | Not replicated |
| 16.35 | 4.277 | | 0.4784 | 1.005 | 31113 | ADD | -0.02293 | 0.007482 | 9.392 | 2.662 | | loci_23 | Not replicated |
| 7.772 | 2.275 | | 0.4511 | 0.9974 | 30062 | ADD | 0.03833 | 0.008127 | 22.24 | 5.619 | | loci_54 | Not replicated |
| 7.889 | 2.303 | | 0.4528 | 0.9978 | 30062 | ADD | 0.03805 | 0.008124 | 21.94 | 5.552 | | loci_54 | Not replicated |
| 7.428 | 2.192 | | 0.4586 | 0.9975 | 30062 | ADD | 0.03765 | 0.008121 | 21.5 | 5.45 | | loci_54 | Not replicated |
| 7.427 | 2.192 | | 0.4586 | 0.9974 | 30062 | ADD | 0.03765 | 0.008121 | 21.5 | 5.45 | | loci_54 | Not replicated |
| 6.436 | 1.952 | | 0.4583 | 0.9941 | 30062 | ADD | 0.03765 | 0.008137 | 21.4 | 5.429 | | loci_54 | Not replicated |
| 12.9 | 3.482 | | 0.1832 | 1.001 | 36195 | ADD | -0.03071 | 0.008944 | 11.79 | 3.225 | | loci_84 | Not replicated |
| 13.73 | 3.676 | | 0.1841 | 0.998 | 36195 | ADD | -0.02906 | 0.008942 | 10.56 | 2.937 | | loci_84 | Not replicated |
| 10.45 | 2.911 | | 0.08339 | 0.9925 | 35703 | ADD | -0.05382 | 0.01323 | 16.56 | 4.326 | | loci_15 | Not replicated |
| 10.15 | 2.84 | | 0.08267 | 0.9932 | 35703 | ADD | -0.05396 | 0.01327 | 16.54 | 4.322 | | loci_15 | Not replicated |
| 10.04 | 2.814 | | 0.08266 | 0.9929 | 35703 | ADD | -0.05381 | 0.01327 | 16.44 | 4.299 | | loci_15 | Not replicated |
| 9.896 | 2.781 | | 0.08287 | 0.993 | 35703 | ADD | -0.05385 | 0.01326 | 16.49 | 4.31 | | loci_15 | Not replicated |
| 9.499 | 2.687 | | 0.08212 | 0.9933 | 35703 | ADD | -0.05387 | 0.01331 | 16.38 | 4.285 | | loci_15 | Not replicated |
| 16.76 | 4.372 | | 0.05871 | 0.9845 | 35703 | ADD | 0.04371 | 0.01559 | 7.859 | 2.296 | | loci_13 | Not replicated |
| 16.76 | 4.373 | | 0.05877 | 0.9871 | 35703 | ADD | 0.04333 | 0.01556 | 7.75 | 2.27 | | loci_13 | Not replicated |
| 16.76 | 4.373 | | 0.05877 | 0.987 | 35703 | ADD | 0.04331 | 0.01556 | 7.743 | 2.268 | | loci_13 | Not replicated |
| 9.345 | 2.651 | | 0.08257 | 0.993 | 35703 | ADD | -0.05277 | 0.01328 | 15.79 | 4.151 | | loci_15 | Not replicated |
| 16.39 | 4.287 | | 0.05829 | 0.9832 | 35703 | ADD | 0.04321 | 0.01564 | 7.629 | 2.241 | | loci_13 | Not replicated |
| 16.29 | 4.264 | | 0.05838 | 0.9857 | 35703 | ADD | 0.04272 | 0.01561 | 7.486 | 2.206 | | loci_13 | Not replicated |
| 16.27 | 4.259 | | 0.05836 | 0.9855 | 35703 | ADD | 0.04266 | 0.01562 | 7.465 | 2.201 | | loci_13 | Not replicated |
| 16.13 | 4.229 | | 0.05839 | 0.9857 | 35703 | ADD | 0.04275 | 0.01561 | 7.5 | 2.21 | | loci_13 | Not replicated |
| 8.902 | 2.545 | | 0.05313 | 1.001 | 35661 | ADD | -0.05865 | 0.01523 | 14.84 | 3.932 | | loci_singleton_117 | Nominal replication |
| 15.28 | 4.034 | | 0.08156 | 0.8941 | 30952 | ADD | 0.05802 | 0.01408 | 16.98 | 4.423 | | loci_singleton_118 | Not replicated |
| 14.65 | 3.888 | | 0.426 | 0.9733 | 35673 | ADD | -0.02314 | 0.006961 | 11.05 | 3.052 | | loci_38 | Not replicated |
| 14.59 | 3.874 | | 0.4162 | 1.004 | 35673 | ADD | -0.02248 | 0.006877 | 10.69 | 2.968 | | loci_38 | Not replicated |
| male_CHISQ | male_LOG10P | male_EXTRA | female_A1FREQ | female_INFO | female_N | female_TEST | female_BETA | female_SE | female_CHISQ | female_LOG10P | female_EXTRA | locus | nominal_replication |
| 10.52 | 2.927 | | 0.05069 | 0.9968 | 35804 | ADD | -0.06369 | 0.01641 | 15.06 | 3.983 | | loci_14 | Not replicated |
| 10.37 | 2.892 | | 0.05073 | 0.9994 | 35804 | ADD | -0.06382 | 0.01638 | 15.17 | 4.009 | | loci_14 | Not replicated |
| 10.16 | 2.842 | | 0.05068 | 0.9997 | 35804 | ADD | -0.0641 | 0.01639 | 15.3 | 4.037 | | loci_14 | Not replicated |
| 9.815 | 2.762 | | 0.05031 | 0.9958 | 35804 | ADD | -0.06441 | 0.01648 | 15.28 | 4.033 | | loci_14 | Not replicated |
| 17.98 | 4.651 | | 0.445 | 0.998 | 35804 | ADD | -0.01882 | 0.007247 | 6.743 | 2.026 | | loci_10 | Not replicated |
| 17.76 | 4.601 | | 0.4459 | 0.9939 | 35804 | ADD | -0.01888 | 0.007261 | 6.759 | 2.03 | | loci_10 | Not replicated |
| 17.53 | 4.548 | | 0.4463 | 0.994 | 35804 | ADD | -0.01894 | 0.00726 | 6.806 | 2.042 | | loci_10 | Not replicated |
| 17.71 | 4.59 | | 0.4435 | 0.9925 | 35804 | ADD | -0.01874 | 0.00727 | 6.648 | 2.003 | | loci_10 | Not replicated |
| 15.03 | 3.976 | | 0.08453 | 1.002 | 35804 | ADD | -0.03723 | 0.0129 | 8.332 | 2.41 | | loci_21 | Not replicated |
| 10.51 | 2.926 | | 0.4964 | 1 | 35804 | ADD | -0.02591 | 0.007186 | 13.01 | 3.508 | | loci_36 | Significant replication |
| 15.14 | 4.002 | | 0.08407 | 0.9986 | 35804 | ADD | -0.03653 | 0.01296 | 7.952 | 2.318 | | loci_21 | Not replicated |
| 10.32 | 2.882 | | 0.4962 | 1.001 | 35804 | ADD | -0.02588 | 0.007184 | 12.98 | 3.502 | | loci_36 | Significant replication |
| 14.85 | 3.933 | | 0.0842 | 0.9988 | 35804 | ADD | -0.0363 | 0.01295 | 7.862 | 2.297 | | loci_21 | Not replicated |
| 14.92 | 3.951 | | 0.08436 | 0.9992 | 35804 | ADD | -0.03601 | 0.01293 | 7.753 | 2.271 | | loci_21 | Not replicated |
| 14.51 | 3.855 | | 0.08514 | 1.004 | 35804 | ADD | -0.03623 | 0.01285 | 7.951 | 2.318 | | loci_21 | Not replicated |
| 12.75 | 3.449 | | 0.1258 | 0.9752 | 21462 | ADD | 0.05543 | 0.01258 | 19.41 | 4.977 | | loci_16 | Not replicated |

| male_CHISQ | male_LOG10P | male_EXTRA | female_A1FREQ | female_INFO | female_N | female_TEST | female_BETA | female_SE | female_CHISQ | female_LOG10P | female_EXTRA | locus | nominal_replication |
|---|---|---|---|---|---|---|---|---|---|---|---|---|---|
| 12.17 | 3.314 | | 0.15 | 0.988 | 21462 | ADD | 0.05025 | 0.01161 | 18.74 | 4.825 | | loci_33 | Not replicated |
| 18.06 | 4.67 | | 0.05739 | 1.004 | 21462 | ADD | -0.05637 | 0.01769 | 10.15 | 2.841 | | loci_22 | Not replicated |
| 15.74 | 4.138 | | 0.06267 | 0.9109 | 21462 | ADD | -0.0607 | 0.01783 | 11.6 | 3.18 | | loci_22 | Not replicated |
| 15.91 | 4.177 | | 0.1099 | 0.9912 | 21462 | ADD | 0.04475 | 0.01324 | 11.42 | 3.139 | | loci_16 | Not replicated |
| 16.18 | 4.24 | | 0.1137 | 0.9874 | 21462 | ADD | 0.04313 | 0.01306 | 10.9 | 3.017 | | loci_16 | Not replicated |
| 10.35 | 2.888 | | 0.1455 | 0.9897 | 21462 | ADD | 0.04806 | 0.01174 | 16.75 | 4.37 | | loci_33 | Not replicated |
| 20.26 | 5.171 | | 0.05618 | 0.9412 | 21462 | ADD | -0.05064 | 0.01845 | 7.532 | 2.217 | | loci_22 | Not replicated |
| 20.06 | 5.126 | | 0.05614 | 0.9404 | 21462 | ADD | -0.05101 | 0.01847 | 7.63 | 2.241 | | loci_22 | Not replicated |
| 11.11 | 3.066 | | 0.1499 | 0.9896 | 21462 | ADD | 0.04571 | 0.0116 | 15.52 | 4.087 | | loci_33 | Not replicated |
| 19.75 | 5.054 | | 0.0528 | 1.005 | 21462 | ADD | -0.05019 | 0.01839 | 7.448 | 2.197 | | loci_22 | Not replicated |
| 32.28 | 7.874 | | 0.1583 | 0.9978 | 21442 | ADD | 0.01902 | 0.01364 | 1.946 | 0.7877 | | loci_6 | No replication available |
| 33.29 | 8.101 | | 0.164 | 1.005 | 21442 | ADD | 0.01685 | 0.01339 | 1.584 | 0.6815 | | loci_6 | No replication available |
| 33.52 | 8.151 | | 0.163 | 0.9976 | 21442 | ADD | 0.01661 | 0.01348 | 1.518 | 0.6616 | | loci_6 | No replication available |
| 31.13 | 7.617 | | 0.1583 | 0.9942 | 21442 | ADD | 0.01939 | 0.01366 | 2.014 | 0.8073 | | loci_6 | No replication available |
| 9.311 | 2.642 | | 0.4738 | 0.9841 | 21442 | ADD | 0.04005 | 0.01003 | 15.94 | 4.185 | | loci_singleton_151 | No replication available |
| 5.657 | 1.76 | | 0.06588 | 0.9796 | 21460 | ADD | 0.09823 | 0.01919 | 26.2 | 6.512 | | loci_12 | No replication available |
| 5.888 | 1.817 | | 0.07103 | 0.9803 | 21460 | ADD | 0.08788 | 0.01853 | 22.5 | 5.677 | | loci_12 | No replication available |
| 15.71 | 4.133 | | 0.08548 | 0.9998 | 21459 | ADD | -0.05283 | 0.01609 | 10.78 | 2.99 | | loci_57 | No replication available |
| 16.85 | 4.394 | | 0.08657 | 0.9993 | 21459 | ADD | -0.04976 | 0.01601 | 9.653 | 2.723 | | loci_57 | No replication available |
| 16.66 | 4.349 | | 0.08698 | 1.005 | 21459 | ADD | -0.04848 | 0.01594 | 9.253 | 2.629 | | loci_57 | No replication available |
| 15.48 | 4.079 | | 0.08528 | 0.9978 | 21459 | ADD | -0.05155 | 0.01613 | 10.22 | 2.857 | | loci_57 | No replication available |
| 15.26 | 4.029 | | 0.08626 | 1.001 | 21459 | ADD | -0.05094 | 0.01601 | 10.12 | 2.834 | | loci_57 | No replication available |
| 12.2 | 3.321 | | 0.1184 | 0.9841 | 21459 | ADD | -0.05013 | 0.01402 | 12.78 | 3.455 | | loci_68 | No replication available |
| 11.96 | 3.265 | | 0.1183 | 0.9843 | 21459 | ADD | -0.05048 | 0.01402 | 12.96 | 3.496 | | loci_68 | No replication available |
| 11.26 | 3.101 | | 0.119 | 0.9868 | 21459 | ADD | -0.05176 | 0.01397 | 13.72 | 3.674 | | loci_68 | No replication available |
| 11.29 | 3.108 | | 0.1186 | 0.9822 | 21459 | ADD | -0.05172 | 0.01403 | 13.59 | 3.644 | | loci_68 | No replication available |
| 11.14 | 3.073 | | 0.1189 | 0.9873 | 21459 | ADD | -0.05181 | 0.01397 | 13.75 | 3.68 | | loci_68 | No replication available |
| 11.14 | 3.073 | | 0.1189 | 0.9873 | 21459 | ADD | -0.05179 | 0.01397 | 13.74 | 3.678 | | loci_68 | No replication available |
| 11.91 | 3.254 | | 0.1194 | 0.9876 | 21459 | ADD | -0.04991 | 0.01394 | 12.82 | 3.464 | | loci_68 | No replication available |
| 11.92 | 3.255 | | 0.1194 | 0.9876 | 21459 | ADD | -0.04979 | 0.01394 | 12.75 | 3.449 | | loci_68 | No replication available |
| 11.91 | 3.253 | | 0.1194 | 0.9876 | 21459 | ADD | -0.0498 | 0.01394 | 12.76 | 3.45 | | loci_68 | No replication available |
| 11.3 | 3.11 | | 0.1189 | 0.9836 | 21459 | ADD | -0.05128 | 0.014 | 13.42 | 3.604 | | loci_68 | No replication available |
| 11.6 | 3.18 | | 0.1182 | 0.9876 | 21459 | ADD | -0.05056 | 0.01401 | 13.03 | 3.514 | | loci_68 | No replication available |
| 30.79 | 7.541 | | 0.1773 | 0.9984 | 21462 | ADD | 0.03487 | 0.01182 | 8.709 | 2.499 | | loci_16 | Not replicated |
| 24.43 | 6.112 | | 0.1524 | 0.9918 | 21462 | ADD | 0.04308 | 0.01259 | 11.7 | 3.205 | | loci_16 | Not replicated |
| 25.14 | 6.272 | | 0.1137 | 0.9874 | 21462 | ADD | 0.04718 | 0.01429 | 10.9 | 3.016 | | loci_16 | Not replicated |
| 26.39 | 6.553 | | 0.09984 | 1.001 | 21462 | ADD | 0.04769 | 0.01504 | 10.06 | 2.82 | | loci_16 | Not replicated |
| 25.71 | 6.401 | | 0.1031 | 1.004 | 21462 | ADD | 0.04789 | 0.0148 | 10.47 | 2.915 | | loci_16 | Not replicated |
| 25.12 | 6.269 | | 0.1057 | 0.9873 | 21462 | ADD | 0.04834 | 0.01476 | 10.72 | 2.974 | | loci_16 | Not replicated |
| 24.61 | 6.155 | | 0.1099 | 0.9912 | 21462 | ADD | 0.04717 | 0.01449 | 10.6 | 2.946 | | loci_16 | Not replicated |
| 25.08 | 6.26 | | 0.1038 | 1 | 21462 | ADD | 0.04696 | 0.01478 | 10.09 | 2.827 | | loci_16 | Not replicated |
| 25.37 | 6.324 | | 0.1038 | 1 | 21462 | ADD | 0.04636 | 0.01478 | 9.839 | 2.767 | | loci_16 | Not replicated |
| 24.62 | 6.157 | | 0.1036 | 1 | 21462 | ADD | 0.0475 | 0.01479 | 10.31 | 2.878 | | loci_16 | Not replicated |
| 25.32 | 6.314 | | 0.1039 | 0.9994 | 21462 | ADD | 0.04605 | 0.01478 | 9.709 | 2.737 | | loci_16 | Not replicated |
| male_CHISQ | male_LOG10P | male_EXTRA | female_A1FREQ | female_INFO | female_N | female_TEST | female_BETA | female_SE | female_CHISQ | female_LOG10P | female_EXTRA | locus | nominal_replication |
| 25.51 | 6.357 | | 0.1039 | 1 | 21462 | ADD | 0.04572 | 0.01477 | 9.576 | 2.705 | | loci_16 | Not replicated |

| male_CHISQ | male_LOG10P | male_EXTRA | female_A1FREQ | female_INFO | female_N | female_TEST | female_BETA | female_SE | female_CHISQ | female_LOG10P | female_EXTRA | locus | nominal_replication |
|---|---|---|---|---|---|---|---|---|---|---|---|---|---|
| 23.46 | 5.894 | | 0.1108 | 0.9943 | 21462 | ADD | 0.04761 | 0.01441 | 10.92 | 3.022 | | loci_16 | Not replicated |
| 24.82 | 6.202 | | 0.1039 | 0.9998 | 21462 | ADD | 0.04656 | 0.01478 | 9.923 | 2.787 | | loci_16 | Not replicated |
| 24.96 | 6.233 | | 0.1102 | 0.9968 | 21462 | ADD | 0.04518 | 0.01442 | 9.811 | 2.761 | | loci_16 | Not replicated |
| 25.49 | 6.351 | | 0.104 | 1 | 21462 | ADD | 0.04549 | 0.01477 | 9.482 | 2.683 | | loci_16 | Not replicated |
| 25.42 | 6.336 | | 0.104 | 1 | 21462 | ADD | 0.04537 | 0.01477 | 9.433 | 2.671 | | loci_16 | Not replicated |
| 22.99 | 5.788 | | 0.1669 | 0.9894 | 21462 | ADD | 0.04002 | 0.01215 | 10.84 | 3.003 | | loci_16 | Not replicated |
| 22.52 | 5.681 | | 0.1076 | 1.004 | 21462 | ADD | 0.04808 | 0.01452 | 10.96 | 3.03 | | loci_16 | Not replicated |
| 17.32 | 4.499 | | 0.1258 | 0.9752 | 21462 | ADD | 0.05393 | 0.01377 | 15.34 | 4.047 | | loci_16 | Not replicated |
| 20.37 | 5.195 | | 0.1043 | 1.002 | 21462 | ADD | 0.05144 | 0.01474 | 12.19 | 3.318 | | loci_16 | Not replicated |
| 22.04 | 5.573 | | 0.1135 | 0.9879 | 21462 | ADD | 0.04479 | 0.01431 | 9.801 | 2.758 | | loci_16 | Not replicated |
| 20.75 | 5.282 | | 0.103 | 1 | 21462 | ADD | 0.04822 | 0.01483 | 10.57 | 2.939 | | loci_16 | Not replicated |
| 19.5 | 4.997 | | 0.1026 | 0.9968 | 21462 | ADD | 0.05027 | 0.01488 | 11.41 | 3.135 | | loci_16 | Not replicated |
| 21.55 | 5.462 | | 0.1224 | 0.9936 | 21462 | ADD | 0.04311 | 0.0138 | 9.755 | 2.747 | | loci_16 | Not replicated |
| 21.76 | 5.511 | | 0.1078 | 0.9942 | 21462 | ADD | 0.045 | 0.01458 | 9.526 | 2.693 | | loci_16 | Not replicated |
| 17.82 | 4.615 | | 0.1157 | 1.001 | 21462 | ADD | 0.0496 | 0.0141 | 12.38 | 3.363 | | loci_16 | Not replicated |
| 20.46 | 5.215 | | 0.1037 | 0.9997 | 21462 | ADD | 0.04627 | 0.01479 | 9.787 | 2.755 | | loci_16 | Not replicated |
| 15.55 | 4.095 | | 0.09603 | 1.004 | 21462 | ADD | 0.05678 | 0.01528 | 13.8 | 3.692 | | loci_16 | Not replicated |
| 16.7 | 4.358 | | 0.1054 | 1.006 | 21462 | ADD | 0.05125 | 0.01465 | 12.23 | 3.329 | | loci_16 | Not replicated |
| 16.77 | 4.376 | | 0.1042 | 0.9952 | 21462 | ADD | 0.05135 | 0.0148 | 12.03 | 3.282 | | loci_16 | Not replicated |
| 17.32 | 4.5 | | 0.08528 | 0.9982 | 21462 | ADD | 0.0545 | 0.01617 | 11.36 | 3.125 | | loci_16 | Not replicated |
| 18.44 | 4.757 | | 0.1009 | 1.007 | 21462 | ADD | 0.04824 | 0.01493 | 10.44 | 2.91 | | loci_16 | Not replicated |
| 18.14 | 4.687 | | 0.1207 | 1.01 | 21462 | ADD | 0.04498 | 0.01378 | 10.66 | 2.961 | | loci_16 | Not replicated |
| 16.55 | 4.324 | | 0.09555 | 0.9878 | 21462 | ADD | 0.05309 | 0.01544 | 11.83 | 3.233 | | loci_16 | Not replicated |
| 18.78 | 4.834 | | 0.085 | 1.001 | 21462 | ADD | 0.05079 | 0.01617 | 9.864 | 2.773 | | loci_16 | Not replicated |
| 15.24 | 4.025 | | 0.0928 | 0.975 | 21462 | ADD | 0.05371 | 0.01574 | 11.64 | 3.189 | | loci_16 | Not replicated |
| 14.59 | 3.874 | | 0.09044 | 1.007 | 21462 | ADD | 0.05186 | 0.01568 | 10.94 | 3.026 | | loci_16 | Not replicated |
| 6.934 | 2.073 | | 0.05902 | 0.9991 | 20991 | ADD | -0.09177 | 0.01908 | 23.14 | 5.822 | | loci_28 | Not replicated |
| 7.929 | 2.313 | | 0.06166 | 0.9926 | 20991 | ADD | -0.08611 | 0.01875 | 21.09 | 5.358 | | loci_28 | Not replicated |
| 5.628 | 1.753 | | 0.05832 | 0.9857 | 20991 | ADD | -0.09341 | 0.01931 | 23.39 | 5.878 | | loci_28 | Not replicated |
| 6.575 | 1.985 | | 0.113 | 0.9794 | 19669 | ADD | 0.08187 | 0.0158 | 26.85 | 6.657 | | loci_69 | Not replicated |
| 6.459 | 1.957 | | 0.1191 | 0.9874 | 19669 | ADD | 0.07982 | 0.01538 | 26.93 | 6.675 | | loci_69 | Not replicated |
| 6.762 | 2.031 | | 0.1191 | 0.9853 | 19669 | ADD | 0.07794 | 0.0154 | 25.62 | 6.381 | | loci_69 | Not replicated |
| 20.34 | 5.188 | | 0.08305 | 0.9969 | 19280 | ADD | -0.04975 | 0.01726 | 8.306 | 2.403 | | loci_8 | Not replicated |
| 19.65 | 5.031 | | 0.08678 | 0.9824 | 19280 | ADD | -0.04662 | 0.01704 | 7.48 | 2.205 | | loci_8 | Not replicated |
| 14.23 | 3.79 | | 0.05835 | 1.004 | 19280 | ADD | -0.06865 | 0.02038 | 11.35 | 3.122 | | loci_50 | Not replicated |
| 20.09 | 5.13 | | 0.08701 | 0.988 | 19280 | ADD | -0.04359 | 0.01698 | 6.594 | 1.99 | | loci_8 | Not replicated |
| 13.99 | 3.737 | | 0.05797 | 1.002 | 19280 | ADD | -0.0686 | 0.02046 | 11.24 | 3.097 | | loci_50 | Not replicated |
| 68.74 | 15.95 | | 0.07899 | 1.015 | 19280 | ADD | -0.2925 | 0.01762 | 275.70 | 61.19 | | loci_46 | Significant replication |
| 19.01 | 4.887 | | 0.08626 | 0.9908 | 19280 | ADD | -0.04511 | 0.01702 | 7.028 | 2.095 | | loci_8 | Not replicated |
| 14.89 | 3.944 | | 0.2346 | 0.9906 | 19280 | ADD | -0.03531 | 0.01134 | 9.694 | 2.733 | | loci_52 | Not replicated |
| 18.04 | 4.665 | | 0.07368 | 0.9798 | 19280 | ADD | -0.04934 | 0.0184 | 7.193 | 2.136 | | loci_8 | Not replicated |
| 14.96 | 3.959 | | 0.2294 | 1.016 | 19280 | ADD | -0.03491 | 0.01128 | 9.575 | 2.705 | | loci_52 | Not replicated |
| 14.62 | 3.882 | | 0.2331 | 0.9887 | 19280 | ADD | -0.03517 | 0.01138 | 9.555 | 2.7 | | loci_52 | Not replicated |
| 14.04 | 3.747 | | 0.2285 | 1.011 | 19280 | ADD | -0.03598 | 0.01133 | 10.09 | 2.827 | | loci_52 | Not replicated |
| 6.606 | 1.993 | | 0.3663 | 1.004 | 19112 | ADD | 0.04471 | 0.01028 | 18.9 | 4.86 | | loci_91 | Not replicated |
| 6.452 | 1.955 | | 0.3643 | 0.9907 | 19112 | ADD | 0.04508 | 0.01037 | 18.91 | 4.863 | | loci_91 | Not replicated |
| 6.46 | 1.957 | | 0.3643 | 0.9908 | 19112 | ADD | 0.04507 | 0.01037 | 18.9 | 4.86 | | loci_91 | Not replicated |

| male_CHISQ | male_LOG10P | male_EXTRA | female_A1FREQ | female_INFO | female_N | female_TEST | female_BETA | female_SE | female_CHISQ | female_LOG10P | female_EXTRA | locus | nominal_replication |
|---|---|---|---|---|---|---|---|---|---|---|---|---|---|
| 6.449 | 1.955 | | 0.3643 | 0.9909 | 19112 | ADD | 0.04489 | 0.01037 | 18.75 | 4.827 | | loci_91 | Not replicated |
| 18.27 | 4.718 | | 0.08622 | 1.014 | 17241 | ADD | -0.0595 | 0.01778 | 11.2 | 3.088 | | loci_61 | Nominal replication |
| 18.27 | 4.718 | | 0.08622 | 1.014 | 17241 | ADD | -0.05948 | 0.01778 | 11.2 | 3.087 | | loci_61 | Nominal replication |
| 18.14 | 4.687 | | 0.08524 | 0.9967 | 17241 | ADD | -0.06003 | 0.01803 | 11.09 | 3.062 | | loci_61 | Nominal replication |
| 12.23 | 3.327 | | 0.07077 | 0.9991 | 17241 | ADD | -0.07694 | 0.0196 | 15.4 | 4.061 | | loci_61 | Nominal replication |
| 11.61 | 3.182 | | 0.07045 | 1.001 | 17241 | ADD | -0.07873 | 0.01963 | 16.09 | 4.219 | | loci_61 | Nominal replication |
| 12.16 | 3.311 | | 0.07077 | 0.9991 | 17241 | ADD | -0.07693 | 0.01961 | 15.4 | 4.06 | | loci_61 | Nominal replication |
| 12.16 | 3.311 | | 0.07077 | 0.9991 | 17241 | ADD | -0.07693 | 0.01961 | 15.4 | 4.06 | | loci_61 | Nominal replication |
| 11.61 | 3.184 | | 0.07052 | 1.001 | 17241 | ADD | -0.07816 | 0.01962 | 15.87 | 4.168 | | loci_61 | Nominal replication |
| 11.6 | 3.181 | | 0.07046 | 1.001 | 17241 | ADD | -0.0782 | 0.01963 | 15.88 | 4.171 | | loci_61 | Nominal replication |
| 11.92 | 3.255 | | 0.07078 | 0.9996 | 17241 | ADD | -0.07701 | 0.0196 | 15.44 | 4.069 | | loci_61 | Nominal replication |
| 12.14 | 3.308 | | 0.07099 | 1.001 | 17241 | ADD | -0.07605 | 0.01956 | 15.12 | 3.995 | | loci_61 | Nominal replication |
| male_CHISQ | male_LOG10P | male_EXTRA | female_A1FREQ | female_INFO | female_N | female_TEST | female_BETA | female_SE | female_CHISQ | female_LOG10P | female_EXTRA | locus | nominal_replication |
| 12.14 | 3.308 | | 0.07098 | 1.001 | 17241 | ADD | -0.07604 | 0.01956 | 15.11 | 3.994 | | loci_61 | Nominal replication |
| 11.61 | 3.184 | | 0.07052 | 1.001 | 17241 | ADD | -0.07778 | 0.01962 | 15.71 | 4.133 | | loci_61 | Nominal replication |
| 12.04 | 3.284 | | 0.07098 | 1.001 | 17241 | ADD | -0.07602 | 0.01956 | 15.11 | 3.994 | | loci_61 | Nominal replication |
| 11.99 | 3.272 | | 0.07098 | 1.002 | 17241 | ADD | -0.076 | 0.01955 | 15.11 | 3.994 | | loci_61 | Nominal replication |
| 13.22 | 3.557 | | 0.06188 | 0.9785 | 17241 | ADD | -0.07745 | 0.02108 | 13.49 | 3.621 | | loci_61 | Nominal replication |
| 12.56 | 3.405 | | 0.06919 | 0.9893 | 17241 | ADD | -0.07381 | 0.01991 | 13.75 | 3.679 | | loci_61 | Nominal replication |
| 11.68 | 3.2 | | 0.07036 | 0.9793 | 17241 | ADD | -0.07615 | 0.01986 | 14.71 | 3.901 | | loci_61 | Nominal replication |
| 12.76 | 3.451 | | 0.06965 | 0.9953 | 17241 | ADD | -0.07232 | 0.01979 | 13.36 | 3.59 | | loci_61 | Nominal replication |
| 11.56 | 3.171 | | 0.07152 | 1 | 17241 | ADD | -0.07482 | 0.0195 | 14.72 | 3.904 | | loci_61 | Nominal replication |
| 11.72 | 3.209 | | 0.07173 | 1.001 | 17241 | ADD | -0.07417 | 0.01946 | 14.52 | 3.858 | | loci_61 | Nominal replication |
| 11.72 | 3.209 | | 0.07173 | 1.001 | 17241 | ADD | -0.07417 | 0.01946 | 14.52 | 3.858 | | loci_61 | Nominal replication |
| 11.56 | 3.17 | | 0.07152 | 1 | 17241 | ADD | -0.07481 | 0.0195 | 14.71 | 3.903 | | loci_61 | Nominal replication |
| 11.56 | 3.17 | | 0.07152 | 1 | 17241 | ADD | -0.07481 | 0.0195 | 14.71 | 3.903 | | loci_61 | Nominal replication |
| 11.69 | 3.201 | | 0.07177 | 1.001 | 17241 | ADD | -0.07426 | 0.01946 | 14.56 | 3.866 | | loci_61 | Nominal replication |
| 11.73 | 3.212 | | 0.07174 | 1.002 | 17241 | ADD | -0.07411 | 0.01946 | 14.5 | 3.854 | | loci_61 | Nominal replication |
| 11.73 | 3.212 | | 0.07174 | 1.001 | 17241 | ADD | -0.07409 | 0.01946 | 14.49 | 3.852 | | loci_61 | Nominal replication |
| 12.76 | 3.45 | | 0.06967 | 0.9953 | 17241 | ADD | -0.07205 | 0.01978 | 13.26 | 3.567 | | loci_61 | Nominal replication |
| 11.71 | 3.207 | | 0.07176 | 1.001 | 17241 | ADD | -0.07385 | 0.01946 | 14.4 | 3.831 | | loci_61 | Nominal replication |
| 11.71 | 3.207 | | 0.07176 | 1.001 | 17241 | ADD | -0.07385 | 0.01946 | 14.4 | 3.831 | | loci_61 | Nominal replication |
| 11.71 | 3.207 | | 0.07176 | 1.001 | 17241 | ADD | -0.07384 | 0.01946 | 14.4 | 3.83 | | loci_61 | Nominal replication |
| 11.74 | 3.213 | | 0.07171 | 1.001 | 17241 | ADD | -0.07368 | 0.01947 | 14.33 | 3.814 | | loci_61 | Nominal replication |
| 11.67 | 3.197 | | 0.07176 | 1.001 | 17241 | ADD | -0.07385 | 0.01946 | 14.4 | 3.83 | | loci_61 | Nominal replication |
| 11.66 | 3.194 | | 0.07176 | 1.001 | 17241 | ADD | -0.07387 | 0.01946 | 14.41 | 3.832 | | loci_61 | Nominal replication |
| 11.65 | 3.192 | | 0.07176 | 1.001 | 17241 | ADD | -0.07385 | 0.01946 | 14.4 | 3.83 | | loci_61 | Nominal replication |
| 11.65 | 3.192 | | 0.07176 | 1.001 | 17241 | ADD | -0.07385 | 0.01946 | 14.4 | 3.83 | | loci_61 | Nominal replication |
| 11.65 | 3.192 | | 0.07176 | 1.001 | 17241 | ADD | -0.07385 | 0.01946 | 14.4 | 3.83 | | loci_61 | Nominal replication |
| 11.65 | 3.192 | | 0.07176 | 1.001 | 17241 | ADD | -0.07385 | 0.01946 | 14.4 | 3.83 | | loci_61 | Nominal replication |
| 11.65 | 3.192 | | 0.07176 | 1.001 | 17241 | ADD | -0.07385 | 0.01946 | 14.4 | 3.83 | | loci_61 | Nominal replication |
| 11.65 | 3.192 | | 0.07176 | 1.001 | 17241 | ADD | -0.07385 | 0.01946 | 14.4 | 3.83 | | loci_61 | Nominal replication |
| 11.65 | 3.192 | | 0.07176 | 1.001 | 17241 | ADD | -0.07384 | 0.01946 | 14.4 | 3.829 | | loci_61 | Nominal replication |
| 13.16 | 3.543 | | 0.06556 | 0.9902 | 17241 | ADD | -0.07214 | 0.02041 | 12.5 | 3.39 | | loci_61 | Nominal replication |
| 12.84 | 3.47 | | 0.06555 | 0.9916 | 17241 | ADD | -0.07291 | 0.02039 | 12.78 | 3.456 | | loci_61 | Nominal replication |
| 12.55 | 3.403 | | 0.06892 | 0.9815 | 17241 | ADD | -0.07218 | 0.02002 | 13 | 3.506 | | loci_61 | Nominal replication |

| male_CHISQ | male_LOG10P | male_EXTRA | female_A1FREQ | female_INFO | female_N | female_TEST | female_BETA | female_SE | female_CHISQ | female_LOG10P | female_EXTRA | locus | nominal_replication |
|---|---|---|---|---|---|---|---|---|---|---|---|---|---|
| 12.55 | 3.402 | | 0.06892 | 0.9815 | 17241 | ADD | -0.07218 | 0.02002 | 13 | 3.506 | | loci_61 | Nominal replication |
| 12.57 | 3.407 | | 0.0689 | 0.9805 | 17241 | ADD | -0.07209 | 0.02003 | 12.95 | 3.494 | | loci_61 | Nominal replication |
| 12.89 | 3.482 | | 0.06564 | 0.9912 | 17241 | ADD | -0.07171 | 0.02038 | 12.38 | 3.362 | | loci_61 | Nominal replication |
| 20.41 | 5.204 | | 0.4818 | 1.008 | 19427 | ADD | -0.02616 | 0.009451 | 7.664 | 2.249 | | loci_singleton_277 | No replication available |
| 25.65 | 6.387 | | 0.1548 | 0.9926 | 19242 | ADD | -0.02803 | 0.01284 | 4.763 | 1.537 | | loci_singleton_278 | Nominal replication |
| 25.95 | 6.455 | | 0.1731 | 0.9959 | 19541 | ADD | -0.02252 | 0.0131 | 2.953 | 1.067 | | loci_81 | Not replicated |
| 25.71 | 6.401 | | 0.173 | 1.003 | 19541 | ADD | -0.02215 | 0.01306 | 2.876 | 1.046 | | loci_81 | Not replicated |
| 5.812 | 1.798 | | 0.4836 | 0.9968 | 19541 | ADD | -0.04747 | 0.009901 | 22.99 | 5.788 | | loci_singleton_281 | Not replicated |
| 11.82 | 3.233 | | 0.1887 | 0.9928 | 18580 | ADD | -0.04799 | 0.01286 | 13.93 | 3.722 | | loci_44 | Not replicated |
| 11.82 | 3.232 | | 0.1886 | 0.993 | 18580 | ADD | -0.04779 | 0.01286 | 13.81 | 3.695 | | loci_44 | Not replicated |
| 11.56 | 3.171 | | 0.1886 | 0.9872 | 18580 | ADD | -0.04838 | 0.0129 | 14.07 | 3.754 | | loci_44 | Not replicated |
| 11.66 | 3.194 | | 0.1887 | 0.9949 | 18580 | ADD | -0.04771 | 0.01284 | 13.8 | 3.691 | | loci_44 | Not replicated |
| 11.85 | 3.24 | | 0.1887 | 0.9926 | 18580 | ADD | -0.04731 | 0.01286 | 13.54 | 3.631 | | loci_44 | Not replicated |
| 11.28 | 3.106 | | 0.1887 | 0.9965 | 18580 | ADD | -0.04659 | 0.01283 | 13.18 | 3.549 | | loci_44 | Not replicated |
| 11.26 | 3.101 | | 0.1898 | 1.003 | 18580 | ADD | -0.04632 | 0.01276 | 13.17 | 3.545 | | loci_44 | Not replicated |
| 233.22 | 51.93 | | 0.07909 | 1.014 | 18095 | ADD | -0.4677 | 0.01843 | 644.29 | 141.41 | | loci_46 | Significant replication |
| 26.51 | 6.582 | | 0.3235 | 0.9946 | 18095 | ADD | 0.02237 | 0.01057 | 4.477 | 1.464 | | loci_singleton_290 | Not replicated |
| 115.29 | 26.17 | | 0.1075 | 1.017 | 18095 | ADD | -0.309 | 0.01603 | 371.49 | 82.05 | | loci_46 | Significant replication |
| 114.75 | 26.05 | | 0.1088 | 1.015 | 18095 | ADD | -0.3052 | 0.01596 | 365.64 | 80.78 | | loci_46 | Significant replication |
| 14.39 | 3.828 | | 0.07261 | 0.9637 | 19052 | ADD | -0.05112 | 0.0187 | 7.473 | 2.203 | | loci_singleton_293 | No replication available |
| 12.27 | 3.336 | | 0.106 | 0.9933 | 27996 | ADD | 0.04303 | 0.01214 | 12.56 | 3.404 | | loci_singleton_294 | Not replicated |
| 15.19 | 4.012 | | 0.4606 | 0.9963 | 15257 | ADD | 0.03954 | 0.01121 | 12.45 | 3.378 | | loci_27 | Not replicated |
| 15.13 | 3.999 | | 0.4652 | 0.9943 | 15257 | ADD | 0.03803 | 0.01121 | 11.5 | 3.158 | | loci_27 | Not replicated |
| 15.13 | 3.999 | | 0.4652 | 0.9943 | 15257 | ADD | 0.03803 | 0.01121 | 11.5 | 3.158 | | loci_27 | Not replicated |
| 14.23 | 3.791 | | 0.4622 | 0.9997 | 15257 | ADD | 0.03893 | 0.01119 | 12.11 | 3.301 | | loci_27 | Not replicated |
| 13.92 | 3.72 | | 0.4719 | 0.9945 | 15257 | ADD | 0.03829 | 0.0112 | 11.68 | 3.2 | | loci_27 | Not replicated |
| 32.7 | 7.967 | | 0.06415 | 0.9591 | 17589 | ADD | -0.02342 | 0.01902 | 1.515 | 0.6608 | | loci_singleton_300 | No replication available |
| male_CHISQ | male_LOG10P | male_EXTRA | female_A1FREQ | female_INFO | female_N | female_TEST | female_BETA | female_SE | female_CHISQ | female_LOG10P | female_EXTRA | locus | nominal_replication |
| 13.57 | 3.639 | | 0.4005 | 1 | 16703 | ADD | 0.03933 | 0.0104 | 14.29 | 3.806 | | loci_48 | Not replicated |
| 13.92 | 3.719 | | 0.4005 | 1.001 | 16703 | ADD | 0.03845 | 0.0104 | 13.67 | 3.661 | | loci_48 | Not replicated |
| 13.31 | 3.579 | | 0.4004 | 0.9998 | 16703 | ADD | 0.03915 | 0.01041 | 14.15 | 3.773 | | loci_48 | Not replicated |
| 13.4 | 3.599 | | 0.4006 | 1.001 | 16703 | ADD | 0.03895 | 0.0104 | 14.02 | 3.743 | | loci_48 | Not replicated |
| 13.73 | 3.676 | | 0.4008 | 1.001 | 16703 | ADD | 0.03826 | 0.0104 | 13.54 | 3.632 | | loci_48 | Not replicated |
| 13.74 | 3.678 | | 0.4008 | 1.001 | 16703 | ADD | 0.03823 | 0.0104 | 13.51 | 3.626 | | loci_48 | Not replicated |
| 13.74 | 3.678 | | 0.4009 | 1.001 | 16703 | ADD | 0.03821 | 0.0104 | 13.51 | 3.624 | | loci_48 | Not replicated |
| 13.75 | 3.68 | | 0.4008 | 1.001 | 16703 | ADD | 0.03819 | 0.0104 | 13.49 | 3.62 | | loci_48 | Not replicated |
| 13.17 | 3.545 | | 0.4007 | 1 | 16703 | ADD | 0.03904 | 0.0104 | 14.08 | 3.756 | | loci_48 | Not replicated |
| 13.43 | 3.607 | | 0.4017 | 1.001 | 16703 | ADD | 0.03852 | 0.0104 | 13.73 | 3.676 | | loci_48 | Not replicated |
| 13.15 | 3.541 | | 0.4011 | 0.9994 | 16703 | ADD | 0.03896 | 0.01041 | 14.02 | 3.742 | | loci_48 | Not replicated |
| 13.22 | 3.557 | | 0.4012 | 0.9996 | 16703 | ADD | 0.03876 | 0.01041 | 13.88 | 3.709 | | loci_48 | Not replicated |
| 13.22 | 3.558 | | 0.4013 | 0.9999 | 16703 | ADD | 0.03869 | 0.0104 | 13.83 | 3.698 | | loci_48 | Not replicated |
| 13.6 | 3.645 | | 0.4002 | 1.001 | 16703 | ADD | 0.03789 | 0.0104 | 13.27 | 3.569 | | loci_48 | Not replicated |
| 12.91 | 3.486 | | 0.4005 | 0.9997 | 16703 | ADD | 0.03895 | 0.01041 | 14 | 3.739 | | loci_48 | Not replicated |
| 13.03 | 3.514 | | 0.4027 | 1.005 | 16703 | ADD | 0.03837 | 0.01037 | 13.69 | 3.666 | | loci_48 | Not replicated |
| 13.1 | 3.529 | | 0.4017 | 1.001 | 16703 | ADD | 0.03835 | 0.0104 | 13.61 | 3.647 | | loci_48 | Not replicated |
| 17.05 | 4.44 | | 0.1346 | 0.9907 | 15677 | ADD | -0.04707 | 0.01601 | 8.644 | 2.484 | | loci_85 | No replication available |

| male_CHISQ | male_LOG10P | male_EXTRA | female_A1FREQ | female_INFO | female_N | female_TEST | female_BETA | female_SE | female_CHISQ | female_LOG10P | female_EXTRA | locus | nominal_replication |
|---|---|---|---|---|---|---|---|---|---|---|---|---|---|
| 16.81 | 4.385 | | 0.1324 | 0.9907 | 15677 | ADD | -0.04627 | 0.01612 | 8.235 | 2.386 | | loci_85 | No replication available |
| 16.86 | 4.394 | | 0.1323 | 0.991 | 15677 | ADD | -0.04617 | 0.01612 | 8.2 | 2.378 | | loci_85 | No replication available |
| 11.43 | 3.14 | | 0.1308 | 0.9778 | 25616 | ADD | 0.04872 | 0.01309 | 13.85 | 3.704 | | loci_4 | Not replicated |
| 15.27 | 4.03 | | 0.4835 | 0.9899 | 25616 | ADD | 0.02693 | 0.00878 | 9.409 | 2.666 | | loci_7 | Not replicated |
| 13.31 | 3.577 | | 0.476 | 0.9921 | 25616 | ADD | 0.02915 | 0.008775 | 11.03 | 3.048 | | loci_7 | Not replicated |
| 8.088 | 2.351 | | 0.1388 | 0.9902 | 25616 | ADD | 0.05211 | 0.01268 | 16.88 | 4.401 | | loci_4 | Not replicated |
| 19.82 | 5.069 | | 0.3752 | 0.9971 | 25672 | ADD | -0.02397 | 0.008732 | 7.534 | 2.218 | | loci_47 | Not replicated |
| 17.35 | 4.507 | | 0.363 | 0.9844 | 25672 | ADD | -0.02504 | 0.008847 | 8.011 | 2.333 | | loci_47 | Not replicated |
| 17.42 | 4.524 | | 0.3628 | 0.9847 | 25672 | ADD | -0.02493 | 0.008848 | 7.938 | 2.315 | | loci_47 | Not replicated |
| 17.01 | 4.43 | | 0.3626 | 0.9844 | 25672 | ADD | -0.02493 | 0.00885 | 7.934 | 2.314 | | loci_47 | Not replicated |
| 12.71 | 3.44 | | 0.07869 | 0.9896 | 14842 | ADD | -0.07032 | 0.02022 | 12.1 | 3.297 | | loci_singleton_329 | Not replicated |
| 17.19 | 4.472 | | 0.1873 | 0.9888 | 14884 | ADD | 0.04572 | 0.01402 | 10.63 | 2.953 | | loci_58 | No replication available |
| 16.27 | 4.259 | | 0.1921 | 0.9943 | 14884 | ADD | 0.0456 | 0.01385 | 10.83 | 3.002 | | loci_58 | No replication available |
| 14.63 | 3.884 | | 0.1414 | 0.9854 | 14884 | ADD | 0.05383 | 0.01573 | 11.71 | 3.207 | | loci_58 | No replication available |
| 12.81 | 3.463 | | 0.243 | 0.9597 | 15127 | ADD | -0.04364 | 0.01258 | 12.03 | 3.281 | | loci_55 | Not replicated |
| 12.75 | 3.449 | | 0.2432 | 0.9601 | 15127 | ADD | -0.0434 | 0.01258 | 11.91 | 3.254 | | loci_55 | Not replicated |
| 11.27 | 3.104 | | 0.1097 | 0.9847 | 14409 | ADD | 0.07418 | 0.01945 | 14.54 | 3.862 | | loci_25 | Not replicated |
| 10.72 | 2.975 | | 0.109 | 0.9869 | 14409 | ADD | 0.07385 | 0.01949 | 14.36 | 3.821 | | loci_25 | Not replicated |
| 10.39 | 2.897 | | 0.1095 | 0.9874 | 14409 | ADD | 0.07417 | 0.01944 | 14.55 | 3.866 | | loci_25 | Not replicated |
| 10.05 | 2.817 | | 0.1095 | 0.9879 | 14409 | ADD | 0.07489 | 0.01944 | 14.85 | 3.934 | | loci_25 | Not replicated |
| 10.02 | 2.811 | | 0.1095 | 0.9874 | 14409 | ADD | 0.07445 | 0.01944 | 14.66 | 3.891 | | loci_25 | Not replicated |
| 10.03 | 2.812 | | 0.1096 | 0.988 | 14409 | ADD | 0.07434 | 0.01943 | 14.64 | 3.885 | | loci_25 | Not replicated |
| 10.02 | 2.81 | | 0.1096 | 0.988 | 14409 | ADD | 0.07434 | 0.01943 | 14.64 | 3.885 | | loci_25 | Not replicated |
| 10.02 | 2.81 | | 0.1096 | 0.988 | 14409 | ADD | 0.07434 | 0.01943 | 14.64 | 3.885 | | loci_25 | Not replicated |
| 10.02 | 2.81 | | 0.1096 | 0.988 | 14409 | ADD | 0.07434 | 0.01943 | 14.64 | 3.885 | | loci_25 | Not replicated |
| 17.91 | 4.636 | | 0.404 | 0.9823 | 13581 | ADD | -0.03513 | 0.01204 | 8.515 | 2.453 | | loci_34 | Not replicated |
| 17.91 | 4.636 | | 0.404 | 0.9822 | 13581 | ADD | -0.03511 | 0.01204 | 8.504 | 2.451 | | loci_34 | Not replicated |
| 17.67 | 4.581 | | 0.4089 | 0.9842 | 13581 | ADD | -0.03541 | 0.01201 | 8.697 | 2.497 | | loci_34 | Not replicated |
| 17.61 | 4.567 | | 0.4089 | 0.9846 | 13581 | ADD | -0.03546 | 0.012 | 8.728 | 2.504 | | loci_34 | Not replicated |
| 17.6 | 4.565 | | 0.4089 | 0.9847 | 13581 | ADD | -0.03546 | 0.012 | 8.727 | 2.504 | | loci_34 | Not replicated |
| 17.6 | 4.565 | | 0.4089 | 0.9847 | 13581 | ADD | -0.03546 | 0.012 | 8.726 | 2.504 | | loci_34 | Not replicated |
| 17.6 | 4.565 | | 0.4089 | 0.9847 | 13581 | ADD | -0.03546 | 0.012 | 8.726 | 2.503 | | loci_34 | Not replicated |
| 17.6 | 4.565 | | 0.4089 | 0.9847 | 13581 | ADD | -0.03546 | 0.012 | 8.726 | 2.503 | | loci_34 | Not replicated |
| 17.7 | 4.586 | | 0.4089 | 0.9842 | 13581 | ADD | -0.03529 | 0.01201 | 8.64 | 2.483 | | loci_34 | Not replicated |
| 17.6 | 4.564 | | 0.4089 | 0.9846 | 13581 | ADD | -0.03546 | 0.012 | 8.728 | 2.504 | | loci_34 | Not replicated |
| 17.59 | 4.562 | | 0.4089 | 0.9846 | 13581 | ADD | -0.03547 | 0.012 | 8.729 | 2.504 | | loci_34 | Not replicated |
| 17.6 | 4.564 | | 0.4089 | 0.9846 | 13581 | ADD | -0.03545 | 0.012 | 8.721 | 2.502 | | loci_34 | Not replicated |
| 17.57 | 4.558 | | 0.4065 | 0.9945 | 13581 | ADD | -0.03532 | 0.01196 | 8.728 | 2.504 | | loci_34 | Not replicated |
| 17.73 | 4.594 | | 0.4095 | 0.9838 | 13581 | ADD | -0.03512 | 0.01201 | 8.555 | 2.463 | | loci_34 | Not replicated |
| 17.73 | 4.594 | | 0.4089 | 0.9844 | 13581 | ADD | -0.0351 | 0.012 | 8.547 | 2.461 | | loci_34 | Not replicated |
| 17.57 | 4.558 | | 0.4089 | 0.9845 | 13581 | ADD | -0.03533 | 0.012 | 8.661 | 2.488 | | loci_34 | Not replicated |
| 17.61 | 4.567 | | 0.4089 | 0.9846 | 13581 | ADD | -0.03521 | 0.012 | 8.606 | 2.475 | | loci_34 | Not replicated |

| male_CHISQ | male_LOG10P | male_EXTRA | female_A1FREQ | female_INFO | female_N | female_TEST | female_BETA | female_SE | female_CHISQ | female_LOG10P | female_EXTRA | locus | nominal_replication |
|---|---|---|---|---|---|---|---|---|---|---|---|---|---|
| 17.61 | 4.567 | | 0.4089 | 0.9846 | 13581 | ADD | -0.03521 | 0.012 | 8.606 | 2.475 | | loci_34 | Not replicated |
| 17.61 | 4.566 | | 0.4089 | 0.9846 | 13581 | ADD | -0.03522 | 0.012 | 8.607 | 2.475 | | loci_34 | Not replicated |
| 17.59 | 4.562 | | 0.4089 | 0.9846 | 13581 | ADD | -0.03522 | 0.012 | 8.608 | 2.475 | | loci_34 | Not replicated |

| male_CHISQ | male_LOG10P | male_EXTRA | female_A1FREQ | female_INFO | female_N | female_TEST | female_BETA | female_SE | female_CHISQ | female_LOG10P | female_EXTRA | locus | nominal_replication |
|---|---|---|---|---|---|---|---|---|---|---|---|---|---|
| 17.58 | 4.561 | | 0.4088 | 0.9839 | 13581 | ADD | -0.03518 | 0.01201 | 8.582 | 2.469 | | loci_34 | Not replicated |
| 17.4 | 4.519 | | 0.4044 | 0.9877 | 13581 | ADD | -0.03541 | 0.01201 | 8.697 | 2.497 | | loci_34 | Not replicated |
| 17.47 | 4.535 | | 0.4044 | 0.9875 | 13581 | ADD | -0.03524 | 0.01201 | 8.617 | 2.477 | | loci_34 | Not replicated |
| 17.47 | 4.535 | | 0.4044 | 0.9875 | 13581 | ADD | -0.03524 | 0.01201 | 8.615 | 2.477 | | loci_34 | Not replicated |
| 17.81 | 4.613 | | 0.4028 | 0.9826 | 13581 | ADD | -0.03466 | 0.01204 | 8.284 | 2.398 | | loci_34 | Not replicated |
| 17.49 | 4.539 | | 0.4093 | 0.9836 | 13581 | ADD | -0.03512 | 0.01201 | 8.554 | 2.463 | | loci_34 | Not replicated |
| 17.42 | 4.523 | | 0.4044 | 0.9878 | 13581 | ADD | -0.03521 | 0.012 | 8.603 | 2.474 | | loci_34 | Not replicated |
| 17.56 | 4.555 | | 0.404 | 0.9869 | 13581 | ADD | -0.03494 | 0.01201 | 8.461 | 2.44 | | loci_34 | Not replicated |
| 18.05 | 4.668 | | 0.4011 | 0.9888 | 13581 | ADD | -0.03404 | 0.01202 | 8.02 | 2.335 | | loci_34 | Not replicated |
| 17.52 | 4.546 | | 0.404 | 0.9872 | 13581 | ADD | -0.03493 | 0.01201 | 8.46 | 2.44 | | loci_34 | Not replicated |
| 17.49 | 4.539 | | 0.404 | 0.9872 | 13581 | ADD | -0.03492 | 0.01201 | 8.455 | 2.439 | | loci_34 | Not replicated |
| 17.39 | 4.517 | | 0.4093 | 0.9834 | 13581 | ADD | -0.03506 | 0.01201 | 8.521 | 2.454 | | loci_34 | Not replicated |
| 17.28 | 4.491 | | 0.406 | 0.9948 | 13581 | ADD | -0.03508 | 0.01195 | 8.611 | 2.476 | | loci_34 | Not replicated |
| 18.11 | 4.68 | | 0.4006 | 0.9885 | 13581 | ADD | -0.03358 | 0.01202 | 7.802 | 2.282 | | loci_34 | Not replicated |
| 17.36 | 4.51 | | 0.4031 | 0.9881 | 13581 | ADD | -0.03488 | 0.01201 | 8.436 | 2.434 | | loci_34 | Not replicated |
| 17.33 | 4.502 | | 0.4039 | 0.9869 | 13581 | ADD | -0.03479 | 0.01201 | 8.387 | 2.423 | | loci_34 | Not replicated |
| 17.25 | 4.484 | | 0.4039 | 0.9868 | 13581 | ADD | -0.03471 | 0.01201 | 8.351 | 2.414 | | loci_34 | Not replicated |
| 17.81 | 4.613 | | 0.4025 | 0.9964 | 13581 | ADD | -0.0334 | 0.01197 | 7.789 | 2.279 | | loci_34 | Not replicated |
| 17.34 | 4.506 | | 0.4026 | 0.9879 | 13581 | ADD | -0.03434 | 0.01201 | 8.175 | 2.372 | | loci_34 | Not replicated |
| 16.76 | 4.371 | | 0.4106 | 0.9835 | 13581 | ADD | -0.03524 | 0.012 | 8.622 | 2.479 | | loci_34 | Not replicated |
| 16.71 | 4.362 | | 0.4106 | 0.9835 | 13581 | ADD | -0.0352 | 0.012 | 8.602 | 2.474 | | loci_34 | Not replicated |
| 16.72 | 4.362 | | 0.4106 | 0.9835 | 13581 | ADD | -0.0352 | 0.012 | 8.601 | 2.474 | | loci_34 | Not replicated |
| 17.22 | 4.479 | | 0.4026 | 0.9881 | 13581 | ADD | -0.03422 | 0.01201 | 8.116 | 2.358 | | loci_34 | Not replicated |
| 17.08 | 4.446 | | 0.1972 | 0.9899 | 26726 | ADD | 0.02579 | 0.009827 | 6.89 | 2.062 | | loci_86 | Not replicated |
| 15.78 | 4.148 | | 0.196 | 0.9893 | 26726 | ADD | 0.02645 | 0.009853 | 7.206 | 2.139 | | loci_86 | Not replicated |
| 15.61 | 4.109 | | 0.1959 | 0.9898 | 26726 | ADD | 0.02662 | 0.009852 | 7.301 | 2.162 | | loci_86 | Not replicated |
| 15.66 | 4.119 | | 0.1959 | 0.9899 | 26726 | ADD | 0.02646 | 0.009852 | 7.213 | 2.14 | | loci_86 | Not replicated |
| 15.61 | 4.109 | | 0.1959 | 0.9898 | 26726 | ADD | 0.02647 | 0.009852 | 7.22 | 2.142 | | loci_86 | Not replicated |
| 15.46 | 4.074 | | 0.1961 | 0.9897 | 26726 | ADD | 0.02667 | 0.009849 | 7.335 | 2.17 | | loci_86 | Not replicated |
| 15.11 | 3.995 | | 0.196 | 0.9898 | 26726 | ADD | 0.02658 | 0.00985 | 7.281 | 2.157 | | loci_86 | Not replicated |
| 12.58 | 3.409 | | 0.2829 | 0.9991 | 26726 | ADD | -0.026 | 0.008646 | 9.041 | 2.578 | | loci_26 | Not replicated |
| 14.07 | 3.755 | | 0.2211 | 0.9872 | 26726 | ADD | 0.02572 | 0.009439 | 7.423 | 2.191 | | loci_86 | Not replicated |
| 14.07 | 3.754 | | 0.221 | 0.987 | 26726 | ADD | 0.02572 | 0.00944 | 7.423 | 2.191 | | loci_86 | Not replicated |
| 14.24 | 3.794 | | 0.221 | 0.9874 | 26726 | ADD | 0.02533 | 0.009437 | 7.206 | 2.139 | | loci_86 | Not replicated |
| 12.42 | 3.372 | | 0.2913 | 0.9918 | 26726 | ADD | -0.02545 | 0.008597 | 8.76 | 2.512 | | loci_26 | Not replicated |
| 14.02 | 3.744 | | 0.2205 | 0.9848 | 26726 | ADD | 0.02537 | 0.009459 | 7.192 | 2.135 | | loci_86 | Not replicated |
| 13.87 | 3.707 | | 0.2212 | 0.9866 | 26726 | ADD | 0.02554 | 0.009439 | 7.32 | 2.166 | | loci_86 | Not replicated |
| 14.04 | 3.748 | | 0.2205 | 0.9848 | 26726 | ADD | 0.02531 | 0.009459 | 7.16 | 2.128 | | loci_86 | Not replicated |
| 15.03 | 3.976 | | 0.1784 | 0.9903 | 26726 | ADD | 0.02559 | 0.01021 | 6.277 | 1.912 | | loci_86 | Not replicated |
| 15.9 | 4.176 | | 0.4819 | 1.003 | 10737 | ADD | -0.04911 | 0.01282 | 14.66 | 3.891 | | loci_67 | Not replicated |
| 15.93 | 4.183 | | 0.4831 | 0.998 | 10737 | ADD | -0.04905 | 0.01285 | 14.57 | 3.869 | | loci_67 | Not replicated |
| 15.98 | 4.193 | | 0.4827 | 0.9975 | 10737 | ADD | -0.04812 | 0.01286 | 14.01 | 3.741 | | loci_67 | Not replicated |
| 14.2 | 3.785 | | 0.4923 | 0.9992 | 10737 | ADD | -0.04753 | 0.01284 | 13.7 | 3.669 | | loci_67 | Not replicated |
| 13.89 | 3.713 | | 0.4923 | 0.9994 | 10737 | ADD | -0.04746 | 0.01284 | 13.66 | 3.659 | | loci_67 | Not replicated |
| 13.99 | 3.735 | | 0.4929 | 0.9981 | 10737 | ADD | -0.04702 | 0.01285 | 13.39 | 3.597 | | loci_67 | Not replicated |
| 16.78 | 4.376 | | 0.4869 | 1.003 | 10737 | ADD | 0.04081 | 0.0128 | 10.16 | 2.843 | | loci_67 | Not replicated |
| 16.52 | 4.317 | | 0.487 | 1.003 | 10737 | ADD | 0.04078 | 0.0128 | 10.15 | 2.84 | | loci_67 | Not replicated |

| male_CHISQ | male_LOG10P | male_EXTRA | female_A1FREQ | female_INFO | female_N | female_TEST | female_BETA | female_SE | female_CHISQ | female_LOG10P | female_EXTRA | locus | nominal_replication |
|---|---|---|---|---|---|---|---|---|---|---|---|---|---|
| 6.04 | 1.854 | | 0.4332 | 1.005 | 8019 | ADD | 0.06934 | 0.01445 | 23.02 | 5.794 | | loci_singleton_411 | No replication available |
| 20.53 | 5.231 | | 0.482 | 1.007 | 8469 | ADD | -0.04028 | 0.01442 | 7.804 | 2.283 | | loci_67 | Not replicated |
| 20.41 | 5.205 | | 0.4807 | 1.012 | 8469 | ADD | -0.04032 | 0.01439 | 7.855 | 2.295 | | loci_67 | Not replicated |
| 20.42 | 5.207 | | 0.4816 | 1.007 | 8469 | ADD | -0.04023 | 0.01442 | 7.782 | 2.278 | | loci_67 | Not replicated |
| 18.67 | 4.808 | | 0.4911 | 1.007 | 8469 | ADD | -0.03989 | 0.01441 | 7.658 | 2.248 | | loci_67 | Not replicated |
| 18.51 | 4.772 | | 0.4905 | 1.008 | 8469 | ADD | -0.04003 | 0.01441 | 7.719 | 2.263 | | loci_67 | Not replicated |
| 19.35 | 4.963 | | 0.1897 | 0.9976 | 7336 | ADD | -0.06123 | 0.01918 | 10.19 | 2.85 | | loci_79 | Not replicated |
| 19.26 | 4.943 | | 0.1897 | 0.9976 | 7336 | ADD | -0.06123 | 0.01918 | 10.19 | 2.85 | | loci_79 | Not replicated |
| 23.33 | 5.865 | | 0.05569 | 0.9877 | 7336 | ADD | -0.07888 | 0.03328 | 5.619 | 1.75 | | loci_singleton_419 | Not replicated |
| 17.02 | 4.433 | | 0.1899 | 0.999 | 7283 | ADD | -0.06734 | 0.01904 | 12.5 | 3.391 | | loci_79 | No replication available |
| male_CHISQ | male_LOG10P | male_EXTRA | female_A1FREQ | female_INFO | female_N | female_TEST | female_BETA | female_SE | female_CHISQ | female_LOG10P | female_EXTRA | locus | nominal_replication |
| 17.01 | 4.43 | | 0.1899 | 0.999 | 7283 | ADD | -0.06734 | 0.01904 | 12.5 | 3.391 | | loci_79 | No replication available |
| 13.74 | 3.678 | | 0.1855 | 1.001 | 7283 | ADD | -0.06685 | 0.01919 | 12.13 | 3.304 | | loci_79 | No replication available |
| 12.79 | 3.459 | | 0.1784 | 1.001 | 25624 | ADD | 0.03464 | 0.009895 | 12.26 | 3.334 | | loci_70 | No replication available |
| 13.24 | 3.563 | | 0.1806 | 1.001 | 25624 | ADD | 0.03376 | 0.00985 | 11.75 | 3.216 | | loci_70 | No replication available |
| 13 | 3.505 | | 0.1779 | 1.001 | 25624 | ADD | 0.03416 | 0.009909 | 11.89 | 3.248 | | loci_70 | No replication available |
| 12.46 | 3.382 | | 0.1784 | 1.001 | 25624 | ADD | 0.03436 | 0.009897 | 12.05 | 3.286 | | loci_70 | No replication available |
| 11.29 | 3.109 | | 0.1643 | 0.98 | 7040 | ADD | -0.09694 | 0.02324 | 17.4 | 4.518 | | loci_75 | No replication available |
| 11.77 | 3.219 | | 0.1641 | 0.9764 | 7040 | ADD | -0.09481 | 0.0233 | 16.56 | 4.326 | | loci_75 | No replication available |
| 11.7 | 3.204 | | 0.2087 | 0.9933 | 7197 | ADD | 0.08215 | 0.02113 | 15.11 | 3.994 | | loci_singleton_429 | No replication available |
| 16.05 | 4.21 | | 0.489 | 1.013 | 7074 | ADD | -0.0591 | 0.01738 | 11.57 | 3.173 | | loci_5 | No replication available |
| 18.09 | 4.675 | | 0.3437 | 0.9902 | 7074 | ADD | -0.05469 | 0.01849 | 8.751 | 2.509 | | loci_5 | No replication available |
| 11.46 | 3.147 | | 0.06056 | 0.9096 | 6926 | ADD | -0.1678 | 0.03809 | 19.4 | 4.975 | | loci_singleton_432 | No replication available |
| 18.38 | 4.742 | | 0.4865 | 1.017 | 7089 | ADD | -0.0512 | 0.0168 | 9.289 | 2.637 | | loci_singleton_433 | No replication available |
| 17.45 | 4.531 | | 0.1612 | 0.9911 | 7224 | ADD | 0.07189 | 0.02295 | 9.814 | 2.762 | | loci_20 | No replication available |
| 15.94 | 4.185 | | 0.1422 | 0.9957 | 7224 | ADD | 0.07814 | 0.0241 | 10.52 | 2.927 | | loci_20 | No replication available |
| 40.1 | 9.619 | | 0.2494 | 1.019 | 6711 | ADD | -0.2602 | 0.01945 | 179.01 | 40.1 | | loci_62 | Significant replication |
| 37.06 | 8.941 | | 0.2425 | 1.017 | 6711 | ADD | -0.2578 | 0.01961 | 172.85 | 38.75 | | loci_62 | Significant replication |
| 37.24 | 8.982 | | 0.2425 | 1.017 | 6711 | ADD | -0.2578 | 0.01961 | 172.85 | 38.75 | | loci_62 | Significant replication |
| 35.8 | 8.66 | | 0.244 | 1.011 | 6711 | ADD | -0.2556 | 0.01962 | 169.58 | 38.04 | | loci_62 | Significant replication |
| 38.56 | 9.275 | | 0.2722 | 1.022 | 6711 | ADD | -0.2496 | 0.01887 | 174.91 | 39.2 | | loci_62 | Significant replication |
| 35.5 | 8.593 | | 0.2555 | 1.019 | 6711 | ADD | -0.2489 | 0.01929 | 166.46 | 37.36 | | loci_62 | Significant replication |
| 35.82 | 8.665 | | 0.255 | 1.02 | 6711 | ADD | -0.248 | 0.01929 | 165.34 | 37.11 | | loci_62 | Significant replication |
| 26.4 | 6.557 | | 0.2212 | 1.019 | 6711 | ADD | -0.2426 | 0.02021 | 144.06 | 32.46 | | loci_62 | Significant replication |
| 36.23 | 8.756 | | 0.2478 | 1.017 | 6711 | ADD | -0.2516 | 0.01948 | 166.88 | 37.45 | | loci_62 | Significant replication |
| 29.31 | 7.209 | | 0.2107 | 1.02 | 6711 | ADD | -0.2505 | 0.02054 | 148.84 | 33.51 | | loci_62 | Significant replication |
| 35.37 | 8.563 | | 0.2385 | 1.018 | 6711 | ADD | -0.2528 | 0.01972 | 164.30 | 36.89 | | loci_62 | Significant replication |
| 34.21 | 8.305 | | 0.2409 | 1.025 | 6711 | ADD | -0.2489 | 0.0196 | 161.27 | 36.22 | | loci_62 | Significant replication |
| 9.073 | 2.586 | | 0.2693 | 1.015 | 5055 | ADD | 0.09418 | 0.02073 | 20.64 | 5.256 | | loci_singleton_448 | No replication available |
| 7.749 | 2.27 | | 0.06105 | 0.986 | 5664 | ADD | 0.1729 | 0.03738 | 21.4 | 5.429 | | loci_43 | Not replicated |
| 7.747 | 2.269 | | 0.06123 | 0.9856 | 5664 | ADD | 0.1722 | 0.03733 | 21.27 | 5.398 | | loci_43 | Not replicated |
| 26.58 | 6.597 | | 0.427 | 0.9712 | 7048 | ADD | -0.0483 | 0.01603 | 9.077 | 2.587 | | loci_77 | Not replicated |
| 26.74 | 6.634 | | 0.4184 | 0.9758 | 7048 | ADD | -0.04738 | 0.01604 | 8.723 | 2.503 | | loci_77 | Not replicated |
| 26.74 | 6.633 | | 0.4186 | 0.975 | 7048 | ADD | -0.04737 | 0.01605 | 8.715 | 2.501 | | loci_77 | Not replicated |
| 26.71 | 6.626 | | 0.4178 | 0.976 | 7048 | ADD | -0.04738 | 0.01604 | 8.723 | 2.503 | | loci_77 | Not replicated |
| 26.73 | 6.63 | | 0.4184 | 0.976 | 7048 | ADD | -0.04732 | 0.01604 | 8.704 | 2.498 | | loci_77 | Not replicated |

| male_CHISQ | male_LOG10P | male_EXTRA | female_A1FREQ | female_INFO | female_N | female_TEST | female_BETA | female_SE | female_CHISQ | female_LOG10P | female_EXTRA | locus | nominal_replication |
|---|---|---|---|---|---|---|---|---|---|---|---|---|---|
| 26.29 | 6.533 | | 0.4197 | 1.001 | 7048 | ADD | -0.04767 | 0.01582 | 9.075 | 2.586 | | loci_77 | Not replicated |
| 26.66 | 6.616 | | 0.4182 | 0.9762 | 7048 | ADD | -0.04729 | 0.01604 | 8.694 | 2.496 | | loci_77 | Not replicated |
| 27.17 | 6.731 | | 0.4182 | 0.9738 | 7048 | ADD | -0.04543 | 0.01606 | 8.003 | 2.331 | | loci_77 | Not replicated |
| 26.92 | 6.673 | | 0.4187 | 0.9722 | 7048 | ADD | -0.04585 | 0.01607 | 8.143 | 2.364 | | loci_77 | Not replicated |
| 26.77 | 6.64 | | 0.4098 | 1.007 | 7048 | ADD | -0.04499 | 0.01584 | 8.066 | 2.346 | | loci_77 | Not replicated |
| 21.03 | 5.344 | | 0.4213 | 0.9994 | 7048 | ADD | -0.04708 | 0.01584 | 8.83 | 2.528 | | loci_77 | Not replicated |
| 12.51 | 3.393 | | 0.07989 | 0.8319 | 3085 | ADD | 0.1953 | 0.04899 | 15.88 | 4.172 | | loci_93 | Not replicated |
| 13.57 | 3.639 | | 0.08111 | 0.8579 | 3085 | ADD | 0.1833 | 0.04792 | 14.63 | 3.884 | | loci_93 | Not replicated |
| 9.029 | 2.576 | | 0.179 | 0.9834 | 2730 | ADD | 0.1364 | 0.03003 | 20.64 | 5.257 | | loci_singleton_464 | No replication available |
| 10.43 | 2.907 | | 0.1569 | 1.012 | 2253 | ADD | -0.1434 | 0.03488 | 16.91 | 4.406 | | loci_76 | No replication available |
| 10.52 | 2.928 | | 0.1569 | 1.007 | 2253 | ADD | -0.1426 | 0.03497 | 16.62 | 4.341 | | loci_76 | No replication available |
| 11.83 | 3.234 | | 0.3847 | 0.9869 | 2156 | ADD | -0.1189 | 0.03087 | 14.84 | 3.932 | | loci_singleton_467 | No replication available |
| 6.645 | 2.003 | | 0.4892 | 1.026 | 2102 | ADD | -0.1382 | 0.02538 | 29.68 | 7.292 | | loci_singleton_468 | No replication available |
| 14.75 | 3.912 | | 0.1819 | 1.012 | 2102 | ADD | -0.1112 | 0.03311 | 11.28 | 3.106 | | loci_singleton_469 | No replication available |
| 12.43 | 3.374 | | 0.4807 | 0.9637 | 2347 | ADD | 0.1098 | 0.02757 | 15.86 | 4.167 | | loci_88 | Not replicated |
| 12.82 | 3.466 | | 0.4915 | 0.9685 | 2347 | ADD | -0.09828 | 0.02752 | 12.75 | 3.449 | | loci_88 | Not replicated |
| 12.66 | 3.427 | | 0.4914 | 0.9716 | 2347 | ADD | -0.09831 | 0.02748 | 12.8 | 3.46 | | loci_88 | Not replicated |
| 12.66 | 3.427 | | 0.4913 | 0.9714 | 2347 | ADD | -0.09831 | 0.02748 | 12.8 | 3.46 | | loci_88 | Not replicated |
| 12.66 | 3.427 | | 0.4914 | 0.9716 | 2347 | ADD | -0.0983 | 0.02748 | 12.8 | 3.46 | | loci_88 | Not replicated |
| 12.66 | 3.427 | | 0.4914 | 0.9716 | 2347 | ADD | -0.0983 | 0.02748 | 12.8 | 3.46 | | loci_88 | Not replicated |
| 5.264 | 1.662 | | 0.3521 | 1.014 | 2056 | ADD | -0.1637 | 0.0292 | 31.44 | 7.687 | | loci_18 | No replication available |
| 3.692 | 1.262 | | 0.3393 | 1.021 | 2056 | ADD | -0.1645 | 0.02938 | 31.36 | 7.669 | | loci_18 | No replication available |
| 3.58 | 1.233 | | 0.337 | 1.035 | 2056 | ADD | -0.1619 | 0.02921 | 30.71 | 7.524 | | loci_18 | No replication available |
| 3.695 | 1.263 | | 0.3389 | 1.02 | 2056 | ADD | -0.1609 | 0.0294 | 29.95 | 7.354 | | loci_18 | No replication available |
| 15.46 | 4.074 | | 0.4399 | 1.037 | 1574 | ADD | 0.1366 | 0.03106 | 19.34 | 4.96 | | loci_42 | No replication available |

| male_CHISQ | male_LOG10P | male_EXTRA | female_A1FREQ | female_INFO | female_N | female_TEST | female_BETA | female_SE | female_CHISQ | female_LOG10P | female_EXTRA | locus | nominal_replication |

| male_CHISQ | male_LOG10P | male_EXTRA | female_A1FREQ | female_INFO | female_N | female_TEST | female_BETA | female_SE | female_CHISQ | female_LOG10P | female_EXTRA | locus | nominal_replication |
|---|---|---|---|---|---|---|---|---|---|---|---|---|---|
| 15.35 | 4.049 | | 0.4403 | 1.026 | 1574 | ADD | 0.1375 | 0.03122 | 19.39 | 4.973 | | loci_42 | No replication available |
| 15.2 | 4.014 | | 0.4384 | 1.035 | 1574 | ADD | 0.1368 | 0.0311 | 19.35 | 4.963 | | loci_42 | No replication available |
| 14.63 | 3.884 | | 0.4239 | 0.9724 | 1988 | ADD | -0.09573 | 0.02767 | 11.97 | 3.267 | | loci_singleton_483 | Not replicated |
| 9.957 | 2.795 | | 0.2598 | 0.9534 | 1686 | ADD | -0.1321 | 0.0304 | 18.89 | 4.859 | | loci_singleton_484 | No replication available |
| 18.67 | 4.808 | | 0.06604 | 0.9498 | 1735 | ADD | -0.1927 | 0.0686 | 7.892 | 2.304 | | loci_singleton_485 | No replication available |
| 24.76 | 6.187 | | 0.4768 | 0.9885 | 1735 | ADD | -0.06239 | 0.02969 | 4.417 | 1.449 | | loci_60 | No replication available |
| 24.24 | 6.071 | | 0.4811 | 0.9828 | 1735 | ADD | -0.0645 | 0.02975 | 4.701 | 1.521 | | loci_60 | No replication available |
| 24.68 | 6.169 | | 0.4766 | 0.9882 | 1735 | ADD | -0.06198 | 0.02969 | 4.359 | 1.434 | | loci_60 | No replication available |
| 24.68 | 6.169 | | 0.4766 | 0.9882 | 1735 | ADD | -0.06198 | 0.02969 | 4.359 | 1.434 | | loci_60 | No replication available |
| 24.52 | 6.133 | | 0.4766 | 0.9882 | 1735 | ADD | -0.06198 | 0.02969 | 4.359 | 1.434 | | loci_60 | No replication available |
| 24.16 | 6.052 | | 0.4766 | 0.9868 | 1735 | ADD | -0.0626 | 0.02971 | 4.439 | 1.454 | | loci_60 | No replication available |
| 24.42 | 6.11 | | 0.4769 | 0.9876 | 1735 | ADD | -0.06154 | 0.02971 | 4.292 | 1.417 | | loci_60 | No replication available |
| 24.45 | 6.118 | | 0.4768 | 0.989 | 1735 | ADD | -0.06124 | 0.02968 | 4.256 | 1.408 | | loci_60 | No replication available |
| 19.31 | 4.954 | | 0.1021 | 1.01 | 1735 | ADD | 0.172 | 0.05431 | 10.03 | 2.812 | | loci_singleton_494 | No replication available |
| 18.98 | 4.88 | | 0.06608 | 0.9497 | 1734 | ADD | -0.1918 | 0.06788 | 7.987 | 2.327 | | loci_singleton_495 | No replication available |
| 21.01 | 5.341 | | 0.3455 | 0.9545 | 2100 | ADD | 0.08757 | 0.03311 | 6.995 | 2.088 | | loci_singleton_496 | No replication available |
| 15.01 | 3.97 | | 0.1311 | 1.031 | 1636 | ADD | 0.1526 | 0.04687 | 10.6 | 2.946 | | loci_singleton_497 | No replication available |
| 5.552 | 1.734 | | 0.5027 | 1.017 | 1312 | ADD | 0.1626 | 0.03306 | 24.2 | 6.062 | | loci_78 | Not replicated |
| 5.471 | 1.714 | | 0.4986 | 1.004 | 1312 | ADD | 0.1637 | 0.03327 | 24.21 | 6.064 | | loci_78 | Not replicated |
| 15.31 | 4.041 | | 0.468 | 1.019 | 1296 | ADD | -0.1152 | 0.03491 | 10.89 | 3.014 | | loci_19 | Not replicated |

| male_CHISQ | male_LOG10P | male_EXTRA | female_A1FREQ | female_INFO | female_N | female_TEST | female_BETA | female_SE | female_CHISQ | female_LOG10P | female_EXTRA | locus | nominal_replication |
|---|---|---|---|---|---|---|---|---|---|---|---|---|---|
| 15.2 | 4.015 | | 0.4682 | 1.021 | 1296 | ADD | -0.1155 | 0.03489 | 10.96 | 3.031 | | loci_19 | Not replicated |
| 15.2 | 4.016 | | 0.4681 | 1.02 | 1296 | ADD | -0.1155 | 0.0349 | 10.96 | 3.031 | | loci_19 | Not replicated |
| 17.09 | 4.449 | | 0.1275 | 1.027 | 1559 | ADD | 0.1789 | 0.05194 | 11.86 | 3.242 | | loci_64 | No replication available |
| 17.43 | 4.525 | | 0.1256 | 1.031 | 1559 | ADD | 0.1763 | 0.05216 | 11.43 | 3.141 | | loci_64 | No replication available |
| 8.389 | 2.423 | | 0.4205 | 1.007 | 1562 | ADD | 0.1428 | 0.02876 | 24.65 | 6.162 | | loci_90 | No replication available |
| 10.04 | 2.814 | | 0.3616 | 0.9923 | 1562 | ADD | 0.1333 | 0.02978 | 20.06 | 5.124 | | loci_90 | No replication available |
| 9.423 | 2.669 | | 0.361 | 0.9948 | 1562 | ADD | 0.1326 | 0.02976 | 19.86 | 5.079 | | loci_90 | No replication available |
| 8.195 | 2.377 | | 0.4205 | 1.007 | 1562 | ADD | 0.1431 | 0.02856 | 25.12 | 6.268 | | loci_90 | No replication available |
| 10.01 | 2.807 | | 0.3616 | 0.9923 | 1562 | ADD | 0.133 | 0.02957 | 20.23 | 5.163 | | loci_90 | No replication available |
| 9.394 | 2.662 | | 0.361 | 0.9948 | 1562 | ADD | 0.1323 | 0.02955 | 20.05 | 5.123 | | loci_90 | No replication available |
| 8.515 | 2.453 | | 0.4277 | 0.9674 | 4794 | ADD | -0.09337 | 0.02057 | 20.59 | 5.246 | | loci_singleton_511 | Not replicated |
| 25.78 | 6.416 | | 0.08533 | 0.9873 | 3623 | ADD | -0.1123 | 0.04061 | 7.642 | 2.244 | | loci_singleton_512 | Not replicated |
| 10.18 | 2.847 | | 0.3881 | 0.9756 | 3623 | ADD | 0.1059 | 0.0234 | 20.47 | 5.217 | | loci_37 | Not replicated |
| 10.17 | 2.845 | | 0.3881 | 0.9756 | 3623 | ADD | 0.1059 | 0.0234 | 20.47 | 5.217 | | loci_37 | Not replicated |
| 10.1 | 2.829 | | 0.3882 | 0.975 | 3623 | ADD | 0.106 | 0.02341 | 20.53 | 5.231 | | loci_37 | Not replicated |
| 8.523 | 2.455 | | 0.3787 | 0.979 | 3623 | ADD | 0.1059 | 0.02346 | 20.38 | 5.197 | | loci_37 | Not replicated |
| 8.512 | 2.452 | | 0.3814 | 0.9793 | 3623 | ADD | 0.1044 | 0.02343 | 19.85 | 5.078 | | loci_37 | Not replicated |
| 8.731 | 2.505 | | 0.3798 | 0.9791 | 3623 | ADD | 0.1036 | 0.02345 | 19.53 | 5.004 | | loci_37 | Not replicated |
| 8.983 | 2.565 | | 0.3939 | 0.9819 | 3623 | ADD | 0.09903 | 0.02326 | 18.13 | 4.686 | | loci_37 | Not replicated |
| 8.2 | 2.378 | | 0.3366 | 0.9676 | 3623 | ADD | 0.1058 | 0.02421 | 19.09 | 4.904 | | loci_37 | Not replicated |
| 15.82 | 4.156 | | 0.2276 | 0.9864 | 2818 | ADD | -0.1142 | 0.03278 | 12.15 | 3.308 | | loci_singleton_521 | Not replicated |
| 22.13 | 5.595 | | 0.09685 | 0.9824 | 3547 | ADD | -0.08814 | 0.03613 | 5.952 | 1.833 | | loci_82 | Nominal replication |
| 21.84 | 5.529 | | 0.09689 | 0.9823 | 3547 | ADD | -0.08895 | 0.03612 | 6.064 | 1.86 | | loci_82 | Nominal replication |
| 21.84 | 5.528 | | 0.09689 | 0.9823 | 3547 | ADD | -0.08895 | 0.03612 | 6.065 | 1.86 | | loci_82 | Nominal replication |
| 17.87 | 4.626 | | 0.2525 | 1.016 | 1806 | ADD | -0.1024 | 0.03563 | 8.267 | 2.394 | | loci_singleton_525 | Not replicated |
| 19.06 | 4.898 | | 0.4834 | 0.9648 | 1168 | ADD | -0.1056 | 0.036 | 8.597 | 2.473 | | loci_singleton_526 | No replication available |
| 18.5 | 4.769 | | 0.07757 | 0.9649 | 1146 | ADD | 0.212 | 0.06999 | 9.175 | 2.61 | | loci_singleton_527 | No replication available |
| 18.31 | 4.726 | | 0.1417 | 0.9729 | 1147 | ADD | -0.162 | 0.05259 | 9.494 | 2.686 | | loci_40 | No replication available |
| 19.11 | 4.908 | | 0.1429 | 0.9651 | 1147 | ADD | -0.1548 | 0.05262 | 8.657 | 2.487 | | loci_40 | No replication available |
| 12.33 | 3.351 | | 0.2271 | 1.036 | 1415 | ADD | 0.1648 | 0.03698 | 19.86 | 5.079 | | loci_66 | Not replicated |
| 12.29 | 3.341 | | 0.2266 | 1.033 | 1415 | ADD | 0.1646 | 0.03706 | 19.74 | 5.052 | | loci_66 | Not replicated |
| 16.83 | 4.389 | | 0.1478 | 0.9987 | 1015 | ADD | 0.1916 | 0.05778 | 11 | 3.039 | | loci_83 | Not replicated |
| 16.56 | 4.326 | | 0.1545 | 0.9491 | 1015 | ADD | 0.1899 | 0.05819 | 10.64 | 2.957 | | loci_83 | Not replicated |
| 13.98 | 3.733 | | 0.3288 | 1.023 | 1061 | ADD | 0.1608 | 0.04195 | 14.68 | 3.896 | | loci_9 | No replication available |
| 13.31 | 3.578 | | 0.3187 | 1.009 | 1061 | ADD | 0.1668 | 0.04259 | 15.33 | 4.045 | | loci_9 | No replication available |
| 13.19 | 3.551 | | 0.2497 | 1.019 | 1396 | ADD | 0.171 | 0.04287 | 15.91 | 4.177 | | loci_singleton_536 | Not replicated |
| 15.34 | 4.047 | | 0.4216 | 1.029 | 925 | ADD | 0.1362 | 0.03625 | 14.12 | 3.766 | | loci_singleton_537 | No replication available |
| 9.216 | 2.62 | | 0.1925 | 0.9882 | 1045 | ADD | 0.2076 | 0.04783 | 18.85 | 4.849 | | loci_singleton_538 | No replication available |
| 10.22 | 2.857 | | 0.2938 | 0.9866 | 878 | ADD | 0.1887 | 0.04578 | 16.99 | 4.424 | | loci_49 | No replication available |
| 13.93 | 3.722 | | 0.3472 | 0.9767 | 878 | ADD | -0.1561 | 0.04358 | 12.83 | 3.466 | | loci_63 | No replication available |
| male_CHISQ | male_LOG10P | male_EXTRA | female_A1FREQ | female_INFO | female_N | female_TEST | female_BETA | female_SE | female_CHISQ | female_LOG10P | female_EXTRA | locus | nominal_replication |
| 9.807 | 2.76 | | 0.3007 | 0.9673 | 878 | ADD | 0.188 | 0.04593 | 16.76 | 4.372 | | loci_49 | No replication available |
| 9.437 | 2.672 | | 0.2994 | 0.9707 | 878 | ADD | 0.1894 | 0.04589 | 17.04 | 4.437 | | loci_49 | No replication available |
| 13.94 | 3.725 | | 0.3461 | 0.9744 | 878 | ADD | -0.1541 | 0.04368 | 12.44 | 3.377 | | loci_63 | No replication available |
| 13.93 | 3.722 | | 0.344 | 0.9848 | 878 | ADD | -0.1534 | 0.0435 | 12.44 | 3.377 | | loci_63 | No replication available |
| 23.34 | 5.868 | | 0.2322 | 1.011 | 1324 | ADD | -0.1242 | 0.04091 | 9.212 | 2.619 | | loci_74 | No replication available |

| male_CHISQ | male_LOG10P | male_EXTRA | female_A1FREQ | female_INFO | female_N | female_TEST | female_BETA | female_SE | female_CHISQ | female_LOG10P | female_EXTRA | locus | nominal_replication |
|---|---|---|---|---|---|---|---|---|---|---|---|---|---|
| 22.62 | 5.704 | | 0.2341 | 1.015 | 1324 | ADD | -0.1258 | 0.04072 | 9.553 | 2.7 | | loci_74 | No replication available |
| 22.62 | 5.705 | | 0.2353 | 1.016 | 1324 | ADD | -0.1251 | 0.04065 | 9.476 | 2.682 | | loci_74 | No replication available |
| 22.61 | 5.702 | | 0.2342 | 1.015 | 1324 | ADD | -0.1254 | 0.0407 | 9.487 | 2.684 | | loci_74 | No replication available |
| 22.62 | 5.706 | | 0.2345 | 1.018 | 1324 | ADD | -0.1248 | 0.04063 | 9.431 | 2.671 | | loci_74 | No replication available |
| 22.7 | 5.722 | | 0.2331 | 1.012 | 1324 | ADD | -0.1246 | 0.04082 | 9.314 | 2.643 | | loci_74 | No replication available |
| 22.62 | 5.706 | | 0.2356 | 1.019 | 1324 | ADD | -0.1239 | 0.04056 | 9.333 | 2.648 | | loci_74 | No replication available |
| 22.38 | 5.65 | | 0.2352 | 1.016 | 1324 | ADD | -0.1245 | 0.04063 | 9.386 | 2.66 | | loci_74 | No replication available |
| 17.97 | 4.648 | | 0.1745 | 1.037 | 1324 | ADD | -0.1345 | 0.04496 | 8.947 | 2.556 | | loci_74 | No replication available |
| 17.97 | 4.65 | | 0.1748 | 1.039 | 1324 | ADD | -0.1334 | 0.04489 | 8.836 | 2.53 | | loci_74 | No replication available |
| 11.09 | 3.061 | | 0.4568 | 0.9503 | 743 | ADD | 0.1881 | 0.04222 | 19.86 | 5.08 | | loci_singleton_555 | Not replicated |
| 17.64 | 4.574 | | 0.3675 | 0.9633 | 743 | ADD | 0.1436 | 0.04338 | 10.96 | 3.031 | | loci_2 | Not replicated |
| 17.64 | 4.574 | | 0.3675 | 0.9635 | 743 | ADD | 0.1435 | 0.04338 | 10.95 | 3.029 | | loci_2 | Not replicated |
| 17.64 | 4.573 | | 0.3675 | 0.9635 | 743 | ADD | 0.1435 | 0.04338 | 10.95 | 3.029 | | loci_2 | Not replicated |
| 17.63 | 4.572 | | 0.3675 | 0.9635 | 743 | ADD | 0.1435 | 0.04338 | 10.94 | 3.027 | | loci_2 | Not replicated |
| 17.63 | 4.572 | | 0.3675 | 0.9635 | 743 | ADD | 0.1435 | 0.04338 | 10.94 | 3.027 | | loci_2 | Not replicated |
| 8.824 | 2.527 | | 0.1866 | 0.9783 | 743 | ADD | -0.2287 | 0.05284 | 18.73 | 4.823 | | loci_singleton_561 | Nominal replication |
| 17.61 | 4.566 | | 0.3677 | 0.9632 | 743 | ADD | 0.1426 | 0.04337 | 10.81 | 2.995 | | loci_2 | Not replicated |
| 17.63 | 4.572 | | 0.3677 | 0.9632 | 743 | ADD | 0.1424 | 0.04337 | 10.78 | 2.99 | | loci_2 | Not replicated |
| 17.63 | 4.572 | | 0.3677 | 0.9632 | 743 | ADD | 0.1424 | 0.04337 | 10.78 | 2.99 | | loci_2 | Not replicated |
| 17.57 | 4.558 | | 0.3677 | 0.9631 | 743 | ADD | 0.1427 | 0.04337 | 10.82 | 2.998 | | loci_2 | Not replicated |
| 17.02 | 4.432 | | 0.3671 | 0.9647 | 743 | ADD | 0.1438 | 0.04336 | 11 | 3.04 | | loci_2 | Not replicated |
| 17.02 | 4.431 | | 0.3671 | 0.965 | 743 | ADD | 0.1438 | 0.04336 | 11 | 3.041 | | loci_2 | Not replicated |
| 17.01 | 4.431 | | 0.3671 | 0.965 | 743 | ADD | 0.1438 | 0.04336 | 11 | 3.041 | | loci_2 | Not replicated |
| 16.87 | 4.397 | | 0.3674 | 0.9644 | 743 | ADD | 0.1438 | 0.04336 | 10.99 | 3.039 | | loci_2 | Not replicated |
| 17 | 4.427 | | 0.3665 | 0.9664 | 743 | ADD | 0.1411 | 0.04334 | 10.59 | 2.945 | | loci_2 | Not replicated |
| 9.015 | 2.572 | | 0.4574 | 0.9984 | 949 | ADD | -0.1969 | 0.04079 | 23.31 | 5.86 | | loci_singleton_571 | No replication available |
| 19.32 | 4.958 | | 0.1771 | 1.048 | 757 | ADD | 0.1514 | 0.05528 | 7.498 | 2.209 | | loci_singleton_572 | Not replicated |
| 13.59 | 3.643 | | 0.0902 | 0.9422 | 757 | ADD | -0.289 | 0.07777 | 13.81 | 3.694 | | loci_71 | Not replicated |
| 13.6 | 3.647 | | 0.09049 | 0.9486 | 757 | ADD | -0.2848 | 0.0774 | 13.54 | 3.632 | | loci_71 | Not replicated |
| 10.36 | 2.89 | | 0.1819 | 1.059 | 769 | ADD | -0.2247 | 0.05381 | 17.44 | 4.527 | | loci_56 | No replication available |
| 10.93 | 3.025 | | 0.1839 | 1.065 | 769 | ADD | -0.2144 | 0.05343 | 16.1 | 4.221 | | loci_56 | No replication available |
| 10.66 | 2.961 | | 0.1857 | 1.071 | 769 | ADD | -0.2148 | 0.05308 | 16.37 | 4.283 | | loci_56 | No replication available |
| 10.22 | 2.857 | | 0.183 | 1.066 | 769 | ADD | -0.2169 | 0.0535 | 16.44 | 4.299 | | loci_56 | No replication available |
| 23.73 | 5.955 | | 0.1598 | 0.9611 | 1059 | ADD | -0.1772 | 0.05757 | 9.472 | 2.681 | | loci_17 | Nominal replication |
| 23.05 | 5.802 | | 0.1606 | 0.9671 | 1059 | ADD | -0.1807 | 0.05729 | 9.948 | 2.793 | | loci_17 | Nominal replication |
| 23.7 | 5.949 | | 0.1624 | 0.972 | 1059 | ADD | -0.1731 | 0.05688 | 9.258 | 2.63 | | loci_17 | Nominal replication |
| 21.92 | 5.546 | | 0.1596 | 0.962 | 1059 | ADD | -0.1772 | 0.05757 | 9.471 | 2.68 | | loci_17 | Nominal replication |
| 20.87 | 5.309 | | 0.1667 | 0.9737 | 1059 | ADD | -0.1781 | 0.05627 | 10.01 | 2.809 | | loci_17 | Nominal replication |
| 18.2 | 4.702 | | 0.1603 | 0.9735 | 1059 | ADD | -0.1957 | 0.05718 | 11.72 | 3.209 | | loci_17 | Nominal replication |
| 18.03 | 4.662 | | 0.1626 | 0.9751 | 1059 | ADD | -0.1954 | 0.05681 | 11.83 | 3.234 | | loci_17 | Nominal replication |
| 18.08 | 4.675 | | 0.1612 | 0.9705 | 1059 | ADD | -0.1954 | 0.05713 | 11.7 | 3.203 | | loci_17 | Nominal replication |
| 18.33 | 4.73 | | 0.1631 | 0.9692 | 1059 | ADD | -0.1909 | 0.05692 | 11.25 | 3.099 | | loci_17 | Nominal replication |
| 19.32 | 4.955 | | 0.1653 | 0.9769 | 1059 | ADD | -0.1809 | 0.05636 | 10.3 | 2.875 | | loci_17 | Nominal replication |
| 18.3 | 4.724 | | 0.1634 | 0.9673 | 1059 | ADD | -0.1899 | 0.05692 | 11.13 | 3.071 | | loci_17 | Nominal replication |
| 17.3 | 4.495 | | 0.1603 | 0.9752 | 1059 | ADD | -0.1946 | 0.05712 | 11.61 | 3.182 | | loci_17 | Nominal replication |
| 17.2 | 4.474 | | 0.1599 | 0.9706 | 1059 | ADD | -0.1954 | 0.05732 | 11.62 | 3.186 | | loci_17 | Nominal replication |
| 16.89 | 4.402 | | 0.1601 | 0.9681 | 1059 | ADD | -0.1886 | 0.05735 | 10.81 | 2.997 | | loci_17 | Nominal replication |

| male_CHISQ | male_LOG10P | male_EXTRA | female_A1FREQ | female_INFO | female_N | female_TEST | female_BETA | female_SE | female_CHISQ | female_LOG10P | female_EXTRA | locus | nominal_replication |
|---|---|---|---|---|---|---|---|---|---|---|---|---|---|
| 16.56 | 4.327 | | 0.05723 | 0.9701 | 1059 | ADD | -0.2861 | 0.08995 | 10.12 | 2.834 | | loci_singleton_593 | Not replicated |
| 6.609 | 1.994 | | 0.2549 | 0.8091 | 643 | ADD | -0.2635 | 0.05881 | 20.07 | 5.128 | | loci_singleton_594 | No replication available |
| 12.1 | 3.297 | | 0.3197 | 0.9504 | 902 | ADD | -0.2099 | 0.04876 | 18.53 | 4.776 | | loci_59 | Not replicated |
| 7.915 | 2.31 | | 0.357 | 0.9616 | 902 | ADD | -0.2357 | 0.04705 | 25.1 | 6.264 | | loci_59 | Not replicated |
| 10.41 | 2.903 | | 0.3165 | 0.9544 | 902 | ADD | -0.2142 | 0.0488 | 19.28 | 4.946 | | loci_59 | Not replicated |
| 10.42 | 2.904 | | 0.3172 | 0.9539 | 902 | ADD | -0.2119 | 0.04877 | 18.87 | 4.854 | | loci_59 | Not replicated |
| 10.45 | 2.91 | | 0.3159 | 0.9506 | 902 | ADD | -0.2111 | 0.04893 | 18.62 | 4.797 | | loci_59 | Not replicated |
| 10.17 | 2.845 | | 0.317 | 0.9518 | 902 | ADD | -0.2118 | 0.04883 | 18.81 | 4.841 | | loci_59 | Not replicated |
| male_CHISQ | male_LOG10P | male_EXTRA | female_A1FREQ | female_INFO | female_N | female_TEST | female_BETA | female_SE | female_CHISQ | female_LOG10P | female_EXTRA | locus | nominal_replication |
| 6.652 | 2.004 | | 0.352 | 0.9543 | 902 | ADD | -0.236 | 0.0474 | 24.79 | 6.194 | | loci_59 | Not replicated |
| 6.405 | 1.944 | | 0.352 | 0.953 | 902 | ADD | -0.2357 | 0.04743 | 24.69 | 6.172 | | loci_59 | Not replicated |
| 6.21 | 1.896 | | 0.3515 | 0.9557 | 902 | ADD | -0.2373 | 0.04738 | 25.08 | 6.259 | | loci_59 | Not replicated |
| 6.154 | 1.882 | | 0.3514 | 0.9551 | 902 | ADD | -0.2372 | 0.0474 | 25.03 | 6.249 | | loci_59 | Not replicated |
| 6.522 | 1.972 | | 0.3512 | 0.9463 | 902 | ADD | -0.2324 | 0.04762 | 23.81 | 5.974 | | loci_59 | Not replicated |
| 6.18 | 1.889 | | 0.3512 | 0.9503 | 902 | ADD | -0.2327 | 0.04753 | 23.97 | 6.009 | | loci_59 | Not replicated |
| 27.98 | 6.911 | | 0.2496 | 0.9575 | 879 | ADD | -0.07581 | 0.04384 | 2.991 | 1.077 | | loci_singleton_607 | No replication available |
| 18.44 | 4.757 | | 0.2437 | 0.9374 | 973 | ADD | -0.1593 | 0.0468 | 11.59 | 3.179 | | loci_30 | No replication available |
| 19.31 | 4.954 | | 0.2574 | 0.9325 | 973 | ADD | -0.1452 | 0.04606 | 9.934 | 2.79 | | loci_30 | No replication available |
| 9.869 | 2.775 | | 0.2656 | 1.011 | 973 | ADD | 0.1955 | 0.04411 | 19.65 | 5.031 | | loci_35 | No replication available |
| 18.1 | 4.679 | | 0.2538 | 0.9473 | 973 | ADD | -0.1447 | 0.04591 | 9.932 | 2.789 | | loci_30 | No replication available |
| 18.09 | 4.675 | | 0.2529 | 0.9417 | 973 | ADD | -0.1453 | 0.04611 | 9.929 | 2.789 | | loci_30 | No replication available |
| 10.2 | 2.852 | | 0.2622 | 1.02 | 973 | ADD | 0.1901 | 0.04406 | 18.63 | 4.798 | | loci_35 | No replication available |
| 10.26 | 2.867 | | 0.2642 | 1.021 | 973 | ADD | 0.1802 | 0.04387 | 16.87 | 4.398 | | loci_35 | No replication available |
| 24.04 | 6.025 | | 0.09405 | 0.9875 | 971 | ADD | -0.2147 | 0.06285 | 11.66 | 3.195 | | loci_3 | No replication available |
| 23.47 | 5.898 | | 0.09456 | 0.9909 | 971 | ADD | -0.2165 | 0.0626 | 11.96 | 3.264 | | loci_3 | No replication available |
| 21.76 | 5.51 | | 0.09549 | 0.9828 | 971 | ADD | -0.1907 | 0.0627 | 9.251 | 2.628 | | loci_3 | No replication available |
| 21.4 | 5.428 | | 0.09505 | 1.001 | 971 | ADD | -0.1867 | 0.06227 | 8.99 | 2.566 | | loci_3 | No replication available |
| 12.19 | 3.318 | | 0.2682 | 0.9701 | 971 | ADD | 0.17 | 0.04233 | 16.14 | 4.23 | | loci_72 | No replication available |
| 12.18 | 3.316 | | 0.2682 | 0.9705 | 971 | ADD | 0.1695 | 0.04232 | 16.04 | 4.208 | | loci_72 | No replication available |
| 10.95 | 3.029 | | 0.3192 | 0.9552 | 971 | ADD | 0.1656 | 0.04054 | 16.69 | 4.357 | | loci_72 | No replication available |
| 19.75 | 5.055 | | 0.07559 | 0.9298 | 936 | ADD | 0.2583 | 0.08225 | 9.863 | 2.773 | | loci_29 | No replication available |
| 18.28 | 4.719 | | 0.07488 | 0.9284 | 936 | ADD | 0.266 | 0.08269 | 10.35 | 2.888 | | loci_29 | No replication available |
| 18.51 | 4.771 | | 0.07392 | 0.9346 | 936 | ADD | 0.2606 | 0.08291 | 9.88 | 2.777 | | loci_29 | No replication available |
| 9.642 | 2.721 | | 0.4307 | 0.9779 | 936 | ADD | 0.1804 | 0.04307 | 17.54 | 4.551 | | loci_65 | No replication available |
| 9.64 | 2.72 | | 0.4307 | 0.9778 | 936 | ADD | 0.1804 | 0.04308 | 17.53 | 4.549 | | loci_65 | No replication available |
| 9.574 | 2.705 | | 0.4299 | 0.978 | 936 | ADD | 0.1793 | 0.04309 | 17.32 | 4.501 | | loci_65 | No replication available |
| 9.562 | 2.702 | | 0.4316 | 0.9852 | 936 | ADD | 0.1786 | 0.04291 | 17.33 | 4.504 | | loci_65 | No replication available |
| 9.574 | 2.705 | | 0.4302 | 0.98 | 936 | ADD | 0.1788 | 0.04304 | 17.25 | 4.485 | | loci_65 | No replication available |
| 9.574 | 2.705 | | 0.4302 | 0.98 | 936 | ADD | 0.1788 | 0.04304 | 17.25 | 4.485 | | loci_65 | No replication available |
| 9.569 | 2.703 | | 0.4302 | 0.98 | 936 | ADD | 0.1788 | 0.04304 | 17.26 | 4.486 | | loci_65 | No replication available |
| 9.562 | 2.702 | | 0.4303 | 0.9799 | 936 | ADD | 0.1788 | 0.04304 | 17.26 | 4.487 | | loci_65 | No replication available |
| 14.78 | 3.917 | | 0.04743 | 1.013 | 829 | ADD | 0.3632 | 0.1018 | 12.74 | 3.446 | | loci_singleton_633 | Not replicated |
| 21.37 | 5.422 | | 0.06318 | 1.045 | 585 | ADD | -0.3537 | 0.1085 | 10.63 | 2.954 | | loci_31 | No replication available |
| 21.38 | 5.424 | | 0.06315 | 1.044 | 585 | ADD | -0.3537 | 0.1085 | 10.62 | 2.951 | | loci_31 | No replication available |
| 8.201 | 2.378 | | 0.04292 | 0.9561 | 587 | ADD | -0.6177 | 0.1434 | 18.56 | 4.784 | | loci_singleton_636 | No replication available |
| 22.44 | 5.663 | | 0.1493 | 0.9661 | 545 | ADD | 0.2248 | 0.08171 | 7.568 | 2.226 | | loci_73 | Not replicated |

| male_CHISQ | male_LOG10P | male_EXTRA | female_A1FREQ | female_INFO | female_N | female_TEST | female_BETA | female_SE | female_CHISQ | female_LOG10P | female_EXTRA | locus | nominal_replication |
|---|---|---|---|---|---|---|---|---|---|---|---|---|---|
| 21.71 | 5.499 | | 0.1438 | 0.9788 | 545 | ADD | 0.2312 | 0.08242 | 7.867 | 2.298 | | loci_73 | Not replicated |
| 21.7 | 5.496 | | 0.1441 | 0.9803 | 545 | ADD | 0.2304 | 0.0823 | 7.84 | 2.292 | | loci_73 | Not replicated |
| 20.05 | 5.122 | | 0.1453 | 0.9674 | 545 | ADD | 0.2373 | 0.08252 | 8.272 | 2.395 | | loci_73 | Not replicated |
| 20.05 | 5.122 | | 0.1455 | 0.9664 | 545 | ADD | 0.2369 | 0.08257 | 8.231 | 2.385 | | loci_73 | Not replicated |
| 19.98 | 5.106 | | 0.1456 | 0.9657 | 545 | ADD | 0.2365 | 0.08261 | 8.195 | 2.377 | | loci_73 | Not replicated |
| 19.98 | 5.106 | | 0.1456 | 0.9657 | 545 | ADD | 0.2365 | 0.08261 | 8.195 | 2.377 | | loci_73 | Not replicated |
| 16.79 | 4.379 | | 0.1723 | 0.9712 | 545 | ADD | 0.2394 | 0.07692 | 9.688 | 2.732 | | loci_73 | Not replicated |
| 19.86 | 5.079 | | 0.1414 | 0.9769 | 545 | ADD | 0.2241 | 0.08322 | 7.251 | 2.149 | | loci_73 | Not replicated |
| 12.62 | 3.419 | | 0.0607 | 1.084 | 569 | ADD | 0.394 | 0.1062 | 13.77 | 3.684 | | loci_singleton_646 | No replication available |
| 19.88 | 5.084 | | 0.07946 | 0.984 | 684 | ADD | -0.2596 | 0.08381 | 9.592 | 2.709 | | loci_89 | No replication available |
| 19.79 | 5.063 | | 0.07582 | 0.9971 | 684 | ADD | -0.2558 | 0.08505 | 9.049 | 2.58 | | loci_89 | No replication available |
| 20.35 | 5.191 | | 0.08535 | 0.9954 | 684 | ADD | -0.2321 | 0.08065 | 8.281 | 2.397 | | loci_89 | No replication available |
| 18.97 | 4.876 | | 0.07588 | 0.9972 | 684 | ADD | -0.2565 | 0.08502 | 9.1 | 2.592 | | loci_89 | No replication available |
| 21.06 | 5.351 | | 0.1006 | 0.9653 | 684 | ADD | -0.2019 | 0.07606 | 7.049 | 2.101 | | loci_89 | No replication available |
| 21.06 | 5.351 | | 0.1006 | 0.9653 | 684 | ADD | -0.2019 | 0.07606 | 7.049 | 2.101 | | loci_89 | No replication available |
| 19.72 | 5.047 | | 0.08449 | 0.9972 | 684 | ADD | -0.2289 | 0.08096 | 7.994 | 2.328 | | loci_89 | No replication available |
| 19.72 | 5.048 | | 0.08449 | 0.9972 | 684 | ADD | -0.2289 | 0.08096 | 7.992 | 2.328 | | loci_89 | No replication available |
| 19.71 | 5.044 | | 0.08348 | 0.9882 | 684 | ADD | -0.2296 | 0.0818 | 7.877 | 2.301 | | loci_89 | No replication available |
| 18.76 | 4.83 | | 0.08261 | 0.9871 | 684 | ADD | -0.2385 | 0.08224 | 8.409 | 2.428 | | loci_89 | No replication available |
| 20.28 | 5.176 | | 0.1012 | 0.9647 | 684 | ADD | -0.2004 | 0.0759 | 6.971 | 2.082 | | loci_89 | No replication available |
| 18.83 | 4.844 | | 0.08462 | 0.9985 | 684 | ADD | -0.227 | 0.08085 | 7.885 | 2.302 | | loci_89 | No replication available |
| 13.36 | 3.591 | | 0.3147 | 0.9204 | 555 | ADD | 0.2004 | 0.05209 | 14.8 | 3.924 | | loci_39 | Not replicated |
| 12.41 | 3.37 | | 0.2924 | 0.9869 | 555 | ADD | 0.1995 | 0.05137 | 15.09 | 3.988 | | loci_39 | Not replicated |

### STable11: Heritability and genetic correlation estimates of per-person median values of clinical quantitative traits in BioVU

508 rows × 60 columns

STable11 — Columns 1–23 of 60

| all_labs | full_name | group | concept_name | LOINC_NAME | heritability_names | selected_unit | sex_specific_lab | genetic.correlation | genetic.correlation.p | gcta.N_female | gcta.N_male | gcta.Variance_Autosomal_h2_female | gcta.Variance_Autosomal_h2_male | gcta.Variance_X_chromosome_h2_female | gcta.Variance_X_chromosome_h2_male | gcta.Variance_h2_female | gcta.Variance_h2_male | gcta.SE_Autosomal_h2_female | gcta.SE_Autosomal_h2_male | gcta.SE_X_chromosome_h2_female | gcta.SE_X_chromosome_h2_male | gcta.SE_h2_female |
|---|---|---|---|---|---|---|---|---|---|---|---|---|---|---|---|---|---|---|---|---|---|---|
| #WBC | # of WBC's | blood | #WBC | | X.WBC | | | 0.08768 | 0.8305 | 925 | 761 | 1.00e-06 | 0.05629 | 0.05765 | 1.00e-06 | 0.05765 | 0.05629 | 0.2948 | 0.3325 | 0.04807 | 0.06032 | 0.3 |
| %SAT | % saturation | other | PERCENT SATURATION | | X_percent_SAT | % | | 0.135 | 0.1349 | 8469 | 6545 | 0.08613 | 0.08734 | 1.00e-06 | 0.001158 | 0.08613 | 0.0885 | 0.03008 | 0.03882 | 0.005535 | 0.005814 | 0.03049 |
| 125DHy | Calcitriol [Mass/volume] in Serum or Plasma | metabolic | VIT D 1,25-DIHYDROXY | Calcitriol [Mass/volume] in Serum or Plasma | X125DHy | pg/mL | | -0.04992 | 0.7165 | 2073 | 897 | 0.005698 | 0.04473 | 1.00e-06 | 0.03086 | 0.005699 | 0.07558 | 0.1181 | 0.256 | 0.01669 | 0.0513 | 0.1194 |
| 25DTot | 25-Hydroxyvitamin D3+25-Hydroxyvitamin D2 [Mass/volume] in Serum or Plasma | metabolic | 25-HYDROXY D TOTAL | 25-Hydroxyvitamin D3+25-Hydroxyvitamin D2 [Mass/volume] in Serum or Plasma | X25DTot | ng/mL | | 0.2122 | 0.03509 | 7048 | 2828 | 0.1424 | 0.1184 | 0.002566 | 0.001877 | 0.145 | 0.1203 | 0.03823 | 0.09154 | 0.005922 | 0.01618 | 0.03862 |
| 25HyD2 | 25-Hydroxyvitamin D2 [Mass/volume] in Serum or Plasma | metabolic | 25-HYDROXY D2 | 25-hydroxyvitamin D2 [Mass/volume] in Serum or Plasma | X25HyD2 | ng/mL | | 0.06622 | 0.6537 | 2276 | 811 | 0.08146 | | 1.00e-06 | | 0.08146 | | 0.1054 | | 0.0192 | | 0.107 |
| 25HyD3 | 25-Hydroxyvitamin D3 [Mass/volume] in Serum or Plasma | metabolic | 25-HYDROXY D3 | 25-hydroxyvitamin D3 [Mass/volume] in Serum or Plasma | X25HyD3 | ng/mL | | 0.1074 | 0.2061 | 7065 | 2839 | 0.07056 | 0.1401 | 1.00e-06 | 1.00e-06 | 0.07056 | 0.1401 | 0.03611 | 0.09069 | 0.005371 | 0.0151 | 0.03644 |
| A-1Gg_ | Alpha 1 globulin [Mass/volume] in Serum or Plasma by Electrophoresis | liver | ALPHA-1 GLOBULIN | Alpha 1 globulin [Mass/volume] in Serum or Plasma by Electrophoresis | A.1Gg_ | g/dL | | 0.07194 | 0.4339 | 3674 | 2884 | | 0.06414 | | 1.00e-06 | | 0.06414 | | 0.08467 | | 0.01493 | |
| A-2Gg_ | Alpha 2 globulin [Mass/volume] in Serum or Plasma by Electrophoresis | liver | ALPHA-2 GLOBULIN | Alpha 2 globulin [Mass/volume] in Serum or Plasma by Electrophoresis | A.2Gg_ | g/dL | | -0.113 | 0.3814 | 3671 | 2892 | 0.02302 | 0.1012 | 0.002673 | 0.01692 | 0.02569 | 0.1181 | 0.06476 | 0.08498 | 0.01105 | 0.01705 | 0.06563 |
| A1Anti | Alpha 1 antitrypsin [Mass/volume] in Serum or Plasma | other | ALPHA -1-ANTITRIPSIN | Alpha 1 antitrypsin [Mass/volume] in Serum or Plasma | A1Anti | mg/dL | | 0.3806 | 0.01305 | 925 | 962 | 0.04841 | 0.2026 | 0.01325 | 0.03654 | 0.06166 | 0.2392 | 0.2684 | 0.2424 | 0.04312 | 0.0481 | 0.2757 |
| ABS_BASO_CT | Basophils [#/volume] in Blood by Automated count | blood | ABS_BASO_CT | Basophils [#/volume] in Blood by Automated count | ABS_BASO_CT | 10*3/uL | | 0.02041 | 0.8924 | 1059 | 638 | 0.427 | | 0.002798 | | 0.4298 | | 0.2224 | | 0.03549 | | 0.2235 |
| ABS_EOS_CT | Eosinophils [#/volume] in Blood by Automated count | blood | ABS_EOS_CT | | ABS_EOS_CT | 10*3/uL | | 0.3115 | 0.08344 | 996 | 598 | 0.235 | | 0.0409 | | 0.2759 | | 0.2558 | | 0.04654 | | 0.2564 |

| all_labs | full_name | group | concept_name | LOINC_NAME | heritability_names | selected_unit | sex_specific_lab | genetic.correlation | genetic.correlation.p | gcta.N_female | gcta.N_male | gcta.Variance_Autosomal_h2_female | gcta.Variance_Autosomal_h2_male | gcta.Variance_X_chromosome_h2_female | gcta.Variance_X_chromosome_h2_male | gcta.Variance_h2_female | gcta.Variance_h2_male | gcta.SE_Autosomal_h2_female | gcta.SE_Autosomal_h2_male | gcta.SE_X_chromosome_h2_female | gcta.SE_X_chromosome_h2_male | gcta.SE_h2_female |
|---|---|---|---|---|---|---|---|---|---|---|---|---|---|---|---|---|---|---|---|---|---|---|
| ABS_IMMATURE_GRANULOCYTES_CT | Immature granulocytes [#/volume] in Blood by Automated count | blood | ABS_IMMATURE_GRANULOCYTES_CT | | ABS_IMMATURE_GRANULOCYTES_CT | 10*3/uL | | 0.1516 | 0.4753 | 1042 | 626 | | 0.3944 | | 1.00e-06 | | 0.3944 | | 0.4033 | | 0.07178 | |
| ABS_LYMPH_CT | Lymphocytes [#/volume] in Blood by Automated count | blood | ABS_LYMPH_CT | | ABS_LYMPH_CT | 10*3/uL | | -0.2954 | 0.08797 | 1060 | 640 | 1.00e-06 | 0.5327 | 0.0445 | 1.00e-06 | 0.0445 | 0.5327 | 0.2285 | 0.3923 | 0.04438 | 0.07966 | 0.2326 |
| ABS_MONO_CT | Monocytes [#/volume] in Blood by Automated count | blood | ABS_MONO_CT | Monocytes [#/volume] in Blood by Automated count | ABS_MONO_CT | 10*3/uL | | 0.02668 | 0.9179 | 1059 | 639 | 0.1182 | 0.05283 | 1.00e-06 | 1.00e-06 | 0.1182 | 0.05283 | 0.2374 | 0.4046 | 0.03747 | 0.07719 | 0.2398 |
| ABS_NEUTRO_CT | Neutrophils [#/volume] in Blood by Automated count | blood | ABS_NEUTRO_CT | | ABS_NEUTRO_CT | 10*3/uL | | 0.03831 | 0.8284 | 1060 | 638 | | 0.3955 | | 1.00e-06 | | 0.3955 | | 0.3846 | | 0.08106 | |
| ACAIgA | Cardiolipin IgA Ab [Units/volume] in Serum by Immunoassay | immune / inflammatory | APA-IGA | Cardiolipin IgA Ab [Units/volume] in Serum by Immunoassay | ACAIgA | | | -0.03264 | 0.851 | 1508 | 548 | 0.03506 | | 0.01978 | | 0.05484 | | 0.1655 | | 0.02957 | | 0.168 |
| ACAIgG | Cardiolipin IgG Ab [Units/volume] in Serum by Immunoassay | immune / inflammatory | APA-IGG | Cardiolipin IgG Ab [Units/volume] in Serum by Immunoassay | ACAIgG | | | 0.03085 | 0.8536 | 1806 | 675 | 1.00e-06 | 0.06546 | 0.02935 | 1.00e-06 | 0.02935 | 0.06546 | 0.132 | 0.3804 | 0.02621 | 0.06722 | 0.1344 |
| ACAIgM | Cardiolipin IgM Ab [Units/volume] in Serum by Immunoassay | immune / inflammatory | APA-IGM | Cardiolipin IgM Ab [Units/volume] in Serum by Immunoassay | ACAIgM | [MPL'U] | | 0.2181 | 0.2013 | 1805 | 675 | 0.2208 | 0.5718 | 1.00e-06 | 0.08289 | 0.2208 | 0.6546 | 0.1343 | 0.3853 | 0.02267 | 0.0759 | 0.136 |
| ACTH | Corticotropin [Mass/volume] in Plasma | endocrine | ADRENOCORTICOTROPIC HORM | Corticotropin [Mass/volume] in Plasma | ACTH | pg/mL | | 0.1858 | 0.1987 | 987 | 598 | | | | | | | | | | | |
| AFP-Ob | Maternal Alpha-1-Feto protein [Mass/volume] in Serum or Plasma | ob / gyn | MATERNAL ALPHAFETO PR0 | Alpha-1-Feto protein [Mass/volume] in Serum or Plasma | AFP.Ob | ng/mL | Sex specific lab | | | 1035 | | 0.2284 | | 0.04385 | | 0.2723 | | 0.2385 | | 0.04603 | | 0.2418 |
| AFPTum | Alpha-1-fetoprotein.tumor marker [Mass/volume] in Serum or Plasma | cancer | AFP TUMOR | Alpha-1-fetoprotein.tumor marker [Mass/volume] in Serum or Plasma | AFPTum | ng/mL | | -0.2388 | 0.02153 | 1312 | 1721 | 0.2278 | | 1.00e-06 | | 0.2278 | | 0.197 | | 0.03024 | | 0.1991 |
| Alb | Albumin [Mass/volume] in Serum or Plasma | liver | ALBUMIN BLOOD | Albumin [Mass/volume] in Serum or Plasma | Alb | g/dL | | 0.3884 | 3.79e-09 | 31325 | 24351 | 0.08774 | 0.07371 | 0.001468 | 4.78e-04 | 0.0892 | 0.07418 | 0.009374 | 0.01133 | 0.001553 | 0.001908 | 0.009486 |

| all_labs | full_name | group | concept_name | LOINC_NAME | heritability_names | selected_unit | sex_specific_lab | genetic.correlation | genetic.correlation.p | gcta.N_female | gcta.N_male | gcta.Variance_Autosomal_h2_female | gcta.Variance_Autosomal_h2_male | gcta.Variance_X_chromosome_h2_female | gcta.Variance_X_chromosome_h2_male | gcta.Variance_h2_female | gcta.Variance_h2_male | gcta.SE_Autosomal_h2_female | gcta.SE_Autosomal_h2_male | gcta.SE_X_chromosome_h2_female | gcta.SE_X_chromosome_h2_male | gcta.SE_h2_female |
|---|---|---|---|---|---|---|---|---|---|---|---|---|---|---|---|---|---|---|---|---|---|---|
| AlbCnc | Albumin [Presence] in Urine | urinary | ALBUMIN URINE | Albumin [Presence] in Urine | AlbCnc | ug/mL | | 0.1771 | 0.2383 | 3319 | 3332 | 0.00231 | 1.00e-06 | 0.004543 | 0.01239 | 0.006853 | 0.01239 | 0.07127 | 0.07711 | 0.01252 | 0.01474 | 0.07201 |
| AlbCre | Microalbumin/Creatinine [Mass Ratio] in 24 hour Urine | liver | ALBUMIN/CREAT RATIO | Microalbumin/Creatinine [Mass Ratio] in 24 hour Urine | AlbCre | ug/mg | | -0.07062 | 0.5024 | 2156 | 2218 | 1.00e-06 | 0.07468 | 0.008779 | 0.004741 | 0.00878 | 0.07942 | 0.1175 | 0.1141 | 0.02068 | 0.02041 | 0.1184 |
| Album | Albumin [Mass/volume] in Serum or Plasma | liver | ALBUMIN | Albumin [Mass/volume] in Serum or Plasma by Electrophoresis | Album | g/dL | | 0.1224 | 0.09429 | 4050 | 3191 | 0.07264 | 0.1089 | 0.005221 | 1.00e-06 | 0.07786 | 0.1089 | 0.05973 | 0.0752 | 0.01031 | 0.01494 | 0.06089 |
| Aldo-P | Aldosterone [Mass/volume] in Serum or Plasma | endocrine | ALDOSTERONE | Aldosterone [Mass/volume] in Serum or Plasma | Aldo.P | ng/dL | | -0.005046 | 0.9822 | 864 | 573 | | 0.001726 | | 1.00e-06 | | 0.001727 | | 0.4196 | | 0.07943 | |
| AlkP | Alkaline phosphatase [Enzymatic activity/volume] in Serum or Plasma | liver | ALKALINE PHOSPHATASE BLD | Alkaline phosphatase [Enzymatic activity/volume] in Serum or Plasma | AlkP | [U]/L | | 0.4754 | 1.51e-13 | 21116 | 15939 | 0.1598 | 0.1346 | 0.001793 | 1.00e-06 | 0.1615 | 0.1346 | 0.0142 | 0.01791 | 0.002252 | 0.002643 | 0.01436 |
| Amm | Ammonia [Moles/volume] in Plasma | metabolic | AMMONIA | Ammonia [Moles/volume] in Plasma | Amm | umol/L | | 0.2736 | 0.4605 | 1426 | 1791 | | 0.3167 | | 1.00e-06 | | 0.3167 | | 0.1428 | | 0.02604 | |
| Amyl | Amylase [Enzymatic activity/volume] in Serum or Plasma | metabolic | AMYLASE BLOOD | Amylase [Enzymatic activity/volume] in Serum or Plasma | Amyl | [iU]/L | | 0.02559 | 0.6756 | 4794 | 3776 | 0.07285 | 0.07553 | 0.004805 | 0.01095 | 0.07765 | 0.08648 | 0.05231 | 0.06404 | 0.00894 | 0.01277 | 0.0532 |
| AN-GAP | Anion gap in Serum or Plasma | metabolic | ANION GAP | Anion gap in Serum or Plasma | AN.GAP | | | 0.2273 | 0.00852 | 34129 | 26892 | 0.0242 | 0.01779 | 0.001673 | 0.001717 | 0.02587 | 0.0195 | 0.007714 | 0.009612 | 0.00152 | 0.001801 | 0.007837 |
| ANESTHESIA PULSE | Heart rate by pulse oximetry | vitals | Heart rate by Pulse oximetry | Heart rate by Pulse oximetry | ANESTHESIA.PULSE | | | 0.014 | 0.8952 | 3746 | 2556 | 0.1344 | 0.03624 | 1.00e-06 | 1.00e-06 | 0.1344 | 0.03624 | 0.06578 | 0.1019 | 0.01312 | 0.01808 | 0.06708 |
| ANION_GAP | Anion gap | metabolic | ANION_GAP | | ANION_GAP | 10*-3.eq/L | | 0.01368 | 0.939 | 619 | 471 | 0.8738 | | 1.00e-06 | | 0.8738 | | 0.3295 | | 0.06624 | | 0.331 |
| AntDNA | DNA double strand Ab [Units/volume] in Serum | immune / inflammatory | ANTI-DNA(SLE)Current | DNA double strand Ab [Units/volume] in Serum | AntDNA | [iU]/mL | | -0.1288 | 0.4023 | 2079 | 642 | 0.1237 | | 0.0251 | | 0.1488 | | 0.1214 | | 0.02324 | | 0.124 |
| APBone | Alkaline phosphatase bone | other | ALKALINE PHOS BONE | Alkaline phosphatase.bone [Mass/volume] in Serum or Plasma | APBone | ug/L | | -0.2937 | 0.4024 | 1045 | 285 | 0.144 | 0.84 | 0.0197 | 0.16 | 0.1637 | 1 | 0.2282 | 0.9779 | 0.03734 | 0.1954 | 0.2304 |
| AT3 | Antithrombin actual/normal in Platelet poor plasma by Chromogenic method | blood | ANTITHROMBIN III ACT | Antithrombin actual/normal in Platelet poor plasma by Chromogenic method | AT3 | % | | -0.1107 | 0.7249 | 625 | 409 | | 0.6675 | | 0.3302 | | 0.9977 | | 0.6239 | | 0.1385 | |

| all_labs | full_name | group | concept_name | LOINC_NAME | heritability_names | selected_unit | sex_specific_lab | genetic.correlation | genetic.correlation.p | gcta.N_female | gcta.N_male | gcta.Variance_Autosomal_h2_female | gcta.Variance_Autosomal_h2_male | gcta.Variance_X_chromosome_h2_female | gcta.Variance_X_chromosome_h2_male | gcta.Variance_h2_female | gcta.Variance_h2_male | gcta.SE_Autosomal_h2_female | gcta.SE_Autosomal_h2_male | gcta.SE_X_chromosome_h2_female | gcta.SE_X_chromosome_h2_male | gcta.SE_h2_female |
|---|---|---|---|---|---|---|---|---|---|---|---|---|---|---|---|---|---|---|---|---|---|---|
| AThyP | Thyroperoxidase Ab | immune / inflammatory | ANTI THYROID PEROXAB | Thyroperoxidase Ab [Units/volume] in Serum or Plasma | AThyP | [U]/mL | | 0.2253 | 0.3643 | 1015 | 231 | 1.00e-06 | 1.00e-06 | 0.004067 | 0.09795 | 0.004068 | 0.09795 | 0.233 | 1.418 | 0.04137 | 0.2741 | 0.2365 |
| ATRIAL RATE | P wave Atrium by EKG | cardiovascular | P wave Atrium by EKG | P wave Atrium by EKG | ATRIAL.RATE | | | 0.03044 | 0.778 | 7227 | 6314 | 0.03217 | 0.04936 | 0.001554 | 1.00e-06 | 0.03372 | 0.04936 | 0.03442 | 0.0406 | 0.006355 | 0.007055 | 0.03493 |
| AtyLym | Variant lymphocytes/100 leukocytes in Blood by Manual count | immune / inflammatory | ATYP LYMPHS % | Variant lymphocytes/100 leukocytes in Blood by Manual count | AtyLym | % | | 0.2202 | 0.0678 | 3888 | 4046 | 0.0803 | | 1.00e-06 | | 0.0803 | | 0.06399 | | 0.01179 | | 0.0651 |
| AutoNt | Neutrophils by automated count | blood | AutoNt | | AutoNt | % | | -0.1164 | 0.2521 | 1354 | 933 | 0.4504 | 0.0995 | 1.00e-06 | 1.00e-06 | 0.4504 | 0.0995 | 0.182 | 0.2748 | 0.03249 | 0.04534 | 0.1851 |
| BAbs | Basophils [#/volume] in Blood by Automated count | immune / inflammatory | BASO (ABS) | Basophils [#/volume] in Blood by Automated count | BAbs | 10*3/uL | | 0.317 | 1.14e-05 | 14842 | 10793 | 0.1159 | 0.1088 | 1.00e-06 | 0.001705 | 0.1159 | 0.1105 | 0.01867 | 0.02495 | 0.002987 | 0.004367 | 0.01893 |
| Baso | Basophils [#/volume] in Blood | immune / inflammatory | BASOPHILS % | Basophils [#/volume] in Blood | Baso | % | | 0.4112 | 5.63e-06 | 15240 | 11083 | 0.1117 | 0.0501 | 1.00e-06 | 5.46e-04 | 0.1117 | 0.05065 | 0.01813 | 0.02318 | 0.003186 | 0.004107 | 0.0184 |
| BASOA | Basophils [#/volume] in Blood | blood | BASO (ABS) | Basophils [#/volume] in Blood | BASOA | 10*3/uL | | 0.081 | 0.5012 | 3882 | 4078 | 1.00e-06 | | 0.008195 | | 0.008196 | | 0.064 | | 0.01101 | | 0.06498 |
| BASOAB | Basophils [#/volume] in Blood by Automated count | blood | BASO (ABS) | Basophils [#/volume] in Blood by Automated count | BASOAB | 10*3/uL | | 0.4055 | 6.24e-07 | 25623 | 19957 | 0.00354 | 0.002488 | 0.001699 | 0.002031 | 0.005239 | 0.004519 | 0.001698 | 0.002032 | 0.001907 | 0.002362 | 0.002545 |
| BASOM | Basophils/100 leukocytes in Blood by Manual count | immune / inflammatory | BASOPHILS % | Basophils/100 leukocytes in Blood by Manual count | BASOM | % | | -0.0212 | 0.8173 | 3717 | 3957 | | 0.009219 | | 1.00e-06 | | 0.00922 | | 0.06148 | | 0.01055 | |
| BASOPHILS | BASOPHILS | immune / inflammatory | BASOPHILS | Basophils [#/volume] in Blood | BASOPHILS | % | | 0.1805 | 0.3879 | 885 | 592 | 0.3363 | 0.03646 | 1.00e-06 | 0.01838 | 0.3363 | 0.05484 | 0.2704 | 0.3755 | 0.04718 | 0.07823 | 0.2757 |
| BASORE | Basophils/100 leukocytes in Blood by Automated count | immune / inflammatory | BASOPHILS % | Basophils/100 leukocytes in Blood by Automated count | BASORE | % | | 0.2758 | 4.11e-04 | 25659 | 19975 | 0.003693 | 0.002168 | 0.002278 | 0.001032 | 0.005971 | 0.0032 | 0.001693 | 0.001984 | 0.002035 | 0.002189 | 0.002629 |
| BBTAb | Borrelia burgdorferi Ab [Units/volume] in Serum by Immunoassay | toxicology / pharmacology | TOTAL Ab BORELIA BURGDORF | Borrelia burgdorferi Ab [Units/volume] in Serum | BBTAb | | | -0.234 | 0.2286 | 684 | 407 | 0.2566 | 1.00e-06 | 1.00e-06 | 0.04613 | 0.2566 | 0.04613 | 0.3463 | 0.6071 | 0.06376 | 0.1325 | 0.351 |

| all_labs | full_name | group | concept_name | LOINC_NAME | heritability_names | selected_unit | sex_specific_lab | genetic.correlation | genetic.correlation.p | gcta.N_female | gcta.N_male | gcta.Variance_Autosomal_h2_female | gcta.Variance_Autosomal_h2_male | gcta.Variance_X_chromosome_h2_female | gcta.Variance_X_chromosome_h2_male | gcta.Variance_h2_female | gcta.Variance_h2_male | gcta.SE_Autosomal_h2_female | gcta.SE_Autosomal_h2_male | gcta.SE_X_chromosome_h2_female | gcta.SE_X_chromosome_h2_male | gcta.SE_h2_female |
|---|---|---|---|---|---|---|---|---|---|---|---|---|---|---|---|---|---|---|---|---|---|---|
| BeGlg_ | Beta globulin [Mass/volume] in Serum or Plasma by Electrophoresis | immune / inflammatory | BETA GLOBULIN | Beta globulin [Mass/volume] in Serum or Plasma by Electrophoresis | BeGlg_ | g/dL | | 0.09405 | 0.4901 | 3676 | 2889 | 0.02769 | 0.07518 | 1.00e-06 | 1.00e-06 | 0.02769 | 0.07518 | 0.05957 | 0.08334 | 0.01183 | 0.01423 | 0.06072 |
| BeHCG | Choriogonadotropin.beta subunit [Moles/volume] in Serum or Plasma | cancer | BETA-HCG SERUM | Choriogonadotropin.beta subunit [Moles/volume] in Serum or Plasma | BeHCG | 10*-3.[iU]/mL | Sex specific lab | | | 1620 | | 1.00e-06 | | 5.98e-04 | | 5.99e-04 | | 0.158 | | 0.02634 | | 0.1605 |
| BEVN | Base excess standard in Venous blood by calculation | metabolic | BASE EXCESS VENOUS | Base excess standard in Venous blood by calculation | BEVN | mmol/L | | -0.05464 | 0.6262 | 3271 | 3698 | 0.03697 | 0.02209 | 0.006819 | 1.00e-06 | 0.04379 | 0.02209 | 0.07182 | 0.06329 | 0.01447 | 0.01231 | 0.07348 |
| BGPIgG | Beta 2 glycoprotein 1 IgG Ab in Serum | immune / inflammatory | IgG BETA 2 GLYCOPROTEIN I | Beta 2 glycoprotein 1 IgG Ab [Units/volume] in Serum | BGPIgG | | | 0.2971 | 0.1821 | 1125 | 449 | 0.08164 | 0.3299 | 1.00e-06 | 0.1492 | 0.08164 | 0.4791 | 0.2139 | 0.5696 | 0.03528 | 0.1106 | 0.2133 |
| BGPIgM | Beta 2 glycoprotein 1 IgM Ab in Serum | immune / inflammatory | IgM BETA 2 GLYCOPROTEIN I | Beta 2 glycoprotein 1 IgM Ab [Units/volume] in Serum | BGPIgM | | | 0.1472 | 0.3999 | 1126 | 452 | 0.0784 | 0.149 | 1.00e-06 | 0.05753 | 0.0784 | 0.2066 | 0.1944 | 0.5807 | 0.03143 | 0.1057 | 0.1978 |
| Blasts | Blasts/100 leukocytes in Blood by Manual count | blood | BLAST CELLS % | Blasts/100 leukocytes in Blood by Manual count | Blasts | | | 0.01029 | 0.9484 | 769 | 867 | 0.7258 | 0.07961 | 1.00e-06 | 1.00e-06 | 0.7258 | 0.07961 | 0.3386 | 0.2999 | 0.05542 | 0.04684 | 0.3438 |
| BLOOD PRESSURE | Diastolic blood pressure | vitals | Diastolic blood pressure | Diastolic blood pressure | BLOOD.PRESSURE | mm[Hg] | | 0.1759 | 0.03298 | 19427 | 12921 | 0.07216 | 0.07651 | 1.00e-06 | 0.00356 | 0.07216 | 0.08007 | 0.01419 | 0.02045 | 0.002507 | 0.00384 | 0.01436 |
| BMI_CLEAN | BMI Clean | vitals | Body mass index (BMI) [Ratio] | Body mass index (BMI) [Ratio] | BMI_CLEAN | kg/m2 | | 0.6079 | 2.97e-38 | 35694 | 26408 | 0.2088 | 0.1691 | 0.003839 | 0.002995 | 0.2127 | 0.1721 | 0.009495 | 0.01174 | 0.00169 | 0.002016 | 0.009617 |
| BNP | Natriuretic peptide B [Mass/volume] in Serum or Plasma | cardiovascular | BRAIN NATRIURETIC PEPTIDE | Natriuretic peptide B [Mass/volume] in Serum or Plasma | BNP | pg/mL | | -0.02993 | 0.7087 | 7050 | 7214 | 0.03918 | 0.02454 | 1.00e-06 | 0.003855 | 0.03918 | 0.02839 | 0.03492 | 0.03525 | 0.005873 | 0.005601 | 0.03522 |
| BUN | Urea nitrogen serum/plasma | metabolic | UREA NITROGEN BLOOD | Ureanitrogen [Mass/volume] in Serum or Plasma | BUN | mg/dL | | 0.4088 | 4.77e-08 | 34401 | 26975 | 0.06104 | 0.07284 | 9.59e-04 | 0.003171 | 0.062 | 0.07601 | 0.008322 | 0.01041 | 0.001378 | 0.001962 | 0.008427 |
| C-Gluc | Glucose spinal fluid | metabolic | GLUCOSE SPINAL FLUID | Glucose [Mass/volume] in Cerebral spinal fluid | C.Gluc | mg/dL | | -0.227 | 0.02269 | 1515 | 1209 | 1.00e-06 | 0.1311 | 0.00148 | 1.00e-06 | 0.001481 | 0.1311 | 0.1682 | 0.2065 | 0.03181 | 0.03545 | 0.1699 |
| C3 | Complement C3 [Mass/volume] in Serum or Plasma | immune / inflammatory | C3 QUANTITATIVE BLOOD | Complement C3 [Mass/volume] in Serum or Plasma | C3 | mg/dL | | -0.06387 | 0.5051 | 2334 | 1372 | 0.05655 | 0.02861 | 1.00e-06 | 0.04217 | 0.05656 | 0.07077 | 0.1088 | 0.1707 | 0.01721 | 0.03795 | 0.1098 |

| all_labs | full_name | group | concept_name | LOINC_NAME | heritability_names | selected_unit | sex_specific_lab | genetic.correlation | genetic.correlation.p | gcta.N_female | gcta.N_male | gcta.Variance_Autosomal_h2_female | gcta.Variance_Autosomal_h2_male | gcta.Variance_X_chromosome_h2_female | gcta.Variance_X_chromosome_h2_male | gcta.Variance_h2_female | gcta.Variance_h2_male | gcta.SE_Autosomal_h2_female | gcta.SE_Autosomal_h2_male | gcta.SE_X_chromosome_h2_female | gcta.SE_X_chromosome_h2_male | gcta.SE_h2_female |
|---|---|---|---|---|---|---|---|---|---|---|---|---|---|---|---|---|---|---|---|---|---|---|
| C4Quan | Complement C4 [Mass/volume] in Serum or Plasma | immune / inflammatory | C4 QUANTITATIVE BLOOD | Complement C4 [Mass/volume] in Serum or Plasma | C4Quan | mg/dL | | 0.07114 | 0.4384 | 2347 | 1374 | 0.5938 | 0.7164 | 1.00e-06 | 1.00e-06 | 0.5938 | 0.7164 | 0.1017 | 0.1713 | 0.01744 | 0.03522 | 0.1034 |

| all_labs | full_name | group | concept_name | LOINC_NAME | heritability_names | selected_unit | sex_specific_lab | genetic.correlation | genetic.correlation.p | gcta.N_female | gcta.N_male | gcta.Variance_Autosomal_h2_female | gcta.Variance_Autosomal_h2_male | gcta.Variance_X_chromosome_h2_female | gcta.Variance_X_chromosome_h2_male | gcta.Variance_h2_female | gcta.Variance_h2_male | gcta.SE_Autosomal_h2_female | gcta.SE_Autosomal_h2_male | gcta.SE_X_chromosome_h2_female | gcta.SE_X_chromosome_h2_male | gcta.SE_h2_female |
|---|---|---|---|---|---|---|---|---|---|---|---|---|---|---|---|---|---|---|---|---|---|---|
| CA-125 | Cancer antigen 125 | cancer | CANCER ANTIGEN 125 | Cancer Ag 125 [Presence] in Serum or Plasma | CA.125 | [U]/mL | | -0.3415 | 0.1666 | 1247 | 119 | 0.07183 | | 1.00e-06 | | 0.07184 | | 0.1846 | | 0.03449 | | 0.1874 |
| Ca | Calcium serum/plasma serum/plasma | metabolic | CALCIUM BLOOD | Calcium [Mass/volume] in Serum or Plasma | Ca | mg/dL | | 0.5067 | 7.64e-12 | 34341 | 26934 | 0.07675 | 0.07265 | 8.32e-04 | 0.001862 | 0.07758 | 0.07451 | 0.008677 | 0.01047 | 0.001411 | 0.001959 | 0.008785 |
| CA19-9 | Cancer Ag 19-9 | cancer | CANCER ANTIGEN 19-9 | Cancer Ag 19-9 [Units/volume] in Serum or Plasma | CA19.9 | [U]/mL | | -0.1913 | 0.2928 | 758 | 749 | | 0.1469 | | 1.00e-06 | | 0.1469 | | 0.3173 | | 0.06449 | |
| CaIo | Calcium.ionized [Mass/volume] in Blood | metabolic | CALCIUM IONIZED | Calcium.ionized [Mass/volume] in Blood | CaIo | mg/dL | | 0.1089 | 0.1867 | 7839 | 9019 | 0.0613 | 0.02832 | 0.001012 | 1.00e-06 | 0.06231 | 0.02832 | 0.03245 | 0.02775 | 0.006326 | 0.004835 | 0.03291 |
| CBil | Bilirubin.conjugated [Mass/volume] in Serum or Plasma | liver | BILIRUBIN CONJUGATED | Bilirubin.conjugated [Mass/volume] in Serum or Plasma | CBil | mg/dL | | 0.2416 | 0.09883 | 14196 | 12855 | 0.01614 | 0.04439 | 1.00e-06 | 0.001956 | 0.01614 | 0.04634 | 0.01725 | 0.02017 | 0.003053 | 0.003436 | 0.01749 |
| CD3# | CD3# | immune / inflammatory | CD3 #/CUMM | CD3 cells [#/volume] in Blood | CD3. | mm3 | | -0.9412 | 0.5801 | 500 | 896 | | 0.04637 | | 7.96e-04 | | 0.04717 | | 0.2544 | | 0.05146 | |
| CD3% | CD3% | immune / inflammatory | PAN T CD3 % | | CD3_percent_ | % | | 0.06177 | 0.7497 | 570 | 926 | | 0.05753 | | 0.06977 | | 0.1273 | | 0.2798 | | 0.05514 | |
| CD4# | CD3+CD4+ (T4 helper) cells [#/volume] in Blood | immune / inflammatory | T HELPER CD4 #/CUMM | CD3+CD4+ (T4 helper) cells [#/volume] in Blood | CD4. | mm3 | | -0.7694 | 0.5594 | 498 | 894 | 1.00e-06 | 1.00e-06 | 0.004691 | 0.02616 | 0.004692 | 0.02616 | 0.5284 | 0.2943 | 0.1013 | 0.05616 | 0.5449 |
| CD4% | CD3+CD4+ (T4 helper) cells/100 cells in Blood | immune / inflammatory | T HELPER CD4 % | CD3+CD4+ (T4 helper) cells/100 cells in Blood | CD4_percent_ | % | | -0.5256 | 0.4184 | 582 | 945 | 0.4898 | 0.04955 | 1.00e-06 | 1.00e-06 | 0.4898 | 0.04955 | 0.4279 | 0.2572 | 0.0852 | 0.04614 | 0.4364 |
| CD8_3# | CD3+CD8+ (T8 suppressor cells) cells [#/volume] in Blood | immune / inflammatory | CD8(CD3+)/CD45 #/CUMM | CD3+CD8+ (T8 suppressor cells) cells [#/volume] in Blood | CD8_3. | mm3 | | -0.2004 | 0.2284 | 500 | 895 | 0.3543 | 0.3388 | 1.00e-06 | 1.00e-06 | 0.3543 | 0.3388 | 0.4877 | 0.2644 | 0.09178 | 0.05254 | 0.494 |

| all_labs | full_name | group | concept_name | LOINC_NAME | heritability_names | selected_unit | sex_specific_lab | genetic.correlation | genetic.correlation.p | gcta.N_female | gcta.N_male | gcta.Variance_Autosomal_h2_female | gcta.Variance_Autosomal_h2_male | gcta.Variance_X_chromosome_h2_female | gcta.Variance_X_chromosome_h2_male | gcta.Variance_h2_female | gcta.Variance_h2_male | gcta.SE_Autosomal_h2_female | gcta.SE_Autosomal_h2_male | gcta.SE_X_chromosome_h2_female | gcta.SE_X_chromosome_h2_male | gcta.SE_h2_female |
|---|---|---|---|---|---|---|---|---|---|---|---|---|---|---|---|---|---|---|---|---|---|---|
| CD8_3% | CD8_3% | immune / inflammatory | CD8(CD3+)/CD45 % | CD3+CD8+ (T8 suppressor cells) cells/100 cells in Specimen | CD8_3_percent_ | % | | -0.2654 | 0.1708 | 584 | 952 | 0.5995 | 0.02831 | 1.00e-06 | 0.01615 | 0.5995 | 0.04447 | 0.4327 | 0.2476 | 0.08462 | 0.04962 | 0.4304 |
| CEA | Carcinoembryonic Ag [Mass/volume] in Serum or Plasma | cancer | CARCINOEMBRYONIC ANTIGEN | Carcinoembryonic Ag [Mass/volume] in Serum or Plasma | CEA | ng/mL | | -0.1405 | 0.1728 | 1255 | 1266 | 0.1239 | | 1.00e-06 | | 0.1239 | | 0.2061 | | 0.03311 | | 0.2079 |
| Cerul | Ceruloplasmin [Mass/volume] in Serum or Plasma | blood | CERULOPLASMIN BLOOD | Ceruloplasmin [Mass/volume] in Serum or Plasma | Cerul | mg/dL | | -0.5308 | 0.527 | 676 | 698 | | 0.1549 | | 1.00e-06 | | 0.1549 | | 0.3796 | | 0.06587 | |
| CHOL_HDL_RATIO | Cholesterol in HDL [Mass/volume] in Serum or Plasma | metabolic | CHOL/HDL_RATIO | Cholesterol in HDL [Mass/volume] in Serum or Plasma | CHOL_HDL_RATIO | | | 0.108 | 0.4342 | 1151 | 1033 | 0.1284 | 0.655 | 1.00e-06 | 1.00e-06 | 0.1284 | 0.655 | 0.2 | 0.2415 | 0.03805 | 0.04244 | 0.2039 |
| Chol | Cholesterol [Mass/volume] in Serum or Plasma | metabolic | CHOLESTEROL BLOOD | Cholesterol [Mass/volume] in Serum or Plasma | Chol | mg/dL | | 0.3032 | 2.06e-05 | 19280 | 15405 | 0.1292 | 0.07666 | 7.44e-04 | 0.005654 | 0.13 | 0.08231 | 0.01487 | 0.01746 | 0.002432 | 0.003713 | 0.01506 |
| CKMBRe | Creatine kinase.MB [Mass/volume] in Serum or Plasma | cardiovascular | CK-MB | Creatine kinase.MB [Mass/volume] in Serum or Plasma | CKMBRe | ng/mL | | 0.09548 | 0.3182 | 6626 | 7983 | 0.04476 | 0.09776 | 1.00e-06 | 1.00e-06 | 0.04476 | 0.09776 | 0.03778 | 0.03208 | 0.005805 | 0.005274 | 0.03816 |
| Cl-WB | Chloride [Moles/volume] in Blood | metabolic | CHLORIDE WHOLE BLOOD | Chloride [Moles/volume] in Blood | Cl.WB | mmol/L | | -0.1274 | 0.1487 | 3199 | 4307 | 0.09024 | | 0.008919 | | 0.09916 | | 0.07601 | | 0.01445 | | 0.07692 |
| Cl | Chloride [Moles/volume] in Serum or Plasma | metabolic | CHLORIDE BLOOD | Chloride [Moles/volume] in Serum or Plasma | Cl | mmol/L | | 0.3462 | 1.33e-06 | 34342 | 26963 | 0.06485 | 0.03875 | 0.002768 | 0.002303 | 0.06762 | 0.04105 | 0.008456 | 0.009969 | 0.001577 | 0.001674 | 0.008589 |
| CLymph | Lymphocytes/100 leukocytes in Cerebral spinal fluid by Manual count | immune / inflammatory | LYMPHS % | Lymphocytes/100 leukocytes in Cerebral spinal fluid by Manual count | CLymph | % | | -0.1137 | 0.3251 | 1015 | 881 | 1.00e-06 | 0.2335 | 0.08221 | 1.00e-06 | 0.08221 | 0.2335 | 0.2592 | 0.2759 | 0.05955 | 0.05667 | 0.2648 |
| CMonMc | Monocytes+Macrophages/100 leukocytes in Cerebral spinal fluid by Manual count | immune / inflammatory | MONO/MACROPHAGE | Monocytes+Macrophages/100 leukocytes in Cerebral spinal fluid by Manual count | CMonMc | % | | 0.01043 | 0.9425 | 980 | 862 | 0.2218 | 0.1269 | 2.21e-04 | 1.00e-06 | 0.2221 | 0.1269 | 0.2592 | 0.297 | 0.04244 | 0.05488 | 0.2622 |
| CNeut | Neutrophils/100 leukocytes in Cerebral spinal fluid by Manual count | immune / inflammatory | NEUTROPHILS | Neutrophils/100 leukocytes in Cerebral spinal fluid by Manual count | CNeut | % | | 0.05092 | 0.7675 | 643 | 622 | 0.3104 | 0.7991 | 1.00e-06 | 1.00e-06 | 0.3104 | 0.7991 | 0.3758 | 0.4181 | 0.06816 | 0.06281 | 0.3799 |

| all_labs | full_name | group | concept_name | LOINC_NAME | heritability_names | selected_unit | sex_specific_lab | genetic.correlation | genetic.correlation.p | gcta.N_female | gcta.N_male | gcta.Variance_Autosomal_h2_female | gcta.Variance_Autosomal_h2_male | gcta.Variance_X_chromosome_h2_female | gcta.Variance_X_chromosome_h2_male | gcta.Variance_h2_female | gcta.Variance_h2_male | gcta.SE_Autosomal_h2_female | gcta.SE_Autosomal_h2_male | gcta.SE_X_chromosome_h2_female | gcta.SE_X_chromosome_h2_male | gcta.SE_h2_female |
|---|---|---|---|---|---|---|---|---|---|---|---|---|---|---|---|---|---|---|---|---|---|---|
| CNucCl | Nucleated cells [#/volume] in Cerebral spinal fluid | immune / inflammatory | NUCLEATED CELL | Nucleated cells [#/volume] in Cerebral spinal fluid by Manual count | CNucCl | /uL | | 0.2045 | 0.07009 | 1294 | 1062 | | 0.04164 | | 1.00e-06 | | 0.04164 | | 0.2254 | | 0.04135 | |
| CO2 | Carbon dioxide serum/plasma | blood | CARBON DIOXIDE BLOOD | Carbon dioxide, total [Moles/volume] in Serum or Plasma | CO2 | mmol/L | | 0.2193 | 3.59e-04 | 34362 | 26971 | 0.06545 | 0.04244 | 8.50e-05 | 0.002947 | 0.06554 | 0.04539 | 0.008437 | 0.01007 | 0.001225 | 0.00197 | 0.008508 |
| CoHgb | Carboxyhemoglobin % in blood | toxicology / pharmacology | CARBOXYHEMOGLOBIN | Carboxyhemoglobin/Hemoglobin.total in Blood | CoHgb | % | | -0.1553 | 0.1308 | 3020 | 4118 | | 0.006286 | | 1.00e-06 | | 0.006287 | | 0.06109 | | 0.01059 | |
| Copper | Copper in Serum or Plasma | toxicology / pharmacology | COPPER SERUM | Copper [Mass/volume] in Serum or Plasma | Copper | ug/dL | | -0.06839 | 0.6308 | 1198 | 893 | | | | | | | | | | | |
| Cort-S | Cortisol [Mass/volume] in Serum or Plasma | endocrine | CORTISOL | Cortisol [Mass/volume] in Serum or Plasma | Cort.S | ug/dL | | 0.02324 | 0.7185 | 4273 | 3588 | 0.05539 | 0.08245 | 0.002967 | 1.00e-06 | 0.05835 | 0.08245 | 0.0596 | 0.07028 | 0.01051 | 0.01234 | 0.06039 |
| CPepti | C peptide [Mass/volume] in Serum or Plasma | endocrine | C-PEPTIDE | C peptide [Mass/volume] in Serum or Plasma | CPepti | ng/mL | | 0.6335 | 0.5067 | 545 | 476 | | | | | | | | | | | |
| CPK | Creatine Kinase | cardiovascular | CK BLOOD | | CPK | [iU]/L | | 0.09418 | 0.2542 | 9073 | 9195 | 0.06263 | 0.06744 | 0.001775 | 1.00e-06 | 0.06441 | 0.06744 | 0.02812 | 0.02748 | 0.005058 | 0.005179 | 0.02852 |
| CPKTot | Creatine kinase [Enzymatic activity/volume] in Serum or Plasma | cardiovascular | CREATINE KINASE TOTAL | Creatine kinase [Enzymatic activity/volume] in Serum or Plasma | CPKTot | [U]/L | | -0.009484 | 0.9135 | 4518 | 4635 | 0.1082 | 0.0251 | 6.09e-04 | 0.007577 | 0.1088 | 0.03267 | 0.05545 | 0.05364 | 0.009955 | 0.009957 | 0.05639 |
| CProt1 | Protein [Mass/volume] in Cerebral spinal fluid | metabolic | PROTEIN CSF | Protein [Mass/volume] in Cerebral spinal fluid | CProt1 | mg/dL | | 0.2802 | 0.0228 | 1540 | 1211 | 0.3832 | 0.006106 | 0.004187 | 1.00e-06 | 0.3874 | 0.006107 | 0.1612 | 0.2098 | 0.02813 | 0.03889 | 0.1624 |
| CR-Day | Creatinine urine per day | kidney | CREATININE URINE PER DAY | Creatinine [Mass/time] in 24 hour Urine | CR.Day | mg/(24.h) | | -0.3417 | 0.07995 | 705 | 423 | | 0.2821 | | 1.00e-06 | | 0.2821 | | 0.5901 | | 0.09775 | |
| Cr24Hr | Creatinine [Mass/time] in 24 hour Urine | kidney | CREATININE G/24 HOUR | Creatinine [Mass/time] in 24 hour Urine | Cr24Hr | g/(24.h) | | 0.1227 | 0.8341 | 870 | 618 | 1.00e-06 | 0.05239 | 0.02831 | 1.00e-06 | 0.02831 | 0.0524 | 0.2937 | 0.3955 | 0.04732 | 0.07011 | 0.2969 |
| CRBC | Erythrocytes in Cerebral spinal fluid | other | RBC | Erythrocytes [#/volume] in Cerebral spinal fluid by Manual count | CRBC | /uL | | 0.08069 | 0.4463 | 1296 | 1072 | 1.00e-06 | 0.1296 | 0.01349 | 0.02802 | 0.01349 | 0.1576 | 0.1821 | 0.2343 | 0.03808 | 0.04276 | 0.1831 |
| Creat | Creatinine [Mass/volume] in Blood Creatinine [Mass/volume] in Blood | kidney | CREATININE BLOOD | Creatinine [Mass/volume] in Blood | Creat | mg/dL | | 0.347 | 7.51e-08 | 34787 | 27025 | 0.07584 | 0.06417 | 0.001228 | 0.003797 | 0.07707 | 0.06797 | 0.00852 | 0.01028 | 0.001331 | 0.002104 | 0.008607 |
| CREATININE | Creatinine | kidney | CREATININE | | CREATININE | mg/dL | | 0.01097 | 0.9387 | 879 | 755 | | 1.00e-06 | | 0.01744 | | 0.01744 | | 0.3562 | | 0.05511 | |

| all_labs | full_name | group | concept_name | LOINC_NAME | heritability_names | selected_unit | sex_specific_lab | genetic.correlation | genetic.correlation.p | gcta.N_female | gcta.N_male | gcta.Variance_Autosomal_h2_female | gcta.Variance_Autosomal_h2_male | gcta.Variance_X_chromosome_h2_female | gcta.Variance_X_chromosome_h2_male | gcta.Variance_h2_female | gcta.Variance_h2_male | gcta.SE_Autosomal_h2_female | gcta.SE_Autosomal_h2_male | gcta.SE_X_chromosome_h2_female | gcta.SE_X_chromosome_h2_male | gcta.SE_h2_female |
|---|---|---|---|---|---|---|---|---|---|---|---|---|---|---|---|---|---|---|---|---|---|---|
| CrePOC | Creatinine (point of care) | kidney | POC CREATININE | | CrePOC | mg/dL | | -0.02487 | 0.7719 | 2951 | 3717 | 0.1637 | 0.02222 | 1.00e-06 | 1.00e-06 | 0.1637 | 0.02222 | 0.08821 | 0.06434 | 0.01545 | 0.01111 | 0.08936 |
| CRP | C reactive protein [Mass/volume] in Serum or Plasma | immune / inflammatory | C-REACTIVE PROTEIN | C reactive protein [Mass/volume] in Serum or Plasma | CRP | mg/L | | 0.01353 | 0.8901 | 11849 | 8464 | 0.0757 | 0.023 | 0.00104 | 0.005726 | 0.07674 | 0.02873 | 0.02198 | 0.03 | 0.003811 | 0.005774 | 0.0223 |
| CTo#Cl | Cells Counted Total in CSF | other | TOTAL # CELLS | Cells Counted Total [#] in Cerebral spinal fluid | CTo.Cl | | | 0.3162 | 0.1004 | 1019 | 888 | 0.34 | | 0.02533 | | 0.3653 | | 0.2431 | | 0.04815 | | 0.2491 |
| D25OHT | Calcitriol [Mass/volume] in Serum or Plasma | metabolic | VITAMIN D 25-OH TOTAL | Vitamin D+Metabolites [Mass/volume] in Serum or Plasma | D25OHT | ng/mL | | 0.2996 | 7.44e-04 | 13921 | 7363 | 0.07689 | 0.08735 | 0.003769 | 0.01107 | 0.08066 | 0.09842 | 0.01932 | 0.03584 | 0.00358 | 0.007344 | 0.01964 |
| DHEA-S | DHEA-S in Serum or Plasma | endocrine | DHEA SULFATE | Dehydroepiandrosterone sulfate (DHEA-S) [Mass/volume] in Serum or Plasma | DHEA.S | ug/dL | | -0.2884 | 0.4191 | 1077 | 119 | 1.00e-06 | | 0.00286 | | 0.002861 | | 0.2123 | | 0.03045 | | 0.2146 |
| DIASTOLIC | Diastolic blood pressure | vitals | Diastolic blood pressure | Diastolic blood pressure | DIASTOLIC | mm[Hg] | | 0.2692 | 4.85e-04 | 35815 | 26957 | 0.06512 | 0.05864 | 0.0039 | 1.00e-06 | 0.06902 | 0.05864 | 0.008127 | 0.01017 | 0.001673 | 0.001787 | 0.008289 |
| Digox | Digoxin [Mass/volume] in Serum or Plasma | toxicology / pharmacology | DIGOXIN | Digoxin [Mass/volume] in Serum or Plasma | Digox | ng/mL | | -0.4863 | 0.5306 | 551 | 794 | 1.00e-06 | 0.1933 | 0.05064 | 0.1153 | 0.05064 | 0.3086 | 0.6071 | 0.3101 | 0.1019 | 0.06978 | 0.6157 |
| Dimer | Dimer | blood | D-DIMER (PATIENT) | Fibrin D-dimer [Units/volume] in Platelet poor plasma | Dimer | ug{FEU}/mL | | 0.01164 | 0.9407 | 962 | 793 | 0.02107 | 0.3454 | 1.00e-06 | 1.00e-06 | 0.02107 | 0.3454 | 0.2401 | 0.324 | 0.04465 | 0.06099 | 0.2456 |
| DirLDL | Cholesterol in LDL [Mass/volume] in Serum or Plasma by Direct assay | metabolic | LDL DIRECT | Cholesterol in LDL [Mass/volume] in Serum or Plasma by Direct assay | DirLDL | mg/dL | | 0.07668 | 0.3952 | 2604 | 2992 | 0.1115 | 0.007763 | 0.01399 | 1.00e-06 | 0.1255 | 0.007764 | 0.09467 | 0.08042 | 0.01939 | 0.01825 | 0.09609 |
| DLCO_COR_PPR | DLCO_COR_PPR | lung | DLCO_COR_PPR | | DLCO_COR_PPR | | | 0.1555 | 0.196 | 1226 | 1268 | | 0.2163 | | 1.00e-06 | | 0.2163 | | 0.2015 | | 0.03543 | |
| DLCO_COR_PRD | DLCO_COR_PRD | lung | DLCO_COR_PRD | | DLCO_COR_PRD | | | -0.01478 | 0.8561 | 2056 | 1955 | 0.4636 | 0.2871 | 0.001732 | 0.0185 | 0.4653 | 0.3056 | 0.1281 | 0.1308 | 0.01929 | 0.02311 | 0.1288 |
| DLCO_COR_PRL | DLCO_COR_PRL | lung | DLCO_COR_PRL | | DLCO_COR_PRL | | | 0.01935 | 0.885 | 1225 | 1269 | 0.362 | 0.3227 | 1.00e-06 | 0.02722 | 0.3621 | 0.3499 | 0.2066 | 0.1943 | 0.0336 | 0.03581 | 0.2078 |
| DLCO_COR_PRU | DLCO_COR_PRU | lung | DLCO_COR_PRU | | DLCO_COR_PRU | | | 0.1692 | 0.1164 | 1225 | 1269 | 0.4477 | 0.3747 | 0.03191 | 0.01623 | 0.4796 | 0.3909 | 0.2186 | 0.1932 | 0.0375 | 0.03367 | 0.2202 |
| DLCO_COR_ZBD | DLCO_COR_ZBD | lung | DLCO_COR_ZBD | Diffusion capacity.carbon monoxide adjusted for hemoglobin by Helium single breath | DLCO_COR_ZBD | | | 0.08097 | 0.5066 | 2056 | 1955 | | 0.06303 | | 1.00e-06 | | 0.06303 | | 0.1288 | | 0.02499 | |

| all_labs | full_name | group | concept_name | LOINC_NAME | heritability_names | selected_unit | sex_specific_lab | genetic.correlation | genetic.correlation.p | gcta.N_female | gcta.N_male | gcta.Variance_Autosomal_h2_female | gcta.Variance_Autosomal_h2_male | gcta.Variance_X_chromosome_h2_female | gcta.Variance_X_chromosome_h2_male | gcta.Variance_h2_female | gcta.Variance_h2_male | gcta.SE_Autosomal_h2_female | gcta.SE_Autosomal_h2_male | gcta.SE_X_chromosome_h2_female | gcta.SE_X_chromosome_h2_male | gcta.SE_h2_female |
|---|---|---|---|---|---|---|---|---|---|---|---|---|---|---|---|---|---|---|---|---|---|---|
| DLCO_UNC_PPR | DLCO_UNC_PPR | lung | DLCO_UNC_PPR | | DLCO_UNC_PPR | | | 0.1828 | 0.1506 | 1227 | 1269 | | 0.2244 | | 1.00e-06 | | 0.2244 | | 0.1992 | | 0.03441 | |
| DLCO_UNC_PRD | DLCO_UNC_PRD | lung | DLCO_UNC_PRD | | DLCO_UNC_PRD | | | -0.01444 | 0.8574 | 2057 | 1955 | 0.4628 | 0.2871 | 0.001573 | 0.0185 | 0.4643 | 0.3056 | 0.1281 | 0.1308 | 0.01929 | 0.02311 | 0.1288 |
| DLCO_UNC_PRL | DLCO_UNC_PRL | lung | DLCO_UNC_PRL | | DLCO_UNC_PRL | | | 0.02033 | 0.8804 | 1226 | 1269 | 0.3584 | 0.3227 | 1.00e-06 | 0.02722 | 0.3584 | 0.3499 | 0.2064 | 0.1943 | 0.03367 | 0.03581 | 0.2077 |
| DLCO_UNC_PRU | DLCO_UNC_PRU | lung | DLCO_UNC_PRU | | DLCO_UNC_PRU | | | 0.17 | 0.1153 | 1226 | 1269 | 0.438 | 0.3747 | 0.03156 | 0.01623 | 0.4695 | 0.3909 | 0.2184 | 0.1932 | 0.03751 | 0.03367 | 0.2201 |
| DLCO_UNC_ZBD | DLCO_UNC_ZBD | lung | DLCO_UNC_ZBD | | DLCO_UNC_ZBD | | | 0.2549 | 0.04728 | 1227 | 1269 | | 0.205 | | 1.00e-06 | | 0.205 | | 0.2005 | | 0.03577 | |
| DLCOOVA_PPR | DLCOOVA_PPR | lung | DLCOOVA_PPR | | DLCOOVA_PPR | % | | 0.2546 | 0.05688 | 1152 | 976 | | 0.1456 | | 1.00e-06 | | 0.1456 | | 0.2469 | | 0.04349 | |
| DLCOOVA_PRD | DLCOOVA_PRD | lung | DLCOOVA_PRD | | DLCOOVA_PRD | | | 0.002167 | 0.9775 | 2056 | 1954 | 0.623 | 0.2066 | 0.006027 | 0.01869 | 0.629 | 0.2253 | 0.1261 | 0.1299 | 0.01942 | 0.02258 | 0.127 |
| DLCOOVA_ZBD | DLCOOVA_ZBD | lung | DLCOOVA_ZBD | | DLCOOVA_ZBD | | | -0.06327 | 0.4926 | 2057 | 1954 | | 0.1901 | | 0.00121 | | 0.1913 | | 0.1285 | | 0.0203 | |
| eGFR | Estimation of glomerular filtration rate | kidney | ESTIMATED GLOMERULAR FILTRATION RATE | | eGFR | mL/min | | 0.7549 | 2.55e-04 | 936 | 680 | 0.3771 | 1.00e-06 | 0.06729 | 0.02675 | 0.4444 | 0.02675 | 0.2607 | 0.4828 | 0.05209 | 0.08653 | 0.2686 |
| EGFR | Estimation of glomerular filtration rate | kidney | Estimation of glomerular filtration rate | | EGFR | mL/min/1.73.m2 | | 0.3224 | 1.43e-08 | 34789 | 27030 | 0.07859 | 0.05722 | 0.001297 | 0.003203 | 0.07989 | 0.06042 | 0.0086 | 0.01011 | 0.001359 | 0.002007 | 0.008692 |
| eGFRcr | Estimation of glomerular filtration rate (eGFRcr) | kidney | eGFRcr | | eGFRcr | | | 0.2322 | 0.05064 | 1649 | 1251 | | 0.1505 | | 1.00e-06 | | 0.1505 | | 0.1976 | | 0.0373 | |

| all_labs | full_name | group | concept_name | LOINC_NAME | heritability_names | selected_unit | sex_specific_lab | genetic.correlation | genetic.correlation.p | gcta.N_female | gcta.N_male | gcta.Variance_Autosomal_h2_female | gcta.Variance_Autosomal_h2_male | gcta.Variance_X_chromosome_h2_female | gcta.Variance_X_chromosome_h2_male | gcta.Variance_h2_female | gcta.Variance_h2_male | gcta.SE_Autosomal_h2_female | gcta.SE_Autosomal_h2_male | gcta.SE_X_chromosome_h2_female | gcta.SE_X_chromosome_h2_male | gcta.SE_h2_female |
|---|---|---|---|---|---|---|---|---|---|---|---|---|---|---|---|---|---|---|---|---|---|---|
| Endomy | Endomysium IgA Ab [Presence] in Serum | immune / inflammatory | ENDOMYSIAL IGA AUTOABS | Endomysium IgA Ab [Presence] in Serum | Endomy | [U] | | -0.04255 | 0.8685 | 817 | 277 | | 0.1904 | | 1.00e-06 | | 0.1904 | | 0.9834 | | 0.2211 | |
| EoAbs | Absolute Eosinophils | immune / inflammatory | EO AUTOMATED ABS | | EoAbs | 10*3/uL | | 0.2937 | 1.62e-05 | 14884 | 10829 | 0.1078 | 0.1338 | 7.24e-04 | 0.004824 | 0.1085 | 0.1387 | 0.0182 | 0.02535 | 0.003067 | 0.004787 | 0.01841 |
| Eos | Eosinophils | immune / inflammatory | EOSINOPHIL % | | Eos | % | | 0.354 | 2.63e-08 | 15677 | 11491 | 0.0746 | 0.1201 | 1.00e-06 | 1.00e-06 | 0.07461 | 0.1201 | 0.01677 | 0.02368 | 0.002941 | 0.004144 | 0.017 |
| EOSIA | Eosinophils [#/volume] in Blood by Automated count | immune / inflammatory | EOSOINOPHIL (ABS) | Eosinophils [#/volume] in Blood | EOSIA | 10*3/uL | | 0.2388 | 0.002057 | 19735 | 15202 | 0.1033 | 0.07505 | 1.00e-06 | 0.004173 | 0.1033 | 0.07922 | 0.01436 | 0.01748 | 0.002427 | 0.003573 | 0.01453 |
| EOSIAB | Eosinophils [#/volume] in Blood by Automated count | immune / inflammatory | EOSOINOPHIL (ABS) | Eosinophils [#/volume] in Blood by Automated count | EOSIAB | 10*3/uL | | 0.2652 | 0.002948 | 15257 | 12637 | 0.08003 | 0.0511 | 1.00e-06 | 0.001801 | 0.08003 | 0.0529 | 0.01755 | 0.02016 | 0.002981 | 0.003698 | 0.01775 |

| all_labs | full_name | group | concept_name | LOINC_NAME | heritability_names | selected_unit | sex_specific_lab | genetic.correlation | genetic.correlation.p | gcta.N_female | gcta.N_male | gcta.Variance_Autosomal_h2_female | gcta.Variance_Autosomal_h2_male | gcta.Variance_X_chromosome_h2_female | gcta.Variance_X_chromosome_h2_male | gcta.Variance_h2_female | gcta.Variance_h2_male | gcta.SE_Autosomal_h2_female | gcta.SE_Autosomal_h2_male | gcta.SE_X_chromosome_h2_female | gcta.SE_X_chromosome_h2_male | gcta.SE_h2_female |
|---|---|---|---|---|---|---|---|---|---|---|---|---|---|---|---|---|---|---|---|---|---|---|
| EOSIM | Eosinophils/100 leukocytes in Blood by Automated count | immune / inflammatory | EOSINOPHILS % | Eosinophils/100 leukocytes in Blood by Automated count | EOSIM | % | | 0.2813 | 8.76e-05 | 19669 | 15117 | 0.09309 | 0.07274 | 1.00e-06 | 0.003368 | 0.09309 | 0.07611 | 0.01423 | 0.01779 | 0.002553 | 0.003404 | 0.0144 |
| EOSINOPHILS | EOSINOPHILS | immune / inflammatory | EOSINOPHILS | Eosinophils/100 leukocytes in Cerebral spinal fluid by Manual count | EOSINOPHILS | % | | -0.07787 | 0.6382 | 1111 | 730 | | 1.00e-06 | | 0.008753 | | 0.008754 | | 0.3324 | | 0.05644 | |
| EOSIRE | Eosinophils % | immune / inflammatory | EOSINOPHIL % | Eosinophils/100 leukocytes in Blood | EOSIRE | % | | 0.3212 | 8.55e-04 | 15398 | 12813 | 0.06993 | 0.04885 | 1.00e-06 | 1.00e-06 | 0.06993 | 0.04885 | 0.01731 | 0.02016 | 0.002753 | 0.003688 | 0.01752 |
| Estrdl | Estradiol (E2) [Mass/volume] in Serum or Plasma | endocrine | ESTRADIOL | Estradiol(E2) [Mass/volume] in Serum or Plasma | Estrdl | pg/mL | | -0.02537 | 0.9366 | 2960 | 169 | | 0.9116 | | 0.0529 | | 0.9645 | | 1.534 | | 0.2751 | |
| F-Gluc | Glucose [Mass/volume] in Body fluid | metabolic | GLUCOSE BODY FLUID | Glucose [Mass/volume] in Body fluid | F.Gluc | mg/dL | | 0.09348 | 0.5878 | 716 | 975 | 0.3607 | 0.04497 | 0.06687 | 0.03798 | 0.4276 | 0.08295 | 0.3353 | 0.2503 | 0.06291 | 0.05035 | 0.3391 |
| Fbrngn | Fibrinogen [Mass/volume] in Platelet poor plasma by Coagulation assay | blood | FIBRINOGEN | Fibrinogen [Mass/volume] in Platelet poor plasma by Coagulation assay | Fbrngn | mg/dL | | 0.01546 | 0.8637 | 2953 | 3225 | | 0.01023 | | 1.00e-06 | | 0.01023 | | 0.07524 | | 0.01177 | |
| FEF25_75_AFD | FEF AFD | lung | FEF25_75_AFD | | FEF25_75_AFD | | | 0.2091 | 0.07991 | 1018 | 810 | 0.1042 | 0.1019 | 0.01744 | 1.00e-06 | 0.1216 | 0.1019 | 0.2516 | 0.3117 | 0.04055 | 0.05641 | 0.2538 |
| FEF25_75_PPR | FEF PPR | lung | FEF25_75_PPR | | FEF25_75_PPR | % | | 0.08566 | 0.4039 | 1559 | 1210 | 0.19 | 0.02528 | 0.03307 | 0.03693 | 0.2231 | 0.06221 | 0.1616 | 0.2004 | 0.02783 | 0.03827 | 0.1647 |
| FEF25_75_PRD | FEF PRD | lung | FEF25_75_PRD | | FEF25_75_PRD | | | 0.1379 | 0.06314 | 2730 | 2375 | 0.3741 | 0.2197 | 0.006857 | 0.009252 | 0.3809 | 0.229 | 0.09429 | 0.1084 | 0.01701 | 0.01887 | 0.09526 |
| FEF25_75_PRL | FEF PRL | lung | FEF25_75_PRL | | FEF25_75_PRL | | | 0.01535 | 0.8737 | 1686 | 1581 | 0.3489 | 0.3044 | 0.0278 | 1.00e-06 | 0.3767 | 0.3044 | 0.1538 | 0.1576 | 0.02664 | 0.0282 | 0.1554 |
| FEF25_75_ZBD | FEF ZBD | lung | FEF25_75_ZBD | | FEF25_75_ZBD | | | 0.2388 | 6.02e-04 | 2730 | 2375 | 0.06348 | 0.3195 | 0.04072 | 0.01044 | 0.1042 | 0.3299 | 0.09028 | 0.1091 | 0.01954 | 0.02037 | 0.09213 |
| FEF25_75_ZPC | FEF ZPC | lung | FEF25_75_ZPC | | FEF25_75_ZPC | | | -0.8538 | 0.6532 | 576 | 497 | 0.4049 | 0.02334 | 0.05905 | 1.00e-06 | 0.464 | 0.02334 | 0.4411 | 0.5282 | 0.0751 | 0.1112 | 0.4452 |
| FEosin | Eosinophils/100 leukocytes in Body fluid by Manual count | immune / inflammatory | EOSINOPHILS | Eosinophils/100 leukocytes in Body fluid by Manual count | FEosin | % | | -0.03721 | 0.9387 | 512 | 669 | 1.00e-06 | 0.3737 | 0.03208 | 1.00e-06 | 0.03208 | 0.3737 | 0.4856 | 0.3671 | 0.08734 | 0.06806 | 0.4875 |
| Ferrit | Ferritin [Mass/volume] in Serum or Plasma | blood | FERRITIN | Ferritin [Mass/volume] in Serum or Plasma | Ferrit | ng/mL | | -0.002081 | 0.9862 | 11119 | 8034 | 0.03239 | 0.009634 | 1.00e-06 | 2.81e-04 | 0.03239 | 0.009915 | 0.02208 | 0.02896 | 0.004215 | 0.005966 | 0.0225 |
| FET_AFD | FET AFD | lung | FET_AFD | | FET_AFD | | | -0.2443 | 0.1989 | 585 | 503 | 0.5229 | | 0.01042 | | 0.5333 | | 0.4482 | | 0.06567 | | 0.454 |
| FET_ZBD | FET ZBD | lung | FET_ZBD | | FET_ZBD | | | 0.1245 | 0.3353 | 1730 | 1595 | 0.06596 | 0.01468 | 1.11e-04 | 1.00e-06 | 0.06607 | 0.01468 | 0.1479 | 0.1606 | 0.02225 | 0.03595 | 0.1493 |
| FET_ZPC | FET ZPC | lung | FET_ZPC | | FET_ZPC | | | 0.2483 | 0.3934 | 580 | 485 | 1.00e-06 | 0.3131 | 0.2574 | 0.04438 | 0.2574 | 0.3575 | 0.4012 | 0.5331 | 0.09876 | 0.1018 | 0.4063 |

| all_labs | full_name | group | concept_name | LOINC_NAME | heritability_names | selected_unit | sex_specific_lab | genetic.correlation | genetic.correlation.p | gcta.N_female | gcta.N_male | gcta.Variance_Autosomal_h2_female | gcta.Variance_Autosomal_h2_male | gcta.Variance_X_chromosome_h2_female | gcta.Variance_X_chromosome_h2_male | gcta.Variance_h2_female | gcta.Variance_h2_male | gcta.SE_Autosomal_h2_female | gcta.SE_Autosomal_h2_male | gcta.SE_X_chromosome_h2_female | gcta.SE_X_chromosome_h2_male | gcta.SE_h2_female |
|---|---|---|---|---|---|---|---|---|---|---|---|---|---|---|---|---|---|---|---|---|---|---|
| FEV1_PPR | FEV1 PPR | lung | FEV1_PPR | FEV1 measured/predicted | FEV1_PPR | % | | 0.1924 | 0.05023 | 1562 | 1214 | 0.1536 | 0.06846 | 1.00e-06 | 0.02213 | 0.1536 | 0.09058 | 0.1591 | 0.2021 | 0.02582 | 0.03481 | 0.1615 |
| FEV1_PRD | FEV1 PRD | lung | FEV1_PRD | | FEV1_PRD | L | | 0.1457 | 0.1504 | 1562 | 1214 | 0.4196 | 0.3023 | 1.00e-06 | 0.02963 | 0.4196 | 0.3319 | 0.1583 | 0.2084 | 0.02704 | 0.04103 | 0.1608 |
| FEV1_PRL | FEV1 PRL | lung | FEV1_PRL | | FEV1_PRL | | | 0.03405 | 0.656 | 1735 | 1604 | 0.4907 | 0.63 | 0.006001 | 1.00e-06 | 0.4967 | 0.63 | 0.1479 | 0.1554 | 0.02271 | 0.02606 | 0.1493 |
| FEV1_ZBD | FEV1 ZBD | lung | FEV1_ZBD | FEV1 --pre bronchodilation | FEV1_ZBD | L | | 0.1316 | 0.2353 | 1562 | 1214 | 0.07979 | 0.04636 | 0.008953 | 0.01637 | 0.08874 | 0.06272 | 0.1572 | 0.1965 | 0.02483 | 0.03381 | 0.1592 |
| FEV1_ZPC | FEV1 ZPC | lung | FEV1_ZPC | | FEV1_ZPC | | | 0.2758 | 0.1888 | 567 | 488 | 0.2213 | 0.5207 | 0.04509 | 1.00e-06 | 0.2664 | 0.5207 | 0.4401 | 0.4835 | 0.0705 | 0.07236 | 0.4452 |
| FEV1OFVC_AFD | FEV1/FVC AFD | lung | FEV1OFVC_AFD | | FEV1OFVC_AFD | | | 0.0499 | 0.8345 | 587 | 506 | 0.4889 | 0.7411 | 0.009198 | 1.00e-06 | 0.4981 | 0.7411 | 0.422 | 0.466 | 0.06451 | 0.1007 | 0.4291 |
| FEV1OFVC_APP | FEV1/FVC APP | lung | FEV1OFVC_APP | | FEV1OFVC_APP | | | 0.07811 | 0.7177 | 587 | 506 | 0.5258 | 0.8707 | 0.01116 | 1.00e-06 | 0.537 | 0.8707 | 0.4217 | 0.4668 | 0.06445 | 0.09857 | 0.4287 |
| FEV1OFVC_PPR | FEV1/FVC PPR | lung | FEV1OFVC_PPR | | FEV1OFVC_PPR | | | 0.2422 | 0.3359 | 1735 | 1604 | 0.152 | 0.3025 | 0.03449 | 1.00e-06 | 0.1865 | 0.3025 | 0.1484 | 0.1533 | 0.02732 | 0.02724 | 0.1493 |
| FEV1OFVC_PRD | FEV1/FVC PRD | lung | FEV1OFVC_PRD | | FEV1OFVC_PRD | | | 0.08869 | 0.3778 | 1735 | 1604 | 0.1403 | 0.192 | 1.00e-06 | 1.00e-06 | 0.1403 | 0.192 | 0.143 | 0.158 | 0.02154 | 0.03182 | 0.1443 |
| FEV1OFVC_PRL | FEV1/FVC PRL | lung | FEV1OFVC_PRL | | FEV1OFVC_PRL | | | 0.1074 | 0.2598 | 1735 | 1604 | 0.2206 | 1.00e-06 | 1.00e-06 | 0.003951 | 0.2206 | 0.003952 | 0.1427 | 0.157 | 0.02357 | 0.02806 | 0.1447 |
| FEV1OFVC_ZBD | FEV1/FVC ZBD | lung | FEV1OFVC_ZBD | | FEV1OFVC_ZBD | | | 0.2075 | 0.0114 | 1734 | 1604 | 0.1484 | 0.279 | 0.04018 | 1.00e-06 | 0.1886 | 0.279 | 0.1487 | 0.153 | 0.02796 | 0.02746 | 0.1499 |
| FIO2 | Oxygen/Inspired gas Respiratory system --on ventilator | blood | FIO2 | Oxygen/Inspired gas Respiratory system --on ventilator | FIO2 | | | 0.2162 | 0.04056 | 3411 | 4836 | | 0.03878 | | 1.00e-06 | | 0.03878 | | 0.05101 | | 0.008696 | |
| FK-506 | Tacrolimus [Mass/volume] in Blood | toxicology / pharmacology | FK-506 (TACROLIMUS ) | Tacrolimus [Mass/volume] in Blood | FK.506 | ng/mL | | -0.2561 | 0.5097 | 673 | 960 | 0.4433 | 0.4256 | 0.0205 | 0.05412 | 0.4638 | 0.4797 | 0.3717 | 0.2587 | 0.06287 | 0.05039 | 0.3767 |
| FLymph | Lymphocytes [#/volume] in Body fluid by Manual count | immune / inflammatory | LYMPHS % | Lymphocytes /100 leukocytes in Blood | FLymph | % | | 0.108 | 0.3819 | 1301 | 1616 | 0.1529 | 1.00e-06 | 0.02467 | 0.00138 | 0.1776 | 0.001381 | 0.2007 | 0.1602 | 0.03363 | 0.0242 | 0.2035 |
| FMoncl | Monocytes+Macrophages/100 leukocytes in Body fluid by Manual count | immune / inflammatory | MONONUCLEAR CELLS | Monocytes+Macrophages/100 leukocytes in Body fluid by Manual count | FMoncl | % | | -0.1158 | 0.5744 | 660 | 793 | | | | | | | | | | | |
| FMonMc | Mononuclear cells [#/volume] in Body fluid by Manual count | immune / inflammatory | MONO/MACROPHAGE | Mononuclear cells [#/volume] in Body fluid by Manual count | FMonMc | % | | 0.05183 | 0.6765 | 744 | 916 | | | | | | | | | | | |

| all_labs | full_name | group | concept_name | LOINC_NAME | heritability_names | selected_unit | sex_specific_lab | genetic.correlation | genetic.correlation.p | gcta.N_female | gcta.N_male | gcta.Variance_Autosomal_h2_female | gcta.Variance_Autosomal_h2_male | gcta.Variance_X_chromosome_h2_female | gcta.Variance_X_chromosome_h2_male | gcta.Variance_h2_female | gcta.Variance_h2_male | gcta.SE_Autosomal_h2_female | gcta.SE_Autosomal_h2_male | gcta.SE_X_chromosome_h2_female | gcta.SE_X_chromosome_h2_male | gcta.SE_h2_female |
|---|---|---|---|---|---|---|---|---|---|---|---|---|---|---|---|---|---|---|---|---|---|---|
| FNeut | Neutrophils/100 leukocytes in Body fluid by Manual count | immune / inflammatory | NEUTROPHILS | Neutrophils/100 leukocytes in Body fluid by Manual count | FNeut | % | | 0.3241 | 0.3685 | 1288 | 1610 | | 1.00e-06 | | 0.03506 | | 0.03506 | | 0.1642 | | 0.03163 | |
| FNucCl | Nucleated cells [#/volume] in Synovial fluid by Manual count | blood | NUCLEATED CELL | Nucleated cells [#/volume] in Body fluid | FNucCl | /uL | | -0.003602 | 0.9667 | 1574 | 1952 | 0.233 | 0.05094 | 0.0539 | 0.03874 | 0.2869 | 0.08969 | 0.1593 | 0.1262 | 0.03214 | 0.02801 | 0.1629 |
| FProt1 | Protein body fluid | metabolic | PROTEIN BODY FLUID | Protein [Mass/volume] in Body fluid | FProt1 | g/dL | | -0.2864 | 0.3649 | 743 | 990 | | 0.1813 | | 1.00e-06 | | 0.1813 | | 0.2458 | | 0.04577 | |
| FRBC1 | Erythrocytes [#/volume] in Blood by Automated count | blood | RBC | Erythrocytes [#/volume] in Body fluid by Automated count | FRBC1 | /uL | | 0.0153 | 0.8872 | 1498 | 1855 | 0.1477 | | 1.00e-06 | | 0.1477 | | 0.1661 | | 0.02851 | | 0.1683 |
| FRC_PPR | FRC PPR | lung | FRC_PPR | | FRC_PPR | | | -0.1254 | 0.2096 | 1144 | 1145 | 0.1857 | 0.3996 | 0.03308 | 1.00e-06 | 0.2188 | 0.3996 | 0.2165 | 0.2064 | 0.04211 | 0.04118 | 0.2181 |
| FRC_PRD | FRC PRD | lung | FRC_PRD | | FRC_PRD | | | 0.1913 | 0.1704 | 1147 | 1144 | 0.4813 | 0.1283 | 1.00e-06 | 0.03008 | 0.4813 | 0.1583 | 0.2217 | 0.2141 | 0.0339 | 0.0406 | 0.2222 |
| FRC_PRL | FRC PRL | lung | FRC_PRL | | FRC_PRL | | | 0.2395 | 0.08195 | 1146 | 1145 | 0.4816 | 0.03562 | 0.02472 | 0.02292 | 0.5063 | 0.05854 | 0.2186 | 0.2113 | 0.0389 | 0.03927 | 0.2203 |
| FRC_PRU | FRC PRU | lung | FRC_PRU | | FRC_PRU | | | 0.1792 | 0.1159 | 1142 | 1144 | 0.4219 | 0.1699 | 1.00e-06 | 0.03735 | 0.4219 | 0.2072 | 0.2234 | 0.2153 | 0.03559 | 0.04229 | 0.2243 |
| FRC_ZBD | FRC ZBD | lung | FRC_ZBD | | FRC_ZBD | | | -0.1671 | 0.2216 | 1145 | 1145 | 0.1409 | 0.3897 | 0.02311 | 1.00e-06 | 0.164 | 0.3897 | 0.2138 | 0.2057 | 0.04068 | 0.03761 | 0.2155 |
| FRONTAL AXIS INITIAL 40 MS | Frontal axis initial 40 ms | cardiovascular | | | FRONTAL.AXIS.INITIAL.40.MS | | | 0.2035 | 0.0318 | 7138 | 6253 | 0.04598 | 0.03653 | 1.00e-06 | 0.009039 | 0.04598 | 0.04557 | 0.03455 | 0.0392 | 0.00553 | 0.008347 | 0.03488 |
| FRONTAL AXIS MEAN QRS | QRS axis | cardiovascular | QRS axis | QRS axis | FRONTAL.AXIS.MEAN.QRS | | | -0.06858 | 0.3828 | 7218 | 6315 | 0.05616 | 0.02772 | 1.00e-06 | 8.13e-04 | 0.05616 | 0.02853 | 0.0354 | 0.03863 | 0.005379 | 0.006147 | 0.03588 |
| FRONTAL AXIS P | P wave axis | cardiovascular | P wave axis | P wave axis | FRONTAL.AXIS.P | | | 0.2207 | 0.002076 | 7040 | 6062 | 0.1242 | 0.006688 | 1.00e-06 | 1.00e-06 | 0.1242 | 0.006689 | 0.03758 | 0.0389 | 0.006995 | 0.007689 | 0.03816 |
| FRONTAL AXIS ST | ST segment axis | cardiovascular | ST segment axis.horizontal plane | ST segment axis.horizontal plane | FRONTAL.AXIS.ST | | | 0.111 | 0.3681 | 7193 | 6301 | 1.00e-06 | 1.00e-06 | 5.46e-04 | 0.001476 | 5.47e-04 | 0.001477 | 0.03554 | 0.03864 | 0.006187 | 0.007039 | 0.03597 |
| FRONTAL AXIS TERMINAL 40 MS | Frontal axis terminal 40 ms | cardiovascular | | | FRONTAL.AXIS.TERMINAL.40.MS | | | 0.08461 | 0.3168 | 7197 | 6296 | 0.04158 | 0.05612 | 1.00e-06 | 0.001193 | 0.04158 | 0.05732 | 0.0346 | 0.04038 | 0.006195 | 0.006384 | 0.03503 |
| FSH1 | Follitropin [Units/volume] in Serum or Plasma | endocrine | FOLLICLE STIMULATING HRM | Follitropin [Units/volume] in Serum or Plasma | FSH1 | 10*-3.[iU]/mL | | -0.2652 | 0.01941 | 3733 | 659 | 0.1732 | 0.9641 | 0.01618 | 0.03587 | 0.1894 | 1 | 0.07105 | 0.3533 | 0.01277 | 0.07178 | 0.07201 |
| FT4 | Thyroxine (T4) free [Mass/volume] in Serum or Plasma | endocrine | F T4 | Thyroxine (T4) free [Mass/volume] in Serum or Plasma | FT4 | ng/dL | | 0.09368 | 0.2646 | 17572 | 10236 | 0.02646 | 0.06222 | 0.002589 | 0.003123 | 0.02905 | 0.06534 | 0.01465 | 0.02497 | 0.002676 | 0.004694 | 0.01489 |
| FTo#Cl | Cells counted in body fluid | other | TOTAL # CELLS | Cells Counted Total [#] in Body fluid | FTo.Cl | | | -0.03535 | 0.7543 | 1347 | 1663 | 0.03623 | 0.1434 | 0.008431 | 1.00e-06 | 0.04466 | 0.1434 | 0.1837 | 0.1552 | 0.03062 | 0.0253 | 0.1862 |

| all_labs | full_name | group | concept_name | LOINC_NAME | heritability_names | selected_unit | sex_specific_lab | genetic.correlation | genetic.correlation.p | gcta.N_female | gcta.N_male | gcta.Variance_Autosomal_h2_female | gcta.Variance_Autosomal_h2_male | gcta.Variance_X_chromosome_h2_female | gcta.Variance_X_chromosome_h2_male | gcta.Variance_h2_female | gcta.Variance_h2_male | gcta.SE_Autosomal_h2_female | gcta.SE_Autosomal_h2_male | gcta.SE_X_chromosome_h2_female | gcta.SE_X_chromosome_h2_male | gcta.SE_h2_female |
|---|---|---|---|---|---|---|---|---|---|---|---|---|---|---|---|---|---|---|---|---|---|---|
| FVC_PPR | FVC PPR | lung | FVC_PPR | | FVC_PPR | % | | 0.07579 | 0.5339 | 1562 | 1214 | 0.2857 | 0.141 | 1.00e-06 | 0.02607 | 0.2857 | 0.1671 | 0.1588 | 0.206 | 0.02604 | 0.03553 | 0.1606 |
| FVC_PRD | FVC PRD | lung | FVC_PRD | | FVC_PRD | L | | 0.1377 | 0.2526 | 1562 | 1214 | 0.4136 | 0.3046 | 1.00e-06 | 0.03055 | 0.4136 | 0.3352 | 0.1583 | 0.2086 | 0.02734 | 0.04128 | 0.1608 |
| FVC_PRL | FVC PRL | lung | FVC_PRL | | FVC_PRL | | | 0.06755 | 0.3461 | 1736 | 1604 | 0.4766 | 0.6378 | 0.001721 | 1.00e-06 | 0.4783 | 0.6378 | 0.1478 | 0.1554 | 0.02198 | 0.02507 | 0.1492 |
| FVC_ZBD | FVC ZBD | lung | FVC_ZBD | Forced vital capacity [Volume] Respiratory system --pre bronchodilation | FVC_ZBD | L | | 0.06121 | 0.6175 | 1562 | 1214 | 0.1907 | 0.229 | 0.004302 | 0.02913 | 0.195 | 0.2582 | 0.1588 | 0.2022 | 0.02445 | 0.03669 | 0.1603 |
| FVC_ZPC | FVC ZPC | lung | FVC_ZPC | | FVC_ZPC | | | 0.01261 | 0.9534 | 550 | 485 | 0.7518 | 0.5914 | 0.03081 | 0.02211 | 0.7826 | 0.6135 | 0.4543 | 0.4894 | 0.06969 | 0.07361 | 0.4608 |
| GamGg_ | Gamma globulin in Serum or Plasma by electrophoresis | immune / inflammatory | GAMMA GLOBULIN | Gamma globulin [Mass/volume] in Serum or Plasma by Electrophoresis | GamGg_ | g/dL | | 0.08935 | 0.3277 | 3670 | 2890 | 0.1599 | 0.1482 | 0.006142 | 1.00e-06 | 0.166 | 0.1482 | 0.07014 | 0.08857 | 0.01119 | 0.01682 | 0.07064 |
| GGT | Gamma glutamyl transferase activity in Serum or Plasma | liver | GAMMA GLUT TRANS BLOOD | Gamma glutamyl transferase [Enzymatic activity/volume] in Serum or Plasma | GGT | [iU]/L | | 0.01087 | 0.9207 | 2363 | 2733 | 0.04698 | 1.00e-06 | 0.06266 | 0.0199 | 0.1096 | 0.0199 | 0.1006 | 0.09449 | 0.02557 | 0.01795 | 0.1031 |
| Glob | Globulin | metabolic | GLOBULIN | | Glob | g/dL | | -0.1285 | 0.3606 | 1060 | 715 | 0.2649 | 0.07729 | 0.01388 | 0.02831 | 0.2788 | 0.1056 | 0.2254 | 0.3485 | 0.03672 | 0.06644 | 0.2273 |
| GluBed | Glucose [Mass/volume] in Blood by Automated test strip | metabolic | BEDSIDE GLUCOSE | Glucose [Mass/volume] in Blood by Automated test strip | GluBed | mg/dL | | 0.4593 | 2.00e-05 | 13645 | 13466 | 0.03676 | 0.06623 | 1.00e-06 | 1.00e-06 | 0.03676 | 0.06623 | 0.01897 | 0.01952 | 0.003376 | 0.003454 | 0.01929 |
| Gluc | Glucose lab | metabolic | GLUCOSE BLOOD | Glucose [Mass/volume] in Serum or Plasma | Gluc | mg/dL | | 0.5594 | 2.33e-14 | 34448 | 26960 | 0.1144 | 0.1194 | 7.74e-04 | 0.001078 | 0.1151 | 0.1204 | 0.009031 | 0.01097 | 0.001398 | 0.001706 | 0.009135 |
| GluWB | Glucose [Mass/volume] in Blood | blood | GLUCOSE WHOLE BLOOD | Glucose [Mass/volume] in Blood | GluWB | mg/dL | | -0.01133 | 0.8973 | 6045 | 7936 | | 0.07712 | | 1.00e-06 | | 0.07712 | | 0.03228 | | 0.005507 | |
| GTT50g | Glucose tolerance 50g (GTT50g) | metabolic | GLUCOSE TOL 50G | | GTT50g | mg/dL | Sex specific lab | | | 2743 | | 0.1626 | | 0.01066 | | 0.1732 | | 0.09472 | | 0.01624 | | 0.09548 |
| Haptgl | Haptoglobin [Mass/volume] in Serum or Plasma | blood | HAPTOGLOBIN BLOOD | Haptoglobin [Mass/volume] in Serum or Plasma | Haptgl | mg/dL | | 0.1429 | 0.8334 | 1516 | 1550 | 0.5659 | 0.2664 | 0.008706 | 0.04293 | 0.5746 | 0.3093 | 0.1673 | 0.1648 | 0.02697 | 0.03538 | 0.1693 |
| HB_PRD | HB_PRD | lung | HB_PRD | | HB_PRD | | | -0.07251 | 0.546 | 1182 | 1215 | 0.1145 | 0.01267 | 1.00e-06 | 1.00e-06 | 0.1145 | 0.01267 | 0.211 | 0.2063 | 0.03971 | 0.03832 | 0.2148 |

| all_labs | full_name | group | concept_name | LOINC_NAME | heritability_names | selected_unit | sex_specific_lab | genetic.correlation | genetic.correlation.p | gcta.N_female | gcta.N_male | gcta.Variance_Autosomal_h2_female | gcta.Variance_Autosomal_h2_male | gcta.Variance_X_chromosome_h2_female | gcta.Variance_X_chromosome_h2_male | gcta.Variance_h2_female | gcta.Variance_h2_male | gcta.SE_Autosomal_h2_female | gcta.SE_Autosomal_h2_male | gcta.SE_X_chromosome_h2_female | gcta.SE_X_chromosome_h2_male | gcta.SE_h2_female |
|---|---|---|---|---|---|---|---|---|---|---|---|---|---|---|---|---|---|---|---|---|---|---|
| HBSA# | Hepatitis B virus surface Ab [Presence] in Serum | immune / inflammatory | HEP B SURFACE AB VALUE | Hepatitis B virus surface Ab [Presence] in Serum | HBSA. | 10*-3.[iU]/mL | | 0.05811 | 0.6143 | 2102 | 2001 | 0.1071 | 0.1114 | 1.00e-06 | 0.001078 | 0.1071 | 0.1125 | 0.1154 | 0.1233 | 0.02271 | 0.02053 | 0.1171 |
| HCO3Ca | Bicarbonate [Moles/volume] in Unspecified specimen | metabolic | BICARBONATE (CALC) | Bicarbonate [Moles/volume] in Venous blood | HCO3Ca | mmol/L | | -0.03506 | 0.7046 | 8404 | 9812 | 0.05831 | 0.01992 | 1.00e-06 | 0.005867 | 0.05831 | 0.02579 | 0.03001 | 0.02526 | 0.005837 | 0.004927 | 0.03059 |
| HCTWB | Hematocrit | blood | PCV BLOOD | Hematocrit [Volume Fraction] of Blood | HCTWB | % | | 0.09765 | 0.3955 | 5311 | 6990 | | 0.05984 | | 1.00e-06 | | 0.05984 | | 0.03588 | | 0.006184 | |
| HDL-C | HDL Cholesterol | metabolic | HDL CHOLESTEROL | Cholesterol in HDL [Mass/volume] in Serum or Plasma | HDL.C | mg/dL | | 0.572 | 2.86e-13 | 18580 | 14828 | 0.2335 | 0.172 | 1.00e-06 | 0.002331 | 0.2335 | 0.1743 | 0.01651 | 0.01962 | 0.001967 | 0.003206 | 0.01661 |
| HEART RATE | Heart rate | vitals | Heart rate | Heart rate | HEART.RATE | | | 0.07289 | 0.5684 | 7229 | 6318 | 0.06487 | 0.02106 | 0.001656 | 1.00e-06 | 0.06652 | 0.02106 | 0.03512 | 0.04006 | 0.006242 | 0.006941 | 0.03565 |
| Height | Height | vitals | Body height | Body height | Height | cm | | 0.7389 | 1.49e-51 | 34577 | 25698 | 0.366 | 0.3345 | 0.007248 | 0.01192 | 0.3733 | 0.3465 | 0.01019 | 0.01288 | 0.001824 | 0.002574 | 0.01029 |
| HEIGHT | Height | vitals | Body height | Body height | HEIGHT | cm | | 0.6233 | 1.58e-30 | 19052 | 12621 | 0.3696 | 0.3181 | 0.006146 | 0.01016 | 0.3758 | 0.3283 | 0.01635 | 0.0237 | 0.002915 | 0.004526 | 0.01654 |
| HEMATOCRIT | HEMATOCRIT | blood | HEMATOCRIT | | HEMATOCRIT | % | | -0.09599 | 0.5964 | 718 | 509 | | 0.2003 | | 1.00e-06 | | 0.2003 | | 0.4735 | | 0.07652 | |
| HEMOGLOBIN | HEMOGLOBIN | blood | HEMOGLOBIN | | HEMOGLOBIN | g/dL | | -0.2115 | 0.2132 | 719 | 509 | 1.00e-06 | 0.2107 | 0.04869 | 0.006259 | 0.04869 | 0.2169 | 0.3803 | 0.4688 | 0.06106 | 0.0773 | 0.3859 |
| Hgb | Hemoglobin | blood | HEMOGLOBIN BLOOD | Hemoglobin [Mass/volume] in Blood | Hgb | g/dL | | 0.3511 | 4.34e-07 | 35678 | 26968 | 0.07945 | 0.07503 | 0.00253 | 1.00e-06 | 0.08198 | 0.07503 | 0.008375 | 0.01045 | 0.001587 | 0.00168 | 0.008509 |
| HGB | Hemoglobin | blood | HEMOGLOBIN BLOOD | Hemoglobin [Mass/volume] in Blood | HGB | | | -0.03 | 0.8761 | 657 | 769 | 0.8445 | 0.3118 | 1.00e-06 | 1.00e-06 | 0.8445 | 0.3118 | 0.4038 | 0.3355 | 0.06563 | 0.05742 | 0.4081 |
| HgbA1C | Hemoglobin A1c (Glycated) Hemoglobin A1c (Glycated) | metabolic | HGB A1C GLYCATED | Hemoglobin A1c/Hemoglobin.total in Blood | HgbA1C | % | | 0.3467 | 1.05e-08 | 14409 | 11801 | 0.1794 | 0.158 | 1.00e-06 | 0.006432 | 0.1794 | 0.1645 | 0.01994 | 0.02327 | 0.00298 | 0.004679 | 0.02015 |
| HomocB | Homocysteine in Serum or Plasma | metabolic | HOMOCYST(E)INE ULTRAQUANT | Homocysteine [Moles/volume] in Serum or Plasma | HomocB | umol/L | | 0.1528 | 0.3076 | 829 | 538 | 0.6224 | | 0.0256 | | 0.648 | | 0.307 | | 0.05107 | | 0.3112 |
| HORIZONTAL AXIS: INITIAL 40 MS | QRS axis | cardiovascular | QRS axis | QRS axis | HORIZONTAL.AXIS..INITIAL.40.MS | | | 0.211 | 0.004133 | 7139 | 6239 | 0.1499 | 0.07963 | 1.00e-06 | 4.44e-04 | 0.1499 | 0.08008 | 0.03712 | 0.04165 | 0.006288 | 0.006795 | 0.03764 |
| HORIZONTALAXIS: TERMINAL 40 MS | QRS axis | cardiovascular | QRS axis | QRS axis | HORIZONTAL.AXIS..TERMINAL.40.MS | | | 0.03657 | 0.7652 | 7074 | 6205 | 0.02914 | 1.00e-06 | 9.11e-04 | 0.01379 | 0.03005 | 0.01379 | 0.03467 | 0.04055 | 0.006595 | 0.009049 | 0.03522 |
| HPIndx | H. pylori EIA index | immune / inflammatory | H PYLORI EIA INDEX | | HPIndx | | | 0.3558 | 0.1492 | 676 | 369 | 0.6299 | 0.2568 | 1.00e-06 | 0.2704 | 0.6299 | 0.5272 | 0.3358 | 0.6539 | 0.06634 | 0.1381 | 0.3416 |
| HR_INTERVAL | Heart rate | vitals | Heart rate | Heart rate | HR_INTERVAL | | | 0.04574 | 0.6432 | 21462 | 19279 | | 0.004569 | | 0.002943 | | 0.007512 | | 0.002262 | | 0.002444 | |

| all_labs | full_name | group | concept_name | LOINC_NAME | heritability_names | selected_unit | sex_specific_lab | genetic.correlation | genetic.correlation.p | gcta.N_female | gcta.N_male | gcta.Variance_Autosomal_h2_female | gcta.Variance_Autosomal_h2_male | gcta.Variance_X_chromosome_h2_female | gcta.Variance_X_chromosome_h2_male | gcta.Variance_h2_female | gcta.Variance_h2_male | gcta.SE_Autosomal_h2_female | gcta.SE_Autosomal_h2_male | gcta.SE_X_chromosome_h2_female | gcta.SE_X_chromosome_h2_male | gcta.SE_h2_female |
|---|---|---|---|---|---|---|---|---|---|---|---|---|---|---|---|---|---|---|---|---|---|---|
| HSCRP | C reactive protein (high sensitivity) | immune / inflammatory | hs-CRP | C reactive protein [Mass/volume] in Serum or Plasma by High sensitivity method | HSCRP | mg/L | | -0.1258 | 0.6039 | 555 | 585 | 0.5718 | | 0.09618 | | 0.6679 | | 0.3867 | | 0.08734 | | 0.3928 |
| IC_PPR | IC PPR | lung | IC_PPR | | IC_PPR | | | 0.0905 | 0.4759 | 1168 | 1168 | 1.00e-06 | 0.1732 | 0.003286 | 1.00e-06 | 0.003287 | 0.1732 | 0.2058 | 0.2124 | 0.03793 | 0.03826 | 0.2101 |
| IC_PRD | IC PRD | lung | IC_PRD | | IC_PRD | | | -0.01854 | 0.8647 | 1168 | 1167 | 0.3328 | 0.01658 | 0.03234 | 1.00e-06 | 0.3651 | 0.01658 | 0.2181 | 0.2072 | 0.03938 | 0.03981 | 0.22 |
| IC_ZBD | IC ZBD | lung | IC_ZBD | | IC_ZBD | | | -0.003825 | 0.976 | 1167 | 1168 | 0.004162 | 0.2605 | 0.03106 | 1.00e-06 | 0.03522 | 0.2605 | 0.2105 | 0.2149 | 0.04208 | 0.03821 | 0.2149 |
| IctIdx | Icteric index of Serum or Plasma | metabolic | ICTERUS INDEX | Icteric index of Serum or Plasma | IctIdx | | | 0.2461 | 0.01789 | 2408 | 1967 | 0.06957 | 0.09529 | 0.006125 | 1.00e-06 | 0.0757 | 0.09529 | 0.09921 | 0.13 | 0.0201 | 0.02341 | 0.1014 |
| IgA | IgA [Mass/volume] in Serum or Plasma | immune / inflammatory | IGA QUANTITATIVE BLOOD | IgA [Mass/volume] in Serum or Plasma | IgA | mg/dL | | 0.3356 | 5.43e-04 | 4327 | 1980 | 0.09325 | 0.08898 | 0.00541 | 1.00e-06 | 0.09866 | 0.08898 | 0.05729 | 0.1207 | 0.01023 | 0.02283 | 0.05819 |
| IGAB | Immature granulocytes [Presence] in Blood by Automated Count | immune / inflammatory | IMM GRANULOCYTES (ABS) | Immature granulocytes [Presence] in Blood by Automated count | IGAB | 10*3/uL | | 0.4159 | 2.59e-06 | 24902 | 19254 | 6.65e-04 | 0.06638 | 1.00e-06 | 0.0028 | 6.66e-04 | 0.06918 | 0.001363 | 0.01366 | 0.001738 | 0.002492 | 0.002207 |
| IgEQnt | IgE [Mass/volume] in Serum | immune / inflammatory | IGE QUANTITATIVE BLOOD | IgE [Mass/volume] in Serum | IgEQnt | [iU]/mL | | 0.1168 | 0.3972 | 1008 | 557 | | 0.9041 | | 0.002934 | | 0.907 | | 0.411 | | 0.07999 | |
| IgG-Q | IgG Quantitative blood | immune / inflammatory | IGG QUANTITATIVE BLOOD | IgG [Mass/volume] in Serum or Plasma | IgG.Q | mg/dL | | 0.1155 | 0.1432 | 2071 | 1520 | 0.1976 | | 0.02346 | | 0.2211 | | 0.1231 | | 0.02276 | | 0.1251 |
| IgM-Q | IgM Quantitative blood | immune / inflammatory | IGM QUANTITATIVE BLOOD | IgM [Mass/volume] in Serum or Plasma | IgM.Q | mg/dL | | 0.3187 | 0.359 | 1272 | 902 | 0.3419 | 0.3613 | 0.0566 | 0.006353 | 0.3985 | 0.3676 | 0.2006 | 0.2837 | 0.03907 | 0.05222 | 0.2043 |
| IGRE | Immature granulocytes/100 leukocytes in Blood | immune / inflammatory | IMM GRANULOCYTES % | Immature granulocytes/100 leukocytes in Blood | IGRE | % | | 0.3376 | 0.003332 | 24933 | 19256 | 0.001549 | 0.04809 | 8.10e-05 | 0.005555 | 0.00163 | 0.05365 | 0.001472 | 0.01348 | 0.00189 | 0.002888 | 0.002396 |
| Infant Apgar 5 M Hr 1 | 5 minute Apgar heart rate | vitals | 5 minute Apgar Heart rate | 5 minute Apgar Heart rate | Infant.Apgar.5.M.Hr.1 | | Sex specific lab | | | 1596 | | 0.3215 | | 0.005935 | | 0.3274 | | 0.1569 | | 0.02602 | | 0.1582 |
| Infant Apgar 5 M Hr 4 | 5 minute Apgar heart rate | vitals | 5 minute Apgar Heart rate | 5 minute Apgar Heart rate | Infant.Apgar.5.M.Hr.4 | | Sex specific lab | | | 1596 | | 0.3215 | | 0.005935 | | 0.3274 | | 0.1569 | | 0.02602 | | 0.1582 |
| Infant Apgar Color 1 M 1 | 1 minute Apgar color | vitals | 1 minute Apgar Color | 1 minute Apgar Color | Infant.Apgar.Color.1.M.1 | | Sex specific lab | | | 1624 | | 1.00e-06 | | 0.03247 | | 0.03247 | | 0.158 | | 0.0288 | | 0.1599 |
| Infant Apgar Color 5 M 1 | 5 minute Apgar color | vitals | 5 minute Apgar Color | 5 minute Apgar Color | Infant.Apgar.Color.5.M.1 | | Sex specific lab | | | 1622 | | | | | | | | | | | | |

| all_labs | full_name | group | concept_name | LOINC_NAME | heritability_names | selected_unit | sex_specific_lab | genetic.correlation | genetic.correlation.p | gcta.N_female | gcta.N_male | gcta.Variance_Autosomal_h2_female | gcta.Variance_Autosomal_h2_male | gcta.Variance_X_chromosome_h2_female | gcta.Variance_X_chromosome_h2_male | gcta.Variance_h2_female | gcta.Variance_h2_male | gcta.SE_Autosomal_h2_female | gcta.SE_Autosomal_h2_male | gcta.SE_X_chromosome_h2_female | gcta.SE_X_chromosome_h2_male | gcta.SE_h2_female |
|---|---|---|---|---|---|---|---|---|---|---|---|---|---|---|---|---|---|---|---|---|---|---|
| Infant Apgar Muscle 1 M 1 | 1 minute Apgar muscle | vitals | 1 minute Apgar Muscle tone | 1 minute Apgar Muscle tone | Infant.Apgar.Muscle.1.M.1 | | Sex specific lab | | | 1628 | | 0.1034 | | 1.00e-06 | | 0.1034 | | 0.1538 | | 0.02755 | | 0.156 |
| Infant Apgar Muscle 5 M 1 | 5 minute Apgar muscle | vitals | 5 minute Apgar Muscle tone | 5 minute Apgar Muscle tone | Infant.Apgar.Muscle.5.M.1 | | Sex specific lab | | | 1588 | | 0.02778 | | 1.00e-06 | | 0.02778 | | 0.1515 | | 0.02811 | | 0.154 |
| Infant Apgar Resp 1 M 1 | 1 minute Apgar respiratory rate | vitals | 1 minute Apgar Respiratory effort | 1 minute Apgar Respiratory effort | Infant.Apgar.Resp.1.M.1 | | Sex specific lab | | | 1629 | | 0.00738 | | 1.00e-06 | | 0.007381 | | 0.1543 | | 0.02835 | | 0.1566 |
| Infant Apgar Resp 5 M 1 | 5 minute Apgar respiratory rate | vitals | 5 minute Apgar Respiratory effort | 5 minute Apgar Respiratory effort | Infant.Apgar.Resp.5.M.1 | | Sex specific lab | | | 1595 | | 0.192 | | 0.003893 | | 0.1959 | | 0.1615 | | 0.02534 | | 0.1631 |
| Infant Apgar Rflx 1 M 1 | 1 minute Apgar reflex | vitals | 1 minute Apgar Reflex irritability | 1 minute Apgar Reflex irritability | Infant.Apgar.Rflx.1.M.1 | | Sex specific lab | | | 1630 | | 0.007483 | | 1.00e-06 | | 0.007484 | | 0.1579 | | 0.02955 | | 0.1601 |
| Infant Apgar Rflx 5 M 1 | 5 minute Apgar reflex | vitals | 5 minute Apgar Reflex irritability | 5 minute Apgar Reflex irritability | Infant.Apgar.Rflx.5.M.1 | | Sex specific lab | | | 1593 | | 0.3458 | | 1.00e-06 | | 0.3458 | | 0.1552 | | 0.02522 | | 0.1575 |
| Infant Apgar Score 1 M 1 | 1 minute Apgar score | vitals | 1 minute Apgar Score | 1 minute Apgar Score | Infant.Apgar.Score.1.M.1 | | Sex specific lab | | | 1619 | | | | | | | | | | | | |
| Infant Apgar Score 5 M 1 | 5 minute Apgar score | vitals | 5 minute Apgar Score | 5 minute Apgar Score | Infant.Apgar.Score.5.M.1 | | Sex specific lab | | | 1594 | | | | | | | | | | | | |
| INITIAL_40_MS | Initial 40 ms | cardiovascular | | | INITIAL_40_MS | | | 0.3851 | 6.77e-08 | 21438 | 19244 | 8.80e-04 | 0.001668 | 0.001689 | 0.002537 | 0.00257 | 0.004205 | 0.001608 | 0.001832 | 0.00226 | 0.002636 | 0.002769 |
| Insuln | Insulin [Units/volume] in Serum or Plasma | endocrine | INSULIN | Insulin [Units/volume] in Serum or Plasma | Insuln | | | 0.4055 | 0.06408 | 1372 | 331 | 0.02332 | 0.4784 | 1.00e-06 | 1.00e-06 | 0.02332 | 0.4784 | 0.1772 | 0.7709 | 0.02861 | 0.1398 | 0.1793 |
| IPF | Platelets reticulated/100 platelets in Blood by Automated count | blood | IMM PLATELET FRACTION | Platelets reticulated/100 platelets in Blood by Automated count | IPF | % | | 0.04316 | 0.6421 | 2403 | 2273 | 0.1139 | 1.00e-06 | 0.006869 | 0.02496 | 0.1208 | 0.02496 | 0.1004 | 0.1096 | 0.01942 | 0.02 | 0.1022 |
| IRF | Immature reticulocytes/Reticulocytes.total in Blood | blood | IMMATURE RETIC FRACTION | Immature reticulocytes/Reticulocytes.total in Blood | IRF | % | | 0.2286 | 0.03311 | 3085 | 2822 | 0.01734 | 0.1263 | 0.02375 | 1.00e-06 | 0.04109 | 0.1263 | 0.07802 | 0.09134 | 0.01791 | 0.01539 | 0.08039 |
| Iron | Iron [Mass/volume] in Serum or Plasma | blood | IRON BLOOD | Iron [Mass/volume] in Serum or Plasma | Iron | ug/dL | | 0.166 | 0.1076 | 10737 | 7897 | 0.06587 | 0.04577 | 1.00e-06 | 5.16e-04 | 0.06587 | 0.04629 | 0.02384 | 0.03155 | 0.004051 | 0.005294 | 0.02411 |
| K-WB | Potassium whole blood | metabolic | POTASSIUM WHOLE BLOOD | Potassium [Moles/volume] in Blood | K.WB | mmol/L | | 0.08361 | 0.4093 | 6320 | 8267 | 0.04827 | 0.007842 | 1.00e-06 | 1.00e-06 | 0.04828 | 0.007843 | 0.03997 | 0.03025 | 0.007271 | 0.005983 | 0.04061 |
| K | Potassium blood | metabolic | POTASSIUM BLOOD | Potassium [Moles/volume] in Serum or Plasma | K | mmol/L | | 0.3142 | 4.32e-05 | 34362 | 26975 | 0.05538 | 0.05862 | 1.27e-04 | 0.002054 | 0.0555 | 0.06067 | 0.008328 | 0.01022 | 0.00142 | 0.00188 | 0.008447 |

| all_labs | full_name | group | concept_name | LOINC_NAME | heritability_names | selected_unit | sex_specific_lab | genetic.correlation | genetic.correlation.p | gcta.N_female | gcta.N_male | gcta.Variance_Autosomal_h2_female | gcta.Variance_Autosomal_h2_male | gcta.Variance_X_chromosome_h2_female | gcta.Variance_X_chromosome_h2_male | gcta.Variance_h2_female | gcta.Variance_h2_male | gcta.SE_Autosomal_h2_female | gcta.SE_Autosomal_h2_male | gcta.SE_X_chromosome_h2_female | gcta.SE_X_chromosome_h2_male | gcta.SE_h2_female |
|---|---|---|---|---|---|---|---|---|---|---|---|---|---|---|---|---|---|---|---|---|---|---|
| KapLCQ | Kappa light chains.free [Mass/volume] in Serum | toxicology / pharmacology | KAPPA LIGHT CHAIN QUANT | Kappa light chains.free [ Mass/volume ] in Serum | KapLCQ | mg/dL | | 0.1638 | 0.2371 | 973 | 640 | 0.4709 | | 0.02368 | | 0.4946 | | 0.2567 | | 0.04078 | | 0.2593 |
| KLRat | Kappa light chains.free/ Lambda light chains.free [Mass Ratio] in Serum | immune / inflammatory | KAPPA/LAMBDA RATIO | Kappa light chains.free/Lambda light chains.free [Mass Ratio] in Serum | KLRat | | | 0.1044 | 0.8504 | 949 | 617 | 0.4549 | 0.9958 | 0.05737 | 1.00e-06 | 0.5123 | 0.9958 | 0.2626 | 0.3893 | 0.04829 | 0.07867 | 0.2689 |
| LA | Lactate [Moles/volume] in Serum or Plasma | metabolic | LACTIC ACID VENOUS | Lactate [Moles/volume] in Serum or Plasma | LA | 10*-3.eq/L | | -0.1187 | 0.361 | 6190 | 6279 | 1.00e-06 | 0.02623 | 0.01705 | 1.00e-06 | 0.01705 | 0.02623 | 0.03957 | 0.03896 | 0.009296 | 0.007452 | 0.04057 |
| LAC | Lactate Whole Blood | metabolic | LACTATE WHOLE BLOOD | Lactate [Moles/volume] in Blood | LAC | mmol/L | | 0.1879 | 0.02261 | 6477 | 8287 | 0.02365 | 0.02113 | 1.00e-06 | 1.00e-06 | 0.02365 | 0.02113 | 0.03594 | 0.0307 | 0.006751 | 0.006078 | 0.03663 |
| all_labs | full_name | group | concept_name | LOINC_NAME | heritability_names | selected_unit | sex_specific_lab | genetic.correlation | genetic.correlation.p | gcta.N_female | gcta.N_male | gcta.Variance_Autosomal_h2_female | gcta.Variance_Autosomal_h2_male | gcta.Variance_X_chromosome_h2_female | gcta.Variance_X_chromosome_h2_male | gcta.Variance_h2_female | gcta.Variance_h2_male | gcta.SE_Autosomal_h2_female | gcta.SE_Autosomal_h2_male | gcta.SE_X_chromosome_h2_female | gcta.SE_X_chromosome_h2_male | gcta.SE_h2_female |
| LamLCQ | Lambda light chains.free [Mass/volume] in Serum or Plasma | immune / inflammatory | LAMBDA LIGHT CHAIN QUANT | Lambda light chains.free [ Mass/volume ] in Serum or Plasma | LamLCQ | mg/dL | | -0.04074 | 0.8421 | 971 | 639 | 0.3967 | 0.1081 | 1.00e-06 | 1.00e-06 | 0.3967 | 0.1081 | 0.2637 | 0.3669 | 0.04146 | 0.07624 | 0.2675 |
| LDH | Lactate dehydrogenase (LDH) | metabolic | LDH BLOOD | | LDH | [iU]/L | | 0.1622 | 0.1515 | 6376 | 5881 | | | | | | | | | | | |
| LDL_CALCULATED | LDL_CALCULATED | metabolic | LDL_CALCULATED | | LDL_CALCULATED | mg/dL | | 0.03532 | 0.7969 | 806 | 700 | | 0.4175 | | 0.06322 | | 0.4807 | | 0.3441 | | 0.0678 | |
| LDL_HDL_RATIO | Cholesterol.total/Cholesterol in HDL [Mass Ratio] in Serum or Plasma | metabolic | LDL/HDL_RATIO | Cholesterol.total/Cholesterol in HDL [Mass Ratio] in Serum or Plasma | LDL_HDL_RATIO | | | 0.193 | 0.3629 | 1151 | 1029 | 0.0977 | 0.4757 | 1.00e-06 | 0.00702 | 0.0977 | 0.4827 | 0.1998 | 0.2409 | 0.03682 | 0.04136 | 0.2038 |
| LDL-C | LDL cholesterol | metabolic | LDL CHOLESTEROL CALC | Cholesterol in LDL [Mass/volume] in Serum or Plasma by calculation | LDL.C | mg/dL | | 0.1574 | 0.1438 | 18095 | 14481 | 0.09522 | 0.1003 | 0.002312 | 0.001305 | 0.09753 | 0.1016 | 0.01544 | 0.01888 | 0.002794 | 0.003524 | 0.01563 |
| LEUKOREDUCED_RANDOM_PLATELETS | LEUKOREDUCED_RANDOM_PLATELETS | immune / inflammatory | LEUKOREDUCED_RANDOM_PLATELETS | | LEUKOREDUCED_RANDOM_PLATELETS | 10*3/uL | | -0.03298 | 0.7635 | 2351 | 1533 | 0.07477 | 1.00e-06 | 0.01171 | 0.0315 | 0.08648 | 0.0315 | 0.1035 | 0.1813 | 0.01961 | 0.03485 | 0.1041 |
| LH1 | Lutropin [Units/volume] in Serum or Plasma | endocrine | LUTEINIZING HOROMONE | Lutropin [Units/volume] in Serum or Plasma | LH1 | 10*-3.[iU]/mL | | -0.04186 | 0.7299 | 1594 | 861 | 0.1223 | 0.1856 | 1.00e-06 | 0.0263 | 0.1223 | 0.2119 | 0.1537 | 0.3075 | 0.02382 | 0.04941 | 0.1564 |

| all_labs | full_name | group | concept_name | LOINC_NAME | heritability_names | selected_unit | sex_specific_lab | genetic.correlation | genetic.correlation.p | gcta.N_female | gcta.N_male | gcta.Variance_Autosomal_h2_female | gcta.Variance_Autosomal_h2_male | gcta.Variance_X_chromosome_h2_female | gcta.Variance_X_chromosome_h2_male | gcta.Variance_h2_female | gcta.Variance_h2_male | gcta.SE_Autosomal_h2_female | gcta.SE_Autosomal_h2_male | gcta.SE_X_chromosome_h2_female | gcta.SE_X_chromosome_h2_male | gcta.SE_h2_female |
|---|---|---|---|---|---|---|---|---|---|---|---|---|---|---|---|---|---|---|---|---|---|---|
| Lipase | Lipase | toxicology / pharmacology | LIPASE BLOOD | Lipase [Enzymatic activity/volume] in Serum or Plasma | Lipase | [iU]/L | | -0.002581 | 0.9764 | 6454 | 5058 | 0.08433 | 0.1425 | 0.008829 | 1.00e-06 | 0.09316 | 0.1425 | 0.03838 | 0.05097 | 0.00756 | 0.00933 | 0.0391 |
| LipIdx | Lipemic index of Serum or Plasma | metabolic | LIPEMIA INDEX | Lipemic index of Serum or Plasma | LipIdx | | | -0.03745 | 0.755 | 2762 | 2114 | 1.00e-06 | 0.01198 | 0.001019 | 8.21e-04 | 0.00102 | 0.0128 | 0.09055 | 0.1076 | 0.01721 | 0.02032 | 0.092 |
| Luc% | Luc% | immune / inflammatory | OTHER CELLS % | Other cells/100 leukocytes in Blood by Manual count | Luc_percent_ | % | | 0.08388 | 0.3529 | 3541 | 2510 | 0.1134 | | 1.00e-06 | | 0.1134 | | 0.06785 | | 0.01372 | | 0.06882 |
| LucAbs | Leukocytes [#/volume] in Blood by Automated count | blood | OTHER CELLS (ABS) | Leukocytes [#/volume] in Blood by Automated count | LucAbs | 10*3/uL | | 0.07815 | 0.4945 | 3547 | 2514 | 0.05433 | | 0.02031 | | 0.07464 | | 0.06714 | | 0.01488 | | 0.06833 |
| Lym | Lymphocytes [#/volume] in Blood | immune / inflammatory | LYMPHS % | Lymphocytes [#/volume] in Blood | Lym | % | | 0.2888 | 4.51e-04 | 15914 | 11637 | 0.06732 | 0.06707 | 0.001368 | 1.00e-06 | 0.06868 | 0.06707 | 0.01692 | 0.02207 | 0.002999 | 0.003891 | 0.01718 |
| LymAbs | Lymphocytes [#/volume] in Blood by Automated count | immune / inflammatory | LYMPHS (ABS) | Lymphocytes [#/volume] in Blood by Automated count | LymAbs | 10*3/uL | | 0.2569 | 3.81e-04 | 15127 | 11025 | 0.1081 | 0.07039 | 1.00e-06 | 1.00e-06 | 0.1081 | 0.07039 | 0.01797 | 0.02343 | 0.00346 | 0.003904 | 0.01826 |
| LYMPA | Lymphocytes [#/volume] in Blood | immune / inflammatory | LYMPHS (ABS) | Lymphocytes [#/volume] in Blood | LYMPA | 10*3/uL | | 0.05299 | 0.6037 | 5177 | 5327 | 0.02876 | 0.005148 | 1.00e-06 | 0.001366 | 0.02877 | 0.006514 | 0.04608 | 0.0419 | 0.009713 | 0.008971 | 0.04712 |
| LYMPAB | Lymphocytes [#/volume] in Blood by Automated count | immune / inflammatory | LYMPHS (ABS) | Lymphocytes [#/volume] in Blood by Automated count | LYMPAB | 10*3/uL | | 0.2556 | 0.001462 | 25616 | 19927 | 0.002403 | 0.00212 | 1.00e-06 | 9.89e-04 | 0.002404 | 0.003109 | 0.001581 | 0.001908 | 0.00174 | 0.00219 | 0.002346 |
| LYMPHOCYTES | LYMPHOCYTES | immune / inflammatory | LYMPHOCYTES | | LYMPHOCYTES | % | | 0.02743 | 0.8307 | 1237 | 795 | | | | | | | | | | | |
| LYMPM | Lymphocytes/100 leukocytes in Blood by Manual count | immune / inflammatory | LYMPS % | Lymphocytes/100 leukocytes in Blood by Manual count | LYMPM | % | | -0.1307 | 0.1405 | 5362 | 5485 | | | | | | | | | | | |
| LYMPRE | Lymphocytes/100 leukocytes in Blood by Automated count | immune / inflammatory | LYMPHS % | Lymphocytes/100 leukocytes in Blood | LYMPRE | % | | 0.2311 | 2.83e-04 | 25619 | 19940 | 0.002736 | 0.00113 | 1.00e-06 | 1.00e-06 | 0.002737 | 0.001131 | 0.001611 | 0.00174 | 0.001647 | 0.001971 | 0.002297 |
| MBRat | Creatine kinase.MB/Creatine kinase.total in Serum or Plasma | cardiovascular | CK-MB RATIO | Creatine kinase.MB/Creatine kinase.total in Serum or Plasma | MBRat | | | 0.224 | 0.01171 | 6477 | 7881 | 0.07989 | 0.12 | 1.00e-06 | 1.00e-06 | 0.07989 | 0.12 | 0.03858 | 0.03308 | 0.006476 | 0.005895 | 0.03902 |

| all_labs | full_name | group | concept_name | LOINC_NAME | heritability_names | selected_unit | sex_specific_lab | genetic.correlation | genetic.correlation.p | gcta.N_female | gcta.N_male | gcta.Variance_Autosomal_h2_female | gcta.Variance_Autosomal_h2_male | gcta.Variance_X_chromosome_h2_female | gcta.Variance_X_chromosome_h2_male | gcta.Variance_h2_female | gcta.Variance_h2_male | gcta.SE_Autosomal_h2_female | gcta.SE_Autosomal_h2_male | gcta.SE_X_chromosome_h2_female | gcta.SE_X_chromosome_h2_male | gcta.SE_h2_female |
|---|---|---|---|---|---|---|---|---|---|---|---|---|---|---|---|---|---|---|---|---|---|---|
| MCH | MCH [Entitic mass] by Automated count | blood | MCH | MCH [Entitic mass] by Automated count | MCH | pg | | 0.5951 | 3.95e-18 | 35668 | 26968 | 0.1806 | 0.1682 | 3.22e-04 | 0.002399 | 0.181 | 0.1706 | 0.009321 | 0.01157 | 0.001128 | 0.001895 | 0.009384 |
| MCHC | MCHC [Mass/volume] by Automated count | blood | MCHC | MCHC [Mass/volume] by Automated count | MCHC | g/dL | | 0.3909 | 2.05e-11 | 35644 | 26945 | 0.1385 | 0.1278 | 0.002656 | 1.00e-06 | 0.1412 | 0.1278 | 0.00888 | 0.01114 | 0.001551 | 0.001739 | 0.008994 |
| MCV | MCV [Entitic volume] by Automated count | blood | MCV | MCV [Entitic volume] by Automated count | MCV | fL | | 0.3249 | 6.38e-04 | 35676 | 26971 | 0.02447 | 0.05712 | 1.00e-06 | 4.85e-04 | 0.02447 | 0.05761 | 0.007477 | 0.01038 | 0.001271 | 0.001834 | 0.00757 |
| MDALab | Atypical lymphocytes | immune / inflammatory | ATYP LYMPHS (ABS) | Variant lymphocytes [#/volume] in Blood | MDALab | | | 0.317 | 0.003513 | 3337 | 3552 | 0.0277 | 0.03555 | 1.00e-06 | 1.00e-06 | 0.0277 | 0.03555 | 0.07069 | 0.07011 | 0.01499 | 0.01373 | 0.07206 |
| MDBlab | Blasts in Blood | immune / inflammatory | BLASTS (ABS) | Blasts [#/volume] in Blood | MDBlab | | | -0.0496 | 0.7742 | 743 | 847 | 0.5723 | 1.00e-06 | 1.00e-06 | 0.02531 | 0.5723 | 0.02531 | 0.3643 | 0.3113 | 0.05083 | 0.05632 | 0.3701 |
| MDMMab | Metamyelocytes (absolute) | immune / inflammatory | METAMYELOCYTES (ABS) | Metamyelocytes [#/volume] in Blood by Manual count | MDMMab | 10*3/uL | | 0.1962 | 0.03493 | 3474 | 3619 | 0.06625 | 0.05906 | 0.006842 | 1.00e-06 | 0.07309 | 0.05906 | 0.07133 | 0.06971 | 0.01343 | 0.01327 | 0.07254 |
| MDMyab | Myelocytes (absolute) | immune / inflammatory | MYELOCYTES (ABS) | Myelocytes [#/volume] in Blood | MDMyab | 10*3/uL | | 0.1757 | 0.03355 | 3632 | 3884 | 0.08927 | 0.1459 | 1.00e-06 | 1.00e-06 | 0.08927 | 0.1459 | 0.07016 | 0.06547 | 0.01362 | 0.01158 | 0.07116 |
| MDPmab | Promyelocytes (absolute) | immune / inflammatory | PROMYELOCYTES (ABS) | Promyelocytes [#/volume] in Blood by Manual count | MDPmab | | | 0.02127 | 0.8402 | 1364 | 1490 | 1.00e-06 | 0.05166 | 0.01488 | 1.00e-06 | 0.01488 | 0.05167 | 0.1917 | 0.1641 | 0.0326 | 0.03327 | 0.1944 |
| MEAN_QRS | QRS axis | cardiovascular | QRS axis | QRS axis | MEAN_QRS | deg | | 0.3783 | 3.95e-08 | 21442 | 19258 | 0.001479 | 0.002961 | 0.002741 | 0.002395 | 0.00422 | 0.005356 | 0.001672 | 0.00198 | 0.002306 | 0.002419 | 0.002841 |
| Meta | Metamyelocytes/100 leukocytes in Blood by Manual count | blood | METAMYELO % | Metamyelocytes/100 leukocytes in Blood by Manual count | Meta | % | | -0.006391 | 0.9581 | 3623 | 3764 | 1.00e-06 | 1.00e-06 | 0.001929 | 0.001381 | 0.00193 | 0.001382 | 0.06671 | 0.06393 | 0.01263 | 0.01332 | 0.06788 |
| MetHgb | Methemoglobin/Hemoglobin.total in Venous blood | blood | METHEMOGLOBIN | Methemoglobin/Hemoglobin.total in Venous blood | MetHgb | % | | 0.003665 | 0.9734 | 2965 | 4068 | 0.04653 | 0.02654 | 0.02704 | 0.006454 | 0.07357 | 0.033 | 0.08501 | 0.05744 | 0.01869 | 0.01004 | 0.08675 |
| Mg | Magnesium [Mass/volume] in Serum or Plasma | metabolic | MAGNESIUM BLOOD | Magnesium [Mass/volume] in Serum or Plasma | Mg | mg/dL | | 0.2871 | 0.005472 | 12679 | 11442 | 0.06068 | 0.02295 | 1.00e-06 | 0.003002 | 0.06068 | 0.02595 | 0.02008 | 0.02199 | 0.003515 | 0.004267 | 0.02037 |
| MmAcid | Methylmalonic acid | metabolic | METHYLMALONIC ACID | Methylmalonate [Moles/volume] in Serum or Plasma | MmAcid | umol/L | | -0.09088 | 0.8499 | 902 | 689 | 0.3188 | 0.388 | 1.00e-06 | 1.00e-06 | 0.3188 | 0.388 | 0.2893 | 0.361 | 0.04095 | 0.06947 | 0.2933 |

| all_labs | full_name | group | concept_name | LOINC_NAME | heritability_names | selected_unit | sex_specific_lab | genetic.correlation | genetic.correlation.p | gcta.N_female | gcta.N_male | gcta.Variance_Autosomal_h2_female | gcta.Variance_Autosomal_h2_male | gcta.Variance_X_chromosome_h2_female | gcta.Variance_X_chromosome_h2_male | gcta.Variance_h2_female | gcta.Variance_h2_male | gcta.SE_Autosomal_h2_female | gcta.SE_Autosomal_h2_male | gcta.SE_X_chromosome_h2_female | gcta.SE_X_chromosome_h2_male | gcta.SE_h2_female |
|---|---|---|---|---|---|---|---|---|---|---|---|---|---|---|---|---|---|---|---|---|---|---|
| MonAbs | Absolute monocytes | immune / inflammatory | MONO (ABS) | | MonAbs | 10*3/uL | | 0.1756 | 0.01278 | 15125 | 11006 | 0.1201 | 0.1142 | 8.53e-04 | 1.00e-06 | 0.1209 | 0.1142 | 0.01824 | 0.02412 | 0.002951 | 0.004449 | 0.01844 |
| MONOA | Monocytes [#/volume] in Blood | immune / inflammatory | MONO (ABS) | Monocytes [#/volume] in Blood | MONOA | 10*3/uL | | -0.01317 | 0.8908 | 5055 | 5259 | 0.02955 | 1.00e-06 | 0.002634 | 0.003195 | 0.03218 | 0.003196 | 0.04847 | 0.04774 | 0.009059 | 0.008402 | 0.04905 |
| MONOAB | Monocytes [#/volume] in Blood by Automated count | immune / inflammatory | MONO (ABS) | Monocytes [#/volume] in Blood by Automated count | MONOAB | 10*3/uL | | 0.4792 | 1.30e-08 | 25647 | 19931 | 0.002642 | 0.00339 | 0.00168 | 0.004086 | 0.004322 | 0.007476 | 0.001728 | 0.002323 | 0.001952 | 0.002577 | 0.002612 |
| Monocy | Monocytes [#/volume] in Blood by Manual count | immune / inflammatory | MONOCYTES % | Monocytes [#/volume] in Blood by Manual count | Monocy | % | | 0.1686 | 0.007845 | 15947 | 11630 | 0.1158 | 0.09369 | 0.001612 | 9.30e-05 | 0.1174 | 0.09378 | 0.01766 | 0.02331 | 0.003118 | 0.003397 | 0.0179 |
| MONOCYTES | MONOCYTES | immune / inflammatory | MONOCYTES | | MONOCYTES | % | | 0.0842 | 0.5351 | 1238 | 794 | 0.07333 | | 0.01886 | | 0.09219 | | 0.1989 | | 0.03401 | | 0.1998 |
| MONOM | Monocytes/100 leukocytes in Blood by Manual count | immune / inflammatory | MONOCYTES % | Monocytes/100 leukocytes in Blood by Manual count | MONOM | % | | -0.024 | 0.8326 | 5096 | 5320 | 0.02532 | 1.00e-06 | 1.00e-06 | 0.004202 | 0.02532 | 0.004203 | 0.04781 | 0.04599 | 0.009473 | 0.008985 | 0.04862 |
| MONORE | Monocytes/100 leukocytes in Blood by Automated count | immune / inflammatory | MONOCYTES % | Monocytes/100 leukocytes in Blood by Automated count | MONORE | % | | 0.6025 | 9.90e-15 | 25672 | 19994 | 0.002384 | 0.005748 | 0.003912 | 0.004789 | 0.006297 | 0.01054 | 0.001671 | 0.002377 | 0.002228 | 0.002626 | 0.002792 |
| MPV | Platelet mean volume [Entitic volume] in Blood by Automated count | blood | MEAN PLATELET VOLUME | Platelet mean volume [Entitic volume] in Blood by Automated count | MPV | fL | | 0.6866 | 1.60e-33 | 26726 | 19784 | 0.3031 | 0.2425 | 0.006951 | 0.00904 | 0.3101 | 0.2515 | 0.01243 | 0.01581 | 0.00212 | 0.002796 | 0.01256 |
| Myelo | Myelocytes/100 leukocytes in Blood by Manual count | immune / inflammatory | MYELOCYTES % | Myelocytes/100 leukocytes in Blood by Manual count | Myelo | % | | 0.1298 | 0.2732 | 3755 | 3995 | 0.02541 | 0.1272 | 0.004846 | 1.00e-06 | 0.03026 | 0.1272 | 0.06559 | 0.06289 | 0.01223 | 0.01075 | 0.06636 |
| Na-WB | Sodium in Blood | metabolic | SODIUM WHOLE BLOOD | Sodium [Moles/volume] in Blood | Na.WB | mmol/L | | 0.0445 | 0.5888 | 5957 | 7792 | 0.08818 | 0.0474 | 0.005373 | 0.007256 | 0.09355 | 0.05465 | 0.04396 | 0.03234 | 0.007532 | 0.006204 | 0.04445 |
| Na | Sodium in Blood | metabolic | SODIUM BLOOD | Sodium [Moles/volume] in Serum or Plasma | Na | mmol/L | | 0.4471 | 8.87e-10 | 34356 | 26971 | 0.06663 | 0.07386 | 0.002206 | 0.003991 | 0.06884 | 0.07785 | 0.008366 | 0.01053 | 0.001575 | 0.001895 | 0.008493 |
| NasIO2 | NasIO2 | blood | NASAL O2 RATE | | NasIO2 | L | | -0.04677 | 0.6318 | 1278 | 1748 | 0.03576 | | 1.00e-06 | | 0.03576 | | 0.1964 | | 0.03156 | | 0.1992 |
| Neut | Neutrophils [#/volume] in Blood | immune / inflammatory | NEUTROPHILS % | Neutrophils [#/volume] in Blood | Neut | % | | 0.2878 | 6.60e-05 | 16047 | 11786 | 0.07582 | 0.06834 | 0.002882 | 1.00e-06 | 0.0787 | 0.06834 | 0.01681 | 0.02183 | 0.003159 | 0.003284 | 0.01707 |

| all_labs | full_name | group | concept_name | LOINC_NAME | heritability_names | selected_unit | sex_specific_lab | genetic.correlation | genetic.correlation.p | gcta.N_female | gcta.N_male | gcta.Variance_Autosomal_h2_female | gcta.Variance_Autosomal_h2_male | gcta.Variance_X_chromosome_h2_female | gcta.Variance_X_chromosome_h2_male | gcta.Variance_h2_female | gcta.Variance_h2_male | gcta.SE_Autosomal_h2_female | gcta.SE_Autosomal_h2_male | gcta.SE_X_chromosome_h2_female | gcta.SE_X_chromosome_h2_male | gcta.SE_h2_female |
|---|---|---|---|---|---|---|---|---|---|---|---|---|---|---|---|---|---|---|---|---|---|---|
| NEUTA | Neutrophils absolute | immune / inflammatory | NEUT (ABS) | Neutrophils [#/volume] in Blood | NEUTA | 10*3/uL | | 0.215 | 0.02268 | 5142 | 5349 | 0.0442 | 1.00e-06 | 1.00e-06 | 0.001467 | 0.0442 | 0.001468 | 0.04937 | 0.0468 | 0.009248 | 0.007941 | 0.05013 |
| NEUTAB | Neutrophils absolute automated count | immune / inflammatory | NEUT (ABS) | Neutrophils [#/volume] in Blood by Automated count | NEUTAB | 10*3/uL | | 0.4018 | 7.80e-07 | 25593 | 19958 | 4.50e-05 | 0.001218 | 0.001764 | 5.25e-04 | 0.00181 | 0.001743 | 0.001362 | 0.001831 | 0.002032 | 0.002083 | 0.002455 |
| NEUTM | Polymorphonuclear cells/100 leukocytes in Body fluid by Manual count | immune / inflammatory | NEUTROPHIL % | Neutrophils [#/volume] in Blood by Manual count | NEUTM | % | | -0.03807 | 0.6869 | 5147 | 5352 | 0.03406 | | 1.00e-06 | | 0.03406 | | 0.04552 | | 0.008926 | | 0.04639 |
| NEUTRE | Neutrophils/100 leukocytes in Blood by Automated count | immune / inflammatory | NEUTROPHILS % | Neutrophils/100 leukocytes in Blood by Automated count | NEUTRE | % | | 0.2696 | 4.44e-07 | 25683 | 20000 | 0.001391 | 9.30e-04 | 1.00e-06 | 1.41e-04 | 0.001392 | 0.001071 | 0.001423 | 0.001673 | 0.001692 | 0.001872 | 0.00221 |
| NonHDL | Cholesterol non HDL [Mass/volume] in Serum or Plasma | metabolic | NON-HDL | Cholesterol non HDL [Mass/volume] in Serum or Plasma | NonHDL | mg/dL | | 0.07192 | 0.4703 | 1846 | 2171 | 0.1838 | 0.2252 | 0.03004 | 0.04738 | 0.2139 | 0.2726 | 0.1331 | 0.1162 | 0.02596 | 0.02553 | 0.1348 |
| NRBC | NRBC | blood | NUCLEATED RBC | Nucleated erythrocytes/100 leukocytes [Ratio] in Blood by Automated count | NRBC | /100.{WBC} | | 0.09594 | 0.2741 | 16758 | 12556 | 0.03578 | 1.00e-06 | 0.004646 | 0.001212 | 0.04043 | 0.001213 | 0.01537 | 0.01993 | 0.003315 | 0.003487 | 0.01571 |
| NRBC# | NRBC # | blood | NUCLEATED RBC# | Nucleated erythrocytes [#/volume] in Blood by Automated count | NRBC. | 10*3/uL | | 0.2639 | 0.02008 | 31113 | 23718 | 0.01732 | 0.05044 | 1.00e-06 | 1.00e-06 | 0.01732 | 0.05044 | 0.00818 | 0.0115 | 0.001562 | 0.002012 | 0.008308 |
| NRBCBC | Nucleated erythrocytes/100 leukocytes [Ratio] in Cerebral spinal fluid by Manual count | blood | NUCLEATED RBC | Nucleated erythrocytes/100 leukocytes [Ratio] in Cerebral spinal fluid by Manual count | NRBCBC | /100.{WBC} | | -0.04033 | 0.7638 | 1048 | 1153 | 0.227 | | 1.00e-06 | | 0.227 | | 0.2445 | | 0.04419 | | 0.2478 |
| NRBCMA | Nucleated erythrocytes [#/volume] in Blood by Automated count | blood | NUCLEATED RBC# | Nucleated erythrocytes [#/volume] in Blood | NRBCMA | 10*3/mL | | 0.2026 | 0.09425 | 1045 | 1142 | | | | | | | | | | | |
| Nt-PTH | NT PTH | endocrine | INTACT-PTH | Parathyrin.intact [Mass/volume] in Serum or Plasma | Nt.PTH | pg/mL | | 0.09407 | 0.3439 | 5143 | 2958 | 0.1349 | 1.00e-06 | 1.00e-06 | 0.01298 | 0.1349 | 0.01298 | 0.05126 | 0.08676 | 0.009021 | 0.01575 | 0.05203 |

| all_labs | full_name | group | concept_name | LOINC_NAME | heritability_names | selected_unit | sex_specific_lab | genetic.correlation | genetic.correlation.p | gcta.N_female | gcta.N_male | gcta.Variance_Autosomal_h2_female | gcta.Variance_Autosomal_h2_male | gcta.Variance_X_chromosome_h2_female | gcta.Variance_X_chromosome_h2_male | gcta.Variance_h2_female | gcta.Variance_h2_male | gcta.SE_Autosomal_h2_female | gcta.SE_Autosomal_h2_male | gcta.SE_X_chromosome_h2_female | gcta.SE_X_chromosome_h2_male | gcta.SE_h2_female |
|---|---|---|---|---|---|---|---|---|---|---|---|---|---|---|---|---|---|---|---|---|---|---|
| NtAbs | Absolute neutrophils | immune / inflammatory | NEUT (ABS) | | NtAbs | 10*3/uL | | 0.2402 | 0.00401 | 15125 | 11008 | 0.1118 | 0.1034 | 0.004151 | 1.00e-06 | 0.1159 | 0.1034 | 0.01824 | 0.0238 | 0.003389 | 0.00345 | 0.01852 |
| NTAuto | Neutrophils | immune / inflammatory | NT AUTOMATED ABS | Neutrophils [#/volume] in Blood by Automated count | NTAuto | 10*3/uL | | 0.4309 | 1.72e-07 | 27996 | 21662 | 0.08304 | 0.079 | 0.001926 | 0.002086 | 0.08496 | 0.08109 | 0.01029 | 0.0127 | 0.001858 | 0.002166 | 0.01043 |
| O2 Liter Flow | O2 liter flow | vitals | Delivered oxygen flow rate | | O2.Liter.Flow | L/min | | -0.01638 | 0.887 | 9833 | 9654 | | 0.0123 | | 0.006611 | | 0.01891 | | 0.025 | | 0.00521 | |
| O2Sat | Oxygen saturation Calculated from oxygen partial pressure in Arterial blood | vitals | O2 SATURATION | Oxygen saturation Calculated from oxygen partial pressure in Arterial blood | O2Sat | % | | -0.07217 | 0.4108 | 4695 | 6419 | 0.06008 | 0.01034 | 0.01339 | 1.00e-06 | 0.07347 | 0.01034 | 0.05457 | 0.03877 | 0.01022 | 0.007214 | 0.05522 |
| O2SAT | Oxygen saturation calculated from oxygen partial pressure in arterial blood | vitals | Oxygen saturation in Blood | Oxygen saturation in Blood | O2SAT | | | 0.4641 | 2.04e-08 | 30062 | 23729 | 0.05654 | 0.05254 | 1.00e-06 | 1.00e-06 | 0.05654 | 0.05254 | 0.009401 | 0.01152 | 0.001562 | 0.001957 | 0.00951 |
| all_labs | full_name | group | concept_name | LOINC_NAME | heritability_names | selected_unit | sex_specific_lab | genetic.correlation | genetic.correlation.p | gcta.N_female | gcta.N_male | gcta.Variance_Autosomal_h2_female | gcta.Variance_Autosomal_h2_male | gcta.Variance_X_chromosome_h2_female | gcta.Variance_X_chromosome_h2_male | gcta.Variance_h2_female | gcta.Variance_h2_male | gcta.SE_Autosomal_h2_female | gcta.SE_Autosomal_h2_male | gcta.SE_X_chromosome_h2_female | gcta.SE_X_chromosome_h2_male | gcta.SE_h2_female |
| O2SatC | O2 saturation (calculated) | vitals | O2 SATURATION (CALC) | | O2SatC | % | | 0.1978 | 0.0522 | 3797 | 4606 | 0.0879 | 0.06898 | 0.01383 | 1.00e-06 | 0.1017 | 0.06898 | 0.067 | 0.05449 | 0.01371 | 0.008651 | 0.06784 |
| O2SatV | Oxygen saturation Calculated from oxygen partial pressure in Venous blood | vitals | O2 SATURATION(VENOUS) | Oxygen saturation Calculated from oxygen partial pressure in Venous blood | O2SatV | % | | 0.0655 | 0.3968 | 3787 | 4535 | 0.04871 | 0.05386 | 6.03e-04 | 1.00e-06 | 0.04931 | 0.05386 | 0.06284 | 0.05353 | 0.01041 | 0.01043 | 0.06368 |
| OB Infant Apgar Score 1 M 1 | 1 minute Apgar score | ob / gyn | 1 minute Apgar Score | 1 minute Apgar Score | OB.Infant.Apgar.Score.1.M.1 | | Sex specific lab | | | 1217 | | | | | | | | | | | | |
| OB Infant Apgar Score 5 M 1 | 5 minute Apgar score | ob / gyn | 5 minute Apgar Score | 5 minute Apgar Score | OB.Infant.Apgar.Score.5.M.1 | | Sex specific lab | | | 1217 | | 0.07405 | | 1.00e-06 | | 0.07405 | | 0.2102 | | 0.03326 | | 0.2106 |
| Osmol | Osmolality of Serum or Plasma | blood | OSMOLALITY BLOOD | Osmolality of Serum or Plasma | Osmol | mosm/kg{H20} | | -0.08377 | 0.3262 | 1780 | 1755 | 0.01329 | 0.03705 | 1.00e-06 | 1.00e-06 | 0.01329 | 0.03705 | 0.1299 | 0.1405 | 0.02441 | 0.0257 | 0.1318 |
| P DURATION | P wave duration | cardiovascular | P wave duration | P wave duration | P.DURATION | | | -0.0656 | 0.3512 | 7220 | 6307 | 0.07306 | 0.04323 | 0.007092 | 1.00e-06 | 0.08015 | 0.04323 | 0.03495 | 0.03833 | 0.006996 | 0.007215 | 0.03557 |

| all_labs | full_name | group | concept_name | LOINC_NAME | heritability_names | selected_unit | sex_specific_lab | genetic.correlation | genetic.correlation.p | gcta.N_female | gcta.N_male | gcta.Variance_Autosomal_h2_female | gcta.Variance_Autosomal_h2_male | gcta.Variance_X_chromosome_h2_female | gcta.Variance_X_chromosome_h2_male | gcta.Variance_h2_female | gcta.Variance_h2_male | gcta.SE_Autosomal_h2_female | gcta.SE_Autosomal_h2_male | gcta.SE_X_chromosome_h2_female | gcta.SE_X_chromosome_h2_male | gcta.SE_h2_female |
|---|---|---|---|---|---|---|---|---|---|---|---|---|---|---|---|---|---|---|---|---|---|---|
| P HORIZONTAL AXIS | P wave axis horizontal plane | cardiovascular | P wave axis horizontal plane Reference beat | P wave axis horizontal plane Reference beat | P.HORIZONTAL.AXIS | | | 0.2467 | 0.007978 | 6926 | 5960 | 0.04173 | 0.01814 | 0.002627 | 1.00e-06 | 0.04436 | 0.01814 | 0.03523 | 0.04081 | 0.006637 | 0.009226 | 0.03584 |
| P_WAVE | P wave axis | cardiovascular | P wave axis | P wave axis | P_WAVE | deg | | 0.2695 | 1.53e-04 | 21451 | 19267 | 2.50e-05 | 1.90e-04 | 1.00e-06 | 0.001691 | 2.60e-05 | 0.001881 | 0.001423 | 0.001785 | 0.00217 | 0.002372 | 0.002599 |
| Pat-PT | Prothrombin time | blood | PATIENT (PT) | Prothrombin time (PT) | Pat.PT | s | | 0.05278 | 0.5258 | 19238 | 18382 | 0.03874 | 0.04948 | 1.00e-06 | 1.00e-06 | 0.03874 | 0.04948 | 0.01382 | 0.01432 | 0.002338 | 0.00245 | 0.01399 |
| Pat-TT | Thrombin time | blood | PATIENT (TT) | Thrombin time | Pat.TT | s | | 0.3732 | 0.1294 | 833 | 419 | 0.0703 | 0.4418 | 0.04364 | 1.00e-06 | 0.1139 | 0.4418 | 0.2929 | 0.633 | 0.05287 | 0.1098 | 0.2993 |
| pC02-V | Carbon dioxide [Partial pressure] in Venous blood | blood | PCO2 VENOUS | Carbon dioxide [Partial pressure] in Venous blood | pC02.V | mm[Hg] | | -0.05581 | 0.5751 | 4142 | 4502 | 0.1056 | 0.002777 | 1.00e-06 | 0.007664 | 0.1056 | 0.01044 | 0.05979 | 0.05201 | 0.01237 | 0.01097 | 0.06102 |
| PCAct | Protein C [ Units/volume] in Platelet poor plasma by Chromogenic method | blood | PROTEIN C ACTIVITY | Protein C actual/normal in Platelet poor plasma by Coagulation assay | PCAct | % | | 0.04979 | 0.856 | 656 | 352 | | 0.2716 | | 1.00e-06 | | 0.2716 | | 0.7062 | | 0.124 | |
| pCO2 | Carbon dioxide [Partial pressure] in Arterial blood | blood | PCO2 ARTERIAL | Carbon dioxide [Partial pressure] in Arterial blood | pCO2 | mm[Hg] | | 0.3588 | 0.004535 | 7132 | 8663 | 0.08042 | 1.00e-06 | 1.00e-06 | 0.005689 | 0.08042 | 0.00569 | 0.03519 | 0.02782 | 0.005697 | 0.005162 | 0.03565 |
| PCR | Protein/Creatinine [Mass ratio] in Urine | kidney | Protein/Creatinine [Mass Ratio] in Urine | Protein/Creatinine [Mass Ratio] in Urine | PCR | g/g | | 0.07167 | 0.5059 | 2818 | 2909 | 0.03738 | | 1.00e-06 | | 0.03738 | | 0.0878 | | 0.01472 | | 0.0892 |
| PCV | Hematocrit [Volume Fraction] of Blood by Automated count | blood | PCV BLOOD | Hematocrit [Volume Fraction] of Blood by Automated count | PCV | % | | 0.4058 | 3.52e-11 | 36195 | 27330 | 0.07716 | 0.0613 | 0.002314 | 1.00e-06 | 0.07948 | 0.0613 | 0.008286 | 0.01021 | 0.001574 | 0.001648 | 0.008429 |
| PEFR_AFD | PEFR_AFD | lung | PEFR_AFD | | PEFR_AFD | | | 0.08174 | 0.7085 | 587 | 506 | 1.00e-06 | 0.94 | 0.01593 | 0.05996 | 0.01593 | 1 | 0.4319 | 0.5488 | 0.06233 | 0.1105 | 0.436 |
| PEFR_APP | PEFR_APP | lung | PEFR_APP | | PEFR_APP | | | -0.5403 | 0.521 | 587 | 506 | | 0.9389 | | 0.06107 | | 1 | | 0.4797 | | 0.09984 | |
| PEFR_PPR | PEFR_PPR | lung | PEFR_PPR | | PEFR_PPR | | | 0.1054 | 0.3497 | 1735 | 1604 | 1.00e-06 | 0.1155 | 0.001415 | 0.04065 | 0.001416 | 0.1562 | 0.1331 | 0.1564 | 0.02264 | 0.03321 | 0.1347 |
| PEFR_PRD | PEFR_PRD | lung | PEFR_PRD | | PEFR_PRD | | | -0.1386 | 0.04224 | 1736 | 1604 | 0.2489 | 0.6655 | 1.00e-06 | 1.00e-06 | 0.2489 | 0.6655 | 0.149 | 0.1584 | 0.02018 | 0.02786 | 0.1502 |
| PEFR_PRL | PEFR_PRL | lung | PEFR_PRL | | PEFR_PRL | | | 0.02479 | 0.8035 | 1540 | 1362 | 0.2316 | 0.5203 | 6.85e-04 | 1.00e-06 | 0.2323 | 0.5203 | 0.1663 | 0.1848 | 0.02434 | 0.03132 | 0.1675 |
| PEFR_ZBD | PEFR_ZBD | lung | PEFR_ZBD | | PEFR_ZBD | | | 0.1179 | 0.2552 | 1736 | 1604 | 1.00e-06 | 0.1843 | 0.005237 | 0.0558 | 0.005238 | 0.2401 | 0.1347 | 0.1562 | 0.02273 | 0.03472 | 0.1361 |
| PEFR_ZPC | PEFR_ZPD | lung | PEFR_ZPC | | PEFR_ZPC | | | -0.3004 | 0.2049 | 569 | 491 | 0.0189 | 0.1717 | 1.00e-06 | 1.00e-06 | 0.0189 | 0.1717 | 0.4246 | 0.521 | 0.07153 | 0.08051 | 0.4359 |
| pH-Ven | Venous pH | blood | VENOUS PH | pH of Venous blood | pH.Ven | | | -0.06683 | 0.5837 | 4438 | 4714 | 0.01085 | 1.00e-06 | 0.001035 | 0.004488 | 0.01188 | 0.004489 | 0.05726 | 0.04952 | 0.008863 | 0.009631 | 0.05797 |

| all_labs | full_name | group | concept_name | LOINC_NAME | heritability_names | selected_unit | sex_specific_lab | genetic.correlation | genetic.correlation.p | gcta.N_female | gcta.N_male | gcta.Variance_Autosomal_h2_female | gcta.Variance_Autosomal_h2_male | gcta.Variance_X_chromosome_h2_female | gcta.Variance_X_chromosome_h2_male | gcta.Variance_h2_female | gcta.Variance_h2_male | gcta.SE_Autosomal_h2_female | gcta.SE_Autosomal_h2_male | gcta.SE_X_chromosome_h2_female | gcta.SE_X_chromosome_h2_male | gcta.SE_h2_female |
|---|---|---|---|---|---|---|---|---|---|---|---|---|---|---|---|---|---|---|---|---|---|---|
| Phos | Phosphate | metabolic | PHOSPHORUS INORGANIC | Phosphate [Mass/volume] in Serum or Plasma | Phos | mg/dL | | 0.247 | 0.0219 | 10326 | 9880 | 0.0462 | 0.05144 | 8.04e-04 | 0.004255 | 0.047 | 0.05569 | 0.02456 | 0.02551 | 0.004387 | 0.005109 | 0.02491 |
| PIFR_AFD | PIFR_AFD | lung | PIFR_AFD | | PIFR_AFD | | | -0.3208 | 0.2187 | 586 | 506 | 0.02634 | 0.8902 | 1.00e-06 | 0.1098 | 0.02635 | 1 | 0.4268 | 0.543 | 0.06374 | 0.1082 | 0.4315 |
| PIFR_APP_APP | PIFR_APP_APP | lung | PIFR_APP_APP | | PIFR_APP_APP | | | -0.3806 | 0.1454 | 587 | 506 | 0.0989 | 0.9618 | 1.00e-06 | 0.03819 | 0.0989 | 1 | 0.4249 | 0.487 | 0.06223 | 0.08791 | 0.4292 |
| PIFR_PPR | PIFR_PPR | lung | PIFR_PPR | | PIFR_PPR | | | 0.02942 | 0.7639 | 1734 | 1604 | 1.00e-06 | 1.00e-06 | 0.03773 | 0.01993 | 0.03773 | 0.01993 | 0.1364 | 0.1516 | 0.02883 | 0.02944 | 0.139 |
| PIFR_PRD | PIFR_PRD | lung | PIFR_PRD | | PIFR_PRD | | | -0.1213 | 0.1321 | 1735 | 1604 | 0.1931 | 0.6098 | 1.00e-06 | 1.00e-06 | 0.1931 | 0.6098 | 0.1497 | 0.1582 | 0.02132 | 0.02882 | 0.1512 |
| PIFR_ZBD | PIFR_ZBD | lung | PIFR_ZBD | | PIFR_ZBD | | | -0.08969 | 0.4283 | 1735 | 1604 | 1.00e-06 | 0.01853 | 0.02718 | 0.03118 | 0.02718 | 0.04971 | 0.134 | 0.153 | 0.02666 | 0.03197 | 0.1363 |
| PIFR_ZPC | PIFR_ZPC | lung | PIFR_ZPC | | PIFR_ZPC | | | 0.6807 | 0.5058 | 570 | 488 | 0.4251 | | 0.02431 | | 0.4494 | | 0.3987 | | 0.07158 | | 0.4021 |
| Plt-Ct | Platelet Count | blood | PLATELET COUNT | Platelets [#/volume] in Blood by Automated count | Plt.Ct | 10*3/uL | | 0.4988 | 2.41e-14 | 35703 | 26981 | 0.1292 | 0.106 | 0.002193 | 0.003846 | 0.1314 | 0.1098 | 0.009053 | 0.01109 | 0.001503 | 0.001969 | 0.009162 |
| pO2_FI | Oxygen [Partial pressure] in Arterial blood | blood | PO2/FIO2 | Oxygen [Partial pressure] in Arterial blood | pO2_FI | mm[Hg] | | -0.1372 | 0.1459 | 3250 | 4659 | 0.03872 | 1.00e-06 | 1.00e-06 | 0.006836 | 0.03872 | 0.006837 | 0.07679 | 0.05511 | 0.01368 | 0.01032 | 0.07826 |
| pO2-V | pO2 venous | blood | PO2 VENOUS | Oxygen [Partial pressure] in Venous blood | pO2.V | mm[Hg] | | -0.06454 | 0.5493 | 4071 | 4442 | 4.37e-04 | 0.0113 | 1.00e-06 | 0.003274 | 4.38e-04 | 0.01458 | 0.05526 | 0.05499 | 0.01191 | 0.01079 | 0.05639 |
| pO2 | pO2 arterial | blood | PO2 ARTERIAL | Oxygen [Partial pressure] in Arterial blood | pO2 | mm[Hg] | | 0.05563 | 0.5911 | 7210 | 8698 | 0.06008 | 0.03877 | 0.005792 | 1.00e-06 | 0.06588 | 0.03877 | 0.03524 | 0.0292 | 0.00577 | 0.005429 | 0.03559 |
| POCA1C | Hemoglobin A1c (Glycated) | blood | HGB A1C GLYCATED POC | Hemoglobin A1c/Hemoglobin.total in Blood | POCA1C | % | | 0.0233 | 0.8003 | 1988 | 1866 | 0.1305 | | 0.01531 | | 0.1458 | | 0.1291 | | 0.02268 | | 0.1306 |
| PR INTERVAL | PR Interval mean | cardiovascular | PP interval mean | PP interval mean | PR.INTERVAL | | | 0.2514 | 0.01074 | 7089 | 6106 | 0.1166 | 0.04342 | 1.00e-06 | 0.00479 | 0.1166 | 0.04821 | 0.03663 | 0.04133 | 0.005975 | 0.007668 | 0.03706 |
| PR_INTERVAL | PR Interval mean | cardiovascular | PP interval mean | PP interval mean | PR_INTERVAL | ms | | 0.4247 | 1.78e-05 | 21460 | 19278 | 2.05e-04 | 0.005278 | 1.00e-06 | 0.009209 | 2.06e-04 | 0.01449 | 0.001506 | 0.002304 | 0.002228 | 0.003176 | 0.002701 |
| Pr24Hr | Protein 24 hour | urinary | PROTEIN /24 HOUR | | Pr24Hr | g/(24.h) | | -0.04208 | 0.822 | 1008 | 529 | 0.303 | | 0.0599 | | 0.3629 | | 0.2579 | | 0.04572 | | 0.2635 |
| Pralbu | Prealbumin [Mass/volume] in Serum or Plasma | metabolic | PREALBUMIN BLOOD | Prealbumin [Mass/volume] in Serum or Plasma | Pralbu | mg/dL | | 0.2071 | 0.1211 | 3276 | 3583 | 0.1154 | | 0.02775 | | 0.1432 | | 0.07699 | | 0.01677 | | 0.0791 |
| PRODUCT BLOOD TYPE | Product blood type | blood | PRODUCT BLOOD TYPE | | PRODUCT.BLOOD.TYPE | | | 0.0769 | 0.7396 | 594 | 598 | | 0.3295 | | 1.00e-06 | | 0.3295 | | 0.4206 | | 0.08098 | |
| Prog-R | Progesterone rapid | ob / gyn | PROGESTERONE RAPID | Progesterone [Mass/volume] in Serum or Plasma | Prog.R | ng/mL | Sex specific lab | | | 1116 | 16 | 0.3836 | | 0.03368 | | 0.4173 | | 0.2305 | | 0.03791 | | 0.2339 |

| all_labs | full_name | group | concept_name | LOINC_NAME | heritability_names | selected_unit | sex_specific_lab | genetic.correlation | genetic.correlation.p | gcta.N_female | gcta.N_male | gcta.Variance_Autosomal_h2_female | gcta.Variance_Autosomal_h2_male | gcta.Variance_X_chromosome_h2_female | gcta.Variance_X_chromosome_h2_male | gcta.Variance_h2_female | gcta.Variance_h2_male | gcta.SE_Autosomal_h2_female | gcta.SE_Autosomal_h2_male | gcta.SE_X_chromosome_h2_female | gcta.SE_X_chromosome_h2_male | gcta.SE_h2_female |
|---|---|---|---|---|---|---|---|---|---|---|---|---|---|---|---|---|---|---|---|---|---|---|
| Prolac | Prolactin [Mass/volume] in Serum or Plasma | endocrine | PROLACTIN | Prolactin [Mass/volume] in Serum or Plasma | Prolac | ng/mL | | 0.0518 | 0.6693 | 2100 | 964 | 0.12 | | 0.02388 | | 0.1439 | | 0.1123 | | 0.02423 | | 0.1147 |
| Promyl | Promyelocytes/100 leukocytes in Blood by Manual count | blood | PROMYELO % | Promyelocytes/100 leukocytes in Blood by Manual count | Promyl | | | 0.03858 | 0.6449 | 1397 | 1537 | | 0.08652 | | 1.00e-06 | | 0.08652 | | 0.1564 | | 0.03126 | |
| PSA | PSA serum/plasma | cancer | PROSTATE SPECIFIC ANTIGEN | Prostate specific Ag [Mass/volume] in Serum or Plasma | PSA | ng/mL | Sex specific lab | | | | 9198 | | 0.05851 | | 0.003299 | | 0.06181 | | 0.02882 | | 0.005086 | |
| PT-inr | International normalized ratio | blood | INR | INR in Platelet poor plasma or blood by Coagulation assay | PT.inr | | | 0.1254 | 0.09714 | 19329 | 18431 | 0.02168 | 0.04394 | 1.00e-06 | 1.00e-06 | 0.02169 | 0.04394 | 0.01336 | 0.01412 | 0.002412 | 0.002477 | 0.01355 |
| PTT-pt | aPTT in Platelet poor plasma | blood | PATIENT PTT | aPTT in Platelet poor plasma by Coagulation assay | PTT.pt | s | | 0.1856 | 0.08897 | 17241 | 16272 | 0.03064 | 0.04533 | 0.001417 | 0.002085 | 0.03206 | 0.04742 | 0.01459 | 0.01581 | 0.002706 | 0.002877 | 0.01483 |
| PTTCtr | aPTT in Platelet poor plasma by Coagulation assay | blood | CONTROL PTT | aPTT in Platelet poor plasma by Coagulation assay | PTTCtr | s | | -0.09515 | 0.2928 | 10228 | 9959 | | 0.02298 | | 0.009319 | | 0.0323 | | 0.02515 | | 0.005466 | |
| PULSE OXIMETRY | Oxygen saturation in Arterial blood by pulse oximetry | vitals | Oxygen saturation in Arterial blood by Pulse oximetry | Oxygen saturation in Arterial blood by Pulse oximetry | PULSE.OXIMETRY | % | | 0.2278 | 0.01598 | 17346 | 11869 | 0.01706 | 0.06756 | 0.003554 | 0.004917 | 0.02062 | 0.07248 | 0.01438 | 0.02221 | 0.002953 | 0.00425 | 0.01467 |
| Pulse | Heart rate | vitals | Heart rate | Heart rate | Pulse | | | 0.374 | 5.46e-11 | 35247 | 26707 | 0.09627 | 0.08775 | 0.001893 | 0.002769 | 0.09816 | 0.09052 | 0.008522 | 0.01083 | 0.001446 | 0.001798 | 0.008634 |
| PULSE | Heart rate | vitals | Heart rate | Heart rate | PULSE | | | 0.3359 | 2.12e-04 | 19242 | 12927 | 0.1087 | 0.07868 | 0.005628 | 1.50e-04 | 0.1143 | 0.07883 | 0.0147 | 0.02065 | 0.003047 | 0.003346 | 0.01499 |
| Q ONSET | Q onset | cardiovascular | | | Q.ONSET | | | -0.04179 | 0.6799 | 7230 | 6313 | 0.02175 | 0.02219 | 9.10e-05 | 0.001762 | 0.02184 | 0.02395 | 0.03243 | 0.03881 | 0.006208 | 0.006738 | 0.03292 |
| QMG-NL | Quantiferon Mitogen assay | immune / inflammatory | QUANTIFERON MITOGEN(-)NIL | Mycobacterium tuberculosis tuberculin stimulated gamma interferon/Mitogen stimulated gamma interferon [Units/volume] in Control Blood | QMG.NL | [iU]/mL | | -0.1746 | 0.2783 | 892 | 747 | 0.1971 | 0.5657 | 0.05167 | 1.00e-06 | 0.2488 | 0.5657 | 0.2759 | 0.3305 | 0.05493 | 0.06243 | 0.2813 |
| QRS DURATION | QRS duration | cardiovascular | QRS duration | QRS duration | QRS.DURATION | | | -0.01112 | 0.8803 | 7224 | 6316 | 0.1556 | 0.07273 | 0.002668 | 0.001791 | 0.1583 | 0.07452 | 0.037 | 0.04107 | 0.005986 | 0.007003 | 0.03751 |

| all_labs | full_name | group | concept_name | LOINC_NAME | heritability_names | selected_unit | sex_specific_lab | genetic.correlation | genetic.correlation.p | gcta.N_female | gcta.N_male | gcta.Variance_Autosomal_h2_female | gcta.Variance_Autosomal_h2_male | gcta.Variance_X_chromosome_h2_female | gcta.Variance_X_chromosome_h2_male | gcta.Variance_h2_female | gcta.Variance_h2_male | gcta.SE_Autosomal_h2_female | gcta.SE_Autosomal_h2_male | gcta.SE_X_chromosome_h2_female | gcta.SE_X_chromosome_h2_male | gcta.SE_h2_female |
|---|---|---|---|---|---|---|---|---|---|---|---|---|---|---|---|---|---|---|---|---|---|---|
| QRS HORIZONTAL AXIS | QRS horizontal axis | cardiovascular | QRS axis.horizontal plane Reference beat | QRS axis.horizontal plane Reference beat | QRS.HORIZONTAL.AXIS | | | -0.007049 | 0.9195 | 7176 | 6237 | | 0.055 | | 0.006515 | | 0.06152 | | 0.03935 | | 0.007432 | |
| QRS_DURATION | QRS duration | cardiovascular | QRS duration | QRS duration | QRS_DURATION | ms | | 0.4398 | 2.03e-08 | 21446 | 19269 | 5.40e-04 | 0.002782 | 3.27e-04 | 0.003668 | 8.67e-04 | 0.006449 | 0.001633 | 0.002054 | 0.002115 | 0.002635 | 0.002667 |
| QT INTERVAL | QT interval | cardiovascular | Q-T interval | Q-T interval | QT.INTERVAL | | | -0.0554 | 0.5992 | 7225 | 6310 | 0.06632 | 0.022 | 0.002165 | 0.002173 | 0.06848 | 0.02417 | 0.03555 | 0.03933 | 0.006654 | 0.007288 | 0.03599 |
| QT_INTERVAL_C | QT interval corrected | cardiovascular | Q-T interval corrected | Q-T interval corrected | QT_INTERVAL_C | ms | | 0.4188 | 4.77e-07 | 21459 | 19274 | 0.001514 | 0.002943 | 0.004413 | 0.002548 | 0.005928 | 0.005491 | 0.001752 | 0.002025 | 0.002533 | 0.002674 | 0.003067 |
| QT_INTERVAL | QT interval | cardiovascular | Q-T interval | Q-T interval | QT_INTERVAL | ms | | 0.2891 | 6.33e-04 | 21462 | 19277 | | 8.01e-04 | | 0.00376 | | 0.004561 | | 0.001918 | | 0.002392 | |
| QT3 | Triiodothyronine (T3) [Mass/volume] in Serum or Plasma | endocrine | TRIIODOTHYRONINE, TOTAL | Triiodothyronine (T3) [Mass/volume] in Serum or Plasma | QT3 | ng/dL | | 0.07653 | 0.5094 | 3383 | 1478 | 0.07178 | 0.06902 | 0.02881 | 0.006319 | 0.1006 | 0.07534 | 0.07147 | 0.17 | 0.0161 | 0.03141 | 0.07305 |
| QTB-NL | Quantiferon TB assay | immune / inflammatory | QUANTIFERON TB(-)NIL | Mycobacterium tuberculosis stimulated gamma interferon release by CD4+ T-cells [Units/volume] corrected for background in Blood | QTB.NL | [iU]/mL | | 0.06498 | 0.6621 | 990 | 827 | 1.00e-06 | | 0.02272 | | 0.02272 | | 0.2492 | | 0.04167 | | 0.2517 |
| QTCB | Q-T interval corrected based on Bazett formula | cardiovascular | Q-T interval corrected based on Bazett formula | Q-T interval corrected based on Bazett formula | QTCB | | | 0.1987 | 0.05202 | 7225 | 6312 | 0.04094 | 0.1055 | 0.00223 | 0.01347 | 0.04317 | 0.119 | 0.03404 | 0.04139 | 0.005813 | 0.008676 | 0.03448 |
| QTCF | Q-T interval corrected based on Fridericia formula | cardiovascular | Q-T interval corrected based on Fridericia formula | Q-T interval corrected based on Fridericia formula | QTCF | | | 0.05251 | 0.6728 | 7224 | 6311 | 0.05805 | 0.06511 | 1.00e-06 | 0.01355 | 0.05805 | 0.07867 | 0.03498 | 0.04033 | 0.005972 | 0.008604 | 0.03537 |
| QtfNIL | Gamma interferon background [Units/volume] in Blood by Immunoassay | immune / inflammatory | QUANTIFERON NIL | Gamma interferon background [Units/volume] in Blood by Immunoassay | QtfNIL | [iU]/mL | | -0.0907 | 0.4214 | 1447 | 1197 | 0.3389 | 0.0217 | 0.04586 | 1.00e-06 | 0.3848 | 0.0217 | 0.1793 | 0.2097 | 0.03477 | 0.04114 | 0.1805 |

| all_labs | full_name | group | concept_name | LOINC_NAME | heritability_names | selected_unit | sex_specific_lab | genetic.correlation | genetic.correlation.p | gcta.N_female | gcta.N_male | gcta.Variance_Autosomal_h2_female | gcta.Variance_Autosomal_h2_male | gcta.Variance_X_chromosome_h2_female | gcta.Variance_X_chromosome_h2_male | gcta.Variance_h2_female | gcta.Variance_h2_male | gcta.SE_Autosomal_h2_female | gcta.SE_Autosomal_h2_male | gcta.SE_X_chromosome_h2_female | gcta.SE_X_chromosome_h2_male | gcta.SE_h2_female |
|---|---|---|---|---|---|---|---|---|---|---|---|---|---|---|---|---|---|---|---|---|---|---|
| Quant_TB1(-)NIL | Quant TB1(-) NIL | immune / inflammatory | Quant_TB1(-)NIL | Mycobacterium tuberculosis stimulated gamma interferon release by CD4+ T-cells [Units/volume] corrected for background in Blood | Quant_TB1...NIL | [iU]/mL | | -0.2762 | 0.4066 | 838 | 639 | 0.1819 | | 1.00e-06 | | 0.1819 | | 0.3004 | | 0.05057 | | 0.3053 |
| Quant_TB2(-)NIL | Quant TB2(-) NIL | immune / inflammatory | Quant_TB2(-)NIL | Mycobacterium tuberculosis stimulated gamma interferon release by CD4+ and CD8+ T-cells [Units/volume] corrected for background in Blood | Quant_TB2...NIL | [iU]/mL | | 0.06621 | 0.9127 | 838 | 639 | 0.4205 | | 1.00e-06 | | 0.4205 | | 0.3024 | | 0.05659 | | 0.307 |
| R AN AIRWAY RESPIRATION RATE | Respiratory rate by airway flow measurement | vitals | Respiratory rate by Airway flow measurement | Respiratory rate by Airway flow measurement | R.AN.AIRWAY.RESPIRATION.RATE | | | -0.008553 | 0.9171 | 6963 | 5423 | 0.02504 | 0.1107 | 1.00e-06 | 0.004251 | 0.02504 | 0.1149 | 0.03643 | 0.04822 | 0.006419 | 0.008386 | 0.03697 |
| R AN ANESTHESIA BLOOD PRESSURE | Diastolic blood pressure by Noninvasive | vitals | Diastolic blood pressure by Noninvasive | Diastolic blood pressure by Noninvasive | R.AN.ANESTHESIA.BLOOD.PRESSURE | mm[Hg] | | 0.01009 | 0.9381 | 7924 | 6029 | 0.03703 | 1.00e-06 | 1.00e-06 | 0.003685 | 0.03703 | 0.003686 | 0.03027 | 0.03845 | 0.006459 | 0.008098 | 0.03087 |
| R AN ANESTHESIA TEMPERATURE | Body temperature | vitals | Body temperature | Body temperature | R.AN.ANESTHESIA.TEMPERATURE | Cel | | 0.09292 | 0.4629 | 1248 | 931 | 0.01151 | 0.2172 | 1.00e-06 | 1.00e-06 | 0.01151 | 0.2172 | 0.2062 | 0.2615 | 0.03368 | 0.04238 | 0.209 |
| R AN CORE BODY TEMPERATURE | Body temperature - Core | vitals | Body temperature - Core | Body temperature - Core | R.AN.CORE.BODY.TEMPERATURE | Cel | | -0.2629 | 0.6827 | 719 | 675 | 0.4622 | 0.4844 | 1.00e-06 | 0.1094 | 0.4622 | 0.5937 | 0.3335 | 0.3893 | 0.05391 | 0.07231 | 0.336 |
| R AN ESOPHAGEAL TEMPERATURE | Temperature - Distal airway | vitals | Temperature Distal airway | Temperature Distal airway | R.AN.ESOPHAGEAL.TEMPERATURE | Cel | | 0.298 | 0.002725 | 1636 | 1304 | 1.00e-06 | | 0.006183 | | 0.006184 | | 0.1557 | | 0.02751 | | 0.1565 |
| R AN HEART RATE MONITOR | Heart rate | vitals | Heart rate | Heart rate | R.AN.HEART.RATE.MONITOR | | | 0.1399 | 0.1796 | 7336 | 5668 | 0.05905 | 7.56e-04 | 0.007223 | 1.00e-06 | 0.06627 | 7.57e-04 | 0.03452 | 0.04271 | 0.006888 | 0.008027 | 0.03507 |
| R AN MEAN ARTERIAL BLOOD PRESSURE | Invasive Mean blood pressure | vitals | Invasive Mean blood pressure | Invasive Mean blood pressure | R.AN.MEAN.ARTERIAL.BLOOD.PRESSURE | mm[Hg] | | -0.04287 | 0.776 | 878 | 1168 | | 0.3449 | | 1.00e-06 | | 0.3449 | | 0.2118 | | 0.03562 | |

| all_labs | full_name | group | concept_name | LOINC_NAME | heritability_names | selected_unit | sex_specific_lab | genetic.correlation | genetic.correlation.p | gcta.N_female | gcta.N_male | gcta.Variance_Autosomal_h2_female | gcta.Variance_Autosomal_h2_male | gcta.Variance_X_chromosome_h2_female | gcta.Variance_X_chromosome_h2_male | gcta.Variance_h2_female | gcta.Variance_h2_male | gcta.SE_Autosomal_h2_female | gcta.SE_Autosomal_h2_male | gcta.SE_X_chromosome_h2_female | gcta.SE_X_chromosome_h2_male | gcta.SE_h2_female |
|---|---|---|---|---|---|---|---|---|---|---|---|---|---|---|---|---|---|---|---|---|---|---|
| R AN NASOPHARYNGEAL TEMPERATURE | Temperature - Distal airway | vitals | Temperature Distal airway | Temperature Distal airway | R.AN.NASOPHARYNGEAL.TEMPERATURE | Cel | | -0.1616 | 0.5131 | 574 | 537 | 0.2986 | 0.5294 | 0.1355 | 0.1376 | 0.4341 | 0.667 | 0.4255 | 0.4703 | 0.08767 | 0.1002 | 0.4306 |
| R AN NIBP MEAN | Mean blood pressure by Noninvasive | vitals | Mean blood pressure by Noninvasive | Mean blood pressure by Noninvasive | R.AN.NIBP.MEAN | mm[Hg] | | -0.06416 | 0.5654 | 7478 | 5736 | 0.02416 | | 1.00e-06 | | 0.02416 | | 0.0311 | | 0.00668 | | 0.03169 |
| R AN PULSE SPO2 | Heart rate by Pulse oximetry | vitals | Heart rate by Pulse oximetry | Heart rate by Pulse oximetry | R.AN.PULSE.SPO2 | | | 0.08988 | 0.2561 | 7283 | 5626 | 0.07424 | 0.01939 | 0.006581 | 1.00e-06 | 0.08082 | 0.01939 | 0.03507 | 0.04339 | 0.006867 | 0.008064 | 0.03563 |
| R AN SKIN TEMPERATURE | Temperature of Skin | vitals | Temperature of Skin | Temperature of Skin | R.AN.SKIN.TEMPERATURE | Cel | | 0.3319 | 4.66e-04 | 1630 | 1261 | | 0.08467 | | 0.04365 | | 0.1283 | | 0.2047 | | 0.04069 | |
| R APACHE TEMPERATURE | Body temperature | vitals | Body temperature | Body temperature | R.APACHE.TEMPERATURE | Cel | | 0.234 | 0.0326 | 17589 | 11789 | 0.01805 | 0.01096 | 0.003037 | 1.00e-06 | 0.02109 | 0.01096 | 0.01439 | 0.02021 | 0.002858 | 0.003633 | 0.01464 |
| R ARTERIAL LINE BLOOD PRESSURE | Invasive Systolic blood pressure | vitals | Invasive Systolic blood pressure | Invasive Systolic blood pressure | R.ARTERIAL.LINE.BLOOD.PRESSURE | mm[Hg] | | 0.1566 | 0.7626 | 512 | 631 | 1.00e-06 | 0.3363 | 0.05778 | 0.1591 | 0.05778 | 0.4954 | 0.516 | 0.4159 | 0.0836 | 0.082 | 0.522 |
| R ED PRE-ARRIVAL BP | Systolic blood pressure by Noninvasive | vitals | Systolic blood pressure by Noninvasive | Systolic blood pressure by Noninvasive | R.ED.PRE.ARRIVAL.BP | mm[Hg] | | -0.1725 | 0.173 | 1097 | 1001 | | 0.2989 | | 1.00e-06 | | 0.2989 | | 0.2624 | | 0.04968 | |
| R ED PRE-ARRIVAL EMS O2 SAT | Oxygen saturation in Arterial blood by Pulse oximetry | vitals | Oxygen saturation in Arterial blood by Pulse oximetry | Oxygen saturation in Arterial blood by Pulse oximetry | R.ED.PRE.ARRIVAL.EMS.O2.SAT | % | | 0.2767 | 0.0482 | 1061 | 981 | 0.09497 | | 0.03443 | | 0.1294 | | 0.2275 | | 0.04534 | | 0.2306 |
| R ED PRE-ARRIVAL PULSE (HEART RATE) | Heart rate | vitals | Heart rate | Heart rate | R.ED.PRE.ARRIVAL.PULSE..HEART.RATE. | | | -0.2237 | 0.1911 | 1100 | 1007 | 0.08837 | 0.1272 | 1.00e-06 | 0.03621 | 0.08837 | 0.1634 | 0.2372 | 0.2491 | 0.04132 | 0.04855 | 0.2415 |
| R ED PRE-ARRIVAL RESP RATE | Respiratory rate | vitals | Respiratory rate | Respiratory rate | R.ED.PRE.ARRIVAL.RESP.RATE | | | -0.1259 | 0.4943 | 1028 | 920 | 1.00e-06 | | 0.03921 | | 0.03921 | | 0.266 | | 0.04811 | | 0.2705 |
| R FN WEIGHT USED FOR EQUATIONS CALCULATIONS | Body weight | vitals | Body weight | Body weight | R.FN.WEIGHT.USED.FOR.EQUATIONS.CALCULATIONS | kg | | 0.6564 | 0.02588 | 522 | 569 | 1.00e-06 | 0.4744 | 0.1068 | 1.00e-06 | 0.1068 | 0.4744 | 0.5346 | 0.4116 | 0.1168 | 0.08604 | 0.5449 |
| R HEART RATE MONITOR | Heart rate | vitals | Heart rate | Heart rate | R.HEART.RATE.MONITOR | | | -0.003951 | 0.9454 | 9190 | 6986 | 0.07575 | 0.06453 | 0.007629 | 1.00e-06 | 0.08338 | 0.06454 | 0.02723 | 0.03606 | 0.006141 | 0.007307 | 0.02782 |
| R IP IDEAL BODY WEIGHT | Ideal body weight | vitals | Ideal body weight | Ideal body weight | R.IP.IDEAL.BODY.WEIGHT | kg | | 0.618 | 5.74e-32 | 19051 | 12623 | 0.3694 | 0.3197 | 0.006133 | 0.01099 | 0.3756 | 0.3307 | 0.01635 | 0.02369 | 0.002915 | 0.004609 | 0.01654 |

| all_labs | full_name | group | concept_name | LOINC_NAME | heritability_names | selected_unit | sex_specific_lab | genetic.correlation | genetic.correlation.p | gcta.N_female | gcta.N_male | gcta.Variance_Autosomal_h2_female | gcta.Variance_Autosomal_h2_male | gcta.Variance_X_chromosome_h2_female | gcta.Variance_X_chromosome_h2_male | gcta.Variance_h2_female | gcta.Variance_h2_male | gcta.SE_Autosomal_h2_female | gcta.SE_Autosomal_h2_male | gcta.SE_X_chromosome_h2_female | gcta.SE_X_chromosome_h2_male | gcta.SE_h2_female |
|---|---|---|---|---|---|---|---|---|---|---|---|---|---|---|---|---|---|---|---|---|---|---|
| R RT (FEMALE ADULT) IDEAL BODY WEIGHT | Ideal body weight — female | vitals | Ideal body weight | Ideal body weight | R.RT..FEMALE.ADULT..IDEAL.BODY.WEIGHT | kg | | 0.6184 | 9.72e-33 | 19052 | 12624 | 0.3695 | 0.3199 | 0.006153 | 0.01102 | 0.3757 | 0.3309 | 0.01635 | 0.02369 | 0.002916 | 0.004611 | 0.01654 |
| R RT (MALE ADULT) IDEAL BODY WEIGHT | Ideal body weight — male | vitals | Ideal body weight | Ideal body weight | R.RT..MALE.ADULT..IDEAL.BODY.WEIGHT | kg | | 0.6184 | 9.72e-33 | 19052 | 12624 | 0.3695 | 0.3199 | 0.006153 | 0.01102 | 0.3757 | 0.3309 | 0.01635 | 0.02369 | 0.002916 | 0.004611 | 0.01654 |
| R VUMC AN RESPIRATION RATE | Respiratory rate | vitals | Respiratory rate | Respiratory rate | R.VUMC.AN.RESPIRATION.RATE | | | 0.07679 | 0.3154 | 5664 | 4501 | 0.01782 | 0.1069 | 1.00e-06 | 0.01657 | 0.01782 | 0.1234 | 0.04359 | 0.05663 | 0.008834 | 0.01141 | 0.04446 |
| R VUMC OXYGEN FLOW DELIVERED | Delivered oxygen flow rate | vitals | Delivered oxygen flow rate | | R.VUMC.OXYGEN.FLOW.DELIVERED | L/min | | 0.0251 | 0.7158 | 5139 | | 0.1103 | | 1.00e-06 | | 0.1103 | | 0.04891 | | 0.009829 | | 0.04971 |
| R_F | Rheumatoid factor [Units/volume] in Serum or Plasma | immune / inflammatory | RHEUMATOID FACTOR | Rheumatoid factor [Units/volume] in Serum or Plasma | R_F | [iU]/mL | | 0.1348 | 0.2559 | 2715 | 1250 | 0.12 | 0.07569 | 1.00e-06 | 0.03206 | 0.12 | 0.1077 | 0.08856 | 0.2066 | 0.01617 | 0.0356 | 0.08942 |
| RATE_C | Heart rate | vitals | Heart rate | Heart rate | RATE_C | | | 0.2622 | 8.66e-05 | 21462 | 19277 | | 0.005439 | | 0.005205 | | 0.01064 | | 0.002343 | | 0.002605 | |
| RBC | Erythrocytes [#/volume] in Blood by Automated count | blood | RBC | Erythrocytes [#/volume] in Blood by Automated count | RBC | 10*6/uL | | 0.4451 | 1.57e-11 | 35638 | 26954 | 0.1006 | 0.08276 | 0.002193 | 0.001202 | 0.1028 | 0.08397 | 0.008697 | 0.01067 | 0.001509 | 0.001749 | 0.008812 |
| RBCFol | Folate [Mass/volume] in Red Blood Cells | metabolic | RBC FOLATE | Folate [Mass/volume] in Red Blood Cells | RBCFol | ng/mL{RBC} | | 0.1008 | 0.2851 | 2842 | 2084 | 0.04303 | 0.03759 | 1.00e-06 | 1.00e-06 | 0.04303 | 0.03759 | 0.08727 | 0.1144 | 0.01382 | 0.02153 | 0.08824 |
| RDW-SD | RDW SD | blood | RDW-SD | Erythrocyte distribution width [Entitic volume] by Automated count | RDW.SD | fL | | 0.002318 | 0.9805 | 3133 | 2143 | 1.00e-06 | 0.0408 | 0.0254 | 0.02377 | 0.0254 | 0.06457 | 0.07724 | 0.1146 | 0.01787 | 0.02232 | 0.07896 |
| RDW | Erythrocyte distribution width [Ratio] by Automated count | blood | RDW | Erythrocyte distribution width [Ratio] by Automated count | RDW | % | | 0.5389 | 5.42e-17 | 35661 | 26955 | 0.1344 | 0.119 | 1.00e-06 | 0.001603 | 0.1344 | 0.1206 | 0.008938 | 0.01099 | 0.001351 | 0.001899 | 0.00902 |
| RDWSD | Erythrocyte distribution width [Entitic volume] by Automated count | blood | RDWSD | Erythrocyte distribution width [Entitic volume] by Automated count | RDWSD | fL | | 0.4728 | 8.04e-19 | 31506 | 24073 | 0.1199 | 0.09522 | 0.00248 | 2.34e-04 | 0.1224 | 0.09545 | 0.009742 | 0.01188 | 0.001691 | 0.001639 | 0.009862 |
| RenAct | Renin [Enzymatic activity/volume] in Plasma | endocrine | RENIN ACTIVITY | Renin [Enzymatic activity/volume] in Plasma | RenAct | ng/mL/h | | 0.3638 | 0.2067 | 701 | 452 | | 1.00e-06 | | 0.09466 | | 0.09466 | | 0.6306 | | 0.1154 | |
| RESPIRATIONS | Respiratory rate | vitals | Respiratory rate | Respiratory rate | RESPIRATIONS | | | 0.1203 | 0.1465 | 16703 | 11399 | 0.01059 | 0.07326 | 1.00e-06 | 0.004164 | 0.01059 | 0.07742 | 0.01484 | 0.02335 | 0.002684 | 0.003942 | 0.01508 |

| all_labs | full_name | group | concept_name | LOINC_NAME | heritability_names | selected_unit | sex_specific_lab | genetic.correlation | genetic.correlation.p | gcta.N_female | gcta.N_male | gcta.Variance_Autosomal_h2_female | gcta.Variance_Autosomal_h2_male | gcta.Variance_X_chromosome_h2_female | gcta.Variance_X_chromosome_h2_male | gcta.Variance_h2_female | gcta.Variance_h2_male | gcta.SE_Autosomal_h2_female | gcta.SE_Autosomal_h2_male | gcta.SE_X_chromosome_h2_female | gcta.SE_X_chromosome_h2_male | gcta.SE_h2_female |
|---|---|---|---|---|---|---|---|---|---|---|---|---|---|---|---|---|---|---|---|---|---|---|
| RespRt | Respiratory rate | vitals | Respiratory rate | Respiratory rate | RespRt | | | 0.1883 | 0.0904 | 30952 | 23979 | 0.01852 | 0.02756 | 5.82e-04 | 0.002297 | 0.0191 | 0.02985 | 0.00829 | 0.01113 | 0.001446 | 0.001916 | 0.008403 |
| RetAbs | Reticulocytes absolute | blood | RET ABS | Reticulocytes [#/volume] in Blood | RetAbs | 10*6/uL | | 0.1684 | 0.03128 | 3978 | 3555 | 0.08721 | 0.01706 | 1.00e-06 | 0.01491 | 0.08721 | 0.03197 | 0.06337 | 0.06873 | 0.01215 | 0.01441 | 0.06455 |
| RETHE | Hemoglobin [Entitic mass] in Reticulocytes | blood | RETIC HGB EQUIV | Hemoglobin [Entitic mass] in Reticulocytes | RETHE | pg | | 0.2041 | 0.09786 | 2574 | 2403 | 1.00e-06 | 0.1938 | 0.004647 | 1.00e-06 | 0.004648 | 0.1938 | 0.09589 | 0.1021 | 0.01389 | 0.01905 | 0.09639 |
| RetiCt | Reticulocytes/100 erythrocytes in Blood by Automated count | blood | RET CT | Reticulocytes /100 erythrocytes in Blood by Automated count | RetiCt | % | | 0.1798 | 0.04162 | 3964 | 3575 | 0.1073 | 0.02348 | 1.00e-06 | 9.55e-04 | 0.1073 | 0.02444 | 0.06473 | 0.06926 | 0.01095 | 0.01298 | 0.06573 |
| RetPal | Retinyl palmitate [ Mass/volume] in Serum or Plasma | metabolic | RETINYL PALMINATE | Retinyl palmitate [Mass/volume] in Serum or Plasma | RetPal | mg/L | | 0.2215 | 0.2875 | 1135 | 631 | 0.01004 | | 1.00e-06 | | 0.01004 | | 0.2179 | | 0.047 | | 0.2215 |
| RR INTERVAL | RR interval | cardiovascular | R-R interval by EKG | R-R interval by EKG | RR.INTERVAL | | | 0.01241 | 0.9093 | 7229 | 6317 | 0.06807 | 0.01663 | 0.004031 | 1.00e-06 | 0.0721 | 0.01663 | 0.03526 | 0.04005 | 0.006537 | 0.007032 | 0.03581 |
| RV_PPR | RV PPR | lung | RV_PPR | | RV_PPR | % | | 0.0456 | 0.7366 | 1045 | 869 | 0.361 | 1.00e-06 | 1.00e-06 | 0.05658 | 0.361 | 0.05658 | 0.2326 | 0.284 | 0.04195 | 0.05372 | 0.2355 |
| RV_PRD | RV PRD | lung | RV_PRD | | RV_PRD | L | | 0.1806 | 0.2462 | 1046 | 868 | 0.2167 | 0.3372 | 1.00e-06 | 1.00e-06 | 0.2167 | 0.3372 | 0.2349 | 0.2927 | 0.04005 | 0.04913 | 0.2369 |
| RV_PRL | RV PRL | lung | RV_PRL | | RV_PRL | | | 0.1877 | 0.1844 | 1147 | 1145 | 0.392 | 0.08622 | 0.02098 | 0.01898 | 0.413 | 0.1052 | 0.2211 | 0.2133 | 0.03898 | 0.03875 | 0.2233 |
| RV_PRU | RV PRU | lung | RV_PRU | | RV_PRU | | | 0.1212 | 0.3615 | 1143 | 1144 | 0.4097 | 0.2328 | 1.00e-06 | 0.02107 | 0.4097 | 0.2539 | 0.223 | 0.2134 | 0.03506 | 0.0385 | 0.2241 |
| RV_ZBD | RV ZBD | lung | RV_ZBD | | RV_ZBD | L | | -0.004587 | 0.9756 | 1043 | 868 | 0.5144 | 1.00e-06 | 1.00e-06 | 0.04972 | 0.5144 | 0.04972 | 0.2339 | 0.2887 | 0.03812 | 0.05398 | 0.2362 |
| RVOTLC_PPR | RVOTLC PPR | lung | RVOTLC_PPR | | RVOTLC_PPR | % | | 0.04417 | 0.7245 | 1046 | 869 | 0.1543 | 1.00e-06 | 1.00e-06 | 0.06718 | 0.1543 | 0.06718 | 0.2328 | 0.2827 | 0.03878 | 0.05539 | 0.2353 |
| RVOTLC_PRD | RVOTLC PRD | lung | RVOTLC_PRD | | RVOTLC_PRD | % | | 0.2809 | 0.7575 | 1046 | 869 | | 0.06605 | | 0.07427 | | 0.1403 | | 0.266 | | 0.06349 | |
| RVOTLC_PRL | RVOTLC PRL | lung | RVOTLC_PRL | | RVOTLC_PRL | | | 0.3515 | 0.00551 | 1147 | 1145 | 0.3532 | | 0.02847 | | 0.3816 | | 0.2231 | | 0.04318 | | 0.2261 |
| RVOTLC_PRU | RVOTLC PRU | lung | RVOTLC_PRU | | RVOTLC_PRU | | | -0.1739 | 0.2337 | 949 | 926 | 1.00e-06 | 1.00e-06 | 0.04484 | 0.01981 | 0.04484 | 0.01981 | 0.2615 | 0.2606 | 0.05374 | 0.05319 | 0.2675 |
| RVOTLC_ZBD | RVOTLC ZBD | lung | RVOTLC_ZBD | | RVOTLC_ZBD | % | | 0.01228 | 0.9122 | 1046 | 869 | 0.1835 | 1.00e-06 | 1.00e-06 | 0.07381 | 0.1835 | 0.07381 | 0.2323 | 0.2829 | 0.03851 | 0.05669 | 0.2348 |
| RVVT | dRVVT actual/normal (normalized LA screen) | immune / inflammatory | DRVVT | dRVVT actual/normal (normalized LA screen) | RVVT | {ratio} | | 0.1636 | 0.2814 | 1518 | 535 | 1.00e-06 | | 0.006789 | | 0.00679 | | 0.1539 | | 0.02727 | | 0.1563 |
| S-T HORIZONTAL AXIS | ST horizontal axis | cardiovascular | ST segment axis.horizontal plane | ST segment axis.horizontal plane | S.T.HORIZONTAL.AXIS | | | 0.2264 | 0.007039 | 7039 | 6224 | 0.08733 | 0.03644 | 0.009308 | 0.002061 | 0.09663 | 0.0385 | 0.03701 | 0.03975 | 0.007418 | 0.007229 | 0.03775 |
| SclAb | SCL-70 extractable nuclear Ab [ Units/volume] in Serum by Immunoassay | immune / inflammatory | SCL-70 AUTOABS EIA | SCL-70 extractable nuclear Ab [ Units/volume] in Serum by Immunoassay | SclAb | | | -0.7959 | 0.02485 | 772 | 237 | 0.3844 | 0.4213 | 1.00e-06 | 1.00e-06 | 0.3844 | 0.4213 | 0.3475 | 1.066 | 0.05948 | 0.2237 | 0.3526 |
| SedRat | Erythrocyte sedimentation rate by Westergren method | blood | SED RATE | Erythrocyte sedimentation rate | SedRat | mm/h | | 0.2726 | 0.002369 | 13581 | 8621 | 0.1175 | 0.1242 | 0.007566 | 0.002591 | 0.125 | 0.1268 | 0.02012 | 0.03087 | 0.003866 | 0.00576 | 0.0204 |

| all_labs | full_name | group | concept_name | LOINC_NAME | heritability_names | selected_unit | sex_specific_lab | genetic.correlation | genetic.correlation.p | gcta.N_female | gcta.N_male | gcta.Variance_Autosomal_h2_female | gcta.Variance_Autosomal_h2_male | gcta.Variance_X_chromosome_h2_female | gcta.Variance_X_chromosome_h2_male | gcta.Variance_h2_female | gcta.Variance_h2_male | gcta.SE_Autosomal_h2_female | gcta.SE_Autosomal_h2_male | gcta.SE_X_chromosome_h2_female | gcta.SE_X_chromosome_h2_male | gcta.SE_h2_female |
|---|---|---|---|---|---|---|---|---|---|---|---|---|---|---|---|---|---|---|---|---|---|---|
| SerFol | Folate [Mass/volume] in Serum or Plasma | metabolic | FOLATE SERUM | Folate [Mass/volume] in Serum or Plasma | SerFol | ng/mL | | 0.2014 | 0.01284 | 4968 | 3545 | 0.1415 | 0.1024 | 0.015 | 1.00e-06 | 0.1564 | 0.1024 | 0.05305 | 0.07317 | 0.01066 | 0.01336 | 0.0541 |
| SGBF | Specific gravity of Body fluid | blood | SPECIFIC GRAVITY BF | Specific gravity of Body fluid | SGBF | | | 0.3321 | 0.1581 | 439 | 581 | 1.00e-06 | | 0.1116 | | 0.1116 | | 0.5478 | | 0.1155 | | 0.5576 |

| all_labs | full_name | group | concept_name | LOINC_NAME | heritability_names | selected_unit | sex_specific_lab | genetic.correlation | genetic.correlation.p | gcta.N_female | gcta.N_male | gcta.Variance_Autosomal_h2_female | gcta.Variance_Autosomal_h2_male | gcta.Variance_X_chromosome_h2_female | gcta.Variance_X_chromosome_h2_male | gcta.Variance_h2_female | gcta.Variance_h2_male | gcta.SE_Autosomal_h2_female | gcta.SE_Autosomal_h2_male | gcta.SE_X_chromosome_h2_female | gcta.SE_X_chromosome_h2_male | gcta.SE_h2_female |
|---|---|---|---|---|---|---|---|---|---|---|---|---|---|---|---|---|---|---|---|---|---|---|
| SGOT | SGOT | liver | AST BLOOD | Aspartate aminotransferase [Enzymatic activity/volume] in Serum or Plasma | SGOT | [U]/L | | 0.2474 | 0.001697 | 20991 | 15836 | 0.09034 | 0.03 | 1.77e-04 | 0.003952 | 0.09052 | 0.03395 | 0.01318 | 0.01591 | 0.001962 | 0.003341 | 0.01332 |
| SGPT | SGPT | liver | ALT BLOOD | Alanine aminotransferase [Enzymatic activity/volume] in Serum or Plasma | SGPT | [U]/L | | 0.2184 | 7.75e-04 | 21170 | 15997 | 0.08697 | 0.02855 | 1.00e-06 | 0.004044 | 0.08697 | 0.03259 | 0.01327 | 0.01614 | 0.001963 | 0.003394 | 0.01341 |
| SSAIgG | Sjogrens syndrome-A extractable nuclear IgG Ab [Units/volume] in Serum by Immunoassay | immune / inflammatory | SSA (Ro)(ENA) Ab, IgG | Sjogrens syndrome-A extractable nuclear IgG Ab [Units/volume] in Serum by Immunoassay | SSAIgG | | | 0.02418 | 0.9023 | 1087 | 325 | 0.1929 | | 0.003276 | | 0.1961 | | 0.2241 | | 0.03911 | | 0.2262 |
| SSBIgG | Sjogrens syndrome-B extractable nuclear IgG Ab [Units/volume] in Serum by Immunoassay | immune / inflammatory | SSB (La)(ENA) AB, IgG | Sjogrens syndrome-B extractable nuclear IgG Ab [Units/volume] in Serum by Immunoassay | SSBIgG | | | -0.06398 | 0.8036 | 1003 | 313 | 7.12e-04 | | 1.00e-06 | | 7.13e-04 | | 0.2417 | | 0.04022 | | 0.2437 |
| ST_SEGMENT | ST segment axis | cardiovascular | ST segment axis.horizontal plane | ST segment axis.horizontal plane | ST_SEGMENT | deg | | 0.274 | 3.27e-04 | 21462 | 19279 | 7.79e-04 | 1.00e-06 | 1.00e-06 | 0.001141 | 7.80e-04 | 0.001142 | 0.001574 | 0.001697 | 0.001983 | 0.002335 | 0.002545 |
| SU-CA | Calcium urine | urinary | CALCIUM URINE | Calcium [Mass/volume] in Urine | SU.CA | mg/dL | | 0.3047 | 0.1409 | 893 | 273 | 0.2186 | 0.03947 | 1.00e-06 | 1.00e-06 | 0.2186 | 0.03948 | 0.2894 | 0.9207 | 0.04488 | 0.1779 | 0.293 |
| SU-Cre | Creatinine urine | urinary | CREATININE URINE | Creatinine [Mass/volume] in Urine | SU.Cre | mg/dL | | -0.005635 | 0.97 | 1130 | 549 | 1.00e-06 | | 0.007394 | | 0.007395 | | 0.2154 | | 0.0431 | | 0.2184 |
| SU-K | Potassium urine | urinary | POTASSIUM URINE SPOT | Potassium [Moles/volume] in Urine collected for unspecified duration | SU.K | mmol/L | | -0.02676 | 0.8883 | 704 | 710 | | | | | | | | | | | |

| all_labs | full_name | group | concept_name | LOINC_NAME | heritability_names | selected_unit | sex_specific_lab | genetic.correlation | genetic.correlation.p | gcta.N_female | gcta.N_male | gcta.Variance_Autosomal_h2_female | gcta.Variance_Autosomal_h2_male | gcta.Variance_X_chromosome_h2_female | gcta.Variance_X_chromosome_h2_male | gcta.Variance_h2_female | gcta.Variance_h2_male | gcta.SE_Autosomal_h2_female | gcta.SE_Autosomal_h2_male | gcta.SE_X_chromosome_h2_female | gcta.SE_X_chromosome_h2_male | gcta.SE_h2_female |
|---|---|---|---|---|---|---|---|---|---|---|---|---|---|---|---|---|---|---|---|---|---|---|
| SU-Na | Sodium urine | urinary | SODIUM URINE SPOT | Sodium [Moles/volume] in Urine collected for unspecified duration | SU.Na | mmol/L | | -0.242 | 0.1021 | 3137 | 3527 | | 0.05311 | | 0.007346 | | 0.06046 | | 0.07209 | | 0.01423 | |
| SU-Osm | Urine osmolality | urinary | OSMOLALITY URINE SPOT | Osmolality of Urine | SU.Osm | mosm/kg{H20} | | 0.2033 | 0.1797 | 1587 | 1602 | 0.123 | 1.00e-06 | 0.01311 | 0.009521 | 0.1361 | 0.009522 | 0.1571 | 0.1684 | 0.02634 | 0.02704 | 0.1592 |
| SU-Pro | Protein urine | urinary | PROTEIN URINE SPOT | Protein [Mass/volume] in 24 hour Urine | SU.Pro | mg/dL | | 0.1347 | 0.1977 | 3560 | 3467 | 0.02878 | 0.01903 | 0.01894 | 1.00e-06 | 0.04772 | 0.01903 | 0.06876 | 0.06916 | 0.01407 | 0.01318 | 0.07043 |
| SU-Ure | Urea urine | urinary | UREA NITROGEN UR SPOT | Ureanitrogen [Mass/volume] in Urine | SU.Ure | mg/dL | | 0.2289 | 0.008379 | 1351 | 1859 | 1.00e-06 | 0.2399 | 0.035 | 0.02105 | 0.035 | 0.2609 | 0.1989 | 0.1406 | 0.03199 | 0.02611 | 0.2007 |
| SUCrea | Creatinine [Mass/volume] in Urine | urinary | CREATININE URINE ""SPOT"" | Creatinine [Mass/volume] in Urine collected for unspecified duration | SUCrea | mg/dL | | 0.185 | 0.005907 | 8019 | 7600 | 0.06271 | 0.1079 | 1.00e-06 | 1.00e-06 | 0.06272 | 0.1079 | 0.03095 | 0.03458 | 0.005586 | 0.006266 | 0.03143 |
| SyMnCl | Mononuclear cells/100 leukocytes in Synovial fluid by Manual count | immune / inflammatory | MONONUCLEAR CELLS SYN FLD | Mononuclear cells/100 leukocytes in Synovial fluid by Manual count | SyMnCl | % | | -0.9978 | 0.5366 | 457 | 584 | 0.2217 | 1.00e-06 | 1.00e-06 | 0.09219 | 0.2217 | 0.09219 | 0.5885 | 0.5388 | 0.09472 | 0.1007 | 0.5878 |
| SYSTOLIC | Systolic blood pressure | vitals | Systolic blood pressure | Systolic blood pressure | SYSTOLIC | mm[Hg] | | 0.3038 | 7.39e-07 | 35815 | 26956 | 0.08041 | 0.05586 | 0.00267 | 0.001651 | 0.08308 | 0.05751 | 0.008391 | 0.01027 | 0.001536 | 0.001744 | 0.008526 |
| T HORIZONTAL AXIS | T horizontal axis | cardiovascular | T wave axis. horizontal plane Reference beat | T wave axis. horizontal plane Reference beat | T.HORIZONTAL.AXIS | | | -0.04719 | 0.5463 | 7147 | 6265 | 0.128 | 0.05295 | 1.00e-06 | 0.006647 | 0.128 | 0.05959 | 0.03679 | 0.04107 | 0.006711 | 0.007912 | 0.0374 |
| T WAVE AXIS | T wave axis | cardiovascular | T wave axis | T wave axis | T.WAVE.AXIS | | | 0.08284 | 0.4388 | 7207 | 6303 | 0.04169 | 1.00e-06 | 0.01033 | 0.005262 | 0.05202 | 0.005263 | 0.03456 | 0.03893 | 0.007381 | 0.007091 | 0.0352 |
| T_WAVE | T wave axis | cardiovascular | T wave axis | T wave axis | T_WAVE | deg | | 0.1126 | 0.2026 | 21462 | 19279 | 4.02e-04 | | 0.002775 | | 0.003177 | | 0.001565 | | 0.002398 | | 0.002864 |
| T3FRE | Triiodothyronine (T3) Free [Mass/volume] in Serum or Plasma | endocrine | TRIIODOTHYRONINE FREE | Triiodothyronine (T3) Free [Mass/volume] in Serum or Plasma | T3FRE | pg/mL | | 0.2741 | 0.1557 | 1107 | 547 | | | | | | | | | | | |
| TBil | Total Bilirubin serum/plasma | metabolic | BILIRUBIN TOTAL BLOOD | Bilirubin.total [Mass/volume] in Serum or Plasma | TBil | mg/dL | | 0.7079 | 2.59e-18 | 31936 | 24820 | 0.1744 | 0.1547 | 1.00e-06 | 7.21e-04 | 0.1744 | 0.1554 | 0.01025 | 0.01247 | 0.001512 | 0.00187 | 0.01036 |
| TC:HDL | TC/HDL ratio | metabolic | TC:HDL RATIO | Cholesterol.total/Cholesterol in HDL [Molar ratio] in Serum or Plasma | TC.HDL | | | 0.01156 | 0.9283 | 2113 | 2451 | 0.216 | 0.1371 | 0.002435 | 0.003308 | 0.2184 | 0.1404 | 0.1173 | 0.1031 | 0.0207 | 0.01881 | 0.1186 |
| Temper | Body temperature | vitals | Body temperature | Body temperature | Temper | [degF] | | 0.3655 | 9.87e-07 | 32524 | 24959 | 0.02623 | 0.03567 | 1.00e-06 | 1.00e-06 | 0.02624 | 0.03567 | 0.008139 | 0.01044 | 0.001434 | 0.001866 | 0.008265 |
| TEMPERATURE | Body temperature | vitals | Body temperature | Body temperature | TEMPERATURE | Cel | | 0.2339 | 0.01579 | 17589 | 11789 | 0.01732 | 0.01054 | 0.002382 | 1.00e-06 | 0.01971 | 0.01054 | 0.0144 | 0.02027 | 0.002789 | 0.003887 | 0.01464 |

| all_labs | full_name | group | concept_name | LOINC_NAME | heritability_names | selected_unit | sex_specific_lab | genetic.correlation | genetic.correlation.p | gcta.N_female | gcta.N_male | gcta.Variance_Autosomal_h2_female | gcta.Variance_Autosomal_h2_male | gcta.Variance_X_chromosome_h2_female | gcta.Variance_X_chromosome_h2_male | gcta.Variance_h2_female | gcta.Variance_h2_male | gcta.SE_Autosomal_h2_female | gcta.SE_Autosomal_h2_male | gcta.SE_X_chromosome_h2_female | gcta.SE_X_chromosome_h2_male | gcta.SE_h2_female |
|---|---|---|---|---|---|---|---|---|---|---|---|---|---|---|---|---|---|---|---|---|---|---|
| TERMINAL_40_MS | Terminal 40 ms | cardiovascular | | | TERMINAL_40_MS | | | 0.2175 | 0.001675 | 21462 | 19279 | 0.001403 | 0.002426 | 0.002802 | 0.002641 | 0.004205 | 0.005067 | 0.001701 | 0.001943 | 0.002491 | 0.002641 | 0.00301 |
| TestFr | Testosterone Free [Mass/volume] in Serum or Plasma | endocrine | TESTOSTERONE FREE | Testosterone Free [Mass/volume] in Serum or Plasma | TestFr | pg/mL | | 0.7492 | 0.348 | 137 | 2215 | 1.00e-06 | 0.1049 | 0.03156 | 1.00e-06 | 0.03156 | 0.1049 | 2.062 | 0.1127 | 0.3835 | 0.01872 | 2.085 |
| Testos | Testosterone total | endocrine | TESTOSTERONE | Testosterone [Mass/volume] in Serum or Plasma | Testos | ng/dL | | 0.008491 | 0.9412 | 1067 | 3626 | 1.00e-06 | 0.04808 | 0.00903 | 0.007843 | 0.009031 | 0.05592 | 0.224 | 0.0699 | 0.04197 | 0.01325 | 0.2284 |
| TGG | Total gamma globulin | immune / inflammatory | TOTAL GAMMA GLOBULIN | Gamma globulin [Mass/volume] in Urine by Electrophoresis | TGG | mg/dL | | 0.5267 | 0.5245 | 434 | 581 | 0.188 | | 0.1468 | | 0.3349 | | 0.5622 | | 0.1071 | | 0.5697 |
| Thiamn | Thiamine pyrophosphate [Moles/volume] in Blood | metabolic | VITAMIN B1 | Thiamine pyrophosphate [Moles/volume] in Blood | Thiamn | nmol/L | | 0.06776 | 0.5898 | 1229 | 696 | 0.1931 | 0.1116 | 0.007723 | 1.00e-06 | 0.2008 | 0.1116 | 0.2029 | 0.37 | 0.03367 | 0.06905 | 0.2049 |
| THM | Total hemoglobin | blood | TOTAL HEMOGLBIN | Hemoglobin [Mass/volume] in Blood | THM | g/dL | | 0.05462 | 0.617 | 4099 | 5720 | 1.00e-06 | 0.03201 | 7.18e-04 | 1.00e-06 | 7.19e-04 | 0.03201 | 0.05985 | 0.04319 | 0.01002 | 0.008485 | 0.06046 |
| Thyrgb | Thyroglobulin [Mass/volume] in Serum or Plasma | endocrine | THYROGLOBULIN SERUM | Thyroglobulin [Mass/volume] in Serum or Plasma | Thyrgb | ng/mL | | 0.1105 | 0.7027 | 813 | 275 | 0.01226 | | 1.00e-06 | | 0.01226 | | 0.31 | | 0.05484 | | 0.3176 |
| TIBC | Total iron binding capacity | blood | TIBC BLOOD | Iron binding capacity [Mass/volume] in Serum or Plasma | TIBC | ug/mL | | 0.06221 | 0.5113 | 8759 | 6750 | | | | | | | | | | | |
| TIGIgG | Tissue transglutaminase IgG Ab [Units/volume] in Serum by Immunoassay | immune / inflammatory | IgG TISS TRNSGLUTAMIN AB | Tissue transglutaminase IgG Ab [Units/volume] in Serum by Immunoassay | TIGIgG | | | 0.2554 | 0.7467 | 757 | 266 | 0.3578 | | 1.00e-06 | | 0.3578 | | 0.3094 | | 0.05202 | | 0.3118 |
| TLC_PPR | TLC PPR | lung | TLC_PPR | | TLC_PPR | % | | 0.03091 | 0.8112 | 1045 | 869 | 0.6356 | 0.003265 | 1.00e-06 | 1.00e-06 | 0.6356 | 0.003266 | 0.2333 | 0.2797 | 0.03993 | 0.0507 | 0.2362 |
| TLC_PRD | TLC PRD | lung | TLC_PRD | | TLC_PRD | L | | 0.1742 | 0.2953 | 1046 | 868 | 0.2802 | 0.354 | 1.00e-06 | 0.002718 | 0.2802 | 0.3567 | 0.2371 | 0.2937 | 0.03943 | 0.04952 | 0.2384 |
| TLC_PRL | TLC PRL | lung | TLC_PRL | | TLC_PRL | | | 0.2334 | 0.3502 | 987 | 955 | 0.1902 | 0.1032 | 0.03015 | 1.00e-06 | 0.2203 | 0.1032 | 0.2553 | 0.2586 | 0.0445 | 0.04452 | 0.2577 |
| TLC_PRU | TLC PRU | lung | TLC_PRU | | TLC_PRU | | | 0.4608 | 0.3343 | 987 | 954 | 0.2396 | 0.3259 | 0.02165 | 1.00e-06 | 0.2613 | 0.3259 | 0.2622 | 0.2621 | 0.04198 | 0.04647 | 0.263 |
| TLC_ZBD | TLC ZBD | lung | TLC_ZBD | | TLC_ZBD | L | | 0.08747 | 0.4798 | 1045 | 869 | 0.6164 | 0.2431 | 1.00e-06 | 1.00e-06 | 0.6165 | 0.2431 | 0.239 | 0.2799 | 0.03825 | 0.05243 | 0.2409 |
| TotVl | Volume in Urine collected for unspecified duration | toxicology / pharmacology | TOTAL VOLUME | Volume in Urine collected for unspecified duration | TotVl | mL | | -0.03451 | 0.7268 | 3739 | 2231 | 0.008163 | 1.00e-06 | 0.002973 | 0.001262 | 0.01114 | 0.001263 | 0.06639 | 0.1143 | 0.01168 | 0.01976 | 0.06733 |
| TProSe | Protein serum/plasma | metabolic | PROTEIN TOTAL BLOOD | Protein [Mass/volume] in Serum or Plasma | TProSe | g/dL | | 0.1785 | 0.04537 | 20773 | 15800 | 0.08697 | 0.06924 | 1.00e-06 | 1.00e-06 | 0.08698 | 0.06924 | 0.01329 | 0.0162 | 0.00204 | 0.003 | 0.01343 |
| TProt | Total protein | blood | PROTEIN TOTAL BLOOD | Tau protein [Presence] in Body fluid | TProt | g/dL | | 0.358 | 1.47e-06 | 25624 | 19923 | 0.08601 | 0.09828 | 9.80e-04 | 1.00e-06 | 0.08699 | 0.09828 | 0.01086 | 0.01385 | 0.001596 | 0.002384 | 0.01096 |

| all_labs | full_name | group | concept_name | LOINC_NAME | heritability_names | selected_unit | sex_specific_lab | genetic.correlation | genetic.correlation.p | gcta.N_female | gcta.N_male | gcta.Variance_Autosomal_h2_female | gcta.Variance_Autosomal_h2_male | gcta.Variance_X_chromosome_h2_female | gcta.Variance_X_chromosome_h2_male | gcta.Variance_h2_female | gcta.Variance_h2_male | gcta.SE_Autosomal_h2_female | gcta.SE_Autosomal_h2_male | gcta.SE_X_chromosome_h2_female | gcta.SE_X_chromosome_h2_male | gcta.SE_h2_female |
|---|---|---|---|---|---|---|---|---|---|---|---|---|---|---|---|---|---|---|---|---|---|---|
| Trigs | Triglyceride [Mass/volume] in Serum or Plasma | metabolic | TRIGLYCERIDES BLOOD | Triglyceride [Mass/volume] in Serum or Plasma | Trigs | mg/dL | | 0.637 | 1.73e-35 | 19112 | 15431 | 0.2236 | 0.1574 | 0.001469 | 0.003917 | 0.225 | 0.1613 | 0.016 | 0.01854 | 0.002561 | 0.003335 | 0.01618 |
| Trnsfr | Transferrin [Mass/volume] in Serum or Plasma | metabolic | TRANSFERRIN BLOOD | Transferrin [Mass/volume] in Serum or Plasma | Trnsfr | mg/dL | | -0.04182 | 0.7242 | 1070 | 1101 | 0.0704 | 0.01137 | 0.03947 | 2.07e-04 | 0.1099 | 0.01158 | 0.2245 | 0.2323 | 0.0429 | 0.03604 | 0.2277 |
| TropnT | Troponin T.cardiac [Mass/volume] in Serum or Plasma | cardiovascular | TROPONIN T | Troponin T.cardiac [Mass/volume] in Serum or Plasma | TropnT | ng/mL | | 0.2426 | 0.1583 | 593 | 820 | 0.05455 | 0.1638 | 0.02305 | 1.00e-06 | 0.07761 | 0.1638 | 0.3811 | 0.3116 | 0.07154 | 0.05964 | 0.3877 |
| TRPI | Troponin I.cardiac [Mass/volume] in Serum or Plasma | cardiovascular | TROPONIN-I | Troponin I.cardiac [Mass/volume] in Serum or Plasma | TRPI | ng/mL | | -0.005314 | 0.9565 | 5855 | 7073 | 0.001342 | 0.05093 | 1.00e-06 | 0.001071 | 0.001343 | 0.052 | 0.03868 | 0.03601 | 0.007094 | 0.006091 | 0.03934 |
| TSH_3RD_GENERATION | Thyroid stimulating hormone (3rd generation) | endocrine | TSH_3RD_GENERATION | | TSH_3RD_GENERATION | 10*-6.[iU]/mL | | 0.1962 | 0.4846 | 831 | 325 | | | | | | | | | | | |
| TSH | Thyroid stimulating hormone | endocrine | THYROID STIM HORMONE | Thyrotropin [Units/volume] in Serum or Plasma | TSH | | | 0.3974 | 5.26e-07 | 26093 | 16812 | 0.107 | 0.1388 | 1.00e-06 | 1.00e-06 | 0.107 | 0.1388 | 0.01124 | 0.01731 | 0.001698 | 0.002882 | 0.01136 |
| TT4 | Thyroxine (T4) [Mass/volume] in Serum or Plasma | endocrine | TOTAL T4 | Thyroxine (T4) [Mass/volume] in Serum or Plasma | TT4 | ug/dL | | -0.004758 | 0.9636 | 3565 | 2286 | | 0.1353 | | 0.03059 | | 0.1659 | | 0.109 | | 0.02309 | |
| TTAIgA | Tissue transglutaminase IgA Ab [Units/volume] in Serum by Immunoassay | immune / inflammatory | IgA TISS TRNSGLUTAMIN AB | Tissue transglutaminase IgA Ab [Units/volume] in Serum by Immunoassay | TTAIgA | | | 0.03576 | 0.8708 | 820 | 294 | 0.516 | | 0.004195 | | 0.5202 | | 0.2901 | | 0.04428 | | 0.2934 |
| TU-Pro | 24 hour urinary protein | urinary | PROTEIN URINE TIMED | Protein [Mass/volume] in 24 hour Urine | TU.Pro | g/L | | -0.09215 | 0.4695 | 1324 | 784 | 0.02356 | | 0.0106 | | 0.03417 | | 0.191 | | 0.02783 | | 0.1928 |
| TUCrea | Creatinine [Mass/volume] in Urine | kidney | CREATININE URINE ""TIMED"" | Creatinine [Mass/volume] in Urine | TUCrea | g/L | | -0.002977 | 0.9788 | 1415 | 1069 | 9.20e-04 | 1.00e-06 | 1.00e-06 | 0.08417 | 9.21e-04 | 0.08418 | 0.1702 | 0.2307 | 0.03162 | 0.05138 | 0.1738 |
| UABM | Urinary albumin | kidney | ALBUMIN | Albumin [Mass/volume] in Urine by Electrophoresis | UABM | mg/dL | | 0.2469 | 0.2092 | 585 | 723 | 0.3967 | 0.07758 | 0.1053 | 1.00e-06 | 0.502 | 0.07758 | 0.4331 | 0.3448 | 0.07729 | 0.04859 | 0.4435 |
| UAlb | Albumin [Presence] in Urine | urinary | ALBUMIN URINE | Albumin [Presence] in Urine | UAlb | | | -0.3195 | 0.538 | 569 | 602 | 0.1051 | 1.00e-06 | 1.00e-06 | 0.09778 | 0.1051 | 0.09778 | 0.4622 | 0.4882 | 0.06542 | 0.08768 | 0.4657 |
| UGluc | Glucose urine dipstick | urinary | GLUCOSE URINE | Glucose [Presence] in Urine by Automated test strip | UGluc | mg/dL | | -0.03033 | 0.7802 | 2253 | 2730 | 1.00e-06 | 1.00e-06 | 0.02056 | 0.001317 | 0.02056 | 0.001318 | 0.105 | 0.08999 | 0.02257 | 0.01575 | 0.107 |

| all_labs | full_name | group | concept_name | LOINC_NAME | heritability_names | selected_unit | sex_specific_lab | genetic.correlation | genetic.correlation.p | gcta.N_female | gcta.N_male | gcta.Variance_Autosomal_h2_female | gcta.Variance_Autosomal_h2_male | gcta.Variance_X_chromosome_h2_female | gcta.Variance_X_chromosome_h2_male | gcta.Variance_h2_female | gcta.Variance_h2_male | gcta.SE_Autosomal_h2_female | gcta.SE_Autosomal_h2_male | gcta.SE_X_chromosome_h2_female | gcta.SE_X_chromosome_h2_male | gcta.SE_h2_female |
|---|---|---|---|---|---|---|---|---|---|---|---|---|---|---|---|---|---|---|---|---|---|---|
| UGrCst | Granular casts [#/area] in Urine sediment by Microscopy high power field | urinary | GRANULAR CAST | Granular casts [#/area] in Urine sediment by Microscopy high power field | UGrCst | /[HPF] | | 0.6848 | 5.39e-04 | 489 | 673 | 0.1487 | 0.4225 | 1.00e-06 | 1.00e-06 | 0.1487 | 0.4225 | 0.4528 | 0.3921 | 0.08278 | 0.06966 | 0.4573 |
| UHyCst | Hyaline casts in urine | urinary | HYALINE CAST | Hyaline casts [#/area] in Urine sediment | UHyCst | /[HPF] | | 0.03158 | 0.7655 | 3554 | 3258 | 0.002642 | 1.00e-06 | 1.00e-06 | 0.01123 | 0.002643 | 0.01123 | 0.06472 | 0.07892 | 0.01393 | 0.01426 | 0.066 |
| UKeto | Ketones [Mass/volume] in Urine by Test strip | urinary | KETONE URINE | Ketones [Mass/volume] in Urine by Test strip | UKeto | mg/dL | | -0.05224 | 0.6295 | 3704 | 2477 | 0.0438 | 1.00e-06 | 0.007139 | 0.01226 | 0.05094 | 0.01227 | 0.06516 | 0.106 | 0.01342 | 0.02032 | 0.06644 |
| UpH | pH of Urine by Test strip | urinary | PH URINE | pH of Specimen | UpH | | | 0.2235 | 0.01134 | 24121 | 18624 | 0.05897 | 0.04872 | 0.003166 | 0.002055 | 0.06214 | 0.05077 | 0.01141 | 0.01417 | 0.002297 | 0.002592 | 0.01161 |
| URBC | Erythrocytes [#/area] in Urine sediment by Microscopy high power field | urinary | RBC URINE MICRO | Erythrocytes [#/area] in Urine sediment by Microscopy high power field | URBC | /[HPF] | | -0.00601 | 0.9513 | 15324 | 11084 | 0.03974 | 0.04426 | 1.00e-06 | 0.003561 | 0.03974 | 0.04782 | 0.01716 | 0.02299 | 0.003352 | 0.00438 | 0.01745 |
| UricA | Urate [Mass/volume] in Serum or Plasma | urinary | URIC ACID BLOOD | Urate [Mass/volume] in Serum or Plasma | UricA | mg/dL | | 0.2997 | 0.07501 | 6711 | 6411 | 0.08796 | 0.06839 | 1.00e-06 | 1.00e-06 | 0.08796 | 0.0684 | 0.03786 | 0.039 | 0.007418 | 0.007127 | 0.03858 |
| UrobUr | Urobilinogen [Units/volume] in Urine by Test strip | urinary | UROBILINOGEN | Urobilinogen [Units/volume] in Urine by Test strip | UrobUr | [EIA'U] | | 0.03048 | 0.7444 | 3025 | 2412 | | | | | | | | | | | |
| UrPro | Protein [Mass/volume] in Urine by Test strip | urinary | PROTEIN URINE | Protein [Mass/volume] in Urine by Test strip | UrPro | mg/dL | | -0.05745 | 0.4668 | 9898 | 8932 | 0.01493 | 0.02664 | 0.002089 | 0.002418 | 0.01702 | 0.02906 | 0.02504 | 0.02647 | 0.004654 | 0.005433 | 0.02537 |
| USpGr | Specific gravity of Urine by Test strip | urinary | SPECIFIC GRAVITY UA | Specific gravity of Urine by Test strip | USpGr | | | 0.2065 | 0.06 | 24468 | 18338 | 0.02303 | 0.04414 | 0.006229 | 0.002005 | 0.02926 | 0.04615 | 0.01071 | 0.01446 | 0.002497 | 0.002678 | 0.01098 |
| USqEpi | Epithelial cells.squamous [#/area] in Urine sediment by Microscopy high power field | urinary | SQUAMOUS EPITHELIAL | Epithelial cells.squamous [Presence] in Urine by Automated | USqEpi | /[HPF] | | -0.1259 | 0.2025 | 14841 | 3944 | 0.03011 | 1.00e-06 | 1.00e-06 | 0.002936 | 0.03011 | 0.002937 | 0.01721 | 0.06301 | 0.002893 | 0.01101 | 0.01745 |
| UTP | Total urine protein | urinary | TOTAL PROTEIN URINE | | UTP | g/L | | 0.2468 | 0.6902 | 869 | 808 | 1.00e-06 | | 0.08006 | | 0.08006 | | 0.2845 | | 0.04992 | | 0.2881 |

| all_labs | full_name | group | concept_name | LOINC_NAME | heritability_names | selected_unit | sex_specific_lab | genetic.correlation | genetic.correlation.p | gcta.N_female | gcta.N_male | gcta.Variance_Autosomal_h2_female | gcta.Variance_Autosomal_h2_male | gcta.Variance_X_chromosome_h2_female | gcta.Variance_X_chromosome_h2_male | gcta.Variance_h2_female | gcta.Variance_h2_male | gcta.SE_Autosomal_h2_female | gcta.SE_Autosomal_h2_male | gcta.SE_X_chromosome_h2_female | gcta.SE_X_chromosome_h2_male | gcta.SE_h2_female |
|---|---|---|---|---|---|---|---|---|---|---|---|---|---|---|---|---|---|---|---|---|---|---|
| UUrob | Urobilinogen [Presence] in Urine by Test strip | urinary | UROBILINOGEN | Urobilinogen [Presence] in Urine by Test strip | UUrob | mg/dL | | -0.1063 | 0.3488 | 4995 | 4911 | | 1.00e-06 | | 0.01348 | | 0.01348 | | 0.04707 | | 0.01028 | |
| UWBC | Leukocytes [#/area] in Urine sediment by Microscopy high power field | urinary | WBC URINE MICRO | Leukocytes [#/volume] in Blood by Automated count | UWBC | /[HPF] | | -0.02362 | 0.8088 | 16299 | 11007 | 0.04802 | 0.01016 | 1.00e-06 | 1.00e-06 | 0.04802 | 0.01016 | 0.01578 | 0.02286 | 0.002921 | 0.004278 | 0.01604 |
| VA_BTPS_PPR | VA BTPS PPR | lung | VA_BTPS_PPR | | VA_BTPS_PPR | | | 0.1444 | 0.1706 | 1227 | 1269 | 0.01161 | 0.02974 | 1.00e-06 | 1.00e-06 | 0.01161 | 0.02974 | 0.2024 | 0.1878 | 0.038 | 0.03692 | 0.2072 |
| VA_BTPS_PRD | VA BTPS PRD | lung | VA_BTPS_PRD | | VA_BTPS_PRD | | | 0.0951 | 0.4412 | 1226 | 1268 | 0.4216 | 0.3765 | 0.008756 | 0.0243 | 0.4304 | 0.4008 | 0.2115 | 0.1927 | 0.03467 | 0.03446 | 0.2123 |
| VA_BTPS_PRL | VA BTPS PRL | lung | VA_BTPS_PRL | | VA_BTPS_PRL | | | 0.03632 | 0.7561 | 1226 | 1268 | 0.5715 | 0.2924 | 0.01255 | 0.01412 | 0.584 | 0.3065 | 0.2097 | 0.191 | 0.03279 | 0.03137 | 0.2102 |
| VA_BTPS_PRU | VA BTPS PRU | lung | VA_BTPS_PRU | | VA_BTPS_PRU | | | 0.1184 | 0.3459 | 1222 | 1267 | 0.4099 | 0.4587 | 0.008425 | 0.04066 | 0.4183 | 0.4994 | 0.2122 | 0.1942 | 0.03588 | 0.03837 | 0.213 |
| VA_BTPS_ZBD | VA BTPS ZBD | lung | VA_BTPS_ZBD | | VA_BTPS_ZBD | | | 0.1284 | 0.1332 | 1227 | 1268 | | 0.05357 | | 1.00e-06 | | 0.05357 | | 0.1922 | | 0.03503 | |
| VC_PPR | VC PPR | lung | VC_PPR | | VC_PPR | | | 0.1437 | 0.2382 | 1146 | 1145 | 1.00e-06 | | 0.01362 | | 0.01362 | | 0.2229 | | 0.04327 | | 0.2272 |
| VC_PRD | VC PRD | lung | VC_PRD | | VC_PRD | | | 0.1662 | 0.1394 | 1147 | 1144 | 0.4254 | 0.2894 | 0.01162 | 0.006329 | 0.4371 | 0.2957 | 0.2216 | 0.2148 | 0.03542 | 0.03734 | 0.2229 |
| VC_PRL | VC PRL | lung | VC_PRL | | VC_PRL | | | 0.1949 | 0.1357 | 1147 | 1144 | 0.4266 | 0.2657 | 0.01109 | 0.01083 | 0.4377 | 0.2766 | 0.2211 | 0.2141 | 0.03507 | 0.03798 | 0.2223 |
| VC_ZBD | VC ZBD | lung | VC_ZBD | | VC_ZBD | | | 0.1707 | 0.1872 | 1147 | 1145 | 0.1039 | 0.02929 | 0.04346 | 1.00e-06 | 0.1473 | 0.02929 | 0.2189 | 0.2173 | 0.04388 | 0.04049 | 0.2234 |
| VD25 | Calcitriol [Mass/volume] in Serum or Plasma | metabolic | VITAMIN D 25-OH | Calcitriol [Mass/volume] in Serum or Plasma | VD25 | ng/mL | | 0.05649 | 0.5745 | 2437 | 949 | 0.2173 | 1.00e-06 | 0.01336 | 0.009517 | 0.2307 | 0.009518 | 0.1051 | 0.2889 | 0.01816 | 0.05153 | 0.1066 |
| Vi-B12 | Vitamin B12 | metabolic | VITAMIN B-12 | Cobalamin (VitaminB12) [Mass/volume] in Serum or Plasma | Vi.B12 | pg/mL | | 0.08931 | 0.3303 | 14107 | 9165 | 0.06299 | 0.01449 | 1.00e-06 | 0.004601 | 0.06299 | 0.01909 | 0.01893 | 0.02765 | 0.003119 | 0.005311 | 0.01918 |
| ViARet | Retinol [Mass/volume] in Serum or Plasma | metabolic | VITAMIN A (RETINOL) | Retinol [Mass/volume] in Serum or Plasma | ViARet | mg/L | | 0.0521 | 0.6762 | 1396 | 795 | 0.02765 | 1.00e-06 | 0.001832 | 0.01322 | 0.02949 | 0.01322 | 0.1671 | 0.3644 | 0.03441 | 0.06666 | 0.1687 |
| VitaB6 | Pyridoxine [Mass/volume] in Serum or Plasma | metabolic | VITAMIN B6 | Pyridoxal phosphate [Moles/volume] in Serum or Plasma | VitaB6 | nmol/L | | -0.1119 | 0.1699 | 1801 | 836 | 0.2136 | 0.1573 | 1.00e-06 | 0.06609 | 0.2136 | 0.2234 | 0.1402 | 0.2888 | 0.0222 | 0.05995 | 0.1414 |
| VLDL | very-low-density lipoprotein | metabolic | VLDL | | VLDL | | | -0.1133 | 0.3949 | 1142 | 1012 | 0.1818 | 0.3065 | 0.01226 | 1.00e-06 | 0.194 | 0.3065 | 0.2099 | 0.2555 | 0.03387 | 0.05554 | 0.2121 |
| WBC | Leukocytes [#/volume] in Blood by Automated count | immune / inflammatory | WBC | Leukocytes [#/volume] in Blood by Automated count | WBC | 10*3/uL | | 0.4822 | 3.00e-20 | 35673 | 26968 | 0.1176 | 0.1168 | 0.002625 | 1.00e-06 | 0.1202 | 0.1168 | 0.008736 | 0.01102 | 0.001526 | 0.001653 | 0.008847 |
| WBCCnt | Leukocytes [#/volume] in Blood by Manual count | immune / inflammatory | # WBC'S COUNTED | Leukocytes [#/volume] in Blood by Manual count | WBCCnt | | | 0.02102 | 0.83 | 5812 | 5932 | 1.00e-06 | 0.007117 | 9.37e-04 | 0.009348 | 9.38e-04 | 0.01647 | 0.04209 | 0.0419 | 0.007304 | 0.008743 | 0.0426 |

| all_labs | full_name | group | concept_name | LOINC_NAME | heritability_names | selected_unit | sex_specific_lab | genetic.correlation | genetic.correlation.p | gcta.N_female | gcta.N_male | gcta.Variance_Autosomal_h2_female | gcta.Variance_Autosomal_h2_male | gcta.Variance_X_chromosome_h2_female | gcta.Variance_X_chromosome_h2_male | gcta.Variance_h2_female | gcta.Variance_h2_male | gcta.SE_Autosomal_h2_female | gcta.SE_Autosomal_h2_male | gcta.SE_X_chromosome_h2_female | gcta.SE_X_chromosome_h2_male | gcta.SE_h2_female |
|---|---|---|---|---|---|---|---|---|---|---|---|---|---|---|---|---|---|---|---|---|---|---|
| WBCrt | Creatinine [Mass/volume] in Blood | blood | ED CREATININE WBLD | Creatinine [Mass/volume] in Blood | WBCrt | mg/dL | | 0.006173 | 0.9266 | 4458 | 4826 | 0.06875 | 0.07374 | 1.00e-06 | 1.00e-06 | 0.06875 | 0.07374 | 0.05749 | 0.05277 | 0.009462 | 0.009746 | 0.05815 |
| WBINR | International normalized ratio | blood | ED INR WBLD | INR in Platelet poor plasma by Coagulation assay | WBINR | | | 0.05764 | 0.6772 | 1253 | 1463 | 0.1661 | | 1.00e-06 | | 0.1661 | | 0.2025 | | 0.03791 | | 0.2054 |
| WBTrpI | Troponin I.cardiac [Mass/volume] in Blood | cardiovascular | ED TROPONIN-I WBLD | Troponin I.cardiac [Mass/volume] in Blood | WBTrpI | ng/mL | | -0.214 | 0.1714 | 906 | 1106 | 0.01747 | | 1.00e-06 | | 0.01747 | | 0.2602 | | 0.05073 | | 0.265 |
| WEIGHT_SCALE | Weight scale | vitals | Body weight | Body weight | WEIGHT_SCALE | kg | | 0.4032 | 2.92e-11 | 19541 | 12994 | 0.174 | 0.2015 | 1.00e-06 | 0.004788 | 0.174 | 0.2063 | 0.0154 | 0.02218 | 0.002415 | 0.004014 | 0.01557 |
| Weight | Weight | vitals | Body weight | Body weight | Weight | kg | | 0.6112 | 9.54e-36 | 35804 | 26877 | 0.209 | 0.1846 | 0.003541 | 0.003827 | 0.2126 | 0.1884 | 0.009537 | 0.01172 | 0.001642 | 0.001996 | 0.009648 |
| Zinc | Zinc | metabolic | ZINC SERUM | Zinc [Mass/volume] in Serum or Plasma | Zinc | ug/dL | | -0.0817 | 0.5867 | 1051 | 681 | 0.4183 | | 0.02451 | | 0.4428 | | 0.2451 | | 0.04062 | | 0.2498 |

| gcta.SE_h2_male | gcta.h2.conf.low_female | gcta.h2.conf.low_male | gcta.h2.conf.high_female | gcta.h2.conf.high_male | gcta.x.h2.conf.low_female | gcta.x.h2.conf.low_male | gcta.x.h2.conf.high_female | gcta.x.h2.conf.high_male | ldsc.int_female | ldsc.int_male | ldsc.int_se_female | ldsc.int_se_male | ldsc.h2_female | ldsc.h2_male | ldsc.h2_se_female | ldsc.h2_se_male | ldsc.int.conf.high_female | ldsc.int.conf.high_male | ldsc.int.conf.low_female | ldsc.int.conf.low_male | ldsc.h2.conf.high_female | ldsc.h2.conf.high_male |
|---|---|---|---|---|---|---|---|---|---|---|---|---|---|---|---|---|---|---|---|---|---|---|
| 0.3357 | -0.5303 | -0.6017 | 0.6456 | 0.7143 | -0.03658 | -0.1182 | 0.1469 | 0.1182 | 1.006 | 0.9954 | 0.00686 | 0.00905 | 0.1115 | 0.2221 | 0.239 | 0.3929 | 1.02 | 1.013 | 0.9928 | 0.9776 | 0.5799 | 0.9922 |
| 0.03918 | 0.02637 | 0.01171 | 0.1459 | 0.1653 | -0.01085 | -0.01024 | 0.01085 | 0.01254 | 1.003 | 0.995 | 0.008523 | 0.009212 | 0.02372 | 0.06967 | 0.0367 | 0.04803 | 1.02 | 1.013 | 0.9867 | 0.9769 | 0.09564 | 0.1638 |
| 0.2624 | -0.2284 | -0.4388 | 0.2398 | 0.59 | -0.03271 | -0.06969 | 0.03271 | 0.129 | 1.013 | 1.016 | 0.007024 | 0.007699 | -0.2013 | -0.542 | 0.1078 | 0.2438 | 1.027 | 1.031 | 0.9995 | 1.001 | 0.009927 | -0.06412 |
| 0.09317 | 0.06929 | -0.06237 | 0.2207 | 0.3029 | -0.009041 | -0.02984 | 0.01398 | 0.03346 | 1.007 | 0.9876 | 0.007934 | 0.005714 | 0.03777 | 0.1767 | 0.04435 | 0.1333 | 1.023 | 0.9988 | 0.9914 | 0.9764 | 0.1247 | 0.438 |
|  | -0.1282 |  | 0.2911 |  | -0.03764 |  | 0.03764 |  | 0.9945 | 1.007 | 0.006593 | 0.006156 | 0.05282 | -0.03617 | 0.1009 | 0.2928 | 1.007 | 1.019 | 0.9815 | 0.995 | 0.2506 | 0.5378 |
| 0.09174 | -8.56e-04 | -0.03971 | 0.142 | 0.3199 | -0.01053 | -0.02959 | 0.01053 | 0.02959 | 1.003 | 0.9945 | 0.005698 | 0.005791 | 0.04205 | 0.1375 | 0.04186 | 0.07138 | 1.014 | 1.006 | 0.9916 | 0.9831 | 0.1241 | 0.2774 |
| 0.08606 |  | -0.1045 |  | 0.2328 |  | -0.02925 |  | 0.02925 | 1.01 | 1.003 | 0.007182 | 0.007576 | -0.1042 | 0.01525 | 0.07709 | 0.08305 | 1.025 | 1.018 | 0.9964 | 0.9883 | 0.04686 | 0.178 |
| 0.08697 | -0.1029 | -0.05237 | 0.1543 | 0.2885 | -0.01898 | -0.0165 | 0.02412 | 0.04934 | 0.9936 | 1.007 | 0.005161 | 0.004184 | 0.05937 | -0.02759 | 0.05023 | 0.05878 | 1.004 | 1.016 | 0.9835 | 0.9992 | 0.1578 | 0.08763 |
| 0.2473 | -0.4787 | -0.2456 | 0.602 | 0.7239 | -0.07127 | -0.05774 | 0.09748 | 0.1292 | 0.9968 | 0.9973 | 0.008828 | 0.009731 | -0.1226 | 0.4357 | 0.2706 | 0.3545 | 1.014 | 1.016 | 0.9795 | 0.9782 | 0.4078 | 1.131 |
|  | -0.00834 |  | 0.8678 |  | -0.06677 |  | 0.07279 |  | 1.001 | 1 | 0.006453 | 0.005264 | 0.1358 | 0.4196 | 0.2623 | 0.3218 | 1.014 | 1.01 | 0.9886 | 0.9897 | 0.6499 | 1.05 |
|  | -0.2268 |  | 0.7785 |  | -0.05031 |  | 0.1396 |  | 1.004 | 1 | 0.005033 | 0.008997 | 0.06431 | -0.5884 | 0.2241 | 0.5758 | 1.014 | 1.018 | 0.9942 | 0.9825 | 0.5035 | 0.5402 |
| 0.4074 |  | -0.4042 |  | 1.193 |  | -0.1407 |  | 0.1407 | 1.005 | 1.003 | 0.005769 | 0.007798 | -0.1603 | -0.0532 | 0.2536 | 0.4482 | 1.017 | 1.018 | 0.9939 | 0.9878 | 0.3368 | 0.8253 |
| 0.3993 | -0.4114 | -0.25 | 0.5004 | 1.315 | -0.04249 | -0.1561 | 0.1346 | 0.1561 | 1.014 | 1.011 | 0.009961 | 0.009633 | -0.2287 | -0.5388 | 0.3378 | 0.4202 | 1.033 | 1.029 | 0.994 | 0.9916 | 0.4333 | 0.2847 |
| 0.4104 | -0.3519 | -0.7516 | 0.5882 | 0.8573 | -0.07344 | -0.1513 | 0.07344 | 0.1513 | 1.029 | 1.008 | 0.005998 | 0.00765 | -0.7572 | -0.5261 | 0.1632 | 0.4795 | 1.041 | 1.023 | 1.017 | 0.9927 | -0.4373 | 0.4137 |
| 0.3928 |  | -0.3744 |  | 1.165 |  | -0.1589 |  | 0.1589 | 1.014 | 1.013 | 0.008298 | 0.00631 | -0.2162 | -0.9645 | 0.202 | 0.3427 | 1.03 | 1.025 | 0.9979 | 1.001 | 0.1798 | -0.2929 |
|  | -0.2745 |  | 0.3841 |  | -0.03817 |  | 0.07819 |  | 1.009 | 0.9974 | 0.006326 | 0.009557 | -0.2331 | 0.01125 | 0.1324 | 0.5695 | 1.022 | 1.016 | 0.9968 | 0.9787 | 0.02653 | 1.127 |
| 0.3853 | -0.234 | -0.6898 | 0.2927 | 0.8207 | -0.02202 | -0.1317 | 0.08251 | 0.1317 | 1.004 | 0.9925 | 0.00799 | 0.007496 | -0.2237 | 0.2632 | 0.1538 | 0.4054 | 1.02 | 1.007 | 0.9886 | 0.9778 | 0.07786 | 1.058 |
| 0.3902 | -0.04574 | -0.1102 | 0.4874 | 1.419 | -0.04444 | -0.06587 | 0.04444 | 0.2268 | 1.002 | 0.9954 | 0.00586 | 0.006943 | 0.1093 | 0.9711 | 0.1009 | 0.4687 | 1.014 | 1.009 | 0.991 | 0.9818 | 0.3071 | 1.89 |
|  |  |  |  |  |  |  |  |  | 1.002 | 1.008 | 0.006418 | 0.007487 | -0.0549 | -0.3269 | 0.2503 | 0.4222 | 1.014 | 1.022 | 0.9891 | 0.9931 | 0.4356 | 0.5007 |
|  | -0.2017 |  | 0.7463 |  | -0.04635 |  | 0.1332 |  | 1.002 |  | 0.005744 |  | 0.06283 |  | 0.2561 |  | 1.014 |  | 0.991 |  | 0.5648 |  |
|  | -0.1624 |  | 0.6181 |  | -0.05927 |  | 0.05927 |  | 1.002 | 1.001 | 0.008275 | 0.0104 | 0.03295 | 0.08772 | 0.2312 | 0.1776 | 1.018 | 1.022 | 0.9858 | 0.9808 | 0.4861 | 0.4358 |
| 0.01148 | 0.07061 | 0.05168 | 0.1078 | 0.09668 | -0.001576 | -0.003262 | 0.004189 | 0.004113 | 0.9982 | 1.018 | 0.008181 | 0.0047 | 0.06044 | 0.03139 | 0.01116 | 0.01104 | 1.014 | 1.028 | 0.9822 | 1.009 | 0.08231 | 0.05303 |
| 0.07855 | -0.1343 | -0.1416 | 0.148 | 0.1664 | -0.01999 | -0.01652 | 0.02973 | 0.04372 | 0.9991 | 1.008 | 0.008366 | 0.006006 | 0.006396 | -0.1188 | 0.08304 | 0.08635 | 1.016 | 1.02 | 0.9827 | 0.9967 | 0.1691 | 0.05044 |
| 0.1159 | -0.2233 | -0.1477 | 0.2409 | 0.3065 | -0.03175 | -0.03526 | 0.05074 | 0.04539 | 1.01 | 1.007 | 0.006816 | 0.006554 | -0.1552 | 0.0318 | 0.1269 | 0.1104 | 1.024 | 1.02 | 0.997 | 0.9946 | 0.09365 | 0.2481 |
| 0.07681 | -0.04149 | -0.04167 | 0.1972 | 0.2594 | -0.01499 | -0.02928 | 0.02499 | 0.02928 | 0.9938 | 1.006 | 0.008924 | 0.006601 | 0.1007 | 0.01325 | 0.06838 | 0.064 | 1.011 | 1.019 | 0.9763 | 0.9931 | 0.2347 | 0.1387 |
| 0.4278 |  | -0.8367 |  | 0.8401 |  | -0.1557 |  | 0.1557 | 0.9889 | 1.005 | 0.006662 | 0.007618 | 0.008598 | 0.008349 | 0.2312 | 0.5058 | 1.002 | 1.02 | 0.9758 | 0.9901 | 0.4617 | 0.9996 |
| 0.0181 | 0.1334 | 0.09917 | 0.1897 | 0.1701 | -0.002621 | -0.005179 | 0.006194 | 0.005181 | 1.013 | 1.008 | 0.01101 | 0.01094 | 0.1075 | 0.1013 | 0.02132 | 0.03124 | 1.034 | 1.03 | 0.9912 | 0.9868 | 0.1493 | 0.1626 |
| 0.1452 |  | 0.03205 |  | 0.6013 |  | -0.05104 |  | 0.05105 | 1.006 | 1.007 | 0.007752 | 0.005668 | -0.02945 | 0.1521 | 0.1994 | 0.1323 | 1.021 | 1.018 | 0.9909 | 0.9957 | 0.3614 | 0.4114 |
| 0.06528 | -0.02663 | -0.04147 | 0.1819 | 0.2144 | -0.01272 | -0.01408 | 0.02238 | 0.03612 | 0.998 | 0.9963 | 0.008951 | 0.005076 | 0.1109 | 0.07519 | 0.07058 | 0.07614 | 1.015 | 1.006 | 0.9804 | 0.9864 | 0.2492 | 0.2244 |

| gcta.SE_h2_male | gcta.h2.conf.low_female | gcta.h2.conf.low_male | gcta.h2.conf.high_female | gcta.h2.conf.high_male | gcta.x.h2.conf.low_female | gcta.x.h2.conf.low_male | gcta.x.h2.conf.high_female | gcta.x.h2.conf.high_male | ldsc.int_female | ldsc.int_male | ldsc.int_se_female | ldsc.int_se_male | ldsc.h2_female | ldsc.h2_male | ldsc.h2_se_female | ldsc.h2_se_male | ldsc.int.conf.high_female | ldsc.int.conf.high_male | ldsc.int.conf.low_female | ldsc.int.conf.low_male | ldsc.h2.conf.high_female | ldsc.h2.conf.high_male |
|---|---|---|---|---|---|---|---|---|---|---|---|---|---|---|---|---|---|---|---|---|---|---|
| 0.00978 | 0.01051 | 3.34e-04 | 0.04124 | 0.03867 | -0.001306 | -0.001813 | 0.004395 | 0.004908 | 1.015 | 1.005 | 0.008132 | 0.008604 | 0.003299 | 0.01552 | 0.008312 | 0.0125 | 1.031 | 1.022 | 0.9992 | 0.9885 | 0.01959 | 0.04002 |
| 0.1034 | 0.002981 | -0.1665 | 0.2659 | 0.239 | -0.02571 | -0.03543 | 0.02572 | 0.03543 | 0.996 | 1.001 | 0.009325 | 0.007673 | 0.1734 | 0.04348 | 0.08207 | 0.1242 | 1.014 | 1.016 | 0.9778 | 0.986 | 0.3343 | 0.287 |
|  | 0.225 |  | 1.523 |  | -0.1298 |  | 0.1298 |  | 1.004 | 0.9923 | 0.00678 | 0.006028 | 0.4492 | 0.1011 | 0.2924 | 0.5225 | 1.017 | 1.004 | 0.9907 | 0.9805 | 1.022 | 1.125 |
|  | -0.09424 |  | 0.3919 |  | -0.02045 |  | 0.07359 |  | 0.989 | 1.009 | 0.005386 | 0.006217 | 0.1305 | -0.4229 | 0.1068 | 0.3782 | 0.9995 | 1.021 | 0.9784 | 0.9967 | 0.3399 | 0.3183 |
| 0.9903 | -0.2878 | -0.9409 | 0.6152 | 2.941 | -0.05349 | -0.223 | 0.08981 | 0.5472 | 1.005 | 1.004 | 0.005169 | 0.007498 | -0.1013 | -0.2 | 0.2071 | 0.8983 | 1.015 | 1.019 | 0.9944 | 0.9897 | 0.3047 | 1.561 |
| 0.6339 |  | -0.2447 |  | 2.24 |  | 0.05886 |  | 0.5834 | 1.009 | 0.9908 | 0.007202 | 0.006378 | -1.19 | 1.445 | 0.3951 | 0.6658 | 1.023 | 1.003 | 0.9949 | 0.9783 | -0.4159 | 2.75 |
| 1.424 | -0.4594 | -2.694 | 0.4676 | 2.89 | -0.07701 | -0.4393 | 0.08437 | 0.6918 | 1.008 | 0.9442 | 0.008606 | 0.005158 | -0.3908 | 1.232 | 0.3528 | 0.9499 | 1.025 | 0.9543 | 0.9914 | 0.9341 | 0.3008 | 3.093 |
| 0.04122 | -0.03475 | -0.03145 | 0.1022 | 0.1302 | -0.0109 | -0.01383 | 0.01388 | 0.01383 | 0.9946 | 1.011 | 0.008003 | 0.006775 | 0.0289 | -0.03362 | 0.04585 | 0.0332 | 1.01 | 1.024 | 0.9789 | 0.9976 | 0.1188 | 0.03146 |
|  | -0.04729 |  | 0.2079 |  | -0.02311 |  | 0.02312 |  | 1.005 | 0.9911 | 0.007991 | 0.007903 | -0.002427 | 0.03707 | 0.0825 | 0.06189 | 1.021 | 1.007 | 0.9894 | 0.9756 | 0.1593 | 0.1584 |
| 0.2779 | 0.08762 | -0.4453 | 0.8131 | 0.6443 | -0.06368 | -0.08886 | 0.06369 | 0.08886 | 1.001 | 0.9993 | 0.006428 | 0.00717 | 0.168 | 0.1519 | 0.1853 | 0.3485 | 1.014 | 1.013 | 0.9886 | 0.9852 | 0.5313 | 0.8349 |
| 0.02535 | 0.07876 | 0.06076 | 0.153 | 0.1602 | -0.005854 | -0.006854 | 0.005856 | 0.01007 | 1.005 | 1.012 | 0.008222 | 0.007563 | 0.05133 | 0.05954 | 0.02219 | 0.03098 | 1.021 | 1.027 | 0.9886 | 0.9972 | 0.09481 | 0.1203 |
| 0.02362 | 0.07564 | 0.004358 | 0.1478 | 0.09694 | -0.006244 | -0.007504 | 0.006246 | 0.008498 | 1 | 1.013 | 0.008128 | 0.006568 | 0.07286 | -1.17e-04 | 0.02453 | 0.03019 | 1.016 | 1.025 | 0.9844 | 0.9997 | 0.1209 | 0.05905 |
|  | -0.1192 |  | 0.1356 |  | -0.01339 |  | 0.03043 |  | 0.9937 | 1.003 | 0.008363 | 0.006639 | 0.0038 | -0.05275 | 0.06035 | 0.05652 | 1.01 | 1.016 | 0.9773 | 0.9897 | 0.1221 | 0.05804 |
| 0.003094 | 2.51e-04 | -0.001545 | 0.01023 | 0.01058 | -0.002039 | -0.002599 | 0.005301 | 0.006418 | 1.014 | 1.027 | 0.008225 | 0.008725 | 0.07706 | 0.01774 | 0.01149 | 0.01508 | 1.03 | 1.044 | 0.9977 | 1.01 | 0.09957 | 0.0473 |
| 0.06229 |  | -0.1129 |  | 0.1313 |  | -0.02067 |  | 0.02067 | 0.9918 | 1 | 0.007774 | 0.006226 | 0.07225 | 0.007888 | 0.08839 | 0.06776 | 1.007 | 1.012 | 0.9765 | 0.988 | 0.2455 | 0.1407 |
| 0.3844 | -0.2041 | -0.6986 | 0.8767 | 0.8083 | -0.09247 | -0.1349 | 0.09247 | 0.1744 | 0.982 | 1.006 | 0.007302 | 0.006894 | 0.8081 | -0.3359 | 0.2808 | 0.4253 | 0.9963 | 1.02 | 0.9677 | 0.9927 | 1.358 | 0.4976 |
| 0.002944 | 8.18e-04 | -0.00257 | 0.01112 | 0.00897 | -0.001711 | -0.003258 | 0.006074 | 0.0052 | 1.009 | 1.013 | 0.006735 | 0.006719 | 0.08036 | 0.04364 | 0.01176 | 0.01385 | 1.023 | 1.026 | 0.9961 | 1 | 0.1034 | 0.0708 |
| 0.6274 | -0.4314 | -1.184 | 0.9445 | 1.276 | -0.125 | -0.2136 | 0.125 | 0.3058 | 1 | 0.999 | 0.008056 | 0.006677 | 0.2167 | -0.0945 | 0.4056 | 0.658 | 1.016 | 1.012 | 0.9844 | 0.9859 | 1.012 | 1.195 |
| 0.08482 | -0.09132 | -0.09106 | 0.1467 | 0.2414 | -0.02319 | -0.02789 | 0.02319 | 0.0279 | 0.9964 | 1.003 | 0.005432 | 0.005973 | -0.04571 | -0.105 | 0.04379 | 0.09452 | 1.007 | 1.015 | 0.9857 | 0.9916 | 0.04013 | 0.08021 |
|  | -0.3139 |  | 0.3151 |  | -0.05102 |  | 0.05208 |  | 0.9984 |  | 0.00896 |  | 0.051 |  | 0.1498 |  | 1.016 |  | 0.9808 |  | 0.3446 |  |
| 0.06437 | -0.1002 | -0.1041 | 0.1878 | 0.1483 | -0.02155 | -0.02413 | 0.0353 | 0.02413 | 0.9999 | 0.999 | 0.006555 | 0.007244 | 0.07713 | -0.04223 | 0.06664 | 0.09469 | 1.013 | 1.013 | 0.987 | 0.9848 | 0.2077 | 0.1434 |
| 0.5781 | -0.3365 | -0.654 | 0.4998 | 1.612 | -0.06914 | -0.06767 | 0.06914 | 0.3576 | 0.9946 | 1.002 | 0.005055 | 0.006246 | 0.2825 | 0.4683 | 0.251 | 0.383 | 1.004 | 1.015 | 0.9846 | 0.9901 | 0.7745 | 1.219 |
| 0.5801 | -0.3092 | -0.9304 | 0.466 | 1.344 | -0.06161 | -0.1497 | 0.06161 | 0.2676 | 1.004 | 1.013 | 0.006988 | 0.009404 | -0.1813 | -0.8189 | 0.2135 | 0.7836 | 1.018 | 1.031 | 0.9907 | 0.9944 | 0.2372 | 0.7169 |
| 0.3048 | 0.05198 | -0.5178 | 1.4 | 0.677 | -0.1086 | -0.0918 | 0.1086 | 0.0918 | 1.003 | 1 | 0.006465 | 0.006089 | -0.05735 | -0.1276 | 0.2624 | 0.2286 | 1.015 | 1.012 | 0.99 | 0.9882 | 0.4569 | 0.3204 |
| 0.02076 | 0.04402 | 0.03937 | 0.1003 | 0.1208 | -0.004913 | -0.003966 | 0.004915 | 0.01053 | 1.009 | 0.9995 | 0.008121 | 0.007131 | 0.04527 | 0.05979 | 0.01645 | 0.02078 | 1.025 | 1.013 | 0.9929 | 0.9855 | 0.07751 | 0.1005 |
| 0.01188 | 0.1938 | 0.1489 | 0.2315 | 0.1954 | 5.27e-04 | -9.56e-04 | 0.007167 | 0.007015 | 1.014 | 1.007 | 0.007059 | 0.00725 | 0.1357 | 0.1148 | 0.01081 | 0.008431 | 1.028 | 1.021 | 1 | 0.9923 | 0.1569 | 0.1313 |
| 0.03571 | -0.02984 | -0.0416 | 0.1082 | 0.09838 | -0.01151 | -0.007123 | 0.01151 | 0.01476 | 0.9945 | 1.001 | 0.009217 | 0.007135 | 0.05775 | 0.02881 | 0.05261 | 0.03736 | 1.013 | 1.015 | 0.9764 | 0.9871 | 0.1609 | 0.102 |
| 0.0106 | 0.04548 | 0.05522 | 0.07851 | 0.09679 | -0.001742 | -6.75e-04 | 0.003676 | 0.007298 | 1 | 1.006 | 0.005767 | 0.005964 | 0.0442 | 0.05569 | 0.007747 | 0.009493 | 1.012 | 1.017 | 0.989 | 0.994 | 0.05938 | 0.07429 |
| 0.21 | -0.3316 | -0.2806 | 0.3346 | 0.5427 | -0.06086 | -0.06947 | 0.0639 | 0.06948 | 1.002 | 1.007 | 0.006888 | 0.006374 | -0.2057 | -0.1212 | 0.1757 | 0.2064 | 1.016 | 1.02 | 0.9888 | 0.9949 | 0.1386 | 0.2834 |

| gcta.SE_h2_male | gcta.h2.conf.low_female | gcta.h2.conf.low_male | gcta.h2.conf.high_female | gcta.h2.conf.high_male | gcta.x.h2.conf.low_female | gcta.x.h2.conf.low_male | gcta.x.h2.conf.high_female | gcta.x.h2.conf.high_male | ldsc.int_female | ldsc.int_male | ldsc.int_se_female | ldsc.int_se_male | ldsc.h2_female | ldsc.h2_male | ldsc.h2_se_female | ldsc.h2_se_male | ldsc.int.conf.high_female | ldsc.int.conf.high_male | ldsc.int.conf.low_female | ldsc.int.conf.low_male | ldsc.h2.conf.high_female | ldsc.h2.conf.high_male |
|---|---|---|---|---|---|---|---|---|---|---|---|---|---|---|---|---|---|---|---|---|---|---|
| 0.1742 | -0.1587 | -0.2706 | 0.2718 | 0.4121 | -0.03372 | -0.03222 | 0.03372 | 0.1131 | 1.01 | 1.006 | 0.009576 | 0.004447 | -0.0255 | -0.2503 | 0.1175 | 0.1528 | 1.029 | 1.014 | 0.9914 | 0.997 | 0.2048 | 0.04913 |
| 0.1736 | 0.391 | 0.3762 | 0.7965 | 1.057 | -0.03419 | -0.06903 | 0.03419 | 0.06903 | 1.015 | 1.006 | 0.005989 | 0.005707 | -0.1972 | 0.02885 | 0.08753 | 0.2024 | 1.027 | 1.017 | 1.003 | 0.9946 | -0.02561 | 0.4256 |
| **gcta.SE_h2_male** | **gcta.h2.conf.low_female** | **gcta.h2.conf.low_male** | **gcta.h2.conf.high_female** | **gcta.h2.conf.high_male** | **gcta.x.h2.conf.low_female** | **gcta.x.h2.conf.low_male** | **gcta.x.h2.conf.high_female** | **gcta.x.h2.conf.high_male** | **ldsc.int_female** | **ldsc.int_male** | **ldsc.int_se_female** | **ldsc.int_se_male** | **ldsc.h2_female** | **ldsc.h2_male** | **ldsc.h2_se_female** | **ldsc.h2_se_male** | **ldsc.int.conf.high_female** | **ldsc.int.conf.high_male** | **ldsc.int.conf.low_female** | **ldsc.int.conf.low_male** | **ldsc.h2.conf.high_female** | **ldsc.h2.conf.high_male** |
|  | -0.2954 |  | 0.4391 |  | -0.06759 |  | 0.06759 |  | 0.983 | 0.7902 | 0.006551 | 0.007822 | 0.5804 | 3.913 | 0.2472 | 1.907 | 0.9958 | 0.8056 | 0.9701 | 0.7749 | 1.065 | 7.65 |
| 0.01062 | 0.06036 | 0.05369 | 0.0948 | 0.09533 | -0.001934 | -0.001978 | 0.003418 | 0.005197 | 1.021 | 1.008 | 0.007767 | 0.007409 | 0.04138 | 0.04271 | 0.01036 | 0.01178 | 1.037 | 1.022 | 1.006 | 0.993 | 0.06168 | 0.0658 |
| 0.3214 |  | -0.4829 |  | 0.7768 |  | -0.1264 |  | 0.1264 | 0.9982 | 0.9863 | 0.006909 | 0.006835 | -0.4697 | 0.7238 | 0.3273 | 0.3273 | 1.012 | 0.9997 | 0.9846 | 0.9729 | 0.1717 | 1.365 |
| 0.0282 | -0.002191 | -0.02696 | 0.1268 | 0.08359 | -0.01139 | -0.009476 | 0.0133 | 0.009478 | 0.994 | 0.9996 | 0.00644 | 0.006457 | 0.05584 | 0.05785 | 0.03593 | 0.02708 | 1.007 | 1.012 | 0.9813 | 0.9869 | 0.1263 | 0.1109 |
| 0.02041 | -0.01813 | 0.006343 | 0.05041 | 0.08634 | -0.005983 | -0.004779 | 0.005985 | 0.008988 | 1.01 | 1.003 | 0.007923 | 0.006515 | 0.009705 | 0.05084 | 0.02341 | 0.02598 | 1.026 | 1.015 | 0.9945 | 0.9898 | 0.05558 | 0.1018 |
| 0.2631 |  | -0.4686 |  | 0.5629 |  | -0.1001 |  | 0.1017 | 0.996 | 1.005 | 0.006127 | 0.007603 | -0.07366 | -0.5481 | 0.3719 | 0.2413 | 1.008 | 1.02 | 0.984 | 0.9904 | 0.6552 | -0.07525 |
| 0.2886 |  | -0.4383 |  | 0.6929 |  | -0.0383 |  | 0.1789 | 0.9979 | 1 | 0.00607 | 0.005817 | -0.6499 | 0.1 | 0.452 | 0.2359 | 1.01 | 1.011 | 0.986 | 0.9886 | 0.236 | 0.5625 |
| 0.3021 | -1.063 | -0.566 | 1.073 | 0.6184 | -0.1939 | -0.08392 | 0.2041 | 0.1367 | 0.9968 | 0.9996 | 0.006021 | 0.009412 | -0.05739 | -0.4546 | 0.4492 | 0.3135 | 1.009 | 1.018 | 0.985 | 0.9811 | 0.823 | 0.1599 |
| 0.2602 | -0.3656 | -0.4604 | 1.345 | 0.5595 | -0.167 | -0.09044 | 0.167 | 0.09044 | 1.007 | 0.995 | 0.006767 | 0.007036 | -0.4673 | 0.06278 | 0.3826 | 0.3042 | 1.021 | 1.009 | 0.9942 | 0.9812 | 0.2826 | 0.659 |
| 0.272 | -0.614 | -0.1943 | 1.323 | 0.8719 | -0.1799 | -0.103 | 0.1799 | 0.103 | 1 | 0.9945 | 0.008457 | 0.00564 | 0.05163 | 0.0837 | 0.5133 | 0.2301 | 1.017 | 1.006 | 0.9835 | 0.9835 | 1.058 | 0.5347 |
| 0.2548 | -0.2441 | -0.4549 | 1.443 | 0.5438 | -0.1659 | -0.08111 | 0.1659 | 0.114 | 0.9937 | 0.9944 | 0.006858 | 0.00638 | 0.376 | -0.04546 | 0.4128 | 0.239 | 1.007 | 1.007 | 0.9803 | 0.9819 | 1.185 | 0.4229 |
|  | -0.2835 |  | 0.5313 |  | -0.0649 |  | 0.0649 |  | 0.994 | 0.9913 | 0.005729 | 0.005748 | 0.09028 | -0.04479 | 0.1809 | 0.1641 | 1.005 | 1.003 | 0.9828 | 0.98 | 0.4448 | 0.2768 |
| 0.386 |  | -0.6017 |  | 0.9115 |  | -0.1291 |  | 0.1291 | 1.012 | 0.9957 | 0.008595 | 0.007064 | -0.8062 | 0.2973 | 0.4726 | 0.3378 | 1.028 | 1.01 | 0.9948 | 0.9819 | 0.1202 | 0.9593 |
| 0.2439 | -0.2711 | 0.1769 | 0.528 | 1.133 | -0.07458 | -0.08318 | 0.07458 | 0.08318 | 0.997 | 1.007 | 0.006655 | 0.006239 | 0.1861 | 0.2235 | 0.2239 | 0.2148 | 1.01 | 1.019 | 0.984 | 0.9949 | 0.625 | 0.6446 |
| 0.01784 | 0.1005 | 0.04735 | 0.1595 | 0.1173 | -0.004023 | -0.001623 | 0.005454 | 0.01217 | 1.009 | 1.013 | 0.01094 | 0.01075 | 0.07101 | 0.04258 | 0.01878 | 0.01944 | 1.03 | 1.034 | 0.9873 | 0.992 | 0.1078 | 0.08069 |
| 0.03257 | -0.03003 | 0.03391 | 0.1195 | 0.1616 | -0.01138 | -0.01034 | 0.01138 | 0.01034 | 1.007 | 0.9976 | 0.007752 | 0.009415 | 0.006965 | 0.07583 | 0.04866 | 0.04163 | 1.022 | 1.016 | 0.9919 | 0.9791 | 0.1023 | 0.1574 |
|  | -0.05161 |  | 0.2499 |  | -0.0194 |  | 0.03674 |  | 1.001 | 1.004 | 0.006609 | 0.005612 | 0.04741 | -0.06469 | 0.09284 | 0.06147 | 1.014 | 1.015 | 0.9884 | 0.9931 | 0.2294 | 0.0558 |
| 0.01011 | 0.05079 | 0.02123 | 0.08446 | 0.06087 | -3.23e-04 | -9.78e-04 | 0.00548 | 0.005248 | 1.001 | 0.9969 | 0.007399 | 0.005641 | 0.05071 | 0.04282 | 0.009001 | 0.007527 | 1.015 | 1.008 | 0.9865 | 0.9859 | 0.06835 | 0.05757 |
| 0.2815 | -0.4368 | -0.3181 | 0.6013 | 0.7852 | -0.03451 | -0.1111 | 0.2093 | 0.1111 | 0.9908 | 1.006 | 0.006407 | 0.008217 | 0.1272 | -0.1102 | 0.271 | 0.3131 | 1.003 | 1.022 | 0.9783 | 0.9901 | 0.6583 | 0.5035 |
| 0.3003 | -0.2919 | -0.4618 | 0.736 | 0.7155 | -0.08296 | -0.1076 | 0.08341 | 0.1076 | 0.9998 | 0.9973 | 0.005863 | 0.007197 | -0.05364 | 0.1732 | 0.2412 | 0.2904 | 1.011 | 1.011 | 0.9883 | 0.9832 | 0.4192 | 0.7425 |
| 0.4195 | -0.4342 | -0.0231 | 1.055 | 1.621 | -0.1336 | -0.1231 | 0.1336 | 0.1231 | 0.9906 | 1.009 | 0.007117 | 0.009577 | 0.344 | -0.5136 | 0.3399 | 0.4667 | 1.005 | 1.028 | 0.9766 | 0.9904 | 1.01 | 0.4011 |
| 0.2263 |  | -0.4019 |  | 0.4851 |  | -0.08105 |  | 0.08105 | 0.9895 | 0.9878 | 0.007987 | 0.008187 | 0.05476 | 0.3525 | 0.2606 | 0.3435 | 1.005 | 1.004 | 0.9739 | 0.9717 | 0.5655 | 1.026 |
| 0.01025 | 0.04886 | 0.0253 | 0.08221 | 0.06548 | -0.002316 | -9.14e-04 | 0.002463 | 0.005973 | 0.9875 | 1.001 | 0.007728 | 0.006978 | 0.05664 | 0.03314 | 0.01038 | 0.008868 | 1.003 | 1.015 | 0.9724 | 0.9878 | 0.07698 | 0.05052 |
| 0.06207 |  | -0.1154 |  | 0.1279 |  | -0.02076 |  | 0.02077 | 0.9902 | 1.006 | 0.007465 | 0.008916 | 0.08898 | 0.00153 | 0.08463 | 0.07495 | 1.005 | 1.024 | 0.9755 | 0.989 | 0.2549 | 0.1484 |
|  |  |  |  |  |  |  |  |  | 0.9887 | 0.9895 | 0.006754 | 0.008137 | 0.02826 | 0.3582 | 0.2156 | 0.3125 | 1.002 | 1.005 | 0.9755 | 0.9735 | 0.4509 | 0.9707 |

| gcta.SE_h2_male | gcta.h2.conf.low_female | gcta.h2.conf.low_male | gcta.h2.conf.high_female | gcta.h2.conf.high_male | gcta.x.h2.conf.low_female | gcta.x.h2.conf.low_male | gcta.x.h2.conf.high_female | gcta.x.h2.conf.high_male | ldsc.int_female | ldsc.int_male | ldsc.int_se_female | ldsc.int_se_male | ldsc.h2_female | ldsc.h2_male | ldsc.h2_se_female | ldsc.h2_se_male | ldsc.int.conf.high_female | ldsc.int.conf.high_male | ldsc.int.conf.low_female | ldsc.int.conf.low_male | ldsc.h2.conf.high_female | ldsc.h2.conf.high_male |
|---|---|---|---|---|---|---|---|---|---|---|---|---|---|---|---|---|---|---|---|---|---|---|
| 0.07118 | -0.06001 | -0.05707 | 0.1767 | 0.222 | -0.01763 | -0.02419 | 0.02366 | 0.02419 | 1.004 | 0.9973 | 0.007935 | 0.007141 | 0.04992 | 0.1175 | 0.06179 | 0.07382 | 1.02 | 1.011 | 0.9887 | 0.9834 | 0.171 | 0.2622 |
| | | | | | | | | | 0.9945 | 0.9928 | 0.006543 | 0.007986 | 0.2522 | 0.8591 | 0.4805 | 0.6878 | 1.007 | 1.008 | 0.9817 | 0.9772 | 1.194 | 2.207 |
| 0.02802 | 0.008514 | 0.01253 | 0.1203 | 0.1224 | -0.008139 | -0.01015 | 0.01152 | 0.01015 | 1.007 | 0.9822 | 0.008107 | 0.006884 | 0.02899 | 0.08258 | 0.03512 | 0.03658 | 1.022 | 0.9957 | 0.9907 | 0.9687 | 0.09783 | 0.1543 |
| 0.0547 | -0.001733 | -0.07453 | 0.2193 | 0.1399 | -0.0189 | -0.01194 | 0.02 | 0.02463 | 1.004 | 0.9991 | 0.004986 | 0.007372 | 0.0648 | 0.01476 | 0.05839 | 0.05744 | 1.014 | 1.014 | 0.9941 | 0.9847 | 0.1792 | 0.1273 |
| 0.2126 | 0.069 | -0.4106 | 0.7057 | 0.4228 | -0.05094 | -0.07622 | 0.05952 | 0.07622 | 0.9933 | 1.01 | 0.007436 | 0.008471 | 0.447 | -0.2093 | 0.1865 | 0.2209 | 1.008 | 1.026 | 0.9788 | 0.9929 | 0.8126 | 0.2236 |
| 0.606 | | -0.9057 | | 1.47 | | -0.1916 | | 0.1916 | 1.007 | 1.005 | 0.008029 | 0.009065 | -0.6596 | -0.1169 | 0.3753 | 0.7721 | 1.022 | 1.023 | 0.9909 | 0.9871 | 0.0761 | 1.396 |
| 0.4032 | -0.5537 | -0.7379 | 0.6103 | 0.8426 | -0.06444 | -0.1374 | 0.1187 | 0.1374 | 0.99 | 1.002 | 0.005587 | 0.007575 | 0.01483 | 0.2891 | 0.2363 | 0.5744 | 1.001 | 1.017 | 0.9791 | 0.9869 | 0.478 | 1.415 |
| 0.2366 | -0.3454 | -0.3061 | 0.3724 | 0.6213 | -0.06114 | -0.05579 | 0.08711 | 0.1083 | 1.001 | 0.9792 | 0.008102 | 0.007569 | -0.188 | 0.7443 | 0.2194 | 0.2961 | 1.017 | 0.994 | 0.9851 | 0.9644 | 0.242 | 1.325 |
| 0.01051 | 0.0602 | 0.04737 | 0.09394 | 0.08856 | -0.001381 | -3.27e-04 | 0.003927 | 0.008366 | 1.01 | 1.013 | 0.0103 | 0.006542 | 0.05519 | 0.03873 | 0.009529 | 0.009466 | 1.03 | 1.025 | 0.9901 | 0.9997 | 0.07387 | 0.05728 |
| 0.3609 | | -0.69 | | 0.7248 | | -0.09058 | | 0.1267 | 0.9953 | 0.9799 | 0.006941 | 0.006321 | 0.143 | 0.7545 | 0.3109 | 0.3526 | 1.009 | 0.9923 | 0.9817 | 0.9675 | 0.7524 | 1.446 |
| 0.06522 | -0.01148 | -0.1056 | 0.3388 | 0.1501 | -0.03029 | -0.02176 | 0.03029 | 0.02177 | 0.9963 | 0.9774 | 0.006201 | 0.00662 | 0.189 | 0.1539 | 0.1129 | 0.06342 | 1.008 | 0.9904 | 0.9841 | 0.9645 | 0.4104 | 0.2782 |
| 0.03052 | 0.03304 | -0.03108 | 0.1204 | 0.08854 | -0.00643 | -0.005591 | 0.008615 | 0.0178 | 0.9995 | 0.992 | 0.008509 | 0.006629 | 0.0562 | 0.0545 | 0.02561 | 0.03647 | 1.016 | 1.005 | 0.9828 | 0.979 | 0.1064 | 0.126 |
| | -0.1229 | | 0.8536 | | -0.06904 | | 0.1147 | | 0.9925 | 1.009 | 0.006353 | 0.007966 | 0.2971 | -0.5629 | 0.2348 | 0.2204 | 1.005 | 1.025 | 0.98 | 0.9937 | 0.7573 | -0.131 |
| 0.03667 | 0.04217 | 0.02654 | 0.1192 | 0.1703 | -0.003248 | -0.003328 | 0.01027 | 0.02394 | 1.01 | 1.012 | 0.00676 | 0.00648 | 0.02742 | 0.005679 | 0.03108 | 0.0343 | 1.023 | 1.024 | 0.9969 | 0.9989 | 0.08834 | 0.07291 |
| | -0.4177 | | 0.4234 | | -0.05683 | | 0.06198 | | 1.006 | 0.9276 | 0.007509 | 0.005035 | -0.4132 | -2.646 | 0.2804 | 1.551 | 1.021 | 0.9375 | 0.9911 | 0.9177 | 0.1364 | 0.3931 |
| 0.01031 | 0.05277 | 0.03843 | 0.08526 | 0.07885 | 6.21e-04 | -0.003502 | 0.0065 | 0.003504 | 1.015 | 0.9984 | 0.008954 | 0.007272 | 0.0367 | 0.04485 | 0.009873 | 0.01033 | 1.032 | 1.013 | 0.9971 | 0.9842 | 0.05605 | 0.06509 |
| 0.3152 | -1.156 | -0.3092 | 1.257 | 0.9263 | -0.1491 | -0.02148 | 0.2631 | 0.2387 | 1.004 | 1.008 | 0.007949 | 0.006531 | -0.4922 | -0.2045 | 0.4568 | 0.3279 | 1.019 | 1.02 | 0.9882 | 0.9948 | 0.4031 | 0.4382 |
| 0.3349 | -0.4603 | -0.311 | 0.5025 | 1.002 | -0.08751 | -0.1195 | 0.08751 | 0.1195 | 0.9939 | 0.9962 | 0.007928 | 0.008531 | 0.04546 | 0.1081 | 0.2351 | 0.3225 | 1.009 | 1.013 | 0.9784 | 0.9795 | 0.5062 | 0.7402 |
| 0.08281 | -0.06282 | -0.1545 | 0.3138 | 0.1701 | -0.02402 | -0.03577 | 0.05048 | 0.03577 | 1.01 | 0.998 | 0.007024 | 0.008426 | -0.0694 | 0.02672 | 0.1154 | 0.08756 | 1.023 | 1.015 | 0.9959 | 0.9815 | 0.1569 | 0.1983 |
| 0.2062 | | -0.1879 | | 0.6204 | | -0.06944 | | 0.06944 | 1.003 | 0.9997 | 0.00594 | 0.006233 | -0.2797 | 0.3256 | 0.1976 | 0.199 | 1.015 | 1.012 | 0.9914 | 0.9875 | 0.1076 | 0.7157 |
| 0.1324 | 0.2129 | 0.04615 | 0.7178 | 0.5651 | -0.03607 | -0.02681 | 0.03943 | 0.06291 | 1.002 | 1.007 | 0.006935 | 0.008856 | 0.279 | 0.08467 | 0.1188 | 0.1484 | 1.016 | 1.024 | 0.9887 | 0.9896 | 0.5119 | 0.3756 |
| 0.1974 | -0.04531 | -0.03694 | 0.7694 | 0.7368 | -0.06585 | -0.04297 | 0.06585 | 0.09467 | 1.008 | 0.9972 | 0.006546 | 0.006702 | -0.05225 | 0.3416 | 0.1643 | 0.1788 | 1.021 | 1.01 | 0.9956 | 0.984 | 0.2697 | 0.692 |
| 0.1962 | 0.04799 | 0.006437 | 0.9112 | 0.7754 | -0.04158 | -0.04978 | 0.1025 | 0.07971 | 1.02 | 0.9959 | 0.006975 | 0.005728 | -0.1803 | 0.3962 | 0.2321 | 0.1714 | 1.033 | 1.007 | 1.006 | 0.9847 | 0.2746 | 0.7322 |
| 0.1327 | | -0.197 | | 0.3231 | | -0.04898 | | 0.04898 | 1.009 | 1.01 | 0.007939 | 0.005175 | -0.1852 | 0.02902 | 0.1249 | 0.1248 | 1.024 | 1.02 | 0.9931 | 0.9995 | 0.05966 | 0.2736 |
| 0.2043 | | -0.176 | | 0.6249 | | -0.06744 | | 0.06744 | 1.004 | 0.9962 | 0.005388 | 0.00691 | -0.3921 | 0.4741 | 0.1871 | 0.1998 | 1.014 | 1.01 | 0.9933 | 0.9827 | -0.0253 | 0.8656 |
| 0.1324 | 0.2119 | 0.04615 | 0.7168 | 0.5651 | -0.03623 | -0.02681 | 0.03928 | 0.06291 | 1.002 | 1.007 | 0.00694 | 0.008856 | 0.2811 | 0.08467 | 0.1196 | 0.1484 | 1.016 | 1.024 | 0.9886 | 0.9896 | 0.5155 | 0.3756 |
| 0.1974 | -0.04868 | -0.03694 | 0.7655 | 0.7368 | -0.06599 | -0.04297 | 0.06599 | 0.09467 | 1.008 | 0.9972 | 0.006483 | 0.006702 | -0.05491 | 0.3416 | 0.1622 | 0.1788 | 1.021 | 1.01 | 0.9958 | 0.984 | 0.263 | 0.692 |
| 0.1962 | 0.03823 | 0.006437 | 0.9009 | 0.7754 | -0.04196 | -0.04978 | 0.1022 | 0.07971 | 1.02 | 0.9959 | 0.006949 | 0.005728 | -0.191 | 0.3962 | 0.2331 | 0.1714 | 1.033 | 1.007 | 1.006 | 0.9847 | 0.2658 | 0.7322 |
| 0.206 | | -0.1987 | | 0.6087 | | -0.07012 | | 0.07012 | 1.005 | 0.9955 | 0.005775 | 0.006561 | -0.3583 | 0.5023 | 0.1945 | 0.1889 | 1.016 | 1.008 | 0.9937 | 0.9826 | 0.02295 | 0.8725 |

| gcta.SE_h2_male | gcta.h2.conf.low_female | gcta.h2.conf.low_male | gcta.h2.conf.high_female | gcta.h2.conf.high_male | gcta.x.h2.conf.low_female | gcta.x.h2.conf.low_male | gcta.x.h2.conf.high_female | gcta.x.h2.conf.high_male | ldsc.int_female | ldsc.int_male | ldsc.int_se_female | ldsc.int_se_male | ldsc.h2_female | ldsc.h2_male | ldsc.h2_se_female | ldsc.h2_se_male | ldsc.int.conf.high_female | ldsc.int.conf.high_male | ldsc.int.conf.low_female | ldsc.int.conf.low_male | ldsc.h2.conf.high_female | ldsc.h2.conf.high_male |
|---|---|---|---|---|---|---|---|---|---|---|---|---|---|---|---|---|---|---|---|---|---|---|
| 0.2501 | | -0.3447 | | 0.6359 | | -0.08524 | | 0.08524 | 1.007 | 1.009 | 0.004828 | 0.006786 | -0.3383 | -0.04715 | 0.1953 | 0.2466 | 1.017 | 1.023 | 0.998 | 0.9959 | 0.0445 | 0.4363 |
| 0.1314 | 0.3801 | -0.03226 | 0.878 | 0.4828 | -0.03204 | -0.02555 | 0.04332 | 0.06054 | 1.003 | 1.013 | 0.007238 | 0.00685 | 0.338 | -0.08019 | 0.1227 | 0.1182 | 1.017 | 1.027 | 0.9891 | 1 | 0.5786 | 0.1515 |
| 0.1304 | | -0.06425 | | 0.4469 | | -0.03858 | | 0.04094 | 1.007 | 1.016 | 0.00898 | 0.008164 | -0.1166 | -0.003048 | 0.1559 | 0.1483 | 1.025 | 1.032 | 0.9898 | 0.9997 | 0.1889 | 0.2877 |
| 0.4941 | -0.08214 | -0.9417 | 0.9709 | 0.9952 | -0.0348 | -0.1428 | 0.1699 | 0.2104 | 1.005 | 1.008 | 0.006502 | 0.007151 | -0.08494 | -0.8258 | 0.3113 | 0.3423 | 1.017 | 1.022 | 0.9919 | 0.9945 | 0.5253 | -0.1548 |
| 0.01031 | 0.06285 | 0.04021 | 0.09692 | 0.08064 | -0.001367 | -7.31e-04 | 0.003908 | 0.007111 | 1.009 | 1.007 | 0.01042 | 0.006716 | 0.06029 | 0.04085 | 0.009398 | 0.01082 | 1.03 | 1.021 | 0.989 | 0.9943 | 0.07871 | 0.06207 |
| 0.2002 | | -0.2419 | | 0.5429 | | -0.07311 | | 0.07312 | 1.004 | 0.9886 | 0.006586 | 0.005772 | -0.2351 | 0.2264 | 0.1584 | 0.2071 | 1.017 | 0.9999 | 0.9914 | 0.9773 | 0.07528 | 0.6322 |
| 1.016 | | -1.802 | | 2.183 | | -0.4333 | | 0.4333 | 1.002 | 0.9154 | 0.006147 | 0.007524 | -0.4001 | 0.3585 | 0.3932 | 0.9635 | 1.014 | 0.9302 | 0.9898 | 0.9007 | 0.3705 | 2.247 |
| 0.02587 | 0.07239 | 0.08796 | 0.1446 | 0.1894 | -0.005287 | -0.004559 | 0.006665 | 0.01358 | 1.007 | 1 | 0.009 | 0.00808 | 0.07423 | 0.1058 | 0.02271 | 0.02789 | 1.024 | 1.016 | 0.9891 | 0.9844 | 0.1187 | 0.1605 |
| 0.02414 | 0.04129 | 0.07283 | 0.1079 | 0.1675 | -0.005763 | -0.008121 | 0.005765 | 0.008123 | 0.9899 | 0.9999 | 0.006873 | 0.009158 | 0.07966 | 0.07787 | 0.0154 | 0.02713 | 1.003 | 1.018 | 0.9764 | 0.9819 | 0.1098 | 0.1311 |
| 0.01783 | 0.07478 | 0.04428 | 0.1317 | 0.1142 | -0.004756 | -0.00283 | 0.004758 | 0.01116 | 1.004 | 1.01 | 0.00762 | 0.009026 | 0.06118 | 0.03185 | 0.01647 | 0.0206 | 1.019 | 1.028 | 0.9893 | 0.9922 | 0.09347 | 0.07223 |
| 0.02045 | 0.04523 | 0.01282 | 0.1148 | 0.09298 | -0.005842 | -0.005447 | 0.005844 | 0.008965 | 0.9899 | 1.003 | 0.008986 | 0.008309 | 0.06386 | 0.04752 | 0.01678 | 0.01765 | 1.008 | 1.02 | 0.9723 | 0.9872 | 0.09675 | 0.08211 |
| 0.01809 | 0.06486 | 0.04065 | 0.1213 | 0.1116 | -0.005003 | -0.003304 | 0.005005 | 0.009913 | 1.006 | 1.001 | 0.007452 | 0.007933 | 0.05587 | 0.06757 | 0.01838 | 0.01683 | 1.021 | 1.016 | 0.9918 | 0.985 | 0.0919 | 0.1005 |
| 0.3386 | | -0.6549 | | 0.6724 | | -0.1019 | | 0.1217 | 1.019 | 0.9955 | 0.007529 | 0.005673 | -0.4884 | -0.2206 | 0.2253 | 0.2967 | 1.034 | 1.007 | 1.004 | 0.9844 | -0.04674 | 0.361 |
| 0.02047 | 0.0356 | 0.008736 | 0.1043 | 0.08897 | -0.005395 | -0.007227 | 0.005397 | 0.007229 | 0.9909 | 1.009 | 0.008682 | 0.008167 | 0.06321 | 0.02755 | 0.01628 | 0.02097 | 1.008 | 1.025 | 0.9739 | 0.9931 | 0.09511 | 0.06866 |
| 1.585 | | -2.142 | | 4.071 | | -0.4863 | | 0.5685 | 1.007 | 0.8293 | 0.00592 | 0.007179 | -0.1466 | 2.395 | 0.06485 | 1.337 | 1.019 | 0.8434 | 0.9957 | 0.8152 | -0.01952 | 5.016 |
| 0.2561 | -0.2371 | -0.4191 | 1.092 | 0.585 | -0.05642 | -0.06071 | 0.1876 | 0.141 | 1.002 | 1.004 | 0.009292 | 0.008283 | -0.2361 | -0.1028 | 0.3971 | 0.354 | 1.02 | 1.02 | 0.9833 | 0.9877 | 0.5421 | 0.5909 |
| 0.07612 | | -0.139 | | 0.1594 | | -0.02308 | | 0.02308 | 0.9997 | 0.9822 | 0.00795 | 0.007192 | -0.02755 | 0.1546 | 0.1097 | 0.07276 | 1.015 | 0.9963 | 0.9841 | 0.9681 | 0.1876 | 0.2972 |
| 0.3169 | -0.3757 | -0.5191 | 0.619 | 0.7229 | -0.06204 | -0.1106 | 0.09638 | 0.1106 | 1.021 | 1 | 0.005108 | 0.00687 | -0.3895 | 0.03903 | 0.217 | 0.2358 | 1.031 | 1.014 | 1.011 | 0.9869 | 0.03578 | 0.5012 |
| 0.2023 | -0.09978 | -0.3344 | 0.546 | 0.4588 | -0.02147 | -0.03807 | 0.08915 | 0.1124 | 0.9937 | 1.001 | 0.007893 | 0.009872 | 0.3463 | 0.1432 | 0.1692 | 0.2206 | 1.009 | 1.021 | 0.9782 | 0.9821 | 0.6779 | 0.5755 |
| 0.1102 | 0.1942 | 0.01305 | 0.5676 | 0.4449 | -0.02649 | -0.02773 | 0.03873 | 0.04735 | 1.001 | 1.017 | 0.008009 | 0.00844 | 0.2648 | -0.07739 | 0.09609 | 0.1422 | 1.016 | 1.033 | 0.9851 | 1 | 0.4531 | 0.2012 |
| 0.1602 | 0.07211 | -0.009583 | 0.6813 | 0.6183 | -0.02441 | -0.05528 | 0.07092 | 0.05528 | 1.005 | 1 | 0.009542 | 0.00743 | 0.1135 | 0.2672 | 0.1902 | 0.1684 | 1.024 | 1.015 | 0.9864 | 0.9858 | 0.4863 | 0.5972 |
| 0.1113 | -0.07637 | 0.1119 | 0.2848 | 0.548 | 0.002423 | -0.02948 | 0.07821 | 0.04956 | 1.01 | 1.009 | 0.007653 | 0.009156 | 0.01998 | 0.1113 | 0.1014 | 0.1219 | 1.025 | 1.027 | 0.9947 | 0.9914 | 0.2187 | 0.3502 |
| 0.5347 | -0.4087 | -1.025 | 1.337 | 1.071 | -0.08814 | -0.2179 | 0.2052 | 0.2179 | 0.9959 | 0.9809 | 0.008639 | 0.008818 | 0.4795 | 1.266 | 0.5481 | 0.7401 | 1.013 | 0.9982 | 0.979 | 0.9636 | 1.554 | 2.717 |
| 0.3727 | -0.9235 | -0.3567 | 0.9876 | 1.104 | -0.1391 | -0.1334 | 0.1998 | 0.1334 | 1.004 | 0.9978 | 0.005123 | 0.009164 | 0.2484 | 0.1306 | 0.4339 | 0.4384 | 1.014 | 1.016 | 0.9942 | 0.9798 | 1.099 | 0.9899 |
| 0.02945 | -0.01172 | -0.04782 | 0.0765 | 0.06765 | -0.00826 | -0.01141 | 0.008262 | 0.01196 | 0.9866 | 0.9966 | 0.009751 | 0.008744 | 0.06709 | 0.01908 | 0.03325 | 0.03595 | 1.006 | 1.014 | 0.9675 | 0.9795 | 0.1323 | 0.08954 |
| | -0.3565 | | 1.423 | | -0.1183 | | 0.1395 | | 0.9914 | 1 | 0.007838 | 0.008412 | 0.002698 | 0.08772 | 0.4132 | 0.6562 | 1.007 | 1.017 | 0.976 | 0.9838 | 0.8126 | 1.374 |
| 0.1664 | -0.2265 | -0.3115 | 0.3586 | 0.3408 | -0.04349 | -0.07046 | 0.04371 | 0.07047 | 0.9964 | 0.9993 | 0.007839 | 0.006329 | 0.008282 | 0.01511 | 0.1424 | 0.1165 | 1.012 | 1.012 | 0.9811 | 0.9869 | 0.2873 | 0.2435 |

| gcta.SE_h2_male | gcta.h2.conf.low_female | gcta.h2.conf.low_male | gcta.h2.conf.high_female | gcta.h2.conf.high_male | gcta.x.h2.conf.low_female | gcta.x.h2.conf.low_male | gcta.x.h2.conf.high_female | gcta.x.h2.conf.high_male | ldsc.int_female | ldsc.int_male | ldsc.int_se_female | ldsc.int_se_male | ldsc.h2_female | ldsc.h2_male | ldsc.h2_se_female | ldsc.h2_se_male | ldsc.int.conf.high_female | ldsc.int.conf.high_male | ldsc.int.conf.low_female | ldsc.int.conf.low_male | ldsc.h2.conf.high_female | ldsc.h2.conf.high_male |
|---|---|---|---|---|---|---|---|---|---|---|---|---|---|---|---|---|---|---|---|---|---|---|
| 0.5428 | -0.5389 | -0.7065 | 1.054 | 1.421 | 0.06379 | -0.1552 | 0.4594 | 0.2382 | 1.004 | 0.9992 | 0.006689 | 0.009195 | 0.1238 | 0.03298 | 0.2429 | 0.6114 | 1.017 | 1.017 | 0.9912 | 0.9812 | 0.5998 | 1.231 |
| 0.2031 | -0.1629 | -0.3076 | 0.47 | 0.4887 | -0.05061 | -0.04611 | 0.05062 | 0.09041 | 1 | 0.994 | 0.007047 | 0.01054 | 0.08095 | 0.3258 | 0.1544 | 0.2591 | 1.014 | 1.015 | 0.9864 | 0.9733 | 0.3836 | 0.8337 |
| 0.2118 | 0.1044 | -0.08327 | 0.7348 | 0.7471 | -0.05299 | -0.05079 | 0.05299 | 0.1056 | 0.9923 | 1.009 | 0.008745 | 0.005246 | 0.488 | -0.05515 | 0.1767 | 0.1757 | 1.009 | 1.019 | 0.9752 | 0.9986 | 0.8343 | 0.2893 |
| 0.1578 | 0.204 | 0.3207 | 0.7894 | 0.9393 | -0.03851 | -0.05107 | 0.04899 | 0.05107 | 1.012 | 1.01 | 0.007809 | 0.008491 | 0.09232 | 0.2646 | 0.1899 | 0.166 | 1.027 | 1.026 | 0.9963 | 0.993 | 0.4646 | 0.59 |
| 0.1978 | -0.2234 | -0.325 | 0.4009 | 0.4504 | -0.03971 | -0.04989 | 0.05723 | 0.08139 | 1.009 | 0.9968 | 0.008541 | 0.0104 | -0.04484 | 0.2984 | 0.1684 | 0.2404 | 1.026 | 1.017 | 0.9922 | 0.9764 | 0.2853 | 0.7697 |
| 0.4882 | -0.6063 | -0.4362 | 1.139 | 1.478 | -0.0931 | -0.1418 | 0.1824 | 0.1418 | 1.002 | 0.9915 | 0.007812 | 0.007362 | 0.6922 | 0.2296 | 0.4585 | 0.6254 | 1.017 | 1.006 | 0.9867 | 0.9771 | 1.591 | 1.455 |
| 0.4743 | -0.3429 | -0.1885 | 1.339 | 1.671 | -0.1172 | -0.1973 | 0.1372 | 0.1973 | 1.012 | 0.9974 | 0.006925 | 0.006742 | -0.7067 | 0.9843 | 0.3567 | 0.421 | 1.026 | 1.011 | 0.9987 | 0.9842 | -0.007672 | 1.809 |
| 0.4738 | -0.3032 | -0.05804 | 1.377 | 1.799 | -0.1152 | -0.1932 | 0.1399 | 0.1932 | 1.012 | 0.9978 | 0.006615 | 0.00687 | -0.689 | 0.9684 | 0.3401 | 0.4379 | 1.025 | 1.011 | 0.9993 | 0.9844 | -0.02243 | 1.827 |
| 0.1564 | -0.1061 | -0.003945 | 0.479 | 0.609 | -0.01905 | -0.0534 | 0.0917 | 0.0534 | 0.9978 | 1.008 | 0.00643 | 0.005484 | 0.1482 | 0.07236 | 0.1814 | 0.1716 | 1.01 | 1.019 | 0.9852 | 0.9973 | 0.5037 | 0.4087 |
| 0.1609 | -0.1425 | -0.1234 | 0.4231 | 0.5074 | -0.04221 | -0.06238 | 0.04221 | 0.06238 | 0.9942 | 0.9984 | 0.007998 | 0.007698 | 0.1415 | 0.03888 | 0.1458 | 0.1832 | 1.01 | 1.013 | 0.9786 | 0.9833 | 0.4273 | 0.3979 |
| 0.1602 | -0.06293 | -0.3101 | 0.5041 | 0.318 | -0.0462 | -0.05105 | 0.0462 | 0.05987 | 1.01 | 0.9973 | 0.006476 | 0.007403 | -0.007529 | -0.01305 | 0.1463 | 0.1588 | 1.023 | 1.012 | 0.9976 | 0.9828 | 0.2791 | 0.2983 |
| 0.1561 | -0.1051 | -0.02689 | 0.4823 | 0.5849 | -0.01461 | -0.05382 | 0.09787 | 0.05382 | 0.999 | 1.008 | 0.006636 | 0.005881 | 0.1107 | 0.05878 | 0.1803 | 0.1932 | 1.012 | 1.02 | 0.986 | 0.9965 | 0.4642 | 0.4374 |
| 0.0518 | | -0.06276 | | 0.1403 | | -0.01704 | | 0.01705 | 1.001 | 1.007 | 0.008373 | 0.007029 | -0.08652 | 0.01304 | 0.08165 | 0.04638 | 1.017 | 1.021 | 0.9842 | 0.9937 | 0.07351 | 0.1039 |
| 0.2636 | -0.2745 | -0.03687 | 1.202 | 0.9962 | -0.1027 | -0.04465 | 0.1419 | 0.1459 | 1.006 | 1.008 | 0.005085 | 0.007312 | -0.1133 | 0.137 | 0.3455 | 0.3073 | 1.016 | 1.022 | 0.9961 | 0.9933 | 0.5639 | 0.7393 |
| 0.162 | -0.2212 | -0.3161 | 0.5765 | 0.3189 | -0.04124 | -0.04606 | 0.09125 | 0.04892 | 1.012 | 1.005 | 0.006871 | 0.004881 | -0.1349 | -0.2448 | 0.2328 | 0.115 | 1.025 | 1.014 | 0.9982 | 0.9949 | 0.3214 | -0.01936 |
| | | | | | | | | | 1.009 | 1.001 | 0.008362 | 0.004751 | -0.7281 | -0.26 | 0.3767 | 0.2943 | 1.025 | 1.011 | 0.9923 | 0.992 | 0.01017 | 0.317 |
| | | | | | | | | | 1.006 | 1.015 | 0.009777 | 0.004991 | 0.1197 | -0.8016 | 0.4882 | 0.2204 | 1.026 | 1.025 | 0.9872 | 1.006 | 1.077 | -0.3697 |
| 0.1668 | | -0.2919 | | 0.362 | | -0.02695 | | 0.102 | 1.015 | 1.01 | 0.006334 | 0.005621 | -0.5384 | -0.4057 | 0.1332 | 0.1728 | 1.027 | 1.021 | 1.002 | 0.9994 | -0.2773 | -0.06694 |
| 0.1288 | -0.03234 | -0.1628 | 0.6062 | 0.3422 | -0.009097 | -0.01616 | 0.1121 | 0.0903 | 0.9925 | 0.9949 | 0.01026 | 0.008482 | 0.1691 | 0.1366 | 0.2422 | 0.1189 | 1.013 | 1.012 | 0.9723 | 0.9783 | 0.6439 | 0.3697 |
| 0.2516 | | -0.3119 | | 0.6746 | | -0.08971 | | 0.08971 | 1.013 | 0.9894 | 0.007272 | 0.008576 | -1.04 | 0.2389 | 0.3306 | 0.2766 | 1.027 | 1.006 | 0.9986 | 0.9726 | -0.392 | 0.7811 |
| | -0.1823 | | 0.4776 | | -0.05588 | | 0.05588 | | 1.004 | 1.002 | 0.008425 | 0.007961 | 0.01671 | -0.0205 | 0.1909 | 0.1252 | 1.02 | 1.017 | 0.9872 | 0.9862 | 0.3908 | 0.2249 |
| 0.208 | -0.2086 | -0.007966 | 0.6462 | 0.8072 | -0.04946 | -0.0807 | 0.1157 | 0.08071 | 0.9856 | 0.9946 | 0.007436 | 0.00558 | 0.4726 | 0.4173 | 0.2339 | 0.2255 | 1 | 1.006 | 0.9711 | 0.9837 | 0.931 | 0.8594 |
| 0.2178 | 0.04569 | -0.2685 | 0.9169 | 0.5852 | -0.06644 | -0.04949 | 0.06645 | 0.109 | 1.022 | 1.002 | 0.007706 | 0.006012 | -0.3925 | -0.07714 | 0.2205 | 0.241 | 1.037 | 1.014 | 1.007 | 0.9904 | 0.03962 | 0.3953 |
| 0.2144 | 0.07448 | -0.3617 | 0.9381 | 0.4788 | -0.05153 | -0.05404 | 0.0975 | 0.09885 | 1.02 | 1.003 | 0.00772 | 0.00607 | -0.3414 | -0.141 | 0.2334 | 0.2159 | 1.035 | 1.015 | 1.005 | 0.9911 | 0.1161 | 0.2821 |
| 0.2189 | -0.01766 | -0.2219 | 0.8614 | 0.6363 | -0.06975 | -0.04555 | 0.06975 | 0.1188 | 1.016 | 1.001 | 0.006205 | 0.005819 | -0.3416 | -0.03842 | 0.2081 | 0.2431 | 1.028 | 1.012 | 1.003 | 0.9896 | 0.06633 | 0.438 |
| 0.2072 | -0.2584 | -0.01644 | 0.5864 | 0.7959 | -0.05661 | -0.07372 | 0.1022 | 0.07372 | 0.9874 | 1.001 | 0.007498 | 0.005719 | 0.4027 | 0.3419 | 0.2304 | 0.2148 | 1.002 | 1.012 | 0.9728 | 0.9895 | 0.8542 | 0.763 |
| 0.04019 | -0.02239 | -0.0332 | 0.1144 | 0.1243 | -0.01084 | -0.007321 | 0.01084 | 0.02704 | 0.9978 | 1.006 | 0.004494 | 0.005578 | 0.02339 | -0.003213 | 0.03405 | 0.02935 | 1.007 | 1.017 | 0.989 | 0.9947 | 0.09012 | 0.05432 |
| 0.03901 | -0.01417 | -0.04792 | 0.1265 | 0.105 | -0.01054 | -0.01124 | 0.01054 | 0.01301 | 1.007 | 1.007 | 0.007019 | 0.008891 | 0.02027 | -0.003361 | 0.03845 | 0.05008 | 1.021 | 1.024 | 0.9932 | 0.9893 | 0.09563 | 0.09479 |
| 0.03947 | 0.04936 | -0.07067 | 0.199 | 0.08405 | -0.01371 | -0.01507 | 0.01371 | 0.01507 | 1.005 | 1.004 | 0.006044 | 0.00811 | 0.09932 | 0.004015 | 0.04698 | 0.04397 | 1.017 | 1.02 | 0.9932 | 0.9881 | 0.1914 | 0.0902 |
| 0.03924 | -0.06995 | -0.07543 | 0.07104 | 0.07839 | -0.01158 | -0.01232 | 0.01265 | 0.01534 | 1.017 | 0.9995 | 0.009324 | 0.008045 | -0.06782 | -0.03768 | 0.05548 | 0.047 | 1.035 | 1.015 | 0.9984 | 0.9837 | 0.04092 | 0.05444 |

| gcta.SE_h2_male | gcta.h2.conf.low_female | gcta.h2.conf.low_male | gcta.h2.conf.high_female | gcta.h2.conf.high_male | gcta.x.h2.conf.low_female | gcta.x.h2.conf.low_male | gcta.x.h2.conf.high_female | gcta.x.h2.conf.high_male | ldsc.int_female | ldsc.int_male | ldsc.int_se_female | ldsc.int_se_male | ldsc.h2_female | ldsc.h2_male | ldsc.h2_se_female | ldsc.h2_se_male | ldsc.int.conf.high_female | ldsc.int.conf.high_male | ldsc.int.conf.low_female | ldsc.int.conf.low_male | ldsc.h2.conf.high_female | ldsc.h2.conf.high_male |
|---|---|---|---|---|---|---|---|---|---|---|---|---|---|---|---|---|---|---|---|---|---|---|
| 0.04086 | -0.02708 | -0.02278 | 0.1102 | 0.1374 | -0.01214 | -0.01132 | 0.01214 | 0.01383 | 1.009 | 1.002 | 0.006402 | 0.00824 | -0.001808 | 0.04858 | 0.02465 | 0.05463 | 1.022 | 1.018 | 0.9969 | 0.9857 | 0.0465 | 0.1557 |
| 0.3523 | 0.04826 | 0.3095 | 0.3305 | 1.69 | -0.008851 | -0.1048 | 0.0442 | 0.1818 | 1.003 | 1.011 | 0.006917 | 0.006182 | 0.09417 | 0.5018 | 0.05942 | 0.377 | 1.017 | 1.023 | 0.9899 | 0.9991 | 0.2106 | 1.241 |
| 0.02543 | -1.27e-04 | 0.01551 | 0.05822 | 0.1152 | -0.002656 | -0.006077 | 0.007797 | 0.01217 | 0.9979 | 0.992 | 0.008176 | 0.006143 | 0.02087 | 0.05822 | 0.01448 | 0.02242 | 1.014 | 1.004 | 0.9819 | 0.9799 | 0.04924 | 0.1022 |
| 0.1558 | -0.3203 | -0.1621 | 0.4096 | 0.4488 | -0.05158 | -0.04959 | 0.06832 | 0.04959 | 1.004 | 0.9916 | 0.00847 | 0.006792 | -0.2866 | 0.1441 | 0.2068 | 0.1555 | 1.02 | 1.005 | 0.9871 | 0.9783 | 0.1187 | 0.449 |
| 0.2066 | -0.02894 | -0.2378 | 0.6004 | 0.572 | -0.05103 | -0.04358 | 0.05103 | 0.09464 | 1.004 | 1.001 | 0.006351 | 0.008926 | 0.1325 | 0.1488 | 0.1913 | 0.2449 | 1.016 | 1.019 | 0.9916 | 0.9837 | 0.5074 | 0.6287 |
| 0.2121 | 0.09845 | -0.08045 | 0.7288 | 0.7509 | -0.05358 | -0.05037 | 0.05358 | 0.1061 | 0.9926 | 1.009 | 0.008504 | 0.005162 | 0.4844 | -0.05483 | 0.1701 | 0.1749 | 1.009 | 1.019 | 0.9759 | 0.9991 | 0.8179 | 0.2879 |
| 0.1577 | 0.1859 | 0.3288 | 0.7707 | 0.9468 | -0.04136 | -0.04914 | 0.0444 | 0.04914 | 1.014 | 1.014 | 0.007903 | 0.007447 | 0.02608 | 0.1601 | 0.189 | 0.1222 | 1.03 | 1.029 | 0.9989 | 0.9994 | 0.3965 | 0.3996 |
| 0.2032 | -0.1192 | -0.1402 | 0.5093 | 0.6565 | -0.04362 | -0.04278 | 0.05178 | 0.09742 | 1.005 | 0.9957 | 0.006727 | 0.009169 | 0.08771 | 0.3258 | 0.1328 | 0.251 | 1.018 | 1.014 | 0.9919 | 0.9778 | 0.348 | 0.8178 |
| 0.4946 | -0.1206 | -0.3559 | 1.686 | 1.583 | -0.1058 | -0.1222 | 0.1697 | 0.1662 | 1.004 | 0.9872 | 0.007169 | 0.005601 | -0.0659 | 0.4895 | 0.4477 | 0.5042 | 1.018 | 0.9982 | 0.9895 | 0.9763 | 0.8116 | 1.478 |
| 0.09024 | 0.02756 | -0.02865 | 0.3045 | 0.3251 | -0.01578 | -0.03297 | 0.02729 | 0.03297 | 0.9915 | 0.998 | 0.007115 | 0.006896 | 0.1971 | 0.08715 | 0.05731 | 0.07496 | 1.005 | 1.011 | 0.9775 | 0.9845 | 0.3095 | 0.2341 |
| 0.09625 | -0.09237 | -0.1687 | 0.3117 | 0.2086 | 0.01255 | -0.01527 | 0.1175 | 0.05744 | 0.9918 | 0.9978 | 0.007968 | 0.006349 | 0.1628 | -0.1268 | 0.136 | 0.06699 | 1.007 | 1.01 | 0.9762 | 0.9854 | 0.4294 | 0.004491 |
| 0.3538 | -0.1666 | -0.5878 | 0.7242 | 0.799 | -0.05809 | -0.1019 | 0.08591 | 0.1591 | 1.013 | 1.014 | 0.006113 | 0.008472 | -0.09668 | -0.3153 | 0.2238 | 0.5608 | 1.025 | 1.031 | 1.001 | 0.9975 | 0.3419 | 0.7838 |
| 0.0198 | -0.00104 | 0.02742 | 0.07457 | 0.105 | -0.006616 | -0.006769 | 0.006618 | 0.006771 | 1.015 | 1.001 | 0.00801 | 0.006661 | -0.001441 | 0.02251 | 0.02389 | 0.01957 | 1.03 | 1.014 | 0.9988 | 0.9879 | 0.04539 | 0.06087 |
| 0.01111 | 0.09723 | 0.09865 | 0.133 | 0.1422 | -0.001966 | -0.002266 | 0.003401 | 0.004264 | 1.019 | 1.02 | 0.006206 | 0.009743 | 0.06399 | 0.05475 | 0.006647 | 0.01146 | 1.031 | 1.039 | 1.007 | 1.001 | 0.07702 | 0.07721 |
| 0.03277 | | 0.01288 | | 0.1414 | | -0.01079 | | 0.01079 | 0.9878 | 0.9963 | 0.007152 | 0.007843 | 0.08694 | 0.05253 | 0.05072 | 0.03601 | 1.002 | 1.012 | 0.9738 | 0.9809 | 0.1864 | 0.1231 |
| | -0.01392 | | 0.3604 | | -0.02118 | | 0.04152 | | 1.019 | | 0.006504 | | -0.01273 | | 0.11 | | 1.031 | | 1.006 | | 0.2029 | |
| 0.1681 | 0.2429 | -0.02024 | 0.9064 | 0.6389 | -0.04415 | -0.02641 | 0.06252 | 0.1173 | 1.002 | 1.006 | 0.008262 | 0.007007 | 0.363 | 0.03056 | 0.2444 | 0.2007 | 1.018 | 1.02 | 0.9855 | 0.9923 | 0.842 | 0.424 |
| 0.2098 | -0.3065 | -0.3985 | 0.5355 | 0.4239 | -0.07783 | -0.07511 | 0.07784 | 0.07511 | 1.024 | 1.003 | 0.007777 | 0.007262 | -0.323 | -0.03894 | 0.2655 | 0.2084 | 1.039 | 1.017 | 1.009 | 0.9883 | 0.1974 | 0.3694 |
| 0.1246 | -0.1225 | -0.1317 | 0.3368 | 0.3566 | -0.04451 | -0.03915 | 0.04451 | 0.0411 | 1.011 | 1.002 | 0.006633 | 0.007739 | -0.02134 | 0.04247 | 0.1364 | 0.1275 | 1.024 | 1.017 | 0.9975 | 0.9868 | 0.246 | 0.2923 |
| 0.02573 | -0.001654 | -0.02463 | 0.1183 | 0.07621 | -0.01144 | -0.00379 | 0.01144 | 0.01474 | 0.9976 | 0.9976 | 0.005865 | 0.007369 | 0.0585 | 0.04821 | 0.02586 | 0.02424 | 1.009 | 1.012 | 0.9861 | 0.9832 | 0.1092 | 0.09571 |
| 0.03647 | | -0.01165 | | 0.1313 | | -0.01212 | | 0.01212 | 1.002 | 1.001 | 0.007496 | 0.007723 | -0.01971 | 0.01024 | 0.05053 | 0.03624 | 1.016 | 1.016 | 0.9869 | 0.9862 | 0.07933 | 0.08126 |
| 0.01988 | 0.2009 | 0.1354 | 0.2661 | 0.2133 | -0.003854 | -0.003953 | 0.003856 | 0.008517 | 1.028 | 1.019 | 0.008105 | 0.008864 | 0.1171 | 0.07311 | 0.02227 | 0.01389 | 1.044 | 1.037 | 1.013 | 1.002 | 0.1608 | 0.1003 |
| 0.04065 | -0.003342 | -0.05862 | 0.1364 | 0.1007 | -0.01058 | -0.0136 | 0.01367 | 0.01361 | 0.9867 | 1.022 | 0.009423 | 0.007573 | 0.06648 | -0.1033 | 0.04333 | 0.03752 | 1.005 | 1.037 | 0.9683 | 1.008 | 0.1514 | -0.02973 |
| 0.01307 | 0.3531 | 0.3209 | 0.3934 | 0.3721 | 0.003673 | 0.006879 | 0.01041 | 0.01622 | 1.029 | 1.03 | 0.01094 | 0.0123 | 0.2527 | 0.1987 | 0.01608 | 0.02274 | 1.05 | 1.055 | 1.008 | 1.006 | 0.2842 | 0.2433 |
| 0.02404 | 0.3434 | 0.2811 | 0.4082 | 0.3754 | 4.33e-04 | 0.001289 | 0.01159 | 0.01865 | 1.002 | 1.031 | 0.007199 | 0.01068 | 0.2671 | 0.1285 | 0.01771 | 0.03975 | 1.016 | 1.052 | 0.9875 | 1.01 | 0.3018 | 0.2064 |
| 0.4786 | | -0.7377 | | 1.138 | | -0.15 | | 0.15 | 1.014 | 1 | 0.006105 | 0.00597 | -0.2027 | 0.4001 | 0.2847 | 0.5973 | 1.026 | 1.012 | 1.002 | 0.9884 | 0.3552 | 1.571 |
| 0.4733 | -0.7077 | -0.7106 | 0.8051 | 1.145 | -0.07099 | -0.1452 | 0.1758 | 0.1576 | 1.013 | 0.9988 | 0.006255 | 0.005519 | -0.3254 | 0.5439 | 0.3139 | 0.5794 | 1.025 | 1.01 | 1.001 | 0.988 | 0.2898 | 1.679 |

| gcta.SE_h2_male | gcta.h2.conf.low_female | gcta.h2.conf.low_male | gcta.h2.conf.high_female | gcta.h2.conf.high_male | gcta.x.h2.conf.low_female | gcta.x.h2.conf.low_male | gcta.x.h2.conf.high_female | gcta.x.h2.conf.high_male | ldsc.int_female | ldsc.int_male | ldsc.int_se_female | ldsc.int_se_male | ldsc.h2_female | ldsc.h2_male | ldsc.h2_se_female | ldsc.h2_se_male | ldsc.int.conf.high_female | ldsc.int.conf.high_male | ldsc.int.conf.low_female | ldsc.int.conf.low_male | ldsc.h2.conf.high_female | ldsc.h2.conf.high_male |
|---|---|---|---|---|---|---|---|---|---|---|---|---|---|---|---|---|---|---|---|---|---|---|
| 0.01058 | 0.0653 | 0.05429 | 0.09865 | 0.09577 | -5.81e-04 | -0.003292 | 0.005411 | 0.003294 | 1.025 | 0.9898 | 0.008066 | 0.008576 | 0.03335 | 0.06354 | 0.006645 | 0.01271 | 1.041 | 1.007 | 1.009 | 0.973 | 0.04637 | 0.08846 |
| 0.344 | 0.04466 | -0.3624 | 1.644 | 0.986 | -0.1286 | -0.1126 | 0.1286 | 0.1126 | 0.9975 | 0.9938 | 0.00984 | 0.007157 | 0.8077 | 0.7135 | 0.5553 | 0.4014 | 1.017 | 1.008 | 0.9783 | 0.9798 | 1.896 | 1.5 |
| 0.02375 | 0.1399 | 0.1179 | 0.2189 | 0.211 | -0.00584 | -0.002739 | 0.005842 | 0.01503 | 1.016 | 0.9984 | 0.009855 | 0.007413 | 0.0843 | 0.09368 | 0.02347 | 0.02468 | 1.035 | 1.013 | 0.9965 | 0.9839 | 0.1303 | 0.1421 |
|  | 0.03797 |  | 1.258 |  | -0.07449 |  | 0.1254 |  | 1.006 | 1.007 | 0.008126 | 0.006165 | 0.1103 | -0.88 | 0.3827 | 0.5234 | 1.022 | 1.019 | 0.9901 | 0.9946 | 0.8603 | 0.146 |
| 0.04222 | 0.07613 | -0.002668 | 0.2237 | 0.1628 | -0.01232 | -0.01287 | 0.01233 | 0.01382 | 1.003 | 1.007 | 0.006633 | 0.008512 | 0.09398 | 0.01545 | 0.03418 | 0.05189 | 1.016 | 1.024 | 0.9897 | 0.9902 | 0.161 | 0.1172 |
| 0.04144 | -0.03899 | -0.06743 | 0.09908 | 0.095 | -0.01202 | -0.003951 | 0.01395 | 0.03251 | 1.004 | 1.004 | 0.005843 | 0.006866 | -0.008144 | 0.004281 | 0.03145 | 0.04156 | 1.016 | 1.017 | 0.993 | 0.9901 | 0.05349 | 0.08574 |
| 0.6677 | -0.03966 | -0.7814 | 1.299 | 1.836 | -0.13 | -2.50e-04 | 0.13 | 0.5456 | 0.9937 | 0.9874 | 0.008341 | 0.006221 | 0.6123 | 1.152 | 0.4937 | 0.6473 | 1.01 | 0.9996 | 0.9774 | 0.9752 | 1.58 | 2.42 |
| 0.003336 |  | 9.73e-04 |  | 0.01405 |  | -0.001847 |  | 0.006892 | 1.004 | 1.004 | 0.005252 | 0.007212 | 0.01167 | 0.0154 | 0.01129 | 0.01203 | 1.014 | 1.019 | 0.9938 | 0.9903 | 0.0338 | 0.03897 |
|  | -0.1018 |  | 1.438 |  | -0.07501 |  | 0.2548 |  | 1.003 | 1.005 | 0.005853 | 0.008304 | 0.2271 | -0.2023 | 0.4593 | 0.4574 | 1.015 | 1.022 | 0.9919 | 0.989 | 1.127 | 0.6943 |
| 0.217 | -0.4085 | -0.2522 | 0.415 | 0.5986 | -0.07106 | -0.07499 | 0.07722 | 0.07499 | 0.9975 | 0.9861 | 0.007925 | 0.008239 | -0.02017 | 0.5873 | 0.161 | 0.2848 | 1.013 | 1.002 | 0.9819 | 0.9699 | 0.2954 | 1.146 |
| 0.2117 | -0.06617 | -0.3983 | 0.7964 | 0.4315 | -0.04484 | -0.07803 | 0.1003 | 0.07804 | 1.021 | 1.006 | 0.007017 | 0.006979 | -0.4351 | -0.05448 | 0.1921 | 0.2682 | 1.035 | 1.02 | 1.007 | 0.9922 | -0.05856 | 0.4713 |
| 0.219 | -0.3861 | -0.1687 | 0.4565 | 0.6896 | -0.05141 | -0.07489 | 0.1108 | 0.07489 | 0.9968 | 0.9911 | 0.006823 | 0.007756 | 0.01381 | 0.4423 | 0.1809 | 0.2468 | 1.01 | 1.006 | 0.9834 | 0.9759 | 0.3684 | 0.926 |
| 0.1316 | -0.123 | -0.1626 | 0.2744 | 0.3532 | -0.03327 | -0.04587 | 0.04431 | 0.04588 | 1.01 | 0.982 | 0.004324 | 0.007154 | -0.004458 | 0.4192 | 0.09105 | 0.1576 | 1.018 | 0.996 | 1.001 | 0.968 | 0.174 | 0.7282 |
| 0.1222 | -0.01539 | -0.1504 | 0.2127 | 0.3284 | -0.01464 | -0.04475 | 0.02527 | 0.04476 | 0.9971 | 1.011 | 0.004369 | 0.008869 | 0.03603 | -0.08534 | 0.04244 | 0.179 | 1.006 | 1.028 | 0.9885 | 0.9933 | 0.1192 | 0.2654 |
| 0.01388 | -0.00366 | 0.04198 | 0.004992 | 0.09639 | -0.003405 | -0.002084 | 0.003407 | 0.007687 | 1.01 | 1.003 | 0.005072 | 0.00747 | 0.03425 | 0.03998 | 0.007809 | 0.01741 | 1.02 | 1.018 | 1 | 0.9886 | 0.04956 | 0.0741 |
| 0.4179 |  | 0.08797 |  | 1.726 |  | -0.1538 |  | 0.1597 | 0.9887 | 0.9986 | 0.005658 | 0.007765 | 0.196 | 0.4182 | 0.2738 | 0.3493 | 0.9997 | 1.014 | 0.9776 | 0.9834 | 0.7326 | 1.103 |
|  | -0.02416 |  | 0.4663 |  | -0.02114 |  | 0.06458 |  | 1.004 | 0.995 | 0.005509 | 0.005499 | -0.02045 | 0.02199 | 0.1442 | 0.1407 | 1.015 | 1.006 | 0.9933 | 0.9842 | 0.2621 | 0.2978 |
| 0.2888 | -0.001989 | -0.1983 | 0.7989 | 0.9336 | -0.01998 | -0.09599 | 0.126 | 0.1081 | 1.005 | 1.005 | 0.005783 | 0.005473 | -0.07416 | 0.2921 | 0.1665 | 0.2998 | 1.016 | 1.015 | 0.9933 | 0.9938 | 0.2521 | 0.8797 |
| 0.01378 | -0.003066 | 0.02664 | 0.006326 | 0.08065 | -0.003623 | -1.05e-04 | 0.003785 | 0.01123 | 1.002 | 0.9973 | 0.007759 | 0.009761 | 0.02843 | 0.04145 | 0.01402 | 0.02121 | 1.017 | 1.016 | 0.9865 | 0.9781 | 0.05592 | 0.08303 |
|  | 0.01733 |  | 0.6375 |  | -0.04507 |  | 0.05691 |  | 1.008 |  | 0.007503 |  | -0.03161 |  | 0.1581 |  | 1.022 |  | 0.993 |  | 0.2782 |  |
|  | 0.01733 |  | 0.6375 |  | -0.04507 |  | 0.05691 |  | 1.008 |  | 0.007503 |  | -0.03161 |  | 0.1581 |  | 1.022 |  | 0.993 |  | 0.2782 |  |
|  | -0.281 |  | 0.3459 |  | -0.02398 |  | 0.09147 |  | 1.007 |  | 0.004939 |  | -0.2547 |  | 0.1218 |  | 1.017 |  | 0.9977 |  | -0.01603 |  |
|  |  |  |  |  |  |  |  |  | 1.002 |  | 0.004396 |  | -0.1156 |  | 0.09089 |  | 1.011 |  | 0.9933 |  | 0.06258 |  |
|  | -0.2023 |  | 0.4092 |  | -0.05401 |  | 0.05401 |  | 1.002 |  | 0.005397 |  | -0.001002 |  | 0.08777 |  | 1.013 |  | 0.9919 |  | 0.171 |  |
|  | -0.2741 |  | 0.3296 |  | -0.0551 |  | 0.0551 |  | 1.011 |  | 0.006071 |  | -0.2777 |  | 0.148 |  | 1.023 |  | 0.9992 |  | 0.01238 |  |
|  | -0.2996 |  | 0.3143 |  | -0.05557 |  | 0.05557 |  | 1.001 |  | 0.00634 |  | -0.03867 |  | 0.1403 |  | 1.013 |  | 0.9885 |  | 0.2363 |  |
|  | -0.1237 |  | 0.5155 |  | -0.04577 |  | 0.05357 |  | 1.002 |  | 0.008625 |  | -0.02449 |  | 0.236 |  | 1.019 |  | 0.9854 |  | 0.438 |  |
|  | -0.3063 |  | 0.3213 |  | -0.05792 |  | 0.05792 |  | 1.007 |  | 0.007516 |  | -0.1692 |  | 0.1351 |  | 1.021 |  | 0.9918 |  | 0.09569 |  |
|  | 0.03716 |  | 0.6545 |  | -0.04943 |  | 0.04943 |  | 1.001 |  | 0.006318 |  | 0.2143 |  | 0.1775 |  | 1.014 |  | 0.989 |  | 0.5623 |  |
|  |  |  |  |  |  |  |  |  | 0.9972 |  | 0.008032 |  | 0.002178 |  | 0.1387 |  | 1.013 |  | 0.9815 |  | 0.2741 |  |
|  |  |  |  |  |  |  |  |  | 1.002 |  | 0.00405 |  | -0.1629 |  | 0.1358 |  | 1.01 |  | 0.9943 |  | 0.1033 |  |

| gcta.SE_h2_male | gcta.h2.conf.low_female | gcta.h2.conf.low_male | gcta.h2.conf.high_female | gcta.h2.conf.high_male | gcta.x.h2.conf.low_female | gcta.x.h2.conf.low_male | gcta.x.h2.conf.high_female | gcta.x.h2.conf.high_male | ldsc.int_female | ldsc.int_male | ldsc.int_se_female | ldsc.int_se_male | ldsc.h2_female | ldsc.h2_male | ldsc.h2_se_female | ldsc.h2_se_male | ldsc.int.conf.high_female | ldsc.int.conf.high_male | ldsc.int.conf.low_female | ldsc.int.conf.low_male | ldsc.h2.conf.high_female | ldsc.h2.conf.high_male |
|---|---|---|---|---|---|---|---|---|---|---|---|---|---|---|---|---|---|---|---|---|---|---|
| 0.003225 | -0.002857 | -0.002116 | 0.007997 | 0.01053 | -0.002741 | -0.00263 | 0.006106 | 0.007793 | 1.001 | 0.992 | 0.009483 | 0.005446 | 0.04955 | 0.07376 | 0.01113 | 0.01337 | 1.02 | 1.003 | 0.9827 | 0.9814 | 0.07137 | 0.09997 |
| 0.7825 | -0.3281 | -1.055 | 0.3747 | 2.012 | -0.05608 | -0.2739 | 0.05608 | 0.2739 | 0.9963 | 1 | 0.008295 | 0.005046 | -0.05613 | -0.7957 | 0.2128 | 0.4941 | 1.013 | 1.01 | 0.98 | 0.9905 | 0.3609 | 0.1727 |
| 0.1116 | -0.07941 | -0.1938 | 0.3211 | 0.2437 | -0.03119 | -0.01424 | 0.04497 | 0.06288 | 0.9893 | 1.007 | 0.006082 | 0.009387 | 0.2149 | -0.1093 | 0.1221 | 0.1555 | 1.001 | 1.026 | 0.9774 | 0.989 | 0.4543 | 0.1955 |
| 0.09234 | -0.1165 | -0.05465 | 0.1987 | 0.3073 | -0.01136 | -0.03017 | 0.05793 | 0.03017 | 1.011 | 0.9854 | 0.008405 | 0.007574 | -0.08953 | 0.2465 | 0.1116 | 0.1026 | 1.027 | 1 | 0.9941 | 0.9706 | 0.1291 | 0.4476 |
| 0.03194 | 0.01861 | -0.01631 | 0.1131 | 0.1089 | -0.007939 | -0.00986 | 0.007941 | 0.01088 | 1.019 | 1.001 | 0.007398 | 0.00792 | -0.01029 | 0.016 | 0.02088 | 0.03369 | 1.034 | 1.016 | 1.005 | 0.9851 | 0.03063 | 0.08203 |
| 0.03077 | -0.03133 | -0.05246 | 0.1279 | 0.06814 | -0.01425 | -0.01173 | 0.01425 | 0.01173 | 0.9935 | 1.011 | 0.004051 | 0.006945 | 0.09652 | -0.009658 | 0.03162 | 0.03889 | 1.001 | 1.024 | 0.9855 | 0.9969 | 0.1585 | 0.06657 |
| 0.0104 | 0.03895 | 0.04029 | 0.07206 | 0.08105 | -0.002656 | -0.001631 | 0.00288 | 0.005272 | 1.005 | 1.005 | 0.007166 | 0.006783 | 0.03293 | 0.03547 | 0.01078 | 0.009497 | 1.019 | 1.018 | 0.9907 | 0.9913 | 0.05406 | 0.05408 |
|  | -0.01357 |  | 1.003 |  | -0.05625 |  | 0.1018 |  | 0.9959 | 0.9927 | 0.006872 | 0.005001 | 0.4269 | 0.5786 | 0.2189 | 0.3932 | 1.009 | 1.002 | 0.9824 | 0.9829 | 0.856 | 1.349 |
| 0.3916 | -0.01482 | 0.2283 | 1.039 | 1.763 | -0.03727 | -0.1542 | 0.1417 | 0.1542 | 0.9989 | 0.9863 | 0.006932 | 0.00646 | 0.3019 | 1.458 | 0.2515 | 0.4536 | 1.012 | 0.999 | 0.9853 | 0.9737 | 0.7949 | 2.347 |
| 0.0397 | -0.06247 | -0.05158 | 0.09657 | 0.104 | -0.001172 | -0.0146 | 0.0338 | 0.01461 | 0.9954 | 1.002 | 0.005342 | 0.007543 | 0.004775 | -0.01756 | 0.03214 | 0.04082 | 1.006 | 1.017 | 0.9849 | 0.9872 | 0.06777 | 0.06245 |
| 0.03122 | -0.04814 | -0.04007 | 0.09545 | 0.08233 | -0.01323 | -0.01191 | 0.01323 | 0.01191 | 0.9914 | 0.9959 | 0.007064 | 0.006814 | 0.06436 | 0.03605 | 0.04616 | 0.03178 | 1.005 | 1.009 | 0.9776 | 0.9825 | 0.1548 | 0.09835 |

| gcta.SE_h2_male | gcta.h2.conf.low_female | gcta.h2.conf.low_male | gcta.h2.conf.high_female | gcta.h2.conf.high_male | gcta.x.h2.conf.low_female | gcta.x.h2.conf.low_male | gcta.x.h2.conf.high_female | gcta.x.h2.conf.high_male | ldsc.int_female | ldsc.int_male | ldsc.int_se_female | ldsc.int_se_male | ldsc.h2_female | ldsc.h2_male | ldsc.h2_se_female | ldsc.h2_se_male | ldsc.int.conf.high_female | ldsc.int.conf.high_male | ldsc.int.conf.low_female | ldsc.int.conf.low_male | ldsc.h2.conf.high_female | ldsc.h2.conf.high_male |
|---|---|---|---|---|---|---|---|---|---|---|---|---|---|---|---|---|---|---|---|---|---|---|
| 0.3745 | -0.1277 | -0.6258 | 0.9211 | 0.8421 | -0.08126 | -0.1494 | 0.08126 | 0.1494 | 0.9981 | 1.002 | 0.008856 | 0.004972 | 0.3389 | 0.04577 | 0.3407 | 0.4212 | 1.015 | 1.011 | 0.9807 | 0.9918 | 1.007 | 0.8714 |
|  |  |  |  |  |  |  |  |  | 1.005 | 1.012 | 0.006933 | 0.007629 | -0.01247 | -0.09483 | 0.05174 | 0.04159 | 1.019 | 1.027 | 0.9914 | 0.9973 | 0.08894 | -0.01332 |
| 0.3514 |  | -0.2081 |  | 1.169 |  | -0.06967 |  | 0.1915 | 1.006 | 0.9912 | 0.007847 | 0.004957 | -0.3939 | 0.6223 | 0.3072 | 0.2773 | 1.021 | 1.001 | 0.9902 | 0.9815 | 0.2081 | 1.166 |
| 0.2443 | -0.3017 | 0.003912 | 0.4972 | 0.9615 | -0.07217 | -0.07404 | 0.07217 | 0.08795 | 0.9921 | 1.011 | 0.004688 | 0.004325 | 0.3924 | -0.1092 | 0.206 | 0.1914 | 1.001 | 1.019 | 0.9829 | 1.002 | 0.7962 | 0.2659 |
| 0.0192 | 0.06689 | 0.06396 | 0.1282 | 0.1392 | -0.003164 | -0.005602 | 0.007651 | 0.008034 | 1.017 | 1.012 | 0.009713 | 0.01018 | 0.03637 | 0.0415 | 0.01628 | 0.0225 | 1.036 | 1.032 | 0.998 | 0.9924 | 0.06828 | 0.0856 |
| 0.1843 | -0.1175 | -0.3296 | 0.2904 | 0.3926 | -0.02673 | -0.0368 | 0.05143 | 0.1162 | 1.011 | 1.001 | 0.005208 | 0.006614 | -0.1446 | -0.2874 | 0.1217 | 0.1752 | 1.021 | 1.014 | 1 | 0.9883 | 0.09399 | 0.05601 |
| 0.3093 | -0.1842 | -0.3943 | 0.4287 | 0.818 | -0.04668 | -0.07053 | 0.04668 | 0.1238 | 0.9824 | 1.009 | 0.007734 | 0.008052 | 0.3112 | -0.1195 | 0.2126 | 0.3802 | 0.9976 | 1.025 | 0.9673 | 0.9933 | 0.728 | 0.6256 |
| 0.05168 | 0.01652 | 0.04126 | 0.1698 | 0.2438 | -0.005989 | -0.01829 | 0.02333 | 0.01829 | 0.9956 | 1.01 | 0.006717 | 0.005816 | 0.1154 | 0.01022 | 0.04517 | 0.04131 | 1.009 | 1.021 | 0.9824 | 0.9984 | 0.2039 | 0.09119 |
| 0.1102 | -0.1793 | -0.2033 | 0.1813 | 0.2289 | -0.03272 | -0.039 | 0.03471 | 0.04053 | 0.9961 | 1.011 | 0.006734 | 0.006308 | 0.08242 | -0.1799 | 0.08021 | 0.1143 | 1.009 | 1.024 | 0.9829 | 0.9991 | 0.2396 | 0.04411 |
|  | -0.02147 |  | 0.2483 |  | -0.02688 |  | 0.02689 |  | 0.9952 | 0.9978 | 0.009516 | 0.008227 | 0.05562 | -0.01829 | 0.0865 | 0.09707 | 1.014 | 1.014 | 0.9765 | 0.9817 | 0.2252 | 0.172 |
|  | -0.05929 |  | 0.2086 |  | -0.00885 |  | 0.04651 |  | 1.002 | 1.007 | 0.007034 | 0.007579 | 0.03185 | -0.1136 | 0.07196 | 0.1172 | 1.015 | 1.022 | 0.9878 | 0.9919 | 0.1729 | 0.1161 |
| 0.02245 | 0.035 | 0.02307 | 0.1024 | 0.1111 | -0.00451 | -0.007625 | 0.006929 | 0.007627 | 1.015 | 1.012 | 0.006898 | 0.006988 | 0.02511 | 0.01908 | 0.01682 | 0.02818 | 1.028 | 1.026 | 1.001 | 0.9988 | 0.05808 | 0.07432 |
| 0.02376 | 0.07231 | 0.02382 | 0.1439 | 0.117 | -0.006781 | -0.007651 | 0.006783 | 0.007653 | 1.003 | 1.008 | 0.006755 | 0.00932 | 0.05946 | 0.04017 | 0.02045 | 0.02714 | 1.016 | 1.026 | 0.9897 | 0.9893 | 0.09954 | 0.09336 |
| 0.0428 | -0.06359 | -0.07737 | 0.1211 | 0.0904 | -0.01904 | -0.01622 | 0.01904 | 0.01879 | 1.003 | 0.9957 | 0.007299 | 0.007958 | -0.007829 | 0.06238 | 0.04767 | 0.05546 | 1.017 | 1.011 | 0.9884 | 0.9802 | 0.0856 | 0.1711 |
| 0.002899 | -0.002194 | -0.002573 | 0.007002 | 0.008791 | -0.003409 | -0.003303 | 0.003411 | 0.00522 | 1.011 | 1.011 | 0.00573 | 0.008285 | 0.06057 | 0.02315 | 0.008893 | 0.01406 | 1.022 | 1.027 | 0.9993 | 0.9947 | 0.078 | 0.0507 |
|  |  |  |  |  |  |  |  |  | 1.006 | 1.003 | 0.007944 | 0.005744 | -0.1388 | -0.7003 | 0.2012 | 0.2199 | 1.021 | 1.015 | 0.9902 | 0.9921 | 0.2555 | -0.2694 |

| gcta.SE_h2_male | gcta.h2.conf.low_female | gcta.h2.conf.low_male | gcta.h2.conf.high_female | gcta.h2.conf.high_male | gcta.x.h2.conf.low_female | gcta.x.h2.conf.low_male | gcta.x.h2.conf.high_female | gcta.x.h2.conf.high_male | ldsc.int_female | ldsc.int_male | ldsc.int_se_female | ldsc.int_se_male | ldsc.h2_female | ldsc.h2_male | ldsc.h2_se_female | ldsc.h2_se_male | ldsc.int.conf.high_female | ldsc.int.conf.high_male | ldsc.int.conf.low_female | ldsc.int.conf.low_male | ldsc.h2.conf.high_female | ldsc.h2.conf.high_male |
|---|---|---|---|---|---|---|---|---|---|---|---|---|---|---|---|---|---|---|---|---|---|---|
| | | | | | | | | | 0.9965 | 0.9892 | 0.006676 | 0.008546 | -0.002443 | 0.09442 | 0.04583 | 0.0446 | 1.01 | 1.006 | 0.9834 | 0.9725 | 0.08739 | 0.1818 |
| 0.002627 | -0.001765 | -0.004018 | 0.007239 | 0.00628 | -0.003227 | -0.003862 | 0.003229 | 0.003864 | 1.007 | 0.9895 | 0.006668 | 0.01016 | 0.04252 | 0.04281 | 0.006635 | 0.01621 | 1.02 | 1.009 | 0.9941 | 0.9696 | 0.05553 | 0.07459 |
| 0.03365 | 0.003424 | 0.05404 | 0.1564 | 0.186 | -0.01269 | -0.01155 | 0.01269 | 0.01156 | 1.007 | 0.9946 | 0.005991 | 0.008738 | 0.02398 | 0.1072 | 0.04021 | 0.03991 | 1.018 | 1.012 | 0.9949 | 0.9775 | 0.1028 | 0.1854 |
| 0.01171 | 0.1626 | 0.1477 | 0.1993 | 0.1936 | -0.001889 | -0.001315 | 0.002487 | 0.006033 | 1.013 | 0.9972 | 0.00826 | 0.006712 | 0.1055 | 0.1122 | 0.01448 | 0.01384 | 1.029 | 1.01 | 0.9963 | 0.984 | 0.1339 | 0.1393 |
| 0.01127 | 0.1235 | 0.1057 | 0.1588 | 0.1499 | -3.84e-04 | -0.003407 | 0.005013 | 0.003409 | 1 | 1.005 | 0.007977 | 0.008369 | 0.09636 | 0.0775 | 0.01263 | 0.01689 | 1.016 | 1.021 | 0.9847 | 0.9881 | 0.1211 | 0.1106 |
| 0.01056 | 0.009637 | 0.03692 | 0.03931 | 0.0783 | -0.00249 | -0.00311 | 0.002492 | 0.004037 | 1.014 | 1.006 | 0.009507 | 0.006776 | -0.003117 | 0.03334 | 0.009122 | 0.007741 | 1.033 | 1.019 | 0.9953 | 0.9927 | 0.01476 | 0.04852 |
| 0.07117 | -0.1135 | -0.1039 | 0.1689 | 0.175 | -0.02938 | -0.02691 | 0.02938 | 0.02691 | 1.001 | 1.011 | 0.007051 | 0.007291 | 0.03282 | -0.04752 | 0.08484 | 0.06929 | 1.015 | 1.026 | 0.9872 | 0.997 | 0.1991 | 0.08828 |
| 0.3176 | -0.1531 | -0.5971 | 1.298 | 0.6478 | -0.09963 | -0.08508 | 0.09963 | 0.1371 | 0.9942 | 0.9959 | 0.008695 | 0.005664 | 0.444 | -0.1932 | 0.4321 | 0.2964 | 1.011 | 1.007 | 0.9772 | 0.9848 | 1.291 | 0.3877 |
| 0.07065 | -0.06908 | -0.07942 | 0.2153 | 0.1975 | -0.01948 | -0.02601 | 0.03237 | 0.02601 | 0.9973 | 1.004 | 0.006214 | 0.01028 | 0.06887 | -0.02105 | 0.07028 | 0.1073 | 1.01 | 1.024 | 0.9851 | 0.9841 | 0.2066 | 0.1893 |
| 0.06648 | -0.0502 | 0.01555 | 0.2287 | 0.2762 | -0.0267 | -0.0227 | 0.0267 | 0.02271 | 1.008 | 0.993 | 0.003308 | 0.008256 | -0.0248 | 0.1686 | 0.06241 | 0.08018 | 1.015 | 1.009 | 1.002 | 0.9769 | 0.09753 | 0.3258 |
| 0.1661 | -0.3662 | -0.2739 | 0.396 | 0.3772 | -0.04902 | -0.0652 | 0.07649 | 0.0652 | 0.9931 | 0.9956 | 0.008414 | 0.008832 | 0.1903 | 0.08445 | 0.2082 | 0.2098 | 1.01 | 1.013 | 0.9766 | 0.9783 | 0.5984 | 0.4957 |
| 0.003126 | -0.001348 | -7.71e-04 | 0.009788 | 0.01148 | -0.001779 | -0.002346 | 0.007501 | 0.007366 | 1.006 | 0.9967 | 0.007987 | 0.008307 | 0.06945 | 0.0632 | 0.01692 | 0.01651 | 1.022 | 1.013 | 0.9903 | 0.9804 | 0.1026 | 0.09557 |
| 0.06508 | -0.1311 | -0.1262 | 0.135 | 0.1289 | -0.02282 | -0.02473 | 0.02671 | 0.02741 | 1.004 | 0.994 | 0.006733 | 0.007983 | -0.05655 | 0.0509 | 0.07083 | 0.07615 | 1.017 | 1.01 | 0.9905 | 0.9783 | 0.08228 | 0.2002 |
| 0.05829 | -0.09646 | -0.08125 | 0.2436 | 0.1472 | -0.009585 | -0.01322 | 0.06793 | 0.02739 | 1.005 | 0.9814 | 0.00807 | 0.008733 | 0.02827 | 0.1398 | 0.09484 | 0.06789 | 1.021 | 0.9986 | 0.9889 | 0.9643 | 0.2141 | 0.2728 |
| 0.0225 | 0.02076 | -0.01815 | 0.1006 | 0.07004 | -0.006888 | -0.005361 | 0.00689 | 0.011 | 1.002 | 0.9991 | 0.00594 | 0.007093 | 0.03365 | 0.02442 | 0.02128 | 0.02734 | 1.014 | 1.013 | 0.9906 | 0.9852 | 0.07536 | 0.07801 |
| 0.3675 | -0.256 | -0.3322 | 0.8936 | 1.108 | -0.08027 | -0.1362 | 0.08027 | 0.1362 | 1.01 | 0.9953 | 0.006914 | 0.01022 | 0.03533 | 0.5455 | 0.2486 | 0.4391 | 1.023 | 1.015 | 0.9964 | 0.9753 | 0.5227 | 1.406 |
| 0.02454 | 0.08477 | 0.0661 | 0.1571 | 0.1623 | -0.004931 | -0.008719 | 0.006464 | 0.008721 | 1.002 | 1.008 | 0.006615 | 0.008641 | 0.07278 | 0.05329 | 0.01647 | 0.03068 | 1.015 | 1.025 | 0.9893 | 0.9911 | 0.1051 | 0.1134 |
| 0.0485 | -0.06396 | -0.09186 | 0.1283 | 0.09825 | -0.01512 | -0.01327 | 0.0202 | 0.0193 | 1.006 | 1.014 | 0.005702 | 0.007817 | -0.0531 | -0.05124 | 0.04384 | 0.05443 | 1.018 | 1.03 | 0.9953 | 0.9989 | 0.03282 | 0.05544 |
| 0.003462 | -7.98e-04 | 6.90e-04 | 0.009442 | 0.01426 | -0.002146 | -9.65e-04 | 0.005364 | 0.008553 | 1.014 | 1.022 | 0.008706 | 0.006852 | 0.08091 | 0.04756 | 0.01253 | 0.01937 | 1.031 | 1.036 | 0.9971 | 1.009 | 0.1055 | 0.08553 |
| 0.02355 | 0.08228 | 0.04763 | 0.1525 | 0.1399 | -0.004499 | -0.006565 | 0.007446 | 0.006736 | 0.9972 | 1.001 | 0.006991 | 0.007297 | 0.08781 | 0.07777 | 0.02672 | 0.02905 | 1.011 | 1.016 | 0.9835 | 0.9869 | 0.1402 | 0.1347 |
| | -0.2995 | | 0.4839 | | -0.04779 | | 0.08833 | | 1.003 | 0.998 | 0.00576 | 0.006777 | 0.0484 | -0.4308 | 0.1892 | 0.316 | 1.015 | 1.011 | 0.9921 | 0.9847 | 0.4193 | 0.1887 |
| 0.04698 | -0.06997 | -0.08788 | 0.1206 | 0.09629 | -0.01857 | -0.01341 | 0.01857 | 0.02165 | 0.9971 | 1.006 | 0.005768 | 0.007463 | 0.03796 | -0.06641 | 0.04768 | 0.05069 | 1.008 | 1.021 | 0.9858 | 0.9913 | 0.1314 | 0.03293 |
| 0.00354 | 8.25e-04 | 0.003599 | 0.01177 | 0.01748 | -4.55e-04 | -3.58e-04 | 0.008168 | 0.009588 | 1.011 | 1.019 | 0.006448 | 0.007936 | 0.08998 | 0.06179 | 0.01097 | 0.01597 | 1.023 | 1.035 | 0.9982 | 1.004 | 0.1115 | 0.0931 |
| 0.016 | 0.2855 | 0.2202 | 0.3347 | 0.2829 | 0.002796 | 0.00356 | 0.01073 | 0.0141 | 1.024 | 1.026 | 0.008008 | 0.005898 | 0.1969 | 0.1434 | 0.02381 | 0.02308 | 1.04 | 1.037 | 1.009 | 1.014 | 0.2435 | 0.1886 |
| 0.06374 | -0.09982 | 0.002221 | 0.1603 | 0.2521 | -0.01913 | -0.02107 | 0.0291 | 0.02107 | 1.015 | 1.003 | 0.006656 | 0.007934 | -0.05532 | 0.03833 | 0.05242 | 0.07359 | 1.028 | 1.019 | 1.002 | 0.9875 | 0.04742 | 0.1826 |
| 0.03293 | 0.006434 | -0.009888 | 0.1807 | 0.1192 | -0.00939 | -0.004904 | 0.01963 | 0.01843 | 0.9975 | 1.002 | 0.00948 | 0.008148 | 0.07476 | 0.03318 | 0.06247 | 0.03973 | 1.016 | 1.018 | 0.9789 | 0.9864 | 0.1972 | 0.111 |
| 0.01069 | 0.05219 | 0.05689 | 0.08549 | 0.0988 | -8.81e-04 | 2.77e-04 | 0.00491 | 0.007031 | 1.003 | 0.9997 | 0.009024 | 0.007085 | 0.04767 | 0.05983 | 0.009788 | 0.01181 | 1.021 | 1.014 | 0.9857 | 0.9858 | 0.06686 | 0.08298 |
| | -0.3547 | | 0.4262 | | -0.06186 | | 0.06186 | | 1.015 | 1 | 0.006671 | 0.006434 | -0.1848 | -0.08242 | 0.221 | 0.1235 | 1.028 | 1.013 | 1.002 | 0.9878 | 0.2483 | 0.1597 |
| 0.02209 | 0.04525 | 0.02503 | 0.1122 | 0.1116 | -0.00331 | -0.006436 | 0.008342 | 0.006438 | 1.007 | 1.012 | 0.005257 | 0.006773 | 0.04227 | 0.02064 | 0.01541 | 0.02467 | 1.017 | 1.026 | 0.9966 | 0.999 | 0.07247 | 0.06898 |

| gcta.SE_h2_male | gcta.h2.conf.low_female | gcta.h2.conf.low_male | gcta.h2.conf.high_female | gcta.h2.conf.high_male | gcta.x.h2.conf.low_female | gcta.x.h2.conf.low_male | gcta.x.h2.conf.high_female | gcta.x.h2.conf.high_male | ldsc.int_female | ldsc.int_male | ldsc.int_se_female | ldsc.int_se_male | ldsc.h2_female | ldsc.h2_male | ldsc.h2_se_female | ldsc.h2_se_male | ldsc.int.conf.high_female | ldsc.int.conf.high_male | ldsc.int.conf.low_female | ldsc.int.conf.low_male | ldsc.h2.conf.high_female | ldsc.h2.conf.high_male |
|---|---|---|---|---|---|---|---|---|---|---|---|---|---|---|---|---|---|---|---|---|---|---|
| 0.04761 | -0.05406 | -0.09184 | 0.1425 | 0.09477 | -0.01813 | -0.0141 | 0.01813 | 0.01681 | 1.016 | 0.9918 | 0.008182 | 0.006169 | -0.07032 | 0.04105 | 0.05488 | 0.04057 | 1.032 | 1.004 | 0.9999 | 0.9797 | 0.03725 | 0.1206 |
| 0.002777 | -0.003002 | -0.0037 | 0.006622 | 0.007186 | -0.002219 | -0.003558 | 0.005469 | 0.004484 | 1.009 | 1.005 | 0.006611 | 0.008855 | 0.04764 | 0.05281 | 0.01162 | 0.01905 | 1.022 | 1.022 | 0.9964 | 0.9878 | 0.07041 | 0.09015 |
| | -0.05686 | | 0.125 | | -0.01749 | | 0.0175 | | 0.999 | 0.9986 | 0.00875 | 0.007526 | 0.01803 | 0.01502 | 0.06142 | 0.05074 | 1.016 | 1.013 | 0.9818 | 0.9838 | 0.1384 | 0.1145 |
| 0.002506 | -0.00294 | -0.003841 | 0.005724 | 0.005983 | -0.003315 | -0.003528 | 0.003317 | 0.003782 | 1.002 | 0.9892 | 0.006697 | 0.00852 | 0.04202 | 0.0488 | 0.00758 | 0.01528 | 1.015 | 1.006 | 0.9891 | 0.9725 | 0.05688 | 0.07875 |
| 0.1195 | -0.05027 | 0.03831 | 0.478 | 0.5069 | -0.02084 | -0.002654 | 0.07843 | 0.09128 | 0.9997 | 0.9921 | 0.007291 | 0.008741 | 0.08942 | 0.2884 | 0.1511 | 0.146 | 1.014 | 1.009 | 0.9854 | 0.975 | 0.3856 | 0.5746 |
| 0.02025 | 0.009636 | -0.03847 | 0.07122 | 0.0409 | -0.001851 | -0.005623 | 0.01297 | 0.008453 | 1.002 | 1.001 | 0.007031 | 0.007813 | 0.03308 | -0.004968 | 0.01416 | 0.01848 | 1.016 | 1.016 | 0.9883 | 0.9852 | 0.06083 | 0.03126 |
| 0.01168 | 0.001038 | 0.02755 | 0.03361 | 0.07333 | -0.003061 | -0.003943 | 0.003063 | 0.003945 | 1.007 | 1.013 | 0.007696 | 0.005854 | -0.009097 | 0.0133 | 0.007408 | 0.008889 | 1.022 | 1.024 | 0.9921 | 1.001 | 0.005423 | 0.03072 |
| | -0.2587 | | 0.7127 | | -0.08662 | | 0.08662 | | 1.001 | 1.004 | 0.006729 | 0.005808 | 0.3087 | -0.2361 | 0.2733 | 0.1416 | 1.014 | 1.015 | 0.9878 | 0.9924 | 0.8442 | 0.04156 |
| | | | | | | | | | 0.9963 | 1.009 | 0.006995 | 0.006147 | -0.03919 | -0.42 | 0.2527 | 0.2161 | 1.01 | 1.021 | 0.9826 | 0.997 | 0.4561 | 0.00359 |
| 0.08735 | 0.03293 | -0.1582 | 0.2369 | 0.1842 | -0.01768 | -0.01788 | 0.01768 | 0.04341 | 1.013 | 1.007 | 0.006654 | 0.00607 | 0.04141 | -0.03562 | 0.04347 | 0.0903 | 1.026 | 1.019 | 0.9997 | 0.9955 | 0.1266 | 0.1414 |
| 0.02407 | 0.07962 | 0.05625 | 0.1522 | 0.1506 | -0.002491 | -0.006761 | 0.009771 | 0.006763 | 0.9967 | 1.011 | 0.006923 | 0.006678 | 0.07782 | 0.04714 | 0.01951 | 0.02425 | 1.01 | 1.024 | 0.9831 | 0.9977 | 0.1161 | 0.09467 |
| 0.01287 | 0.06451 | 0.05586 | 0.1054 | 0.1063 | -0.001716 | -0.002159 | 0.005179 | 0.005791 | 1.009 | 1.007 | 0.007481 | 0.00669 | 0.046 | 0.05616 | 0.008885 | 0.01362 | 1.024 | 1.02 | 0.9942 | 0.9934 | 0.06342 | 0.08285 |
| 0.02557 | | -0.03122 | | 0.06903 | | -0.003601 | | 0.01601 | 1.005 | 1.002 | 0.006948 | 0.008734 | -0.007156 | -3.30e-04 | 0.02683 | 0.02672 | 1.019 | 1.019 | 0.9913 | 0.9852 | 0.04543 | 0.05204 |
| 0.03925 | -0.03477 | -0.06658 | 0.1817 | 0.08726 | -0.006636 | -0.01414 | 0.03282 | 0.01414 | 1.008 | 1.005 | 0.007264 | 0.006989 | -0.01817 | 0.001001 | 0.06222 | 0.04109 | 1.022 | 1.019 | 0.994 | 0.9912 | 0.1038 | 0.08154 |
| 0.0117 | 0.03791 | 0.02962 | 0.07518 | 0.07546 | -0.003061 | -0.003835 | 0.003063 | 0.003837 | 1.003 | 1.009 | 0.01005 | 0.006437 | 0.03833 | 0.03607 | 0.01191 | 0.01292 | 1.023 | 1.021 | 0.9832 | 0.9963 | 0.06167 | 0.0614 |

| gcta.SE_h2_male | gcta.h2.conf.low_female | gcta.h2.conf.low_male | gcta.h2.conf.high_female | gcta.h2.conf.high_male | gcta.x.h2.conf.low_female | gcta.x.h2.conf.low_male | gcta.x.h2.conf.high_female | gcta.x.h2.conf.high_male | ldsc.int_female | ldsc.int_male | ldsc.int_se_female | ldsc.int_se_male | ldsc.h2_female | ldsc.h2_male | ldsc.h2_se_female | ldsc.h2_se_male | ldsc.int.conf.high_female | ldsc.int.conf.high_male | ldsc.int.conf.low_female | ldsc.int.conf.low_male | ldsc.h2.conf.high_female | ldsc.h2.conf.high_male |
|---|---|---|---|---|---|---|---|---|---|---|---|---|---|---|---|---|---|---|---|---|---|---|
| 0.05519 | -0.03124 | -0.03919 | 0.2347 | 0.1772 | -0.01304 | -0.01695 | 0.03701 | 0.01696 | 1 | 1.019 | 0.003932 | 0.00777 | 0.04874 | -0.04857 | 0.03622 | 0.06196 | 1.008 | 1.034 | 0.9923 | 1.003 | 0.1197 | 0.07288 |
| 0.05451 | -0.0755 | -0.05298 | 0.1741 | 0.1607 | -0.0198 | -0.02045 | 0.021 | 0.02045 | 0.9948 | 1.002 | 0.009406 | 0.005446 | 0.06492 | 0.04247 | 0.08721 | 0.06208 | 1.013 | 1.013 | 0.9764 | 0.9916 | 0.2359 | 0.1642 |
| | | | | | | | | | 1.005 | | 0.005304 | | -0.2263 | | 0.2023 | | 1.016 | | 0.9948 | | 0.1702 | |
| | -0.3387 | | 0.4868 | | -0.06519 | | 0.06519 | | 0.9934 | | 0.008258 | | 0.01645 | | 0.3038 | | 1.01 | | 0.9772 | | 0.6118 | |
| 0.1438 | -0.2451 | -0.2447 | 0.2717 | 0.3188 | -0.04784 | -0.05037 | 0.04784 | 0.05037 | 1.005 | 1.013 | 0.007452 | 0.006048 | 0.01117 | -0.1858 | 0.1479 | 0.1013 | 1.019 | 1.025 | 0.9902 | 1.002 | 0.3011 | 0.01285 |
| 0.03903 | 0.01044 | -0.03326 | 0.1499 | 0.1197 | -0.00662 | -0.01414 | 0.02136 | 0.01414 | 0.9762 | 0.998 | 0.006541 | 0.007801 | 0.1604 | 0.05053 | 0.03054 | 0.03852 | 0.989 | 1.013 | 0.9634 | 0.9827 | 0.2202 | 0.126 |
| 0.04196 | -0.0259 | -0.06411 | 0.1146 | 0.1004 | -0.01038 | -0.01808 | 0.01585 | 0.01808 | 0.9988 | 1.003 | 0.00627 | 0.005903 | 0.03258 | 0.004579 | 0.03337 | 0.0367 | 1.011 | 1.014 | 0.9865 | 0.9912 | 0.09799 | 0.07652 |
| 0.00297 | -0.005068 | -0.00394 | 0.00512 | 0.007702 | -0.004252 | -0.002958 | 0.004254 | 0.006273 | 1.022 | 0.9953 | 0.006673 | 0.006651 | 0.01606 | 0.05648 | 0.008321 | 0.01551 | 1.035 | 1.008 | 1.009 | 0.9823 | 0.03237 | 0.08688 |
| 0.01449 | 0.01131 | 0.02108 | 0.06617 | 0.07788 | -0.004581 | -0.004801 | 0.004583 | 0.004803 | 1 | 0.993 | 0.0106 | 0.007756 | 0.03531 | 0.05291 | 0.02079 | 0.01298 | 1.021 | 1.008 | 0.9795 | 0.9777 | 0.07606 | 0.07834 |
| 0.647 | -0.4726 | -0.8262 | 0.7005 | 1.71 | -0.05999 | -0.2152 | 0.1519 | 0.2152 | 1.01 | 1.007 | 0.008165 | 0.006058 | -0.1264 | -0.3977 | 0.3794 | 0.5701 | 1.026 | 1.018 | 0.9938 | 0.9947 | 0.6172 | 0.7198 |
| 0.05283 | -0.01395 | -0.0931 | 0.2252 | 0.114 | -0.02424 | -0.01384 | 0.02424 | 0.02884 | 0.9903 | 0.996 | 0.006843 | 0.009056 | 0.1248 | 0.00667 | 0.05795 | 0.06481 | 1.004 | 1.014 | 0.9769 | 0.9782 | 0.2383 | 0.1337 |
| 0.7239 | | -1.147 | | 1.69 | | -0.243 | | 0.243 | 1.008 | 0.9999 | 0.008951 | 0.007103 | -0.8649 | 0.5386 | 0.5625 | 0.5993 | 1.026 | 1.014 | 0.9909 | 0.986 | 0.2376 | 1.713 |

| gcta.SE_h2_male | gcta.h2.conf.low_female | gcta.h2.conf.low_male | gcta.h2.conf.high_female | gcta.h2.conf.high_male | gcta.x.h2.conf.low_female | gcta.x.h2.conf.low_male | gcta.x.h2.conf.high_female | gcta.x.h2.conf.high_male | ldsc.int_female | ldsc.int_male | ldsc.int_se_female | ldsc.int_se_male | ldsc.h2_female | ldsc.h2_male | ldsc.h2_se_female | ldsc.h2_se_male | ldsc.int.conf.high_female | ldsc.int.conf.high_male | ldsc.int.conf.low_female | ldsc.int.conf.low_male | ldsc.h2.conf.high_female | ldsc.h2.conf.high_male |
|---|---|---|---|---|---|---|---|---|---|---|---|---|---|---|---|---|---|---|---|---|---|---|
| 0.0283 | 0.01056 | -0.04978 | 0.1503 | 0.06116 | -0.01117 | -0.004429 | 0.01117 | 0.01507 | 0.9978 | 1.003 | 0.007116 | 0.006256 | 0.09149 | -0.01741 | 0.02648 | 0.03457 | 1.012 | 1.015 | 0.9839 | 0.9903 | 0.1434 | 0.05035 |
| | -0.1374 | | 0.2122 | | -0.02885 | | 0.02886 | | 1.006 | 0.9916 | 0.008941 | 0.006305 | -0.009249 | 0.08129 | 0.128 | 0.09896 | 1.024 | 1.004 | 0.9889 | 0.9793 | 0.2416 | 0.2752 |
| 0.01034 | 0.06295 | 0.04104 | 0.096 | 0.08156 | -7.71e-04 | -0.003229 | 0.005148 | 0.003231 | 1.017 | 0.9865 | 0.007121 | 0.009494 | 0.03886 | 0.05892 | 0.006566 | 0.01407 | 1.031 | 1.005 | 1.003 | 0.9679 | 0.05173 | 0.0865 |
| 0.5504 | -0.8386 | -0.07876 | 0.8704 | 2.079 | -0.1062 | -0.1566 | 0.1386 | 0.282 | 1.014 | 0.9937 | 0.006168 | 0.007755 | -0.4765 | 0.9021 | 0.432 | 0.5585 | 1.027 | 1.009 | 1.002 | 0.9785 | 0.3703 | 1.997 |
| 0.484 | | 0.05127 | | 1.949 | | -0.1346 | | 0.2573 | 1.015 | 0.997 | 0.007608 | 0.008012 | -0.4816 | 0.857 | 0.581 | 0.6482 | 1.03 | 1.013 | 0.9997 | 0.9813 | 0.6571 | 2.128 |
| 0.1596 | -0.2625 | -0.1567 | 0.2654 | 0.469 | -0.04295 | -0.02444 | 0.04588 | 0.1073 | 1.019 | 0.9941 | 0.004499 | 0.007334 | -0.5442 | 0.393 | 0.09226 | 0.2063 | 1.028 | 1.008 | 1.01 | 0.9797 | -0.3634 | 0.7973 |
| 0.1607 | -0.04557 | 0.3507 | 0.5433 | 0.9804 | -0.03955 | -0.0546 | 0.03955 | 0.0546 | 1.013 | 1.017 | 0.006677 | 0.008155 | -0.06525 | 0.1153 | 0.1536 | 0.1427 | 1.026 | 1.033 | 0.9999 | 1.001 | 0.2359 | 0.3951 |
| 0.188 | -0.09601 | 0.1519 | 0.5606 | 0.8888 | -0.04703 | -0.06139 | 0.04821 | 0.0614 | 1.011 | 1.013 | 0.007024 | 0.007609 | -0.08043 | -0.06818 | 0.2216 | 0.1278 | 1.025 | 1.028 | 0.997 | 0.9979 | 0.354 | 0.1824 |
| 0.1593 | -0.2616 | -0.07218 | 0.2721 | 0.5523 | -0.03932 | -0.01226 | 0.04996 | 0.1233 | 1.015 | 0.9925 | 0.004906 | 0.006443 | -0.3823 | 0.47 | 0.101 | 0.1851 | 1.025 | 1.005 | 1.005 | 0.9799 | -0.1843 | 0.8328 |
| 0.5282 | -0.8356 | -0.8635 | 0.8734 | 1.207 | -0.1402 | -0.1578 | 0.1402 | 0.1578 | 0.9965 | 0.9923 | 0.008524 | 0.007087 | 0.3089 | 0.4604 | 0.4917 | 0.5724 | 1.013 | 1.006 | 0.9798 | 0.9784 | 1.273 | 1.582 |
| 0.0503 | -0.1017 | -0.0941 | 0.1255 | 0.1031 | -0.01634 | -0.01439 | 0.01836 | 0.02311 | 1.003 | 1.008 | 0.005849 | 0.007135 | 0.01792 | -0.08516 | 0.05644 | 0.06239 | 1.014 | 1.022 | 0.9915 | 0.9943 | 0.1285 | 0.03712 |
| 0.0259 | -0.001822 | 0.004923 | 0.09582 | 0.1065 | -0.007795 | -0.005759 | 0.00928 | 0.01351 | 1.008 | 1.014 | 0.004898 | 0.007903 | 0.01147 | -0.01194 | 0.02199 | 0.02575 | 1.017 | 1.029 | 0.998 | 0.9982 | 0.05457 | 0.03853 |
| 0.5518 | -0.8194 | -0.08153 | 0.8721 | 2.082 | -0.1249 | -0.1023 | 0.1249 | 0.3152 | 1.011 | 0.9937 | 0.005534 | 0.005724 | -0.3762 | 0.7503 | 0.3926 | 0.4487 | 1.022 | 1.005 | 1 | 0.9825 | 0.3932 | 1.63 |
| 0.4986 | -0.7423 | 0.02273 | 0.9401 | 1.977 | -0.122 | -0.1341 | 0.122 | 0.2054 | 1.006 | 0.9953 | 0.004729 | 0.006581 | -0.3338 | 0.612 | 0.3514 | 0.4998 | 1.015 | 1.008 | 0.9966 | 0.9824 | 0.3551 | 1.592 |
| 0.155 | -0.2346 | -0.2839 | 0.3101 | 0.3237 | -0.01877 | -0.03778 | 0.0959 | 0.07665 | 1.007 | 0.9978 | 0.005113 | 0.007202 | -0.2154 | 0.132 | 0.08863 | 0.1737 | 1.017 | 1.012 | 0.9966 | 0.9837 | -0.04165 | 0.4726 |
| 0.1605 | -0.1032 | 0.2953 | 0.4895 | 0.9243 | -0.04178 | -0.05649 | 0.04178 | 0.05649 | 1.01 | 1.013 | 0.006469 | 0.007941 | -0.04353 | 0.1955 | 0.1467 | 0.162 | 1.023 | 1.029 | 0.9975 | 0.9974 | 0.244 | 0.513 |
| 0.157 | -0.2399 | -0.258 | 0.2943 | 0.3574 | -0.02506 | -0.03148 | 0.07825 | 0.09103 | 1.008 | 0.9987 | 0.0051 | 0.008024 | -0.1642 | 0.1278 | 0.0946 | 0.2015 | 1.018 | 1.014 | 0.998 | 0.983 | 0.02116 | 0.5228 |
| | -0.3388 | | 1.238 | | -0.116 | | 0.1617 | | 0.9982 | 1.014 | 0.007099 | 0.00593 | 0.6038 | -0.9671 | 0.4588 | 0.5394 | 1.012 | 1.026 | 0.9843 | 1.003 | 1.503 | 0.09013 |
| 0.01125 | 0.1134 | 0.08779 | 0.1493 | 0.1319 | -7.53e-04 | -1.32e-05 | 0.005109 | 0.007727 | 1.027 | 1.011 | 0.01083 | 0.007793 | 0.06251 | 0.06842 | 0.009853 | 0.0145 | 1.048 | 1.027 | 1.006 | 0.9961 | 0.08182 | 0.09684 |
| 0.05597 | -0.1147 | -0.1029 | 0.1921 | 0.1165 | -0.02681 | -0.0134 | 0.02682 | 0.02683 | 1.012 | 0.993 | 0.007103 | 0.01004 | -0.0236 | -0.0103 | 0.0723 | 0.05864 | 1.026 | 1.013 | 0.998 | 0.9733 | 0.1181 | 0.1046 |
| 0.05593 | -0.1101 | -0.09505 | 0.111 | 0.1242 | -0.02333 | -0.01787 | 0.02334 | 0.02398 | 1 | 1.006 | 0.008949 | 0.004994 | 0.001841 | -0.04091 | 0.08569 | 0.04446 | 1.018 | 1.016 | 0.9828 | 0.9961 | 0.1698 | 0.04624 |
| 0.02975 | -0.003886 | -0.01954 | 0.1356 | 0.09707 | -0.005517 | -0.01064 | 0.01679 | 0.01064 | 1.001 | 1 | 0.005997 | 0.006948 | 0.002064 | 0.01691 | 0.03937 | 0.02878 | 1.013 | 1.014 | 0.9892 | 0.9868 | 0.07922 | 0.07331 |
| | -0.1102 | | 0.4017 | | -0.02915 | | 0.05705 | | 0.9986 | 0.9939 | 0.007281 | 0.008117 | 0.09291 | 0.03316 | 0.1306 | 0.1359 | 1.013 | 1.01 | 0.9844 | 0.978 | 0.3488 | 0.2995 |
| 0.04194 | 0.044 | -0.03398 | 0.1893 | 0.1304 | -0.01171 | -0.01024 | 0.01171 | 0.02089 | 1.017 | 1.003 | 0.00713 | 0.005482 | -0.02039 | 0.02249 | 0.04054 | 0.03214 | 1.031 | 1.014 | 1.003 | 0.9922 | 0.05908 | 0.08548 |
| 0.003913 | -0.005088 | 0.006817 | 0.0055 | 0.02216 | -0.004366 | 0.002984 | 0.004368 | 0.01566 | 1.009 | 1.006 | 0.00602 | 0.006826 | 0.06329 | 0.04955 | 0.01464 | 0.01273 | 1.021 | 1.019 | 0.9974 | 0.9927 | 0.092 | 0.07449 |
| | -0.1537 | | 0.8794 | | -0.02971 | | 0.1461 | | 0.995 | 0.9872 | 0.006865 | 0.007209 | 0.2082 | 1.178 | 0.2927 | 0.5402 | 1.008 | 1.001 | 0.9815 | 0.9731 | 0.7818 | 2.236 |
| | -0.01186 | | 0.2982 | | -0.005125 | | 0.06106 | | 1.007 | 1.016 | 0.006951 | 0.007941 | -0.01666 | -0.1729 | 0.08208 | 0.07209 | 1.02 | 1.032 | 0.993 | 1.001 | 0.1442 | -0.03166 |
| 0.4285 | | -0.5104 | | 1.169 | | -0.1587 | | 0.1587 | 1.009 | 1.014 | 0.008151 | 0.007468 | -0.5164 | -0.2658 | 0.5707 | 0.3403 | 1.025 | 1.029 | 0.9931 | 0.9993 | 0.6022 | 0.4012 |
| | -0.0411 | | 0.8757 | | -0.04063 | | 0.1012 | | 1.003 | | 0.006118 | | 0.1224 | | 0.2431 | | 1.015 | | 0.9913 | | 0.599 | |

| gcta.SE_h2_male | gcta.h2.conf.low_female | gcta.h2.conf.low_male | gcta.h2.conf.high_female | gcta.h2.conf.high_male | gcta.x.h2.conf.low_female | gcta.x.h2.conf.low_male | gcta.x.h2.conf.high_female | gcta.x.h2.conf.high_male | ldsc.int_female | ldsc.int_male | ldsc.int_se_female | ldsc.int_se_male | ldsc.h2_female | ldsc.h2_male | ldsc.h2_se_female | ldsc.h2_se_male | ldsc.int.conf.high_female | ldsc.int.conf.high_male | ldsc.int.conf.low_female | ldsc.int.conf.low_male | ldsc.h2.conf.high_female | ldsc.h2.conf.high_male |
|---|---|---|---|---|---|---|---|---|---|---|---|---|---|---|---|---|---|---|---|---|---|---|
| | -0.08092 | | 0.3687 | | -0.02362 | | 0.07386 | | 0.9961 | 0.9985 | 0.007224 | 0.006729 | 0.1907 | -0.1542 | 0.138 | 0.1585 | 1.01 | 1.012 | 0.9819 | 0.9853 | 0.4613 | 0.1563 |
| 0.1596 | | -0.2263 | | 0.3993 | | -0.06128 | | 0.06128 | 0.9849 | 0.993 | 0.007381 | 0.006588 | 0.2742 | 0.1527 | 0.2112 | 0.1482 | 0.9994 | 1.006 | 0.9705 | 0.9801 | 0.6882 | 0.4432 |
| 0.02939 | | 0.004198 | | 0.1194 | | -0.00667 | | 0.01314 | | 1.007 | | 0.008806 | | 0.02176 | | 0.03156 | | 1.025 | | 0.9902 | | 0.08362 |
| 0.0143 | -0.004864 | 0.01592 | 0.04824 | 0.07196 | -0.004727 | -0.004854 | 0.004729 | 0.004856 | 0.9961 | 1.001 | 0.01074 | 0.007765 | 0.02517 | 0.03105 | 0.01836 | 0.01277 | 1.017 | 1.016 | 0.9751 | 0.9857 | 0.06116 | 0.05608 |
| 0.01605 | 0.002991 | 0.01596 | 0.06112 | 0.07888 | -0.003887 | -0.003554 | 0.006601 | 0.007607 | 1.002 | 1.018 | 0.005298 | 0.006155 | 0.01338 | -0.001177 | 0.01501 | 0.01465 | 1.013 | 1.031 | 0.9917 | 1.006 | 0.04281 | 0.02754 |
| 0.02572 | | -0.01811 | | 0.0827 | | -0.001394 | | 0.01599 | 1.003 | 1.002 | 0.008246 | 0.005855 | 0.003347 | 0.01266 | 0.02909 | 0.01992 | 1.019 | 1.014 | 0.9869 | 0.9906 | 0.06036 | 0.0517 |
| 0.0226 | -0.008141 | 0.02819 | 0.04937 | 0.1168 | -0.002234 | -0.003413 | 0.009768 | 0.01344 | 1.007 | 1.008 | 0.007511 | 0.008341 | -1.19e-04 | 0.04166 | 0.01651 | 0.03015 | 1.022 | 1.024 | 0.9927 | 0.9916 | 0.03223 | 0.1007 |
| 0.01097 | 0.08124 | 0.06902 | 0.1151 | 0.112 | -9.41e-04 | -7.55e-04 | 0.004338 | 0.005679 | 0.9967 | 1.002 | 0.008335 | 0.007442 | 0.07979 | 0.0707 | 0.01077 | 0.01006 | 1.013 | 1.017 | 0.9804 | 0.9875 | 0.1009 | 0.09043 |
| 0.02091 | 0.08497 | 0.03785 | 0.1437 | 0.1198 | -3.44e-04 | -0.006408 | 0.01074 | 0.006679 | 0.9927 | 1.005 | 0.007472 | 0.006221 | 0.08458 | 0.03969 | 0.01274 | 0.0204 | 1.007 | 1.018 | 0.9781 | 0.9932 | 0.1096 | 0.07967 |
| 0.03938 | -0.04268 | -0.05323 | 0.08637 | 0.1011 | -0.01208 | -0.01144 | 0.01224 | 0.01478 | 1.002 | 1.002 | 0.009609 | 0.008076 | 0.03971 | 0.007024 | 0.03701 | 0.0453 | 1.021 | 1.017 | 0.9836 | 0.9858 | 0.1123 | 0.09581 |
| 0.3391 | -0.3025 | -0.09889 | 0.8001 | 1.23 | -0.056 | -0.1224 | 0.1551 | 0.1224 | 1.008 | 1.008 | 0.006503 | 0.005519 | -0.2156 | -0.02252 | 0.2913 | 0.1641 | 1.021 | 1.019 | 0.9956 | 0.9969 | 0.3554 | 0.2991 |
| 0.0414 | 0.08475 | -0.00662 | 0.2318 | 0.1557 | -0.009065 | -0.01193 | 0.01462 | 0.01582 | 1.001 | 1.004 | 0.00557 | 0.006322 | 0.1197 | 0.04508 | 0.03618 | 0.0519 | 1.012 | 1.016 | 0.9903 | 0.9912 | 0.1906 | 0.1468 |
| 0.03998 | | -0.01684 | | 0.1399 | | -0.008052 | | 0.0227 | 0.993 | 0.9983 | 0.007256 | 0.006794 | 0.04542 | 0.0156 | 0.04155 | 0.03966 | 1.007 | 1.012 | 0.9788 | 0.985 | 0.1269 | 0.09332 |
| 0.003355 | -0.00436 | -1.27e-04 | 0.006094 | 0.01302 | -0.003818 | -0.001497 | 0.004471 | 0.009022 | 1.012 | 1.008 | 0.007741 | 0.006988 | 0.06613 | 0.08866 | 0.01608 | 0.01207 | 1.027 | 1.022 | 0.9964 | 0.9943 | 0.09764 | 0.1123 |
| 0.03988 | -0.002049 | -0.054 | 0.139 | 0.1023 | -0.01088 | -0.01211 | 0.01517 | 0.01642 | 0.9998 | 1.016 | 0.009061 | 0.00761 | 0.04503 | -0.06242 | 0.04993 | 0.04584 | 1.018 | 1.031 | 0.982 | 1.001 | 0.1429 | 0.02742 |
| 0.003349 | -8.33e-05 | -0.001073 | 0.01194 | 0.01206 | -5.52e-04 | -0.002693 | 0.008902 | 0.007552 | 0.9962 | 1.017 | 0.008507 | 0.006639 | 0.07277 | 0.04135 | 0.01836 | 0.01754 | 1.013 | 1.03 | 0.9795 | 1.004 | 0.1088 | 0.07573 |
| 0.003069 | | -0.001454 | | 0.01058 | | -9.28e-04 | | 0.008163 | 1.009 | 0.9929 | 0.007049 | 0.008902 | 0.05488 | 0.07145 | 0.01424 | 0.01619 | 1.023 | 1.01 | 0.9957 | 0.9754 | 0.0828 | 0.1032 |
| 0.1739 | -0.04258 | -0.2655 | 0.2438 | 0.4162 | -0.002741 | -0.05524 | 0.05578 | 0.0677 | 1.011 | 0.999 | 0.005061 | 0.006953 | -0.06838 | 0.1598 | 0.06815 | 0.1537 | 1.021 | 1.013 | 1.001 | 0.9854 | 0.0652 | 0.461 |
| | -0.4706 | | 0.516 | | -0.05896 | | 0.1023 | | 1.008 | 0.9962 | 0.005231 | 0.005888 | -0.3927 | -0.08071 | 0.1937 | 0.2611 | 1.019 | 1.008 | 0.998 | 0.9846 | -0.01305 | 0.4311 |
| gcta.SE_h2_male | gcta.h2.conf.low_female | gcta.h2.conf.low_male | gcta.h2.conf.high_female | gcta.h2.conf.high_male | gcta.x.h2.conf.low_female | gcta.x.h2.conf.low_male | gcta.x.h2.conf.high_female | gcta.x.h2.conf.high_male | ldsc.int_female | ldsc.int_male | ldsc.int_se_female | ldsc.int_se_male | ldsc.h2_female | ldsc.h2_male | ldsc.h2_se_female | ldsc.h2_se_male | ldsc.int.conf.high_female | ldsc.int.conf.high_male | ldsc.int.conf.low_female | ldsc.int.conf.low_male | ldsc.h2.conf.high_female | ldsc.h2.conf.high_male |
| 0.04208 | -0.0244 | 0.03655 | 0.1108 | 0.2015 | -0.009163 | -0.003537 | 0.01358 | 0.03113 | 1.002 | 0.9993 | 0.0089 | 0.00675 | 0.03798 | 0.0825 | 0.0493 | 0.04602 | 1.02 | 1.013 | 0.9848 | 0.9861 | 0.1346 | 0.1727 |
| 0.04101 | -0.01127 | -0.001707 | 0.1274 | 0.159 | -0.0117 | -0.003309 | 0.01171 | 0.03126 | 1.008 | 0.9998 | 0.007004 | 0.006902 | 0.02547 | 0.04499 | 0.04505 | 0.03909 | 1.021 | 1.013 | 0.9938 | 0.9863 | 0.1138 | 0.1216 |
| 0.2127 | 0.03108 | -0.3952 | 0.7385 | 0.4386 | -0.0223 | -0.08063 | 0.1099 | 0.08063 | 1.003 | 0.9896 | 0.008156 | 0.006492 | 0.1255 | 0.1634 | 0.203 | 0.2112 | 1.019 | 1.002 | 0.9867 | 0.9769 | 0.5233 | 0.5774 |
| | -0.4163 | | 0.7802 | | -0.09912 | | 0.09912 | | 1.005 | 1.005 | 0.007449 | 0.01027 | -0.03706 | -0.7819 | 0.2645 | 0.5404 | 1.02 | 1.025 | 0.9904 | 0.9851 | 0.4813 | 0.2773 |
| | -0.1811 | | 1.022 | | -0.1109 | | 0.1109 | | 1.014 | 1.005 | 0.009024 | 0.005139 | -0.4132 | -0.7437 | 0.3077 | 0.2852 | 1.032 | 1.015 | 0.9966 | 0.9947 | 0.1899 | -0.1848 |
| 0.04894 | -0.04743 | 0.019 | 0.09751 | 0.2109 | -0.01258 | -0.01219 | 0.01258 | 0.02086 | 1.011 | 1.002 | 0.007367 | 0.006305 | -0.02154 | 0.07517 | 0.02817 | 0.0392 | 1.025 | 1.015 | 0.9965 | 0.9898 | 0.03367 | 0.152 |
| 0.03931 | -0.02347 | -0.07336 | 0.09753 | 0.08073 | -0.01266 | -0.01219 | 0.01266 | 0.01919 | 1.013 | 1.019 | 0.00852 | 0.005955 | -0.03388 | -0.08453 | 0.04244 | 0.0305 | 1.03 | 1.03 | 0.9968 | 1.007 | 0.0493 | -0.02474 |
| 0.2668 | -0.3982 | -0.3058 | 0.4212 | 0.7402 | -0.06601 | -0.08307 | 0.06602 | 0.08307 | 1.013 | 0.9961 | 0.0079 | 0.006289 | -0.1529 | 0.2827 | 0.2076 | 0.2373 | 1.028 | 1.008 | 0.9971 | 0.9838 | 0.2541 | 0.7478 |

| gcta.SE_h2_male | gcta.h2.conf.low_female | gcta.h2.conf.low_male | gcta.h2.conf.high_female | gcta.h2.conf.high_male | gcta.x.h2.conf.low_female | gcta.x.h2.conf.low_male | gcta.x.h2.conf.high_female | gcta.x.h2.conf.high_male | ldsc.int_female | ldsc.int_male | ldsc.int_se_female | ldsc.int_se_male | ldsc.h2_female | ldsc.h2_male | ldsc.h2_se_female | ldsc.h2_se_male | ldsc.int.conf.high_female | ldsc.int.conf.high_male | ldsc.int.conf.low_female | ldsc.int.conf.low_male | ldsc.h2.conf.high_female | ldsc.h2.conf.high_male |
|---|---|---|---|---|---|---|---|---|---|---|---|---|---|---|---|---|---|---|---|---|---|---|
| 0.3962 | -0.1964 | -0.1829 | 1.121 | 1.37 | -0.1057 | -0.03235 | 0.1057 | 0.2425 | 0.991 | 1.007 | 0.008204 | 0.008171 | 0.4197 | 0.003843 | 0.5002 | 0.4224 | 1.007 | 1.023 | 0.9749 | 0.991 | 1.4 | 0.8318 |
| | -0.3006 | | 0.313 | | -0.04773 | | 0.06017 | | 0.9866 | 1.001 | 0.005128 | 0.007171 | 0.2407 | -0.2483 | 0.1389 | 0.1792 | 0.9967 | 1.015 | 0.9766 | 0.9868 | 0.5131 | 0.1029 |
| 0.04344 | -0.002469 | -0.08438 | 0.135 | 0.08589 | -0.006277 | -0.01573 | 0.01985 | 0.01573 | 1.01 | 1 | 0.008167 | 0.007177 | -0.01517 | 1.53e-04 | 0.03557 | 0.04245 | 1.026 | 1.014 | 0.994 | 0.9861 | 0.05454 | 0.08336 |
| 0.2146 | | -0.07572 | | 0.7655 | | -0.06981 | | 0.06982 | 0.9872 | 1.004 | 0.007124 | 0.007278 | 0.8611 | 0.1487 | 0.1884 | 0.2274 | 1.001 | 1.018 | 0.9732 | 0.9896 | 1.23 | 0.5944 |
| 0.4734 | -0.4098 | -0.261 | 1.278 | 1.595 | -0.03631 | -0.05879 | 0.2985 | 0.3183 | 1.012 | 1 | 0.007418 | 0.007285 | -0.2463 | 0.07704 | 0.5369 | 0.4851 | 1.027 | 1.014 | 0.9979 | 0.9859 | 0.8059 | 1.028 |
| | -0.03796 | | 0.08628 | | -0.01309 | | 0.01309 | | 1.015 | 1.017 | 0.007374 | 0.005641 | -0.04273 | -0.1015 | 0.02354 | 0.03015 | 1.029 | 1.028 | 1 | 1.006 | 0.003415 | -0.04243 |
| 0.04414 | 0.01099 | -0.06711 | 0.1507 | 0.1059 | -0.006878 | -0.0158 | 0.01923 | 0.01581 | 1.007 | 1.002 | 0.007054 | 0.00844 | -0.002532 | 0.002421 | 0.03031 | 0.05487 | 1.02 | 1.018 | 0.9928 | 0.9852 | 0.05688 | 0.11 |
| 0.208 | | -0.2793 | | 0.536 | | -0.03611 | | 0.1183 | 0.9971 | 1.019 | 0.005896 | 0.00639 | -0.157 | -0.3861 | 0.1254 | 0.1513 | 1.009 | 1.032 | 0.9855 | 1.007 | 0.08885 | -0.08956 |
| 0.02052 | -0.007611 | -0.02925 | 0.04979 | 0.05117 | -0.002565 | -0.00712 | 0.007867 | 0.007122 | 1 | 0.9945 | 0.008078 | 0.006809 | 0.01478 | 0.03234 | 0.01643 | 0.01764 | 1.016 | 1.008 | 0.9846 | 0.9812 | 0.04699 | 0.06691 |
| 0.4195 | -0.9653 | -0.3268 | 1.081 | 1.318 | -0.1061 | -0.001657 | 0.2259 | 0.3027 | 1.001 | 0.991 | 0.007201 | 0.005546 | 0.1778 | 0.4885 | 0.5288 | 0.3707 | 1.015 | 1.002 | 0.9868 | 0.9801 | 1.214 | 1.215 |
| 0.269 | | -0.2284 | | 0.8262 | | -0.09737 | | 0.09738 | 0.9942 | 0.9938 | 0.007056 | 0.007782 | 0.1054 | 0.2202 | 0.196 | 0.2816 | 1.008 | 1.009 | 0.9804 | 0.9785 | 0.4896 | 0.7722 |
| | -0.3226 | | 0.5814 | | -0.05445 | | 0.1239 | | 0.9966 | 1.003 | 0.004967 | 0.005945 | 0.1126 | -0.177 | 0.2211 | 0.3127 | 1.006 | 1.015 | 0.9869 | 0.9916 | 0.5458 | 0.4359 |
| 0.2551 | -0.385 | -0.3365 | 0.5618 | 0.6634 | -0.08099 | -0.05896 | 0.08099 | 0.1292 | 1.009 | 1.003 | 0.006437 | 0.008387 | 0.07911 | -0.05813 | 0.2473 | 0.3554 | 1.022 | 1.019 | 0.9965 | 0.9865 | 0.5639 | 0.6385 |
| | -0.4909 | | 0.5693 | | -0.0551 | | 0.1382 | | 1.014 | 0.9982 | 0.006543 | 0.007655 | -0.5996 | -0.134 | 0.2813 | 0.2483 | 1.026 | 1.013 | 1.001 | 0.9832 | -0.04822 | 0.3528 |
| 0.4157 | -0.9612 | -0.3403 | 1.175 | 1.289 | -0.1221 | -0.1686 | 0.3467 | 0.1686 | 0.9925 | 1.001 | 0.005804 | 0.008509 | 0.0425 | 0.518 | 0.4241 | 0.5547 | 1.004 | 1.017 | 0.9811 | 0.9838 | 0.8737 | 1.605 |
| 0.03678 | 0.02885 | -0.007557 | 0.1379 | 0.1366 | -0.004407 | -0.01432 | 0.01908 | 0.01432 | 0.9786 | 0.9866 | 0.00726 | 0.009384 | 0.1282 | 0.1222 | 0.03193 | 0.05441 | 0.9928 | 1.005 | 0.9643 | 0.9682 | 0.1908 | 0.2289 |
| 0.02404 | 0.3432 | 0.2836 | 0.408 | 0.3779 | 4.20e-04 | 0.001959 | 0.01158 | 0.01961 | 1.002 | 1.031 | 0.0073 | 0.01105 | 0.2664 | 0.1277 | 0.01765 | 0.04016 | 1.016 | 1.053 | 0.9877 | 1.01 | 0.3009 | 0.2064 |
| 0.02404 | 0.3433 | 0.2838 | 0.4081 | 0.378 | 4.38e-04 | 0.001983 | 0.0116 | 0.01965 | 1.002 | 1.031 | 0.007271 | 0.01109 | 0.2668 | 0.1278 | 0.01763 | 0.04023 | 1.016 | 1.053 | 0.9875 | 1.009 | 0.3013 | 0.2067 |
| 0.02404 | 0.3433 | 0.2838 | 0.4081 | 0.378 | 4.38e-04 | 0.001983 | 0.0116 | 0.01965 | 1.002 | 1.031 | 0.007271 | 0.01109 | 0.2668 | 0.1278 | 0.01763 | 0.04023 | 1.016 | 1.053 | 0.9875 | 1.009 | 0.3013 | 0.2067 |
| 0.05764 | -0.06931 | 0.01045 | 0.105 | 0.2364 | -0.01731 | -0.005794 | 0.01732 | 0.0393 | 0.9981 | 0.9953 | 0.009098 | 0.005851 | 0.009254 | 0.1371 | 0.04898 | 0.05247 | 1.016 | 1.007 | 0.9803 | 0.9838 | 0.1053 | 0.24 |
| | 0.01282 | | 0.2077 | | -0.01926 | | 0.01927 | | 0.9906 | | 0.008262 | | 0.1454 | | 0.06187 | | 1.007 | | 0.9744 | | 0.2666 | |
| 0.2094 | -0.05522 | -0.3028 | 0.2953 | 0.5182 | -0.0317 | -0.03771 | 0.0317 | 0.1034 | 1.001 | 1.003 | 0.008068 | 0.007739 | 0.009383 | -0.07199 | 0.1038 | 0.2088 | 1.017 | 1.018 | 0.9853 | 0.9882 | 0.2128 | 0.3372 |
| 0.003509 | | 0.003766 | | 0.01752 | | 9.92e-05 | | 0.009645 | 1.003 | 0.9919 | 0.008269 | 0.006446 | 0.05721 | 0.06373 | 0.01136 | 0.01172 | 1.019 | 1.005 | 0.9866 | 0.9793 | 0.07947 | 0.08669 |
| 0.01081 | 0.08553 | 0.06278 | 0.1201 | 0.1052 | -7.65e-04 | -0.002226 | 0.004986 | 0.004493 | 1.026 | 0.9898 | 0.00917 | 0.008639 | 0.0525 | 0.07048 | 0.008474 | 0.01078 | 1.044 | 1.007 | 1.008 | 0.9729 | 0.06911 | 0.0916 |
| 0.1173 | -0.1299 | -0.1924 | 0.216 | 0.2675 | -0.02709 | -0.0422 | 0.02709 | 0.04221 | 1.004 | 1.003 | 0.006594 | 0.007788 | 0.02578 | -0.0181 | 0.1231 | 0.1315 | 1.016 | 1.018 | 0.9906 | 0.9876 | 0.267 | 0.2396 |
| 0.1163 | -0.1294 | -0.1634 | 0.1802 | 0.2925 | -0.009626 | -0.01997 | 0.0622 | 0.06836 | 0.9985 | 1.008 | 0.004741 | 0.007594 | 0.06651 | -0.005229 | 0.06007 | 0.1522 | 1.008 | 1.023 | 0.9892 | 0.993 | 0.1842 | 0.2931 |
| 0.01113 | 0.1167 | 0.09881 | 0.1521 | 0.1424 | -0.002647 | -0.002119 | 0.002649 | 0.005269 | 1.021 | 1 | 0.007657 | 0.007433 | 0.06523 | 0.0877 | 0.01064 | 0.01531 | 1.036 | 1.015 | 1.006 | 0.9856 | 0.08609 | 0.1177 |
| 0.01198 | 0.1031 | 0.07198 | 0.1417 | 0.1189 | -8.34e-04 | -0.002978 | 0.005766 | 0.003445 | 1.014 | 0.9951 | 0.00753 | 0.007264 | 0.07171 | 0.077 | 0.009674 | 0.01462 | 1.028 | 1.009 | 0.9989 | 0.9809 | 0.09067 | 0.1057 |
| 0.641 | | -1.162 | | 1.351 | | -0.1316 | | 0.3381 | 1.01 | 1.008 | 0.007915 | 0.006718 | -0.3695 | -1.064 | 0.3525 | 0.4393 | 1.026 | 1.021 | 0.9949 | 0.995 | 0.3215 | -0.2033 |

| gcta.SE_h2_male | gcta.h2.conf.low_female | gcta.h2.conf.low_male | gcta.h2.conf.high_female | gcta.h2.conf.high_male | gcta.x.h2.conf.low_female | gcta.x.h2.conf.low_male | gcta.x.h2.conf.high_female | gcta.x.h2.conf.high_male | ldsc.int_female | ldsc.int_male | ldsc.int_se_female | ldsc.int_se_male | ldsc.h2_female | ldsc.h2_male | ldsc.h2_se_female | ldsc.h2_se_male | ldsc.int.conf.high_female | ldsc.int.conf.high_male | ldsc.int.conf.low_female | ldsc.int.conf.low_male | ldsc.h2.conf.high_female | ldsc.h2.conf.high_male |
|---|---|---|---|---|---|---|---|---|---|---|---|---|---|---|---|---|---|---|---|---|---|---|
| 0.0237 | -0.01896 | 0.03098 | 0.04014 | 0.1239 | -0.00526 | -0.003562 | 0.005262 | 0.01178 | 0.9918 | 1.008 | 0.009918 | 0.009473 | 0.03206 | 0.03347 | 0.02167 | 0.02385 | 1.011 | 1.027 | 0.9724 | 0.9898 | 0.07453 | 0.08021 |
| 0.01129 | 0.002631 | 0.007727 | 0.03557 | 0.05198 | -0.002252 | -0.001458 | 0.003326 | 0.005591 | 1.019 | 1.003 | 0.00671 | 0.008542 | -0.002883 | 0.006047 | 0.007252 | 0.00988 | 1.032 | 1.02 | 1.006 | 0.9866 | 0.01133 | 0.02541 |
| 0.07004 | -0.03931 | -0.1053 | 0.2137 | 0.1692 | -0.02382 | -0.01332 | 0.02382 | 0.04145 | 1.002 | 0.9873 | 0.008302 | 0.005452 | 0.07094 | 0.09901 | 0.06951 | 0.0583 | 1.018 | 0.998 | 0.9857 | 0.9767 | 0.2072 | 0.2133 |
| 0.1038 | -0.1843 | -0.009696 | 0.1936 | 0.3973 | -0.02258 | -0.03735 | 0.03138 | 0.03735 | 0.9924 | 1.006 | 0.005592 | 0.005942 | 0.07049 | 0.01888 | 0.08359 | 0.1185 | 1.003 | 1.017 | 0.9815 | 0.9942 | 0.2343 | 0.2511 |
| 0.0703 | -0.02155 | -0.1134 | 0.2361 | 0.1622 | -0.02146 | -0.02448 | 0.02146 | 0.0263 | 0.9996 | 0.992 | 0.007279 | 0.007321 | 0.1002 | 0.07884 | 0.07282 | 0.07919 | 1.014 | 1.006 | 0.9854 | 0.9776 | 0.243 | 0.2341 |
|  | -0.4241 |  | 0.4442 |  | -0.09213 |  | 0.09213 |  | 1.002 | 1.015 | 0.007969 | 0.006339 | -0.1818 | -1.475 | 0.2372 | 0.3768 | 1.018 | 1.028 | 0.9867 | 1.003 | 0.2832 | -0.7365 |
| 0.04066 | 0.001924 | -0.06307 | 0.1423 | 0.09633 | -0.008782 | -0.01378 | 0.01648 | 0.01378 | 0.9845 | 1.02 | 0.01013 | 0.008133 | 0.08872 | -0.0738 | 0.04433 | 0.0402 | 1.004 | 1.036 | 0.9646 | 1.004 | 0.1756 | 0.004994 |
| 0.2896 | -0.1006 | -0.511 | 0.8226 | 0.6242 | -0.08222 | -0.04872 | 0.08223 | 0.159 | 0.9991 | 1.004 | 0.006453 | 0.006658 | 0.1861 | -0.2897 | 0.2048 | 0.2487 | 1.012 | 1.017 | 0.9864 | 0.9911 | 0.5876 | 0.1978 |
| 0.2964 | -0.2475 | -0.2437 | 0.681 | 0.9182 | -0.0785 | -0.0963 | 0.0785 | 0.0963 | 0.9881 | 1.004 | 0.007119 | 0.008819 | 0.3442 | 0.07659 | 0.2583 | 0.2832 | 1.002 | 1.021 | 0.9742 | 0.9866 | 0.8505 | 0.6316 |
| 0.2159 | -0.0248 | -0.318 | 0.8507 | 0.5284 | -0.05542 | -0.05696 | 0.09254 | 0.09083 | 1.02 | 1.003 | 0.008649 | 0.007008 | -0.4304 | -0.01022 | 0.2288 | 0.1661 | 1.037 | 1.017 | 1.003 | 0.9897 | 0.018 | 0.3154 |
| 0.216 | -0.02952 | -0.1695 | 0.8489 | 0.6773 | -0.06873 | -0.05439 | 0.06873 | 0.09311 | 1.016 | 1.007 | 0.005934 | 0.006772 | -0.3619 | -0.07439 | 0.2027 | 0.1832 | 1.027 | 1.021 | 1.004 | 0.9942 | 0.03534 | 0.2846 |
| 0.2942 | 0.05155 | -0.5268 | 0.9773 | 0.6263 | -0.07471 | -0.05608 | 0.07471 | 0.1543 | 0.9929 | 1.009 | 0.00591 | 0.007359 | 0.3489 | -0.4439 | 0.1971 | 0.3016 | 1.004 | 1.023 | 0.9813 | 0.9943 | 0.7353 | 0.1473 |
| 0.2863 | -0.3068 | -0.494 | 0.6155 | 0.6284 | -0.07602 | -0.0414 | 0.07602 | 0.1782 | 1.007 | 1.001 | 0.005884 | 0.005695 | -0.1255 | -0.3811 | 0.2424 | 0.294 | 1.018 | 1.012 | 0.9954 | 0.9896 | 0.3497 | 0.1951 |
| 0.2748 |  | -0.3983 |  | 0.6789 |  | -0.05017 |  | 0.1871 | 1.001 | 0.9909 | 0.009218 | 0.006423 | -0.1949 | 0.716 | 0.2355 | 0.2284 | 1.019 | 1.003 | 0.9829 | 0.9783 | 0.2667 | 1.164 |
|  | -0.06147 |  | 0.8248 |  | -0.05617 |  | 0.1072 |  | 1.01 | 1.004 | 0.008116 | 0.007348 | 0.005706 | -0.1642 | 0.2215 | 0.2171 | 1.026 | 1.019 | 0.9945 | 0.99 | 0.4399 | 0.2613 |
| 0.2652 | -0.4794 | -0.5 | 0.5691 | 0.5397 | -0.06049 | -0.08445 | 0.1503 | 0.1221 | 0.9962 | 0.9895 | 0.007049 | 0.006567 | -0.1154 | 0.3619 | 0.2185 | 0.3657 | 1.01 | 1.002 | 0.9823 | 0.9766 | 0.3129 | 1.079 |
| 0.2869 | -0.2768 | -0.4884 | 0.6437 | 0.6361 | -0.07548 | -0.03731 | 0.07549 | 0.1922 | 1.012 | 0.9995 | 0.006721 | 0.005689 | -0.2592 | -0.3225 | 0.2446 | 0.3018 | 1.025 | 1.011 | 0.9988 | 0.9883 | 0.2202 | 0.2692 |
|  | -0.2995 |  | 0.3131 |  | -0.04666 |  | 0.06045 |  | 1.004 | 1.015 | 0.008674 | 0.005657 | -0.1528 | -0.6567 | 0.166 | 0.3199 | 1.021 | 1.027 | 0.9873 | 1.004 | 0.1726 | -0.02957 |
| 0.04033 | 0.02265 | -0.04055 | 0.1706 | 0.1176 | -0.005231 | -0.01211 | 0.02476 | 0.0167 | 1.01 | 1.007 | 0.006831 | 0.008233 | 0.04008 | 0.01018 | 0.03161 | 0.04092 | 1.024 | 1.023 | 0.9969 | 0.9905 | 0.102 | 0.09039 |
| 1.102 | -0.3068 | -1.739 | 1.076 | 2.582 | -0.1166 | -0.4384 | 0.1166 | 0.4384 | 0.995 | 0.9442 | 0.007114 | 0.007106 | 0.5654 | 2.463 | 0.3501 | 1.251 | 1.009 | 0.9582 | 0.9811 | 0.9303 | 1.252 | 4.915 |
| 0.03131 | 0.08506 | 0.06546 | 0.165 | 0.1882 | -1.14e-05 | -0.008699 | 0.01491 | 0.01395 | 1.01 | 1.002 | 0.005427 | 0.00795 | 0.06413 | 0.0779 | 0.02142 | 0.03603 | 1.021 | 1.018 | 0.9992 | 0.9865 | 0.1061 | 0.1485 |
| 0.0742 | 0.05041 | -0.04302 | 0.2625 | 0.2478 | -0.005903 | -0.02618 | 0.03648 | 0.02619 | 0.9888 | 0.9995 | 0.006068 | 0.005581 | 0.1987 | 0.08716 | 0.04578 | 0.05897 | 1.001 | 1.01 | 0.9769 | 0.9886 | 0.2884 | 0.2027 |
|  | -0.9814 |  | 1.204 |  | -0.1148 |  | 0.3321 |  | 0.9888 | 0.9859 | 0.007372 | 0.005856 | -0.04703 | 0.3036 | 0.468 | 0.4152 | 1.003 | 0.9974 | 0.9743 | 0.9744 | 0.8703 | 1.117 |

| gcta.SE_h2_male | gcta.h2.conf.low_female | gcta.h2.conf.low_male | gcta.h2.conf.high_female | gcta.h2.conf.high_male | gcta.x.h2.conf.low_female | gcta.x.h2.conf.low_male | gcta.x.h2.conf.high_female | gcta.x.h2.conf.high_male | ldsc.int_female | ldsc.int_male | ldsc.int_se_female | ldsc.int_se_male | ldsc.h2_female | ldsc.h2_male | ldsc.h2_se_female | ldsc.h2_se_male | ldsc.int.conf.high_female | ldsc.int.conf.high_male | ldsc.int.conf.low_female | ldsc.int.conf.low_male | ldsc.h2.conf.high_female | ldsc.h2.conf.high_male |
|---|---|---|---|---|---|---|---|---|---|---|---|---|---|---|---|---|---|---|---|---|---|---|
| 0.01624 | 0.06442 | 0.002112 | 0.1166 | 0.06578 | -0.003669 | -0.002596 | 0.004002 | 0.009754 | 1.005 | 1.002 | 0.007445 | 0.005906 | 0.05772 | 0.02597 | 0.01789 | 0.01692 | 1.02 | 1.014 | 0.9909 | 0.9909 | 0.09278 | 0.05914 |
| 0.01648 | 0.06069 | 2.84e-04 | 0.1132 | 0.0649 | -0.003846 | -0.002608 | 0.003848 | 0.009875 | 1.002 | 1 | 0.005825 | 0.007655 | 0.06619 | 0.03568 | 0.0144 | 0.01814 | 1.014 | 1.015 | 0.9907 | 0.985 | 0.09441 | 0.07123 |
|  | -0.2473 |  | 0.6396 |  | -0.07338 |  | 0.08002 |  | 1.002 | 0.9723 | 0.008992 | 0.007609 | -0.2221 | 1.847 | 0.2703 | 0.6538 | 1.019 | 0.9872 | 0.984 | 0.9574 | 0.3078 | 3.128 |
|  | -0.477 |  | 0.4784 |  | -0.07883 |  | 0.07883 |  | 1.01 | 1 | 0.008023 | 0.005013 | -0.4658 | -0.6307 | 0.2249 | 0.6656 | 1.025 | 1.01 | 0.9938 | 0.9904 | -0.02499 | 0.6738 |

| gcta.SE_h2_male | gcta.h2.conf.low_female | gcta.h2.conf.low_male | gcta.h2.conf.high_female | gcta.h2.conf.high_male | gcta.x.h2.conf.low_female | gcta.x.h2.conf.low_male | gcta.x.h2.conf.high_female | gcta.x.h2.conf.high_male | ldsc.int_female | ldsc.int_male | ldsc.int_se_female | ldsc.int_se_male | ldsc.h2_female | ldsc.h2_male | ldsc.h2_se_female | ldsc.h2_se_male | ldsc.int.conf.high_female | ldsc.int.conf.high_male | ldsc.int.conf.low_female | ldsc.int.conf.low_male | ldsc.h2.conf.high_female | ldsc.h2.conf.high_male |
|---|---|---|---|---|---|---|---|---|---|---|---|---|---|---|---|---|---|---|---|---|---|---|
| 0.002899 | -0.004208 | -0.00454 | 0.005768 | 0.006824 | -0.003886 | -0.003436 | 0.003888 | 0.005683 | 1.01 | 1.001 | 0.00832 | 0.00773 | 0.0142 | 0.03005 | 0.01373 | 0.01456 | 1.026 | 1.017 | 0.9935 | 0.9862 | 0.04111 | 0.05859 |
| 0.9449 | -0.3557 | -1.813 | 0.7929 | 1.891 | -0.08796 | -0.3487 | 0.08796 | 0.3487 | 1.009 | 1.003 | 0.006958 | 0.007097 | 0.04191 | -0.1495 | 0.2658 | 0.8723 | 1.023 | 1.017 | 0.9956 | 0.9895 | 0.5628 | 1.56 |
| | -0.4207 | | 0.4355 | | -0.07707 | | 0.09161 | | 1.013 | 0.9934 | 0.008265 | 0.008094 | -0.2457 | 0.6393 | 0.231 | 0.5057 | 1.029 | 1.009 | 0.9963 | 0.9776 | 0.207 | 1.63 |
| | | | | | | | | | 1.008 | 0.9982 | 0.007279 | 0.008165 | -0.5584 | -0.02647 | 0.3186 | 0.2904 | 1.022 | 1.014 | 0.9936 | 0.9822 | 0.06609 | 0.5428 |
| 0.07359 | | -0.08377 | | 0.2047 | | -0.02055 | | 0.03456 | 1.009 | 1.001 | 0.007166 | 0.008093 | -0.1986 | 0.04158 | 0.06662 | 0.08709 | 1.023 | 1.016 | 0.9949 | 0.9846 | -0.06802 | 0.2123 |
| 0.1716 | -0.1758 | -0.3269 | 0.4481 | 0.3459 | -0.03852 | -0.04347 | 0.06452 | 0.06155 | 0.9881 | 1.002 | 0.006301 | 0.009283 | 0.3174 | -0.166 | 0.1782 | 0.1853 | 1 | 1.02 | 0.9757 | 0.9837 | 0.6666 | 0.1973 |
| 0.07063 | -0.09032 | -0.1194 | 0.1858 | 0.1575 | -0.008631 | -0.02583 | 0.04501 | 0.02584 | 0.9961 | 0.9878 | 0.007547 | 0.005715 | 0.04162 | 0.07658 | 0.07601 | 0.05182 | 1.011 | 0.999 | 0.9814 | 0.9766 | 0.1906 | 0.1782 |
| 0.1421 | -0.3584 | -0.0176 | 0.4284 | 0.5394 | -0.02769 | -0.03012 | 0.1007 | 0.06928 | 1.005 | 1.012 | 0.006811 | 0.00704 | -0.1985 | -0.03623 | 0.1718 | 0.09748 | 1.019 | 1.026 | 0.9919 | 0.998 | 0.1383 | 0.1548 |
| 0.03504 | 0.001108 | 0.03918 | 0.1243 | 0.1765 | -0.01095 | -0.01228 | 0.01095 | 0.01228 | 0.9973 | 0.9923 | 0.007483 | 0.007531 | 0.05673 | 0.1387 | 0.03075 | 0.04097 | 1.012 | 1.007 | 0.9827 | 0.9775 | 0.117 | 0.219 |
| 0.5505 | -0.9303 | -0.9867 | 1.374 | 1.171 | -0.1857 | -0.1052 | 0.1857 | 0.3283 | 0.9863 | 1.008 | 0.006857 | 0.009556 | 0.924 | -1.062 | 0.6753 | 0.5861 | 0.9998 | 1.027 | 0.9729 | 0.9893 | 2.248 | 0.08726 |
| 0.0104 | 0.06637 | 0.03712 | 0.09979 | 0.0779 | -3.41e-04 | -0.001767 | 0.005181 | 0.004748 | 1.011 | 1.013 | 0.008932 | 0.007226 | 0.04387 | 0.02874 | 0.007511 | 0.01306 | 1.028 | 1.027 | 0.9932 | 0.9986 | 0.05859 | 0.05434 |
| 0.04168 | 0.05467 | -0.02211 | 0.2013 | 0.1413 | -0.01315 | -0.008861 | 0.01315 | 0.02297 | 1.007 | 0.9977 | 0.008392 | 0.007634 | 0.06543 | 0.05315 | 0.03869 | 0.03759 | 1.023 | 1.013 | 0.9904 | 0.9827 | 0.1413 | 0.1268 |
| 0.03945 | -0.01698 | -0.07207 | 0.121 | 0.08259 | -0.004138 | -0.008636 | 0.02494 | 0.01952 | 1.01 | 1.009 | 0.007134 | 0.007801 | -0.03136 | -0.03991 | 0.03394 | 0.0471 | 1.024 | 1.025 | 0.996 | 0.9939 | 0.03516 | 0.05241 |
| | -0.002436 | | 0.00879 | | -0.001925 | | 0.00742 | | 1.021 | 1.006 | 0.0087 | 0.007393 | -0.004361 | 0.03234 | 0.01359 | 0.01178 | 1.038 | 1.021 | 1.004 | 0.9916 | 0.02227 | 0.05543 |
| | | | | | | | | | 1.013 | 1.011 | 0.007616 | 0.007582 | -0.4566 | -1.1 | 0.2155 | 0.5365 | 1.028 | 1.026 | 0.9984 | 0.9961 | -0.03435 | -0.04851 |
| 0.01261 | 0.1541 | 0.1307 | 0.1947 | 0.1801 | -0.002963 | -0.002944 | 0.002965 | 0.004331 | 1.013 | 1.002 | 0.005224 | 0.007267 | 0.1835 | 0.1705 | 0.08512 | 0.07739 | 1.023 | 1.017 | 1.003 | 0.988 | 0.3503 | 0.3222 |
| 0.1047 | -0.01403 | -0.06468 | 0.4509 | 0.3456 | -0.03813 | -0.03356 | 0.04281 | 0.03968 | 0.997 | 1.006 | 0.00778 | 0.007814 | 0.165 | -0.005999 | 0.1124 | 0.06895 | 1.012 | 1.021 | 0.9817 | 0.9903 | 0.3854 | 0.1291 |
| 0.0106 | 0.01004 | 0.01489 | 0.04243 | 0.05645 | -0.00281 | -0.003656 | 0.002812 | 0.003658 | 0.996 | 1.009 | 0.009519 | 0.007432 | 0.03082 | 0.02038 | 0.01126 | 0.0102 | 1.015 | 1.024 | 0.9773 | 0.9949 | 0.05289 | 0.04039 |
| 0.02062 | -0.008987 | -0.02988 | 0.0484 | 0.05096 | -0.003084 | -0.007618 | 0.007205 | 0.00762 | 1 | 0.9975 | 0.008591 | 0.007617 | 0.01377 | 0.02044 | 0.01705 | 0.02498 | 1.017 | 1.012 | 0.9833 | 0.9826 | 0.04719 | 0.0694 |
| 0.003268 | -0.001695 | -0.001338 | 0.0101 | 0.01147 | -0.00208 | -0.002535 | 0.007902 | 0.007994 | 1.007 | 1.002 | 0.005588 | 0.006734 | 0.05083 | 0.04454 | 0.01332 | 0.01268 | 1.018 | 1.015 | 0.9964 | 0.9888 | 0.07693 | 0.06939 |
| 0.1142 | -4.055 | -0.1189 | 4.118 | 0.3287 | -0.7201 | -0.0367 | 0.7883 | 0.0367 | 0.9812 | 1.01 | 0.005585 | 0.00533 | -0.1903 | -0.1299 | 1.284 | 0.09547 | 0.9922 | 1.021 | 0.9703 | 0.9996 | 2.325 | 0.05721 |
| 0.07092 | -0.4387 | -0.08308 | 0.4567 | 0.1949 | -0.07322 | -0.01813 | 0.0898 | 0.03288 | 1.008 | 0.993 | 0.006318 | 0.007606 | -0.123 | 0.1179 | 0.1631 | 0.07585 | 1.021 | 1.008 | 0.9958 | 0.9781 | 0.1967 | 0.2665 |
| | -0.7817 | | 1.451 | | -0.06313 | | 0.3349 | | 0.9896 | 1.011 | 0.008381 | 0.00739 | 1.417 | -0.5908 | 0.6189 | 0.5234 | 1.006 | 1.025 | 0.9731 | 0.9965 | 2.63 | 0.435 |
| 0.3779 | -0.2008 | -0.6292 | 0.6025 | 0.8524 | -0.05826 | -0.1353 | 0.07387 | 0.1353 | 1.006 | 1.003 | 0.006307 | 0.006008 | -0.03121 | 0.03505 | 0.1717 | 0.3539 | 1.019 | 1.015 | 0.994 | 0.9913 | 0.3052 | 0.7288 |
| 0.04386 | -0.1178 | -0.05396 | 0.1192 | 0.118 | -0.01893 | -0.01663 | 0.02033 | 0.01663 | 1 | 0.9818 | 0.009745 | 0.007394 | -0.07765 | 0.1617 | 0.08011 | 0.05537 | 1.02 | 0.9963 | 0.9814 | 0.9673 | 0.07937 | 0.2702 |
| | -0.6102 | | 0.6347 | | -0.1075 | | 0.1075 | | 1.007 | 0.9925 | 0.009258 | 0.007686 | -0.05995 | 1.026 | 0.4436 | 0.9437 | 1.026 | 1.008 | 0.9892 | 0.9774 | 0.8096 | 2.876 |
| | | | | | | | | | 1.002 | 1.006 | 0.006766 | 0.01066 | -0.008713 | -0.02144 | 0.02354 | 0.04552 | 1.015 | 1.027 | 0.9883 | 0.9855 | 0.03743 | 0.06779 |
| | -0.2532 | | 0.9689 | | -0.102 | | 0.102 | | 0.9882 | 0.9944 | 0.005293 | 0.005839 | 0.5978 | -2.312 | 0.2602 | 0.897 | 0.9986 | 1.006 | 0.9778 | 0.983 | 1.108 | -0.5536 |
| 0.2856 | 0.1727 | -0.5565 | 1.099 | 0.563 | -0.07826 | -0.09937 | 0.07826 | 0.09937 | 0.9967 | 1.015 | 0.008149 | 0.008349 | 0.4214 | -0.5614 | 0.2862 | 0.3351 | 1.013 | 1.032 | 0.9808 | 0.9988 | 0.9824 | 0.09545 |

| gcta.SE_h2_male | gcta.h2.conf.low_female | gcta.h2.conf.low_male | gcta.h2.conf.high_female | gcta.h2.conf.high_male | gcta.x.h2.conf.low_female | gcta.x.h2.conf.low_male | gcta.x.h2.conf.high_female | gcta.x.h2.conf.high_male | ldsc.int_female | ldsc.int_male | ldsc.int_se_female | ldsc.int_se_male | ldsc.h2_female | ldsc.h2_male | ldsc.h2_se_female | ldsc.h2_se_male | ldsc.int.conf.high_female | ldsc.int.conf.high_male | ldsc.int.conf.low_female | ldsc.int.conf.low_male | ldsc.h2.conf.high_female | ldsc.h2.conf.high_male |
|---|---|---|---|---|---|---|---|---|---|---|---|---|---|---|---|---|---|---|---|---|---|---|
| 0.2976 | -0.187 | -0.2267 | 0.7475 | 0.9401 | -0.07728 | -0.09434 | 0.07728 | 0.09923 | 0.9868 | 1.007 | 0.007316 | 0.009094 | 0.4376 | -0.004415 | 0.2592 | 0.289 | 1.001 | 1.025 | 0.9724 | 0.9893 | 0.9456 | 0.5621 |
| 0.262 | -0.2848 | -0.4103 | 0.7255 | 0.6167 | -0.05707 | -0.08726 | 0.1118 | 0.08726 | 1.028 | 1.012 | 0.00708 | 0.00757 | -0.7609 | -0.3073 | 0.2099 | 0.2513 | 1.042 | 1.027 | 1.014 | 0.9973 | -0.3495 | 0.1853 |
| 0.2651 | -0.2543 | -0.1936 | 0.7768 | 0.8455 | -0.06064 | -0.09109 | 0.1004 | 0.09109 | 1.026 | 1.01 | 0.006125 | 0.007735 | -0.7307 | -0.1654 | 0.19 | 0.2637 | 1.038 | 1.025 | 1.014 | 0.9947 | -0.3583 | 0.3515 |
| 0.2853 | 0.1443 | -0.3161 | 1.089 | 0.8024 | -0.07497 | -0.1028 | 0.07497 | 0.1028 | 0.9947 | 1.012 | 0.007888 | 0.008569 | 0.3852 | -0.3487 | 0.2768 | 0.3106 | 1.01 | 1.029 | 0.9792 | 0.9951 | 0.9278 | 0.2601 |
| 0.1155 | -0.1208 | -0.2251 | 0.1431 | 0.2276 | -0.01992 | -0.03746 | 0.02586 | 0.04001 | 0.9959 | 0.9991 | 0.009163 | 0.006425 | 0.04066 | 0.03824 | 0.07726 | 0.109 | 1.014 | 1.012 | 0.978 | 0.9865 | 0.1921 | 0.2519 |
| 0.01646 | 0.06065 | 0.03697 | 0.1133 | 0.1015 | -0.003997 | -0.005879 | 0.003999 | 0.005881 | 1.015 | 1.026 | 0.006516 | 0.007457 | 0.04064 | -0.01645 | 0.01103 | 0.02182 | 1.028 | 1.041 | 1.002 | 1.012 | 0.06225 | 0.02632 |
| 0.01406 | 0.0655 | 0.07071 | 0.1085 | 0.1258 | -0.002148 | -0.004672 | 0.003873 | 0.004674 | 1.005 | 1.018 | 0.008381 | 0.008067 | 0.05648 | 0.03108 | 0.01044 | 0.01702 | 1.021 | 1.034 | 0.9881 | 1.002 | 0.07695 | 0.06444 |
| 0.01885 | 0.1933 | 0.1244 | 0.2568 | 0.1983 | -0.003551 | -0.00262 | 0.006496 | 0.01005 | 1.016 | 0.9935 | 0.009833 | 0.008159 | 0.1335 | 0.133 | 0.02291 | 0.0357 | 1.036 | 1.01 | 0.9972 | 0.9775 | 0.1784 | 0.203 |
| 0.2359 | -0.3365 | -0.4509 | 0.5562 | 0.474 | -0.04461 | -0.07043 | 0.1243 | 0.07084 | 1.015 | 1.005 | 0.009054 | 0.005159 | -0.0967 | -0.1191 | 0.3264 | 0.1614 | 1.033 | 1.015 | 0.9976 | 0.9948 | 0.5429 | 0.1973 |
| 0.3171 | -0.6823 | -0.4577 | 0.8375 | 0.7853 | -0.1172 | -0.1169 | 0.1624 | 0.1169 | 0.9906 | 0.9957 | 0.005618 | 0.005057 | 0.7474 | 0.2463 | 0.4321 | 0.2673 | 1.002 | 1.006 | 0.9796 | 0.9858 | 1.594 | 0.7701 |
| 0.03659 | -0.07576 | -0.01972 | 0.07845 | 0.1237 | -0.0139 | -0.01087 | 0.01391 | 0.01295 | 1.004 | 0.9827 | 0.007384 | 0.00629 | -0.08704 | 0.1378 | 0.05392 | 0.02687 | 1.018 | 0.995 | 0.9895 | 0.9704 | 0.01864 | 0.1905 |
| | | | | | | | | | 0.9937 | 1.003 | 0.007775 | 0.006409 | -0.2253 | -0.145 | 0.3197 | 0.6962 | 1.009 | 1.016 | 0.9784 | 0.9908 | 0.4013 | 1.22 |
| 0.01757 | 0.08471 | 0.1044 | 0.1292 | 0.1733 | -0.003327 | -0.005648 | 0.003329 | 0.00565 | 1.017 | 1.012 | 0.007018 | 0.008891 | 0.05295 | 0.07562 | 0.01121 | 0.01981 | 1.03 | 1.029 | 1.003 | 0.9942 | 0.07492 | 0.1145 |
| 0.112 | | -0.05353 | | 0.3853 | | -0.01467 | | 0.07459 | 1.009 | 1.009 | 0.007179 | 0.007134 | -0.06726 | -0.03531 | 0.1014 | 0.1225 | 1.023 | 1.023 | 0.9954 | 0.9954 | 0.1314 | 0.2047 |
| | -0.05485 | | 1.095 | | -0.08258 | | 0.09096 | | 0.9982 | 0.9568 | 0.00759 | 0.008481 | 0.2334 | 0.9186 | 0.3552 | 0.963 | 1.013 | 0.9735 | 0.9834 | 0.9402 | 0.9297 | 2.806 |
| | -0.3437 | | 0.412 | | -0.04395 | | 0.06517 | | 0.9906 | 1.005 | 0.009196 | 0.005102 | 0.33 | -0.05191 | 0.2599 | 0.2289 | 1.009 | 1.015 | 0.9725 | 0.9946 | 0.8394 | 0.3968 |
| 0.2363 | -0.3398 | -0.3789 | 0.3416 | 0.5472 | -0.06197 | -0.01653 | 0.06197 | 0.1779 | 0.99 | 1.001 | 0.007004 | 0.008351 | 0.1611 | 0.04791 | 0.1577 | 0.2786 | 1.004 | 1.018 | 0.9763 | 0.985 | 0.4702 | 0.5941 |
| 0.3454 | -0.3672 | -0.5993 | 1.371 | 0.7545 | -0.04617 | -0.09523 | 0.2419 | 0.09523 | 0.9865 | 1.01 | 0.007574 | 0.008604 | 0.5846 | -0.315 | 0.4174 | 0.3207 | 1.001 | 1.027 | 0.9717 | 0.9929 | 1.403 | 0.3135 |
| 0.4922 | -0.8076 | -0.867 | 1.018 | 1.063 | -0.1282 | -0.07408 | 0.1282 | 0.2878 | 0.9874 | 1.01 | 0.008456 | 0.006968 | 0.8791 | -0.3999 | 0.632 | 0.3501 | 1.004 | 1.024 | 0.9709 | 0.9966 | 2.118 | 0.2864 |
| 0.09138 | -0.1892 | -0.1778 | 0.2303 | 0.1804 | -0.02369 | -0.02955 | 0.06198 | 0.03199 | 0.9991 | 0.9937 | 0.009908 | 0.006545 | 0.07727 | 0.07431 | 0.1502 | 0.0895 | 1.018 | 1.006 | 0.9796 | 0.9808 | 0.3717 | 0.2497 |
| 0.402 | -0.7476 | -0.3654 | 1.045 | 1.211 | -0.1623 | -0.1365 | 0.1623 | 0.1365 | 1.001 | 0.9931 | 0.006866 | 0.005551 | -0.4162 | 0.7886 | 0.4938 | 0.348 | 1.015 | 1.004 | 0.988 | 0.9822 | 0.5516 | 1.471 |
| 0.08077 | -0.1267 | -0.1471 | 0.132 | 0.1695 | -0.02729 | -0.01672 | 0.0273 | 0.03722 | 0.9982 | 1.009 | 0.008078 | 0.006964 | -0.01718 | -0.1502 | 0.06148 | 0.0801 | 1.014 | 1.022 | 0.9824 | 0.9951 | 0.1033 | 0.00679 |
| 0.1077 | -0.07928 | -0.1989 | 0.1812 | 0.2234 | -0.01917 | -0.02757 | 0.03275 | 0.05095 | 0.9906 | 1 | 0.006393 | 0.008255 | 0.08998 | -0.09125 | 0.06004 | 0.1199 | 1.003 | 1.016 | 0.9781 | 0.9839 | 0.2077 | 0.1437 |
| 0.0144 | 0.03938 | 0.02254 | 0.08489 | 0.07901 | -0.001336 | -0.003025 | 0.006993 | 0.006597 | 0.9998 | 0.9884 | 0.006042 | 0.007617 | 0.03831 | 0.05558 | 0.009888 | 0.01519 | 1.012 | 1.003 | 0.988 | 0.9734 | 0.05769 | 0.08535 |
| 0.02341 | 0.005542 | 0.001936 | 0.07394 | 0.09371 | -0.006569 | -0.005024 | 0.006571 | 0.01261 | 1.004 | 0.995 | 0.007934 | 0.006419 | 0.0137 | 0.03532 | 0.02145 | 0.02591 | 1.02 | 1.008 | 0.9885 | 0.9824 | 0.05574 | 0.08611 |
| 0.03965 | 0.01234 | -0.009326 | 0.1636 | 0.1461 | -0.01454 | -0.01397 | 0.01454 | 0.01397 | 0.9949 | 1.009 | 0.007427 | 0.00706 | 0.1362 | -0.03276 | 0.07069 | 0.04054 | 1.009 | 1.023 | 0.9804 | 0.9952 | 0.2747 | 0.04669 |
| | | | | | | | | | 0.9993 | 1.016 | 0.004532 | 0.008776 | -0.05883 | -0.2192 | 0.04878 | 0.1312 | 1.008 | 1.034 | 0.9904 | 0.9992 | 0.03678 | 0.03798 |

| gcta.SE_h2_male | gcta.h2.conf.low_female | gcta.h2.conf.low_male | gcta.h2.conf.high_female | gcta.h2.conf.high_male | gcta.x.h2.conf.low_female | gcta.x.h2.conf.low_male | gcta.x.h2.conf.high_female | gcta.x.h2.conf.high_male | ldsc.int_female | ldsc.int_male | ldsc.int_se_female | ldsc.int_se_male | ldsc.h2_female | ldsc.h2_male | ldsc.h2_se_female | ldsc.h2_se_male | ldsc.int.conf.high_female | ldsc.int.conf.high_male | ldsc.int.conf.low_female | ldsc.int.conf.low_male | ldsc.h2.conf.high_female | ldsc.h2.conf.high_male |
|---|---|---|---|---|---|---|---|---|---|---|---|---|---|---|---|---|---|---|---|---|---|---|
| 0.02709 | -0.0327 | -0.02403 | 0.06674 | 0.08215 | -0.007033 | -0.008231 | 0.01116 | 0.01299 | 1.007 | 0.9879 | 0.007348 | 0.009365 | 0.007439 | 0.06237 | 0.02242 | 0.03816 | 1.022 | 1.006 | 0.993 | 0.9695 | 0.05138 | 0.1372 |
| 0.01469 | 0.007737 | 0.01736 | 0.05079 | 0.07494 | 0.001335 | -0.003244 | 0.009385 | 0.006633 | 1.015 | 1.02 | 0.006927 | 0.006747 | 0.001774 | -4.83e-04 | 0.01113 | 0.01126 | 1.028 | 1.033 | 1.001 | 1.006 | 0.02358 | 0.02158 |
| 0.06376 | -0.004098 | -0.122 | 0.06433 | 0.1279 | -0.005669 | -0.01865 | 0.005671 | 0.02452 | 1.004 | 0.9982 | 0.008658 | 0.009444 | 0.02603 | 0.03111 | 0.02192 | 0.06456 | 1.021 | 1.017 | 0.9866 | 0.9796 | 0.06899 | 0.1576 |
| | -0.4846 | | 0.6447 | | -0.01779 | | 0.1663 | | 0.9908 | 0.993 | 0.005996 | 0.007519 | 0.1317 | 0.07274 | 0.3836 | 0.2875 | 1.003 | 1.008 | 0.9791 | 0.9783 | 0.8835 | 0.6362 |
| 0.04806 | | -0.08072 | | 0.1077 | | -0.006675 | | 0.03246 | 0.9939 | 1.007 | 0.007297 | 0.007862 | 0.03599 | -0.07943 | 0.06028 | 0.06701 | 1.008 | 1.023 | 0.9796 | 0.9917 | 0.1541 | 0.05192 |
| 0.02322 | 0.01659 | -0.03535 | 0.07945 | 0.05567 | -0.005724 | -0.008384 | 0.005726 | 0.008386 | 1.006 | 0.9973 | 0.00773 | 0.007601 | 0.01247 | 0.01159 | 0.01739 | 0.03219 | 1.021 | 1.012 | 0.9904 | 0.9824 | 0.04656 | 0.07467 |
| 0.1921 | -0.3945 | -0.3467 | 0.4177 | 0.4062 | -0.07448 | -0.07236 | 0.07448 | 0.07236 | 0.9919 | 0.9799 | 0.006218 | 0.006293 | 0.2718 | 0.6178 | 0.1741 | 0.1992 | 1.004 | 0.9922 | 0.9797 | 0.9675 | 0.613 | 1.008 |
| 0.196 | 0.01429 | 0.01671 | 0.8464 | 0.7849 | -0.05919 | -0.04324 | 0.07644 | 0.09011 | 1.018 | 0.9966 | 0.007214 | 0.006771 | -0.2314 | 0.3825 | 0.1777 | 0.1657 | 1.032 | 1.01 | 1.004 | 0.9833 | 0.117 | 0.7073 |
| 0.1937 | 0.1721 | -0.07319 | 0.996 | 0.6863 | -0.05172 | -0.04736 | 0.07656 | 0.07419 | 1.018 | 0.9969 | 0.00708 | 0.006317 | -0.07484 | 0.4027 | 0.1971 | 0.1232 | 1.032 | 1.009 | 1.004 | 0.9845 | 0.3115 | 0.6441 |
| 0.1974 | 8.35e-04 | 0.1124 | 0.8357 | 0.8864 | -0.0619 | -0.03454 | 0.07865 | 0.1123 | 1.02 | 1.001 | 0.007045 | 0.006963 | -0.3222 | 0.308 | 0.1682 | 0.1904 | 1.034 | 1.014 | 1.007 | 0.9871 | 0.007452 | 0.6812 |
| 0.1964 | | -0.3314 | | 0.4385 | | -0.06865 | | 0.06865 | 0.9912 | 0.9891 | 0.007378 | 0.005887 | 0.1141 | 0.465 | 0.2266 | 0.2036 | 1.006 | 1.001 | 0.9768 | 0.9775 | 0.5582 | 0.864 |
| | -0.4318 | | 0.459 | | -0.07119 | | 0.0984 | | 1.001 | 0.9745 | 0.006844 | 0.006028 | -0.1061 | 0.8224 | 0.2204 | 0.2662 | 1.014 | 0.9863 | 0.9876 | 0.9626 | 0.3259 | 1.344 |
| 0.2175 | 1.83e-04 | -0.1306 | 0.8739 | 0.722 | -0.05779 | -0.06686 | 0.07843 | 0.07837 | 1.024 | 1.003 | 0.007449 | 0.006445 | -0.4919 | 0.06888 | 0.2036 | 0.2073 | 1.038 | 1.016 | 1.009 | 0.9908 | -0.09284 | 0.4753 |
| 0.217 | 0.002071 | -0.1487 | 0.8734 | 0.7018 | -0.05764 | -0.06361 | 0.07683 | 0.08302 | 1.024 | 1.004 | 0.007622 | 0.005602 | -0.5199 | 0.006874 | 0.2111 | 0.1833 | 1.039 | 1.015 | 1.009 | 0.9926 | -0.1061 | 0.3661 |
| 0.2231 | -0.2906 | -0.4079 | 0.5853 | 0.4665 | -0.04254 | -0.07936 | 0.1261 | 0.07936 | 1.004 | 0.985 | 0.008006 | 0.005927 | -0.1524 | 0.6048 | 0.2704 | 0.2327 | 1.02 | 0.9966 | 0.9886 | 0.9734 | 0.3776 | 1.061 |
| 0.2923 | 0.02178 | -0.5634 | 0.4396 | 0.5824 | -0.02222 | -0.09149 | 0.04733 | 0.1109 | 0.9964 | 1.007 | 0.005532 | 0.00539 | 0.1587 | -0.3529 | 0.1072 | 0.2204 | 1.007 | 1.017 | 0.9856 | 0.9961 | 0.3689 | 0.07906 |
| 0.02815 | 0.0254 | -0.03608 | 0.1006 | 0.07426 | -0.006112 | -0.005809 | 0.006114 | 0.01512 | 0.9995 | 1 | 0.005959 | 0.009339 | 0.0548 | 0.01054 | 0.02108 | 0.02439 | 1.011 | 1.018 | 0.9878 | 0.9819 | 0.09611 | 0.05835 |
| 0.3664 | -0.3013 | -0.705 | 0.3602 | 0.7314 | -0.06562 | -0.1174 | 0.06956 | 0.1494 | 1.004 | 1.004 | 0.006555 | 0.005568 | -0.07722 | -0.337 | 0.1686 | 0.2461 | 1.017 | 1.015 | 0.9912 | 0.9931 | 0.2532 | 0.1453 |
| 0.2924 | -0.06357 | -0.3497 | 0.4908 | 0.7965 | -0.04351 | -0.05141 | 0.04351 | 0.1863 | 0.9894 | 1.007 | 0.006257 | 0.006548 | 0.3152 | 0.1935 | 0.1572 | 0.3424 | 1.002 | 1.02 | 0.9772 | 0.9939 | 0.6234 | 0.8646 |
| 0.2616 | -0.2217 | -0.2062 | 0.6097 | 0.8193 | -0.05413 | -0.1089 | 0.07907 | 0.1089 | 1.001 | 0.9958 | 0.006518 | 0.006232 | 0.08676 | 0.6038 | 0.183 | 0.3023 | 1.014 | 1.008 | 0.9886 | 0.9836 | 0.4455 | 1.196 |
| 0.01115 | 0.1028 | 0.09498 | 0.1375 | 0.1387 | -3.66e-04 | -0.003239 | 0.005235 | 0.003241 | 1.021 | 1.012 | 0.007139 | 0.00804 | 0.05974 | 0.07334 | 0.009984 | 0.009848 | 1.035 | 1.028 | 1.007 | 0.9962 | 0.07931 | 0.09264 |
| 0.04294 | -0.08255 | -0.0677 | 0.08442 | 0.1006 | -0.01338 | -0.007788 | 0.01531 | 0.02657 | 0.9977 | 1.003 | 0.009017 | 0.00619 | 0.02094 | -0.02933 | 0.0532 | 0.03533 | 1.015 | 1.015 | 0.98 | 0.9906 | 0.1252 | 0.03991 |
| 0.05376 | -0.04523 | -0.03162 | 0.1827 | 0.1791 | -0.01854 | -0.0191 | 0.01855 | 0.0191 | 1.001 | 0.9902 | 0.008003 | 0.005445 | 0.07852 | 0.1075 | 0.04815 | 0.05412 | 1.017 | 1.001 | 0.9856 | 0.9795 | 0.1729 | 0.2136 |
| | -0.2365 | | 0.5686 | | -0.07429 | | 0.0743 | | 0.9907 | 1 | 0.008219 | 0.004563 | 0.3668 | -0.11 | 0.2514 | 0.1235 | 1.007 | 1.009 | 0.9746 | 0.9914 | 0.8595 | 0.1322 |
| | -0.5019 | | 0.5369 | | -0.09944 | | 0.09944 | | 1.004 | 1.002 | 0.006783 | 0.007464 | 0.06871 | -0.03199 | 0.2165 | 0.2024 | 1.017 | 1.017 | 0.9909 | 0.9877 | 0.493 | 0.3647 |
| 0.02248 | 0.1435 | 0.1622 | 0.2045 | 0.2504 | -0.004732 | -0.003079 | 0.004734 | 0.01264 | 1.011 | 1.009 | 0.007162 | 0.008841 | 0.111 | 0.1363 | 0.01629 | 0.02601 | 1.025 | 1.026 | 0.9965 | 0.9913 | 0.143 | 0.1873 |
| 0.01185 | 0.1937 | 0.1652 | 0.2315 | 0.2116 | 3.23e-04 | -8.52e-05 | 0.00668 | 0.007739 | 1.008 | 1.006 | 0.007933 | 0.007858 | 0.1445 | 0.1238 | 0.01213 | 0.009747 | 1.023 | 1.022 | 0.9922 | 0.991 | 0.1683 | 0.1429 |
| | -0.04674 | | 0.9323 | | -0.05511 | | 0.1012 | | 0.9999 | 0.9936 | 0.005438 | 0.00722 | 0.2755 | -0.3284 | 0.2323 | 0.4204 | 1.011 | 1.008 | 0.9893 | 0.9794 | 0.7307 | 0.4957 |

| ldsc.h2.conf.low_female | ldsc.h2.conf.low_male | ldsc.int.flag_female | ldsc.int.flag_male | ldsc.N_female | ldsc.N_male | HDL.h2_female | HDL.h2_male | HDL.h2.se_female | HDL.h2.se_male | HDL.conf.high_female | HDL.conf.high_male | HDL.conf.low_female | HDL.conf.low_male |
|---|---|---|---|---|---|---|---|---|---|---|---|---|---|
| -0.357 | -0.548 | | | 925 | 761 | 1 | 1 | 0 | 0 | 1 | 1 | 1 | 1 |
| -0.04821 | -0.02447 | | | 8469 | 6545 | 0.179 | 0.2814 | 0.03322 | 0.03341 | 0.2441 | 0.3469 | 0.1139 | 0.2159 |
| -0.4126 | -1.02 | | flag | 2073 | 897 | 0.6245 | 1 | 0.0486 | 0 | 0.7197 | 1 | 0.5292 | 1 |
| -0.04914 | -0.08464 | | flag | 7048 | 2828 | 0.2927 | 0.605 | 0.02955 | 0.05904 | 0.3506 | 0.7207 | 0.2347 | 0.4893 |
| -0.145 | -0.6101 | | | 2276 | 811 | 0.6311 | 1 | 0.05925 | 0 | 0.7472 | 1 | 0.515 | 1 |
| -0.04 | -0.002373 | | | 7065 | 2839 | 0.2528 | 0.6006 | 0.03492 | 0.04079 | 0.3212 | 0.6806 | 0.1843 | 0.5207 |
| -0.2553 | -0.1475 | | | 3674 | 2884 | 0.3913 | 0.5014 | 0.04733 | 0.03087 | 0.4841 | 0.5619 | 0.2985 | 0.4409 |
| -0.03908 | -0.1428 | | | 3671 | 2892 | 0.4732 | 0.356 | 0.02871 | 0.02818 | 0.5295 | 0.4112 | 0.4169 | 0.3007 |
| -0.653 | -0.2591 | | | 925 | 962 | 1 | 1 | 0 | 0 | 1 | 1 | 1 | 1 |
| -0.3784 | -0.2112 | | | 1059 | 638 | 1 | 1 | 0 | 0 | 1 | 1 | 1 | 1 |
| -0.3748 | -1.717 | | | 996 | 598 | 1 | 1 | 0 | 0 | 1 | 1 | 1 | 1 |
| -0.6574 | -0.9317 | | | 1042 | 626 | 1 | 1 | 0 | 0 | 1 | 1 | 1 | 1 |
| -0.8907 | -1.362 | | | 1060 | 640 | 1 | 1 | 0 | 0 | 1 | 1 | 1 | 1 |
| -1.077 | -1.466 | flag | | 1059 | 639 | 1 | 1 | 0 | 0 | 1 | 1 | 1 | 1 |
| -0.6121 | -1.636 | | flag | 1060 | 638 | 1 | 1 | 0 | 0 | 1 | 1 | 1 | 1 |
| -0.4926 | -1.105 | | | 1508 | 548 | 1 | 1 | 0.05651 | 0 | 1.111 | 1 | 0.8892 | 1 |
| -0.5252 | -0.5315 | | | 1806 | 675 | 0.4653 | 1 | 0.1144 | 0 | 0.6894 | 1 | 0.2411 | 1 |
| -0.08846 | 0.05255 | | | 1805 | 675 | 0.742 | 1 | 0.07921 | 0 | 0.8973 | 1 | 0.5868 | 1 |
| -0.5454 | -1.155 | | | 987 | 598 | 1 | 1 | 0 | 0 | 1 | 1 | 1 | 1 |
| -0.4392 | | | | 1035 | | | | | | | | | |
| -0.4202 | -0.2604 | | | 1312 | 1721 | 1 | 0.9053 | 0 | 0.06187 | 1 | 1.027 | 1 | 0.784 |
| 0.03857 | 0.009752 | | flag | 31325 | 24351 | 0.0887 | 0.09682 | 0.006953 | 0.01007 | 0.1023 | 0.1166 | 0.07507 | 0.07708 |
| -0.1564 | -0.2881 | | | 3319 | 3332 | 0.3765 | 0.3206 | 0.04107 | 0.05008 | 0.457 | 0.4187 | 0.296 | 0.2224 |
| -0.404 | -0.1846 | | | 2156 | 2218 | 0.6452 | 0.6883 | 0.07391 | 0.06215 | 0.79 | 0.8101 | 0.5003 | 0.5665 |
| -0.0333 | -0.1122 | | | 4050 | 3191 | 0.3651 | 0.4803 | 0.0412 | 0.04691 | 0.4458 | 0.5722 | 0.2843 | 0.3884 |
| -0.4445 | -0.9829 | | | 864 | 573 | 1 | 1 | 0 | 0 | 1 | 1 | 1 | 1 |
| 0.06572 | 0.0401 | | | 21116 | 15939 | 0.144 | 0.1538 | 0.01291 | 0.0187 | 0.1693 | 0.1905 | 0.1187 | 0.1172 |
| -0.4204 | -0.1072 | | | 1426 | 1791 | 1 | 0.9107 | 0.02027 | 0.06528 | 1.04 | 1.039 | 0.9603 | 0.7827 |
| -0.02746 | -0.07405 | | | 4794 | 3776 | 0.411 | 0.4687 | 0.04275 | 0.05127 | 0.4948 | 0.5692 | 0.3272 | 0.3683 |
| -0.01299 | -0.008986 | | | 34129 | 26892 | 0.05189 | 0.0719 | 0.00845 | 0.008161 | 0.06845 | 0.08789 | 0.03532 | 0.0559 |
| 0.01256 | -0.2 | | | 3746 | 2556 | 0.5233 | 0.4493 | 0.06343 | 0.06432 | 0.6476 | 0.5754 | 0.3989 | 0.3233 |
| -0.1238 | -0.9231 | | | 619 | 471 | 1 | 1 | 0 | 0 | 1 | 1 | 1 | 1 |
| -0.0788 | -1.164 | flag | | 2079 | 642 | 0.8768 | 1 | 0.05766 | 0 | 0.9898 | 1 | 0.7637 | 1 |
| -0.5072 | -1.961 | | | 1045 | 285 | 0.9345 | 1 | 0.1198 | 0 | 1.169 | 1 | 0.6996 | 1 |
| -1.965 | 0.1401 | | | 625 | 409 | 1 | 1 | 0 | 0 | 1 | 1 | 1 | 1 |
| -1.082 | -0.63 | | flag | 1015 | 231 | 1 | 1 | 0.003478 | 0 | 1.007 | 1 | 0.9932 | 1 |
| -0.06098 | -0.0987 | | | 7227 | 6314 | 0.1925 | 0.2075 | 0.0297 | 0.02688 | 0.2507 | 0.2602 | 0.1342 | 0.1548 |
| -0.1641 | -0.08424 | | | 3888 | 4046 | 0.3194 | 0.3718 | 0.03456 | 0.04406 | 0.3872 | 0.4582 | 0.2517 | 0.2855 |
| -0.1953 | -0.5311 | | | 1354 | 933 | 1 | 1 | 0 | 0 | 1 | 1 | 1 | 1 |
| 0.007844 | -0.00118 | | | 14842 | 10793 | 0.1251 | 0.1891 | 0.01784 | 0.02669 | 0.1601 | 0.2414 | 0.09016 | 0.1368 |
| 0.02478 | -0.05929 | | | 15240 | 11083 | 0.1456 | 0.1388 | 0.02073 | 0.02658 | 0.1863 | 0.1909 | 0.105 | 0.08667 |
| -0.1145 | -0.1635 | | | 3882 | 4078 | 0.2846 | 0.2762 | 0.03597 | 0.03512 | 0.3551 | 0.345 | 0.2141 | 0.2074 |
| 0.05454 | -0.01182 | | flag | 25623 | 19957 | 0.1247 | 0.1002 | 0.01247 | 0.01454 | 0.1492 | 0.1287 | 0.1003 | 0.07168 |
| -0.101 | -0.1249 | | | 3717 | 3957 | 0.3836 | 0.3736 | 0.04027 | 0.03067 | 0.4625 | 0.4337 | 0.3046 | 0.3135 |
| 0.2578 | -1.169 | flag | | 885 | 592 | 1 | 1 | 0 | 0 | 1 | 1 | 1 | 1 |
| 0.0573 | 0.01649 | | | 25659 | 19975 | 0.1191 | 0.1128 | 0.01594 | 0.01474 | 0.1503 | 0.1416 | 0.08782 | 0.08388 |

| ldsc.h2.conf.low_female | ldsc.h2.conf.low_male | ldsc.int.flag_female | ldsc.int.flag_male | ldsc.N_female | ldsc.N_male | HDL.h2_female | HDL.h2_male | HDL.h2.se_female | HDL.h2.se_male | HDL.conf.high_female | HDL.conf.high_male | HDL.conf.low_female | HDL.conf.low_male |
|---|---|---|---|---|---|---|---|---|---|---|---|---|---|
| -0.5783 | -1.384 | | | 684 | 407 | 1 | 1 | 0 | 0 | 1 | 1 | 1 | 1 |
| -0.1315 | -0.2903 | | | 3676 | 2889 | 0.2767 | 0.3619 | 0.03768 | 0.04312 | 0.3506 | 0.4464 | 0.2029 | 0.2774 |
| -0.2426 | | | | 1620 | | | | | | | | | |
| -0.05348 | -0.2278 | | | 3271 | 3698 | 0.4461 | 0.3421 | 0.03774 | 0.0553 | 0.52 | 0.4505 | 0.3721 | 0.2337 |
| -0.2095 | -0.2824 | | | 1125 | 449 | 1 | 1 | 0 | 0 | 1 | 1 | 1 | 1 |
| -0.5998 | -2.355 | | | 1126 | 452 | 1 | 1 | 0 | 0 | 1 | 1 | 1 | 1 |
| -0.5716 | -0.5755 | | | 769 | 867 | 1 | 1 | 0 | 0 | 1 | 1 | 1 | 1 |
| 0.01303 | 0.01907 | | | 19427 | 12921 | 0.1133 | 0.1495 | 0.01478 | 0.01927 | 0.1423 | 0.1873 | 0.08432 | 0.1118 |
| 0.1145 | 0.09825 | flag | | 35694 | 26408 | 0.1483 | 0.1562 | 0.009514 | 0.01155 | 0.167 | 0.1789 | 0.1297 | 0.1336 |
| -0.04537 | -0.04441 | | | 7050 | 7214 | 0.2444 | 0.205 | 0.03558 | 0.02571 | 0.3141 | 0.2554 | 0.1747 | 0.1546 |
| 0.02902 | 0.03708 | | | 34401 | 26975 | 0.07694 | 0.1126 | 0.008464 | 0.01148 | 0.09353 | 0.1351 | 0.06035 | 0.0901 |
| -0.55 | -0.5257 | | | 1515 | 1209 | 0.6942 | 1 | 0.04737 | 0.001982 | 0.7871 | 1.004 | 0.6014 | 0.9961 |
| -0.2558 | -0.5498 | | | 2334 | 1372 | 0.6335 | 0.8135 | 0.05376 | 0.07759 | 0.7389 | 0.9656 | 0.5282 | 0.6614 |
| -0.3687 | -0.3679 | flag | | 2347 | 1374 | 0.5456 | 1 | 0.03226 | 0 | 0.6088 | 1 | 0.4824 | 1 |
| ldsc.h2.conf.low_female | ldsc.h2.conf.low_male | ldsc.int.flag_female | ldsc.int.flag_male | ldsc.N_female | ldsc.N_male | HDL.h2_female | HDL.h2_male | HDL.h2.se_female | HDL.h2.se_male | HDL.conf.high_female | HDL.conf.high_male | HDL.conf.low_female | HDL.conf.low_male |
| 0.09586 | 0.1758 | flag | flag | 1247 | 119 | 1 | 1 | 0 | 0 | 1 | 1 | 1 | 1 |
| 0.02107 | 0.01963 | flag | | 34341 | 26934 | 0.07575 | 0.09989 | 0.009891 | 0.01096 | 0.09514 | 0.1214 | 0.05637 | 0.07841 |
| -1.111 | 0.08239 | | flag | 758 | 749 | 1 | 1 | 0 | 0 | 1 | 1 | 1 | 1 |
| -0.01458 | 0.004782 | | | 7839 | 9019 | 0.2079 | 0.218 | 0.03049 | 0.02802 | 0.2677 | 0.273 | 0.1482 | 0.1631 |
| -0.03617 | -8.17e-05 | | | 14196 | 12855 | 0.1317 | 0.1555 | 0.03165 | 0.02648 | 0.1937 | 0.2074 | 0.06962 | 0.1036 |
| -0.8025 | -1.021 | | | 500 | 896 | 1 | 1 | 0 | 0 | 1 | 1 | 1 | 1 |
| -1.536 | -0.3624 | | | 570 | 926 | 1 | 1 | 0 | 0 | 1 | 1 | 1 | 1 |
| -0.9378 | -1.069 | | | 498 | 894 | 1 | 1 | 0 | 0 | 1 | 1 | 1 | 1 |
| -1.217 | -0.5334 | | | 582 | 945 | 1 | 1 | 0 | 0 | 1 | 1 | 1 | 1 |
| -0.9544 | -0.3673 | | | 500 | 895 | 1 | 1 | 0 | 0 | 1 | 1 | 1 | 1 |
| -0.4331 | -0.5138 | | | 584 | 952 | 1 | 1 | 0 | 0 | 1 | 1 | 1 | 1 |
| -0.2643 | -0.3664 | | | 1255 | 1266 | 0.9304 | 1 | 0.1476 | 0.0291 | 1.22 | 1.057 | 0.6411 | 0.943 |
| -1.733 | -0.3647 | | | 676 | 698 | 1 | 1 | 0 | 0 | 1 | 1 | 1 | 1 |
| -0.2527 | -0.1976 | | | 1151 | 1033 | 1 | 1 | 0 | 0 | 1 | 1 | 1 | 1 |
| 0.0342 | 0.004473 | | | 19280 | 15405 | 0.125 | 0.1345 | 0.0171 | 0.0184 | 0.1585 | 0.1706 | 0.09146 | 0.09845 |
| -0.08841 | -0.005762 | | | 6626 | 7983 | 0.2465 | 0.2179 | 0.03215 | 0.02968 | 0.3096 | 0.2761 | 0.1835 | 0.1598 |
| -0.1346 | -0.1852 | | | 3199 | 4307 | 0.5122 | 0.2919 | 0.04036 | 0.02886 | 0.5913 | 0.3485 | 0.4331 | 0.2353 |
| 0.03306 | 0.02806 | | | 34342 | 26963 | 0.08501 | 0.08455 | 0.008388 | 0.01021 | 0.1015 | 0.1046 | 0.06857 | 0.06453 |
| -0.404 | -0.7238 | | | 1015 | 881 | 1 | 1 | 0 | 0 | 1 | 1 | 1 | 1 |
| -0.5265 | -0.396 | | | 980 | 862 | 1 | 1 | 0 | 0 | 1 | 1 | 1 | 1 |
| -0.3222 | -1.428 | | | 643 | 622 | 1 | 1 | 0 | 0 | 1 | 1 | 1 | 1 |
| -0.456 | -0.3207 | | | 1294 | 1062 | 1 | 1 | 0 | 0 | 1 | 1 | 1 | 1 |
| 0.03629 | 0.01576 | | | 34362 | 26971 | 0.07557 | 0.08254 | 0.006295 | 0.008895 | 0.08791 | 0.09997 | 0.06323 | 0.06511 |
| -0.0769 | -0.1454 | | | 3020 | 4118 | 0.4678 | 0.3904 | 0.02359 | 0.03152 | 0.514 | 0.4522 | 0.4215 | 0.3286 |
| -0.3944 | -0.2543 | | | 1198 | 893 | 0.9319 | 1 | 0.1011 | 0 | 1.13 | 1 | 0.7337 | 1 |
| -0.07119 | -0.02718 | | | 4273 | 3588 | 0.3906 | 0.5485 | 0.0404 | 0.03837 | 0.4698 | 0.6237 | 0.3114 | 0.4733 |
| -0.6895 | -0.4889 | | | 545 | 476 | 1 | 1 | 0 | 0 | 1 | 1 | 1 | 1 |
| -0.03984 | 0.01088 | | flag | 9073 | 9195 | 0.2126 | 0.1857 | 0.02914 | 0.0183 | 0.2697 | 0.2215 | 0.1555 | 0.1498 |
| -0.04965 | -0.09782 | | | 4518 | 4635 | 0.3788 | 0.3396 | 0.04432 | 0.0419 | 0.4657 | 0.4217 | 0.2919 | 0.2575 |
| 0.08138 | -0.6421 | | | 1540 | 1211 | 1 | 1 | 0 | 0 | 1 | 1 | 1 | 1 |
| -1.395 | -1.63 | | | 705 | 423 | 1 | 1 | 0 | 0 | 1 | 1 | 1 | 1 |

| ldsc.h2.conf.low_female | ldsc.h2.conf.low_male | ldsc.int.flag_female | ldsc.int.flag_male | ldsc.N_female | ldsc.N_male | HDL.h2_female | HDL.h2_male | HDL.h2.se_female | HDL.h2.se_male | HDL.conf.high_female | HDL.conf.high_male | HDL.conf.low_female | HDL.conf.low_male |
|---|---|---|---|---|---|---|---|---|---|---|---|---|---|
| -0.4483 | -0.8367 | | | 870 | 618 | 1 | 1 | 0 | 0 | 1 | 1 | 1 | 1 |
| -0.6181 | 0.164 | | flag | 1296 | 1072 | 0.8948 | 1 | 0.1039 | 0 | 1.098 | 1 | 0.6912 | 1 |
| 0.03651 | 0.02017 | | | 34787 | 27025 | 0.09653 | 0.08731 | 0.008153 | 0.01042 | 0.1125 | 0.1077 | 0.08055 | 0.06689 |
| -0.4665 | 0.06338 | | flag | 879 | 755 | 1 | 1 | 0 | 0 | 1 | 1 | 1 | 1 |
| -0.03233 | 0.02957 | | flag | 2951 | 3717 | 0.566 | 0.4462 | 0.05146 | 0.03991 | 0.6668 | 0.5244 | 0.4651 | 0.368 |
| 0.006 | -0.01698 | | | 11849 | 8464 | 0.1385 | 0.1906 | 0.02111 | 0.02745 | 0.1799 | 0.2444 | 0.09712 | 0.1368 |
| -0.163 | -0.9948 | | | 1019 | 888 | 1 | 1 | 0 | 0 | 1 | 1 | 1 | 1 |
| -0.0335 | -0.06155 | | | 13921 | 7363 | 0.1509 | 0.1957 | 0.0222 | 0.02309 | 0.1944 | 0.241 | 0.1073 | 0.1505 |
| -0.9627 | -5.686 | | flag | 1077 | 119 | 0.8155 | 1 | 0.1318 | 0 | 1.074 | 1 | 0.5571 | 1 |
| 0.01735 | 0.0246 | | | 35815 | 26957 | 0.07718 | 0.08748 | 0.007431 | 0.007857 | 0.09175 | 0.1029 | 0.06262 | 0.07208 |
| -1.388 | -0.8471 | | | 551 | 794 | 1 | 1 | 0 | 0 | 1 | 1 | 1 | 1 |
| -0.4152 | -0.524 | | | 962 | 793 | 1 | 1 | 0 | 0 | 1 | 1 | 1 | 1 |
| -0.2956 | -0.1449 | | | 2604 | 2992 | 0.5154 | 0.4175 | 0.07394 | 0.04126 | 0.6603 | 0.4984 | 0.3704 | 0.3366 |
| -0.6669 | -0.06452 | | | 1226 | 1268 | 1 | 1 | 0.02932 | 0 | 1.057 | 1 | 0.9425 | 1 |
| 0.04607 | -0.2063 | | | 2056 | 1955 | 0.9928 | 0.776 | 0.04082 | 0.06231 | 1.073 | 0.8981 | 0.9128 | 0.6538 |
| -0.3742 | -0.008848 | | | 1225 | 1269 | 1 | 1 | 0 | 0 | 1 | 1 | 1 | 1 |
| -0.6353 | 0.06019 | flag | | 1225 | 1269 | 1 | 1 | 0 | 0 | 1 | 1 | 1 | 1 |
| -0.43 | -0.2155 | | | 2056 | 1955 | 0.5005 | 0.7587 | 0.07163 | 0.08059 | 0.6409 | 0.9166 | 0.3601 | 0.6007 |
| -0.7588 | 0.08251 | | | 1227 | 1269 | 0.8732 | 1 | 0.1162 | 0 | 1.101 | 1 | 0.6455 | 1 |
| 0.0467 | -0.2063 | | | 2057 | 1955 | 0.9951 | 0.776 | 0.03919 | 0.06231 | 1.072 | 0.8981 | 0.9183 | 0.6538 |
| -0.3728 | -0.008848 | | | 1226 | 1269 | 1 | 1 | 0 | 0 | 1 | 1 | 1 | 1 |
| -0.6477 | 0.06019 | flag | | 1226 | 1269 | 1 | 1 | 7.99e-04 | 0 | 1.002 | 1 | 0.9984 | 1 |
| -0.7396 | 0.132 | | | 1227 | 1269 | 0.8626 | 1 | 0.1048 | 0 | 1.068 | 1 | 0.6571 | 1 |
| -0.7211 | -0.5306 | | | 1152 | 976 | 1 | 1 | 0.05271 | 0 | 1.103 | 1 | 0.8967 | 1 |
| 0.09746 | -0.3119 | | flag | 2056 | 1954 | 0.9785 | 0.6341 | 0.04632 | 0.05114 | 1.069 | 0.7343 | 0.8877 | 0.5338 |
| -0.4221 | -0.2938 | | | 2057 | 1954 | 0.7596 | 0.8527 | 0.05123 | 0.06695 | 0.86 | 0.984 | 0.6592 | 0.7215 |
| -0.6952 | -1.497 | | | 936 | 680 | 1 | 1 | 0 | 0 | 1 | 1 | 1 | 1 |
| 0.04187 | 0.01964 | | | 34789 | 27030 | 0.0966 | 0.0862 | 0.007791 | 0.009684 | 0.1119 | 0.1052 | 0.08133 | 0.06722 |
| -0.5455 | -0.1795 | | flag | 1649 | 1251 | 0.676 | 1 | 0.07829 | 0 | 0.8294 | 1 | 0.5225 | 1 |
| ldsc.h2.conf.low_female | ldsc.h2.conf.low_male | ldsc.int.flag_female | ldsc.int.flag_male | ldsc.N_female | ldsc.N_male | HDL.h2_female | HDL.h2_male | HDL.h2.se_female | HDL.h2.se_male | HDL.conf.high_female | HDL.conf.high_male | HDL.conf.low_female | HDL.conf.low_male |
| -1.171 | -1.53 | | flag | 817 | 277 | 1 | 1 | 0 | 0 | 1 | 1 | 1 | 1 |
| 0.02972 | 0.05112 | | | 14884 | 10829 | 0.1716 | 0.2058 | 0.02003 | 0.02657 | 0.2109 | 0.2579 | 0.1324 | 0.1538 |
| 0.04948 | 0.0247 | | | 15677 | 11491 | 0.1449 | 0.169 | 0.0168 | 0.02133 | 0.1778 | 0.2108 | 0.1119 | 0.1272 |
| 0.02889 | -0.008527 | | | 19735 | 15202 | 0.1219 | 0.1335 | 0.01219 | 0.0126 | 0.1458 | 0.1582 | 0.09801 | 0.1088 |
| 0.03098 | 0.01292 | | | 15257 | 12637 | 0.1201 | 0.1432 | 0.01407 | 0.0147 | 0.1477 | 0.172 | 0.09254 | 0.1144 |
| 0.01984 | 0.03459 | | | 19669 | 15117 | 0.1169 | 0.1449 | 0.01443 | 0.01537 | 0.1452 | 0.175 | 0.08862 | 0.1148 |
| -0.9301 | -0.8021 | flag | | 1111 | 730 | 1 | 1 | 0.001879 | 0 | 1.004 | 1 | 0.9963 | 1 |
| 0.03131 | -0.01355 | | | 15398 | 12813 | 0.1182 | 0.1387 | 0.01535 | 0.01981 | 0.1483 | 0.1775 | 0.08809 | 0.09983 |
| -0.2737 | -0.2268 | | flag | 2960 | 169 | 0.3312 | 1 | 0.03767 | 0 | 0.4051 | 1 | 0.2574 | 1 |
| -1.014 | -0.7966 | | | 716 | 975 | 1 | 1 | 0 | 0 | 1 | 1 | 1 | 1 |
| -0.2427 | 0.01195 | | flag | 2953 | 3225 | 0.4297 | 0.5312 | 0.04096 | 0.03891 | 0.51 | 0.6075 | 0.3495 | 0.4549 |
| -0.8148 | -0.4232 | flag | | 1018 | 810 | 1 | 1 | 0 | 0 | 1 | 1 | 1 | 1 |
| 0.01469 | -0.289 | | | 1559 | 1210 | 1 | 1 | 0.002889 | 0 | 1.006 | 1 | 0.9943 | 1 |
| 0.07643 | -0.356 | | flag | 2730 | 2375 | 0.7506 | 0.6095 | 0.03516 | 0.04335 | 0.8195 | 0.6944 | 0.6816 | 0.5245 |
| -0.2593 | -0.06284 | | | 1686 | 1581 | 0.9443 | 1 | 0.07558 | 0 | 1.092 | 1 | 0.7962 | 1 |
| -0.1787 | -0.1275 | | | 2730 | 2375 | 0.6214 | 0.5491 | 0.05357 | 0.04251 | 0.7264 | 0.6324 | 0.5164 | 0.4658 |

| ldsc.h2.conf.low_female | ldsc.h2.conf.low_male | ldsc.int.flag_female | ldsc.int.flag_male | ldsc.N_female | ldsc.N_male | HDL.h2_female | HDL.h2_male | HDL.h2.se_female | HDL.h2.se_male | HDL.conf.high_female | HDL.conf.high_male | HDL.conf.low_female | HDL.conf.low_male |
|---|---|---|---|---|---|---|---|---|---|---|---|---|---|
| -0.5947 | -0.1843 | | flag | 576 | 497 | 1 | 1 | 0 | 0 | 1 | 1 | 1 | 1 |
| -0.602 | -0.7287 | | | 512 | 669 | 1 | 1 | 0 | 0 | 1 | 1 | 1 | 1 |
| 0.001923 | -0.05139 | | | 11119 | 8034 | 0.1484 | 0.182 | 0.02141 | 0.02673 | 0.1904 | 0.2344 | 0.1064 | 0.1297 |
| -0.8072 | -1.199 | | | 585 | 503 | 1 | 1 | 0 | 0 | 1 | 1 | 1 | 1 |
| -0.2708 | -0.2133 | | | 1730 | 1595 | 0.7831 | 0.827 | 0.05609 | 0.05669 | 0.893 | 0.9382 | 0.6731 | 0.7159 |
| -0.3523 | -1.165 | | | 580 | 485 | 1 | 1 | 0 | 0 | 1 | 1 | 1 | 1 |
| -0.2217 | -0.1821 | | | 1562 | 1214 | 0.8587 | 1 | 0.08924 | 0 | 1.034 | 1 | 0.6838 | 1 |
| 0.1416 | -0.3996 | | | 1562 | 1214 | 1 | 1 | 0 | 0 | 1 | 1 | 1 | 1 |
| -0.2799 | -0.06076 | | | 1735 | 1604 | 0.9928 | 1 | 0.06931 | 0 | 1.129 | 1 | 0.857 | 1 |
| -0.375 | -0.1728 | | | 1562 | 1214 | 0.8337 | 1 | 0.09037 | 0 | 1.011 | 1 | 0.6566 | 1 |
| -0.2064 | -0.9962 | | | 567 | 488 | 1 | 1 | 0 | 0 | 1 | 1 | 1 | 1 |
| -1.406 | 0.1591 | | | 587 | 506 | 1 | 1 | 0 | 0 | 1 | 1 | 1 | 1 |
| -1.356 | 0.1102 | | | 587 | 506 | 1 | 1 | 0 | 0 | 1 | 1 | 1 | 1 |
| -0.2074 | -0.2639 | | | 1735 | 1604 | 0.9796 | 0.9296 | 0.09971 | 0.08515 | 1.175 | 1.096 | 0.7842 | 0.7627 |
| -0.1443 | -0.3202 | | | 1735 | 1604 | 0.8945 | 0.7707 | 0.08174 | 0.1083 | 1.055 | 0.983 | 0.7343 | 0.5585 |
| -0.2942 | -0.3244 | | | 1735 | 1604 | 0.7632 | 0.7547 | 0.03451 | 0.1013 | 0.8309 | 0.9532 | 0.6956 | 0.5561 |
| -0.2427 | -0.3199 | | | 1734 | 1604 | 0.9391 | 0.9106 | 0.0967 | 0.0794 | 1.129 | 1.066 | 0.7496 | 0.7549 |
| -0.2466 | -0.07787 | | | 3411 | 4836 | 0.3087 | 0.3901 | 0.02472 | 0.03945 | 0.3572 | 0.4674 | 0.2603 | 0.3128 |
| -0.7905 | -0.4652 | | | 673 | 960 | 1 | 1 | 0 | 0 | 1 | 1 | 1 | 1 |
| -0.5913 | -0.4703 | | | 1301 | 1616 | 1 | 0.8575 | 0 | 0.08142 | 1 | 1.017 | 1 | 0.6979 |
| -1.466 | -0.8369 | | | 660 | 793 | 1 | 1 | 0 | 0 | 1 | 1 | 1 | 1 |
| -0.8372 | -1.234 | | flag | 744 | 916 | 1 | 1 | 0 | 0 | 1 | 1 | 1 | 1 |
| -0.7994 | -0.7445 | flag | | 1288 | 1610 | 0.8259 | 0.7757 | 0.08923 | 0.05812 | 1.001 | 0.8896 | 0.651 | 0.6617 |
| -0.3057 | -0.09653 | | | 1574 | 1952 | 1 | 0.8764 | 0 | 0.06373 | 1 | 1.001 | 1 | 0.7515 |
| -1.688 | -0.3032 | | | 743 | 990 | 1 | 1 | 0 | 0 | 1 | 1 | 1 | 1 |
| -0.3574 | -0.2659 | | | 1498 | 1855 | 0.8224 | 0.7373 | 0.1197 | 0.05703 | 1.057 | 0.8491 | 0.5878 | 0.6255 |
| 0.0141 | -0.0247 | | | 1144 | 1145 | 1 | 1 | 0 | 0 | 1 | 1 | 1 | 1 |
| -0.8246 | -0.5495 | flag | | 1147 | 1144 | 1 | 1 | 0.001089 | 0 | 1.002 | 1 | 0.9979 | 1 |
| -0.7989 | -0.5641 | flag | | 1146 | 1145 | 1 | 1 | 0.00165 | 0 | 1.003 | 1 | 0.9968 | 1 |
| -0.7495 | -0.5148 | flag | | 1142 | 1144 | 1 | 1 | 0.001138 | 0 | 1.002 | 1 | 0.9978 | 1 |
| -0.04884 | -0.0792 | | | 1145 | 1145 | 1 | 1 | 0 | 0 | 1 | 1 | 1 | 1 |
| -0.04334 | -0.06075 | | | 7138 | 6253 | 0.2103 | 0.2176 | 0.03114 | 0.03062 | 0.2713 | 0.2776 | 0.1492 | 0.1576 |
| -0.05509 | -0.1015 | | | 7218 | 6315 | 0.2377 | 0.2576 | 0.04029 | 0.03242 | 0.3166 | 0.3211 | 0.1587 | 0.194 |
| 0.007235 | -0.08217 | | | 7040 | 6062 | 0.3461 | 0.2294 | 0.04504 | 0.03506 | 0.4343 | 0.2982 | 0.2578 | 0.1607 |
| -0.1766 | -0.1298 | | | 7193 | 6301 | 0.1432 | 0.2341 | 0.0282 | 0.03507 | 0.1985 | 0.3028 | 0.08795 | 0.1653 |
| -0.05012 | -0.05851 | | | 7197 | 6296 | 0.2021 | 0.2384 | 0.02287 | 0.03302 | 0.247 | 0.3032 | 0.1573 | 0.1737 |
| -0.0223 | -0.2372 | | | 3733 | 659 | 0.4164 | 1 | 0.03184 | 0 | 0.4788 | 1 | 0.354 | 1 |
| -0.007504 | 0.01427 | | | 17572 | 10236 | 0.09089 | 0.175 | 0.01073 | 0.01951 | 0.1119 | 0.2132 | 0.06985 | 0.1368 |
| -0.6919 | -0.1608 | | | 1347 | 1663 | 0.9689 | 0.8722 | 0.07876 | 0.08465 | 1.123 | 1.038 | 0.8146 | 0.7062 |
| -0.2424 | -0.3312 | | | 1562 | 1214 | 0.9927 | 1 | 0.03851 | 0 | 1.068 | 1 | 0.9172 | 1 |
| 0.151 | -0.3976 | | | 1562 | 1214 | 1 | 1 | 0 | 0 | 1 | 1 | 1 | 1 |
| -0.3444 | -0.07948 | | | 1736 | 1604 | 0.9362 | 1 | 0.07135 | 0 | 1.076 | 1 | 0.7964 | 1 |
| -0.1726 | -0.1661 | | | 1562 | 1214 | 0.926 | 1 | 0.06717 | 0 | 1.058 | 1 | 0.7943 | 1 |
| -0.9434 | -0.4988 | | flag | 550 | 485 | 1 | 1 | 0 | 0 | 1 | 1 | 1 | 1 |
| ldsc.h2.conf.low_female | ldsc.h2.conf.low_male | ldsc.int.flag_female | ldsc.int.flag_male | ldsc.N_female | ldsc.N_male | HDL.h2_female | HDL.h2_male | HDL.h2.se_female | HDL.h2.se_male | HDL.conf.high_female | HDL.conf.high_male | HDL.conf.low_female | HDL.conf.low_male |
| 0.0848 | -0.05977 | | | 3670 | 2890 | 0.5336 | 0.52 | 0.03918 | 0.03897 | 0.6104 | 0.5964 | 0.4568 | 0.4436 |

| ldsc.h2.conf.low_female | ldsc.h2.conf.low_male | ldsc.int.flag_female | ldsc.int.flag_male | ldsc.N_female | ldsc.N_male | HDL.h2_female | HDL.h2_male | HDL.h2.se_female | HDL.h2.se_male | HDL.conf.high_female | HDL.conf.high_male | HDL.conf.low_female | HDL.conf.low_male |
|---|---|---|---|---|---|---|---|---|---|---|---|---|---|
| -0.1038 | -0.2581 | | | 2363 | 2733 | 0.6783 | 0.4478 | 0.08011 | 0.04057 | 0.8353 | 0.5273 | 0.5213 | 0.3683 |
| -0.5352 | -1.414 | flag | | 1060 | 715 | 1 | 1 | 0 | 0 | 1 | 1 | 1 | 1 |
| -0.04827 | -0.01586 | | | 13645 | 13466 | 0.1332 | 0.1375 | 0.02062 | 0.02026 | 0.1736 | 0.1772 | 0.09274 | 0.09775 |
| 0.05096 | 0.03229 | flag | flag | 34448 | 26960 | 0.09859 | 0.1074 | 0.009425 | 0.01048 | 0.1171 | 0.128 | 0.08012 | 0.08689 |
| -0.01247 | -0.01804 | | | 6045 | 7936 | 0.2931 | 0.2198 | 0.03407 | 0.02946 | 0.3599 | 0.2775 | 0.2263 | 0.162 |
| -0.2283 | | flag | | 2743 | | | | | | | | | |
| -0.1161 | -0.3629 | | | 1516 | 1550 | 1 | 1 | 0 | 1.95e-04 | 1 | 1 | 1 | 0.9996 |
| -0.8434 | -0.4473 | flag | | 1182 | 1215 | 1 | 1 | 0 | 0 | 1 | 1 | 1 | 1 |
| -0.2887 | -0.2074 | | | 2102 | 2001 | 0.5922 | 0.6837 | 0.08279 | 0.06454 | 0.7544 | 0.8102 | 0.4299 | 0.5572 |
| 0.007814 | 7.02e-04 | | | 8404 | 9812 | 0.1938 | 0.1738 | 0.02338 | 0.02833 | 0.2397 | 0.2293 | 0.148 | 0.1183 |
| -0.1188 | -0.06079 | | | 5311 | 6990 | 0.2274 | 0.227 | 0.03119 | 0.03338 | 0.2885 | 0.2925 | 0.1662 | 0.1616 |
| 0.07349 | 0.04589 | flag | flag | 18580 | 14828 | 0.1808 | 0.1558 | 0.01717 | 0.01926 | 0.2145 | 0.1936 | 0.1472 | 0.1181 |
| -0.01845 | -0.1768 | | flag | 7229 | 6318 | 0.2017 | 0.1388 | 0.03868 | 0.028 | 0.2775 | 0.1937 | 0.1259 | 0.08394 |
| 0.2211 | 0.1542 | flag | flag | 34577 | 25698 | 0.248 | 0.2275 | 0.01751 | 0.01716 | 0.2823 | 0.2611 | 0.2137 | 0.1939 |
| 0.2323 | 0.05063 | | flag | 19052 | 12621 | 0.2821 | 0.2325 | 0.02039 | 0.02488 | 0.322 | 0.2813 | 0.2421 | 0.1838 |
| -0.7606 | -0.7706 | flag | | 718 | 509 | 1 | 1 | 0 | 0 | 1 | 1 | 1 | 1 |
| -0.9406 | -0.5918 | flag | | 719 | 509 | 1 | 1 | 0 | 0 | 1 | 1 | 1 | 1 |
| 0.02032 | 0.03862 | flag | | 35678 | 26968 | 0.07219 | 0.09779 | 0.006745 | 0.01095 | 0.08541 | 0.1192 | 0.05897 | 0.07632 |
| -0.2807 | -0.07335 | | | 657 | 769 | 1 | 1 | 0 | 0 | 1 | 1 | 1 | 1 |
| 0.03829 | 0.04531 | | | 14409 | 11801 | 0.1589 | 0.2115 | 0.01243 | 0.01692 | 0.1833 | 0.2446 | 0.1346 | 0.1783 |
| -0.6398 | -1.906 | | | 829 | 538 | 1 | 1 | 0 | 0 | 1 | 1 | 1 | 1 |
| 0.02699 | -0.08625 | | | 7139 | 6239 | 0.311 | 0.2844 | 0.03793 | 0.04275 | 0.3854 | 0.3681 | 0.2367 | 0.2006 |
| -0.06978 | -0.07718 | | | 7074 | 6205 | 0.1515 | 0.2266 | 0.02236 | 0.02968 | 0.1953 | 0.2848 | 0.1077 | 0.1684 |
| -0.3553 | -0.1171 | | flag | 676 | 369 | 1 | 1 | 0 | 0 | 1 | 1 | 1 | 1 |
| -0.01047 | -0.00817 | | | 21462 | 19279 | 0.08494 | 0.08785 | 0.01228 | 0.01277 | 0.109 | 0.1129 | 0.06087 | 0.06282 |
| -0.6732 | -1.099 | | | 555 | 585 | 1 | 1 | 0 | 0 | 1 | 1 | 1 | 1 |
| -0.3357 | 0.02901 | | | 1168 | 1168 | 1 | 1 | 0 | 0 | 1 | 1 | 1 | 1 |
| -0.8115 | -0.5802 | flag | | 1168 | 1167 | 0.9391 | 1 | 0.07101 | 0 | 1.078 | 1 | 0.7999 | 1 |
| -0.3407 | -0.04145 | | | 1167 | 1168 | 1 | 1 | 0 | 0 | 1 | 1 | 1 | 1 |
| -0.1829 | 0.1102 | flag | flag | 2408 | 1967 | 0.5216 | 0.9449 | 0.06579 | 0.05642 | 0.6505 | 1.055 | 0.3926 | 0.8343 |
| -0.04716 | -0.4361 | | | 4327 | 1980 | 0.372 | 0.6818 | 0.04283 | 0.08833 | 0.4559 | 0.855 | 0.288 | 0.5087 |
| 0.01895 | 0.005864 | flag | | 24902 | 19254 | 0.09347 | 0.0921 | 0.009951 | 0.01173 | 0.113 | 0.1151 | 0.07396 | 0.0691 |
| -0.3407 | -0.2663 | flag | | 1008 | 557 | 1 | 1 | 0 | 0 | 1 | 1 | 1 | 1 |
| -0.303 | -0.2539 | | | 2071 | 1520 | 0.697 | 0.9616 | 0.05953 | 0.09656 | 0.8136 | 1.151 | 0.5803 | 0.7723 |
| -0.4004 | -0.2955 | | | 1272 | 902 | 1 | 1 | 0.08212 | 0 | 1.161 | 1 | 0.839 | 1 |
| 9.48e-04 | -1.30e-04 | | | 24933 | 19256 | 0.06698 | 0.08448 | 0.01 | 0.01185 | 0.08658 | 0.1077 | 0.04738 | 0.06125 |
| -0.3415 | | | | 1596 | | | | | | | | | |
| -0.3415 | | | | 1596 | | | | | | | | | |
| -0.4934 | | | | 1624 | | | | | | | | | |
| -0.2937 | | | | 1622 | | | | | | | | | |
| -0.173 | | | | 1628 | | | | | | | | | |
| -0.5677 | | | | 1588 | | | | | | | | | |
| -0.3136 | | | | 1629 | | | | | | | | | |
| -0.487 | | | | 1595 | | | | | | | | | |
| -0.4341 | | | | 1630 | | | | | | | | | |
| -0.1336 | | | | 1593 | | | | | | | | | |
| -0.2697 | | | | 1619 | | | | | | | | | |

| ldsc.h2.conf.low_female | ldsc.h2.conf.low_male | ldsc.int.flag_female | ldsc.int.flag_male | ldsc.N_female | ldsc.N_male | HDL.h2_female | HDL.h2_male | HDL.h2.se_female | HDL.h2.se_male | HDL.conf.high_female | HDL.conf.high_male | HDL.conf.low_female | HDL.conf.low_male |
|---|---|---|---|---|---|---|---|---|---|---|---|---|---|
| -0.429 | | | | 1594 | | | | | | | | | |
| 0.02773 | 0.04755 | | | 21438 | 19244 | 0.1025 | 0.1309 | 0.01142 | 0.01518 | 0.1249 | 0.1607 | 0.0801 | 0.1012 |
| -0.4731 | -1.764 | | | 1372 | 331 | 0.8866 | 1 | 0.06489 | 0 | 1.014 | 1 | 0.7595 | 1 |
| -0.02441 | -0.4141 | | | 2403 | 2273 | 0.673 | 0.6195 | 0.06491 | 0.06758 | 0.8002 | 0.7519 | 0.5457 | 0.487 |
| -0.3082 | 0.04535 | | | 3085 | 2822 | 0.4348 | 0.6153 | 0.04138 | 0.05112 | 0.5159 | 0.7155 | 0.3537 | 0.5151 |
| -0.05122 | -0.05003 | flag | | 10737 | 7897 | 0.1372 | 0.2128 | 0.02456 | 0.02433 | 0.1854 | 0.2605 | 0.0891 | 0.1651 |
| 0.03455 | -0.08588 | | | 6320 | 8267 | 0.2853 | 0.2136 | 0.02636 | 0.02738 | 0.337 | 0.2673 | 0.2337 | 0.16 |
| 0.01181 | 0.01686 | | | 34362 | 26975 | 0.06621 | 0.08598 | 0.008159 | 0.008045 | 0.0822 | 0.1017 | 0.05022 | 0.07021 |
| -0.002247 | -0.1921 | | | 973 | 640 | 1 | 1 | 0 | 0 | 1 | 1 | 1 | 1 |
| -0.1911 | 0.5689 | | flag | 949 | 617 | 1 | 1 | 0 | 0 | 1 | 1 | 1 | 1 |
| -0.05822 | -0.09756 | | | 6190 | 6279 | 0.2043 | 0.1962 | 0.03637 | 0.03706 | 0.2756 | 0.2689 | 0.133 | 0.1236 |
| -0.02611 | -0.02624 | | | 6477 | 8287 | 0.2538 | 0.1922 | 0.03449 | 0.02499 | 0.3214 | 0.2411 | 0.1862 | 0.1432 |
| ldsc.h2.conf.low_female | ldsc.h2.conf.low_male | ldsc.int.flag_female | ldsc.int.flag_male | ldsc.N_female | ldsc.N_male | HDL.h2_female | HDL.h2_male | HDL.h2.se_female | HDL.h2.se_male | HDL.conf.high_female | HDL.conf.high_male | HDL.conf.low_female | HDL.conf.low_male |
| -0.3289 | -0.7798 | | | 971 | 639 | 1 | 1 | 0 | 0 | 1 | 1 | 1 | 1 |
| -0.1139 | -0.1763 | | | 6376 | 5881 | 0.2276 | 0.2152 | 0.03636 | 0.03536 | 0.2989 | 0.2845 | 0.1563 | 0.1459 |
| -0.9959 | 0.07875 | | | 806 | 700 | 1 | 1 | 0 | 0 | 1 | 1 | 1 | 1 |
| -0.01144 | -0.4842 | | flag | 1151 | 1029 | 1 | 1 | 0 | 0 | 1 | 1 | 1 | 1 |
| 0.00445 | -0.002594 | | | 18095 | 14481 | 0.09841 | 0.15 | 0.01917 | 0.01928 | 0.136 | 0.1878 | 0.06084 | 0.1122 |
| -0.3833 | -0.6308 | flag | | 2351 | 1533 | 0.6035 | 0.8549 | 0.0483 | 0.08011 | 0.6982 | 1.012 | 0.5088 | 0.6979 |
| -0.1055 | -0.8647 | flag | | 1594 | 861 | 0.8232 | 1 | 0.09083 | 0 | 1.001 | 1 | 0.6452 | 1 |
| 0.02685 | -0.07075 | | | 6454 | 5058 | 0.3147 | 0.2622 | 0.03575 | 0.03695 | 0.3848 | 0.3346 | 0.2446 | 0.1898 |
| -0.07479 | -0.4039 | | | 2762 | 2114 | 0.4888 | 0.5238 | 0.04521 | 0.06183 | 0.5774 | 0.645 | 0.4002 | 0.4027 |
| -0.1139 | -0.2085 | | | 3541 | 2510 | 0.3718 | 0.5263 | 0.04086 | 0.05324 | 0.4519 | 0.6306 | 0.2918 | 0.4219 |
| -0.1092 | -0.3432 | | | 3547 | 2514 | 0.4359 | 0.3525 | 0.04926 | 0.05245 | 0.5324 | 0.4553 | 0.3393 | 0.2497 |
| -0.00786 | -0.03616 | flag | | 15914 | 11637 | 0.1107 | 0.157 | 0.01609 | 0.02005 | 0.1423 | 0.1963 | 0.0792 | 0.1177 |
| 0.01939 | -0.01303 | | | 15127 | 11025 | 0.1374 | 0.1764 | 0.01798 | 0.02528 | 0.1726 | 0.226 | 0.1022 | 0.1269 |
| -0.1013 | -0.04633 | | | 5177 | 5327 | 0.236 | 0.3415 | 0.02089 | 0.03315 | 0.277 | 0.4065 | 0.1951 | 0.2765 |
| 0.04314 | -0.004406 | | | 25616 | 19927 | 0.108 | 0.1001 | 0.01014 | 0.01188 | 0.1279 | 0.1234 | 0.08815 | 0.07679 |
| -0.5331 | -1.131 | | | 1237 | 795 | 1 | 1 | 0 | 0 | 1 | 1 | 1 | 1 |
| -0.09227 | 0.006999 | | | 5362 | 5485 | 0.2572 | 0.3252 | 0.0401 | 0.02449 | 0.3358 | 0.3732 | 0.1786 | 0.2772 |
| 0.02952 | 0.01104 | | | 25619 | 19940 | 0.09455 | 0.09965 | 0.01005 | 0.01159 | 0.1142 | 0.1224 | 0.07486 | 0.07693 |
| -0.05484 | 0.02899 | | | 6477 | 7881 | 0.294 | 0.2426 | 0.03053 | 0.02492 | 0.3539 | 0.2914 | 0.2342 | 0.1937 |
| 0.07713 | 0.08502 | | | 35668 | 26968 | 0.1252 | 0.1347 | 0.01434 | 0.0139 | 0.1533 | 0.1619 | 0.09711 | 0.1074 |
| 0.0716 | 0.04438 | | | 35644 | 26945 | 0.1163 | 0.1215 | 0.009713 | 0.01301 | 0.1353 | 0.147 | 0.09727 | 0.09599 |
| -0.021 | 0.01817 | | | 35676 | 26971 | 0.04022 | 0.08765 | 0.007397 | 0.00934 | 0.05472 | 0.106 | 0.02572 | 0.06934 |
| -0.1335 | -0.1833 | | | 3337 | 3552 | 0.4243 | 0.3284 | 0.03706 | 0.03452 | 0.4969 | 0.3961 | 0.3516 | 0.2607 |
| -0.4029 | -0.7742 | | | 743 | 847 | 1 | 1 | 0 | 0 | 1 | 1 | 1 | 1 |
| -0.06888 | -0.2314 | | | 3474 | 3619 | 0.4633 | 0.4234 | 0.03211 | 0.05939 | 0.5262 | 0.5398 | 0.4004 | 0.307 |
| -0.1471 | 0.01147 | flag | | 3632 | 3884 | 0.3971 | 0.3997 | 0.03341 | 0.03296 | 0.4626 | 0.4643 | 0.3316 | 0.3351 |
| -0.2178 | -0.3268 | | | 1364 | 1490 | 1 | 0.9449 | 0 | 0.08346 | 1 | 1.108 | 1 | 0.7813 |
| 0.03629 | 0.03084 | | | 21442 | 19258 | 0.1334 | 0.1265 | 0.01449 | 0.01232 | 0.1618 | 0.1507 | 0.105 | 0.1024 |
| -0.1954 | -0.09836 | | | 3623 | 3764 | 0.3519 | 0.4113 | 0.03233 | 0.04749 | 0.4153 | 0.5044 | 0.2886 | 0.3183 |
| -0.1576 | 0.006728 | | flag | 2965 | 4068 | 0.5076 | 0.3387 | 0.05691 | 0.03253 | 0.6191 | 0.4025 | 0.396 | 0.2749 |
| -0.00806 | -0.02916 | | | 12679 | 11442 | 0.1341 | 0.1296 | 0.01563 | 0.02323 | 0.1647 | 0.1751 | 0.1034 | 0.08409 |
| -0.452 | -0.3151 | | | 902 | 689 | 1 | 1 | 0 | 0 | 1 | 1 | 1 | 1 |
| 0.0405 | -0.006855 | | | 15125 | 11006 | 0.1425 | 0.1887 | 0.01931 | 0.02365 | 0.1803 | 0.235 | 0.1046 | 0.1423 |

| ldsc.h2.conf.low_female | ldsc.h2.conf.low_male | ldsc.int.flag_female | ldsc.int.flag_male | ldsc.N_female | ldsc.N_male | HDL.h2_female | HDL.h2_male | HDL.h2.se_female | HDL.h2.se_male | HDL.conf.high_female | HDL.conf.high_male | HDL.conf.low_female | HDL.conf.low_male |
|---|---|---|---|---|---|---|---|---|---|---|---|---|---|
| -0.139 | -0.1579 | | | 5055 | 5259 | 0.2518 | 0.2822 | 0.03395 | 0.04267 | 0.3184 | 0.3659 | 0.1853 | 0.1986 |
| 0.05634 | 0.009598 | | flag | 25647 | 19931 | 0.1199 | 0.1191 | 0.01259 | 0.01402 | 0.1445 | 0.1466 | 0.09519 | 0.09159 |
| 0.03543 | 0.02082 | | | 15947 | 11630 | 0.1532 | 0.178 | 0.02193 | 0.02435 | 0.1961 | 0.2257 | 0.1102 | 0.1303 |
| -0.3225 | -1.05 | | | 1238 | 794 | 1 | 1 | 0 | 0 | 1 | 1 | 1 | 1 |
| -0.05549 | -0.1658 | | | 5096 | 5320 | 0.3077 | 0.2282 | 0.02906 | 0.03166 | 0.3647 | 0.2903 | 0.2508 | 0.1662 |
| 0.06847 | 0.03048 | | flag | 25672 | 19994 | 0.1267 | 0.1311 | 0.01476 | 0.01711 | 0.1556 | 0.1646 | 0.09779 | 0.09753 |
| 0.1502 | 0.09814 | flag | flag | 26726 | 19784 | 0.203 | 0.2016 | 0.02718 | 0.02363 | 0.2563 | 0.2479 | 0.1497 | 0.1553 |
| -0.1581 | -0.1059 | flag | | 3755 | 3995 | 0.4048 | 0.3492 | 0.02775 | 0.03561 | 0.4592 | 0.419 | 0.3504 | 0.2794 |
| -0.04769 | -0.04469 | | | 5957 | 7792 | 0.2851 | 0.2146 | 0.03106 | 0.02797 | 0.346 | 0.2694 | 0.2242 | 0.1598 |
| 0.02849 | 0.03667 | | | 34356 | 26971 | 0.07973 | 0.09891 | 0.009223 | 0.01057 | 0.09781 | 0.1196 | 0.06166 | 0.07819 |
| -0.6178 | -0.3246 | flag | | 1278 | 1748 | 1 | 0.7926 | 0 | 0.0501 | 1 | 0.8908 | 1 | 0.6944 |
| 0.01207 | -0.0277 | | | 16047 | 11786 | 0.118 | 0.1622 | 0.01415 | 0.02072 | 0.1457 | 0.2028 | 0.09028 | 0.1216 |
| -0.1779 | -0.03847 | | | 5142 | 5349 | 0.2668 | 0.2581 | 0.01913 | 0.03499 | 0.3043 | 0.3267 | 0.2293 | 0.1896 |
| 0.02488 | 0.01546 | | | 25593 | 19958 | 0.1009 | 0.1249 | 0.009324 | 0.01518 | 0.1191 | 0.1546 | 0.08258 | 0.09514 |
| -0.1024 | -0.08443 | | | 5147 | 5352 | 0.2838 | 0.2665 | 0.03447 | 0.02699 | 0.3513 | 0.3194 | 0.2162 | 0.2136 |
| 0.02717 | 0.01885 | | | 25683 | 20000 | 0.09272 | 0.1084 | 0.01044 | 0.01296 | 0.1132 | 0.1339 | 0.07226 | 0.08304 |
| -0.2068 | 0.002121 | | | 1846 | 2171 | 0.918 | 0.7959 | 0.04902 | 0.06407 | 1.014 | 0.9215 | 0.8219 | 0.6703 |
| 0.005337 | -0.0412 | | | 16758 | 12556 | 0.09866 | 0.1047 | 0.01485 | 0.01741 | 0.1278 | 0.1388 | 0.06956 | 0.07053 |
| -0.02362 | -0.004124 | | flag | 31113 | 23718 | 0.03813 | 0.09268 | 0.007779 | 0.009599 | 0.05338 | 0.1115 | 0.02289 | 0.07387 |
| -0.2269 | -0.5137 | | | 1048 | 1153 | 1 | 1 | 0 | 1.49e-04 | 1 | 1 | 1 | 0.9997 |
| -0.5345 | -0.8435 | | | 1045 | 1142 | 1 | 1 | 0 | 4.32e-04 | 1 | 1.001 | 1 | 0.9992 |
| -0.0438 | -0.2126 | | | 5143 | 2958 | 0.3581 | 0.4816 | 0.02975 | 0.04763 | 0.4164 | 0.575 | 0.2998 | 0.3883 |
| 0.03957 | -3.88e-04 | | | 15125 | 11008 | 0.1488 | 0.1788 | 0.01541 | 0.0208 | 0.179 | 0.2195 | 0.1186 | 0.138 |
| 0.02859 | 0.02946 | | | 27996 | 21662 | 0.09361 | 0.1116 | 0.007531 | 0.01288 | 0.1084 | 0.1369 | 0.07885 | 0.08641 |
| -0.05974 | -0.0527 | | | 9833 | 9654 | 0.1223 | 0.1346 | 0.02085 | 0.02046 | 0.1632 | 0.1747 | 0.08146 | 0.09447 |
| -0.1401 | -0.07954 | | | 4695 | 6419 | 0.3051 | 0.2351 | 0.04453 | 0.02893 | 0.3924 | 0.2918 | 0.2178 | 0.1784 |
| 0.01499 | 0.01074 | | | 30062 | 23729 | 0.06556 | 0.0971 | 0.008721 | 0.01041 | 0.08265 | 0.1175 | 0.04847 | 0.0767 |

| ldsc.h2.conf.low_female | ldsc.h2.conf.low_male | ldsc.int.flag_female | ldsc.int.flag_male | ldsc.N_female | ldsc.N_male | HDL.h2_female | HDL.h2_male | HDL.h2.se_female | HDL.h2.se_male | HDL.conf.high_female | HDL.conf.high_male | HDL.conf.low_female | HDL.conf.low_male |
|---|---|---|---|---|---|---|---|---|---|---|---|---|---|
| -0.02226 | -0.17 | | flag | 3797 | 4606 | 0.3722 | 0.3679 | 0.02318 | 0.0399 | 0.4176 | 0.4461 | 0.3267 | 0.2896 |
| -0.106 | -0.07922 | | | 3787 | 4535 | 0.467 | 0.3585 | 0.04065 | 0.03355 | 0.5467 | 0.4243 | 0.3873 | 0.2928 |
| -0.6229 | | | | 1217 | | | | | | | | | |
| -0.5789 | | | | 1217 | | | | | | | | | |
| -0.2788 | -0.3844 | | flag | 1780 | 1755 | 0.7998 | 0.8386 | 0.05482 | 0.04334 | 0.9073 | 0.9235 | 0.6924 | 0.7536 |
| 0.1005 | -0.02496 | flag | | 7220 | 6307 | 0.208 | 0.2754 | 0.03475 | 0.02975 | 0.2761 | 0.3337 | 0.1399 | 0.2171 |
| -0.03282 | -0.06736 | | | 6926 | 5960 | 0.212 | 0.2517 | 0.01945 | 0.02828 | 0.2501 | 0.3071 | 0.1739 | 0.1962 |
| -2.51e-04 | 0.02608 | flag | | 21451 | 19267 | 0.08763 | 0.131 | 0.01282 | 0.01519 | 0.1128 | 0.1608 | 0.0625 | 0.1013 |
| -0.005449 | 0.02747 | | | 19238 | 18382 | 0.09331 | 0.1013 | 0.01293 | 0.01303 | 0.1187 | 0.1269 | 0.06798 | 0.07581 |
| -0.8701 | -1.515 | | | 833 | 419 | 1 | 1 | 0 | 0 | 1 | 1 | 1 | 1 |
| 0.01119 | -0.1204 | | | 4142 | 4502 | 0.3632 | 0.3577 | 0.03422 | 0.03331 | 0.4303 | 0.423 | 0.2962 | 0.2924 |
| -1.967 | -0.6361 | | | 656 | 352 | 1 | 1 | 0 | 0 | 1 | 1 | 1 | 1 |
| 0.03958 | -0.08518 | | | 7132 | 8663 | 0.2528 | 0.1438 | 0.02367 | 0.03159 | 0.2992 | 0.2058 | 0.2064 | 0.08193 |
| -0.2601 | -0.1127 | | | 2818 | 2909 | 0.453 | 0.4263 | 0.0566 | 0.03037 | 0.5639 | 0.4858 | 0.342 | 0.3668 |
| 0.02599 | 0.03133 | flag | | 36195 | 27330 | 0.07141 | 0.08856 | 0.007617 | 0.01079 | 0.08634 | 0.1097 | 0.05648 | 0.06742 |
| -1.323 | -0.1926 | flag | | 587 | 506 | 1 | 1 | 0 | 0 | 1 | 1 | 1 | 1 |
| -1.62 | -0.4135 | | | 587 | 506 | 1 | 1 | 0 | 0 | 1 | 1 | 1 | 1 |
| -0.725 | -0.01125 | flag | | 1735 | 1604 | 0.5225 | 1 | 0.0741 | 0.0253 | 0.6678 | 1.05 | 0.3773 | 0.9504 |

| ldsc.h2.conf.low_female | ldsc.h2.conf.low_male | ldsc.int.flag_female | ldsc.int.flag_male | ldsc.N_female | ldsc.N_male | HDL.h2_female | HDL.h2_male | HDL.h2.se_female | HDL.h2.se_male | HDL.conf.high_female | HDL.conf.high_male | HDL.conf.low_female | HDL.conf.low_male |
|---|---|---|---|---|---|---|---|---|---|---|---|---|---|
| -0.3664 | -0.1644 | | flag | 1736 | 1604 | 0.869 | 1 | 0.07054 | 0 | 1.007 | 1 | 0.7308 | 1 |
| -0.5149 | -0.3187 | | | 1540 | 1362 | 1 | 1 | 0.003055 | 0.001211 | 1.006 | 1.002 | 0.994 | 0.9976 |
| -0.5803 | 0.1071 | flag | | 1736 | 1604 | 0.6406 | 1 | 0.07336 | 0.001413 | 0.7844 | 1.003 | 0.4968 | 0.9972 |
| -0.6547 | -0.6616 | | | 569 | 491 | 1 | 1 | 0 | 0 | 1 | 1 | 1 | 1 |
| -0.0927 | -0.2074 | | | 4438 | 4714 | 0.3005 | 0.2515 | 0.03116 | 0.05006 | 0.3616 | 0.3496 | 0.2394 | 0.1534 |
| -0.03162 | -0.06241 | | | 10326 | 9880 | 0.1338 | 0.1481 | 0.02136 | 0.02088 | 0.1757 | 0.189 | 0.09194 | 0.1072 |
| -1.146 | -0.129 | flag | | 586 | 506 | 1 | 1 | 0 | 0 | 1 | 1 | 1 | 1 |
| -1.023 | -0.3676 | | | 587 | 506 | 1 | 1 | 0 | 0 | 1 | 1 | 1 | 1 |
| -0.3891 | -0.2085 | | | 1734 | 1604 | 0.5411 | 0.9888 | 0.08764 | 0.04301 | 0.7129 | 1.073 | 0.3693 | 0.9045 |
| -0.3311 | -0.122 | | | 1735 | 1604 | 0.8576 | 1 | 0.08336 | 0 | 1.021 | 1 | 0.6942 | 1 |
| -0.3497 | -0.2671 | | | 1735 | 1604 | 0.6099 | 0.948 | 0.08754 | 0.05335 | 0.7815 | 1.053 | 0.4383 | 0.8434 |
| -0.2955 | -2.024 | | flag | 570 | 488 | 1 | 1 | 0 | 0 | 1 | 1 | 1 | 1 |
| 0.0432 | 0.04 | flag | | 35703 | 26981 | 0.09542 | 0.1219 | 0.007137 | 0.01186 | 0.1094 | 0.1451 | 0.08143 | 0.09862 |
| -0.1653 | -0.1252 | | | 3250 | 4659 | 0.4907 | 0.2695 | 0.03364 | 0.02826 | 0.5566 | 0.3248 | 0.4248 | 0.2141 |
| -0.1661 | -0.1281 | | | 4071 | 4442 | 0.3666 | 0.2722 | 0.04597 | 0.03279 | 0.4567 | 0.3365 | 0.2765 | 0.208 |
| -0.07509 | -0.03949 | | | 7210 | 8698 | 0.1652 | 0.1833 | 0.02008 | 0.02228 | 0.2045 | 0.227 | 0.1258 | 0.1397 |
| -0.163 | -0.2332 | | | 1988 | 1866 | 0.7263 | 0.7438 | 0.07726 | 0.04247 | 0.8777 | 0.827 | 0.5748 | 0.6606 |
| -0.09985 | -0.04049 | flag | | 7089 | 6106 | 0.2074 | 0.2404 | 0.03295 | 0.03276 | 0.272 | 0.3046 | 0.1428 | 0.1762 |
| 0.03459 | 0.02461 | | | 21460 | 19278 | 0.1178 | 0.1231 | 0.01585 | 0.01644 | 0.1489 | 0.1553 | 0.08678 | 0.09084 |
| -0.3654 | 0.1188 | | | 1008 | 529 | 1 | 1 | 0.02497 | 0 | 1.049 | 1 | 0.9511 | 1 |
| -0.1775 | -0.3142 | | flag | 3276 | 3583 | 0.444 | 0.2724 | 0.05111 | 0.03703 | 0.5442 | 0.345 | 0.3438 | 0.1998 |
| -1.635 | -0.9328 | | | 594 | 598 | 1 | 1 | 0 | 0 | 1 | 1 | 1 | 1 |
| -0.3541 | | | | 1116 | 16 | | | | | | | | |
| -0.07985 | -0.4648 | | | 2100 | 964 | 0.7447 | 1 | 0.06338 | 0 | 0.869 | 1 | 0.6205 | 1 |
| -0.1397 | -0.1377 | flag | | 1397 | 1537 | 1 | 1 | 0.0284 | 4.94e-04 | 1.056 | 1.001 | 0.9443 | 0.999 |
| | -0.04009 | | | | 9198 | | | | | | | | |
| -0.01082 | 0.006011 | | | 19329 | 18431 | 0.09079 | 0.08348 | 0.01457 | 0.01069 | 0.1193 | 0.1044 | 0.06223 | 0.06253 |
| -0.01604 | -0.02989 | | flag | 17241 | 16272 | 0.1067 | 0.09982 | 0.01365 | 0.0168 | 0.1335 | 0.1328 | 0.07997 | 0.06689 |
| -0.05367 | -0.02639 | | | 10228 | 9959 | 0.1464 | 0.1622 | 0.02097 | 0.01996 | 0.1875 | 0.2013 | 0.1053 | 0.1231 |
| -0.03247 | -0.01743 | | | 17346 | 11869 | 0.08269 | 0.1668 | 0.01384 | 0.02346 | 0.1098 | 0.2128 | 0.05556 | 0.1209 |
| 0.05868 | 0.05098 | | | 35247 | 26707 | 0.1015 | 0.1071 | 0.01028 | 0.01107 | 0.1216 | 0.1288 | 0.08136 | 0.08539 |
| 0.05961 | -2.99e-04 | | | 19242 | 12927 | 0.1238 | 0.1355 | 0.015 | 0.01995 | 0.1532 | 0.1746 | 0.0944 | 0.09635 |
| -0.03283 | -0.08176 | | | 7230 | 6313 | 0.2183 | 0.2152 | 0.01985 | 0.03401 | 0.2572 | 0.2819 | 0.1794 | 0.1486 |
| -0.7866 | -0.3441 | | | 892 | 747 | 1 | 1 | 0 | 0 | 1 | 1 | 1 | 1 |
| 0.04875 | -0.05664 | | | 7224 | 6316 | 0.2901 | 0.2816 | 0.0349 | 0.02456 | 0.3585 | 0.3297 | 0.2216 | 0.2335 |
| -0.03602 | -0.06213 | | | 7176 | 6237 | 0.2437 | 0.273 | 0.03095 | 0.03295 | 0.3043 | 0.3376 | 0.183 | 0.2084 |
| 0.03461 | 0.065 | | | 21446 | 19269 | 0.1332 | 0.1624 | 0.01637 | 0.0137 | 0.1653 | 0.1892 | 0.1012 | 0.1355 |
| -0.05284 | -0.1523 | | flag | 7225 | 6310 | 0.2225 | 0.186 | 0.03715 | 0.03106 | 0.2953 | 0.2469 | 0.1496 | 0.1252 |
| 0.03678 | 0.006978 | | flag | 21459 | 19274 | 0.1262 | 0.1219 | 0.01635 | 0.0143 | 0.1583 | 0.1499 | 0.09416 | 0.09387 |
| 0.02696 | 0.03973 | | | 21462 | 19277 | 0.1234 | 0.1267 | 0.01519 | 0.0147 | 0.1532 | 0.1555 | 0.09365 | 0.09791 |
| -0.202 | -0.1414 | flag | | 3383 | 1478 | 0.3455 | 1 | 0.03166 | 0 | 0.4075 | 1 | 0.2834 | 1 |
| -0.7724 | -0.5925 | | | 990 | 827 | 1 | 1 | 0 | 0 | 1 | 1 | 1 | 1 |

| ldsc.h2.conf.low_female | ldsc.h2.conf.low_male | ldsc.int.flag_female | ldsc.int.flag_male | ldsc.N_female | ldsc.N_male | HDL.h2_female | HDL.h2_male | HDL.h2.se_female | HDL.h2.se_male | HDL.conf.high_female | HDL.conf.high_male | HDL.conf.low_female | HDL.conf.low_male |
|---|---|---|---|---|---|---|---|---|---|---|---|---|---|
| -0.05865 | -0.007704 | | | 7225 | 6312 | 0.2023 | 0.2526 | 0.03605 | 0.03489 | 0.2729 | 0.3209 | 0.1316 | 0.1842 |
| -0.06283 | -0.03163 | | | 7224 | 6311 | 0.2091 | 0.248 | 0.02836 | 0.02842 | 0.2647 | 0.3037 | 0.1535 | 0.1923 |
| -0.2723 | -0.2506 | | | 1447 | 1197 | 1 | 0.8898 | 0 | 0.09519 | 1 | 1.076 | 1 | 0.7032 |

| ldsc.h2.conf.low_female | ldsc.h2.conf.low_male | ldsc.int.flag_female | ldsc.int.flag_male | ldsc.N_female | ldsc.N_male | HDL.h2_female | HDL.h2_male | HDL.h2.se_female | HDL.h2.se_male | HDL.conf.high_female | HDL.conf.high_male | HDL.conf.low_female | HDL.conf.low_male |
|---|---|---|---|---|---|---|---|---|---|---|---|---|---|
| -0.5554 | -1.841 | | | 838 | 639 | 1 | 1 | 0 | 0 | 1 | 1 | 1 | 1 |
| -1.016 | -1.303 | | | 838 | 639 | 1 | 1 | 0 | 0 | 1 | 1 | 1 | 1 |
| -0.07676 | -0.001672 | | | 6963 | 5423 | 0.2174 | 0.3167 | 0.02353 | 0.0341 | 0.2635 | 0.3835 | 0.1713 | 0.2498 |
| -0.1171 | -0.1443 | | flag | 7924 | 6029 | 0.1902 | 0.1949 | 0.02666 | 0.02819 | 0.2425 | 0.2502 | 0.138 | 0.1397 |
| -0.5599 | -0.1823 | | | 1248 | 931 | 1 | 1 | 0 | 0 | 1 | 1 | 1 | 1 |
| -0.5607 | -0.8241 | | | 719 | 675 | 1 | 1 | 0 | 0 | 1 | 1 | 1 | 1 |
| -0.03158 | -0.5994 | flag | | 1636 | 1304 | 1 | 0.9738 | 0 | 0.1214 | 1 | 1.212 | 1 | 0.7359 |
| -0.08488 | -0.08306 | | | 7336 | 5668 | 0.1885 | 0.2411 | 0.02197 | 0.03923 | 0.2315 | 0.318 | 0.1454 | 0.1642 |
| 0.4919 | -0.2971 | | | 878 | 1168 | 1 | 1 | 0 | 0 | 1 | 1 | 1 | 1 |
| -1.299 | -0.8738 | | | 574 | 537 | 1 | 1 | 0 | 0 | 1 | 1 | 1 | 1 |
| -0.08888 | -0.1606 | flag | flag | 7478 | 5736 | 0.1895 | 0.215 | 0.02217 | 0.0314 | 0.233 | 0.2766 | 0.1461 | 0.1535 |
| -0.06194 | -0.1051 | | | 7283 | 5626 | 0.1989 | 0.2509 | 0.01992 | 0.04104 | 0.2379 | 0.3313 | 0.1598 | 0.1705 |
| -0.4029 | -0.6827 | | flag | 1630 | 1261 | 0.7464 | 1 | 0.08301 | 0.05574 | 0.9092 | 1.109 | 0.5837 | 0.8908 |
| -0.01743 | -0.002228 | | | 17589 | 11789 | 0.08051 | 0.1234 | 0.01631 | 0.01746 | 0.1125 | 0.1576 | 0.04854 | 0.08916 |
| -0.8586 | -0.2381 | | | 512 | 631 | 1 | 1 | 0 | 0 | 1 | 1 | 1 | 1 |
| -0.2789 | -0.3317 | | | 1097 | 1001 | 1 | 1 | 0 | 0 | 1 | 1 | 1 | 1 |
| -0.3207 | -0.7899 | | | 1061 | 981 | 1 | 1 | 0 | 0 | 1 | 1 | 1 | 1 |
| -0.4057 | -0.7548 | | | 1100 | 1007 | 1 | 1 | 0 | 0 | 1 | 1 | 1 | 1 |
| -1.151 | -0.6207 | flag | | 1028 | 920 | 0.9736 | 1 | 0.07729 | 0 | 1.125 | 1 | 0.8221 | 1 |
| -0.7887 | -0.5693 | | | 522 | 569 | 1 | 1 | 0 | 0 | 1 | 1 | 1 | 1 |
| 0.06562 | 0.01561 | flag | | 9190 | 6986 | 0.2209 | 0.2652 | 0.02976 | 0.03388 | 0.2792 | 0.3316 | 0.1626 | 0.1988 |
| 0.2318 | 0.04896 | | flag | 19051 | 12623 | 0.2818 | 0.2353 | 0.02024 | 0.0254 | 0.3215 | 0.2851 | 0.2422 | 0.1855 |
| 0.2322 | 0.04899 | | flag | 19052 | 12624 | 0.2821 | 0.2353 | 0.02041 | 0.02526 | 0.322 | 0.2849 | 0.2421 | 0.1858 |
| 0.2322 | 0.04899 | | flag | 19052 | 12624 | 0.2821 | 0.2353 | 0.02041 | 0.02526 | 0.322 | 0.2849 | 0.2421 | 0.1858 |
| -0.08675 | 0.03427 | | | 5664 | 4501 | 0.2617 | 0.4177 | 0.03735 | 0.03009 | 0.3349 | 0.4767 | 0.1885 | 0.3587 |
| 0.02409 | | | | 5139 | | 0.3088 | | 0.04366 | | 0.3944 | | 0.2232 | |
| -0.194 | -0.4812 | | | 2715 | 1250 | 0.5301 | 0.9652 | 0.04669 | 0.06427 | 0.6216 | 1.091 | 0.4386 | 0.8393 |
| 0.03494 | 0.04076 | | | 21462 | 19277 | 0.1171 | 0.1237 | 0.01199 | 0.01517 | 0.1406 | 0.1534 | 0.09356 | 0.09396 |
| 0.03589 | 0.04936 | flag | | 35638 | 26954 | 0.08361 | 0.09841 | 0.009635 | 0.01081 | 0.1025 | 0.1196 | 0.06472 | 0.07723 |
| -0.2154 | -0.2758 | | | 2842 | 2084 | 0.5078 | 0.6681 | 0.06172 | 0.09259 | 0.6288 | 0.8496 | 0.3868 | 0.4866 |
| -0.05123 | -0.3036 | | | 3133 | 2143 | 0.5992 | 0.5607 | 0.02151 | 0.05614 | 0.6413 | 0.6708 | 0.557 | 0.4507 |
| 0.04436 | 0.0577 | flag | | 35661 | 26955 | 0.09723 | 0.1263 | 0.008492 | 0.01257 | 0.1139 | 0.1509 | 0.08058 | 0.1016 |
| 0.05275 | 0.04835 | | | 31506 | 24073 | 0.1037 | 0.1224 | 0.00947 | 0.00975 | 0.1222 | 0.1415 | 0.08512 | 0.1033 |
| -1.06 | -1.925 | | | 701 | 452 | 1 | 1 | 0 | 0 | 1 | 1 | 1 | 1 |
| -0.01042 | -0.01328 | | | 16703 | 11399 | 0.1044 | 0.1462 | 0.0143 | 0.01879 | 0.1324 | 0.183 | 0.07635 | 0.1094 |
| -0.0171 | -0.01332 | flag | | 30952 | 23979 | 0.04752 | 0.05623 | 0.008426 | 0.01147 | 0.06404 | 0.07871 | 0.03101 | 0.03374 |
| -0.0653 | -0.01525 | | flag | 3978 | 3555 | 0.4189 | 0.3805 | 0.03184 | 0.04412 | 0.4813 | 0.467 | 0.3565 | 0.294 |
| -0.09334 | -0.2134 | | | 2574 | 2403 | 0.602 | 0.6601 | 0.05063 | 0.05953 | 0.7012 | 0.7768 | 0.5027 | 0.5435 |
| -0.0425 | -0.07637 | | | 3964 | 3575 | 0.4389 | 0.4124 | 0.02722 | 0.04951 | 0.4923 | 0.5094 | 0.3856 | 0.3154 |
| -0.6468 | -2.214 | | flag | 1135 | 631 | 0.6979 | 1 | 0.1059 | 0.08283 | 0.9056 | 1.162 | 0.4903 | 0.8377 |
| 0.001831 | -0.1526 | | flag | 7229 | 6317 | 0.2204 | 0.1629 | 0.03975 | 0.02595 | 0.2983 | 0.2137 | 0.1425 | 0.112 |
| -0.2154 | -0.7772 | | | 1045 | 869 | 1 | 1 | 0 | 0 | 1 | 1 | 1 | 1 |
| -0.1622 | -0.4785 | | | 1046 | 868 | 1 | 1 | 0 | 0 | 1 | 1 | 1 | 1 |
| -0.8789 | -0.3358 | flag | | 1147 | 1145 | 0.9758 | 1 | 0.06895 | 0 | 1.111 | 1 | 0.8407 | 1 |
| -0.7591 | -0.4334 | flag | | 1143 | 1144 | 1 | 1 | 0.003438 | 0 | 1.007 | 1 | 0.9933 | 1 |
| -0.03744 | -1.035 | | | 1043 | 868 | 1 | 1 | 0 | 0 | 1 | 1 | 1 | 1 |
| -0.6007 | -0.9573 | | | 1046 | 869 | 1 | 1 | 0 | 0 | 1 | 1 | 1 | 1 |

| ldsc.h2.conf.low_female | ldsc.h2.conf.low_male | ldsc.int.flag_female | ldsc.int.flag_male | ldsc.N_female | ldsc.N_male | HDL.h2_female | HDL.h2_male | HDL.h2.se_female | HDL.h2.se_male | HDL.conf.high_female | HDL.conf.high_male | HDL.conf.low_female | HDL.conf.low_male |
|---|---|---|---|---|---|---|---|---|---|---|---|---|---|
| -0.6564 | 0.2683 | | | 1046 | 869 | 1 | 1 | 0 | 0 | 1 | 1 | 1 | 1 |
| -0.4285 | -0.5897 | | | 1147 | 1145 | 1 | 1 | 0 | 0.00436 | 1 | 1.009 | 1 | 0.9915 |
| -0.5437 | -0.3549 | | | 949 | 926 | 1 | 1 | 0 | 0 | 1 | 1 | 1 | 1 |
| -0.7386 | -0.9141 | | | 1046 | 869 | 1 | 1 | 0 | 0 | 1 | 1 | 1 | 1 |
| -0.4782 | -1.284 | | flag | 1518 | 535 | 0.7525 | 1 | 0.08315 | 0 | 0.9155 | 1 | 0.5896 | 1 |
| -0.02187 | -0.07003 | | | 7039 | 6224 | 0.2338 | 0.2854 | 0.03144 | 0.0312 | 0.2954 | 0.3465 | 0.1722 | 0.2242 |
| -0.1208 | 0.01132 | | flag | 772 | 237 | 1 | 1 | 0 | 0 | 1 | 1 | 1 | 1 |
| 0.02214 | 0.007286 | | | 13581 | 8621 | 0.1776 | 0.2031 | 0.02271 | 0.024 | 0.2221 | 0.2502 | 0.1331 | 0.1561 |
| 0.1089 | -0.02842 | | | 4968 | 3545 | 0.493 | 0.417 | 0.03938 | 0.05258 | 0.5702 | 0.52 | 0.4158 | 0.3139 |
| -0.9644 | -0.5102 | | flag | 439 | 581 | 1 | 1 | 0 | 0 | 1 | 1 | 1 | 1 |
| ldsc.h2.conf.low_female | ldsc.h2.conf.low_male | ldsc.int.flag_female | ldsc.int.flag_male | ldsc.N_female | ldsc.N_male | HDL.h2_female | HDL.h2_male | HDL.h2.se_female | HDL.h2.se_male | HDL.conf.high_female | HDL.conf.high_male | HDL.conf.low_female | HDL.conf.low_male |
| 0.02266 | -0.007205 | | | 20991 | 15836 | 0.1192 | 0.1179 | 0.0117 | 0.01454 | 0.1421 | 0.1464 | 0.09628 | 0.08941 |
| 0.03797 | 1.31e-04 | | | 21170 | 15997 | 0.1251 | 0.1248 | 0.009946 | 0.01387 | 0.1446 | 0.152 | 0.1056 | 0.09763 |
| -0.7519 | 0.5652 | | flag | 1087 | 325 | 1 | 1 | 0.05161 | 0 | 1.101 | 1 | 0.8989 | 1 |
| -0.9067 | -1.935 | | | 1003 | 313 | 0.9649 | 1 | 0.1474 | 0 | 1.254 | 1 | 0.6761 | 1 |
| -0.01271 | 0.001512 | | | 21462 | 19279 | 0.08149 | 0.1064 | 0.01096 | 0.0146 | 0.103 | 0.135 | 0.06001 | 0.07781 |
| -0.479 | -1.859 | | | 893 | 273 | 1 | 1 | 0 | 0 | 1 | 1 | 1 | 1 |
| -0.6984 | -0.3519 | | | 1130 | 549 | 1 | 1 | 0 | 0 | 1 | 1 | 1 | 1 |
| -1.183 | -0.5958 | | | 704 | 710 | 1 | 1 | 0 | 0 | 1 | 1 | 1 | 1 |
| -0.3292 | -0.1291 | | | 3137 | 3527 | 0.2761 | 0.4372 | 0.04648 | 0.05012 | 0.3672 | 0.5354 | 0.185 | 0.339 |
| -0.03179 | -0.5292 | | | 1587 | 1602 | 1 | 0.7817 | 8.94e-04 | 0.08805 | 1.002 | 0.9543 | 0.9982 | 0.6091 |
| -0.1073 | -0.025 | | flag | 3560 | 3467 | 0.3752 | 0.3476 | 0.04688 | 0.03877 | 0.467 | 0.4236 | 0.2833 | 0.2716 |
| -0.5352 | -0.2273 | | | 1351 | 1859 | 0.921 | 0.9041 | 0.07209 | 0.0686 | 1.062 | 1.039 | 0.7797 | 0.7696 |
| -0.003549 | 0.05844 | | | 8019 | 7600 | 0.2175 | 0.2826 | 0.02611 | 0.02668 | 0.2686 | 0.3349 | 0.1663 | 0.2303 |
| -0.3995 | -2.21 | flag | | 457 | 584 | 1 | 1 | 0 | 0 | 1 | 1 | 1 | 1 |
| 0.02915 | 0.003133 | | | 35815 | 26956 | 0.08587 | 0.08547 | 0.00735 | 0.009768 | 0.1003 | 0.1046 | 0.07146 | 0.06633 |
| -0.01039 | -0.02053 | | | 7147 | 6265 | 0.281 | 0.2932 | 0.0264 | 0.02598 | 0.3328 | 0.3441 | 0.2293 | 0.2423 |
| -0.09788 | -0.1322 | | | 7207 | 6303 | 0.208 | 0.2313 | 0.02444 | 0.03055 | 0.2559 | 0.2912 | 0.1601 | 0.1714 |
| -0.03099 | 0.009253 | flag | | 21462 | 19279 | 0.0651 | 0.1077 | 0.01138 | 0.01226 | 0.0874 | 0.1317 | 0.04279 | 0.08369 |
| -0.8789 | -2.152 | | | 1107 | 547 | 1 | 1 | 0.00432 | 0 | 1.008 | 1 | 0.9915 | 1 |
| 0.01664 | 0.01878 | flag | | 31936 | 24820 | 0.2339 | 0.2301 | 0.1691 | 0.1378 | 0.5653 | 0.5002 | -0.09746 | -0.03995 |
| -0.05535 | -0.1411 | | | 2113 | 2451 | 0.7183 | 0.536 | 0.05692 | 0.08252 | 0.8299 | 0.6977 | 0.6068 | 0.3742 |
| 0.008756 | 3.83e-04 | | | 32524 | 24959 | 0.06454 | 0.06833 | 0.008068 | 0.01089 | 0.08035 | 0.08968 | 0.04873 | 0.04698 |
| -0.01964 | -0.02851 | | | 17589 | 11789 | 0.08133 | 0.1237 | 0.01577 | 0.01914 | 0.1122 | 0.1613 | 0.05041 | 0.08622 |
| 0.02473 | 0.01969 | | | 21462 | 19279 | 0.1076 | 0.1097 | 0.01278 | 0.0136 | 0.1327 | 0.1363 | 0.08256 | 0.083 |
| -2.706 | -0.317 | flag | | 137 | 2215 | 1 | 0.5948 | 0 | 0.05753 | 1 | 0.7075 | 1 | 0.482 |
| -0.4427 | -0.03081 | | | 1067 | 3626 | 1 | 0.4516 | 0 | 0.03042 | 1 | 0.5112 | 1 | 0.392 |
| 0.2039 | -1.617 | | | 434 | 581 | 1 | 1 | 0 | 0 | 1 | 1 | 1 | 1 |
| -0.3677 | -0.6587 | | | 1229 | 696 | 1 | 1 | 0 | 0 | 1 | 1 | 1 | 1 |
| -0.2347 | 0.05316 | | flag | 4099 | 5720 | 0.2737 | 0.3542 | 0.03855 | 0.04076 | 0.3492 | 0.434 | 0.1981 | 0.2743 |
| -0.9295 | -0.8238 | | | 813 | 275 | 1 | 1 | 0 | 0 | 1 | 1 | 1 | 1 |
| -0.05486 | -0.1107 | | | 8759 | 6750 | 0.154 | 0.1758 | 0.02148 | 0.03133 | 0.1961 | 0.2372 | 0.1119 | 0.1144 |
| 0.08772 | -4.07 | flag | | 757 | 266 | 1 | 0.03061 | 0 | 2.534 | 1 | 4.998 | 1 | -4.937 |
| -0.1396 | -1.218 | | | 1045 | 869 | 1 | 1 | 0 | 0 | 1 | 1 | 1 | 1 |
| -0.07041 | -0.571 | | | 1046 | 868 | 1 | 1 | 0 | 0 | 1 | 1 | 1 | 1 |
| -1.172 | -0.7999 | flag | | 987 | 955 | 1 | 1 | 0 | 0 | 1 | 1 | 1 | 1 |

| ldsc.h2.conf.low_female | ldsc.h2.conf.low_male | ldsc.int.flag_female | ldsc.int.flag_male | ldsc.N_female | ldsc.N_male | HDL.h2_female | HDL.h2_male | HDL.h2.se_female | HDL.h2.se_male | HDL.conf.high_female | HDL.conf.high_male | HDL.conf.low_female | HDL.conf.low_male |
|---|---|---|---|---|---|---|---|---|---|---|---|---|---|
| -1.103 | -0.6823 | flag | | 987 | 954 | 1 | 1 | 0.001374 | 0 | 1.003 | 1 | 0.9973 | 1 |
| -0.1574 | -0.9575 | | | 1045 | 869 | 1 | 1 | 0 | 0 | 1 | 1 | 1 | 1 |
| -0.1108 | -0.1754 | | | 3739 | 2231 | 0.3969 | 0.6739 | 0.04472 | 0.03458 | 0.4845 | 0.7417 | 0.3092 | 0.6061 |
| 0.01903 | -0.05922 | flag | flag | 20773 | 15800 | 0.1072 | 0.08524 | 0.01215 | 0.01467 | 0.131 | 0.114 | 0.08339 | 0.05648 |
| 0.03601 | -0.002282 | | flag | 25624 | 19923 | 0.08444 | 0.1231 | 0.009507 | 0.01415 | 0.1031 | 0.1509 | 0.0658 | 0.09541 |
| 0.08859 | 0.06308 | | | 19112 | 15431 | 0.1925 | 0.1853 | 0.02168 | 0.02122 | 0.235 | 0.2269 | 0.15 | 0.1437 |
| -0.7363 | -0.4355 | | | 1070 | 1101 | 1 | 1 | 0 | 0 | 1 | 1 | 1 | 1 |
| -0.09953 | -0.2775 | | | 593 | 820 | 1 | 1 | 0 | 0 | 1 | 1 | 1 | 1 |
| -0.1927 | 0.08517 | | flag | 5855 | 7073 | 0.1976 | 0.2732 | 0.04208 | 0.02545 | 0.2801 | 0.3231 | 0.1151 | 0.2233 |
| -0.8519 | -1.51 | | | 831 | 325 | 1 | 1 | 0 | 0 | 1 | 1 | 1 | 1 |
| 0.03097 | 0.0368 | flag | | 26093 | 16812 | 0.109 | 0.151 | 0.01177 | 0.01541 | 0.1321 | 0.1812 | 0.08593 | 0.1208 |
| -0.2659 | -0.2753 | | | 3565 | 2286 | 0.3418 | 0.7704 | 0.04718 | 0.05513 | 0.4342 | 0.8785 | 0.2493 | 0.6624 |
| -0.4628 | -0.9689 | | flag | 820 | 294 | 1 | 1 | 0 | 0 | 1 | 1 | 1 | 1 |
| -0.1794 | -0.5006 | | | 1324 | 784 | 1 | 1 | 0 | 0 | 1 | 1 | 1 | 1 |
| -0.148 | -0.4982 | | | 1415 | 1069 | 1 | 1 | 0 | 0 | 1 | 1 | 1 | 1 |
| -0.2335 | -0.9435 | | | 585 | 723 | 1 | 1 | 0 | 0 | 1 | 1 | 1 | 1 |
| -0.3596 | -1.086 | | | 569 | 602 | 1 | 1 | 0 | 0 | 1 | 1 | 1 | 1 |
| -0.2172 | -0.1011 | | | 2253 | 2730 | 0.7184 | 0.483 | 0.06152 | 0.04674 | 0.839 | 0.5746 | 0.5978 | 0.3914 |
| -1.384 | 0.1065 | | | 489 | 673 | 1 | 1 | 0 | 0 | 1 | 1 | 1 | 1 |
| -0.1377 | -0.3072 | | | 3554 | 3258 | 0.3516 | 0.3573 | 0.02994 | 0.02638 | 0.4103 | 0.409 | 0.2929 | 0.3056 |
| -0.0277 | -0.3262 | | | 3704 | 2477 | 0.4095 | 0.4656 | 0.03513 | 0.07298 | 0.4783 | 0.6087 | 0.3406 | 0.3226 |
| 0.01893 | 0.02581 | | | 24121 | 18624 | 0.08265 | 0.1092 | 0.01029 | 0.01458 | 0.1028 | 0.1378 | 0.06247 | 0.08062 |
| -0.02835 | -0.01547 | | | 15324 | 11084 | 0.09265 | 0.1694 | 0.01711 | 0.0191 | 0.1262 | 0.2069 | 0.05912 | 0.132 |
| -0.002378 | -0.1122 | | | 6711 | 6411 | 0.36 | 0.2137 | 0.09899 | 0.03707 | 0.554 | 0.2864 | 0.166 | 0.1411 |
| -0.1544 | -0.4765 | | | 3025 | 2412 | 0.3416 | 0.526 | 0.02805 | 0.05735 | 0.3966 | 0.6384 | 0.2867 | 0.4136 |
| ldsc.h2.conf.low_female | ldsc.h2.conf.low_male | ldsc.int.flag_female | ldsc.int.flag_male | ldsc.N_female | ldsc.N_male | HDL.h2_female | HDL.h2_male | HDL.h2.se_female | HDL.h2.se_male | HDL.conf.high_female | HDL.conf.high_male | HDL.conf.low_female | HDL.conf.low_male |
| -0.03651 | -0.01242 | | | 9898 | 8932 | 0.1803 | 0.1789 | 0.0233 | 0.02301 | 0.226 | 0.224 | 0.1346 | 0.1338 |
| -0.02004 | -0.02255 | flag | flag | 24468 | 18338 | 0.06388 | 0.09702 | 0.01095 | 0.01051 | 0.08534 | 0.1176 | 0.04242 | 0.07643 |
| -0.01694 | -0.09542 | | | 14841 | 3944 | 0.1143 | 0.4037 | 0.01774 | 0.03968 | 0.1491 | 0.4815 | 0.07955 | 0.3259 |
| -0.6201 | -0.4907 | | | 869 | 808 | 1 | 1 | 0 | 0 | 1 | 1 | 1 | 1 |
| -0.08216 | -0.2108 | | | 4995 | 4911 | 0.2516 | 0.2334 | 0.03548 | 0.02428 | 0.3211 | 0.281 | 0.182 | 0.1858 |
| -0.02163 | -0.05149 | | | 16299 | 11007 | 0.08766 | 0.1515 | 0.01614 | 0.02269 | 0.1193 | 0.1959 | 0.05603 | 0.107 |
| -0.06933 | 0.2274 | | flag | 1227 | 1269 | 1 | 1 | 0 | 0 | 1 | 1 | 1 | 1 |
| -0.5797 | 0.05766 | flag | | 1226 | 1268 | 1 | 1 | 0 | 0 | 1 | 1 | 1 | 1 |
| -0.4612 | 0.1613 | flag | | 1226 | 1268 | 1 | 1 | 0 | 0 | 1 | 1 | 1 | 1 |
| -0.6519 | -0.06516 | flag | | 1222 | 1267 | 1 | 1 | 9.51e-05 | 0 | 1 | 1 | 0.9998 | 1 |
| -0.3299 | 0.06593 | | | 1227 | 1268 | 1 | 1 | 0 | 0 | 1 | 1 | 1 | 1 |
| -0.538 | 0.3006 | | flag | 1146 | 1145 | 1 | 1 | 0 | 0 | 1 | 1 | 1 | 1 |
| -0.8911 | -0.3375 | flag | | 1147 | 1144 | 0.9841 | 1 | 0.06053 | 0 | 1.103 | 1 | 0.8655 | 1 |
| -0.9337 | -0.3523 | flag | | 1147 | 1144 | 0.9656 | 1 | 0.08864 | 0 | 1.139 | 1 | 0.7919 | 1 |
| -0.6824 | 0.1487 | | flag | 1147 | 1145 | 1 | 1 | 0 | 0 | 1 | 1 | 1 | 1 |
| -0.05153 | -0.7849 | | | 2437 | 949 | 0.5893 | 0.9949 | 0.04839 | 0.1019 | 0.6841 | 1.195 | 0.4945 | 0.7951 |
| 0.01349 | -0.03726 | | | 14107 | 9165 | 0.1396 | 0.1573 | 0.01737 | 0.02712 | 0.1737 | 0.2104 | 0.1056 | 0.1041 |
| -0.4077 | -0.8193 | | | 1396 | 795 | 1 | 1 | 0.001097 | 0 | 1.002 | 1 | 0.9978 | 1 |
| 0.007015 | -0.4776 | | | 1801 | 836 | 0.9805 | 1 | 0.06067 | 0 | 1.099 | 1 | 0.8616 | 1 |
| -0.272 | 0.01126 | | | 1142 | 1012 | 1 | 1 | 0 | 0 | 1 | 1 | 1 | 1 |

| ldsc.h2.conf.low_female | ldsc.h2.conf.low_male | ldsc.int.flag_female | ldsc.int.flag_male | ldsc.N_female | ldsc.N_male | HDL.h2_female | HDL.h2_male | HDL.h2.se_female | HDL.h2.se_male | HDL.conf.high_female | HDL.conf.high_male | HDL.conf.low_female | HDL.conf.low_male |
|---|---|---|---|---|---|---|---|---|---|---|---|---|---|
| 0.04018 | 0.05403 | flag | | 35673 | 26968 | 0.09555 | 0.1254 | 0.008631 | 0.01084 | 0.1125 | 0.1466 | 0.07864 | 0.1041 |
| -0.08333 | -0.09857 | | | 5812 | 5932 | 0.2536 | 0.2413 | 0.03417 | 0.0293 | 0.3206 | 0.2987 | 0.1866 | 0.1839 |
| -0.01587 | 0.001462 | | | 4458 | 4826 | 0.3747 | 0.3379 | 0.04168 | 0.03554 | 0.4564 | 0.4076 | 0.293 | 0.2683 |
| -0.1258 | -0.3521 | | | 1253 | 1463 | 1 | 0.9463 | 0 | 0.09328 | 1 | 1.129 | 1 | 0.7634 |
| -0.3556 | -0.4287 | | | 906 | 1106 | 1 | 1 | 0 | 0 | 1 | 1 | 1 | 1 |
| 0.07911 | 0.08534 | | | 19541 | 12994 | 0.1618 | 0.2354 | 0.01416 | 0.02231 | 0.1896 | 0.2791 | 0.1341 | 0.1917 |
| 0.1208 | 0.1047 | | | 35804 | 26877 | 0.1502 | 0.1622 | 0.01163 | 0.01255 | 0.173 | 0.1868 | 0.1274 | 0.1376 |
| -0.1798 | -1.152 | | | 1051 | 681 | 1 | 1 | 0 | 0 | 1 | 1 | 1 | 1 |

#### STable12: Regression estimates from regressions of replicable sex difference trait-loci pairs (APOE and LDL cholesterol; uric acid and SLC2A9) with control for comorbidities and medication use

12 rows × 6 columns

| term | estimate | std.error | statistic | p.value | type |
|---|---|---|---|---|---|
| hardcalled_rs7412_T | -15.13 | 0.5732 | -26.39 | 7.15e-152 | Median LDL - ever statin - all participants |
| SEXmale:hardcalled_rs7412_T | 4.526 | 0.8631 | 5.243 | 1.59e-07 | Median LDL - ever statin - all participants |
| hardcalled_rs7412_T | -0.4824 | 0.0175 | -27.56 | 2.62e-165 | Median LDL inverse normalized within sex - ever statin - all participants |
| SEXmale:hardcalled_rs7412_T | 0.1553 | 0.02635 | 5.892 | 3.85e-09 | Median LDL inverse normalized within sex - ever statin - all participants |
| hardcalled_rs7412_T | -17.28 | 0.7212 | -23.96 | 2.80e-125 | First LDL - current statin - IHD cohort |
| SEXmale:hardcalled_rs7412_T | 5.324 | 1.17 | 4.549 | 5.42e-06 | First LDL - current statin - IHD cohort |
| hardcalled_rs1014290_G | -0.428 | 0.03478 | -12.3 | 1.33e-34 | Median uric acid - ever uric acid drugs - all participants |
| SEXmale:hardcalled_rs1014290_G | 0.2302 | 0.05049 | 4.56 | 5.15e-06 | Median uric acid - ever uric acid drugs - all participants |
| hardcalled_rs1014290_G | -0.2103 | 0.02794 | -7.53 | 5.89e-14 | Median uric acid inverse normalized within sex - ever uric acid drugs - all participants |
| SEXmale:hardcalled_rs1014290_G | 0.08449 | 0.03795 | 2.226 | 0.02605 | Median uric acid inverse normalized within sex - ever uric acid drugs - all participants |
| hardcalled_rs1014290_G | -0.39 | 0.03754 | -10.39 | 3.66e-25 | First uric acid - current uric acid drugs - gout cohort |
| SEXmale:hardcalled_rs1014290_G | 0.2241 | 0.05648 | 3.968 | 7.31e-05 | First uric acid - current uric acid drugs - gout cohort |

#### STable13: Causal mediation estimates from gout, uric acid, and SLC2A9 analysis

21 rows × 7 columns

| Estimate | Std.error | 95% CIL | 95% CIU | P.val | effect_type | Sex |
|---|---|---|---|---|---|---|
| 0.7445 | 0.05415 | 0.6522 | 0.8556 | 0 | Controlled direct effect | all |
| 0.7548 | 0.05238 | 0.6625 | 0.86 | 0 | Pure natural direct effect | all |
| 0.7542 | 0.05251 | 0.6622 | 0.8611 | 0 | Total natural direct effect | all |
| 0.9189 | 0.009752 | 0.9084 | 0.9451 | 0 | Pure natural indirect effect | all |
| 0.9182 | 0.01026 | 0.9062 | 0.9452 | 0 | Natural indirect effect | all |
| 0.693 | 0.04921 | 0.6119 | 0.7981 | 0 | Total effect | all |
| 0.2011 | 0.05364 | 0.1175 | 0.3282 | 0 | Proportion mediated | all |
| 0.7592 | 0.08345 | 0.5942 | 0.906 | 0 | Controlled direct effect | female |
| 0.7593 | 0.08325 | 0.5964 | 0.9064 | 0.01 | Pure natural direct effect | female |
| 0.761 | 0.08341 | 0.5955 | 0.9057 | 0.01 | Total natural direct effect | female |
| 0.9163 | 0.01501 | 0.8863 | 0.9433 | 0 | Pure natural indirect effect | female |
| 0.9185 | 0.01555 | 0.8834 | 0.9418 | 0 | Natural indirect effect | female |
| 0.6974 | 0.07756 | 0.5477 | 0.8321 | 0 | Total effect | female |
| 0.2045 | 0.1111 | 0.09867 | 0.427 | 0 | Proportion mediated | female |
| 0.7475 | 0.06299 | 0.6331 | 0.8631 | 0 | Controlled direct effect | male |
| 0.7582 | 0.06105 | 0.6452 | 0.8712 | 0 | Pure natural direct effect | male |
| 0.758 | 0.06119 | 0.6463 | 0.8705 | 0 | Total natural direct effect | male |
| 0.9499 | 0.01316 | 0.9256 | 0.9748 | 0.01 | Pure natural indirect effect | male |
| 0.9496 | 0.01348 | 0.9235 | 0.9735 | 0.01 | Natural indirect effect | male |
| 0.72 | 0.05909 | 0.6086 | 0.8238 | 0 | Total effect | male |
| 0.1366 | 0.05774 | 0.05482 | 0.2845 | 0.01 | Proportion mediated | male |

#### STable14: Causal mediation estimates from ischemic heart disease, LDL cholesterol, and APOE analysis

21 rows × 7 columns

| Estimate | Std.error | 95% CIL | 95% CIU | P.val | effect_type | Sex |
|---|---|---|---|---|---|---|
| 0.9769 | 0.04652 | 0.8977 | 1.073 | 0.68 | Controlled direct effect | all |
| 0.9778 | 0.04573 | 0.9003 | 1.073 | 0.68 | Pure natural direct effect | all |
| 0.9778 | 0.04586 | 0.8997 | 1.073 | 0.68 | Total natural direct effect | all |
| 0.9136 | 0.007746 | 0.8999 | 0.9287 | 0 | Pure natural indirect effect | all |
| 0.9136 | 0.007735 | 0.8999 | 0.9288 | 0 | Natural indirect effect | all |
| 0.8933 | 0.04133 | 0.8167 | 0.9843 | 0 | Total effect | all |
| 0.7919 | 1.534 | 0.4157 | 5.099 | 0 | Proportion mediated | all |
| 1.013 | 0.06891 | 0.8941 | 1.149 | 0.82 | Controlled direct effect | female |
| 1.013 | 0.06782 | 0.8953 | 1.147 | 0.82 | Pure natural direct effect | female |
| 1.013 | 0.06801 | 0.8948 | 1.147 | 0.82 | Total natural direct effect | female |
| 0.907 | 0.01279 | 0.8801 | 0.9299 | 0 | Pure natural indirect effect | female |
| 0.9071 | 0.01274 | 0.8798 | 0.9297 | 0 | Natural indirect effect | female |
| 0.9186 | 0.06093 | 0.8132 | 1.038 | 0.24 | Total effect | female |
| 1.157 | 80.1 | -17.25 | 10.56 | 0.24 | Proportion mediated | female |
| 0.9455 | 0.0644 | 0.831 | 1.064 | 0.41 | Controlled direct effect | male |
| 0.9467 | 0.06378 | 0.8326 | 1.063 | 0.41 | Pure natural direct effect | male |
| 0.9465 | 0.06382 | 0.8324 | 1.063 | 0.41 | Total natural direct effect | male |
| 0.9337 | 0.008793 | 0.915 | 0.9476 | 0 | Pure natural indirect effect | male |
| 0.9335 | 0.008814 | 0.9146 | 0.9474 | 0 | Natural indirect effect | male |
| 0.8838 | 0.05849 | 0.7774 | 0.9848 | 0.03 | Total effect | male |
| 0.5415 | 4.623 | 0.2103 | 3.76 | 0.03 | Proportion mediated | male |

### STable15: Ascertainment PheWAS of having a lab ever measured for vitamin D, LDL cholesterol, heart rate from EKG, and uric acid

*Notes: cells with blank regression estimates were not run due to low sample size in phewas*

7268 rows × 21 columns

| phenotype | male_beta | male_SE | male_OR | male_p | male_n_total | male_n_cases | male_n_controls | female_beta | female_SE | female_OR | female_p | female_n_total | female_n_cases | female_n_controls | interaction_p | interaction_beta | description | group | lab | label |
|---|---|---|---|---|---|---|---|---|---|---|---|---|---|---|---|---|---|---|---|---|
| 008 | 0.7471 | 0.1076 | 2.111 | 3.88e-12 | 27060 | 574 | 26486 | 0.5235 | 0.08636 | 1.688 | 1.34e-09 | 35831 | 704 | 35127 | 0.2385 | -0.1579 | Intestinal infection | infectious diseases | 25DTot | |
| 008.5 | | | | | 26875 | 389 | 26486 | | | | | 35533 | 406 | 35127 | 0.6817 | 0.07244 | Bacterial enteritis | infectious diseases | 25DTot | |
| 008.51 | | | | | 26487 | 1 | 26486 | | | | | 35130 | 3 | 35127 | | | Intestinal e.coli | infectious diseases | 25DTot | |
| 008.52 | | | | | 26824 | 338 | 26486 | | | | | 35475 | 348 | 35127 | 0.9023 | 0.02333 | Intestinal infection due to C. difficile | infectious diseases | 25DTot | |
| 008.6 | | | | | 26592 | 106 | 26486 | | | | | 35263 | 136 | 35127 | | | Viral Enteritis | infectious diseases | 25DTot | |
| 008.7 | | | | | 26495 | 9 | 26486 | | | | | 35133 | 6 | 35127 | | | Intestinal infection due to protozoa | infectious diseases | 25DTot | |
| 010 | | | | | 21692 | 51 | 21641 | | | | | 30214 | 54 | 30160 | | | Tuberculosis | infectious diseases | 25DTot | |
| 031 | | | | | 21701 | 60 | 21641 | | | | | 30222 | 62 | 30160 | | | Diseases due to other mycobacteria | infectious diseases | 25DTot | |
| 031.1 | | | | | 21641 | | | | | | | 30160 | | | | | Leprosy | infectious diseases | 25DTot | |
| 038 | 0.4696 | 0.05806 | 1.599 | 6.07e-16 | 24701 | 3060 | 21641 | 0.1834 | 0.0519 | 1.201 | 4.10e-04 | 32656 | 2496 | 30160 | 0.002308 | -0.2308 | Septicemia | infectious diseases | 25DTot | |
| 038.1 | | | | | 22136 | 495 | 21641 | | | | | 30550 | 390 | 30160 | 0.3417 | -0.1527 | Gram negative septicemia | infectious diseases | 25DTot | |
| 038.2 | | | | | 22055 | 414 | 21641 | | | | | 30433 | 273 | 30160 | 0.005365 | -0.5513 | Gram positive septicemia | infectious diseases | 25DTot | |
| 038.3 | 0.584 | 0.07847 | 1.793 | 9.90e-14 | 23021 | 1380 | 21641 | 0.2624 | 0.07752 | 1.3 | 7.13e-04 | 31182 | 1022 | 30160 | 0.006332 | -0.2924 | Bacteremia | infectious diseases | 25DTot | |
| 041 | 0.4878 | 0.05711 | 1.629 | 1.33e-17 | 24639 | 2998 | 21641 | 0.2268 | 0.04542 | 1.255 | 5.94e-07 | 33323 | 3163 | 30160 | 0.01127 | -0.1805 | Bacterial infection NOS | infectious diseases | 25DTot | |
| 041.1 | 0.5335 | 0.07966 | 1.705 | 2.11e-11 | 22993 | 1352 | 21641 | 0.1268 | 0.07738 | 1.135 | 0.1014 | 31259 | 1099 | 30160 | 6.86e-04 | -0.3672 | Staphylococcus infections | infectious diseases | 25DTot | |
| 041.11 | 0.5479 | 0.1168 | 1.73 | 2.72e-06 | 22231 | 590 | 21641 | | | | | 30598 | 438 | 30160 | 0.09778 | -0.2684 | Methicillin sensitive Staphylococcus aureus | infectious diseases | 25DTot | |
| 041.12 | 0.5752 | 0.1149 | 1.777 | 5.60e-07 | 22233 | 592 | 21641 | | | | | 30654 | 494 | 30160 | 0.007487 | -0.4214 | Methicillin resistant Staphylococcus aureus | infectious diseases | 25DTot | |
| 041.2 | 0.4631 | 0.1113 | 1.589 | 3.18e-05 | 22331 | 690 | 21641 | 0.3452 | 0.09228 | 1.412 | 1.83e-04 | 30829 | 669 | 30160 | 0.5715 | -0.07972 | Streptococcus infection | infectious diseases | 25DTot | |
| 041.21 | | | | | 21646 | 5 | 21641 | | | | | 30170 | 10 | 30160 | | | Rheumatic fever / chorea | infectious diseases | 25DTot | |
| 041.4 | | | | | 22126 | 485 | 21641 | 0.3749 | 0.07705 | 1.455 | 1.14e-06 | 31066 | 906 | 30160 | 0.7057 | -0.05505 | E. coli | infectious diseases | 25DTot | |
| 041.8 | | | | | 21691 | 50 | 21641 | | | | | 30238 | 78 | 30160 | | | H. pylori | infectious diseases | 25DTot | |
| 041.9 | | | | | 21990 | 349 | 21641 | | | | | 30526 | 366 | 30160 | 0.1963 | -0.231 | Infection with drug-resistant microorganisms | infectious diseases | 25DTot | |
| 053 | | | | | 22267 | 352 | 21915 | 0.775 | 0.09074 | 2.171 | 1.32e-17 | 29621 | 557 | 29064 | 0.08936 | 0.2759 | Herpes zoster | infectious diseases | 25DTot | |
| 053.1 | | | | | 21986 | 71 | 21915 | | | | | 29192 | 128 | 29064 | | | Herpes zoster with nervous system complications | infectious diseases | 25DTot | |
| 054 | | | | | 22206 | 291 | 21915 | 0.4984 | 0.09945 | 1.646 | 5.39e-07 | 29622 | 558 | 29064 | 0.7017 | -0.06965 | Herpes simplex | infectious diseases | 25DTot | |
| 070 | 0.4546 | 0.09449 | 1.576 | 1.50e-06 | 22942 | 1027 | 21915 | 0.1427 | 0.1057 | 1.153 | 0.1768 | 29665 | 601 | 29064 | 0.08952 | -0.2348 | Viral hepatitis | infectious diseases | 25DTot | |

| phenotype | male_beta | male_SE | male_OR | male_p | male_n_total | male_n_cases | male_n_controls | female_beta | female_SE | female_OR | female_p | female_n_total | female_n_cases | female_n_controls | interaction_p | interaction_beta | description | group | lab | label |
|---|---|---|---|---|---|---|---|---|---|---|---|---|---|---|---|---|---|---|---|---|
| 070.1 | | | | | 21931 | 16 | 21915 | | | | | 29069 | 5 | 29064 | | | Viral hepatitis A | infectious diseases | 25DTot | |
| 070.2 | | | | | 22087 | 172 | 21915 | | | | | 29138 | 74 | 29064 | | | Viral hepatitis B | infectious diseases | 25DTot | |
| 070.3 | 0.4464 | 0.1019 | 1.563 | 1.18e-05 | 22806 | 891 | 21915 | 0.07045 | 0.1171 | 1.073 | 0.5473 | 29579 | 515 | 29064 | 0.0476 | -0.3 | Viral hepatitis C | infectious diseases | 25DTot | |
| 070.4 | | | | | 22042 | 127 | 21915 | | | | | 29294 | 230 | 29064 | | | Chronic hepatitis | infectious diseases | 25DTot | |
| 070.9 | | | | | 22131 | 216 | 21915 | | | | | 29270 | 206 | 29064 | | | Hepatitis NOS | infectious diseases | 25DTot | |
| 071 | 0.1004 | 0.1333 | 1.106 | 0.4515 | 22529 | 614 | 21915 | | | | | 29177 | 113 | 29064 | 0.5737 | 0.1498 | Human immunodeficiency virus [HIV] disease | infectious diseases | 25DTot | |
| 071.1 | 0.1158 | 0.1334 | 1.123 | 0.3854 | 22521 | 606 | 21915 | | | | | 29170 | 106 | 29064 | 0.8547 | 0.05096 | HIV infection, symptomatic | infectious diseases | 25DTot | |
| 078 | 0.7733 | 0.09608 | 2.167 | 8.32e-16 | 22638 | 723 | 21915 | 0.5604 | 0.08241 | 1.751 | 1.04e-11 | 29855 | 791 | 29064 | 0.1941 | -0.1606 | Viral warts & HPV | infectious diseases | 25DTot | |
| 079 | 0.4704 | 0.1091 | 1.601 | 1.61e-05 | 22615 | 700 | 21915 | 0.5181 | 0.07839 | 1.679 | 3.84e-11 | 30057 | 993 | 29064 | 0.6098 | 0.06708 | Viral infection | infectious diseases | 25DTot | |
| 079.1 | | | | | 21924 | 9 | 21915 | | | | | 29075 | 11 | 29064 | | | Varicella infection | infectious diseases | 25DTot | |
| 079.2 | | | | | 21970 | 55 | 21915 | | | | | 29142 | 78 | 29064 | | | Infectious mononucleosis | infectious diseases | 25DTot | |
| 079.9 | | | | | 21977 | 62 | 21915 | | | | | 29164 | 100 | 29064 | | | Viremia, NOS | infectious diseases | 25DTot | |
| 080 | 0.2168 | 0.1233 | 1.242 | 0.07875 | 26108 | 681 | 25427 | 0.1553 | 0.09353 | 1.168 | 0.09673 | 35329 | 748 | 34581 | 0.9292 | 0.01343 | Postoperative infection | infectious diseases | 25DTot | |
| 081 | 0.4021 | 0.1077 | 1.495 | 1.88e-04 | 26193 | 766 | 25427 | 0.2019 | 0.09228 | 1.224 | 0.02869 | 35303 | 722 | 34581 | 0.4628 | -0.1014 | Infection/inflammation of internal prosthetic device; implant; and graft | infectious diseases | 25DTot | |
| 081.1 | | | | | 25468 | 41 | 25427 | | | | | 34621 | 40 | 34581 | | | Graft-versus-host disease | infectious diseases | 25DTot | |
| 081.11 | | | | | 25439 | 12 | 25427 | | | | | 34597 | 16 | 34581 | | | Acute graft-versus-host disease | infectious diseases | 25DTot | |
| 081.12 | | | | | 25458 | 31 | 25427 | | | | | 34611 | 30 | 34581 | | | Chronic graft-versus-host disease | infectious diseases | 25DTot | |
| 090 | | | | | 27678 | 201 | 27477 | | | | | 36786 | 64 | 36722 | | | Sexually transmitted infections (not HIV or hepatitis) | infectious diseases | 25DTot | |
| 090.2 | | | | | 27533 | 56 | 27477 | | | | | 36735 | 13 | 36722 | | | Gonococcal infections | infectious diseases | 25DTot | |
| 090.3 | | | | | 27479 | 2 | 27477 | | | | | 36731 | 9 | 36722 | | | Venereal diseases due to Chlamydia trachomatis | infectious diseases | 25DTot | |
| 110 | 0.7252 | 0.07075 | 2.065 | 1.17e-24 | 24822 | 1390 | 23432 | 0.6729 | 0.06291 | 1.96 | 1.05e-26 | 31711 | 1224 | 30487 | 0.5179 | -0.06026 | Dermatophytosis / Dermatomycosis | infectious diseases | 25DTot | |
| 110.1 | 0.759 | 0.07229 | 2.136 | 8.62e-26 | 24733 | 1301 | 23432 | 0.7139 | 0.06451 | 2.042 | 1.82e-28 | 31638 | 1151 | 30487 | 0.6238 | -0.0468 | Dermatophytosis | infectious diseases | 25DTot | |
| 110.11 | 0.8132 | 0.08066 | 2.255 | 6.63e-24 | 24428 | 996 | 23432 | 0.7727 | 0.07131 | 2.166 | 2.32e-27 | 31407 | 920 | 30487 | 0.6915 | -0.04206 | Dermatophytosis of nail | infectious diseases | 25DTot | |
| 110.12 | | | | | 23675 | 243 | 23432 | | | | | 30615 | 128 | 30487 | | | Althete's foot | infectious diseases | 25DTot | |
| 110.13 | | | | | 23580 | 148 | 23432 | | | | | 30581 | 94 | 30487 | | | Dermatophytosis of the body | infectious diseases | 25DTot | |
| 110.2 | | | | | 23511 | 79 | 23432 | | | | | 30547 | 60 | 30487 | | | Dermatomycoses | infectious diseases | 25DTot | |
| 112 | 0.6882 | 0.09675 | 1.99 | 1.13e-12 | 24237 | 805 | 23432 | 0.4918 | 0.05897 | 1.635 | 7.42e-17 | 32100 | 1613 | 30487 | 0.4686 | -0.08053 | Candidiasis | infectious diseases | 25DTot | |
| 112.3 | | | | | 23511 | 79 | 23432 | | | | | 30667 | 180 | 30487 | | | Candidiasis of skin and nails | infectious diseases | 25DTot | |
| 117 | | | | | 23791 | 359 | 23432 | | | | | 30831 | 344 | 30487 | 0.2273 | -0.2184 | Mycoses | infectious diseases | 25DTot | |

| phenotype | male_beta | male_SE | male_OR | male_p | male_n_total | male_n_cases | male_n_controls | female_beta | female_SE | female_OR | female_p | female_n_total | female_n_cases | female_n_controls | interaction_p | interaction_beta | description | group | lab | label |
|---|---|---|---|---|---|---|---|---|---|---|---|---|---|---|---|---|---|---|---|---|
| 117.1 | | | | | 23530 | 98 | 23432 | | | | | 30601 | 114 | 30487 | | | Histoplasmosis | infectious diseases | 25DTot | |
| 117.2 | | | | | 23434 | 2 | 23432 | | | | | 30487 | | | | | Coccidioidomycosis | infectious diseases | 25DTot | |
| 117.3 | | | | | 23448 | 16 | 23432 | | | | | 30500 | 13 | 30487 | | | Blastomycotic infection | infectious diseases | 25DTot | |
| 117.4 | | | | | 23501 | 69 | 23432 | | | | | 30544 | 57 | 30487 | | | Aspergillosis | infectious diseases | 25DTot | |
| 130 | | | | | 26354 | 10 | 26344 | | | | | 35233 | 9 | 35224 | | | Spirochetal infection | infectious diseases | 25DTot | |
| phenotype | male_beta | male_SE | male_OR | male_p | male_n_total | male_n_cases | male_n_controls | female_beta | female_SE | female_OR | female_p | female_n_total | female_n_cases | female_n_controls | interaction_p | interaction_beta | description | group | lab | label |
| 130.1 | | | | | 26349 | 5 | 26344 | | | | | 35227 | 3 | 35224 | | | Lyme disease | infectious diseases | 25DTot | |
| 131 | | | | | 26353 | 9 | 26344 | | | | | 35256 | 32 | 35224 | | | Protozoan infection | infectious diseases | 25DTot | |
| 132 | | | | | 26380 | 36 | 26344 | | | | | 35274 | 50 | 35224 | | | Infestation (lice, mites) | infectious diseases | 25DTot | |
| 132.1 | | | | | 26348 | 4 | 26344 | | | | | 35245 | 21 | 35224 | | | Pediculosis and phthirus infestation | infectious diseases | 25DTot | |
| 133 | | | | | 26352 | 8 | 26344 | | | | | 35226 | 2 | 35224 | | | Arthropod-borne diseases | infectious diseases | 25DTot | |
| 134 | | | | | 26349 | 5 | 26344 | | | | | 35248 | 24 | 35224 | | | Helminthiases | infectious diseases | 25DTot | |
| 134.1 | | | | | 26347 | 3 | 26344 | | | | | 35236 | 12 | 35224 | | | Intestinal helminthiases | infectious diseases | 25DTot | |
| 136 | | | | | 26733 | 389 | 26344 | | | | | 35599 | 375 | 35224 | 0.7164 | -0.0609 | Other infectious and parasitic diseases | infectious diseases | 25DTot | |
| 145 | -0.5192 | 0.175 | 0.595 | 0.003001 | 26903 | 624 | 26279 | | | | | 36280 | 243 | 36037 | 0.3343 | -0.2702 | Cancer of mouth | neoplasms | 25DTot | |
| 145.1 | | | | | 26299 | 20 | 26279 | | | | | 36044 | 7 | 36037 | | | Cancer of lip | neoplasms | 25DTot | |
| 145.2 | | | | | 26660 | 381 | 26279 | | | | | 36136 | 99 | 36037 | | | Cancer of tongue | neoplasms | 25DTot | |
| 145.3 | | | | | 26384 | 105 | 26279 | | | | | 36087 | 50 | 36037 | | | Cancer of major salivary glands | neoplasms | 25DTot | |
| 145.4 | | | | | 26326 | 47 | 26279 | | | | | 36068 | 31 | 36037 | | | Cancer of the gums | neoplasms | 25DTot | |
| 145.5 | | | | | 26338 | 59 | 26279 | | | | | 36058 | 21 | 36037 | | | Cancer of the mouth floor | neoplasms | 25DTot | |
| 149 | -0.7111 | 0.1574 | 0.4911 | 6.28e-06 | 27197 | 918 | 26279 | | | | | 36384 | 347 | 36037 | 0.2117 | -0.3214 | Cancer of larynx, pharynx, nasal cavities | neoplasms | 25DTot | |
| 149.1 | | | | | 26708 | 429 | 26279 | | | | | 36147 | 110 | 36037 | 0.5267 | -0.2492 | Cancer of oropharynx | neoplasms | 25DTot | |
| 149.2 | | | | | 26347 | 68 | 26279 | | | | | 36075 | 38 | 36037 | | | Cancer of nasopharynx | neoplasms | 25DTot | |
| 149.3 | | | | | 26333 | 54 | 26279 | | | | | 36064 | 27 | 36037 | | | Cancer of hypopharynx | neoplasms | 25DTot | |
| 149.4 | | | | | 26607 | 328 | 26279 | | | | | 36132 | 95 | 36037 | | | Cancer of larynx | neoplasms | 25DTot | |
| 149.5 | | | | | 26723 | 444 | 26279 | | | | | 36197 | 160 | 36037 | 0.2124 | -0.4414 | Hx of malignant neoplasm of oral cavity and pharynx | neoplasms | 25DTot | |
| 149.9 | | | | | 26377 | 98 | 26279 | | | | | 36109 | 72 | 36037 | | | Cancer of of nasal cavities | neoplasms | 25DTot | |
| 150 | | | | | 23056 | 399 | 22657 | | | | | 32735 | 70 | 32665 | | | Cancer of esophagus | neoplasms | 25DTot | |
| 151 | | | | | 23072 | 415 | 22657 | | | | | 32805 | 140 | 32665 | 0.361 | 0.3131 | Cancer of stomach | neoplasms | 25DTot | |
| 153 | -0.1668 | 0.1124 | 0.8463 | 0.1377 | 21472 | 1270 | 20202 | -0.304 | 0.1004 | 0.7378 | 0.002459 | 30905 | 948 | 29957 | 0.3188 | -0.1477 | Colorectal cancer | neoplasms | 25DTot | |
| 153.2 | -0.1733 | 0.1349 | 0.8409 | 0.1987 | 21102 | 900 | 20202 | -0.2705 | 0.1088 | 0.763 | 0.01288 | 30731 | 774 | 29957 | 0.641 | -0.07942 | Colon cancer | neoplasms | 25DTot | |
| 153.3 | -0.2259 | 0.1485 | 0.7978 | 0.1282 | 20940 | 738 | 20202 | | | | | 30403 | 446 | 29957 | 0.03131 | -0.4791 | Malignant neoplasm of rectum, rectosigmoid junction, and anus | neoplasms | 25DTot | |
| 155 | | | | | 23097 | 440 | 22657 | | | | | 32892 | 227 | 32665 | 0.1072 | -0.3745 | Cancer of liver and intrahepatic bile duct | neoplasms | 25DTot | |
| 155.1 | | | | | 23033 | 376 | 22657 | | | | | 32838 | 173 | 32665 | 0.2262 | -0.301 | Malignant neoplasm of liver, primary | neoplasms | 25DTot | |
| 157 | | | | | 23046 | 389 | 22657 | | | | | 32990 | 325 | 32665 | 0.0138 | -0.666 | Pancreatic cancer | neoplasms | 25DTot | |

| phenotype | male_beta | male_SE | male_OR | male_p | male_n_total | male_n_cases | male_n_controls | female_beta | female_SE | female_OR | female_p | female_n_total | female_n_cases | female_n_controls | interaction_p | interaction_beta | description | group | lab | label |
|---|---|---|---|---|---|---|---|---|---|---|---|---|---|---|---|---|---|---|---|---|
| 158 | | | | | 22882 | 225 | 22657 | | | | | 32904 | 239 | 32665 | | | Neoplasm of unspecified nature of digestive system | neoplasms | 25DTot | |
| 159 | -0.02049 | 0.1599 | 0.9797 | 0.8981 | 23177 | 520 | 22657 | -0.544 | 0.1448 | 0.5804 | 1.72e-04 | 33167 | 502 | 32665 | 0.03348 | -0.4497 | Malignant neoplasm of other and ill-defined sites within the digestive organs and peritoneum | neoplasms | 25DTot | |
| 159.2 | | | | | 22805 | 148 | 22657 | | | | | 32791 | 126 | 32665 | | | Malignant neoplasm of small intestine, including duodenum | neoplasms | 25DTot | |
| 159.3 | | | | | 22758 | 101 | 22657 | | | | | 32760 | 95 | 32665 | | | Malignant neoplasm of gallbladder and extrahepatic bile ducts | neoplasms | 25DTot | |
| 159.4 | | | | | 22743 | 86 | 22657 | | | | | 32783 | 118 | 32665 | | | Malignant neoplasm of retroperitoneum and peritoneum | neoplasms | 25DTot | |
| 164 | | | | | 25778 | 71 | 25707 | | | | | 35298 | 55 | 35243 | | | Cancer of intrathoracic organs | neoplasms | 25DTot | |
| 165 | -0.5287 | 0.1195 | 0.5894 | 9.59e-06 | 27233 | 1526 | 25707 | -0.4954 | 0.09765 | 0.6093 | 3.91e-07 | 36502 | 1259 | 35243 | 0.695 | -0.05931 | Cancer within the respiratory system | neoplasms | 25DTot | |
| 165.1 | -0.5346 | 0.1209 | 0.5859 | 9.79e-06 | 27209 | 1502 | 25707 | -0.5032 | 0.09833 | 0.6046 | 3.09e-07 | 36492 | 1249 | 35243 | 0.7039 | -0.05808 | Cancer of bronchus; lung | neoplasms | 25DTot | |
| 170 | -0.577 | 0.2009 | 0.5616 | 0.004075 | 27372 | 508 | 26864 | | | | | 36581 | 434 | 36147 | 0.5575 | -0.1516 | Cancer of bone and connective tissue | neoplasms | 25DTot | |
| 170.1 | | | | | 27106 | 242 | 26864 | | | | | 36353 | 206 | 36147 | | | Bone cancer | neoplasms | 25DTot | |
| 170.2 | | | | | 27193 | 329 | 26864 | | | | | 36433 | 286 | 36147 | 0.772 | 0.09879 | Cancer of connective tissue | neoplasms | 25DTot | |
| 172 | 0.4027 | 0.05643 | 1.496 | 9.58e-13 | 25415 | 3454 | 21961 | 0.3281 | 0.04768 | 1.388 | 5.90e-12 | 33247 | 2675 | 30572 | 0.4157 | -0.05879 | Skin cancer | neoplasms | 25DTot | |
| 172.1 | -0.06682 | 0.1186 | 0.9354 | 0.5731 | 22929 | 968 | 21961 | -0.08541 | 0.102 | 0.9181 | 0.4025 | 31295 | 723 | 30572 | 0.6785 | -0.06341 | Melanomas of skin, dx or hx | neoplasms | 25DTot | |
| 172.11 | -0.3332 | 0.1462 | 0.7166 | 0.02265 | 22766 | 805 | 21961 | -0.2337 | 0.1239 | 0.7916 | 0.05917 | 31138 | 566 | 30572 | 0.8799 | 0.02841 | Melanomas of skin | neoplasms | 25DTot | |
| 172.2 | 0.4971 | 0.05958 | 1.644 | 7.19e-17 | 24804 | 2843 | 21961 | 0.4244 | 0.05133 | 1.529 | 1.37e-16 | 32709 | 2137 | 30572 | 0.6093 | -0.03948 | Other non-epithelial cancer of skin | neoplasms | 25DTot | |
| 172.21 | 0.4799 | 0.08984 | 1.616 | 9.21e-08 | 22933 | 972 | 21961 | 0.489 | 0.08219 | 1.631 | 2.70e-09 | 31280 | 708 | 30572 | 0.8836 | -0.01758 | Basal cell carcinoma | neoplasms | 25DTot | |
| 172.22 | 0.6113 | 0.09452 | 1.843 | 9.95e-11 | 22797 | 836 | 21961 | | | | | 30972 | 400 | 30572 | 0.9652 | 0.006121 | Squamous cell carcinoma | neoplasms | 25DTot | |
| 172.3 | | | | | 22359 | 398 | 21961 | | | | | 30807 | 235 | 30572 | 0.3436 | -0.1747 | Carcinoma in situ of skin | neoplasms | 25DTot | |
| 173 | 0.5974 | 0.06451 | 1.817 | 2.02e-20 | 23987 | 2026 | 21961 | 0.5809 | 0.04806 | 1.788 | 1.24e-33 | 32915 | 2343 | 30572 | 0.4314 | -0.06207 | Neoplasm of uncertain behavior of skin | neoplasms | 25DTot | |
| 174 | | | | | 27350 | 45 | 27305 | -0.138 | 0.04956 | 0.8711 | 0.005377 | 31827 | 3093 | 28734 | 0.7428 | 0.1728 | Breast cancer | neoplasms | 25DTot | |
| 174.1 | | | | | 7 | | | -0.1617 | 0.05067 | 0.8507 | 0.00142 | 31729 | 2998 | 28731 | 0.9995 | -0.2235 | Breast cancer [female] | neoplasms | 25DTot | |
| 174.11 | | | | | 7 | | | -0.1655 | 0.05095 | 0.8474 | 0.001158 | 31698 | 2967 | 28731 | 0.9995 | -0.2254 | Malignant neoplasm of female breast | neoplasms | 25DTot | |
| 174.2 | | | | | 27313 | 15 | 27298 | | | | | 3 | | | | | Breast cancer [male] | neoplasms | 25DTot | |
| 174.3 | | | | | 27310 | 5 | 27305 | | | | | 28889 | 155 | 28734 | | | Neoplasm of uncertain behavior of breast | neoplasms | 25DTot | |
| 175 | | | | | 7 | | | -0.0891 | 0.07574 | 0.9148 | 0.2394 | 27358 | 1237 | 26121 | 0.9994 | -0.2637 | Acquired absence of breast | neoplasms | 25DTot | |
| 180 | | | | | 7 | | | 0.07151 | 0.08788 | 1.074 | 0.4158 | 28736 | 897 | 27839 | 0.9997 | -0.1961 | Cervical cancer and dysplasia | neoplasms | 25DTot | |
| 180.1 | | | | | 7 | | | | | | | 28134 | 295 | 27839 | | | Cervical cancer | neoplasms | 25DTot | |
| 180.3 | | | | | 7 | | | 0.281 | 0.1037 | 1.324 | 0.006725 | 28457 | 618 | 27839 | 0.9999 | -0.1027 | Cervical intraepithelial neoplasia [CIN] [Cervical dysplasia] | neoplasms | 25DTot | |
| 182 | | | | | 7 | | | -0.3396 | 0.1216 | 0.7121 | 0.005215 | 29788 | 534 | 29254 | 0.9995 | -0.3399 | Malignant neoplasm of uterus | neoplasms | 25DTot | |
| 184 | | | | | 7 | | | -0.3249 | 0.1038 | 0.7226 | 0.001748 | 30877 | 771 | 30106 | 0.9996 | -0.2643 | Cancer of other female genital organs | neoplasms | 25DTot | |
| 184.1 | | | | | 7 | | | -0.4068 | 0.1245 | 0.6658 | 0.001083 | 30679 | 573 | 30106 | 0.9996 | -0.314 | Malignant neoplasm of ovary and other uterine adnexa | neoplasms | 25DTot | |

| phenotype | male_beta | male_SE | male_OR | male_p | male_n_total | male_n_cases | male_n_controls | female_beta | female_SE | female_OR | female_p | female_n_total | female_n_cases | female_n_controls | interaction_p | interaction_beta | description | group | lab | label |
|---|---|---|---|---|---|---|---|---|---|---|---|---|---|---|---|---|---|---|---|---|
| 184.11 | | | | | 7 | | | | | | | 30568 | 462 | 30106 | | | Malignant neoplasm of ovary | neoplasms | 25DTot | |
| 184.2 | | | | | 7 | | | | | | | 30333 | 227 | 30106 | | | Cancer of other female genital organs (excluding uterus and ovary) | neoplasms | 25DTot | |
| 185 | 0.08596 | 0.08671 | 1.09 | 0.3215 | 21441 | 2053 | 19388 | | | | | 4 | | | | | Cancer of prostate | neoplasms | 25DTot | |
| 187 | | | | | 25045 | 181 | 24864 | | | | | 4 | | | | | Cancer of other male genital organs | neoplasms | 25DTot | |
| 187.1 | | | | | 24907 | 43 | 24864 | | | | | 4 | | | | | Malignant neoplasm of unspecified male genital organ | neoplasms | 25DTot | |
| 187.2 | | | | | 24988 | 124 | 24864 | | | | | 4 | | | | | Malignant neoplasm of testis | neoplasms | 25DTot | |
| 187.8 | | | | | 24880 | 16 | 24864 | | | | | 4 | | | | | Neoplasm of uncertain behavior of male genital organs | neoplasms | 25DTot | |
| 189 | -0.08229 | 0.09891 | 0.921 | 0.4054 | 27414 | 1343 | 26071 | -0.2859 | 0.1167 | 0.7513 | 0.01428 | 36667 | 553 | 36114 | 0.409 | -0.1224 | Cancer of urinary organs (incl. kidney and bladder) | neoplasms | 25DTot | |
| 189.1 | -0.01166 | 0.1293 | 0.9884 | 0.9281 | 26801 | 730 | 26071 | | | | | 36472 | 358 | 36114 | 0.4429 | -0.1444 | Cancer of kidney and renal pelvis | neoplasms | 25DTot | |
| 189.11 | 0.02509 | 0.1371 | 1.025 | 0.8548 | 26706 | 635 | 26071 | | | | | 36422 | 308 | 36114 | 0.4236 | -0.1611 | Malignant neoplasm of kidney, except pelvis | neoplasms | 25DTot | |
| 189.12 | | | | | 26217 | 146 | 26071 | | | | | 36178 | 64 | 36114 | | | Malignant neoplasm of renal pelvis | neoplasms | 25DTot | |
| 189.2 | -0.1732 | 0.1364 | 0.841 | 0.2041 | 26790 | 719 | 26071 | | | | | 36340 | 226 | 36114 | 0.8218 | -0.04909 | Cancer of bladder | neoplasms | 25DTot | |
| 189.21 | -0.1888 | 0.1384 | 0.8279 | 0.1724 | 26780 | 709 | 26071 | | | | | 36336 | 222 | 36114 | 0.9735 | -0.007299 | Malignant neoplasm of bladder | neoplasms | 25DTot | |
| 189.4 | | | | | 26367 | 296 | 26071 | | | | | 36227 | 113 | 36114 | | | Malignant neoplasm of other urinary organs | neoplasms | 25DTot | |
| 190 | | | | | 26618 | 60 | 26558 | | | | | 35577 | 58 | 35519 | | | Cancer of eye | neoplasms | 25DTot | |
| 191 | | | | | 27023 | 465 | 26558 | | | | | 35973 | 454 | 35519 | 0.2638 | -0.2629 | Manlignant and unknown neoplasms of brain and nervous system | neoplasms | 25DTot | |
| 191.1 | | | | | 26929 | 371 | 26558 | | | | | 35854 | 335 | 35519 | 0.1526 | -0.4082 | Cancer of brain and nervous system | neoplasms | 25DTot | |
| 191.11 | | | | | 26893 | 335 | 26558 | | | | | 35801 | 282 | 35519 | 0.3547 | -0.292 | Cancer of brain | neoplasms | 25DTot | |
| 193 | | | | | 27161 | 215 | 26946 | | | | | 35853 | 482 | 35371 | 0.2317 | -0.2423 | Thyroid cancer | neoplasms | 25DTot | |
| 194 | | | | | 26993 | 47 | 26946 | | | | | 35425 | 54 | 35371 | | | Cancer of other endocrine glands | neoplasms | 25DTot | |
| 195 | -0.2581 | 0.08338 | 0.7725 | 0.001965 | 20735 | 2450 | 18285 | -0.4401 | 0.07265 | 0.644 | 1.39e-09 | 28047 | 1903 | 26144 | 0.2 | -0.1393 | Cancer, suspected or other | neoplasms | 25DTot | |
| 195.1 | -0.3523 | 0.1178 | 0.7031 | 0.002776 | 19494 | 1209 | 18285 | -0.4691 | 0.1007 | 0.6255 | 3.17e-06 | 27102 | 958 | 26144 | 0.5649 | -0.08793 | Malignant neoplasm, other | neoplasms | 25DTot | |
| 195.3 | -0.6843 | 0.1616 | 0.5045 | 2.29e-05 | 19155 | 870 | 18285 | | | | | 26453 | 309 | 26144 | 0.9781 | 0.006847 | Malignant neoplasm of head, face, and neck | neoplasms | 25DTot | |
| 196 | 0.03696 | 0.09126 | 1.038 | 0.6855 | 19459 | 1174 | 18285 | 0.08705 | 0.06422 | 1.091 | 0.1752 | 27555 | 1411 | 26144 | 0.6715 | 0.04656 | Radiotherapy | neoplasms | 25DTot | |
| 197 | -0.2619 | 0.06193 | 0.7696 | 2.35e-05 | 22676 | 4391 | 18285 | -0.2122 | 0.04219 | 0.8088 | 4.92e-07 | 30992 | 4848 | 26144 | 0.05337 | 0.142 | Chemotherapy | neoplasms | 25DTot | |
| 198 | -0.4594 | 0.07909 | 0.6317 | 6.29e-09 | 22039 | 3754 | 18285 | -0.5609 | 0.06102 | 0.5707 | 3.83e-20 | 29550 | 3406 | 26144 | 0.5663 | -0.05644 | Secondary malignant neoplasm | neoplasms | 25DTot | |
| 198.1 | -0.5849 | 0.1071 | 0.5571 | 4.76e-08 | 20292 | 2007 | 18285 | -0.5169 | 0.08048 | 0.5963 | 1.34e-10 | 27825 | 1681 | 26144 | 0.2873 | 0.1407 | Secondary malignancy of lymph nodes | neoplasms | 25DTot | |
| 198.2 | -0.5912 | 0.1376 | 0.5536 | 1.73e-05 | 19626 | 1341 | 18285 | -0.883 | 0.1205 | 0.4136 | 2.35e-13 | 27296 | 1152 | 26144 | 0.1371 | -0.2691 | Secondary malignancy of respiratory organs | neoplasms | 25DTot | |
| 198.3 | -0.6347 | 0.2121 | 0.5301 | 0.002763 | 18851 | 566 | 18285 | -0.5596 | 0.1439 | 0.5714 | 1.01e-04 | 26726 | 582 | 26144 | 0.5151 | 0.1651 | Secondary malignant neoplasm of digestive systems | neoplasms | 25DTot | |
| 198.4 | -0.371 | 0.1445 | 0.69 | 0.01023 | 19347 | 1062 | 18285 | -0.6602 | 0.1209 | 0.5168 | 4.77e-08 | 27121 | 977 | 26144 | 0.2457 | -0.216 | Secondary malignant neoplasm of liver | neoplasms | 25DTot | |
| 198.5 | -0.3844 | 0.2017 | 0.6809 | 0.05674 | 18799 | 514 | 18285 | -0.8446 | 0.1677 | 0.4297 | 4.71e-07 | 26737 | 593 | 26144 | 0.08276 | -0.4504 | Secondary malignancy of brain/spine | neoplasms | 25DTot | |
| 198.6 | -0.3858 | 0.1409 | 0.6799 | 0.006171 | 19284 | 999 | 18285 | -0.7263 | 0.1291 | 0.4837 | 1.86e-08 | 26962 | 818 | 26144 | 0.07609 | -0.3346 | Secondary malignancy of bone | neoplasms | 25DTot | |

| phenotype | male_beta | male_SE | male_OR | male_p | male_n_total | male_n_cases | male_n_controls | female_beta | female_SE | female_OR | female_p | female_n_total | female_n_cases | female_n_controls | interaction_p | interaction_beta | description | group | lab | label |
|---|---|---|---|---|---|---|---|---|---|---|---|---|---|---|---|---|---|---|---|---|
| 198.7 | | | | | 18508 | 223 | 18285 | | | | | 26369 | 225 | 26144 | | | Secondary malignant neoplasm of skin | neoplasms | 25DTot | |
| 199 | 0.3494 | 0.08035 | 1.418 | 1.37e-05 | 19599 | 1314 | 18285 | 0.1561 | 0.06191 | 1.169 | 0.01167 | 27705 | 1561 | 26144 | 0.05361 | -0.1918 | Neoplasm of uncertain behavior | neoplasms | 25DTot | |
| 199.4 | | | | | 18315 | 30 | 18285 | | | | | 26186 | 42 | 26144 | | | Neurofibromatosis | neoplasms | 25DTot | |
| 200 | 0.1269 | 0.1285 | 1.135 | 0.3237 | 26538 | 642 | 25896 | -0.1207 | 0.1101 | 0.8863 | 0.273 | 35761 | 577 | 35184 | 0.1642 | -0.2283 | Myeloproliferative disease | neoplasms | 25DTot | |
| 200.1 | | | | | 21974 | 134 | 21840 | | | | | 30355 | 101 | 30254 | | | Polycythemia vera | neoplasms | 25DTot | |
| 201 | | | | | 25974 | 78 | 25896 | | | | | 35242 | 58 | 35184 | | | Hodgkin's disease | neoplasms | 25DTot | |
| 202 | | | | | 26283 | 387 | 25896 | | | | | 35476 | 292 | 35184 | 0.1195 | -0.3375 | Cancer of other lymphoid, histiocytic tissue | neoplasms | 25DTot | |
| 202.2 | | | | | 26223 | 327 | 25896 | | | | | 35425 | 241 | 35184 | 0.1221 | -0.3659 | Non-Hodgkins lymphoma | neoplasms | 25DTot | |
| 202.21 | | | | | 25957 | 61 | 25896 | | | | | 35235 | 51 | 35184 | | | Nodular lymphoma | neoplasms | 25DTot | |
| 202.22 | | | | | 25981 | 85 | 25896 | | | | | 35237 | 53 | 35184 | | | Reticulosarcoma | neoplasms | 25DTot | |
| 202.23 | | | | | 25947 | 51 | 25896 | | | | | 35208 | 24 | 35184 | | | Lymphosarcoma | neoplasms | 25DTot | |
| 202.24 | | | | | 25960 | 64 | 25896 | | | | | 35217 | 33 | 35184 | | | Large cell lymphoma | neoplasms | 25DTot | |
| 204 | -0.2906 | 0.1465 | 0.7478 | 0.04734 | 26591 | 695 | 25896 | -0.3412 | 0.1293 | 0.7109 | 0.008322 | 35705 | 521 | 35184 | 0.6486 | -0.08694 | Leukemia | neoplasms | 25DTot | |
| 204.1 | | | | | 26113 | 217 | 25896 | | | | | 35327 | 143 | 35184 | | | Lymphoid leukemia | neoplasms | 25DTot | |
| 204.11 | | | | | 25996 | 100 | 25896 | | | | | 35260 | 76 | 35184 | | | Lymphoid leukemia, acute | neoplasms | 25DTot | |
| 204.12 | | | | | 26033 | 137 | 25896 | | | | | 35270 | 86 | 35184 | | | Lymphoid leukemia, chronic | neoplasms | 25DTot | |
| 204.2 | | | | | 26331 | 435 | 25896 | | | | | 35537 | 353 | 35184 | 0.5526 | -0.1492 | Myeloid leukemia | neoplasms | 25DTot | |
| 204.21 | | | | | 26263 | 367 | 25896 | | | | | 35482 | 298 | 35184 | 0.3667 | -0.2479 | Myeloid leukemia, acute | neoplasms | 25DTot | |
| 204.22 | | | | | 26069 | 173 | 25896 | | | | | 35327 | 143 | 35184 | | | Myeloid leukemia, chronic | neoplasms | 25DTot | |
| 204.3 | | | | | 25933 | 37 | 25896 | | | | | 35214 | 30 | 35184 | | | Monocytic leukemia | neoplasms | 25DTot | |
| 204.4 | | | | | 26060 | 164 | 25896 | | | | | 35291 | 107 | 35184 | | | Multiple myeloma | neoplasms | 25DTot | |
| 208 | 0.5275 | 0.05758 | 1.695 | 5.16e-20 | 24481 | 2858 | 21623 | 0.6212 | 0.04615 | 1.861 | 2.64e-41 | 33540 | 2573 | 30967 | 0.3896 | 0.06265 | Benign neoplasm of colon | neoplasms | 25DTot | |
| 209 | | | | | 27760 | 86 | 27674 | | | | | 36911 | 107 | 36804 | | | Neuroendocrine tumors | neoplasms | 25DTot | |
| 210 | | | | | 26386 | 107 | 26279 | | | | | 36167 | 130 | 36037 | | | Benign neoplasm of lip, oral cavity, and pharynx | neoplasms | 25DTot | |
| 211 | | | | | 24095 | 236 | 23859 | | | | | 33779 | 379 | 33400 | 0.8479 | 0.03946 | Benign neoplasm of other parts of digestive system | neoplasms | 25DTot | |
| 212 | | | | | 27637 | 101 | 27536 | | | | | 36801 | 112 | 36689 | | | Benign neoplasm of respiratory and intrathoracic organs | neoplasms | 25DTot | |
| 213 | | | | | 21553 | 68 | 21485 | | | | | 26105 | 131 | 25974 | | | Benign neoplasm of bone and articular cartilage | neoplasms | 25DTot | |
| 214 | | | | | 27025 | 277 | 26748 | | | | | 35990 | 359 | 35631 | 0.7043 | -0.06994 | Lipoma | neoplasms | 25DTot | |
| 214.1 | | | | | 26919 | 171 | 26748 | | | | | 35867 | 236 | 35631 | | | Lipoma of skin and subcutaneous tissue | neoplasms | 25DTot | |
| 215 | | | | | 26820 | 72 | 26748 | | | | | 35793 | 162 | 35631 | | | Other benign neoplasm of connective and other soft tissue | neoplasms | 25DTot | |
| 216 | 0.455 | 0.06996 | 1.576 | 7.87e-11 | 25936 | 1607 | 24329 | 0.6895 | 0.04589 | 1.993 | 4.96e-51 | 34007 | 2621 | 31386 | 0.04735 | 0.1631 | Benign neoplasm of skin | neoplasms | 25DTot | |
| 216.1 | | | | | 24408 | 79 | 24329 | | | | | 31484 | 98 | 31386 | | | Screening for malignant neoplasms of the skin | neoplasms | 25DTot | |
| 217 | | | | | 24477 | 148 | 24329 | | | | | 31698 | 312 | 31386 | | | Vascular hamartomas and non-neoplastic nevi | neoplasms | 25DTot | |
| 217.1 | | | | | 24444 | 115 | 24329 | | | | | 31637 | 251 | 31386 | | | Nevus, non-neoplastic | neoplasms | 25DTot | |
| 218 | | | | | 7 | | | 0.3274 | 0.0793 | 1.387 | 3.66e-05 | 31913 | 869 | 31044 | 0.9999 | -0.05367 | Benign neoplasm of uterus | neoplasms | 25DTot | |
| 218.1 | | | | | 7 | | | 0.3406 | 0.08116 | 1.406 | 2.71e-05 | 31868 | 824 | 31044 | 0.9999 | -0.04522 | Uterine leiomyoma | neoplasms | 25DTot | |
| 218.2 | | | | | 7 | | | | | | | 31064 | 20 | 31044 | | | Other benign neoplasm of uterus | neoplasms | 25DTot | |
| 220 | | | | | 7 | | | | | | | 28308 | 209 | 28099 | | | Benign neoplasm of ovary | neoplasms | 25DTot | |

| phenotype | male_beta | male_SE | male_OR | male_p | male_n_total | male_n_cases | male_n_controls | female_beta | female_SE | female_OR | female_p | female_n_total | female_n_cases | female_n_controls | interaction_p | interaction_beta | description | group | lab | label |
|---|---|---|---|---|---|---|---|---|---|---|---|---|---|---|---|---|---|---|---|---|
| 221 | | | | | 7 | | | | | | | 28086 | 59 | 28027 | | | Benign neoplasm of other female genital organs | neoplasms | 25DTot | |
| 222 | | | | | 24885 | 21 | 24864 | | | | | 4 | | | | | Benign neoplasm of male genital organs | neoplasms | 25DTot | |
| 223 | | | | | 21509 | 24 | 21485 | | | | | 26009 | 35 | 25974 | | | Benign neoplasm of kidney and other urinary organs | neoplasms | 25DTot | |
| 224 | | | | | 26668 | 110 | 26558 | | | | | 35659 | 140 | 35519 | | | Benign neoplasm of eye | neoplasms | 25DTot | |
| 224.1 | | | | | 26643 | 85 | 26558 | | | | | 35621 | 102 | 35519 | | | Benign neoplasm of eye, uveal | neoplasms | 25DTot | |
| 225 | | | | | 26749 | 191 | 26558 | | | | | 35909 | 390 | 35519 | 0.4425 | -0.1976 | Benign neoplasm of brain and other parts of nervous system | neoplasms | 25DTot | |
| 225.1 | | | | | 26730 | 172 | 26558 | | | | | 35878 | 359 | 35519 | 0.4333 | -0.215 | Benign neoplasm of brain, cranial nerves, meninges | neoplasms | 25DTot | |
| 225.2 | | | | | 26574 | 16 | 26558 | | | | | 35551 | 32 | 35519 | | | Benign neoplasm of spinal cord, meninges | neoplasms | 25DTot | |
| 226 | | | | | 26971 | 25 | 26946 | | | | | 35500 | 129 | 35371 | | | Benign neoplasm of thyroid glands | neoplasms | 25DTot | |
| 227 | | | | | 27177 | 231 | 26946 | | | | | 35717 | 346 | 35371 | 0.2353 | -0.2332 | Benign neoplasm of other endocrine glands and related structures | neoplasms | 25DTot | |
| 227.1 | | | | | 27032 | 86 | 26946 | | | | | 35495 | 124 | 35371 | | | Benign neoplasm of adrenal gland | neoplasms | 25DTot | |
| 227.2 | | | | | 26972 | 26 | 26946 | | | | | 35433 | 62 | 35371 | | | Benign neoplasm of parathyroid gland | neoplasms | 25DTot | |
| 227.3 | | | | | 27062 | 116 | 26946 | | | | | 35525 | 154 | 35371 | | | Benign neoplasm of pituitary gland and craniopharyngeal duct (pouch) | neoplasms | 25DTot | |
| 228 | 0.131 | 0.1201 | 1.14 | 0.2757 | 26936 | 519 | 26417 | 0.5814 | 0.0784 | 1.789 | 1.21e-13 | 35731 | 759 | 34972 | 0.007405 | 0.3776 | Hemangioma and lymphangioma, any site | neoplasms | 25DTot | |
| 228.1 | | | | | 26853 | 436 | 26417 | 0.6451 | 0.08566 | 1.906 | 5.05e-14 | 35591 | 619 | 34972 | 0.01929 | 0.3507 | Hemangioma of skin and subcutaneous tissue | neoplasms | 25DTot | |
| 229 | | | | | 27551 | 83 | 27468 | | | | | 36445 | 69 | 36376 | | | Benign neoplasm of unspecified sites | neoplasms | 25DTot | |
| 229.1 | | | | | 27477 | 9 | 27468 | | | | | 36386 | 10 | 36376 | | | Benign neoplasm of lymph nodes | neoplasms | 25DTot | |
| 230 | | | | | 27785 | 15 | 27770 | | | | | 36952 | 4 | 36948 | | | Kaposi's sarcoma | neoplasms | 25DTot | |
| 240 | | | | | 23255 | 80 | 23175 | | | | | 26401 | 346 | 26055 | | | Simple and unspecified goiter | endocrine/metabolic | 25DTot | |
| 241 | 0.4424 | 0.1215 | 1.556 | 2.73e-04 | 23679 | 504 | 23175 | 0.4961 | 0.0537 | 1.642 | 2.47e-20 | 28029 | 1974 | 26055 | 0.7241 | 0.04604 | Nontoxic nodular goiter | endocrine/metabolic | 25DTot | |
| 241.1 | | | | | 23492 | 317 | 23175 | 0.4935 | 0.07374 | 1.638 | 2.20e-11 | 27019 | 964 | 26055 | 0.9941 | 0.001209 | Nontoxic uninodular goiter | endocrine/metabolic | 25DTot | |
| 241.2 | | | | | 23428 | 253 | 23175 | 0.5586 | 0.06432 | 1.748 | 3.79e-18 | 27333 | 1278 | 26055 | 0.8322 | -0.03624 | Nontoxic multinodular goiter | endocrine/metabolic | 25DTot | |
| 242 | | | | | 23461 | 286 | 23175 | 0.4558 | 0.08329 | 1.577 | 4.42e-08 | 26848 | 793 | 26055 | 0.7114 | -0.06476 | Thyrotoxicosis with or without goiter | endocrine/metabolic | 25DTot | |
| 242.1 | | | | | 23301 | 126 | 23175 | | | | | 26434 | 379 | 26055 | 0.8495 | 0.05066 | Graves' disease | endocrine/metabolic | 25DTot | |
| 242.2 | | | | | 23196 | 21 | 23175 | | | | | 26178 | 123 | 26055 | | | Toxic multinodular goiter | endocrine/metabolic | 25DTot | |
| 242.3 | | | | | 23231 | 56 | 23175 | | | | | 26122 | 67 | 26055 | | | Exophthalmos | endocrine/metabolic | 25DTot | |
| 242.31 | | | | | 23186 | 11 | 23175 | | | | | 26071 | 16 | 26055 | | | Thyrotoxic exophthalmos | endocrine/metabolic | 25DTot | |
| 244 | 0.5179 | 0.05898 | 1.678 | 1.64e-18 | 25712 | 2537 | 23175 | 0.4512 | 0.03394 | 1.57 | 2.48e-40 | 32658 | 6603 | 26055 | 0.5365 | -0.04139 | Hypothyroidism | endocrine/metabolic | 25DTot | |
| 244.1 | 0.5429 | 0.1191 | 1.721 | 5.21e-06 | 23694 | 519 | 23175 | 0.5968 | 0.06303 | 1.816 | 2.82e-21 | 27402 | 1347 | 26055 | 0.4241 | 0.1058 | Secondary hypothyroidism | endocrine/metabolic | 25DTot | |
| 244.2 | | | | | 23522 | 347 | 23175 | 0.6889 | 0.07794 | 1.992 | 9.66e-19 | 26876 | 821 | 26055 | 0.02125 | 0.3801 | Acquired hypothyroidism | endocrine/metabolic | 25DTot | |
| 244.3 | | | | | 23175 | | | | | | | 26060 | 5 | 26055 | | | Iodine hypothyroidism | endocrine/metabolic | 25DTot | |

| phenotype | male_beta | male_SE | male_OR | male_p | male_n_total | male_n_cases | male_n_controls | female_beta | female_SE | female_OR | female_p | female_n_total | female_n_cases | female_n_controls | interaction_p | interaction_beta | description | group | lab | label |
|---|---|---|---|---|---|---|---|---|---|---|---|---|---|---|---|---|---|---|---|---|
| 244.4 | 0.5358 | 0.06149 | 1.709 | 2.95e-18 | 25441 | 2266 | 23175 | 0.4502 | 0.03501 | 1.569 | 7.59e-38 | 32096 | 6041 | 26055 | 0.4246 | -0.05559 | Hypothyroidism NOS | endocrine/metabolic | 25DTot | |
| 244.5 | | | | | 23199 | 24 | 23175 | | | | | 26089 | 34 | 26055 | | | Congenital hypothyroidism | endocrine/metabolic | 25DTot | |
| 245 | | | | | 23436 | 261 | 23175 | 0.6009 | 0.07826 | 1.824 | 1.63e-14 | 26942 | 887 | 26055 | 0.7484 | -0.0556 | Thyroiditis | endocrine/metabolic | 25DTot | |
| 245.1 | | | | | 23196 | 21 | 23175 | | | | | 26097 | 42 | 26055 | | | Thyroiditis, acute and subacute | endocrine/metabolic | 25DTot | |
| 245.2 | | | | | 23326 | 151 | 23175 | 0.6426 | 0.09039 | 1.901 | 1.17e-12 | 26707 | 652 | 26055 | 0.7375 | -0.07339 | Chronic thyroiditis | endocrine/metabolic | 25DTot | |
| 245.21 | | | | | 23324 | 149 | 23175 | 0.6384 | 0.0909 | 1.894 | 2.16e-12 | 26701 | 646 | 26055 | 0.9476 | -0.0147 | Chronic lymphocytic thyroiditis | endocrine/metabolic | 25DTot | |
| 246 | | | | | 23388 | 213 | 23175 | | | | | 26539 | 484 | 26055 | 6.53e-04 | -0.6188 | Other disorders of thyroid | endocrine/metabolic | 25DTot | |
| 246.2 | | | | | 23182 | 7 | 23175 | | | | | 26065 | 10 | 26055 | | | Thyroid cyst | endocrine/metabolic | 25DTot | |
| 246.7 | | | | | 23294 | 119 | 23175 | | | | | 26304 | 249 | 26055 | | | Abnormal results of function study of thyroid | endocrine/metabolic | 25DTot | |
| 249 | | | | | 17130 | 307 | 16823 | | | | | 26073 | 265 | 25808 | 0.01035 | -0.4751 | Secondary diabetes mellitus | endocrine/metabolic | 25DTot | |
| 250 | 0.4479 | 0.04834 | 1.565 | 1.92e-20 | 22885 | 6062 | 16823 | 0.09419 | 0.03739 | 1.099 | 0.01176 | 31483 | 5675 | 25808 | 2.26e-10 | -0.3794 | Diabetes mellitus | endocrine/metabolic | 25DTot | |
| 250.1 | 1.099 | 0.07339 | 3.003 | 9.65e-51 | 18010 | 1187 | 16823 | 0.3524 | 0.06974 | 1.422 | 4.36e-07 | 27021 | 1213 | 25808 | 2.23e-16 | -0.8111 | Type 1 diabetes | endocrine/metabolic | 25DTot | |
| 250.11 | | | | | 16945 | 122 | 16823 | | | | | 25971 | 163 | 25808 | | | Type 1 diabetes with ketoacidosis | endocrine/metabolic | 25DTot | |
| 250.12 | | | | | 17020 | 197 | 16823 | | | | | 25984 | 176 | 25808 | | | Type 1 diabetes with renal manifestations | endocrine/metabolic | 25DTot | |
| 250.13 | | | | | 16957 | 134 | 16823 | | | | | 25957 | 149 | 25808 | | | Type 1 diabetes with ophthalmic manifestations | endocrine/metabolic | 25DTot | |
| 250.14 | | | | | 17123 | 300 | 16823 | | | | | 26082 | 274 | 25808 | 9.30e-04 | -0.6153 | Type 1 diabetes with neurological manifestations | endocrine/metabolic | 25DTot | |
| 250.15 | | | | | 16873 | 50 | 16823 | | | | | 25855 | 47 | 25808 | | | Diabetes type 1 with peripheral circulatory disorders | endocrine/metabolic | 25DTot | |
| 250.2 | 0.4462 | 0.04914 | 1.562 | 1.09e-19 | 22677 | 5854 | 16823 | 0.09634 | 0.03801 | 1.101 | 0.01127 | 31244 | 5436 | 25808 | 8.16e-10 | -0.3736 | Type 2 diabetes | endocrine/metabolic | 25DTot | |
| 250.21 | | | | | 16879 | 56 | 16823 | | | | | 25884 | 76 | 25808 | | | Type 2 diabetes with ketoacidosis | endocrine/metabolic | 25DTot | |
| 250.22 | 1.054 | 0.07082 | 2.869 | 4.19e-50 | 18226 | 1403 | 16823 | 0.4195 | 0.07487 | 1.521 | 2.11e-08 | 26727 | 919 | 25808 | 1.34e-10 | -0.6485 | Type 2 diabetes with renal manifestations | endocrine/metabolic | 25DTot | |
| 250.23 | | | | | 17271 | 448 | 16823 | | | | | 26204 | 396 | 25808 | 0.01923 | -0.3743 | Type 2 diabetes with ophthalmic manifestations | endocrine/metabolic | 25DTot | |
| 250.24 | 0.7939 | 0.0718 | 2.212 | 2.02e-28 | 18366 | 1543 | 16823 | 0.4132 | 0.06479 | 1.512 | 1.79e-10 | 27084 | 1276 | 25808 | 2.76e-05 | -0.3973 | Type 2 diabetes with neurological manifestations | endocrine/metabolic | 25DTot | |
| 250.25 | | | | | 17307 | 484 | 16823 | | | | | 26109 | 301 | 25808 | 0.4127 | -0.1389 | Diabetes type 2 with peripheral circulatory disorders | endocrine/metabolic | 25DTot | |
| 250.3 | 0.7468 | 0.06495 | 2.11 | 1.34e-30 | 18803 | 1980 | 16823 | 0.18 | 0.05891 | 1.197 | 0.002247 | 27587 | 1779 | 25808 | 3.97e-12 | -0.5958 | Insulin pump user | endocrine/metabolic | 25DTot | |
| 250.4 | 0.2554 | 0.05782 | 1.291 | 1.00e-05 | 20734 | 3911 | 16823 | 0.1842 | 0.04322 | 1.202 | 2.03e-05 | 29488 | 3680 | 25808 | 0.3651 | -0.06407 | Abnormal glucose | endocrine/metabolic | 25DTot | |
| 250.41 | 0.4555 | 0.1139 | 1.577 | 6.35e-05 | 17402 | 579 | 16823 | 0.6122 | 0.08488 | 1.845 | 5.47e-13 | 26465 | 657 | 25808 | 0.6614 | 0.06098 | Impaired fasting glucose | endocrine/metabolic | 25DTot | |
| 250.42 | 0.1898 | 0.06177 | 1.209 | 0.002124 | 20295 | 3472 | 16823 | 0.09514 | 0.04712 | 1.1 | 0.04348 | 28947 | 3139 | 25808 | 0.3346 | -0.07347 | Other abnormal glucose | endocrine/metabolic | 25DTot | |
| 250.5 | | | | | 16841 | 18 | 16823 | | | | | 25844 | 36 | 25808 | | | Glycosuria or Acetonuria | endocrine/metabolic | 25DTot | |

| phenotype | male_beta | male_SE | male_OR | male_p | male_n_total | male_n_cases | male_n_controls | female_beta | female_SE | female_OR | female_p | female_n_total | female_n_cases | female_n_controls | interaction_p | interaction_beta | description | group | lab | label |
|---|---|---|---|---|---|---|---|---|---|---|---|---|---|---|---|---|---|---|---|---|
| 250.6 | 0.8012 | 0.07865 | 2.228 | 2.26e-24 | 18032 | 1209 | 16823 | 0.3992 | 0.07353 | 1.491 | 5.64e-08 | 26769 | 961 | 25808 | 1.14e-04 | -0.4068 | Polyneuropathy in diabetes | endocrine/metabolic | 25DTot | |
| 250.7 | | | | | 24804 | 450 | 24354 | | | | | 33128 | 398 | 32730 | 0.1867 | -0.2115 | Diabetic retinopathy | endocrine/metabolic | 25DTot | |
| 251 | | | | | 23451 | 170 | 23281 | | | | | 31202 | 167 | 31035 | | | Other disorders of pancreatic internal secretion | endocrine/metabolic | 25DTot | |
| 251.1 | | | | | 15397 | 246 | 15151 | | | | | 23345 | 400 | 22945 | 0.2319 | -0.2449 | Hypoglycemia | endocrine/metabolic | 25DTot | |
| 251.8 | | | | | 23287 | 6 | 23281 | | | | | 31042 | 7 | 31035 | | | Abnormality of secretion of glucagon or gastrin | endocrine/metabolic | 25DTot | |
| 252 | | | | | 23548 | 267 | 23281 | 1.137 | 0.07865 | 3.118 | 2.17e-47 | 31785 | 750 | 31035 | 0.008567 | -0.3961 | Disorders of parathyroid gland | endocrine/metabolic | 25DTot | |
| 252.1 | | | | | 23519 | 238 | 23281 | 1.18 | 0.0838 | 3.255 | 4.81e-45 | 31693 | 658 | 31035 | 0.02297 | -0.3626 | Hyperparathyroidism | endocrine/metabolic | 25DTot | |
| 252.2 | | | | | 23316 | 35 | 23281 | | | | | 31163 | 128 | 31035 | | | Hypoparathyroidism | endocrine/metabolic | 25DTot | |
| 253 | | | | | 23665 | 384 | 23281 | | | | | 31431 | 396 | 31035 | 0.04552 | -0.3323 | Disorders of the pituitary gland and its hypothalamic control | endocrine/metabolic | 25DTot | |
| 253.1 | | | | | 23343 | 62 | 23281 | | | | | 31171 | 136 | 31035 | | | Pituitary hyperfunction | endocrine/metabolic | 25DTot | |
| 253.11 | | | | | 23302 | 21 | 23281 | | | | | 31046 | 11 | 31035 | | | Acromegaly and gigantism | endocrine/metabolic | 25DTot | |
| 253.2 | | | | | 23400 | 119 | 23281 | | | | | 31125 | 90 | 31035 | | | Pituitary hypofunction | endocrine/metabolic | 25DTot | |
| 253.3 | | | | | 23305 | 24 | 23281 | | | | | 31086 | 51 | 31035 | | | Diabetes insipidus | endocrine/metabolic | 25DTot | |
| 253.4 | | | | | 23423 | 142 | 23281 | | | | | 31110 | 75 | 31035 | | | Anterior pituitary disorders | endocrine/metabolic | 25DTot | |
| 253.5 | | | | | 23359 | 78 | 23281 | | | | | 31085 | 50 | 31035 | | | Pituitary dwarfism | endocrine/metabolic | 25DTot | |
| 253.7 | | | | | 23345 | 64 | 23281 | | | | | 31102 | 67 | 31035 | | | Other disorders of neurohypophysis | endocrine/metabolic | 25DTot | |
| 254 | | | | | 23286 | 5 | 23281 | | | | | 31042 | 7 | 31035 | | | Diseases of thymus gland | endocrine/metabolic | 25DTot | |
| 255 | 0.7848 | 0.1035 | 2.192 | 3.41e-14 | 23957 | 676 | 23281 | 0.5348 | 0.07951 | 1.707 | 1.75e-11 | 31897 | 862 | 31035 | 0.09339 | -0.2135 | Disorders of adrenal glands | endocrine/metabolic | 25DTot | |
| 255.1 | | | | | 23371 | 90 | 23281 | | | | | 31211 | 176 | 31035 | | | Adrenal hyperfunction | endocrine/metabolic | 25DTot | |
| 255.11 | | | | | 23313 | 32 | 23281 | | | | | 31129 | 94 | 31035 | | | Cushing's syndrome | endocrine/metabolic | 25DTot | |
| 255.12 | | | | | 23315 | 34 | 23281 | | | | | 31064 | 29 | 31035 | | | Hyperaldosteronism | endocrine/metabolic | 25DTot | |
| 255.13 | | | | | 23297 | 16 | 23281 | | | | | 31065 | 30 | 31035 | | | Medulloadrenal hyperfunction | endocrine/metabolic | 25DTot | |
| 255.2 | | | | | 23716 | 435 | 23281 | 0.6434 | 0.1023 | 1.903 | 3.20e-10 | 31535 | 500 | 31035 | 0.6881 | -0.06454 | Adrenal hypofunction | endocrine/metabolic | 25DTot | |
| 255.21 | | | | | 23674 | 393 | 23281 | | | | | 31474 | 439 | 31035 | 0.7464 | -0.05499 | Glucocorticoid deficiency | endocrine/metabolic | 25DTot | |
| 255.22 | | | | | 23296 | 15 | 23281 | | | | | 31067 | 32 | 31035 | | | Mineralocorticoid deficiency | endocrine/metabolic | 25DTot | |
| 255.3 | | | | | 23289 | 8 | 23281 | | | | | 31065 | 30 | 31035 | | | Adrenogenital disorders | endocrine/metabolic | 25DTot | |
| 256 | | | | | 4 | | | 0.8228 | 0.09364 | 2.277 | 1.53e-18 | 31698 | 666 | 31032 | 1.53e-18 | 0.8228 | Ovarian dysfunction | endocrine/metabolic | 25DTot | |
| 256.1 | | | | | 23283 | 2 | 23281 | | | | | 31037 | 2 | 31035 | | | Hyperestrogenism | endocrine/metabolic | 25DTot | |
| 256.4 | | | | | 4 | | | 0.6229 | 0.1128 | 1.864 | 3.34e-08 | 31569 | 537 | 31032 | 3.34e-08 | 0.6229 | Polycystic ovaries | endocrine/metabolic | 25DTot | |

| phenotype | male_beta | male_SE | male_OR | male_p | male_n_total | male_n_cases | male_n_controls | female_beta | female_SE | female_OR | female_p | female_n_total | female_n_cases | female_n_controls | interaction_p | interaction_beta | description | group | lab | label |
|---|---|---|---|---|---|---|---|---|---|---|---|---|---|---|---|---|---|---|---|---|
| 257 | 0.9242 | 0.07939 | 2.52 | 2.55e-31 | 24271 | 994 | 23277 | | | | | 3 | | | | | Testicular dysfunction | endocrine/metabolic | 25DTot | |
| 257.1 | 0.9264 | 0.07986 | 2.525 | 4.12e-31 | 24255 | 978 | 23277 | | | | | 3 | | | | | Testicular hypofunction | endocrine/metabolic | 25DTot | |
| 258 | | | | | 23304 | 23 | 23281 | | | | | 31068 | 33 | 31035 | | | Iatrogenic endocrine disorders | endocrine/metabolic | 25DTot | |
| 258.1 | | | | | 4 | | | | | | | 31087 | 55 | 31032 | | | Postablative ovarian failure | endocrine/metabolic | 25DTot | |
| 259 | | | | | 23461 | 180 | 23281 | | | | | 31392 | 357 | 31035 | 0.6626 | 0.09085 | Other endocrine disorders | endocrine/metabolic | 25DTot | |
| 259.1 | | | | | 23352 | 71 | 23281 | | | | | 31134 | 99 | 31035 | | | Nonspecific abnormal results of other endocrine function study | endocrine/metabolic | 25DTot | |
| 259.2 | | | | | 23320 | 39 | 23281 | | | | | 31092 | 57 | 31035 | | | Carcinoid syndrome | endocrine/metabolic | 25DTot | |
| 259.3 | | | | | 23324 | 43 | 23281 | | | | | 31063 | 28 | 31035 | | | Delay in sexual development and puberty NEC | endocrine/metabolic | 25DTot | |
| 259.4 | | | | | 23291 | 10 | 23281 | | | | | 31071 | 36 | 31035 | | | Precocious sexual development and puberty NEC | endocrine/metabolic | 25DTot | |
| 259.8 | | | | | 23297 | 16 | 23281 | | | | | 31067 | 32 | 31035 | | | Polyglandular activity in multiple endocrine adenomatosis | endocrine/metabolic | 25DTot | |
| 260 | 1.008 | 0.06246 | 2.741 | 1.22e-58 | 22122 | 2837 | 19285 | 0.7931 | 0.05276 | 2.21 | 4.39e-51 | 27318 | 2532 | 24786 | 0.07527 | -0.1429 | Protein-calorie malnutrition | endocrine/metabolic | 25DTot | |
| 260.1 | | | | | 19435 | 150 | 19285 | | | | | 24879 | 93 | 24786 | | | Cachexia | endocrine/metabolic | 25DTot | |
| 260.2 | 1.037 | 0.09101 | 2.821 | 4.53e-30 | 20371 | 1086 | 19285 | 0.7717 | 0.08085 | 2.163 | 1.37e-21 | 25774 | 988 | 24786 | 0.06652 | -0.2196 | severe protein-calorie malnutrition | endocrine/metabolic | 25DTot | |
| 260.21 | | | | | 19301 | 16 | 19285 | | | | | 24805 | 19 | 24786 | | | Kwashiorkor | endocrine/metabolic | 25DTot | |
| 260.22 | | | | | 19399 | 114 | 19285 | | | | | 24929 | 143 | 24786 | | | Nutritional marasmus | endocrine/metabolic | 25DTot | |
| 260.3 | 1.307 | 0.1091 | 3.695 | 4.34e-33 | 19914 | 629 | 19285 | 0.9737 | 0.09789 | 2.648 | 2.61e-23 | 25366 | 580 | 24786 | 0.08084 | -0.25 | Adult failure to thrive | endocrine/metabolic | 25DTot | |
| 260.6 | 0.811 | 0.1352 | 2.25 | 1.97e-09 | 19875 | 590 | 19285 | | | | | 25233 | 447 | 24786 | 0.735 | -0.06064 | Anorexia | endocrine/metabolic | 25DTot | |
| 260.7 | | | | | 19286 | 1 | 19285 | | | | | 24804 | 18 | 24786 | | | Polyphagia | endocrine/metabolic | 25DTot | |
| 261 | 2.032 | 0.05405 | 7.633 | 0 | 21716 | 2431 | 19285 | 1.695 | 0.03562 | 5.449 | 0 | 30360 | 5574 | 24786 | 1.84e-06 | -0.3087 | Vitamin deficiency | endocrine/metabolic | 25DTot | |
| 261.1 | | | | | 19312 | 27 | 19285 | | | | | 24829 | 43 | 24786 | | | Vitamin A deficiency | endocrine/metabolic | 25DTot | |
| 261.2 | 1.885 | 0.08854 | 6.585 | 1.44e-100 | 19973 | 688 | 19285 | 1.613 | 0.05676 | 5.016 | 1.57e-177 | 26376 | 1590 | 24786 | 0.004644 | -0.2948 | Vitamin B-complex deficiencies | endocrine/metabolic | 25DTot | |
| 261.3 | | | | | 19292 | 7 | 19285 | | | | | 24807 | 21 | 24786 | | | Vitamin C deficiencies | endocrine/metabolic | 25DTot | |
| 261.4 | 2.158 | 0.05783 | 8.651 | 1.19e-304 | 21216 | 1931 | 19285 | 1.796 | 0.03787 | 6.027 | 0 | 29414 | 4628 | 24786 | 4.74e-06 | -0.3156 | Vitamin D deficiency | endocrine/metabolic | 25DTot | Vitamin D deficiency |
| 261.41 | | | | | 19297 | 12 | 19285 | | | | | 24819 | 33 | 24786 | | | Rickets or osteomalacia | endocrine/metabolic | 25DTot | |
| 262 | | | | | 19493 | 208 | 19285 | | | | | 25217 | 431 | 24786 | 0.104 | -0.3154 | Mineral deficiency NEC | endocrine/metabolic | 25DTot | |
| 263 | | | | | 19569 | 284 | 19285 | | | | | 25175 | 389 | 24786 | 0.4872 | -0.1249 | Other nutritional deficiency | endocrine/metabolic | 25DTot | |
| 264 | | | | | 19689 | 404 | 19285 | | | | | 25083 | 297 | 24786 | 0.5646 | -0.174 | Lack of normal physiological development | endocrine/metabolic | 25DTot | |

| phenotype | male_beta | male_SE | male_OR | male_p | male_n_total | male_n_cases | male_n_controls | female_beta | female_SE | female_OR | female_p | female_n_total | female_n_cases | female_n_controls | interaction_p | interaction_beta | description | group | lab | label |
|---|---|---|---|---|---|---|---|---|---|---|---|---|---|---|---|---|---|---|---|---|
| 264.1 | | | | | 19424 | 139 | 19285 | | | | | 24879 | 93 | 24786 | | | Short stature | endocrine/metabolic | 25DTot | |
| 264.2 | | | | | 19409 | 124 | 19285 | | | | | 24883 | 97 | 24786 | | | Failure to thrive (childhood) | endocrine/metabolic | 25DTot | |
| 264.3 | | | | | 19388 | 103 | 19285 | | | | | 24857 | 71 | 24786 | | | Delayed milestones | endocrine/metabolic | 25DTot | |
| 264.9 | | | | | 19465 | 180 | 19285 | | | | | 24925 | 139 | 24786 | | | Lack of normal physiological development, unspecified | endocrine/metabolic | 25DTot | |
| 269 | 1.045 | 0.07879 | 2.845 | 3.54e-40 | 26340 | 917 | 25423 | 0.6162 | 0.08221 | 1.852 | 6.59e-14 | 35408 | 726 | 34682 | 4.77e-05 | -0.4507 | Proteinuria | endocrine/metabolic | 25DTot | |
| 270 | | | | | 25887 | 464 | 25423 | 0.3406 | 0.103 | 1.406 | 9.44e-04 | 35187 | 505 | 34682 | 0.01186 | -0.3872 | Disorders of protein plasma/amino-acid transport and metabolism | endocrine/metabolic | 25DTot | |
| 270.1 | | | | | 25452 | 29 | 25423 | | | | | 34732 | 50 | 34682 | | | Disturbances of amino-acid transport | endocrine/metabolic | 25DTot | |
| 270.11 | | | | | 25431 | 8 | 25423 | | | | | 34711 | 29 | 34682 | | | Disturbances of sulphur-bearing amino-acid metabolism | endocrine/metabolic | 25DTot | |
| 270.12 | | | | | 25434 | 11 | 25423 | | | | | 34696 | 14 | 34682 | | | Phenylketonuria [PKU] | endocrine/metabolic | 25DTot | |
| 270.2 | | | | | 25465 | 42 | 25423 | | | | | 34719 | 37 | 34682 | | | Disorders of amino-acid metabolism | endocrine/metabolic | 25DTot | |
| 270.21 | | | | | 25457 | 34 | 25423 | | | | | 34703 | 21 | 34682 | | | Disorders of urea cycle metabolism | endocrine/metabolic | 25DTot | |
| 270.3 | | | | | 25814 | 391 | 25423 | | | | | 35096 | 414 | 34682 | 0.0123 | -0.4174 | Disorders of plasma protein metabolism | endocrine/metabolic | 25DTot | |
| 270.31 | | | | | 25425 | 2 | 25423 | | | | | 34689 | 7 | 34682 | | | Polyclonal hypergammaglobulinemia | endocrine/metabolic | 25DTot | |
| 270.32 | | | | | 25665 | 242 | 25423 | | | | | 34920 | 238 | 34682 | | | Paraproteinemia | endocrine/metabolic | 25DTot | |
| 270.33 | | | | | 25484 | 61 | 25423 | | | | | 34731 | 49 | 34682 | | | Amyloidosis | endocrine/metabolic | 25DTot | |
| 270.34 | | | | | 25480 | 57 | 25423 | | | | | 34726 | 44 | 34682 | | | Alpha-1-antitrypsin deficiency | endocrine/metabolic | 25DTot | |
| 270.35 | | | | | 25440 | 17 | 25423 | | | | | 34692 | 10 | 34682 | | | Macroglobulinemia | endocrine/metabolic | 25DTot | |
| 270.38 | | | | | 25509 | 86 | 25423 | | | | | 34792 | 110 | 34682 | | | Other specified disorders of plasma protein metabolism | endocrine/metabolic | 25DTot | |
| 271 | | | | | 27474 | 374 | 27100 | 0.5055 | 0.08344 | 1.658 | 1.38e-09 | 36489 | 676 | 35813 | 0.04021 | 0.3287 | Disorders of carbohydrate transport and metabolism | endocrine/metabolic | 25DTot | |
| 271.3 | | | | | 27455 | 355 | 27100 | 0.5209 | 0.08412 | 1.683 | 5.96e-10 | 36474 | 661 | 35813 | 0.02877 | 0.3591 | Intestinal disaccharidase deficiencies and disaccharide malabsorption | endocrine/metabolic | 25DTot | |
| 271.9 | | | | | 27119 | 19 | 27100 | | | | | 35828 | 15 | 35813 | | | Other disorders of carbohydrate transport and metabolism | endocrine/metabolic | 25DTot | |
| 272 | 0.4167 | 0.05014 | 1.517 | 9.50e-17 | 25396 | 11129 | 14267 | 0.4845 | 0.03355 | 1.623 | 2.77e-47 | 34085 | 10701 | 23384 | 0.6769 | 0.02494 | Disorders of lipoid metabolism | endocrine/metabolic | 25DTot | |
| 272.1 | 0.4172 | 0.05015 | 1.518 | 8.94e-17 | 25384 | 11117 | 14267 | 0.4839 | 0.03358 | 1.622 | 4.39e-47 | 34065 | 10681 | 23384 | 0.6939 | 0.02357 | Hyperlipidemia | endocrine/metabolic | 25DTot | |
| 272.11 | 0.7022 | 0.06502 | 2.018 | 3.44e-27 | 18174 | 3907 | 14267 | 0.7653 | 0.04477 | 2.15 | 1.62e-65 | 27356 | 3972 | 23384 | 0.9678 | 0.003166 | Hypercholesterolemia | endocrine/metabolic | 25DTot | |
| 272.12 | | | | | 14667 | 400 | 14267 | | | | | 23678 | 294 | 23384 | 0.7123 | -0.06483 | Hyperglyceridemia | endocrine/metabolic | 25DTot | |
| 272.13 | 0.6215 | 0.05774 | 1.862 | 5.11e-27 | 20087 | 5820 | 14267 | 0.6531 | 0.04044 | 1.922 | 1.10e-58 | 28806 | 5422 | 23384 | 0.9095 | -0.007987 | Mixed hyperlipidemia | endocrine/metabolic | 25DTot | |
| 272.14 | | | | | 14272 | 5 | 14267 | | | | | 23388 | 4 | 23384 | | | Hyperchylomicronemia | endocrine/metabolic | 25DTot | |
| 272.9 | | | | | 14286 | 19 | 14267 | | | | | 23403 | 19 | 23384 | | | Unspecified disorder of lipoid metabolism | endocrine/metabolic | 25DTot | |

| phenotype | male_beta | male_SE | male_OR | male_p | male_n_total | male_n_cases | male_n_controls | female_beta | female_SE | female_OR | female_p | female_n_total | female_n_cases | female_n_controls | interaction_p | interaction_beta | description | group | lab | label |
|---|---|---|---|---|---|---|---|---|---|---|---|---|---|---|---|---|---|---|---|---|
| 274 | 0.8767 | 0.06045 | 2.403 | 1.19e-47 | 26980 | 1926 | 25054 | 0.4776 | 0.07521 | 1.612 | 2.15e-10 | 36355 | 864 | 35491 | 5.09e-05 | -0.3802 | Gout and other crystal arthropathies | endocrine/metabolic | 25DTot | |
| 274.1 | 0.875 | 0.06175 | 2.399 | 1.44e-45 | 26882 | 1828 | 25054 | 0.4578 | 0.08225 | 1.581 | 2.61e-08 | 36217 | 726 | 35491 | 1.81e-05 | -0.4278 | Gout | endocrine/metabolic | 25DTot | |
| 274.11 | 0.9469 | 0.09388 | 2.578 | 6.40e-24 | 25702 | 648 | 25054 | | | | | 35744 | 253 | 35491 | 0.0782 | -0.2782 | Gouty arthropathy | endocrine/metabolic | 25DTot | |
| 274.2 | | | | | 25175 | 121 | 25054 | | | | | 35648 | 157 | 35491 | | | Crystal arthropathies | endocrine/metabolic | 25DTot | |
| 274.21 | | | | | 25165 | 111 | 25054 | | | | | 35627 | 136 | 35491 | | | Chondrocalcinosis | endocrine/metabolic | 25DTot | |
| 275 | 0.8006 | 0.06017 | 2.227 | 2.14e-40 | 25825 | 2131 | 23694 | 0.4759 | 0.04808 | 1.61 | 4.18e-23 | 34866 | 2480 | 32386 | 0.001876 | -0.2331 | Disorders of mineral metabolism | endocrine/metabolic | 25DTot | |
| 275.1 | | | | | 23853 | 159 | 23694 | | | | | 32511 | 125 | 32386 | | | Disorders of iron metabolism | hematopoietic | 25DTot | |
| 275.11 | | | | | 23755 | 61 | 23694 | | | | | 32420 | 34 | 32386 | | | Hereditary hemochromatosis | hematopoietic | 25DTot | |
| 275.2 | | | | | 23712 | 18 | 23694 | | | | | 32400 | 14 | 32386 | | | Disorders of copper metabolism | endocrine/metabolic | 25DTot | |
| 275.3 | 0.7452 | 0.09456 | 2.107 | 3.25e-15 | 24542 | 848 | 23694 | 0.2208 | 0.08213 | 1.247 | 0.007163 | 33312 | 926 | 32386 | 2.11e-04 | -0.4514 | Disorders of magnesium metabolism | endocrine/metabolic | 25DTot | |
| 275.5 | 1.076 | 0.08122 | 2.933 | 4.72e-40 | 24610 | 916 | 23694 | 0.6981 | 0.07386 | 2.01 | 3.33e-21 | 33307 | 921 | 32386 | 0.003904 | -0.3072 | Disorders of calcium/phosphorus metabolism | endocrine/metabolic | 25DTot | |
| 275.51 | | | | | 23996 | 302 | 23694 | | | | | 32803 | 417 | 32386 | 0.4964 | -0.1187 | Hypocalcemia | endocrine/metabolic | 25DTot | |
| 275.53 | 1.201 | 0.1011 | 3.322 | 1.59e-32 | 24238 | 544 | 23694 | | | | | 32769 | 383 | 32386 | 0.001654 | -0.4637 | Disorders of phosphorus metabolism | endocrine/metabolic | 25DTot | |
| 275.6 | | | | | 24110 | 416 | 23694 | 0.8326 | 0.08188 | 2.299 | 2.74e-24 | 33089 | 703 | 32386 | 0.7639 | -0.04234 | Hypercalcemia | endocrine/metabolic | 25DTot | |
| 276 | 0.5291 | 0.04447 | 1.697 | 1.20e-32 | 24148 | 8143 | 16005 | 0.2304 | 0.03235 | 1.259 | 1.06e-12 | 32644 | 8450 | 24194 | 8.67e-06 | -0.24 | Disorders of fluid, electrolyte, and acid-base balance | endocrine/metabolic | 25DTot | |
| 276.1 | 0.6026 | 0.04776 | 1.827 | 1.69e-36 | 22066 | 6061 | 16005 | 0.3037 | 0.0355 | 1.355 | 1.18e-17 | 30513 | 6319 | 24194 | 3.30e-05 | -0.2424 | Electrolyte imbalance | endocrine/metabolic | 25DTot | |
| 276.11 | | | | | 16449 | 444 | 16005 | | | | | 24509 | 315 | 24194 | 0.01251 | -0.492 | Hyperosmolality and/or hypernatremia | endocrine/metabolic | 25DTot | |
| 276.12 | 0.5694 | 0.06924 | 1.767 | 1.96e-16 | 18173 | 2168 | 16005 | 0.3285 | 0.05438 | 1.389 | 1.53e-09 | 26321 | 2127 | 24194 | 0.08627 | -0.148 | Hyposmolality and/or hyponatremia | endocrine/metabolic | 25DTot | |
| 276.13 | 0.8708 | 0.06354 | 2.389 | 9.47e-43 | 18167 | 2162 | 16005 | 0.5407 | 0.05867 | 1.717 | 3.07e-20 | 25804 | 1610 | 24194 | 0.001682 | -0.2654 | Hyperpotassemia | endocrine/metabolic | 25DTot | |
| 276.14 | 0.637 | 0.0628 | 1.891 | 3.57e-24 | 18672 | 2667 | 16005 | 0.3431 | 0.0439 | 1.409 | 5.53e-15 | 27767 | 3573 | 24194 | 7.12e-04 | -0.2543 | Hypopotassemia | endocrine/metabolic | 25DTot | |
| 276.4 | 0.8376 | 0.06494 | 2.311 | 4.61e-38 | 18109 | 2104 | 16005 | 0.2427 | 0.05925 | 1.275 | 4.19e-05 | 26022 | 1828 | 24194 | 1.45e-10 | -0.5502 | Acid-base balance disorder | endocrine/metabolic | 25DTot | |
| 276.41 | 0.872 | 0.06721 | 2.392 | 1.72e-38 | 17879 | 1874 | 16005 | 0.2797 | 0.06209 | 1.323 | 6.66e-06 | 25798 | 1604 | 24194 | 1.37e-09 | -0.5413 | Acidosis | endocrine/metabolic | 25DTot | |
| 276.42 | | | | | 16255 | 250 | 16005 | | | | | 24456 | 262 | 24194 | 8.00e-04 | -0.7636 | Alkalosis | endocrine/metabolic | 25DTot | |
| 276.5 | 0.6393 | 0.05813 | 1.895 | 3.92e-28 | 19178 | 3173 | 16005 | 0.244 | 0.04534 | 1.276 | 7.36e-08 | 27587 | 3393 | 24194 | 4.86e-06 | -0.3303 | Hypovolemia | endocrine/metabolic | 25DTot | |
| 276.6 | 0.8615 | 0.07128 | 2.367 | 1.24e-33 | 17709 | 1704 | 16005 | 0.2912 | 0.06675 | 1.338 | 1.29e-05 | 25588 | 1394 | 24194 | 3.72e-09 | -0.5613 | Fluid overload | endocrine/metabolic | 25DTot | |
| 276.8 | | | | | 16014 | 9 | 16005 | | | | | 24216 | 22 | 24194 | | | Polydipsia | endocrine/metabolic | 25DTot | |
| 277 | | | | | 26131 | 420 | 25711 | 0.4981 | 0.09574 | 1.646 | 1.96e-07 | 35099 | 554 | 34545 | 0.007102 | -0.3947 | Other disorders of metabolism | endocrine/metabolic | 25DTot | |
| phenotype | male_beta | male_SE | male_OR | male_p | male_n_total | male_n_cases | male_n_controls | female_beta | female_SE | female_OR | female_p | female_n_total | female_n_cases | female_n_controls | interaction_p | interaction_beta | description | group | lab | label |
| 277.1 | | | | | 25722 | 11 | 25711 | | | | | 34554 | 9 | 34545 | | | Disorders of porphyrin metabolism | endocrine/metabolic | 25DTot | |
| 277.2 | | | | | 25713 | 2 | 25711 | | | | | 34547 | 2 | 34545 | | | Other disorders of purine and pyrimidine metabolism | endocrine/metabolic | 25DTot | |

| phenotype | male_beta | male_SE | male_OR | male_p | male_n_total | male_n_cases | male_n_controls | female_beta | female_SE | female_OR | female_p | female_n_total | female_n_cases | female_n_controls | interaction_p | interaction_beta | description | group | lab | label |
|---|---|---|---|---|---|---|---|---|---|---|---|---|---|---|---|---|---|---|---|---|
| 277.4 | | | | | 25910 | 199 | 25711 | | | | | 34665 | 120 | 34545 | | | Disorders of bilirubin excretion | endocrine/metabolic | 25DTot | |
| 277.5 | | | | | 25845 | 134 | 25711 | | | | | 34656 | 111 | 34545 | | | Other disorders of lipoid metabolism | endocrine/metabolic | 25DTot | |
| 277.51 | | | | | 25790 | 79 | 25711 | | | | | 34590 | 45 | 34545 | | | Lipoprotein disorders | endocrine/metabolic | 25DTot | |
| 277.6 | | | | | 25723 | 12 | 25711 | | | | | 34560 | 15 | 34545 | | | Other deficiencies of circulating enzymes | endocrine/metabolic | 25DTot | |
| 277.7 | | | | | 25918 | 207 | 25711 | | | | | 34828 | 283 | 34545 | | | Dysmetabolic syndrome X | endocrine/metabolic | 25DTot | |
| 277.8 | | | | | 25715 | 4 | 25711 | | | | | 34546 | 1 | 34545 | | | Carnitine deficiencies | endocrine/metabolic | 25DTot | |
| 278 | 0.4893 | 0.05021 | 1.631 | 1.93e-22 | 24739 | 3939 | 20800 | 0.2719 | 0.03397 | 1.312 | 1.20e-15 | 32627 | 6840 | 25787 | 1.57e-04 | -0.225 | Overweight, obesity and other hyperalimentation | endocrine/metabolic | 25DTot | |
| 278.1 | 0.5162 | 0.0532 | 1.676 | 2.89e-22 | 24116 | 3316 | 20800 | 0.2327 | 0.03699 | 1.262 | 3.16e-10 | 31306 | 5519 | 25787 | 2.35e-06 | -0.3003 | Obesity | endocrine/metabolic | 25DTot | |
| 278.11 | 0.5083 | 0.0727 | 1.662 | 2.72e-12 | 22371 | 1571 | 20800 | 0.3297 | 0.04554 | 1.391 | 4.49e-13 | 28985 | 3198 | 25787 | 0.02398 | -0.1902 | Morbid obesity | endocrine/metabolic | 25DTot | |
| 278.3 | | | | | 20837 | 37 | 20800 | | | | | 25988 | 201 | 25787 | | | Localized adiposity | endocrine/metabolic | 25DTot | |
| 278.4 | | | | | 21218 | 418 | 20800 | 0.5832 | 0.0617 | 1.792 | 3.34e-21 | 27263 | 1476 | 25787 | 0.1813 | -0.1799 | Abnormal weight gain | endocrine/metabolic | 25DTot | |
| 279 | 1.112 | 0.07118 | 3.04 | 5.33e-55 | 25738 | 1171 | 24567 | 0.7667 | 0.05755 | 2.153 | 1.72e-40 | 33388 | 1505 | 31883 | 6.22e-04 | -0.307 | Disorders involving the immune mechanism | endocrine/metabolic | 25DTot | |
| 279.1 | 1.109 | 0.07309 | 3.032 | 4.92e-52 | 25668 | 1101 | 24567 | 0.7383 | 0.05968 | 2.092 | 3.71e-35 | 33274 | 1391 | 31883 | 4.21e-04 | -0.3263 | Immunity deficiency | endocrine/metabolic | 25DTot | |
| 279.11 | | | | | 24666 | 99 | 24567 | | | | | 32052 | 169 | 31883 | | | Deficiency of humoral immunity | endocrine/metabolic | 25DTot | |
| 279.2 | | | | | 24578 | 11 | 24567 | | | | | 31914 | 31 | 31883 | | | Autoimmune disease NEC | endocrine/metabolic | 25DTot | |
| 279.7 | | | | | 24823 | 256 | 24567 | 0.7557 | 0.07341 | 2.129 | 7.56e-25 | 32824 | 941 | 31883 | 0.8359 | -0.03556 | Other immunological findings | endocrine/metabolic | 25DTot | |
| 279.8 | | | | | 24598 | 31 | 24567 | | | | | 31935 | 52 | 31883 | | | Other specified disorders involving the immune mechanism | endocrine/metabolic | 25DTot | |
| 280 | 1.075 | 0.0661 | 2.929 | 1.98e-59 | 18083 | 1688 | 16395 | 0.7083 | 0.04521 | 2.03 | 2.65e-55 | 25891 | 2832 | 23059 | 1.31e-05 | -0.3429 | Iron deficiency anemias | hematopoietic | 25DTot | |
| 280.1 | 1.137 | 0.0713 | 3.117 | 3.08e-57 | 17740 | 1345 | 16395 | 0.7604 | 0.04816 | 2.139 | 3.69e-56 | 25454 | 2395 | 23059 | 1.76e-05 | -0.3628 | Iron deficiency anemias, unspecified or not due to blood loss | hematopoietic | 25DTot | |
| 280.2 | 0.9357 | 0.1128 | 2.549 | 1.09e-16 | 16909 | 514 | 16395 | 0.594 | 0.08194 | 1.811 | 4.20e-13 | 23821 | 762 | 23059 | 0.01316 | -0.3379 | Iron deficiency anemia secondary to blood loss (chronic) | hematopoietic | 25DTot | |
| 281 | | | | | 16870 | 475 | 16395 | 0.9903 | 0.07097 | 2.692 | 3.01e-44 | 24011 | 952 | 23059 | 0.245 | -0.1511 | Other deficiency anemia | hematopoietic | 25DTot | |
| 281.1 | | | | | 16669 | 274 | 16395 | 1.161 | 0.08283 | 3.193 | 1.26e-44 | 23736 | 677 | 23059 | 0.1594 | -0.2211 | Megaloblastic anemia | hematopoietic | 25DTot | |
| 281.11 | | | | | 16464 | 69 | 16395 | | | | | 23278 | 219 | 23059 | | | Pernicious anemia | hematopoietic | 25DTot | |
| 281.12 | | | | | 16590 | 195 | 16395 | 1.288 | 0.09549 | 3.627 | 1.74e-41 | 23561 | 502 | 23059 | 0.2288 | -0.2176 | Other vitamin B12 deficiency anemia | hematopoietic | 25DTot | |
| 281.13 | | | | | 16415 | 20 | 16395 | | | | | 23107 | 48 | 23059 | | | Folate-deficiency anemia | hematopoietic | 25DTot | |
| 281.9 | | | | | 16581 | 186 | 16395 | | | | | 23292 | 233 | 23059 | | | Deficiency anemias | hematopoietic | 25DTot | |
| 282 | | | | | 16442 | 47 | 16395 | | | | | 23190 | 131 | 23059 | | | Hereditary hemolytic anemias | hematopoietic | 25DTot | |
| 282.5 | | | | | 16399 | 4 | 16395 | | | | | 23069 | 10 | 23059 | | | Sickle cell anemia | hematopoietic | 25DTot | |
| 282.8 | | | | | 16410 | 15 | 16395 | | | | | 23102 | 43 | 23059 | | | Other hemoglobinopathies | hematopoietic | 25DTot | |
| 282.9 | | | | | 16427 | 32 | 16395 | | | | | 23140 | 81 | 23059 | | | Other hereditary hemolytic anemias | hematopoietic | 25DTot | |
| 283 | | | | | 16455 | 60 | 16395 | | | | | 23168 | 109 | 23059 | | | Acquired hemolytic anemias | hematopoietic | 25DTot | |
| 283.1 | | | | | 16418 | 23 | 16395 | | | | | 23095 | 36 | 23059 | | | Autoimmune hemolytic anemias | hematopoietic | 25DTot | |
| 283.2 | | | | | 16419 | 24 | 16395 | | | | | 23100 | 41 | 23059 | | | Non-autoimmune hemolytic anemias | hematopoietic | 25DTot | |

| phenotype | male_beta | male_SE | male_OR | male_p | male_n_total | male_n_cases | male_n_controls | female_beta | female_SE | female_OR | female_p | female_n_total | female_n_cases | female_n_controls | interaction_p | interaction_beta | description | group | lab | label |
|---|---|---|---|---|---|---|---|---|---|---|---|---|---|---|---|---|---|---|---|---|
| 283.21 | | | | | 16402 | 7 | 16395 | | | | | 23073 | 14 | 23059 | | | Hemolytic-uremic syndrome | hematopoietic | 25DTot | |
| 284 | 0.7422 | 0.1006 | 2.1 | 1.58e-13 | 17250 | 855 | 16395 | 0.0756 | 0.101 | 1.079 | 0.4541 | 23771 | 712 | 23059 | 1.24e-06 | -0.6737 | Aplastic anemia | hematopoietic | 25DTot | |
| 284.1 | 0.7972 | 0.1064 | 2.219 | 6.90e-14 | 17146 | 751 | 16395 | 0.08686 | 0.1058 | 1.091 | 0.4117 | 23702 | 643 | 23059 | 3.11e-06 | -0.6819 | Pancytopenia | hematopoietic | 25DTot | |
| 284.2 | | | | | 16406 | 11 | 16395 | | | | | 23065 | 6 | 23059 | | | Constitutional aplastic anemia | hematopoietic | 25DTot | |
| 285 | 0.5523 | 0.04549 | 1.737 | 6.46e-34 | 23977 | 7582 | 16395 | 0.2533 | 0.03352 | 1.288 | 4.15e-14 | 30991 | 7932 | 23059 | 5.51e-07 | -0.2769 | Other anemias | hematopoietic | 25DTot | |
| 285.1 | 0.4747 | 0.06421 | 1.608 | 1.44e-13 | 19293 | 2898 | 16395 | 0.01836 | 0.05768 | 1.019 | 0.7502 | 25340 | 2281 | 23059 | 1.93e-07 | -0.4396 | Acute posthemorrhagic anemia | hematopoietic | 25DTot | |
| 285.2 | 0.8203 | 0.06433 | 2.271 | 3.02e-37 | 18710 | 2315 | 16395 | 0.2159 | 0.0569 | 1.241 | 1.48e-04 | 25252 | 2193 | 23059 | 8.47e-14 | -0.6239 | Anemia of chronic disease | hematopoietic | 25DTot | |
| 285.21 | 1.359 | 0.08117 | 3.891 | 6.70e-63 | 17283 | 888 | 16395 | 0.6569 | 0.08289 | 1.929 | 2.30e-15 | 23775 | 716 | 23059 | 4.43e-10 | -0.7079 | Anemia in chronic kidney disease | hematopoietic | 25DTot | |
| 285.22 | -0.1882 | 0.1324 | 0.8284 | 0.1549 | 17510 | 1115 | 16395 | -0.3749 | 0.09355 | 0.6874 | 6.15e-05 | 24273 | 1214 | 23059 | 0.2951 | -0.1672 | Anemia in neoplastic disease | hematopoietic | 25DTot | |
| 285.3 | | | | | 16401 | 6 | 16395 | | | | | 23062 | 3 | 23059 | | | Sideroblastic anemia | hematopoietic | 25DTot | |
| 285.8 | | | | | 16399 | 4 | 16395 | | | | | 23072 | 13 | 23059 | | | Hemoglobinuria | hematopoietic | 25DTot | |
| 286 | 0.5116 | 0.07036 | 1.668 | 3.58e-13 | 21087 | 1817 | 19270 | 0.06053 | 0.0586 | 1.062 | 0.3017 | 30311 | 1937 | 28374 | 3.91e-07 | -0.4539 | Coagulation defects | hematopoietic | 25DTot | |
| 286.1 | | | | | 19382 | 112 | 19270 | | | | | 28488 | 114 | 28374 | | | Congenital coagulation defects | hematopoietic | 25DTot | |
| 286.11 | | | | | 19306 | 36 | 19270 | | | | | 28422 | 48 | 28374 | | | Von willebrand's disease | hematopoietic | 25DTot | |
| 286.12 | | | | | 19305 | 35 | 19270 | | | | | 28432 | 58 | 28374 | | | Congenital deficiency of other clotting factors (including factor VII) | hematopoietic | 25DTot | |
| 286.13 | | | | | 19324 | 54 | 19270 | | | | | 28385 | 11 | 28374 | | | Congenital factor VIII disorder | hematopoietic | 25DTot | |
| 286.2 | 0.3193 | 0.05965 | 1.376 | 8.65e-08 | 22410 | 3140 | 19270 | 0.08712 | 0.04868 | 1.091 | 0.07349 | 31115 | 2741 | 28374 | 0.001579 | -0.2385 | Encounter for long-term (current) use of anticoagulants | hematopoietic | 25DTot | |
| 286.3 | | | | | 6 | | | | | | | 28478 | 106 | 28372 | | | Coagulation defects complicating pregnancy or postpartum | hematopoietic | 25DTot | |
| 286.4 | | | | | 19353 | 83 | 19270 | | | | | 28429 | 55 | 28374 | | | Acquired coagulation factor deficiency | hematopoietic | 25DTot | |
| 286.5 | | | | | 19388 | 118 | 19270 | | | | | 28511 | 137 | 28374 | | | Hemorrhagic disorder due to intrinsic circulating anticoagulants | hematopoietic | 25DTot | |
| 286.6 | | | | | 19312 | 42 | 19270 | | | | | 28420 | 46 | 28374 | | | Defibrination syndrome | hematopoietic | 25DTot | |
| 286.7 | 0.5614 | 0.09617 | 1.753 | 5.28e-09 | 20156 | 886 | 19270 | 0.1064 | 0.08218 | 1.112 | 0.1955 | 29323 | 949 | 28374 | 3.67e-04 | -0.4401 | Other and unspecified coagulation defects | hematopoietic | 25DTot | |
| 286.8 | | | | | 19460 | 190 | 19270 | | | | | 28789 | 415 | 28374 | 0.4626 | -0.1848 | Hypercoagulable state | hematopoietic | 25DTot | |
| 286.81 | | | | | 19445 | 175 | 19270 | | | | | 28747 | 373 | 28374 | 0.7352 | -0.09016 | Primary hypercoagulable state | hematopoietic | 25DTot | |
| 286.9 | | | | | 19741 | 471 | 19270 | | | | | 28784 | 410 | 28374 | 0.002307 | -0.514 | Abnormal coagulation profile | hematopoietic | 25DTot | |
| phenotype | male_beta | male_SE | male_OR | male_p | male_n_total | male_n_cases | male_n_controls | female_beta | female_SE | female_OR | female_p | female_n_total | female_n_cases | female_n_controls | interaction_p | interaction_beta | description | group | lab | label |
| 287 | 0.5029 | 0.06505 | 1.654 | 1.06e-14 | 21610 | 2340 | 19270 | 0.06713 | 0.06085 | 1.069 | 0.2699 | 30271 | 1897 | 28374 | 4.44e-08 | -0.4742 | Purpura and other hemorrhagic conditions | hematopoietic | 25DTot | |
| 287.1 | | | | | 19326 | 56 | 19270 | | | | | 28485 | 111 | 28374 | | | Spontaneous ecchymoses | hematopoietic | 25DTot | |
| 287.2 | | | | | 19288 | 18 | 19270 | | | | | 28390 | 16 | 28374 | | | Allergic purpura | hematopoietic | 25DTot | |
| 287.3 | 0.4852 | 0.06818 | 1.624 | 1.11e-12 | 21432 | 2162 | 19270 | -0.009351 | 0.06548 | 0.9907 | 0.8864 | 30102 | 1728 | 28374 | 9.58e-09 | -0.5282 | Thrombocytopenia | hematopoietic | 25DTot | |
| 287.31 | | | | | 19391 | 121 | 19270 | | | | | 28538 | 164 | 28374 | | | Primary thrombocytopenia | hematopoietic | 25DTot | |
| 287.32 | | | | | 19594 | 324 | 19270 | | | | | 28675 | 301 | 28374 | 0.004249 | -0.6253 | Secondary thrombocytopenia | hematopoietic | 25DTot | |
| 287.4 | | | | | 19290 | 20 | 19270 | | | | | 28393 | 19 | 28374 | | | Qualitative platelet defects | hematopoietic | 25DTot | |
| 288 | 0.4533 | 0.0605 | 1.574 | 6.73e-14 | 21474 | 2790 | 18684 | 0.1905 | 0.04938 | 1.21 | 1.15e-04 | 29549 | 2753 | 26796 | 0.009684 | -0.1975 | Diseases of white blood cells | hematopoietic | 25DTot | |
| 288.1 | 0.3471 | 0.09572 | 1.415 | 2.88e-04 | 19788 | 1104 | 18684 | 0.05666 | 0.07148 | 1.058 | 0.428 | 28156 | 1360 | 26796 | 0.07196 | -0.2105 | Decreased white blood cell count | hematopoietic | 25DTot | |
| 288.11 | 0.04477 | 0.1254 | 1.046 | 0.721 | 19506 | 822 | 18684 | -0.08187 | 0.08666 | 0.9214 | 0.3448 | 27815 | 1019 | 26796 | 0.7661 | -0.04455 | Neutropenia | hematopoietic | 25DTot | |

| phenotype | male_beta | male_SE | male_OR | male_p | male_n_total | male_n_cases | male_n_controls | female_beta | female_SE | female_OR | female_p | female_n_total | female_n_cases | female_n_controls | interaction_p | interaction_beta | description | group | lab | label |
|---|---|---|---|---|---|---|---|---|---|---|---|---|---|---|---|---|---|---|---|---|
| 288.2 | 0.4062 | 0.06643 | 1.501 | 9.66e-10 | 21008 | 2324 | 18684 | 0.1887 | 0.05407 | 1.208 | 4.85e-04 | 29055 | 2259 | 26796 | 0.07532 | -0.1489 | Elevated white blood cell count | hematopoietic | 25DTot | |
| 288.3 | | | | | 18778 | 94 | 18684 | | | | | 26909 | 113 | 26796 | | | Eosinophilia | hematopoietic | 25DTot | |
| 289 | | | | | 19172 | 488 | 18684 | 0.2589 | 0.09449 | 1.296 | 0.006136 | 27421 | 625 | 26796 | 0.1178 | -0.2397 | Other diseases of blood and blood-forming organs | hematopoietic | 25DTot | |
| 289.1 | | | | | 18755 | 71 | 18684 | | | | | 26856 | 60 | 26796 | | | Myelofibrosis | hematopoietic | 25DTot | |
| 289.3 | | | | | 18767 | 83 | 18684 | | | | | 26919 | 123 | 26796 | | | Personal history of diseases of blood and blood-forming organs | hematopoietic | 25DTot | |
| 289.4 | 0.09918 | 0.08894 | 1.104 | 0.2648 | 20171 | 1487 | 18684 | 0.1739 | 0.06228 | 1.19 | 0.005245 | 28443 | 1647 | 26796 | 0.1086 | 0.1709 | Lymphadenitis | hematopoietic | 25DTot | |
| 289.5 | | | | | 18840 | 156 | 18684 | | | | | 26948 | 152 | 26796 | | | Diseases of spleen | hematopoietic | 25DTot | |
| 289.8 | | | | | 18685 | 237 | 18448 | | | | | 26687 | 169 | 26518 | | | Polycythemia, secondary | hematopoietic | 25DTot | |
| 289.9 | | | | | 18788 | 104 | 18684 | | | | | 26914 | 118 | 26796 | | | Abnormality of red blood cells | hematopoietic | 25DTot | |
| 290 | 0.8612 | 0.0794 | 2.366 | 2.09e-27 | 21683 | 1272 | 20411 | 0.432 | 0.06959 | 1.54 | 5.36e-10 | 30218 | 1222 | 28996 | 4.62e-05 | -0.4168 | Delirium dementia and amnestic and other cognitive disorders | mental disorders | 25DTot | |
| 290.1 | 0.8533 | 0.09894 | 2.347 | 6.46e-18 | 21205 | 794 | 20411 | 0.4786 | 0.08481 | 1.614 | 1.67e-08 | 29834 | 838 | 28996 | 3.88e-04 | -0.4481 | Dementias | mental disorders | 25DTot | |
| 290.11 | | | | | 20682 | 271 | 20411 | | | | | 29352 | 356 | 28996 | 0.04465 | -0.3875 | Alzheimer's disease | mental disorders | 25DTot | |
| 290.12 | | | | | 20506 | 95 | 20411 | | | | | 29043 | 47 | 28996 | | | Dementia with cerebral degenerations | mental disorders | 25DTot | |
| 290.13 | | | | | 20487 | 76 | 20411 | | | | | 29129 | 133 | 28996 | | | Senile dementia | mental disorders | 25DTot | |
| 290.16 | | | | | 20543 | 132 | 20411 | | | | | 29107 | 111 | 28996 | | | Vascular dementia | mental disorders | 25DTot | |
| 290.2 | | | | | 20877 | 466 | 20411 | | | | | 29341 | 345 | 28996 | 0.06297 | -0.312 | Delirium due to conditions classified elsewhere | mental disorders | 25DTot | |
| 290.3 | | | | | 20762 | 351 | 20411 | | | | | 29366 | 370 | 28996 | 3.53e-05 | -0.703 | Other persistent mental disorders due to conditions classified elsewhere | mental disorders | 25DTot | |
| 291 | 0.7804 | 0.07882 | 2.182 | 4.12e-23 | 21730 | 1319 | 20411 | 0.2602 | 0.06947 | 1.297 | 1.80e-04 | 30293 | 1297 | 28996 | 1.20e-05 | -0.4463 | Other specified nonpsychotic and/or transient mental disorders | mental disorders | 25DTot | |
| 291.1 | | | | | 20440 | 29 | 20411 | | | | | 29037 | 41 | 28996 | | | Transient mental disorders due to conditions classified elsewhere | mental disorders | 25DTot | |
| 291.4 | | | | | 20576 | 165 | 20411 | | | | | 29194 | 198 | 28996 | | | Specific nonpsychotic mental disorders due to brain damage | mental disorders | 25DTot | |
| 291.8 | 0.8321 | 0.08538 | 2.298 | 1.93e-22 | 21537 | 1126 | 20411 | 0.2432 | 0.07847 | 1.275 | 0.001938 | 30045 | 1049 | 28996 | 3.48e-06 | -0.5207 | Alteration of consciousness | mental disorders | 25DTot | |
| 292 | 0.5798 | 0.0511 | 1.786 | 7.76e-30 | 24322 | 3911 | 20411 | 0.2407 | 0.04035 | 1.272 | 2.45e-09 | 33173 | 4177 | 28996 | 2.89e-06 | -0.2983 | Neurological disorders | mental disorders | 25DTot | |
| 292.1 | 0.4942 | 0.1052 | 1.639 | 2.63e-06 | 21162 | 751 | 20411 | 0.3028 | 0.08518 | 1.354 | 3.78e-04 | 29761 | 765 | 28996 | 0.2792 | -0.143 | Aphasia/speech disturbance | mental disorders | 25DTot | |
| 292.11 | | | | | 20684 | 273 | 20411 | | | | | 29270 | 274 | 28996 | 0.9458 | -0.01511 | Aphasia | mental disorders | 25DTot | |
| 292.12 | | | | | 20439 | 28 | 20411 | | | | | 29020 | 24 | 28996 | | | Symbolic dysfunction | mental disorders | 25DTot | |
| 292.2 | | | | | 20778 | 367 | 20411 | | | | | 29334 | 338 | 28996 | 0.008117 | -0.461 | Mild cognitive impairment | mental disorders | 25DTot | |
| 292.3 | 0.7986 | 0.08609 | 2.222 | 1.77e-20 | 21327 | 916 | 20411 | 0.566 | 0.06668 | 1.761 | 2.09e-17 | 30127 | 1131 | 28996 | 0.02109 | -0.2458 | Memory loss | mental disorders | 25DTot | |
| 292.4 | 0.6747 | 0.06247 | 1.963 | 3.40e-27 | 22762 | 2351 | 20411 | 0.2429 | 0.05359 | 1.275 | 5.83e-06 | 31254 | 2258 | 28996 | 1.17e-06 | -0.3894 | Altered mental status | mental disorders | 25DTot | |
| 292.5 | | | | | 20547 | 136 | 20411 | | | | | 29184 | 188 | 28996 | | | Transient alteration of awareness | mental disorders | 25DTot | |
| 292.6 | | | | | 20491 | 80 | 20411 | | | | | 29129 | 133 | 28996 | | | Hallucinations | mental disorders | 25DTot | |
| 293 | | | | | 25542 | 80 | 25462 | | | | | 34286 | 108 | 34178 | | | Symptoms involving head and neck | mental disorders | 25DTot | |

| phenotype | male_beta | male_SE | male_OR | male_p | male_n_total | male_n_cases | male_n_controls | female_beta | female_SE | female_OR | female_p | female_n_total | female_n_cases | female_n_controls | interaction_p | interaction_beta | description | group | lab | label |
|---|---|---|---|---|---|---|---|---|---|---|---|---|---|---|---|---|---|---|---|---|
| 293.1 | 0.1032 | 0.1073 | 1.109 | 0.336 | 26351 | 889 | 25462 | 0.1232 | 0.08247 | 1.131 | 0.1352 | 35053 | 875 | 34178 | 0.2441 | 0.1539 | Swelling, mass, or lump in head and neck [Space-occupying lesion, intracranial NOS] | mental disorders | 25DTot | |
| 295 | | | | | 17073 | 479 | 16594 | 0.3796 | 0.1058 | 1.462 | 3.32e-04 | 19345 | 524 | 18821 | 0.9199 | -0.01675 | Schizophrenia and other psychotic disorders | mental disorders | 25DTot | |
| 295.1 | | | | | 16740 | 146 | 16594 | | | | | 18981 | 160 | 18821 | | | Schizophrenia | mental disorders | 25DTot | |
| 295.2 | | | | | 16700 | 106 | 16594 | | | | | 18968 | 147 | 18821 | | | Paranoid disorders | mental disorders | 25DTot | |
| 295.3 | | | | | 16878 | 284 | 16594 | | | | | 19094 | 273 | 18821 | 0.1906 | -0.2777 | Psychosis | mental disorders | 25DTot | |
| 296 | 0.6394 | 0.0502 | 1.895 | 3.71e-37 | 21203 | 4609 | 16594 | 0.4977 | 0.03437 | 1.645 | 1.56e-47 | 27169 | 8348 | 18821 | 0.4001 | -0.05053 | Mood disorders | mental disorders | 25DTot | |
| 296.1 | 0.5857 | 0.102 | 1.796 | 9.48e-09 | 17404 | 810 | 16594 | 0.2832 | 0.07197 | 1.327 | 8.31e-05 | 20203 | 1382 | 18821 | 0.1039 | -0.2001 | Bipolar | mental disorders | 25DTot | |
| 296.2 | 0.6805 | 0.05223 | 1.975 | 8.31e-39 | 20515 | 3921 | 16594 | 0.5453 | 0.0353 | 1.725 | 7.92e-54 | 26233 | 7412 | 18821 | 0.3348 | -0.06006 | Depression | mental disorders | 25DTot | |
| 296.22 | 0.64 | 0.06408 | 1.896 | 1.73e-23 | 18825 | 2231 | 16594 | 0.5525 | 0.04219 | 1.738 | 3.51e-39 | 23232 | 4411 | 18821 | 0.5877 | -0.0411 | Major depressive disorder | mental disorders | 25DTot | |
| 297 | | | | | 17005 | 411 | 16594 | 0.4459 | 0.1248 | 1.562 | 3.53e-04 | 19321 | 500 | 18821 | 0.812 | 0.04659 | Suicidal ideation or attempt | mental disorders | 25DTot | |
| 297.1 | | | | | 16944 | 350 | 16594 | | | | | 19222 | 401 | 18821 | 0.58 | 0.1176 | Suicidal ideation | mental disorders | 25DTot | |
| 297.2 | | | | | 16666 | 72 | 16594 | | | | | 18942 | 121 | 18821 | | | Suicide or self-inflicted injury | mental disorders | 25DTot | |
| 300 | 0.515 | 0.05412 | 1.674 | 1.80e-21 | 20603 | 4009 | 16594 | 0.399 | 0.03531 | 1.49 | 1.29e-29 | 27035 | 8214 | 18821 | 0.6484 | -0.02906 | Anxiety disorders | mental disorders | 25DTot | |
| 300.1 | 0.4813 | 0.05721 | 1.618 | 4.02e-17 | 20057 | 3463 | 16594 | 0.4097 | 0.03653 | 1.506 | 3.34e-29 | 26048 | 7227 | 18821 | 0.7718 | 0.01943 | Anxiety disorder | mental disorders | 25DTot | |
| 300.11 | 0.6591 | 0.1056 | 1.933 | 4.27e-10 | 17284 | 690 | 16594 | 0.4705 | 0.0664 | 1.601 | 1.39e-12 | 20323 | 1502 | 18821 | 0.2614 | -0.138 | Generalized anxiety disorder | mental disorders | 25DTot | |
| 300.12 | | | | | 16882 | 288 | 16594 | 0.4667 | 0.1033 | 1.595 | 6.21e-06 | 19405 | 584 | 18821 | 0.6113 | 0.1006 | Agorophobia, social phobia, and panic disorder | mental disorders | 25DTot | |
| 300.13 | | | | | 16650 | 56 | 16594 | | | | | 18930 | 109 | 18821 | | | Phobia | mental disorders | 25DTot | |
| 300.2 | | | | | 16594 | | | | | | | 18821 | | | | | Generalized anxiety & phobic disorders | mental disorders | 25DTot | |
| 300.3 | | | | | 16794 | 200 | 16594 | | | | | 19097 | 276 | 18821 | | | Obsessive-compulsive disorders | mental disorders | 25DTot | |
| phenotype | male_beta | male_SE | male_OR | male_p | male_n_total | male_n_cases | male_n_controls | female_beta | female_SE | female_OR | female_p | female_n_total | female_n_cases | female_n_controls | interaction_p | interaction_beta | description | group | lab | label |
| 300.4 | | | | | 16995 | 401 | 16594 | 0.6882 | 0.08224 | 1.99 | 5.84e-17 | 19615 | 794 | 18821 | 8.49e-06 | -0.6144 | Dysthymic disorder | mental disorders | 25DTot | |
| 300.8 | | | | | 16639 | 45 | 16594 | | | | | 18944 | 123 | 18821 | | | Acute reaction to stress | mental disorders | 25DTot | |
| 300.9 | | | | | 16936 | 342 | 16594 | 0.2583 | 0.09143 | 1.295 | 0.00473 | 19768 | 947 | 18821 | 0.1405 | -0.2633 | Posttraumatic stress disorder | mental disorders | 25DTot | |
| 301 | | | | | 16724 | 130 | 16594 | | | | | 19101 | 280 | 18821 | | | Personality disorders | mental disorders | 25DTot | |
| 301.1 | | | | | 16605 | 11 | 16594 | | | | | 18829 | 8 | 18821 | | | Schizoid personality disorder | mental disorders | 25DTot | |
| 301.2 | | | | | 16645 | 51 | 16594 | | | | | 19000 | 179 | 18821 | | | Antisocial/borderline personality disorder | mental disorders | 25DTot | |
| 302 | | | | | 16787 | 193 | 16594 | | | | | 18953 | 132 | 18821 | | | Sexual and gender identity disorders | mental disorders | 25DTot | |
| 302.1 | | | | | 16728 | 134 | 16594 | | | | | 18905 | 84 | 18821 | | | Decreased libido | mental disorders | 25DTot | |

| phenotype | male_beta | male_SE | male_OR | male_p | male_n_total | male_n_cases | male_n_controls | female_beta | female_SE | female_OR | female_p | female_n_total | female_n_cases | female_n_controls | interaction_p | interaction_beta | description | group | lab | label |
|---|---|---|---|---|---|---|---|---|---|---|---|---|---|---|---|---|---|---|---|---|
| 303 | | | | | 16730 | 136 | 16594 | | | | | 19170 | 349 | 18821 | | | Psychogenic and somatoform disorders | mental disorders | 25DTot | |
| 303.1 | | | | | 16603 | 9 | 16594 | | | | | 18849 | 28 | 18821 | | | Dissociative disorder | mental disorders | 25DTot | |
| 303.3 | | | | | 16631 | 37 | 16594 | | | | | 18918 | 97 | 18821 | | | Psychogenic disorder | mental disorders | 25DTot | |
| 303.31 | | | | | 16597 | 3 | 16594 | | | | | 18830 | 9 | 18821 | | | Gastrointestinal malfunction arising from mental factors | mental disorders | 25DTot | |
| 303.4 | | | | | 16689 | 95 | 16594 | | | | | 19064 | 243 | 18821 | | | Somatoform disorder | mental disorders | 25DTot | |
| 304 | 0.5086 | 0.09627 | 1.663 | 1.27e-07 | 17547 | 953 | 16594 | 0.6119 | 0.05921 | 1.844 | 4.91e-25 | 20609 | 1788 | 18821 | 0.04174 | 0.2271 | Adjustment reaction | mental disorders | 25DTot | |
| 305.2 | | | | | 16644 | 50 | 16594 | | | | | 19240 | 419 | 18821 | | | Eating disorder | mental disorders | 25DTot | |
| 305.21 | | | | | 16607 | 13 | 16594 | | | | | 18929 | 108 | 18821 | | | Anorexia nervosa | mental disorders | 25DTot | |
| 306 | 0.2532 | 0.09415 | 1.288 | 0.00715 | 17668 | 1074 | 16594 | 0.4829 | 0.06096 | 1.621 | 2.34e-15 | 20481 | 1660 | 18821 | 0.03711 | 0.231 | Other mental disorder | mental disorders | 25DTot | |
| 306.1 | | | | | 0 | | | | | | | 19216 | 395 | 18821 | | | Mental disorders durring/after pregnancy | mental disorders | 25DTot | |
| 306.9 | | | | | 16626 | 32 | 16594 | | | | | 18928 | 107 | 18821 | | | Tension headache | mental disorders | 25DTot | |
| 312 | | | | | 25880 | 213 | 25667 | | | | | 34967 | 157 | 34810 | | | Conduct disorders | mental disorders | 25DTot | |
| 312.3 | | | | | 25729 | 62 | 25667 | | | | | 34851 | 41 | 34810 | | | Impulse control disorder | mental disorders | 25DTot | |
| 313 | -0.174 | 0.1473 | 0.8403 | 0.2374 | 26378 | 711 | 25667 | 0.1912 | 0.1065 | 1.211 | 0.0726 | 35536 | 726 | 34810 | 0.002455 | 0.5359 | Pervasive developmental disorders | mental disorders | 25DTot | |
| 313.1 | -0.1885 | 0.1712 | 0.8282 | 0.2708 | 26178 | 511 | 25667 | 0.237 | 0.1214 | 1.267 | 0.05094 | 35352 | 542 | 34810 | 0.005557 | 0.5671 | Attention deficit hyperactivity disorder | mental disorders | 25DTot | |
| 313.2 | | | | | 25726 | 59 | 25667 | | | | | 34849 | 39 | 34810 | | | Tics and stuttering | mental disorders | 25DTot | |
| 313.3 | | | | | 25854 | 187 | 25667 | | | | | 34891 | 81 | 34810 | | | Autism | mental disorders | 25DTot | |
| 315 | -0.1311 | 0.144 | 0.8771 | 0.3624 | 26237 | 570 | 25667 | | | | | 35246 | 436 | 34810 | 0.4907 | -0.1376 | Develomental delays and disorders | mental disorders | 25DTot | |
| 315.1 | | | | | 25693 | 26 | 25667 | | | | | 34830 | 20 | 34810 | | | Learning disorder | mental disorders | 25DTot | |
| 315.2 | | | | | 25753 | 86 | 25667 | | | | | 34881 | 71 | 34810 | | | Speech and language disorder | mental disorders | 25DTot | |
| 315.3 | | | | | 25849 | 182 | 25667 | | | | | 34981 | 171 | 34810 | | | Mental retardation | mental disorders | 25DTot | |
| 316 | 0.3459 | 0.08057 | 1.413 | 1.76e-05 | 19622 | 1399 | 18223 | -0.07782 | 0.06476 | 0.9251 | 0.2295 | 29618 | 1741 | 27877 | 9.69e-05 | -0.3947 | Substance addiction and disorders | mental disorders | 25DTot | |
| 316.1 | | | | | 18355 | 132 | 18223 | | | | | 28169 | 292 | 27877 | | | Polyneuropathy due to drugs | mental disorders | 25DTot | |
| 317 | 0.2476 | 0.08493 | 1.281 | 0.003558 | 19570 | 1347 | 18223 | -0.01344 | 0.101 | 0.9866 | 0.8941 | 28525 | 648 | 27877 | 0.1046 | -0.2086 | Alcohol-related disorders | mental disorders | 25DTot | |
| 317.1 | 0.3024 | 0.09815 | 1.353 | 0.002065 | 19193 | 970 | 18223 | | | | | 28321 | 444 | 27877 | 0.1187 | -0.2378 | Alcoholism | mental disorders | 25DTot | |
| 317.11 | | | | | 18678 | 455 | 18223 | | | | | 28037 | 160 | 27877 | 0.002414 | -0.7541 | Alcoholic liver damage | mental disorders | 25DTot | |
| 318 | 0.02857 | 0.05083 | 1.029 | 0.5741 | 23438 | 5215 | 18223 | -0.1197 | 0.04063 | 0.8872 | 0.003218 | 32604 | 4727 | 27877 | 0.003879 | -0.1839 | Tobacco use disorder | mental disorders | 25DTot | |
| 320 | | | | | 27253 | 157 | 27096 | | | | | 36416 | 155 | 36261 | | | Meningitis | neurological | 25DTot | |
| 323 | | | | | 27190 | 94 | 27096 | | | | | 36377 | 116 | 36261 | | | Encephalitis | neurological | 25DTot | |
| 323.2 | | | | | 27111 | 15 | 27096 | | | | | 36289 | 28 | 36261 | | | Acute (transverse) myelitis | neurological | 25DTot | |
| 323.8 | | | | | 27165 | 69 | 27096 | | | | | 36347 | 86 | 36261 | | | Encephalitis, non-infectious | neurological | 25DTot | |

| phenotype | male_beta | male_SE | male_OR | male_p | male_n_total | male_n_cases | male_n_controls | female_beta | female_SE | female_OR | female_p | female_n_total | female_n_cases | female_n_controls | interaction_p | interaction_beta | description | group | lab | label |
|---|---|---|---|---|---|---|---|---|---|---|---|---|---|---|---|---|---|---|---|---|
| 324 | | | | | 27283 | 187 | 27096 | | | | | 36381 | 120 | 36261 | | | Other CNS infection and poliomyelitis | neurological | 25DTot | |
| 324.1 | | | | | 27097 | 1 | 27096 | | | | | 36261 | | | | | Jakob-Creutzfeldt disease | neurological | 25DTot | |
| 325 | | | | | 27112 | 16 | 27096 | | | | | 36306 | 45 | 36261 | | | Phlebitis and thrombophlebitis of intracranial venous sinuses | neurological | 25DTot | |
| 327 | 0.5994 | 0.05471 | 1.821 | 6.19e-28 | 22679 | 3201 | 19478 | 0.6914 | 0.03701 | 1.997 | 7.12e-78 | 31789 | 4772 | 27017 | 0.02862 | 0.143 | Sleep disorders | neurological | 25DTot | |
| 327.1 | | | | | 19663 | 185 | 19478 | | | | | 27358 | 341 | 27017 | 0.1338 | 0.3339 | Hypersomnia | neurological | 25DTot | |
| 327.3 | 0.4804 | 0.05272 | 1.617 | 8.09e-20 | 23346 | 3868 | 19478 | 0.5097 | 0.04396 | 1.665 | 4.42e-31 | 30225 | 3208 | 27017 | 0.3444 | 0.06403 | Sleep apnea | neurological | 25DTot | |
| 327.31 | | | | | 19588 | 110 | 19478 | | | | | 27080 | 63 | 27017 | | | Central/nonobstructive sleep apnea | neurological | 25DTot | |
| 327.32 | 0.5283 | 0.05469 | 1.696 | 4.48e-22 | 22855 | 3377 | 19478 | 0.5398 | 0.0469 | 1.716 | 1.20e-30 | 29718 | 2701 | 27017 | 0.5744 | 0.03991 | Obstructive sleep apnea | neurological | 25DTot | |
| 327.4 | 0.6657 | 0.0651 | 1.946 | 1.53e-24 | 21429 | 1951 | 19478 | 0.7383 | 0.04319 | 2.092 | 1.64e-65 | 30146 | 3129 | 27017 | 0.05345 | 0.1493 | Insomnia | neurological | 25DTot | |
| 327.41 | | | | | 19695 | 217 | 19478 | | | | | 27448 | 431 | 27017 | 0.5213 | 0.1183 | Organic or persistent insomnia | neurological | 25DTot | |
| 327.5 | | | | | 19615 | 137 | 19478 | | | | | 27116 | 99 | 27017 | | | Parasomnia | neurological | 25DTot | |
| 327.6 | | | | | 19505 | 27 | 19478 | | | | | 27055 | 38 | 27017 | | | Circadian rhythm sleep disorder | neurological | 25DTot | |
| 327.7 | 0.8326 | 0.1106 | 2.299 | 5.27e-14 | 19996 | 518 | 19478 | 0.7072 | 0.07084 | 2.028 | 1.81e-23 | 28011 | 994 | 27017 | 0.3723 | -0.1153 | Sleep related movement disorders | neurological | 25DTot | |
| 327.71 | | | | | 19923 | 445 | 19478 | 0.6656 | 0.07527 | 1.946 | 9.38e-19 | 27896 | 879 | 27017 | 0.1746 | -0.1868 | Restless legs syndrome | neurological | 25DTot | |
| 327.72 | | | | | 19495 | 17 | 19478 | | | | | 27043 | 26 | 27017 | | | Sleep related leg cramps | neurological | 25DTot | |
| 331 | 0.5862 | 0.1239 | 1.797 | 2.22e-06 | 20649 | 514 | 20135 | | | | | 28193 | 478 | 27715 | 0.007495 | -0.4422 | Other cerebral degenerations | neurological | 25DTot | |
| 331.1 | | | | | 20475 | 340 | 20135 | | | | | 28036 | 321 | 27715 | 0.03583 | -0.5538 | Hydrocephalus | neurological | 25DTot | |
| 331.9 | | | | | 20281 | 146 | 20135 | | | | | 27836 | 121 | 27715 | | | Cerebral degeneration, unspecified | neurological | 25DTot | |
| 332 | | | | | 20573 | 438 | 20135 | | | | | 27974 | 259 | 27715 | 0.1405 | -0.2791 | Parkinson's disease | neurological | 25DTot | |
| 333 | 0.5642 | 0.09561 | 1.758 | 3.60e-09 | 20955 | 820 | 20135 | 0.3592 | 0.07537 | 1.432 | 1.88e-06 | 28634 | 919 | 27715 | 0.1279 | -0.1818 | Extrapyramidal disease and abnormal movement disorders | neurological | 25DTot | |
| 333.1 | | | | | 20501 | 366 | 20135 | | | | | 28134 | 419 | 27715 | 0.192 | -0.2239 | Essential tremor | neurological | 25DTot | |

| phenotype | male_beta | male_SE | male_OR | male_p | male_n_total | male_n_cases | male_n_controls | female_beta | female_SE | female_OR | female_p | female_n_total | female_n_cases | female_n_controls | interaction_p | interaction_beta | description | group | lab | label |
|---|---|---|---|---|---|---|---|---|---|---|---|---|---|---|---|---|---|---|---|---|
| 333.2 | | | | | 20258 | 123 | 20135 | | | | | 27820 | 105 | 27715 | | | Myoclonus | neurological | 25DTot | |
| 333.3 | | | | | 20164 | 29 | 20135 | | | | | 27748 | 33 | 27715 | | | Tics and choreas | neurological | 25DTot | |
| 333.4 | | | | | 20284 | 149 | 20135 | | | | | 28005 | 290 | 27715 | | | Torsion dystonia | neurological | 25DTot | |
| 333.8 | | | | | 20218 | 83 | 20135 | | | | | 27746 | 31 | 27715 | | | Other degenerative diseases of the basal ganglia | neurological | 25DTot | |
| 334 | | | | | 20526 | 391 | 20135 | | | | | 28145 | 430 | 27715 | 0.4501 | -0.1389 | Degenerative disease of the spinal cord | neurological | 25DTot | |
| 334.1 | | | | | 20181 | 46 | 20135 | | | | | 27786 | 71 | 27715 | | | Spinocerebellar disease | neurological | 25DTot | |
| 334.2 | | | | | 20201 | 66 | 20135 | | | | | 27768 | 53 | 27715 | | | Anterior horn cell disease | neurological | 25DTot | |
| 334.21 | | | | | 20183 | 48 | 20135 | | | | | 27759 | 44 | 27715 | | | Amyotrophic Lateral Sclerosis | neurological | 25DTot | |
| 335 | | | | | 20445 | 310 | 20135 | 1.205 | 0.0705 | 3.338 | 1.56e-65 | 28694 | 979 | 27715 | 1.12e-06 | -0.6636 | Multiple sclerosis | neurological | 25DTot | |
| 337 | | | | | 20390 | 255 | 20135 | | | | | 28028 | 313 | 27715 | 0.005071 | -0.5516 | Disorders of the autonomic nervous system | neurological | 25DTot | |
| 337.1 | | | | | 20203 | 68 | 20135 | | | | | 27793 | 78 | 27715 | | | Peripheral autonomic neuropathy | neurological | 25DTot | |
| 338 | 0.1922 | 0.04757 | 1.212 | 5.33e-05 | 24967 | 5863 | 19104 | 0.2354 | 0.03302 | 1.265 | 1.03e-12 | 33179 | 7691 | 25488 | 0.0066 | 0.155 | Pain | neurological | 25DTot | |
| 338.1 | 0.1397 | 0.05626 | 1.15 | 0.013 | 23126 | 4022 | 19104 | 0.1321 | 0.04009 | 1.141 | 9.82e-04 | 30282 | 4794 | 25488 | 0.07349 | 0.1216 | Acute pain | neurological | 25DTot | |
| 338.2 | 0.3694 | 0.05949 | 1.447 | 5.33e-10 | 21816 | 2712 | 19104 | 0.4424 | 0.0394 | 1.556 | 2.93e-29 | 29799 | 4311 | 25488 | 0.07341 | 0.1263 | Chronic pain | neurological | 25DTot | |
| 339 | 0.196 | 0.1115 | 1.217 | 0.07872 | 25998 | 701 | 25297 | 0.3986 | 0.06266 | 1.49 | 2.01e-10 | 31803 | 1472 | 30331 | 0.03615 | 0.2634 | Other headache syndromes | neurological | 25DTot | |

| phenotype | male_beta | male_SE | male_OR | male_p | male_n_total | male_n_cases | male_n_controls | female_beta | female_SE | female_OR | female_p | female_n_total | female_n_cases | female_n_controls | interaction_p | interaction_beta | description | group | lab | label |
|---|---|---|---|---|---|---|---|---|---|---|---|---|---|---|---|---|---|---|---|---|
| 340 | 0.5223 | 0.1034 | 1.686 | 4.36e-07 | 25980 | 683 | 25297 | 0.4686 | 0.04743 | 1.598 | 4.99e-23 | 33186 | 2855 | 30331 | 0.7185 | -0.04021 | Migraine | neurological | 25DTot | |
| 340.1 | | | | | 25434 | 137 | 25297 | 0.6378 | 0.08591 | 1.892 | 1.14e-13 | 31063 | 732 | 30331 | 0.1691 | 0.3415 | Migrain with aura | neurological | 25DTot | |
| 341 | | | | | 20276 | 141 | 20135 | | | | | 28206 | 491 | 27715 | 0.08171 | -0.34 | Other demyelinating diseases of central nervous system | neurological | 25DTot | |
| 342 | | | | | 20493 | 358 | 20135 | | | | | 28011 | 296 | 27715 | 0.6314 | -0.09994 | Hemiplegia | neurological | 25DTot | |
| 343 | | | | | 20279 | 144 | 20135 | | | | | 27844 | 129 | 27715 | | | Infantile cerebral palsy | neurological | 25DTot | |
| 344 | | | | | 20460 | 325 | 20135 | | | | | 27997 | 282 | 27715 | 0.3374 | -0.2239 | Other paralytic syndromes | neurological | 25DTot | |
| 345 | 0.2482 | 0.08219 | 1.282 | 0.002533 | 21729 | 1594 | 20135 | -0.01214 | 0.06401 | 0.9879 | 0.8496 | 29581 | 1866 | 27715 | 0.0618 | -0.1906 | Epilepsy, recurrent seizures, convulsions | neurological | 25DTot | |
| 345.1 | 0.03816 | 0.1284 | 1.039 | 0.7663 | 20847 | 712 | 20135 | -0.1551 | 0.1032 | 0.8564 | 0.1329 | 28454 | 739 | 27715 | 0.7778 | -0.04561 | Epilepsy | neurological | 25DTot | |
| 345.11 | | | | | 20451 | 316 | 20135 | | | | | 28000 | 285 | 27715 | 0.009081 | -0.6512 | Generalized convulsive epilepsy | neurological | 25DTot | |
| 345.12 | -0.07864 | 0.1569 | 0.9244 | 0.6162 | 20638 | 503 | 20135 | | | | | 28179 | 464 | 27715 | 0.4946 | 0.1353 | Partial epilepsy | neurological | 25DTot | |
| 345.3 | 0.293 | 0.08951 | 1.34 | 0.001064 | 21418 | 1283 | 20135 | -0.01442 | 0.07158 | 0.9857 | 0.8403 | 29200 | 1485 | 27715 | 0.03657 | -0.2347 | Convulsions | neurological | 25DTot | |
| 346 | | | | | 20172 | 37 | 20135 | | | | | 27787 | 72 | 27715 | | | Abnormal findings on study of brain and/or nervous system | neurological | 25DTot | |
| 346.1 | | | | | 20298 | 163 | 20135 | | | | | 27984 | 269 | 27715 | | | Nonspecific abnormal findings on radiological and other examination of skull and head | neurological | 25DTot | |
| 346.2 | | | | | 20163 | 28 | 20135 | | | | | 27765 | 50 | 27715 | | | Nonspecific abnormal results of function study of brain and central nervous system | neurological | 25DTot | |
| 346.3 | | | | | 20173 | 38 | 20135 | | | | | 27758 | 43 | 27715 | | | Nonspecific abnormal findings in cerebrospinal fluid | neurological | 25DTot | |
| 347 | | | | | 20161 | 26 | 20135 | | | | | 27775 | 60 | 27715 | | | Cataplexy and narcolepsy | neurological | 25DTot | |
| 348 | 0.491 | 0.07819 | 1.634 | 3.40e-10 | 21644 | 1509 | 20135 | 0.1206 | 0.06673 | 1.128 | 0.07083 | 29223 | 1508 | 27715 | 8.07e-04 | -0.3362 | Other conditions of brain | neurological | 25DTot | |
| 348.2 | -0.09749 | 0.1702 | 0.9071 | 0.5669 | 20647 | 512 | 20135 | -0.235 | 0.1227 | 0.7906 | 0.05555 | 28310 | 595 | 27715 | 0.698 | -0.08015 | Cerebral edema and compression of brain | neurological | 25DTot | |
| 348.4 | | | | | 20194 | 59 | 20135 | | | | | 27769 | 54 | 27715 | | | Cerebral cysts | neurological | 25DTot | |
| 348.7 | | | | | 20262 | 127 | 20135 | | | | | 27823 | 108 | 27715 | | | Coma | neurological | 25DTot | |
| 348.8 | 0.7207 | 0.08834 | 2.056 | 3.40e-16 | 21082 | 947 | 20135 | 0.3015 | 0.07953 | 1.352 | 1.50e-04 | 28610 | 895 | 27715 | 9.48e-04 | -0.3831 | Encephalopathy, not elsewhere classified | neurological | 25DTot | |
| 348.9 | 0.2823 | 0.1176 | 1.326 | 0.01633 | 20916 | 781 | 20135 | -0.02465 | 0.09973 | 0.9757 | 0.8048 | 28459 | 744 | 27715 | 0.1177 | -0.2359 | Other conditions of brain, NOS | neurological | 25DTot | |
| 349 | | | | | 20369 | 234 | 20135 | | | | | 27969 | 254 | 27715 | | | Other and unspecified disorders of the nervous system | neurological | 25DTot | |
| 350 | 0.6259 | 0.05573 | 1.87 | 2.90e-29 | 25783 | 2709 | 23074 | 0.6112 | 0.04059 | 1.843 | 3.13e-51 | 34411 | 3494 | 30917 | 0.8602 | 0.01197 | Abnormal movement | neurological | 25DTot | |
| 350.1 | 0.4423 | 0.09823 | 1.556 | 6.70e-06 | 23884 | 810 | 23074 | 0.5489 | 0.06812 | 1.731 | 7.80e-16 | 32013 | 1096 | 30917 | 0.1474 | 0.1706 | Abnormal involuntary movements | neurological | 25DTot | |
| 350.2 | 0.7367 | 0.06463 | 2.089 | 4.28e-30 | 24868 | 1794 | 23074 | 0.631 | 0.04818 | 1.88 | 3.38e-39 | 33227 | 2310 | 30917 | 0.3204 | -0.07895 | Abnormality of gait | neurological | 25DTot | |
| 350.3 | 1.083 | 0.1025 | 2.955 | 4.32e-26 | 23605 | 531 | 23074 | 1.117 | 0.07444 | 3.055 | 7.19e-51 | 31754 | 837 | 30917 | 0.7935 | 0.03249 | Lack of coordination | neurological | 25DTot | |
| 350.5 | | | | | 23103 | 29 | 23074 | | | | | 30953 | 36 | 30917 | | | Abnormal reflex | neurological | 25DTot | |
| 350.6 | | | | | 23183 | 109 | 23074 | | | | | 31041 | 124 | 30917 | | | Disturbances of sensation of smell and taste | neurological | 25DTot | |
| 351 | 0.5301 | 0.07512 | 1.699 | 1.71e-12 | 25090 | 1362 | 23728 | 0.4592 | 0.04988 | 1.583 | 3.36e-20 | 32761 | 2244 | 30517 | 0.9554 | -0.004949 | Other peripheral nerve disorders | neurological | 25DTot | |
| 352 | | | | | 24026 | 298 | 23728 | 0.3652 | 0.09841 | 1.441 | 2.07e-04 | 31061 | 544 | 30517 | 0.9654 | 0.007961 | Disorders of other cranial nerves | neurological | 25DTot | |
| 352.1 | | | | | 23876 | 148 | 23728 | | | | | 30870 | 353 | 30517 | 0.1944 | 0.362 | Trigeminal nerve disorders [CN5] | neurological | 25DTot | |
| 352.2 | | | | | 23861 | 133 | 23728 | | | | | 30696 | 179 | 30517 | | | Facial nerve disorders [CN7] | neurological | 25DTot | |

| phenotype | male_beta | male_SE | male_OR | male_p | male_n_total | male_n_cases | male_n_controls | female_beta | female_SE | female_OR | female_p | female_n_total | female_n_cases | female_n_controls | interaction_p | interaction_beta | description | group | lab | label |
|---|---|---|---|---|---|---|---|---|---|---|---|---|---|---|---|---|---|---|---|---|
| 353 | | | | | 23928 | 200 | 23728 | | | | | 30746 | 229 | 30517 | | | Nerve root and plexus disorders | neurological | 25DTot | |
| 353.1 | | | | | 23805 | 77 | 23728 | | | | | 30605 | 88 | 30517 | | | Nerve plexus lesions | neurological | 25DTot | |
| 353.2 | | | | | 23800 | 72 | 23728 | | | | | 30603 | 86 | 30517 | | | Nerve root lesions | neurological | 25DTot | |
| 355 | | | | | 23869 | 141 | 23728 | | | | | 30825 | 308 | 30517 | | | Complex regional/central pain syndrome | neurological | 25DTot | |
| 355.1 | 0.6377 | 0.1048 | 1.892 | 1.18e-09 | 24316 | 588 | 23728 | 0.7199 | 0.06877 | 2.054 | 1.22e-25 | 31540 | 1023 | 30517 | 0.4442 | 0.09426 | Chronic pain syndrome | neurological | 25DTot | |
| 356 | 0.9879 | 0.07914 | 2.686 | 9.23e-36 | 24919 | 1003 | 23916 | 0.687 | 0.0697 | 1.988 | 6.48e-23 | 33295 | 1002 | 32293 | 0.003489 | -0.3001 | Hereditary and idiopathic peripheral neuropathy | neurological | 25DTot | |
| 357 | 0.5501 | 0.08255 | 1.733 | 2.66e-11 | 24993 | 1077 | 23916 | 0.446 | 0.0631 | 1.562 | 1.57e-12 | 33576 | 1283 | 32293 | 0.1252 | -0.1561 | Inflammatory and toxic neuropathy | neurological | 25DTot | |
| 358 | | | | | 24025 | 109 | 23916 | | | | | 32462 | 169 | 32293 | | | Myoneural disorders | neurological | 25DTot | |
| 358.1 | | | | | 23987 | 71 | 23916 | | | | | 32384 | 91 | 32293 | | | Myasthenia gravis | neurological | 25DTot | |
| 359 | 0.9635 | 0.102 | 2.621 | 3.40e-21 | 24562 | 646 | 23916 | 0.565 | 0.09355 | 1.759 | 1.55e-09 | 32892 | 599 | 32293 | 0.002412 | -0.4054 | Muscular dystrophies and other myopathies | neurological | 25DTot | |
| 359.1 | | | | | 23949 | 33 | 23916 | | | | | 32328 | 35 | 32293 | | | Muscular dystrophies | neurological | 25DTot | |
| 359.2 | 1.035 | 0.1029 | 2.816 | 7.93e-24 | 24538 | 622 | 23916 | 0.6137 | 0.09535 | 1.847 | 1.22e-10 | 32864 | 571 | 32293 | 0.001721 | -0.4236 | Myopathy | neurological | 25DTot | |

| phenotype | male_beta | male_SE | male_OR | male_p | male_n_total | male_n_cases | male_n_controls | female_beta | female_SE | female_OR | female_p | female_n_total | female_n_cases | female_n_controls | interaction_p | interaction_beta | description | group | lab | label |
|---|---|---|---|---|---|---|---|---|---|---|---|---|---|---|---|---|---|---|---|---|
| 360 | | | | | 24749 | 89 | 24660 | | | | | 33075 | 87 | 32988 | | | Disorders of the globe | sense organs | 25DTot | |
| 360.2 | | | | | 24707 | 47 | 24660 | | | | | 33042 | 54 | 32988 | | | Progressive myopia | sense organs | 25DTot | |
| 360.3 | | | | | 24670 | 10 | 24660 | | | | | 33001 | 13 | 32988 | | | Hypotony of eye | sense organs | 25DTot | |
| 361 | | | | | 24928 | 268 | 24660 | | | | | 33163 | 175 | 32988 | | | Retinal detachments and defects | sense organs | 25DTot | |
| 361.1 | | | | | 24821 | 161 | 24660 | | | | | 33064 | 76 | 32988 | | | Retinal detachment with retinal defect | sense organs | 25DTot | |
| 361.2 | | | | | 24678 | 18 | 24660 | | | | | 32996 | 8 | 32988 | | | Retinoschisis and retinal cysts | sense organs | 25DTot | |
| 362 | 0.5535 | 0.0821 | 1.739 | 1.56e-11 | 25441 | 1087 | 24354 | 0.5192 | 0.06096 | 1.681 | 1.62e-17 | 34109 | 1379 | 32730 | 0.6209 | -0.04971 | Other retinal disorders | sense organs | 25DTot | |
| 362.1 | | | | | 24369 | 15 | 24354 | | | | | 32743 | 13 | 32730 | | | Retinopathy of prematurity | sense organs | 25DTot | |
| 362.2 | 0.6118 | 0.1058 | 1.844 | 7.38e-09 | 24974 | 620 | 24354 | 0.4609 | 0.07906 | 1.586 | 5.54e-09 | 33549 | 819 | 32730 | 0.1666 | -0.1793 | Degeneration of macula and posterior pole of retina | sense organs | 25DTot | |
| 362.21 | | | | | 24463 | 109 | 24354 | | | | | 32881 | 151 | 32730 | | | Macular degeneration, dry | sense organs | 25DTot | |
| 362.22 | | | | | 24410 | 56 | 24354 | | | | | 32819 | 89 | 32730 | | | Macular degeneration, wet | sense organs | 25DTot | |
| 362.23 | | | | | 24511 | 157 | 24354 | | | | | 32900 | 170 | 32730 | | | Cystoid macular degeneration of retina | sense organs | 25DTot | |
| 362.26 | | | | | 24590 | 236 | 24354 | | | | | 32963 | 233 | 32730 | | | Macular puckering of retina | sense organs | 25DTot | |
| 362.27 | | | | | 24433 | 79 | 24354 | | | | | 32875 | 145 | 32730 | | | Drusen (degenerative) of retina | sense organs | 25DTot | |
| 362.29 | | | | | 24520 | 166 | 24354 | | | | | 33004 | 274 | 32730 | | | Macular degeneration (senile) of retina NOS | sense organs | 25DTot | |
| 362.3 | | | | | 24535 | 181 | 24354 | | | | | 32906 | 176 | 32730 | | | Other nondiabetic retinopathy | sense organs | 25DTot | |
| 362.31 | | | | | 24393 | 39 | 24354 | | | | | 32753 | 23 | 32730 | | | Separation of retinal layers | sense organs | 25DTot | |
| 362.4 | | | | | 24528 | 174 | 24354 | | | | | 32939 | 209 | 32730 | | | Retinal vascular changes and abnormalities | sense organs | 25DTot | |
| 362.5 | | | | | 24369 | 15 | 24354 | | | | | 32823 | 93 | 32730 | | | Toxic maculopathy of retina | sense organs | 25DTot | |
| 362.6 | | | | | 24414 | 60 | 24354 | | | | | 32796 | 66 | 32730 | | | Peripheral retinal degenerations | sense organs | 25DTot | |
| 362.7 | | | | | 24401 | 47 | 24354 | | | | | 32775 | 45 | 32730 | | | Hereditary retinal dystrophies | sense organs | 25DTot | |
| 362.8 | | | | | 24401 | 47 | 24354 | | | | | 32774 | 44 | 32730 | | | Retinal hemorrhage/ischemia | sense organs | 25DTot | |
| 362.9 | | | | | 24469 | 115 | 24354 | | | | | 32862 | 132 | 32730 | | | Retinal edema | sense organs | 25DTot | |
| 363 | | | | | 24770 | 110 | 24660 | | | | | 33134 | 146 | 32988 | | | Chorioretinal inflammations, scars, and other disorders of choroid | sense organs | 25DTot | |
| 363.3 | | | | | 24721 | 61 | 24660 | | | | | 33072 | 84 | 32988 | | | Chorioretinal scars | sense organs | 25DTot | |
| 363.4 | | | | | 24662 | 2 | 24660 | | | | | 32995 | 7 | 32988 | | | Choroidal degenerations | sense organs | 25DTot | |
| 364 | | | | | 24841 | 181 | 24660 | | | | | 33258 | 270 | 32988 | | | Corneal opacity and other disorders of cornea | sense organs | 25DTot | |

| phenotype | male_beta | male_SE | male_OR | male_p | male_n_total | male_n_cases | male_n_controls | female_beta | female_SE | female_OR | female_p | female_n_total | female_n_cases | female_n_controls | interaction_p | interaction_beta | description | group | lab | label |
|---|---|---|---|---|---|---|---|---|---|---|---|---|---|---|---|---|---|---|---|---|
| 364.1 | | | | | 24753 | 93 | 24660 | | | | | 33113 | 125 | 32988 | | | Corneal opacity | sense organs | 25DTot | |
| 364.2 | | | | | 24712 | 52 | 24660 | | | | | 33031 | 43 | 32988 | | | Corneal edema | sense organs | 25DTot | |
| 364.4 | | | | | 24715 | 55 | 24660 | | | | | 33057 | 69 | 32988 | | | Corneal degenerations | sense organs | 25DTot | |
| 364.41 | | | | | 24684 | 24 | 24660 | | | | | 33006 | 18 | 32988 | | | Keratoconus | sense organs | 25DTot | |
| 364.5 | | | | | 24720 | 60 | 24660 | | | | | 33111 | 123 | 32988 | | | Corneal dystrophy | sense organs | 25DTot | |
| 364.51 | | | | | 24699 | 39 | 24660 | | | | | 33061 | 73 | 32988 | | | Fuchs' dystrophy | sense organs | 25DTot | |
| 364.9 | | | | | 24688 | 28 | 24660 | | | | | 33020 | 32 | 32988 | | | Cornea replaced by transplant | sense organs | 25DTot | |
| 365 | 0.5555 | 0.08779 | 1.743 | 2.49e-10 | 25576 | 916 | 24660 | 0.5054 | 0.06603 | 1.658 | 1.95e-14 | 34125 | 1137 | 32988 | 0.6089 | -0.05521 | Glaucoma | sense organs | 25DTot | |
| 365.1 | 0.685 | 0.1064 | 1.984 | 1.21e-10 | 25218 | 558 | 24660 | 0.6563 | 0.0828 | 1.928 | 2.26e-15 | 33666 | 678 | 32988 | 0.7503 | -0.04213 | Open-angle glaucoma | sense organs | 25DTot | |
| 365.11 | | | | | 24878 | 218 | 24660 | | | | | 33217 | 229 | 32988 | | | Primary open angle glaucoma | sense organs | 25DTot | |
| 365.2 | | | | | 24753 | 93 | 24660 | | | | | 33132 | 144 | 32988 | | | Primary angle-closure glaucoma | sense organs | 25DTot | |
| 365.5 | | | | | 24671 | 11 | 24660 | | | | | 33004 | 16 | 32988 | | | Pseudoexfoliation glaucoma | sense organs | 25DTot | |
| 366 | 0.4717 | 0.05811 | 1.603 | 4.76e-16 | 26713 | 2883 | 23830 | 0.6524 | 0.04124 | 1.92 | 2.34e-56 | 35649 | 3722 | 31927 | 0.03149 | 0.1518 | Cataract | sense organs | 25DTot | |
| 366.1 | | | | | 23845 | 15 | 23830 | | | | | 31936 | 9 | 31927 | | | Nonsenile Cataract | sense organs | 25DTot | |
| 366.2 | 0.5099 | 0.06087 | 1.665 | 5.44e-17 | 26293 | 2463 | 23830 | 0.6698 | 0.04366 | 1.954 | 4.01e-53 | 35061 | 3134 | 31927 | 0.064 | 0.1374 | Senile cataract | sense organs | 25DTot | |
| 366.3 | | | | | 23835 | 5 | 23830 | | | | | 31932 | 5 | 31927 | | | Traumatic cataract | sense organs | 25DTot | |
| 367 | 0.4921 | 0.07725 | 1.636 | 1.89e-10 | 26602 | 1195 | 25407 | 0.5842 | 0.05094 | 1.794 | 1.88e-30 | 35278 | 2096 | 33182 | 0.5435 | 0.05514 | Disorders of refraction and accommodation; blindness and low vision | sense organs | 25DTot | |
| 367.1 | 0.6034 | 0.1052 | 1.828 | 9.70e-09 | 25991 | 584 | 25407 | 0.7352 | 0.06485 | 2.086 | 8.70e-30 | 34437 | 1255 | 33182 | 0.6028 | 0.06289 | Myopia | sense organs | 25DTot | |
| 367.2 | | | | | 25731 | 324 | 25407 | 0.6402 | 0.09571 | 1.897 | 2.24e-11 | 33701 | 519 | 33182 | 0.8817 | -0.02408 | Astigmatism | sense organs | 25DTot | |
| 367.8 | | | | | 25506 | 99 | 25407 | | | | | 33431 | 249 | 33182 | | | Hypermetropia | sense organs | 25DTot | |
| 367.9 | | | | | 25707 | 300 | 25407 | | | | | 33524 | 342 | 33182 | 0.07953 | -0.3306 | Blindness and low vision | sense organs | 25DTot | |
| 368 | 0.5552 | 0.08367 | 1.742 | 3.24e-11 | 26249 | 990 | 25259 | 0.4954 | 0.05955 | 1.641 | 8.85e-17 | 34737 | 1481 | 33256 | 0.3495 | -0.09402 | Visual disturbances | sense organs | 25DTot | |
| 368.1 | | | | | 25311 | 52 | 25259 | | | | | 33326 | 70 | 33256 | | | Amblyopia | sense organs | 25DTot | |
| 368.2 | | | | | 25592 | 333 | 25259 | | | | | 33627 | 371 | 33256 | 0.6726 | -0.07403 | Diplopia and disorders of binocular vision | sense organs | 25DTot | |
| 368.3 | | | | | 25277 | 18 | 25259 | | | | | 33272 | 16 | 33256 | | | Anisometropia | sense organs | 25DTot | |
| 368.4 | | | | | 25410 | 151 | 25259 | | | | | 33464 | 208 | 33256 | | | Visual field defects | sense organs | 25DTot | |
| 368.5 | | | | | 25262 | 3 | 25259 | | | | | 33257 | 1 | 33256 | | | Color vision deficiencies | sense organs | 25DTot | |
| 368.7 | | | | | 25259 | | | | | | | 33257 | 1 | 33256 | | | Disorders of accommodation | sense organs | 25DTot | |
| 368.9 | | | | | 25451 | 192 | 25259 | | | | | 33633 | 377 | 33256 | 0.5223 | -0.1383 | Subjective visual disturbances | sense organs | 25DTot | |
| 368.91 | | | | | 25299 | 40 | 25259 | | | | | 33323 | 67 | 33256 | | | Psychophysical visual disturbances | sense organs | 25DTot | |
| 369 | 0.1973 | 0.1242 | 1.218 | 0.1123 | 24317 | 512 | 23805 | 0.5512 | 0.07396 | 1.735 | 9.19e-14 | 31511 | 916 | 30595 | 0.01849 | 0.3355 | Infection of the eye | sense organs | 25DTot | |
| 369.2 | | | | | 23885 | 80 | 23805 | | | | | 30724 | 129 | 30595 | | | Eye infection, viral | sense organs | 25DTot | |
| 369.5 | | | | | 24187 | 382 | 23805 | 0.6647 | 0.08176 | 1.944 | 4.28e-16 | 31314 | 719 | 30595 | 5.37e-04 | 0.5781 | Conjunctivitis, infectious | sense organs | 25DTot | |
| 370 | | | | | 24074 | 269 | 23805 | 0.8675 | 0.08264 | 2.381 | 8.86e-26 | 31271 | 676 | 30595 | 0.02414 | 0.3958 | Keratitis | sense organs | 25DTot | |
| 370.1 | | | | | 23880 | 75 | 23805 | | | | | 30694 | 99 | 30595 | | | Corneal ulcer | sense organs | 25DTot | |
| 370.2 | | | | | 23830 | 25 | 23805 | | | | | 30656 | 61 | 30595 | | | Superficial keratitis | sense organs | 25DTot | |
| 370.3 | | | | | 23964 | 159 | 23805 | | | | | 31063 | 468 | 30595 | 0.01185 | 0.5529 | Keratoconjunctivitis | sense organs | 25DTot | |
| 370.31 | | | | | 23923 | 118 | 23805 | | | | | 31009 | 414 | 30595 | 0.1036 | 0.3886 | Keratoconjunctivitis sicca | sense organs | 25DTot | |
| 371 | 0.3569 | 0.08857 | 1.429 | 5.60e-05 | 24762 | 957 | 23805 | 0.6711 | 0.05721 | 1.956 | 9.06e-32 | 32133 | 1538 | 30595 | 0.008808 | 0.2717 | Inflammation of the eye | sense organs | 25DTot | |

| phenotype | male_beta | male_SE | male_OR | male_p | male_n_total | male_n_cases | male_n_controls | female_beta | female_SE | female_OR | female_p | female_n_total | female_n_cases | female_n_controls | interaction_p | interaction_beta | description | group | lab | label |
|---|---|---|---|---|---|---|---|---|---|---|---|---|---|---|---|---|---|---|---|---|
| 371.1 | | | | | 23925 | 120 | 23805 | | | | | 30815 | 220 | 30595 | | | Uveitis, noninfectious or NOS | sense organs | 25DTot | |
| 371.2 | | | | | 24253 | 448 | 23805 | 0.7343 | 0.07774 | 2.084 | 3.52e-21 | 31368 | 773 | 30595 | 0.1048 | 0.2312 | Conjunctivitis, noninfectious | sense organs | 25DTot | |
| 371.21 | | | | | 24201 | 396 | 23805 | 0.7547 | 0.08223 | 2.127 | 4.41e-20 | 31279 | 684 | 30595 | 0.2047 | 0.1893 | Allergic conjunctivitis | sense organs | 25DTot | |
| 371.3 | | | | | 24165 | 360 | 23805 | 0.7702 | 0.09027 | 2.16 | 1.44e-17 | 31154 | 559 | 30595 | 0.0433 | 0.3235 | Inflammation of eyelids | sense organs | 25DTot | |
| 371.33 | | | | | 23808 | 3 | 23805 | | | | | 30603 | 8 | 30595 | | | Noninfectious dermatoses of eyelid | sense organs | 25DTot | |
| 371.9 | | | | | 23817 | 12 | 23805 | | | | | 30607 | 12 | 30595 | | | Chronic inflammatory disorders of orbit | sense organs | 25DTot | |
| 372 | | | | | 23933 | 128 | 23805 | | | | | 30728 | 133 | 30595 | | | Disorders of conjunctiva | sense organs | 25DTot | |
| 374 | 0.1295 | 0.1051 | 1.138 | 0.218 | 24581 | 776 | 23805 | 0.5575 | 0.06876 | 1.746 | 5.16e-16 | 31646 | 1051 | 30595 | 8.25e-04 | 0.413 | Other disorders of eyelids | sense organs | 25DTot | |
| 374.1 | | | | | 23920 | 115 | 23805 | | | | | 30673 | 78 | 30595 | | | Ectropion or entropion | sense organs | 25DTot | |
| 374.2 | | | | | 23850 | 45 | 23805 | | | | | 30619 | 24 | 30595 | | | Lagophthalmos | sense organs | 25DTot | |
| 374.3 | | | | | 24001 | 196 | 23805 | | | | | 30970 | 375 | 30595 | 0.1531 | 0.3089 | Ptosis of eyelid | sense organs | 25DTot | |
| 374.6 | | | | | 24250 | 445 | 23805 | 0.528 | 0.08829 | 1.695 | 2.23e-09 | 31205 | 610 | 30595 | 0.02589 | 0.3514 | Dermatochalasis | sense organs | 25DTot | |
| 375 | | | | | 23884 | 86 | 23798 | | | | | 31096 | 146 | 30950 | | | Disorders of lacrimal system | sense organs | 25DTot | |
| 375.1 | 0.2926 | 0.08689 | 1.34 | 7.57e-04 | 24860 | 1062 | 23798 | 0.6786 | 0.05145 | 1.971 | 1.00e-39 | 32921 | 1971 | 30950 | 4.18e-04 | 0.3512 | Dry eyes | sense organs | 25DTot | |
| 375.2 | | | | | 23848 | 50 | 23798 | | | | | 31036 | 86 | 30950 | | | Epiphora | sense organs | 25DTot | |
| 376 | | | | | 23842 | 44 | 23798 | | | | | 30982 | 32 | 30950 | | | Disorders of the orbit | sense organs | 25DTot | |
| 377 | | | | | 24004 | 206 | 23798 | | | | | 31323 | 373 | 30950 | 0.6985 | 0.08305 | Disorders of optic nerve and visual pathways | sense organs | 25DTot | |
| 377.1 | | | | | 23872 | 74 | 23798 | | | | | 31057 | 107 | 30950 | | | Optic atrophy | sense organs | 25DTot | |
| 377.3 | | | | | 23899 | 101 | 23798 | | | | | 31129 | 179 | 30950 | | | Optic neuritis/neuropathy | sense organs | 25DTot | |
| 378 | | | | | 24117 | 319 | 23798 | | | | | 31299 | 349 | 30950 | 0.5106 | -0.133 | Strabismus and other disorders of binocular eye movements | sense organs | 25DTot | |
| 378.1 | | | | | 23973 | 175 | 23798 | | | | | 31136 | 186 | 30950 | | | Strabismus (not specified as paralytic) | sense organs | 25DTot | |
| 378.2 | | | | | 23835 | 37 | 23798 | | | | | 31001 | 51 | 30950 | | | Nystagmus and other irregular eye movements | sense organs | 25DTot | |
| 378.5 | | | | | 23889 | 91 | 23798 | | | | | 31030 | 80 | 30950 | | | Paralytic strabismus | sense organs | 25DTot | |
| 379 | 0.327 | 0.08299 | 1.387 | 8.14e-05 | 24956 | 1158 | 23798 | 0.5594 | 0.05662 | 1.75 | 5.10e-23 | 32559 | 1609 | 30950 | 0.03074 | 0.2139 | Other disorders of eye | sense organs | 25DTot | |
| 379.1 | | | | | 23846 | 48 | 23798 | | | | | 31085 | 135 | 30950 | | | Scleritis and episcleritis | sense organs | 25DTot | |
| 379.2 | 0.3702 | 0.0889 | 1.448 | 3.12e-05 | 24762 | 964 | 23798 | 0.6265 | 0.06158 | 1.871 | 2.62e-24 | 32269 | 1319 | 30950 | 0.02212 | 0.2442 | Disorders of vitreous body | sense organs | 25DTot | |
| 379.3 | | | | | 23855 | 57 | 23798 | | | | | 30995 | 45 | 30950 | | | Aphakia and other disorders of lens | sense organs | 25DTot | |
| 379.4 | | | | | 23835 | 37 | 23798 | | | | | 31014 | 64 | 30950 | | | Anomalies of pupillary function | sense organs | 25DTot | |
| 379.5 | | | | | 23914 | 116 | 23798 | | | | | 31022 | 72 | 30950 | | | Disorders of iris and ciliary body | sense organs | 25DTot | |
| 379.51 | | | | | 23816 | 18 | 23798 | | | | | 30958 | 8 | 30950 | | | Pigmentary iris degeneration | sense organs | 25DTot | |
| 379.9 | | | | | 23893 | 95 | 23798 | | | | | 31121 | 171 | 30950 | | | Pain, swelling or discharge of eye | sense organs | 25DTot | |
| 380 | | | | | 25280 | 65 | 25215 | | | | | 34053 | 71 | 33982 | | | Disorders of external ear | sense organs | 25DTot | |
| 380.1 | | | | | 25546 | 331 | 25215 | | | | | 34343 | 361 | 33982 | 0.307 | 0.1997 | Otitis externa | sense organs | 25DTot | |
| 380.4 | 0.2902 | 0.08934 | 1.337 | 0.001158 | 26150 | 935 | 25215 | 0.441 | 0.07134 | 1.554 | 6.34e-10 | 34929 | 947 | 33982 | 0.2487 | 0.1298 | Impacted cerumen | sense organs | 25DTot | |
| 381 | 0.1673 | 0.0854 | 1.182 | 0.05014 | 25736 | 1209 | 24527 | 0.3932 | 0.05232 | 1.482 | 5.73e-14 | 33271 | 2155 | 31116 | 0.01035 | 0.2525 | Otitis media and Eustachian tube disorders | sense organs | 25DTot | |
| 381.1 | 0.1201 | 0.1194 | 1.128 | 0.3145 | 25160 | 633 | 24527 | 0.4267 | 0.07213 | 1.532 | 3.28e-09 | 32235 | 1119 | 31116 | 0.01111 | 0.3474 | Otitis media | sense organs | 25DTot | |
| 381.11 | | | | | 24939 | 412 | 24527 | 0.4858 | 0.08878 | 1.625 | 4.46e-08 | 31847 | 731 | 31116 | 0.003743 | 0.5021 | Suppurative and unspecified otitis media | sense organs | 25DTot | |
| 381.2 | 0.2269 | 0.112 | 1.255 | 0.04277 | 25149 | 622 | 24527 | 0.4706 | 0.06785 | 1.601 | 4.05e-12 | 32238 | 1122 | 31116 | 0.03983 | 0.2652 | Eustachian tube disorders | sense organs | 25DTot | |

| phenotype | male_beta | male_SE | male_OR | male_p | male_n_total | male_n_cases | male_n_controls | female_beta | female_SE | female_OR | female_p | female_n_total | female_n_cases | female_n_controls | interaction_p | interaction_beta | description | group | lab | label |
|---|---|---|---|---|---|---|---|---|---|---|---|---|---|---|---|---|---|---|---|---|
| 381.3 | | | | | 24579 | 52 | 24527 | | | | | 31179 | 63 | 31116 | | | Mastoiditis & related conditions | sense organs | 25DTot | |
| 381.9 | | | | | 24645 | 118 | 24527 | | | | | 31231 | 115 | 31116 | | | Otorrhea | sense organs | 25DTot | |
| 382 | | | | | 24849 | 322 | 24527 | 0.5215 | 0.07863 | 1.685 | 3.30e-11 | 31925 | 809 | 31116 | 0.07137 | 0.3049 | Otalgia | sense organs | 25DTot | |
| 383 | | | | | 24540 | 13 | 24527 | | | | | 31138 | 22 | 31116 | | | Otosclerosis | sense organs | 25DTot | |
| 384 | | | | | 24691 | 164 | 24527 | | | | | 31314 | 198 | 31116 | | | Other disorders of tympanic membrane | sense organs | 25DTot | |
| 384.1 | | | | | 24551 | 24 | 24527 | | | | | 31124 | 8 | 31116 | | | Myringitis | sense organs | 25DTot | |
| 384.4 | | | | | 24667 | 140 | 24527 | | | | | 31294 | 178 | 31116 | | | Perforation of tympanic membrane | sense organs | 25DTot | |
| 385 | | | | | 24625 | 98 | 24527 | | | | | 31245 | 129 | 31116 | | | Other disorders of middle ear and mastoid | sense organs | 25DTot | |
| 385.3 | | | | | 24606 | 79 | 24527 | | | | | 31204 | 88 | 31116 | | | Cholesteatoma | sense organs | 25DTot | |
| 385.5 | | | | | 24556 | 29 | 24527 | | | | | 31165 | 49 | 31116 | | | Tympanosclerosis and middle ear disease related to otitis media | sense organs | 25DTot | |
| 386 | | | | | 23483 | 489 | 22994 | 0.6383 | 0.07007 | 1.893 | 8.29e-20 | 30272 | 992 | 29280 | 0.5112 | 0.08853 | Vertiginous syndromes and other disorders of vestibular system | sense organs | 25DTot | |
| 386.1 | | | | | 23077 | 83 | 22994 | | | | | 29457 | 177 | 29280 | | | Meniere's disease | sense organs | 25DTot | |
| 386.2 | | | | | 23328 | 334 | 22994 | 0.6396 | 0.08357 | 1.896 | 1.96e-14 | 29955 | 675 | 29280 | 0.2258 | 0.1986 | Peripheral or central vertigo | sense organs | 25DTot | |
| 386.21 | | | | | 23041 | 47 | 22994 | | | | | 29385 | 105 | 29280 | | | Central origin vertigo | sense organs | 25DTot | |
| 386.3 | | | | | 23038 | 44 | 22994 | | | | | 29344 | 64 | 29280 | | | Labyrinthitis | sense organs | 25DTot | |
| 386.9 | 0.4626 | 0.06237 | 1.588 | 1.20e-13 | 25211 | 2217 | 22994 | 0.532 | 0.04062 | 1.702 | 3.36e-39 | 32967 | 3687 | 29280 | 0.3464 | 0.06905 | Dizziness and giddiness (Light-headedness and vertigo) | sense organs | 25DTot | |

| phenotype | male_beta | male_SE | male_OR | male_p | male_n_total | male_n_cases | male_n_controls | female_beta | female_SE | female_OR | female_p | female_n_total | female_n_cases | female_n_controls | interaction_p | interaction_beta | description | group | lab | label |
|---|---|---|---|---|---|---|---|---|---|---|---|---|---|---|---|---|---|---|---|---|
| 388 | | | | | 23411 | 34 | 23377 | | | | | 32030 | 47 | 31983 | | | Other disorders of ear | sense organs | 25DTot | |
| 389 | 0.4756 | 0.05793 | 1.609 | 2.24e-16 | 26031 | 2654 | 23377 | 0.4669 | 0.04393 | 1.595 | 2.23e-26 | 34938 | 2955 | 31983 | 0.6823 | -0.02935 | Hearing loss | sense organs | 25DTot | |
| 389.1 | 0.5571 | 0.06983 | 1.746 | 1.49e-15 | 25007 | 1630 | 23377 | 0.5194 | 0.05555 | 1.681 | 8.73e-21 | 33675 | 1692 | 31983 | 0.4622 | -0.06469 | Sensorineural hearing loss | sense organs | 25DTot | |
| 389.2 | | | | | 23633 | 256 | 23377 | | | | | 32346 | 363 | 31983 | 0.2635 | 0.2459 | Conductive hearing loss | sense organs | 25DTot | |
| 389.3 | | | | | 23398 | 21 | 23377 | | | | | 31995 | 12 | 31983 | | | Degenerative and vascular disorders of ear | sense organs | 25DTot | |
| 389.4 | 0.4818 | 0.1073 | 1.619 | 7.08e-06 | 23983 | 606 | 23377 | 0.6418 | 0.07997 | 1.9 | 1.02e-15 | 32714 | 731 | 31983 | 0.2165 | 0.1631 | Tinnitus | sense organs | 25DTot | |
| 389.5 | | | | | 23394 | 17 | 23377 | | | | | 32002 | 19 | 31983 | | | Disorders of acoustic nerve | sense organs | 25DTot | |
| 394 | -0.1138 | 0.09071 | 0.8924 | 0.2095 | 22330 | 1378 | 20952 | -0.1414 | 0.06633 | 0.8681 | 0.03304 | 31197 | 1468 | 29729 | 0.5961 | -0.0584 | Rheumatic disease of the heart valves | circulatory system | 25DTot | |
| 394.1 | | | | | 21305 | 353 | 20952 | | | | | 30068 | 339 | 29729 | 0.6152 | -0.1067 | Mitral valve stenosis and aortic valve stenosis | circulatory system | 25DTot | |
| 394.2 | | | | | 21121 | 169 | 20952 | | | | | 29965 | 236 | 29729 | | | Mitral valve disease | circulatory system | 25DTot | |
| 394.3 | | | | | 20969 | 17 | 20952 | | | | | 29742 | 13 | 29729 | | | Aortic valve disease | circulatory system | 25DTot | |
| 394.4 | | | | | 20972 | 20 | 20952 | | | | | 29771 | 42 | 29729 | | | Acute rheumatic heart disease | circulatory system | 25DTot | |
| 394.7 | -0.03194 | 0.1453 | 0.9686 | 0.8261 | 21461 | 509 | 20952 | -0.2727 | 0.1048 | 0.7613 | 0.009241 | 30357 | 628 | 29729 | 0.2136 | -0.2186 | Disease of tricuspid valve | circulatory system | 25DTot | |
| 395 | -0.01637 | 0.06113 | 0.9838 | 0.7889 | 24383 | 3431 | 20952 | -0.04257 | 0.04756 | 0.9583 | 0.3707 | 32852 | 3123 | 29729 | 0.5802 | -0.04186 | Heart valve disorders | circulatory system | 25DTot | |
| 395.1 | -0.05619 | 0.08045 | 0.9454 | 0.4849 | 22776 | 1824 | 20952 | -0.03089 | 0.05856 | 0.9696 | 0.5979 | 31647 | 1918 | 29729 | 0.9336 | 0.008115 | Nonrheumatic mitral valve disorders | circulatory system | 25DTot | |
| 395.2 | 0.04253 | 0.0784 | 1.043 | 0.5875 | 22773 | 1821 | 20952 | -0.147 | 0.06856 | 0.8633 | 0.03203 | 31147 | 1418 | 29729 | 0.04002 | -0.2086 | Nonrheumatic aortic valve disorders | circulatory system | 25DTot | |
| 395.3 | | | | | 21269 | 317 | 20952 | | | | | 30053 | 324 | 29729 | 0.556 | -0.1273 | Nonrheumatic tricuspid valve disorders | circulatory system | 25DTot | |

| phenotype | male_beta | male_SE | male_OR | male_p | male_n_total | male_n_cases | male_n_controls | female_beta | female_SE | female_OR | female_p | female_n_total | female_n_cases | female_n_controls | interaction_p | interaction_beta | description | group | lab | label |
|---|---|---|---|---|---|---|---|---|---|---|---|---|---|---|---|---|---|---|---|---|
| 395.4 | | | | | 21053 | 101 | 20952 | | | | | 29855 | 126 | 29729 | | | Nonrheumatic pulmonary valve disorders | circulatory system | 25DTot | |
| 395.6 | -0.1716 | 0.1156 | 0.8423 | 0.1377 | 21815 | 863 | 20952 | -0.3821 | 0.1089 | 0.6824 | 4.48e-04 | 30347 | 618 | 29729 | 0.06347 | -0.2881 | Heart valve replaced | circulatory system | 25DTot | |
| 396 | 0.1832 | 0.1194 | 1.201 | 0.125 | 21534 | 582 | 20952 | 0.2206 | 0.07622 | 1.247 | 0.0038 | 30632 | 903 | 29729 | 0.6282 | 0.06742 | Abnormal heart sounds | circulatory system | 25DTot | |
| 401 | 0.3377 | 0.05219 | 1.402 | 9.77e-11 | 25158 | 14670 | 10488 | 0.2054 | 0.03327 | 1.228 | 6.69e-10 | 33895 | 14782 | 19113 | 0.004542 | -0.1741 | Hypertension | circulatory system | 25DTot | |
| 401.1 | 0.3414 | 0.05249 | 1.407 | 7.80e-11 | 24789 | 14301 | 10488 | 0.2141 | 0.03341 | 1.239 | 1.46e-10 | 33585 | 14472 | 19113 | 0.005918 | -0.1697 | Essential hypertension | circulatory system | 25DTot | |
| 401.2 | 0.8844 | 0.06714 | 2.422 | 1.25e-39 | 14837 | 4349 | 10488 | 0.396 | 0.05132 | 1.486 | 1.19e-14 | 22464 | 3351 | 19113 | 2.84e-10 | -0.5289 | Hypertensive heart and/or renal disease | circulatory system | 25DTot | |
| 401.21 | 0.7321 | 0.0845 | 2.079 | 4.59e-18 | 12798 | 2310 | 10488 | 0.4201 | 0.06289 | 1.522 | 2.39e-11 | 21086 | 1973 | 19113 | 2.35e-04 | -0.3853 | Hypertensive heart disease | circulatory system | 25DTot | |
| 401.22 | 1.168 | 0.07297 | 3.215 | 1.24e-57 | 12900 | 2412 | 10488 | 0.5596 | 0.06384 | 1.75 | 1.87e-18 | 20743 | 1630 | 19113 | 9.05e-12 | -0.6511 | Hypertensive chronic kidney disease | circulatory system | 25DTot | |
| 401.3 | 1.225 | 0.09481 | 3.404 | 3.51e-38 | 11324 | 836 | 10488 | 0.5411 | 0.07749 | 1.718 | 2.89e-12 | 20074 | 961 | 19113 | 3.56e-10 | -0.7629 | Other hypertensive complications | circulatory system | 25DTot | |
| 411 | 0.1112 | 0.0497 | 1.118 | 0.02533 | 24924 | 8227 | 16697 | -0.06395 | 0.04092 | 0.9381 | 0.1181 | 33484 | 5060 | 28424 | 0.004102 | -0.1802 | Ischemic Heart Disease | circulatory system | 25DTot | |
| 411.1 | 0.2131 | 0.08509 | 1.238 | 0.01225 | 18153 | 1456 | 16697 | 0.05282 | 0.08641 | 1.054 | 0.541 | 29191 | 767 | 28424 | 0.05564 | -0.2266 | Unstable angina (intermediate coronary syndrome) | circulatory system | 25DTot | |
| 411.2 | 0.1486 | 0.06682 | 1.16 | 0.02612 | 19713 | 3016 | 16697 | -0.05889 | 0.06359 | 0.9428 | 0.3544 | 30089 | 1665 | 28424 | 0.007273 | -0.2414 | Myocardial infarction | circulatory system | 25DTot | |
| 411.3 | 0.1498 | 0.08096 | 1.162 | 0.06436 | 18373 | 1676 | 16697 | 0.004635 | 0.07535 | 1.005 | 0.951 | 29445 | 1021 | 28424 | 0.06536 | -0.1995 | Angina pectoris | circulatory system | 25DTot | |
| 411.4 | 0.1093 | 0.05158 | 1.115 | 0.03417 | 24205 | 7508 | 16697 | -0.06811 | 0.04437 | 0.9342 | 0.1248 | 32583 | 4159 | 28424 | 0.003962 | -0.1911 | Coronary atherosclerosis | circulatory system | 25DTot | |
| 411.41 | | | | | 16754 | 57 | 16697 | | | | | 28492 | 68 | 28424 | | | Aneurysm and dissection of heart | circulatory system | 25DTot | |
| 411.8 | 0.1994 | 0.0691 | 1.221 | 0.003912 | 19412 | 2715 | 16697 | -0.01692 | 0.07697 | 0.9832 | 0.826 | 29458 | 1034 | 28424 | 0.02087 | -0.2329 | Other chronic ischemic heart disease, unspecified | circulatory system | 25DTot | |
| 411.9 | | | | | 16791 | 94 | 16697 | | | | | 28492 | 68 | 28424 | | | Other acute and subacute forms of ischemic heart disease | circulatory system | 25DTot | |
| 414 | 0.1191 | 0.07486 | 1.127 | 0.1116 | 18860 | 2163 | 16697 | -0.06578 | 0.06622 | 0.9363 | 0.3205 | 29887 | 1463 | 28424 | 0.03252 | -0.2085 | Other forms of chronic heart disease | circulatory system | 25DTot | |
| 414.2 | | | | | 16930 | 233 | 16697 | | | | | 28541 | 117 | 28424 | | | ASCVD | circulatory system | 25DTot | |
| 415 | 0.4273 | 0.06943 | 1.533 | 7.53e-10 | 22610 | 2006 | 20604 | -0.08577 | 0.06049 | 0.9178 | 0.1562 | 32928 | 1974 | 30954 | 2.91e-08 | -0.4972 | Pulmonary heart disease | circulatory system | 25DTot | |
| 415.1 | 0.321 | 0.1047 | 1.378 | 0.002163 | 21476 | 872 | 20604 | -0.01635 | 0.09084 | 0.9838 | 0.8572 | 31762 | 808 | 30954 | 0.01434 | -0.3297 | Acute pulmonary heart disease | circulatory system | 25DTot | |
| 415.11 | 0.3574 | 0.1108 | 1.43 | 0.00125 | 21366 | 762 | 20604 | 0.004424 | 0.09379 | 1.004 | 0.9624 | 31710 | 756 | 30954 | 0.01598 | -0.3396 | Pulmonary embolism and infarction, acute | circulatory system | 25DTot | |
| 415.2 | 0.6206 | 0.08856 | 1.86 | 2.43e-12 | 21649 | 1045 | 20604 | -0.188 | 0.0821 | 0.8286 | 0.02201 | 32069 | 1115 | 30954 | 7.88e-12 | -0.8036 | Chronic pulmonary heart disease | circulatory system | 25DTot | |
| 415.21 | | | | | 20763 | 159 | 20604 | | | | | 31201 | 247 | 30954 | | | Primary pulmonary hypertension | circulatory system | 25DTot | |
| 416 | 0.2125 | 0.06191 | 1.237 | 5.98e-04 | 23564 | 2960 | 20604 | -0.02677 | 0.05688 | 0.9736 | 0.6379 | 32994 | 2040 | 30954 | 0.01705 | -0.1955 | Cardiomegaly | circulatory system | 25DTot | |
| 418 | 0.3876 | 0.04621 | 1.473 | 4.96e-17 | 23574 | 7662 | 15912 | 0.3614 | 0.03258 | 1.435 | 1.33e-28 | 31720 | 9073 | 22647 | 0.8363 | 0.01155 | Nonspecific chest pain | circulatory system | 25DTot | |
| 418.1 | 0.441 | 0.08259 | 1.554 | 9.31e-08 | 17223 | 1311 | 15912 | 0.2305 | 0.06537 | 1.259 | 4.22e-04 | 24061 | 1414 | 22647 | 0.07381 | -0.1863 | Precordial pain | circulatory system | 25DTot | |
| 420 | 0.08032 | 0.1046 | 1.084 | 0.4427 | 24630 | 934 | 23696 | 0.0963 | 0.08281 | 1.101 | 0.2449 | 34594 | 895 | 33699 | 0.6984 | 0.05052 | Carditis | circulatory system | 25DTot | |
| 420.1 | | | | | 23727 | 31 | 23696 | | | | | 33725 | 26 | 33699 | | | Myocarditis | circulatory system | 25DTot | |

| phenotype | male_beta | male_SE | male_OR | male_p | male_n_total | male_n_cases | male_n_controls | female_beta | female_SE | female_OR | female_p | female_n_total | female_n_cases | female_n_controls | interaction_p | interaction_beta | description | group | lab | label |
|---|---|---|---|---|---|---|---|---|---|---|---|---|---|---|---|---|---|---|---|---|
| 420.2 | 0.07753 | 0.1284 | 1.081 | 0.546 | 24311 | 615 | 23696 | 0.1714 | 0.09495 | 1.187 | 0.071 | 34349 | 650 | 33699 | 0.3562 | 0.144 | Pericarditis | circulatory system | 25DTot | |
| 420.21 | | | | | 23792 | 96 | 23696 | | | | | 33781 | 82 | 33699 | | | Acute pericarditis | circulatory system | 25DTot | |
| 420.22 | | | | | 23800 | 104 | 23696 | | | | | 33807 | 108 | 33699 | | | Chronic pericarditis | circulatory system | 25DTot | |
| 420.3 | | | | | 23990 | 294 | 23696 | | | | | 33932 | 233 | 33699 | 0.4922 | -0.1744 | Endocarditis | circulatory system | 25DTot | |
| 425 | -0.03949 | 0.07461 | 0.9613 | 0.5966 | 25754 | 2058 | 23696 | -0.1527 | 0.07262 | 0.8584 | 0.03555 | 34958 | 1259 | 33699 | 0.3792 | -0.08936 | Cardiomyopathy | circulatory system | 25DTot | |
| 425.1 | -0.06454 | 0.07856 | 0.9375 | 0.4113 | 25538 | 1842 | 23696 | -0.1866 | 0.07953 | 0.8298 | 0.01898 | 34754 | 1055 | 33699 | 0.3091 | -0.111 | Primary/intrinsic cardiomyopathies | circulatory system | 25DTot | |
| 425.11 | | | | | 23809 | 113 | 23696 | | | | | 33810 | 111 | 33699 | | | Hypertrophic obstructive cardiomyopathy | circulatory system | 25DTot | |
| 425.12 | | | | | 23788 | 92 | 23696 | | | | | 33777 | 78 | 33699 | | | Other hypertrophic cardiomyopathy | circulatory system | 25DTot | |
| 425.2 | -0.08576 | 0.1291 | 0.9178 | 0.5066 | 24351 | 655 | 23696 | | | | | 34066 | 367 | 33699 | 0.8303 | -0.03856 | Secondary/extrinsic cardiomyopathies | circulatory system | 25DTot | |
| 425.8 | | | | | 23738 | 42 | 23696 | | | | | 33802 | 103 | 33699 | | | Other cardiomyopathy | circulatory system | 25DTot | |
| 426 | 0.08105 | 0.06335 | 1.084 | 0.2007 | 18055 | 3856 | 14199 | 0.06298 | 0.05065 | 1.065 | 0.2138 | 24927 | 2812 | 22115 | 0.8465 | 0.01541 | Cardiac conduction disorders | circulatory system | 25DTot | |
| 426.2 | 0.1415 | 0.1096 | 1.152 | 0.1969 | 15129 | 930 | 14199 | -0.08531 | 0.1083 | 0.9182 | 0.4309 | 22650 | 535 | 22115 | 0.03443 | -0.3187 | Atrioventricular [AV] block | circulatory system | 25DTot | |
| 426.21 | | | | | 14391 | 192 | 14199 | | | | | 22185 | 70 | 22115 | | | First degree AV block | circulatory system | 25DTot | |
| 426.22 | | | | | 14228 | 29 | 14199 | | | | | 22135 | 20 | 22115 | | | Mobitz II AV block | circulatory system | 25DTot | |
| 426.23 | | | | | 14388 | 189 | 14199 | | | | | 22218 | 103 | 22115 | | | Second degree AV block | circulatory system | 25DTot | |
| 426.24 | -0.0879 | 0.1409 | 0.9159 | 0.5327 | 14798 | 599 | 14199 | | | | | 22478 | 363 | 22115 | 0.7225 | -0.06648 | Atrioventricular block, complete | circulatory system | 25DTot | |
| 426.25 | | | | | 14243 | 44 | 14199 | | | | | 22136 | 21 | 22115 | | | Other heart block | circulatory system | 25DTot | |
| 426.3 | 0.2035 | 0.1087 | 1.226 | 0.06121 | 15070 | 871 | 14199 | -0.06852 | 0.1026 | 0.9338 | 0.5043 | 22678 | 563 | 22115 | 0.1032 | -0.2392 | Bundle branch block | circulatory system | 25DTot | |
| 426.31 | | | | | 14547 | 348 | 14199 | | | | | 22262 | 147 | 22115 | | | Right bundle branch block | circulatory system | 25DTot | |
| 426.32 | | | | | 14638 | 439 | 14199 | | | | | 22500 | 385 | 22115 | 0.6506 | -0.09035 | Left bundle branch block | circulatory system | 25DTot | |
| 426.4 | | | | | 14250 | 51 | 14199 | | | | | 22148 | 33 | 22115 | | | Anomalous atrioventricular excitation | circulatory system | 25DTot | |
| 426.7 | 0.137 | 0.1208 | 1.147 | 0.2566 | 14888 | 689 | 14199 | 0.3695 | 0.08367 | 1.447 | 1.00e-05 | 22857 | 742 | 22115 | 0.01883 | 0.3405 | Abnormal electrocardiogram [ECG] [EKG] | circulatory system | 25DTot | |
| 426.8 | | | | | 14301 | 102 | 14199 | | | | | 22230 | 115 | 22115 | | | Other cardiac conduction disorders | circulatory system | 25DTot | |
| 426.9 | 0.02995 | 0.08237 | 1.03 | 0.7161 | 16283 | 2084 | 14199 | -0.07993 | 0.07456 | 0.9232 | 0.2837 | 23352 | 1237 | 22115 | 0.4204 | -0.08782 | Cardiac pacemaker/device in situ | circulatory system | 25DTot | |
| 426.91 | 0.226 | 0.09437 | 1.254 | 0.01662 | 15508 | 1309 | 14199 | -0.0294 | 0.08421 | 0.971 | 0.727 | 23040 | 925 | 22115 | 0.02575 | -0.2766 | Cardiac pacemaker in situ | circulatory system | 25DTot | |
| 426.92 | -0.1073 | 0.1121 | 0.8983 | 0.3384 | 15274 | 1075 | 14199 | | | | | 22521 | 406 | 22115 | 0.4949 | -0.1154 | Cardiac defibrillator in situ | circulatory system | 25DTot | |
| 427 | 0.167 | 0.04633 | 1.182 | 3.11e-04 | 23930 | 9731 | 14199 | 0.126 | 0.03181 | 1.134 | 7.49e-05 | 32189 | 10074 | 22115 | 0.608 | -0.02818 | Cardiac dysrhythmias | circulatory system | 25DTot | |
| 427.1 | 0.08282 | 0.08167 | 1.086 | 0.3106 | 16046 | 1847 | 14199 | 0.1373 | 0.0656 | 1.147 | 0.03634 | 23534 | 1419 | 22115 | 0.3404 | 0.09793 | Paroxysmal tachycardia, unspecified | circulatory system | 25DTot | |

| phenotype | male_beta | male_SE | male_OR | male_p | male_n_total | male_n_cases | male_n_controls | female_beta | female_SE | female_OR | female_p | female_n_total | female_n_cases | female_n_controls | interaction_p | interaction_beta | description | group | lab | label |
|---|---|---|---|---|---|---|---|---|---|---|---|---|---|---|---|---|---|---|---|---|
| 427.11 | 0.1178 | 0.1221 | 1.125 | 0.3345 | 14852 | 653 | 14199 | 0.2656 | 0.07984 | 1.304 | 8.79e-04 | 22978 | 863 | 22115 | 0.3709 | 0.1283 | Paroxysmal supraventricular tachycardia | circulatory system | 25DTot | |
| 427.12 | 0.02149 | 0.09781 | 1.022 | 0.8261 | 15495 | 1296 | 14199 | -0.04217 | 0.1039 | 0.9587 | 0.6849 | 22690 | 575 | 22115 | 0.8225 | -0.03137 | Paroxysmal ventricular tachycardia | circulatory system | 25DTot | |
| 427.2 | 0.1314 | 0.06238 | 1.14 | 0.03523 | 18883 | 4684 | 14199 | -0.007939 | 0.05287 | 0.9921 | 0.8807 | 25226 | 3111 | 22115 | 0.1912 | -0.1047 | Atrial fibrillation and flutter | circulatory system | 25DTot | |
| 427.21 | 0.14 | 0.06299 | 1.15 | 0.02626 | 18758 | 4559 | 14199 | 7.75e-05 | 0.05371 | 1 | 0.9988 | 25130 | 3015 | 22115 | 0.1839 | -0.1077 | Atrial fibrillation | circulatory system | 25DTot | |
| 427.22 | 0.05686 | 0.09328 | 1.059 | 0.5422 | 15660 | 1461 | 14199 | 0.018 | 0.08639 | 1.018 | 0.8349 | 22935 | 820 | 22115 | 0.9077 | 0.01448 | Atrial flutter | circulatory system | 25DTot | |
| 427.3 | 0.3531 | 0.07515 | 1.423 | 2.62e-06 | 16128 | 1929 | 14199 | 0.211 | 0.06024 | 1.235 | 4.61e-04 | 23822 | 1707 | 22115 | 0.0642 | -0.1743 | Other specified cardiac dysrhythmias | circulatory system | 25DTot | |
| 427.4 | | | | | 14682 | 483 | 14199 | | | | | 22371 | 256 | 22115 | 0.4483 | 0.1631 | Cardiac arrest and ventricular fibrillation | circulatory system | 25DTot | |
| 427.41 | | | | | 14406 | 207 | 14199 | | | | | 22188 | 73 | 22115 | | | Ventricular fibrillation and flutter | circulatory system | 25DTot | |
| 427.42 | | | | | 14543 | 344 | 14199 | | | | | 22324 | 209 | 22115 | 0.439 | 0.1901 | Cardiac arrest | circulatory system | 25DTot | |
| 427.5 | 0.236 | 0.06591 | 1.266 | 3.43e-04 | 17123 | 2924 | 14199 | 0.1474 | 0.05379 | 1.159 | 0.006135 | 24375 | 2260 | 22115 | 0.4035 | -0.06976 | Arrhythmia (cardiac) NOS | circulatory system | 25DTot | |
| 427.6 | 0.1184 | 0.1109 | 1.126 | 0.2857 | 15021 | 822 | 14199 | 0.2203 | 0.09305 | 1.246 | 0.01789 | 22723 | 608 | 22115 | 0.5916 | 0.07607 | Premature beats | circulatory system | 25DTot | |
| 427.61 | | | | | 14305 | 106 | 14199 | | | | | 22243 | 128 | 22115 | | | Supraventricular premature beats | circulatory system | 25DTot | |
| 427.7 | 0.4027 | 0.06457 | 1.496 | 4.46e-10 | 16928 | 2729 | 14199 | 0.239 | 0.04745 | 1.27 | 4.73e-07 | 25298 | 3183 | 22115 | 0.09232 | -0.132 | Tachycardia NOS | circulatory system | 25DTot | |
| 427.8 | 0.1487 | 0.1039 | 1.16 | 0.1523 | 15296 | 1097 | 14199 | -0.05251 | 0.09022 | 0.9488 | 0.5606 | 22910 | 795 | 22115 | 0.08426 | -0.2331 | Sinoatrial node dysfunction (Bradycardia) | circulatory system | 25DTot | |
| 427.9 | 0.2965 | 0.07322 | 1.345 | 5.13e-05 | 16102 | 1903 | 14199 | 0.3665 | 0.04227 | 1.443 | 4.35e-18 | 25899 | 3784 | 22115 | 0.6298 | 0.04012 | Palpitations | circulatory system | 25DTot | |
| 428 | 0.2214 | 0.05469 | 1.248 | 5.17e-05 | 24029 | 4495 | 19534 | -0.04037 | 0.04565 | 0.9604 | 0.3764 | 33140 | 3590 | 29550 | 3.05e-04 | -0.2514 | Congestive heart failure; nonhypertensive | circulatory system | 25DTot | |
| 428.1 | 0.2846 | 0.06 | 1.329 | 2.11e-06 | 22921 | 3387 | 19534 | -0.02762 | 0.0527 | 0.9728 | 0.6002 | 32151 | 2601 | 29550 | 7.14e-05 | -0.3095 | Congestive heart failure (CHF) NOS | circulatory system | 25DTot | |
| 428.2 | 0.3559 | 0.08909 | 1.427 | 6.48e-05 | 20752 | 1218 | 19534 | 0.01345 | 0.09248 | 1.014 | 0.8844 | 30286 | 736 | 29550 | 0.006059 | -0.3431 | Heart failure NOS | circulatory system | 25DTot | |
| 428.3 | 0.1391 | 0.07194 | 1.149 | 0.05309 | 21811 | 2277 | 19534 | -0.1978 | 0.07211 | 0.8205 | 0.006082 | 30905 | 1355 | 29550 | 3.65e-04 | -0.3542 | Heart failure with reduced EF [Systolic or combined heart failure] | circulatory system | 25DTot | |
| 428.4 | 0.4659 | 0.08154 | 1.593 | 1.11e-08 | 20846 | 1312 | 19534 | 0.1098 | 0.06295 | 1.116 | 0.08107 | 31066 | 1516 | 29550 | 2.80e-04 | -0.3673 | Heart failure with preserved EF [Diastolic heart failure] | circulatory system | 25DTot | |
| 429 | 0.4071 | 0.07544 | 1.503 | 6.79e-08 | 21072 | 1538 | 19534 | 0.07804 | 0.06481 | 1.081 | 0.2285 | 30921 | 1371 | 29550 | 0.001564 | -0.3077 | Ill-defined descriptions and complications of heart disease | circulatory system | 25DTot | |
| 429.1 | | | | | 19984 | 450 | 19534 | | | | | 29770 | 220 | 29550 | 0.005494 | -0.5559 | Heart transplant/surgery | circulatory system | 25DTot | |
| 429.2 | 0.3993 | 0.1221 | 1.491 | 0.001071 | 20041 | 507 | 19534 | | | | | 29943 | 393 | 29550 | 0.01434 | -0.4053 | Abnormal function study of cardiovascular system | circulatory system | 25DTot | |
| 429.3 | 0.5286 | 0.1099 | 1.697 | 1.50e-06 | 20117 | 583 | 19534 | 0.221 | 0.08697 | 1.247 | 0.01106 | 30233 | 683 | 29550 | 0.02831 | -0.3011 | Symptoms involving cardiovascular system | circulatory system | 25DTot | |
| 429.9 | | | | | 19772 | 238 | 19534 | | | | | 29698 | 148 | 29550 | | | Cardiac complications, not elsewhere classified | circulatory system | 25DTot | |
| 430 | 0.419 | 0.117 | 1.521 | 3.43e-04 | 22249 | 653 | 21596 | -0.1123 | 0.1151 | 0.8938 | 0.3296 | 31365 | 529 | 30836 | 0.001135 | -0.5202 | Intracranial hemorrhage | circulatory system | 25DTot | |
| 430.1 | | | | | 21793 | 197 | 21596 | | | | | 31023 | 187 | 30836 | | | Subarachnoid hemorrhage | circulatory system | 25DTot | |
| 430.2 | | | | | 21911 | 315 | 21596 | | | | | 31093 | 257 | 30836 | 0.003348 | -0.6725 | Intracerebral hemorrhage | circulatory system | 25DTot | |
| 430.3 | | | | | 21798 | 202 | 21596 | | | | | 30969 | 133 | 30836 | | | Subdural hemorrhage | circulatory system | 25DTot | |

| phenotype | male_beta | male_SE | male_OR | male_p | male_n_total | male_n_cases | male_n_controls | female_beta | female_SE | female_OR | female_p | female_n_total | female_n_cases | female_n_controls | interaction_p | interaction_beta | description | group | lab | label |
|---|---|---|---|---|---|---|---|---|---|---|---|---|---|---|---|---|---|---|---|---|
| 433 | 0.3987 | 0.0553 | 1.49 | 5.59e-13 | 25148 | 3552 | 21596 | 0.1408 | 0.04384 | 1.151 | 0.001321 | 34408 | 3572 | 30836 | 1.00e-04 | -0.2685 | Cerebrovascular disease | circulatory system | 25DTot | |
| 433.1 | 0.4821 | 0.0781 | 1.619 | 6.73e-10 | 23051 | 1455 | 21596 | 0.2602 | 0.0698 | 1.297 | 1.94e-04 | 32011 | 1175 | 30836 | 0.01052 | -0.2608 | Occlusion and stenosis of precerebral arteries | circulatory system | 25DTot | |
| 433.11 | | | | | 21806 | 210 | 21596 | | | | | 31009 | 173 | 30836 | | | Occlusion of cerebral arteries, with cerebral infarction | circulatory system | 25DTot | |
| 433.12 | | | | | 21766 | 170 | 21596 | | | | | 30990 | 154 | 30836 | | | Cerebral atherosclerosis | circulatory system | 25DTot | |
| 433.2 | 0.3421 | 0.08619 | 1.408 | 7.23e-05 | 22816 | 1220 | 21596 | 0.1201 | 0.07123 | 1.128 | 0.09181 | 32003 | 1167 | 30836 | 0.04804 | -0.2162 | Occlusion of cerebral arteries | circulatory system | 25DTot | |
| 433.21 | 0.3166 | 0.08862 | 1.372 | 3.54e-04 | 22764 | 1168 | 21596 | 0.1301 | 0.07278 | 1.139 | 0.07378 | 31945 | 1109 | 30836 | 0.1141 | -0.1772 | Cerebral artery occlusion, with cerebral infarction | circulatory system | 25DTot | |
| 433.3 | 0.4836 | 0.07124 | 1.622 | 1.13e-11 | 23331 | 1735 | 21596 | 0.173 | 0.05806 | 1.189 | 0.002886 | 32595 | 1759 | 30836 | 3.54e-04 | -0.3213 | Cerebral ischemia | circulatory system | 25DTot | |
| 433.31 | 0.4591 | 0.07407 | 1.583 | 5.69e-10 | 23194 | 1598 | 21596 | 0.1587 | 0.06025 | 1.172 | 0.008434 | 32452 | 1616 | 30836 | 0.001048 | -0.3063 | Transient cerebral ischemia | circulatory system | 25DTot | |
| 433.32 | | | | | 21602 | 6 | 21596 | | | | | 30850 | 14 | 30836 | | | Moyamoya disease | circulatory system | 25DTot | |
| 433.5 | | | | | 21737 | 141 | 21596 | | | | | 31155 | 319 | 30836 | | | Cerebral aneurysm | circulatory system | 25DTot | |
| 433.6 | | | | | 21926 | 330 | 21596 | | | | | 31171 | 335 | 30836 | 0.01167 | -0.4524 | Acute, but ill-defined cerebrovascular disease | circulatory system | 25DTot | |
| 433.8 | 0.7314 | 0.1031 | 2.078 | 1.28e-12 | 22246 | 650 | 21596 | 0.356 | 0.09112 | 1.428 | 9.34e-05 | 31470 | 634 | 30836 | 0.007797 | -0.3566 | Late effects of cerebrovascular disease | circulatory system | 25DTot | |
| 440 | 0.5896 | 0.07614 | 1.803 | 9.67e-15 | 21894 | 1543 | 20351 | 0.3732 | 0.06691 | 1.452 | 2.43e-08 | 31649 | 1262 | 30387 | 0.007877 | -0.2619 | Atherosclerosis | circulatory system | 25DTot | |
| 440.1 | | | | | 20496 | 145 | 20351 | | | | | 30601 | 214 | 30387 | | | Atherosclerosis of renal artery | circulatory system | 25DTot | |
| 440.2 | 0.5575 | 0.1069 | 1.746 | 1.84e-07 | 21065 | 714 | 20351 | | | | | 30809 | 422 | 30387 | 6.82e-04 | -0.5277 | Atherosclerosis of the extremities | circulatory system | 25DTot | |
| 440.21 | | | | | 20544 | 193 | 20351 | | | | | 30472 | 85 | 30387 | | | Atherosclerosis of native arteries of the extremities with ulceration or gangrene | circulatory system | 25DTot | |
| 440.22 | | | | | 20765 | 414 | 20351 | | | | | 30616 | 229 | 30387 | 0.006681 | -0.5681 | Atherosclerosis of native arteries of the extremities with intermittent claudication | circulatory system | 25DTot | |

| phenotype | male_beta | male_SE | male_OR | male_p | male_n_total | male_n_cases | male_n_controls | female_beta | female_SE | female_OR | female_p | female_n_total | female_n_cases | female_n_controls | interaction_p | interaction_beta | description | group | lab | label |
|---|---|---|---|---|---|---|---|---|---|---|---|---|---|---|---|---|---|---|---|---|
| 440.9 | | | | | 20820 | 469 | 20351 | | | | | 30841 | 454 | 30387 | 0.3972 | -0.133 | Atherosclerosis of aorta | circulatory system | 25DTot | |
| 441 | | | | | 20539 | 188 | 20351 | | | | | 30626 | 239 | 30387 | | | Vascular insufficiency of intestine | circulatory system | 25DTot | |
| 441.1 | | | | | 20421 | 70 | 20351 | | | | | 30462 | 75 | 30387 | | | Acute vascular insufficiency of intestine | circulatory system | 25DTot | |
| 441.2 | | | | | 20385 | 34 | 20351 | | | | | 30454 | 67 | 30387 | | | Chronic vascular insufficiency of intestine | circulatory system | 25DTot | |
| 442 | 0.1865 | 0.09087 | 1.205 | 0.04014 | 21609 | 1258 | 20351 | 0.2048 | 0.09129 | 1.227 | 0.02488 | 31061 | 674 | 30387 | 0.6257 | -0.06109 | Other aneurysm | circulatory system | 25DTot | |
| 442.1 | 0.1647 | 0.1015 | 1.179 | 0.1046 | 21366 | 1015 | 20351 | | | | | 30810 | 423 | 30387 | 0.3905 | -0.1288 | Aortic aneurysm | circulatory system | 25DTot | |
| 442.11 | 0.2882 | 0.1273 | 1.334 | 0.02356 | 20965 | 614 | 20351 | | | | | 30570 | 183 | 30387 | 0.8645 | -0.035 | Abdominal aortic aneurysm | circulatory system | 25DTot | |
| 442.2 | | | | | 20421 | 70 | 20351 | | | | | 30397 | 10 | 30387 | | | Aneurysm of iliac artery | circulatory system | 25DTot | |
| 442.3 | | | | | 20442 | 91 | 20351 | | | | | 30441 | 54 | 30387 | | | Aneurysm of artery of lower extremity | circulatory system | 25DTot | |
| 442.4 | | | | | 20394 | 43 | 20351 | | | | | 30441 | 54 | 30387 | | | Arterial dissection | circulatory system | 25DTot | |

| phenotype | male_beta | male_SE | male_OR | male_p | male_n_total | male_n_cases | male_n_controls | female_beta | female_SE | female_OR | female_p | female_n_total | female_n_cases | female_n_controls | interaction_p | interaction_beta | description | group | lab | label |
|---|---|---|---|---|---|---|---|---|---|---|---|---|---|---|---|---|---|---|---|---|
| 442.8 | | | | | 20454 | 103 | 20351 | | | | | 30512 | 125 | 30387 | | | Aneurysm of other specified artery | circulatory system | 25DTot | |
| 443 | 0.3795 | 0.07608 | 1.462 | 6.10e-07 | 22049 | 1698 | 20351 | 0.3794 | 0.06045 | 1.461 | 3.48e-10 | 31903 | 1516 | 30387 | 0.6161 | -0.04746 | Peripheral vascular disease | circulatory system | 25DTot | |
| 443.1 | | | | | 20444 | 93 | 20351 | | | | | 30852 | 465 | 30387 | 0.3222 | 0.2967 | Raynaud's syndrome | circulatory system | 25DTot | |
| 443.7 | | | | | 20611 | 260 | 20351 | | | | | 30517 | 130 | 30387 | | | Peripheral angiopathy in diseases classified elsewhere | circulatory system | 25DTot | |
| 443.8 | | | | | 20448 | 97 | 20351 | | | | | 30453 | 66 | 30387 | | | Other specified peripheral vascular diseases | circulatory system | 25DTot | |
| 443.9 | 0.4153 | 0.08119 | 1.515 | 3.12e-07 | 21791 | 1440 | 20351 | 0.2291 | 0.07684 | 1.257 | 0.002871 | 31349 | 962 | 30387 | 0.03873 | -0.2254 | Peripheral vascular disease, unspecified | circulatory system | 25DTot | |
| 444 | | | | | 20661 | 310 | 20351 | | | | | 30633 | 246 | 30387 | 0.132 | -0.3296 | Arterial embolism and thrombosis | circulatory system | 25DTot | |
| 444.1 | | | | | 20465 | 114 | 20351 | | | | | 30462 | 75 | 30387 | | | Arterial embolism and thrombosis of lower extremity artery | circulatory system | 25DTot | |
| 444.2 | | | | | 20391 | 40 | 20351 | | | | | 30424 | 37 | 30387 | | | Embolism and thrombosis of abdominal aorta | circulatory system | 25DTot | |
| 444.5 | | | | | 20357 | 6 | 20351 | | | | | 30392 | 5 | 30387 | | | Atheroembolism | circulatory system | 25DTot | |
| 446 | | | | | 20562 | 211 | 20351 | | | | | 30776 | 389 | 30387 | 0.2078 | -0.253 | Polyarteritis nodosa and allied conditions | circulatory system | 25DTot | |
| 446.1 | | | | | 20353 | 2 | 20351 | | | | | 30391 | 4 | 30387 | | | Thromboangiitis obliterans | circulatory system | 25DTot | |
| 446.2 | | | | | 20351 | | | | | | | 30393 | 6 | 30387 | | | Acute febrile mucocutaneous lymph node syndrome (Kawasaki disease) | circulatory system | 25DTot | |
| 446.3 | | | | | 20377 | 26 | 20351 | | | | | 30419 | 32 | 30387 | | | Hypersensitivity angiitis | circulatory system | 25DTot | |
| 446.4 | | | | | 20429 | 78 | 20351 | | | | | 30467 | 80 | 30387 | | | Wegener's granulomatosis | circulatory system | 25DTot | |
| 446.5 | | | | | 20386 | 35 | 20351 | | | | | 30500 | 113 | 30387 | | | Giant cell arteritis | circulatory system | 25DTot | |
| 446.6 | | | | | 20380 | 29 | 20351 | | | | | 30423 | 36 | 30387 | | | Polyarteritis nodosa | circulatory system | 25DTot | |
| 446.7 | | | | | 20353 | 2 | 20351 | | | | | 30403 | 16 | 30387 | | | Takayasu's disease | circulatory system | 25DTot | |
| 446.8 | | | | | 20362 | 11 | 20351 | | | | | 30409 | 22 | 30387 | | | Thrombotic microangiopathy | circulatory system | 25DTot | |
| 446.9 | | | | | 20443 | 92 | 20351 | | | | | 30554 | 167 | 30387 | | | Arteritis NOS | circulatory system | 25DTot | |
| 447 | 0.09668 | 0.09615 | 1.102 | 0.3146 | 21399 | 1048 | 20351 | 0.16 | 0.09657 | 1.174 | 0.09757 | 30938 | 551 | 30387 | 0.7354 | 0.04522 | Other disorders of arteries and arterioles | circulatory system | 25DTot | |
| 447.1 | | | | | 20462 | 111 | 20351 | | | | | 30487 | 100 | 30387 | | | Stricture of artery | circulatory system | 25DTot | |
| 447.7 | -0.09854 | 0.1211 | 0.9062 | 0.4157 | 21056 | 705 | 20351 | | | | | 30596 | 209 | 30387 | 0.8457 | 0.0381 | Aortic ectasia | circulatory system | 25DTot | |
| 448 | | | | | 20358 | 7 | 20351 | | | | | 30410 | 23 | 30387 | | | Disease of capillaries | circulatory system | 25DTot | |
| 450 | | | | | 27464 | 390 | 27074 | 0.06398 | 0.08778 | 1.066 | 0.4661 | 36525 | 739 | 35786 | 0.8321 | -0.036 | Noninfectious disorders of lymphatic channels | circulatory system | 25DTot | |
| 451 | | | | | 17675 | 229 | 17446 | | | | | 26197 | 265 | 25932 | | | Phlebitis and thrombophlebitis | circulatory system | 25DTot | |
| 451.2 | | | | | 17551 | 105 | 17446 | | | | | 26069 | 137 | 25932 | | | Phlebitis and thrombophlebitis of lower extremities | circulatory system | 25DTot | |
| 452 | 0.4204 | 0.07304 | 1.523 | 8.63e-09 | 19497 | 2051 | 17446 | 0.2169 | 0.05696 | 1.242 | 1.40e-04 | 28016 | 2084 | 25932 | 0.05973 | -0.1711 | Other venous embolism and thrombosis | circulatory system | 25DTot | |
| 452.1 | | | | | 17477 | 31 | 17446 | | | | | 25960 | 28 | 25932 | | | Iatrogenic pulmonary embolism and infarction | circulatory system | 25DTot | |

| phenotype | male_beta | male_SE | male_OR | male_p | male_n_total | male_n_cases | male_n_controls | female_beta | female_SE | female_OR | female_p | female_n_total | female_n_cases | female_n_controls | interaction_p | interaction_beta | description | group | lab | label |
|---|---|---|---|---|---|---|---|---|---|---|---|---|---|---|---|---|---|---|---|---|
| 452.2 | 0.4168 | 0.09741 | 1.517 | 1.88e-05 | 18523 | 1077 | 17446 | 0.2261 | 0.07751 | 1.254 | 0.003528 | 26996 | 1064 | 25932 | 0.2312 | -0.1459 | Deep vein thrombosis [DVT] | circulatory system | 25DTot | |
| 452.8 | | | | | 17477 | 31 | 17446 | | | | | 25956 | 24 | 25932 | | | Postphlebitic syndrome | circulatory system | 25DTot | |
| 453 | | | | | 17473 | 27 | 17446 | | | | | 25945 | 13 | 25932 | | | Chronic venous hypertension | circulatory system | 25DTot | |
| 454 | | | | | 17905 | 459 | 17446 | 0.6251 | 0.08858 | 1.868 | 1.71e-12 | 26542 | 610 | 25932 | 0.7608 | -0.04491 | Varicose veins | circulatory system | 25DTot | |
| 454.1 | | | | | 17712 | 266 | 17446 | 0.6164 | 0.09633 | 1.852 | 1.56e-10 | 26438 | 506 | 25932 | 0.4589 | -0.132 | Varicose veins of lower extremity | circulatory system | 25DTot | |
| 454.11 | | | | | 17657 | 211 | 17446 | | | | | 26297 | 365 | 25932 | 0.3652 | -0.1834 | Varicose veins of lower extremity, symptomtic | circulatory system | 25DTot | |
| 455 | 0.842 | 0.0781 | 2.321 | 4.27e-27 | 18658 | 1212 | 17446 | 0.7556 | 0.06184 | 2.129 | 2.47e-34 | 27261 | 1329 | 25932 | 0.1401 | -0.1443 | Hemorrhoids | circulatory system | 25DTot | |
| 456 | | | | | 17849 | 403 | 17446 | | | | | 26402 | 470 | 25932 | 0.3429 | -0.1574 | Chronic venous insufficiency [CVI] | circulatory system | 25DTot | |
| 457 | 0.4632 | 0.06626 | 1.589 | 2.75e-12 | 19943 | 2497 | 17446 | 0.2521 | 0.05743 | 1.287 | 1.14e-05 | 27763 | 1831 | 25932 | 0.0114 | -0.2182 | Encounter for long-term (current) use of anticoagulants, antithrombotics, aspirin | circulatory system | 25DTot | |
| 457.2 | 0.4637 | 0.1235 | 1.59 | 1.73e-04 | 17977 | 531 | 17446 | | | | | 26262 | 330 | 25932 | 0.2647 | -0.1915 | Encounter for long-term (current) use of antiplatelets/antithrombotics | circulatory system | 25DTot | |
| 457.3 | 0.4774 | 0.06808 | 1.612 | 2.34e-12 | 19733 | 2287 | 17446 | 0.2578 | 0.05971 | 1.294 | 1.57e-05 | 27596 | 1664 | 25932 | 0.01047 | -0.228 | Encounter for long-term (current) use of aspirin | circulatory system | 25DTot | |
| 458 | 0.4856 | 0.05459 | 1.625 | 5.89e-19 | 23987 | 3579 | 20408 | 0.1642 | 0.04763 | 1.178 | 5.66e-04 | 32936 | 2891 | 30045 | 2.01e-05 | -0.3007 | Hypotension | circulatory system | 25DTot | |
| 458.1 | 0.7908 | 0.09777 | 2.205 | 6.06e-16 | 21124 | 716 | 20408 | 0.4199 | 0.09494 | 1.522 | 9.75e-06 | 30599 | 554 | 30045 | 0.008758 | -0.3465 | Orthostatic hypotension | circulatory system | 25DTot | |
| 458.2 | | | | | 20542 | 134 | 20408 | | | | | 30168 | 123 | 30045 | | | Iatrogenic hypotension | circulatory system | 25DTot | |
| 458.9 | 0.4515 | 0.06091 | 1.571 | 1.24e-13 | 23196 | 2788 | 20408 | 0.1438 | 0.05412 | 1.155 | 0.007884 | 32272 | 2227 | 30045 | 2.01e-04 | -0.2948 | Hypotension NOS | circulatory system | 25DTot | |
| 459 | 0.2785 | 0.09485 | 1.321 | 0.003322 | 21377 | 969 | 20408 | 0.1085 | 0.07933 | 1.115 | 0.1714 | 30928 | 883 | 30045 | 0.202 | -0.1545 | Other disorders of circulatory system | circulatory system | 25DTot | |
| 459.1 | | | | | 20541 | 133 | 20408 | | | | | 30175 | 130 | 30045 | | | Hemorrhage NOS | circulatory system | 25DTot | |
| 459.7 | | | | | 20566 | 158 | 20408 | | | | | 30162 | 117 | 30045 | | | Blood vessel replaced | circulatory system | 25DTot | |
| 459.9 | 0.3723 | 0.1148 | 1.451 | 0.001184 | 21001 | 593 | 20408 | 0.1398 | 0.1004 | 1.15 | 0.1638 | 30563 | 518 | 30045 | 0.1702 | -0.2049 | Circulatory disease NEC | circulatory system | 25DTot | |
| 464 | 0.5532 | 0.07376 | 1.739 | 6.40e-14 | 22311 | 1470 | 20841 | 0.8795 | 0.04307 | 2.41 | 1.17e-92 | 28085 | 3592 | 24493 | 1.58e-04 | 0.3196 | Acute sinusitis | respiratory | 25DTot | |
| 465 | 0.2727 | 0.06118 | 1.314 | 8.29e-06 | 23812 | 2971 | 20841 | 0.655 | 0.03725 | 1.925 | 3.33e-69 | 30856 | 6363 | 24493 | 9.99e-08 | 0.3767 | Acute upper respiratory infections of multiple or unspecified sites | respiratory | 25DTot | |
| phenotype | male_beta | male_SE | male_OR | male_p | male_n_total | male_n_cases | male_n_controls | female_beta | female_SE | female_OR | female_p | female_n_total | female_n_cases | female_n_controls | interaction_p | interaction_beta | description | group | lab | label |
| 465.2 | 0.3396 | 0.09687 | 1.404 | 4.56e-04 | 21831 | 990 | 20841 | 0.7338 | 0.05199 | 2.083 | 3.10e-45 | 27189 | 2696 | 24493 | 1.39e-04 | 0.4134 | Acute pharyngitis | respiratory | 25DTot | |
| 465.4 | | | | | 20905 | 64 | 20841 | | | | | 24590 | 97 | 24493 | | | Acute laryngitis and tracheitis | respiratory | 25DTot | |
| 470 | 0.5407 | 0.09194 | 1.717 | 4.09e-09 | 19858 | 932 | 18926 | 0.7417 | 0.06812 | 2.099 | 1.30e-27 | 25321 | 1191 | 24130 | 0.1304 | 0.1704 | Septal Deviations/Turbinate Hypertrophy | respiratory | 25DTot | |
| 471 | | | | | 19079 | 153 | 18926 | | | | | 24245 | 115 | 24130 | | | Nasal polyps | respiratory | 25DTot | |
| 472 | 0.7566 | 0.09838 | 2.131 | 1.46e-14 | 19621 | 695 | 18926 | 0.9621 | 0.06543 | 2.617 | 6.01e-49 | 25328 | 1198 | 24130 | 0.1127 | 0.1849 | Chronic pharyngitis and nasopharyngitis | respiratory | 25DTot | |
| 473 | 0.4004 | 0.09471 | 1.492 | 2.37e-05 | 19940 | 1014 | 18926 | 0.5332 | 0.0669 | 1.704 | 1.59e-15 | 25397 | 1267 | 24130 | 0.0609 | 0.2138 | Diseases of the larynx and vocal cords | respiratory | 25DTot | |
| 473.1 | | | | | 19013 | 87 | 18926 | | | | | 24250 | 120 | 24130 | | | Chronic laryngitis | respiratory | 25DTot | |
| 473.3 | | | | | 19140 | 214 | 18926 | | | | | 24368 | 238 | 24130 | | | Paralysis/spasm of vocal cords or larynx | respiratory | 25DTot | |

| phenotype | male_beta | male_SE | male_OR | male_p | male_n_total | male_n_cases | male_n_controls | female_beta | female_SE | female_OR | female_p | female_n_total | female_n_cases | female_n_controls | interaction_p | interaction_beta | description | group | lab | label |
|---|---|---|---|---|---|---|---|---|---|---|---|---|---|---|---|---|---|---|---|---|
| 473.4 | 0.5373 | 0.1033 | 1.711 | 1.95e-07 | 19690 | 764 | 18926 | 0.5911 | 0.07355 | 1.806 | 9.20e-16 | 25123 | 993 | 24130 | 0.2074 | 0.1572 | Voice disturbance | respiratory | 25DTot | |
| 474 | | | | | 19193 | 267 | 18926 | | | | | 24615 | 485 | 24130 | 0.1859 | 0.3084 | Acute and chronic tonsillitis | respiratory | 25DTot | |
| 474.1 | | | | | 18953 | 27 | 18926 | | | | | 24211 | 81 | 24130 | | | Acute tonsillitis | respiratory | 25DTot | |
| 474.2 | | | | | 19156 | 230 | 18926 | | | | | 24527 | 397 | 24130 | 0.136 | 0.3781 | Chronic tonsillitis and adenoiditis | respiratory | 25DTot | |
| 475 | 0.7386 | 0.07414 | 2.093 | 2.23e-23 | 20308 | 1382 | 18926 | 0.8421 | 0.05125 | 2.321 | 1.13e-60 | 26319 | 2189 | 24130 | 0.1671 | 0.1231 | Chronic sinusitis | respiratory | 25DTot | |
| 475.9 | | | | | 19118 | 192 | 18926 | | | | | 24485 | 355 | 24130 | 0.1482 | 0.3204 | Postnasal drip | respiratory | 25DTot | |
| 476 | 0.6129 | 0.05931 | 1.846 | 4.95e-25 | 21597 | 2671 | 18926 | 0.8161 | 0.03836 | 2.262 | 1.93e-100 | 28998 | 4868 | 24130 | 0.002028 | 0.2154 | Allergic rhinitis | respiratory | 25DTot | |
| 477 | | | | | 19327 | 401 | 18926 | | | | | 24561 | 431 | 24130 | 0.1729 | 0.2345 | Epistaxis or throat hemorrhage | respiratory | 25DTot | |
| 478 | | | | | 19150 | 224 | 18926 | | | | | 24407 | 277 | 24130 | 0.001432 | 0.8681 | Throat pain | respiratory | 25DTot | |
| 479 | 0.4766 | 0.08099 | 1.611 | 4.00e-09 | 20272 | 1346 | 18926 | 0.6906 | 0.05565 | 1.995 | 2.28e-35 | 26030 | 1900 | 24130 | 0.002793 | 0.2901 | Other upper respiratory disease | respiratory | 25DTot | |
| 480 | 0.4904 | 0.05344 | 1.633 | 4.50e-20 | 23852 | 3704 | 20148 | 0.3155 | 0.04364 | 1.371 | 4.85e-13 | 31060 | 3539 | 27521 | 0.132 | -0.1017 | Pneumonia | respiratory | 25DTot | |
| 480.1 | 0.4664 | 0.09419 | 1.594 | 7.34e-07 | 21107 | 959 | 20148 | 0.4265 | 0.08074 | 1.532 | 1.28e-07 | 28349 | 828 | 27521 | 0.498 | 0.0822 | Bacterial pneumonia | respiratory | 25DTot | |
| 480.11 | | | | | 20392 | 244 | 20148 | | | | | 27735 | 214 | 27521 | | | Pneumococcal pneumonia | respiratory | 25DTot | |
| 480.12 | | | | | 20313 | 165 | 20148 | | | | | 27625 | 104 | 27521 | | | Pseudomonal pneumonia | respiratory | 25DTot | |
| 480.13 | | | | | 20243 | 95 | 20148 | | | | | 27596 | 75 | 27521 | | | MRSA pneumonia | respiratory | 25DTot | |
| 480.2 | | | | | 20252 | 104 | 20148 | | | | | 27629 | 108 | 27521 | | | Viral pneumonia | respiratory | 25DTot | |
| 480.3 | | | | | 20261 | 113 | 20148 | | | | | 27595 | 74 | 27521 | | | Pneumonia due to fungus (mycoses) | respiratory | 25DTot | |
| 480.5 | | | | | 20273 | 125 | 20148 | | | | | 27647 | 126 | 27521 | | | Bronchopneumonia and lung abscess | respiratory | 25DTot | |
| 481 | | | | | 20437 | 289 | 20148 | | | | | 27934 | 413 | 27521 | 0.7208 | 0.07035 | Influenza | respiratory | 25DTot | |
| 483 | 0.402 | 0.08739 | 1.495 | 4.23e-06 | 21180 | 1032 | 20148 | 0.5724 | 0.0541 | 1.773 | 3.68e-26 | 29347 | 1826 | 27521 | 0.1029 | 0.1655 | Acute bronchitis and bronchiolitis | respiratory | 25DTot | |
| 495 | 0.4154 | 0.0735 | 1.515 | 1.58e-08 | 22371 | 1521 | 20850 | 0.3141 | 0.04419 | 1.369 | 1.18e-12 | 31168 | 3223 | 27945 | 0.3874 | -0.07274 | Asthma | respiratory | 25DTot | |
| 495.1 | | | | | 21010 | 160 | 20850 | | | | | 28173 | 228 | 27945 | | | Chronic obstructive asthma | respiratory | 25DTot | |
| 495.11 | | | | | 20869 | 19 | 20850 | | | | | 27991 | 46 | 27945 | | | Chronic obstructive asthma with exacerbation | respiratory | 25DTot | |
| 495.2 | | | | | 21125 | 275 | 20850 | 0.4968 | 0.08305 | 1.643 | 2.21e-09 | 28693 | 748 | 27945 | 0.7846 | -0.04684 | Asthma with exacerbation | respiratory | 25DTot | |
| 496 | 0.3345 | 0.05883 | 1.397 | 1.30e-08 | 23991 | 3141 | 20850 | 0.1133 | 0.04885 | 1.12 | 0.02032 | 30852 | 2907 | 27945 | 0.009821 | -0.1924 | Chronic airway obstruction | respiratory | 25DTot | |
| 496.1 | 0.1604 | 0.1021 | 1.174 | 0.116 | 21816 | 966 | 20850 | 0.1496 | 0.09062 | 1.161 | 0.09872 | 28671 | 726 | 27945 | 0.9184 | -0.01359 | Emphysema | respiratory | 25DTot | |
| 496.2 | 0.4073 | 0.09816 | 1.503 | 3.33e-05 | 21707 | 857 | 20850 | 0.3339 | 0.07605 | 1.396 | 1.13e-05 | 28880 | 935 | 27945 | 0.7039 | -0.04602 | Chronic bronchitis | respiratory | 25DTot | |
| 496.21 | 0.4751 | 0.1072 | 1.608 | 9.43e-06 | 21544 | 694 | 20850 | 0.3215 | 0.08563 | 1.379 | 1.74e-04 | 28686 | 741 | 27945 | 0.3031 | -0.1375 | Obstructive chronic bronchitis | respiratory | 25DTot | |
| 496.3 | | | | | 21106 | 256 | 20850 | | | | | 28305 | 360 | 27945 | 0.1281 | -0.2727 | Bronchiectasis | respiratory | 25DTot | |
| 497 | | | | | 21118 | 268 | 20850 | | | | | 28430 | 485 | 27945 | 0.7818 | -0.05075 | Bronchitis | respiratory | 25DTot | |
| 498 | | | | | 20873 | 23 | 20850 | | | | | 27982 | 37 | 27945 | | | Acute bronchospasm | respiratory | 25DTot | |
| 499 | | | | | 27770 | 89 | 27681 | | | | | 36921 | 77 | 36844 | | | Cystic fibrosis | respiratory | 25DTot | |
| 500 | | | | | 16503 | 145 | 16358 | | | | | 26637 | 93 | 26544 | | | Lung disease due to external agents | respiratory | 25DTot | |
| 500.1 | | | | | 16369 | 11 | 16358 | | | | | 26562 | 18 | 26544 | | | Extrinsic allergic alveolitis | respiratory | 25DTot | |
| 500.2 | | | | | 16384 | 26 | 16358 | | | | | 26547 | 3 | 26544 | | | Pneumoconiosis | respiratory | 25DTot | |
| 501 | 0.4871 | 0.1221 | 1.628 | 6.63e-05 | 16908 | 550 | 16358 | | | | | 26905 | 361 | 26544 | 0.001018 | -0.5799 | Pneumonitis due to inhalation of food or vomitus | respiratory | 25DTot | |
| 502 | | | | | 16775 | 417 | 16358 | | | | | 26948 | 404 | 26544 | 0.1396 | -0.2573 | Postinflammatory pulmonary fibrosis | respiratory | 25DTot | |

| phenotype | male_beta | male_SE | male_OR | male_p | male_n_total | male_n_cases | male_n_controls | female_beta | female_SE | female_OR | female_p | female_n_total | female_n_cases | female_n_controls | interaction_p | interaction_beta | description | group | lab | label |
|---|---|---|---|---|---|---|---|---|---|---|---|---|---|---|---|---|---|---|---|---|
| 503 | 0.5019 | 0.09346 | 1.652 | 7.86e-08 | 17413 | 1055 | 16358 | 0.1323 | 0.08684 | 1.141 | 0.1277 | 27379 | 835 | 26544 | 0.007428 | -0.3321 | Pulmonary congestion and hypostasis | respiratory | 25DTot | |
| 504 | | | | | 16686 | 328 | 16358 | | | | | 26850 | 306 | 26544 | 0.9068 | 0.02418 | Other alveolar and parietoalveolar pneumonopathy | respiratory | 25DTot | |
| 504.1 | | | | | 16528 | 170 | 16358 | | | | | 26674 | 130 | 26544 | | | Idiopathic fibrosing alveolitis | respiratory | 25DTot | |
| 505 | | | | | 16583 | 225 | 16358 | | | | | 26786 | 242 | 26544 | | | Other pulmonary inflamation or edema | respiratory | 25DTot | |
| 506 | -0.005474 | 0.09876 | 0.9945 | 0.9558 | 17754 | 1396 | 16358 | -0.4384 | 0.09244 | 0.645 | 2.11e-06 | 27619 | 1075 | 26544 | 0.001105 | -0.4319 | Empyema and pneumothorax | respiratory | 25DTot | |
| 507 | 0.3187 | 0.05686 | 1.375 | 2.07e-08 | 20407 | 4049 | 16358 | -0.03154 | 0.04843 | 0.969 | 0.5149 | 29890 | 3346 | 26544 | 1.81e-06 | -0.3475 | Pleurisy; pleural effusion | respiratory | 25DTot | |
| 508 | 0.2963 | 0.04987 | 1.345 | 2.81e-09 | 22186 | 5828 | 16358 | -0.1085 | 0.04395 | 0.8972 | 0.01359 | 30908 | 4364 | 26544 | 1.10e-10 | -0.4182 | Pulmonary collapse; interstitial and compensatory emphysema | respiratory | 25DTot | |
| 509 | 0.2551 | 0.05599 | 1.291 | 5.21e-06 | 20689 | 4331 | 16358 | -0.08406 | 0.04691 | 0.9194 | 0.07318 | 30174 | 3630 | 26544 | 1.40e-05 | -0.3101 | Respiratory failure, insufficiency, arrest | respiratory | 25DTot | |
| 509.1 | 0.3496 | 0.06318 | 1.418 | 3.15e-08 | 19312 | 2954 | 16358 | 0.01037 | 0.05275 | 1.01 | 0.8442 | 29134 | 2590 | 26544 | 4.11e-04 | -0.2839 | Respiratory failure | respiratory | 25DTot | |
| 509.2 | 0.2076 | 0.1047 | 1.231 | 0.04734 | 17397 | 1039 | 16358 | -0.1328 | 0.09778 | 0.8756 | 0.1743 | 27339 | 795 | 26544 | 0.007703 | -0.3725 | Respiratory insufficiency | respiratory | 25DTot | |
| 509.3 | 0.2982 | 0.1062 | 1.347 | 0.004988 | 17365 | 1007 | 16358 | -0.22 | 0.1117 | 0.8026 | 0.04897 | 27216 | 672 | 26544 | 0.0017 | -0.4724 | Pulmonary insufficiency or respiratory failure following trauma and surgery | respiratory | 25DTot | |
| 509.5 | | | | | 16395 | 37 | 16358 | | | | | 26570 | 26 | 26544 | | | Respiratory arrest | respiratory | 25DTot | |
| 509.8 | 0.2991 | 0.1127 | 1.349 | 0.007943 | 17139 | 781 | 16358 | 0.08246 | 0.09136 | 1.086 | 0.3668 | 27262 | 718 | 26544 | 0.4398 | -0.1094 | Dependence on respirator [Ventilator] or supplemental oxygen | respiratory | 25DTot | |
| 510 | 0.353 | 0.06279 | 1.423 | 1.89e-08 | 24918 | 2670 | 22248 | 0.1571 | 0.0541 | 1.17 | 0.003681 | 34185 | 2275 | 31910 | 0.04826 | -0.1592 | Other diseases of lung | respiratory | 25DTot | |
| 510.2 | | | | | 22355 | 107 | 22248 | | | | | 32017 | 107 | 31910 | | | Lung transplant | respiratory | 25DTot | |

| phenotype | male_beta | male_SE | male_OR | male_p | male_n_total | male_n_cases | male_n_controls | female_beta | female_SE | female_OR | female_p | female_n_total | female_n_cases | female_n_controls | interaction_p | interaction_beta | description | group | lab | label |
|---|---|---|---|---|---|---|---|---|---|---|---|---|---|---|---|---|---|---|---|---|
| 512 | 0.3364 | 0.04698 | 1.4 | 8.02e-13 | 23558 | 11275 | 12283 | 0.3115 | 0.03204 | 1.365 | 2.41e-22 | 31474 | 13478 | 17996 | 0.5639 | 0.03248 | Other symptoms of respiratory system | respiratory | 25DTot | |
| 512.1 | | | | | 12736 | 453 | 12283 | 0.4744 | 0.08786 | 1.607 | 6.67e-08 | 18716 | 720 | 17996 | 0.8992 | 0.02021 | Wheezing | respiratory | 25DTot | |
| 512.2 | | | | | 12736 | 453 | 12283 | 0.6342 | 0.09481 | 1.886 | 2.24e-11 | 18591 | 595 | 17996 | 0.05375 | -0.2908 | Painful respiration | respiratory | 25DTot | |
| 512.3 | | | | | 12290 | 7 | 12283 | | | | | 18006 | 10 | 17996 | | | Abnormal chest sounds | respiratory | 25DTot | |
| 512.7 | 0.4382 | 0.05282 | 1.55 | 1.06e-16 | 19002 | 6719 | 12283 | 0.369 | 0.03745 | 1.446 | 6.62e-23 | 25374 | 7378 | 17996 | 0.712 | -0.02365 | Shortness of breath | respiratory | 25DTot | |
| 512.8 | 0.5861 | 0.0575 | 1.797 | 2.15e-24 | 16541 | 4258 | 12283 | 0.5964 | 0.03806 | 1.816 | 2.53e-55 | 24362 | 6366 | 17996 | 0.5622 | 0.03973 | Cough | respiratory | 25DTot | |
| 512.9 | 0.475 | 0.05585 | 1.608 | 1.82e-17 | 17400 | 5117 | 12283 | 0.3523 | 0.04133 | 1.422 | 1.55e-17 | 23359 | 5363 | 17996 | 0.2404 | -0.08064 | Other dyspnea | respiratory | 25DTot | |
| 513 | 0.3733 | 0.09735 | 1.453 | 1.26e-04 | 26022 | 945 | 25077 | 0.3457 | 0.08392 | 1.413 | 3.79e-05 | 35312 | 811 | 34501 | 0.5893 | 0.06741 | Respiratory abnormalities | respiratory | 25DTot | |
| 513.3 | | | | | 25286 | 209 | 25077 | | | | | 34707 | 206 | 34501 | | | Hypoventilation | respiratory | 25DTot | |
| 513.31 | | | | | 25126 | 49 | 25077 | | | | | 34541 | 40 | 34501 | | | Apnea | respiratory | 25DTot | |
| 513.32 | | | | | 25149 | 72 | 25077 | | | | | 34549 | 48 | 34501 | | | Orthopnea | respiratory | 25DTot | |
| 513.4 | | | | | 25445 | 368 | 25077 | | | | | 34837 | 336 | 34501 | 0.04425 | 0.457 | Hyperventilation | respiratory | 25DTot | |
| 513.8 | | | | | 25261 | 184 | 25077 | | | | | 34580 | 79 | 34501 | | | Disorders of diaphragm | respiratory | 25DTot | |
| 514 | 0.08819 | 0.06184 | 1.092 | 0.1538 | 24679 | 2921 | 21758 | 0.07328 | 0.04988 | 1.076 | 0.1418 | 33770 | 2584 | 31186 | 0.721 | 0.02776 | Abnormal findings examination of lungs | respiratory | 25DTot | |
| 514.1 | | | | | 21848 | 90 | 21758 | | | | | 31294 | 108 | 31186 | | | Abnormal results of function study of pulmonary system | respiratory | 25DTot | |
| 514.2 | 0.1955 | 0.09396 | 1.216 | 0.03751 | 22786 | 1028 | 21758 | 0.01977 | 0.07631 | 1.02 | 0.7956 | 32192 | 1006 | 31186 | 0.3042 | -0.1218 | Solitary pulmonary nodule | respiratory | 25DTot | |

| phenotype | male_beta | male_SE | male_OR | male_p | male_n_total | male_n_cases | male_n_controls | female_beta | female_SE | female_OR | female_p | female_n_total | female_n_cases | female_n_controls | interaction_p | interaction_beta | description | group | lab | label |
|---|---|---|---|---|---|---|---|---|---|---|---|---|---|---|---|---|---|---|---|---|
| 516 | 0.5435 | 0.1187 | 1.722 | 4.67e-06 | 27219 | 552 | 26667 | | | | | 36488 | 334 | 36154 | 0.2543 | -0.1944 | Abnormal sputum | respiratory | 25DTot | |
| 516.1 | | | | | 27145 | 478 | 26667 | | | | | 36446 | 292 | 36154 | 0.5817 | -0.1017 | Hemoptysis | respiratory | 25DTot | |
| 519 | 0.2955 | 0.08471 | 1.344 | 4.87e-04 | 25596 | 1340 | 24256 | 0.2644 | 0.06605 | 1.303 | 6.24e-05 | 34332 | 1351 | 32981 | 0.4868 | 0.07272 | Other diseases of respiratory system, not elsewhere classified | respiratory | 25DTot | |
| 519.1 | | | | | 24327 | 71 | 24256 | | | | | 33033 | 52 | 32981 | | | Tracheostomy complications | respiratory | 25DTot | |
| 519.2 | | | | | 24383 | 127 | 24256 | | | | | 33036 | 55 | 32981 | | | Respiratory complications | respiratory | 25DTot | |
| 519.8 | | | | | 24432 | 176 | 24256 | | | | | 33153 | 172 | 32981 | | | Other diseases of respiratory system, NEC | respiratory | 25DTot | |
| 519.9 | 0.2912 | 0.102 | 1.338 | 0.004328 | 25196 | 940 | 24256 | 0.3063 | 0.07538 | 1.358 | 4.83e-05 | 34006 | 1025 | 32981 | 0.3332 | 0.1194 | Symptoms involving respiratory system and other chest symptoms | respiratory | 25DTot | |
| 520 | | | | | 25518 | 159 | 25359 | | | | | 33516 | 257 | 33259 | | | Disorders of tooth development | digestive | 25DTot | |
| 520.1 | | | | | 25370 | 11 | 25359 | | | | | 33270 | 11 | 33259 | | | Hereditary disturbances in tooth structure | digestive | 25DTot | |
| 520.2 | | | | | 25503 | 144 | 25359 | | | | | 33503 | 244 | 33259 | | | Disturbances in tooth eruption | digestive | 25DTot | |
| 521 | 0.4155 | 0.1042 | 1.515 | 6.63e-05 | 26066 | 707 | 25359 | 0.4821 | 0.07752 | 1.62 | 4.99e-10 | 34146 | 887 | 33259 | 0.2308 | 0.1522 | Diseases of hard tissues of teeth | digestive | 25DTot | |
| 521.1 | 0.401 | 0.1057 | 1.493 | 1.49e-04 | 26050 | 691 | 25359 | 0.4938 | 0.07845 | 1.638 | 3.09e-10 | 34122 | 863 | 33259 | 0.17 | 0.1767 | Dental caries | digestive | 25DTot | |
| 521.2 | | | | | 25360 | 1 | 25359 | | | | | 33261 | 2 | 33259 | | | Dental abrasion, erosion and attrition | digestive | 25DTot | |
| 521.4 | | | | | 25359 | | | | | | | 33267 | 8 | 33259 | | | Tooth complications likely association with other diseases | digestive | 25DTot | |
| 522 | | | | | 25451 | 92 | 25359 | | | | | 33377 | 118 | 33259 | | | Diseases of pulp and periapical tissues | digestive | 25DTot | |
| 522.1 | | | | | 25361 | 2 | 25359 | | | | | 33263 | 4 | 33259 | | | Pulpitis and necrosis of tooth pulp | digestive | 25DTot | |
| 522.5 | | | | | 25445 | 86 | 25359 | | | | | 33365 | 106 | 33259 | | | Periapical abscess | digestive | 25DTot | |
| 523 | | | | | 25698 | 339 | 25359 | 0.5959 | 0.09495 | 1.815 | 3.49e-10 | 33819 | 560 | 33259 | 0.482 | 0.117 | Gingival and periodontal diseases | digestive | 25DTot | |
| 523.1 | | | | | 25536 | 177 | 25359 | | | | | 33688 | 429 | 33259 | 0.08829 | 0.3784 | Gingivitis | digestive | 25DTot | |
| 523.3 | | | | | 25492 | 133 | 25359 | | | | | 33407 | 148 | 33259 | | | Periodontitis (acute or chronic) | digestive | 25DTot | |
| 523.31 | | | | | 25454 | 95 | 25359 | | | | | 33368 | 109 | 33259 | | | Acute periodontitis | digestive | 25DTot | |
| 523.32 | | | | | 25382 | 23 | 25359 | | | | | 33285 | 26 | 33259 | | | Chronic periodontitis | digestive | 25DTot | |
| 524 | | | | | 25495 | 136 | 25359 | | | | | 33518 | 259 | 33259 | | | Dentofacial anomalies, including malocclusion | digestive | 25DTot | |
| 524.3 | | | | | 25481 | 122 | 25359 | | | | | 33493 | 234 | 33259 | | | Anomalies of tooth position/malocclusion | digestive | 25DTot | |
| 525 | | | | | 25595 | 236 | 25359 | | | | | 33641 | 382 | 33259 | 0.9774 | 0.005959 | Other diseases of the teeth and supporting structures | digestive | 25DTot | |
| 525.1 | | | | | 25495 | 136 | 25359 | | | | | 33466 | 207 | 33259 | | | Loss of teeth or edentulism | digestive | 25DTot | |
| 525.2 | | | | | 25366 | 7 | 25359 | | | | | 33279 | 20 | 33259 | | | Atrophy of edentulous alveolar ridge | digestive | 25DTot | |
| 526 | | | | | 25636 | 277 | 25359 | 0.4183 | 0.09553 | 1.519 | 1.19e-05 | 33838 | 579 | 33259 | 0.007906 | 0.5534 | Diseases of the jaws | digestive | 25DTot | |
| 526.1 | | | | | 25372 | 13 | 25359 | | | | | 33270 | 11 | 33259 | | | Cysts of the jaws | digestive | 25DTot | |
| 526.3 | | | | | 25393 | 34 | 25359 | | | | | 33336 | 77 | 33259 | | | Anomalies of jaw size/symmetry | digestive | 25DTot | |
| 526.4 | | | | | 25426 | 67 | 25359 | | | | | 33523 | 264 | 33259 | | | Temporomandibular joint disorders | digestive | 25DTot | |
| 526.41 | | | | | 25407 | 48 | 25359 | | | | | 33417 | 158 | 33259 | | | Temporomandibular joint disorder, unspecified | digestive | 25DTot | |
| 526.42 | | | | | 25364 | 5 | 25359 | | | | | 33300 | 41 | 33259 | | | Arthralgia/ankylosis of temporomandibular joint | digestive | 25DTot | |
| 526.5 | | | | | 25416 | 57 | 25359 | | | | | 33320 | 61 | 33259 | | | Inflammatory conditions of jaw | digestive | 25DTot | |
| 526.8 | | | | | 25363 | 4 | 25359 | | | | | 33267 | 8 | 33259 | | | Exostosis of jaw | digestive | 25DTot | |
| 526.9 | | | | | 25414 | 55 | 25359 | | | | | 33304 | 45 | 33259 | | | Jaw disease NOS | digestive | 25DTot | |
| 527 | | | | | 25962 | 420 | 25542 | | | | | 34324 | 353 | 33971 | 0.001673 | 0.637 | Diseases of the salivary glands | digestive | 25DTot | |
| 527.1 | | | | | 25545 | 3 | 25542 | | | | | 33976 | 5 | 33971 | | | Hypertrophy of salivary gland | digestive | 25DTot | |

| phenotype | male_beta | male_SE | male_OR | male_p | male_n_total | male_n_cases | male_n_controls | female_beta | female_SE | female_OR | female_p | female_n_total | female_n_cases | female_n_controls | interaction_p | interaction_beta | description | group | lab | label |
|---|---|---|---|---|---|---|---|---|---|---|---|---|---|---|---|---|---|---|---|---|
| 527.2 | | | | | 25596 | 54 | 25542 | | | | | 34069 | 98 | 33971 | | | Sialoadenitis | digestive | 25DTot | |
| 527.7 | | | | | 25871 | 329 | 25542 | | | | | 34176 | 205 | 33971 | 0.004005 | 0.7529 | Disturbance of salivary secretion | digestive | 25DTot | |
| 527.8 | | | | | 25561 | 19 | 25542 | | | | | 34003 | 32 | 33971 | | | Other specified diseases of the salivary glands | digestive | 25DTot | |
| 528 | 0.158 | 0.1116 | 1.171 | 0.157 | 26317 | 775 | 25542 | 0.3109 | 0.07983 | 1.365 | 9.82e-05 | 34849 | 878 | 33971 | 0.04209 | 0.273 | Diseases of the oral soft tissues, excluding lesions specific for gingiva and tongue | digestive | 25DTot | |
| 528.1 | | | | | 25938 | 396 | 25542 | | | | | 34341 | 370 | 33971 | 0.5483 | 0.1289 | Stomatitis and mucositis | digestive | 25DTot | |
| 528.11 | | | | | 25631 | 89 | 25542 | | | | | 34056 | 85 | 33971 | | | Stomatitis and mucositis (ulcerative) | digestive | 25DTot | |
| 528.12 | | | | | 25583 | 41 | 25542 | | | | | 34026 | 55 | 33971 | | | Oral aphthae | digestive | 25DTot | |
| 528.3 | | | | | 25579 | 37 | 25542 | | | | | 34020 | 49 | 33971 | | | Cellulitis and abscess of oral soft tissues | digestive | 25DTot | |
| 528.4 | | | | | 25551 | 9 | 25542 | | | | | 33984 | 13 | 33971 | | | Cysts of oral soft tissues | digestive | 25DTot | |
| 528.41 | | | | | 25551 | 9 | 25542 | | | | | 33982 | 11 | 33971 | | | Cyst of the salivary gland | digestive | 25DTot | |
| 528.5 | | | | | 25587 | 45 | 25542 | | | | | 34050 | 79 | 33971 | | | Diseases of lips | digestive | 25DTot | |
| 528.6 | | | | | 25578 | 36 | 25542 | | | | | 34015 | 44 | 33971 | | | Leukoplakia of oral mucosa | digestive | 25DTot | |
| 528.7 | | | | | 25551 | 9 | 25542 | | | | | 33984 | 13 | 33971 | | | Sialolithiasis | digestive | 25DTot | |
| 529 | | | | | 25647 | 105 | 25542 | | | | | 34108 | 137 | 33971 | | | Diseases and other conditions of the tongue | digestive | 25DTot | |
| 529.1 | | | | | 25565 | 23 | 25542 | | | | | 34012 | 41 | 33971 | | | Glossitis | digestive | 25DTot | |
| 529.6 | | | | | 25565 | 23 | 25542 | | | | | 34004 | 33 | 33971 | | | Glossodynia | digestive | 25DTot | |
| 530 | 0.6164 | 0.04627 | 1.852 | 1.75e-40 | 22713 | 7010 | 15703 | 0.511 | 0.03225 | 1.667 | 1.54e-56 | 30970 | 8921 | 22049 | 0.2808 | -0.06032 | Diseases of esophagus | digestive | 25DTot | |
| 530.1 | 0.5899 | 0.04771 | 1.804 | 4.11e-35 | 22053 | 6350 | 15703 | 0.5133 | 0.03282 | 1.671 | 3.97e-55 | 30445 | 8396 | 22049 | 0.4715 | -0.04134 | Esophagitis, GERD and related diseases | digestive | 25DTot | |
| 530.11 | 0.5931 | 0.04886 | 1.81 | 6.62e-34 | 21565 | 5862 | 15703 | 0.509 | 0.0333 | 1.664 | 9.82e-53 | 30082 | 8033 | 22049 | 0.2939 | -0.06151 | GERD | digestive | 25DTot | |
| 530.12 | | | | | 15778 | 75 | 15703 | | | | | 22113 | 64 | 22049 | | | Ulcer of esophagus | digestive | 25DTot | |
| 530.13 | | | | | 16021 | 318 | 15703 | | | | | 22193 | 144 | 22049 | | | Barrett's esophagus | digestive | 25DTot | |
| 530.14 | | | | | 16000 | 297 | 15703 | | | | | 22411 | 362 | 22049 | 0.4743 | 0.1334 | Reflux esophagitis | digestive | 25DTot | |
| 530.15 | | | | | 15777 | 74 | 15703 | | | | | 22097 | 48 | 22049 | | | Eosinophilic esophagitis | digestive | 25DTot | |
| 530.2 | 1.074 | 0.1045 | 2.927 | 8.89e-25 | 16327 | 624 | 15703 | | | | | 22465 | 416 | 22049 | 0.09248 | -0.2506 | Esophageal bleeding (varices/hemorrhage) | digestive | 25DTot | |
| 530.3 | | | | | 16140 | 437 | 15703 | | | | | 22429 | 380 | 22049 | 0.297 | 0.1848 | Stricture and stenosis of esophagus | digestive | 25DTot | |
| 530.5 | | | | | 15823 | 120 | 15703 | | | | | 22257 | 208 | 22049 | | | Disorders of esophageal motility | digestive | 25DTot | |
| 530.6 | | | | | 15740 | 37 | 15703 | | | | | 22084 | 35 | 22049 | | | Diverticulum of esophagus, acquired | digestive | 25DTot | |
| 530.7 | | | | | 15735 | 32 | 15703 | | | | | 22067 | 18 | 22049 | | | Gastroesophageal laceration-hemorrhage syndrome | digestive | 25DTot | |
| 530.9 | | | | | 15920 | 217 | 15703 | | | | | 22322 | 273 | 22049 | | | Heartburn | digestive | 25DTot | |
| 531 | 0.5505 | 0.09667 | 1.734 | 1.24e-08 | 26644 | 757 | 25887 | 0.6051 | 0.07952 | 1.831 | 2.74e-14 | 35675 | 773 | 34902 | 0.6279 | 0.05905 | Peptic ulcer (excl. esophageal) | digestive | 25DTot | |
| 531.1 | | | | | 26046 | 159 | 25887 | | | | | 35004 | 102 | 34902 | | | Hemorrhage from gastrointestinal ulcer | digestive | 25DTot | |
| 531.2 | | | | | 26063 | 176 | 25887 | | | | | 35135 | 233 | 34902 | | | Gastric ulcer | digestive | 25DTot | |
| 531.3 | | | | | 26049 | 162 | 25887 | | | | | 35017 | 115 | 34902 | | | Duodenal ulcer | digestive | 25DTot | |
| 531.4 | | | | | 26128 | 241 | 25887 | | | | | 35173 | 271 | 34902 | 0.598 | 0.1127 | Peptic ulcer, site unspecified | digestive | 25DTot | |
| 531.5 | | | | | 25904 | 17 | 25887 | | | | | 34952 | 50 | 34902 | | | Gastrojejunal ulcer | digestive | 25DTot | |
| 532 | 0.4291 | 0.0641 | 1.536 | 2.17e-11 | 18697 | 2994 | 15703 | 0.4756 | 0.04679 | 1.609 | 2.87e-24 | 25127 | 3078 | 22049 | 0.03868 | 0.1616 | Dysphagia | digestive | 25DTot | |
| 535 | 0.8576 | 0.09996 | 2.357 | 9.54e-18 | 24211 | 698 | 23513 | 0.8243 | 0.07733 | 2.28 | 1.58e-26 | 32091 | 888 | 31203 | 0.5513 | 0.07296 | Gastritis and duodenitis | digestive | 25DTot | |

| phenotype | male_beta | male_SE | male_OR | male_p | male_n_total | male_n_cases | male_n_controls | female_beta | female_SE | female_OR | female_p | female_n_total | female_n_cases | female_n_controls | interaction_p | interaction_beta | description | group | lab | label |
|---|---|---|---|---|---|---|---|---|---|---|---|---|---|---|---|---|---|---|---|---|
| 535.1 | | | | | 23554 | 41 | 23513 | | | | | 31256 | 53 | 31203 | | | Acute gastritis | digestive | 25DTot | |
| 535.2 | | | | | 23753 | 240 | 23513 | | | | | 31519 | 316 | 31203 | 0.0179 | 0.4802 | Atrophic gastritis | digestive | 25DTot | |
| 535.6 | | | | | 23634 | 121 | 23513 | | | | | 31294 | 91 | 31203 | | | Duodenitis | digestive | 25DTot | |
| 535.8 | | | | | 23673 | 160 | 23513 | | | | | 31440 | 237 | 31203 | | | Other specified gastritis | digestive | 25DTot | |
| 535.9 | | | | | 23653 | 140 | 23513 | | | | | 31383 | 180 | 31203 | | | Gastritis and duodenitis, NOS | digestive | 25DTot | |
| 536 | 0.6652 | 0.1109 | 1.945 | 1.98e-09 | 24111 | 598 | 23513 | 0.6612 | 0.07053 | 1.937 | 6.89e-21 | 32274 | 1071 | 31203 | 0.586 | 0.0697 | Disorders of function of stomach | digestive | 25DTot | |
| 536.3 | | | | | 23695 | 182 | 23513 | | | | | 31598 | 395 | 31203 | 0.6495 | -0.101 | Gastroparesis | digestive | 25DTot | |
| 536.7 | | | | | 23722 | 209 | 23513 | | | | | 31426 | 223 | 31203 | | | Complications of gastrostomy, colostomy and enterostomy | digestive | 25DTot | |
| 536.8 | | | | | 23738 | 225 | 23513 | 0.9328 | 0.09781 | 2.542 | 1.48e-21 | 31704 | 501 | 31203 | 0.5234 | 0.1189 | Dyspepsia and other specified disorders of function of stomach | digestive | 25DTot | |
| 537 | 0.8694 | 0.0982 | 2.385 | 8.53e-19 | 24195 | 682 | 23513 | 0.6651 | 0.08489 | 1.945 | 4.68e-15 | 31926 | 723 | 31203 | 0.1277 | -0.1915 | Other disorders of stomach and duodenum | digestive | 25DTot | |
| 537.1 | | | | | 23574 | 61 | 23513 | | | | | 31261 | 58 | 31203 | | | Lesions of stomach and duodenum | digestive | 25DTot | |
| 539 | | | | | 27703 | 290 | 27413 | 1.004 | 0.0721 | 2.728 | 4.84e-44 | 36747 | 945 | 35802 | 0.1876 | -0.1944 | Bariatric surgery | digestive | 25DTot | |
| 540 | | | | | 27635 | 152 | 27483 | | | | | 36729 | 193 | 36536 | | | Appendiceal conditions | digestive | 25DTot | |
| 540.1 | | | | | 27611 | 128 | 27483 | | | | | 36714 | 178 | 36536 | | | Appendicitis | digestive | 25DTot | |
| 540.11 | | | | | 27592 | 109 | 27483 | | | | | 36692 | 156 | 36536 | | | Acute appendicitis | digestive | 25DTot | |
| 550 | 0.4458 | 0.05622 | 1.562 | 2.18e-15 | 25722 | 2883 | 22839 | 0.2513 | 0.04832 | 1.286 | 1.99e-07 | 34582 | 2531 | 32051 | 8.16e-04 | -0.2427 | Abdominal hernia | digestive | 25DTot | |
| 550.1 | 0.2466 | 0.09188 | 1.28 | 0.007267 | 23813 | 974 | 22839 | | | | | 32167 | 116 | 32051 | 0.09268 | -0.3895 | Inguinal hernia | digestive | 25DTot | |
| 550.2 | 0.5351 | 0.08801 | 1.708 | 1.20e-09 | 23793 | 954 | 22839 | 0.3753 | 0.06106 | 1.455 | 7.90e-10 | 33474 | 1423 | 32051 | 0.08247 | -0.1821 | Diaphragmatic hernia | digestive | 25DTot | |
| 550.3 | | | | | 22846 | 7 | 22839 | | | | | 32067 | 16 | 32051 | | | Femoral hernia | digestive | 25DTot | |
| 550.4 | | | | | 23250 | 411 | 22839 | | | | | 32268 | 217 | 32051 | 4.21e-05 | -0.8052 | Umbilical hernia | digestive | 25DTot | |
| 550.5 | | | | | 23322 | 483 | 22839 | 0.08369 | 0.1046 | 1.087 | 0.4238 | 32584 | 533 | 32051 | 0.04166 | -0.3279 | Ventral hernia | digestive | 25DTot | |
| 550.6 | 0.408 | 0.124 | 1.504 | 0.001003 | 23364 | 525 | 22839 | 0.112 | 0.1016 | 1.118 | 0.2702 | 32610 | 559 | 32051 | 0.109 | -0.2514 | Incisional hernia | digestive | 25DTot | |
| 555 | 2.19 | 0.07488 | 8.935 | 5.10e-188 | 18550 | 1092 | 17458 | 1.742 | 0.06116 | 5.709 | 2.05e-178 | 24902 | 1438 | 23464 | 8.02e-05 | -0.3639 | Inflammatory bowel disease and other gastroenteritis and colitis | digestive | 25DTot | |
| 555.1 | 2.447 | 0.08239 | 11.56 | 6.54e-194 | 18342 | 884 | 17458 | 1.992 | 0.06668 | 7.329 | 4.55e-196 | 24671 | 1207 | 23464 | 4.26e-04 | -0.352 | Regional enteritis | digestive | 25DTot | |
| 555.2 | 2.139 | 0.09817 | 8.488 | 3.28e-105 | 18001 | 543 | 17458 | 1.641 | 0.08723 | 5.16 | 6.23e-79 | 24116 | 652 | 23464 | 9.48e-04 | -0.4159 | Ulcerative colitis | digestive | 25DTot | |
| 555.21 | | | | | 17689 | 231 | 17458 | | | | | 23710 | 246 | 23464 | | | Ulcerative colitis (chronic) | digestive | 25DTot | |
| 556 | | | | | 17670 | 212 | 17458 | | | | | 23667 | 203 | 23464 | | | Ulceration of the lower GI tract | digestive | 25DTot | |
| 556.1 | | | | | 17633 | 175 | 17458 | | | | | 23632 | 168 | 23464 | | | Ulceration of intestine | digestive | 25DTot | |
| 556.11 | | | | | 17498 | 40 | 17458 | | | | | 23504 | 40 | 23464 | | | Angiodysplasia of intestine (without mention of hemorrhage) | digestive | 25DTot | |

| phenotype | male_beta | male_SE | male_OR | male_p | male_n_total | male_n_cases | male_n_controls | female_beta | female_SE | female_OR | female_p | female_n_total | female_n_cases | female_n_controls | interaction_p | interaction_beta | description | group | lab | label |
|---|---|---|---|---|---|---|---|---|---|---|---|---|---|---|---|---|---|---|---|---|
| 557 | | | | | 17645 | 187 | 17458 | | | | | 23820 | 356 | 23464 | 0.48 | -0.1387 | Intestinal malabsorption (non-celiac) | digestive | 25DTot | |
| 557.1 | | | | | 17538 | 80 | 17458 | | | | | 23734 | 270 | 23464 | | | Celiac disease | digestive | 25DTot | |
| 558 | 1.828 | 0.07596 | 6.221 | 5.64e-128 | 18499 | 1041 | 17458 | 1.325 | 0.05863 | 3.761 | 5.03e-113 | 25010 | 1546 | 23464 | 3.30e-05 | -0.3863 | Noninfectious gastroenteritis | digestive | 25DTot | |
| 559 | | | | | 17873 | 415 | 17458 | | | | | 23811 | 347 | 23464 | 0.1879 | -0.2202 | Ileostomy status | digestive | 25DTot | |
| 560 | 0.9932 | 0.0785 | 2.7 | 1.08e-36 | 18805 | 1347 | 17458 | 0.7245 | 0.06998 | 2.064 | 4.07e-25 | 24662 | 1198 | 23464 | 0.02715 | -0.2268 | Intestinal obstruction without mention of hernia | digestive | 25DTot | |
| 560.1 | 0.7354 | 0.1129 | 2.086 | 7.31e-11 | 18153 | 695 | 17458 | | | | | 23849 | 385 | 23464 | 0.2893 | -0.1727 | Paralytic ileus | digestive | 25DTot | |
| 560.2 | | | | | 17514 | 56 | 17458 | | | | | 23568 | 104 | 23464 | | | Impaction of intestine | digestive | 25DTot | |
| 560.3 | | | | | 17538 | 80 | 17458 | | | | | 23554 | 90 | 23464 | | | Peritoneal or intestinal adhesions | digestive | 25DTot | |

| phenotype | male_beta | male_SE | male_OR | male_p | male_n_total | male_n_cases | male_n_controls | female_beta | female_SE | female_OR | female_p | female_n_total | female_n_cases | female_n_controls | interaction_p | interaction_beta | description | group | lab | label |
|---|---|---|---|---|---|---|---|---|---|---|---|---|---|---|---|---|---|---|---|---|
| 560.4 | 1.218 | 0.09948 | 3.381 | 1.76e-34 | 18149 | 691 | 17458 | 0.8234 | 0.08564 | 2.278 | 6.94e-22 | 24229 | 765 | 23464 | 0.003615 | -0.3721 | Other intestinal obstruction | digestive | 25DTot | |
| 561 | 0.8623 | 0.07063 | 2.369 | 2.77e-34 | 19227 | 1769 | 17458 | 0.7259 | 0.04621 | 2.067 | 1.32e-55 | 26443 | 2979 | 23464 | 0.3824 | -0.07269 | Symptoms involving digestive system | digestive | 25DTot | |
| 562 | 0.687 | 0.06713 | 1.988 | 1.40e-24 | 19778 | 2320 | 17458 | 0.7733 | 0.0523 | 2.167 | 1.82e-49 | 25685 | 2221 | 23464 | 0.2963 | 0.08806 | Diverticulosis and diverticulitis | digestive | 25DTot | |
| 562.1 | 0.7002 | 0.06923 | 2.014 | 4.76e-24 | 19570 | 2112 | 17458 | 0.8271 | 0.05499 | 2.287 | 3.96e-51 | 25403 | 1939 | 23464 | 0.1292 | 0.133 | Diverticulosis | digestive | 25DTot | |
| 562.2 | | | | | 17837 | 379 | 17458 | 0.7795 | 0.0982 | 2.18 | 2.06e-15 | 23966 | 502 | 23464 | 0.7354 | 0.05646 | Diverticulitis | digestive | 25DTot | |
| 564 | 1.099 | 0.0739 | 3.003 | 4.61e-50 | 18857 | 1399 | 17458 | 0.8878 | 0.04829 | 2.43 | 1.76e-75 | 26038 | 2574 | 23464 | 0.08135 | -0.1515 | Functional digestive disorders | digestive | 25DTot | |
| 564.1 | | | | | 17914 | 456 | 17458 | 1.009 | 0.05845 | 2.743 | 9.20e-67 | 25043 | 1579 | 23464 | 1.13e-04 | -0.4689 | Irritable Bowel Syndrome | digestive | 25DTot | |
| 564.9 | | | | | 17664 | 206 | 17458 | | | | | 23673 | 209 | 23464 | | | Personal history of diseases of digestive system | digestive | 25DTot | |
| 565 | 1.093 | 0.07777 | 2.983 | 7.27e-45 | 23018 | 1093 | 21925 | 0.9649 | 0.06796 | 2.625 | 9.36e-46 | 31069 | 1106 | 29963 | 0.7985 | -0.02547 | Anal and rectal conditions | digestive | 25DTot | |
| 565.1 | | | | | 22052 | 127 | 21925 | | | | | 30077 | 114 | 29963 | | | Anal and rectal polyp | digestive | 25DTot | |
| 567 | 0.9528 | 0.1067 | 2.593 | 4.28e-19 | 22577 | 652 | 21925 | 0.6143 | 0.09903 | 1.848 | 5.54e-10 | 30541 | 578 | 29963 | 0.168 | -0.1936 | Peritonitis and retroperitoneal infections | digestive | 25DTot | |
| 568 | | | | | 22412 | 487 | 21925 | 0.4435 | 0.1069 | 1.558 | 3.35e-05 | 30476 | 513 | 29963 | 0.9092 | -0.01908 | Other disorders of peritoneum | digestive | 25DTot | |
| 568.1 | | | | | 22164 | 239 | 21925 | | | | | 30277 | 314 | 29963 | 0.8763 | 0.03573 | Peritoneal adhesions (postoperative) (postinfection) | digestive | 25DTot | |
| 569 | 1.175 | 0.06691 | 3.237 | 5.67e-69 | 23419 | 1494 | 21925 | 0.9864 | 0.05148 | 2.682 | 7.76e-82 | 31975 | 2012 | 29963 | 0.1621 | -0.1143 | Other disorders of intestine | digestive | 25DTot | |
| 569.1 | | | | | 21954 | 29 | 21925 | | | | | 30021 | 58 | 29963 | | | Toxic gastroenteritis and colitis | digestive | 25DTot | |
| 569.2 | 1.399 | 0.09895 | 4.052 | 2.15e-45 | 22496 | 571 | 21925 | 1.133 | 0.07163 | 3.104 | 2.58e-56 | 30942 | 979 | 29963 | 0.04062 | -0.2406 | Gastrointestinal complications | digestive | 25DTot | |
| 571 | 0.7585 | 0.06088 | 2.135 | 1.26e-35 | 21552 | 2387 | 19165 | 0.3733 | 0.0524 | 1.452 | 1.05e-12 | 29934 | 2235 | 27699 | 1.61e-04 | -0.2957 | Chronic liver disease and cirrhosis | digestive | 25DTot | |
| 571.5 | 0.7664 | 0.06244 | 2.152 | 1.25e-34 | 21382 | 2217 | 19165 | 0.3468 | 0.05448 | 1.415 | 1.93e-10 | 29754 | 2055 | 27699 | 6.27e-05 | -0.3236 | Other chronic nonalcoholic liver disease | digestive | 25DTot | |
| 571.51 | 0.8838 | 0.07743 | 2.42 | 3.58e-30 | 20477 | 1312 | 19165 | 0.4704 | 0.07813 | 1.601 | 1.73e-09 | 28658 | 959 | 27699 | 4.50e-04 | -0.3737 | Cirrhosis of liver without mention of alcohol | digestive | 25DTot | |
| 571.6 | | | | | 19242 | 77 | 19165 | | | | | 27883 | 184 | 27699 | | | Primary biliary cirrhosis | digestive | 25DTot | |
| 571.8 | 1.006 | 0.08513 | 2.735 | 3.18e-32 | 20161 | 996 | 19165 | 0.5258 | 0.0884 | 1.692 | 2.71e-09 | 28437 | 738 | 27699 | 1.81e-04 | -0.4445 | Liver abscess and sequelae of chronic liver disease | digestive | 25DTot | |
| 571.81 | 1.13 | 0.09701 | 3.095 | 2.43e-31 | 19856 | 691 | 19165 | | | | | 28168 | 469 | 27699 | 0.001368 | -0.447 | Portal hypertension | digestive | 25DTot | |
| 572 | 0.8246 | 0.07968 | 2.281 | 4.23e-25 | 20559 | 1394 | 19165 | 0.352 | 0.07419 | 1.422 | 2.10e-06 | 28923 | 1224 | 27699 | 7.27e-05 | -0.4188 | Ascites (non malignant) | digestive | 25DTot | |
| 573 | 0.6014 | 0.09483 | 1.825 | 2.26e-10 | 20126 | 961 | 19165 | 0.2932 | 0.08001 | 1.341 | 2.48e-04 | 28655 | 956 | 27699 | 0.02352 | -0.2737 | Other disorders of liver | digestive | 25DTot | |
| 573.1 | | | | | 19195 | 30 | 19165 | | | | | 27731 | 32 | 27699 | | | Chronic passive congestion of liver | digestive | 25DTot | |
| 573.2 | | | | | 19596 | 431 | 19165 | | | | | 27945 | 246 | 27699 | 0.1122 | -0.2701 | Liver replaced by transplant | digestive | 25DTot | |
| 573.3 | | | | | 19292 | 127 | 19165 | | | | | 27860 | 161 | 27699 | | | Hepatomegaly | digestive | 25DTot | |
| 573.4 | | | | | 19356 | 191 | 19165 | | | | | 27881 | 182 | 27699 | | | Acute and subacute necrosis of liver | digestive | 25DTot | |
| 573.5 | | | | | 19606 | 441 | 19165 | | | | | 28006 | 307 | 27699 | 0.5467 | -0.1195 | Jaundice (not of newborn) | digestive | 25DTot | |
| 573.6 | 0.8595 | 0.1042 | 2.362 | 1.59e-16 | 19797 | 632 | 19165 | 0.6001 | 0.09623 | 1.822 | 4.49e-10 | 28268 | 569 | 27699 | 0.1727 | -0.1878 | Nonspecific elevation of levels of transaminase or lactic acid dehydrogenase [LDH] | digestive | 25DTot | |
| 573.7 | 0.8309 | 0.08582 | 2.295 | 3.59e-22 | 20119 | 954 | 19165 | 0.6698 | 0.06742 | 1.954 | 2.91e-23 | 28828 | 1129 | 27699 | 0.3514 | -0.09922 | Abnormal results of function study of liver | digestive | 25DTot | |
| 573.9 | 0.7926 | 0.09562 | 2.209 | 1.14e-16 | 19944 | 779 | 19165 | 0.3503 | 0.0789 | 1.42 | 8.98e-06 | 28587 | 888 | 27699 | 0.002137 | -0.3717 | Abnormal serum enzyme levels | digestive | 25DTot | |

| phenotype | male_beta | male_SE | male_OR | male_p | male_n_total | male_n_cases | male_n_controls | female_beta | female_SE | female_OR | female_p | female_n_total | female_n_cases | female_n_controls | interaction_p | interaction_beta | description | group | lab | label |
|---|---|---|---|---|---|---|---|---|---|---|---|---|---|---|---|---|---|---|---|---|
| 574 | 0.5722 | 0.07596 | 1.772 | 4.95e-14 | 25697 | 1283 | 24414 | 0.3309 | 0.06061 | 1.392 | 4.77e-08 | 34656 | 1503 | 33153 | 0.0161 | -0.228 | Cholelithiasis and cholecystitis | digestive | 25DTot | |
| 574.1 | 0.593 | 0.08451 | 1.809 | 2.27e-12 | 25404 | 990 | 24414 | 0.337 | 0.0682 | 1.401 | 7.78e-07 | 34320 | 1167 | 33153 | 0.01251 | -0.2643 | Cholelithiasis | digestive | 25DTot | |
| 574.11 | | | | | 24503 | 89 | 24414 | | | | | 33224 | 71 | 33153 | | | Cholelithiasis with acute cholecystitis | digestive | 25DTot | |
| 574.12 | | | | | 24528 | 114 | 24414 | | | | | 33340 | 187 | 33153 | | | Cholelithiasis with other cholecystitis | digestive | 25DTot | |
| 574.2 | | | | | 24625 | 211 | 24414 | | | | | 33383 | 230 | 33153 | | | Calculus of bile duct | digestive | 25DTot | |
| 574.3 | | | | | 24711 | 297 | 24414 | | | | | 33444 | 291 | 33153 | 0.759 | 0.06295 | Cholecystitis without cholelithiasis | digestive | 25DTot | |
| 575 | 0.7146 | 0.09176 | 2.043 | 6.81e-15 | 25285 | 871 | 24414 | 0.3526 | 0.07789 | 1.423 | 5.97e-06 | 34100 | 947 | 33153 | 0.01379 | -0.2877 | Other biliary tract disease | digestive | 25DTot | |
| 575.1 | | | | | 24612 | 198 | 24414 | | | | | 33277 | 124 | 33153 | | | Cholangitis | digestive | 25DTot | |
| 575.2 | | | | | 24699 | 285 | 24414 | | | | | 33392 | 239 | 33153 | 0.02825 | -0.493 | Obstruction of bile duct | digestive | 25DTot | |
| 575.6 | | | | | 24435 | 21 | 24414 | | | | | 33185 | 32 | 33153 | | | Cholesterolosis of gallbladder | digestive | 25DTot | |
| 575.7 | | | | | 24611 | 197 | 24414 | | | | | 33370 | 217 | 33153 | | | Other disorders of gallbladder | digestive | 25DTot | |
| 575.8 | | | | | 24783 | 369 | 24414 | | | | | 33584 | 431 | 33153 | 0.009421 | -0.4366 | Other disorders of biliary tract | digestive | 25DTot | |
| 575.9 | | | | | 24488 | 74 | 24414 | | | | | 33244 | 91 | 33153 | | | Nonspecific abnormal findings on radiological and other examination of biliary tract | digestive | 25DTot | |
| 577 | 0.8337 | 0.08733 | 2.302 | 1.34e-21 | 27246 | 863 | 26383 | 0.334 | 0.0821 | 1.397 | 4.73e-05 | 36384 | 840 | 35544 | 1.18e-04 | -0.4469 | Diseases of pancreas | digestive | 25DTot | |
| 577.1 | | | | | 26693 | 310 | 26383 | | | | | 35828 | 284 | 35544 | 0.3271 | -0.2016 | Acute pancreatitis | digestive | 25DTot | |
| 577.2 | | | | | 26568 | 185 | 26383 | | | | | 35726 | 182 | 35544 | | | Chronic pancreatitis | digestive | 25DTot | |
| 577.3 | | | | | 26620 | 237 | 26383 | | | | | 35810 | 266 | 35544 | 0.09453 | -0.3655 | Cyst and pseudocyst of pancreas | digestive | 25DTot | |
| 578 | 0.5712 | 0.06049 | 1.77 | 3.65e-21 | 23398 | 2404 | 20994 | 0.4307 | 0.04825 | 1.538 | 4.38e-19 | 31110 | 2548 | 28562 | 0.1659 | -0.105 | Gastrointestinal hemorrhage | digestive | 25DTot | |
| 578.1 | | | | | 21273 | 279 | 20994 | | | | | 28819 | 257 | 28562 | 0.7413 | -0.07297 | Hematemesis | digestive | 25DTot | |
| 578.2 | 0.6511 | 0.08228 | 1.918 | 2.50e-15 | 22097 | 1103 | 20994 | 0.5098 | 0.06722 | 1.665 | 3.35e-14 | 29747 | 1185 | 28562 | 0.2859 | -0.1109 | Blood in stool | digestive | 25DTot | |

| phenotype | male_beta | male_SE | male_OR | male_p | male_n_total | male_n_cases | male_n_controls | female_beta | female_SE | female_OR | female_p | female_n_total | female_n_cases | female_n_controls | interaction_p | interaction_beta | description | group | lab | label |
|---|---|---|---|---|---|---|---|---|---|---|---|---|---|---|---|---|---|---|---|---|
| 578.8 | 0.6132 | 0.1066 | 1.846 | 8.82e-09 | 21622 | 628 | 20994 | 0.4613 | 0.08547 | 1.586 | 6.77e-08 | 29288 | 726 | 28562 | 0.3696 | -0.1199 | Hemorrhage of rectum and anus | digestive | 25DTot | |
| 578.9 | 0.5772 | 0.08813 | 1.781 | 5.78e-11 | 22034 | 1040 | 20994 | 0.2709 | 0.08401 | 1.311 | 0.001261 | 29380 | 818 | 28562 | 0.008006 | -0.3143 | Hemorrhage of gastrointestinal tract | digestive | 25DTot | |
| 579 | 0.5135 | 0.07628 | 1.671 | 1.67e-11 | 22498 | 1504 | 20994 | 0.3209 | 0.05549 | 1.378 | 7.37e-09 | 30586 | 2024 | 28562 | 0.1239 | -0.1421 | Other symptoms involving abdomen and pelvis | digestive | 25DTot | |
| 579.2 | 0.7748 | 0.1106 | 2.17 | 2.46e-12 | 21574 | 580 | 20994 | | | | | 28950 | 388 | 28562 | 0.006193 | -0.4467 | Splenomegaly | digestive | 25DTot | |
| 579.8 | | | | | 21190 | 196 | 20994 | | | | | 28810 | 248 | 28562 | | | Nonspecific abnormal findings in stool contents | digestive | 25DTot | |
| 580 | 1.438 | 0.09081 | 4.212 | 1.78e-56 | 18292 | 656 | 17636 | 0.5133 | 0.09346 | 1.671 | 3.98e-08 | 28613 | 588 | 28025 | 6.10e-13 | -0.9158 | Nephritis; nephrosis; renal sclerosis | genitourinary | 25DTot | |
| 580.1 | | | | | 17811 | 175 | 17636 | | | | | 28185 | 160 | 28025 | | | Glomerulonephritis | genitourinary | 25DTot | |
| 580.11 | | | | | 17690 | 54 | 17636 | | | | | 28073 | 48 | 28025 | | | Proliferative glomerulonephritis | genitourinary | 25DTot | |
| 580.12 | | | | | 17715 | 79 | 17636 | | | | | 28084 | 59 | 28025 | | | Non-proliferative glomerulonephritis | genitourinary | 25DTot | |
| 580.13 | | | | | 17654 | 18 | 17636 | | | | | 28048 | 23 | 28025 | | | Acute glomerulonephritis, NOS | genitourinary | 25DTot | |
| 580.14 | | | | | 17676 | 40 | 17636 | | | | | 28076 | 51 | 28025 | | | Chronic glomerulonephritis, NOS | genitourinary | 25DTot | |
| 580.2 | | | | | 17776 | 140 | 17636 | | | | | 28143 | 118 | 28025 | | | Nephrotic syndrome without mention of glomerulonephritis | genitourinary | 25DTot | |
| 580.3 | | | | | 17973 | 337 | 17636 | | | | | 28368 | 343 | 28025 | 1.63e-09 | -1.003 | Nephritis and nephropathy without mention of glomerulonephritis | genitourinary | 25DTot | |
| 580.31 | | | | | 17810 | 174 | 17636 | | | | | 28242 | 217 | 28025 | | | Nephritis and nephropathy in diseases classified elsewhere | genitourinary | 25DTot | |
| 580.32 | | | | | 17795 | 159 | 17636 | | | | | 28162 | 137 | 28025 | | | Nephritis and nephropathy with pathological lesion | genitourinary | 25DTot | |

| phenotype | male_beta | male_SE | male_OR | male_p | male_n_total | male_n_cases | male_n_controls | female_beta | female_SE | female_OR | female_p | female_n_total | female_n_cases | female_n_controls | interaction_p | interaction_beta | description | group | lab | label |
|---|---|---|---|---|---|---|---|---|---|---|---|---|---|---|---|---|---|---|---|---|
| 580.4 | | | | | 17734 | 98 | 17636 | | | | | 28125 | 100 | 28025 | | | Renal sclerosis, NOS | genitourinary | 25DTot | |
| 585 | 0.7457 | 0.04627 | 2.108 | 1.96e-58 | 24023 | 6387 | 17636 | 0.2518 | 0.03844 | 1.286 | 5.78e-11 | 32893 | 4868 | 28025 | 7.17e-17 | -0.4908 | Renal failure | genitourinary | 25DTot | |
| 585.1 | 0.7285 | 0.05094 | 2.072 | 2.11e-46 | 22356 | 4720 | 17636 | 0.2806 | 0.04362 | 1.324 | 1.25e-10 | 31559 | 3534 | 28025 | 2.73e-11 | -0.4358 | Acute renal failure | genitourinary | 25DTot | |
| 585.2 | 1.211 | 0.1091 | 3.358 | 1.21e-28 | 18192 | 556 | 17636 | | | | | 28438 | 413 | 28025 | 1.39e-06 | -0.7462 | Renal failure NOS | genitourinary | 25DTot | |
| 585.3 | 0.989 | 0.05276 | 2.689 | 2.15e-78 | 21314 | 3678 | 17636 | 0.3243 | 0.04918 | 1.383 | 4.27e-11 | 30569 | 2544 | 28025 | 5.92e-21 | -0.6617 | Chronic renal failure [CKD] | genitourinary | 25DTot | |
| 585.31 | 1.014 | 0.09818 | 2.758 | 5.03e-25 | 18326 | 690 | 17636 | | | | | 28496 | 471 | 28025 | 3.64e-06 | -0.6579 | Renal dialysis | genitourinary | 25DTot | |
| 585.32 | 1.216 | 0.08189 | 3.372 | 7.50e-50 | 18620 | 984 | 17636 | 0.3291 | 0.09164 | 1.39 | 3.29e-04 | 28699 | 674 | 28025 | 1.19e-12 | -0.8479 | End stage renal disease | genitourinary | 25DTot | |
| 585.33 | 1.289 | 0.06266 | 3.627 | 5.89e-94 | 19559 | 1923 | 17636 | 0.6197 | 0.06191 | 1.858 | 1.38e-23 | 29378 | 1353 | 28025 | 5.98e-15 | -0.6723 | Chronic Kidney Disease, Stage III | genitourinary | 25DTot | |
| 585.34 | 1.659 | 0.07792 | 5.252 | 1.55e-100 | 18575 | 939 | 17636 | 0.6123 | 0.08379 | 1.845 | 2.73e-13 | 28729 | 704 | 28025 | 2.11e-21 | -1.056 | Chronic Kidney Disease, Stage IV | genitourinary | 25DTot | Chronic Kidney Disease, Stage IV |
| 585.4 | 1.394 | 0.08312 | 4.031 | 4.04e-63 | 18446 | 810 | 17636 | 0.7222 | 0.08739 | 2.059 | 1.41e-16 | 28650 | 625 | 28025 | 5.38e-10 | -0.73 | Chronic kidney disease, Stage I or II | genitourinary | 25DTot | |
| 586 | 0.7545 | 0.05556 | 2.127 | 5.35e-42 | 21065 | 3429 | 17636 | 0.2214 | 0.05151 | 1.248 | 1.72e-05 | 30360 | 2335 | 28025 | 1.22e-14 | -0.5695 | Other disorders of the kidney and ureters | genitourinary | 25DTot | |
| 586.1 | | | | | 17687 | 51 | 17636 | | | | | 28103 | 78 | 28025 | | | Anatomical abnormatilies of kidney and ureters | genitourinary | 25DTot | |
| 586.11 | | | | | 17638 | 2 | 17636 | | | | | 28033 | 8 | 28025 | | | Small kidney | genitourinary | 25DTot | |
| 586.12 | | | | | 17684 | 48 | 17636 | | | | | 28097 | 72 | 28025 | | | Vesicoureteral reflux | genitourinary | 25DTot | |
| 586.2 | 0.7268 | 0.09413 | 2.068 | 1.15e-14 | 18502 | 866 | 17636 | 0.331 | 0.09743 | 1.392 | 6.82e-04 | 28545 | 520 | 28025 | 0.001005 | -0.435 | Cyst of kidney, acquired | genitourinary | 25DTot | |
| 586.3 | | | | | 17685 | 49 | 17636 | | | | | 28057 | 32 | 28025 | | | Vascular disorders of kidney/hypertrophy | genitourinary | 25DTot | |
| 586.4 | 0.6368 | 0.1064 | 1.89 | 2.15e-09 | 18326 | 690 | 17636 | | | | | 28313 | 288 | 28025 | 4.02e-12 | -1.31 | Stricture/obstruction of ureter | genitourinary | 25DTot | |
| 587 | 1.32 | 0.09884 | 3.745 | 1.05e-40 | 18201 | 565 | 17636 | | | | | 28416 | 391 | 28025 | 1.29e-07 | -0.7751 | Kidney replaced by transpant | genitourinary | 25DTot | |
| 588 | 1.277 | 0.08078 | 3.585 | 2.95e-56 | 18557 | 921 | 17636 | 0.6787 | 0.08304 | 1.971 | 3.00e-16 | 28718 | 693 | 28025 | 1.17e-07 | -0.5993 | Disorders resulting from impaired renal function | genitourinary | 25DTot | |
| 588.1 | 1.273 | 0.08876 | 3.572 | 1.19e-46 | 18381 | 745 | 17636 | 0.6387 | 0.09465 | 1.894 | 1.50e-11 | 28555 | 530 | 28025 | 8.49e-07 | -0.6233 | Renal osteodystrophy | genitourinary | 25DTot | |
| 588.2 | | | | | 17925 | 289 | 17636 | | | | | 28246 | 221 | 28025 | 0.002002 | -0.5861 | Secondary hyperparathyroidism (of renal origin) | genitourinary | 25DTot | |
| 589 | | | | | 17733 | 97 | 17636 | | | | | 28140 | 115 | 28025 | | | Abnormal results of function study of kidney | genitourinary | 25DTot | |
| 590 | | | | | 21698 | 323 | 21375 | 0.4044 | 0.08882 | 1.498 | 5.30e-06 | 24682 | 746 | 23936 | 0.0364 | -0.3438 | Pyelonephritis | genitourinary | 25DTot | |
| 591 | 0.6603 | 0.05933 | 1.935 | 8.88e-29 | 23909 | 2534 | 21375 | 0.5186 | 0.03433 | 1.68 | 1.45e-51 | 30676 | 6740 | 23936 | 0.3553 | -0.06219 | Urinary tract infection | genitourinary | 25DTot | |
| 592 | | | | | 21768 | 393 | 21375 | 0.6153 | 0.06024 | 1.85 | 1.73e-24 | 25433 | 1497 | 23936 | 0.05758 | 0.2852 | Cystitis and urethritis | genitourinary | 25DTot | |
| 592.1 | | | | | 21707 | 332 | 21375 | 0.6353 | 0.06107 | 1.888 | 2.41e-25 | 25375 | 1439 | 23936 | 0.03709 | 0.3395 | Cystitis | genitourinary | 25DTot | |
| 592.11 | | | | | 21543 | 168 | 21375 | 0.7132 | 0.07439 | 2.041 | 9.05e-22 | 24834 | 898 | 23936 | 0.03493 | 0.47 | Acute cystitis | genitourinary | 25DTot | |
| 592.12 | | | | | 21411 | 36 | 21375 | | | | | 23989 | 53 | 23936 | | | Chronic cystitis | genitourinary | 25DTot | |
| 592.13 | | | | | 21383 | 8 | 21375 | | | | | 24169 | 233 | 23936 | | | Chronic interstitial cystitis | genitourinary | 25DTot | |

| phenotype | male_beta | male_SE | male_OR | male_p | male_n_total | male_n_cases | male_n_controls | female_beta | female_SE | female_OR | female_p | female_n_total | female_n_cases | female_n_controls | interaction_p | interaction_beta | description | group | lab | label |
|---|---|---|---|---|---|---|---|---|---|---|---|---|---|---|---|---|---|---|---|---|
| 592.2 | | | | | 21408 | 33 | 21375 | | | | | 23967 | 31 | 23936 | | | Urethritis and urethral syndrome | genitourinary | 25DTot | |
| 592.21 | | | | | 21378 | 3 | 21375 | | | | | 23938 | 2 | 23936 | | | Urethral syndrome | genitourinary | 25DTot | |
| 592.3 | | | | | 21378 | 3 | 21375 | | | | | 23938 | 2 | 23936 | | | Urethral stricture due to infecton | genitourinary | 25DTot | |
| 593 | 0.4448 | 0.07004 | 1.56 | 2.13e-10 | 23190 | 1815 | 21375 | 0.4908 | 0.05653 | 1.634 | 3.88e-18 | 25739 | 1803 | 23936 | 0.2574 | 0.1002 | Hematuria | genitourinary | 25DTot | |
| 593.1 | 0.3717 | 0.1118 | 1.45 | 8.89e-04 | 22020 | 645 | 21375 | | | | | 24248 | 312 | 23936 | 0.8061 | 0.04069 | Gross hematuria | genitourinary | 25DTot | |
| 593.2 | | | | | 21568 | 193 | 21375 | | | | | 24188 | 252 | 23936 | | | Microscopic hematuria | genitourinary | 25DTot | |
| 594 | 0.4773 | 0.06592 | 1.612 | 4.45e-13 | 25965 | 1823 | 24142 | 0.2678 | 0.05927 | 1.307 | 6.22e-06 | 35229 | 1634 | 33595 | 0.002152 | -0.2654 | Urinary calculus | genitourinary | 25DTot | |
| 594.1 | 0.5332 | 0.07154 | 1.704 | 9.09e-14 | 25599 | 1457 | 24142 | 0.2938 | 0.06467 | 1.342 | 5.53e-06 | 34942 | 1347 | 33595 | 0.001727 | -0.2948 | Calculus of kidney | genitourinary | 25DTot | |
| 594.2 | | | | | 24276 | 134 | 24142 | | | | | 33649 | 54 | 33595 | | | Calculus of lower urinary tract | genitourinary | 25DTot | |
| 594.3 | | | | | 24598 | 456 | 24142 | | | | | 33995 | 400 | 33595 | 0.7413 | -0.05279 | Calculus of ureter | genitourinary | 25DTot | |
| 594.8 | | | | | 24179 | 37 | 24142 | | | | | 33649 | 54 | 33595 | | | Renal colic | genitourinary | 25DTot | |
| 595 | 0.26 | 0.1084 | 1.297 | 0.01648 | 24902 | 760 | 24142 | 0.1539 | 0.09091 | 1.166 | 0.09039 | 34327 | 732 | 33595 | 0.3095 | -0.1402 | Hydronephrosis | genitourinary | 25DTot | |
| 596 | 0.4066 | 0.08342 | 1.502 | 1.09e-06 | 25451 | 1131 | 24320 | 0.2347 | 0.06869 | 1.264 | 6.35e-04 | 34046 | 1171 | 32875 | 0.07368 | -0.189 | Other disorders of bladder | genitourinary | 25DTot | |
| 596.1 | | | | | 24500 | 180 | 24320 | | | | | 32903 | 28 | 32875 | | | Bladder neck obstruction | genitourinary | 25DTot | |
| 596.5 | 0.5662 | 0.107 | 1.762 | 1.20e-07 | 24918 | 598 | 24320 | 0.2619 | 0.07663 | 1.299 | 6.32e-04 | 33798 | 923 | 32875 | 0.03121 | -0.2778 | Functional disorders of bladder | genitourinary | 25DTot | |
| 597 | | | | | 24709 | 389 | 24320 | | | | | 33264 | 389 | 32875 | 0.2413 | 0.2064 | Other disorders of urethra and urinary tract | genitourinary | 25DTot | |
| 597.1 | | | | | 24499 | 179 | 24320 | | | | | 32898 | 23 | 32875 | | | Urethral stricture (not specified as infectious) | genitourinary | 25DTot | |
| 597.2 | | | | | 24431 | 111 | 24320 | | | | | 32951 | 76 | 32875 | | | Urinary complications NEC | genitourinary | 25DTot | |
| 597.8 | | | | | 24446 | 126 | 24320 | | | | | 32996 | 121 | 32875 | | | Urethral hypermobility/ISD | genitourinary | 25DTot | |
| 598 | | | | | 27202 | 318 | 26884 | | | | | 36132 | 426 | 35706 | 0.2294 | -0.2056 | Abnormal findings on examination of urine | genitourinary | 25DTot | |
| 598.4 | | | | | 26884 | | | | | | | 35709 | 3 | 35706 | | | Other cells and casts in urine | genitourinary | 25DTot | |
| 599 | 0.7052 | 0.05238 | 2.024 | 2.57e-41 | 23589 | 4209 | 19380 | 0.5827 | 0.03397 | 1.791 | 6.18e-66 | 31705 | 6851 | 24854 | 0.006434 | -0.1664 | Other symptoms/disorders or the urinary system | genitourinary | 25DTot | |
| 599.1 | | | | | 19615 | 235 | 19380 | | | | | 24927 | 73 | 24854 | | | Urinary obstruction | genitourinary | 25DTot | |
| 599.2 | 0.7075 | 0.07437 | 2.029 | 1.85e-21 | 21042 | 1662 | 19380 | 0.5656 | 0.06939 | 1.76 | 3.63e-16 | 25979 | 1125 | 24854 | 0.1545 | -0.1419 | Retention of urine | genitourinary | 25DTot | |
| 599.3 | 0.8791 | 0.09552 | 2.409 | 3.48e-20 | 20130 | 750 | 19380 | 0.7288 | 0.04427 | 2.073 | 6.95e-61 | 27990 | 3136 | 24854 | 0.07644 | -0.1837 | Dysuria | genitourinary | 25DTot | |
| 599.4 | 0.5114 | 0.1078 | 1.668 | 2.09e-06 | 20122 | 742 | 19380 | 0.5395 | 0.05219 | 1.715 | 4.84e-25 | 27034 | 2180 | 24854 | 0.7072 | 0.04451 | Urinary incontinence | genitourinary | 25DTot | |
| 599.5 | 0.9234 | 0.06841 | 2.518 | 1.61e-41 | 21290 | 1910 | 19380 | 0.7805 | 0.05199 | 2.183 | 6.06e-51 | 26913 | 2059 | 24854 | 0.01271 | -0.2098 | Frequency of urination and polyuria | genitourinary | 25DTot | |
| 599.6 | | | | | 19476 | 96 | 19380 | | | | | 24972 | 118 | 24854 | | | Oliguria and anuria | genitourinary | 25DTot | |
| 599.7 | | | | | 19390 | 10 | 19380 | | | | | 24855 | 1 | 24854 | | | Urethral discharge | genitourinary | 25DTot | |
| 599.8 | | | | | 19615 | 235 | 19380 | | | | | 25086 | 232 | 24854 | | | Other symptoms involving urinary system | genitourinary | 25DTot | |
| 599.9 | 1.17 | 0.08991 | 3.221 | 1.08e-38 | 20173 | 793 | 19380 | 0.728 | 0.0647 | 2.071 | 2.26e-29 | 26091 | 1237 | 24854 | 2.05e-06 | -0.5155 | Other abnormality of urination | genitourinary | 25DTot | |
| 600 | 0.707 | 0.05981 | 2.028 | 3.07e-32 | 23028 | 3640 | 19388 | | | | | 4 | | | | | Hyperplasia of prostate | genitourinary | 25DTot | |
| 601 | 0.7008 | 0.1009 | 2.015 | 3.73e-12 | 20083 | 695 | 19388 | | | | | 4 | | | | | Inflammatory diseases of prostate | genitourinary | 25DTot | |
| 601.1 | | | | | 19781 | 393 | 19388 | | | | | 4 | | | | | Prostatitis | genitourinary | 25DTot | |
| 601.11 | | | | | 19525 | 137 | 19388 | | | | | 4 | | | | | Acute prostatitis | genitourinary | 25DTot | |

| phenotype | male_beta | male_SE | male_OR | male_p | male_n_total | male_n_cases | male_n_controls | female_beta | female_SE | female_OR | female_p | female_n_total | female_n_cases | female_n_controls | interaction_p | interaction_beta | description | group | lab | label |
|---|---|---|---|---|---|---|---|---|---|---|---|---|---|---|---|---|---|---|---|---|
| 601.12 | | | | | 19502 | 114 | 19388 | | | | | 4 | | | | | Chronic prostatitis | genitourinary | 25DTot | |
| 601.3 | | | | | 19577 | 189 | 19388 | | | | | 4 | | | | | Orchitis and epididymitis | genitourinary | 25DTot | |
| 601.4 | | | | | 19435 | 47 | 19388 | | | | | 4 | | | | | Balanoposthitis | genitourinary | 25DTot | |
| 601.8 | | | | | 19460 | 72 | 19388 | | | | | 4 | | | | | Other inflammatory disorders of male genital organs | genitourinary | 25DTot | |
| 602 | | | | | 19490 | 102 | 19388 | | | | | 4 | | | | | Other disorders of prostate | genitourinary | 25DTot | |
| 602.3 | | | | | 19390 | 2 | 19388 | | | | | 4 | | | | | Dysplasia of prostate | genitourinary | 25DTot | |
| 603 | | | | | 19102 | 243 | 18859 | | | | | 4 | | | | | Other disorders of testis | genitourinary | 25DTot | |
| 603.1 | | | | | 19051 | 192 | 18859 | | | | | 4 | | | | | Hydrocele | genitourinary | 25DTot | |
| 603.2 | | | | | 18915 | 56 | 18859 | | | | | 4 | | | | | Spermatocele | genitourinary | 25DTot | |
| 604 | | | | | 19178 | 319 | 18859 | | | | | 4 | | | | | Disorders of penis | genitourinary | 25DTot | |
| 604.1 | | | | | 18955 | 96 | 18859 | | | | | 4 | | | | | Redundant prepuce and phimosis/BXO | genitourinary | 25DTot | |
| 604.2 | | | | | 18859 | | | | | | | 4 | | | | | Vascular disorders of penis | genitourinary | 25DTot | |
| 604.3 | | | | | 18985 | 126 | 18859 | | | | | 4 | | | | | Peyronie's disease | genitourinary | 25DTot | |
| 605 | 0.7006 | 0.06695 | 2.015 | 1.27e-25 | 21005 | 2146 | 18859 | | | | | 4 | | | | | Erectile dysfunction [ED] | genitourinary | 25DTot | |
| 608 | 0.5612 | 0.1215 | 1.753 | 3.86e-06 | 19398 | 539 | 18859 | | | | | 4 | | | | | Other disorders of male genital organs | genitourinary | 25DTot | |
| 609 | | | | | 18929 | 70 | 18859 | | | | | 4 | | | | | Male infertility and abnormal spermatozoa | genitourinary | 25DTot | |
| 609.1 | | | | | 18884 | 25 | 18859 | | | | | 4 | | | | | Infertility, male | genitourinary | 25DTot | |
| 609.11 | | | | | 18861 | 2 | 18859 | | | | | 4 | | | | | Azoospermia and oligospermia | genitourinary | 25DTot | |
| 609.2 | | | | | 18902 | 43 | 18859 | | | | | 4 | | | | | Abnormal spermatozoa | genitourinary | 25DTot | |
| 610 | | | | | 5 | | | 0.5885 | 0.05793 | 1.801 | 3.01e-24 | 29977 | 1611 | 28366 | 0.9998 | 0.08459 | Benign mammary dysplasias | genitourinary | 25DTot | |
| 610.1 | | | | | 5 | | | 0.6918 | 0.06447 | 1.997 | 7.31e-27 | 29598 | 1232 | 28366 | 0.9997 | 0.1494 | Cystic mastopathy | genitourinary | 25DTot | |
| 610.2 | | | | | 5 | | | | | | | 28425 | 59 | 28366 | | | Fibroadenosis of breast | genitourinary | 25DTot | |
| 610.3 | | | | | 5 | | | | | | | 28501 | 135 | 28366 | | | Fibrosclerosis of breast | genitourinary | 25DTot | |
| 610.4 | | | | | 5 | | | | | | | 28633 | 267 | 28366 | | | Benign neoplasm of breast | genitourinary | 25DTot | |
| 610.8 | | | | | 5 | | | | | | | 28599 | 233 | 28366 | | | Other specified benign mammary dysplasias | genitourinary | 25DTot | |
| 611 | | | | | 27275 | 62 | 27213 | 0.541 | 0.03665 | 1.718 | 2.62e-49 | 33455 | 5088 | 28367 | 0.6269 | -0.1427 | Abnormal findings on mammogram or breast exam | genitourinary | 25DTot | |
| 611.1 | | | | | 27225 | 12 | 27213 | 0.5463 | 0.03942 | 1.727 | 1.11e-43 | 32548 | 4181 | 28367 | 0.5664 | 0.445 | Abnormal mammogram | genitourinary | 25DTot | |
| 611.11 | | | | | 27213 | | | 0.5879 | 0.0923 | 1.8 | 1.89e-10 | 28926 | 559 | 28367 | 0.999 | 1.19 | Mammographic microcalcification | genitourinary | 25DTot | |
| 611.3 | | | | | 27266 | 53 | 27213 | 0.61 | 0.0562 | 1.84 | 1.91e-27 | 30081 | 1714 | 28367 | 0.296 | -0.3191 | Lump or mass in breast | genitourinary | 25DTot | |
| 612 | | | | | 27326 | 113 | 27213 | | | | | 28757 | 390 | 28367 | 0.03887 | -0.4833 | Breast conditions, congenital or relating to hormones | genitourinary | 25DTot | |
| 612.1 | | | | | 27213 | | | | | | | 28446 | 79 | 28367 | | | Galactorrhea | genitourinary | 25DTot | |
| 612.2 | | | | | 27325 | 112 | 27213 | | | | | 28664 | 297 | 28367 | | | Hypertrophy of breast (Gynecomastia) | genitourinary | 25DTot | |
| 612.3 | | | | | 27214 | 1 | 27213 | | | | | 28379 | 12 | 28367 | | | Congenital anomalies of breast | genitourinary | 25DTot | |
| 613 | | | | | 27667 | 60 | 27607 | 0.4476 | 0.04781 | 1.565 | 7.89e-21 | 34784 | 2435 | 32349 | 0.727 | -0.1082 | Other nonmalignant breast conditions | genitourinary | 25DTot | |
| 613.1 | | | | | 27607 | | | | | | | 32616 | 267 | 32349 | | | Inflammatory disease of breast | genitourinary | 25DTot | |
| 613.5 | | | | | 27649 | 42 | 27607 | 0.5745 | 0.07111 | 1.776 | 6.54e-16 | 33336 | 987 | 32349 | 0.4463 | -0.2661 | Mastodynia | genitourinary | 25DTot | |
| 613.7 | | | | | 27616 | 9 | 27607 | | | | | 32785 | 436 | 32349 | | | Other signs and symptoms in breast | genitourinary | 25DTot | |
| 613.8 | | | | | 27608 | 1 | 27607 | 0.3164 | 0.08552 | 1.372 | 2.16e-04 | 33034 | 685 | 32349 | 0.9581 | 10.2 | Other specified disorders of breast | genitourinary | 25DTot | |

| phenotype | male_beta | male_SE | male_OR | male_p | male_n_total | male_n_cases | male_n_controls | female_beta | female_SE | female_OR | female_p | female_n_total | female_n_cases | female_n_controls | interaction_p | interaction_beta | description | group | lab | label |
|---|---|---|---|---|---|---|---|---|---|---|---|---|---|---|---|---|---|---|---|---|
| 613.9 | | | | | 27609 | 2 | 27607 | | | | | 32422 | 73 | 32349 | | | Breast disorder NOS | genitourinary | 25DTot | |
| 614 | | | | | 7 | | | 0.3261 | 0.06273 | 1.386 | 2.01e-07 | 33657 | 1625 | 32032 | 1 | -0.01432 | Inflammatory diseases of female pelvic organs | genitourinary | 25DTot | |
| 614.1 | | | | | 7 | | | | | | | 32176 | 144 | 32032 | | | Pelvic peritoneal adhesions, female (postoperative) (postinfection) | genitourinary | 25DTot | |
| 614.3 | | | | | 7 | | | | | | | 32232 | 200 | 32032 | | | Pelvic inflammatory disease (PID) | genitourinary | 25DTot | |
| 614.31 | | | | | 7 | | | | | | | 32055 | 23 | 32032 | | | Acute inflammatory pelvic disease | genitourinary | 25DTot | |
| 614.32 | | | | | 7 | | | | | | | 32140 | 108 | 32032 | | | Chronic inflammatory pelvic disease | genitourinary | 25DTot | |
| 614.33 | | | | | 7 | | | | | | | 32100 | 68 | 32032 | | | Pelvic inflammatory disease, NOS | genitourinary | 25DTot | |
| 614.4 | | | | | 7 | | | | | | | 32069 | 37 | 32032 | | | Inflammatory diseases of uterus, except cervix | genitourinary | 25DTot | |
| 614.5 | | | | | 7 | | | 0.3937 | 0.07099 | 1.482 | 2.92e-08 | 33209 | 1177 | 32032 | 0.9999 | 0.02209 | Inflammatory disease of cervix, vagina, and vulva | genitourinary | 25DTot | |
| 614.51 | | | | | 7 | | | | | | | 32101 | 69 | 32032 | | | Cervicitis and endocervicitis | genitourinary | 25DTot | |
| 614.52 | | | | | 7 | | | 0.3999 | 0.08315 | 1.492 | 1.52e-06 | 32877 | 845 | 32032 | 1 | 0.02115 | Vaginitis and vulvovaginitis | genitourinary | 25DTot | |
| 614.53 | | | | | 7 | | | | | | | 32093 | 61 | 32032 | | | Cyst or abscess of Bartholin's gland | genitourinary | 25DTot | |
| 614.54 | | | | | 7 | | | | | | | 32097 | 65 | 32032 | | | Abscess or ulceration of vulva | genitourinary | 25DTot | |
| 615 | | | | | 7 | | | | | | | 32416 | 384 | 32032 | | | Endometriosis | genitourinary | 25DTot | |
| 618 | | | | | 7 | | | 0.06566 | 0.06915 | 1.068 | 0.3424 | 36315 | 1140 | 35175 | 0.9993 | -0.3235 | Genital prolapse | genitourinary | 25DTot | |
| 618.1 | | | | | 7 | | | 0.07352 | 0.07716 | 1.076 | 0.3406 | 36071 | 896 | 35175 | 0.9996 | -0.3176 | Prolapse of vaginal walls | genitourinary | 25DTot | |
| 618.2 | | | | | 7 | | | | | | | 35515 | 340 | 35175 | | | Uterine/Uterovaginal prolapse | genitourinary | 25DTot | |
| 618.5 | | | | | 7 | | | | | | | 35494 | 319 | 35175 | | | Prolapse of vaginal vault after hysterectomy | genitourinary | 25DTot | |
| 618.6 | | | | | 7 | | | | | | | 35376 | 201 | 35175 | | | Vaginal enterocele, congenital or acquired | genitourinary | 25DTot | |
| 619 | | | | | 7 | | | 0.2772 | 0.0551 | 1.319 | 4.86e-07 | 33857 | 2126 | 31731 | 0.9999 | -0.05799 | Noninflammatory female genital disorders | genitourinary | 25DTot | |
| 619.1 | | | | | 7 | | | | | | | 32002 | 271 | 31731 | | | Noninflammatory disorders of ovary, fallopian tube, and broad ligament | genitourinary | 25DTot | |
| 619.2 | | | | | 7 | | | | | | | 32192 | 461 | 31731 | | | Disorders of uterus, NEC | genitourinary | 25DTot | |
| 619.3 | | | | | 7 | | | | | | | 31864 | 133 | 31731 | | | Noninflammatory disorders of cervix | genitourinary | 25DTot | |
| 619.4 | | | | | 7 | | | 0.1313 | 0.1092 | 1.14 | 0.2293 | 32259 | 528 | 31731 | 0.9997 | -0.209 | Noninflammatory disorders of vagina | genitourinary | 25DTot | |
| 619.5 | | | | | 7 | | | | | | | 31969 | 238 | 31731 | | | Noninflammatory disorders of vulva and perineum | genitourinary | 25DTot | |
| 620 | | | | | 7 | | | | | | | 35251 | 113 | 35138 | | | Dysplasia of female genital organs | genitourinary | 25DTot | |
| 620.1 | | | | | 7 | | | | | | | 35152 | 14 | 35138 | | | Dysplasia of cervix | genitourinary | 25DTot | |
| 621 | | | | | 7 | | | | | | | 35309 | 171 | 35138 | | | Endometrial hyperplasia | genitourinary | 25DTot | |
| 622 | | | | | 7 | | | | | | | 35534 | 396 | 35138 | | | Polyp of female genital organs | genitourinary | 25DTot | |
| 622.1 | | | | | 7 | | | | | | | 35431 | 293 | 35138 | | | Polyp of corpus uteri | genitourinary | 25DTot | |
| 622.2 | | | | | 7 | | | | | | | 35215 | 77 | 35138 | | | Mucous polyp of cervix | genitourinary | 25DTot | |
| 623 | | | | | 7 | | | | | | | 35299 | 161 | 35138 | | | Hypertrophy of female genital organs | genitourinary | 25DTot | |
| 624 | | | | | 7 | | | | | | | 31709 | 227 | 31482 | | | Symptoms involving female genital tract | genitourinary | 25DTot | |
| 624.1 | | | | | 7 | | | | | | | 31539 | 57 | 31482 | | | Dystrophy of female genital tract | genitourinary | 25DTot | |
| 624.2 | | | | | 7 | | | | | | | 31503 | 21 | 31482 | | | Atrophy of female genital tract | genitourinary | 25DTot | |
| 624.9 | | | | | 7 | | | 0.38 | 0.07142 | 1.462 | 1.04e-07 | 32466 | 984 | 31482 | 1 | 0.00532 | stress incontinence, female | genitourinary | 25DTot | |

| phenotype | male_beta | male_SE | male_OR | male_p | male_n_total | male_n_cases | male_n_controls | female_beta | female_SE | female_OR | female_p | female_n_total | female_n_cases | female_n_controls | interaction_p | interaction_beta | description | group | lab | label |
|---|---|---|---|---|---|---|---|---|---|---|---|---|---|---|---|---|---|---|---|---|
| 625 | | | | | 7 | | | 0.395 | 0.06559 | 1.484 | 1.71e-09 | 32899 | 1417 | 31482 | 0.9998 | 0.08347 | Pain and other symptoms associated with female genital organs | genitourinary | 25DTot | |
| 625.1 | | | | | 7 | | | | | | | 31749 | 267 | 31482 | | | Dyspareunia | genitourinary | 25DTot | |
| 626 | | | | | 5 | | | 0.4926 | 0.04921 | 1.637 | 1.39e-23 | 27458 | 3874 | 23584 | 0.9998 | -0.06169 | Disorders of menstruation and other abnormal bleeding from female genital tract | genitourinary | 25DTot | |
| 626.1 | | | | | 5 | | | 0.5369 | 0.05314 | 1.711 | 5.31e-24 | 26685 | 3101 | 23584 | 0.9999 | -0.02501 | Irregular menstrual cycle/bleeding | genitourinary | 25DTot | |
| 626.11 | | | | | 5 | | | 0.6384 | 0.1133 | 1.893 | 1.78e-08 | 24169 | 585 | 23584 | 0.9999 | 0.1094 | Absent or infrequent menstruation | genitourinary | 25DTot | |
| 626.12 | | | | | 5 | | | 0.5641 | 0.0776 | 1.758 | 3.59e-13 | 24734 | 1150 | 23584 | 0.9999 | 0.02961 | Excessive or frequent menstruation | genitourinary | 25DTot | |
| 626.13 | | | | | 5 | | | 0.7242 | 0.07953 | 2.063 | 8.55e-20 | 24711 | 1127 | 23584 | 0.9997 | 0.1171 | Irregular menstrual cycle | genitourinary | 25DTot | |
| 626.14 | | | | | 5 | | | | | | | 23850 | 266 | 23584 | | | Irregular menstrual bleeding | genitourinary | 25DTot | |
| 626.15 | | | | | 5 | | | | | | | 23629 | 45 | 23584 | | | Infertility, female, associated with anovulation | genitourinary | 25DTot | |
| 626.2 | | | | | 5 | | | | | | | 24029 | 445 | 23584 | | | Dysmenorrhea | genitourinary | 25DTot | |
| 626.21 | | | | | 5 | | | | | | | 23591 | 7 | 23584 | | | Mittelschmerz | genitourinary | 25DTot | |
| 626.4 | | | | | 5 | | | | | | | 23636 | 52 | 23584 | | | Premenstrual tension syndromes | genitourinary | 25DTot | |
| 626.8 | | | | | 5 | | | | | | | 23771 | 187 | 23584 | | | Infertility, female | genitourinary | 25DTot | |
| 627 | | | | | 6 | | | 0.8167 | 0.03765 | 2.263 | 2.49e-104 | 28947 | 5363 | 23584 | 0.9367 | 9.487 | Menopausal and postmenopausal disorders | genitourinary | 25DTot | |
| 627.1 | | | | | 5 | | | 0.7013 | 0.07997 | 2.016 | 1.80e-18 | 24382 | 798 | 23584 | 0.9998 | 0.1381 | Postmenopausal bleeding | genitourinary | 25DTot | |
| 627.2 | | | | | 6 | | | 0.8877 | 0.04388 | 2.429 | 5.23e-91 | 26962 | 3378 | 23584 | 0.9365 | 9.519 | Symptomatic menopause | genitourinary | 25DTot | |
| 627.21 | | | | | 5 | | | | | | | 23934 | 350 | 23584 | | | Symptomatic artificial menopause | genitourinary | 25DTot | |
| 627.22 | | | | | 6 | | | 0.7855 | 0.07546 | 2.193 | 2.26e-25 | 24465 | 881 | 23584 | 0.9578 | 10.41 | Need for Hormone replacement therapy (postmenopausal) | genitourinary | 25DTot | |
| 627.3 | | | | | 5 | | | 0.8558 | 0.06273 | 2.353 | 2.25e-42 | 24943 | 1359 | 23584 | 0.9994 | 0.265 | Postmenopausal atrophic vaginitis | genitourinary | 25DTot | |
| 627.4 | | | | | 5 | | | | | | | 23687 | 103 | 23584 | | | Premenopausal menorrhagia | genitourinary | 25DTot | |
| 627.5 | | | | | 5 | | | | | | | 23742 | 158 | 23584 | | | Premature menopause and other ovarian failure | genitourinary | 25DTot | |
| 628 | | | | | 5 | | | 0.7032 | 0.07872 | 2.02 | 4.13e-19 | 24586 | 1002 | 23584 | 0.9994 | 0.2943 | Ovarian cyst | genitourinary | 25DTot | |
| 634 | | | | | 7 | | | -0.2933 | 0.1483 | 0.7458 | 0.04793 | 34235 | 519 | 33716 | 0.9992 | -0.6063 | Miscarriage; stillbirth | pregnancy complications | 25DTot | |
| 634.1 | | | | | 7 | | | | | | | 33945 | 229 | 33716 | | | Missed abortion/Hydatidiform mole | pregnancy complications | 25DTot | |
| 634.3 | | | | | 7 | | | | | | | 33757 | 41 | 33716 | | | Ectopic pregnancy | pregnancy complications | 25DTot | |
| 635 | | | | | 7 | | | | | | | 34113 | 397 | 33716 | | | Hemorrhage during pregnancy; childbirth and postpartum | pregnancy complications | 25DTot | |
| 635.2 | | | | | 7 | | | | | | | 33989 | 273 | 33716 | | | Antepartum hemorrhage, abruptio placentae, and placenta previa | pregnancy complications | 25DTot | |
| phenotype | male_beta | male_SE | male_OR | male_p | male_n_total | male_n_cases | male_n_controls | female_beta | female_SE | female_OR | female_p | female_n_total | female_n_cases | female_n_controls | interaction_p | interaction_beta | description | group | lab | label |
| 635.3 | | | | | 7 | | | | | | | 33851 | 135 | 33716 | | | Placenta previa and abruptio placenta | pregnancy complications | 25DTot | |
| 636 | | | | | 7 | | | -0.5351 | 0.09595 | 0.5856 | 2.45e-08 | 35417 | 1701 | 33716 | 0.998 | -0.886 | Early or threatened labor; hemorrhage in early pregnancy | pregnancy complications | 25DTot | |
| 636.1 | | | | | 7 | | | | | | | 34209 | 493 | 33716 | | | Threatened premature labor | pregnancy complications | 25DTot | |
| 636.2 | | | | | 7 | | | | | | | 34111 | 395 | 33716 | | | Early onset of delivery | pregnancy complications | 25DTot | |

| phenotype | male_beta | male_SE | male_OR | male_p | male_n_total | male_n_cases | male_n_controls | female_beta | female_SE | female_OR | female_p | female_n_total | female_n_cases | female_n_controls | interaction_p | interaction_beta | description | group | lab | label |
|---|---|---|---|---|---|---|---|---|---|---|---|---|---|---|---|---|---|---|---|---|
| 636.3 | | | | | 7 | | | -0.2734 | 0.1476 | 0.7608 | 0.06403 | 34236 | 520 | 33716 | 0.9991 | -0.6518 | Hemorrhage in early pregnancy | pregnancy complications | 25DTot | |
| 636.8 | | | | | 7 | | | | | | | 33814 | 98 | 33716 | | | Cervical incompetence | pregnancy complications | 25DTot | |
| 637 | | | | | 27759 | 8 | 27751 | | | | | 33709 | 19 | 33690 | | | Short gestation; low birth weight; and fetal growth retardation | pregnancy complications | 25DTot | |
| 642 | | | | | 7 | | | -0.478 | 0.139 | 0.62 | 5.86e-04 | 36632 | 759 | 35873 | 0.9989 | -0.7877 | Hypertension complicating pregnancy, childbirth, and the puerperium | pregnancy complications | 25DTot | |
| 642.1 | | | | | 7 | | | | | | | 36229 | 356 | 35873 | | | Preeclampsia and eclampsia | pregnancy complications | 25DTot | |
| 643 | | | | | 7 | | | | | | | 36714 | 156 | 36558 | | | Excessive vomiting in pregnancy | pregnancy complications | 25DTot | |
| 643.1 | | | | | 7 | | | | | | | 36608 | 50 | 36558 | | | Hyperemesis gravidarum | pregnancy complications | 25DTot | |
| 644 | | | | | 7 | | | | | | | 36546 | 186 | 36360 | | | Anemia during pregnancy | pregnancy complications | 25DTot | |
| 646 | | | | | 7 | | | -0.4063 | 0.1624 | 0.6661 | 0.01233 | 36367 | 578 | 35789 | 0.9988 | -0.8403 | Other complications of pregnancy NEC | pregnancy complications | 25DTot | |
| 647 | | | | | 7 | | | | | | | 36080 | 179 | 35901 | | | Infectious and parasitic complications affecting pregnancy | pregnancy complications | 25DTot | |
| 647.1 | | | | | 7 | | | | | | | 36102 | 201 | 35901 | | | Infections of genitourinary tract during pregnancy | pregnancy complications | 25DTot | |
| 647.3 | | | | | 7 | | | | | | | 35950 | 49 | 35901 | | | Major puerperal infection | pregnancy complications | 25DTot | |
| 649 | | | | | 7 | | | -0.3087 | 0.09078 | 0.7344 | 6.74e-04 | 35652 | 1664 | 33988 | 0.9985 | -0.6481 | Other conditions or status of the mother complicating pregnancy, childbirth, or the puerperium | pregnancy complications | 25DTot | |
| 649.1 | | | | | 7 | | | -0.4432 | 0.1475 | 0.642 | 0.002653 | 34593 | 605 | 33988 | 0.9989 | -0.7656 | Diabetes or abnormal glucose tolerance complicating pregnancy | pregnancy complications | 25DTot | |
| 653 | | | | | 7 | | | -0.6913 | 0.1743 | 0.5009 | 7.28e-05 | 35753 | 642 | 35111 | 0.9987 | -0.9345 | Problems associated with amniotic cavity and membranes | pregnancy complications | 25DTot | |
| 654 | | | | | 7 | | | | | | | 35408 | 297 | 35111 | | | Other and unspecified complications of birth; puerperium affecting management of mother | pregnancy complications | 25DTot | |
| 654.1 | | | | | 7 | | | | | | | 35312 | 201 | 35111 | | | Abnormality of organs and soft tissues of pelvis complicating pregnancy, childbirth, or the puerperium | pregnancy complications | 25DTot | |
| 654.2 | | | | | 7 | | | | | | | 35201 | 90 | 35111 | | | Rhesus isoimmunization in pregnancy | pregnancy complications | 25DTot | |
| 655 | | | | | 7 | | | -0.5451 | 0.08545 | 0.5798 | 1.78e-10 | 36061 | 2235 | 33826 | 0.998 | -0.8655 | Known or suspected fetal abnormality affecting management of mother | pregnancy complications | 25DTot | |
| 655.1 | | | | | 7 | | | -0.4885 | 0.1398 | 0.6135 | 4.75e-04 | 34615 | 789 | 33826 | 0.999 | -0.7554 | Abnormality in fetal heart rate or rhythm | pregnancy complications | 25DTot | |
| 656 | | | | | 27627 | 68 | 27559 | | | | | 36677 | 120 | 36557 | | | Other perinatal conditions of fetus or newborn | pregnancy complications | 25DTot | |
| 656.1 | | | | | 27562 | 3 | 27559 | | | | | 36558 | 1 | 36557 | | | Isoimmunization of fetus or newborn | pregnancy complications | 25DTot | |
| 656.2 | | | | | 27597 | 38 | 27559 | | | | | 36585 | 28 | 36557 | | | Respiratory conditions of fetus and newborn | pregnancy complications | 25DTot | |
| 656.22 | | | | | 27562 | 3 | 27559 | | | | | 36559 | 2 | 36557 | | | Interstitial emphysema and related conditions of newborn | pregnancy complications | 25DTot | |
| 656.26 | | | | | 27567 | 8 | 27559 | | | | | 36561 | 4 | 36557 | | | Transitory tachypnea or apnea of newborn | pregnancy complications | 25DTot | |
| 656.3 | | | | | 27567 | 8 | 27559 | | | | | 36567 | 10 | 36557 | | | Endocrine and metabolic disturbances of fetus and newborn | pregnancy complications | 25DTot | |

| phenotype | male_beta | male_SE | male_OR | male_p | male_n_total | male_n_cases | male_n_controls | female_beta | female_SE | female_OR | female_p | female_n_total | female_n_cases | female_n_controls | interaction_p | interaction_beta | description | group | lab | label |
|---|---|---|---|---|---|---|---|---|---|---|---|---|---|---|---|---|---|---|---|---|
| 656.4 | | | | | 27568 | 9 | 27559 | | | | | 36563 | 6 | 36557 | | | Hemorrhage of fetus or newborn | pregnancy complications | 25DTot | |
| 656.5 | | | | | 27565 | 6 | 27559 | | | | | 36562 | 5 | 36557 | | | Hematological disorders of newborn | pregnancy complications | 25DTot | |
| 656.6 | | | | | 27567 | 8 | 27559 | | | | | 36562 | 5 | 36557 | | | Perinatal disorders of digestive system | pregnancy complications | 25DTot | |
| 656.7 | | | | | 27566 | 7 | 27559 | | | | | 36566 | 9 | 36557 | | | Conditions involving the integument and temperature regulation of fetus and newborn | pregnancy complications | 25DTot | |
| 656.8 | | | | | 27575 | 16 | 27559 | | | | | 36572 | 15 | 36557 | | | Perinatal jaundice | pregnancy complications | 25DTot | |
| 656.9 | | | | | 27559 | | | | | | | 36559 | 2 | 36557 | | | Neonatal bradycardia or tachycardia | pregnancy complications | 25DTot | |
| 657 | | | | | 27745 | 7 | 27738 | | | | | 36919 | 6 | 36913 | | | Infections specific to the perinatal period | pregnancy complications | 25DTot | |
| 669 | | | | | 7 | | | | | | | 36606 | 168 | 36438 | | | Complications of labor and delivery NEC | pregnancy complications | 25DTot | |
| 671 | | | | | 7 | | | | | | | 36547 | 117 | 36430 | | | Venous/cerebrovascular complications embolism in pregnancy and the puerperium | pregnancy complications | 25DTot | |
| 674 | | | | | 7 | | | | | | | 36509 | 79 | 36430 | | | Other complications of the puerperium NEC | pregnancy complications | 25DTot | |
| 676 | | | | | 7 | | | | | | | 36440 | 10 | 36430 | | | Other disorders of the breast associated with childbirth and disorders of lactation | pregnancy complications | 25DTot | |
| 681 | 0.5041 | 0.0571 | 1.656 | 1.06e-18 | 24732 | 2737 | 21995 | 0.3872 | 0.04447 | 1.473 | 3.17e-18 | 32934 | 3053 | 29881 | 0.2043 | -0.09012 | Superficial cellulitis and abscess | dermatologic | 25DTot | |
| 681.1 | | | | | 22394 | 399 | 21995 | | | | | 30304 | 423 | 29881 | 0.4925 | -0.1125 | Cellulitis and abscess of fingers/toes | dermatologic | 25DTot | |
| 681.2 | | | | | 22204 | 209 | 21995 | | | | | 30123 | 242 | 29881 | | | Cellulitis and abscess of face/neck | dermatologic | 25DTot | |
| 681.3 | | | | | 22375 | 380 | 21995 | | | | | 30260 | 379 | 29881 | 0.3497 | -0.1742 | Cellulitis and abscess of arm/hand | dermatologic | 25DTot | |
| 681.5 | 0.6605 | 0.08476 | 1.936 | 6.61e-15 | 22978 | 983 | 21995 | 0.4082 | 0.07632 | 1.504 | 8.89e-08 | 30774 | 893 | 29881 | 0.05895 | -0.2108 | Cellulitis and abscess of leg, except foot | dermatologic | 25DTot | |
| 681.6 | | | | | 22233 | 238 | 21995 | | | | | 30029 | 148 | 29881 | | | Cellulitis and abscess of foot, toe | dermatologic | 25DTot | |
| 681.7 | | | | | 22491 | 496 | 21995 | 0.379 | 0.09677 | 1.461 | 9.00e-05 | 30505 | 624 | 29881 | 0.1172 | -0.2378 | Cellulitis and abscess of trunk | dermatologic | 25DTot | |
| 686 | 0.4022 | 0.1141 | 1.495 | 4.22e-04 | 22609 | 614 | 21995 | 0.3009 | 0.09118 | 1.351 | 9.66e-04 | 30530 | 649 | 29881 | 0.8827 | -0.0211 | Other local infections of skin and subcutaneous tissue | dermatologic | 25DTot | |
| 686.1 | | | | | 22123 | 128 | 21995 | | | | | 30019 | 138 | 29881 | | | Carbuncle and furuncle | dermatologic | 25DTot | |
| 686.2 | | | | | 22049 | 54 | 21995 | | | | | 29945 | 64 | 29881 | | | Impetigo | dermatologic | 25DTot | |
| 686.3 | | | | | 22049 | 54 | 21995 | | | | | 29925 | 44 | 29881 | | | Pilonidal cyst | dermatologic | 25DTot | |
| 686.4 | | | | | 22031 | 36 | 21995 | | | | | 29904 | 23 | 29881 | | | Pyogenic granuloma | dermatologic | 25DTot | |
| 686.5 | | | | | 22021 | 26 | 21995 | | | | | 29916 | 35 | 29881 | | | Pyoderma | dermatologic | 25DTot | |
| 687 | | | | | 21481 | 302 | 21179 | 0.6895 | 0.0937 | 1.993 | 1.87e-13 | 26852 | 564 | 26288 | 0.02447 | 0.4219 | Symptoms affecting skin | dermatologic | 25DTot | |
| 687.1 | 0.6381 | 0.06996 | 1.893 | 7.39e-20 | 22861 | 1682 | 21179 | 0.8264 | 0.04553 | 2.285 | 1.25e-73 | 29124 | 2836 | 26288 | 0.00246 | 0.2494 | Rash and other nonspecific skin eruption | dermatologic | 25DTot | |
| 687.3 | | | | | 21208 | 29 | 21179 | | | | | 26359 | 71 | 26288 | | | Changes in skin texture | dermatologic | 25DTot | |
| 687.4 | 0.8225 | 0.07235 | 2.276 | 6.00e-30 | 22559 | 1380 | 21179 | 0.8617 | 0.04775 | 2.367 | 8.37e-73 | 28776 | 2488 | 26288 | 0.5817 | 0.04705 | Disturbance of skin sensation | dermatologic | 25DTot | |
| 689 | 0.2355 | 0.09684 | 1.265 | 0.01504 | 26203 | 833 | 25370 | 0.532 | 0.06819 | 1.702 | 6.07e-15 | 34664 | 1062 | 33602 | 0.01596 | 0.2798 | Disorder of skin and subcutaneous tissue NOS | dermatologic | 25DTot | |
| 690 | | | | | 23839 | 493 | 23346 | | | | | 29843 | 385 | 29458 | 0.6074 | -0.07861 | Erythematosquamous dermatosis | dermatologic | 25DTot | |
| phenotype | male_beta | male_SE | male_OR | male_p | male_n_total | male_n_cases | male_n_controls | female_beta | female_SE | female_OR | female_p | female_n_total | female_n_cases | female_n_controls | interaction_p | interaction_beta | description | group | lab | label |
| 690.1 | | | | | 23835 | 489 | 23346 | | | | | 29839 | 381 | 29458 | 0.5485 | -0.09227 | Seborrheic dermatitis | dermatologic | 25DTot | |
| 691 | | | | | 23418 | 72 | 23346 | | | | | 29595 | 137 | 29458 | | | Congenital anomalies of skin | dermatologic | 25DTot | |

| phenotype | male_beta | male_SE | male_OR | male_p | male_n_total | male_n_cases | male_n_controls | female_beta | female_SE | female_OR | female_p | female_n_total | female_n_cases | female_n_controls | interaction_p | interaction_beta | description | group | lab | label |
|---|---|---|---|---|---|---|---|---|---|---|---|---|---|---|---|---|---|---|---|---|
| 691.1 | | | | | 23351 | 5 | 23346 | | | | | 29460 | 2 | 29458 | | | Ichthyosis congenita | dermatologic | 25DTot | |
| 691.3 | | | | | 23350 | 4 | 23346 | | | | | 29482 | 24 | 29458 | | | Congenital pigmentary anomalies of skin | dermatologic | 25DTot | |
| 694 | 0.4249 | 0.0944 | 1.529 | 6.77e-06 | 24181 | 835 | 23346 | 0.6913 | 0.0591 | 1.996 | 1.30e-31 | 30929 | 1471 | 29458 | 0.05541 | 0.2101 | Dyschromia and Vitiligo | dermatologic | 25DTot | |
| 694.1 | | | | | 23374 | 28 | 23346 | | | | | 29501 | 43 | 29458 | | | Vitiligo | dermatologic | 25DTot | |
| 694.2 | 0.4618 | 0.0951 | 1.587 | 1.20e-06 | 24148 | 802 | 23346 | 0.6884 | 0.06005 | 1.991 | 1.98e-30 | 30877 | 1419 | 29458 | 0.1364 | 0.1649 | Other dyschromia | dermatologic | 25DTot | |
| 694.3 | | | | | 23359 | 13 | 23346 | | | | | 29477 | 19 | 29458 | | | Vascular disorders of skin | dermatologic | 25DTot | |
| 695 | 0.6408 | 0.09082 | 1.898 | 1.71e-12 | 24180 | 834 | 23346 | 0.7669 | 0.04876 | 2.153 | 9.72e-56 | 31756 | 2298 | 29458 | 0.6732 | 0.04262 | Erythematous conditions | dermatologic | 25DTot | |
| 695.1 | | | | | 23358 | 12 | 23346 | | | | | 29480 | 22 | 29458 | | | Toxic erythema | dermatologic | 25DTot | |
| 695.2 | | | | | 23377 | 31 | 23346 | | | | | 29513 | 55 | 29458 | | | Bullous dermatoses | dermatologic | 25DTot | |
| 695.21 | | | | | 23356 | 10 | 23346 | | | | | 29480 | 22 | 29458 | | | Dermatitis herpetiformis | dermatologic | 25DTot | |
| 695.22 | | | | | 23363 | 17 | 23346 | | | | | 29486 | 28 | 29458 | | | Pemphigus and pemphigoid | dermatologic | 25DTot | |
| 695.3 | | | | | 23613 | 267 | 23346 | 0.7801 | 0.08457 | 2.182 | 2.87e-20 | 30121 | 663 | 29458 | 0.09935 | 0.2939 | Rosacea | dermatologic | 25DTot | |
| 695.4 | | | | | 23118 | 140 | 22978 | 1.038 | 0.07407 | 2.822 | 1.40e-44 | 29377 | 925 | 28452 | 0.7425 | 0.07135 | Lupus (localized and systemic) | dermatologic | 25DTot | |
| 695.41 | | | | | 23008 | 30 | 22978 | | | | | 28621 | 169 | 28452 | | | Cutaneous lupus erythematosus | dermatologic | 25DTot | |
| 695.42 | | | | | 23104 | 126 | 22978 | 1.057 | 0.0762 | 2.878 | 8.92e-44 | 29322 | 870 | 28452 | 0.8175 | -0.05099 | Systemic lupus erythematosus | dermatologic | 25DTot | |
| 695.7 | | | | | 23445 | 99 | 23346 | | | | | 29666 | 208 | 29458 | | | Prurigo and Lichen | dermatologic | 25DTot | |
| 695.8 | | | | | 23469 | 123 | 23346 | | | | | 29759 | 301 | 29458 | | | Other specified erythematous conditions | dermatologic | 25DTot | |
| 695.81 | | | | | 23357 | 11 | 23346 | | | | | 29501 | 43 | 29458 | | | Erythema nodosum | dermatologic | 25DTot | |
| 695.9 | | | | | 23478 | 132 | 23346 | | | | | 29596 | 138 | 29458 | | | Unspecified erythematous condition | dermatologic | 25DTot | |
| 696 | | | | | 23091 | 490 | 22601 | 0.5955 | 0.08664 | 1.814 | 6.26e-12 | 28469 | 689 | 27780 | 0.8519 | -0.02689 | Psoriasis and related disorders | dermatologic | 25DTot | |
| 696.2 | | | | | 23352 | 6 | 23346 | | | | | 29465 | 7 | 29458 | | | Parapsoriasis | dermatologic | 25DTot | |
| 696.3 | | | | | 23354 | 8 | 23346 | | | | | 29466 | 8 | 29458 | | | Pityriasis | dermatologic | 25DTot | |
| 696.4 | | | | | 23086 | 485 | 22601 | 0.5948 | 0.08768 | 1.813 | 1.17e-11 | 28453 | 673 | 27780 | 0.822 | -0.03265 | Psoriasis | dermatologic | 25DTot | |
| 696.41 | | | | | 23001 | 400 | 22601 | 0.5825 | 0.09752 | 1.791 | 2.32e-09 | 28318 | 538 | 27780 | 0.9483 | -0.01041 | Psoriasis vulgaris | dermatologic | 25DTot | |
| 696.42 | | | | | 22782 | 181 | 22601 | | | | | 28051 | 271 | 27780 | | | Psoriatic arthropathy | dermatologic | 25DTot | |
| 697 | | | | | 23461 | 115 | 23346 | | | | | 29654 | 196 | 29458 | | | Sarcoidosis | dermatologic | 25DTot | |
| 698 | | | | | 27077 | 348 | 26729 | 0.4987 | 0.07978 | 1.647 | 4.07e-10 | 35552 | 771 | 34781 | 0.4696 | -0.1097 | Pruritus and related conditions | dermatologic | 25DTot | |
| 700 | | | | | 24764 | 207 | 24557 | | | | | 32043 | 272 | 31771 | | | Corns and callosities | dermatologic | 25DTot | |
| 701 | 0.3258 | 0.08258 | 1.385 | 7.98e-05 | 25713 | 1156 | 24557 | 0.3711 | 0.05138 | 1.449 | 5.10e-13 | 33879 | 2108 | 31771 | 0.8693 | 0.01574 | Other hypertrophic and atrophic conditions of skin | dermatologic | 25DTot | |
| 701.1 | | | | | 24638 | 81 | 24557 | | | | | 31856 | 85 | 31771 | | | Keratoderma, acquired | dermatologic | 25DTot | |
| 701.2 | | | | | 24996 | 439 | 24557 | 0.1588 | 0.07935 | 1.172 | 0.04543 | 32665 | 894 | 31771 | 0.15 | -0.216 | Scar conditions and fibrosis of skin | dermatologic | 25DTot | |
| 701.3 | | | | | 24568 | 11 | 24557 | | | | | 31968 | 197 | 31771 | | | Circumscribed scleroderma | dermatologic | 25DTot | |
| 701.4 | | | | | 24632 | 75 | 24557 | | | | | 31928 | 157 | 31771 | | | Keloid scar | dermatologic | 25DTot | |
| 701.5 | | | | | 24606 | 49 | 24557 | | | | | 31836 | 65 | 31771 | | | Abnormal granulation tissue | dermatologic | 25DTot | |
| 701.6 | | | | | 24575 | 18 | 24557 | | | | | 31812 | 41 | 31771 | | | Acquired acanthosis nigricans | dermatologic | 25DTot | |
| 702 | 0.5898 | 0.05893 | 1.804 | 1.40e-23 | 24883 | 2887 | 21996 | 0.758 | 0.04396 | 2.134 | 1.27e-66 | 32069 | 3086 | 28983 | 0.02219 | 0.1665 | Degenerative skin conditions and other dermatoses | dermatologic | 25DTot | |
| 702.1 | 0.5858 | 0.06512 | 1.796 | 2.36e-19 | 25602 | 2131 | 23471 | 0.7975 | 0.05811 | 2.22 | 7.46e-43 | 33264 | 1502 | 31762 | 0.0235 | 0.1955 | Actinic keratosis | dermatologic | 25DTot | |
| 702.2 | 0.5762 | 0.06513 | 1.779 | 8.94e-19 | 25554 | 2083 | 23471 | 0.7611 | 0.04639 | 2.141 | 1.68e-60 | 34321 | 2559 | 31762 | 0.04704 | 0.1569 | Seborrheic keratosis | dermatologic | 25DTot | |
| 702.4 | | | | | 21998 | 2 | 21996 | | | | | 28993 | 10 | 28983 | | | Degenerative skin disorders | dermatologic | 25DTot | |

| phenotype | male_beta | male_SE | male_OR | male_p | male_n_total | male_n_cases | male_n_controls | female_beta | female_SE | female_OR | female_p | female_n_total | female_n_cases | female_n_controls | interaction_p | interaction_beta | description | group | lab | label |
|---|---|---|---|---|---|---|---|---|---|---|---|---|---|---|---|---|---|---|---|---|
| 703 | | | | | 25804 | 185 | 25619 | | | | | 33096 | 363 | 32733 | 0.1736 | -0.266 | Diseases of nail, NOS | dermatologic | 25DTot | |
| 703.1 | | | | | 25920 | 301 | 25619 | | | | | 33093 | 360 | 32733 | 0.3054 | 0.1796 | Ingrowing nail | dermatologic | 25DTot | |
| 704 | 0.4466 | 0.1173 | 1.563 | 1.41e-04 | 26130 | 511 | 25619 | 0.5753 | 0.06754 | 1.778 | 1.61e-17 | 33832 | 1099 | 32733 | 0.1675 | 0.1838 | Diseases of hair and hair follicles | dermatologic | 25DTot | |
| 704.1 | | | | | 25706 | 87 | 25619 | 0.6396 | 0.08322 | 1.896 | 1.52e-14 | 33424 | 691 | 32733 | 0.331 | 0.2813 | Alopecia | dermatologic | 25DTot | |
| 704.11 | | | | | 25639 | 20 | 25619 | | | | | 32796 | 63 | 32733 | | | Alopecia Areata | dermatologic | 25DTot | |
| 704.12 | | | | | 25622 | 3 | 25619 | | | | | 32824 | 91 | 32733 | | | Telogen effluvium | dermatologic | 25DTot | |
| 704.2 | | | | | 25621 | 2 | 25619 | | | | | 32916 | 183 | 32733 | | | Hirsutism | dermatologic | 25DTot | |
| 704.8 | | | | | 25903 | 284 | 25619 | | | | | 32966 | 233 | 32733 | 0.595 | 0.1102 | Other specified diseases of hair and hair follicles | dermatologic | 25DTot | |
| 705 | | | | | 24635 | 110 | 24525 | | | | | 32184 | 194 | 31990 | | | Disorders of sweat glands | dermatologic | 25DTot | |
| 705.1 | | | | | 24588 | 63 | 24525 | | | | | 32070 | 80 | 31990 | | | Dyshidrosis | dermatologic | 25DTot | |
| 705.3 | | | | | 24563 | 38 | 24525 | | | | | 32095 | 105 | 31990 | | | Hidradenitis | dermatologic | 25DTot | |
| 705.8 | | | | | 24661 | 136 | 24525 | | | | | 32208 | 218 | 31990 | | | Hyperhidrosis | dermatologic | 25DTot | |
| 706 | 0.5099 | 0.07459 | 1.665 | 8.14e-12 | 25847 | 1322 | 24525 | 0.6755 | 0.05293 | 1.965 | 2.63e-37 | 33934 | 1944 | 31990 | 0.7584 | 0.02751 | Diseases of sebaceous glands | dermatologic | 25DTot | |
| 706.1 | | | | | 24797 | 272 | 24525 | 0.6238 | 0.08991 | 1.866 | 3.98e-12 | 32819 | 829 | 31990 | 0.3382 | 0.1898 | Acne | dermatologic | 25DTot | |
| 706.2 | 0.4748 | 0.09782 | 1.608 | 1.21e-06 | 25246 | 721 | 24525 | 0.7201 | 0.07484 | 2.055 | 6.43e-22 | 32825 | 835 | 31990 | 0.08255 | 0.2104 | Sebaceous cyst | dermatologic | 25DTot | |
| 706.3 | | | | | 24526 | 1 | 24525 | | | | | 31991 | 1 | 31990 | | | Seborrhea | dermatologic | 25DTot | |
| 706.8 | | | | | 24813 | 288 | 24525 | | | | | 32225 | 235 | 31990 | 0.7042 | -0.07419 | Other specified diseases of sebaceous glands | dermatologic | 25DTot | |
| 707 | 0.4551 | 0.07974 | 1.576 | 1.15e-08 | 26784 | 1253 | 25531 | 0.3197 | 0.07354 | 1.377 | 1.38e-05 | 36008 | 999 | 35009 | 0.2766 | -0.1148 | Chronic ulcer of skin | dermatologic | 25DTot | |
| 707.1 | 0.4978 | 0.1161 | 1.645 | 1.80e-05 | 26112 | 581 | 25531 | | | | | 35488 | 479 | 35009 | 0.3643 | -0.1391 | Decubitus ulcer | dermatologic | 25DTot | |
| 707.2 | 0.4706 | 0.1036 | 1.601 | 5.55e-06 | 26220 | 689 | 25531 | | | | | 35483 | 474 | 35009 | 0.5549 | -0.08381 | Chronic ulcer of leg or foot | dermatologic | 25DTot | |
| 707.3 | | | | | 25727 | 196 | 25531 | | | | | 35188 | 179 | 35009 | | | Chronic ulcer of unspecified site | dermatologic | 25DTot | |
| 709 | | | | | 23344 | 211 | 23133 | 1.013 | 0.06509 | 2.754 | 1.32e-54 | 29862 | 1162 | 28700 | 0.03582 | 0.4021 | Diffuse diseases of connective tissue | dermatologic | 25DTot | |
| 709.2 | | | | | 23183 | 50 | 23133 | | | | | 29154 | 454 | 28700 | 0.06368 | 0.7394 | Sicca syndrome | dermatologic | 25DTot | |
| 709.3 | | | | | 23192 | 59 | 23133 | | | | | 28935 | 235 | 28700 | | | Systemic sclerosis | dermatologic | 25DTot | |
| 709.4 | | | | | 23156 | 23 | 23133 | | | | | 28760 | 60 | 28700 | | | Polymyositis | dermatologic | 25DTot | |
| 709.5 | | | | | 23161 | 28 | 23133 | | | | | 28793 | 93 | 28700 | | | Dermatomyositis | dermatologic | 25DTot | |
| 709.6 | | | | | 23165 | 32 | 23133 | | | | | 28946 | 246 | 28700 | | | Other specified diffuse diseases of connective tissue | dermatologic | 25DTot | |
| 709.7 | | | | | 23162 | 29 | 23133 | | | | | 28907 | 207 | 28700 | | | Unspecified diffuse connective tissue disease | dermatologic | 25DTot | |
| 710 | 0.3854 | 0.1032 | 1.47 | 1.87e-04 | 23512 | 830 | 22682 | 0.1791 | 0.09498 | 1.196 | 0.05937 | 30097 | 684 | 29413 | 0.2492 | -0.1579 | Osteomyelitis, periostitis, and other infections involving bone | musculoskeletal | 25DTot | |
| 710.1 | 0.3551 | 0.11 | 1.426 | 0.00125 | 23427 | 745 | 22682 | 0.09685 | 0.1095 | 1.102 | 0.3765 | 29958 | 545 | 29413 | 0.1137 | -0.2399 | Osteomyelitis | musculoskeletal | 25DTot | |
| 710.11 | | | | | 22957 | 275 | 22682 | | | | | 29563 | 150 | 29413 | | | Acute osteomyelitis | musculoskeletal | 25DTot | |
| 710.12 | | | | | 23048 | 366 | 22682 | | | | | 29661 | 248 | 29413 | 0.1763 | -0.2903 | Chronic osteomyelitis | musculoskeletal | 25DTot | |
| 710.19 | 0.373 | 0.1298 | 1.452 | 0.004058 | 23198 | 516 | 22682 | | | | | 29784 | 371 | 29413 | 0.1501 | -0.2602 | Unspecified osteomyelitis | musculoskeletal | 25DTot | |
| 710.2 | | | | | 22682 | | | | | | | 29416 | 3 | 29413 | | | Periostitis | musculoskeletal | 25DTot | |
| 710.3 | | | | | 22684 | 2 | 22682 | | | | | 29414 | 1 | 29413 | | | Osteopathy resulting from poliomyelitis | musculoskeletal | 25DTot | |

| phenotype | male_beta | male_SE | male_OR | male_p | male_n_total | male_n_cases | male_n_controls | female_beta | female_SE | female_OR | female_p | female_n_total | female_n_cases | female_n_controls | interaction_p | interaction_beta | description | group | lab | label |
|---|---|---|---|---|---|---|---|---|---|---|---|---|---|---|---|---|---|---|---|---|
| 711 | | | | | 22939 | 257 | 22682 | | | | | 29633 | 220 | 29413 | | | Arthropathy associated with infections | musculoskeletal | 25DTot | |
| 711.1 | | | | | 22893 | 211 | 22682 | | | | | 29566 | 153 | 29413 | | | Pyogenic arthritis | musculoskeletal | 25DTot | |
| 711.2 | | | | | 22696 | 14 | 22682 | | | | | 29424 | 11 | 29413 | | | Reiter's disease | musculoskeletal | 25DTot | |
| 711.3 | | | | | 22695 | 13 | 22682 | | | | | 29451 | 38 | 29413 | | | Behcet's syndrome | musculoskeletal | 25DTot | |
| 712 | | | | | 22798 | 116 | 22682 | | | | | 29481 | 68 | 29413 | | | Infective connective tissue disorders | musculoskeletal | 25DTot | |
| 713 | | | | | 22821 | 139 | 22682 | | | | | 29546 | 133 | 29413 | | | Arthropathy associated with other disorders classified elsewhere | musculoskeletal | 25DTot | |
| 713.5 | | | | | 22789 | 107 | 22682 | | | | | 29502 | 89 | 29413 | | | Arthropathy associated with neurological disorders | musculoskeletal | 25DTot | |
| 714 | 0.8363 | 0.09363 | 2.308 | 4.19e-19 | 24325 | 732 | 23593 | 0.6586 | 0.05331 | 1.932 | 4.60e-35 | 31860 | 1880 | 29980 | 0.1397 | -0.1559 | Rheumatoid arthritis and other inflammatory polyarthropathies | musculoskeletal | 25DTot | |
| 714.1 | 0.9278 | 0.1092 | 2.529 | 1.91e-17 | 24107 | 514 | 23593 | 0.67 | 0.06053 | 1.954 | 1.79e-28 | 31387 | 1407 | 29980 | 0.09145 | -0.206 | Rheumatoid arthritis | musculoskeletal | 25DTot | |
| 714.2 | | | | | 23636 | 43 | 23593 | | | | | 30097 | 117 | 29980 | | | Juvenile rheumatoid arthritis | musculoskeletal | 25DTot | |
| 715 | | | | | 23907 | 314 | 23593 | 0.661 | 0.08626 | 1.937 | 1.82e-14 | 30615 | 635 | 29980 | 0.6261 | 0.07986 | Other inflammatory spondylopathies | musculoskeletal | 25DTot | |
| 715.1 | | | | | 23774 | 181 | 23593 | | | | | 30461 | 481 | 29980 | 0.6523 | -0.09027 | Sacroiliitis NEC | musculoskeletal | 25DTot | |
| 715.2 | | | | | 23683 | 90 | 23593 | | | | | 30051 | 71 | 29980 | | | Ankylosing spondylitis | musculoskeletal | 25DTot | |
| 715.3 | | | | | 23620 | 27 | 23593 | | | | | 30038 | 58 | 29980 | | | Spinal enthesopathy | musculoskeletal | 25DTot | |
| 716 | 0.6828 | 0.08781 | 1.979 | 7.48e-15 | 23562 | 880 | 22682 | 0.5856 | 0.06133 | 1.796 | 1.33e-21 | 30774 | 1361 | 29413 | 0.4726 | -0.07552 | Other arthropathies | musculoskeletal | 25DTot | |
| 716.1 | | | | | 22700 | 18 | 22682 | | | | | 29437 | 24 | 29413 | | | Unspecified polyarthropathy or polyarthritis | musculoskeletal | 25DTot | |
| 716.2 | | | | | 22688 | 6 | 22682 | | | | | 29420 | 7 | 29413 | | | Unspecified monoarthritis | musculoskeletal | 25DTot | |
| 716.3 | | | | | 22683 | 1 | 22682 | | | | | 29414 | 1 | 29413 | | | Kaschin-Beck disease | musculoskeletal | 25DTot | |
| 716.8 | | | | | 22688 | 6 | 22682 | | | | | 29426 | 13 | 29413 | | | Palindromic rheumatism | musculoskeletal | 25DTot | |
| 716.9 | 0.6731 | 0.09303 | 1.96 | 4.65e-13 | 23461 | 779 | 22682 | 0.5805 | 0.06436 | 1.787 | 1.89e-19 | 30638 | 1225 | 29413 | 0.5309 | -0.06955 | Arthropathy NOS | musculoskeletal | 25DTot | |
| 717 | | | | | 27682 | 166 | 27516 | | | | | 36769 | 263 | 36506 | | | Polymyalgia Rheumatica | musculoskeletal | 25DTot | |
| 720 | 0.6855 | 0.07483 | 1.985 | 5.10e-20 | 22346 | 1436 | 20910 | 0.5525 | 0.05725 | 1.738 | 4.93e-22 | 29466 | 1690 | 27776 | 0.1435 | -0.1359 | Spinal stenosis | musculoskeletal | 25DTot | |
| 720.1 | 0.7789 | 0.08798 | 2.179 | 8.52e-19 | 21846 | 936 | 20910 | 0.5753 | 0.06855 | 1.778 | 4.79e-17 | 28903 | 1127 | 27776 | 0.04581 | -0.2193 | Spinal stenosis of lumbar region | musculoskeletal | 25DTot | |
| 721 | 0.743 | 0.06253 | 2.102 | 1.47e-32 | 23062 | 2152 | 20910 | 0.5864 | 0.0452 | 1.797 | 1.72e-38 | 30738 | 2962 | 27776 | 0.05168 | -0.1486 | Spondylosis and allied disorders | musculoskeletal | 25DTot | |
| 721.1 | 0.7482 | 0.06426 | 2.113 | 2.44e-31 | 22908 | 1998 | 20910 | 0.6098 | 0.04607 | 1.84 | 5.39e-40 | 30580 | 2804 | 27776 | 0.08291 | -0.1357 | Spondylosis without myelopathy | musculoskeletal | 25DTot | |
| 721.2 | | | | | 21155 | 245 | 20910 | | | | | 28026 | 250 | 27776 | | | Spondylosis with myelopathy | musculoskeletal | 25DTot | |
| 721.8 | | | | | 20961 | 51 | 20910 | | | | | 27797 | 21 | 27776 | | | Other allied disorders of spine | musculoskeletal | 25DTot | |
| 722 | 0.6301 | 0.06015 | 1.878 | 1.12e-25 | 23387 | 2477 | 20910 | 0.5476 | 0.04262 | 1.729 | 8.62e-38 | 31200 | 3424 | 27776 | 0.241 | -0.0855 | Intervertebral disc disorders | musculoskeletal | 25DTot | |
| 722.1 | 0.4884 | 0.1145 | 1.63 | 1.99e-05 | 21491 | 581 | 20910 | 0.5623 | 0.08049 | 1.755 | 2.84e-12 | 28557 | 781 | 27776 | 0.637 | 0.06507 | Displacement of intervertebral disc | musculoskeletal | 25DTot | |

| phenotype | male_beta | male_SE | male_OR | male_p | male_n_total | male_n_cases | male_n_controls | female_beta | female_SE | female_OR | female_p | female_n_total | female_n_cases | female_n_controls | interaction_p | interaction_beta | description | group | lab | label |
|---|---|---|---|---|---|---|---|---|---|---|---|---|---|---|---|---|---|---|---|---|
| 722.3 | | | | | 20912 | 2 | 20910 | | | | | 27777 | 1 | 27776 | | | Schmorl's nodes | musculoskeletal | 25DTot | |
| 722.6 | 0.7268 | 0.06682 | 2.068 | 1.47e-27 | 22720 | 1810 | 20910 | 0.593 | 0.0469 | 1.809 | 1.24e-36 | 30454 | 2678 | 27776 | 0.0735 | -0.1448 | Degeneration of intervertebral disc | musculoskeletal | 25DTot | |
| 722.7 | | | | | 21018 | 108 | 20910 | | | | | 27881 | 105 | 27776 | | | Intervertebral disc disorder with myelopathy | musculoskeletal | 25DTot | |
| 722.8 | | | | | 21238 | 328 | 20910 | | | | | 28155 | 379 | 27776 | 0.2637 | -0.2098 | Postlaminectomy syndrome | musculoskeletal | 25DTot | |
| 722.9 | | | | | 21012 | 102 | 20910 | | | | | 27883 | 107 | 27776 | | | Other and unspecified disc disorder | musculoskeletal | 25DTot | |
| 723 | | | | | 21252 | 342 | 20910 | | | | | 28186 | 410 | 27776 | 0.05101 | 0.3673 | Other disorders of cervical region | musculoskeletal | 25DTot | |
| 723.1 | | | | | 20943 | 33 | 20910 | | | | | 27812 | 36 | 27776 | | | Torticollis | musculoskeletal | 25DTot | |
| 724 | | | | | 21393 | 483 | 20910 | 0.528 | 0.07594 | 1.695 | 3.59e-12 | 28648 | 872 | 27776 | 0.8358 | -0.02925 | Other and unspecified disorders of back | musculoskeletal | 25DTot | |
| 724.1 | | | | | 21038 | 128 | 20910 | | | | | 28127 | 351 | 27776 | | | Disorders of sacrum | musculoskeletal | 25DTot | |
| 724.2 | | | | | 21034 | 124 | 20910 | | | | | 28108 | 332 | 27776 | | | Disorders of coccyx | musculoskeletal | 25DTot | |
| 724.8 | | | | | 21013 | 103 | 20910 | | | | | 27951 | 175 | 27776 | | | Other symptoms referable to back | musculoskeletal | 25DTot | |
| 724.9 | | | | | 20981 | 71 | 20910 | | | | | 27892 | 116 | 27776 | | | Other unspecified back disorders | musculoskeletal | 25DTot | |
| 726 | 0.5953 | 0.05846 | 1.813 | 2.40e-24 | 23098 | 2817 | 20281 | 0.7913 | 0.03907 | 2.206 | 3.32e-91 | 30370 | 4595 | 25775 | 0.00377 | 0.2027 | Peripheral enthesopathies and allied syndromes | musculoskeletal | 25DTot | |
| 726.1 | 0.6836 | 0.08099 | 1.981 | 3.15e-17 | 21421 | 1140 | 20281 | 0.8438 | 0.0468 | 2.325 | 1.16e-72 | 28550 | 2775 | 25775 | 0.08633 | 0.1596 | Enthesopathy | musculoskeletal | 25DTot | |
| 726.2 | | | | | 20365 | 84 | 20281 | | | | | 25894 | 119 | 25775 | | | Synoviopathy | musculoskeletal | 25DTot | |
| 726.3 | | | | | 20728 | 447 | 20281 | | | | | 26194 | 419 | 25775 | 0.8965 | 0.02023 | Bursitis | musculoskeletal | 25DTot | |
| 726.4 | | | | | 20361 | 80 | 20281 | | | | | 25949 | 174 | 25775 | | | Calcaneal spur; Exostosis NOS | musculoskeletal | 25DTot | |
| 727 | 0.4883 | 0.07268 | 1.63 | 1.83e-11 | 21945 | 1664 | 20281 | 0.7391 | 0.04687 | 2.094 | 5.19e-56 | 28564 | 2789 | 25775 | 0.002165 | 0.2632 | Other disorders of synovium, tendon, and bursa | musculoskeletal | 25DTot | |
| 727.1 | 0.5018 | 0.1024 | 1.652 | 9.55e-07 | 21009 | 728 | 20281 | 0.7943 | 0.06085 | 2.213 | 6.18e-39 | 27213 | 1438 | 25775 | 0.01675 | 0.2821 | Synovitis and tenosynovitis | musculoskeletal | 25DTot | |
| 727.2 | | | | | 20360 | 79 | 20281 | | | | | 25927 | 152 | 25775 | | | Bursitis disorders | musculoskeletal | 25DTot | |
| 727.4 | | | | | 20471 | 190 | 20281 | | | | | 26261 | 486 | 25775 | 0.02772 | 0.5136 | Ganglion and cyst of synovium, tendon, and bursa | musculoskeletal | 25DTot | |
| 727.5 | | | | | 20324 | 43 | 20281 | | | | | 25904 | 129 | 25775 | | | Rupture of synovium | musculoskeletal | 25DTot | |
| 727.6 | | | | | 20660 | 379 | 20281 | | | | | 26104 | 329 | 25775 | 0.01611 | 0.4377 | Rupture of tendon, nontraumatic | musculoskeletal | 25DTot | |
| 727.7 | | | | | 20347 | 66 | 20281 | | | | | 25864 | 89 | 25775 | | | Contracture of tendon (sheath) | musculoskeletal | 25DTot | |
| 727.8 | | | | | 20310 | 29 | 20281 | | | | | 25829 | 54 | 25775 | | | Plica syndrome | musculoskeletal | 25DTot | |
| 728 | | | | | 20364 | 83 | 20281 | | | | | 25899 | 124 | 25775 | | | Disorders of muscle, ligament, and fascia | musculoskeletal | 25DTot | |
| 728.1 | | | | | 20313 | 32 | 20281 | | | | | 25798 | 23 | 25775 | | | Muscular calcification and ossification | musculoskeletal | 25DTot | |
| 728.2 | | | | | 20304 | 23 | 20281 | | | | | 25866 | 91 | 25775 | | | Laxity of ligament or hypermobility syndrome | musculoskeletal | 25DTot | |

| phenotype | male_beta | male_SE | male_OR | male_p | male_n_total | male_n_cases | male_n_controls | female_beta | female_SE | female_OR | female_p | female_n_total | female_n_cases | female_n_controls | interaction_p | interaction_beta | description | group | lab | label |
|---|---|---|---|---|---|---|---|---|---|---|---|---|---|---|---|---|---|---|---|---|
| 728.7 | | | | | 20721 | 440 | 20281 | 0.784 | 0.08202 | 2.19 | 1.19e-21 | 26509 | 734 | 25775 | 0.02996 | 0.3302 | Fasciitis | musculoskeletal | 25DTot | |
| 728.71 | | | | | 20411 | 130 | 20281 | | | | | 25848 | 73 | 25775 | | | Contracture of palmar fascia [Dupuytren's disease] | musculoskeletal | 25DTot | |
| 729 | 0.5035 | 0.1057 | 1.655 | 1.90e-06 | 20964 | 683 | 20281 | 0.666 | 0.07433 | 1.946 | 3.25e-19 | 26711 | 936 | 25775 | 0.1638 | 0.1781 | Other disorders of soft tissues | musculoskeletal | 25DTot | |
| 729.1 | | | | | 20284 | 3 | 20281 | | | | | 25782 | 7 | 25775 | | | Rheumatism, unspecified and fibrositis | musculoskeletal | 25DTot | |
| 729.3 | | | | | 20292 | 11 | 20281 | | | | | 25825 | 50 | 25775 | | | Panniculitis | musculoskeletal | 25DTot | |
| 729.7 | | | | | 20307 | 26 | 20281 | | | | | 25785 | 10 | 25775 | | | Nontraumatic compartment syndrome | musculoskeletal | 25DTot | |
| 731 | | | | | 22403 | 94 | 22309 | | | | | 27658 | 148 | 27510 | | | Osteitis deformans and osteopathies associated with other disorders classified elsewhere | musculoskeletal | 25DTot | |
| 731.1 | | | | | 22320 | 11 | 22309 | | | | | 27517 | 7 | 27510 | | | Osteitis deformans [Paget's disease of bone] | musculoskeletal | 25DTot | |
| 732 | | | | | 22384 | 75 | 22309 | | | | | 27583 | 73 | 27510 | | | Osteochondropathies | musculoskeletal | 25DTot | |
| 732.1 | | | | | 22341 | 32 | 22309 | | | | | 27549 | 39 | 27510 | | | Juvenile osteochondrosis | musculoskeletal | 25DTot | |
| 732.7 | | | | | 22348 | 39 | 22309 | | | | | 27539 | 29 | 27510 | | | Osteochondritis dissecans | musculoskeletal | 25DTot | |
| 733 | 1.129 | 0.09437 | 3.093 | 5.47e-33 | 22918 | 609 | 22309 | 0.8499 | 0.04918 | 2.339 | 6.59e-67 | 29872 | 2362 | 27510 | 0.1687 | -0.1474 | Other disorders of bone and cartilage | musculoskeletal | 25DTot | |
| 733.2 | | | | | 22336 | 27 | 22309 | | | | | 27554 | 44 | 27510 | | | Cyst of bone | musculoskeletal | 25DTot | |
| 733.4 | | | | | 22516 | 207 | 22309 | | | | | 27762 | 252 | 27510 | | | Aseptic necrosis of bone | musculoskeletal | 25DTot | |
| 733.6 | | | | | 22334 | 25 | 22309 | | | | | 27574 | 64 | 27510 | | | Costochondritis | musculoskeletal | 25DTot | |
| 733.8 | | | | | 22558 | 249 | 22309 | | | | | 27858 | 348 | 27510 | 0.116 | 0.3442 | Malunion and nonunion of fracture | musculoskeletal | 25DTot | |
| 733.9 | | | | | 22350 | 41 | 22309 | | | | | 27565 | 55 | 27510 | | | Chondromalacia | musculoskeletal | 25DTot | |
| 735 | 0.5734 | 0.09 | 1.774 | 1.88e-10 | 24919 | 827 | 24092 | 0.6594 | 0.05938 | 1.934 | 1.19e-28 | 32235 | 1405 | 30830 | 0.2793 | 0.1153 | Acquired foot deformities | musculoskeletal | 25DTot | |
| 735.1 | | | | | 24190 | 98 | 24092 | | | | | 30970 | 140 | 30830 | | | Flat foot | musculoskeletal | 25DTot | |
| 735.2 | | | | | 24498 | 406 | 24092 | 0.6819 | 0.08132 | 1.978 | 5.06e-17 | 31534 | 704 | 30830 | 0.5084 | -0.09405 | Acquired toe deformities | musculoskeletal | 25DTot | |
| 735.21 | | | | | 24374 | 282 | 24092 | 0.6594 | 0.09263 | 1.934 | 1.09e-12 | 31365 | 535 | 30830 | 0.3578 | -0.1523 | Hammer toe (acquired) | musculoskeletal | 25DTot | |
| 735.22 | | | | | 24107 | 15 | 24092 | | | | | 30844 | 14 | 30830 | | | Claw toe (acquired) | musculoskeletal | 25DTot | |
| 735.23 | | | | | 24184 | 92 | 24092 | | | | | 30968 | 138 | 30830 | | | Hallux rigidus | musculoskeletal | 25DTot | |
| 735.3 | | | | | 24207 | 115 | 24092 | 0.7504 | 0.09449 | 2.118 | 2.00e-15 | 31337 | 507 | 30830 | 0.4859 | -0.159 | Hallux valgus (Bunion) | musculoskeletal | 25DTot | |
| 736 | | | | | 24293 | 201 | 24092 | | | | | 31078 | 248 | 30830 | | | Other acquired deformities of limbs | musculoskeletal | 25DTot | |
| 736.1 | | | | | 24103 | 11 | 24092 | | | | | 30840 | 10 | 30830 | | | Acquired deformities of forearm | musculoskeletal | 25DTot | |
| 736.2 | | | | | 24158 | 66 | 24092 | | | | | 30902 | 72 | 30830 | | | Acquired deformities of finger | musculoskeletal | 25DTot | |
| 736.3 | | | | | 24100 | 8 | 24092 | | | | | 30841 | 11 | 30830 | | | Acquired deformities of hip | musculoskeletal | 25DTot | |
| 736.4 | | | | | 24104 | 12 | 24092 | | | | | 30857 | 27 | 30830 | | | Genu valgum or varum (acquired) | musculoskeletal | 25DTot | |

| phenotype | male_beta | male_SE | male_OR | male_p | male_n_total | male_n_cases | male_n_controls | female_beta | female_SE | female_OR | female_p | female_n_total | female_n_cases | female_n_controls | interaction_p | interaction_beta | description | group | lab | label |
|---|---|---|---|---|---|---|---|---|---|---|---|---|---|---|---|---|---|---|---|---|
| 736.5 | | | | | 24095 | 3 | 24092 | | | | | 30840 | 10 | 30830 | | | Acquired deformities of knee | musculoskeletal | 25DTot | |
| 736.6 | | | | | 24114 | 22 | 24092 | | | | | 30876 | 46 | 30830 | | | Unequal leg length (acquired) | musculoskeletal | 25DTot | |
| 737 | | | | | 24514 | 422 | 24092 | 0.3129 | 0.07762 | 1.367 | 5.55e-05 | 31760 | 930 | 30830 | 0.2313 | 0.1898 | Curvature of spine | musculoskeletal | 25DTot | |
| 737.1 | | | | | 24226 | 134 | 24092 | | | | | 31005 | 175 | 30830 | | | Kyphosis (acquired) | musculoskeletal | 25DTot | |
| 737.2 | | | | | 24093 | 1 | 24092 | | | | | 30832 | 2 | 30830 | | | Lordosis (acquired) | musculoskeletal | 25DTot | |
| 737.3 | | | | | 24413 | 321 | 24092 | 0.2867 | 0.08363 | 1.332 | 6.08e-04 | 31641 | 811 | 30830 | 0.1026 | 0.3025 | Kyphoscoliosis and scoliosis | musculoskeletal | 25DTot | |
| 738 | 0.4133 | 0.1151 | 1.512 | 3.29e-04 | 24637 | 545 | 24092 | 0.2796 | 0.07402 | 1.323 | 1.58e-04 | 31798 | 968 | 30830 | 0.5164 | -0.08746 | Other acquired musculoskeletal deformity | musculoskeletal | 25DTot | |
| 738.4 | | | | | 24467 | 375 | 24092 | 0.3293 | 0.0807 | 1.39 | 4.48e-05 | 31608 | 778 | 30830 | 0.251 | -0.1751 | Acquired spondylolisthesis | musculoskeletal | 25DTot | |
| 739 | | | | | 24307 | 215 | 24092 | | | | | 31047 | 217 | 30830 | | | Contracture of joint | musculoskeletal | 25DTot | |
| 740 | 0.6015 | 0.05146 | 1.825 | 1.45e-31 | 25263 | 4641 | 20622 | 0.5885 | 0.03544 | 1.801 | 6.13e-62 | 33672 | 7527 | 26145 | 0.6022 | -0.0325 | Osteoarthrosis | musculoskeletal | 25DTot | |
| 740.1 | 0.5387 | 0.06004 | 1.714 | 2.88e-19 | 23687 | 3065 | 20622 | 0.587 | 0.04129 | 1.799 | 7.19e-46 | 30839 | 4694 | 26145 | 0.6269 | 0.03535 | Osteoarthritis; localized | musculoskeletal | 25DTot | |
| 740.11 | 0.5467 | 0.06198 | 1.727 | 1.14e-18 | 23439 | 2817 | 20622 | 0.5789 | 0.04246 | 1.784 | 2.52e-42 | 30486 | 4341 | 26145 | 0.849 | 0.01427 | Osteoarthrosis, localized, primary | musculoskeletal | 25DTot | |
| 740.12 | | | | | 20717 | 95 | 20622 | | | | | 26285 | 140 | 26145 | | | Osteoarthrosis, localized, secondary | musculoskeletal | 25DTot | |
| 740.2 | 1.099 | 0.09972 | 3 | 3.19e-28 | 21264 | 642 | 20622 | 0.8363 | 0.05599 | 2.308 | 1.87e-50 | 28076 | 1931 | 26145 | 0.00505 | -0.3174 | Osteoarthrosis, generalized | musculoskeletal | 25DTot | |
| 740.3 | | | | | 20669 | 47 | 20622 | | | | | 26282 | 137 | 26145 | | | Osteoarthrosis involving more than one site, but not specified as generalized | musculoskeletal | 25DTot | |
| 740.9 | 0.7285 | 0.06272 | 2.072 | 3.42e-31 | 23067 | 2445 | 20622 | 0.6821 | 0.04279 | 1.978 | 3.32e-57 | 30322 | 4177 | 26145 | 0.4464 | -0.05766 | Osteoarthrosis NOS | musculoskeletal | 25DTot | |
| 741 | 0.4896 | 0.06024 | 1.632 | 4.41e-16 | 24701 | 2405 | 22296 | 0.5665 | 0.04081 | 1.762 | 8.13e-44 | 32920 | 3637 | 29283 | 0.1586 | 0.1016 | Symptoms and disorders of the joints | musculoskeletal | 25DTot | |
| 741.1 | | | | | 22321 | 25 | 22296 | | | | | 29334 | 51 | 29283 | | | Ankylosis of joint | musculoskeletal | 25DTot | |
| 741.2 | 0.2061 | 0.1177 | 1.229 | 0.08002 | 22901 | 605 | 22296 | 0.5396 | 0.06473 | 1.715 | 7.63e-17 | 30510 | 1227 | 29283 | 0.0122 | 0.3331 | Stiffness of joint | musculoskeletal | 25DTot | |
| 741.3 | 0.6236 | 0.1028 | 1.866 | 1.33e-09 | 22942 | 646 | 22296 | 0.7766 | 0.0727 | 2.174 | 1.24e-26 | 30187 | 904 | 29283 | 0.2137 | 0.1544 | Difficulty in walking | musculoskeletal | 25DTot | |
| 741.4 | 0.6167 | 0.1009 | 1.853 | 9.80e-10 | 22964 | 668 | 22296 | 0.5447 | 0.07241 | 1.724 | 5.38e-14 | 30241 | 958 | 29283 | 0.7584 | -0.0377 | Joint effusions | musculoskeletal | 25DTot | |
| 741.5 | | | | | 22328 | 32 | 22296 | | | | | 29301 | 18 | 29283 | | | Hemarthrosis | musculoskeletal | 25DTot | |
| 741.6 | | | | | 22307 | 11 | 22296 | | | | | 29300 | 17 | 29283 | | | Villonodular synovitis | musculoskeletal | 25DTot | |
| 742 | | | | | 22735 | 439 | 22296 | 0.4867 | 0.08902 | 1.627 | 4.58e-08 | 29934 | 651 | 29283 | 0.5182 | 0.1003 | Derangement of joint, non-traumatic | musculoskeletal | 25DTot | |
| 742.1 | | | | | 22319 | 23 | 22296 | | | | | 29301 | 18 | 29283 | | | Loose body in joint | musculoskeletal | 25DTot | |
| 742.2 | | | | | 22327 | 31 | 22296 | | | | | 29340 | 57 | 29283 | | | Pathological, developmental or recurrent dislocation | musculoskeletal | 25DTot | |
| 742.8 | | | | | 22322 | 26 | 22296 | | | | | 29329 | 46 | 29283 | | | Articular cartilage disorder | musculoskeletal | 25DTot | |

| phenotype | male_beta | male_SE | male_OR | male_p | male_n_total | male_n_cases | male_n_controls | female_beta | female_SE | female_OR | female_p | female_n_total | female_n_cases | female_n_controls | interaction_p | interaction_beta | description | group | lab | label |
|---|---|---|---|---|---|---|---|---|---|---|---|---|---|---|---|---|---|---|---|---|
| 742.9 | | | | | 22643 | 347 | 22296 | 0.5513 | 0.09722 | 1.736 | 1.42e-08 | 29808 | 525 | 29283 | 0.3965 | 0.1453 | Other derangement of joint | musculoskeletal | 25DTot | |
| 743 | 1.503 | 0.05578 | 4.493 | 8.39e-160 | 26032 | 1946 | 24086 | 1.25 | 0.03558 | 3.491 | 1.86e-270 | 33952 | 6028 | 27924 | 0.003822 | -0.1913 | Osteoporosis, osteopenia and pathological fracture | musculoskeletal | 25DTot | |
| 743.1 | 1.843 | 0.06962 | 6.318 | 1.78e-154 | 25110 | 1024 | 24086 | 1.416 | 0.04171 | 4.12 | 1.56e-252 | 31886 | 3962 | 27924 | 9.34e-09 | -0.4673 | Osteoporosis | musculoskeletal | 25DTot | |
| 743.11 | 1.757 | 0.0799 | 5.794 | 3.60e-107 | 24850 | 764 | 24086 | 1.465 | 0.04514 | 4.327 | 5.22e-231 | 31204 | 3280 | 27924 | 3.96e-04 | -0.3256 | Osteoporosis NOS | musculoskeletal | 25DTot | |
| 743.12 | | | | | 24196 | 110 | 24086 | 2.374 | 0.08407 | 10.74 | 2.16e-175 | 28758 | 834 | 27924 | 0.01327 | -0.5533 | Senile osteoporosis | musculoskeletal | 25DTot | |
| 743.13 | | | | | 24546 | 460 | 24086 | 1.636 | 0.06711 | 5.133 | 3.17e-131 | 29013 | 1089 | 27924 | 1.30e-04 | -0.4502 | Other specified osteoporosis | musculoskeletal | 25DTot | |
| 743.2 | | | | | 24445 | 359 | 24086 | 1.102 | 0.08936 | 3.012 | 5.74e-35 | 28551 | 627 | 27924 | 0.03017 | 0.3566 | Pathologic fracture | musculoskeletal | 25DTot | |
| 743.21 | | | | | 24187 | 101 | 24086 | | | | | 28089 | 165 | 27924 | | | Pathologic fracture of vertebrae | musculoskeletal | 25DTot | |
| 743.22 | | | | | 24160 | 74 | 24086 | | | | | 28032 | 108 | 27924 | | | Pathologic fracture of femur | musculoskeletal | 25DTot | |
| 743.4 | | | | | 24126 | 40 | 24086 | | | | | 28053 | 129 | 27924 | | | Stress fracture | musculoskeletal | 25DTot | |
| 743.9 | 1.613 | 0.08428 | 5.019 | 1.16e-81 | 24819 | 733 | 24086 | 1.556 | 0.04805 | 4.742 | 3.57e-230 | 30516 | 2592 | 27924 | 0.2565 | 0.1088 | Osteopenia or other disorder of bone and cartilage | musculoskeletal | 25DTot | |
| 745 | 0.5683 | 0.04714 | 1.765 | 1.85e-33 | 24606 | 7378 | 17228 | 0.8945 | 0.03306 | 2.446 | 3.29e-161 | 32439 | 12064 | 20375 | 5.73e-10 | 0.3548 | Pain in joint | musculoskeletal | 25DTot | |
| 747 | -0.1612 | 0.08973 | 0.8511 | 0.07234 | 26877 | 1323 | 25554 | -0.3423 | 0.08046 | 0.7102 | 2.10e-05 | 36150 | 1183 | 34967 | 0.02378 | -0.2668 | Cardiac and circulatory congenital anomalies | congenital anomalies | 25DTot | |
| 747.1 | -0.1389 | 0.09226 | 0.8703 | 0.1321 | 26767 | 1213 | 25554 | -0.3265 | 0.08494 | 0.7215 | 1.21e-04 | 36008 | 1041 | 34967 | 0.0208 | -0.2838 | Cardiac congenital anomalies | congenital anomalies | 25DTot | |
| 747.11 | | | | | 25997 | 443 | 25554 | -0.4429 | 0.1219 | 0.6421 | 2.80e-04 | 35539 | 572 | 34967 | 0.9496 | 0.01338 | Cardiac shunt/ heart septal defect | congenital anomalies | 25DTot | |
| 747.12 | | | | | 25876 | 322 | 25554 | | | | | 35219 | 252 | 34967 | 0.06823 | -0.5303 | Valvular heart disease/ heart chambers | congenital anomalies | 25DTot | |
| 747.13 | | | | | 25697 | 143 | 25554 | | | | | 35131 | 164 | 34967 | | | Congenital anomalies of great vessels | congenital anomalies | 25DTot | |
| 747.2 | | | | | 25638 | 84 | 25554 | | | | | 35084 | 117 | 34967 | | | Congenital anomalies of peripheral vascular system | congenital anomalies | 25DTot | |
| 748 | | | | | 27528 | 39 | 27489 | | | | | 36621 | 44 | 36577 | | | Anomalies of respiratory system, congenital | congenital anomalies | 25DTot | |
| 749 | | | | | 27561 | 72 | 27489 | | | | | 36671 | 94 | 36577 | | | Congenital anomalies of face and neck | congenital anomalies | 25DTot | |
| 749.1 | | | | | 22492 | 16 | 22476 | | | | | 31338 | 20 | 31318 | | | Cleft palate | congenital anomalies | 25DTot | |
| 749.2 | | | | | 27520 | 31 | 27489 | | | | | 36613 | 36 | 36577 | | | Congenital anomalies of skull and face bones | congenital anomalies | 25DTot | |
| 750 | | | | | 26052 | 143 | 25909 | | | | | 35186 | 190 | 34996 | | | Digestive congenital anomalies | congenital anomalies | 25DTot | |
| 750.1 | | | | | 25951 | 42 | 25909 | | | | | 35050 | 54 | 34996 | | | Upper gastrointestinal congenital anomalies | congenital anomalies | 25DTot | |
| 750.11 | | | | | 25920 | 11 | 25909 | | | | | 35005 | 9 | 34996 | | | Esophageal atresia/tracheoesophageal fistula | congenital anomalies | 25DTot | |
| 750.13 | | | | | 25928 | 19 | 25909 | | | | | 35009 | 13 | 34996 | | | Congenital anomalies of mouth/tongue | congenital anomalies | 25DTot | |
| 750.14 | | | | | 25914 | 5 | 25909 | | | | | 35012 | 16 | 34996 | | | Congenital anomalies of esophagus | congenital anomalies | 25DTot | |
| 750.15 | | | | | 25911 | 2 | 25909 | | | | | 34997 | 1 | 34996 | | | Congenital anomalies of stomach | congenital anomalies | 25DTot | |
| 750.2 | | | | | 26004 | 95 | 25909 | | | | | 35124 | 128 | 34996 | | | Lower gastrointestinal congenital anomalies | congenital anomalies | 25DTot | |

| phenotype | male_beta | male_SE | male_OR | male_p | male_n_total | male_n_cases | male_n_controls | female_beta | female_SE | female_OR | female_p | female_n_total | female_n_cases | female_n_controls | interaction_p | interaction_beta | description | group | lab | label |
|---|---|---|---|---|---|---|---|---|---|---|---|---|---|---|---|---|---|---|---|---|
| 750.21 | | | | | 25974 | 65 | 25909 | | | | | 35061 | 65 | 34996 | | | Congenital anomalies of intestine | congenital anomalies | 25DTot | |
| 750.22 | | | | | 25936 | 27 | 25909 | | | | | 35055 | 59 | 34996 | | | Congenital anomaly of gallbladder, bile ducts, liver, pancreas | congenital anomalies | 25DTot | |
| 750.5 | | | | | 25913 | 4 | 25909 | | | | | 34999 | 3 | 34996 | | | Congenital hypertrophic pyloric stenosis | congenital anomalies | 25DTot | |
| 751 | | | | | 26398 | 489 | 25909 | | | | | 35443 | 447 | 34996 | 0.005003 | -0.4537 | Genitourinary congenital anomalies | congenital anomalies | 25DTot | |
| 751.1 | | | | | 26010 | 101 | 25909 | | | | | 35087 | 91 | 34996 | | | Congenital anomalies of genital organs | congenital anomalies | 25DTot | |
| 751.11 | | | | | 7 | | | | | | | 35071 | 79 | 34992 | | | Congenital anomalies of female genital organs | congenital anomalies | 25DTot | |
| 751.12 | | | | | 25992 | 90 | 25902 | | | | | 4 | | | | | Congenital anomalies of male genital organs | congenital anomalies | 25DTot | |
| 751.2 | | | | | 26267 | 358 | 25909 | | | | | 35304 | 308 | 34996 | 0.001433 | -0.5844 | Congenital anomalies of urinary system | congenital anomalies | 25DTot | |
| 751.21 | | | | | 26140 | 231 | 25909 | | | | | 35199 | 203 | 34996 | | | Cystic kidney disease | congenital anomalies | 25DTot | |
| 751.22 | | | | | 26007 | 98 | 25909 | | | | | 35074 | 78 | 34996 | | | Other specified congenital anomalies of kidney | congenital anomalies | 25DTot | |
| 751.3 | | | | | 25949 | 40 | 25909 | | | | | 35047 | 51 | 34996 | | | Obstructive genitourinary defect | congenital anomalies | 25DTot | |
| 752 | | | | | 27335 | 160 | 27175 | | | | | 36299 | 223 | 36076 | | | Nervous system congenital anomalies | congenital anomalies | 25DTot | |
| 752.1 | | | | | 27234 | 59 | 27175 | | | | | 36188 | 112 | 36076 | | | Neural tube defects | congenital anomalies | 25DTot | |
| 752.11 | | | | | 27225 | 50 | 27175 | | | | | 36167 | 91 | 36076 | | | Spina bifida | congenital anomalies | 25DTot | |
| 752.2 | | | | | 27279 | 104 | 27175 | | | | | 36200 | 124 | 36076 | | | Other specified congenital anomalies of nervous system | congenital anomalies | 25DTot | |
| 753 | | | | | 27198 | 23 | 27175 | | | | | 36127 | 51 | 36076 | | | Congenital anomalies of the eye | congenital anomalies | 25DTot | |
| 753.1 | | | | | 27183 | 8 | 27175 | | | | | 36087 | 11 | 36076 | | | Congenital cataract and lens anomalies | congenital anomalies | 25DTot | |
| 753.2 | | | | | 27181 | 6 | 27175 | | | | | 36095 | 19 | 36076 | | | Congenital anomalies of posterior segment of eye | congenital anomalies | 25DTot | |
| 754 | | | | | 27083 | 58 | 27025 | | | | | 35886 | 111 | 35775 | | | Congenital musculoskeletal deformities of spine | congenital anomalies | 25DTot | |
| 754.1 | | | | | 27033 | 8 | 27025 | | | | | 35793 | 18 | 35775 | | | Lumbosacral spondylolysis, congenital | congenital anomalies | 25DTot | |
| 754.2 | | | | | 27059 | 34 | 27025 | | | | | 35837 | 62 | 35775 | | | Spondylolisthesis, congenital | congenital anomalies | 25DTot | |
| 755 | | | | | 27119 | 94 | 27025 | | | | | 35920 | 145 | 35775 | | | Congenital anomalies of limbs | congenital anomalies | 25DTot | |
| phenotype | male_beta | male_SE | male_OR | male_p | male_n_total | male_n_cases | male_n_controls | female_beta | female_SE | female_OR | female_p | female_n_total | female_n_cases | female_n_controls | interaction_p | interaction_beta | description | group | lab | label |
| 755.1 | | | | | 27058 | 33 | 27025 | | | | | 35834 | 59 | 35775 | | | Congenital deformities of feet | congenital anomalies | 25DTot | |
| 755.3 | | | | | 27031 | 6 | 27025 | | | | | 35781 | 6 | 35775 | | | Congenital anomaly of fingers/toes | congenital anomalies | 25DTot | |
| 755.4 | | | | | 27034 | 9 | 27025 | | | | | 35795 | 20 | 35775 | | | Congenital anomalies of upper limb, including shoulder girdle | congenital anomalies | 25DTot | |
| 755.6 | | | | | 27065 | 40 | 27025 | | | | | 35837 | 62 | 35775 | | | Other congenital anomalies of lower limb, including pelvic girdle | congenital anomalies | 25DTot | |
| 755.61 | | | | | 27060 | 35 | 27025 | | | | | 35828 | 53 | 35775 | | | Congenital hip dysplasia and deformity | congenital anomalies | 25DTot | |
| 756 | | | | | 27125 | 100 | 27025 | | | | | 35939 | 164 | 35775 | | | Other congenital musculoskeletal anomalies | congenital anomalies | 25DTot | |

| phenotype | male_beta | male_SE | male_OR | male_p | male_n_total | male_n_cases | male_n_controls | female_beta | female_SE | female_OR | female_p | female_n_total | female_n_cases | female_n_controls | interaction_p | interaction_beta | description | group | lab | label |
|---|---|---|---|---|---|---|---|---|---|---|---|---|---|---|---|---|---|---|---|---|
| 756.1 | | | | | 27036 | 11 | 27025 | | | | | 35791 | 16 | 35775 | | | Congenital anomalies of abdominal wall; diaphram | congenital anomalies | 25DTot | |
| 756.2 | | | | | 27064 | 39 | 27025 | | | | | 35801 | 26 | 35775 | | | Pectus and other congenital anomalies of ribs/sternum | congenital anomalies | 25DTot | |
| 756.21 | | | | | 27054 | 29 | 27025 | | | | | 35798 | 23 | 35775 | | | Pectus excavatum | congenital anomalies | 25DTot | |
| 756.22 | | | | | 27036 | 11 | 27025 | | | | | 35776 | 1 | 35775 | | | Pectus carinatum | congenital anomalies | 25DTot | |
| 756.3 | | | | | 27041 | 16 | 27025 | | | | | 35840 | 65 | 35775 | | | Congenital anomalies of muscle, tendon, fascia, and connective tissue | congenital anomalies | 25DTot | |
| 756.5 | | | | | 27043 | 18 | 27025 | | | | | 35804 | 29 | 35775 | | | Congenital osteodystrophies | congenital anomalies | 25DTot | |
| 757 | | | | | 26746 | 13 | 26733 | | | | | 35965 | 21 | 35944 | | | Congenital anomalies of the integument | congenital anomalies | 25DTot | |
| 758 | | | | | 26835 | 102 | 26733 | | | | | 36032 | 88 | 35944 | | | Chromosomal anomalies and genetic disorders | congenital anomalies | 25DTot | |
| 758.1 | | | | | 26801 | 68 | 26733 | | | | | 36013 | 69 | 35944 | | | Chromosomal anomalies | congenital anomalies | 25DTot | |
| 759 | | | | | 27017 | 284 | 26733 | | | | | 36141 | 197 | 35944 | | | Other and unspecified congenital anomalies | congenital anomalies | 25DTot | |
| 759.1 | | | | | 26741 | 8 | 26733 | | | | | 35957 | 13 | 35944 | | | Anomalies of endocrine glands, congenital | congenital anomalies | 25DTot | |
| 760 | 0.6243 | 0.04826 | 1.867 | 2.81e-38 | 24911 | 4715 | 20196 | 0.6631 | 0.03278 | 1.941 | 5.66e-91 | 32596 | 7782 | 24814 | 0.1829 | 0.07699 | Back pain | symptoms | 25DTot | |
| 761 | 0.5134 | 0.06339 | 1.671 | 5.54e-16 | 26084 | 1972 | 24112 | 0.6379 | 0.04004 | 1.893 | 3.84e-57 | 34098 | 3706 | 30392 | 0.03044 | 0.1603 | Cervicalgia | symptoms | 25DTot | |
| 763 | 0.5908 | 0.07673 | 1.806 | 1.35e-14 | 25089 | 1208 | 23881 | 0.525 | 0.05322 | 1.69 | 5.97e-23 | 32717 | 1862 | 30855 | 0.6144 | -0.0465 | Thoracic or lumbosacral neuritis or radiculitis, unspecified | symptoms | 25DTot | |
| 764 | 0.4275 | 0.1012 | 1.533 | 2.41e-05 | 24586 | 705 | 23881 | 0.5886 | 0.06168 | 1.802 | 1.39e-21 | 32170 | 1315 | 30855 | 0.1469 | 0.1699 | Sciatica | symptoms | 25DTot | |
| 765 | 0.456 | 0.1077 | 1.578 | 2.31e-05 | 24494 | 613 | 23881 | 0.548 | 0.07532 | 1.73 | 3.45e-13 | 31724 | 869 | 30855 | 0.453 | 0.09722 | Cervical radiculitis | symptoms | 25DTot | |
| 766 | 0.8134 | 0.09527 | 2.256 | 1.37e-17 | 24563 | 682 | 23881 | 0.6833 | 0.06758 | 1.98 | 4.98e-24 | 31922 | 1067 | 30855 | 0.5351 | -0.07119 | Neuralgia, neuritis, and radiculitis NOS | symptoms | 25DTot | |
| 767 | | | | | 23904 | 23 | 23881 | | | | | 30882 | 27 | 30855 | | | Cervicocranial/Cervicobrachial syndrome | symptoms | 25DTot | |
| 769 | | | | | 27744 | 17 | 27727 | | | | | 36864 | 59 | 36805 | | | Nonallopathic lesions NEC | symptoms | 25DTot | |
| 770 | 1.049 | 0.08954 | 2.854 | 1.12e-31 | 26643 | 686 | 25957 | 0.9026 | 0.04232 | 2.466 | 6.07e-101 | 34609 | 3011 | 31598 | 0.05714 | -0.1846 | Myalgia and myositis unspecified | symptoms | 25DTot | |
| 771 | 0.6889 | 0.1075 | 1.992 | 1.48e-10 | 23704 | 575 | 23129 | 0.3326 | 0.09064 | 1.395 | 2.43e-04 | 31912 | 628 | 31284 | 0.02445 | -0.3091 | Musculoskeletal symptoms referable to limbs | symptoms | 25DTot | |
| 771.1 | 0.6293 | 0.07785 | 1.876 | 6.27e-16 | 24349 | 1220 | 23129 | 0.3768 | 0.06148 | 1.458 | 8.91e-10 | 32708 | 1424 | 31284 | 0.07807 | -0.1712 | Swelling of limb | symptoms | 25DTot | |
| 771.2 | | | | | 23196 | 67 | 23129 | | | | | 31394 | 110 | 31284 | | | Cramp of limb | symptoms | 25DTot | |
| 772 | | | | | 23175 | 134 | 23041 | | | | | 30713 | 257 | 30456 | | | Symptoms of the muscles | symptoms | 25DTot | |
| 772.1 | | | | | 23094 | 53 | 23041 | | | | | 30516 | 60 | 30456 | | | Muscular wasting and disuse atrophy | symptoms | 25DTot | |
| 772.2 | 0.4472 | 0.1084 | 1.564 | 3.68e-05 | 23676 | 635 | 23041 | 0.7134 | 0.06764 | 2.041 | 5.24e-26 | 31533 | 1077 | 30456 | 0.01334 | 0.3108 | Spasm of muscle | symptoms | 25DTot | |
| 772.3 | 0.5605 | 0.06932 | 1.751 | 6.24e-16 | 24642 | 1601 | 23041 | 0.3692 | 0.04977 | 1.447 | 1.19e-13 | 32724 | 2268 | 30456 | 0.03901 | -0.1732 | Muscle weakness | symptoms | 25DTot | |
| 772.4 | | | | | 23175 | 134 | 23041 | | | | | 30527 | 71 | 30456 | | | Rhabdomyolysis | symptoms | 25DTot | |
| 772.6 | | | | | 23142 | 101 | 23041 | | | | | 30584 | 128 | 30456 | | | Facial weakness | symptoms | 25DTot | |
| 780 | | | | | 27429 | 77 | 27352 | | | | | 36521 | 68 | 36453 | | | Hypothermia/Chills | symptoms | 25DTot | |
| 781 | 0.5095 | 0.1107 | 1.664 | 4.21e-06 | 26628 | 527 | 26101 | 0.472 | 0.08289 | 1.603 | 1.24e-08 | 35384 | 686 | 34698 | 0.9517 | -0.008223 | Symptoms involving nervous and musculoskeletal systems | symptoms | 25DTot | |
| 781.1 | | | | | 26112 | 11 | 26101 | | | | | 34735 | 37 | 34698 | | | Loss of height | symptoms | 25DTot | |

| phenotype | male_beta | male_SE | male_OR | male_p | male_n_total | male_n_cases | male_n_controls | female_beta | female_SE | female_OR | female_p | female_n_total | female_n_cases | female_n_controls | interaction_p | interaction_beta | description | group | lab | label |
|---|---|---|---|---|---|---|---|---|---|---|---|---|---|---|---|---|---|---|---|---|
| 781.2 | | | | | 26298 | 197 | 26101 | | | | | 34933 | 235 | 34698 | | | Abnormal posture | symptoms | 25DTot | |
| 782 | | | | | 22002 | 106 | 21896 | | | | | 29294 | 158 | 29136 | | | Symptoms involving skin and other integumentary tissue | symptoms | 25DTot | |
| 782.3 | 0.6507 | 0.05499 | 1.917 | 2.66e-32 | 24907 | 3011 | 21896 | 0.5213 | 0.04003 | 1.684 | 9.07e-39 | 33039 | 3903 | 29136 | 0.2057 | -0.08459 | Edema | symptoms | 25DTot | |
| 782.6 | | | | | 21947 | 51 | 21896 | | | | | 29472 | 336 | 29136 | | | Pallor and flushing | symptoms | 25DTot | |
| 783 | 0.424 | 0.0514 | 1.528 | 1.60e-16 | 24799 | 4188 | 20611 | 0.3037 | 0.04041 | 1.355 | 5.65e-14 | 32997 | 4266 | 28731 | 0.3614 | -0.05835 | Fever of unknown origin | symptoms | 25DTot | |
| 783.1 | | | | | 20666 | 55 | 20611 | | | | | 28768 | 37 | 28731 | | | Postprocedural fever | symptoms | 25DTot | |
| 785 | 0.6573 | 0.04544 | 1.93 | 2.07e-47 | 24378 | 6486 | 17892 | 0.5188 | 0.03109 | 1.68 | 1.56e-62 | 32108 | 11436 | 20672 | 0.06813 | -0.09913 | Abdominal pain | symptoms | 25DTot | |
| 788 | 0.3664 | 0.06376 | 1.443 | 9.10e-09 | 26167 | 2188 | 23979 | 0.2827 | 0.04768 | 1.327 | 3.04e-09 | 34916 | 2566 | 32350 | 0.2136 | -0.09686 | Syncope and collapse | symptoms | 25DTot | |
| 789 | 0.3992 | 0.05203 | 1.491 | 1.69e-14 | 24975 | 4146 | 20829 | 0.2784 | 0.03443 | 1.321 | 6.12e-16 | 32311 | 6758 | 25553 | 0.9942 | 4.42e-04 | Nausea and vomiting | symptoms | 25DTot | |
| 789.1 | | | | | 20853 | 24 | 20829 | | | | | 25611 | 58 | 25553 | | | Persistent vomiting | symptoms | 25DTot | |
| 790 | | | | | 22915 | 183 | 22732 | | | | | 30897 | 312 | 30585 | | | Nonspecific findings on examination of blood | symptoms | 25DTot | |
| 790.1 | | | | | 22812 | 80 | 22732 | | | | | 30855 | 270 | 30585 | | | Elevated sedimentation rate | symptoms | 25DTot | |
| 790.6 | 0.6263 | 0.06005 | 1.871 | 1.79e-25 | 25006 | 2274 | 22732 | 0.6738 | 0.04503 | 1.962 | 1.30e-50 | 33298 | 2713 | 30585 | 0.2049 | 0.09381 | Other abnormal blood chemistry | symptoms | 25DTot | |
| 790.8 | | | | | 22867 | 135 | 22732 | | | | | 30869 | 284 | 30585 | | | Elevated C-reactive protein (CRP) | symptoms | 25DTot | |
| 790.9 | | | | | 22737 | 5 | 22732 | | | | | 30591 | 6 | 30585 | | | Abnormal arterial blood gases | symptoms | 25DTot | |
| 791 | | | | | 27648 | 165 | 27483 | | | | | 36840 | 92 | 36748 | | | Gangrene | symptoms | 25DTot | |
| 792 | | | | | 7 | | | 0.3334 | 0.06459 | 1.396 | 2.45e-07 | 29446 | 1607 | 27839 | 1 | -0.01771 | Abnormal Papanicolaou smear of cervix and cervical HPV | genitourinary | 25DTot | |
| 792.1 | | | | | 7 | | | 0.3113 | 0.07177 | 1.365 | 1.45e-05 | 29183 | 1344 | 27839 | 1 | -0.01891 | Papanicolaou smear of cervix or vagina with atypical squamous cells | genitourinary | 25DTot | |
| 794 | | | | | 27589 | 45 | 27544 | | | | | 36771 | 38 | 36733 | | | Abnormal results of other function studies (bladder, pancreas, placenta, spleen, etc) | symptoms | 25DTot | |
| 795 | | | | | 27272 | 12 | 27260 | | | | | 36232 | 106 | 36126 | | | Other and nonspecific abnormal cytological, histological and immunological findings | symptoms | 25DTot | |
| 795.8 | | | | | 27502 | 242 | 27260 | | | | | 36401 | 275 | 36126 | 0.9135 | -0.03045 | Abnormal tumor markers | symptoms | 25DTot | |
| 795.81 | | | | | 27386 | 126 | 27260 | | | | | 36227 | 101 | 36126 | | | Elevated carcinoembryonic antigen [CEA] | symptoms | 25DTot | |
| 795.82 | | | | | 27261 | 1 | 27260 | | | | | 36221 | 95 | 36126 | | | Elevated cancer antigen 125 [CA 125] | symptoms | 25DTot | |
| 796 | 0.6064 | 0.08143 | 1.834 | 9.59e-14 | 20872 | 1484 | 19388 | | | | | 4 | | | | | Elevated prostate specific antigen [PSA] | genitourinary | 25DTot | |
| 797 | 0.3254 | 0.09762 | 1.385 | 8.57e-04 | 26901 | 1035 | 25866 | 0.0568 | 0.09561 | 1.058 | 0.5525 | 36273 | 725 | 35548 | 0.1502 | -0.1907 | Shock | symptoms | 25DTot | |
| 797.1 | | | | | 26238 | 372 | 25866 | | | | | 35746 | 198 | 35548 | 0.4065 | -0.2279 | Cardiogenic shock | symptoms | 25DTot | |
| 798 | 1.017 | 0.04654 | 2.764 | 8.93e-106 | 23475 | 7649 | 15826 | 0.9607 | 0.03249 | 2.614 | 3.35e-192 | 30980 | 11865 | 19115 | 0.9719 | -0.001989 | Malaise and fatigue | symptoms | 25DTot | |
| 798.1 | | | | | 16119 | 293 | 15826 | 1.431 | 0.08293 | 4.185 | 9.12e-67 | 19839 | 724 | 19115 | 0.3361 | -0.1509 | Chronic fatigue syndrome | symptoms | 25DTot | |
| 800 | 0.1722 | 0.09454 | 1.188 | 0.06849 | 22951 | 1064 | 21887 | 0.428 | 0.05661 | 1.534 | 4.06e-14 | 30522 | 1698 | 28824 | 2.15e-04 | 0.402 | Fracture of lower limb | injuries & poisonings | 25DTot | |
| 800.1 | | | | | 22264 | 377 | 21887 | 0.5016 | 0.09411 | 1.651 | 9.80e-08 | 29394 | 570 | 28824 | 0.2948 | 0.1781 | Fracture of neck of femur | injuries & poisonings | 25DTot | |
| 800.2 | | | | | 22107 | 220 | 21887 | | | | | 29062 | 238 | 28824 | | | Fracture of unspecified part of femur | injuries & poisonings | 25DTot | |

| phenotype | male_beta | male_SE | male_OR | male_p | male_n_total | male_n_cases | male_n_controls | female_beta | female_SE | female_OR | female_p | female_n_total | female_n_cases | female_n_controls | interaction_p | interaction_beta | description | group | lab | label |
|---|---|---|---|---|---|---|---|---|---|---|---|---|---|---|---|---|---|---|---|---|
| 800.3 | | | | | 22240 | 353 | 21887 | | | | | 29291 | 467 | 28824 | 0.02901 | 0.425 | Fracture of tibia and fibula | injuries & poisonings | 25DTot | |
| 800.4 | | | | | 22024 | 137 | 21887 | | | | | 29015 | 191 | 28824 | | | Fracture of patella | injuries & poisonings | 25DTot | |
| 801 | 0.3535 | 0.1183 | 1.424 | 0.002814 | 22470 | 583 | 21887 | 0.4972 | 0.06492 | 1.644 | 1.88e-14 | 30057 | 1233 | 28824 | 0.01028 | 0.3421 | Fracture of ankle and foot | injuries & poisonings | 25DTot | |
| 801.1 | | | | | 22222 | 335 | 21887 | 0.2353 | 0.102 | 1.265 | 0.02105 | 29349 | 525 | 28824 | 0.5104 | 0.1222 | Fracture of foot | injuries & poisonings | 25DTot | |
| 802 | | | | | 22131 | 244 | 21887 | | | | | 29104 | 280 | 28824 | 0.04481 | 0.4786 | Fracture of pelvis | injuries & poisonings | 25DTot | |
| 803 | 0.2518 | 0.09362 | 1.286 | 0.007156 | 22909 | 1022 | 21887 | 0.3393 | 0.05882 | 1.404 | 8.00e-09 | 30429 | 1605 | 28824 | 0.01384 | 0.2679 | Fracture of upper limb | injuries & poisonings | 25DTot | |
| 803.1 | | | | | 22122 | 235 | 21887 | | | | | 29293 | 469 | 28824 | 0.3556 | 0.1913 | Fracture of humerus | injuries & poisonings | 25DTot | |
| 803.2 | | | | | 22365 | 478 | 21887 | 0.27 | 0.07936 | 1.31 | 6.69e-04 | 29684 | 860 | 28824 | 0.004134 | 0.471 | Fracture of radius and ulna | injuries & poisonings | 25DTot | |
| 803.21 | | | | | 21931 | 44 | 21887 | | | | | 28974 | 150 | 28824 | | | Colles' fracture | injuries & poisonings | 25DTot | |
| 803.3 | | | | | 22141 | 254 | 21887 | | | | | 28997 | 173 | 28824 | | | Fracture of clavicle or scapula | injuries & poisonings | 25DTot | |
| 804 | | | | | 22374 | 487 | 21887 | | | | | 29317 | 493 | 28824 | 0.008713 | 0.4438 | Fracture of hand or wrist | injuries & poisonings | 25DTot | |
| 805 | 0.4569 | 0.0971 | 1.579 | 2.53e-06 | 22729 | 842 | 21887 | 0.3458 | 0.07408 | 1.413 | 3.04e-06 | 29817 | 993 | 28824 | 0.8847 | -0.01736 | Fracture of vertebral column without mention of spinal cord injury | injuries & poisonings | 25DTot | |
| 807 | | | | | 22286 | 399 | 21887 | | | | | 29056 | 232 | 28824 | 0.2108 | 0.2807 | Fracture of ribs | injuries & poisonings | 25DTot | |
| 809 | 0.15 | 0.08016 | 1.162 | 0.06139 | 23404 | 1517 | 21887 | 0.3337 | 0.0555 | 1.396 | 1.83e-09 | 30670 | 1846 | 28824 | 0.002174 | 0.2944 | Fracture of unspecified bones | injuries & poisonings | 25DTot | |
| 816 | | | | | 26400 | 43 | 26357 | | | | | 35690 | 21 | 35669 | | | Cerebral laceration and contusion | injuries & poisonings | 25DTot | |
| 817 | | | | | 26422 | 65 | 26357 | | | | | 35734 | 65 | 35669 | | | Concussion | injuries & poisonings | 25DTot | |
| 818 | | | | | 26606 | 249 | 26357 | | | | | 35816 | 147 | 35669 | | | Intracranial hemorrhage (injury) | injuries & poisonings | 25DTot | |
| 818.1 | | | | | 26475 | 118 | 26357 | | | | | 35737 | 68 | 35669 | | | Subdural hemorrhage (injury) | injuries & poisonings | 25DTot | |
| 818.2 | | | | | 26451 | 94 | 26357 | | | | | 35727 | 58 | 35669 | | | Subarachnoid hemorrhage (injury) | injuries & poisonings | 25DTot | |
| 819 | -0.0969 | 0.1466 | 0.9076 | 0.5086 | 26908 | 551 | 26357 | | | | | 36031 | 362 | 35669 | 0.2647 | 0.2178 | Skull and face fracture and other intercranial injury | injuries & poisonings | 25DTot | |
| 823 | | | | | 21319 | | | | | | | 28267 | 2 | 28265 | | | Torus fracture | injuries & poisonings | 25DTot | |
| 830 | 0.1083 | 0.09778 | 1.114 | 0.2679 | 26492 | 928 | 25564 | 0.2938 | 0.06631 | 1.341 | 9.40e-06 | 34859 | 1219 | 33640 | 0.01365 | 0.2861 | Dislocation | injuries & poisonings | 25DTot | |
| 835 | | | | | 26013 | 449 | 25564 | 0.3364 | 0.0784 | 1.4 | 1.78e-05 | 34495 | 855 | 33640 | 0.09667 | 0.255 | Internal derangement of knee | injuries & poisonings | 25DTot | |
| 836 | | | | | 25643 | 79 | 25564 | | | | | 33724 | 84 | 33640 | | | Traumatic arthropathy | injuries & poisonings | 25DTot | |
| 840 | 0.3491 | 0.07689 | 1.418 | 5.62e-06 | 25492 | 1396 | 24096 | 0.4292 | 0.05212 | 1.536 | 1.80e-16 | 33198 | 2125 | 31073 | 0.2641 | 0.1021 | Sprains and strains | injuries & poisonings | 25DTot | |
| 840.1 | | | | | 24130 | 34 | 24096 | | | | | 31116 | 43 | 31073 | | | Muscle/tendon sprain | injuries & poisonings | 25DTot | |
| 840.2 | | | | | 24369 | 273 | 24096 | | | | | 31341 | 268 | 31073 | 0.3589 | 0.1869 | Rotator cuff (capsule) sprain | injuries & poisonings | 25DTot | |
| 840.3 | | | | | 24387 | 291 | 24096 | | | | | 31485 | 412 | 31073 | 0.7906 | 0.0536 | Joint/ligament sprain | injuries & poisonings | 25DTot | |
| 841 | | | | | 24387 | 291 | 24096 | 0.7998 | 0.08552 | 2.225 | 8.63e-21 | 31714 | 641 | 31073 | 0.5203 | 0.1054 | Sprains and strains of back and neck | injuries & poisonings | 25DTot | |

| phenotype | male_beta | male_SE | male_OR | male_p | male_n_total | male_n_cases | male_n_controls | female_beta | female_SE | female_OR | female_p | female_n_total | female_n_cases | female_n_controls | interaction_p | interaction_beta | description | group | lab | label |
|---|---|---|---|---|---|---|---|---|---|---|---|---|---|---|---|---|---|---|---|---|
| 842 | | | | | 24179 | 83 | 24096 | | | | | 31195 | 122 | 31073 | | | Other sprains and strains | injuries & poisonings | 25DTot | |
| 850 | | | | | 23223 | 456 | 22767 | | | | | 32260 | 465 | 31795 | 0.1003 | -0.2843 | Hemorrhage or hematoma complicating a procedure | injuries & poisonings | 25DTot | |
| 851 | | | | | 23157 | 390 | 22767 | | | | | 32086 | 291 | 31795 | 3.50e-04 | -0.5974 | Complications of transplants and reattached limbs | injuries & poisonings | 25DTot | |
| 853 | | | | | 22918 | 151 | 22767 | | | | | 31956 | 161 | 31795 | | | Complication of colostomy or enterostomy | injuries & poisonings | 25DTot | |
| 854 | 0.315 | 0.1136 | 1.37 | 0.00556 | 23435 | 668 | 22767 | 0.1704 | 0.1064 | 1.186 | 0.1092 | 32312 | 517 | 31795 | 0.3199 | -0.1507 | Complications of cardiac/vascular device, implant, and graft | injuries & poisonings | 25DTot | |
| 855 | | | | | 22967 | 200 | 22767 | | | | | 32019 | 224 | 31795 | | | Complication of nervous system device, implant, and graft | injuries & poisonings | 25DTot | |
| 856 | | | | | 22823 | 56 | 22767 | | | | | 31840 | 45 | 31795 | | | Vascular complications of surgery and medical procedures | injuries & poisonings | 25DTot | |
| 857 | | | | | 22852 | 85 | 22767 | | | | | 31886 | 91 | 31795 | | | Mechanical complication of unspecified genitourinary device, implant, and graft | injuries & poisonings | 25DTot | |
| 858 | | | | | 23191 | 424 | 22767 | -0.07456 | 0.09994 | 0.9282 | 0.4556 | 32396 | 601 | 31795 | 0.2087 | -0.2153 | Complication of internal orthopedic device | injuries & poisonings | 25DTot | |
| 859 | | | | | 23165 | 398 | 22767 | | | | | 32227 | 432 | 31795 | 0.2222 | -0.2422 | Complication due to other implant and internal device | injuries & poisonings | 25DTot | |
| 860 | | | | | 27702 | 127 | 27575 | | | | | 36911 | 120 | 36791 | | | Bone marrow or stem cell transplant | neoplasms | 25DTot | |
| 870 | 0.06994 | 0.1179 | 1.072 | 0.553 | 24342 | 736 | 23606 | 0.09605 | 0.09298 | 1.101 | 0.3016 | 33149 | 715 | 32434 | 0.5363 | 0.09095 | Open wounds of head; neck; and trunk | injuries & poisonings | 25DTot | |
| 870.1 | | | | | 23654 | 48 | 23606 | | | | | 32454 | 20 | 32434 | | | Open wound or laceration of eye or eyelid | injuries & poisonings | 25DTot | |
| 870.2 | | | | | 23622 | 16 | 23606 | | | | | 32443 | 9 | 32434 | | | Open wound of ear | injuries & poisonings | 25DTot | |
| 870.3 | | | | | 23791 | 185 | 23606 | | | | | 32553 | 119 | 32434 | | | Other open wound of head and face | injuries & poisonings | 25DTot | |
| 870.4 | | | | | 23626 | 20 | 23606 | | | | | 32447 | 13 | 32434 | | | Open wound of nose and sinus | injuries & poisonings | 25DTot | |
| 870.5 | | | | | 23652 | 46 | 23606 | | | | | 32464 | 30 | 32434 | | | Open wound of lip and mouth | injuries & poisonings | 25DTot | |
| 870.6 | | | | | 23625 | 19 | 23606 | | | | | 32440 | 6 | 32434 | | | Open wound of neck | injuries & poisonings | 25DTot | |
| 870.8 | | | | | 23613 | 7 | 23606 | | | | | 32442 | 8 | 32434 | | | Open wound of genital organs | injuries & poisonings | 25DTot | |
| 871 | 0.2246 | 0.0915 | 1.252 | 0.01413 | 24632 | 1026 | 23606 | 0.401 | 0.07402 | 1.493 | 6.04e-08 | 33363 | 929 | 32434 | 0.01225 | 0.2889 | Open wounds of extremities | injuries & poisonings | 25DTot | |
| 871.1 | | | | | 23768 | 162 | 23606 | | | | | 32546 | 112 | 32434 | | | Open wound of hand except finger(s) | injuries & poisonings | 25DTot | |
| 871.2 | | | | | 23775 | 169 | 23606 | | | | | 32574 | 140 | 32434 | | | Open wound of finger(s) | injuries & poisonings | 25DTot | |
| phenotype | male_beta | male_SE | male_OR | male_p | male_n_total | male_n_cases | male_n_controls | female_beta | female_SE | female_OR | female_p | female_n_total | female_n_cases | female_n_controls | interaction_p | interaction_beta | description | group | lab | label |
| 871.3 | | | | | 23720 | 114 | 23606 | | | | | 32510 | 76 | 32434 | | | Open wound of foot except toe(s) alone | injuries & poisonings | 25DTot | |
| 871.4 | | | | | 23638 | 32 | 23606 | | | | | 32453 | 19 | 32434 | | | Open wound of toe(s) | injuries & poisonings | 25DTot | |
| 872 | | | | | 23730 | 124 | 23606 | | | | | 32497 | 63 | 32434 | | | Traumatic amputation | injuries & poisonings | 25DTot | |
| 874 | | | | | 23727 | 121 | 23606 | | | | | 32492 | 58 | 32434 | | | Complication of amputation stump | injuries & poisonings | 25DTot | |
| 875 | | | | | 23705 | 99 | 23606 | | | | | 32557 | 123 | 32434 | | | Non-healing surgical wound | injuries & poisonings | 25DTot | |

| phenotype | male_beta | male_SE | male_OR | male_p | male_n_total | male_n_cases | male_n_controls | female_beta | female_SE | female_OR | female_p | female_n_total | female_n_cases | female_n_controls | interaction_p | interaction_beta | description | group | lab | label |
|---|---|---|---|---|---|---|---|---|---|---|---|---|---|---|---|---|---|---|---|---|
| 876 | | | | | 23630 | 24 | 23606 | | | | | 32475 | 41 | 32434 | | | Posttraumatic wound infection not elsewhere classified | injuries & poisonings | 25DTot | |
| 905 | | | | | 27300 | 4 | 27296 | | | | | 36496 | 6 | 36490 | | | Late effects of musculoskeletal and connective tissue injuries | injuries & poisonings | 25DTot | |
| 907 | | | | | 27494 | 198 | 27296 | | | | | 36638 | 148 | 36490 | | | Injuries to the nervous system | injuries & poisonings | 25DTot | |
| 910 | | | | | 27755 | 6 | 27749 | | | | | 36903 | 8 | 36895 | | | Superficial injury, infected | injuries & poisonings | 25DTot | |
| 911 | | | | | 27688 | 32 | 27656 | | | | | 36846 | 32 | 36814 | | | Blister | injuries & poisonings | 25DTot | |
| 912 | | | | | 27054 | 122 | 26932 | | | | | 35677 | 201 | 35476 | | | Insect bite | injuries & poisonings | 25DTot | |
| 913 | | | | | 26968 | 36 | 26932 | | | | | 35544 | 68 | 35476 | | | Toxic effect of venom | injuries & poisonings | 25DTot | |
| 916 | 0.4467 | 0.1108 | 1.563 | 5.55e-05 | 26060 | 586 | 25474 | 0.2755 | 0.07115 | 1.317 | 1.08e-04 | 34442 | 1043 | 33399 | 0.6583 | -0.05717 | Contusion | injuries & poisonings | 25DTot | |
| 930 | | | | | 22109 | 280 | 21829 | 0.4796 | 0.09348 | 1.615 | 2.89e-07 | 27961 | 619 | 27342 | 0.8117 | 0.04416 | Allergic reaction to food | injuries & poisonings | 25DTot | |
| 931 | | | | | 21895 | 66 | 21829 | | | | | 27431 | 89 | 27342 | | | Contact dermatitis and other eczema due to plants [except food] | dermatologic | 25DTot | |
| 938 | 0.6046 | 0.0887 | 1.83 | 9.37e-12 | 22767 | 938 | 21829 | 0.7997 | 0.07656 | 2.225 | 1.55e-25 | 28152 | 810 | 27342 | 0.1109 | 0.1846 | Dermatitis due to solar radiation | dermatologic | 25DTot | |
| 938.1 | | | | | 21840 | 11 | 21829 | | | | | 27356 | 14 | 27342 | | | Acute dermatitis due to solar radiation | dermatologic | 25DTot | |
| 938.2 | 0.6561 | 0.09264 | 1.927 | 1.42e-12 | 22665 | 836 | 21829 | 0.8635 | 0.08031 | 2.371 | 5.78e-27 | 28065 | 723 | 27342 | 0.1369 | 0.1802 | Chronic dermatitis due to solar radiation | dermatologic | 25DTot | |
| 939 | 0.561 | 0.07455 | 1.752 | 5.29e-14 | 23219 | 1390 | 21829 | 0.6124 | 0.05069 | 1.845 | 1.33e-33 | 29539 | 2197 | 27342 | 0.8847 | 0.0129 | Atopic/contact dermatitis due to other or unspecified | dermatologic | 25DTot | |
| 939.1 | | | | | 21831 | 2 | 21829 | | | | | 27354 | 12 | 27342 | | | Contact and allergic dermatitis of eyelid | dermatologic | 25DTot | |
| 941 | | | | | 21871 | 42 | 21829 | | | | | 27432 | 90 | 27342 | | | Adverse reaction to serum or vaccine | injuries & poisonings | 25DTot | |
| 942 | | | | | 21868 | 39 | 21829 | | | | | 27392 | 50 | 27342 | | | Infusion and transfusion reaction | injuries & poisonings | 25DTot | |
| 946 | | | | | 21895 | 66 | 21829 | | | | | 27494 | 152 | 27342 | | | Anaphylactic shock NOS | injuries & poisonings | 25DTot | |
| 947 | | | | | 21986 | 157 | 21829 | | | | | 27830 | 488 | 27342 | 0.3941 | 0.1983 | Urticaria | dermatologic | 25DTot | |
| 949 | 0.2568 | 0.1265 | 1.293 | 0.04239 | 22350 | 521 | 21829 | 0.3708 | 0.06853 | 1.449 | 6.26e-08 | 28540 | 1198 | 27342 | 0.48 | 0.1005 | Allergies, other | injuries & poisonings | 25DTot | |
| 949.1 | | | | | 21836 | 7 | 21829 | | | | | 27350 | 8 | 27342 | | | Diaper or napkin rash | injuries & poisonings | 25DTot | |
| 952 | | | | | 27725 | 66 | 27659 | | | | | 36923 | 32 | 36891 | | | Spinal cord injury without evidence of spinal bone injury | injuries & poisonings | 25DTot | |
| 957 | | | | | 27659 | | | | | | | 36893 | 2 | 36891 | | | Injury to other and unspecified nerves | injuries & poisonings | 25DTot | |
| 958 | | | | | 27212 | 374 | 26838 | | | | | 36518 | 232 | 36286 | 0.7986 | 0.06139 | Certain early complications of trauma or procedure | injuries & poisonings | 25DTot | |
| 958.1 | | | | | 26905 | 67 | 26838 | | | | | 36331 | 45 | 36286 | | | Postoperative shock | injuries & poisonings | 25DTot | |
| 958.2 | | | | | 26974 | 136 | 26838 | | | | | 36364 | 78 | 36286 | | | Traumatic and surgical subcutaneous emphysema | injuries & poisonings | 25DTot | |
| 960 | 0.5818 | 0.07859 | 1.789 | 1.33e-13 | 21000 | 1222 | 19778 | 0.2517 | 0.05279 | 1.286 | 1.87e-06 | 28211 | 2139 | 26072 | 0.008157 | -0.2474 | Poisoning by antibiotics | injuries & poisonings | 25DTot | |
| 960.1 | | | | | 19779 | 1 | 19778 | | | | | 26073 | 1 | 26072 | | | Adverse effects of antibacterials (not penicillins) | injuries & poisonings | 25DTot | |
| 960.2 | 0.4954 | 0.104 | 1.641 | 1.91e-06 | 20476 | 698 | 19778 | 0.274 | 0.06792 | 1.315 | 5.49e-05 | 27268 | 1196 | 26072 | 0.2716 | -0.1346 | Allergy/adverse effect of penicillin | injuries & poisonings | 25DTot | |

| phenotype | male_beta | male_SE | male_OR | male_p | male_n_total | male_n_cases | male_n_controls | female_beta | female_SE | female_OR | female_p | female_n_total | female_n_cases | female_n_controls | interaction_p | interaction_beta | description | group | lab | label |
|---|---|---|---|---|---|---|---|---|---|---|---|---|---|---|---|---|---|---|---|---|
| 960.3 | | | | | 19779 | 1 | 19778 | | | | | 26073 | 1 | 26072 | | | Poisoning by antifungal antibiotics | injuries & poisonings | 25DTot | |
| 961 | | | | | 19843 | 65 | 19778 | | | | | 26198 | 126 | 26072 | | | Poisoning by other anti-infectives | injuries & poisonings | 25DTot | |
| 961.1 | | | | | 20163 | 385 | 19778 | 0.3249 | 0.07013 | 1.384 | 3.61e-06 | 27149 | 1077 | 26072 | 0.3972 | -0.1274 | Poisoning/allergy of sulfonamides | injuries & poisonings | 25DTot | |
| 962 | | | | | 19885 | 107 | 19778 | | | | | 26193 | 121 | 26072 | | | Poisoning by hormones and synthetic substitutes | injuries & poisonings | 25DTot | |
| 962.1 | | | | | 19862 | 84 | 19778 | | | | | 26166 | 94 | 26072 | | | Adrenal cortical steroids causing adverse effects in therapeutic use | injuries & poisonings | 25DTot | |
| 962.2 | | | | | 19800 | 22 | 19778 | | | | | 26094 | 22 | 26072 | | | Insulins and antidiabetic agents causing adverse effects in therapeutic use | injuries & poisonings | 25DTot | |
| 962.3 | | | | | 19778 | | | | | | | 26078 | 6 | 26072 | | | Hormones and synthetic substitutes causing adverse effects in therapeutic use | injuries & poisonings | 25DTot | |
| 963 | 0.06868 | 0.1519 | 1.071 | 0.6512 | 20349 | 571 | 19778 | -0.05881 | 0.119 | 0.9429 | 0.6213 | 26631 | 559 | 26072 | 0.8542 | -0.03484 | Poisoning by primarily systemic agents | injuries & poisonings | 25DTot | |
| 963.1 | 0.06641 | 0.1534 | 1.069 | 0.6651 | 20340 | 562 | 19778 | -0.09273 | 0.1219 | 0.9114 | 0.447 | 26617 | 545 | 26072 | 0.7184 | -0.06943 | Antineoplastic and immunosuppressive drugs causing adverse effects | injuries & poisonings | 25DTot | |
| 964 | | | | | 19833 | 55 | 19778 | | | | | 26136 | 64 | 26072 | | | Poisoning by agents primarily affecting blood constituents | injuries & poisonings | 25DTot | |
| 964.1 | | | | | 19833 | 55 | 19778 | | | | | 26131 | 59 | 26072 | | | Anticoagulants causing adverse effects | injuries & poisonings | 25DTot | |
| 965 | | | | | 20187 | 409 | 19778 | 0.3299 | 0.08161 | 1.391 | 5.28e-05 | 26882 | 810 | 26072 | 0.4815 | -0.1083 | Poisoning by analgesics, antipyretics, and antirheumatics | injuries & poisonings | 25DTot | |
| 965.1 | 0.5008 | 0.1119 | 1.65 | 7.67e-06 | 20350 | 572 | 19778 | 0.2045 | 0.06537 | 1.227 | 0.001758 | 27386 | 1314 | 26072 | 0.05979 | -0.2412 | Opiates and related narcotics causing adverse effects in therapeutic use | injuries & poisonings | 25DTot | |
| 965.2 | | | | | 19780 | 2 | 19778 | | | | | 26073 | 1 | 26072 | | | Antirheumatics causing adverse effects in therapeutic use | injuries & poisonings | 25DTot | |
| 965.3 | | | | | 19783 | 5 | 19778 | | | | | 26085 | 13 | 26072 | | | Salicylates causing adverse effects in therapeutic use | injuries & poisonings | 25DTot | |
| 966 | | | | | 19797 | 19 | 19778 | | | | | 26089 | 17 | 26072 | | | Poisoning by anticonvulsants and anti-Parkinsonism drugs | injuries & poisonings | 25DTot | |
| 967 | | | | | 19831 | 53 | 19778 | | | | | 26222 | 150 | 26072 | | | Adverse effects of sedatives or other central nervous system depressants and anesthetics | injuries & poisonings | 25DTot | |
| 969 | | | | | 19838 | 60 | 19778 | | | | | 26169 | 97 | 26072 | | | Poisoning by psychotropic agents | injuries & poisonings | 25DTot | |
| 971 | | | | | 19780 | 2 | 19778 | | | | | 26075 | 3 | 26072 | | | Poisoning by drugs primarily affecting the autonomic nervous system | injuries & poisonings | 25DTot | |
| 972 | | | | | 19849 | 71 | 19778 | | | | | 26139 | 67 | 26072 | | | Poisoning by agents primarily affecting the cardiovascular system | injuries & poisonings | 25DTot | |
| 972.1 | | | | | 19792 | 14 | 19778 | | | | | 26083 | 11 | 26072 | | | Cardiac rhythm regulators causing adverse effects in therapeutic use | injuries & poisonings | 25DTot | |
| 972.2 | | | | | 19778 | | | | | | | 26072 | | | | | Antilipemic and antiarteriosclerotic drugs causing adverse effects in therapeutic use | injuries & poisonings | 25DTot | |
| 972.6 | | | | | 19783 | 5 | 19778 | | | | | 26077 | 5 | 26072 | | | Antihypertensive agents causing adverse effects | injuries & poisonings | 25DTot | |
| 973 | | | | | 19785 | 7 | 19778 | | | | | 26074 | 2 | 26072 | | | Poisoning by agents primarily affecting the gastrointestinal system | injuries & poisonings | 25DTot | |
| 974 | | | | | 19842 | 64 | 19778 | | | | | 26094 | 22 | 26072 | | | Poisoning by water, mineral, and uric acid metabolism drugs | injuries & poisonings | 25DTot | |

| phenotype | male_beta | male_SE | male_OR | male_p | male_n_total | male_n_cases | male_n_controls | female_beta | female_SE | female_OR | female_p | female_n_total | female_n_cases | female_n_controls | interaction_p | interaction_beta | description | group | lab | label |
|---|---|---|---|---|---|---|---|---|---|---|---|---|---|---|---|---|---|---|---|---|
| 975 | | | | | 19784 | 6 | 19778 | | | | | 26079 | 7 | 26072 | | | Poisoning by agents primarily acting on the smooth and skeletal muscles and respiratory system | injuries & poisonings | 25DTot | |
| phenotype | male_beta | male_SE | male_OR | male_p | male_n_total | male_n_cases | male_n_controls | female_beta | female_SE | female_OR | female_p | female_n_total | female_n_cases | female_n_controls | interaction_p | interaction_beta | description | group | lab | label |
| 976 | | | | | 19780 | 2 | 19778 | | | | | 26075 | 3 | 26072 | | | Poisoning by agents primarily affecting skin & mucous membrane, ophthalmological, otorhinolaryngological, & dental drugs | injuries & poisonings | 25DTot | |
| 977 | 0.5827 | 0.07359 | 1.791 | 2.42e-15 | 21166 | 1388 | 19778 | 0.338 | 0.05119 | 1.402 | 4.04e-11 | 28281 | 2209 | 26072 | 0.01351 | -0.219 | Personal history of allergy to medicinal agents | injuries & poisonings | 25DTot | |
| 979 | 0.3511 | 0.103 | 1.421 | 6.51e-04 | 20676 | 898 | 19778 | 0.2454 | 0.07892 | 1.278 | 0.001878 | 27006 | 934 | 26072 | 0.7584 | 0.03909 | Adverse drug events and drug allergies | injuries & poisonings | 25DTot | |
| 980 | 0.4429 | 0.08365 | 1.557 | 1.19e-07 | 26597 | 1176 | 25421 | 0.3325 | 0.07477 | 1.394 | 8.72e-06 | 35862 | 1000 | 34862 | 0.7346 | -0.03702 | Encounter for long-term (current) use of antibiotics | infectious diseases | 25DTot | |
| 981 | | | | | 27615 | 2 | 27613 | | | | | 36741 | 3 | 36738 | | | Toxic effect of (non-ethyl) alcohol and petroleum and other solvents | injuries & poisonings | 25DTot | |
| 983 | | | | | 27616 | 3 | 27613 | | | | | 36738 | | | | | Toxic effect of corrosive aromatics, acids, and caustic alkalis | injuries & poisonings | 25DTot | |
| 984 | | | | | 27616 | 3 | 27613 | | | | | 36739 | 1 | 36738 | | | Toxic effect of lead and its compounds (including fumes) | injuries & poisonings | 25DTot | |
| 985 | | | | | 27622 | 9 | 27613 | | | | | 36745 | 7 | 36738 | | | Toxic effect of other metals | injuries & poisonings | 25DTot | |
| 986 | | | | | 27616 | 3 | 27613 | | | | | 36740 | 2 | 36738 | | | Toxic effect of carbon monoxide | injuries & poisonings | 25DTot | |
| 987 | | | | | 27617 | 4 | 27613 | | | | | 36745 | 7 | 36738 | | | Toxic effect of other gases, fumes, or vapors | injuries & poisonings | 25DTot | |
| 988 | | | | | 27613 | | | | | | | 36740 | 2 | 36738 | | | Toxic effect of noxious substances eaten as food | injuries & poisonings | 25DTot | |
| 989 | | | | | 27632 | 19 | 27613 | | | | | 36758 | 20 | 36738 | | | Toxic effect of other substances, chiefly nonmedicinal as to source | injuries & poisonings | 25DTot | |
| 990 | -0.3144 | 0.1299 | 0.7302 | 0.01551 | 23446 | 892 | 22554 | -0.4348 | 0.1032 | 0.6474 | 2.50e-05 | 33242 | 782 | 32460 | 0.6861 | -0.06587 | Effects radiation NOS | injuries & poisonings | 25DTot | |
| 994 | 0.3971 | 0.06197 | 1.487 | 1.47e-10 | 26534 | 2690 | 23844 | 0.1533 | 0.05633 | 1.166 | 0.006498 | 35853 | 2103 | 33750 | 0.03335 | -0.173 | Sepsis and SIRS | injuries & poisonings | 25DTot | |
| 994.1 | 0.4621 | 0.1296 | 1.587 | 3.65e-04 | 24423 | 579 | 23844 | | | | | 34199 | 449 | 33750 | 0.1333 | -0.2644 | Systemic inflammatory response syndrome (SIRS) | injuries & poisonings | 25DTot | |
| 994.2 | 0.4232 | 0.06468 | 1.527 | 6.03e-11 | 26190 | 2346 | 23844 | 0.192 | 0.05857 | 1.212 | 0.001048 | 35609 | 1859 | 33750 | 0.05578 | -0.1621 | Sepsis | injuries & poisonings | 25DTot | |
| 994.21 | 0.5886 | 0.0986 | 1.802 | 2.37e-09 | 24740 | 896 | 23844 | 0.2164 | 0.09615 | 1.242 | 0.02438 | 34424 | 674 | 33750 | 0.05204 | -0.2588 | Septic shock | injuries & poisonings | 25DTot | |
| 008 | 0.6885 | 0.1011 | 1.991 | 9.92e-12 | 27060 | 574 | 26486 | 0.808 | 0.092 | 2.244 | 1.59e-18 | 35831 | 704 | 35127 | 0.08026 | 0.2178 | Intestinal infection | infectious diseases | LDL-C | |
| 008.5 | | | | | 26875 | 389 | 26486 | | | | | 35533 | 406 | 35127 | 0.2782 | 0.1627 | Bacterial enteritis | infectious diseases | LDL-C | |
| 008.51 | | | | | 26487 | 1 | 26486 | | | | | 35130 | 3 | 35127 | | | Intestinal e.coli | infectious diseases | LDL-C | |
| 008.52 | | | | | 26824 | 338 | 26486 | | | | | 35475 | 348 | 35127 | 0.2807 | 0.1716 | Intestinal infection due to C. difficile | infectious diseases | LDL-C | |
| 008.6 | | | | | 26592 | 106 | 26486 | | | | | 35263 | 136 | 35127 | | | Viral Enteritis | infectious diseases | LDL-C | |
| 008.7 | | | | | 26495 | 9 | 26486 | | | | | 35133 | 6 | 35127 | | | Intestinal infection due to protozoa | infectious diseases | LDL-C | |
| 010 | | | | | 21692 | 51 | 21641 | | | | | 30214 | 54 | 30160 | | | Tuberculosis | infectious diseases | LDL-C | |
| 031 | | | | | 21701 | 60 | 21641 | | | | | 30222 | 62 | 30160 | | | Diseases due to other mycobacteria | infectious diseases | LDL-C | |

| phenotype | male_beta | male_SE | male_OR | male_p | male_n_total | male_n_cases | male_n_controls | female_beta | female_SE | female_OR | female_p | female_n_total | female_n_cases | female_n_controls | interaction_p | interaction_beta | description | group | lab | label |
|---|---|---|---|---|---|---|---|---|---|---|---|---|---|---|---|---|---|---|---|---|
| 031.1 | | | | | 21641 | | | | | | | 30160 | | | | | Leprosy | infectious diseases | LDL-C | |
| 038 | 0.3814 | 0.04412 | 1.464 | 5.41e-18 | 24701 | 3060 | 21641 | 0.3193 | 0.04716 | 1.376 | 1.29e-11 | 32656 | 2496 | 30160 | 0.6885 | 0.0232 | Septicemia | infectious diseases | LDL-C | |
| 038.1 | | | | | 22136 | 495 | 21641 | | | | | 30550 | 390 | 30160 | 0.4617 | 0.1086 | Gram negative septicemia | infectious diseases | LDL-C | |
| 038.2 | | | | | 22055 | 414 | 21641 | | | | | 30433 | 273 | 30160 | 0.1402 | -0.2445 | Gram positive septicemia | infectious diseases | LDL-C | |
| 038.3 | 0.5934 | 0.06462 | 1.81 | 4.20e-20 | 23021 | 1380 | 21641 | 0.505 | 0.07278 | 1.657 | 3.96e-12 | 31182 | 1022 | 30160 | 0.6522 | -0.0395 | Bacteremia | infectious diseases | LDL-C | |
| 041 | 0.3075 | 0.04434 | 1.36 | 4.03e-12 | 24639 | 2998 | 21641 | 0.3437 | 0.04262 | 1.41 | 7.40e-16 | 33323 | 3163 | 30160 | 0.007246 | 0.1489 | Bacterial infection NOS | infectious diseases | LDL-C | |
| 041.1 | 0.24 | 0.06307 | 1.271 | 1.42e-04 | 22993 | 1352 | 21641 | 0.3208 | 0.069 | 1.378 | 3.34e-06 | 31259 | 1099 | 30160 | 0.2118 | 0.1054 | Staphylococcus infections | infectious diseases | LDL-C | |
| 041.11 | 0.2295 | 0.09346 | 1.258 | 0.01407 | 22231 | 590 | 21641 | | | | | 30598 | 438 | 30160 | 0.06619 | 0.2385 | Methicillin sensitive Staphylococcus aureus | infectious diseases | LDL-C | |
| 041.12 | 0.2599 | 0.094 | 1.297 | 0.005696 | 22233 | 592 | 21641 | | | | | 30654 | 494 | 30160 | 0.6847 | 0.05089 | Methicillin resistant Staphylococcus aureus | infectious diseases | LDL-C | |
| 041.2 | 0.3907 | 0.08857 | 1.478 | 1.03e-05 | 22331 | 690 | 21641 | 0.4637 | 0.08926 | 1.59 | 2.05e-07 | 30829 | 669 | 30160 | 0.2437 | 0.1327 | Streptococcus infection | infectious diseases | LDL-C | |
| 041.21 | | | | | 21646 | 5 | 21641 | | | | | 30170 | 10 | 30160 | | | Rheumatic fever / chorea | infectious diseases | LDL-C | |
| 041.4 | | | | | 22126 | 485 | 21641 | 0.5622 | 0.08001 | 1.754 | 2.12e-12 | 31066 | 906 | 30160 | 0.01173 | 0.3049 | E. coli | infectious diseases | LDL-C | |
| 041.8 | | | | | 21691 | 50 | 21641 | | | | | 30238 | 78 | 30160 | | | H. pylori | infectious diseases | LDL-C | |
| 041.9 | | | | | 21990 | 349 | 21641 | | | | | 30526 | 366 | 30160 | 0.13 | 0.2396 | Infection with drug-resistant microorganisms | infectious diseases | LDL-C | |
| 053 | | | | | 22267 | 352 | 21915 | 1.58 | 0.1379 | 4.853 | 2.15e-30 | 29621 | 557 | 29064 | 0.1823 | 0.2743 | Herpes zoster | infectious diseases | LDL-C | |
| 053.1 | | | | | 21986 | 71 | 21915 | | | | | 29192 | 128 | 29064 | | | Herpes zoster with nervous system complications | infectious diseases | LDL-C | |
| 054 | | | | | 22206 | 291 | 21915 | 1.291 | 0.1089 | 3.635 | 2.17e-32 | 29622 | 558 | 29064 | 0.001152 | -0.6974 | Herpes simplex | infectious diseases | LDL-C | |
| 070 | 0.2638 | 0.07057 | 1.302 | 1.85e-04 | 22942 | 1027 | 21915 | 0.3089 | 0.09092 | 1.362 | 6.79e-04 | 29665 | 601 | 29064 | 0.1944 | 0.136 | Viral hepatitis | infectious diseases | LDL-C | |
| 070.1 | | | | | 21931 | 16 | 21915 | | | | | 29069 | 5 | 29064 | | | Viral hepatitis A | infectious diseases | LDL-C | |
| 070.2 | | | | | 22087 | 172 | 21915 | | | | | 29138 | 74 | 29064 | | | Viral hepatitis B | infectious diseases | LDL-C | |
| 070.3 | 0.1927 | 0.07521 | 1.212 | 0.01042 | 22806 | 891 | 21915 | 0.2754 | 0.09783 | 1.317 | 0.00488 | 29579 | 515 | 29064 | 0.1595 | 0.1582 | Viral hepatitis C | infectious diseases | LDL-C | |
| 070.4 | | | | | 22042 | 127 | 21915 | | | | | 29294 | 230 | 29064 | | | Chronic hepatitis | infectious diseases | LDL-C | |
| 070.9 | | | | | 22131 | 216 | 21915 | | | | | 29270 | 206 | 29064 | | | Hepatitis NOS | infectious diseases | LDL-C | |
| 071 | 2.398 | 0.1355 | 11 | 4.87e-70 | 22529 | 614 | 21915 | | | | | 29177 | 113 | 29064 | 0.9409 | 0.02327 | Human immunodeficiency virus [HIV] disease | infectious diseases | LDL-C | |
| 071.1 | 2.437 | 0.1381 | 11.44 | 1.21e-69 | 22521 | 606 | 21915 | | | | | 29170 | 106 | 29064 | 0.9904 | -0.003913 | HIV infection, symptomatic | infectious diseases | LDL-C | |
| 078 | 1.813 | 0.1174 | 6.126 | 8.55e-54 | 22638 | 723 | 21915 | 1.429 | 0.09823 | 4.174 | 6.10e-48 | 29855 | 791 | 29064 | 0.1001 | -0.2385 | Viral warts & HPV | infectious diseases | LDL-C | |
| 079 | 1.416 | 0.1022 | 4.119 | 1.13e-43 | 22615 | 700 | 21915 | 1.151 | 0.07958 | 3.162 | 2.01e-47 | 30057 | 993 | 29064 | 0.1426 | -0.1762 | Viral infection | infectious diseases | LDL-C | |
| 079.1 | | | | | 21924 | 9 | 21915 | | | | | 29075 | 11 | 29064 | | | Varicella infection | infectious diseases | LDL-C | |
| 079.2 | | | | | 21970 | 55 | 21915 | | | | | 29142 | 78 | 29064 | | | Infectious mononucleosis | infectious diseases | LDL-C | |

| phenotype | male_beta | male_SE | male_OR | male_p | male_n_total | male_n_cases | male_n_controls | female_beta | female_SE | female_OR | female_p | female_n_total | female_n_cases | female_n_controls | interaction_p | interaction_beta | description | group | lab | label |
|---|---|---|---|---|---|---|---|---|---|---|---|---|---|---|---|---|---|---|---|---|
| 079.9 | | | | | 21977 | 62 | 21915 | | | | | 29164 | 100 | 29064 | | | Viremia, NOS | infectious diseases | LDL-C | |
| 080 | -0.3667 | 0.08692 | 0.693 | 2.46e-05 | 26108 | 681 | 25427 | -0.1626 | 0.08267 | 0.8499 | 0.04922 | 35329 | 748 | 34581 | 0.009125 | 0.2812 | Postoperative infection | infectious diseases | LDL-C | |
| 081 | -0.2798 | 0.0814 | 0.756 | 5.88e-04 | 26193 | 766 | 25427 | -0.05655 | 0.08409 | 0.945 | 0.5013 | 35303 | 722 | 34581 | 0.001944 | 0.3263 | Infection/inflammation of internal prosthetic device; implant; and graft | infectious diseases | LDL-C | |
| 081.1 | | | | | 25468 | 41 | 25427 | | | | | 34621 | 40 | 34581 | | | Graft-versus-host disease | infectious diseases | LDL-C | |
| 081.11 | | | | | 25439 | 12 | 25427 | | | | | 34597 | 16 | 34581 | | | Acute graft-versus-host disease | infectious diseases | LDL-C | |
| 081.12 | | | | | 25458 | 31 | 25427 | | | | | 34611 | 30 | 34581 | | | Chronic graft-versus-host disease | infectious diseases | LDL-C | |
| phenotype | male_beta | male_SE | male_OR | male_p | male_n_total | male_n_cases | male_n_controls | female_beta | female_SE | female_OR | female_p | female_n_total | female_n_cases | female_n_controls | interaction_p | interaction_beta | description | group | lab | label |
| 090 | | | | | 27678 | 201 | 27477 | | | | | 36786 | 64 | 36722 | | | Sexually transmitted infections (not HIV or hepatitis) | infectious diseases | LDL-C | |
| 090.2 | | | | | 27533 | 56 | 27477 | | | | | 36735 | 13 | 36722 | | | Gonococcal infections | infectious diseases | LDL-C | |
| 090.3 | | | | | 27479 | 2 | 27477 | | | | | 36731 | 9 | 36722 | | | Venereal diseases due to Chlamydia trachomatis | infectious diseases | LDL-C | |
| 110 | 1.914 | 0.107 | 6.783 | 1.44e-71 | 24822 | 1390 | 23432 | 1.897 | 0.1074 | 6.664 | 9.05e-70 | 31711 | 1224 | 30487 | 0.9496 | -0.009339 | Dermatophytosis / Dermatomycosis | infectious diseases | LDL-C | |
| 110.1 | 1.946 | 0.1123 | 6.999 | 2.85e-67 | 24733 | 1301 | 23432 | 2.007 | 0.1167 | 7.441 | 2.65e-66 | 31638 | 1151 | 30487 | 0.6354 | 0.0749 | Dermatophytosis | infectious diseases | LDL-C | |
| 110.11 | 2.313 | 0.1505 | 10.11 | 2.80e-53 | 24428 | 996 | 23432 | 2.229 | 0.1455 | 9.289 | 5.63e-53 | 31407 | 920 | 30487 | 0.754 | -0.06431 | Dermatophytosis of nail | infectious diseases | LDL-C | |
| 110.12 | | | | | 23675 | 243 | 23432 | | | | | 30615 | 128 | 30487 | | | Althete's foot | infectious diseases | LDL-C | |
| 110.13 | | | | | 23580 | 148 | 23432 | | | | | 30581 | 94 | 30487 | | | Dermatophytosis of the body | infectious diseases | LDL-C | |
| 110.2 | | | | | 23511 | 79 | 23432 | | | | | 30547 | 60 | 30487 | | | Dermatomycoses | infectious diseases | LDL-C | |
| 112 | 0.6305 | 0.08247 | 1.879 | 2.09e-14 | 24237 | 805 | 23432 | 0.6861 | 0.05958 | 1.986 | 1.09e-30 | 32100 | 1613 | 30487 | 0.008329 | 0.2467 | Candidiasis | infectious diseases | LDL-C | |
| 112.3 | | | | | 23511 | 79 | 23432 | | | | | 30667 | 180 | 30487 | | | Candidiasis of skin and nails | infectious diseases | LDL-C | |
| 117 | | | | | 23791 | 359 | 23432 | | | | | 30831 | 344 | 30487 | 0.6779 | 0.06478 | Mycoses | infectious diseases | LDL-C | |
| 117.1 | | | | | 23530 | 98 | 23432 | | | | | 30601 | 114 | 30487 | | | Histoplasmosis | infectious diseases | LDL-C | |
| 117.2 | | | | | 23434 | 2 | 23432 | | | | | 30487 | | | | | Coccidioidomycosis | infectious diseases | LDL-C | |
| 117.3 | | | | | 23448 | 16 | 23432 | | | | | 30500 | 13 | 30487 | | | Blastomycotic infection | infectious diseases | LDL-C | |
| 117.4 | | | | | 23501 | 69 | 23432 | | | | | 30544 | 57 | 30487 | | | Aspergillosis | infectious diseases | LDL-C | |
| 130 | | | | | 26354 | 10 | 26344 | | | | | 35233 | 9 | 35224 | | | Spirochetal infection | infectious diseases | LDL-C | |
| 130.1 | | | | | 26349 | 5 | 26344 | | | | | 35227 | 3 | 35224 | | | Lyme disease | infectious diseases | LDL-C | |
| 131 | | | | | 26353 | 9 | 26344 | | | | | 35256 | 32 | 35224 | | | Protozoan infection | infectious diseases | LDL-C | |
| 132 | | | | | 26380 | 36 | 26344 | | | | | 35274 | 50 | 35224 | | | Infestation (lice, mites) | infectious diseases | LDL-C | |
| 132.1 | | | | | 26348 | 4 | 26344 | | | | | 35245 | 21 | 35224 | | | Pediculosis and phthirus infestation | infectious diseases | LDL-C | |
| 133 | | | | | 26352 | 8 | 26344 | | | | | 35226 | 2 | 35224 | | | Arthropod-borne diseases | infectious diseases | LDL-C | |

| phenotype | male_beta | male_SE | male_OR | male_p | male_n_total | male_n_cases | male_n_controls | female_beta | female_SE | female_OR | female_p | female_n_total | female_n_cases | female_n_controls | interaction_p | interaction_beta | description | group | lab | label |
|---|---|---|---|---|---|---|---|---|---|---|---|---|---|---|---|---|---|---|---|---|
| 134 | | | | | 26349 | 5 | 26344 | | | | | 35248 | 24 | 35224 | | | Helminthiases | infectious diseases | LDL-C | |
| 134.1 | | | | | 26347 | 3 | 26344 | | | | | 35236 | 12 | 35224 | | | Intestinal helminthiases | infectious diseases | LDL-C | |
| 136 | | | | | 26733 | 389 | 26344 | | | | | 35599 | 375 | 35224 | 0.4453 | 0.1293 | Other infectious and parasitic diseases | infectious diseases | LDL-C | |
| 145 | -0.7999 | 0.09469 | 0.4494 | 2.97e-17 | 26903 | 624 | 26279 | | | | | 36280 | 243 | 36037 | 0.5524 | -0.09775 | Cancer of mouth | neoplasms | LDL-C | |
| 145.1 | | | | | 26299 | 20 | 26279 | | | | | 36044 | 7 | 36037 | | | Cancer of lip | neoplasms | LDL-C | |
| 145.2 | | | | | 26660 | 381 | 26279 | | | | | 36136 | 99 | 36037 | | | Cancer of tongue | neoplasms | LDL-C | |
| 145.3 | | | | | 26384 | 105 | 26279 | | | | | 36087 | 50 | 36037 | | | Cancer of major salivary glands | neoplasms | LDL-C | |
| 145.4 | | | | | 26326 | 47 | 26279 | | | | | 36068 | 31 | 36037 | | | Cancer of the gums | neoplasms | LDL-C | |
| 145.5 | | | | | 26338 | 59 | 26279 | | | | | 36058 | 21 | 36037 | | | Cancer of the mouth floor | neoplasms | LDL-C | |
| 149 | -0.7824 | 0.0784 | 0.4573 | 1.87e-23 | 27197 | 918 | 26279 | | | | | 36384 | 347 | 36037 | 0.07209 | -0.2535 | Cancer of larynx, pharynx, nasal cavities | neoplasms | LDL-C | |
| 149.1 | | | | | 26708 | 429 | 26279 | | | | | 36147 | 110 | 36037 | 0.5485 | 0.1338 | Cancer of oropharynx | neoplasms | LDL-C | |
| 149.2 | | | | | 26347 | 68 | 26279 | | | | | 36075 | 38 | 36037 | | | Cancer of nasopharynx | neoplasms | LDL-C | |
| 149.3 | | | | | 26333 | 54 | 26279 | | | | | 36064 | 27 | 36037 | | | Cancer of hypopharynx | neoplasms | LDL-C | |
| 149.4 | | | | | 26607 | 328 | 26279 | | | | | 36132 | 95 | 36037 | | | Cancer of larynx | neoplasms | LDL-C | |
| 149.5 | | | | | 26723 | 444 | 26279 | | | | | 36197 | 160 | 36037 | 0.3823 | -0.1696 | Hx of malignant neoplasm of oral cavity and pharynx | neoplasms | LDL-C | |
| 149.9 | | | | | 26377 | 98 | 26279 | | | | | 36109 | 72 | 36037 | | | Cancer of of nasal cavities | neoplasms | LDL-C | |
| 150 | | | | | 23056 | 399 | 22657 | | | | | 32735 | 70 | 32665 | | | Cancer of esophagus | neoplasms | LDL-C | |
| 151 | | | | | 23072 | 415 | 22657 | | | | | 32805 | 140 | 32665 | 0.08067 | 0.3747 | Cancer of stomach | neoplasms | LDL-C | |
| 153 | 0.1673 | 0.06331 | 1.182 | 0.00824 | 21472 | 1270 | 20202 | -0.2452 | 0.0745 | 0.7826 | 9.99e-04 | 30905 | 948 | 29957 | 8.88e-05 | -0.35 | Colorectal cancer | neoplasms | LDL-C | |
| 153.2 | -0.1983 | 0.07597 | 0.8201 | 0.009047 | 21102 | 900 | 20202 | -0.1469 | 0.08158 | 0.8634 | 0.07174 | 30731 | 774 | 29957 | 0.4576 | 0.07552 | Colon cancer | neoplasms | LDL-C | |
| 153.3 | 0.3572 | 0.08124 | 1.429 | 1.10e-05 | 20940 | 738 | 20202 | | | | | 30403 | 446 | 29957 | 1.41e-09 | -0.7669 | Malignant neoplasm of rectum, rectosigmoid junction, and anus | neoplasms | LDL-C | |
| 155 | | | | | 23097 | 440 | 22657 | | | | | 32892 | 227 | 32665 | 0.06715 | 0.3034 | Cancer of liver and intrahepatic bile duct | neoplasms | LDL-C | |
| 155.1 | | | | | 23033 | 376 | 22657 | | | | | 32838 | 173 | 32665 | 0.02165 | 0.4255 | Malignant neoplasm of liver, primary | neoplasms | LDL-C | |
| 157 | | | | | 23046 | 389 | 22657 | | | | | 32990 | 325 | 32665 | 0.3944 | 0.1408 | Pancreatic cancer | neoplasms | LDL-C | |
| 158 | | | | | 22882 | 225 | 22657 | | | | | 32904 | 239 | 32665 | | | Neoplasm of unspecified nature of digestive system | neoplasms | LDL-C | |
| 159 | -0.579 | 0.102 | 0.5604 | 1.38e-08 | 23177 | 520 | 22657 | -0.4559 | 0.1042 | 0.6339 | 1.21e-05 | 33167 | 502 | 32665 | 0.1853 | 0.1737 | Malignant neoplasm of other and ill-defined sites within the digestive organs and peritoneum | neoplasms | LDL-C | |
| 159.2 | | | | | 22805 | 148 | 22657 | | | | | 32791 | 126 | 32665 | | | Malignant neoplasm of small intestine, including duodenum | neoplasms | LDL-C | |
| 159.3 | | | | | 22758 | 101 | 22657 | | | | | 32760 | 95 | 32665 | | | Malignant neoplasm of gallbladder and extrahepatic bile ducts | neoplasms | LDL-C | |
| 159.4 | | | | | 22743 | 86 | 22657 | | | | | 32783 | 118 | 32665 | | | Malignant neoplasm of retroperitoneum and peritoneum | neoplasms | LDL-C | |
| 164 | | | | | 25778 | 71 | 25707 | | | | | 35298 | 55 | 35243 | | | Cancer of intrathoracic organs | neoplasms | LDL-C | |
| 165 | -0.4046 | 0.06184 | 0.6673 | 6.04e-11 | 27233 | 1526 | 25707 | -0.3295 | 0.06916 | 0.7193 | 1.90e-06 | 36502 | 1259 | 35243 | 0.2928 | -0.08641 | Cancer within the respiratory system | neoplasms | LDL-C | |
| 165.1 | -0.3999 | 0.06235 | 0.6704 | 1.42e-10 | 27209 | 1502 | 25707 | -0.3345 | 0.06949 | 0.7157 | 1.48e-06 | 36492 | 1249 | 35243 | 0.2734 | -0.09052 | Cancer of bronchus; lung | neoplasms | LDL-C | |
| 170 | -0.8449 | 0.1062 | 0.4296 | 1.82e-15 | 27372 | 508 | 26864 | | | | | 36581 | 434 | 36147 | 0.7014 | 0.05449 | Cancer of bone and connective tissue | neoplasms | LDL-C | |
| 170.1 | | | | | 27106 | 242 | 26864 | | | | | 36353 | 206 | 36147 | | | Bone cancer | neoplasms | LDL-C | |
| 170.2 | | | | | 27193 | 329 | 26864 | | | | | 36433 | 286 | 36147 | 0.9868 | 0.002937 | Cancer of connective tissue | neoplasms | LDL-C | |
| 172 | 0.3168 | 0.04507 | 1.373 | 2.08e-12 | 25415 | 3454 | 21961 | 0.5015 | 0.04934 | 1.651 | 2.87e-24 | 33247 | 2675 | 30572 | 0.002647 | 0.1832 | Skin cancer | neoplasms | LDL-C | |

| phenotype | male_beta | male_SE | male_OR | male_p | male_n_total | male_n_cases | male_n_controls | female_beta | female_SE | female_OR | female_p | female_n_total | female_n_cases | female_n_controls | interaction_p | interaction_beta | description | group | lab | label |
|---|---|---|---|---|---|---|---|---|---|---|---|---|---|---|---|---|---|---|---|---|
| 172.1 | -0.3159 | 0.07396 | 0.7291 | 1.94e-05 | 22929 | 968 | 21961 | -0.08279 | 0.08416 | 0.9205 | 0.3252 | 31295 | 723 | 30572 | 0.2812 | 0.1086 | Melanomas of skin, dx or hx | neoplasms | LDL-C | |
| 172.11 | -0.4916 | 0.08129 | 0.6117 | 1.47e-09 | 22766 | 805 | 21961 | -0.2281 | 0.09623 | 0.796 | 0.01775 | 31138 | 566 | 30572 | 0.3992 | 0.0958 | Melanomas of skin | neoplasms | LDL-C | |
| phenotype | male_beta | male_SE | male_OR | male_p | male_n_total | male_n_cases | male_n_controls | female_beta | female_SE | female_OR | female_p | female_n_total | female_n_cases | female_n_controls | interaction_p | interaction_beta | description | group | lab | label |
| 172.2 | 0.5079 | 0.05144 | 1.662 | 5.38e-23 | 24804 | 2843 | 21961 | 0.7106 | 0.0582 | 2.035 | 2.75e-34 | 32709 | 2137 | 30572 | 6.51e-04 | 0.2478 | Other non-epithelial cancer of skin | neoplasms | LDL-C | |
| 172.21 | 1.028 | 0.1009 | 2.797 | 2.10e-24 | 22933 | 972 | 21961 | 1.097 | 0.1126 | 2.996 | 1.94e-22 | 31280 | 708 | 30572 | 0.8955 | 0.01921 | Basal cell carcinoma | neoplasms | LDL-C | |
| 172.22 | 0.5078 | 0.09099 | 1.662 | 2.39e-08 | 22797 | 836 | 21961 | | | | | 30972 | 400 | 30572 | 0.002677 | 0.4754 | Squamous cell carcinoma | neoplasms | LDL-C | |
| 172.3 | | | | | 22359 | 398 | 21961 | | | | | 30807 | 235 | 30572 | 0.5092 | 0.1548 | Carcinoma in situ of skin | neoplasms | LDL-C | |
| 173 | 1.186 | 0.07265 | 3.273 | 6.99e-60 | 23987 | 2026 | 21961 | 1.415 | 0.06578 | 4.115 | 1.34e-102 | 32915 | 2343 | 30572 | 0.1536 | 0.1349 | Neoplasm of uncertain behavior of skin | neoplasms | LDL-C | |
| 174 | | | | | 27350 | 45 | 27305 | 0.02606 | 0.04288 | 1.026 | 0.5434 | 31827 | 3093 | 28734 | 0.1265 | 0.46 | Breast cancer | neoplasms | LDL-C | |
| 174.1 | | | | | 7 | | | 0.009079 | 0.04342 | 1.009 | 0.8344 | 31729 | 2998 | 28731 | 0.9999 | 0.05969 | Breast cancer [female] | neoplasms | LDL-C | |
| 174.11 | | | | | 7 | | | 0.004001 | 0.04361 | 1.004 | 0.9269 | 31698 | 2967 | 28731 | 0.9999 | 0.05297 | Malignant neoplasm of female breast | neoplasms | LDL-C | |
| 174.2 | | | | | 27313 | 15 | 27298 | | | | | 3 | | | | | Breast cancer [male] | neoplasms | LDL-C | |
| 174.3 | | | | | 27310 | 5 | 27305 | | | | | 28889 | 155 | 28734 | | | Neoplasm of uncertain behavior of breast | neoplasms | LDL-C | |
| 175 | | | | | 7 | | | 0.2578 | 0.06405 | 1.294 | 5.72e-05 | 27358 | 1237 | 26121 | 0.9991 | 0.4032 | Acquired absence of breast | neoplasms | LDL-C | |
| 180 | | | | | 7 | | | 0.5421 | 0.07685 | 1.72 | 1.74e-12 | 28736 | 897 | 27839 | 0.9989 | 0.7803 | Cervical cancer and dysplasia | neoplasms | LDL-C | |
| 180.1 | | | | | 7 | | | | | | | 28134 | 295 | 27839 | | | Cervical cancer | neoplasms | LDL-C | |
| 180.3 | | | | | 7 | | | 0.9386 | 0.09557 | 2.556 | 9.11e-23 | 28457 | 618 | 27839 | 0.9982 | 1.295 | Cervical intraepithelial neoplasia [CIN] [Cervical dysplasia] | neoplasms | LDL-C | |
| 182 | | | | | 7 | | | -0.1336 | 0.09883 | 0.875 | 0.1766 | 29788 | 534 | 29254 | 0.9998 | -0.1394 | Malignant neoplasm of uterus | neoplasms | LDL-C | |
| 184 | | | | | 7 | | | -0.2147 | 0.08268 | 0.8068 | 0.009411 | 30877 | 771 | 30106 | 0.9996 | -0.2698 | Cancer of other female genital organs | neoplasms | LDL-C | |
| 184.1 | | | | | 7 | | | -0.2384 | 0.09605 | 0.7879 | 0.01307 | 30679 | 573 | 30106 | 0.9995 | -0.3272 | Malignant neoplasm of ovary and other uterine adnexa | neoplasms | LDL-C | |
| 184.11 | | | | | 7 | | | | | | | 30568 | 462 | 30106 | | | Malignant neoplasm of ovary | neoplasms | LDL-C | |
| 184.2 | | | | | 7 | | | | | | | 30333 | 227 | 30106 | | | Cancer of other female genital organs (excluding uterus and ovary) | neoplasms | LDL-C | |
| 185 | -0.07378 | 0.05551 | 0.9289 | 0.1838 | 21441 | 2053 | 19388 | | | | | 4 | | | 0.9974 | -1.19 | Cancer of prostate | neoplasms | LDL-C | |
| 187 | | | | | 25045 | 181 | 24864 | | | | | 4 | | | | | Cancer of other male genital organs | neoplasms | LDL-C | |
| 187.1 | | | | | 24907 | 43 | 24864 | | | | | 4 | | | | | Malignant neoplasm of unspecified male genital organ | neoplasms | LDL-C | |
| 187.2 | | | | | 24988 | 124 | 24864 | | | | | 4 | | | | | Malignant neoplasm of testis | neoplasms | LDL-C | |
| 187.8 | | | | | 24880 | 16 | 24864 | | | | | 4 | | | | | Neoplasm of uncertain behavior of male genital organs | neoplasms | LDL-C | |
| 189 | -0.2212 | 0.06503 | 0.8016 | 6.72e-04 | 27414 | 1343 | 26071 | 0.01707 | 0.09843 | 1.017 | 0.8623 | 36667 | 553 | 36114 | 0.008854 | 0.2704 | Cancer of urinary organs (incl. kidney and bladder) | neoplasms | LDL-C | |
| 189.1 | -0.08614 | 0.08553 | 0.9175 | 0.3138 | 26801 | 730 | 26071 | | | | | 36472 | 358 | 36114 | 0.1057 | 0.2115 | Cancer of kidney and renal pelvis | neoplasms | LDL-C | |
| 189.11 | -0.02018 | 0.09143 | 0.98 | 0.8254 | 26706 | 635 | 26071 | | | | | 36422 | 308 | 36114 | 0.1304 | 0.2126 | Malignant neoplasm of kidney, except pelvis | neoplasms | LDL-C | |
| 189.12 | | | | | 26217 | 146 | 26071 | | | | | 36178 | 64 | 36114 | | | Malignant neoplasm of renal pelvis | neoplasms | LDL-C | |
| 189.2 | -0.407 | 0.08865 | 0.6656 | 4.40e-06 | 26790 | 719 | 26071 | | | | | 36340 | 226 | 36114 | 0.05855 | 0.2923 | Cancer of bladder | neoplasms | LDL-C | |

| phenotype | male_beta | male_SE | male_OR | male_p | male_n_total | male_n_cases | male_n_controls | female_beta | female_SE | female_OR | female_p | female_n_total | female_n_cases | female_n_controls | interaction_p | interaction_beta | description | group | lab | label |
|---|---|---|---|---|---|---|---|---|---|---|---|---|---|---|---|---|---|---|---|---|
| 189.21 | -0.4161 | 0.0893 | 0.6596 | 3.17e-06 | 26780 | 709 | 26071 | | | | | 36336 | 222 | 36114 | 0.0634 | 0.2893 | Malignant neoplasm of bladder | neoplasms | LDL-C | |
| 189.4 | | | | | 26367 | 296 | 26071 | | | | | 36227 | 113 | 36114 | | | Malignant neoplasm of other urinary organs | neoplasms | LDL-C | |
| 190 | | | | | 26618 | 60 | 26558 | | | | | 35577 | 58 | 35519 | | | Cancer of eye | neoplasms | LDL-C | |
| 191 | | | | | 27023 | 465 | 26558 | | | | | 35973 | 454 | 35519 | 0.9185 | -0.01432 | Manlignant and unknown neoplasms of brain and nervous system | neoplasms | LDL-C | |
| 191.1 | | | | | 26929 | 371 | 26558 | | | | | 35854 | 335 | 35519 | 0.809 | -0.03922 | Cancer of brain and nervous system | neoplasms | LDL-C | |
| 191.11 | | | | | 26893 | 335 | 26558 | | | | | 35801 | 282 | 35519 | 0.9726 | -0.005958 | Cancer of brain | neoplasms | LDL-C | |
| 193 | | | | | 27161 | 215 | 26946 | | | | | 35853 | 482 | 35371 | 0.173 | 0.2252 | Thyroid cancer | neoplasms | LDL-C | |
| 194 | | | | | 26993 | 47 | 26946 | | | | | 35425 | 54 | 35371 | | | Cancer of other endocrine glands | neoplasms | LDL-C | |
| 195 | -0.5446 | 0.049 | 0.5801 | 1.07e-28 | 20735 | 2450 | 18285 | -0.2715 | 0.05389 | 0.7622 | 4.70e-07 | 28047 | 1903 | 26144 | 7.56e-05 | 0.2613 | Cancer, suspected or other | neoplasms | LDL-C | |
| 195.1 | -0.5108 | 0.06638 | 0.6 | 1.41e-14 | 19494 | 1209 | 18285 | -0.2818 | 0.0736 | 0.7544 | 1.29e-04 | 27102 | 958 | 26144 | 0.01746 | 0.2151 | Malignant neoplasm, other | neoplasms | LDL-C | |
| 195.3 | -0.7959 | 0.08022 | 0.4512 | 3.36e-23 | 19155 | 870 | 18285 | | | | | 26453 | 309 | 26144 | 0.4988 | -0.09978 | Malignant neoplasm of head, face, and neck | neoplasms | LDL-C | |
| 196 | 0.01362 | 0.07006 | 1.014 | 0.8458 | 19459 | 1174 | 18285 | 0.4314 | 0.0648 | 1.539 | 2.79e-11 | 27555 | 1411 | 26144 | 3.29e-05 | 0.368 | Radiotherapy | neoplasms | LDL-C | |
| 197 | -0.2827 | 0.03829 | 0.7538 | 1.55e-13 | 22676 | 4391 | 18285 | -0.01065 | 0.03556 | 0.9894 | 0.7645 | 30992 | 4848 | 26144 | 1.11e-13 | 0.3471 | Chemotherapy | neoplasms | LDL-C | |
| 198 | -0.5721 | 0.04226 | 0.5643 | 9.40e-42 | 22039 | 3754 | 18285 | -0.3553 | 0.04314 | 0.701 | 1.79e-16 | 29550 | 3406 | 26144 | 8.36e-05 | 0.2152 | Secondary malignant neoplasm | neoplasms | LDL-C | |
| 198.1 | -0.5399 | 0.054 | 0.5828 | 1.54e-23 | 20292 | 2007 | 18285 | -0.2505 | 0.05721 | 0.7784 | 1.19e-05 | 27825 | 1681 | 26144 | 6.90e-06 | 0.322 | Secondary malignancy of lymph nodes | neoplasms | LDL-C | |
| 198.2 | -0.5018 | 0.06615 | 0.6055 | 3.32e-14 | 19626 | 1341 | 18285 | -0.3154 | 0.07124 | 0.7295 | 9.53e-06 | 27296 | 1152 | 26144 | 0.06423 | 0.165 | Secondary malignancy of respiratory organs | neoplasms | LDL-C | |
| 198.3 | -0.504 | 0.09894 | 0.6041 | 3.51e-07 | 18851 | 566 | 18285 | -0.3224 | 0.0971 | 0.7244 | 8.99e-04 | 26726 | 582 | 26144 | 0.0314 | 0.2742 | Secondary malignant neoplasm of digestive systems | neoplasms | LDL-C | |
| 198.4 | -0.4634 | 0.07454 | 0.6291 | 5.05e-10 | 19347 | 1062 | 18285 | -0.2606 | 0.07692 | 0.7706 | 7.05e-04 | 27121 | 977 | 26144 | 0.01082 | 0.2503 | Secondary malignant neoplasm of liver | neoplasms | LDL-C | |
| 198.5 | -0.249 | 0.1014 | 0.7796 | 0.01404 | 18799 | 514 | 18285 | -0.4065 | 0.09958 | 0.666 | 4.46e-05 | 26737 | 593 | 26144 | 0.2727 | -0.1436 | Secondary malignancy of brain/spine | neoplasms | LDL-C | |
| 198.6 | -0.3847 | 0.0742 | 0.6807 | 2.17e-07 | 19284 | 999 | 18285 | -0.2197 | 0.08165 | 0.8027 | 0.007119 | 26962 | 818 | 26144 | 0.2326 | 0.1201 | Secondary malignancy of bone | neoplasms | LDL-C | |
| 198.7 | | | | | 18508 | 223 | 18285 | | | | | 26369 | 225 | 26144 | | | Secondary malignant neoplasm of skin | neoplasms | LDL-C | |
| 199 | -0.1864 | 0.06445 | 0.83 | 0.003829 | 19599 | 1314 | 18285 | 0.07863 | 0.05894 | 1.082 | 0.1821 | 27705 | 1561 | 26144 | 0.006372 | 0.2168 | Neoplasm of uncertain behavior | neoplasms | LDL-C | |
| 199.4 | | | | | 18315 | 30 | 18285 | | | | | 26186 | 42 | 26144 | | | Neurofibromatosis | neoplasms | LDL-C | |
| 200 | 0.07474 | 0.09287 | 1.078 | 0.4209 | 26538 | 642 | 25896 | -0.01877 | 0.09606 | 0.9814 | 0.8451 | 35761 | 577 | 35184 | 0.6291 | -0.0564 | Myeloproliferative disease | neoplasms | LDL-C | |
| 200.1 | | | | | 21974 | 134 | 21840 | | | | | 30355 | 101 | 30254 | | | Polycythemia vera | neoplasms | LDL-C | |
| 201 | | | | | 25974 | 78 | 25896 | | | | | 35242 | 58 | 35184 | | | Hodgkin's disease | neoplasms | LDL-C | |
| 202 | | | | | 26283 | 387 | 25896 | | | | | 35476 | 292 | 35184 | 0.3793 | -0.138 | Cancer of other lymphoid, histiocytic tissue | neoplasms | LDL-C | |
| 202.2 | | | | | 26223 | 327 | 25896 | | | | | 35425 | 241 | 35184 | 0.1446 | -0.251 | Non-Hodgkins lymphoma | neoplasms | LDL-C | |
| 202.21 | | | | | 25957 | 61 | 25896 | | | | | 35235 | 51 | 35184 | | | Nodular lymphoma | neoplasms | LDL-C | |
| 202.22 | | | | | 25981 | 85 | 25896 | | | | | 35237 | 53 | 35184 | | | Reticulosarcoma | neoplasms | LDL-C | |
| 202.23 | | | | | 25947 | 51 | 25896 | | | | | 35208 | 24 | 35184 | | | Lymphosarcoma | neoplasms | LDL-C | |
| phenotype | male_beta | male_SE | male_OR | male_p | male_n_total | male_n_cases | male_n_controls | female_beta | female_SE | female_OR | female_p | female_n_total | female_n_cases | female_n_controls | interaction_p | interaction_beta | description | group | lab | label |
| 202.24 | | | | | 25960 | 64 | 25896 | | | | | 35217 | 33 | 35184 | | | Large cell lymphoma | neoplasms | LDL-C | |
| 204 | -0.3239 | 0.08795 | 0.7233 | 2.31e-04 | 26591 | 695 | 25896 | -0.264 | 0.102 | 0.768 | 0.009644 | 35705 | 521 | 35184 | 0.8183 | -0.02717 | Leukemia | neoplasms | LDL-C | |

| phenotype | male_beta | male_SE | male_OR | male_p | male_n_total | male_n_cases | male_n_controls | female_beta | female_SE | female_OR | female_p | female_n_total | female_n_cases | female_n_controls | interaction_p | interaction_beta | description | group | lab | label |
|---|---|---|---|---|---|---|---|---|---|---|---|---|---|---|---|---|---|---|---|---|
| 204.1 | | | | | 26113 | 217 | 25896 | | | | | 35327 | 143 | 35184 | | | Lymphoid leukemia | neoplasms | LDL-C | |
| 204.11 | | | | | 25996 | 100 | 25896 | | | | | 35260 | 76 | 35184 | | | Lymphoid leukemia, acute | neoplasms | LDL-C | |
| 204.12 | | | | | 26033 | 137 | 25896 | | | | | 35270 | 86 | 35184 | | | Lymphoid leukemia, chronic | neoplasms | LDL-C | |
| 204.2 | | | | | 26331 | 435 | 25896 | | | | | 35537 | 353 | 35184 | 0.3954 | 0.1275 | Myeloid leukemia | neoplasms | LDL-C | |
| 204.21 | | | | | 26263 | 367 | 25896 | | | | | 35482 | 298 | 35184 | 0.7206 | 0.0584 | Myeloid leukemia, acute | neoplasms | LDL-C | |
| 204.22 | | | | | 26069 | 173 | 25896 | | | | | 35327 | 143 | 35184 | | | Myeloid leukemia, chronic | neoplasms | LDL-C | |
| 204.3 | | | | | 25933 | 37 | 25896 | | | | | 35214 | 30 | 35184 | | | Monocytic leukemia | neoplasms | LDL-C | |
| 204.4 | | | | | 26060 | 164 | 25896 | | | | | 35291 | 107 | 35184 | | | Multiple myeloma | neoplasms | LDL-C | |
| 208 | 1.611 | 0.06556 | 5.006 | 2.95e-133 | 24481 | 2858 | 21623 | 1.339 | 0.06197 | 3.815 | 1.51e-103 | 33540 | 2573 | 30967 | 0.001273 | -0.2811 | Benign neoplasm of colon | neoplasms | LDL-C | |
| 209 | | | | | 27760 | 86 | 27674 | | | | | 36911 | 107 | 36804 | | | Neuroendocrine tumors | neoplasms | LDL-C | |
| 210 | | | | | 26386 | 107 | 26279 | | | | | 36167 | 130 | 36037 | | | Benign neoplasm of lip, oral cavity, and pharynx | neoplasms | LDL-C | |
| 211 | | | | | 24095 | 236 | 23859 | | | | | 33779 | 379 | 33400 | 0.6593 | 0.07784 | Benign neoplasm of other parts of digestive system | neoplasms | LDL-C | |
| 212 | | | | | 27637 | 101 | 27536 | | | | | 36801 | 112 | 36689 | | | Benign neoplasm of respiratory and intrathoracic organs | neoplasms | LDL-C | |
| 213 | | | | | 21553 | 68 | 21485 | | | | | 26105 | 131 | 25974 | | | Benign neoplasm of bone and articular cartilage | neoplasms | LDL-C | |
| 214 | | | | | 27025 | 277 | 26748 | | | | | 35990 | 359 | 35631 | 0.9208 | 0.0247 | Lipoma | neoplasms | LDL-C | |
| 214.1 | | | | | 26919 | 171 | 26748 | | | | | 35867 | 236 | 35631 | | | Lipoma of skin and subcutaneous tissue | neoplasms | LDL-C | |
| 215 | | | | | 26820 | 72 | 26748 | | | | | 35793 | 162 | 35631 | | | Other benign neoplasm of connective and other soft tissue | neoplasms | LDL-C | |
| 216 | 1.505 | 0.08587 | 4.505 | 8.69e-69 | 25936 | 1607 | 24329 | 1.778 | 0.06529 | 5.916 | 3.20e-163 | 34007 | 2621 | 31386 | 0.1813 | 0.1397 | Benign neoplasm of skin | neoplasms | LDL-C | |
| 216.1 | | | | | 24408 | 79 | 24329 | | | | | 31484 | 98 | 31386 | | | Screening for malignant neoplasms of the skin | neoplasms | LDL-C | |
| 217 | | | | | 24477 | 148 | 24329 | | | | | 31698 | 312 | 31386 | | | Vascular hamartomas and non-neoplastic nevi | neoplasms | LDL-C | |
| 217.1 | | | | | 24444 | 115 | 24329 | | | | | 31637 | 251 | 31386 | | | Nevus, non-neoplastic | neoplasms | LDL-C | |
| 218 | | | | | 7 | | | 1.095 | 0.08953 | 2.99 | 2.10e-34 | 31913 | 869 | 31044 | 0.998 | 1.418 | Benign neoplasm of uterus | neoplasms | LDL-C | |
| 218.1 | | | | | 7 | | | 1.149 | 0.09304 | 3.155 | 4.83e-35 | 31868 | 824 | 31044 | 0.998 | 1.476 | Uterine leiomyoma | neoplasms | LDL-C | |
| 218.2 | | | | | 7 | | | | | | | 31064 | 20 | 31044 | | | Other benign neoplasm of uterus | neoplasms | LDL-C | |
| 220 | | | | | 7 | | | | | | | 28308 | 209 | 28099 | | | Benign neoplasm of ovary | neoplasms | LDL-C | |
| 221 | | | | | 7 | | | | | | | 28086 | 59 | 28027 | | | Benign neoplasm of other female genital organs | neoplasms | LDL-C | |
| 222 | | | | | 24885 | 21 | 24864 | | | | | 4 | | | | | Benign neoplasm of male genital organs | neoplasms | LDL-C | |
| 223 | | | | | 21509 | 24 | 21485 | | | | | 26009 | 35 | 25974 | | | Benign neoplasm of kidney and other urinary organs | neoplasms | LDL-C | |
| 224 | | | | | 26668 | 110 | 26558 | | | | | 35659 | 140 | 35519 | | | Benign neoplasm of eye | neoplasms | LDL-C | |
| 224.1 | | | | | 26643 | 85 | 26558 | | | | | 35621 | 102 | 35519 | | | Benign neoplasm of eye, uveal | neoplasms | LDL-C | |
| 225 | | | | | 26749 | 191 | 26558 | | | | | 35909 | 390 | 35519 | 0.1527 | 0.255 | Benign neoplasm of brain and other parts of nervous system | neoplasms | LDL-C | |
| 225.1 | | | | | 26730 | 172 | 26558 | | | | | 35878 | 359 | 35519 | 0.1112 | 0.2979 | Benign neoplasm of brain, cranial nerves, meninges | neoplasms | LDL-C | |
| 225.2 | | | | | 26574 | 16 | 26558 | | | | | 35551 | 32 | 35519 | | | Benign neoplasm of spinal cord, meninges | neoplasms | LDL-C | |
| 226 | | | | | 26971 | 25 | 26946 | | | | | 35500 | 129 | 35371 | | | Benign neoplasm of thyroid glands | neoplasms | LDL-C | |
| 227 | | | | | 27177 | 231 | 26946 | | | | | 35717 | 346 | 35371 | 0.8032 | 0.04416 | Benign neoplasm of other endocrine glands and related structures | neoplasms | LDL-C | |
| 227.1 | | | | | 27032 | 86 | 26946 | | | | | 35495 | 124 | 35371 | | | Benign neoplasm of adrenal gland | neoplasms | LDL-C | |

| phenotype | male_beta | male_SE | male_OR | male_p | male_n_total | male_n_cases | male_n_controls | female_beta | female_SE | female_OR | female_p | female_n_total | female_n_cases | female_n_controls | interaction_p | interaction_beta | description | group | lab | label |
|---|---|---|---|---|---|---|---|---|---|---|---|---|---|---|---|---|---|---|---|---|
| 227.2 | | | | | 26972 | 26 | 26946 | | | | | 35433 | 62 | 35371 | | | Benign neoplasm of parathyroid gland | neoplasms | LDL-C | |
| 227.3 | | | | | 27062 | 116 | 26946 | | | | | 35525 | 154 | 35371 | | | Benign neoplasm of pituitary gland and craniopharyngeal duct (pouch) | neoplasms | LDL-C | |
| 228 | 1.272 | 0.1516 | 3.568 | 4.90e-17 | 26936 | 519 | 26417 | 1.344 | 0.1168 | 3.836 | 1.14e-30 | 35731 | 759 | 34972 | 0.8539 | -0.03418 | Hemangioma and lymphangioma, any site | neoplasms | LDL-C | |
| 228.1 | | | | | 26853 | 436 | 26417 | 1.812 | 0.1588 | 6.123 | 3.73e-30 | 35591 | 619 | 34972 | 0.771 | 0.07223 | Hemangioma of skin and subcutaneous tissue | neoplasms | LDL-C | |
| 229 | | | | | 27551 | 83 | 27468 | | | | | 36445 | 69 | 36376 | | | Benign neoplasm of unspecified sites | neoplasms | LDL-C | |
| 229.1 | | | | | 27477 | 9 | 27468 | | | | | 36386 | 10 | 36376 | | | Benign neoplasm of lymph nodes | neoplasms | LDL-C | |
| 230 | | | | | 27785 | 15 | 27770 | | | | | 36952 | 4 | 36948 | | | Kaposi's sarcoma | neoplasms | LDL-C | |
| 240 | | | | | 23255 | 80 | 23175 | | | | | 26401 | 346 | 26055 | | | Simple and unspecified goiter | endocrine/metabolic | LDL-C | |
| 241 | 0.7222 | 0.117 | 2.059 | 6.75e-10 | 23679 | 504 | 23175 | 0.8735 | 0.05781 | 2.395 | 1.41e-51 | 28029 | 1974 | 26055 | 0.262 | 0.1361 | Nontoxic nodular goiter | endocrine/metabolic | LDL-C | |
| 241.1 | | | | | 23492 | 317 | 23175 | 1.051 | 0.08528 | 2.862 | 6.28e-35 | 27019 | 964 | 26055 | 0.3123 | 0.1669 | Nontoxic uninodular goiter | endocrine/metabolic | LDL-C | |
| 241.2 | | | | | 23428 | 253 | 23175 | 0.8777 | 0.07166 | 2.405 | 1.72e-34 | 27333 | 1278 | 26055 | 0.2194 | 0.2034 | Nontoxic multinodular goiter | endocrine/metabolic | LDL-C | |
| 242 | | | | | 23461 | 286 | 23175 | 0.7389 | 0.08479 | 2.094 | 2.92e-18 | 26848 | 793 | 26055 | 0.565 | 0.0903 | Thyrotoxicosis with or without goiter | endocrine/metabolic | LDL-C | |
| 242.1 | | | | | 23301 | 126 | 23175 | | | | | 26434 | 379 | 26055 | 0.03651 | 0.4559 | Graves' disease | endocrine/metabolic | LDL-C | |
| 242.2 | | | | | 23196 | 21 | 23175 | | | | | 26178 | 123 | 26055 | | | Toxic multinodular goiter | endocrine/metabolic | LDL-C | |
| 242.3 | | | | | 23231 | 56 | 23175 | | | | | 26122 | 67 | 26055 | | | Exophthalmos | endocrine/metabolic | LDL-C | |
| 242.31 | | | | | 23186 | 11 | 23175 | | | | | 26071 | 16 | 26055 | | | Thyrotoxic exophthalmos | endocrine/metabolic | LDL-C | |
| 244 | 0.6949 | 0.05183 | 2.004 | 5.36e-41 | 25712 | 2537 | 23175 | 0.8417 | 0.03331 | 2.32 | 7.38e-141 | 32658 | 6603 | 26055 | 0.001116 | 0.1853 | Hypothyroidism | endocrine/metabolic | LDL-C | |
| 244.1 | 0.2777 | 0.1028 | 1.32 | 0.006912 | 23694 | 519 | 23175 | 0.6143 | 0.06536 | 1.848 | 5.55e-21 | 27402 | 1347 | 26055 | 3.58e-04 | 0.4016 | Secondary hypothyroidism | endocrine/metabolic | LDL-C | |
| 244.2 | | | | | 23522 | 347 | 23175 | 1.747 | 0.1118 | 5.737 | 4.45e-55 | 26876 | 821 | 26055 | 1.25e-04 | 0.6915 | Acquired hypothyroidism | endocrine/metabolic | LDL-C | |
| 244.3 | | | | | 23175 | | | | | | | 26060 | 5 | 26055 | | | Iodine hypothyroidism | endocrine/metabolic | LDL-C | |
| 244.4 | 0.8024 | 0.05587 | 2.231 | 9.00e-47 | 25441 | 2266 | 23175 | 0.9534 | 0.03531 | 2.595 | 1.54e-160 | 32096 | 6041 | 26055 | 0.001245 | 0.1973 | Hypothyroidism NOS | endocrine/metabolic | LDL-C | |
| 244.5 | | | | | 23199 | 24 | 23175 | | | | | 26089 | 34 | 26055 | | | Congenital hypothyroidism | endocrine/metabolic | LDL-C | |

| phenotype | male_beta | male_SE | male_OR | male_p | male_n_total | male_n_cases | male_n_controls | female_beta | female_SE | female_OR | female_p | female_n_total | female_n_cases | female_n_controls | interaction_p | interaction_beta | description | group | lab | label |
|---|---|---|---|---|---|---|---|---|---|---|---|---|---|---|---|---|---|---|---|---|
| 245 | | | | | 23436 | 261 | 23175 | 1.303 | 0.08788 | 3.679 | 1.02e-49 | 26942 | 887 | 26055 | 0.5807 | -0.1066 | Thyroiditis | endocrine/metabolic | LDL-C | |
| 245.1 | | | | | 23196 | 21 | 23175 | | | | | 26097 | 42 | 26055 | | | Thyroiditis, acute and subacute | endocrine/metabolic | LDL-C | |
| 245.2 | | | | | 23326 | 151 | 23175 | 1.316 | 0.1019 | 3.729 | 3.85e-38 | 26707 | 652 | 26055 | 0.243 | -0.3083 | Chronic thyroiditis | endocrine/metabolic | LDL-C | |
| 245.21 | | | | | 23324 | 149 | 23175 | 1.308 | 0.1021 | 3.698 | 1.48e-37 | 26701 | 646 | 26055 | 0.2485 | -0.305 | Chronic lymphocytic thyroiditis | endocrine/metabolic | LDL-C | |
| 246 | | | | | 23388 | 213 | 23175 | | | | | 26539 | 484 | 26055 | 0.006194 | 0.5458 | Other disorders of thyroid | endocrine/metabolic | LDL-C | |
| 246.2 | | | | | 23182 | 7 | 23175 | | | | | 26065 | 10 | 26055 | | | Thyroid cyst | endocrine/metabolic | LDL-C | |
| 246.7 | | | | | 23294 | 119 | 23175 | | | | | 26304 | 249 | 26055 | | | Abnormal results of function study of thyroid | endocrine/metabolic | LDL-C | |

| phenotype | male_beta | male_SE | male_OR | male_p | male_n_total | male_n_cases | male_n_controls | female_beta | female_SE | female_OR | female_p | female_n_total | female_n_cases | female_n_controls | interaction_p | interaction_beta | description | group | lab | label |
|---|---|---|---|---|---|---|---|---|---|---|---|---|---|---|---|---|---|---|---|---|
| 249 | | | | | 17130 | 307 | 16823 | | | | | 26073 | 265 | 25808 | 0.8076 | -0.05169 | Secondary diabetes mellitus | endocrine/metabolic | LDL-C | |
| 250 | 1.143 | 0.03702 | 3.136 | 2.18e-209 | 22885 | 6062 | 16823 | 1.119 | 0.03577 | 3.062 | 7.35e-215 | 31483 | 5675 | 25808 | 0.1318 | -0.07147 | Diabetes mellitus | endocrine/metabolic | LDL-C | |
| 250.1 | 1.97 | 0.09416 | 7.171 | 3.27e-97 | 18010 | 1187 | 16823 | 2.034 | 0.08831 | 7.646 | 2.12e-117 | 27021 | 1213 | 25808 | 0.5655 | -0.07115 | Type 1 diabetes | endocrine/metabolic | LDL-C | |
| 250.11 | | | | | 16945 | 122 | 16823 | | | | | 25971 | 163 | 25808 | | | Type 1 diabetes with ketoacidosis | endocrine/metabolic | LDL-C | |
| 250.12 | | | | | 17020 | 197 | 16823 | | | | | 25984 | 176 | 25808 | | | Type 1 diabetes with renal manifestations | endocrine/metabolic | LDL-C | |
| 250.13 | | | | | 16957 | 134 | 16823 | | | | | 25957 | 149 | 25808 | | | Type 1 diabetes with ophthalmic manifestations | endocrine/metabolic | LDL-C | |
| 250.14 | | | | | 17123 | 300 | 16823 | | | | | 26082 | 274 | 25808 | 0.2767 | 0.3402 | Type 1 diabetes with neurological manifestations | endocrine/metabolic | LDL-C | |
| 250.15 | | | | | 16873 | 50 | 16823 | | | | | 25855 | 47 | 25808 | | | Diabetes type 1 with peripheral circulatory disorders | endocrine/metabolic | LDL-C | |
| 250.2 | 1.157 | 0.03776 | 3.179 | 4.94e-206 | 22677 | 5854 | 16823 | 1.109 | 0.03659 | 3.031 | 1.00e-201 | 31244 | 5436 | 25808 | 0.06345 | -0.08989 | Type 2 diabetes | endocrine/metabolic | LDL-C | |
| 250.21 | | | | | 16879 | 56 | 16823 | | | | | 25884 | 76 | 25808 | | | Type 2 diabetes with ketoacidosis | endocrine/metabolic | LDL-C | |
| 250.22 | 2.269 | 0.1044 | 9.67 | 1.04e-104 | 18226 | 1403 | 16823 | 2.273 | 0.1256 | 9.708 | 3.15e-73 | 26727 | 919 | 25808 | 0.8022 | -0.03967 | Type 2 diabetes with renal manifestations | endocrine/metabolic | LDL-C | |
| 250.23 | | | | | 17271 | 448 | 16823 | | | | | 26204 | 396 | 25808 | 0.5653 | 0.1392 | Type 2 diabetes with ophthalmic manifestations | endocrine/metabolic | LDL-C | |
| 250.24 | 1.948 | 0.08726 | 7.012 | 2.28e-110 | 18366 | 1543 | 16823 | 1.734 | 0.08796 | 5.663 | 1.70e-86 | 27084 | 1276 | 25808 | 0.05216 | -0.2299 | Type 2 diabetes with neurological manifestations | endocrine/metabolic | LDL-C | |
| 250.25 | | | | | 17307 | 484 | 16823 | | | | | 26109 | 301 | 25808 | 0.9111 | 0.04006 | Diabetes type 2 with peripheral circulatory disorders | endocrine/metabolic | LDL-C | |
| 250.3 | 1.849 | 0.07172 | 6.353 | 1.51e-146 | 18803 | 1980 | 16823 | 1.729 | 0.06965 | 5.638 | 4.22e-136 | 27587 | 1779 | 25808 | 0.06943 | -0.1726 | Insulin pump user | endocrine/metabolic | LDL-C | |
| 250.4 | 0.917 | 0.04313 | 2.502 | 2.79e-100 | 20734 | 3911 | 16823 | 1.008 | 0.04359 | 2.74 | 2.68e-118 | 29488 | 3680 | 25808 | 0.1112 | 0.08916 | Abnormal glucose | endocrine/metabolic | LDL-C | |
| 250.41 | 3.075 | 0.2642 | 21.65 | 2.65e-31 | 17402 | 579 | 16823 | 2.436 | 0.1713 | 11.43 | 6.92e-46 | 26465 | 657 | 25808 | 0.01037 | -0.7992 | Impaired fasting glucose | endocrine/metabolic | LDL-C | |
| 250.42 | 0.8226 | 0.04421 | 2.276 | 3.00e-77 | 20295 | 3472 | 16823 | 0.8807 | 0.04555 | 2.413 | 2.77e-83 | 28947 | 3139 | 25808 | 0.2095 | 0.07247 | Other abnormal glucose | endocrine/metabolic | LDL-C | |
| 250.5 | | | | | 16841 | 18 | 16823 | | | | | 25844 | 36 | 25808 | | | Glycosuria or Acetonuria | endocrine/metabolic | LDL-C | |
| 250.6 | 2.09 | 0.1035 | 8.087 | 1.24e-90 | 18032 | 1209 | 16823 | 1.789 | 0.1037 | 5.983 | 1.18e-66 | 26769 | 961 | 25808 | 0.04244 | -0.2854 | Polyneuropathy in diabetes | endocrine/metabolic | LDL-C | |
| 250.7 | | | | | 24804 | 450 | 24354 | | | | | 33128 | 398 | 32730 | 0.7724 | 0.07154 | Diabetic retinopathy | endocrine/metabolic | LDL-C | |
| 251 | | | | | 23451 | 170 | 23281 | | | | | 31202 | 167 | 31035 | | | Other disorders of pancreatic internal secretion | endocrine/metabolic | LDL-C | |
| 251.1 | | | | | 15397 | 246 | 15151 | | | | | 23345 | 400 | 22945 | 0.122 | 0.2961 | Hypoglycemia | endocrine/metabolic | LDL-C | |
| 251.8 | | | | | 23287 | 6 | 23281 | | | | | 31042 | 7 | 31035 | | | Abnormality of secretion of glucagon or gastrin | endocrine/metabolic | LDL-C | |
| 252 | | | | | 23548 | 267 | 23281 | 0.5261 | 0.08991 | 1.692 | 4.86e-09 | 31785 | 750 | 31035 | 0.2162 | -0.209 | Disorders of parathyroid gland | endocrine/metabolic | LDL-C | |
| 252.1 | | | | | 23519 | 238 | 23281 | 0.5586 | 0.09689 | 1.748 | 8.13e-09 | 31693 | 658 | 31035 | 0.07238 | -0.3346 | Hyperparathyroidism | endocrine/metabolic | LDL-C | |
| 252.2 | | | | | 23316 | 35 | 23281 | | | | | 31163 | 128 | 31035 | | | Hypoparathyroidism | endocrine/metabolic | LDL-C | |
| 253 | | | | | 23665 | 384 | 23281 | | | | | 31431 | 396 | 31035 | 0.01273 | 0.3663 | Disorders of the pituitary gland and its hypothalamic control | endocrine/metabolic | LDL-C | |
| 253.1 | | | | | 23343 | 62 | 23281 | | | | | 31171 | 136 | 31035 | | | Pituitary hyperfunction | endocrine/metabolic | LDL-C | |

| phenotype | male_beta | male_SE | male_OR | male_p | male_n_total | male_n_cases | male_n_controls | female_beta | female_SE | female_OR | female_p | female_n_total | female_n_cases | female_n_controls | interaction_p | interaction_beta | description | group | lab | label |
|---|---|---|---|---|---|---|---|---|---|---|---|---|---|---|---|---|---|---|---|---|
| 253.11 | | | | | 23302 | 21 | 23281 | | | | | 31046 | 11 | 31035 | | | Acromegaly and gigantism | endocrine/metabolic | LDL-C | |
| 253.2 | | | | | 23400 | 119 | 23281 | | | | | 31125 | 90 | 31035 | | | Pituitary hypofunction | endocrine/metabolic | LDL-C | |
| 253.3 | | | | | 23305 | 24 | 23281 | | | | | 31086 | 51 | 31035 | | | Diabetes insipidus | endocrine/metabolic | LDL-C | |
| 253.4 | | | | | 23423 | 142 | 23281 | | | | | 31110 | 75 | 31035 | | | Anterior pituitary disorders | endocrine/metabolic | LDL-C | |
| 253.5 | | | | | 23359 | 78 | 23281 | | | | | 31085 | 50 | 31035 | | | Pituitary dwarfism | endocrine/metabolic | LDL-C | |
| 253.7 | | | | | 23345 | 64 | 23281 | | | | | 31102 | 67 | 31035 | | | Other disorders of neurohypophysis | endocrine/metabolic | LDL-C | |
| 254 | | | | | 23286 | 5 | 23281 | | | | | 31042 | 7 | 31035 | | | Diseases of thymus gland | endocrine/metabolic | LDL-C | |
| 255 | 0.3569 | 0.08861 | 1.429 | 5.62e-05 | 23957 | 676 | 23281 | 0.4334 | 0.07875 | 1.542 | 3.74e-08 | 31897 | 862 | 31035 | 0.2257 | 0.1305 | Disorders of adrenal glands | endocrine/metabolic | LDL-C | |
| 255.1 | | | | | 23371 | 90 | 23281 | | | | | 31211 | 176 | 31035 | | | Adrenal hyperfunction | endocrine/metabolic | LDL-C | |
| 255.11 | | | | | 23313 | 32 | 23281 | | | | | 31129 | 94 | 31035 | | | Cushing's syndrome | endocrine/metabolic | LDL-C | |
| 255.12 | | | | | 23315 | 34 | 23281 | | | | | 31064 | 29 | 31035 | | | Hyperaldosteronism | endocrine/metabolic | LDL-C | |
| 255.13 | | | | | 23297 | 16 | 23281 | | | | | 31065 | 30 | 31035 | | | Medulloadrenal hyperfunction | endocrine/metabolic | LDL-C | |
| 255.2 | | | | | 23716 | 435 | 23281 | 0.4377 | 0.1023 | 1.549 | 1.87e-05 | 31535 | 500 | 31035 | 0.05427 | 0.2606 | Adrenal hypofunction | endocrine/metabolic | LDL-C | |
| 255.21 | | | | | 23674 | 393 | 23281 | | | | | 31474 | 439 | 31035 | 0.05615 | 0.2753 | Glucocorticoid deficiency | endocrine/metabolic | LDL-C | |
| 255.22 | | | | | 23296 | 15 | 23281 | | | | | 31067 | 32 | 31035 | | | Mineralocorticoid deficiency | endocrine/metabolic | LDL-C | |
| 255.3 | | | | | 23289 | 8 | 23281 | | | | | 31065 | 30 | 31035 | | | Adrenogenital disorders | endocrine/metabolic | LDL-C | |
| 256 | | | | | 4 | | | 1.388 | 0.09627 | 4.008 | 3.83e-47 | 31698 | 666 | 31032 | 3.83e-47 | 1.388 | Ovarian dysfunction | endocrine/metabolic | LDL-C | |
| 256.1 | | | | | 23283 | 2 | 23281 | | | | | 31037 | 2 | 31035 | | | Hyperestrogenism | endocrine/metabolic | LDL-C | |
| 256.4 | | | | | 4 | | | 1.606 | 0.1073 | 4.983 | 1.07e-50 | 31569 | 537 | 31032 | 1.07e-50 | 1.606 | Polycystic ovaries | endocrine/metabolic | LDL-C | |
| 257 | 1.48 | 0.09982 | 4.393 | 9.77e-50 | 24271 | 994 | 23277 | | | | | 3 | | | 0.9962 | -1.863 | Testicular dysfunction | endocrine/metabolic | LDL-C | |
| 257.1 | 1.497 | 0.1014 | 4.47 | 2.36e-49 | 24255 | 978 | 23277 | | | | | 3 | | | 0.9962 | -1.88 | Testicular hypofunction | endocrine/metabolic | LDL-C | |
| 258 | | | | | 23304 | 23 | 23281 | | | | | 31068 | 33 | 31035 | | | Iatrogenic endocrine disorders | endocrine/metabolic | LDL-C | |
| 258.1 | | | | | 4 | | | | | | | 31087 | 55 | 31032 | | | Postablative ovarian failure | endocrine/metabolic | LDL-C | |
| 259 | | | | | 23461 | 180 | 23281 | | | | | 31392 | 357 | 31035 | 0.6153 | 0.1115 | Other endocrine disorders | endocrine/metabolic | LDL-C | |
| phenotype | male_beta | male_SE | male_OR | male_p | male_n_total | male_n_cases | male_n_controls | female_beta | female_SE | female_OR | female_p | female_n_total | female_n_cases | female_n_controls | interaction_p | interaction_beta | description | group | lab | label |
| 259.1 | | | | | 23352 | 71 | 23281 | | | | | 31134 | 99 | 31035 | | | Nonspecific abnormal results of other endocrine function study | endocrine/metabolic | LDL-C | |
| 259.2 | | | | | 23320 | 39 | 23281 | | | | | 31092 | 57 | 31035 | | | Carcinoid syndrome | endocrine/metabolic | LDL-C | |
| 259.3 | | | | | 23324 | 43 | 23281 | | | | | 31063 | 28 | 31035 | | | Delay in sexual development and puberty NEC | endocrine/metabolic | LDL-C | |
| 259.4 | | | | | 23291 | 10 | 23281 | | | | | 31071 | 36 | 31035 | | | Precocious sexual development and puberty NEC | endocrine/metabolic | LDL-C | |

| phenotype | male_beta | male_SE | male_OR | male_p | male_n_total | male_n_cases | male_n_controls | female_beta | female_SE | female_OR | female_p | female_n_total | female_n_cases | female_n_controls | interaction_p | interaction_beta | description | group | lab | label |
|---|---|---|---|---|---|---|---|---|---|---|---|---|---|---|---|---|---|---|---|---|
| 259.8 | | | | | 23297 | 16 | 23281 | | | | | 31067 | 32 | 31035 | | | Polyglandular activity in multiple endocrine adenomatosis | endocrine/metabolic | LDL-C | |
| 260 | 0.1106 | 0.04475 | 1.117 | 0.01344 | 22122 | 2837 | 19285 | 0.4136 | 0.04628 | 1.512 | 4.06e-19 | 27318 | 2532 | 24786 | 1.38e-11 | 0.3949 | Protein-calorie malnutrition | endocrine/metabolic | LDL-C | |
| 260.1 | | | | | 19435 | 150 | 19285 | | | | | 24879 | 93 | 24786 | | | Cachexia | endocrine/metabolic | LDL-C | |
| 260.2 | 0.07225 | 0.06912 | 1.075 | 0.2959 | 20371 | 1086 | 19285 | 0.2906 | 0.07081 | 1.337 | 4.05e-05 | 25774 | 988 | 24786 | 0.001478 | 0.287 | severe protein-calorie malnutrition | endocrine/metabolic | LDL-C | |
| 260.21 | | | | | 19301 | 16 | 19285 | | | | | 24805 | 19 | 24786 | | | Kwashiorkor | endocrine/metabolic | LDL-C | |
| 260.22 | | | | | 19399 | 114 | 19285 | | | | | 24929 | 143 | 24786 | | | Nutritional marasmus | endocrine/metabolic | LDL-C | |
| 260.3 | 0.3894 | 0.09204 | 1.476 | 2.33e-05 | 19914 | 629 | 19285 | 0.7163 | 0.09595 | 2.047 | 8.35e-14 | 25366 | 580 | 24786 | 2.54e-04 | 0.4407 | Adult failure to thrive | endocrine/metabolic | LDL-C | |
| 260.6 | 0.126 | 0.09302 | 1.134 | 0.1754 | 19875 | 590 | 19285 | | | | | 25233 | 447 | 24786 | 0.00127 | 0.4101 | Anorexia | endocrine/metabolic | LDL-C | |
| 260.7 | | | | | 19286 | 1 | 19285 | | | | | 24804 | 18 | 24786 | | | Polyphagia | endocrine/metabolic | LDL-C | |
| 261 | 1.13 | 0.0595 | 3.095 | 2.12e-80 | 21716 | 2431 | 19285 | 1.252 | 0.03915 | 3.497 | 1.98e-224 | 30360 | 5574 | 24786 | 4.44e-04 | 0.2385 | Vitamin deficiency | endocrine/metabolic | LDL-C | |
| 261.1 | | | | | 19312 | 27 | 19285 | | | | | 24829 | 43 | 24786 | | | Vitamin A deficiency | endocrine/metabolic | LDL-C | |
| 261.2 | 1.207 | 0.116 | 3.345 | 2.30e-25 | 19973 | 688 | 19285 | 1.469 | 0.07332 | 4.343 | 2.98e-89 | 26376 | 1590 | 24786 | 0.09968 | 0.2172 | Vitamin B-complex deficiencies | endocrine/metabolic | LDL-C | |
| 261.3 | | | | | 19292 | 7 | 19285 | | | | | 24807 | 21 | 24786 | | | Vitamin C deficiencies | endocrine/metabolic | LDL-C | |
| 261.4 | 1.161 | 0.06672 | 3.192 | 8.82e-68 | 21216 | 1931 | 19285 | 1.27 | 0.04274 | 3.561 | 5.67e-194 | 29414 | 4628 | 24786 | 7.56e-04 | 0.2547 | Vitamin D deficiency | endocrine/metabolic | LDL-C | |
| 261.41 | | | | | 19297 | 12 | 19285 | | | | | 24819 | 33 | 24786 | | | Rickets or osteomalacia | endocrine/metabolic | LDL-C | |
| 262 | | | | | 19493 | 208 | 19285 | | | | | 25217 | 431 | 24786 | 0.9162 | -0.02626 | Mineral deficiency NEC | endocrine/metabolic | LDL-C | |
| 263 | | | | | 19569 | 284 | 19285 | | | | | 25175 | 389 | 24786 | 3.37e-08 | 0.9509 | Other nutritional deficiency | endocrine/metabolic | LDL-C | |
| 264 | | | | | 19689 | 404 | 19285 | | | | | 25083 | 297 | 24786 | 0.8842 | -0.02751 | Lack of normal physiological development | endocrine/metabolic | LDL-C | |
| 264.1 | | | | | 19424 | 139 | 19285 | | | | | 24879 | 93 | 24786 | | | Short stature | endocrine/metabolic | LDL-C | |
| 264.2 | | | | | 19409 | 124 | 19285 | | | | | 24883 | 97 | 24786 | | | Failure to thrive (childhood) | endocrine/metabolic | LDL-C | |
| 264.3 | | | | | 19388 | 103 | 19285 | | | | | 24857 | 71 | 24786 | | | Delayed milestones | endocrine/metabolic | LDL-C | |
| 264.9 | | | | | 19465 | 180 | 19285 | | | | | 24925 | 139 | 24786 | | | Lack of normal physiological development, unspecified | endocrine/metabolic | LDL-C | |
| 269 | 1.108 | 0.09365 | 3.028 | 2.75e-32 | 26340 | 917 | 25423 | 1.403 | 0.107 | 4.069 | 2.67e-39 | 35408 | 726 | 34682 | 0.09423 | 0.2256 | Proteinuria | endocrine/metabolic | LDL-C | |
| 270 | | | | | 25887 | 464 | 25423 | 0.278 | 0.1028 | 1.321 | 0.006822 | 35187 | 505 | 34682 | 0.5 | -0.09242 | Disorders of protein plasma/amino-acid transport and metabolism | endocrine/metabolic | LDL-C | |
| 270.1 | | | | | 25452 | 29 | 25423 | | | | | 34732 | 50 | 34682 | | | Disturbances of amino-acid transport | endocrine/metabolic | LDL-C | |
| 270.11 | | | | | 25431 | 8 | 25423 | | | | | 34711 | 29 | 34682 | | | Disturbances of sulphur-bearing amino-acid metabolism | endocrine/metabolic | LDL-C | |
| 270.12 | | | | | 25434 | 11 | 25423 | | | | | 34696 | 14 | 34682 | | | Phenylketonuria [PKU] | endocrine/metabolic | LDL-C | |
| 270.2 | | | | | 25465 | 42 | 25423 | | | | | 34719 | 37 | 34682 | | | Disorders of amino-acid metabolism | endocrine/metabolic | LDL-C | |
| 270.21 | | | | | 25457 | 34 | 25423 | | | | | 34703 | 21 | 34682 | | | Disorders of urea cycle metabolism | endocrine/metabolic | LDL-C | |

| phenotype | male_beta | male_SE | male_OR | male_p | male_n_total | male_n_cases | male_n_controls | female_beta | female_SE | female_OR | female_p | female_n_total | female_n_cases | female_n_controls | interaction_p | interaction_beta | description | group | lab | label |
|---|---|---|---|---|---|---|---|---|---|---|---|---|---|---|---|---|---|---|---|---|
| 270.3 | | | | | 25814 | 391 | 25423 | | | | | 35096 | 414 | 34682 | 0.1789 | -0.2059 | Disorders of plasma protein metabolism | endocrine/metabolic | LDL-C | |
| 270.31 | | | | | 25425 | 2 | 25423 | | | | | 34689 | 7 | 34682 | | | Polyclonal hypergammaglobulinemia | endocrine/metabolic | LDL-C | |
| 270.32 | | | | | 25665 | 242 | 25423 | | | | | 34920 | 238 | 34682 | | | Paraproteinemia | endocrine/metabolic | LDL-C | |
| 270.33 | | | | | 25484 | 61 | 25423 | | | | | 34731 | 49 | 34682 | | | Amyloidosis | endocrine/metabolic | LDL-C | |
| 270.34 | | | | | 25480 | 57 | 25423 | | | | | 34726 | 44 | 34682 | | | Alpha-1-antitrypsin deficiency | endocrine/metabolic | LDL-C | |
| 270.35 | | | | | 25440 | 17 | 25423 | | | | | 34692 | 10 | 34682 | | | Macroglobulinemia | endocrine/metabolic | LDL-C | |
| 270.38 | | | | | 25509 | 86 | 25423 | | | | | 34792 | 110 | 34682 | | | Other specified disorders of plasma protein metabolism | endocrine/metabolic | LDL-C | |
| 271 | | | | | 27474 | 374 | 27100 | 2.389 | 0.1826 | 10.91 | 3.82e-39 | 36489 | 676 | 35813 | 0.7811 | -0.09069 | Disorders of carbohydrate transport and metabolism | endocrine/metabolic | LDL-C | |
| 271.3 | | | | | 27455 | 355 | 27100 | 2.565 | 0.2003 | 13 | 1.56e-37 | 36474 | 661 | 35813 | 0.4279 | -0.3102 | Intestinal disaccharidase deficiencies and disaccharide malabsorption | endocrine/metabolic | LDL-C | |
| 271.9 | | | | | 27119 | 19 | 27100 | | | | | 35828 | 15 | 35813 | | | Other disorders of carbohydrate transport and metabolism | endocrine/metabolic | LDL-C | |
| 272 | 2.412 | 0.03815 | 11.16 | 0 | 25396 | 11129 | 14267 | 2.32 | 0.03674 | 10.18 | 0 | 34085 | 10701 | 23384 | 5.03e-05 | -0.2 | Disorders of lipoid metabolism | endocrine/metabolic | LDL-C | |
| 272.1 | 2.412 | 0.03816 | 11.15 | 0 | 25384 | 11117 | 14267 | 2.323 | 0.0368 | 10.2 | 0 | 34065 | 10681 | 23384 | 5.66e-05 | -0.1988 | Hyperlipidemia | endocrine/metabolic | LDL-C | |
| 272.11 | 2.969 | 0.07003 | 19.47 | 0 | 18174 | 3907 | 14267 | 2.898 | 0.06899 | 18.13 | 0 | 27356 | 3972 | 23384 | 0.01632 | -0.2279 | Hypercholesterolemia | endocrine/metabolic | LDL-C | |
| 272.12 | | | | | 14667 | 400 | 14267 | | | | | 23678 | 294 | 23384 | 0.02762 | -0.6853 | Hyperglyceridemia | endocrine/metabolic | LDL-C | |
| 272.13 | 3.389 | 0.06612 | 29.65 | 0 | 20087 | 5820 | 14267 | 3.311 | 0.0681 | 27.42 | 0 | 28806 | 5422 | 23384 | 0.01908 | -0.2158 | Mixed hyperlipidemia | endocrine/metabolic | LDL-C | Mixed hyperlipidemia |
| 272.14 | | | | | 14272 | 5 | 14267 | | | | | 23388 | 4 | 23384 | | | Hyperchylomicronemia | endocrine/metabolic | LDL-C | |
| 272.9 | | | | | 14286 | 19 | 14267 | | | | | 23403 | 19 | 23384 | | | Unspecified disorder of lipoid metabolism | endocrine/metabolic | LDL-C | |
| 274 | 1.056 | 0.06629 | 2.876 | 3.67e-57 | 26980 | 1926 | 25054 | 1.276 | 0.1043 | 3.581 | 2.12e-34 | 36355 | 864 | 35491 | 0.03776 | 0.2403 | Gout and other crystal arthropathies | endocrine/metabolic | LDL-C | |
| 274.1 | 1.07 | 0.06785 | 2.916 | 4.99e-56 | 26882 | 1828 | 25054 | 1.266 | 0.1106 | 3.547 | 2.36e-30 | 36217 | 726 | 35491 | 0.1801 | 0.1624 | Gout | endocrine/metabolic | LDL-C | |
| 274.11 | 1.523 | 0.1383 | 4.587 | 3.19e-28 | 25702 | 648 | 25054 | | | | | 35744 | 253 | 35491 | 0.1166 | -0.3546 | Gouty arthropathy | endocrine/metabolic | LDL-C | |
| 274.2 | | | | | 25175 | 121 | 25054 | | | | | 35648 | 157 | 35491 | | | Crystal arthropathies | endocrine/metabolic | LDL-C | |
| 274.21 | | | | | 25165 | 111 | 25054 | | | | | 35627 | 136 | 35491 | | | Chondrocalcinosis | endocrine/metabolic | LDL-C | |
| 275 | 0.4564 | 0.05256 | 1.578 | 3.81e-18 | 25825 | 2131 | 23694 | 0.589 | 0.04942 | 1.802 | 9.53e-33 | 34866 | 2480 | 32386 | 3.49e-05 | 0.2695 | Disorders of mineral metabolism | endocrine/metabolic | LDL-C | |
| 275.1 | | | | | 23853 | 159 | 23694 | | | | | 32511 | 125 | 32386 | | | Disorders of iron metabolism | hematopoietic | LDL-C | |
| 275.11 | | | | | 23755 | 61 | 23694 | | | | | 32420 | 34 | 32386 | | | Hereditary hemochromatosis | hematopoietic | LDL-C | |
| 275.2 | | | | | 23712 | 18 | 23694 | | | | | 32400 | 14 | 32386 | | | Disorders of copper metabolism | endocrine/metabolic | LDL-C | |
| 275.3 | 0.3805 | 0.07939 | 1.463 | 1.65e-06 | 24542 | 848 | 23694 | 0.4539 | 0.07566 | 1.574 | 1.99e-09 | 33312 | 926 | 32386 | 0.09962 | 0.1628 | Disorders of magnesium metabolism | endocrine/metabolic | LDL-C | |

| phenotype | male_beta | male_SE | male_OR | male_p | male_n_total | male_n_cases | male_n_controls | female_beta | female_SE | female_OR | female_p | female_n_total | female_n_cases | female_n_controls | interaction_p | interaction_beta | description | group | lab | label |
|---|---|---|---|---|---|---|---|---|---|---|---|---|---|---|---|---|---|---|---|---|
| 275.5 | 0.6366 | 0.0803 | 1.89 | 2.23e-15 | 24610 | 916 | 23694 | 0.5575 | 0.07858 | 1.746 | 1.30e-12 | 33307 | 921 | 32386 | 0.5692 | 0.05825 | Disorders of calcium/phosphorus metabolism | endocrine/metabolic | LDL-C | |
| 275.51 | | | | | 23996 | 302 | 23694 | | | | | 32803 | 417 | 32386 | 0.1767 | 0.2121 | Hypocalcemia | endocrine/metabolic | LDL-C | |
| 275.53 | 0.9136 | 0.1075 | 2.493 | 1.94e-17 | 24238 | 544 | 23694 | | | | | 32769 | 383 | 32386 | 0.9033 | 0.01827 | Disorders of phosphorus metabolism | endocrine/metabolic | LDL-C | |
| 275.6 | | | | | 24110 | 416 | 23694 | 0.9162 | 0.1019 | 2.5 | 2.45e-19 | 33089 | 703 | 32386 | 3.90e-06 | 0.653 | Hypercalcemia | endocrine/metabolic | LDL-C | |
| 276 | 0.4829 | 0.03142 | 1.621 | 2.62e-53 | 24148 | 8143 | 16005 | 0.5332 | 0.02937 | 1.704 | 1.11e-73 | 32644 | 8450 | 24194 | 0.001291 | 0.1246 | Disorders of fluid, electrolyte, and acid-base balance | endocrine/metabolic | LDL-C | |
| 276.1 | 0.613 | 0.03522 | 1.846 | 7.65e-68 | 22066 | 6061 | 16005 | 0.6497 | 0.03349 | 1.915 | 8.04e-84 | 30513 | 6319 | 24194 | 0.01211 | 0.1098 | Electrolyte imbalance | endocrine/metabolic | LDL-C | |
| 276.11 | | | | | 16449 | 444 | 16005 | | | | | 24509 | 315 | 24194 | 0.312 | 0.1545 | Hyperosmolality and/or hypernatremia | endocrine/metabolic | LDL-C | |
| 276.12 | 0.6076 | 0.05325 | 1.836 | 3.74e-30 | 18173 | 2168 | 16005 | 0.7701 | 0.05471 | 2.16 | 5.21e-45 | 26321 | 2127 | 24194 | 5.01e-05 | 0.2789 | Hyposmolality and/or hyponatremia | endocrine/metabolic | LDL-C | |
| 276.13 | 1.156 | 0.05938 | 3.177 | 2.10e-84 | 18167 | 2162 | 16005 | 1.304 | 0.06893 | 3.684 | 8.10e-80 | 25804 | 1610 | 24194 | 0.007864 | 0.2222 | Hyperpotassemia | endocrine/metabolic | LDL-C | |
| 276.14 | 0.5901 | 0.04836 | 1.804 | 2.98e-34 | 18672 | 2667 | 16005 | 0.5995 | 0.04153 | 1.821 | 3.13e-47 | 27767 | 3573 | 24194 | 0.2302 | 0.06905 | Hypopotassemia | endocrine/metabolic | LDL-C | |
| 276.4 | 0.7795 | 0.05487 | 2.18 | 8.46e-46 | 18109 | 2104 | 16005 | 0.7417 | 0.05642 | 2.099 | 1.83e-39 | 26022 | 1828 | 24194 | 0.9196 | 0.007217 | Acid-base balance disorder | endocrine/metabolic | LDL-C | |
| 276.41 | 0.8258 | 0.05834 | 2.284 | 1.73e-45 | 17879 | 1874 | 16005 | 0.808 | 0.06064 | 2.243 | 1.67e-40 | 25798 | 1604 | 24194 | 0.6406 | 0.03579 | Acidosis | endocrine/metabolic | LDL-C | |
| 276.42 | | | | | 16255 | 250 | 16005 | | | | | 24456 | 262 | 24194 | 0.4828 | -0.127 | Alkalosis | endocrine/metabolic | LDL-C | |
| 276.5 | 0.435 | 0.04426 | 1.545 | 8.58e-23 | 19178 | 3173 | 16005 | 0.5587 | 0.04172 | 1.748 | 6.89e-41 | 27587 | 3393 | 24194 | 3.33e-04 | 0.198 | Hypovolemia | endocrine/metabolic | LDL-C | |
| 276.6 | 1.093 | 0.06386 | 2.985 | 9.73e-66 | 17709 | 1704 | 16005 | 1.056 | 0.06701 | 2.874 | 6.26e-56 | 25588 | 1394 | 24194 | 0.7086 | -0.03143 | Fluid overload | endocrine/metabolic | LDL-C | |
| 276.8 | | | | | 16014 | 9 | 16005 | | | | | 24216 | 22 | 24194 | | | Polydipsia | endocrine/metabolic | LDL-C | |
| 277 | | | | | 26131 | 420 | 25711 | 0.9512 | 0.1101 | 2.589 | 5.69e-18 | 35099 | 554 | 34545 | 0.08581 | 0.265 | Other disorders of metabolism | endocrine/metabolic | LDL-C | |
| 277.1 | | | | | 25722 | 11 | 25711 | | | | | 34554 | 9 | 34545 | | | Disorders of porphyrin metabolism | endocrine/metabolic | LDL-C | |
| 277.2 | | | | | 25713 | 2 | 25711 | | | | | 34547 | 2 | 34545 | | | Other disorders of purine and pyrimidine metabolism | endocrine/metabolic | LDL-C | |
| 277.4 | | | | | 25910 | 199 | 25711 | | | | | 34665 | 120 | 34545 | | | Disorders of bilirubin excretion | endocrine/metabolic | LDL-C | |
| 277.5 | | | | | 25845 | 134 | 25711 | | | | | 34656 | 111 | 34545 | | | Other disorders of lipoid metabolism | endocrine/metabolic | LDL-C | |
| 277.51 | | | | | 25790 | 79 | 25711 | | | | | 34590 | 45 | 34545 | | | Lipoprotein disorders | endocrine/metabolic | LDL-C | |
| 277.6 | | | | | 25723 | 12 | 25711 | | | | | 34560 | 15 | 34545 | | | Other deficiencies of circulating enzymes | endocrine/metabolic | LDL-C | |
| 277.7 | | | | | 25918 | 207 | 25711 | | | | | 34828 | 283 | 34545 | | | Dysmetabolic syndrome X | endocrine/metabolic | LDL-C | |
| 277.8 | | | | | 25715 | 4 | 25711 | | | | | 34546 | 1 | 34545 | | | Carnitine deficiencies | endocrine/metabolic | LDL-C | |
| 278 | 0.9927 | 0.04375 | 2.698 | 5.91e-114 | 24739 | 3939 | 20800 | 1.005 | 0.03276 | 2.731 | 1.85e-206 | 32627 | 6840 | 25787 | 0.7561 | -0.01572 | Overweight, obesity and other hyperalimentation | endocrine/metabolic | LDL-C | |
| 278.1 | 1.074 | 0.0479 | 2.927 | 2.61e-111 | 24116 | 3316 | 20800 | 0.9835 | 0.03553 | 2.674 | 1.25e-168 | 31306 | 5519 | 25787 | 0.02505 | -0.1239 | Obesity | endocrine/metabolic | LDL-C | |
| 278.11 | 1.003 | 0.06567 | 2.726 | 1.22e-52 | 22371 | 1571 | 20800 | 1.009 | 0.04482 | 2.743 | 3.30e-112 | 28985 | 3198 | 25787 | 0.695 | -0.02889 | Morbid obesity | endocrine/metabolic | LDL-C | |
| 278.3 | | | | | 20837 | 37 | 20800 | | | | | 25988 | 201 | 25787 | | | Localized adiposity | endocrine/metabolic | LDL-C | |

| phenotype | male_beta | male_SE | male_OR | male_p | male_n_total | male_n_cases | male_n_controls | female_beta | female_SE | female_OR | female_p | female_n_total | female_n_cases | female_n_controls | interaction_p | interaction_beta | description | group | lab | label |
|---|---|---|---|---|---|---|---|---|---|---|---|---|---|---|---|---|---|---|---|---|
| 278.4 | | | | | 21218 | 418 | 20800 | 2.002 | 0.08367 | 7.4 | 1.82e-126 | 27263 | 1476 | 25787 | 0.7774 | 0.05028 | Abnormal weight gain | endocrine/metabolic | LDL-C | |
| 279 | 1.125 | 0.08337 | 3.079 | 1.80e-41 | 25738 | 1171 | 24567 | 0.808 | 0.06782 | 2.243 | 1.01e-32 | 33388 | 1505 | 31883 | 0.03889 | -0.2089 | Disorders involving the immune mechanism | endocrine/metabolic | LDL-C | |
| 279.1 | 1.147 | 0.08697 | 3.147 | 1.10e-39 | 25668 | 1101 | 24567 | 0.8879 | 0.07231 | 2.43 | 1.18e-34 | 33274 | 1391 | 31883 | 0.1511 | -0.1532 | Immunity deficiency | endocrine/metabolic | LDL-C | |
| 279.11 | | | | | 24666 | 99 | 24567 | | | | | 32052 | 169 | 31883 | | | Deficiency of humoral immunity | endocrine/metabolic | LDL-C | |
| 279.2 | | | | | 24578 | 11 | 24567 | | | | | 31914 | 31 | 31883 | | | Autoimmune disease NEC | endocrine/metabolic | LDL-C | |
| 279.7 | | | | | 24823 | 256 | 24567 | 0.2289 | 0.07435 | 1.257 | 0.002074 | 32824 | 941 | 31883 | 8.72e-05 | -0.6613 | Other immunological findings | endocrine/metabolic | LDL-C | |
| 279.8 | | | | | 24598 | 31 | 24567 | | | | | 31935 | 52 | 31883 | | | Other specified disorders involving the immune mechanism | endocrine/metabolic | LDL-C | |
| 280 | 0.8765 | 0.06414 | 2.402 | 1.63e-42 | 18083 | 1688 | 16395 | 0.9958 | 0.04941 | 2.707 | 2.40e-90 | 25891 | 2832 | 23059 | 0.03863 | 0.1557 | Iron deficiency anemias | hematopoietic | LDL-C | |
| 280.1 | 0.9342 | 0.07233 | 2.545 | 3.69e-38 | 17740 | 1345 | 16395 | 1.068 | 0.05413 | 2.91 | 1.17e-86 | 25454 | 2395 | 23059 | 0.07434 | 0.1505 | Iron deficiency anemias, unspecified or not due to blood loss | hematopoietic | LDL-C | |
| 280.2 | 0.8293 | 0.1134 | 2.292 | 2.65e-13 | 16909 | 514 | 16395 | 0.943 | 0.09137 | 2.568 | 5.66e-25 | 23821 | 762 | 23059 | 0.2949 | 0.1422 | Iron deficiency anemia secondary to blood loss (chronic) | hematopoietic | LDL-C | |
| 281 | | | | | 16870 | 475 | 16395 | 1.195 | 0.09018 | 3.302 | 4.71e-40 | 24011 | 952 | 23059 | 0.4106 | 0.1214 | Other deficiency anemia | hematopoietic | LDL-C | |
| 281.1 | | | | | 16669 | 274 | 16395 | 1.336 | 0.1119 | 3.803 | 7.38e-33 | 23736 | 677 | 23059 | 0.9874 | -0.003214 | Megaloblastic anemia | hematopoietic | LDL-C | |
| 281.11 | | | | | 16464 | 69 | 16395 | | | | | 23278 | 219 | 23059 | | | Pernicious anemia | hematopoietic | LDL-C | |
| 281.12 | | | | | 16590 | 195 | 16395 | 1.34 | 0.1293 | 3.818 | 3.73e-25 | 23561 | 502 | 23059 | 0.9187 | -0.02474 | Other vitamin B12 deficiency anemia | hematopoietic | LDL-C | |
| 281.13 | | | | | 16415 | 20 | 16395 | | | | | 23107 | 48 | 23059 | | | Folate-deficiency anemia | hematopoietic | LDL-C | |
| 281.9 | | | | | 16581 | 186 | 16395 | | | | | 23292 | 233 | 23059 | | | Deficiency anemias | hematopoietic | LDL-C | |
| 282 | | | | | 16442 | 47 | 16395 | | | | | 23190 | 131 | 23059 | | | Hereditary hemolytic anemias | hematopoietic | LDL-C | |
| 282.5 | | | | | 16399 | 4 | 16395 | | | | | 23069 | 10 | 23059 | | | Sickle cell anemia | hematopoietic | LDL-C | |
| 282.8 | | | | | 16410 | 15 | 16395 | | | | | 23102 | 43 | 23059 | | | Other hemoglobinopathies | hematopoietic | LDL-C | |
| 282.9 | | | | | 16427 | 32 | 16395 | | | | | 23140 | 81 | 23059 | | | Other hereditary hemolytic anemias | hematopoietic | LDL-C | |
| 283 | | | | | 16455 | 60 | 16395 | | | | | 23168 | 109 | 23059 | | | Acquired hemolytic anemias | hematopoietic | LDL-C | |
| 283.1 | | | | | 16418 | 23 | 16395 | | | | | 23095 | 36 | 23059 | | | Autoimmune hemolytic anemias | hematopoietic | LDL-C | |
| 283.2 | | | | | 16419 | 24 | 16395 | | | | | 23100 | 41 | 23059 | | | Non-autoimmune hemolytic anemias | hematopoietic | LDL-C | |
| 283.21 | | | | | 16402 | 7 | 16395 | | | | | 23073 | 14 | 23059 | | | Hemolytic-uremic syndrome | hematopoietic | LDL-C | |
| 284 | 0.3939 | 0.07899 | 1.483 | 6.15e-07 | 17250 | 855 | 16395 | 0.3796 | 0.08486 | 1.462 | 7.70e-06 | 23771 | 712 | 23059 | 0.6055 | -0.05398 | Aplastic anemia | hematopoietic | LDL-C | |
| 284.1 | 0.4861 | 0.08417 | 1.626 | 7.69e-09 | 17146 | 751 | 16395 | 0.4051 | 0.08931 | 1.499 | 5.74e-06 | 23702 | 643 | 23059 | 0.583 | -0.06073 | Pancytopenia | hematopoietic | LDL-C | |
| 284.2 | | | | | 16406 | 11 | 16395 | | | | | 23065 | 6 | 23059 | | | Constitutional aplastic anemia | hematopoietic | LDL-C | |
| 285 | 0.4455 | 0.03242 | 1.561 | 5.86e-43 | 23977 | 7582 | 16395 | 0.5857 | 0.0303 | 1.796 | 3.19e-83 | 30991 | 7932 | 23059 | 2.58e-04 | 0.1459 | Other anemias | hematopoietic | LDL-C | |
| 285.1 | 0.3358 | 0.04608 | 1.399 | 3.17e-13 | 19293 | 2898 | 16395 | 0.4317 | 0.04987 | 1.54 | 4.84e-18 | 25340 | 2281 | 23059 | 0.154 | 0.08737 | Acute posthemorrhagic anemia | hematopoietic | LDL-C | |
| 285.2 | 0.6267 | 0.05197 | 1.871 | 1.74e-33 | 18710 | 2315 | 16395 | 0.6765 | 0.05145 | 1.967 | 1.71e-39 | 25252 | 2193 | 23059 | 0.8861 | -0.009423 | Anemia of chronic disease | hematopoietic | LDL-C | |
| 285.21 | 1.656 | 0.1109 | 5.238 | 2.10e-50 | 17283 | 888 | 16395 | 1.784 | 0.1195 | 5.952 | 2.26e-50 | 23775 | 716 | 23059 | 0.5779 | 0.08684 | Anemia in chronic kidney disease | hematopoietic | LDL-C | |
| phenotype | male_beta | male_SE | male_OR | male_p | male_n_total | male_n_cases | male_n_controls | female_beta | female_SE | female_OR | female_p | female_n_total | female_n_cases | female_n_controls | interaction_p | interaction_beta | description | group | lab | label |
| 285.22 | 0.02441 | 0.07014 | 1.025 | 0.7278 | 17510 | 1115 | 16395 | 0.2373 | 0.06628 | 1.268 | 3.44e-04 | 24273 | 1214 | 23059 | 0.0323 | 0.1858 | Anemia in neoplastic disease | hematopoietic | LDL-C | |
| 285.3 | | | | | 16401 | 6 | 16395 | | | | | 23062 | 3 | 23059 | | | Sideroblastic anemia | hematopoietic | LDL-C | |
| 285.8 | | | | | 16399 | 4 | 16395 | | | | | 23072 | 13 | 23059 | | | Hemoglobinuria | hematopoietic | LDL-C | |

| phenotype | male_beta | male_SE | male_OR | male_p | male_n_total | male_n_cases | male_n_controls | female_beta | female_SE | female_OR | female_p | female_n_total | female_n_cases | female_n_controls | interaction_p | interaction_beta | description | group | lab | label |
|---|---|---|---|---|---|---|---|---|---|---|---|---|---|---|---|---|---|---|---|---|
| 286 | 0.6866 | 0.05822 | 1.987 | 4.23e-32 | 21087 | 1817 | 19270 | 0.5881 | 0.05404 | 1.8 | 1.41e-27 | 30311 | 1937 | 28374 | 0.1637 | -0.1008 | Coagulation defects | hematopoietic | LDL-C | |
| 286.1 | | | | | 19382 | 112 | 19270 | | | | | 28488 | 114 | 28374 | | | Congenital coagulation defects | hematopoietic | LDL-C | |
| 286.11 | | | | | 19306 | 36 | 19270 | | | | | 28422 | 48 | 28374 | | | Von willebrand's disease | hematopoietic | LDL-C | |
| 286.12 | | | | | 19305 | 35 | 19270 | | | | | 28432 | 58 | 28374 | | | Congenital deficiency of other clotting factors (including factor VII) | hematopoietic | LDL-C | |
| 286.13 | | | | | 19324 | 54 | 19270 | | | | | 28385 | 11 | 28374 | | | Congenital factor VIII disorder | hematopoietic | LDL-C | |
| 286.2 | 0.9255 | 0.05024 | 2.523 | 9.07e-76 | 22410 | 3140 | 19270 | 0.7721 | 0.05045 | 2.164 | 7.29e-53 | 31115 | 2741 | 28374 | 0.01926 | -0.1533 | Encounter for long-term (current) use of anticoagulants | hematopoietic | LDL-C | |
| 286.3 | | | | | 6 | | | | | | | 28478 | 106 | 28372 | | | Coagulation defects complicating pregnancy or postpartum | hematopoietic | LDL-C | |
| 286.4 | | | | | 19353 | 83 | 19270 | | | | | 28429 | 55 | 28374 | | | Acquired coagulation factor deficiency | hematopoietic | LDL-C | |
| 286.5 | | | | | 19388 | 118 | 19270 | | | | | 28511 | 137 | 28374 | | | Hemorrhagic disorder due to intrinsic circulating anticoagulants | hematopoietic | LDL-C | |
| 286.6 | | | | | 19312 | 42 | 19270 | | | | | 28420 | 46 | 28374 | | | Defibrination syndrome | hematopoietic | LDL-C | |
| 286.7 | 0.7115 | 0.08086 | 2.037 | 1.38e-18 | 20156 | 886 | 19270 | 0.5456 | 0.07518 | 1.726 | 3.95e-13 | 29323 | 949 | 28374 | 0.2027 | -0.128 | Other and unspecified coagulation defects | hematopoietic | LDL-C | |
| 286.8 | | | | | 19460 | 190 | 19270 | | | | | 28789 | 415 | 28374 | 0.111 | -0.3167 | Hypercoagulable state | hematopoietic | LDL-C | |
| 286.81 | | | | | 19445 | 175 | 19270 | | | | | 28747 | 373 | 28374 | 0.2557 | -0.2327 | Primary hypercoagulable state | hematopoietic | LDL-C | |
| 286.9 | | | | | 19741 | 471 | 19270 | | | | | 28784 | 410 | 28374 | 0.06151 | -0.2901 | Abnormal coagulation profile | hematopoietic | LDL-C | |
| 287 | 0.465 | 0.05044 | 1.592 | 2.98e-20 | 21610 | 2340 | 19270 | 0.4479 | 0.05385 | 1.565 | 8.96e-17 | 30271 | 1897 | 28374 | 0.1543 | -0.09419 | Purpura and other hemorrhagic conditions | hematopoietic | LDL-C | |
| 287.1 | | | | | 19326 | 56 | 19270 | | | | | 28485 | 111 | 28374 | | | Spontaneous ecchymoses | hematopoietic | LDL-C | |
| 287.2 | | | | | 19288 | 18 | 19270 | | | | | 28390 | 16 | 28374 | | | Allergic purpura | hematopoietic | LDL-C | |
| 287.3 | 0.4404 | 0.05178 | 1.553 | 1.79e-17 | 21432 | 2162 | 19270 | 0.4031 | 0.05588 | 1.496 | 5.47e-13 | 30102 | 1728 | 28374 | 0.1123 | -0.1085 | Thrombocytopenia | hematopoietic | LDL-C | |
| 287.31 | | | | | 19391 | 121 | 19270 | | | | | 28538 | 164 | 28374 | | | Primary thrombocytopenia | hematopoietic | LDL-C | |
| 287.32 | | | | | 19594 | 324 | 19270 | | | | | 28675 | 301 | 28374 | 0.3228 | 0.1603 | Secondary thrombocytopenia | hematopoietic | LDL-C | |
| 287.4 | | | | | 19290 | 20 | 19270 | | | | | 28393 | 19 | 28374 | | | Qualitative platelet defects | hematopoietic | LDL-C | |
| 288 | 0.365 | 0.04611 | 1.44 | 2.46e-15 | 21474 | 2790 | 18684 | 0.5239 | 0.04583 | 1.689 | 2.90e-30 | 29549 | 2753 | 26796 | 1.45e-04 | 0.2235 | Diseases of white blood cells | hematopoietic | LDL-C | |
| 288.1 | 0.2844 | 0.06941 | 1.329 | 4.17e-05 | 19788 | 1104 | 18684 | 0.3996 | 0.06237 | 1.491 | 1.49e-10 | 28156 | 1360 | 26796 | 0.01584 | 0.2033 | Decreased white blood cell count | hematopoietic | LDL-C | |
| 288.11 | 0.08569 | 0.0791 | 1.089 | 0.2786 | 19506 | 822 | 18684 | 0.2247 | 0.07088 | 1.252 | 0.001527 | 27815 | 1019 | 26796 | 0.01784 | 0.2272 | Neutropenia | hematopoietic | LDL-C | |
| 288.2 | 0.3608 | 0.04991 | 1.434 | 4.90e-13 | 21008 | 2324 | 18684 | 0.5567 | 0.05025 | 1.745 | 1.58e-28 | 29055 | 2259 | 26796 | 3.97e-05 | 0.2629 | Elevated white blood cell count | hematopoietic | LDL-C | |
| 288.3 | | | | | 18778 | 94 | 18684 | | | | | 26909 | 113 | 26796 | | | Eosinophilia | hematopoietic | LDL-C | |
| 289 | | | | | 19172 | 488 | 18684 | 0.6469 | 0.09623 | 1.91 | 1.79e-11 | 27421 | 625 | 26796 | 0.979 | -0.003568 | Other diseases of blood and blood-forming organs | hematopoietic | LDL-C | |
| 289.1 | | | | | 18755 | 71 | 18684 | | | | | 26856 | 60 | 26796 | | | Myelofibrosis | hematopoietic | LDL-C | |
| 289.3 | | | | | 18767 | 83 | 18684 | | | | | 26919 | 123 | 26796 | | | Personal history of diseases of blood and blood-forming organs | hematopoietic | LDL-C | |
| 289.4 | 0.1754 | 0.06046 | 1.192 | 0.003712 | 20171 | 1487 | 18684 | 0.4506 | 0.05778 | 1.569 | 6.32e-15 | 28443 | 1647 | 26796 | 4.36e-07 | 0.3824 | Lymphadenitis | hematopoietic | LDL-C | |
| 289.5 | | | | | 18840 | 156 | 18684 | | | | | 26948 | 152 | 26796 | | | Diseases of spleen | hematopoietic | LDL-C | |
| 289.8 | | | | | 18685 | 237 | 18448 | | | | | 26687 | 169 | 26518 | | | Polycythemia, secondary | hematopoietic | LDL-C | |
| 289.9 | | | | | 18788 | 104 | 18684 | | | | | 26914 | 118 | 26796 | | | Abnormality of red blood cells | hematopoietic | LDL-C | |
| 290 | 0.8215 | 0.07383 | 2.274 | 9.31e-29 | 21683 | 1272 | 20411 | 0.9899 | 0.07785 | 2.691 | 4.79e-37 | 30218 | 1222 | 28996 | 0.09689 | 0.1613 | Delirium dementia and amnestic and other cognitive disorders | mental disorders | LDL-C | |
| 290.1 | 0.8676 | 0.09623 | 2.381 | 1.96e-19 | 21205 | 794 | 20411 | 1.172 | 0.09886 | 3.23 | 1.93e-32 | 29834 | 838 | 28996 | 0.2239 | 0.1534 | Dementias | mental disorders | LDL-C | |
| 290.11 | | | | | 20682 | 271 | 20411 | | | | | 29352 | 356 | 28996 | 0.7289 | -0.06919 | Alzheimer's disease | mental disorders | LDL-C | |

| phenotype | male_beta | male_SE | male_OR | male_p | male_n_total | male_n_cases | male_n_controls | female_beta | female_SE | female_OR | female_p | female_n_total | female_n_cases | female_n_controls | interaction_p | interaction_beta | description | group | lab | label |
|---|---|---|---|---|---|---|---|---|---|---|---|---|---|---|---|---|---|---|---|---|
| 290.12 | | | | | 20506 | 95 | 20411 | | | | | 29043 | 47 | 28996 | | | Dementia with cerebral degenerations | mental disorders | LDL-C | |
| 290.13 | | | | | 20487 | 76 | 20411 | | | | | 29129 | 133 | 28996 | | | Senile dementia | mental disorders | LDL-C | |
| 290.16 | | | | | 20543 | 132 | 20411 | | | | | 29107 | 111 | 28996 | | | Vascular dementia | mental disorders | LDL-C | |
| 290.2 | | | | | 20877 | 466 | 20411 | | | | | 29341 | 345 | 28996 | 0.4492 | 0.1181 | Delirium due to conditions classified elsewhere | mental disorders | LDL-C | |
| 290.3 | | | | | 20762 | 351 | 20411 | | | | | 29366 | 370 | 28996 | 0.6757 | -0.06973 | Other persistent mental disorders due to conditions classified elsewhere | mental disorders | LDL-C | |
| 291 | 0.4352 | 0.06511 | 1.545 | 2.33e-11 | 21730 | 1319 | 20411 | 0.4674 | 0.06493 | 1.596 | 6.08e-13 | 30293 | 1297 | 28996 | 0.1414 | 0.1211 | Other specified nonpsychotic and/or transient mental disorders | mental disorders | LDL-C | |
| 291.1 | | | | | 20440 | 29 | 20411 | | | | | 29037 | 41 | 28996 | | | Transient mental disorders due to conditions classified elsewhere | mental disorders | LDL-C | |
| 291.4 | | | | | 20576 | 165 | 20411 | | | | | 29194 | 198 | 28996 | | | Specific nonpsychotic mental disorders due to brain damage | mental disorders | LDL-C | |
| 291.8 | 0.4915 | 0.07015 | 1.635 | 2.47e-12 | 21537 | 1126 | 20411 | 0.4668 | 0.07171 | 1.595 | 7.57e-11 | 30045 | 1049 | 28996 | 0.5028 | 0.06006 | Alteration of consciousness | mental disorders | LDL-C | |
| 292 | 0.5563 | 0.04176 | 1.744 | 1.77e-40 | 24322 | 3911 | 20411 | 0.6047 | 0.03975 | 1.831 | 2.95e-52 | 33173 | 4177 | 28996 | 0.05602 | 0.09978 | Neurological disorders | mental disorders | LDL-C | |
| 292.1 | 0.8366 | 0.09186 | 2.309 | 8.45e-20 | 21162 | 751 | 20411 | 0.9375 | 0.09235 | 2.554 | 3.24e-24 | 29761 | 765 | 28996 | 0.2275 | 0.1438 | Aphasia/speech disturbance | mental disorders | LDL-C | |
| 292.11 | | | | | 20684 | 273 | 20411 | | | | | 29270 | 274 | 28996 | 0.384 | 0.1796 | Aphasia | mental disorders | LDL-C | |
| 292.12 | | | | | 20439 | 28 | 20411 | | | | | 29020 | 24 | 28996 | | | Symbolic dysfunction | mental disorders | LDL-C | |
| 292.2 | | | | | 20778 | 367 | 20411 | | | | | 29334 | 338 | 28996 | 0.5867 | 0.1086 | Mild cognitive impairment | mental disorders | LDL-C | |
| 292.3 | 0.7887 | 0.09149 | 2.201 | 6.62e-18 | 21327 | 916 | 20411 | 0.9621 | 0.08293 | 2.617 | 4.08e-31 | 30127 | 1131 | 28996 | 0.1808 | 0.1535 | Memory loss | mental disorders | LDL-C | |
| 292.4 | 0.5398 | 0.0511 | 1.716 | 4.32e-26 | 22762 | 2351 | 20411 | 0.5464 | 0.0513 | 1.727 | 1.72e-26 | 31254 | 2258 | 28996 | 0.3626 | 0.05917 | Altered mental status | mental disorders | LDL-C | |
| 292.5 | | | | | 20547 | 136 | 20411 | | | | | 29184 | 188 | 28996 | | | Transient alteration of awareness | mental disorders | LDL-C | |
| 292.6 | | | | | 20491 | 80 | 20411 | | | | | 29129 | 133 | 28996 | | | Hallucinations | mental disorders | LDL-C | |
| 293 | | | | | 25542 | 80 | 25462 | | | | | 34286 | 108 | 34178 | | | Symptoms involving head and neck | mental disorders | LDL-C | |
| 293.1 | -0.03045 | 0.07683 | 0.97 | 0.6919 | 26351 | 889 | 25462 | 0.218 | 0.07822 | 1.244 | 0.005313 | 35053 | 875 | 34178 | 3.41e-05 | 0.4078 | Swelling, mass, or lump in head and neck [Space-occupying lesion, intracranial NOS] | mental disorders | LDL-C | |
| 295 | | | | | 17073 | 479 | 16594 | 1.143 | 0.1039 | 3.135 | 4.11e-28 | 19345 | 524 | 18821 | 0.05426 | 0.2733 | Schizophrenia and other psychotic disorders | mental disorders | LDL-C | |
| 295.1 | | | | | 16740 | 146 | 16594 | | | | | 18981 | 160 | 18821 | | | Schizophrenia | mental disorders | LDL-C | |
| 295.2 | | | | | 16700 | 106 | 16594 | | | | | 18968 | 147 | 18821 | | | Paranoid disorders | mental disorders | LDL-C | |
| 295.3 | | | | | 16878 | 284 | 16594 | | | | | 19094 | 273 | 18821 | 0.9172 | 0.01878 | Psychosis | mental disorders | LDL-C | |
| 296 | 1.038 | 0.04054 | 2.824 | 1.24e-144 | 21203 | 4609 | 16594 | 1.114 | 0.03134 | 3.047 | 8.69e-277 | 27169 | 8348 | 18821 | 1.49e-07 | 0.2482 | Mood disorders | mental disorders | LDL-C | |
| 296.1 | 1.274 | 0.08994 | 3.576 | 1.47e-45 | 17404 | 810 | 16594 | 0.9427 | 0.06349 | 2.567 | 7.25e-50 | 20203 | 1382 | 18821 | 0.418 | -0.08248 | Bipolar | mental disorders | LDL-C | |
| 296.2 | 1.064 | 0.04371 | 2.898 | 6.70e-131 | 20515 | 3921 | 16594 | 1.182 | 0.03304 | 3.26 | 3.31e-280 | 26233 | 7412 | 18821 | 2.27e-07 | 0.2623 | Depression | mental disorders | LDL-C | |

| phenotype | male_beta | male_SE | male_OR | male_p | male_n_total | male_n_cases | male_n_controls | female_beta | female_SE | female_OR | female_p | female_n_total | female_n_cases | female_n_controls | interaction_p | interaction_beta | description | group | lab | label |
|---|---|---|---|---|---|---|---|---|---|---|---|---|---|---|---|---|---|---|---|---|
| 296.22 | 1.225 | 0.05951 | 3.403 | 4.02e-94 | 18825 | 2231 | 16594 | 1.268 | 0.04184 | 3.553 | 1.08e-201 | 23232 | 4411 | 18821 | 0.0268 | 0.1503 | Major depressive disorder | mental disorders | LDL-C | |
| 297 | | | | | 17005 | 411 | 16594 | 0.7494 | 0.09964 | 2.116 | 5.44e-14 | 19321 | 500 | 18821 | 0.04918 | -0.2795 | Suicidal ideation or attempt | mental disorders | LDL-C | |
| 297.1 | | | | | 16944 | 350 | 16594 | | | | | 19222 | 401 | 18821 | 0.09401 | -0.263 | Suicidal ideation | mental disorders | LDL-C | |
| 297.2 | | | | | 16666 | 72 | 16594 | | | | | 18942 | 121 | 18821 | | | Suicide or self-inflicted injury | mental disorders | LDL-C | |
| 300 | 1.062 | 0.04364 | 2.891 | 1.04e-130 | 20603 | 4009 | 16594 | 1.154 | 0.03186 | 3.17 | 2.81e-287 | 27035 | 8214 | 18821 | 2.16e-07 | 0.2572 | Anxiety disorders | mental disorders | LDL-C | |
| 300.1 | 1.103 | 0.04671 | 3.014 | 2.36e-123 | 20057 | 3463 | 16594 | 1.217 | 0.03364 | 3.376 | 2.05e-286 | 26048 | 7227 | 18821 | 3.08e-08 | 0.2935 | Anxiety disorder | mental disorders | LDL-C | |
| 300.11 | 1.58 | 0.1077 | 4.857 | 8.69e-49 | 17284 | 690 | 16594 | 1.54 | 0.06819 | 4.663 | 6.75e-113 | 20323 | 1502 | 18821 | 0.3757 | 0.105 | Generalized anxiety disorder | mental disorders | LDL-C | |
| 300.12 | | | | | 16882 | 288 | 16594 | 1.265 | 0.09937 | 3.543 | 3.97e-37 | 19405 | 584 | 18821 | 0.09401 | -0.3033 | Agorophobia, social phobia, and panic disorder | mental disorders | LDL-C | |
| 300.13 | | | | | 16650 | 56 | 16594 | | | | | 18930 | 109 | 18821 | | | Phobia | mental disorders | LDL-C | |
| 300.2 | | | | | 16594 | | | | | | | 18821 | | | | | Generalized anxiety & phobic disorders | mental disorders | LDL-C | |
| 300.3 | | | | | 16794 | 200 | 16594 | | | | | 19097 | 276 | 18821 | | | Obsessive-compulsive disorders | mental disorders | LDL-C | |
| 300.4 | | | | | 16995 | 401 | 16594 | 1.457 | 0.09072 | 4.295 | 4.46e-58 | 19615 | 794 | 18821 | 0.1663 | 0.2099 | Dysthymic disorder | mental disorders | LDL-C | |
| 300.8 | | | | | 16639 | 45 | 16594 | | | | | 18944 | 123 | 18821 | | | Acute reaction to stress | mental disorders | LDL-C | |
| 300.9 | | | | | 16936 | 342 | 16594 | 0.8356 | 0.07449 | 2.306 | 3.32e-29 | 19768 | 947 | 18821 | 0.5536 | -0.08062 | Posttraumatic stress disorder | mental disorders | LDL-C | |
| 301 | | | | | 16724 | 130 | 16594 | | | | | 19101 | 280 | 18821 | | | Personality disorders | mental disorders | LDL-C | |
| 301.1 | | | | | 16605 | 11 | 16594 | | | | | 18829 | 8 | 18821 | | | Schizoid personality disorder | mental disorders | LDL-C | |
| 301.2 | | | | | 16645 | 51 | 16594 | | | | | 19000 | 179 | 18821 | | | Antisocial/borderline personality disorder | mental disorders | LDL-C | |
| 302 | | | | | 16787 | 193 | 16594 | | | | | 18953 | 132 | 18821 | | | Sexual and gender identity disorders | mental disorders | LDL-C | |
| 302.1 | | | | | 16728 | 134 | 16594 | | | | | 18905 | 84 | 18821 | | | Decreased libido | mental disorders | LDL-C | |
| 303 | | | | | 16730 | 136 | 16594 | | | | | 19170 | 349 | 18821 | | | Psychogenic and somatoform disorders | mental disorders | LDL-C | |
| 303.1 | | | | | 16603 | 9 | 16594 | | | | | 18849 | 28 | 18821 | | | Dissociative disorder | mental disorders | LDL-C | |
| 303.3 | | | | | 16631 | 37 | 16594 | | | | | 18918 | 97 | 18821 | | | Psychogenic disorder | mental disorders | LDL-C | |
| 303.31 | | | | | 16597 | 3 | 16594 | | | | | 18830 | 9 | 18821 | | | Gastrointestinal malfunction arising from mental factors | mental disorders | LDL-C | |
| 303.4 | | | | | 16689 | 95 | 16594 | | | | | 19064 | 243 | 18821 | | | Somatoform disorder | mental disorders | LDL-C | |
| 304 | 0.9516 | 0.07959 | 2.59 | 6.06e-33 | 17547 | 953 | 16594 | 1.362 | 0.06115 | 3.903 | 7.20e-110 | 20609 | 1788 | 18821 | 1.84e-11 | 0.6231 | Adjustment reaction | mental disorders | LDL-C | |
| 305.2 | | | | | 16644 | 50 | 16594 | | | | | 19240 | 419 | 18821 | | | Eating disorder | mental disorders | LDL-C | |
| 305.21 | | | | | 16607 | 13 | 16594 | | | | | 18929 | 108 | 18821 | | | Anorexia nervosa | mental disorders | LDL-C | |
| 306 | 0.7988 | 0.07947 | 2.223 | 9.03e-24 | 17668 | 1074 | 16594 | 1.15 | 0.06327 | 3.16 | 6.79e-74 | 20481 | 1660 | 18821 | 3.15e-04 | 0.3455 | Other mental disorder | mental disorders | LDL-C | |
| 306.1 | | | | | 0 | | | | | | | 19216 | 395 | 18821 | | | Mental disorders durring/after pregnancy | mental disorders | LDL-C | |

| phenotype | male_beta | male_SE | male_OR | male_p | male_n_total | male_n_cases | male_n_controls | female_beta | female_SE | female_OR | female_p | female_n_total | female_n_cases | female_n_controls | interaction_p | interaction_beta | description | group | lab | label |
|---|---|---|---|---|---|---|---|---|---|---|---|---|---|---|---|---|---|---|---|---|
| 306.9 | | | | | 16626 | 32 | 16594 | | | | | 18928 | 107 | 18821 | | | Tension headache | mental disorders | LDL-C | |
| 312 | | | | | 25880 | 213 | 25667 | | | | | 34967 | 157 | 34810 | | | Conduct disorders | mental disorders | LDL-C | |
| 312.3 | | | | | 25729 | 62 | 25667 | | | | | 34851 | 41 | 34810 | | | Impulse control disorder | mental disorders | LDL-C | |
| 313 | 0.786 | 0.09455 | 2.194 | 9.37e-17 | 26378 | 711 | 25667 | 0.7485 | 0.0855 | 2.114 | 2.04e-18 | 35536 | 726 | 34810 | 0.03986 | 0.234 | Pervasive developmental disorders | mental disorders | LDL-C | |
| 313.1 | 0.8254 | 0.109 | 2.283 | 3.68e-14 | 26178 | 511 | 25667 | 0.6914 | 0.09754 | 1.996 | 1.36e-12 | 35352 | 542 | 34810 | 0.3698 | 0.1174 | Attention deficit hyperactivity disorder | mental disorders | LDL-C | |
| 313.2 | | | | | 25726 | 59 | 25667 | | | | | 34849 | 39 | 34810 | | | Tics and stuttering | mental disorders | LDL-C | |
| 313.3 | | | | | 25854 | 187 | 25667 | | | | | 34891 | 81 | 34810 | | | Autism | mental disorders | LDL-C | |
| 315 | -0.09533 | 0.09629 | 0.9091 | 0.3221 | 26237 | 570 | 25667 | | | | | 35246 | 436 | 34810 | 0.4728 | -0.0936 | Develomental delays and disorders | mental disorders | LDL-C | |
| 315.1 | | | | | 25693 | 26 | 25667 | | | | | 34830 | 20 | 34810 | | | Learning disorder | mental disorders | LDL-C | |
| 315.2 | | | | | 25753 | 86 | 25667 | | | | | 34881 | 71 | 34810 | | | Speech and language disorder | mental disorders | LDL-C | |
| 315.3 | | | | | 25849 | 182 | 25667 | | | | | 34981 | 171 | 34810 | | | Mental retardation | mental disorders | LDL-C | |
| 316 | 0.6185 | 0.06457 | 1.856 | 9.76e-22 | 19622 | 1399 | 18223 | 0.1052 | 0.05461 | 1.111 | 0.05395 | 29618 | 1741 | 27877 | 1.68e-07 | -0.4051 | Substance addiction and disorders | mental disorders | LDL-C | |
| 316.1 | | | | | 18355 | 132 | 18223 | | | | | 28169 | 292 | 27877 | | | Polyneuropathy due to drugs | mental disorders | LDL-C | |
| 317 | 0.5243 | 0.06471 | 1.689 | 5.43e-16 | 19570 | 1347 | 18223 | 0.4334 | 0.08981 | 1.542 | 1.40e-06 | 28525 | 648 | 27877 | 0.9667 | 0.004228 | Alcohol-related disorders | mental disorders | LDL-C | |
| 317.1 | 0.4476 | 0.07454 | 1.565 | 1.92e-09 | 19193 | 970 | 18223 | | | | | 28321 | 444 | 27877 | 0.5166 | 0.07747 | Alcoholism | mental disorders | LDL-C | |
| 317.11 | | | | | 18678 | 455 | 18223 | | | | | 28037 | 160 | 27877 | 0.6343 | 0.08819 | Alcoholic liver damage | mental disorders | LDL-C | |
| 318 | 0.5012 | 0.03679 | 1.651 | 2.94e-42 | 23438 | 5215 | 18223 | 0.2817 | 0.03562 | 1.325 | 2.57e-15 | 32604 | 4727 | 27877 | 1.25e-07 | -0.2467 | Tobacco use disorder | mental disorders | LDL-C | |
| 320 | | | | | 27253 | 157 | 27096 | | | | | 36416 | 155 | 36261 | | | Meningitis | neurological | LDL-C | |
| 323 | | | | | 27190 | 94 | 27096 | | | | | 36377 | 116 | 36261 | | | Encephalitis | neurological | LDL-C | |
| 323.2 | | | | | 27111 | 15 | 27096 | | | | | 36289 | 28 | 36261 | | | Acute (transverse) myelitis | neurological | LDL-C | |
| 323.8 | | | | | 27165 | 69 | 27096 | | | | | 36347 | 86 | 36261 | | | Encephalitis, non-infectious | neurological | LDL-C | |
| 324 | | | | | 27283 | 187 | 27096 | | | | | 36381 | 120 | 36261 | | | Other CNS infection and poliomyelitis | neurological | LDL-C | |
| 324.1 | | | | | 27097 | 1 | 27096 | | | | | 36261 | | | | | Jakob-Creutzfeldt disease | neurological | LDL-C | |
| 325 | | | | | 27112 | 16 | 27096 | | | | | 36306 | 45 | 36261 | | | Phlebitis and thrombophlebitis of intracranial venous sinuses | neurological | LDL-C | |
| 327 | 1.11 | 0.05011 | 3.036 | 7.81e-109 | 22679 | 3201 | 19478 | 1.324 | 0.04236 | 3.76 | 1.21e-214 | 31789 | 4772 | 27017 | 1.85e-07 | 0.3225 | Sleep disorders | neurological | LDL-C | |
| phenotype | male_beta | male_SE | male_OR | male_p | male_n_total | male_n_cases | male_n_controls | female_beta | female_SE | female_OR | female_p | female_n_total | female_n_cases | female_n_controls | interaction_p | interaction_beta | description | group | lab | label |
| 327.1 | | | | | 19663 | 185 | 19478 | | | | | 27358 | 341 | 27017 | 0.3058 | 0.2446 | Hypersomnia | neurological | LDL-C | |
| 327.3 | 1.065 | 0.04526 | 2.902 | 1.53e-122 | 23346 | 3868 | 19478 | 1.205 | 0.04849 | 3.337 | 2.46e-136 | 30225 | 3208 | 27017 | 0.001607 | 0.1973 | Sleep apnea | neurological | LDL-C | |
| 327.31 | | | | | 19588 | 110 | 19478 | | | | | 27080 | 63 | 27017 | | | Central/nonobstructive sleep apnea | neurological | LDL-C | |
| 327.32 | 1.139 | 0.04923 | 3.125 | 1.61e-118 | 22855 | 3377 | 19478 | 1.257 | 0.05347 | 3.516 | 2.73e-122 | 29718 | 2701 | 27017 | 0.01455 | 0.168 | Obstructive sleep apnea | neurological | LDL-C | |
| 327.4 | 1.083 | 0.06197 | 2.952 | 2.41e-68 | 21429 | 1951 | 19478 | 1.421 | 0.05292 | 4.14 | 9.70e-159 | 30146 | 3129 | 27017 | 1.32e-10 | 0.4934 | Insomnia | neurological | LDL-C | |
| 327.41 | | | | | 19695 | 217 | 19478 | | | | | 27448 | 431 | 27017 | 0.01438 | 0.6201 | Organic or persistent insomnia | neurological | LDL-C | |
| 327.5 | | | | | 19615 | 137 | 19478 | | | | | 27116 | 99 | 27017 | | | Parasomnia | neurological | LDL-C | |

| phenotype | male_beta | male_SE | male_OR | male_p | male_n_total | male_n_cases | male_n_controls | female_beta | female_SE | female_OR | female_p | female_n_total | female_n_cases | female_n_controls | interaction_p | interaction_beta | description | group | lab | label |
|---|---|---|---|---|---|---|---|---|---|---|---|---|---|---|---|---|---|---|---|---|
| 327.6 | | | | | 19505 | 27 | 19478 | | | | | 27055 | 38 | 27017 | | | Circadian rhythm sleep disorder | neurological | LDL-C | |
| 327.7 | 1.383 | 0.132 | 3.988 | 1.12e-25 | 19996 | 518 | 19478 | 1.223 | 0.08734 | 3.396 | 1.60e-44 | 28011 | 994 | 27017 | 0.4609 | -0.1109 | Sleep related movement disorders | neurological | LDL-C | |
| 327.71 | | | | | 19923 | 445 | 19478 | 1.208 | 0.09238 | 3.347 | 4.44e-39 | 27896 | 879 | 27017 | 0.6547 | -0.0709 | Restless legs syndrome | neurological | LDL-C | |
| 327.72 | | | | | 19495 | 17 | 19478 | | | | | 27043 | 26 | 27017 | | | Sleep related leg cramps | neurological | LDL-C | |
| 331 | -0.09678 | 0.09957 | 0.9078 | 0.3311 | 20649 | 514 | 20135 | | | | | 28193 | 478 | 27715 | 0.9128 | -0.01412 | Other cerebral degenerations | neurological | LDL-C | |
| 331.1 | | | | | 20475 | 340 | 20135 | | | | | 28036 | 321 | 27715 | 0.9456 | 0.01091 | Hydrocephalus | neurological | LDL-C | |
| 331.9 | | | | | 20281 | 146 | 20135 | | | | | 27836 | 121 | 27715 | | | Cerebral degeneration, unspecified | neurological | LDL-C | |
| 332 | | | | | 20573 | 438 | 20135 | | | | | 27974 | 259 | 27715 | 0.07067 | 0.3046 | Parkinson's disease | neurological | LDL-C | |
| 333 | 0.3606 | 0.08693 | 1.434 | 3.36e-05 | 20955 | 820 | 20135 | 0.3944 | 0.08098 | 1.484 | 1.11e-06 | 28634 | 919 | 27715 | 0.4852 | 0.07659 | Extrapyramidal disease and abnormal movement disorders | neurological | LDL-C | |
| 333.1 | | | | | 20501 | 366 | 20135 | | | | | 28134 | 419 | 27715 | 0.641 | 0.08167 | Essential tremor | neurological | LDL-C | |
| 333.2 | | | | | 20258 | 123 | 20135 | | | | | 27820 | 105 | 27715 | | | Myoclonus | neurological | LDL-C | |
| 333.3 | | | | | 20164 | 29 | 20135 | | | | | 27748 | 33 | 27715 | | | Tics and choreas | neurological | LDL-C | |
| 333.4 | | | | | 20284 | 149 | 20135 | | | | | 28005 | 290 | 27715 | | | Torsion dystonia | neurological | LDL-C | |
| 333.8 | | | | | 20218 | 83 | 20135 | | | | | 27746 | 31 | 27715 | | | Other degenerative diseases of the basal ganglia | neurological | LDL-C | |
| 334 | | | | | 20526 | 391 | 20135 | | | | | 28145 | 430 | 27715 | 0.03016 | 0.3072 | Degenerative disease of the spinal cord | neurological | LDL-C | |
| 334.1 | | | | | 20181 | 46 | 20135 | | | | | 27786 | 71 | 27715 | | | Spinocerebellar disease | neurological | LDL-C | |
| 334.2 | | | | | 20201 | 66 | 20135 | | | | | 27768 | 53 | 27715 | | | Anterior horn cell disease | neurological | LDL-C | |
| 334.21 | | | | | 20183 | 48 | 20135 | | | | | 27759 | 44 | 27715 | | | Amyotrophic Lateral Sclerosis | neurological | LDL-C | |
| 335 | | | | | 20445 | 310 | 20135 | -0.9279 | 0.07443 | 0.3954 | 1.15e-35 | 28694 | 979 | 27715 | 0.2834 | 0.148 | Multiple sclerosis | neurological | LDL-C | |
| 337 | | | | | 20390 | 255 | 20135 | | | | | 28028 | 313 | 27715 | 0.7889 | -0.04973 | Disorders of the autonomic nervous system | neurological | LDL-C | |
| 337.1 | | | | | 20203 | 68 | 20135 | | | | | 27793 | 78 | 27715 | | | Peripheral autonomic neuropathy | neurological | LDL-C | |
| 338 | 0.09984 | 0.03432 | 1.105 | 0.003619 | 24967 | 5863 | 19104 | 0.3952 | 0.03033 | 1.485 | 8.49e-39 | 33179 | 7691 | 25488 | 4.90e-26 | 0.4397 | Pain | neurological | LDL-C | Pain |
| 338.1 | -0.1473 | 0.03884 | 0.863 | 1.49e-04 | 23126 | 4022 | 19104 | 0.1718 | 0.03543 | 1.187 | 1.24e-06 | 30282 | 4794 | 25488 | 1.57e-22 | 0.4661 | Acute pain | neurological | LDL-C | |
| 338.2 | 0.7024 | 0.05242 | 2.019 | 6.17e-41 | 21816 | 2712 | 19104 | 0.7949 | 0.04129 | 2.214 | 1.32e-82 | 29799 | 4311 | 25488 | 0.001044 | 0.2057 | Chronic pain | neurological | LDL-C | |
| 339 | 0.8165 | 0.09558 | 2.263 | 1.31e-17 | 25998 | 701 | 25297 | 0.9435 | 0.06595 | 2.569 | 1.99e-46 | 31803 | 1472 | 30331 | 0.02717 | 0.2379 | Other headache syndromes | neurological | LDL-C | |
| 340 | 0.7384 | 0.09556 | 2.093 | 1.10e-14 | 25980 | 683 | 25297 | 0.9718 | 0.04813 | 2.643 | 1.16e-90 | 33186 | 2855 | 30331 | 0.01001 | 0.2528 | Migraine | neurological | LDL-C | |
| 340.1 | | | | | 25434 | 137 | 25297 | 1.273 | 0.09795 | 3.571 | 1.29e-38 | 31063 | 732 | 30331 | 0.4305 | 0.1826 | Migrain with aura | neurological | LDL-C | |
| 341 | | | | | 20276 | 141 | 20135 | | | | | 28206 | 491 | 27715 | 0.02928 | 0.4392 | Other demyelinating diseases of central nervous system | neurological | LDL-C | |
| 342 | | | | | 20493 | 358 | 20135 | | | | | 28011 | 296 | 27715 | 0.4097 | 0.1454 | Hemiplegia | neurological | LDL-C | |
| 343 | | | | | 20279 | 144 | 20135 | | | | | 27844 | 129 | 27715 | | | Infantile cerebral palsy | neurological | LDL-C | |
| 344 | | | | | 20460 | 325 | 20135 | | | | | 27997 | 282 | 27715 | 0.3479 | 0.1547 | Other paralytic syndromes | neurological | LDL-C | |
| 345 | -0.1743 | 0.05854 | 0.8401 | 0.00291 | 21729 | 1594 | 20135 | -0.1561 | 0.05319 | 0.8555 | 0.003336 | 29581 | 1866 | 27715 | 0.1567 | 0.1013 | Epilepsy, recurrent seizures, convulsions | neurological | LDL-C | |
| 345.1 | -0.5054 | 0.08694 | 0.6033 | 6.15e-09 | 20847 | 712 | 20135 | -0.3985 | 0.08259 | 0.6713 | 1.40e-06 | 28454 | 739 | 27715 | 0.01034 | 0.2787 | Epilepsy | neurological | LDL-C | |
| 345.11 | | | | | 20451 | 316 | 20135 | | | | | 28000 | 285 | 27715 | 0.4624 | 0.1264 | Generalized convulsive epilepsy | neurological | LDL-C | |
| 345.12 | -0.6274 | 0.103 | 0.534 | 1.11e-09 | 20638 | 503 | 20135 | | | | | 28179 | 464 | 27715 | 0.001535 | 0.4199 | Partial epilepsy | neurological | LDL-C | |
| 345.3 | -0.1324 | 0.06477 | 0.876 | 0.04097 | 21418 | 1283 | 20135 | -0.1242 | 0.05918 | 0.8832 | 0.03581 | 29200 | 1485 | 27715 | 0.2335 | 0.0944 | Convulsions | neurological | LDL-C | |

| phenotype | male_beta | male_SE | male_OR | male_p | male_n_total | male_n_cases | male_n_controls | female_beta | female_SE | female_OR | female_p | female_n_total | female_n_cases | female_n_controls | interaction_p | interaction_beta | description | group | lab | label |
|---|---|---|---|---|---|---|---|---|---|---|---|---|---|---|---|---|---|---|---|---|
| 346 | | | | | 20172 | 37 | 20135 | | | | | 27787 | 72 | 27715 | | | Abnormal findings on study of brain and/or nervous system | neurological | LDL-C | |
| 346.1 | | | | | 20298 | 163 | 20135 | | | | | 27984 | 269 | 27715 | | | Nonspecific abnormal findings on radiological and other examination of skull and head | neurological | LDL-C | |
| 346.2 | | | | | 20163 | 28 | 20135 | | | | | 27765 | 50 | 27715 | | | Nonspecific abnormal results of function study of brain and central nervous system | neurological | LDL-C | |
| 346.3 | | | | | 20173 | 38 | 20135 | | | | | 27758 | 43 | 27715 | | | Nonspecific abnormal findings in cerebrospinal fluid | neurological | LDL-C | |
| 347 | | | | | 20161 | 26 | 20135 | | | | | 27775 | 60 | 27715 | | | Cataplexy and narcolepsy | neurological | LDL-C | |
| 348 | 0.2853 | 0.06102 | 1.33 | 2.93e-06 | 21644 | 1509 | 20135 | 0.1204 | 0.05962 | 1.128 | 0.04346 | 29223 | 1508 | 27715 | 0.1948 | -0.09933 | Other conditions of brain | neurological | LDL-C | |
| 348.2 | 0.1147 | 0.0996 | 1.122 | 0.2493 | 20647 | 512 | 20135 | -0.121 | 0.09313 | 0.8861 | 0.1939 | 28310 | 595 | 27715 | 0.2908 | -0.1294 | Cerebral edema and compression of brain | neurological | LDL-C | |
| 348.4 | | | | | 20194 | 59 | 20135 | | | | | 27769 | 54 | 27715 | | | Cerebral cysts | neurological | LDL-C | |
| 348.7 | | | | | 20262 | 127 | 20135 | | | | | 27823 | 108 | 27715 | | | Coma | neurological | LDL-C | |
| 348.8 | 0.4134 | 0.07869 | 1.512 | 1.49e-07 | 21082 | 947 | 20135 | 0.3639 | 0.07926 | 1.439 | 4.41e-06 | 28610 | 895 | 27715 | 0.9694 | 0.003879 | Encephalopathy, not elsewhere classified | neurological | LDL-C | |
| 348.9 | -0.05094 | 0.08141 | 0.9503 | 0.5315 | 20916 | 781 | 20135 | -0.01716 | 0.08335 | 0.983 | 0.8369 | 28459 | 744 | 27715 | 0.2666 | 0.1158 | Other conditions of brain, NOS | neurological | LDL-C | |
| 349 | | | | | 20369 | 234 | 20135 | | | | | 27969 | 254 | 27715 | | | Other and unspecified disorders of the nervous system | neurological | LDL-C | |
| 350 | 0.353 | 0.04879 | 1.423 | 4.60e-13 | 25783 | 2709 | 23074 | 0.3956 | 0.04272 | 1.485 | 2.02e-20 | 34411 | 3494 | 30917 | 0.03522 | 0.1267 | Abnormal movement | neurological | LDL-C | |
| 350.1 | 0.3794 | 0.0843 | 1.461 | 6.76e-06 | 23884 | 810 | 23074 | 0.4171 | 0.07202 | 1.518 | 6.97e-09 | 32013 | 1096 | 30917 | 0.06391 | 0.1914 | Abnormal involuntary movements | neurological | LDL-C | |
| 350.2 | 0.4063 | 0.06041 | 1.501 | 1.74e-11 | 24868 | 1794 | 23074 | 0.4177 | 0.0528 | 1.519 | 2.54e-15 | 33227 | 2310 | 30917 | 0.2075 | 0.0942 | Abnormality of gait | neurological | LDL-C | |
| 350.3 | -0.03186 | 0.09775 | 0.9686 | 0.7445 | 23605 | 531 | 23074 | -0.1769 | 0.07661 | 0.8378 | 0.02091 | 31754 | 837 | 30917 | 0.5779 | -0.06387 | Lack of coordination | neurological | LDL-C | |
| 350.5 | | | | | 23103 | 29 | 23074 | | | | | 30953 | 36 | 30917 | | | Abnormal reflex | neurological | LDL-C | |

| phenotype | male_beta | male_SE | male_OR | male_p | male_n_total | male_n_cases | male_n_controls | female_beta | female_SE | female_OR | female_p | female_n_total | female_n_cases | female_n_controls | interaction_p | interaction_beta | description | group | lab | label |
|---|---|---|---|---|---|---|---|---|---|---|---|---|---|---|---|---|---|---|---|---|
| 350.6 | | | | | 23183 | 109 | 23074 | | | | | 31041 | 124 | 30917 | | | Disturbances of sensation of smell and taste | neurological | LDL-C | |
| 351 | 0.4996 | 0.06782 | 1.648 | 1.76e-13 | 25090 | 1362 | 23728 | 0.7107 | 0.05411 | 2.035 | 2.11e-39 | 32761 | 2244 | 30517 | 4.55e-05 | 0.3259 | Other peripheral nerve disorders | neurological | LDL-C | |
| 352 | | | | | 24026 | 298 | 23728 | 0.2854 | 0.09987 | 1.33 | 0.004271 | 31061 | 544 | 30517 | 3.02e-04 | 0.5398 | Disorders of other cranial nerves | neurological | LDL-C | |
| 352.1 | | | | | 23876 | 148 | 23728 | | | | | 30870 | 353 | 30517 | 0.004194 | 0.5707 | Trigeminal nerve disorders [CN5] | neurological | LDL-C | |
| 352.2 | | | | | 23861 | 133 | 23728 | | | | | 30696 | 179 | 30517 | | | Facial nerve disorders [CN7] | neurological | LDL-C | |
| 353 | | | | | 23928 | 200 | 23728 | | | | | 30746 | 229 | 30517 | | | Nerve root and plexus disorders | neurological | LDL-C | |
| 353.1 | | | | | 23805 | 77 | 23728 | | | | | 30605 | 88 | 30517 | | | Nerve plexus lesions | neurological | LDL-C | |
| 353.2 | | | | | 23800 | 72 | 23728 | | | | | 30603 | 86 | 30517 | | | Nerve root lesions | neurological | LDL-C | |
| 355 | | | | | 23869 | 141 | 23728 | | | | | 30825 | 308 | 30517 | | | Complex regional/central pain syndrome | neurological | LDL-C | |
| 355.1 | 0.7343 | 0.1092 | 2.084 | 1.78e-11 | 24316 | 588 | 23728 | 0.8823 | 0.08245 | 2.416 | 1.01e-26 | 31540 | 1023 | 30517 | 0.1021 | 0.2108 | Chronic pain syndrome | neurological | LDL-C | |
| 356 | 0.6523 | 0.08237 | 1.92 | 2.40e-15 | 24919 | 1003 | 23916 | 0.799 | 0.08357 | 2.223 | 1.16e-21 | 33295 | 1002 | 32293 | 0.1765 | 0.1466 | Hereditary and idiopathic peripheral neuropathy | neurological | LDL-C | |
| 357 | 1.007 | 0.08953 | 2.738 | 2.34e-29 | 24993 | 1077 | 23916 | 0.6721 | 0.07201 | 1.958 | 1.03e-20 | 33576 | 1283 | 32293 | 6.65e-04 | -0.3685 | Inflammatory and toxic neuropathy | neurological | LDL-C | |
| 358 | | | | | 24025 | 109 | 23916 | | | | | 32462 | 169 | 32293 | | | Myoneural disorders | neurological | LDL-C | |
| 358.1 | | | | | 23987 | 71 | 23916 | | | | | 32384 | 91 | 32293 | | | Myasthenia gravis | neurological | LDL-C | |
| 359 | 0.6341 | 0.0958 | 1.885 | 3.61e-11 | 24562 | 646 | 23916 | 0.5872 | 0.09728 | 1.799 | 1.58e-09 | 32892 | 599 | 32293 | 0.825 | -0.02726 | Muscular dystrophies and other myopathies | neurological | LDL-C | |

| phenotype | male_beta | male_SE | male_OR | male_p | male_n_total | male_n_cases | male_n_controls | female_beta | female_SE | female_OR | female_p | female_n_total | female_n_cases | female_n_controls | interaction_p | interaction_beta | description | group | lab | label |
|---|---|---|---|---|---|---|---|---|---|---|---|---|---|---|---|---|---|---|---|---|
| 359.1 | | | | | 23949 | 33 | 23916 | | | | | 32328 | 35 | 32293 | | | Muscular dystrophies | neurological | LDL-C | |
| 359.2 | 0.73 | 0.0987 | 2.075 | 1.40e-13 | 24538 | 622 | 23916 | 0.6659 | 0.1006 | 1.946 | 3.60e-11 | 32864 | 571 | 32293 | 0.8137 | -0.03001 | Myopathy | neurological | LDL-C | |
| 360 | | | | | 24749 | 89 | 24660 | | | | | 33075 | 87 | 32988 | | | Disorders of the globe | sense organs | LDL-C | |
| 360.2 | | | | | 24707 | 47 | 24660 | | | | | 33042 | 54 | 32988 | | | Progressive myopia | sense organs | LDL-C | |
| 360.3 | | | | | 24670 | 10 | 24660 | | | | | 33001 | 13 | 32988 | | | Hypotony of eye | sense organs | LDL-C | |
| 361 | | | | | 24928 | 268 | 24660 | | | | | 33163 | 175 | 32988 | | | Retinal detachments and defects | sense organs | LDL-C | |
| 361.1 | | | | | 24821 | 161 | 24660 | | | | | 33064 | 76 | 32988 | | | Retinal detachment with retinal defect | sense organs | LDL-C | |
| 361.2 | | | | | 24678 | 18 | 24660 | | | | | 32996 | 8 | 32988 | | | Retinoschisis and retinal cysts | sense organs | LDL-C | |
| 362 | 0.9208 | 0.08716 | 2.511 | 4.33e-26 | 25441 | 1087 | 24354 | 0.9208 | 0.0754 | 2.511 | 2.66e-34 | 34109 | 1379 | 32730 | 0.9816 | -0.002519 | Other retinal disorders | sense organs | LDL-C | |
| 362.1 | | | | | 24369 | 15 | 24354 | | | | | 32743 | 13 | 32730 | | | Retinopathy of prematurity | sense organs | LDL-C | |
| 362.2 | 0.7859 | 0.1141 | 2.194 | 5.75e-12 | 24974 | 620 | 24354 | 0.9004 | 0.09981 | 2.461 | 1.85e-19 | 33549 | 819 | 32730 | 0.6106 | 0.07294 | Degeneration of macula and posterior pole of retina | sense organs | LDL-C | |
| 362.21 | | | | | 24463 | 109 | 24354 | | | | | 32881 | 151 | 32730 | | | Macular degeneration, dry | sense organs | LDL-C | |
| 362.22 | | | | | 24410 | 56 | 24354 | | | | | 32819 | 89 | 32730 | | | Macular degeneration, wet | sense organs | LDL-C | |
| 362.23 | | | | | 24511 | 157 | 24354 | | | | | 32900 | 170 | 32730 | | | Cystoid macular degeneration of retina | sense organs | LDL-C | |
| 362.26 | | | | | 24590 | 236 | 24354 | | | | | 32963 | 233 | 32730 | | | Macular puckering of retina | sense organs | LDL-C | |
| 362.27 | | | | | 24433 | 79 | 24354 | | | | | 32875 | 145 | 32730 | | | Drusen (degenerative) of retina | sense organs | LDL-C | |
| 362.29 | | | | | 24520 | 166 | 24354 | | | | | 33004 | 274 | 32730 | | | Macular degeneration (senile) of retina NOS | sense organs | LDL-C | |
| 362.3 | | | | | 24535 | 181 | 24354 | | | | | 32906 | 176 | 32730 | | | Other nondiabetic retinopathy | sense organs | LDL-C | |
| 362.31 | | | | | 24393 | 39 | 24354 | | | | | 32753 | 23 | 32730 | | | Separation of retinal layers | sense organs | LDL-C | |
| 362.4 | | | | | 24528 | 174 | 24354 | | | | | 32939 | 209 | 32730 | | | Retinal vascular changes and abnomalities | sense organs | LDL-C | |
| 362.5 | | | | | 24369 | 15 | 24354 | | | | | 32823 | 93 | 32730 | | | Toxic maculopathy of retina | sense organs | LDL-C | |
| 362.6 | | | | | 24414 | 60 | 24354 | | | | | 32796 | 66 | 32730 | | | Peripheral retinal degenerations | sense organs | LDL-C | |
| 362.7 | | | | | 24401 | 47 | 24354 | | | | | 32775 | 45 | 32730 | | | Hereditary retinal dystrophies | sense organs | LDL-C | |
| 362.8 | | | | | 24401 | 47 | 24354 | | | | | 32774 | 44 | 32730 | | | Retinal hemorrhage/ischemia | sense organs | LDL-C | |
| 362.9 | | | | | 24469 | 115 | 24354 | | | | | 32862 | 132 | 32730 | | | Retinal edema | sense organs | LDL-C | |
| 363 | | | | | 24770 | 110 | 24660 | | | | | 33134 | 146 | 32988 | | | Chorioretinal inflammations, scars, and other disorders of choroid | sense organs | LDL-C | |
| 363.3 | | | | | 24721 | 61 | 24660 | | | | | 33072 | 84 | 32988 | | | Chorioretinal scars | sense organs | LDL-C | |
| 363.4 | | | | | 24662 | 2 | 24660 | | | | | 32995 | 7 | 32988 | | | Choroidal degenerations | sense organs | LDL-C | |
| 364 | | | | | 24841 | 181 | 24660 | | | | | 33258 | 270 | 32988 | | | Corneal opacity and other disorders of cornea | sense organs | LDL-C | |
| 364.1 | | | | | 24753 | 93 | 24660 | | | | | 33113 | 125 | 32988 | | | Corneal opacity | sense organs | LDL-C | |
| 364.2 | | | | | 24712 | 52 | 24660 | | | | | 33031 | 43 | 32988 | | | Corneal edema | sense organs | LDL-C | |
| 364.4 | | | | | 24715 | 55 | 24660 | | | | | 33057 | 69 | 32988 | | | Corneal degenerations | sense organs | LDL-C | |
| 364.41 | | | | | 24684 | 24 | 24660 | | | | | 33006 | 18 | 32988 | | | Keratoconus | sense organs | LDL-C | |
| 364.5 | | | | | 24720 | 60 | 24660 | | | | | 33111 | 123 | 32988 | | | Corneal dystrophy | sense organs | LDL-C | |
| 364.51 | | | | | 24699 | 39 | 24660 | | | | | 33061 | 73 | 32988 | | | Fuchs' dystrophy | sense organs | LDL-C | |
| 364.9 | | | | | 24688 | 28 | 24660 | | | | | 33020 | 32 | 32988 | | | Cornea replaced by transplant | sense organs | LDL-C | |
| 365 | 0.9051 | 0.09579 | 2.472 | 3.42e-21 | 25576 | 916 | 24660 | 0.9358 | 0.08372 | 2.549 | 5.22e-29 | 34125 | 1137 | 32988 | 0.7558 | 0.03745 | Glaucoma | sense organs | LDL-C | |
| 365.1 | 1.078 | 0.1305 | 2.938 | 1.45e-16 | 25218 | 558 | 24660 | 0.9663 | 0.1105 | 2.628 | 2.26e-18 | 33666 | 678 | 32988 | 0.4882 | -0.1134 | Open-angle glaucoma | sense organs | LDL-C | |
| 365.11 | | | | | 24878 | 218 | 24660 | | | | | 33217 | 229 | 32988 | | | Primary open angle glaucoma | sense organs | LDL-C | |
| 365.2 | | | | | 24753 | 93 | 24660 | | | | | 33132 | 144 | 32988 | | | Primary angle-closure glaucoma | sense organs | LDL-C | |
| 365.5 | | | | | 24671 | 11 | 24660 | | | | | 33004 | 16 | 32988 | | | Pseudoexfoliation glaucoma | sense organs | LDL-C | |
| 366 | 1.348 | 0.06354 | 3.849 | 7.23e-100 | 26713 | 2883 | 23830 | 1.369 | 0.05426 | 3.932 | 1.81e-140 | 35649 | 3722 | 31927 | 0.9585 | -0.004216 | Cataract | sense organs | LDL-C | |
| 366.1 | | | | | 23845 | 15 | 23830 | | | | | 31936 | 9 | 31927 | | | Nonsenile Cataract | sense organs | LDL-C | |

| phenotype | male_beta | male_SE | male_OR | male_p | male_n_total | male_n_cases | male_n_controls | female_beta | female_SE | female_OR | female_p | female_n_total | female_n_cases | female_n_controls | interaction_p | interaction_beta | description | group | lab | label |
|---|---|---|---|---|---|---|---|---|---|---|---|---|---|---|---|---|---|---|---|---|
| 366.2 | 1.421 | 0.07015 | 4.139 | 3.51e-91 | 26293 | 2463 | 23830 | 1.443 | 0.06014 | 4.235 | 2.97e-127 | 35061 | 3134 | 31927 | 0.9331 | 0.007555 | Senile cataract | sense organs | LDL-C | |
| 366.3 | | | | | 23835 | 5 | 23830 | | | | | 31932 | 5 | 31927 | | | Traumatic cataract | sense organs | LDL-C | |
| phenotype | male_beta | male_SE | male_OR | male_p | male_n_total | male_n_cases | male_n_controls | female_beta | female_SE | female_OR | female_p | female_n_total | female_n_cases | female_n_controls | interaction_p | interaction_beta | description | group | lab | label |
| 367 | 1.107 | 0.08488 | 3.025 | 7.27e-39 | 26602 | 1195 | 25407 | 1.401 | 0.06442 | 4.061 | 6.46e-105 | 35278 | 2096 | 33182 | 0.02465 | 0.2264 | Disorders of refraction and accommodation; blindness and low vision | sense organs | LDL-C | |
| 367.1 | 1.47 | 0.1292 | 4.349 | 5.61e-30 | 25991 | 584 | 25407 | 1.724 | 0.08638 | 5.61 | 1.16e-88 | 34437 | 1255 | 33182 | 0.3205 | 0.1465 | Myopia | sense organs | LDL-C | |
| 367.2 | | | | | 25731 | 324 | 25407 | 1.687 | 0.1465 | 5.405 | 1.06e-30 | 33701 | 519 | 33182 | 0.08185 | 0.3823 | Astigmatism | sense organs | LDL-C | |
| 367.8 | | | | | 25506 | 99 | 25407 | | | | | 33431 | 249 | 33182 | | | Hypermetropia | sense organs | LDL-C | |
| 367.9 | | | | | 25707 | 300 | 25407 | | | | | 33524 | 342 | 33182 | 0.9695 | 0.006724 | Blindness and low vision | sense organs | LDL-C | |
| 368 | 0.6318 | 0.08227 | 1.881 | 1.60e-14 | 26249 | 990 | 25259 | 0.7343 | 0.06555 | 2.084 | 4.01e-29 | 34737 | 1481 | 33256 | 0.6453 | 0.04491 | Visual disturbances | sense organs | LDL-C | |
| 368.1 | | | | | 25311 | 52 | 25259 | | | | | 33326 | 70 | 33256 | | | Amblyopia | sense organs | LDL-C | |
| 368.2 | | | | | 25592 | 333 | 25259 | | | | | 33627 | 371 | 33256 | 0.7487 | 0.05405 | Diplopia and disorders of binocular vision | sense organs | LDL-C | |
| 368.3 | | | | | 25277 | 18 | 25259 | | | | | 33272 | 16 | 33256 | | | Anisometropia | sense organs | LDL-C | |
| 368.4 | | | | | 25410 | 151 | 25259 | | | | | 33464 | 208 | 33256 | | | Visual field defects | sense organs | LDL-C | |
| 368.5 | | | | | 25262 | 3 | 25259 | | | | | 33257 | 1 | 33256 | | | Color vision deficiencies | sense organs | LDL-C | |
| 368.7 | | | | | 25259 | | | | | | | 33257 | 1 | 33256 | | | Disorders of accommodation | sense organs | LDL-C | |
| 368.9 | | | | | 25451 | 192 | 25259 | | | | | 33633 | 377 | 33256 | 0.1347 | -0.3275 | Subjective visual disturbances | sense organs | LDL-C | |
| 368.91 | | | | | 25299 | 40 | 25259 | | | | | 33323 | 67 | 33256 | | | Psychophysical visual disturbances | sense organs | LDL-C | |
| 369 | 1.085 | 0.127 | 2.961 | 1.25e-17 | 24317 | 512 | 23805 | 1.159 | 0.09104 | 3.186 | 4.19e-37 | 31511 | 916 | 30595 | 0.6605 | 0.06576 | Infection of the eye | sense organs | LDL-C | |
| 369.2 | | | | | 23885 | 80 | 23805 | | | | | 30724 | 129 | 30595 | | | Eye infection, viral | sense organs | LDL-C | |
| 369.5 | | | | | 24187 | 382 | 23805 | 1.319 | 0.1081 | 3.738 | 3.27e-34 | 31314 | 719 | 30595 | 0.9444 | 0.0129 | Conjunctivitis, infectious | sense organs | LDL-C | |
| 370 | | | | | 24074 | 269 | 23805 | 1.076 | 0.1049 | 2.934 | 1.03e-24 | 31271 | 676 | 30595 | 0.01253 | 0.4411 | Keratitis | sense organs | LDL-C | |
| 370.1 | | | | | 23880 | 75 | 23805 | | | | | 30694 | 99 | 30595 | | | Corneal ulcer | sense organs | LDL-C | |
| 370.2 | | | | | 23830 | 25 | 23805 | | | | | 30656 | 61 | 30595 | | | Superficial keratitis | sense organs | LDL-C | |
| 370.3 | | | | | 23964 | 159 | 23805 | | | | | 31063 | 468 | 30595 | 0.05343 | 0.4403 | Keratoconjunctivitis | sense organs | LDL-C | |
| 370.31 | | | | | 23923 | 118 | 23805 | | | | | 31009 | 414 | 30595 | 0.6244 | 0.1366 | Keratoconjunctivitis sicca | sense organs | LDL-C | |
| 371 | 1.399 | 0.1081 | 4.051 | 2.54e-38 | 24762 | 957 | 23805 | 1.303 | 0.07633 | 3.679 | 2.58e-65 | 32133 | 1538 | 30595 | 0.266 | -0.1426 | Inflammation of the eye | sense organs | LDL-C | |
| 371.1 | | | | | 23925 | 120 | 23805 | | | | | 30815 | 220 | 30595 | | | Uveitis, noninfectious or NOS | sense organs | LDL-C | |
| 371.2 | | | | | 24253 | 448 | 23805 | 1.63 | 0.1195 | 5.105 | 2.22e-42 | 31368 | 773 | 30595 | 0.006541 | -0.6754 | Conjunctivitis, noninfectious | sense organs | LDL-C | |
| 371.21 | | | | | 24201 | 396 | 23805 | 1.683 | 0.1292 | 5.383 | 8.21e-39 | 31279 | 684 | 30595 | 0.009619 | -0.7062 | Allergic conjunctivitis | sense organs | LDL-C | |
| 371.3 | | | | | 24165 | 360 | 23805 | 1.586 | 0.1417 | 4.883 | 4.60e-29 | 31154 | 559 | 30595 | 0.1155 | 0.3347 | Inflammation of eyelids | sense organs | LDL-C | |
| 371.33 | | | | | 23808 | 3 | 23805 | | | | | 30603 | 8 | 30595 | | | Noninfectious dermatoses of eyelid | sense organs | LDL-C | |
| 371.9 | | | | | 23817 | 12 | 23805 | | | | | 30607 | 12 | 30595 | | | Chronic inflammatory disorders of orbit | sense organs | LDL-C | |
| 372 | | | | | 23933 | 128 | 23805 | | | | | 30728 | 133 | 30595 | | | Disorders of conjunctiva | sense organs | LDL-C | |
| 374 | 0.9313 | 0.1041 | 2.538 | 3.76e-19 | 24581 | 776 | 23805 | 1.319 | 0.09549 | 3.738 | 2.29e-43 | 31646 | 1051 | 30595 | 0.006482 | 0.3675 | Other disorders of eyelids | sense organs | LDL-C | |
| 374.1 | | | | | 23920 | 115 | 23805 | | | | | 30673 | 78 | 30595 | | | Ectropion or entropion | sense organs | LDL-C | |
| 374.2 | | | | | 23850 | 45 | 23805 | | | | | 30619 | 24 | 30595 | | | Lagophthalmos | sense organs | LDL-C | |
| 374.3 | | | | | 24001 | 196 | 23805 | | | | | 30970 | 375 | 30595 | 0.339 | 0.2229 | Ptosis of eyelid | sense organs | LDL-C | |
| 374.6 | | | | | 24250 | 445 | 23805 | 1.915 | 0.1595 | 6.786 | 3.36e-33 | 31205 | 610 | 30595 | 0.7264 | 0.08547 | Dermatochalasis | sense organs | LDL-C | |

| phenotype | male_beta | male_SE | male_OR | male_p | male_n_total | male_n_cases | male_n_controls | female_beta | female_SE | female_OR | female_p | female_n_total | female_n_cases | female_n_controls | interaction_p | interaction_beta | description | group | lab | label |
|---|---|---|---|---|---|---|---|---|---|---|---|---|---|---|---|---|---|---|---|---|
| 375 | | | | | 23884 | 86 | 23798 | | | | | 31096 | 146 | 30950 | | | Disorders of lacrimal system | sense organs | LDL-C | |
| 375.1 | 1.557 | 0.114 | 4.743 | 1.86e-42 | 24860 | 1062 | 23798 | 1.508 | 0.07603 | 4.518 | 1.49e-87 | 32921 | 1971 | 30950 | 0.5085 | -0.08855 | Dry eyes | sense organs | LDL-C | |
| 375.2 | | | | | 23848 | 50 | 23798 | | | | | 31036 | 86 | 30950 | | | Epiphora | sense organs | LDL-C | |
| 376 | | | | | 23842 | 44 | 23798 | | | | | 30982 | 32 | 30950 | | | Disorders of the orbit | sense organs | LDL-C | |
| 377 | | | | | 24004 | 206 | 23798 | | | | | 31323 | 373 | 30950 | 0.6112 | 0.09066 | Disorders of optic nerve and visual pathways | sense organs | LDL-C | |
| 377.1 | | | | | 23872 | 74 | 23798 | | | | | 31057 | 107 | 30950 | | | Optic atrophy | sense organs | LDL-C | |
| 377.3 | | | | | 23899 | 101 | 23798 | | | | | 31129 | 179 | 30950 | | | Optic neuritis/neuropathy | sense organs | LDL-C | |
| 378 | | | | | 24117 | 319 | 23798 | | | | | 31299 | 349 | 30950 | 0.1592 | 0.2252 | Strabismus and other disorders of binocular eye movements | sense organs | LDL-C | |
| 378.1 | | | | | 23973 | 175 | 23798 | | | | | 31136 | 186 | 30950 | | | Strabismus (not specified as paralytic) | sense organs | LDL-C | |
| 378.2 | | | | | 23835 | 37 | 23798 | | | | | 31001 | 51 | 30950 | | | Nystagmus and other irregular eye movements | sense organs | LDL-C | |
| 378.5 | | | | | 23889 | 91 | 23798 | | | | | 31030 | 80 | 30950 | | | Paralytic strabismus | sense organs | LDL-C | |
| 379 | 1.085 | 0.09195 | 2.96 | 3.86e-32 | 24956 | 1158 | 23798 | 1.062 | 0.07283 | 2.893 | 3.36e-48 | 32559 | 1609 | 30950 | 0.812 | -0.02689 | Other disorders of eye | sense organs | LDL-C | |
| 379.1 | | | | | 23846 | 48 | 23798 | | | | | 31085 | 135 | 30950 | | | Scleritis and episcleritis | sense organs | LDL-C | |
| 379.2 | 1.319 | 0.1102 | 3.741 | 4.98e-33 | 24762 | 964 | 23798 | 1.225 | 0.08614 | 3.406 | 6.25e-46 | 32269 | 1319 | 30950 | 0.5289 | -0.08575 | Disorders of vitreous body | sense organs | LDL-C | |
| 379.3 | | | | | 23855 | 57 | 23798 | | | | | 30995 | 45 | 30950 | | | Aphakia and other disorders of lens | sense organs | LDL-C | |
| 379.4 | | | | | 23835 | 37 | 23798 | | | | | 31014 | 64 | 30950 | | | Anomalies of pupillary function | sense organs | LDL-C | |
| 379.5 | | | | | 23914 | 116 | 23798 | | | | | 31022 | 72 | 30950 | | | Disorders of iris and ciliary body | sense organs | LDL-C | |
| 379.51 | | | | | 23816 | 18 | 23798 | | | | | 30958 | 8 | 30950 | | | Pigmentary iris degeneration | sense organs | LDL-C | |
| 379.9 | | | | | 23893 | 95 | 23798 | | | | | 31121 | 171 | 30950 | | | Pain, swelling or discharge of eye | sense organs | LDL-C | |
| 380 | | | | | 25280 | 65 | 25215 | | | | | 34053 | 71 | 33982 | | | Disorders of external ear | sense organs | LDL-C | |
| 380.1 | | | | | 25546 | 331 | 25215 | | | | | 34343 | 361 | 33982 | 0.2779 | -0.2109 | Otitis externa | sense organs | LDL-C | |
| 380.4 | 1.481 | 0.1169 | 4.395 | 8.79e-37 | 26150 | 935 | 25215 | 1.521 | 0.1117 | 4.578 | 3.23e-42 | 34929 | 947 | 33982 | 0.9473 | 0.01039 | Impacted cerumen | sense organs | LDL-C | |
| 381 | 0.8907 | 0.07727 | 2.437 | 9.60e-31 | 25736 | 1209 | 24527 | 1.172 | 0.05858 | 3.228 | 4.70e-89 | 33271 | 2155 | 31116 | 2.12e-04 | 0.3362 | Otitis media and Eustachian tube disorders | sense organs | LDL-C | |
| 381.1 | 0.853 | 0.1022 | 2.347 | 6.99e-17 | 25160 | 633 | 24527 | 1.179 | 0.07831 | 3.252 | 2.98e-51 | 32235 | 1119 | 31116 | 7.21e-04 | 0.4035 | Otitis media | sense organs | LDL-C | |
| 381.11 | | | | | 24939 | 412 | 24527 | 1.195 | 0.09478 | 3.303 | 1.95e-36 | 31847 | 731 | 31116 | 7.22e-04 | 0.483 | Suppurative and unspecified otitis media | sense organs | LDL-C | |
| phenotype | male_beta | male_SE | male_OR | male_p | male_n_total | male_n_cases | male_n_controls | female_beta | female_SE | female_OR | female_p | female_n_total | female_n_cases | female_n_controls | interaction_p | interaction_beta | description | group | lab | label |
| 381.2 | 0.9179 | 0.1095 | 2.504 | 5.32e-17 | 25149 | 622 | 24527 | 1.199 | 0.08306 | 3.317 | 3.09e-47 | 32238 | 1122 | 31116 | 0.01383 | 0.3216 | Eustachian tube disorders | sense organs | LDL-C | |
| 381.3 | | | | | 24579 | 52 | 24527 | | | | | 31179 | 63 | 31116 | | | Mastoiditis & related conditions | sense organs | LDL-C | |
| 381.9 | | | | | 24645 | 118 | 24527 | | | | | 31231 | 115 | 31116 | | | Otorrhea | sense organs | LDL-C | |
| 382 | | | | | 24849 | 322 | 24527 | 1.386 | 0.1038 | 3.998 | 1.15e-40 | 31925 | 809 | 31116 | 0.7838 | 0.05354 | Otalgia | sense organs | LDL-C | |
| 383 | | | | | 24540 | 13 | 24527 | | | | | 31138 | 22 | 31116 | | | Otosclerosis | sense organs | LDL-C | |
| 384 | | | | | 24691 | 164 | 24527 | | | | | 31314 | 198 | 31116 | | | Other disorders of tympanic membrane | sense organs | LDL-C | |
| 384.1 | | | | | 24551 | 24 | 24527 | | | | | 31124 | 8 | 31116 | | | Myringitis | sense organs | LDL-C | |
| 384.4 | | | | | 24667 | 140 | 24527 | | | | | 31294 | 178 | 31116 | | | Perforation of tympanic membrane | sense organs | LDL-C | |
| 385 | | | | | 24625 | 98 | 24527 | | | | | 31245 | 129 | 31116 | | | Other disorders of middle ear and mastoid | sense organs | LDL-C | |
| 385.3 | | | | | 24606 | 79 | 24527 | | | | | 31204 | 88 | 31116 | | | Cholesteatoma | sense organs | LDL-C | |
| 385.5 | | | | | 24556 | 29 | 24527 | | | | | 31165 | 49 | 31116 | | | Tympanosclerosis and middle ear disease related to otitis media | sense organs | LDL-C | |
| 386 | | | | | 23483 | 489 | 22994 | 1.321 | 0.09471 | 3.745 | 3.46e-44 | 30272 | 992 | 29280 | 0.0402 | 0.3167 | Vertiginous syndromes and other disorders of vestibular system | sense organs | LDL-C | |

| phenotype | male_beta | male_SE | male_OR | male_p | male_n_total | male_n_cases | male_n_controls | female_beta | female_SE | female_OR | female_p | female_n_total | female_n_cases | female_n_controls | interaction_p | interaction_beta | description | group | lab | label |
|---|---|---|---|---|---|---|---|---|---|---|---|---|---|---|---|---|---|---|---|---|
| 386.1 | | | | | 23077 | 83 | 22994 | | | | | 29457 | 177 | 29280 | | | Meniere's disease | sense organs | LDL-C | |
| 386.2 | | | | | 23328 | 334 | 22994 | 1.473 | 0.122 | 4.362 | 1.47e-33 | 29955 | 675 | 29280 | 0.1767 | 0.2721 | Peripheral or central vertigo | sense organs | LDL-C | |
| 386.21 | | | | | 23041 | 47 | 22994 | | | | | 29385 | 105 | 29280 | | | Central origin vertigo | sense organs | LDL-C | |
| 386.3 | | | | | 23038 | 44 | 22994 | | | | | 29344 | 64 | 29280 | | | Labyrinthitis | sense organs | LDL-C | |
| 386.9 | 1.249 | 0.06554 | 3.487 | 5.85e-81 | 25211 | 2217 | 22994 | 1.278 | 0.04872 | 3.59 | 1.16e-151 | 32967 | 3687 | 29280 | 0.5964 | 0.0413 | Dizziness and giddiness (Light-headedness and vertigo) | sense organs | LDL-C | |
| 388 | | | | | 23411 | 34 | 23377 | | | | | 32030 | 47 | 31983 | | | Other disorders of ear | sense organs | LDL-C | |
| 389 | 0.8229 | 0.05628 | 2.277 | 2.09e-48 | 26031 | 2654 | 23377 | 0.8809 | 0.0516 | 2.413 | 2.35e-65 | 34938 | 2955 | 31983 | 0.7181 | 0.02634 | Hearing loss | sense organs | LDL-C | |
| 389.1 | 0.965 | 0.07523 | 2.625 | 1.16e-37 | 25007 | 1630 | 23377 | 0.934 | 0.07025 | 2.545 | 2.50e-40 | 33675 | 1692 | 31983 | 0.4993 | -0.06683 | Sensorineural hearing loss | sense organs | LDL-C | |
| 389.2 | | | | | 23633 | 256 | 23377 | | | | | 32346 | 363 | 31983 | 0.03096 | 0.3632 | Conductive hearing loss | sense organs | LDL-C | |
| 389.3 | | | | | 23398 | 21 | 23377 | | | | | 31995 | 12 | 31983 | | | Degenerative and vascular disorders of ear | sense organs | LDL-C | |
| 389.4 | 1.444 | 0.1346 | 4.238 | 7.60e-27 | 23983 | 606 | 23377 | 1.222 | 0.11 | 3.395 | 1.17e-28 | 32714 | 731 | 31983 | 0.2808 | -0.1818 | Tinnitus | sense organs | LDL-C | |
| 389.5 | | | | | 23394 | 17 | 23377 | | | | | 32002 | 19 | 31983 | | | Disorders of acoustic nerve | sense organs | LDL-C | |
| 394 | 0.8915 | 0.07258 | 2.439 | 1.12e-34 | 22330 | 1378 | 20952 | 0.8002 | 0.06822 | 2.226 | 9.05e-32 | 31197 | 1468 | 29729 | 0.06948 | -0.167 | Rheumatic disease of the heart valves | circulatory system | LDL-C | |
| 394.1 | | | | | 21305 | 353 | 20952 | | | | | 30068 | 339 | 29729 | 0.09666 | -0.2931 | Mitral valve stenosis and aortic valve stenosis | circulatory system | LDL-C | |
| 394.2 | | | | | 21121 | 169 | 20952 | | | | | 29965 | 236 | 29729 | | | Mitral valve disease | circulatory system | LDL-C | |
| 394.3 | | | | | 20969 | 17 | 20952 | | | | | 29742 | 13 | 29729 | | | Aortic valve disease | circulatory system | LDL-C | |
| 394.4 | | | | | 20972 | 20 | 20952 | | | | | 29771 | 42 | 29729 | | | Acute rheumatic heart disease | circulatory system | LDL-C | |
| 394.7 | 0.9001 | 0.1105 | 2.46 | 3.81e-16 | 21461 | 509 | 20952 | 0.783 | 0.09786 | 2.188 | 1.23e-15 | 30357 | 628 | 29729 | 0.4465 | -0.1026 | Disease of tricuspid valve | circulatory system | LDL-C | |
| 395 | 0.8797 | 0.04695 | 2.41 | 2.44e-78 | 24383 | 3431 | 20952 | 0.7585 | 0.0472 | 2.135 | 4.22e-58 | 32852 | 3123 | 29729 | 0.01791 | -0.1434 | Heart valve disorders | circulatory system | LDL-C | |
| 395.1 | 1.098 | 0.06435 | 2.998 | 2.92e-65 | 22776 | 1824 | 20952 | 0.911 | 0.0603 | 2.487 | 1.46e-51 | 31647 | 1918 | 29729 | 0.01206 | -0.2023 | Nonrheumatic mitral valve disorders | circulatory system | LDL-C | |
| 395.2 | 0.8212 | 0.06274 | 2.273 | 3.76e-39 | 22773 | 1821 | 20952 | 0.6899 | 0.06831 | 1.993 | 5.60e-24 | 31147 | 1418 | 29729 | 0.04279 | -0.17 | Nonrheumatic aortic valve disorders | circulatory system | LDL-C | |
| 395.3 | | | | | 21269 | 317 | 20952 | | | | | 30053 | 324 | 29729 | 0.3113 | -0.1976 | Nonrheumatic tricuspid valve disorders | circulatory system | LDL-C | |
| 395.4 | | | | | 21053 | 101 | 20952 | | | | | 29855 | 126 | 29729 | | | Nonrheumatic pulmonary valve disorders | circulatory system | LDL-C | |
| 395.6 | 0.5002 | 0.08283 | 1.649 | 1.56e-09 | 21815 | 863 | 20952 | 0.7037 | 0.09802 | 2.021 | 6.98e-13 | 30347 | 618 | 29729 | 0.9627 | -0.005442 | Heart valve replaced | circulatory system | LDL-C | |
| 396 | 1.107 | 0.1155 | 3.026 | 9.46e-22 | 21534 | 582 | 20952 | 1.14 | 0.09404 | 3.127 | 7.99e-34 | 30632 | 903 | 29729 | 0.5284 | 0.08795 | Abnormal heart sounds | circulatory system | LDL-C | |
| 401 | 1.275 | 0.03354 | 3.58 | 0 | 25158 | 14670 | 10488 | 1.218 | 0.02968 | 3.382 | 0 | 33895 | 14782 | 19113 | 2.42e-04 | -0.1518 | Hypertension | circulatory system | LDL-C | |
| 401.1 | 1.303 | 0.03391 | 3.679 | 0 | 24789 | 14301 | 10488 | 1.243 | 0.02998 | 3.465 | 0 | 33585 | 14472 | 19113 | 2.20e-04 | -0.1544 | Essential hypertension | circulatory system | LDL-C | |
| 401.2 | 2.278 | 0.05969 | 9.76 | 0 | 14837 | 4349 | 10488 | 2.167 | 0.06296 | 8.731 | 1.47e-259 | 22464 | 3351 | 19113 | 0.003439 | -0.2325 | Hypertensive heart and/or renal disease | circulatory system | LDL-C | |
| 401.21 | 2.705 | 0.09029 | 14.96 | 2.99e-197 | 12798 | 2310 | 10488 | 2.68 | 0.09545 | 14.59 | 1.82e-173 | 21086 | 1973 | 19113 | 0.05403 | -0.2367 | Hypertensive heart disease | circulatory system | LDL-C | |
| 401.22 | 2.231 | 0.06997 | 9.312 | 3.97e-223 | 12900 | 2412 | 10488 | 2.056 | 0.07833 | 7.814 | 7.93e-152 | 20743 | 1630 | 19113 | 0.004745 | -0.2712 | Hypertensive chronic kidney disease | circulatory system | LDL-C | |
| 401.3 | 2.05 | 0.1101 | 7.77 | 1.84e-77 | 11324 | 836 | 10488 | 1.92 | 0.09872 | 6.823 | 2.79e-84 | 20074 | 961 | 19113 | 0.006974 | -0.3763 | Other hypertensive complications | circulatory system | LDL-C | |
| 411 | 1.34 | 0.03734 | 3.819 | 5.75e-282 | 24924 | 8227 | 16697 | 1.225 | 0.04174 | 3.404 | 2.65e-189 | 33484 | 5060 | 28424 | 0.03307 | -0.1068 | Ischemic Heart Disease | circulatory system | LDL-C | |

| phenotype | male_beta | male_SE | male_OR | male_p | male_n_total | male_n_cases | male_n_controls | female_beta | female_SE | female_OR | female_p | female_n_total | female_n_cases | female_n_controls | interaction_p | interaction_beta | description | group | lab | label |
|---|---|---|---|---|---|---|---|---|---|---|---|---|---|---|---|---|---|---|---|---|
| 411.1 | 2.106 | 0.09336 | 8.216 | 1.11e-112 | 18153 | 1456 | 16697 | 2.117 | 0.1279 | 8.308 | 1.67e-61 | 29191 | 767 | 28424 | 0.3985 | -0.1264 | Unstable angina (intermediate coronary syndrome) | circulatory system | LDL-C | |
| 411.2 | 1.727 | 0.0574 | 5.624 | 6.91e-199 | 19713 | 3016 | 16697 | 1.578 | 0.07198 | 4.843 | 1.85e-106 | 30089 | 1665 | 28424 | 0.03801 | -0.1742 | Myocardial infarction | circulatory system | LDL-C | |
| 411.3 | 1.917 | 0.08515 | 6.802 | 2.89e-112 | 18373 | 1676 | 16697 | 1.721 | 0.1016 | 5.591 | 2.11e-64 | 29445 | 1021 | 28424 | 0.0375 | -0.26 | Angina pectoris | circulatory system | LDL-C | |
| 411.4 | 1.406 | 0.03926 | 4.081 | 5.23e-281 | 24205 | 7508 | 16697 | 1.311 | 0.04613 | 3.708 | 1.63e-177 | 32583 | 4159 | 28424 | 0.05591 | -0.1035 | Coronary atherosclerosis | circulatory system | LDL-C | |
| 411.41 | | | | | 16754 | 57 | 16697 | | | | | 28492 | 68 | 28424 | | | Aneurysm and dissection of heart | circulatory system | LDL-C | |
| 411.8 | 2.092 | 0.06698 | 8.1 | 4.10e-214 | 19412 | 2715 | 16697 | 1.94 | 0.1028 | 6.956 | 1.79e-79 | 29458 | 1034 | 28424 | 0.2779 | -0.1237 | Other chronic ischemic heart disease, unspecified | circulatory system | LDL-C | |
| 411.9 | | | | | 16791 | 94 | 16697 | | | | | 28492 | 68 | 28424 | | | Other acute and subacute forms of ischemic heart disease | circulatory system | LDL-C | |
| 414 | 1.364 | 0.06207 | 3.912 | 4.95e-107 | 18860 | 2163 | 16697 | 1.163 | 0.07073 | 3.198 | 1.00e-60 | 29887 | 1463 | 28424 | 0.01218 | -0.2152 | Other forms of chronic heart disease | circulatory system | LDL-C | |
| 414.2 | | | | | 16930 | 233 | 16697 | | | | | 28541 | 117 | 28424 | | | ASCVD | circulatory system | LDL-C | |
| 415 | 0.8985 | 0.05625 | 2.456 | 1.98e-57 | 22610 | 2006 | 20604 | 0.5636 | 0.05422 | 1.757 | 2.62e-25 | 32928 | 1974 | 30954 | 3.77e-05 | -0.2881 | Pulmonary heart disease | circulatory system | LDL-C | |
| 415.1 | 0.6789 | 0.08161 | 1.972 | 8.86e-17 | 21476 | 872 | 20604 | 0.5287 | 0.08296 | 1.697 | 1.85e-10 | 31762 | 808 | 30954 | 0.184 | -0.1372 | Acute pulmonary heart disease | circulatory system | LDL-C | |
| 415.11 | 0.5288 | 0.08569 | 1.697 | 6.79e-10 | 21366 | 762 | 20604 | 0.4796 | 0.08521 | 1.615 | 1.82e-08 | 31710 | 756 | 30954 | 0.6739 | -0.04501 | Pulmonary embolism and infarction, acute | circulatory system | LDL-C | |
| 415.2 | 1.336 | 0.08109 | 3.803 | 5.69e-61 | 21649 | 1045 | 20604 | 0.7392 | 0.07176 | 2.094 | 6.95e-25 | 32069 | 1115 | 30954 | 3.34e-08 | -0.5403 | Chronic pulmonary heart disease | circulatory system | LDL-C | |
| 415.21 | | | | | 20763 | 159 | 20604 | | | | | 31201 | 247 | 30954 | | | Primary pulmonary hypertension | circulatory system | LDL-C | |
| 416 | 1.048 | 0.04982 | 2.851 | 3.67e-98 | 23564 | 2960 | 20604 | 0.8838 | 0.0574 | 2.42 | 1.71e-53 | 32994 | 2040 | 30954 | 0.1756 | -0.093 | Cardiomegaly | circulatory system | LDL-C | |
| 418 | 1.28 | 0.03447 | 3.598 | 5.69e-302 | 23574 | 7662 | 15912 | 1.218 | 0.03081 | 3.379 | 0 | 31720 | 9073 | 22647 | 0.825 | 0.009577 | Nonspecific chest pain | circulatory system | LDL-C | |
| phenotype | male_beta | male_SE | male_OR | male_p | male_n_total | male_n_cases | male_n_controls | female_beta | female_SE | female_OR | female_p | female_n_total | female_n_cases | female_n_controls | interaction_p | interaction_beta | description | group | lab | label |
| 418.1 | 1.762 | 0.08917 | 5.822 | 7.24e-87 | 17223 | 1311 | 15912 | 1.478 | 0.07632 | 4.382 | 1.71e-83 | 24061 | 1414 | 22647 | 0.03454 | -0.2409 | Precordial pain | circulatory system | LDL-C | |
| 420 | 0.6726 | 0.07893 | 1.959 | 1.58e-17 | 24630 | 934 | 23696 | 0.8502 | 0.08198 | 2.34 | 3.35e-25 | 34594 | 895 | 33699 | 0.06596 | 0.1897 | Carditis | circulatory system | LDL-C | |
| 420.1 | | | | | 23727 | 31 | 23696 | | | | | 33725 | 26 | 33699 | | | Myocarditis | circulatory system | LDL-C | |
| 420.2 | 0.6337 | 0.09593 | 1.885 | 3.96e-11 | 24311 | 615 | 23696 | 0.882 | 0.09679 | 2.416 | 8.02e-20 | 34349 | 650 | 33699 | 0.0242 | 0.2784 | Pericarditis | circulatory system | LDL-C | |
| 420.21 | | | | | 23792 | 96 | 23696 | | | | | 33781 | 82 | 33699 | | | Acute pericarditis | circulatory system | LDL-C | |
| 420.22 | | | | | 23800 | 104 | 23696 | | | | | 33807 | 108 | 33699 | | | Chronic pericarditis | circulatory system | LDL-C | |
| 420.3 | | | | | 23990 | 294 | 23696 | | | | | 33932 | 233 | 33699 | 0.7608 | -0.05758 | Endocarditis | circulatory system | LDL-C | |
| 425 | 1.147 | 0.06051 | 3.148 | 4.42e-80 | 25754 | 2058 | 23696 | 1.065 | 0.0739 | 2.9 | 4.71e-47 | 34958 | 1259 | 33699 | 0.5119 | -0.05741 | Cardiomyopathy | circulatory system | LDL-C | |
| 425.1 | 1.142 | 0.06413 | 3.134 | 5.58e-71 | 25538 | 1842 | 23696 | 1.075 | 0.08067 | 2.93 | 1.68e-40 | 34754 | 1055 | 33699 | 0.4553 | -0.07069 | Primary/intrinsic cardiomyopathies | circulatory system | LDL-C | |
| 425.11 | | | | | 23809 | 113 | 23696 | | | | | 33810 | 111 | 33699 | | | Hypertrophic obstructive cardiomyopathy | circulatory system | LDL-C | |
| 425.12 | | | | | 23788 | 92 | 23696 | | | | | 33777 | 78 | 33699 | | | Other hypertrophic cardiomyopathy | circulatory system | LDL-C | |
| 425.2 | 1.332 | 0.1095 | 3.789 | 5.01e-34 | 24351 | 655 | 23696 | | | | | 34066 | 367 | 33699 | 0.59 | -0.08835 | Secondary/extrinsic cardiomyopathies | circulatory system | LDL-C | |

| phenotype | male_beta | male_SE | male_OR | male_p | male_n_total | male_n_cases | male_n_controls | female_beta | female_SE | female_OR | female_p | female_n_total | female_n_cases | female_n_controls | interaction_p | interaction_beta | description | group | lab | label |
|---|---|---|---|---|---|---|---|---|---|---|---|---|---|---|---|---|---|---|---|---|
| 425.8 | | | | | 23738 | 42 | 23696 | | | | | 33802 | 103 | 33699 | | | Other cardiomyopathy | circulatory system | LDL-C | |
| 426 | 1.034 | 0.04639 | 2.811 | 5.76e-110 | 18055 | 3856 | 14199 | 1.015 | 0.05067 | 2.759 | 2.94e-89 | 24927 | 2812 | 22115 | 0.6771 | 0.02641 | Cardiac conduction disorders | circulatory system | LDL-C | |
| 426.2 | 1.075 | 0.09096 | 2.931 | 2.99e-32 | 15129 | 930 | 14199 | 0.8944 | 0.1072 | 2.446 | 7.09e-17 | 22650 | 535 | 22115 | 0.01675 | -0.3093 | Atrioventricular [AV] block | circulatory system | LDL-C | |
| 426.21 | | | | | 14391 | 192 | 14199 | | | | | 22185 | 70 | 22115 | | | First degree AV block | circulatory system | LDL-C | |
| 426.22 | | | | | 14228 | 29 | 14199 | | | | | 22135 | 20 | 22115 | | | Mobitz II AV block | circulatory system | LDL-C | |
| 426.23 | | | | | 14388 | 189 | 14199 | | | | | 22218 | 103 | 22115 | | | Second degree AV block | circulatory system | LDL-C | |
| 426.24 | 0.8844 | 0.107 | 2.422 | 1.38e-16 | 14798 | 599 | 14199 | | | | | 22478 | 363 | 22115 | 0.09857 | -0.2518 | Atrioventricular block, complete | circulatory system | LDL-C | |
| 426.25 | | | | | 14243 | 44 | 14199 | | | | | 22136 | 21 | 22115 | | | Other heart block | circulatory system | LDL-C | |
| 426.3 | 1.443 | 0.1001 | 4.231 | 4.82e-47 | 15070 | 871 | 14199 | 1.158 | 0.1135 | 3.184 | 1.98e-24 | 22678 | 563 | 22115 | 0.1618 | -0.198 | Bundle branch block | circulatory system | LDL-C | |
| 426.31 | | | | | 14547 | 348 | 14199 | | | | | 22262 | 147 | 22115 | | | Right bundle branch block | circulatory system | LDL-C | |
| 426.32 | | | | | 14638 | 439 | 14199 | | | | | 22500 | 385 | 22115 | 0.1992 | -0.2389 | Left bundle branch block | circulatory system | LDL-C | |
| 426.4 | | | | | 14250 | 51 | 14199 | | | | | 22148 | 33 | 22115 | | | Anomalous atrioventricular excitation | circulatory system | LDL-C | |
| 426.7 | 1.086 | 0.09832 | 2.963 | 2.24e-28 | 14888 | 689 | 14199 | 1.32 | 0.1018 | 3.742 | 2.04e-38 | 22857 | 742 | 22115 | 0.003381 | 0.3891 | Abnormal electrocardiogram [ECG] [EKG] | circulatory system | LDL-C | |
| 426.8 | | | | | 14301 | 102 | 14199 | | | | | 22230 | 115 | 22115 | | | Other cardiac conduction disorders | circulatory system | LDL-C | |
| 426.9 | 1.222 | 0.06201 | 3.395 | 1.75e-86 | 16283 | 2084 | 14199 | 1.007 | 0.07377 | 2.736 | 2.15e-42 | 23352 | 1237 | 22115 | 0.04635 | -0.1761 | Cardiac pacemaker/device in situ | circulatory system | LDL-C | |
| 426.91 | 1.107 | 0.0768 | 3.027 | 3.90e-47 | 15508 | 1309 | 14199 | 0.9822 | 0.08557 | 2.67 | 1.68e-30 | 23040 | 925 | 22115 | 0.121 | -0.1635 | Cardiac pacemaker in situ | circulatory system | LDL-C | |
| 426.92 | 1.484 | 0.08434 | 4.409 | 2.89e-69 | 15274 | 1075 | 14199 | | | | | 22521 | 406 | 22115 | 0.5392 | -0.0874 | Cardiac defibrillator in situ | circulatory system | LDL-C | |
| 427 | 0.6832 | 0.03155 | 1.98 | 5.53e-104 | 23930 | 9731 | 14199 | 0.7928 | 0.02847 | 2.21 | 1.27e-170 | 32189 | 10074 | 22115 | 0.002356 | 0.1179 | Cardiac dysrhythmias | circulatory system | LDL-C | |
| 427.1 | 1.178 | 0.06376 | 3.247 | 3.69e-76 | 16046 | 1847 | 14199 | 0.9587 | 0.06732 | 2.608 | 5.18e-46 | 23534 | 1419 | 22115 | 0.3001 | -0.08937 | Paroxysmal tachycardia, unspecified | circulatory system | LDL-C | |
| 427.11 | 0.8843 | 0.1005 | 2.421 | 1.35e-18 | 14852 | 653 | 14199 | 0.8859 | 0.08462 | 2.425 | 1.20e-25 | 22978 | 863 | 22115 | 0.9574 | -0.006558 | Paroxysmal supraventricular tachycardia | circulatory system | LDL-C | |
| 427.12 | 1.357 | 0.07652 | 3.885 | 2.16e-70 | 15495 | 1296 | 14199 | 1.037 | 0.1033 | 2.821 | 1.06e-23 | 22690 | 575 | 22115 | 0.08167 | -0.2081 | Paroxysmal ventricular tachycardia | circulatory system | LDL-C | |
| 427.2 | 0.8994 | 0.04399 | 2.458 | 6.37e-93 | 18883 | 4684 | 14199 | 0.8934 | 0.0505 | 2.443 | 4.77e-70 | 25226 | 3111 | 22115 | 0.9926 | 5.54e-04 | Atrial fibrillation and flutter | circulatory system | LDL-C | |
| 427.21 | 0.9074 | 0.04457 | 2.478 | 4.01e-92 | 18758 | 4559 | 14199 | 0.9037 | 0.05145 | 2.469 | 4.52e-69 | 25130 | 3015 | 22115 | 0.9651 | -0.002664 | Atrial fibrillation | circulatory system | LDL-C | |
| 427.22 | 0.9959 | 0.07158 | 2.707 | 5.28e-44 | 15660 | 1461 | 14199 | 1.16 | 0.09551 | 3.189 | 6.32e-34 | 22935 | 820 | 22115 | 0.06469 | 0.2033 | Atrial flutter | circulatory system | LDL-C | |
| 427.3 | 0.8948 | 0.06145 | 2.447 | 4.95e-48 | 16128 | 1929 | 14199 | 0.9342 | 0.06119 | 2.545 | 1.27e-52 | 23822 | 1707 | 22115 | 0.9439 | -0.005614 | Other specified cardiac dysrhythmias | circulatory system | LDL-C | |
| 427.4 | | | | | 14682 | 483 | 14199 | | | | | 22371 | 256 | 22115 | 0.06891 | -0.319 | Cardiac arrest and ventricular fibrillation | circulatory system | LDL-C | |
| 427.41 | | | | | 14406 | 207 | 14199 | | | | | 22188 | 73 | 22115 | | | Ventricular fibrillation and flutter | circulatory system | LDL-C | |
| 427.42 | | | | | 14543 | 344 | 14199 | | | | | 22324 | 209 | 22115 | 0.3389 | -0.1833 | Cardiac arrest | circulatory system | LDL-C | |
| 427.5 | 0.9543 | 0.05169 | 2.597 | 4.21e-76 | 17123 | 2924 | 14199 | 0.991 | 0.05527 | 2.694 | 6.73e-72 | 24375 | 2260 | 22115 | 0.3254 | 0.06885 | Arrhythmia (cardiac) NOS | circulatory system | LDL-C | |

| phenotype | male_beta | male_SE | male_OR | male_p | male_n_total | male_n_cases | male_n_controls | female_beta | female_SE | female_OR | female_p | female_n_total | female_n_cases | female_n_controls | interaction_p | interaction_beta | description | group | lab | label |
|---|---|---|---|---|---|---|---|---|---|---|---|---|---|---|---|---|---|---|---|---|
| 427.6 | 1.177 | 0.1007 | 3.243 | 1.61e-31 | 15021 | 822 | 14199 | 1.045 | 0.107 | 2.843 | 1.52e-22 | 22723 | 608 | 22115 | 0.5271 | -0.0879 | Premature beats | circulatory system | LDL-C | |
| 427.61 | | | | | 14305 | 106 | 14199 | | | | | 22243 | 128 | 22115 | | | Supraventricular premature beats | circulatory system | LDL-C | |
| 427.7 | 0.5132 | 0.04706 | 1.671 | 1.10e-27 | 16928 | 2729 | 14199 | 0.6749 | 0.04266 | 1.964 | 2.23e-56 | 25298 | 3183 | 22115 | 0.001269 | 0.1867 | Tachycardia NOS | circulatory system | LDL-C | |
| 427.8 | 1.065 | 0.08559 | 2.9 | 1.57e-35 | 15296 | 1097 | 14199 | 0.9078 | 0.09296 | 2.479 | 1.58e-22 | 22910 | 795 | 22115 | 0.08034 | -0.2046 | Sinoatrial node dysfunction (Bradycardia) | circulatory system | LDL-C | |
| 427.9 | 1.297 | 0.06819 | 3.658 | 1.17e-80 | 16102 | 1903 | 14199 | 1.347 | 0.04722 | 3.845 | 6.03e-179 | 25899 | 3784 | 22115 | 0.9919 | 8.01e-04 | Palpitations | circulatory system | LDL-C | |
| 428 | 1.179 | 0.04303 | 3.25 | 3.90e-165 | 24029 | 4495 | 19534 | 0.9629 | 0.04518 | 2.619 | 9.07e-101 | 33140 | 3590 | 29550 | 5.05e-04 | -0.1967 | Congestive heart failure; nonhypertensive | circulatory system | LDL-C | |
| 428.1 | 1.329 | 0.04892 | 3.776 | 1.96e-162 | 22921 | 3387 | 19534 | 1.066 | 0.05216 | 2.903 | 9.01e-93 | 32151 | 2601 | 29550 | 1.15e-04 | -0.2499 | Congestive heart failure (CHF) NOS | circulatory system | LDL-C | |
| 428.2 | 1.878 | 0.08721 | 6.538 | 7.97e-103 | 20752 | 1218 | 19534 | 1.525 | 0.1021 | 4.594 | 1.77e-50 | 30286 | 736 | 29550 | 0.01769 | -0.2951 | Heart failure NOS | circulatory system | LDL-C | |
| 428.3 | 1.5 | 0.06113 | 4.48 | 7.27e-133 | 21811 | 2277 | 19534 | 1.328 | 0.07388 | 3.773 | 3.21e-72 | 30905 | 1355 | 29550 | 0.02646 | -0.1947 | Heart failure with reduced EF [Systolic or combined heart failure] | circulatory system | LDL-C | |
| 428.4 | 1.568 | 0.08672 | 4.795 | 4.97e-73 | 20846 | 1312 | 19534 | 1.393 | 0.07742 | 4.025 | 2.39e-72 | 31066 | 1516 | 29550 | 0.03994 | -0.223 | Heart failure with preserved EF [Diastolic heart failure] | circulatory system | LDL-C | |
| 429 | 1.289 | 0.07465 | 3.629 | 8.18e-67 | 21072 | 1538 | 19534 | 1.323 | 0.07898 | 3.755 | 5.44e-63 | 30921 | 1371 | 29550 | 0.6588 | 0.04463 | Ill-defined descriptions and complications of heart disease | circulatory system | LDL-C | |
| 429.1 | | | | | 19984 | 450 | 19534 | | | | | 29770 | 220 | 29550 | 0.162 | -0.3271 | Heart transplant/surgery | circulatory system | LDL-C | |
| 429.2 | 1.408 | 0.1357 | 4.089 | 3.10e-25 | 20041 | 507 | 19534 | | | | | 29943 | 393 | 29550 | 0.629 | -0.0908 | Abnormal function study of cardiovascular system | circulatory system | LDL-C | |
| 429.3 | 1.486 | 0.1293 | 4.421 | 1.37e-30 | 20117 | 583 | 19534 | 1.481 | 0.118 | 4.397 | 4.02e-36 | 30233 | 683 | 29550 | 0.8519 | -0.03102 | Symptoms involving cardiovascular system | circulatory system | LDL-C | |
| 429.9 | | | | | 19772 | 238 | 19534 | | | | | 29698 | 148 | 29550 | | | Cardiac complications, not elsewhere classified | circulatory system | LDL-C | |
| 430 | 0.8176 | 0.09483 | 2.265 | 6.63e-18 | 22249 | 653 | 21596 | 0.5992 | 0.1021 | 1.821 | 4.39e-09 | 31365 | 529 | 30836 | 0.09686 | -0.2063 | Intracranial hemorrhage | circulatory system | LDL-C | |
| 430.1 | | | | | 21793 | 197 | 21596 | | | | | 31023 | 187 | 30836 | | | Subarachnoid hemorrhage | circulatory system | LDL-C | |
| 430.2 | | | | | 21911 | 315 | 21596 | | | | | 31093 | 257 | 30836 | 0.5289 | -0.1123 | Intracerebral hemorrhage | circulatory system | LDL-C | |
| 430.3 | | | | | 21798 | 202 | 21596 | | | | | 30969 | 133 | 30836 | | | Subdural hemorrhage | circulatory system | LDL-C | |
| phenotype | male_beta | male_SE | male_OR | male_p | male_n_total | male_n_cases | male_n_controls | female_beta | female_SE | female_OR | female_p | female_n_total | female_n_cases | female_n_controls | interaction_p | interaction_beta | description | group | lab | label |
| 433 | 1.121 | 0.04851 | 3.069 | 3.19e-118 | 25148 | 3552 | 21596 | 0.998 | 0.04644 | 2.713 | 2.02e-102 | 34408 | 3572 | 30836 | 0.02448 | -0.1375 | Cerebrovascular disease | circulatory system | LDL-C | |
| 433.1 | 1.618 | 0.08397 | 5.041 | 1.09e-82 | 23051 | 1455 | 21596 | 1.657 | 0.09146 | 5.245 | 2.24e-73 | 32011 | 1175 | 30836 | 0.7185 | -0.04146 | Occlusion and stenosis of precerebral arteries | circulatory system | LDL-C | |
| 433.11 | | | | | 21806 | 210 | 21596 | | | | | 31009 | 173 | 30836 | | | Occlusion of cerebral arteries, with cerebral infarction | circulatory system | LDL-C | |
| 433.12 | | | | | 21766 | 170 | 21596 | | | | | 30990 | 154 | 30836 | | | Cerebral atherosclerosis | circulatory system | LDL-C | |
| 433.2 | 1.817 | 0.09189 | 6.152 | 5.36e-87 | 22816 | 1220 | 21596 | 1.842 | 0.09332 | 6.306 | 1.12e-86 | 32003 | 1167 | 30836 | 0.9465 | -0.008171 | Occlusion of cerebral arteries | circulatory system | LDL-C | |
| 433.21 | 1.879 | 0.09536 | 6.548 | 1.92e-86 | 22764 | 1168 | 21596 | 1.867 | 0.09645 | 6.47 | 1.67e-83 | 31945 | 1109 | 30836 | 0.7581 | -0.03893 | Cerebral artery occlusion, with cerebral infarction | circulatory system | LDL-C | |
| 433.3 | 1.448 | 0.07399 | 4.256 | 2.51e-85 | 23331 | 1735 | 21596 | 1.227 | 0.0685 | 3.41 | 1.02e-71 | 32595 | 1759 | 30836 | 0.01466 | -0.2269 | Cerebral ischemia | circulatory system | LDL-C | |
| 433.31 | 1.439 | 0.07689 | 4.218 | 3.31e-78 | 23194 | 1598 | 21596 | 1.264 | 0.07203 | 3.539 | 6.59e-69 | 32452 | 1616 | 30836 | 0.07649 | -0.1722 | Transient cerebral ischemia | circulatory system | LDL-C | |
| 433.32 | | | | | 21602 | 6 | 21596 | | | | | 30850 | 14 | 30836 | | | Moyamoya disease | circulatory system | LDL-C | |

| phenotype | male_beta | male_SE | male_OR | male_p | male_n_total | male_n_cases | male_n_controls | female_beta | female_SE | female_OR | female_p | female_n_total | female_n_cases | female_n_controls | interaction_p | interaction_beta | description | group | lab | label |
|---|---|---|---|---|---|---|---|---|---|---|---|---|---|---|---|---|---|---|---|---|
| 433.5 | | | | | 21737 | 141 | 21596 | | | | | 31155 | 319 | 30836 | | | Cerebral aneurysm | circulatory system | LDL-C | |
| 433.6 | | | | | 21926 | 330 | 21596 | | | | | 31171 | 335 | 30836 | 0.0492 | -0.5026 | Acute, but ill-defined cerebrovascular disease | circulatory system | LDL-C | |
| 433.8 | 1.379 | 0.1112 | 3.97 | 2.64e-35 | 22246 | 650 | 21596 | 0.9405 | 0.1023 | 2.561 | 3.95e-20 | 31470 | 634 | 30836 | 0.006262 | -0.38 | Late effects of cerebrovascular disease | circulatory system | LDL-C | |
| 440 | 1.405 | 0.07603 | 4.077 | 2.77e-76 | 21894 | 1543 | 20351 | 1.229 | 0.08035 | 3.419 | 7.79e-53 | 31649 | 1262 | 30387 | 0.02457 | -0.2284 | Atherosclerosis | circulatory system | LDL-C | |
| 440.1 | | | | | 20496 | 145 | 20351 | | | | | 30601 | 214 | 30387 | | | Atherosclerosis of renal artery | circulatory system | LDL-C | |
| 440.2 | 1.496 | 0.1079 | 4.466 | 9.45e-44 | 21065 | 714 | 20351 | | | | | 30809 | 422 | 30387 | 0.006189 | -0.4214 | Atherosclerosis of the extremities | circulatory system | LDL-C | |
| 440.21 | | | | | 20544 | 193 | 20351 | | | | | 30472 | 85 | 30387 | | | Atherosclerosis of native arteries of the extremities with ulceration or gangrene | circulatory system | LDL-C | |
| 440.22 | | | | | 20765 | 414 | 20351 | | | | | 30616 | 229 | 30387 | 0.2844 | -0.2281 | Atherosclerosis of native arteries of the extremities with intermittent claudication | circulatory system | LDL-C | |
| 440.9 | | | | | 20820 | 469 | 20351 | | | | | 30841 | 454 | 30387 | 0.1529 | -0.2982 | Atherosclerosis of aorta | circulatory system | LDL-C | |
| 441 | | | | | 20539 | 188 | 20351 | | | | | 30626 | 239 | 30387 | | | Vascular insufficiency of intestine | circulatory system | LDL-C | |
| 441.1 | | | | | 20421 | 70 | 20351 | | | | | 30462 | 75 | 30387 | | | Acute vascular insufficiency of intestine | circulatory system | LDL-C | |
| 441.2 | | | | | 20385 | 34 | 20351 | | | | | 30454 | 67 | 30387 | | | Chronic vascular insufficiency of intestine | circulatory system | LDL-C | |
| 442 | 0.7163 | 0.07272 | 2.047 | 6.88e-23 | 21609 | 1258 | 20351 | 0.7429 | 0.09632 | 2.102 | 1.22e-14 | 31061 | 674 | 30387 | 0.3167 | -0.1094 | Other aneurysm | circulatory system | LDL-C | |
| 442.1 | 0.7864 | 0.08138 | 2.195 | 4.35e-22 | 21366 | 1015 | 20351 | | | | | 30810 | 423 | 30387 | 0.03201 | -0.2791 | Aortic aneurysm | circulatory system | LDL-C | |
| 442.11 | 0.7451 | 0.1021 | 2.107 | 2.98e-13 | 20965 | 614 | 20351 | | | | | 30570 | 183 | 30387 | 0.7613 | -0.05813 | Abdominal aortic aneurysm | circulatory system | LDL-C | |
| 442.2 | | | | | 20421 | 70 | 20351 | | | | | 30397 | 10 | 30387 | | | Aneurysm of iliac artery | circulatory system | LDL-C | |
| 442.3 | | | | | 20442 | 91 | 20351 | | | | | 30441 | 54 | 30387 | | | Aneurysm of artery of lower extremity | circulatory system | LDL-C | |
| 442.4 | | | | | 20394 | 43 | 20351 | | | | | 30441 | 54 | 30387 | | | Arterial dissection | circulatory system | LDL-C | |
| 442.8 | | | | | 20454 | 103 | 20351 | | | | | 30512 | 125 | 30387 | | | Aneurysm of other specified artery | circulatory system | LDL-C | |
| 443 | 1.352 | 0.07174 | 3.865 | 3.08e-79 | 22049 | 1698 | 20351 | 0.9565 | 0.06792 | 2.603 | 4.88e-45 | 31903 | 1516 | 30387 | 1.05e-05 | -0.4031 | Peripheral vascular disease | circulatory system | LDL-C | |
| 443.1 | | | | | 20444 | 93 | 20351 | | | | | 30852 | 465 | 30387 | 0.1104 | -0.4008 | Raynaud's syndrome | circulatory system | LDL-C | |
| 443.7 | | | | | 20611 | 260 | 20351 | | | | | 30517 | 130 | 30387 | | | Peripheral angiopathy in diseases classified elsewhere | circulatory system | LDL-C | |
| 443.8 | | | | | 20448 | 97 | 20351 | | | | | 30453 | 66 | 30387 | | | Other specified peripheral vascular diseases | circulatory system | LDL-C | |
| 443.9 | 1.414 | 0.07855 | 4.112 | 1.95e-72 | 21791 | 1440 | 20351 | 1.365 | 0.09354 | 3.914 | 3.32e-48 | 31349 | 962 | 30387 | 0.2835 | -0.1218 | Peripheral vascular disease, unspecified | circulatory system | LDL-C | |
| 444 | | | | | 20661 | 310 | 20351 | | | | | 30633 | 246 | 30387 | 0.3948 | -0.1633 | Arterial embolism and thrombosis | circulatory system | LDL-C | |
| 444.1 | | | | | 20465 | 114 | 20351 | | | | | 30462 | 75 | 30387 | | | Arterial embolism and thrombosis of lower extremity artery | circulatory system | LDL-C | |
| 444.2 | | | | | 20391 | 40 | 20351 | | | | | 30424 | 37 | 30387 | | | Embolism and thrombosis of abdominal aorta | circulatory system | LDL-C | |
| 444.5 | | | | | 20357 | 6 | 20351 | | | | | 30392 | 5 | 30387 | | | Atheroembolism | circulatory system | LDL-C | |

| phenotype | male_beta | male_SE | male_OR | male_p | male_n_total | male_n_cases | male_n_controls | female_beta | female_SE | female_OR | female_p | female_n_total | female_n_cases | female_n_controls | interaction_p | interaction_beta | description | group | lab | label |
|---|---|---|---|---|---|---|---|---|---|---|---|---|---|---|---|---|---|---|---|---|
| 446 | | | | | 20562 | 211 | 20351 | | | | | 30776 | 389 | 30387 | 0.009277 | -0.4691 | Polyarteritis nodosa and allied conditions | circulatory system | LDL-C | |
| 446.1 | | | | | 20353 | 2 | 20351 | | | | | 30391 | 4 | 30387 | | | Thromboangiitis obliterans | circulatory system | LDL-C | |
| 446.2 | | | | | 20351 | | | | | | | 30393 | 6 | 30387 | | | Acute febrile mucocutaneous lymph node syndrome (Kawasaki disease) | circulatory system | LDL-C | |
| 446.3 | | | | | 20377 | 26 | 20351 | | | | | 30419 | 32 | 30387 | | | Hypersensitivity angiitis | circulatory system | LDL-C | |
| 446.4 | | | | | 20429 | 78 | 20351 | | | | | 30467 | 80 | 30387 | | | Wegener's granulomatosis | circulatory system | LDL-C | |
| 446.5 | | | | | 20386 | 35 | 20351 | | | | | 30500 | 113 | 30387 | | | Giant cell arteritis | circulatory system | LDL-C | |
| 446.6 | | | | | 20380 | 29 | 20351 | | | | | 30423 | 36 | 30387 | | | Polyarteritis nodosa | circulatory system | LDL-C | |
| 446.7 | | | | | 20353 | 2 | 20351 | | | | | 30403 | 16 | 30387 | | | Takayasu's disease | circulatory system | LDL-C | |
| 446.8 | | | | | 20362 | 11 | 20351 | | | | | 30409 | 22 | 30387 | | | Thrombotic microangiopathy | circulatory system | LDL-C | |
| 446.9 | | | | | 20443 | 92 | 20351 | | | | | 30554 | 167 | 30387 | | | Arteritis NOS | circulatory system | LDL-C | |
| 447 | 1.342 | 0.09718 | 3.825 | 2.37e-43 | 21399 | 1048 | 20351 | 0.875 | 0.1155 | 2.399 | 3.54e-14 | 30938 | 551 | 30387 | 0.002 | -0.4418 | Other disorders of arteries and arterioles | circulatory system | LDL-C | |
| 447.1 | | | | | 20462 | 111 | 20351 | | | | | 30487 | 100 | 30387 | | | Stricture of artery | circulatory system | LDL-C | |
| 447.7 | 1.364 | 0.1226 | 3.911 | 9.13e-29 | 21056 | 705 | 20351 | | | | | 30596 | 209 | 30387 | 6.96e-04 | -0.6994 | Aortic ectasia | circulatory system | LDL-C | |
| 448 | | | | | 20358 | 7 | 20351 | | | | | 30410 | 23 | 30387 | | | Disease of capillaries | circulatory system | LDL-C | |
| 450 | | | | | 27464 | 390 | 27074 | 0.2607 | 0.08596 | 1.298 | 0.002421 | 36525 | 739 | 35786 | 0.08325 | 0.2294 | Noninfectious disorders of lymphatic channels | circulatory system | LDL-C | |
| 451 | | | | | 17675 | 229 | 17446 | | | | | 26197 | 265 | 25932 | | | Phlebitis and thrombophlebitis | circulatory system | LDL-C | |
| 451.2 | | | | | 17551 | 105 | 17446 | | | | | 26069 | 137 | 25932 | | | Phlebitis and thrombophlebitis of lower extremities | circulatory system | LDL-C | |
| 452 | 0.5814 | 0.0522 | 1.789 | 8.15e-29 | 19497 | 2051 | 17446 | 0.6374 | 0.05161 | 1.892 | 4.79e-35 | 28016 | 2084 | 25932 | 0.1981 | 0.08688 | Other venous embolism and thrombosis | circulatory system | LDL-C | |
| 452.1 | | | | | 17477 | 31 | 17446 | | | | | 25960 | 28 | 25932 | | | Iatrogenic pulmonary embolism and infarction | circulatory system | LDL-C | |
| 452.2 | 0.6738 | 0.07066 | 1.962 | 1.48e-21 | 18523 | 1077 | 17446 | 0.6659 | 0.07096 | 1.946 | 6.32e-21 | 26996 | 1064 | 25932 | 0.5907 | 0.04935 | Deep vein thrombosis [DVT] | circulatory system | LDL-C | |
| 452.8 | | | | | 17477 | 31 | 17446 | | | | | 25956 | 24 | 25932 | | | Postphlebitic syndrome | circulatory system | LDL-C | |
| 453 | | | | | 17473 | 27 | 17446 | | | | | 25945 | 13 | 25932 | | | Chronic venous hypertension | circulatory system | LDL-C | |
| 454 | | | | | 17905 | 459 | 17446 | 1.629 | 0.1289 | 5.098 | 1.28e-36 | 26542 | 610 | 25932 | 7.37e-04 | 0.5831 | Varicose veins | circulatory system | LDL-C | |

| phenotype | male_beta | male_SE | male_OR | male_p | male_n_total | male_n_cases | male_n_controls | female_beta | female_SE | female_OR | female_p | female_n_total | female_n_cases | female_n_controls | interaction_p | interaction_beta | description | group | lab | label |
|---|---|---|---|---|---|---|---|---|---|---|---|---|---|---|---|---|---|---|---|---|
| 454.1 | | | | | 17712 | 266 | 17446 | 2.021 | 0.1695 | 7.549 | 8.82e-33 | 26438 | 506 | 25932 | 0.7522 | 0.08799 | Varicose veins of lower extremity | circulatory system | LDL-C | |
| 454.11 | | | | | 17657 | 211 | 17446 | | | | | 26297 | 365 | 25932 | 0.6334 | -0.1522 | Varicose veins of lower extremity, symptomtic | circulatory system | LDL-C | |
| 455 | 1.565 | 0.09112 | 4.782 | 4.15e-66 | 18658 | 1212 | 17446 | 1.279 | 0.07625 | 3.594 | 3.56e-63 | 27261 | 1329 | 25932 | 0.003082 | -0.3379 | Hemorrhoids | circulatory system | LDL-C | |
| 456 | | | | | 17849 | 403 | 17446 | | | | | 26402 | 470 | 25932 | 0.0726 | -0.3797 | Chronic venous insufficiency [CVI] | circulatory system | LDL-C | |

| phenotype | male_beta | male_SE | male_OR | male_p | male_n_total | male_n_cases | male_n_controls | female_beta | female_SE | female_OR | female_p | female_n_total | female_n_cases | female_n_controls | interaction_p | interaction_beta | description | group | lab | label |
|---|---|---|---|---|---|---|---|---|---|---|---|---|---|---|---|---|---|---|---|---|
| 457 | 1.503 | 0.06349 | 4.497 | 5.72e-124 | 19943 | 2497 | 17446 | 1.389 | 0.06958 | 4.011 | 1.21e-88 | 27763 | 1831 | 25932 | 0.1719 | -0.1229 | Encounter for long-term (current) use of anticoagulants, antithrombotics, aspirin | circulatory system | LDL-C | |
| 457.2 | 1.798 | 0.1452 | 6.036 | 3.16e-35 | 17977 | 531 | 17446 | | | | | 26262 | 330 | 25932 | 0.999 | -2.68e-04 | Encounter for long-term (current) use of antiplatelets/antithrombotics | circulatory system | LDL-C | |
| 457.3 | 1.529 | 0.0665 | 4.613 | 5.66e-117 | 19733 | 2287 | 17446 | 1.377 | 0.07254 | 3.962 | 2.61e-80 | 27596 | 1664 | 25932 | 0.08088 | -0.1642 | Encounter for long-term (current) use of aspirin | circulatory system | LDL-C | |
| 458 | 0.7014 | 0.04351 | 2.017 | 1.84e-58 | 23987 | 3579 | 20408 | 0.6702 | 0.0462 | 1.955 | 1.10e-47 | 32936 | 2891 | 30045 | 0.9269 | -0.005246 | Hypotension | circulatory system | LDL-C | |
| 458.1 | 0.9148 | 0.09903 | 2.496 | 2.52e-20 | 21124 | 716 | 20408 | 1.081 | 0.1142 | 2.948 | 2.93e-21 | 30599 | 554 | 30045 | 0.1565 | 0.1981 | Orthostatic hypotension | circulatory system | LDL-C | |
| 458.2 | | | | | 20542 | 134 | 20408 | | | | | 30168 | 123 | 30045 | | | Iatrogenic hypotension | circulatory system | LDL-C | |
| 458.9 | 0.696 | 0.04802 | 2.006 | 1.34e-47 | 23196 | 2788 | 20408 | 0.6492 | 0.05163 | 1.914 | 2.96e-36 | 32272 | 2227 | 30045 | 0.6234 | -0.03117 | Hypotension NOS | circulatory system | LDL-C | |
| 459 | 0.897 | 0.08608 | 2.452 | 2.00e-25 | 21377 | 969 | 20408 | 0.9662 | 0.08926 | 2.628 | 2.65e-27 | 30928 | 883 | 30045 | 0.5056 | 0.07676 | Other disorders of circulatory system | circulatory system | LDL-C | |
| 459.1 | | | | | 20541 | 133 | 20408 | | | | | 30175 | 130 | 30045 | | | Hemorrhage NOS | circulatory system | LDL-C | |
| 459.7 | | | | | 20566 | 158 | 20408 | | | | | 30162 | 117 | 30045 | | | Blood vessel replaced | circulatory system | LDL-C | |
| 459.9 | 1.215 | 0.1206 | 3.37 | 7.37e-24 | 21001 | 593 | 20408 | 1.234 | 0.1272 | 3.435 | 3.03e-22 | 30563 | 518 | 30045 | 0.7586 | 0.05085 | Circulatory disease NEC | circulatory system | LDL-C | |
| 464 | 1.975 | 0.09206 | 7.207 | 4.11e-102 | 22311 | 1470 | 20841 | 2.206 | 0.0571 | 9.077 | 0 | 28085 | 3592 | 24493 | 0.02393 | 0.2354 | Acute sinusitis | respiratory | LDL-C | |
| 465 | 1.642 | 0.05897 | 5.166 | 1.10e-170 | 23812 | 2971 | 20841 | 1.925 | 0.04097 | 6.856 | 0 | 30856 | 6363 | 24493 | 3.97e-05 | 0.2727 | Acute upper respiratory infections of multiple or unspecified sites | respiratory | LDL-C | |
| 465.2 | 1.661 | 0.09218 | 5.264 | 1.40e-72 | 21831 | 990 | 20841 | 2.019 | 0.05852 | 7.533 | 6.55e-261 | 27189 | 2696 | 24493 | 1.54e-04 | 0.3815 | Acute pharyngitis | respiratory | LDL-C | |
| 465.4 | | | | | 20905 | 64 | 20841 | | | | | 24590 | 97 | 24493 | | | Acute laryngitis and tracheitis | respiratory | LDL-C | |
| 470 | 1.088 | 0.08738 | 2.967 | 1.43e-35 | 19858 | 932 | 18926 | 1.277 | 0.0739 | 3.586 | 6.71e-67 | 25321 | 1191 | 24130 | 0.1638 | 0.1514 | Septal Deviations/Turbinate Hypertrophy | respiratory | LDL-C | |
| 471 | | | | | 19079 | 153 | 18926 | | | | | 24245 | 115 | 24130 | | | Nasal polyps | respiratory | LDL-C | |
| 472 | 1.113 | 0.1048 | 3.043 | 2.35e-26 | 19621 | 695 | 18926 | 1.237 | 0.07609 | 3.444 | 2.15e-59 | 25328 | 1198 | 24130 | 0.3411 | 0.1177 | Chronic pharyngitis and nasopharyngitis | respiratory | LDL-C | |
| 473 | 0.2641 | 0.07238 | 1.302 | 2.64e-04 | 19940 | 1014 | 18926 | 0.6944 | 0.06603 | 2.003 | 7.23e-26 | 25397 | 1267 | 24130 | 6.84e-09 | 0.5277 | Diseases of the larynx and vocal cords | respiratory | LDL-C | |
| 473.1 | | | | | 19013 | 87 | 18926 | | | | | 24250 | 120 | 24130 | | | Chronic laryngitis | respiratory | LDL-C | |
| 473.3 | | | | | 19140 | 214 | 18926 | | | | | 24368 | 238 | 24130 | | | Paralysis/spasm of vocal cords or larynx | respiratory | LDL-C | |
| 473.4 | 0.3295 | 0.0834 | 1.39 | 7.77e-05 | 19690 | 764 | 18926 | 0.8017 | 0.07571 | 2.229 | 3.36e-26 | 25123 | 993 | 24130 | 7.42e-09 | 0.6075 | Voice disturbance | respiratory | LDL-C | |
| 474 | | | | | 19193 | 267 | 18926 | | | | | 24615 | 485 | 24130 | 0.5756 | 0.09075 | Acute and chronic tonsillitis | respiratory | LDL-C | |
| 474.1 | | | | | 18953 | 27 | 18926 | | | | | 24211 | 81 | 24130 | | | Acute tonsillitis | respiratory | LDL-C | |
| 474.2 | | | | | 19156 | 230 | 18926 | | | | | 24527 | 397 | 24130 | 0.2039 | 0.2215 | Chronic tonsillitis and adenoiditis | respiratory | LDL-C | |
| 475 | 0.8969 | 0.06972 | 2.452 | 7.19e-38 | 20308 | 1382 | 18926 | 1.207 | 0.05614 | 3.342 | 1.79e-102 | 26319 | 2189 | 24130 | 1.89e-05 | 0.3627 | Chronic sinusitis | respiratory | LDL-C | |
| 475.9 | | | | | 19118 | 192 | 18926 | | | | | 24485 | 355 | 24130 | 0.6086 | 0.1342 | Postnasal drip | respiratory | LDL-C | |
| 476 | 1.49 | 0.0602 | 4.436 | 3.51e-135 | 21597 | 2671 | 18926 | 1.75 | 0.04482 | 5.753 | 0 | 28998 | 4868 | 24130 | 1.24e-05 | 0.3129 | Allergic rhinitis | respiratory | LDL-C | |
| 477 | | | | | 19327 | 401 | 18926 | | | | | 24561 | 431 | 24130 | 0.6638 | 0.07153 | Epistaxis or throat hemorrhage | respiratory | LDL-C | |
| 478 | | | | | 19150 | 224 | 18926 | | | | | 24407 | 277 | 24130 | 3.63e-06 | 0.9312 | Throat pain | respiratory | LDL-C | |
| 479 | 0.8213 | 0.06749 | 2.273 | 4.48e-34 | 20272 | 1346 | 18926 | 1.176 | 0.05871 | 3.242 | 2.97e-89 | 26030 | 1900 | 24130 | 8.38e-09 | 0.484 | Other upper respiratory disease | respiratory | LDL-C | |
| 480 | 0.6045 | 0.04116 | 1.83 | 7.75e-49 | 23852 | 3704 | 20148 | 0.7541 | 0.04175 | 2.126 | 6.47e-73 | 31060 | 3539 | 27521 | 4.94e-06 | 0.2447 | Pneumonia | respiratory | LDL-C | |
| 480.1 | 0.5297 | 0.0755 | 1.698 | 2.29e-12 | 21107 | 959 | 20148 | 0.799 | 0.08343 | 2.223 | 9.90e-22 | 28349 | 828 | 27521 | 2.54e-05 | 0.4382 | Bacterial pneumonia | respiratory | LDL-C | |

| phenotype | male_beta | male_SE | male_OR | male_p | male_n_total | male_n_cases | male_n_controls | female_beta | female_SE | female_OR | female_p | female_n_total | female_n_cases | female_n_controls | interaction_p | interaction_beta | description | group | lab | label |
|---|---|---|---|---|---|---|---|---|---|---|---|---|---|---|---|---|---|---|---|---|
| 480.11 | | | | | 20392 | 244 | 20148 | | | | | 27735 | 214 | 27521 | | | Pneumococcal pneumonia | respiratory | LDL-C | |
| 480.12 | | | | | 20313 | 165 | 20148 | | | | | 27625 | 104 | 27521 | | | Pseudomonal pneumonia | respiratory | LDL-C | |
| 480.13 | | | | | 20243 | 95 | 20148 | | | | | 27596 | 75 | 27521 | | | MRSA pneumonia | respiratory | LDL-C | |
| 480.2 | | | | | 20252 | 104 | 20148 | | | | | 27629 | 108 | 27521 | | | Viral pneumonia | respiratory | LDL-C | |
| 480.3 | | | | | 20261 | 113 | 20148 | | | | | 27595 | 74 | 27521 | | | Pneumonia due to fungus (mycoses) | respiratory | LDL-C | |
| 480.5 | | | | | 20273 | 125 | 20148 | | | | | 27647 | 126 | 27521 | | | Bronchopneumonia and lung abscess | respiratory | LDL-C | |
| 481 | | | | | 20437 | 289 | 20148 | | | | | 27934 | 413 | 27521 | 0.3684 | 0.1681 | Influenza | respiratory | LDL-C | |
| 483 | 1.592 | 0.1049 | 4.913 | 5.44e-52 | 21180 | 1032 | 20148 | 1.823 | 0.0788 | 6.188 | 2.25e-118 | 29347 | 1826 | 27521 | 0.06014 | 0.2395 | Acute bronchitis and bronchiolitis | respiratory | LDL-C | |
| 495 | 0.5488 | 0.06388 | 1.731 | 8.68e-18 | 22371 | 1521 | 20850 | 0.6667 | 0.04406 | 1.948 | 1.01e-51 | 31168 | 3223 | 27945 | 0.01583 | 0.1724 | Asthma | respiratory | LDL-C | |
| 495.1 | | | | | 21010 | 160 | 20850 | | | | | 28173 | 228 | 27945 | | | Chronic obstructive asthma | respiratory | LDL-C | |
| 495.11 | | | | | 20869 | 19 | 20850 | | | | | 27991 | 46 | 27945 | | | Chronic obstructive asthma with exacerbation | respiratory | LDL-C | |
| 495.2 | | | | | 21125 | 275 | 20850 | 1.232 | 0.1002 | 3.428 | 1.04e-34 | 28693 | 748 | 27945 | 0.07203 | 0.3206 | Asthma with exacerbation | respiratory | LDL-C | |
| 496 | 0.4899 | 0.04551 | 1.632 | 5.07e-27 | 23991 | 3141 | 20850 | 0.5889 | 0.0466 | 1.802 | 1.34e-36 | 30852 | 2907 | 27945 | 0.04783 | 0.1157 | Chronic airway obstruction | respiratory | LDL-C | |
| 496.1 | 0.4755 | 0.07858 | 1.609 | 1.44e-09 | 21816 | 966 | 20850 | 0.863 | 0.09316 | 2.37 | 1.98e-20 | 28671 | 726 | 27945 | 0.004897 | 0.3079 | Emphysema | respiratory | LDL-C | |
| 496.2 | 0.9979 | 0.09196 | 2.713 | 1.97e-27 | 21707 | 857 | 20850 | 1.102 | 0.08815 | 3.009 | 7.86e-36 | 28880 | 935 | 27945 | 0.2135 | 0.1463 | Chronic bronchitis | respiratory | LDL-C | |
| 496.21 | 1.102 | 0.1033 | 3.011 | 1.50e-26 | 21544 | 694 | 20850 | 1.15 | 0.09852 | 3.157 | 1.82e-31 | 28686 | 741 | 27945 | 0.5405 | 0.08064 | Obstructive chronic bronchitis | respiratory | LDL-C | |
| 496.3 | | | | | 21106 | 256 | 20850 | | | | | 28305 | 360 | 27945 | 0.2063 | 0.2167 | Bronchiectasis | respiratory | LDL-C | |
| 497 | | | | | 21118 | 268 | 20850 | | | | | 28430 | 485 | 27945 | 0.3165 | 0.241 | Bronchitis | respiratory | LDL-C | |
| 498 | | | | | 20873 | 23 | 20850 | | | | | 27982 | 37 | 27945 | | | Acute bronchospasm | respiratory | LDL-C | |
| 499 | | | | | 27770 | 89 | 27681 | | | | | 36921 | 77 | 36844 | | | Cystic fibrosis | respiratory | LDL-C | |
| 500 | | | | | 16503 | 145 | 16358 | | | | | 26637 | 93 | 26544 | | | Lung disease due to external agents | respiratory | LDL-C | |
| 500.1 | | | | | 16369 | 11 | 16358 | | | | | 26562 | 18 | 26544 | | | Extrinsic allergic alveolitis | respiratory | LDL-C | |
| 500.2 | | | | | 16384 | 26 | 16358 | | | | | 26547 | 3 | 26544 | | | Pneumoconiosis | respiratory | LDL-C | |
| phenotype | male_beta | male_SE | male_OR | male_p | male_n_total | male_n_cases | male_n_controls | female_beta | female_SE | female_OR | female_p | female_n_total | female_n_cases | female_n_controls | interaction_p | interaction_beta | description | group | lab | label |
| 501 | 0.1616 | 0.09976 | 1.175 | 0.1053 | 16908 | 550 | 16358 | | | | | 26905 | 361 | 26544 | 0.2922 | 0.1484 | Pneumonitis due to inhalation of food or vomitus | respiratory | LDL-C | |
| 502 | | | | | 16775 | 417 | 16358 | | | | | 26948 | 404 | 26544 | 0.135 | -0.2175 | Postinflammatory pulmonary fibrosis | respiratory | LDL-C | |
| 503 | 1.016 | 0.07786 | 2.762 | 6.69e-39 | 17413 | 1055 | 16358 | 0.8782 | 0.08417 | 2.406 | 1.74e-25 | 27379 | 835 | 26544 | 0.4093 | -0.0852 | Pulmonary congestion and hypostasis | respiratory | LDL-C | |
| 504 | | | | | 16686 | 328 | 16358 | | | | | 26850 | 306 | 26544 | 0.04342 | -0.3421 | Other alveolar and parietoalveolar pneumonopathy | respiratory | LDL-C | |
| 504.1 | | | | | 16528 | 170 | 16358 | | | | | 26674 | 130 | 26544 | | | Idiopathic fibrosing alveolitis | respiratory | LDL-C | |
| 505 | | | | | 16583 | 225 | 16358 | | | | | 26786 | 242 | 26544 | | | Other pulmonary inflamation or edema | respiratory | LDL-C | |
| 506 | 0.003961 | 0.06338 | 1.004 | 0.9502 | 17754 | 1396 | 16358 | 0.07174 | 0.0706 | 1.074 | 0.3095 | 27619 | 1075 | 26544 | 0.968 | 0.003377 | Empyema and pneumothorax | respiratory | LDL-C | |
| 507 | 0.3744 | 0.041 | 1.454 | 6.77e-20 | 20407 | 4049 | 16358 | 0.3785 | 0.04281 | 1.46 | 9.46e-19 | 29890 | 3346 | 26544 | 0.6404 | -0.02452 | Pleurisy; pleural effusion | respiratory | LDL-C | |
| 508 | 0.2134 | 0.03542 | 1.238 | 1.69e-09 | 22186 | 5828 | 16358 | 0.2882 | 0.0379 | 1.334 | 2.87e-14 | 30908 | 4364 | 26544 | 0.9073 | 0.005354 | Pulmonary collapse; interstitial and compensatory emphysema | respiratory | LDL-C | |
| 509 | 0.3113 | 0.0393 | 1.365 | 2.33e-15 | 20689 | 4331 | 16358 | 0.2871 | 0.04069 | 1.333 | 1.73e-12 | 30174 | 3630 | 26544 | 0.9616 | -0.002433 | Respiratory failure, insufficiency, arrest | respiratory | LDL-C | |
| 509.1 | 0.4051 | 0.04607 | 1.5 | 1.43e-18 | 19312 | 2954 | 16358 | 0.3941 | 0.04753 | 1.483 | 1.13e-16 | 29134 | 2590 | 26544 | 0.4179 | 0.04791 | Respiratory failure | respiratory | LDL-C | |

| phenotype | male_beta | male_SE | male_OR | male_p | male_n_total | male_n_cases | male_n_controls | female_beta | female_SE | female_OR | female_p | female_n_total | female_n_cases | female_n_controls | interaction_p | interaction_beta | description | group | lab | label |
|---|---|---|---|---|---|---|---|---|---|---|---|---|---|---|---|---|---|---|---|---|
| 509.2 | 0.6334 | 0.07481 | 1.884 | 2.53e-17 | 17397 | 1039 | 16358 | 0.4519 | 0.08184 | 1.571 | 3.36e-08 | 27339 | 795 | 26544 | 0.03406 | -0.2095 | Respiratory insufficiency | respiratory | LDL-C | |
| 509.3 | -0.01491 | 0.07328 | 0.9852 | 0.8387 | 17365 | 1007 | 16358 | 0.06267 | 0.08741 | 1.065 | 0.4733 | 27216 | 672 | 26544 | 0.3225 | 0.1012 | Pulmonary insufficiency or respiratory failure following trauma and surgery | respiratory | LDL-C | |
| 509.5 | | | | | 16395 | 37 | 16358 | | | | | 26570 | 26 | 26544 | | | Respiratory arrest | respiratory | LDL-C | |
| 509.8 | 0.3761 | 0.08464 | 1.457 | 8.84e-06 | 17139 | 781 | 16358 | 0.4519 | 0.08797 | 1.571 | 2.80e-07 | 27262 | 718 | 26544 | 0.03836 | 0.2268 | Dependence on respirator [Ventilator] or supplemental oxygen | respiratory | LDL-C | |
| 510 | 0.2702 | 0.0467 | 1.31 | 7.25e-09 | 24918 | 2670 | 22248 | 0.3878 | 0.04997 | 1.474 | 8.48e-15 | 34185 | 2275 | 31910 | 0.02282 | 0.1391 | Other diseases of lung | respiratory | LDL-C | |
| 510.2 | | | | | 22355 | 107 | 22248 | | | | | 32017 | 107 | 31910 | | | Lung transplant | respiratory | LDL-C | |
| 512 | 1.01 | 0.03084 | 2.746 | 2.51e-235 | 23558 | 11275 | 12283 | 1.146 | 0.02733 | 3.146 | 0 | 31474 | 13478 | 17996 | 5.80e-08 | 0.2091 | Other symptoms of respiratory system | respiratory | LDL-C | |
| 512.1 | | | | | 12736 | 453 | 12283 | 1.575 | 0.09802 | 4.83 | 4.43e-58 | 18716 | 720 | 17996 | 0.005113 | 0.4124 | Wheezing | respiratory | LDL-C | |
| 512.2 | | | | | 12736 | 453 | 12283 | 1.534 | 0.1042 | 4.638 | 4.44e-49 | 18591 | 595 | 17996 | 0.04507 | 0.2968 | Painful respiration | respiratory | LDL-C | |
| 512.3 | | | | | 12290 | 7 | 12283 | | | | | 18006 | 10 | 17996 | | | Abnormal chest sounds | respiratory | LDL-C | |
| 512.7 | 1.18 | 0.03684 | 3.253 | 7.02e-225 | 19002 | 6719 | 12283 | 1.205 | 0.03364 | 3.336 | 6.64e-281 | 25374 | 7378 | 17996 | 0.03814 | 0.09714 | Shortness of breath | respiratory | LDL-C | |
| 512.8 | 1.481 | 0.04707 | 4.398 | 2.43e-217 | 16541 | 4258 | 12283 | 1.634 | 0.0387 | 5.126 | 0 | 24362 | 6366 | 17996 | 3.32e-04 | 0.2103 | Cough | respiratory | LDL-C | |
| 512.9 | 1.147 | 0.03986 | 3.15 | 3.34e-182 | 17400 | 5117 | 12283 | 1.223 | 0.03775 | 3.398 | 2.29e-230 | 23359 | 5363 | 17996 | 0.01607 | 0.1239 | Other dyspnea | respiratory | LDL-C | |
| 513 | 0.4024 | 0.07574 | 1.495 | 1.08e-07 | 26022 | 945 | 25077 | 0.4591 | 0.08213 | 1.583 | 2.27e-08 | 35312 | 811 | 34501 | 0.0521 | 0.1946 | Respiratory abnormalities | respiratory | LDL-C | |
| 513.3 | | | | | 25286 | 209 | 25077 | | | | | 34707 | 206 | 34501 | | | Hypoventilation | respiratory | LDL-C | |
| 513.31 | | | | | 25126 | 49 | 25077 | | | | | 34541 | 40 | 34501 | | | Apnea | respiratory | LDL-C | |
| 513.32 | | | | | 25149 | 72 | 25077 | | | | | 34549 | 48 | 34501 | | | Orthopnea | respiratory | LDL-C | |
| 513.4 | | | | | 25445 | 368 | 25077 | | | | | 34837 | 336 | 34501 | 0.04372 | 0.3068 | Hyperventilation | respiratory | LDL-C | |
| 513.8 | | | | | 25261 | 184 | 25077 | | | | | 34580 | 79 | 34501 | | | Disorders of diaphragm | respiratory | LDL-C | |
| 514 | 0.3496 | 0.04585 | 1.418 | 2.45e-14 | 24679 | 2921 | 21758 | 0.446 | 0.04787 | 1.562 | 1.20e-20 | 33770 | 2584 | 31186 | 0.01273 | 0.1502 | Abnormal findings examination of lungs | respiratory | LDL-C | |
| 514.1 | | | | | 21848 | 90 | 21758 | | | | | 31294 | 108 | 31186 | | | Abnormal results of function study of pulmonary system | respiratory | LDL-C | |
| 514.2 | 0.408 | 0.07606 | 1.504 | 8.15e-08 | 22786 | 1028 | 21758 | 0.394 | 0.07533 | 1.483 | 1.70e-07 | 32192 | 1006 | 31186 | 0.457 | 0.07293 | Solitary pulmonary nodule | respiratory | LDL-C | |
| 516 | 0.4515 | 0.09992 | 1.571 | 6.24e-06 | 27219 | 552 | 26667 | | | | | 36488 | 334 | 36154 | 0.9753 | 0.004465 | Abnormal sputum | respiratory | LDL-C | |
| 516.1 | | | | | 27145 | 478 | 26667 | | | | | 36446 | 292 | 36154 | 0.6836 | 0.06353 | Hemoptysis | respiratory | LDL-C | |
| 519 | 0.2711 | 0.06393 | 1.311 | 2.24e-05 | 25596 | 1340 | 24256 | 0.447 | 0.06477 | 1.564 | 5.17e-12 | 34332 | 1351 | 32981 | 2.49e-04 | 0.298 | Other diseases of respiratory system, not elsewhere classified | respiratory | LDL-C | |
| 519.1 | | | | | 24327 | 71 | 24256 | | | | | 33033 | 52 | 32981 | | | Tracheostomy complications | respiratory | LDL-C | |
| 519.2 | | | | | 24383 | 127 | 24256 | | | | | 33036 | 55 | 32981 | | | Respiratory complications | respiratory | LDL-C | |
| 519.8 | | | | | 24432 | 176 | 24256 | | | | | 33153 | 172 | 32981 | | | Other diseases of respiratory system, NEC | respiratory | LDL-C | |
| 519.9 | 0.2819 | 0.07544 | 1.326 | 1.87e-04 | 25196 | 940 | 24256 | 0.4104 | 0.07374 | 1.507 | 2.62e-08 | 34006 | 1025 | 32981 | 0.007322 | 0.2523 | Symptoms involving respiratory system and other chest symptoms | respiratory | LDL-C | |
| 520 | | | | | 25518 | 159 | 25359 | | | | | 33516 | 257 | 33259 | | | Disorders of tooth development | digestive | LDL-C | |
| 520.1 | | | | | 25370 | 11 | 25359 | | | | | 33270 | 11 | 33259 | | | Hereditary disturbances in tooth structure | digestive | LDL-C | |
| 520.2 | | | | | 25503 | 144 | 25359 | | | | | 33503 | 244 | 33259 | | | Disturbances in tooth eruption | digestive | LDL-C | |
| 521 | 0.6075 | 0.09134 | 1.836 | 2.92e-11 | 26066 | 707 | 25359 | 1.005 | 0.086 | 2.733 | 1.44e-31 | 34146 | 887 | 33259 | 9.64e-06 | 0.5126 | Diseases of hard tissues of teeth | digestive | LDL-C | |
| 521.1 | 0.6093 | 0.09236 | 1.839 | 4.21e-11 | 26050 | 691 | 25359 | 1.017 | 0.0873 | 2.765 | 2.35e-31 | 34122 | 863 | 33259 | 9.56e-06 | 0.5198 | Dental caries | digestive | LDL-C | |

| phenotype | male_beta | male_SE | male_OR | male_p | male_n_total | male_n_cases | male_n_controls | female_beta | female_SE | female_OR | female_p | female_n_total | female_n_cases | female_n_controls | interaction_p | interaction_beta | description | group | lab | label |
|---|---|---|---|---|---|---|---|---|---|---|---|---|---|---|---|---|---|---|---|---|
| 521.2 | | | | | 25360 | 1 | 25359 | | | | | 33261 | 2 | 33259 | | | Dental abrasion, erosion and attrition | digestive | LDL-C | |
| 521.4 | | | | | 25359 | | | | | | | 33267 | 8 | 33259 | | | Tooth complications likely association with other diseases | digestive | LDL-C | |
| 522 | | | | | 25451 | 92 | 25359 | | | | | 33377 | 118 | 33259 | | | Diseases of pulp and periapical tissues | digestive | LDL-C | |
| 522.1 | | | | | 25361 | 2 | 25359 | | | | | 33263 | 4 | 33259 | | | Pulpitis and necrosis of tooth pulp | digestive | LDL-C | |
| 522.5 | | | | | 25445 | 86 | 25359 | | | | | 33365 | 106 | 33259 | | | Periapical abscess | digestive | LDL-C | |
| 523 | | | | | 25698 | 339 | 25359 | 1.545 | 0.1245 | 4.688 | 2.20e-35 | 33819 | 560 | 33259 | 0.01415 | 0.4479 | Gingival and periodontal diseases | digestive | LDL-C | |
| 523.1 | | | | | 25536 | 177 | 25359 | | | | | 33688 | 429 | 33259 | 0.7894 | 0.08014 | Gingivitis | digestive | LDL-C | |
| 523.3 | | | | | 25492 | 133 | 25359 | | | | | 33407 | 148 | 33259 | | | Periodontitis (acute or chronic) | digestive | LDL-C | |
| 523.31 | | | | | 25454 | 95 | 25359 | | | | | 33368 | 109 | 33259 | | | Acute periodontitis | digestive | LDL-C | |
| 523.32 | | | | | 25382 | 23 | 25359 | | | | | 33285 | 26 | 33259 | | | Chronic periodontitis | digestive | LDL-C | |
| 524 | | | | | 25495 | 136 | 25359 | | | | | 33518 | 259 | 33259 | | | Dentofacial anomalies, including malocclusion | digestive | LDL-C | |
| 524.3 | | | | | 25481 | 122 | 25359 | | | | | 33493 | 234 | 33259 | | | Anomalies of tooth position/malocclusion | digestive | LDL-C | |
| 525 | | | | | 25595 | 236 | 25359 | | | | | 33641 | 382 | 33259 | 0.1091 | 0.3127 | Other diseases of the teeth and supporting structures | digestive | LDL-C | |
| 525.1 | | | | | 25495 | 136 | 25359 | | | | | 33466 | 207 | 33259 | | | Loss of teeth or edentulism | digestive | LDL-C | |
| 525.2 | | | | | 25366 | 7 | 25359 | | | | | 33279 | 20 | 33259 | | | Atrophy of edentulous alveolar ridge | digestive | LDL-C | |
| 526 | | | | | 25636 | 277 | 25359 | 0.3721 | 0.09706 | 1.451 | 1.26e-04 | 33838 | 579 | 33259 | 4.49e-07 | 0.7615 | Diseases of the jaws | digestive | LDL-C | |
| 526.1 | | | | | 25372 | 13 | 25359 | | | | | 33270 | 11 | 33259 | | | Cysts of the jaws | digestive | LDL-C | |
| 526.3 | | | | | 25393 | 34 | 25359 | | | | | 33336 | 77 | 33259 | | | Anomalies of jaw size/symmetry | digestive | LDL-C | |
| 526.4 | | | | | 25426 | 67 | 25359 | | | | | 33523 | 264 | 33259 | | | Temporomandibular joint disorders | digestive | LDL-C | |
| 526.41 | | | | | 25407 | 48 | 25359 | | | | | 33417 | 158 | 33259 | | | Temporomandibular joint disorder, unspecified | digestive | LDL-C | |
| 526.42 | | | | | 25364 | 5 | 25359 | | | | | 33300 | 41 | 33259 | | | Arthralgia/ankylosis of temporomandibular joint | digestive | LDL-C | |
| 526.5 | | | | | 25416 | 57 | 25359 | | | | | 33320 | 61 | 33259 | | | Inflammatory conditions of jaw | digestive | LDL-C | |
| 526.8 | | | | | 25363 | 4 | 25359 | | | | | 33267 | 8 | 33259 | | | Exostosis of jaw | digestive | LDL-C | |
| 526.9 | | | | | 25414 | 55 | 25359 | | | | | 33304 | 45 | 33259 | | | Jaw disease NOS | digestive | LDL-C | |
| 527 | | | | | 25962 | 420 | 25542 | | | | | 34324 | 353 | 33971 | 5.66e-07 | 0.7597 | Diseases of the salivary glands | digestive | LDL-C | |
| 527.1 | | | | | 25545 | 3 | 25542 | | | | | 33976 | 5 | 33971 | | | Hypertrophy of salivary gland | digestive | LDL-C | |
| 527.2 | | | | | 25596 | 54 | 25542 | | | | | 34069 | 98 | 33971 | | | Sialoadenitis | digestive | LDL-C | |
| 527.7 | | | | | 25871 | 329 | 25542 | | | | | 34176 | 205 | 33971 | 1.08e-06 | 0.901 | Disturbance of salivary secretion | digestive | LDL-C | |
| 527.8 | | | | | 25561 | 19 | 25542 | | | | | 34003 | 32 | 33971 | | | Other specified diseases of the salivary glands | digestive | LDL-C | |
| 528 | 0.01568 | 0.08141 | 1.016 | 0.8473 | 26317 | 775 | 25542 | 0.3756 | 0.07883 | 1.456 | 1.89e-06 | 34849 | 878 | 33971 | 1.43e-06 | 0.4951 | Diseases of the oral soft tissues, excluding lesions specific for gingiva and tongue | digestive | LDL-C | |
| 528.1 | | | | | 25938 | 396 | 25542 | | | | | 34341 | 370 | 33971 | 2.17e-04 | 0.5412 | Stomatitis and mucositis | digestive | LDL-C | |
| 528.11 | | | | | 25631 | 89 | 25542 | | | | | 34056 | 85 | 33971 | | | Stomatitis and mucositis (ulcerative) | digestive | LDL-C | |
| 528.12 | | | | | 25583 | 41 | 25542 | | | | | 34026 | 55 | 33971 | | | Oral aphthae | digestive | LDL-C | |
| 528.3 | | | | | 25579 | 37 | 25542 | | | | | 34020 | 49 | 33971 | | | Cellulitis and abscess of oral soft tissues | digestive | LDL-C | |
| 528.4 | | | | | 25551 | 9 | 25542 | | | | | 33984 | 13 | 33971 | | | Cysts of oral soft tissues | digestive | LDL-C | |
| 528.41 | | | | | 25551 | 9 | 25542 | | | | | 33982 | 11 | 33971 | | | Cyst of the salivary gland | digestive | LDL-C | |
| 528.5 | | | | | 25587 | 45 | 25542 | | | | | 34050 | 79 | 33971 | | | Diseases of lips | digestive | LDL-C | |
| 528.6 | | | | | 25578 | 36 | 25542 | | | | | 34015 | 44 | 33971 | | | Leukoplakia of oral mucosa | digestive | LDL-C | |

| phenotype | male_beta | male_SE | male_OR | male_p | male_n_total | male_n_cases | male_n_controls | female_beta | female_SE | female_OR | female_p | female_n_total | female_n_cases | female_n_controls | interaction_p | interaction_beta | description | group | lab | label |
|---|---|---|---|---|---|---|---|---|---|---|---|---|---|---|---|---|---|---|---|---|
| 528.7 | | | | | 25551 | 9 | 25542 | | | | | 33984 | 13 | 33971 | | | Sialolithiasis | digestive | LDL-C | |
| 529 | | | | | 25647 | 105 | 25542 | | | | | 34108 | 137 | 33971 | | | Diseases and other conditions of the tongue | digestive | LDL-C | |
| 529.1 | | | | | 25565 | 23 | 25542 | | | | | 34012 | 41 | 33971 | | | Glossitis | digestive | LDL-C | |
| 529.6 | | | | | 25565 | 23 | 25542 | | | | | 34004 | 33 | 33971 | | | Glossodynia | digestive | LDL-C | |
| 530 | 0.5894 | 0.03415 | 1.803 | 9.01e-67 | 22713 | 7010 | 15703 | 0.769 | 0.03021 | 2.158 | 6.84e-143 | 30970 | 8921 | 22049 | 1.13e-09 | 0.2574 | Diseases of esophagus | digestive | LDL-C | |
| 530.1 | 0.6585 | 0.03585 | 1.932 | 2.31e-75 | 22053 | 6350 | 15703 | 0.8135 | 0.03111 | 2.256 | 1.02e-150 | 30445 | 8396 | 22049 | 6.72e-07 | 0.2196 | Esophagitis, GERD and related diseases | digestive | LDL-C | |
| 530.11 | 0.7144 | 0.03738 | 2.043 | 2.02e-81 | 21565 | 5862 | 15703 | 0.8413 | 0.03181 | 2.319 | 4.21e-154 | 30082 | 8033 | 22049 | 1.83e-04 | 0.1715 | GERD | digestive | LDL-C | |
| 530.12 | | | | | 15778 | 75 | 15703 | | | | | 22113 | 64 | 22049 | | | Ulcer of esophagus | digestive | LDL-C | |
| 530.13 | | | | | 16021 | 318 | 15703 | | | | | 22193 | 144 | 22049 | | | Barrett's esophagus | digestive | LDL-C | |
| 530.14 | | | | | 16000 | 297 | 15703 | | | | | 22411 | 362 | 22049 | 0.1773 | 0.2411 | Reflux esophagitis | digestive | LDL-C | |
| 530.15 | | | | | 15777 | 74 | 15703 | | | | | 22097 | 48 | 22049 | | | Eosinophilic esophagitis | digestive | LDL-C | |
| 530.2 | 0.2483 | 0.08983 | 1.282 | 0.005714 | 16327 | 624 | 15703 | | | | | 22465 | 416 | 22049 | 0.001997 | 0.4061 | Esophageal bleeding (varices/hemorrhage) | digestive | LDL-C | |
| 530.3 | | | | | 16140 | 437 | 15703 | | | | | 22429 | 380 | 22049 | 1.77e-04 | 0.5797 | Stricture and stenosis of esophagus | digestive | LDL-C | |
| 530.5 | | | | | 15823 | 120 | 15703 | | | | | 22257 | 208 | 22049 | | | Disorders of esophageal motility | digestive | LDL-C | |
| 530.6 | | | | | 15740 | 37 | 15703 | | | | | 22084 | 35 | 22049 | | | Diverticulum of esophagus, acquired | digestive | LDL-C | |
| 530.7 | | | | | 15735 | 32 | 15703 | | | | | 22067 | 18 | 22049 | | | Gastroesophageal laceration-hemorrhage syndrome | digestive | LDL-C | |
| 530.9 | | | | | 15920 | 217 | 15703 | | | | | 22322 | 273 | 22049 | | | Heartburn | digestive | LDL-C | |
| 531 | 0.5488 | 0.0908 | 1.731 | 1.50e-09 | 26644 | 757 | 25887 | 0.7273 | 0.09093 | 2.069 | 1.26e-15 | 35675 | 773 | 34902 | 0.1108 | 0.1882 | Peptic ulcer (excl. esophageal) | digestive | LDL-C | |
| 531.1 | | | | | 26046 | 159 | 25887 | | | | | 35004 | 102 | 34902 | | | Hemorrhage from gastrointestinal ulcer | digestive | LDL-C | |
| 531.2 | | | | | 26063 | 176 | 25887 | | | | | 35135 | 233 | 34902 | | | Gastric ulcer | digestive | LDL-C | |
| 531.3 | | | | | 26049 | 162 | 25887 | | | | | 35017 | 115 | 34902 | | | Duodenal ulcer | digestive | LDL-C | |
| 531.4 | | | | | 26128 | 241 | 25887 | | | | | 35173 | 271 | 34902 | 0.4548 | 0.1556 | Peptic ulcer, site unspecified | digestive | LDL-C | |
| 531.5 | | | | | 25904 | 17 | 25887 | | | | | 34952 | 50 | 34902 | | | Gastrojejunal ulcer | digestive | LDL-C | |
| 532 | 0.2819 | 0.04561 | 1.326 | 6.38e-10 | 18697 | 2994 | 15703 | 0.6565 | 0.04564 | 1.928 | 6.63e-47 | 25127 | 3078 | 22049 | 9.30e-19 | 0.5209 | Dysphagia | digestive | LDL-C | |
| 535 | 0.1443 | 0.08554 | 1.155 | 0.0917 | 24211 | 698 | 23513 | 0.3681 | 0.07624 | 1.445 | 1.37e-06 | 32091 | 888 | 31203 | 4.17e-04 | 0.3708 | Gastritis and duodenitis | digestive | LDL-C | |
| 535.1 | | | | | 23554 | 41 | 23513 | | | | | 31256 | 53 | 31203 | | | Acute gastritis | digestive | LDL-C | |
| 535.2 | | | | | 23753 | 240 | 23513 | | | | | 31519 | 316 | 31203 | 0.001118 | 0.5665 | Atrophic gastritis | digestive | LDL-C | |
| 535.6 | | | | | 23634 | 121 | 23513 | | | | | 31294 | 91 | 31203 | | | Duodenitis | digestive | LDL-C | |
| 535.8 | | | | | 23673 | 160 | 23513 | | | | | 31440 | 237 | 31203 | | | Other specified gastritis | digestive | LDL-C | |
| 535.9 | | | | | 23653 | 140 | 23513 | | | | | 31383 | 180 | 31203 | | | Gastritis and duodenitis, NOS | digestive | LDL-C | |
| 536 | 0.4178 | 0.09546 | 1.519 | 1.20e-05 | 24111 | 598 | 23513 | 0.553 | 0.07207 | 1.739 | 1.67e-14 | 32274 | 1071 | 31203 | 0.01879 | 0.2572 | Disorders of function of stomach | digestive | LDL-C | |
| 536.3 | | | | | 23695 | 182 | 23513 | | | | | 31598 | 395 | 31203 | 0.6553 | -0.09014 | Gastroparesis | digestive | LDL-C | |
| 536.7 | | | | | 23722 | 209 | 23513 | | | | | 31426 | 223 | 31203 | | | Complications of gastrostomy, colostomy and enterostomy | digestive | LDL-C | |
| 536.8 | | | | | 23738 | 225 | 23513 | 0.7402 | 0.1095 | 2.096 | 1.35e-11 | 31704 | 501 | 31203 | 0.2427 | 0.2115 | Dyspepsia and other specified disorders of function of stomach | digestive | LDL-C | |
| 537 | 0.3352 | 0.08907 | 1.398 | 1.68e-04 | 24195 | 682 | 23513 | 0.5073 | 0.0865 | 1.661 | 4.48e-09 | 31926 | 723 | 31203 | 0.1361 | 0.1692 | Other disorders of stomach and duodenum | digestive | LDL-C | |
| 537.1 | | | | | 23574 | 61 | 23513 | | | | | 31261 | 58 | 31203 | | | Lesions of stomach and duodenum | digestive | LDL-C | |
| 539 | | | | | 27703 | 290 | 27413 | 0.8648 | 0.07983 | 2.375 | 2.38e-27 | 36747 | 945 | 35802 | 0.3514 | 0.1397 | Bariatric surgery | digestive | LDL-C | |

| phenotype | male_beta | male_SE | male_OR | male_p | male_n_total | male_n_cases | male_n_controls | female_beta | female_SE | female_OR | female_p | female_n_total | female_n_cases | female_n_controls | interaction_p | interaction_beta | description | group | lab | label |
|---|---|---|---|---|---|---|---|---|---|---|---|---|---|---|---|---|---|---|---|---|
| 540 | | | | | 27635 | 152 | 27483 | | | | | 36729 | 193 | 36536 | | | Appendiceal conditions | digestive | LDL-C | |
| 540.1 | | | | | 27611 | 128 | 27483 | | | | | 36714 | 178 | 36536 | | | Appendicitis | digestive | LDL-C | |
| 540.11 | | | | | 27592 | 109 | 27483 | | | | | 36692 | 156 | 36536 | | | Acute appendicitis | digestive | LDL-C | |
| 550 | 0.4282 | 0.04837 | 1.535 | 8.53e-19 | 25722 | 2883 | 22839 | 0.4368 | 0.0493 | 1.548 | 7.93e-19 | 34582 | 2531 | 32051 | 0.2943 | -0.06666 | Abdominal hernia | digestive | LDL-C | |
| 550.1 | 0.6937 | 0.08783 | 2.001 | 2.82e-15 | 23813 | 974 | 22839 | | | | | 32167 | 116 | 32051 | 0.9682 | 0.009745 | Inguinal hernia | digestive | LDL-C | |
| 550.2 | 0.4203 | 0.08082 | 1.522 | 1.98e-07 | 23793 | 954 | 22839 | 0.5669 | 0.0674 | 1.763 | 4.04e-17 | 33474 | 1423 | 32051 | 0.2757 | 0.1057 | Diaphragmatic hernia | digestive | LDL-C | |
| 550.3 | | | | | 22846 | 7 | 22839 | | | | | 32067 | 16 | 32051 | | | Femoral hernia | digestive | LDL-C | |
| 550.4 | | | | | 23250 | 411 | 22839 | | | | | 32268 | 217 | 32051 | 0.07633 | -0.3647 | Umbilical hernia | digestive | LDL-C | |
| 550.5 | | | | | 23322 | 483 | 22839 | 0.2195 | 0.09897 | 1.245 | 0.02655 | 32584 | 533 | 32051 | 0.6446 | 0.06104 | Ventral hernia | digestive | LDL-C | |
| 550.6 | -0.0915 | 0.09749 | 0.9126 | 0.348 | 23364 | 525 | 22839 | 0.1621 | 0.09626 | 1.176 | 0.09222 | 32610 | 559 | 32051 | 0.03067 | 0.272 | Incisional hernia | digestive | LDL-C | |
| 555 | -0.08354 | 0.06866 | 0.9199 | 0.2237 | 18550 | 1092 | 17458 | -0.1138 | 0.05982 | 0.8925 | 0.05718 | 24902 | 1438 | 23464 | 0.4802 | 0.06032 | Inflammatory bowel disease and other gastroenteritis and colitis | digestive | LDL-C | |
| 555.1 | -0.1642 | 0.07685 | 0.8486 | 0.03269 | 18342 | 884 | 17458 | -0.1816 | 0.06585 | 0.8339 | 0.005816 | 24671 | 1207 | 23464 | 0.409 | 0.07861 | Regional enteritis | digestive | LDL-C | |
| 555.2 | -0.001248 | 0.09331 | 0.9988 | 0.9893 | 18001 | 543 | 17458 | 0.1214 | 0.08471 | 1.129 | 0.1517 | 24116 | 652 | 23464 | 0.06296 | 0.2213 | Ulcerative colitis | digestive | LDL-C | |
| 555.21 | | | | | 17689 | 231 | 17458 | | | | | 23710 | 246 | 23464 | | | Ulcerative colitis (chronic) | digestive | LDL-C | |
| 556 | | | | | 17670 | 212 | 17458 | | | | | 23667 | 203 | 23464 | | | Ulceration of the lower GI tract | digestive | LDL-C | |
| 556.1 | | | | | 17633 | 175 | 17458 | | | | | 23632 | 168 | 23464 | | | Ulceration of intestine | digestive | LDL-C | |
| 556.11 | | | | | 17498 | 40 | 17458 | | | | | 23504 | 40 | 23464 | | | Angiodysplasia of intestine (without mention of hemorrhage) | digestive | LDL-C | |
| 557 | | | | | 17645 | 187 | 17458 | | | | | 23820 | 356 | 23464 | 0.005025 | 0.5611 | Intestinal malabsorption (non-celiac) | digestive | LDL-C | |
| 557.1 | | | | | 17538 | 80 | 17458 | | | | | 23734 | 270 | 23464 | | | Celiac disease | digestive | LDL-C | |
| 558 | 0.3445 | 0.06994 | 1.411 | 8.38e-07 | 18499 | 1041 | 17458 | 0.5195 | 0.05781 | 1.681 | 2.57e-19 | 25010 | 1546 | 23464 | 2.40e-04 | 0.3128 | Noninfectious gastroenteritis | digestive | LDL-C | |
| 559 | | | | | 17873 | 415 | 17458 | | | | | 23811 | 347 | 23464 | 0.02948 | 0.3244 | Ileostomy status | digestive | LDL-C | |
| 560 | 0.04006 | 0.061 | 1.041 | 0.5114 | 18805 | 1347 | 17458 | 0.1488 | 0.06404 | 1.16 | 0.02018 | 24662 | 1198 | 23464 | 0.05086 | 0.1608 | Intestinal obstruction without mention of hernia | digestive | LDL-C | |
| 560.1 | 0.03106 | 0.0836 | 1.032 | 0.7103 | 18153 | 695 | 17458 | | | | | 23849 | 385 | 23464 | 0.2197 | 0.1583 | Paralytic ileus | digestive | LDL-C | |
| 560.2 | | | | | 17514 | 56 | 17458 | | | | | 23568 | 104 | 23464 | | | Impaction of intestine | digestive | LDL-C | |
| 560.3 | | | | | 17538 | 80 | 17458 | | | | | 23554 | 90 | 23464 | | | Peritoneal or intestinal adhesions | digestive | LDL-C | |
| 560.4 | 0.0408 | 0.08305 | 1.042 | 0.6233 | 18149 | 691 | 17458 | 0.1295 | 0.07903 | 1.138 | 0.1013 | 24229 | 765 | 23464 | 0.3081 | 0.1093 | Other intestinal obstruction | digestive | LDL-C | |
| 561 | 0.7105 | 0.05889 | 2.035 | 1.62e-33 | 19227 | 1769 | 17458 | 0.8971 | 0.04671 | 2.452 | 3.35e-82 | 26443 | 2979 | 23464 | 9.25e-05 | 0.2795 | Symptoms involving digestive system | digestive | LDL-C | |
| 562 | 1.289 | 0.06431 | 3.628 | 2.54e-89 | 19778 | 2320 | 17458 | 1.326 | 0.06459 | 3.766 | 1.22e-93 | 25685 | 2221 | 23464 | 0.6457 | 0.04024 | Diverticulosis and diverticulitis | digestive | LDL-C | |
| 562.1 | 1.371 | 0.06918 | 3.941 | 1.86e-87 | 19570 | 2112 | 17458 | 1.328 | 0.0697 | 3.773 | 6.29e-81 | 25403 | 1939 | 23464 | 0.8155 | -0.02211 | Diverticulosis | digestive | LDL-C | |
| 562.2 | | | | | 17837 | 379 | 17458 | 1.196 | 0.1227 | 3.307 | 1.92e-22 | 23966 | 502 | 23464 | 0.2186 | 0.2132 | Diverticulitis | digestive | LDL-C | |
| 564 | 0.5435 | 0.06364 | 1.722 | 1.35e-17 | 18857 | 1399 | 17458 | 0.794 | 0.04875 | 2.212 | 1.22e-59 | 26038 | 2574 | 23464 | 1.53e-05 | 0.3285 | Functional digestive disorders | digestive | LDL-C | |
| 564.1 | | | | | 17914 | 456 | 17458 | 1.014 | 0.06346 | 2.757 | 1.81e-57 | 25043 | 1579 | 23464 | 0.3916 | 0.109 | Irritable Bowel Syndrome | digestive | LDL-C | |
| 564.9 | | | | | 17664 | 206 | 17458 | | | | | 23673 | 209 | 23464 | | | Personal history of diseases of digestive system | digestive | LDL-C | |
| 565 | 0.5979 | 0.07246 | 1.818 | 1.56e-16 | 23018 | 1093 | 21925 | 0.3076 | 0.0688 | 1.36 | 7.81e-06 | 31069 | 1106 | 29963 | 0.4643 | -0.0672 | Anal and rectal conditions | digestive | LDL-C | |
| 565.1 | | | | | 22052 | 127 | 21925 | | | | | 30077 | 114 | 29963 | | | Anal and rectal polyp | digestive | LDL-C | |

| phenotype | male_beta | male_SE | male_OR | male_p | male_n_total | male_n_cases | male_n_controls | female_beta | female_SE | female_OR | female_p | female_n_total | female_n_cases | female_n_controls | interaction_p | interaction_beta | description | group | lab | label |
|---|---|---|---|---|---|---|---|---|---|---|---|---|---|---|---|---|---|---|---|---|
| 567 | 0.08739 | 0.08735 | 1.091 | 0.3171 | 22577 | 652 | 21925 | 0.219 | 0.09291 | 1.245 | 0.01839 | 30541 | 578 | 29963 | 0.009337 | 0.3018 | Peritonitis and retroperitoneal infections | digestive | LDL-C | |
| 568 | | | | | 22412 | 487 | 21925 | -0.06343 | 0.09722 | 0.9385 | 0.5141 | 30476 | 513 | 29963 | 0.06622 | 0.2352 | Other disorders of peritoneum | digestive | LDL-C | |
| 568.1 | | | | | 22164 | 239 | 21925 | | | | | 30277 | 314 | 29963 | 0.0704 | 0.3132 | Peritoneal adhesions (postoperative) (postinfection) | digestive | LDL-C | |
| 569 | 0.0632 | 0.05929 | 1.065 | 0.2864 | 23419 | 1494 | 21925 | 0.4179 | 0.05195 | 1.519 | 8.73e-16 | 31975 | 2012 | 29963 | 1.40e-09 | 0.436 | Other disorders of intestine | digestive | LDL-C | |
| 569.1 | | | | | 21954 | 29 | 21925 | | | | | 30021 | 58 | 29963 | | | Toxic gastroenteritis and colitis | digestive | LDL-C | |
| 569.2 | 0.2234 | 0.09473 | 1.25 | 0.01837 | 22496 | 571 | 21925 | 0.7162 | 0.07505 | 2.047 | 1.37e-21 | 30942 | 979 | 29963 | 1.17e-05 | 0.4849 | Gastrointestinal complications | digestive | LDL-C | |
| 571 | 0.4662 | 0.04932 | 1.594 | 3.33e-21 | 21552 | 2387 | 19165 | 0.5469 | 0.05064 | 1.728 | 3.47e-27 | 29934 | 2235 | 27699 | 0.002679 | 0.1924 | Chronic liver disease and cirrhosis | digestive | LDL-C | |
| 571.5 | 0.4886 | 0.05112 | 1.63 | 1.19e-21 | 21382 | 2217 | 19165 | 0.5907 | 0.05296 | 1.805 | 6.95e-29 | 29754 | 2055 | 27699 | 0.00114 | 0.2176 | Other chronic nonalcoholic liver disease | digestive | LDL-C | |
| 571.51 | 0.2522 | 0.06363 | 1.287 | 7.36e-05 | 20477 | 1312 | 19165 | 0.4025 | 0.07422 | 1.496 | 5.87e-08 | 28658 | 959 | 27699 | 0.04645 | 0.1753 | Cirrhosis of liver without mention of alcohol | digestive | LDL-C | |
| 571.6 | | | | | 19242 | 77 | 19165 | | | | | 27883 | 184 | 27699 | | | Primary biliary cirrhosis | digestive | LDL-C | |
| 571.8 | 0.318 | 0.0722 | 1.374 | 1.06e-05 | 20161 | 996 | 19165 | 0.4562 | 0.08406 | 1.578 | 5.73e-08 | 28437 | 738 | 27699 | 0.1374 | 0.1487 | Liver abscess and sequelae of chronic liver disease | digestive | LDL-C | |
| 571.81 | 0.3284 | 0.08599 | 1.389 | 1.34e-04 | 19856 | 691 | 19165 | | | | | 28168 | 469 | 27699 | 0.03583 | 0.2597 | Portal hypertension | digestive | LDL-C | |
| 572 | 0.2949 | 0.06151 | 1.343 | 1.64e-06 | 20559 | 1394 | 19165 | 0.3192 | 0.06517 | 1.376 | 9.66e-07 | 28923 | 1224 | 27699 | 0.3074 | 0.08241 | Ascites (non malignant) | digestive | LDL-C | |
| 573 | 0.4438 | 0.07512 | 1.559 | 3.47e-09 | 20126 | 961 | 19165 | 0.5138 | 0.07504 | 1.672 | 7.56e-12 | 28655 | 956 | 27699 | 0.3031 | 0.09885 | Other disorders of liver | digestive | LDL-C | |
| 573.1 | | | | | 19195 | 30 | 19165 | | | | | 27731 | 32 | 27699 | | | Chronic passive congestion of liver | digestive | LDL-C | |
| 573.2 | | | | | 19596 | 431 | 19165 | | | | | 27945 | 246 | 27699 | 9.35e-04 | 0.616 | Liver replaced by transplant | digestive | LDL-C | |
| 573.3 | | | | | 19292 | 127 | 19165 | | | | | 27860 | 161 | 27699 | | | Hepatomegaly | digestive | LDL-C | |
| 573.4 | | | | | 19356 | 191 | 19165 | | | | | 27881 | 182 | 27699 | | | Acute and subacute necrosis of liver | digestive | LDL-C | |
| 573.5 | | | | | 19606 | 441 | 19165 | | | | | 28006 | 307 | 27699 | 0.566 | -0.08624 | Jaundice (not of newborn) | digestive | LDL-C | |
| 573.6 | 1.215 | 0.1048 | 3.371 | 4.15e-31 | 19797 | 632 | 19165 | 1.007 | 0.1043 | 2.737 | 4.86e-22 | 28268 | 569 | 27699 | 0.6176 | -0.06871 | Nonspecific elevation of levels of transaminase or lactic acid dehydrogenase [LDH] | digestive | LDL-C | |
| 573.7 | 1.079 | 0.08578 | 2.943 | 2.62e-36 | 20119 | 954 | 19165 | 0.8709 | 0.07499 | 2.389 | 3.54e-31 | 28828 | 1129 | 27699 | 0.4516 | -0.07952 | Abnormal results of function study of liver | digestive | LDL-C | |
| 573.9 | 1.058 | 0.09326 | 2.881 | 7.79e-30 | 19944 | 779 | 19165 | 1.019 | 0.08561 | 2.769 | 1.22e-32 | 28587 | 888 | 27699 | 0.4894 | 0.08129 | Abnormal serum enzyme levels | digestive | LDL-C | |
| 574 | 0.6055 | 0.07111 | 1.832 | 1.66e-17 | 25697 | 1283 | 24414 | 0.7411 | 0.06507 | 2.098 | 4.69e-30 | 34656 | 1503 | 33153 | 0.0923 | 0.1489 | Cholelithiasis and cholecystitis | digestive | LDL-C | |
| 574.1 | 0.6609 | 0.08198 | 1.937 | 7.54e-16 | 25404 | 990 | 24414 | 0.7543 | 0.07382 | 2.126 | 1.64e-24 | 34320 | 1167 | 33153 | 0.4726 | 0.07293 | Cholelithiasis | digestive | LDL-C | |
| 574.11 | | | | | 24503 | 89 | 24414 | | | | | 33224 | 71 | 33153 | | | Cholelithiasis with acute cholecystitis | digestive | LDL-C | |

| phenotype | male_beta | male_SE | male_OR | male_p | male_n_total | male_n_cases | male_n_controls | female_beta | female_SE | female_OR | female_p | female_n_total | female_n_cases | female_n_controls | interaction_p | interaction_beta | description | group | lab | label |
|---|---|---|---|---|---|---|---|---|---|---|---|---|---|---|---|---|---|---|---|---|
| 574.12 | | | | | 24528 | 114 | 24414 | | | | | 33340 | 187 | 33153 | | | Cholelithiasis with other cholecystitis | digestive | LDL-C | |
| 574.2 | | | | | 24625 | 211 | 24414 | | | | | 33383 | 230 | 33153 | | | Calculus of bile duct | digestive | LDL-C | |
| 574.3 | | | | | 24711 | 297 | 24414 | | | | | 33444 | 291 | 33153 | 0.3793 | 0.1692 | Cholecystitis without cholelithiasis | digestive | LDL-C | |
| 575 | 0.2998 | 0.07859 | 1.35 | 1.37e-04 | 25285 | 871 | 24414 | 0.4087 | 0.07531 | 1.505 | 5.73e-08 | 34100 | 947 | 33153 | 0.04018 | 0.201 | Other biliary tract disease | digestive | LDL-C | |
| 575.1 | | | | | 24612 | 198 | 24414 | | | | | 33277 | 124 | 33153 | | | Cholangitis | digestive | LDL-C | |
| 575.2 | | | | | 24699 | 285 | 24414 | | | | | 33392 | 239 | 33153 | 0.6379 | 0.08324 | Obstruction of bile duct | digestive | LDL-C | |
| 575.6 | | | | | 24435 | 21 | 24414 | | | | | 33185 | 32 | 33153 | | | Cholesterolosis of gallbladder | digestive | LDL-C | |
| 575.7 | | | | | 24611 | 197 | 24414 | | | | | 33370 | 217 | 33153 | | | Other disorders of gallbladder | digestive | LDL-C | |
| 575.8 | | | | | 24783 | 369 | 24414 | | | | | 33584 | 431 | 33153 | 0.7091 | 0.05453 | Other disorders of biliary tract | digestive | LDL-C | |

| phenotype | male_beta | male_SE | male_OR | male_p | male_n_total | male_n_cases | male_n_controls | female_beta | female_SE | female_OR | female_p | female_n_total | female_n_cases | female_n_controls | interaction_p | interaction_beta | description | group | lab | label |
|---|---|---|---|---|---|---|---|---|---|---|---|---|---|---|---|---|---|---|---|---|
| 575.9 | | | | | 24488 | 74 | 24414 | | | | | 33244 | 91 | 33153 | | | Nonspecific abnormal findings on radiological and other examination of biliary tract | digestive | LDL-C | |
| 577 | 0.4364 | 0.08043 | 1.547 | 5.78e-08 | 27246 | 863 | 26383 | 0.3768 | 0.08055 | 1.458 | 2.90e-06 | 36384 | 840 | 35544 | 0.9215 | 0.01009 | Diseases of pancreas | digestive | LDL-C | |
| 577.1 | | | | | 26693 | 310 | 26383 | | | | | 35828 | 284 | 35544 | 0.596 | 0.09508 | Acute pancreatitis | digestive | LDL-C | |
| 577.2 | | | | | 26568 | 185 | 26383 | | | | | 35726 | 182 | 35544 | | | Chronic pancreatitis | digestive | LDL-C | |
| 577.3 | | | | | 26620 | 237 | 26383 | | | | | 35810 | 266 | 35544 | 0.7663 | -0.05604 | Cyst and pseudocyst of pancreas | digestive | LDL-C | |
| 578 | 0.7268 | 0.05171 | 2.068 | 7.13e-45 | 23398 | 2404 | 20994 | 0.801 | 0.04947 | 2.228 | 5.63e-59 | 31110 | 2548 | 28562 | 0.03988 | 0.1357 | Gastrointestinal hemorrhage | digestive | LDL-C | |
| 578.1 | | | | | 21273 | 279 | 20994 | | | | | 28819 | 257 | 28562 | 0.3636 | 0.1646 | Hematemesis | digestive | LDL-C | |
| 578.2 | 0.8208 | 0.07592 | 2.272 | 3.06e-27 | 22097 | 1103 | 20994 | 0.7678 | 0.07099 | 2.155 | 2.90e-27 | 29747 | 1185 | 28562 | 0.8488 | 0.01841 | Blood in stool | digestive | LDL-C | |
| 578.8 | 0.7846 | 0.09986 | 2.192 | 3.93e-15 | 21622 | 628 | 20994 | 0.8006 | 0.09057 | 2.227 | 9.65e-19 | 29288 | 726 | 28562 | 0.4935 | 0.08631 | Hemorrhage of rectum and anus | digestive | LDL-C | |
| 578.9 | 0.6875 | 0.07641 | 1.989 | 2.31e-19 | 22034 | 1040 | 20994 | 0.7715 | 0.08466 | 2.163 | 8.00e-20 | 29380 | 818 | 28562 | 0.5348 | 0.06475 | Hemorrhage of gastrointestinal tract | digestive | LDL-C | |
| 579 | 0.4598 | 0.0605 | 1.584 | 2.96e-14 | 22498 | 1504 | 20994 | 0.5362 | 0.05226 | 1.709 | 1.06e-24 | 30586 | 2024 | 28562 | 0.05366 | 0.1414 | Other symptoms involving abdomen and pelvis | digestive | LDL-C | |
| 579.2 | 0.5094 | 0.09447 | 1.664 | 6.97e-08 | 21574 | 580 | 20994 | | | | | 28950 | 388 | 28562 | 0.1579 | 0.1945 | Splenomegaly | digestive | LDL-C | |
| 579.8 | | | | | 21190 | 196 | 20994 | | | | | 28810 | 248 | 28562 | | | Nonspecific abnormal findings in stool contents | digestive | LDL-C | |
| 580 | 1.476 | 0.1123 | 4.375 | 1.92e-39 | 18292 | 656 | 17636 | 1.295 | 0.1098 | 3.65 | 4.33e-32 | 28613 | 588 | 28025 | 0.3143 | -0.1495 | Nephritis; nephrosis; renal sclerosis | genitourinary | LDL-C | |
| 580.1 | | | | | 17811 | 175 | 17636 | | | | | 28185 | 160 | 28025 | | | Glomerulonephritis | genitourinary | LDL-C | |
| 580.11 | | | | | 17690 | 54 | 17636 | | | | | 28073 | 48 | 28025 | | | Proliferative glomerulonephritis | genitourinary | LDL-C | |
| 580.12 | | | | | 17715 | 79 | 17636 | | | | | 28084 | 59 | 28025 | | | Non-proliferative glomerulonephritis | genitourinary | LDL-C | |
| 580.13 | | | | | 17654 | 18 | 17636 | | | | | 28048 | 23 | 28025 | | | Acute glomerulonephritis, NOS | genitourinary | LDL-C | |
| 580.14 | | | | | 17676 | 40 | 17636 | | | | | 28076 | 51 | 28025 | | | Chronic glomerulonephritis, NOS | genitourinary | LDL-C | |
| 580.2 | | | | | 17776 | 140 | 17636 | | | | | 28143 | 118 | 28025 | | | Nephrotic syndrome without mention of glomerulonephritis | genitourinary | LDL-C | |
| 580.3 | | | | | 17973 | 337 | 17636 | | | | | 28368 | 343 | 28025 | 0.7268 | -0.07208 | Nephritis and nephropathy without mention of glomerulonephritis | genitourinary | LDL-C | |
| 580.31 | | | | | 17810 | 174 | 17636 | | | | | 28242 | 217 | 28025 | | | Nephritis and nephropathy in diseases classified elsewhere | genitourinary | LDL-C | |
| 580.32 | | | | | 17795 | 159 | 17636 | | | | | 28162 | 137 | 28025 | | | Nephritis and nephropathy with pathological lesion | genitourinary | LDL-C | |
| 580.4 | | | | | 17734 | 98 | 17636 | | | | | 28125 | 100 | 28025 | | | Renal sclerosis, NOS | genitourinary | LDL-C | |
| 585 | 0.8144 | 0.03536 | 2.258 | 2.46e-117 | 24023 | 6387 | 17636 | 0.8377 | 0.03789 | 2.311 | 2.46e-108 | 32893 | 4868 | 28025 | 0.8103 | 0.01121 | Renal failure | genitourinary | LDL-C | |
| 585.1 | 0.7761 | 0.03934 | 2.173 | 1.24e-86 | 22356 | 4720 | 17636 | 0.8036 | 0.04314 | 2.233 | 1.97e-77 | 31559 | 3534 | 28025 | 0.5507 | 0.03126 | Acute renal failure | genitourinary | LDL-C | |
| 585.2 | 1.298 | 0.1071 | 3.663 | 8.53e-34 | 18192 | 556 | 17636 | | | | | 28438 | 413 | 28025 | 0.5025 | 0.1004 | Renal failure NOS | genitourinary | LDL-C | |
| 585.3 | 1.312 | 0.04861 | 3.714 | 1.59e-160 | 21314 | 3678 | 17636 | 1.249 | 0.05516 | 3.488 | 1.36e-113 | 30569 | 2544 | 28025 | 0.3472 | -0.0633 | Chronic renal failure [CKD] | genitourinary | LDL-C | |
| 585.31 | 1.736 | 0.1137 | 5.677 | 1.20e-52 | 18326 | 690 | 17636 | | | | | 28496 | 471 | 28025 | 0.4401 | 0.1315 | Renal dialysis | genitourinary | LDL-C | |
| 585.32 | 1.818 | 0.09424 | 6.162 | 5.89e-83 | 18620 | 984 | 17636 | 1.781 | 0.112 | 5.936 | 6.78e-57 | 28699 | 674 | 28025 | 0.9055 | -0.01614 | End stage renal disease | genitourinary | LDL-C | |
| 585.33 | 1.556 | 0.07234 | 4.741 | 1.19e-102 | 19559 | 1923 | 17636 | 1.399 | 0.08127 | 4.052 | 2.03e-66 | 29378 | 1353 | 28025 | 0.1677 | -0.14 | Chronic Kidney Disease, Stage III | genitourinary | LDL-C | |
| 585.34 | 1.779 | 0.1065 | 5.921 | 1.25e-62 | 18575 | 939 | 17636 | 1.563 | 0.113 | 4.774 | 1.53e-43 | 28729 | 704 | 28025 | 0.1102 | -0.2321 | Chronic Kidney Disease, Stage IV | genitourinary | LDL-C | |
| 585.4 | 1.443 | 0.1072 | 4.234 | 2.65e-41 | 18446 | 810 | 17636 | 1.228 | 0.1088 | 3.413 | 1.65e-29 | 28650 | 625 | 28025 | 0.04122 | -0.2951 | Chronic kidney disease, Stage I or II | genitourinary | LDL-C | |

| phenotype | male_beta | male_SE | male_OR | male_p | male_n_total | male_n_cases | male_n_controls | female_beta | female_SE | female_OR | female_p | female_n_total | female_n_cases | female_n_controls | interaction_p | interaction_beta | description | group | lab | label |
|---|---|---|---|---|---|---|---|---|---|---|---|---|---|---|---|---|---|---|---|---|
| 586 | 0.7787 | 0.04653 | 2.179 | 7.06e-63 | 21065 | 3429 | 17636 | 0.6671 | 0.05161 | 1.949 | 3.23e-38 | 30360 | 2335 | 28025 | 0.01085 | -0.1602 | Other disorders of the kidney and ureters | genitourinary | LDL-C | |
| 586.1 | | | | | 17687 | 51 | 17636 | | | | | 28103 | 78 | 28025 | | | Anatomical abnormatilies of kidney and ureters | genitourinary | LDL-C | |
| 586.11 | | | | | 17638 | 2 | 17636 | | | | | 28033 | 8 | 28025 | | | Small kidney | genitourinary | LDL-C | |
| 586.12 | | | | | 17684 | 48 | 17636 | | | | | 28097 | 72 | 28025 | | | Vesicoureteral reflux | genitourinary | LDL-C | |
| 586.2 | 0.7226 | 0.09209 | 2.06 | 4.25e-15 | 18502 | 866 | 17636 | 0.7776 | 0.1147 | 2.176 | 1.20e-11 | 28545 | 520 | 28025 | 0.9887 | 0.00195 | Cyst of kidney, acquired | genitourinary | LDL-C | |
| 586.3 | | | | | 17685 | 49 | 17636 | | | | | 28057 | 32 | 28025 | | | Vascular disorders of kidney/hypertrophy | genitourinary | LDL-C | |
| 586.4 | 0.4669 | 0.0989 | 1.595 | 2.34e-06 | 18326 | 690 | 17636 | | | | | 28313 | 288 | 28025 | 3.18e-19 | -1.343 | Stricture/obstruction of ureter | genitourinary | LDL-C | Stricture/obstruction of ureter |
| 587 | 2.639 | 0.1779 | 14 | 8.28e-50 | 18201 | 565 | 17636 | | | | | 28416 | 391 | 28025 | 0.6794 | 0.1128 | Kidney replaced by transpant | genitourinary | LDL-C | |
| 588 | 1.462 | 0.09972 | 4.313 | 1.21e-48 | 18557 | 921 | 17636 | 1.535 | 0.1146 | 4.64 | 7.10e-41 | 28718 | 693 | 28025 | 0.6288 | 0.06947 | Disorders resulting from impaired renal function | genitourinary | LDL-C | |
| 588.1 | 1.65 | 0.1161 | 5.207 | 8.42e-46 | 18381 | 745 | 17636 | 1.599 | 0.1334 | 4.948 | 4.28e-33 | 28555 | 530 | 28025 | 0.9143 | -0.01807 | Renal osteodystrophy | genitourinary | LDL-C | |
| 588.2 | | | | | 17925 | 289 | 17636 | | | | | 28246 | 221 | 28025 | 0.1648 | 0.3641 | Secondary hyperparathyroidism (of renal origin) | genitourinary | LDL-C | |
| 589 | | | | | 17733 | 97 | 17636 | | | | | 28140 | 115 | 28025 | | | Abnormal results of function study of kidney | genitourinary | LDL-C | |
| 590 | | | | | 21698 | 323 | 21375 | 0.655 | 0.08371 | 1.925 | 5.09e-15 | 24682 | 746 | 23936 | 0.5795 | -0.08305 | Pyelonephritis | genitourinary | LDL-C | |
| 591 | 0.6323 | 0.05103 | 1.882 | 2.92e-35 | 23909 | 2534 | 21375 | 0.8571 | 0.03303 | 2.356 | 1.76e-148 | 30676 | 6740 | 23936 | 5.83e-09 | 0.3253 | Urinary tract infection | genitourinary | LDL-C | |
| 592 | | | | | 21768 | 393 | 21375 | 1.138 | 0.07079 | 3.121 | 3.75e-58 | 25433 | 1497 | 23936 | 0.01116 | 0.367 | Cystitis and urethritis | genitourinary | LDL-C | |
| 592.1 | | | | | 21707 | 332 | 21375 | 1.193 | 0.07333 | 3.297 | 1.71e-59 | 25375 | 1439 | 23936 | 0.02482 | 0.3558 | Cystitis | genitourinary | LDL-C | |
| 592.11 | | | | | 21543 | 168 | 21375 | 1.527 | 0.1048 | 4.607 | 4.45e-48 | 24834 | 898 | 23936 | 0.4006 | 0.2228 | Acute cystitis | genitourinary | LDL-C | |
| 592.12 | | | | | 21411 | 36 | 21375 | | | | | 23989 | 53 | 23936 | | | Chronic cystitis | genitourinary | LDL-C | |
| 592.13 | | | | | 21383 | 8 | 21375 | | | | | 24169 | 233 | 23936 | | | Chronic interstitial cystitis | genitourinary | LDL-C | |
| 592.2 | | | | | 21408 | 33 | 21375 | | | | | 23967 | 31 | 23936 | | | Urethritis and urethral syndrome | genitourinary | LDL-C | |
| 592.21 | | | | | 21378 | 3 | 21375 | | | | | 23938 | 2 | 23936 | | | Urethral syndrome | genitourinary | LDL-C | |
| 592.3 | | | | | 21378 | 3 | 21375 | | | | | 23938 | 2 | 23936 | | | Urethral stricture due to infecton | genitourinary | LDL-C | |
| 593 | 0.8334 | 0.0637 | 2.301 | 4.11e-39 | 23190 | 1815 | 21375 | 1.139 | 0.06362 | 3.123 | 1.20e-71 | 25739 | 1803 | 23936 | 3.01e-06 | 0.3963 | Hematuria | genitourinary | LDL-C | |
| 593.1 | 0.706 | 0.105 | 2.026 | 1.74e-11 | 22020 | 645 | 21375 | | | | | 24248 | 312 | 23936 | 0.02271 | 0.3915 | Gross hematuria | genitourinary | LDL-C | |
| 593.2 | | | | | 21568 | 193 | 21375 | | | | | 24188 | 252 | 23936 | | | Microscopic hematuria | genitourinary | LDL-C | |
| 594 | 0.4919 | 0.05951 | 1.635 | 1.39e-16 | 25965 | 1823 | 24142 | 0.3854 | 0.05907 | 1.47 | 6.78e-11 | 35229 | 1634 | 33595 | 0.02147 | -0.1772 | Urinary calculus | genitourinary | LDL-C | |
| 594.1 | 0.5368 | 0.06656 | 1.71 | 7.34e-16 | 25599 | 1457 | 24142 | 0.3951 | 0.06482 | 1.484 | 1.10e-09 | 34942 | 1347 | 33595 | 0.01324 | -0.2119 | Calculus of kidney | genitourinary | LDL-C | |
| 594.2 | | | | | 24276 | 134 | 24142 | | | | | 33649 | 54 | 33595 | | | Calculus of lower urinary tract | genitourinary | LDL-C | |
| 594.3 | | | | | 24598 | 456 | 24142 | | | | | 33995 | 400 | 33595 | 0.7326 | -0.05204 | Calculus of ureter | genitourinary | LDL-C | |
| 594.8 | | | | | 24179 | 37 | 24142 | | | | | 33649 | 54 | 33595 | | | Renal colic | genitourinary | LDL-C | |
| 595 | -0.03049 | 0.0831 | 0.97 | 0.7137 | 24902 | 760 | 24142 | -0.1114 | 0.08363 | 0.8946 | 0.1827 | 34327 | 732 | 33595 | 0.2621 | -0.119 | Hydronephrosis | genitourinary | LDL-C | |

| phenotype | male_beta | male_SE | male_OR | male_p | male_n_total | male_n_cases | male_n_controls | female_beta | female_SE | female_OR | female_p | female_n_total | female_n_cases | female_n_controls | interaction_p | interaction_beta | description | group | lab | label |
|---|---|---|---|---|---|---|---|---|---|---|---|---|---|---|---|---|---|---|---|---|
| 596 | -0.1161 | 0.0699 | 0.8903 | 0.09659 | 25451 | 1131 | 24320 | -0.08936 | 0.06694 | 0.9145 | 0.182 | 34046 | 1171 | 32875 | 0.9933 | 7.40e-04 | Other disorders of bladder | genitourinary | LDL-C | |
| 596.1 | | | | | 24500 | 180 | 24320 | | | | | 32903 | 28 | 32875 | | | Bladder neck obstruction | genitourinary | LDL-C | |
| 596.5 | -0.2158 | 0.09291 | 0.8059 | 0.02021 | 24918 | 598 | 24320 | -0.1404 | 0.07453 | 0.869 | 0.05955 | 33798 | 923 | 32875 | 0.2906 | 0.1154 | Functional disorders of bladder | genitourinary | LDL-C | |
| 597 | | | | | 24709 | 389 | 24320 | | | | | 33264 | 389 | 32875 | 1.81e-07 | 0.7968 | Other disorders of urethra and urinary tract | genitourinary | LDL-C | |
| 597.1 | | | | | 24499 | 179 | 24320 | | | | | 32898 | 23 | 32875 | | | Urethral stricture (not specified as infectious) | genitourinary | LDL-C | |
| 597.2 | | | | | 24431 | 111 | 24320 | | | | | 32951 | 76 | 32875 | | | Urinary complications NEC | genitourinary | LDL-C | |
| 597.8 | | | | | 24446 | 126 | 24320 | | | | | 32996 | 121 | 32875 | | | Urethral hypermobility/ISD | genitourinary | LDL-C | |
| 598 | | | | | 27202 | 318 | 26884 | | | | | 36132 | 426 | 35706 | 0.8822 | -0.03104 | Abnormal findings on examination of urine | genitourinary | LDL-C | |
| 598.4 | | | | | 26884 | | | | | | | 35709 | 3 | 35706 | | | Other cells and casts in urine | genitourinary | LDL-C | |
| 599 | 0.9612 | 0.04553 | 2.615 | 6.09e-99 | 23589 | 4209 | 19380 | 0.9965 | 0.03399 | 2.709 | 5.69e-189 | 31705 | 6851 | 24854 | 0.3735 | -0.0478 | Other symptoms/disorders or the urinary system | genitourinary | LDL-C | |
| 599.1 | | | | | 19615 | 235 | 19380 | | | | | 24927 | 73 | 24854 | | | Urinary obstruction | genitourinary | LDL-C | |
| 599.2 | 0.5817 | 0.0616 | 1.789 | 3.60e-21 | 21042 | 1662 | 19380 | 0.5258 | 0.0694 | 1.692 | 3.56e-14 | 25979 | 1125 | 24854 | 0.7021 | -0.0332 | Retention of urine | genitourinary | LDL-C | |
| 599.3 | 1.502 | 0.1154 | 4.492 | 9.95e-39 | 20130 | 750 | 19380 | 1.331 | 0.05117 | 3.787 | 2.78e-149 | 27990 | 3136 | 24854 | 0.05313 | -0.2364 | Dysuria | genitourinary | LDL-C | |
| 599.4 | 0.3283 | 0.08615 | 1.389 | 1.38e-04 | 20122 | 742 | 19380 | 0.8215 | 0.05514 | 2.274 | 3.36e-50 | 27034 | 2180 | 24854 | 3.26e-07 | 0.4932 | Urinary incontinence | genitourinary | LDL-C | |
| 599.5 | 1.811 | 0.08952 | 6.119 | 4.82e-91 | 21290 | 1910 | 19380 | 1.35 | 0.06495 | 3.857 | 6.51e-96 | 26913 | 2059 | 24854 | 4.67e-08 | -0.5895 | Frequency of urination and polyuria | genitourinary | LDL-C | |
| 599.6 | | | | | 19476 | 96 | 19380 | | | | | 24972 | 118 | 24854 | | | Oliguria and anuria | genitourinary | LDL-C | |
| 599.7 | | | | | 19390 | 10 | 19380 | | | | | 24855 | 1 | 24854 | | | Urethral discharge | genitourinary | LDL-C | |
| 599.8 | | | | | 19615 | 235 | 19380 | | | | | 25086 | 232 | 24854 | | | Other symptoms involving urinary system | genitourinary | LDL-C | |
| 599.9 | 1.441 | 0.1168 | 4.224 | 5.68e-35 | 20173 | 793 | 19380 | 0.9984 | 0.07461 | 2.714 | 7.75e-41 | 26091 | 1237 | 24854 | 1.40e-05 | -0.5834 | Other abnormality of urination | genitourinary | LDL-C | |
| 600 | 1.344 | 0.05601 | 3.833 | 3.27e-127 | 23028 | 3640 | 19388 | | | | | 4 | | | 0.994 | -2.717 | Hyperplasia of prostate | genitourinary | LDL-C | |
| 601 | 1.329 | 0.1127 | 3.776 | 4.59e-32 | 20083 | 695 | 19388 | | | | | 4 | | | 0.9962 | -1.796 | Inflammatory diseases of prostate | genitourinary | LDL-C | |
| 601.1 | | | | | 19781 | 393 | 19388 | | | | | 4 | | | | | Prostatitis | genitourinary | LDL-C | |
| 601.11 | | | | | 19525 | 137 | 19388 | | | | | 4 | | | | | Acute prostatitis | genitourinary | LDL-C | |
| 601.12 | | | | | 19502 | 114 | 19388 | | | | | 4 | | | | | Chronic prostatitis | genitourinary | LDL-C | |
| 601.3 | | | | | 19577 | 189 | 19388 | | | | | 4 | | | | | Orchitis and epididymitis | genitourinary | LDL-C | |
| 601.4 | | | | | 19435 | 47 | 19388 | | | | | 4 | | | | | Balanoposthitis | genitourinary | LDL-C | |
| 601.8 | | | | | 19460 | 72 | 19388 | | | | | 4 | | | | | Other inflammatory disorders of male genital organs | genitourinary | LDL-C | |
| 602 | | | | | 19490 | 102 | 19388 | | | | | 4 | | | | | Other disorders of prostate | genitourinary | LDL-C | |
| 602.3 | | | | | 19390 | 2 | 19388 | | | | | 4 | | | | | Dysplasia of prostate | genitourinary | LDL-C | |
| 603 | | | | | 19102 | 243 | 18859 | | | | | 4 | | | | | Other disorders of testis | genitourinary | LDL-C | |
| 603.1 | | | | | 19051 | 192 | 18859 | | | | | 4 | | | | | Hydrocele | genitourinary | LDL-C | |
| 603.2 | | | | | 18915 | 56 | 18859 | | | | | 4 | | | | | Spermatocele | genitourinary | LDL-C | |
| 604 | | | | | 19178 | 319 | 18859 | | | | | 4 | | | | | Disorders of penis | genitourinary | LDL-C | |
| 604.1 | | | | | 18955 | 96 | 18859 | | | | | 4 | | | | | Redundant prepuce and phimosis/BXO | genitourinary | LDL-C | |
| 604.2 | | | | | 18859 | | | | | | | 4 | | | | | Vascular disorders of penis | genitourinary | LDL-C | |
| 604.3 | | | | | 18985 | 126 | 18859 | | | | | 4 | | | | | Peyronie's disease | genitourinary | LDL-C | |
| 605 | 1.175 | 0.06293 | 3.237 | 9.61e-78 | 21005 | 2146 | 18859 | | | | | 4 | | | 0.9963 | -1.735 | Erectile dysfunction [ED] | genitourinary | LDL-C | |
| 608 | 1.075 | 0.1088 | 2.93 | 5.07e-23 | 19398 | 539 | 18859 | | | | | 4 | | | 0.9981 | -1.481 | Other disorders of male genital organs | genitourinary | LDL-C | |

| phenotype | male_beta | male_SE | male_OR | male_p | male_n_total | male_n_cases | male_n_controls | female_beta | female_SE | female_OR | female_p | female_n_total | female_n_cases | female_n_controls | interaction_p | interaction_beta | description | group | lab | label |
|---|---|---|---|---|---|---|---|---|---|---|---|---|---|---|---|---|---|---|---|---|
| 609 | | | | | 18929 | 70 | 18859 | | | | | 4 | | | | | Male infertility and abnormal spermatozoa | genitourinary | LDL-C | |
| 609.1 | | | | | 18884 | 25 | 18859 | | | | | 4 | | | | | Infertility, male | genitourinary | LDL-C | |
| 609.11 | | | | | 18861 | 2 | 18859 | | | | | 4 | | | | | Azoospermia and oligospermia | genitourinary | LDL-C | |
| 609.2 | | | | | 18902 | 43 | 18859 | | | | | 4 | | | | | Abnormal spermatozoa | genitourinary | LDL-C | |
| 610 | | | | | 5 | | | 1.03 | 0.06559 | 2.802 | 1.35e-55 | 29977 | 1611 | 28366 | 0.9967 | 1.489 | Benign mammary dysplasias | genitourinary | LDL-C | |
| 610.1 | | | | | 5 | | | 1.053 | 0.07548 | 2.866 | 3.23e-44 | 29598 | 1232 | 28366 | 0.9966 | 1.556 | Cystic mastopathy | genitourinary | LDL-C | |
| 610.2 | | | | | 5 | | | | | | | 28425 | 59 | 28366 | | | Fibroadenosis of breast | genitourinary | LDL-C | |
| 610.3 | | | | | 5 | | | | | | | 28501 | 135 | 28366 | | | Fibrosclerosis of breast | genitourinary | LDL-C | |
| 610.4 | | | | | 5 | | | | | | | 28633 | 267 | 28366 | | | Benign neoplasm of breast | genitourinary | LDL-C | |
| 610.8 | | | | | 5 | | | | | | | 28599 | 233 | 28366 | | | Other specified benign mammary dysplasias | genitourinary | LDL-C | |
| 611 | | | | | 27275 | 62 | 27213 | 1.215 | 0.04054 | 3.37 | 2.56e-197 | 33455 | 5088 | 28367 | 0.296 | 0.3633 | Abnormal findings on mammogram or breast exam | genitourinary | LDL-C | |
| 611.1 | | | | | 27225 | 12 | 27213 | 1.231 | 0.04501 | 3.423 | 1.38e-164 | 32548 | 4181 | 28367 | 0.5316 | 0.4854 | Abnormal mammogram | genitourinary | LDL-C | |
| 611.11 | | | | | 27213 | | | 0.8756 | 0.111 | 2.4 | 3.15e-15 | 28926 | 559 | 28367 | 0.9954 | 1.932 | Mammographic microcalcification | genitourinary | LDL-C | |
| 611.3 | | | | | 27266 | 53 | 27213 | 1.117 | 0.06392 | 3.057 | 2.03e-68 | 30081 | 1714 | 28367 | 0.7332 | 0.1325 | Lump or mass in breast | genitourinary | LDL-C | |
| 612 | | | | | 27326 | 113 | 27213 | | | | | 28757 | 390 | 28367 | 0.1085 | -0.4686 | Breast conditions, congenital or relating to hormones | genitourinary | LDL-C | |
| 612.1 | | | | | 27213 | | | | | | | 28446 | 79 | 28367 | | | Galactorrhea | genitourinary | LDL-C | |
| 612.2 | | | | | 27325 | 112 | 27213 | | | | | 28664 | 297 | 28367 | | | Hypertrophy of breast (Gynecomastia) | genitourinary | LDL-C | |
| 612.3 | | | | | 27214 | 1 | 27213 | | | | | 28379 | 12 | 28367 | | | Congenital anomalies of breast | genitourinary | LDL-C | |
| 613 | | | | | 27667 | 60 | 27607 | 0.9499 | 0.05505 | 2.585 | 1.01e-66 | 34784 | 2435 | 32349 | 0.07521 | -0.9254 | Other nonmalignant breast conditions | genitourinary | LDL-C | |
| 613.1 | | | | | 27607 | | | | | | | 32616 | 267 | 32349 | | | Inflammatory disease of breast | genitourinary | LDL-C | |
| 613.5 | | | | | 27649 | 42 | 27607 | 1.068 | 0.08623 | 2.909 | 3.27e-35 | 33336 | 987 | 32349 | 0.2168 | -0.7471 | Mastodynia | genitourinary | LDL-C | |
| 613.7 | | | | | 27616 | 9 | 27607 | | | | | 32785 | 436 | 32349 | | | Other signs and symptoms in breast | genitourinary | LDL-C | |
| 613.8 | | | | | 27608 | 1 | 27607 | 0.9868 | 0.1065 | 2.683 | 1.94e-20 | 33034 | 685 | 32349 | 0.9661 | -9.983 | Other specified disorders of breast | genitourinary | LDL-C | |
| 613.9 | | | | | 27609 | 2 | 27607 | | | | | 32422 | 73 | 32349 | | | Breast disorder NOS | genitourinary | LDL-C | |
| 614 | | | | | 7 | | | 0.5559 | 0.05884 | 1.743 | 3.47e-21 | 33657 | 1625 | 32032 | 0.998 | 0.8748 | Inflammatory diseases of female pelvic organs | genitourinary | LDL-C | |
| 614.1 | | | | | 7 | | | | | | | 32176 | 144 | 32032 | | | Pelvic peritoneal adhesions, female (postoperative) (postinfection) | genitourinary | LDL-C | |
| 614.3 | | | | | 7 | | | | | | | 32232 | 200 | 32032 | | | Pelvic inflammatory disease (PID) | genitourinary | LDL-C | |
| 614.31 | | | | | 7 | | | | | | | 32055 | 23 | 32032 | | | Acute inflammatory pelvic disease | genitourinary | LDL-C | |
| 614.32 | | | | | 7 | | | | | | | 32140 | 108 | 32032 | | | Chronic inflammatory pelvic disease | genitourinary | LDL-C | |
| 614.33 | | | | | 7 | | | | | | | 32100 | 68 | 32032 | | | Pelvic inflammatory disease, NOS | genitourinary | LDL-C | |
| 614.4 | | | | | 7 | | | | | | | 32069 | 37 | 32032 | | | Inflammatory diseases of uterus, except cervix | genitourinary | LDL-C | |
| 614.5 | | | | | 7 | | | 0.728 | 0.07052 | 2.071 | 5.49e-25 | 33209 | 1177 | 32032 | 0.9976 | 1.072 | Inflammatory disease of cervix, vagina, and vulva | genitourinary | LDL-C | |
| 614.51 | | | | | 7 | | | | | | | 32101 | 69 | 32032 | | | Cervicitis and endocervicitis | genitourinary | LDL-C | |
| 614.52 | | | | | 7 | | | 0.7575 | 0.0828 | 2.133 | 5.77e-20 | 32877 | 845 | 32032 | 0.9985 | 1.108 | Vaginitis and vulvovaginitis | genitourinary | LDL-C | |
| 614.53 | | | | | 7 | | | | | | | 32093 | 61 | 32032 | | | Cyst or abscess of Bartholin's gland | genitourinary | LDL-C | |

| phenotype | male_beta | male_SE | male_OR | male_p | male_n_total | male_n_cases | male_n_controls | female_beta | female_SE | female_OR | female_p | female_n_total | female_n_cases | female_n_controls | interaction_p | interaction_beta | description | group | lab | label |
|---|---|---|---|---|---|---|---|---|---|---|---|---|---|---|---|---|---|---|---|---|
| 614.54 | | | | | 7 | | | | | | | 32097 | 65 | 32032 | | | Abscess or ulceration of vulva | genitourinary | LDL-C | |
| 615 | | | | | 7 | | | | | | | 32416 | 384 | 32032 | | | Endometriosis | genitourinary | LDL-C | |
| 618 | | | | | 7 | | | 0.09643 | 0.07186 | 1.101 | 0.1796 | 36315 | 1140 | 35175 | 0.9989 | 0.4794 | Genital prolapse | genitourinary | LDL-C | |
| 618.1 | | | | | 7 | | | 0.1145 | 0.08094 | 1.121 | 0.1573 | 36071 | 896 | 35175 | 0.9993 | 0.4979 | Prolapse of vaginal walls | genitourinary | LDL-C | |
| 618.2 | | | | | 7 | | | | | | | 35515 | 340 | 35175 | | | Uterine/Uterovaginal prolapse | genitourinary | LDL-C | |
| 618.5 | | | | | 7 | | | | | | | 35494 | 319 | 35175 | | | Prolapse of vaginal vault after hysterectomy | genitourinary | LDL-C | |
| 618.6 | | | | | 7 | | | | | | | 35376 | 201 | 35175 | | | Vaginal enterocele, congenital or acquired | genitourinary | LDL-C | |
| 619 | | | | | 7 | | | 0.4662 | 0.05172 | 1.594 | 1.98e-19 | 33857 | 2126 | 31731 | 0.9982 | 0.7819 | Noninflammatory female genital disorders | genitourinary | LDL-C | |
| 619.1 | | | | | 7 | | | | | | | 32002 | 271 | 31731 | | | Noninflammatory disorders of ovary, fallopian tube, and broad ligament | genitourinary | LDL-C | |
| 619.2 | | | | | 7 | | | | | | | 32192 | 461 | 31731 | | | Disorders of uterus, NEC | genitourinary | LDL-C | |
| 619.3 | | | | | 7 | | | | | | | 31864 | 133 | 31731 | | | Noninflammatory disorders of cervix | genitourinary | LDL-C | |
| 619.4 | | | | | 7 | | | 0.2655 | 0.09881 | 1.304 | 0.007213 | 32259 | 528 | 31731 | 0.9992 | 0.5932 | Noninflammatory disorders of vagina | genitourinary | LDL-C | |
| 619.5 | | | | | 7 | | | | | | | 31969 | 238 | 31731 | | | Noninflammatory disorders of vulva and perineum | genitourinary | LDL-C | |
| 620 | | | | | 7 | | | | | | | 35251 | 113 | 35138 | | | Dysplasia of female genital organs | genitourinary | LDL-C | |
| 620.1 | | | | | 7 | | | | | | | 35152 | 14 | 35138 | | | Dysplasia of cervix | genitourinary | LDL-C | |
| 621 | | | | | 7 | | | | | | | 35309 | 171 | 35138 | | | Endometrial hyperplasia | genitourinary | LDL-C | |
| 622 | | | | | 7 | | | | | | | 35534 | 396 | 35138 | | | Polyp of female genital organs | genitourinary | LDL-C | |
| 622.1 | | | | | 7 | | | | | | | 35431 | 293 | 35138 | | | Polyp of corpus uteri | genitourinary | LDL-C | |
| 622.2 | | | | | 7 | | | | | | | 35215 | 77 | 35138 | | | Mucous polyp of cervix | genitourinary | LDL-C | |
| 623 | | | | | 7 | | | | | | | 35299 | 161 | 35138 | | | Hypertrophy of female genital organs | genitourinary | LDL-C | |
| 624 | | | | | 7 | | | | | | | 31709 | 227 | 31482 | | | Symptoms involving female genital tract | genitourinary | LDL-C | |
| 624.1 | | | | | 7 | | | | | | | 31539 | 57 | 31482 | | | Dystrophy of female genital tract | genitourinary | LDL-C | |
| 624.2 | | | | | 7 | | | | | | | 31503 | 21 | 31482 | | | Atrophy of female genital tract | genitourinary | LDL-C | |
| 624.9 | | | | | 7 | | | 0.4905 | 0.08004 | 1.633 | 8.94e-10 | 32466 | 984 | 31482 | 0.9988 | 0.8427 | stress incontinence, female | genitourinary | LDL-C | |
| 625 | | | | | 7 | | | 0.418 | 0.0616 | 1.519 | 1.15e-11 | 32899 | 1417 | 31482 | 0.9984 | 0.7169 | Pain and other symptoms associated with female genital organs | genitourinary | LDL-C | |
| 625.1 | | | | | 7 | | | | | | | 31749 | 267 | 31482 | | | Dyspareunia | genitourinary | LDL-C | |
| 626 | | | | | 5 | | | 1.365 | 0.04472 | 3.914 | 1.88e-204 | 27458 | 3874 | 23584 | 0.9958 | 1.916 | Disorders of menstruation and other abnormal bleeding from female genital tract | genitourinary | LDL-C | |
| 626.1 | | | | | 5 | | | 1.47 | 0.04944 | 4.348 | 3.32e-194 | 26685 | 3101 | 23584 | 0.9955 | 2.033 | Irregular menstrual cycle/bleeding | genitourinary | LDL-C | |
| 626.11 | | | | | 5 | | | 1.595 | 0.1 | 4.93 | 2.72e-57 | 24169 | 585 | 23584 | 0.9971 | 2.156 | Absent or infrequent menstruation | genitourinary | LDL-C | |
| 626.12 | | | | | 5 | | | 1.379 | 0.07402 | 3.971 | 1.84e-77 | 24734 | 1150 | 23584 | 0.9958 | 1.904 | Excessive or frequent menstruation | genitourinary | LDL-C | |
| 626.13 | | | | | 5 | | | 1.841 | 0.08084 | 6.302 | 8.96e-115 | 24711 | 1127 | 23584 | 0.9946 | 2.471 | Irregular menstrual cycle | genitourinary | LDL-C | |
| 626.14 | | | | | 5 | | | | | | | 23850 | 266 | 23584 | | | Irregular menstrual bleeding | genitourinary | LDL-C | |
| 626.15 | | | | | 5 | | | | | | | 23629 | 45 | 23584 | | | Infertility, female, associated with anovulation | genitourinary | LDL-C | |
| 626.2 | | | | | 5 | | | | | | | 24029 | 445 | 23584 | | | Dysmenorrhea | genitourinary | LDL-C | |
| phenotype | male_beta | male_SE | male_OR | male_p | male_n_total | male_n_cases | male_n_controls | female_beta | female_SE | female_OR | female_p | female_n_total | female_n_cases | female_n_controls | interaction_p | interaction_beta | description | group | lab | label |
| 626.21 | | | | | 5 | | | | | | | 23591 | 7 | 23584 | | | Mittelschmerz | genitourinary | LDL-C | |

| phenotype | male_beta | male_SE | male_OR | male_p | male_n_total | male_n_cases | male_n_controls | female_beta | female_SE | female_OR | female_p | female_n_total | female_n_cases | female_n_controls | interaction_p | interaction_beta | description | group | lab | label |
|---|---|---|---|---|---|---|---|---|---|---|---|---|---|---|---|---|---|---|---|---|
| 626.4 | | | | | 5 | | | | | | | 23636 | 52 | 23584 | | | Premenstrual tension syndromes | genitourinary | LDL-C | |
| 626.8 | | | | | 5 | | | | | | | 23771 | 187 | 23584 | | | Infertility, female | genitourinary | LDL-C | |
| 627 | | | | | 6 | | | 1.472 | 0.04345 | 4.358 | 1.57e-251 | 28947 | 5363 | 23584 | 0.9515 | -7.26 | Menopausal and postmenopausal disorders | genitourinary | LDL-C | |
| 627.1 | | | | | 5 | | | 1.455 | 0.1018 | 4.284 | 2.36e-46 | 24382 | 798 | 23584 | 0.9974 | 1.953 | Postmenopausal bleeding | genitourinary | LDL-C | |
| 627.2 | | | | | 6 | | | 1.641 | 0.05589 | 5.163 | 1.45e-189 | 26962 | 3378 | 23584 | 0.953 | -7.047 | Symptomatic menopause | genitourinary | LDL-C | |
| 627.21 | | | | | 5 | | | | | | | 23934 | 350 | 23584 | | | Symptomatic artificial menopause | genitourinary | LDL-C | |
| 627.22 | | | | | 6 | | | 1.436 | 0.09847 | 4.202 | 3.86e-48 | 24465 | 881 | 23584 | 0.9666 | -8.24 | Need for Hormone replacement therapy (postmenopausal) | genitourinary | LDL-C | |
| 627.3 | | | | | 5 | | | 1.221 | 0.07802 | 3.39 | 3.38e-55 | 24943 | 1359 | 23584 | 0.9961 | 1.757 | Postmenopausal atrophic vaginitis | genitourinary | LDL-C | |
| 627.4 | | | | | 5 | | | | | | | 23687 | 103 | 23584 | | | Premenopausal menorrhagia | genitourinary | LDL-C | |
| 627.5 | | | | | 5 | | | | | | | 23742 | 158 | 23584 | | | Premature menopause and other ovarian failure | genitourinary | LDL-C | |
| 628 | | | | | 5 | | | 0.9089 | 0.07274 | 2.482 | 7.91e-36 | 24586 | 1002 | 23584 | 0.9971 | 1.302 | Ovarian cyst | genitourinary | LDL-C | |
| 634 | | | | | 7 | | | 0.2652 | 0.09841 | 1.304 | 0.007053 | 34235 | 519 | 33716 | 0.9992 | 0.581 | Miscarriage; stillbirth | pregnancy complications | LDL-C | |
| 634.1 | | | | | 7 | | | | | | | 33945 | 229 | 33716 | | | Missed abortion/Hydatidiform mole | pregnancy complications | LDL-C | |
| 634.3 | | | | | 7 | | | | | | | 33757 | 41 | 33716 | | | Ectopic pregnancy | pregnancy complications | LDL-C | |
| 635 | | | | | 7 | | | | | | | 34113 | 397 | 33716 | | | Hemorrhage during pregnancy; childbirth and postpartum | pregnancy complications | LDL-C | |
| 635.2 | | | | | 7 | | | | | | | 33989 | 273 | 33716 | | | Antepartum hemorrhage, abruptio placentae, and placenta previa | pregnancy complications | LDL-C | |
| 635.3 | | | | | 7 | | | | | | | 33851 | 135 | 33716 | | | Placenta previa and abruptio placenta | pregnancy complications | LDL-C | |
| 636 | | | | | 7 | | | -0.03779 | 0.05876 | 0.9629 | 0.5201 | 35417 | 1701 | 33716 | 0.9993 | 0.3189 | Early or threatened labor; hemorrhage in early pregnancy | pregnancy complications | LDL-C | |
| 636.1 | | | | | 7 | | | | | | | 34209 | 493 | 33716 | | | Threatened premature labor | pregnancy complications | LDL-C | |
| 636.2 | | | | | 7 | | | | | | | 34111 | 395 | 33716 | | | Early onset of delivery | pregnancy complications | LDL-C | |
| 636.3 | | | | | 7 | | | 0.3989 | 0.09873 | 1.49 | 5.33e-05 | 34236 | 520 | 33716 | 0.9989 | 0.7827 | Hemorrhage in early pregnancy | pregnancy complications | LDL-C | |
| 636.8 | | | | | 7 | | | | | | | 33814 | 98 | 33716 | | | Cervical incompetence | pregnancy complications | LDL-C | |
| 637 | | | | | 27759 | 8 | 27751 | | | | | 33709 | 19 | 33690 | | | Short gestation; low birth weight; and fetal growth retardation | pregnancy complications | LDL-C | |
| 642 | | | | | 7 | | | 0.2741 | 0.08252 | 1.315 | 8.95e-04 | 36632 | 759 | 35873 | 0.9992 | 0.5891 | Hypertension complicating pregnancy, childbirth, and the puerperium | pregnancy complications | LDL-C | |
| 642.1 | | | | | 7 | | | | | | | 36229 | 356 | 35873 | | | Preeclampsia and eclampsia | pregnancy complications | LDL-C | |
| 643 | | | | | 7 | | | | | | | 36714 | 156 | 36558 | | | Excessive vomiting in pregnancy | pregnancy complications | LDL-C | |
| 643.1 | | | | | 7 | | | | | | | 36608 | 50 | 36558 | | | Hyperemesis gravidarum | pregnancy complications | LDL-C | |
| 644 | | | | | 7 | | | | | | | 36546 | 186 | 36360 | | | Anemia during pregnancy | pregnancy complications | LDL-C | |
| 646 | | | | | 7 | | | -0.1478 | 0.09767 | 0.8626 | 0.1301 | 36367 | 578 | 35789 | 0.9996 | 0.2912 | Other complications of pregnancy NEC | pregnancy complications | LDL-C | |
| 647 | | | | | 7 | | | | | | | 36080 | 179 | 35901 | | | Infectious and parasitic complications affecting pregnancy | pregnancy complications | LDL-C | |

| phenotype | male_beta | male_SE | male_OR | male_p | male_n_total | male_n_cases | male_n_controls | female_beta | female_SE | female_OR | female_p | female_n_total | female_n_cases | female_n_controls | interaction_p | interaction_beta | description | group | lab | label |
|---|---|---|---|---|---|---|---|---|---|---|---|---|---|---|---|---|---|---|---|---|
| 647.1 | | | | | 7 | | | | | | | 36102 | 201 | 35901 | | | Infections of genitourinary tract during pregnancy | pregnancy complications | LDL-C | |
| 647.3 | | | | | 7 | | | | | | | 35950 | 49 | 35901 | | | Major puerperal infection | pregnancy complications | LDL-C | |
| 649 | | | | | 7 | | | 0.02819 | 0.05907 | 1.029 | 0.6332 | 35652 | 1664 | 33988 | 0.9991 | 0.3723 | Other conditions or status of the mother complicating pregnancy, childbirth, or the puerperium | pregnancy complications | LDL-C | |
| 649.1 | | | | | 7 | | | 0.5635 | 0.09159 | 1.757 | 7.60e-10 | 34593 | 605 | 33988 | 0.9988 | 0.8889 | Diabetes or abnormal glucose tolerance complicating pregnancy | pregnancy complications | LDL-C | |
| 653 | | | | | 7 | | | -0.2651 | 0.09581 | 0.7672 | 0.005666 | 35753 | 642 | 35111 | 1 | -0.01495 | Problems associated with amniotic cavity and membranes | pregnancy complications | LDL-C | |
| 654 | | | | | 7 | | | | | | | 35408 | 297 | 35111 | | | Other and unspecified complications of birth; puerperium affecting management of mother | pregnancy complications | LDL-C | |
| 654.1 | | | | | 7 | | | | | | | 35312 | 201 | 35111 | | | Abnormality of organs and soft tissues of pelvis complicating pregnancy, childbirth, or the puerperium | pregnancy complications | LDL-C | |
| 654.2 | | | | | 7 | | | | | | | 35201 | 90 | 35111 | | | Rhesus isoimmunization in pregnancy | pregnancy complications | LDL-C | |
| 655 | | | | | 7 | | | -0.01551 | 0.05215 | 0.9846 | 0.7662 | 36061 | 2235 | 33826 | 0.9993 | 0.3104 | Known or suspected fetal abnormality affecting management of mother | pregnancy complications | LDL-C | |
| 655.1 | | | | | 7 | | | -0.1023 | 0.08388 | 0.9028 | 0.2226 | 34615 | 789 | 33826 | 0.9998 | 0.1704 | Abnormality in fetal heart rate or rhythm | pregnancy complications | LDL-C | |
| 656 | | | | | 27627 | 68 | 27559 | | | | | 36677 | 120 | 36557 | | | Other perinatal conditions of fetus or newborn | pregnancy complications | LDL-C | |
| 656.1 | | | | | 27562 | 3 | 27559 | | | | | 36558 | 1 | 36557 | | | Isoimmunization of fetus or newborn | pregnancy complications | LDL-C | |
| 656.2 | | | | | 27597 | 38 | 27559 | | | | | 36585 | 28 | 36557 | | | Respiratory conditions of fetus and newborn | pregnancy complications | LDL-C | |
| 656.22 | | | | | 27562 | 3 | 27559 | | | | | 36559 | 2 | 36557 | | | Interstitial emphysema and related conditions of newborn | pregnancy complications | LDL-C | |
| 656.26 | | | | | 27567 | 8 | 27559 | | | | | 36561 | 4 | 36557 | | | Transitory tachypnea or apnea of newborn | pregnancy complications | LDL-C | |
| 656.3 | | | | | 27567 | 8 | 27559 | | | | | 36567 | 10 | 36557 | | | Endocrine and metabolic disturbances of fetus and newborn | pregnancy complications | LDL-C | |
| 656.4 | | | | | 27568 | 9 | 27559 | | | | | 36563 | 6 | 36557 | | | Hemorrhage of fetus or newborn | pregnancy complications | LDL-C | |
| 656.5 | | | | | 27565 | 6 | 27559 | | | | | 36562 | 5 | 36557 | | | Hematological disorders of newborn | pregnancy complications | LDL-C | |
| 656.6 | | | | | 27567 | 8 | 27559 | | | | | 36562 | 5 | 36557 | | | Perinatal disorders of digestive system | pregnancy complications | LDL-C | |
| 656.7 | | | | | 27566 | 7 | 27559 | | | | | 36566 | 9 | 36557 | | | Conditions involving the integument and temperature regulation of fetus and newborn | pregnancy complications | LDL-C | |
| 656.8 | | | | | 27575 | 16 | 27559 | | | | | 36572 | 15 | 36557 | | | Perinatal jaundice | pregnancy complications | LDL-C | |
| 656.9 | | | | | 27559 | | | | | | | 36559 | 2 | 36557 | | | Neonatal bradycardia or tachycardia | pregnancy complications | LDL-C | |
| 657 | | | | | 27745 | 7 | 27738 | | | | | 36919 | 6 | 36913 | | | Infections specific to the perinatal period | pregnancy complications | LDL-C | |
| 669 | | | | | 7 | | | | | | | 36606 | 168 | 36438 | | | Complications of labor and delivery NEC | pregnancy complications | LDL-C | |
| 671 | | | | | 7 | | | | | | | 36547 | 117 | 36430 | | | Venous/cerebrovascular complications embolism in pregnancy and the puerperium | pregnancy complications | LDL-C | |
| 674 | | | | | 7 | | | | | | | 36509 | 79 | 36430 | | | Other complications of the puerperium NEC | pregnancy complications | LDL-C | |

| phenotype | male_beta | male_SE | male_OR | male_p | male_n_total | male_n_cases | male_n_controls | female_beta | female_SE | female_OR | female_p | female_n_total | female_n_cases | female_n_controls | interaction_p | interaction_beta | description | group | lab | label |
|---|---|---|---|---|---|---|---|---|---|---|---|---|---|---|---|---|---|---|---|---|
| 676 | | | | | 7 | | | | | | | 36440 | 10 | 36430 | | | Other disorders of the breast associated with childbirth and disorders of lactation | pregnancy complications | LDL-C | |
| 681 | 0.7605 | 0.04954 | 2.139 | 3.61e-53 | 24732 | 2737 | 21995 | 0.7881 | 0.04584 | 2.199 | 3.05e-66 | 32934 | 3053 | 29881 | 0.2229 | 0.07651 | Superficial cellulitis and abscess | dermatologic | LDL-C | |
| 681.1 | | | | | 22394 | 399 | 21995 | | | | | 30304 | 423 | 29881 | 0.05872 | 0.3906 | Cellulitis and abscess of fingers/toes | dermatologic | LDL-C | |
| phenotype | male_beta | male_SE | male_OR | male_p | male_n_total | male_n_cases | male_n_controls | female_beta | female_SE | female_OR | female_p | female_n_total | female_n_cases | female_n_controls | interaction_p | interaction_beta | description | group | lab | label |
| 681.2 | | | | | 22204 | 209 | 21995 | | | | | 30123 | 242 | 29881 | | | Cellulitis and abscess of face/neck | dermatologic | LDL-C | |
| 681.3 | | | | | 22375 | 380 | 21995 | | | | | 30260 | 379 | 29881 | 0.1133 | -0.2574 | Cellulitis and abscess of arm/hand | dermatologic | LDL-C | |
| 681.5 | 0.977 | 0.08439 | 2.657 | 5.39e-31 | 22978 | 983 | 21995 | 0.8168 | 0.08391 | 2.263 | 2.16e-22 | 30774 | 893 | 29881 | 0.5717 | -0.06305 | Cellulitis and abscess of leg, except foot | dermatologic | LDL-C | |
| 681.6 | | | | | 22233 | 238 | 21995 | | | | | 30029 | 148 | 29881 | | | Cellulitis and abscess of foot, toe | dermatologic | LDL-C | |
| 681.7 | | | | | 22491 | 496 | 21995 | 0.4299 | 0.08973 | 1.537 | 1.66e-06 | 30505 | 624 | 29881 | 0.5498 | 0.07468 | Cellulitis and abscess of trunk | dermatologic | LDL-C | |
| 686 | 0.7717 | 0.09998 | 2.163 | 1.18e-14 | 22609 | 614 | 21995 | 0.8136 | 0.09678 | 2.256 | 4.23e-17 | 30530 | 649 | 29881 | 0.1467 | 0.1879 | Other local infections of skin and subcutaneous tissue | dermatologic | LDL-C | |
| 686.1 | | | | | 22123 | 128 | 21995 | | | | | 30019 | 138 | 29881 | | | Carbuncle and furuncle | dermatologic | LDL-C | |
| 686.2 | | | | | 22049 | 54 | 21995 | | | | | 29945 | 64 | 29881 | | | Impetigo | dermatologic | LDL-C | |
| 686.3 | | | | | 22049 | 54 | 21995 | | | | | 29925 | 44 | 29881 | | | Pilonidal cyst | dermatologic | LDL-C | |
| 686.4 | | | | | 22031 | 36 | 21995 | | | | | 29904 | 23 | 29881 | | | Pyogenic granuloma | dermatologic | LDL-C | |
| 686.5 | | | | | 22021 | 26 | 21995 | | | | | 29916 | 35 | 29881 | | | Pyoderma | dermatologic | LDL-C | |
| 687 | | | | | 21481 | 302 | 21179 | 1.465 | 0.1206 | 4.328 | 5.89e-34 | 26852 | 564 | 26288 | 0.1039 | 0.3183 | Symptoms affecting skin | dermatologic | LDL-C | |
| 687.1 | 0.9546 | 0.06356 | 2.598 | 5.61e-51 | 22861 | 1682 | 21179 | 1.29 | 0.05149 | 3.631 | 2.10e-138 | 29124 | 2836 | 26288 | 1.99e-09 | 0.4599 | Rash and other nonspecific skin eruption | dermatologic | LDL-C | |
| 687.3 | | | | | 21208 | 29 | 21179 | | | | | 26359 | 71 | 26288 | | | Changes in skin texture | dermatologic | LDL-C | |
| 687.4 | 0.7915 | 0.06855 | 2.207 | 7.69e-31 | 22559 | 1380 | 21179 | 0.9666 | 0.05072 | 2.629 | 5.81e-81 | 28776 | 2488 | 26288 | 0.008141 | 0.2121 | Disturbance of skin sensation | dermatologic | LDL-C | |
| 689 | 1.13 | 0.1027 | 3.094 | 3.87e-28 | 26203 | 833 | 25370 | 1.236 | 0.08925 | 3.441 | 1.32e-43 | 34664 | 1062 | 33602 | 0.4696 | 0.09316 | Disorder of skin and subcutaneous tissue NOS | dermatologic | LDL-C | |
| 690 | | | | | 23839 | 493 | 23346 | | | | | 29843 | 385 | 29458 | 0.5137 | -0.1438 | Erythematosquamous dermatosis | dermatologic | LDL-C | |
| 690.1 | | | | | 23835 | 489 | 23346 | | | | | 29839 | 381 | 29458 | 0.3802 | -0.1953 | Seborrheic dermatitis | dermatologic | LDL-C | |
| 691 | | | | | 23418 | 72 | 23346 | | | | | 29595 | 137 | 29458 | | | Congenital anomalies of skin | dermatologic | LDL-C | |
| 691.1 | | | | | 23351 | 5 | 23346 | | | | | 29460 | 2 | 29458 | | | Ichthyosis congenita | dermatologic | LDL-C | |
| 691.3 | | | | | 23350 | 4 | 23346 | | | | | 29482 | 24 | 29458 | | | Congenital pigmentary anomalies of skin | dermatologic | LDL-C | |
| 694 | 1.796 | 0.1372 | 6.025 | 3.66e-39 | 24181 | 835 | 23346 | 1.985 | 0.09966 | 7.28 | 2.74e-88 | 30929 | 1471 | 29458 | 0.5957 | 0.0881 | Dyschromia and Vitiligo | dermatologic | LDL-C | |
| 694.1 | | | | | 23374 | 28 | 23346 | | | | | 29501 | 43 | 29458 | | | Vitiligo | dermatologic | LDL-C | |
| 694.2 | 1.781 | 0.1403 | 5.937 | 6.21e-37 | 24148 | 802 | 23346 | 2.031 | 0.1033 | 7.623 | 4.42e-86 | 30877 | 1419 | 29458 | 0.4564 | 0.1273 | Other dyschromia | dermatologic | LDL-C | |
| 694.3 | | | | | 23359 | 13 | 23346 | | | | | 29477 | 19 | 29458 | | | Vascular disorders of skin | dermatologic | LDL-C | |
| 695 | 0.8579 | 0.09273 | 2.358 | 2.21e-20 | 24180 | 834 | 23346 | 0.6889 | 0.05157 | 1.991 | 1.05e-40 | 31756 | 2298 | 29458 | 0.004855 | -0.281 | Erythematous conditions | dermatologic | LDL-C | |
| 695.1 | | | | | 23358 | 12 | 23346 | | | | | 29480 | 22 | 29458 | | | Toxic erythema | dermatologic | LDL-C | |
| 695.2 | | | | | 23377 | 31 | 23346 | | | | | 29513 | 55 | 29458 | | | Bullous dermatoses | dermatologic | LDL-C | |
| 695.21 | | | | | 23356 | 10 | 23346 | | | | | 29480 | 22 | 29458 | | | Dermatitis herpetiformis | dermatologic | LDL-C | |
| 695.22 | | | | | 23363 | 17 | 23346 | | | | | 29486 | 28 | 29458 | | | Pemphigus and pemphigoid | dermatologic | LDL-C | |
| 695.3 | | | | | 23613 | 267 | 23346 | 1.744 | 0.1322 | 5.722 | 8.91e-40 | 30121 | 663 | 29458 | 0.7988 | -0.06568 | Rosacea | dermatologic | LDL-C | |
| 695.4 | | | | | 23118 | 140 | 22978 | -0.02025 | 0.07293 | 0.9799 | 0.7812 | 29377 | 925 | 28452 | 0.5281 | 0.1154 | Lupus (localized and systemic) | dermatologic | LDL-C | |
| 695.41 | | | | | 23008 | 30 | 22978 | | | | | 28621 | 169 | 28452 | | | Cutaneous lupus erythematosus | dermatologic | LDL-C | |
| 695.42 | | | | | 23104 | 126 | 22978 | -0.05013 | 0.07501 | 0.9511 | 0.5039 | 29322 | 870 | 28452 | 0.6176 | 0.09588 | Systemic lupus erythematosus | dermatologic | LDL-C | |
| 695.7 | | | | | 23445 | 99 | 23346 | | | | | 29666 | 208 | 29458 | | | Prurigo and Lichen | dermatologic | LDL-C | |

| phenotype | male_beta | male_SE | male_OR | male_p | male_n_total | male_n_cases | male_n_controls | female_beta | female_SE | female_OR | female_p | female_n_total | female_n_cases | female_n_controls | interaction_p | interaction_beta | description | group | lab | label |
|---|---|---|---|---|---|---|---|---|---|---|---|---|---|---|---|---|---|---|---|---|
| 695.8 | | | | | 23469 | 123 | 23346 | | | | | 29759 | 301 | 29458 | | | Other specified erythematous conditions | dermatologic | LDL-C | |
| 695.81 | | | | | 23357 | 11 | 23346 | | | | | 29501 | 43 | 29458 | | | Erythema nodosum | dermatologic | LDL-C | |
| 695.9 | | | | | 23478 | 132 | 23346 | | | | | 29596 | 138 | 29458 | | | Unspecified erythematous condition | dermatologic | LDL-C | |
| 696 | | | | | 23091 | 490 | 22601 | 0.6515 | 0.09155 | 1.918 | 1.11e-12 | 28469 | 689 | 27780 | 0.1922 | -0.1832 | Psoriasis and related disorders | dermatologic | LDL-C | |
| 696.2 | | | | | 23352 | 6 | 23346 | | | | | 29465 | 7 | 29458 | | | Parapsoriasis | dermatologic | LDL-C | |
| 696.3 | | | | | 23354 | 8 | 23346 | | | | | 29466 | 8 | 29458 | | | Pityriasis | dermatologic | LDL-C | |
| 696.4 | | | | | 23086 | 485 | 22601 | 0.6211 | 0.09205 | 1.861 | 1.50e-11 | 28453 | 673 | 27780 | 0.1486 | -0.2034 | Psoriasis | dermatologic | LDL-C | |
| 696.41 | | | | | 23001 | 400 | 22601 | 0.8455 | 0.1076 | 2.329 | 3.79e-15 | 28318 | 538 | 27780 | 0.05599 | -0.3248 | Psoriasis vulgaris | dermatologic | LDL-C | |
| 696.42 | | | | | 22782 | 181 | 22601 | | | | | 28051 | 271 | 27780 | | | Psoriatic arthropathy | dermatologic | LDL-C | |
| 697 | | | | | 23461 | 115 | 23346 | | | | | 29654 | 196 | 29458 | | | Sarcoidosis | dermatologic | LDL-C | |
| 698 | | | | | 27077 | 348 | 26729 | 1.001 | 0.09787 | 2.721 | 1.52e-24 | 35552 | 771 | 34781 | 0.1107 | 0.257 | Pruritus and related conditions | dermatologic | LDL-C | |
| 700 | | | | | 24764 | 207 | 24557 | | | | | 32043 | 272 | 31771 | | | Corns and callosities | dermatologic | LDL-C | |
| 701 | 0.6436 | 0.07785 | 1.903 | 1.37e-16 | 25713 | 1156 | 24557 | 0.7399 | 0.05616 | 2.096 | 1.22e-39 | 33879 | 2108 | 31771 | 0.6348 | 0.04302 | Other hypertrophic and atrophic conditions of skin | dermatologic | LDL-C | |
| 701.1 | | | | | 24638 | 81 | 24557 | | | | | 31856 | 85 | 31771 | | | Keratoderma, acquired | dermatologic | LDL-C | |
| 701.2 | | | | | 24996 | 439 | 24557 | 0.3439 | 0.07797 | 1.41 | 1.03e-05 | 32665 | 894 | 31771 | 0.2522 | 0.1449 | Scar conditions and fibrosis of skin | dermatologic | LDL-C | |
| 701.3 | | | | | 24568 | 11 | 24557 | | | | | 31968 | 197 | 31771 | | | Circumscribed scleroderma | dermatologic | LDL-C | |
| 701.4 | | | | | 24632 | 75 | 24557 | | | | | 31928 | 157 | 31771 | | | Keloid scar | dermatologic | LDL-C | |
| 701.5 | | | | | 24606 | 49 | 24557 | | | | | 31836 | 65 | 31771 | | | Abnormal granulation tissue | dermatologic | LDL-C | |
| 701.6 | | | | | 24575 | 18 | 24557 | | | | | 31812 | 41 | 31771 | | | Acquired acanthosis nigricans | dermatologic | LDL-C | |
| 702 | 1.499 | 0.06586 | 4.478 | 1.10e-114 | 24883 | 2887 | 21996 | 1.736 | 0.06535 | 5.675 | 1.83e-155 | 32069 | 3086 | 28983 | 0.006872 | 0.2459 | Degenerative skin conditions and other dermatoses | dermatologic | LDL-C | |
| 702.1 | 1.398 | 0.07503 | 4.049 | 1.58e-77 | 25602 | 2131 | 23471 | 1.597 | 0.09054 | 4.937 | 1.29e-69 | 33264 | 1502 | 31762 | 0.107 | 0.1851 | Actinic keratosis | dermatologic | LDL-C | |
| 702.2 | 1.627 | 0.08318 | 5.089 | 3.27e-85 | 25554 | 2083 | 23471 | 1.73 | 0.07301 | 5.642 | 3.72e-124 | 34321 | 2559 | 31762 | 0.609 | 0.05563 | Seborrheic keratosis | dermatologic | LDL-C | |
| 702.4 | | | | | 21998 | 2 | 21996 | | | | | 28993 | 10 | 28983 | | | Degenerative skin disorders | dermatologic | LDL-C | |
| 703 | | | | | 25804 | 185 | 25619 | | | | | 33096 | 363 | 32733 | 0.5422 | -0.1984 | Diseases of nail, NOS | dermatologic | LDL-C | |
| 703.1 | | | | | 25920 | 301 | 25619 | | | | | 33093 | 360 | 32733 | 0.1026 | 0.5012 | Ingrowing nail | dermatologic | LDL-C | |

| phenotype | male_beta | male_SE | male_OR | male_p | male_n_total | male_n_cases | male_n_controls | female_beta | female_SE | female_OR | female_p | female_n_total | female_n_cases | female_n_controls | interaction_p | interaction_beta | description | group | lab | label |
|---|---|---|---|---|---|---|---|---|---|---|---|---|---|---|---|---|---|---|---|---|
| 704 | 1.53 | 0.1403 | 4.618 | 1.09e-27 | 26130 | 511 | 25619 | 1.459 | 0.09095 | 4.301 | 6.68e-58 | 33832 | 1099 | 32733 | 0.5944 | 0.08495 | Diseases of hair and hair follicles | dermatologic | LDL-C | |
| 704.1 | | | | | 25706 | 87 | 25619 | 1.247 | 0.1077 | 3.479 | 5.17e-31 | 33424 | 691 | 32733 | 0.7916 | 0.08402 | Alopecia | dermatologic | LDL-C | |
| 704.11 | | | | | 25639 | 20 | 25619 | | | | | 32796 | 63 | 32733 | | | Alopecia Areata | dermatologic | LDL-C | |
| 704.12 | | | | | 25622 | 3 | 25619 | | | | | 32824 | 91 | 32733 | | | Telogen effluvium | dermatologic | LDL-C | |
| 704.2 | | | | | 25621 | 2 | 25619 | | | | | 32916 | 183 | 32733 | | | Hirsutism | dermatologic | LDL-C | |
| 704.8 | | | | | 25903 | 284 | 25619 | | | | | 32966 | 233 | 32733 | 0.1561 | 0.416 | Other specified diseases of hair and hair follicles | dermatologic | LDL-C | |
| 705 | | | | | 24635 | 110 | 24525 | | | | | 32184 | 194 | 31990 | | | Disorders of sweat glands | dermatologic | LDL-C | |
| 705.1 | | | | | 24588 | 63 | 24525 | | | | | 32070 | 80 | 31990 | | | Dyshidrosis | dermatologic | LDL-C | |
| 705.3 | | | | | 24563 | 38 | 24525 | | | | | 32095 | 105 | 31990 | | | Hidradenitis | dermatologic | LDL-C | |
| 705.8 | | | | | 24661 | 136 | 24525 | | | | | 32208 | 218 | 31990 | | | Hyperhidrosis | dermatologic | LDL-C | |
| 706 | 1.489 | 0.09105 | 4.431 | 4.42e-60 | 25847 | 1322 | 24525 | 1.671 | 0.06907 | 5.318 | 2.52e-129 | 33934 | 1944 | 31990 | 0.4967 | -0.07447 | Diseases of sebaceous glands | dermatologic | LDL-C | |
| 706.1 | | | | | 24797 | 272 | 24525 | 1.708 | 0.09037 | 5.516 | 1.21e-79 | 32819 | 829 | 31990 | 0.06895 | 0.296 | Acne | dermatologic | LDL-C | |
| 706.2 | 1.86 | 0.1479 | 6.425 | 2.90e-36 | 25246 | 721 | 24525 | 2.014 | 0.1333 | 7.495 | 1.26e-51 | 32825 | 835 | 31990 | 0.5961 | 0.1033 | Sebaceous cyst | dermatologic | LDL-C | |

| phenotype | male_beta | male_SE | male_OR | male_p | male_n_total | male_n_cases | male_n_controls | female_beta | female_SE | female_OR | female_p | female_n_total | female_n_cases | female_n_controls | interaction_p | interaction_beta | description | group | lab | label |
|---|---|---|---|---|---|---|---|---|---|---|---|---|---|---|---|---|---|---|---|---|
| 706.3 | | | | | 24526 | 1 | 24525 | | | | | 31991 | 1 | 31990 | | | Seborrhea | dermatologic | LDL-C | |
| 706.8 | | | | | 24813 | 288 | 24525 | | | | | 32225 | 235 | 31990 | 0.2381 | 0.3944 | Other specified diseases of sebaceous glands | dermatologic | LDL-C | |
| 707 | 0.6512 | 0.07117 | 1.918 | 5.69e-20 | 26784 | 1253 | 25531 | 0.4629 | 0.07636 | 1.589 | 1.35e-09 | 36008 | 999 | 35009 | 0.2486 | -0.1097 | Chronic ulcer of skin | dermatologic | LDL-C | |
| 707.1 | 0.3167 | 0.09646 | 1.373 | 0.001024 | 26112 | 581 | 25531 | | | | | 35488 | 479 | 35009 | 0.813 | 0.03042 | Decubitus ulcer | dermatologic | LDL-C | |
| 707.2 | 1.189 | 0.1093 | 3.284 | 1.46e-27 | 26220 | 689 | 25531 | | | | | 35483 | 474 | 35009 | 0.1807 | -0.2036 | Chronic ulcer of leg or foot | dermatologic | LDL-C | |
| 707.3 | | | | | 25727 | 196 | 25531 | | | | | 35188 | 179 | 35009 | | | Chronic ulcer of unspecified site | dermatologic | LDL-C | |
| 709 | | | | | 23344 | 211 | 23133 | -0.126 | 0.06532 | 0.8817 | 0.05382 | 29862 | 1162 | 28700 | 0.4807 | 0.1069 | Diffuse diseases of connective tissue | dermatologic | LDL-C | |
| 709.2 | | | | | 23183 | 50 | 23133 | | | | | 29154 | 454 | 28700 | 0.1012 | -0.5665 | Sicca syndrome | dermatologic | LDL-C | |
| 709.3 | | | | | 23192 | 59 | 23133 | | | | | 28935 | 235 | 28700 | | | Systemic sclerosis | dermatologic | LDL-C | |
| 709.4 | | | | | 23156 | 23 | 23133 | | | | | 28760 | 60 | 28700 | | | Polymyositis | dermatologic | LDL-C | |
| 709.5 | | | | | 23161 | 28 | 23133 | | | | | 28793 | 93 | 28700 | | | Dermatomyositis | dermatologic | LDL-C | |
| 709.6 | | | | | 23165 | 32 | 23133 | | | | | 28946 | 246 | 28700 | | | Other specified diffuse diseases of connective tissue | dermatologic | LDL-C | |
| 709.7 | | | | | 23162 | 29 | 23133 | | | | | 28907 | 207 | 28700 | | | Unspecified diffuse connective tissue disease | dermatologic | LDL-C | |
| 710 | 0.0323 | 0.07925 | 1.033 | 0.6835 | 23512 | 830 | 22682 | 0.0408 | 0.08633 | 1.042 | 0.6365 | 30097 | 684 | 29413 | 0.4564 | 0.0791 | Osteomyelitis, periostitis, and other infections involving bone | musculoskeletal | LDL-C | |
| 710.1 | -0.02981 | 0.08303 | 0.9706 | 0.7196 | 23427 | 745 | 22682 | -0.1163 | 0.09582 | 0.8902 | 0.2249 | 29958 | 545 | 29413 | 0.6543 | -0.05144 | Osteomyelitis | musculoskeletal | LDL-C | |
| 710.11 | | | | | 22957 | 275 | 22682 | | | | | 29563 | 150 | 29413 | | | Acute osteomyelitis | musculoskeletal | LDL-C | |
| 710.12 | | | | | 23048 | 366 | 22682 | | | | | 29661 | 248 | 29413 | 0.2811 | -0.1793 | Chronic osteomyelitis | musculoskeletal | LDL-C | |
| 710.19 | 0.0974 | 0.1002 | 1.102 | 0.3311 | 23198 | 516 | 22682 | | | | | 29784 | 371 | 29413 | 0.721 | -0.04961 | Unspecified osteomyelitis | musculoskeletal | LDL-C | |
| 710.2 | | | | | 22682 | | | | | | | 29416 | 3 | 29413 | | | Periostitis | musculoskeletal | LDL-C | |
| 710.3 | | | | | 22684 | 2 | 22682 | | | | | 29414 | 1 | 29413 | | | Osteopathy resulting from poliomyelitis | musculoskeletal | LDL-C | |
| 711 | | | | | 22939 | 257 | 22682 | | | | | 29633 | 220 | 29413 | | | Arthropathy associated with infections | musculoskeletal | LDL-C | |
| 711.1 | | | | | 22893 | 211 | 22682 | | | | | 29566 | 153 | 29413 | | | Pyogenic arthritis | musculoskeletal | LDL-C | |
| 711.2 | | | | | 22696 | 14 | 22682 | | | | | 29424 | 11 | 29413 | | | Reiter's disease | musculoskeletal | LDL-C | |
| 711.3 | | | | | 22695 | 13 | 22682 | | | | | 29451 | 38 | 29413 | | | Behcet's syndrome | musculoskeletal | LDL-C | |
| 712 | | | | | 22798 | 116 | 22682 | | | | | 29481 | 68 | 29413 | | | Infective connective tissue disorders | musculoskeletal | LDL-C | |
| 713 | | | | | 22821 | 139 | 22682 | | | | | 29546 | 133 | 29413 | | | Arthropathy associated with other disorders classified elsewhere | musculoskeletal | LDL-C | |
| 713.5 | | | | | 22789 | 107 | 22682 | | | | | 29502 | 89 | 29413 | | | Arthropathy associated with neurological disorders | musculoskeletal | LDL-C | |
| 714 | -0.2161 | 0.08321 | 0.8056 | 0.009395 | 24325 | 732 | 23593 | -0.1351 | 0.05304 | 0.8736 | 0.01086 | 31860 | 1880 | 29980 | 0.1499 | 0.1304 | Rheumatoid arthritis and other inflammatory polyarthropathies | musculoskeletal | LDL-C | |
| 714.1 | -0.2205 | 0.09887 | 0.8021 | 0.02574 | 24107 | 514 | 23593 | -0.1168 | 0.06097 | 0.8897 | 0.05531 | 31387 | 1407 | 29980 | 0.08502 | 0.1835 | Rheumatoid arthritis | musculoskeletal | LDL-C | |
| 714.2 | | | | | 23636 | 43 | 23593 | | | | | 30097 | 117 | 29980 | | | Juvenile rheumatoid arthritis | musculoskeletal | LDL-C | |
| 715 | | | | | 23907 | 314 | 23593 | 0.874 | 0.1083 | 2.396 | 6.84e-16 | 30615 | 635 | 29980 | 0.01889 | 0.4014 | Other inflammatory spondylopathies | musculoskeletal | LDL-C | |
| 715.1 | | | | | 23774 | 181 | 23593 | | | | | 30461 | 481 | 29980 | 0.9391 | 0.01909 | Sacroiliitis NEC | musculoskeletal | LDL-C | |

| phenotype | male_beta | male_SE | male_OR | male_p | male_n_total | male_n_cases | male_n_controls | female_beta | female_SE | female_OR | female_p | female_n_total | female_n_cases | female_n_controls | interaction_p | interaction_beta | description | group | lab | label |
|---|---|---|---|---|---|---|---|---|---|---|---|---|---|---|---|---|---|---|---|---|
| 715.2 | | | | | 23683 | 90 | 23593 | | | | | 30051 | 71 | 29980 | | | Ankylosing spondylitis | musculoskeletal | LDL-C | |
| 715.3 | | | | | 23620 | 27 | 23593 | | | | | 30038 | 58 | 29980 | | | Spinal enthesopathy | musculoskeletal | LDL-C | |
| 716 | 0.1831 | 0.08303 | 1.201 | 0.02747 | 23562 | 880 | 22682 | 0.3992 | 0.06884 | 1.491 | 6.68e-09 | 30774 | 1361 | 29413 | 0.01083 | 0.2546 | Other arthropathies | musculoskeletal | LDL-C | |
| 716.1 | | | | | 22700 | 18 | 22682 | | | | | 29437 | 24 | 29413 | | | Unspecified polyarthropathy or polyarthritis | musculoskeletal | LDL-C | |
| 716.2 | | | | | 22688 | 6 | 22682 | | | | | 29420 | 7 | 29413 | | | Unspecified monoarthritis | musculoskeletal | LDL-C | |
| 716.3 | | | | | 22683 | 1 | 22682 | | | | | 29414 | 1 | 29413 | | | Kaschin-Beck disease | musculoskeletal | LDL-C | |
| 716.8 | | | | | 22688 | 6 | 22682 | | | | | 29426 | 13 | 29413 | | | Palindromic rheumatism | musculoskeletal | LDL-C | |
| 716.9 | 0.2154 | 0.08865 | 1.24 | 0.0151 | 23461 | 779 | 22682 | 0.3864 | 0.07255 | 1.472 | 1.01e-07 | 30638 | 1225 | 29413 | 0.03932 | 0.2186 | Arthropathy NOS | musculoskeletal | LDL-C | |
| 717 | | | | | 27682 | 166 | 27516 | | | | | 36769 | 263 | 36506 | | | Polymyalgia Rheumatica | musculoskeletal | LDL-C | |
| 720 | 0.5107 | 0.06835 | 1.666 | 7.91e-14 | 22346 | 1436 | 20910 | 0.6557 | 0.06263 | 1.926 | 1.20e-25 | 29466 | 1690 | 27776 | 0.07661 | 0.1542 | Spinal stenosis | musculoskeletal | LDL-C | |
| 720.1 | 0.4962 | 0.08367 | 1.642 | 3.02e-09 | 21846 | 936 | 20910 | 0.6618 | 0.0762 | 1.938 | 3.76e-18 | 28903 | 1127 | 27776 | 0.1787 | 0.1428 | Spinal stenosis of lumbar region | musculoskeletal | LDL-C | |
| 721 | 0.6949 | 0.05948 | 2.004 | 1.55e-31 | 23062 | 2152 | 20910 | 0.8699 | 0.05113 | 2.387 | 6.39e-65 | 30738 | 2962 | 27776 | 0.007116 | 0.1997 | Spondylosis and allied disorders | musculoskeletal | LDL-C | |
| 721.1 | 0.7607 | 0.06271 | 2.14 | 7.28e-34 | 22908 | 1998 | 20910 | 0.9221 | 0.05313 | 2.515 | 1.76e-67 | 30580 | 2804 | 27776 | 0.02206 | 0.1787 | Spondylosis without myelopathy | musculoskeletal | LDL-C | |
| 721.2 | | | | | 21155 | 245 | 20910 | | | | | 28026 | 250 | 27776 | | | Spondylosis with myelopathy | musculoskeletal | LDL-C | |
| 721.8 | | | | | 20961 | 51 | 20910 | | | | | 27797 | 21 | 27776 | | | Other allied disorders of spine | musculoskeletal | LDL-C | |
| 722 | 0.7323 | 0.05538 | 2.08 | 6.38e-40 | 23387 | 2477 | 20910 | 0.9077 | 0.04709 | 2.479 | 8.75e-83 | 31200 | 3424 | 27776 | 0.009061 | 0.18 | Intervertebral disc disorders | musculoskeletal | LDL-C | |
| 722.1 | 0.6744 | 0.1053 | 1.963 | 1.51e-10 | 21491 | 581 | 20910 | 1 | 0.09275 | 2.719 | 4.11e-27 | 28557 | 781 | 27776 | 0.0175 | 0.3151 | Displacement of intervertebral disc | musculoskeletal | LDL-C | |
| 722.3 | | | | | 20912 | 2 | 20910 | | | | | 27777 | 1 | 27776 | | | Schmorl's nodes | musculoskeletal | LDL-C | |
| 722.6 | 0.975 | 0.06939 | 2.651 | 7.62e-45 | 22720 | 1810 | 20910 | 1.108 | 0.05638 | 3.027 | 6.29e-86 | 30454 | 2678 | 27776 | 0.1507 | 0.1234 | Degeneration of intervertebral disc | musculoskeletal | LDL-C | |
| 722.7 | | | | | 21018 | 108 | 20910 | | | | | 27881 | 105 | 27776 | | | Intervertebral disc disorder with myelopathy | musculoskeletal | LDL-C | |
| 722.8 | | | | | 21238 | 328 | 20910 | | | | | 28155 | 379 | 27776 | 0.0998 | 0.2655 | Postlaminectomy syndrome | musculoskeletal | LDL-C | |
| 722.9 | | | | | 21012 | 102 | 20910 | | | | | 27883 | 107 | 27776 | | | Other and unspecified disc disorder | musculoskeletal | LDL-C | |
| 723 | | | | | 21252 | 342 | 20910 | | | | | 28186 | 410 | 27776 | 1.33e-05 | 0.6905 | Other disorders of cervical region | musculoskeletal | LDL-C | |
| 723.1 | | | | | 20943 | 33 | 20910 | | | | | 27812 | 36 | 27776 | | | Torticollis | musculoskeletal | LDL-C | |
| 724 | | | | | 21393 | 483 | 20910 | 1.066 | 0.09362 | 2.903 | 4.96e-30 | 28648 | 872 | 27776 | 0.5736 | 0.08656 | Other and unspecified disorders of back | musculoskeletal | LDL-C | |
| 724.1 | | | | | 21038 | 128 | 20910 | | | | | 28127 | 351 | 27776 | | | Disorders of sacrum | musculoskeletal | LDL-C | |
| 724.2 | | | | | 21034 | 124 | 20910 | | | | | 28108 | 332 | 27776 | | | Disorders of coccyx | musculoskeletal | LDL-C | |
| 724.8 | | | | | 21013 | 103 | 20910 | | | | | 27951 | 175 | 27776 | | | Other symptoms referable to back | musculoskeletal | LDL-C | |

| phenotype | male_beta | male_SE | male_OR | male_p | male_n_total | male_n_cases | male_n_controls | female_beta | female_SE | female_OR | female_p | female_n_total | female_n_cases | female_n_controls | interaction_p | interaction_beta | description | group | lab | label |
|---|---|---|---|---|---|---|---|---|---|---|---|---|---|---|---|---|---|---|---|---|
| 724.9 | | | | | 20981 | 71 | 20910 | | | | | 27892 | 116 | 27776 | | | Other unspecified back disorders | musculoskeletal | LDL-C | |
| 726 | 1.398 | 0.06059 | 4.048 | 7.28e-118 | 23098 | 2817 | 20281 | 1.445 | 0.04561 | 4.244 | 2.31e-220 | 30370 | 4595 | 25775 | 0.2252 | 0.08967 | Peripheral enthesopathies and allied syndromes | musculoskeletal | LDL-C | |
| 726.1 | 1.597 | 0.1002 | 4.937 | 3.72e-57 | 21421 | 1140 | 20281 | 1.442 | 0.05783 | 4.23 | 2.74e-137 | 28550 | 2775 | 25775 | 0.2642 | -0.1265 | Enthesopathy | musculoskeletal | LDL-C | |
| 726.2 | | | | | 20365 | 84 | 20281 | | | | | 25894 | 119 | 25775 | | | Synoviopathy | musculoskeletal | LDL-C | |
| 726.3 | | | | | 20728 | 447 | 20281 | | | | | 26194 | 419 | 25775 | 0.2919 | 0.2494 | Bursitis | musculoskeletal | LDL-C | |
| 726.4 | | | | | 20361 | 80 | 20281 | | | | | 25949 | 174 | 25775 | | | Calcaneal spur; Exostosis NOS | musculoskeletal | LDL-C | |
| 727 | 1.063 | 0.06932 | 2.895 | 4.68e-53 | 21945 | 1664 | 20281 | 1.336 | 0.05477 | 3.803 | 2.09e-131 | 28564 | 2789 | 25775 | 2.11e-04 | 0.3158 | Other disorders of synovium, tendon, and bursa | musculoskeletal | LDL-C | |
| 727.1 | 1.281 | 0.1097 | 3.601 | 1.61e-31 | 21009 | 728 | 20281 | 1.417 | 0.07557 | 4.123 | 2.05e-78 | 27213 | 1438 | 25775 | 0.2658 | 0.1433 | Synovitis and tenosynovitis | musculoskeletal | LDL-C | |
| 727.2 | | | | | 20360 | 79 | 20281 | | | | | 25927 | 152 | 25775 | | | Bursitis disorders | musculoskeletal | LDL-C | |
| 727.4 | | | | | 20471 | 190 | 20281 | | | | | 26261 | 486 | 25775 | 0.09272 | 0.3866 | Ganglion and cyst of synovium, tendon, and bursa | musculoskeletal | LDL-C | |
| 727.5 | | | | | 20324 | 43 | 20281 | | | | | 25904 | 129 | 25775 | | | Rupture of synovium | musculoskeletal | LDL-C | |
| 727.6 | | | | | 20660 | 379 | 20281 | | | | | 26104 | 329 | 25775 | 0.01176 | 0.4875 | Rupture of tendon, nontraumatic | musculoskeletal | LDL-C | |
| 727.7 | | | | | 20347 | 66 | 20281 | | | | | 25864 | 89 | 25775 | | | Contracture of tendon (sheath) | musculoskeletal | LDL-C | |
| 727.8 | | | | | 20310 | 29 | 20281 | | | | | 25829 | 54 | 25775 | | | Plica syndrome | musculoskeletal | LDL-C | |
| 728 | | | | | 20364 | 83 | 20281 | | | | | 25899 | 124 | 25775 | | | Disorders of muscle, ligament, and fascia | musculoskeletal | LDL-C | |
| 728.1 | | | | | 20313 | 32 | 20281 | | | | | 25798 | 23 | 25775 | | | Muscular calcification and ossification | musculoskeletal | LDL-C | |
| 728.2 | | | | | 20304 | 23 | 20281 | | | | | 25866 | 91 | 25775 | | | Laxity of ligament or hypermobility syndrome | musculoskeletal | LDL-C | |
| 728.7 | | | | | 20721 | 440 | 20281 | 2.231 | 0.1394 | 9.309 | 1.26e-57 | 26509 | 734 | 25775 | 0.007205 | 0.5609 | Fasciitis | musculoskeletal | LDL-C | |
| 728.71 | | | | | 20411 | 130 | 20281 | | | | | 25848 | 73 | 25775 | | | Contracture of palmar fascia [Dupuytren's disease] | musculoskeletal | LDL-C | |
| 729 | 1.13 | 0.1051 | 3.095 | 5.79e-27 | 20964 | 683 | 20281 | 1.31 | 0.08868 | 3.706 | 2.21e-49 | 26711 | 936 | 25775 | 0.08033 | 0.232 | Other disorders of soft tissues | musculoskeletal | LDL-C | |
| 729.1 | | | | | 20284 | 3 | 20281 | | | | | 25782 | 7 | 25775 | | | Rheumatism, unspecified and fibrositis | musculoskeletal | LDL-C | |
| 729.3 | | | | | 20292 | 11 | 20281 | | | | | 25825 | 50 | 25775 | | | Panniculitis | musculoskeletal | LDL-C | |
| 729.7 | | | | | 20307 | 26 | 20281 | | | | | 25785 | 10 | 25775 | | | Nontraumatic compartment syndrome | musculoskeletal | LDL-C | |
| 731 | | | | | 22403 | 94 | 22309 | | | | | 27658 | 148 | 27510 | | | Osteitis deformans and osteopathies associated with other disorders classified elsewhere | musculoskeletal | LDL-C | |
| 731.1 | | | | | 22320 | 11 | 22309 | | | | | 27517 | 7 | 27510 | | | Osteitis deformans [Paget's disease of bone] | musculoskeletal | LDL-C | |
| 732 | | | | | 22384 | 75 | 22309 | | | | | 27583 | 73 | 27510 | | | Osteochondropathies | musculoskeletal | LDL-C | |
| 732.1 | | | | | 22341 | 32 | 22309 | | | | | 27549 | 39 | 27510 | | | Juvenile osteochondrosis | musculoskeletal | LDL-C | |
| 732.7 | | | | | 22348 | 39 | 22309 | | | | | 27539 | 29 | 27510 | | | Osteochondritis dissecans | musculoskeletal | LDL-C | |
| 733 | 0.8101 | 0.1081 | 2.248 | 6.85e-14 | 22918 | 609 | 22309 | 1.389 | 0.06718 | 4.011 | 5.54e-95 | 29872 | 2362 | 27510 | 4.63e-15 | 0.9612 | Other disorders of bone and cartilage | musculoskeletal | LDL-C | |

| phenotype | male_beta | male_SE | male_OR | male_p | male_n_total | male_n_cases | male_n_controls | female_beta | female_SE | female_OR | female_p | female_n_total | female_n_cases | female_n_controls | interaction_p | interaction_beta | description | group | lab | label |
|---|---|---|---|---|---|---|---|---|---|---|---|---|---|---|---|---|---|---|---|---|
| 733.2 | | | | | 22336 | 27 | 22309 | | | | | 27554 | 44 | 27510 | | | Cyst of bone | musculoskeletal | LDL-C | |
| 733.4 | | | | | 22516 | 207 | 22309 | | | | | 27762 | 252 | 27510 | | | Aseptic necrosis of bone | musculoskeletal | LDL-C | |
| 733.6 | | | | | 22334 | 25 | 22309 | | | | | 27574 | 64 | 27510 | | | Costochondritis | musculoskeletal | LDL-C | |
| 733.8 | | | | | 22558 | 249 | 22309 | | | | | 27858 | 348 | 27510 | 3.62e-06 | 0.7846 | Malunion and nonunion of fracture | musculoskeletal | LDL-C | |
| 733.9 | | | | | 22350 | 41 | 22309 | | | | | 27565 | 55 | 27510 | | | Chondromalacia | musculoskeletal | LDL-C | |
| 735 | 1.057 | 0.1033 | 2.878 | 1.43e-24 | 24919 | 827 | 24092 | 1.164 | 0.07879 | 3.203 | 2.13e-49 | 32235 | 1405 | 30830 | 0.1381 | 0.184 | Acquired foot deformities | musculoskeletal | LDL-C | |
| 735.1 | | | | | 24190 | 98 | 24092 | | | | | 30970 | 140 | 30830 | | | Flat foot | musculoskeletal | LDL-C | |
| 735.2 | | | | | 24498 | 406 | 24092 | 1.254 | 0.1164 | 3.504 | 4.75e-27 | 31534 | 704 | 30830 | 0.346 | -0.1914 | Acquired toe deformities | musculoskeletal | LDL-C | |
| 735.21 | | | | | 24374 | 282 | 24092 | 1.398 | 0.1423 | 4.048 | 8.58e-23 | 31365 | 535 | 30830 | 0.1518 | -0.3945 | Hammer toe (acquired) | musculoskeletal | LDL-C | |
| 735.22 | | | | | 24107 | 15 | 24092 | | | | | 30844 | 14 | 30830 | | | Claw toe (acquired) | musculoskeletal | LDL-C | |
| 735.23 | | | | | 24184 | 92 | 24092 | | | | | 30968 | 138 | 30830 | | | Hallux rigidus | musculoskeletal | LDL-C | |
| 735.3 | | | | | 24207 | 115 | 24092 | 1.466 | 0.1448 | 4.333 | 4.35e-24 | 31337 | 507 | 30830 | 0.5798 | 0.1807 | Hallux valgus (Bunion) | musculoskeletal | LDL-C | |
| 736 | | | | | 24293 | 201 | 24092 | | | | | 31078 | 248 | 30830 | | | Other acquired deformities of limbs | musculoskeletal | LDL-C | |
| 736.1 | | | | | 24103 | 11 | 24092 | | | | | 30840 | 10 | 30830 | | | Acquired deformities of forearm | musculoskeletal | LDL-C | |
| 736.2 | | | | | 24158 | 66 | 24092 | | | | | 30902 | 72 | 30830 | | | Acquired deformities of finger | musculoskeletal | LDL-C | |
| 736.3 | | | | | 24100 | 8 | 24092 | | | | | 30841 | 11 | 30830 | | | Acquired deformities of hip | musculoskeletal | LDL-C | |
| 736.4 | | | | | 24104 | 12 | 24092 | | | | | 30857 | 27 | 30830 | | | Genu valgum or varum (acquired) | musculoskeletal | LDL-C | |
| 736.5 | | | | | 24095 | 3 | 24092 | | | | | 30840 | 10 | 30830 | | | Acquired deformities of knee | musculoskeletal | LDL-C | |
| phenotype | male_beta | male_SE | male_OR | male_p | male_n_total | male_n_cases | male_n_controls | female_beta | female_SE | female_OR | female_p | female_n_total | female_n_cases | female_n_controls | interaction_p | interaction_beta | description | group | lab | label |
| 736.6 | | | | | 24114 | 22 | 24092 | | | | | 30876 | 46 | 30830 | | | Unequal leg length (acquired) | musculoskeletal | LDL-C | |
| 737 | | | | | 24514 | 422 | 24092 | -0.052 | 0.07341 | 0.9493 | 0.4787 | 31760 | 930 | 30830 | 0.008352 | 0.3182 | Curvature of spine | musculoskeletal | LDL-C | |
| 737.1 | | | | | 24226 | 134 | 24092 | | | | | 31005 | 175 | 30830 | | | Kyphosis (acquired) | musculoskeletal | LDL-C | |
| 737.2 | | | | | 24093 | 1 | 24092 | | | | | 30832 | 2 | 30830 | | | Lordosis (acquired) | musculoskeletal | LDL-C | |
| 737.3 | | | | | 24413 | 321 | 24092 | -0.08808 | 0.07816 | 0.9157 | 0.2598 | 31641 | 811 | 30830 | 3.43e-04 | 0.4815 | Kyphoscoliosis and scoliosis | musculoskeletal | LDL-C | |
| 738 | 0.2573 | 0.1031 | 1.293 | 0.01258 | 24637 | 545 | 24092 | 0.5767 | 0.08066 | 1.78 | 8.69e-13 | 31798 | 968 | 30830 | 0.001894 | 0.3792 | Other acquired musculoskeletal deformity | musculoskeletal | LDL-C | |
| 738.4 | | | | | 24467 | 375 | 24092 | 0.7452 | 0.0952 | 2.107 | 4.98e-15 | 31608 | 778 | 30830 | 0.1422 | 0.2293 | Acquired spondylolisthesis | musculoskeletal | LDL-C | |
| 739 | | | | | 24307 | 215 | 24092 | | | | | 31047 | 217 | 30830 | | | Contracture of joint | musculoskeletal | LDL-C | |
| 740 | 0.8219 | 0.04367 | 2.275 | 5.20e-79 | 25263 | 4641 | 20622 | 0.8325 | 0.03524 | 2.299 | 2.36e-123 | 33672 | 7527 | 26145 | 0.7877 | 0.01442 | Osteoarthrosis | musculoskeletal | LDL-C | |
| 740.1 | 0.9278 | 0.05492 | 2.529 | 5.08e-64 | 23687 | 3065 | 20622 | 1.144 | 0.04602 | 3.14 | 1.80e-136 | 30839 | 4694 | 26145 | 0.002639 | 0.2088 | Osteoarthritis; localized | musculoskeletal | LDL-C | |

| phenotype | male_beta | male_SE | male_OR | male_p | male_n_total | male_n_cases | male_n_controls | female_beta | female_SE | female_OR | female_p | female_n_total | female_n_cases | female_n_controls | interaction_p | interaction_beta | description | group | lab | label |
|---|---|---|---|---|---|---|---|---|---|---|---|---|---|---|---|---|---|---|---|---|
| 740.11 | 1.027 | 0.05895 | 2.793 | 5.20e-68 | 23439 | 2817 | 20622 | 1.155 | 0.04792 | 3.173 | 2.91e-128 | 30486 | 4341 | 26145 | 0.1275 | 0.1126 | Osteoarthrosis, localized, primary | musculoskeletal | LDL-C | |
| 740.12 | | | | | 20717 | 95 | 20622 | | | | | 26285 | 140 | 26145 | | | Osteoarthrosis, localized, secondary | musculoskeletal | LDL-C | |
| 740.2 | 0.6286 | 0.1032 | 1.875 | 1.11e-09 | 21264 | 642 | 20622 | 0.6142 | 0.05979 | 1.848 | 9.41e-25 | 28076 | 1931 | 26145 | 0.5819 | -0.06278 | Osteoarthrosis, generalized | musculoskeletal | LDL-C | |
| 740.3 | | | | | 20669 | 47 | 20622 | | | | | 26282 | 137 | 26145 | | | Osteoarthrosis involving more than one site, but not specified as generalized | musculoskeletal | LDL-C | |
| 740.9 | 0.91 | 0.05828 | 2.484 | 5.87e-55 | 23067 | 2445 | 20622 | 0.9306 | 0.04583 | 2.536 | 1.12e-91 | 30322 | 4177 | 26145 | 0.434 | 0.05536 | Osteoarthrosis NOS | musculoskeletal | LDL-C | |
| 741 | 0.7899 | 0.05616 | 2.203 | 6.24e-45 | 24701 | 2405 | 22296 | 1.017 | 0.04673 | 2.765 | 5.55e-105 | 32920 | 3637 | 29283 | 5.64e-05 | 0.28 | Symptoms and disorders of the joints | musculoskeletal | LDL-C | |
| 741.1 | | | | | 22321 | 25 | 22296 | | | | | 29334 | 51 | 29283 | | | Ankylosis of joint | musculoskeletal | LDL-C | |
| 741.2 | 0.8826 | 0.111 | 2.417 | 1.88e-15 | 22901 | 605 | 22296 | 1.242 | 0.08214 | 3.462 | 1.19e-51 | 30510 | 1227 | 29283 | 0.005343 | 0.3687 | Stiffness of joint | musculoskeletal | LDL-C | |
| 741.3 | 1.062 | 0.1161 | 2.893 | 5.72e-20 | 22942 | 646 | 22296 | 1.39 | 0.1035 | 4.014 | 4.29e-41 | 30187 | 904 | 29283 | 0.02761 | 0.3293 | Difficulty in walking | musculoskeletal | LDL-C | |
| 741.4 | 0.8283 | 0.103 | 2.289 | 8.93e-16 | 22964 | 668 | 22296 | 1.019 | 0.08658 | 2.77 | 5.71e-32 | 30241 | 958 | 29283 | 0.03319 | 0.2724 | Joint effusions | musculoskeletal | LDL-C | |
| 741.5 | | | | | 22328 | 32 | 22296 | | | | | 29301 | 18 | 29283 | | | Hemarthrosis | musculoskeletal | LDL-C | |
| 741.6 | | | | | 22307 | 11 | 22296 | | | | | 29300 | 17 | 29283 | | | Villonodular synovitis | musculoskeletal | LDL-C | |
| 742 | | | | | 22735 | 439 | 22296 | 0.8503 | 0.09777 | 2.34 | 3.42e-18 | 29934 | 651 | 29283 | 9.87e-04 | 0.4665 | Derangement of joint, non-traumatic | musculoskeletal | LDL-C | |
| 742.1 | | | | | 22319 | 23 | 22296 | | | | | 29301 | 18 | 29283 | | | Loose body in joint | musculoskeletal | LDL-C | |
| 742.2 | | | | | 22327 | 31 | 22296 | | | | | 29340 | 57 | 29283 | | | Pathological, developmental or recurrent dislocation | musculoskeletal | LDL-C | |
| 742.8 | | | | | 22322 | 26 | 22296 | | | | | 29329 | 46 | 29283 | | | Articular cartilage disorder | musculoskeletal | LDL-C | |
| 742.9 | | | | | 22643 | 347 | 22296 | 1.009 | 0.1135 | 2.742 | 6.19e-19 | 29808 | 525 | 29283 | 4.83e-04 | 0.5684 | Other derangement of joint | musculoskeletal | LDL-C | |
| 743 | 0.5093 | 0.05687 | 1.664 | 3.39e-19 | 26032 | 1946 | 24086 | 0.939 | 0.03793 | 2.557 | 2.78e-135 | 33952 | 6028 | 27924 | 6.14e-31 | 0.7268 | Osteoporosis, osteopenia and pathological fracture | musculoskeletal | LDL-C | Osteoporosis, osteopenia and pathologic al fracture |
| 743.1 | 0.8286 | 0.08481 | 2.29 | 1.52e-22 | 25110 | 1024 | 24086 | 0.9764 | 0.0463 | 2.655 | 1.01e-98 | 31886 | 3962 | 27924 | 1.73e-04 | 0.3424 | Osteoporosis | musculoskeletal | LDL-C | |
| 743.11 | 0.7609 | 0.09821 | 2.14 | 9.36e-15 | 24850 | 764 | 24086 | 1.142 | 0.05275 | 3.133 | 6.06e-104 | 31204 | 3280 | 27924 | 1.63e-07 | 0.5512 | Osteoporosis NOS | musculoskeletal | LDL-C | |
| 743.12 | | | | | 24196 | 110 | 24086 | 0.6854 | 0.08808 | 1.985 | 7.15e-15 | 28758 | 834 | 27924 | 0.886 | -0.03898 | Senile osteoporosis | musculoskeletal | LDL-C | |
| 743.13 | | | | | 24546 | 460 | 24086 | 1.092 | 0.08357 | 2.98 | 5.03e-39 | 29013 | 1089 | 27924 | 0.05321 | 0.2909 | Other specified osteoporosis | musculoskeletal | LDL-C | |
| 743.2 | | | | | 24445 | 359 | 24086 | 0.5163 | 0.09229 | 1.676 | 2.21e-08 | 28551 | 627 | 27924 | 2.31e-13 | 0.9951 | Pathologic fracture | musculoskeletal | LDL-C | |
| 743.21 | | | | | 24187 | 101 | 24086 | | | | | 28089 | 165 | 27924 | | | Pathologic fracture of vertebrae | musculoskeletal | LDL-C | |
| 743.22 | | | | | 24160 | 74 | 24086 | | | | | 28032 | 108 | 27924 | | | Pathologic fracture of femur | musculoskeletal | LDL-C | |
| 743.4 | | | | | 24126 | 40 | 24086 | | | | | 28053 | 129 | 27924 | | | Stress fracture | musculoskeletal | LDL-C | |

| phenotype | male_beta | male_SE | male_OR | male_p | male_n_total | male_n_cases | male_n_controls | female_beta | female_SE | female_OR | female_p | female_n_total | female_n_cases | female_n_controls | interaction_p | interaction_beta | description | group | lab | label |
|---|---|---|---|---|---|---|---|---|---|---|---|---|---|---|---|---|---|---|---|---|
| 743.9 | 0.4401 | 0.08959 | 1.553 | 9.01e-07 | 24819 | 733 | 24086 | 1.386 | 0.06176 | 3.999 | 1.49e-111 | 30516 | 2592 | 27924 | 1.78e-42 | 1.363 | Osteopenia or other disorder of bone and cartilage | musculoskeletal | LDL-C | |
| 745 | 1.087 | 0.03543 | 2.967 | 6.71e-207 | 24606 | 7378 | 17228 | 1.162 | 0.02852 | 3.198 | 0 | 32439 | 12064 | 20375 | 1.26e-04 | 0.1665 | Pain in joint | musculoskeletal | LDL-C | |
| 747 | 0.5364 | 0.06889 | 1.71 | 6.90e-15 | 26877 | 1323 | 25554 | 0.2923 | 0.06761 | 1.34 | 1.54e-05 | 36150 | 1183 | 34967 | 2.12e-05 | -0.3741 | Cardiac and circulatory congenital anomalies | congenital anomalies | LDL-C | |
| 747.1 | 0.6027 | 0.07305 | 1.827 | 1.58e-16 | 26767 | 1213 | 25554 | 0.3536 | 0.0725 | 1.424 | 1.08e-06 | 36008 | 1041 | 34967 | 2.25e-05 | -0.3981 | Cardiac congenital anomalies | congenital anomalies | LDL-C | |
| 747.11 | | | | | 25997 | 443 | 25554 | 0.2187 | 0.09517 | 1.244 | 0.02154 | 35539 | 572 | 34967 | 0.6581 | -0.05783 | Cardiac shunt/ heart septal defect | congenital anomalies | LDL-C | |
| 747.12 | | | | | 25876 | 322 | 25554 | | | | | 35219 | 252 | 34967 | 0.1262 | -0.2642 | Valvular heart disease/ heart chambers | congenital anomalies | LDL-C | |
| 747.13 | | | | | 25697 | 143 | 25554 | | | | | 35131 | 164 | 34967 | | | Congenital anomalies of great vessels | congenital anomalies | LDL-C | |
| 747.2 | | | | | 25638 | 84 | 25554 | | | | | 35084 | 117 | 34967 | | | Congenital anomalies of peripheral vascular system | congenital anomalies | LDL-C | |
| 748 | | | | | 27528 | 39 | 27489 | | | | | 36621 | 44 | 36577 | | | Anomalies of respiratory system, congenital | congenital anomalies | LDL-C | |
| 749 | | | | | 27561 | 72 | 27489 | | | | | 36671 | 94 | 36577 | | | Congenital anomalies of face and neck | congenital anomalies | LDL-C | |
| 749.1 | | | | | 22492 | 16 | 22476 | | | | | 31338 | 20 | 31318 | | | Cleft palate | congenital anomalies | LDL-C | |
| 749.2 | | | | | 27520 | 31 | 27489 | | | | | 36613 | 36 | 36577 | | | Congenital anomalies of skull and face bones | congenital anomalies | LDL-C | |
| 750 | | | | | 26052 | 143 | 25909 | | | | | 35186 | 190 | 34996 | | | Digestive congenital anomalies | congenital anomalies | LDL-C | |
| 750.1 | | | | | 25951 | 42 | 25909 | | | | | 35050 | 54 | 34996 | | | Upper gastrointestinal congenital anomalies | congenital anomalies | LDL-C | |
| 750.11 | | | | | 25920 | 11 | 25909 | | | | | 35005 | 9 | 34996 | | | Esophageal atresia/tracheoesophageal fistula | congenital anomalies | LDL-C | |
| 750.13 | | | | | 25928 | 19 | 25909 | | | | | 35009 | 13 | 34996 | | | Congenital anomalies of mouth/tongue | congenital anomalies | LDL-C | |
| 750.14 | | | | | 25914 | 5 | 25909 | | | | | 35012 | 16 | 34996 | | | Congenital anomalies of esophagus | congenital anomalies | LDL-C | |
| 750.15 | | | | | 25911 | 2 | 25909 | | | | | 34997 | 1 | 34996 | | | Congenital anomalies of stomach | congenital anomalies | LDL-C | |
| 750.2 | | | | | 26004 | 95 | 25909 | | | | | 35124 | 128 | 34996 | | | Lower gastrointestinal congenital anomalies | congenital anomalies | LDL-C | |
| 750.21 | | | | | 25974 | 65 | 25909 | | | | | 35061 | 65 | 34996 | | | Congenital anomalies of intestine | congenital anomalies | LDL-C | |
| 750.22 | | | | | 25936 | 27 | 25909 | | | | | 35055 | 59 | 34996 | | | Congenital anomaly of gallbladder, bile ducts, liver, pancreas | congenital anomalies | LDL-C | |
| 750.5 | | | | | 25913 | 4 | 25909 | | | | | 34999 | 3 | 34996 | | | Congenital hypertrophic pyloric stenosis | congenital anomalies | LDL-C | |
| 751 | | | | | 26398 | 489 | 25909 | | | | | 35443 | 447 | 34996 | 0.5351 | -0.08529 | Genitourinary congenital anomalies | congenital anomalies | LDL-C | |
| 751.1 | | | | | 26010 | 101 | 25909 | | | | | 35087 | 91 | 34996 | | | Congenital anomalies of genital organs | congenital anomalies | LDL-C | |

| phenotype | male_beta | male_SE | male_OR | male_p | male_n_total | male_n_cases | male_n_controls | female_beta | female_SE | female_OR | female_p | female_n_total | female_n_cases | female_n_controls | interaction_p | interaction_beta | description | group | lab | label |
|---|---|---|---|---|---|---|---|---|---|---|---|---|---|---|---|---|---|---|---|---|
| 751.11 | | | | | 7 | | | | | | | 35071 | 79 | 34992 | | | Congenital anomalies of female genital organs | congenital anomalies | LDL-C | |
| 751.12 | | | | | 25992 | 90 | 25902 | | | | | 4 | | | | | Congenital anomalies of male genital organs | congenital anomalies | LDL-C | |
| 751.2 | | | | | 26267 | 358 | 25909 | | | | | 35304 | 308 | 34996 | 0.6497 | -0.07724 | Congenital anomalies of urinary system | congenital anomalies | LDL-C | |
| 751.21 | | | | | 26140 | 231 | 25909 | | | | | 35199 | 203 | 34996 | | | Cystic kidney disease | congenital anomalies | LDL-C | |

| phenotype | male_beta | male_SE | male_OR | male_p | male_n_total | male_n_cases | male_n_controls | female_beta | female_SE | female_OR | female_p | female_n_total | female_n_cases | female_n_controls | interaction_p | interaction_beta | description | group | lab | label |
|---|---|---|---|---|---|---|---|---|---|---|---|---|---|---|---|---|---|---|---|---|
| 751.22 | | | | | 26007 | 98 | 25909 | | | | | 35074 | 78 | 34996 | | | Other specified congenital anomalies of kidney | congenital anomalies | LDL-C | |
| 751.3 | | | | | 25949 | 40 | 25909 | | | | | 35047 | 51 | 34996 | | | Obstructive genitourinary defect | congenital anomalies | LDL-C | |
| 752 | | | | | 27335 | 160 | 27175 | | | | | 36299 | 223 | 36076 | | | Nervous system congenital anomalies | congenital anomalies | LDL-C | |
| 752.1 | | | | | 27234 | 59 | 27175 | | | | | 36188 | 112 | 36076 | | | Neural tube defects | congenital anomalies | LDL-C | |
| 752.11 | | | | | 27225 | 50 | 27175 | | | | | 36167 | 91 | 36076 | | | Spina bifida | congenital anomalies | LDL-C | |
| 752.2 | | | | | 27279 | 104 | 27175 | | | | | 36200 | 124 | 36076 | | | Other specified congenital anomalies of nervous system | congenital anomalies | LDL-C | |
| 753 | | | | | 27198 | 23 | 27175 | | | | | 36127 | 51 | 36076 | | | Congenital anomalies of the eye | congenital anomalies | LDL-C | |
| 753.1 | | | | | 27183 | 8 | 27175 | | | | | 36087 | 11 | 36076 | | | Congenital cataract and lens anomalies | congenital anomalies | LDL-C | |
| 753.2 | | | | | 27181 | 6 | 27175 | | | | | 36095 | 19 | 36076 | | | Congenital anomalies of posterior segment of eye | congenital anomalies | LDL-C | |
| 754 | | | | | 27083 | 58 | 27025 | | | | | 35886 | 111 | 35775 | | | Congenital musculoskeletal deformities of spine | congenital anomalies | LDL-C | |
| 754.1 | | | | | 27033 | 8 | 27025 | | | | | 35793 | 18 | 35775 | | | Lumbosacral spondylolysis, congenital | congenital anomalies | LDL-C | |
| 754.2 | | | | | 27059 | 34 | 27025 | | | | | 35837 | 62 | 35775 | | | Spondylolisthesis, congenital | congenital anomalies | LDL-C | |
| 755 | | | | | 27119 | 94 | 27025 | | | | | 35920 | 145 | 35775 | | | Congenital anomalies of limbs | congenital anomalies | LDL-C | |
| 755.1 | | | | | 27058 | 33 | 27025 | | | | | 35834 | 59 | 35775 | | | Congenital deformities of feet | congenital anomalies | LDL-C | |
| 755.3 | | | | | 27031 | 6 | 27025 | | | | | 35781 | 6 | 35775 | | | Congenital anomaly of fingers/toes | congenital anomalies | LDL-C | |
| 755.4 | | | | | 27034 | 9 | 27025 | | | | | 35795 | 20 | 35775 | | | Congenital anomalies of upper limb, including shoulder girdle | congenital anomalies | LDL-C | |
| 755.6 | | | | | 27065 | 40 | 27025 | | | | | 35837 | 62 | 35775 | | | Other congenital anomalies of lower limb, including pelvic girdle | congenital anomalies | LDL-C | |
| 755.61 | | | | | 27060 | 35 | 27025 | | | | | 35828 | 53 | 35775 | | | Congenital hip dysplasia and deformity | congenital anomalies | LDL-C | |
| 756 | | | | | 27125 | 100 | 27025 | | | | | 35939 | 164 | 35775 | | | Other congenital musculoskeletal anomalies | congenital anomalies | LDL-C | |
| 756.1 | | | | | 27036 | 11 | 27025 | | | | | 35791 | 16 | 35775 | | | Congenital anomalies of abdominal wall; diaphram | congenital anomalies | LDL-C | |
| 756.2 | | | | | 27064 | 39 | 27025 | | | | | 35801 | 26 | 35775 | | | Pectus and other congenital anomalies of ribs/sternum | congenital anomalies | LDL-C | |
| 756.21 | | | | | 27054 | 29 | 27025 | | | | | 35798 | 23 | 35775 | | | Pectus excavatum | congenital anomalies | LDL-C | |
| 756.22 | | | | | 27036 | 11 | 27025 | | | | | 35776 | 1 | 35775 | | | Pectus carinatum | congenital anomalies | LDL-C | |
| 756.3 | | | | | 27041 | 16 | 27025 | | | | | 35840 | 65 | 35775 | | | Congenital anomalies of muscle, tendon, fascia, and connective tissue | congenital anomalies | LDL-C | |
| 756.5 | | | | | 27043 | 18 | 27025 | | | | | 35804 | 29 | 35775 | | | Congenital osteodystrophies | congenital anomalies | LDL-C | |
| 757 | | | | | 26746 | 13 | 26733 | | | | | 35965 | 21 | 35944 | | | Congenital anomalies of the integument | congenital anomalies | LDL-C | |
| 758 | | | | | 26835 | 102 | 26733 | | | | | 36032 | 88 | 35944 | | | Chromosomal anomalies and genetic disorders | congenital anomalies | LDL-C | |
| 758.1 | | | | | 26801 | 68 | 26733 | | | | | 36013 | 69 | 35944 | | | Chromosomal anomalies | congenital anomalies | LDL-C | |
| 759 | | | | | 27017 | 284 | 26733 | | | | | 36141 | 197 | 35944 | | | Other and unspecified congenital anomalies | congenital anomalies | LDL-C | |

| phenotype | male_beta | male_SE | male_OR | male_p | male_n_total | male_n_cases | male_n_controls | female_beta | female_SE | female_OR | female_p | female_n_total | female_n_cases | female_n_controls | interaction_p | interaction_beta | description | group | lab | label |
|---|---|---|---|---|---|---|---|---|---|---|---|---|---|---|---|---|---|---|---|---|
| 759.1 | | | | | 26741 | 8 | 26733 | | | | | 35957 | 13 | 35944 | | | Anomalies of endocrine glands, congenital | congenital anomalies | LDL-C | |
| 760 | 0.951 | 0.04095 | 2.588 | 2.56e-119 | 24911 | 4715 | 20196 | 1.089 | 0.03244 | 2.971 | 3.81e-247 | 32596 | 7782 | 24814 | 5.49e-06 | 0.224 | Back pain | symptoms | LDL-C | |
| 761 | 0.7717 | 0.06102 | 2.163 | 1.16e-36 | 26084 | 1972 | 24112 | 1.134 | 0.04663 | 3.108 | 1.26e-130 | 34098 | 3706 | 30392 | 4.13e-09 | 0.4258 | Cervicalgia | symptoms | LDL-C | |
| 763 | 0.7881 | 0.08021 | 2.199 | 8.74e-23 | 25089 | 1208 | 23881 | 1.077 | 0.06663 | 2.935 | 9.36e-59 | 32717 | 1862 | 30855 | 0.001075 | 0.3264 | Thoracic or lumbosacral neuritis or radiculitis, unspecified | symptoms | LDL-C | |
| 764 | 1.291 | 0.1218 | 3.636 | 3.03e-26 | 24586 | 705 | 23881 | 1.436 | 0.08809 | 4.204 | 9.45e-60 | 32170 | 1315 | 30855 | 0.2198 | 0.1783 | Sciatica | symptoms | LDL-C | |
| 765 | 1.082 | 0.1184 | 2.951 | 6.20e-20 | 24494 | 613 | 23881 | 1.106 | 0.09415 | 3.021 | 7.76e-32 | 31724 | 869 | 30855 | 0.6963 | 0.05655 | Cervical radiculitis | symptoms | LDL-C | |
| 766 | 0.6149 | 0.09731 | 1.849 | 2.64e-10 | 24563 | 682 | 23881 | 0.8726 | 0.08044 | 2.393 | 2.04e-27 | 31922 | 1067 | 30855 | 0.001972 | 0.3663 | Neuralgia, neuritis, and radiculitis NOS | symptoms | LDL-C | |
| 767 | | | | | 23904 | 23 | 23881 | | | | | 30882 | 27 | 30855 | | | Cervicocranial/Cervicobrachial syndrome | symptoms | LDL-C | |
| 769 | | | | | 27744 | 17 | 27727 | | | | | 36864 | 59 | 36805 | | | Nonallopathic lesions NEC | symptoms | LDL-C | |
| 770 | 0.7508 | 0.09922 | 2.119 | 3.82e-14 | 26643 | 686 | 25957 | 0.374 | 0.04418 | 1.453 | 2.56e-17 | 34609 | 3011 | 31598 | 3.14e-04 | -0.3651 | Myalgia and myositis unspecified | symptoms | LDL-C | |
| 771 | 1.097 | 0.1166 | 2.996 | 4.90e-21 | 23704 | 575 | 23129 | 0.8175 | 0.1023 | 2.265 | 1.31e-15 | 31912 | 628 | 31284 | 0.1922 | -0.1879 | Musculoskeletal symptoms referable to limbs | symptoms | LDL-C | |
| 771.1 | 0.8432 | 0.07415 | 2.324 | 5.76e-30 | 24349 | 1220 | 23129 | 0.8807 | 0.06824 | 2.413 | 4.14e-38 | 32708 | 1424 | 31284 | 0.0461 | 0.1865 | Swelling of limb | symptoms | LDL-C | |
| 771.2 | | | | | 23196 | 67 | 23129 | | | | | 31394 | 110 | 31284 | | | Cramp of limb | symptoms | LDL-C | |
| 772 | | | | | 23175 | 134 | 23041 | | | | | 30713 | 257 | 30456 | | | Symptoms of the muscles | symptoms | LDL-C | |
| 772.1 | | | | | 23094 | 53 | 23041 | | | | | 30516 | 60 | 30456 | | | Muscular wasting and disuse atrophy | symptoms | LDL-C | |
| 772.2 | 0.4385 | 0.09618 | 1.55 | 5.14e-06 | 23676 | 635 | 23041 | 0.7277 | 0.07613 | 2.07 | 1.18e-21 | 31533 | 1077 | 30456 | 8.25e-04 | 0.3832 | Spasm of muscle | symptoms | LDL-C | |
| 772.3 | 0.6832 | 0.0667 | 1.98 | 1.27e-24 | 24642 | 1601 | 23041 | 0.8174 | 0.05562 | 2.265 | 6.72e-49 | 32724 | 2268 | 30456 | 0.04201 | 0.1658 | Muscle weakness | symptoms | LDL-C | |
| 772.4 | | | | | 23175 | 134 | 23041 | | | | | 30527 | 71 | 30456 | | | Rhabdomyolysis | symptoms | LDL-C | |
| 772.6 | | | | | 23142 | 101 | 23041 | | | | | 30584 | 128 | 30456 | | | Facial weakness | symptoms | LDL-C | |
| 780 | | | | | 27429 | 77 | 27352 | | | | | 36521 | 68 | 36453 | | | Hypothermia/Chills | symptoms | LDL-C | |
| 781 | 0.6016 | 0.1173 | 1.825 | 2.94e-07 | 26628 | 527 | 26101 | 0.7074 | 0.1034 | 2.029 | 7.77e-12 | 35384 | 686 | 34698 | 0.2082 | 0.1852 | Symptoms involving nervous and musculoskeletal systems | symptoms | LDL-C | |
| 781.1 | | | | | 26112 | 11 | 26101 | | | | | 34735 | 37 | 34698 | | | Loss of height | symptoms | LDL-C | |
| 781.2 | | | | | 26298 | 197 | 26101 | | | | | 34933 | 235 | 34698 | | | Abnormal posture | symptoms | LDL-C | |
| 782 | | | | | 22002 | 106 | 21896 | | | | | 29294 | 158 | 29136 | | | Symptoms involving skin and other integumentary tissue | symptoms | LDL-C | |
| 782.3 | 0.9232 | 0.04958 | 2.517 | 2.20e-77 | 24907 | 3011 | 21896 | 1.19 | 0.04521 | 3.289 | 7.61e-153 | 33039 | 3903 | 29136 | 1.12e-07 | 0.3301 | Edema | symptoms | LDL-C | |
| 782.6 | | | | | 21947 | 51 | 21896 | | | | | 29472 | 336 | 29136 | | | Pallor and flushing | symptoms | LDL-C | |
| 783 | 0.3949 | 0.0387 | 1.484 | 1.91e-24 | 24799 | 4188 | 20611 | 0.5846 | 0.03764 | 1.794 | 2.15e-54 | 32997 | 4266 | 28731 | 1.45e-07 | 0.2578 | Fever of unknown origin | symptoms | LDL-C | |
| 783.1 | | | | | 20666 | 55 | 20611 | | | | | 28768 | 37 | 28731 | | | Postprocedural fever | symptoms | LDL-C | |
| 785 | 0.5572 | 0.0337 | 1.746 | 2.01e-61 | 24378 | 6486 | 17892 | 0.7959 | 0.02757 | 2.217 | 3.17e-183 | 32108 | 11436 | 20672 | 1.30e-12 | 0.2825 | Abdominal pain | symptoms | LDL-C | |
| 788 | 0.8394 | 0.05747 | 2.315 | 2.56e-48 | 26167 | 2188 | 23979 | 0.7686 | 0.0506 | 2.157 | 4.18e-52 | 34916 | 2566 | 32350 | 0.2676 | -0.0786 | Syncope and collapse | symptoms | LDL-C | |
| 789 | 0.1227 | 0.03816 | 1.131 | 0.001304 | 24975 | 4146 | 20829 | 0.4714 | 0.03094 | 1.602 | 1.97e-52 | 32311 | 6758 | 25553 | 4.24e-27 | 0.4779 | Nausea and vomiting | symptoms | LDL-C | |
| 789.1 | | | | | 20853 | 24 | 20829 | | | | | 25611 | 58 | 25553 | | | Persistent vomiting | symptoms | LDL-C | |
| 790 | | | | | 22915 | 183 | 22732 | | | | | 30897 | 312 | 30585 | | | Nonspecific findings on examination of blood | symptoms | LDL-C | |

| phenotype | male_beta | male_SE | male_OR | male_p | male_n_total | male_n_cases | male_n_controls | female_beta | female_SE | female_OR | female_p | female_n_total | female_n_cases | female_n_controls | interaction_p | interaction_beta | description | group | lab | label |
|---|---|---|---|---|---|---|---|---|---|---|---|---|---|---|---|---|---|---|---|---|
| 790.1 | | | | | 22812 | 80 | 22732 | | | | | 30855 | 270 | 30585 | | | Elevated sedimentation rate | symptoms | LDL-C | |
| 790.6 | 1.33 | 0.06259 | 3.783 | 2.90e-100 | 25006 | 2274 | 22732 | 1.495 | 0.05892 | 4.461 | 4.07e-142 | 33298 | 2713 | 30585 | 0.001037 | 0.2683 | Other abnormal blood chemistry | symptoms | LDL-C | |
| 790.8 | | | | | 22867 | 135 | 22732 | | | | | 30869 | 284 | 30585 | | | Elevated C-reactive protein (CRP) | symptoms | LDL-C | |
| 790.9 | | | | | 22737 | 5 | 22732 | | | | | 30591 | 6 | 30585 | | | Abnormal arterial blood gases | symptoms | LDL-C | |
| 791 | | | | | 27648 | 165 | 27483 | | | | | 36840 | 92 | 36748 | | | Gangrene | symptoms | LDL-C | |
| 792 | | | | | 7 | | | 1.006 | 0.06178 | 2.735 | 1.31e-59 | 29446 | 1607 | 27839 | 0.997 | 1.324 | Abnormal Papanicolaou smear of cervix and cervical HPV | genitourinary | LDL-C | |
| 792.1 | | | | | 7 | | | 1.003 | 0.06631 | 2.727 | 1.05e-51 | 29183 | 1344 | 27839 | 0.997 | 1.304 | Papanicolaou smear of cervix or vagina with atypical squamous cells | genitourinary | LDL-C | |
| 794 | | | | | 27589 | 45 | 27544 | | | | | 36771 | 38 | 36733 | | | Abnormal results of other function studies (bladder, pancreas, placenta, spleen, etc) | symptoms | LDL-C | |
| 795 | | | | | 27272 | 12 | 27260 | | | | | 36232 | 106 | 36126 | | | Other and nonspecific abnormal cytological, histological and immunological findings | symptoms | LDL-C | |
| 795.8 | | | | | 27502 | 242 | 27260 | | | | | 36401 | 275 | 36126 | 0.07333 | 0.3201 | Abnormal tumor markers | symptoms | LDL-C | |
| 795.81 | | | | | 27386 | 126 | 27260 | | | | | 36227 | 101 | 36126 | | | Elevated carcinoembryonic antigen [CEA] | symptoms | LDL-C | |
| 795.82 | | | | | 27261 | 1 | 27260 | | | | | 36221 | 95 | 36126 | | | Elevated cancer antigen 125 [CA 125] | symptoms | LDL-C | |
| 796 | 1.333 | 0.08506 | 3.791 | 2.52e-55 | 20872 | 1484 | 19388 | | | | | 4 | | | 0.9942 | -2.653 | Elevated prostate specific antigen [PSA] | genitourinary | LDL-C | |
| 797 | 0.7294 | 0.07445 | 2.074 | 1.16e-22 | 26901 | 1035 | 25866 | 0.7062 | 0.08798 | 2.026 | 9.97e-16 | 36273 | 725 | 35548 | 0.4124 | 0.08453 | Shock | symptoms | LDL-C | |
| 797.1 | | | | | 26238 | 372 | 25866 | | | | | 35746 | 198 | 35548 | 0.7745 | -0.05723 | Cardiogenic shock | symptoms | LDL-C | |
| 798 | 0.8318 | 0.03284 | 2.297 | 1.53e-141 | 23475 | 7649 | 15826 | 1.125 | 0.02779 | 3.081 | 0 | 30980 | 11865 | 19115 | 1.92e-22 | 0.3911 | Malaise and fatigue | symptoms | LDL-C | |
| 798.1 | | | | | 16119 | 293 | 15826 | 1.506 | 0.09938 | 4.511 | 6.61e-52 | 19839 | 724 | 19115 | 0.6153 | 0.09604 | Chronic fatigue syndrome | symptoms | LDL-C | |
| 800 | -0.1786 | 0.07028 | 0.8365 | 0.01106 | 22951 | 1064 | 21887 | 0.4309 | 0.05852 | 1.539 | 1.80e-13 | 30522 | 1698 | 28824 | 1.93e-23 | 0.8374 | Fracture of lower limb | injuries & poisonings | LDL-C | |
| 800.1 | | | | | 22264 | 377 | 21887 | 0.3696 | 0.0986 | 1.447 | 1.78e-04 | 29394 | 570 | 28824 | 8.45e-07 | 0.6883 | Fracture of neck of femur | injuries & poisonings | LDL-C | |
| 800.2 | | | | | 22107 | 220 | 21887 | | | | | 29062 | 238 | 28824 | | | Fracture of unspecified part of femur | injuries & poisonings | LDL-C | |
| 800.3 | | | | | 22240 | 353 | 21887 | | | | | 29291 | 467 | 28824 | 1.92e-09 | 0.8693 | Fracture of tibia and fibula | injuries & poisonings | LDL-C | |
| 800.4 | | | | | 22024 | 137 | 21887 | | | | | 29015 | 191 | 28824 | | | Fracture of patella | injuries & poisonings | LDL-C | |
| 801 | 0.3053 | 0.09636 | 1.357 | 0.001533 | 22470 | 583 | 21887 | 0.798 | 0.07237 | 2.221 | 2.83e-28 | 30057 | 1233 | 28824 | 1.82e-14 | 0.8545 | Fracture of ankle and foot | injuries & poisonings | LDL-C | |
| 801.1 | | | | | 22222 | 335 | 21887 | 0.4617 | 0.1016 | 1.587 | 5.47e-06 | 29349 | 525 | 28824 | 5.20e-07 | 0.7362 | Fracture of foot | injuries & poisonings | LDL-C | |
| 802 | | | | | 22131 | 244 | 21887 | | | | | 29104 | 280 | 28824 | 2.05e-07 | 0.9294 | Fracture of pelvis | injuries & poisonings | LDL-C | |
| 803 | 0.05451 | 0.07258 | 1.056 | 0.4526 | 22909 | 1022 | 21887 | 0.5682 | 0.06151 | 1.765 | 2.55e-20 | 30429 | 1605 | 28824 | 4.22e-20 | 0.8023 | Fracture of upper limb | injuries & poisonings | LDL-C | |
| 803.1 | | | | | 22122 | 235 | 21887 | | | | | 29293 | 469 | 28824 | 3.10e-08 | 0.9368 | Fracture of humerus | injuries & poisonings | LDL-C | |
| 803.2 | | | | | 22365 | 478 | 21887 | 0.5156 | 0.08189 | 1.675 | 3.05e-10 | 29684 | 860 | 28824 | 1.09e-10 | 0.786 | Fracture of radius and ulna | injuries & poisonings | LDL-C | |
| 803.21 | | | | | 21931 | 44 | 21887 | | | | | 28974 | 150 | 28824 | | | Colles' fracture | injuries & poisonings | LDL-C | |
| 803.3 | | | | | 22141 | 254 | 21887 | | | | | 28997 | 173 | 28824 | | | Fracture of clavicle or scapula | injuries & poisonings | LDL-C | |
| 804 | | | | | 22374 | 487 | 21887 | | | | | 29317 | 493 | 28824 | 5.48e-06 | 0.6368 | Fracture of hand or wrist | injuries & poisonings | LDL-C | |

| phenotype | male_beta | male_SE | male_OR | male_p | male_n_total | male_n_cases | male_n_controls | female_beta | female_SE | female_OR | female_p | female_n_total | female_n_cases | female_n_controls | interaction_p | interaction_beta | description | group | lab | label |
|---|---|---|---|---|---|---|---|---|---|---|---|---|---|---|---|---|---|---|---|---|
| 805 | -0.1462 | 0.07858 | 0.8639 | 0.06273 | 22729 | 842 | 21887 | 0.3485 | 0.07448 | 1.417 | 2.88e-06 | 29817 | 993 | 28824 | 3.44e-10 | 0.622 | Fracture of vertebral column without mention of spinal cord injury | injuries & poisonings | LDL-C | |
| 807 | | | | | 22286 | 399 | 21887 | | | | | 29056 | 232 | 28824 | 0.003272 | 0.4902 | Fracture of ribs | injuries & poisonings | LDL-C | |
| 809 | 0.01543 | 0.06041 | 1.016 | 0.7984 | 23404 | 1517 | 21887 | 0.4558 | 0.056 | 1.577 | 4.01e-16 | 30670 | 1846 | 28824 | 1.21e-15 | 0.6079 | Fracture of unspecified bones | injuries & poisonings | LDL-C | |
| 816 | | | | | 26400 | 43 | 26357 | | | | | 35690 | 21 | 35669 | | | Cerebral laceration and contusion | injuries & poisonings | LDL-C | |
| 817 | | | | | 26422 | 65 | 26357 | | | | | 35734 | 65 | 35669 | | | Concussion | injuries & poisonings | LDL-C | |
| 818 | | | | | 26606 | 249 | 26357 | | | | | 35816 | 147 | 35669 | | | Intracranial hemorrhage (injury) | injuries & poisonings | LDL-C | |
| 818.1 | | | | | 26475 | 118 | 26357 | | | | | 35737 | 68 | 35669 | | | Subdural hemorrhage (injury) | injuries & poisonings | LDL-C | |
| 818.2 | | | | | 26451 | 94 | 26357 | | | | | 35727 | 58 | 35669 | | | Subarachnoid hemorrhage (injury) | injuries & poisonings | LDL-C | |
| 819 | -0.3409 | 0.09611 | 0.7112 | 3.90e-04 | 26908 | 551 | 26357 | | | | | 36031 | 362 | 35669 | 1.21e-07 | 0.7313 | Skull and face fracture and other intercranial injury | injuries & poisonings | LDL-C | |
| 823 | | | | | 21319 | | | | | | | 28267 | 2 | 28265 | | | Torus fracture | injuries & poisonings | LDL-C | |
| 830 | 0.5117 | 0.08145 | 1.668 | 3.33e-10 | 26492 | 928 | 25564 | 0.7077 | 0.07259 | 2.029 | 1.88e-22 | 34859 | 1219 | 33640 | 1.36e-04 | 0.3856 | Dislocation | injuries & poisonings | LDL-C | |
| 835 | | | | | 26013 | 449 | 25564 | 1.045 | 0.09257 | 2.842 | 1.57e-29 | 34495 | 855 | 33640 | 0.3509 | 0.1411 | Internal derangement of knee | injuries & poisonings | LDL-C | |
| 836 | | | | | 25643 | 79 | 25564 | | | | | 33724 | 84 | 33640 | | | Traumatic arthropathy | injuries & poisonings | LDL-C | |
| 840 | 0.8956 | 0.07215 | 2.449 | 2.23e-35 | 25492 | 1396 | 24096 | 1.135 | 0.05886 | 3.11 | 8.56e-83 | 33198 | 2125 | 31073 | 0.001283 | 0.2815 | Sprains and strains | injuries & poisonings | LDL-C | |
| 840.1 | | | | | 24130 | 34 | 24096 | | | | | 31116 | 43 | 31073 | | | Muscle/tendon sprain | injuries & poisonings | LDL-C | |
| 840.2 | | | | | 24369 | 273 | 24096 | | | | | 31341 | 268 | 31073 | 0.1334 | 0.3432 | Rotator cuff (capsule) sprain | injuries & poisonings | LDL-C | |
| 840.3 | | | | | 24387 | 291 | 24096 | | | | | 31485 | 412 | 31073 | 1.40e-04 | 0.6501 | Joint/ligament sprain | injuries & poisonings | LDL-C | |
| 841 | | | | | 24387 | 291 | 24096 | 1.184 | 0.1096 | 3.267 | 3.21e-27 | 31714 | 641 | 31073 | 0.5345 | 0.118 | Sprains and strains of back and neck | injuries & poisonings | LDL-C | |
| 842 | | | | | 24179 | 83 | 24096 | | | | | 31195 | 122 | 31073 | | | Other sprains and strains | injuries & poisonings | LDL-C | |
| 850 | | | | | 23223 | 456 | 22767 | | | | | 32260 | 465 | 31795 | 0.08413 | 0.2395 | Hemorrhage or hematoma complicating a procedure | injuries & poisonings | LDL-C | |
| 851 | | | | | 23157 | 390 | 22767 | | | | | 32086 | 291 | 31795 | 0.8996 | 0.03541 | Complications of transplants and reattached limbs | injuries & poisonings | LDL-C | |
| 853 | | | | | 22918 | 151 | 22767 | | | | | 31956 | 161 | 31795 | | | Complication of colostomy or enterostomy | injuries & poisonings | LDL-C | |
| 854 | 0.8996 | 0.09959 | 2.459 | 1.67e-19 | 23435 | 668 | 22767 | 0.6166 | 0.1047 | 1.853 | 3.91e-09 | 32312 | 517 | 31795 | 0.06912 | -0.2386 | Complications of cardiac/vascular device, implant, and graft | injuries & poisonings | LDL-C | |
| phenotype | male_beta | male_SE | male_OR | male_p | male_n_total | male_n_cases | male_n_controls | female_beta | female_SE | female_OR | female_p | female_n_total | female_n_cases | female_n_controls | interaction_p | interaction_beta | description | group | lab | label |
| 855 | | | | | 22967 | 200 | 22767 | | | | | 32019 | 224 | 31795 | | | Complication of nervous system device, implant, and graft | injuries & poisonings | LDL-C | |
| 856 | | | | | 22823 | 56 | 22767 | | | | | 31840 | 45 | 31795 | | | Vascular complications of surgery and medical procedures | injuries & poisonings | LDL-C | |
| 857 | | | | | 22852 | 85 | 22767 | | | | | 31886 | 91 | 31795 | | | Mechanical complication of unspecified genitourinary device, implant, and graft | injuries & poisonings | LDL-C | |
| 858 | | | | | 23191 | 424 | 22767 | -0.345 | 0.09169 | 0.7082 | 1.68e-04 | 32396 | 601 | 31795 | 2.53e-05 | 0.5447 | Complication of internal orthopedic device | injuries & poisonings | LDL-C | |

| phenotype | male_beta | male_SE | male_OR | male_p | male_n_total | male_n_cases | male_n_controls | female_beta | female_SE | female_OR | female_p | female_n_total | female_n_cases | female_n_controls | interaction_p | interaction_beta | description | group | lab | label |
|---|---|---|---|---|---|---|---|---|---|---|---|---|---|---|---|---|---|---|---|---|
| 859 | | | | | 23165 | 398 | 22767 | | | | | 32227 | 432 | 31795 | 0.02663 | 0.3113 | Complication due to other implant and internal device | injuries & poisonings | LDL-C | |
| 860 | | | | | 27702 | 127 | 27575 | | | | | 36911 | 120 | 36791 | | | Bone marrow or stem cell transplant | neoplasms | LDL-C | |
| 870 | -0.3102 | 0.08275 | 0.7333 | 1.77e-04 | 24342 | 736 | 23606 | 0.1041 | 0.08438 | 1.11 | 0.2173 | 33149 | 715 | 32434 | 1.89e-05 | 0.4584 | Open wounds of head; neck; and trunk | injuries & poisonings | LDL-C | |
| 870.1 | | | | | 23654 | 48 | 23606 | | | | | 32454 | 20 | 32434 | | | Open wound or laceration of eye or eyelid | injuries & poisonings | LDL-C | |
| 870.2 | | | | | 23622 | 16 | 23606 | | | | | 32443 | 9 | 32434 | | | Open wound of ear | injuries & poisonings | LDL-C | |
| 870.3 | | | | | 23791 | 185 | 23606 | | | | | 32553 | 119 | 32434 | | | Other open wound of head and face | injuries & poisonings | LDL-C | |
| 870.4 | | | | | 23626 | 20 | 23606 | | | | | 32447 | 13 | 32434 | | | Open wound of nose and sinus | injuries & poisonings | LDL-C | |
| 870.5 | | | | | 23652 | 46 | 23606 | | | | | 32464 | 30 | 32434 | | | Open wound of lip and mouth | injuries & poisonings | LDL-C | |
| 870.6 | | | | | 23625 | 19 | 23606 | | | | | 32440 | 6 | 32434 | | | Open wound of neck | injuries & poisonings | LDL-C | |
| 870.8 | | | | | 23613 | 7 | 23606 | | | | | 32442 | 8 | 32434 | | | Open wound of genital organs | injuries & poisonings | LDL-C | |
| 871 | 0.2971 | 0.0749 | 1.346 | 7.29e-05 | 24632 | 1026 | 23606 | 0.6402 | 0.08221 | 1.897 | 6.81e-15 | 33363 | 929 | 32434 | 5.66e-07 | 0.5129 | Open wounds of extremities | injuries & poisonings | LDL-C | |
| 871.1 | | | | | 23768 | 162 | 23606 | | | | | 32546 | 112 | 32434 | | | Open wound of hand except finger(s) | injuries & poisonings | LDL-C | |
| 871.2 | | | | | 23775 | 169 | 23606 | | | | | 32574 | 140 | 32434 | | | Open wound of finger(s) | injuries & poisonings | LDL-C | |
| 871.3 | | | | | 23720 | 114 | 23606 | | | | | 32510 | 76 | 32434 | | | Open wound of foot except toe(s) alone | injuries & poisonings | LDL-C | |
| 871.4 | | | | | 23638 | 32 | 23606 | | | | | 32453 | 19 | 32434 | | | Open wound of toe(s) | injuries & poisonings | LDL-C | |
| 872 | | | | | 23730 | 124 | 23606 | | | | | 32497 | 63 | 32434 | | | Traumatic amputation | injuries & poisonings | LDL-C | |
| 874 | | | | | 23727 | 121 | 23606 | | | | | 32492 | 58 | 32434 | | | Complication of amputation stump | injuries & poisonings | LDL-C | |
| 875 | | | | | 23705 | 99 | 23606 | | | | | 32557 | 123 | 32434 | | | Non-healing surgical wound | injuries & poisonings | LDL-C | |
| 876 | | | | | 23630 | 24 | 23606 | | | | | 32475 | 41 | 32434 | | | Posttraumatic wound infection not elsewhere classified | injuries & poisonings | LDL-C | |
| 905 | | | | | 27300 | 4 | 27296 | | | | | 36496 | 6 | 36490 | | | Late effects of musculoskeletal and connective tissue injuries | injuries & poisonings | LDL-C | |
| 907 | | | | | 27494 | 198 | 27296 | | | | | 36638 | 148 | 36490 | | | Injuries to the nervous system | injuries & poisonings | LDL-C | |
| 910 | | | | | 27755 | 6 | 27749 | | | | | 36903 | 8 | 36895 | | | Superficial injury, infected | injuries & poisonings | LDL-C | |
| 911 | | | | | 27688 | 32 | 27656 | | | | | 36846 | 32 | 36814 | | | Blister | injuries & poisonings | LDL-C | |
| 912 | | | | | 27054 | 122 | 26932 | | | | | 35677 | 201 | 35476 | | | Insect bite | injuries & poisonings | LDL-C | |
| 913 | | | | | 26968 | 36 | 26932 | | | | | 35544 | 68 | 35476 | | | Toxic effect of venom | injuries & poisonings | LDL-C | |
| 916 | 0.6403 | 0.1036 | 1.897 | 6.31e-10 | 26060 | 586 | 25474 | 0.8947 | 0.08181 | 2.447 | 7.69e-28 | 34442 | 1043 | 33399 | 2.39e-04 | 0.4496 | Contusion | injuries & poisonings | LDL-C | |
| 930 | | | | | 22109 | 280 | 21829 | 0.8838 | 0.0967 | 2.42 | 6.30e-20 | 27961 | 619 | 27342 | 0.6715 | 0.07131 | Allergic reaction to food | injuries & poisonings | LDL-C | |
| 931 | | | | | 21895 | 66 | 21829 | | | | | 27431 | 89 | 27342 | | | Contact dermatitis and other eczema due to plants [except food] | dermatologic | LDL-C | |
| 938 | 1.355 | 0.1101 | 3.879 | 8.40e-35 | 22767 | 938 | 21829 | 1.596 | 0.1182 | 4.932 | 1.59e-41 | 28152 | 810 | 27342 | 0.1663 | 0.2181 | Dermatitis due to solar radiation | dermatologic | LDL-C | |

| phenotype | male_beta | male_SE | male_OR | male_p | male_n_total | male_n_cases | male_n_controls | female_beta | female_SE | female_OR | female_p | female_n_total | female_n_cases | female_n_controls | interaction_p | interaction_beta | description | group | lab | label |
|---|---|---|---|---|---|---|---|---|---|---|---|---|---|---|---|---|---|---|---|---|
| 938.1 | | | | | 21840 | 11 | 21829 | | | | | 27356 | 14 | 27342 | | | Acute dermatitis due to solar radiation | dermatologic | LDL-C | |
| 938.2 | 1.762 | 0.1393 | 5.825 | 1.11e-36 | 22665 | 836 | 21829 | 1.77 | 0.135 | 5.871 | 2.99e-39 | 28065 | 723 | 27342 | 0.8137 | -0.045 | Chronic dermatitis due to solar radiation | dermatologic | LDL-C | |
| 939 | 1.028 | 0.07653 | 2.795 | 3.96e-41 | 23219 | 1390 | 21829 | 1.34 | 0.0606 | 3.819 | 2.54e-108 | 29539 | 2197 | 27342 | 0.01624 | 0.2242 | Atopic/contact dermatitis due to other or unspecified | dermatologic | LDL-C | |
| 939.1 | | | | | 21831 | 2 | 21829 | | | | | 27354 | 12 | 27342 | | | Contact and allergic dermatitis of eyelid | dermatologic | LDL-C | |
| 941 | | | | | 21871 | 42 | 21829 | | | | | 27432 | 90 | 27342 | | | Adverse reaction to serum or vaccine | injuries & poisonings | LDL-C | |
| 942 | | | | | 21868 | 39 | 21829 | | | | | 27392 | 50 | 27342 | | | Infusion and transfusion reaction | injuries & poisonings | LDL-C | |
| 946 | | | | | 21895 | 66 | 21829 | | | | | 27494 | 152 | 27342 | | | Anaphylactic shock NOS | injuries & poisonings | LDL-C | |
| 947 | | | | | 21986 | 157 | 21829 | | | | | 27830 | 488 | 27342 | 0.1127 | 0.3475 | Urticaria | dermatologic | LDL-C | |
| 949 | 0.6601 | 0.1091 | 1.935 | 1.46e-09 | 22350 | 521 | 21829 | 0.9185 | 0.0722 | 2.506 | 4.48e-37 | 28540 | 1198 | 27342 | 0.05284 | 0.2403 | Allergies, other | injuries & poisonings | LDL-C | |
| 949.1 | | | | | 21836 | 7 | 21829 | | | | | 27350 | 8 | 27342 | | | Diaper or napkin rash | injuries & poisonings | LDL-C | |
| 952 | | | | | 27725 | 66 | 27659 | | | | | 36923 | 32 | 36891 | | | Spinal cord injury without evidence of spinal bone injury | injuries & poisonings | LDL-C | |
| 957 | | | | | 27659 | | | | | | | 36893 | 2 | 36891 | | | Injury to other and unspecified nerves | injuries & poisonings | LDL-C | |
| 958 | | | | | 27212 | 374 | 26838 | | | | | 36518 | 232 | 36286 | 0.04046 | 0.3507 | Certain early complications of trauma or procedure | injuries & poisonings | LDL-C | |
| 958.1 | | | | | 26905 | 67 | 26838 | | | | | 36331 | 45 | 36286 | | | Postoperative shock | injuries & poisonings | LDL-C | |
| 958.2 | | | | | 26974 | 136 | 26838 | | | | | 36364 | 78 | 36286 | | | Traumatic and surgical subcutaneous emphysema | injuries & poisonings | LDL-C | |
| 960 | 0.7288 | 0.07459 | 2.073 | 1.50e-22 | 21000 | 1222 | 19778 | 0.5828 | 0.05473 | 1.791 | 1.77e-26 | 28211 | 2139 | 26072 | 0.7679 | 0.02555 | Poisoning by antibiotics | injuries & poisonings | LDL-C | |
| 960.1 | | | | | 19779 | 1 | 19778 | | | | | 26073 | 1 | 26072 | | | Adverse effects of antibacterials (not penicillins) | injuries & poisonings | LDL-C | |
| 960.2 | 0.7637 | 0.09762 | 2.146 | 5.15e-15 | 20476 | 698 | 19778 | 0.6027 | 0.07179 | 1.827 | 4.63e-17 | 27268 | 1196 | 26072 | 0.8273 | 0.02466 | Allergy/adverse effect of penicillin | injuries & poisonings | LDL-C | |
| 960.3 | | | | | 19779 | 1 | 19778 | | | | | 26073 | 1 | 26072 | | | Poisoning by antifungal antibiotics | injuries & poisonings | LDL-C | |
| 961 | | | | | 19843 | 65 | 19778 | | | | | 26198 | 126 | 26072 | | | Poisoning by other anti-infectives | injuries & poisonings | LDL-C | |
| 961.1 | | | | | 20163 | 385 | 19778 | 0.5748 | 0.07653 | 1.777 | 5.89e-14 | 27149 | 1077 | 26072 | 0.4729 | 0.1013 | Poisoning/allergy of sulfonamides | injuries & poisonings | LDL-C | |
| 962 | | | | | 19885 | 107 | 19778 | | | | | 26193 | 121 | 26072 | | | Poisoning by hormones and synthetic substitutes | injuries & poisonings | LDL-C | |
| 962.1 | | | | | 19862 | 84 | 19778 | | | | | 26166 | 94 | 26072 | | | Adrenal cortical steroids causing adverse effects in therapeutic use | injuries & poisonings | LDL-C | |
| 962.2 | | | | | 19800 | 22 | 19778 | | | | | 26094 | 22 | 26072 | | | Insulins and antidiabetic agents causing adverse effects in therapeutic use | injuries & poisonings | LDL-C | |
| 962.3 | | | | | 19778 | | | | | | | 26078 | 6 | 26072 | | | Hormones and synthetic substitutes causing adverse effects in therapeutic use | injuries & poisonings | LDL-C | |
| 963 | -0.1479 | 0.09442 | 0.8625 | 0.1172 | 20349 | 571 | 19778 | 0.1893 | 0.09442 | 1.208 | 0.04501 | 26631 | 559 | 26072 | 8.90e-04 | 0.4034 | Poisoning by primarily systemic agents | injuries & poisonings | LDL-C | |
| 963.1 | -0.1212 | 0.09501 | 0.8859 | 0.202 | 20340 | 562 | 19778 | 0.17 | 0.0957 | 1.185 | 0.07575 | 26617 | 545 | 26072 | 0.003527 | 0.3576 | Antineoplastic and immunosuppressive drugs causing adverse effects | injuries & poisonings | LDL-C | |

| phenotype | male_beta | male_SE | male_OR | male_p | male_n_total | male_n_cases | male_n_controls | female_beta | female_SE | female_OR | female_p | female_n_total | female_n_cases | female_n_controls | interaction_p | interaction_beta | description | group | lab | label |
|---|---|---|---|---|---|---|---|---|---|---|---|---|---|---|---|---|---|---|---|---|
| 964 | | | | | 19833 | 55 | 19778 | | | | | 26136 | 64 | 26072 | | | Poisoning by agents primarily affecting blood constituents | injuries & poisonings | LDL-C | |
| 964.1 | | | | | 19833 | 55 | 19778 | | | | | 26131 | 59 | 26072 | | | Anticoagulants causing adverse effects | injuries & poisonings | LDL-C | |
| 965 | | | | | 20187 | 409 | 19778 | 0.6369 | 0.08563 | 1.891 | 1.02e-13 | 26882 | 810 | 26072 | 0.9422 | -0.0104 | Poisoning by analgesics, antipyretics, and antirheumatics | injuries & poisonings | LDL-C | |
| 965.1 | 0.5446 | 0.106 | 1.724 | 2.78e-07 | 20350 | 572 | 19778 | 0.6908 | 0.07085 | 1.995 | 1.85e-22 | 27386 | 1314 | 26072 | 0.05651 | 0.2291 | Opiates and related narcotics causing adverse effects in therapeutic use | injuries & poisonings | LDL-C | |
| 965.2 | | | | | 19780 | 2 | 19778 | | | | | 26073 | 1 | 26072 | | | Antirheumatics causing adverse effects in therapeutic use | injuries & poisonings | LDL-C | |
| 965.3 | | | | | 19783 | 5 | 19778 | | | | | 26085 | 13 | 26072 | | | Salicylates causing adverse effects in therapeutic use | injuries & poisonings | LDL-C | |
| 966 | | | | | 19797 | 19 | 19778 | | | | | 26089 | 17 | 26072 | | | Poisoning by anticonvulsants and anti-Parkinsonism drugs | injuries & poisonings | LDL-C | |
| 967 | | | | | 19831 | 53 | 19778 | | | | | 26222 | 150 | 26072 | | | Adverse effects of sedatives or other central nervous system depressants and anesthetics | injuries & poisonings | LDL-C | |
| 969 | | | | | 19838 | 60 | 19778 | | | | | 26169 | 97 | 26072 | | | Poisoning by psychotropic agents | injuries & poisonings | LDL-C | |
| 971 | | | | | 19780 | 2 | 19778 | | | | | 26075 | 3 | 26072 | | | Poisoning by drugs primarily affecting the autonomic nervous system | injuries & poisonings | LDL-C | |
| 972 | | | | | 19849 | 71 | 19778 | | | | | 26139 | 67 | 26072 | | | Poisoning by agents primarily affecting the cardiovascular system | injuries & poisonings | LDL-C | |
| 972.1 | | | | | 19792 | 14 | 19778 | | | | | 26083 | 11 | 26072 | | | Cardiac rhythm regulators causing adverse effects in therapeutic use | injuries & poisonings | LDL-C | |
| 972.2 | | | | | 19778 | | | | | | | 26072 | | | | | Antilipemic and antiarteriosclerotic drugs causing adverse effects in therapeutic use | injuries & poisonings | LDL-C | |
| 972.6 | | | | | 19783 | 5 | 19778 | | | | | 26077 | 5 | 26072 | | | Antihypertensive agents causing adverse effects | injuries & poisonings | LDL-C | |
| 973 | | | | | 19785 | 7 | 19778 | | | | | 26074 | 2 | 26072 | | | Poisoning by agents primarily affecting the gastrointestinal system | injuries & poisonings | LDL-C | |
| 974 | | | | | 19842 | 64 | 19778 | | | | | 26094 | 22 | 26072 | | | Poisoning by water, mineral, and uric acid metabolism drugs | injuries & poisonings | LDL-C | |
| 975 | | | | | 19784 | 6 | 19778 | | | | | 26079 | 7 | 26072 | | | Poisoning by agents primarily acting on the smooth and skeletal muscles and respiratory system | injuries & poisonings | LDL-C | |
| 976 | | | | | 19780 | 2 | 19778 | | | | | 26075 | 3 | 26072 | | | Poisoning by agents primarily affecting skin & mucous membrane, ophthalmological, otorhinolaryngological, & dental drugs | injuries & poisonings | LDL-C | |
| 977 | 0.983 | 0.07833 | 2.672 | 4.02e-36 | 21166 | 1388 | 19778 | 0.7383 | 0.05663 | 2.092 | 7.33e-39 | 28281 | 2209 | 26072 | 0.04953 | -0.1812 | Personal history of allergy to medicinal agents | injuries & poisonings | LDL-C | |
| 979 | 0.449 | 0.07821 | 1.567 | 9.44e-09 | 20676 | 898 | 19778 | 0.4821 | 0.07628 | 1.619 | 2.62e-10 | 27006 | 934 | 26072 | 0.005895 | 0.2737 | Adverse drug events and drug allergies | injuries & poisonings | LDL-C | |
| 980 | 0.3222 | 0.06846 | 1.38 | 2.53e-06 | 26597 | 1176 | 25421 | 0.3448 | 0.07337 | 1.412 | 2.61e-06 | 35862 | 1000 | 34862 | 0.1197 | 0.1419 | Encounter for long-term (current) use of antibiotics | infectious diseases | LDL-C | |
| 981 | | | | | 27615 | 2 | 27613 | | | | | 36741 | 3 | 36738 | | | Toxic effect of (non-ethyl) alcohol and petroleum and other solvents | injuries & poisonings | LDL-C | |
| 983 | | | | | 27616 | 3 | 27613 | | | | | 36738 | | | | | Toxic effect of corrosive aromatics, acids, and caustic alkalis | injuries & poisonings | LDL-C | |
| 984 | | | | | 27616 | 3 | 27613 | | | | | 36739 | 1 | 36738 | | | Toxic effect of lead and its compounds (including fumes) | injuries & poisonings | LDL-C | |
| 985 | | | | | 27622 | 9 | 27613 | | | | | 36745 | 7 | 36738 | | | Toxic effect of other metals | injuries & poisonings | LDL-C | |

| phenotype | male_beta | male_SE | male_OR | male_p | male_n_total | male_n_cases | male_n_controls | female_beta | female_SE | female_OR | female_p | female_n_total | female_n_cases | female_n_controls | interaction_p | interaction_beta | description | group | lab | label |
|---|---|---|---|---|---|---|---|---|---|---|---|---|---|---|---|---|---|---|---|---|
| 986 | | | | | 27616 | 3 | 27613 | | | | | 36740 | 2 | 36738 | | | Toxic effect of carbon monoxide | injuries & poisonings | LDL-C | |
| 987 | | | | | 27617 | 4 | 27613 | | | | | 36745 | 7 | 36738 | | | Toxic effect of other gases, fumes, or vapors | injuries & poisonings | LDL-C | |
| 988 | | | | | 27613 | | | | | | | 36740 | 2 | 36738 | | | Toxic effect of noxious substances eaten as food | injuries & poisonings | LDL-C | |
| 989 | | | | | 27632 | 19 | 27613 | | | | | 36758 | 20 | 36738 | | | Toxic effect of other substances, chiefly nonmedicinal as to source | injuries & poisonings | LDL-C | |
| 990 | -0.2765 | 0.07628 | 0.7584 | 2.88e-04 | 23446 | 892 | 22554 | -0.05832 | 0.08064 | 0.9433 | 0.4696 | 33242 | 782 | 32460 | 0.02112 | 0.2307 | Effects radiation NOS | injuries & poisonings | LDL-C | |
| 994 | 0.2385 | 0.04607 | 1.269 | 2.27e-07 | 26534 | 2690 | 23844 | 0.2145 | 0.05082 | 1.239 | 2.43e-05 | 35853 | 2103 | 33750 | 0.2093 | 0.07684 | Sepsis and SIRS | injuries & poisonings | LDL-C | |
| 994.1 | 0.0101 | 0.09395 | 1.01 | 0.9144 | 24423 | 579 | 23844 | | | | | 34199 | 449 | 33750 | 0.05879 | 0.2406 | Systemic inflammatory response syndrome (SIRS) | injuries & poisonings | LDL-C | |
| 994.2 | 0.3007 | 0.04933 | 1.351 | 1.10e-09 | 26190 | 2346 | 23844 | 0.2878 | 0.05413 | 1.333 | 1.06e-07 | 35609 | 1859 | 33750 | 0.1704 | 0.08978 | Sepsis | injuries & poisonings | LDL-C | |
| 994.21 | 0.3146 | 0.07701 | 1.37 | 4.41e-05 | 24740 | 896 | 23844 | 0.3656 | 0.08886 | 1.441 | 3.88e-05 | 34424 | 674 | 33750 | 0.07724 | 0.185 | Septic shock | injuries & poisonings | LDL-C | |
| 008 | 1.138 | 0.08956 | 3.122 | 5.08e-37 | 27060 | 574 | 26486 | 0.8521 | 0.08215 | 2.345 | 3.31e-25 | 35831 | 704 | 35127 | 0.1621 | -0.1631 | Intestinal infection | infectious diseases | UricA | |
| 008.5 | | | | | 26875 | 389 | 26486 | | | | | 35533 | 406 | 35127 | 0.6127 | -0.075 | Bacterial enteritis | infectious diseases | UricA | |
| 008.51 | | | | | 26487 | 1 | 26486 | | | | | 35130 | 3 | 35127 | | | Intestinal e.coli | infectious diseases | UricA | |
| 008.52 | | | | | 26824 | 338 | 26486 | | | | | 35475 | 348 | 35127 | 0.6576 | 0.07032 | Intestinal infection due to C. difficile | infectious diseases | UricA | |
| 008.6 | | | | | 26592 | 106 | 26486 | | | | | 35263 | 136 | 35127 | | | Viral Enteritis | infectious diseases | UricA | |
| 008.7 | | | | | 26495 | 9 | 26486 | | | | | 35133 | 6 | 35127 | | | Intestinal infection due to protozoa | infectious diseases | UricA | |
| 010 | | | | | 21692 | 51 | 21641 | | | | | 30214 | 54 | 30160 | | | Tuberculosis | infectious diseases | UricA | |
| 031 | | | | | 21701 | 60 | 21641 | | | | | 30222 | 62 | 30160 | | | Diseases due to other mycobacteria | infectious diseases | UricA | |
| 031.1 | | | | | 21641 | | | | | | | 30160 | | | | | Leprosy | infectious diseases | UricA | |
| 038 | 0.9024 | 0.04264 | 2.466 | 2.03e-99 | 24701 | 3060 | 21641 | 1.054 | 0.04614 | 2.869 | 1.66e-115 | 32656 | 2496 | 30160 | 6.31e-04 | 0.2078 | Septicemia | infectious diseases | UricA | |
| 038.1 | | | | | 22136 | 495 | 21641 | | | | | 30550 | 390 | 30160 | 8.76e-04 | 0.4585 | Gram negative septicemia | infectious diseases | UricA | |
| 038.2 | | | | | 22055 | 414 | 21641 | | | | | 30433 | 273 | 30160 | 0.07628 | 0.2833 | Gram positive septicemia | infectious diseases | UricA | |
| 038.3 | 1.115 | 0.05946 | 3.05 | 1.85e-78 | 23021 | 1380 | 21641 | 1.31 | 0.06748 | 3.707 | 5.68e-84 | 31182 | 1022 | 30160 | 0.006062 | 0.2385 | Bacteremia | infectious diseases | UricA | |
| 041 | 0.8154 | 0.0432 | 2.26 | 1.78e-79 | 24639 | 2998 | 21641 | 0.9677 | 0.04174 | 2.632 | 6.56e-119 | 33323 | 3163 | 30160 | 4.32e-05 | 0.2378 | Bacterial infection NOS | infectious diseases | UricA | |
| 041.1 | 0.8418 | 0.06192 | 2.32 | 4.30e-42 | 22993 | 1352 | 21641 | 1.05 | 0.06686 | 2.859 | 1.30e-55 | 31259 | 1099 | 30160 | 0.003749 | 0.2551 | Staphylococcus infections | infectious diseases | UricA | |
| 041.11 | 0.8605 | 0.09219 | 2.364 | 1.02e-20 | 22231 | 590 | 21641 | | | | | 30598 | 438 | 30160 | 0.1409 | 0.1983 | Methicillin sensitive Staphylococcus aureus | infectious diseases | UricA | |
| 041.12 | 0.6632 | 0.0929 | 1.941 | 9.43e-13 | 22233 | 592 | 21641 | | | | | 30654 | 494 | 30160 | 0.001063 | 0.4264 | Methicillin resistant Staphylococcus aureus | infectious diseases | UricA | |
| 041.2 | 0.7841 | 0.08424 | 2.19 | 1.30e-20 | 22331 | 690 | 21641 | 1.209 | 0.08294 | 3.35 | 3.95e-48 | 30829 | 669 | 30160 | 4.70e-05 | 0.465 | Streptococcus infection | infectious diseases | UricA | |
| 041.21 | | | | | 21646 | 5 | 21641 | | | | | 30170 | 10 | 30160 | | | Rheumatic fever / chorea | infectious diseases | UricA | |
| 041.4 | | | | | 22126 | 485 | 21641 | 1.11 | 0.07205 | 3.035 | 1.41e-53 | 31066 | 906 | 30160 | 4.14e-04 | 0.4163 | E. coli | infectious diseases | UricA | |

| phenotype | male_beta | male_SE | male_OR | male_p | male_n_total | male_n_cases | male_n_controls | female_beta | female_SE | female_OR | female_p | female_n_total | female_n_cases | female_n_controls | interaction_p | interaction_beta | description | group | lab | label |
|---|---|---|---|---|---|---|---|---|---|---|---|---|---|---|---|---|---|---|---|---|
| 041.8 | | | | | 21691 | 50 | 21641 | | | | | 30238 | 78 | 30160 | | | H. pylori | infectious diseases | UricA | |
| 041.9 | | | | | 21990 | 349 | 21641 | | | | | 30526 | 366 | 30160 | 0.00143 | 0.4909 | Infection with drug-resistant microorganisms | infectious diseases | UricA | |
| 053 | | | | | 22267 | 352 | 21915 | 0.7022 | 0.09259 | 2.018 | 3.37e-14 | 29621 | 557 | 29064 | 0.06644 | -0.2592 | Herpes zoster | infectious diseases | UricA | |
| 053.1 | | | | | 21986 | 71 | 21915 | | | | | 29192 | 128 | 29064 | | | Herpes zoster with nervous system complications | infectious diseases | UricA | |
| 054 | | | | | 22206 | 291 | 21915 | 0.6898 | 0.09512 | 1.993 | 4.11e-13 | 29622 | 558 | 29064 | 1.53e-06 | -0.7309 | Herpes simplex | infectious diseases | UricA | |
| 070 | 0.6012 | 0.07315 | 1.824 | 2.05e-16 | 22942 | 1027 | 21915 | 0.5448 | 0.0963 | 1.724 | 1.54e-08 | 29665 | 601 | 29064 | 0.8232 | 0.02626 | Viral hepatitis | infectious diseases | UricA | |
| phenotype | male_beta | male_SE | male_OR | male_p | male_n_total | male_n_cases | male_n_controls | female_beta | female_SE | female_OR | female_p | female_n_total | female_n_cases | female_n_controls | interaction_p | interaction_beta | description | group | lab | label |
| 070.1 | | | | | 21931 | 16 | 21915 | | | | | 29069 | 5 | 29064 | | | Viral hepatitis A | infectious diseases | UricA | |
| 070.2 | | | | | 22087 | 172 | 21915 | | | | | 29138 | 74 | 29064 | | | Viral hepatitis B | infectious diseases | UricA | |
| 070.3 | 0.4705 | 0.08021 | 1.601 | 4.47e-09 | 22806 | 891 | 21915 | 0.5436 | 0.1043 | 1.722 | 1.86e-07 | 29579 | 515 | 29064 | 0.2693 | 0.1414 | Viral hepatitis C | infectious diseases | UricA | |
| 070.4 | | | | | 22042 | 127 | 21915 | | | | | 29294 | 230 | 29064 | | | Chronic hepatitis | infectious diseases | UricA | |
| 070.9 | | | | | 22131 | 216 | 21915 | | | | | 29270 | 206 | 29064 | | | Hepatitis NOS | infectious diseases | UricA | |
| 071 | 2.224 | 0.09468 | 9.243 | 5.45e-122 | 22529 | 614 | 21915 | | | | | 29177 | 113 | 29064 | 0.156 | 0.3141 | Human immunodeficiency virus [HIV] disease | infectious diseases | UricA | |
| 071.1 | 2.27 | 0.09583 | 9.684 | 4.18e-124 | 22521 | 606 | 21915 | | | | | 29170 | 106 | 29064 | 0.3459 | 0.2131 | HIV infection, symptomatic | infectious diseases | UricA | |
| 078 | 1.062 | 0.08427 | 2.892 | 2.06e-36 | 22638 | 723 | 21915 | 0.4433 | 0.08358 | 1.558 | 1.14e-07 | 29855 | 791 | 29064 | 4.73e-05 | -0.4622 | Viral warts & HPV | infectious diseases | UricA | |
| 079 | 1.483 | 0.0878 | 4.408 | 4.92e-64 | 22615 | 700 | 21915 | 0.8862 | 0.07228 | 2.426 | 1.49e-34 | 30057 | 993 | 29064 | 4.67e-05 | -0.436 | Viral infection | infectious diseases | UricA | |
| 079.1 | | | | | 21924 | 9 | 21915 | | | | | 29075 | 11 | 29064 | | | Varicella infection | infectious diseases | UricA | |
| 079.2 | | | | | 21970 | 55 | 21915 | | | | | 29142 | 78 | 29064 | | | Infectious mononucleosis | infectious diseases | UricA | |
| 079.9 | | | | | 21977 | 62 | 21915 | | | | | 29164 | 100 | 29064 | | | Viremia, NOS | infectious diseases | UricA | |
| 080 | 0.0803 | 0.0962 | 1.084 | 0.4038 | 26108 | 681 | 25427 | 0.343 | 0.08982 | 1.409 | 1.34e-04 | 35329 | 748 | 34581 | 0.008084 | 0.3387 | Postoperative infection | infectious diseases | UricA | |
| 081 | 0.6932 | 0.08091 | 2 | 1.06e-17 | 26193 | 766 | 25427 | 0.836 | 0.08223 | 2.307 | 2.81e-24 | 35303 | 722 | 34581 | 0.01962 | 0.2598 | Infection/inflammation of internal prosthetic device; implant; and graft | infectious diseases | UricA | |
| 081.1 | | | | | 25468 | 41 | 25427 | | | | | 34621 | 40 | 34581 | | | Graft-versus-host disease | infectious diseases | UricA | |
| 081.11 | | | | | 25439 | 12 | 25427 | | | | | 34597 | 16 | 34581 | | | Acute graft-versus-host disease | infectious diseases | UricA | |
| 081.12 | | | | | 25458 | 31 | 25427 | | | | | 34611 | 30 | 34581 | | | Chronic graft-versus-host disease | infectious diseases | UricA | |
| 090 | | | | | 27678 | 201 | 27477 | | | | | 36786 | 64 | 36722 | | | Sexually transmitted infections (not HIV or hepatitis) | infectious diseases | UricA | |
| 090.2 | | | | | 27533 | 56 | 27477 | | | | | 36735 | 13 | 36722 | | | Gonococcal infections | infectious diseases | UricA | |
| 090.3 | | | | | 27479 | 2 | 27477 | | | | | 36731 | 9 | 36722 | | | Venereal diseases due to Chlamydia trachomatis | infectious diseases | UricA | |
| 110 | 0.6658 | 0.05982 | 1.946 | 9.08e-29 | 24822 | 1390 | 23432 | 0.5647 | 0.06529 | 1.759 | 5.20e-18 | 31711 | 1224 | 30487 | 0.3335 | -0.08332 | Dermatophytosis / Dermatomycosis | infectious diseases | UricA | |
| 110.1 | 0.6615 | 0.06161 | 1.938 | 6.79e-27 | 24733 | 1301 | 23432 | 0.548 | 0.06728 | 1.73 | 3.79e-16 | 31638 | 1151 | 30487 | 0.3354 | -0.08548 | Dermatophytosis | infectious diseases | UricA | |

| phenotype | male_beta | male_SE | male_OR | male_p | male_n_total | male_n_cases | male_n_controls | female_beta | female_SE | female_OR | female_p | female_n_total | female_n_cases | female_n_controls | interaction_p | interaction_beta | description | group | lab | label |
|---|---|---|---|---|---|---|---|---|---|---|---|---|---|---|---|---|---|---|---|---|
| 110.11 | 0.7149 | 0.0694 | 2.044 | 7.03e-25 | 24428 | 996 | 23432 | 0.5897 | 0.07414 | 1.803 | 1.82e-15 | 31407 | 920 | 30487 | 0.3225 | -0.09769 | Dermatophytosis of nail | infectious diseases | UricA | |
| 110.12 | | | | | 23675 | 243 | 23432 | | | | | 30615 | 128 | 30487 | | | Althete's foot | infectious diseases | UricA | |
| 110.13 | | | | | 23580 | 148 | 23432 | | | | | 30581 | 94 | 30487 | | | Dermatophytosis of the body | infectious diseases | UricA | |
| 110.2 | | | | | 23511 | 79 | 23432 | | | | | 30547 | 60 | 30487 | | | Dermatomycoses | infectious diseases | UricA | |
| 112 | 1.045 | 0.07646 | 2.842 | 1.73e-42 | 24237 | 805 | 23432 | 0.8855 | 0.05604 | 2.424 | 3.00e-56 | 32100 | 1613 | 30487 | 0.9038 | -0.01106 | Candidiasis | infectious diseases | UricA | |
| 112.3 | | | | | 23511 | 79 | 23432 | | | | | 30667 | 180 | 30487 | | | Candidiasis of skin and nails | infectious diseases | UricA | |
| 117 | | | | | 23791 | 359 | 23432 | | | | | 30831 | 344 | 30487 | 0.3144 | -0.1587 | Mycoses | infectious diseases | UricA | |
| 117.1 | | | | | 23530 | 98 | 23432 | | | | | 30601 | 114 | 30487 | | | Histoplasmosis | infectious diseases | UricA | |
| 117.2 | | | | | 23434 | 2 | 23432 | | | | | 30487 | | | | | Coccidioidomycosis | infectious diseases | UricA | |
| 117.3 | | | | | 23448 | 16 | 23432 | | | | | 30500 | 13 | 30487 | | | Blastomycotic infection | infectious diseases | UricA | |
| 117.4 | | | | | 23501 | 69 | 23432 | | | | | 30544 | 57 | 30487 | | | Aspergillosis | infectious diseases | UricA | |
| 130 | | | | | 26354 | 10 | 26344 | | | | | 35233 | 9 | 35224 | | | Spirochetal infection | infectious diseases | UricA | |
| 130.1 | | | | | 26349 | 5 | 26344 | | | | | 35227 | 3 | 35224 | | | Lyme disease | infectious diseases | UricA | |
| 131 | | | | | 26353 | 9 | 26344 | | | | | 35256 | 32 | 35224 | | | Protozoan infection | infectious diseases | UricA | |
| 132 | | | | | 26380 | 36 | 26344 | | | | | 35274 | 50 | 35224 | | | Infestation (lice, mites) | infectious diseases | UricA | |
| 132.1 | | | | | 26348 | 4 | 26344 | | | | | 35245 | 21 | 35224 | | | Pediculosis and phthirus infestation | infectious diseases | UricA | |
| 133 | | | | | 26352 | 8 | 26344 | | | | | 35226 | 2 | 35224 | | | Arthropod-borne diseases | infectious diseases | UricA | |
| 134 | | | | | 26349 | 5 | 26344 | | | | | 35248 | 24 | 35224 | | | Helminthiases | infectious diseases | UricA | |
| 134.1 | | | | | 26347 | 3 | 26344 | | | | | 35236 | 12 | 35224 | | | Intestinal helminthiases | infectious diseases | UricA | |
| 136 | | | | | 26733 | 389 | 26344 | | | | | 35599 | 375 | 35224 | 0.1934 | 0.1925 | Other infectious and parasitic diseases | infectious diseases | UricA | |
| 145 | -0.2074 | 0.1053 | 0.8127 | 0.04895 | 26903 | 624 | 26279 | | | | | 36280 | 243 | 36037 | 0.4567 | -0.1628 | Cancer of mouth | neoplasms | UricA | |
| 145.1 | | | | | 26299 | 20 | 26279 | | | | | 36044 | 7 | 36037 | | | Cancer of lip | neoplasms | UricA | |
| 145.2 | | | | | 26660 | 381 | 26279 | | | | | 36136 | 99 | 36037 | | | Cancer of tongue | neoplasms | UricA | |
| 145.3 | | | | | 26384 | 105 | 26279 | | | | | 36087 | 50 | 36037 | | | Cancer of major salivary glands | neoplasms | UricA | |
| 145.4 | | | | | 26326 | 47 | 26279 | | | | | 36068 | 31 | 36037 | | | Cancer of the gums | neoplasms | UricA | |
| 145.5 | | | | | 26338 | 59 | 26279 | | | | | 36058 | 21 | 36037 | | | Cancer of the mouth floor | neoplasms | UricA | |
| 149 | -0.1426 | 0.08611 | 0.8671 | 0.09763 | 27197 | 918 | 26279 | | | | | 36384 | 347 | 36037 | 0.4129 | -0.1463 | Cancer of larynx, pharynx, nasal cavities | neoplasms | UricA | |
| 149.1 | | | | | 26708 | 429 | 26279 | | | | | 36147 | 110 | 36037 | 0.1259 | -0.5018 | Cancer of oropharynx | neoplasms | UricA | |
| 149.2 | | | | | 26347 | 68 | 26279 | | | | | 36075 | 38 | 36037 | | | Cancer of nasopharynx | neoplasms | UricA | |
| 149.3 | | | | | 26333 | 54 | 26279 | | | | | 36064 | 27 | 36037 | | | Cancer of hypopharynx | neoplasms | UricA | |
| 149.4 | | | | | 26607 | 328 | 26279 | | | | | 36132 | 95 | 36037 | | | Cancer of larynx | neoplasms | UricA | |
| 149.5 | | | | | 26723 | 444 | 26279 | | | | | 36197 | 160 | 36037 | 0.1477 | -0.3922 | Hx of malignant neoplasm of oral cavity and pharynx | neoplasms | UricA | |
| 149.9 | | | | | 26377 | 98 | 26279 | | | | | 36109 | 72 | 36037 | | | Cancer of of nasal cavities | neoplasms | UricA | |
| 150 | | | | | 23056 | 399 | 22657 | | | | | 32735 | 70 | 32665 | | | Cancer of esophagus | neoplasms | UricA | |

| phenotype | male_beta | male_SE | male_OR | male_p | male_n_total | male_n_cases | male_n_controls | female_beta | female_SE | female_OR | female_p | female_n_total | female_n_cases | female_n_controls | interaction_p | interaction_beta | description | group | lab | label |
|---|---|---|---|---|---|---|---|---|---|---|---|---|---|---|---|---|---|---|---|---|
| 151 | | | | | 23072 | 415 | 22657 | | | | | 32805 | 140 | 32665 | 0.07554 | 0.4966 | Cancer of stomach | neoplasms | UricA | |
| 153 | 0.5158 | 0.06631 | 1.675 | 7.35e-15 | 21472 | 1270 | 20202 | 0.04318 | 0.08913 | 1.044 | 0.6281 | 30905 | 948 | 29957 | 4.21e-05 | -0.4453 | Colorectal cancer | neoplasms | UricA | |
| 153.2 | 0.2878 | 0.08102 | 1.333 | 3.82e-04 | 21102 | 900 | 20202 | 0.08288 | 0.09673 | 1.086 | 0.3916 | 30731 | 774 | 29957 | 0.1393 | -0.1828 | Colon cancer | neoplasms | UricA | |
| 153.3 | 0.652 | 0.08436 | 1.919 | 1.09e-14 | 20940 | 738 | 20202 | | | | | 30403 | 446 | 29957 | 1.52e-05 | -0.6715 | Malignant neoplasm of rectum, rectosigmoid junction, and anus | neoplasms | UricA | |
| phenotype | male_beta | male_SE | male_OR | male_p | male_n_total | male_n_cases | male_n_controls | female_beta | female_SE | female_OR | female_p | female_n_total | female_n_cases | female_n_controls | interaction_p | interaction_beta | description | group | lab | label |
| 155 | | | | | 23097 | 440 | 22657 | | | | | 32892 | 227 | 32665 | 0.4515 | 0.1494 | Cancer of liver and intrahepatic bile duct | neoplasms | UricA | |
| 155.1 | | | | | 23033 | 376 | 22657 | | | | | 32838 | 173 | 32665 | 0.6106 | 0.115 | Malignant neoplasm of liver, primary | neoplasms | UricA | |
| 157 | | | | | 23046 | 389 | 22657 | | | | | 32990 | 325 | 32665 | 0.2834 | 0.2153 | Pancreatic cancer | neoplasms | UricA | |
| 158 | | | | | 22882 | 225 | 22657 | | | | | 32904 | 239 | 32665 | | | Neoplasm of unspecified nature of digestive system | neoplasms | UricA | |
| 159 | 0.01933 | 0.1112 | 1.02 | 0.862 | 23177 | 520 | 22657 | 0.1577 | 0.117 | 1.171 | 0.178 | 33167 | 502 | 32665 | 0.2253 | 0.1914 | Malignant neoplasm of other and ill-defined sites within the digestive organs and peritoneum | neoplasms | UricA | |
| 159.2 | | | | | 22805 | 148 | 22657 | | | | | 32791 | 126 | 32665 | | | Malignant neoplasm of small intestine, including duodenum | neoplasms | UricA | |
| 159.3 | | | | | 22758 | 101 | 22657 | | | | | 32760 | 95 | 32665 | | | Malignant neoplasm of gallbladder and extrahepatic bile ducts | neoplasms | UricA | |
| 159.4 | | | | | 22743 | 86 | 22657 | | | | | 32783 | 118 | 32665 | | | Malignant neoplasm of retroperitoneum and peritoneum | neoplasms | UricA | |
| 164 | | | | | 25778 | 71 | 25707 | | | | | 35298 | 55 | 35243 | | | Cancer of intrathoracic organs | neoplasms | UricA | |
| 165 | 0.1721 | 0.06461 | 1.188 | 0.007719 | 27233 | 1526 | 25707 | 0.3724 | 0.07441 | 1.451 | 5.61e-07 | 36502 | 1259 | 35243 | 0.2094 | 0.1204 | Cancer within the respiratory system | neoplasms | UricA | |
| 165.1 | 0.1777 | 0.06507 | 1.195 | 0.006304 | 27209 | 1502 | 25707 | 0.379 | 0.0746 | 1.461 | 3.78e-07 | 36492 | 1249 | 35243 | 0.2006 | 0.1233 | Cancer of bronchus; lung | neoplasms | UricA | |
| 170 | -0.03557 | 0.1129 | 0.9651 | 0.7528 | 27372 | 508 | 26864 | | | | | 36581 | 434 | 36147 | 0.726 | 0.05926 | Cancer of bone and connective tissue | neoplasms | UricA | |
| 170.1 | | | | | 27106 | 242 | 26864 | | | | | 36353 | 206 | 36147 | | | Bone cancer | neoplasms | UricA | |
| 170.2 | | | | | 27193 | 329 | 26864 | | | | | 36433 | 286 | 36147 | 0.6612 | 0.09093 | Cancer of connective tissue | neoplasms | UricA | |
| 172 | 0.5178 | 0.04224 | 1.678 | 1.52e-34 | 25415 | 3454 | 21961 | 0.3238 | 0.04878 | 1.382 | 3.20e-11 | 33247 | 2675 | 30572 | 0.01048 | -0.1611 | Skin cancer | neoplasms | UricA | |
| 172.1 | 0.4328 | 0.07421 | 1.542 | 5.46e-09 | 22929 | 968 | 21961 | 0.3749 | 0.09105 | 1.455 | 3.83e-05 | 31295 | 723 | 30572 | 0.3926 | -0.09779 | Melanomas of skin, dx or hx | neoplasms | UricA | |
| 172.11 | 0.3887 | 0.0821 | 1.475 | 2.20e-06 | 22766 | 805 | 21961 | 0.4544 | 0.1023 | 1.575 | 8.92e-06 | 31138 | 566 | 30572 | 0.9719 | 0.004497 | Melanomas of skin | neoplasms | UricA | |
| 172.2 | 0.5322 | 0.046 | 1.703 | 5.89e-31 | 24804 | 2843 | 21961 | 0.2945 | 0.0537 | 1.342 | 4.16e-08 | 32709 | 2137 | 30572 | 0.006617 | -0.1874 | Other non-epithelial cancer of skin | neoplasms | UricA | |
| 172.21 | 0.4415 | 0.07262 | 1.555 | 1.21e-09 | 22933 | 972 | 21961 | 0.1636 | 0.08821 | 1.178 | 0.06362 | 31280 | 708 | 30572 | 0.008281 | -0.294 | Basal cell carcinoma | neoplasms | UricA | |
| 172.22 | 0.7191 | 0.07634 | 2.052 | 4.57e-21 | 22797 | 836 | 21961 | | | | | 30972 | 400 | 30572 | 0.07134 | -0.2364 | Squamous cell carcinoma | neoplasms | UricA | |
| 172.3 | | | | | 22359 | 398 | 21961 | | | | | 30807 | 235 | 30572 | 0.006303 | -0.4744 | Carcinoma in situ of skin | neoplasms | UricA | |
| 173 | 0.5376 | 0.0524 | 1.712 | 1.09e-24 | 23987 | 2026 | 21961 | 0.2396 | 0.05143 | 1.271 | 3.18e-06 | 32915 | 2343 | 30572 | 2.28e-06 | -0.339 | Neoplasm of uncertain behavior of skin | neoplasms | UricA | |
| 174 | | | | | 27350 | 45 | 27305 | -0.05008 | 0.04992 | 0.9512 | 0.3158 | 31827 | 3093 | 28734 | 0.7018 | -0.1275 | Breast cancer | neoplasms | UricA | |
| 174.1 | | | | | 7 | | | -0.04801 | 0.05068 | 0.9531 | 0.3435 | 31729 | 2998 | 28731 | 0.9991 | 0.2914 | Breast cancer [female] | neoplasms | UricA | |
| 174.11 | | | | | 7 | | | -0.04545 | 0.05088 | 0.9556 | 0.3717 | 31698 | 2967 | 28731 | 0.9991 | 0.293 | Malignant neoplasm of female breast | neoplasms | UricA | |
| 174.2 | | | | | 27313 | 15 | 27298 | | | | | 3 | | | | | Breast cancer [male] | neoplasms | UricA | |
| 174.3 | | | | | 27310 | 5 | 27305 | | | | | 28889 | 155 | 28734 | | | Neoplasm of uncertain behavior of breast | neoplasms | UricA | |

| phenotype | male_beta | male_SE | male_OR | male_p | male_n_total | male_n_cases | male_n_controls | female_beta | female_SE | female_OR | female_p | female_n_total | female_n_cases | female_n_controls | interaction_p | interaction_beta | description | group | lab | label |
|---|---|---|---|---|---|---|---|---|---|---|---|---|---|---|---|---|---|---|---|---|
| 175 | | | | | 7 | | | -0.1401 | 0.07814 | 0.8692 | 0.07287 | 27358 | 1237 | 26121 | 0.9996 | 0.1455 | Acquired absence of breast | neoplasms | UricA | |
| 180 | | | | | 7 | | | 0.6243 | 0.07779 | 1.867 | 1.01e-15 | 28736 | 897 | 27839 | 0.9991 | 0.508 | Cervical cancer and dysplasia | neoplasms | UricA | |
| 180.1 | | | | | 7 | | | | | | | 28134 | 295 | 27839 | | | Cervical cancer | neoplasms | UricA | |
| 180.3 | | | | | 7 | | | 0.796 | 0.09142 | 2.217 | 3.14e-18 | 28457 | 618 | 27839 | 0.999 | 0.5432 | Cervical intraepithelial neoplasia [CIN] [Cervical dysplasia] | neoplasms | UricA | |
| 182 | | | | | 7 | | | -0.05281 | 0.1157 | 0.9486 | 0.6481 | 29788 | 534 | 29254 | 0.9995 | 0.2937 | Malignant neoplasm of uterus | neoplasms | UricA | |
| 184 | | | | | 7 | | | 0.04799 | 0.09581 | 1.049 | 0.6164 | 30877 | 771 | 30106 | 0.9995 | 0.2578 | Cancer of other female genital organs | neoplasms | UricA | |
| 184.1 | | | | | 7 | | | -0.04125 | 0.1144 | 0.9596 | 0.7184 | 30679 | 573 | 30106 | 0.9997 | 0.1871 | Malignant neoplasm of ovary and other uterine adnexa | neoplasms | UricA | |
| 184.11 | | | | | 7 | | | | | | | 30568 | 462 | 30106 | | | Malignant neoplasm of ovary | neoplasms | UricA | |
| 184.2 | | | | | 7 | | | | | | | 30333 | 227 | 30106 | | | Cancer of other female genital organs (excluding uterus and ovary) | neoplasms | UricA | |
| 185 | -0.04304 | 0.06045 | 0.9579 | 0.4765 | 21441 | 2053 | 19388 | | | | | 4 | | | | | Cancer of prostate | neoplasms | UricA | |
| 187 | | | | | 25045 | 181 | 24864 | | | | | 4 | | | | | Cancer of other male genital organs | neoplasms | UricA | |
| 187.1 | | | | | 24907 | 43 | 24864 | | | | | 4 | | | | | Malignant neoplasm of unspecified male genital organ | neoplasms | UricA | |
| 187.2 | | | | | 24988 | 124 | 24864 | | | | | 4 | | | | | Malignant neoplasm of testis | neoplasms | UricA | |
| 187.8 | | | | | 24880 | 16 | 24864 | | | | | 4 | | | | | Neoplasm of uncertain behavior of male genital organs | neoplasms | UricA | |
| 189 | 0.1444 | 0.06615 | 1.155 | 0.02907 | 27414 | 1343 | 26071 | 0.2259 | 0.1046 | 1.253 | 0.03076 | 36667 | 553 | 36114 | 0.3048 | 0.1232 | Cancer of urinary organs (incl. kidney and bladder) | neoplasms | UricA | |
| 189.1 | 0.3334 | 0.08542 | 1.396 | 9.49e-05 | 26801 | 730 | 26071 | | | | | 36472 | 358 | 36114 | 0.6581 | 0.06566 | Cancer of kidney and renal pelvis | neoplasms | UricA | |
| 189.11 | 0.3724 | 0.09102 | 1.451 | 4.29e-05 | 26706 | 635 | 26071 | | | | | 36422 | 308 | 36114 | 0.4991 | 0.1065 | Malignant neoplasm of kidney, except pelvis | neoplasms | UricA | |
| 189.12 | | | | | 26217 | 146 | 26071 | | | | | 36178 | 64 | 36114 | | | Malignant neoplasm of renal pelvis | neoplasms | UricA | |
| 189.2 | -0.05013 | 0.09217 | 0.9511 | 0.5865 | 26790 | 719 | 26071 | | | | | 36340 | 226 | 36114 | 0.6429 | 0.0869 | Cancer of bladder | neoplasms | UricA | |
| 189.21 | -0.08391 | 0.09358 | 0.9195 | 0.3699 | 26780 | 709 | 26071 | | | | | 36336 | 222 | 36114 | 0.6421 | 0.08865 | Malignant neoplasm of bladder | neoplasms | UricA | |
| 189.4 | | | | | 26367 | 296 | 26071 | | | | | 36227 | 113 | 36114 | | | Malignant neoplasm of other urinary organs | neoplasms | UricA | |
| 190 | | | | | 26618 | 60 | 26558 | | | | | 35577 | 58 | 35519 | | | Cancer of eye | neoplasms | UricA | |
| 191 | | | | | 27023 | 465 | 26558 | | | | | 35973 | 454 | 35519 | 0.8885 | -0.02334 | Manlignant and unknown neoplasms of brain and nervous system | neoplasms | UricA | |
| 191.1 | | | | | 26929 | 371 | 26558 | | | | | 35854 | 335 | 35519 | 0.9689 | 0.007523 | Cancer of brain and nervous system | neoplasms | UricA | |
| 191.11 | | | | | 26893 | 335 | 26558 | | | | | 35801 | 282 | 35519 | 0.9613 | 0.0103 | Cancer of brain | neoplasms | UricA | |
| 193 | | | | | 27161 | 215 | 26946 | | | | | 35853 | 482 | 35371 | 0.7142 | 0.07275 | Thyroid cancer | neoplasms | UricA | |
| 194 | | | | | 26993 | 47 | 26946 | | | | | 35425 | 54 | 35371 | | | Cancer of other endocrine glands | neoplasms | UricA | |
| 195 | 0.475 | 0.05161 | 1.608 | 3.45e-20 | 20735 | 2450 | 18285 | 0.5768 | 0.05873 | 1.78 | 9.19e-23 | 28047 | 1903 | 26144 | 0.09468 | 0.127 | Cancer, suspected or other | neoplasms | UricA | |
| 195.1 | 0.4124 | 0.07037 | 1.51 | 4.60e-09 | 19494 | 1209 | 18285 | 0.4429 | 0.08247 | 1.557 | 7.85e-08 | 27102 | 958 | 26144 | 0.6473 | 0.04836 | Malignant neoplasm, other | neoplasms | UricA | |
| 195.3 | 0.09626 | 0.08892 | 1.101 | 0.279 | 19155 | 870 | 18285 | | | | | 26453 | 309 | 26144 | 0.2661 | -0.2112 | Malignant neoplasm of head, face, and neck | neoplasms | UricA | |
| 196 | 0.3897 | 0.06703 | 1.476 | 6.12e-09 | 19459 | 1174 | 18285 | 0.4883 | 0.06291 | 1.63 | 8.38e-15 | 27555 | 1411 | 26144 | 0.2906 | 0.09441 | Radiotherapy | neoplasms | UricA | |
| 197 | 0.6914 | 0.03853 | 1.996 | 5.31e-72 | 22676 | 4391 | 18285 | 0.5803 | 0.03795 | 1.787 | 8.58e-53 | 30992 | 4848 | 26144 | 0.5939 | -0.02805 | Chemotherapy | neoplasms | UricA | |

| phenotype | male_beta | male_SE | male_OR | male_p | male_n_total | male_n_cases | male_n_controls | female_beta | female_SE | female_OR | female_p | female_n_total | female_n_cases | female_n_controls | interaction_p | interaction_beta | description | group | lab | label |
|---|---|---|---|---|---|---|---|---|---|---|---|---|---|---|---|---|---|---|---|---|
| 198 | 0.381 | 0.04556 | 1.464 | 6.09e-17 | 22039 | 3754 | 18285 | 0.3912 | 0.04937 | 1.479 | 2.31e-15 | 29550 | 3406 | 26144 | 0.5827 | 0.03602 | Secondary malignant neoplasm | neoplasms | UricA | |
| 198.1 | 0.4215 | 0.05728 | 1.524 | 1.86e-13 | 20292 | 2007 | 18285 | 0.3811 | 0.06549 | 1.464 | 5.90e-09 | 27825 | 1681 | 26144 | 0.8607 | 0.01492 | Secondary malignancy of lymph nodes | neoplasms | UricA | |
| 198.2 | 0.7529 | 0.06652 | 2.123 | 1.07e-29 | 19626 | 1341 | 18285 | 0.9257 | 0.07294 | 2.524 | 6.58e-37 | 27296 | 1152 | 26144 | 0.0725 | 0.1727 | Secondary malignancy of respiratory organs | neoplasms | UricA | |
| 198.3 | 0.7354 | 0.09929 | 2.086 | 1.30e-13 | 18851 | 566 | 18285 | 0.6682 | 0.1036 | 1.951 | 1.13e-10 | 26726 | 582 | 26144 | 0.8881 | 0.0197 | Secondary malignant neoplasm of digestive systems | neoplasms | UricA | |
| 198.4 | 0.882 | 0.07369 | 2.416 | 5.21e-33 | 19347 | 1062 | 18285 | 0.7848 | 0.08097 | 2.192 | 3.27e-22 | 27121 | 977 | 26144 | 0.7435 | -0.03489 | Secondary malignant neoplasm of liver | neoplasms | UricA | |
| 198.5 | 0.9055 | 0.1011 | 2.473 | 3.44e-19 | 18799 | 514 | 18285 | 0.8456 | 0.1009 | 2.329 | 5.35e-17 | 26737 | 593 | 26144 | 0.7028 | -0.05321 | Secondary malignancy of brain/spine | neoplasms | UricA | |
| 198.6 | 0.6222 | 0.07652 | 1.863 | 4.28e-16 | 19284 | 999 | 18285 | 0.8663 | 0.08452 | 2.378 | 1.19e-24 | 26962 | 818 | 26144 | 0.04129 | 0.2267 | Secondary malignancy of bone | neoplasms | UricA | |
| 198.7 | | | | | 18508 | 223 | 18285 | | | | | 26369 | 225 | 26144 | | | Secondary malignant neoplasm of skin | neoplasms | UricA | |
| 199 | 0.4102 | 0.06434 | 1.507 | 1.83e-10 | 19599 | 1314 | 18285 | 0.3043 | 0.06326 | 1.356 | 1.51e-06 | 27705 | 1561 | 26144 | 0.2946 | -0.09154 | Neoplasm of uncertain behavior | neoplasms | UricA | |
| 199.4 | | | | | 18315 | 30 | 18285 | | | | | 26186 | 42 | 26144 | | | Neurofibromatosis | neoplasms | UricA | |
| 200 | 1.856 | 0.08714 | 6.395 | 1.28e-100 | 26538 | 642 | 25896 | 1.593 | 0.08836 | 4.92 | 1.11e-72 | 35761 | 577 | 35184 | 0.07896 | -0.2083 | Myeloproliferative disease | neoplasms | UricA | |
| 200.1 | | | | | 21974 | 134 | 21840 | | | | | 30355 | 101 | 30254 | | | Polycythemia vera | neoplasms | UricA | |
| 201 | | | | | 25974 | 78 | 25896 | | | | | 35242 | 58 | 35184 | | | Hodgkin's disease | neoplasms | UricA | |
| 202 | | | | | 26283 | 387 | 25896 | | | | | 35476 | 292 | 35184 | 0.5462 | -0.09857 | Cancer of other lymphoid, histiocytic tissue | neoplasms | UricA | |
| 202.2 | | | | | 26223 | 327 | 25896 | | | | | 35425 | 241 | 35184 | 0.4022 | -0.1552 | Non-Hodgkins lymphoma | neoplasms | UricA | |
| 202.21 | | | | | 25957 | 61 | 25896 | | | | | 35235 | 51 | 35184 | | | Nodular lymphoma | neoplasms | UricA | |
| 202.22 | | | | | 25981 | 85 | 25896 | | | | | 35237 | 53 | 35184 | | | Reticulosarcoma | neoplasms | UricA | |
| 202.23 | | | | | 25947 | 51 | 25896 | | | | | 35208 | 24 | 35184 | | | Lymphosarcoma | neoplasms | UricA | |
| 202.24 | | | | | 25960 | 64 | 25896 | | | | | 35217 | 33 | 35184 | | | Large cell lymphoma | neoplasms | UricA | |
| 204 | 2.571 | 0.0917 | 13.08 | 5.62e-173 | 26591 | 695 | 25896 | 2.596 | 0.09961 | 13.41 | 9.86e-150 | 35705 | 521 | 35184 | 0.9391 | 0.009948 | Leukemia | neoplasms | UricA | |
| 204.1 | | | | | 26113 | 217 | 25896 | | | | | 35327 | 143 | 35184 | | | Lymphoid leukemia | neoplasms | UricA | |
| 204.11 | | | | | 25996 | 100 | 25896 | | | | | 35260 | 76 | 35184 | | | Lymphoid leukemia, acute | neoplasms | UricA | |
| 204.12 | | | | | 26033 | 137 | 25896 | | | | | 35270 | 86 | 35184 | | | Lymphoid leukemia, chronic | neoplasms | UricA | |
| 204.2 | | | | | 26331 | 435 | 25896 | | | | | 35537 | 353 | 35184 | 0.9755 | -0.005401 | Myeloid leukemia | neoplasms | UricA | |
| 204.21 | | | | | 26263 | 367 | 25896 | | | | | 35482 | 298 | 35184 | 0.5078 | -0.136 | Myeloid leukemia, acute | neoplasms | UricA | |
| 204.22 | | | | | 26069 | 173 | 25896 | | | | | 35327 | 143 | 35184 | | | Myeloid leukemia, chronic | neoplasms | UricA | |
| 204.3 | | | | | 25933 | 37 | 25896 | | | | | 35214 | 30 | 35184 | | | Monocytic leukemia | neoplasms | UricA | |
| 204.4 | | | | | 26060 | 164 | 25896 | | | | | 35291 | 107 | 35184 | | | Multiple myeloma | neoplasms | UricA | |
| 208 | 0.4845 | 0.0459 | 1.623 | 4.73e-26 | 24481 | 2858 | 21623 | 0.1573 | 0.04995 | 1.17 | 0.001636 | 33540 | 2573 | 30967 | 9.53e-07 | -0.325 | Benign neoplasm of colon | neoplasms | UricA | |
| 209 | | | | | 27760 | 86 | 27674 | | | | | 36911 | 107 | 36804 | | | Neuroendocrine tumors | neoplasms | UricA | |
| 210 | | | | | 26386 | 107 | 26279 | | | | | 36167 | 130 | 36037 | | | Benign neoplasm of lip, oral cavity, and pharynx | neoplasms | UricA | |
| 211 | | | | | 24095 | 236 | 23859 | | | | | 33779 | 379 | 33400 | 0.1905 | 0.2377 | Benign neoplasm of other parts of digestive system | neoplasms | UricA | |
| 212 | | | | | 27637 | 101 | 27536 | | | | | 36801 | 112 | 36689 | | | Benign neoplasm of respiratory and intrathoracic organs | neoplasms | UricA | |
| 213 | | | | | 21553 | 68 | 21485 | | | | | 26105 | 131 | 25974 | | | Benign neoplasm of bone and articular cartilage | neoplasms | UricA | |
| 214 | | | | | 27025 | 277 | 26748 | | | | | 35990 | 359 | 35631 | 0.681 | -0.06985 | Lipoma | neoplasms | UricA | |
| 214.1 | | | | | 26919 | 171 | 26748 | | | | | 35867 | 236 | 35631 | | | Lipoma of skin and subcutaneous tissue | neoplasms | UricA | |

| phenotype | male_beta | male_SE | male_OR | male_p | male_n_total | male_n_cases | male_n_controls | female_beta | female_SE | female_OR | female_p | female_n_total | female_n_cases | female_n_controls | interaction_p | interaction_beta | description | group | lab | label |
|---|---|---|---|---|---|---|---|---|---|---|---|---|---|---|---|---|---|---|---|---|
| 215 | | | | | 26820 | 72 | 26748 | | | | | 35793 | 162 | 35631 | | | Other benign neoplasm of connective and other soft tissue | neoplasms | UricA | |
| 216 | 0.4723 | 0.05706 | 1.604 | 1.26e-16 | 25936 | 1607 | 24329 | 0.227 | 0.04901 | 1.255 | 3.60e-06 | 34007 | 2621 | 31386 | 4.21e-05 | -0.2994 | Benign neoplasm of skin | neoplasms | UricA | |
| 216.1 | | | | | 24408 | 79 | 24329 | | | | | 31484 | 98 | 31386 | | | Screening for malignant neoplasms of the skin | neoplasms | UricA | |
| 217 | | | | | 24477 | 148 | 24329 | | | | | 31698 | 312 | 31386 | | | Vascular hamartomas and non-neoplastic nevi | neoplasms | UricA | |
| 217.1 | | | | | 24444 | 115 | 24329 | | | | | 31637 | 251 | 31386 | | | Nevus, non-neoplastic | neoplasms | UricA | |
| 218 | | | | | 7 | | | 0.07002 | 0.08477 | 1.073 | 0.4088 | 31913 | 869 | 31044 | 0.9996 | 0.2199 | Benign neoplasm of uterus | neoplasms | UricA | |
| 218.1 | | | | | 7 | | | 0.09078 | 0.08648 | 1.095 | 0.2939 | 31868 | 824 | 31044 | 0.9996 | 0.2378 | Uterine leiomyoma | neoplasms | UricA | |
| 218.2 | | | | | 7 | | | | | | | 31064 | 20 | 31044 | | | Other benign neoplasm of uterus | neoplasms | UricA | |
| 220 | | | | | 7 | | | | | | | 28308 | 209 | 28099 | | | Benign neoplasm of ovary | neoplasms | UricA | |
| 221 | | | | | 7 | | | | | | | 28086 | 59 | 28027 | | | Benign neoplasm of other female genital organs | neoplasms | UricA | |
| 222 | | | | | 24885 | 21 | 24864 | | | | | 4 | | | | | Benign neoplasm of male genital organs | neoplasms | UricA | |
| 223 | | | | | 21509 | 24 | 21485 | | | | | 26009 | 35 | 25974 | | | Benign neoplasm of kidney and other urinary organs | neoplasms | UricA | |
| 224 | | | | | 26668 | 110 | 26558 | | | | | 35659 | 140 | 35519 | | | Benign neoplasm of eye | neoplasms | UricA | |
| 224.1 | | | | | 26643 | 85 | 26558 | | | | | 35621 | 102 | 35519 | | | Benign neoplasm of eye, uveal | neoplasms | UricA | |
| 225 | | | | | 26749 | 191 | 26558 | | | | | 35909 | 390 | 35519 | 0.1989 | 0.2959 | Benign neoplasm of brain and other parts of nervous system | neoplasms | UricA | |
| 225.1 | | | | | 26730 | 172 | 26558 | | | | | 35878 | 359 | 35519 | 0.2918 | 0.2516 | Benign neoplasm of brain, cranial nerves, meninges | neoplasms | UricA | |
| 225.2 | | | | | 26574 | 16 | 26558 | | | | | 35551 | 32 | 35519 | | | Benign neoplasm of spinal cord, meninges | neoplasms | UricA | |
| 226 | | | | | 26971 | 25 | 26946 | | | | | 35500 | 129 | 35371 | | | Benign neoplasm of thyroid glands | neoplasms | UricA | |
| 227 | | | | | 27177 | 231 | 26946 | | | | | 35717 | 346 | 35371 | 0.2869 | -0.202 | Benign neoplasm of other endocrine glands and related structures | neoplasms | UricA | |
| 227.1 | | | | | 27032 | 86 | 26946 | | | | | 35495 | 124 | 35371 | | | Benign neoplasm of adrenal gland | neoplasms | UricA | |
| 227.2 | | | | | 26972 | 26 | 26946 | | | | | 35433 | 62 | 35371 | | | Benign neoplasm of parathyroid gland | neoplasms | UricA | |
| 227.3 | | | | | 27062 | 116 | 26946 | | | | | 35525 | 154 | 35371 | | | Benign neoplasm of pituitary gland and craniopharyngeal duct (pouch) | neoplasms | UricA | |
| 228 | 0.2284 | 0.0954 | 1.257 | 0.01667 | 26936 | 519 | 26417 | 0.03074 | 0.08612 | 1.031 | 0.7212 | 35731 | 759 | 34972 | 0.04626 | -0.2479 | Hemangioma and lymphangioma, any site | neoplasms | UricA | |
| 228.1 | | | | | 26853 | 436 | 26417 | -0.05087 | 0.09555 | 0.9504 | 0.5945 | 35591 | 619 | 34972 | 0.02291 | -0.3104 | Hemangioma of skin and subcutaneous tissue | neoplasms | UricA | |
| 229 | | | | | 27551 | 83 | 27468 | | | | | 36445 | 69 | 36376 | | | Benign neoplasm of unspecified sites | neoplasms | UricA | |
| 229.1 | | | | | 27477 | 9 | 27468 | | | | | 36386 | 10 | 36376 | | | Benign neoplasm of lymph nodes | neoplasms | UricA | |
| 230 | | | | | 27785 | 15 | 27770 | | | | | 36952 | 4 | 36948 | | | Kaposi's sarcoma | neoplasms | UricA | |
| 240 | | | | | 23255 | 80 | 23175 | | | | | 26401 | 346 | 26055 | | | Simple and unspecified goiter | endocrine/metabolic | UricA | |
| 241 | 0.5314 | 0.0966 | 1.701 | 3.78e-08 | 23679 | 504 | 23175 | 0.2463 | 0.05703 | 1.279 | 1.57e-05 | 28029 | 1974 | 26055 | 0.01388 | -0.2671 | Nontoxic nodular goiter | endocrine/metabolic | UricA | |
| 241.1 | | | | | 23492 | 317 | 23175 | 0.3609 | 0.07657 | 1.435 | 2.43e-06 | 27019 | 964 | 26055 | 0.6887 | 0.0563 | Nontoxic uninodular goiter | endocrine/metabolic | UricA | |
| 241.2 | | | | | 23428 | 253 | 23175 | 0.2963 | 0.06844 | 1.345 | 1.50e-05 | 27333 | 1278 | 26055 | 0.002702 | -0.4329 | Nontoxic multinodular goiter | endocrine/metabolic | UricA | |
| 242 | | | | | 23461 | 286 | 23175 | 0.2447 | 0.08739 | 1.277 | 0.005108 | 26848 | 793 | 26055 | 0.07207 | -0.2702 | Thyrotoxicosis with or without goiter | endocrine/metabolic | UricA | |

| phenotype | male_beta | male_SE | male_OR | male_p | male_n_total | male_n_cases | male_n_controls | female_beta | female_SE | female_OR | female_p | female_n_total | female_n_cases | female_n_controls | interaction_p | interaction_beta | description | group | lab | label |
|---|---|---|---|---|---|---|---|---|---|---|---|---|---|---|---|---|---|---|---|---|
| 242.1 | | | | | 23301 | 126 | 23175 | | | | | 26434 | 379 | 26055 | 0.1964 | -0.3069 | Graves' disease | endocrine/metabolic | UricA | |
| 242.2 | | | | | 23196 | 21 | 23175 | | | | | 26178 | 123 | 26055 | | | Toxic multinodular goiter | endocrine/metabolic | UricA | |
| 242.3 | | | | | 23231 | 56 | 23175 | | | | | 26122 | 67 | 26055 | | | Exophthalmos | endocrine/metabolic | UricA | |
| 242.31 | | | | | 23186 | 11 | 23175 | | | | | 26071 | 16 | 26055 | | | Thyrotoxic exophthalmos | endocrine/metabolic | UricA | |
| 244 | 0.5491 | 0.04625 | 1.732 | 1.65e-32 | 25712 | 2537 | 23175 | 0.3487 | 0.03496 | 1.417 | 1.98e-23 | 32658 | 6603 | 26055 | 0.009855 | -0.146 | Hypothyroidism | endocrine/metabolic | UricA | |
| 244.1 | 0.4657 | 0.09675 | 1.593 | 1.49e-06 | 23694 | 519 | 23175 | 0.2414 | 0.06802 | 1.273 | 3.86e-04 | 27402 | 1347 | 26055 | 0.2336 | -0.1368 | Secondary hypothyroidism | endocrine/metabolic | UricA | |
| 244.2 | | | | | 23522 | 347 | 23175 | 0.4986 | 0.08026 | 1.646 | 5.23e-10 | 26876 | 821 | 26055 | 0.563 | 0.07929 | Acquired hypothyroidism | endocrine/metabolic | UricA | |
| 244.3 | | | | | 23175 | | | | | | | 26060 | 5 | 26055 | | | Iodine hypothyroidism | endocrine/metabolic | UricA | |
| 244.4 | 0.5718 | 0.04842 | 1.771 | 3.43e-32 | 25441 | 2266 | 23175 | 0.3876 | 0.0359 | 1.473 | 3.58e-27 | 32096 | 6041 | 26055 | 0.03183 | -0.1263 | Hypothyroidism NOS | endocrine/metabolic | UricA | |
| 244.5 | | | | | 23199 | 24 | 23175 | | | | | 26089 | 34 | 26055 | | | Congenital hypothyroidism | endocrine/metabolic | UricA | |
| 245 | | | | | 23436 | 261 | 23175 | 0.2903 | 0.08222 | 1.337 | 4.14e-04 | 26942 | 887 | 26055 | 0.07098 | -0.2748 | Thyroiditis | endocrine/metabolic | UricA | |
| 245.1 | | | | | 23196 | 21 | 23175 | | | | | 26097 | 42 | 26055 | | | Thyroiditis, acute and subacute | endocrine/metabolic | UricA | |
| 245.2 | | | | | 23326 | 151 | 23175 | 0.2101 | 0.09711 | 1.234 | 0.03047 | 26707 | 652 | 26055 | 0.1675 | -0.2703 | Chronic thyroiditis | endocrine/metabolic | UricA | |
| 245.21 | | | | | 23324 | 149 | 23175 | 0.2243 | 0.09725 | 1.251 | 0.02107 | 26701 | 646 | 26055 | 0.2022 | -0.2513 | Chronic lymphocytic thyroiditis | endocrine/metabolic | UricA | |
| 246 | | | | | 23388 | 213 | 23175 | | | | | 26539 | 484 | 26055 | 1.66e-04 | -0.6491 | Other disorders of thyroid | endocrine/metabolic | UricA | |
| 246.2 | | | | | 23182 | 7 | 23175 | | | | | 26065 | 10 | 26055 | | | Thyroid cyst | endocrine/metabolic | UricA | |
| 246.7 | | | | | 23294 | 119 | 23175 | | | | | 26304 | 249 | 26055 | | | Abnormal results of function study of thyroid | endocrine/metabolic | UricA | |
| 249 | | | | | 17130 | 307 | 16823 | | | | | 26073 | 265 | 25808 | 0.392 | 0.1461 | Secondary diabetes mellitus | endocrine/metabolic | UricA | |
| 250 | 0.6847 | 0.03569 | 1.983 | 4.92e-82 | 22885 | 6062 | 16823 | 0.7341 | 0.03565 | 2.084 | 3.48e-94 | 31483 | 5675 | 25808 | 0.5888 | 0.02675 | Diabetes mellitus | endocrine/metabolic | UricA | |
| 250.1 | 0.92 | 0.06631 | 2.509 | 8.99e-44 | 18010 | 1187 | 16823 | 1.014 | 0.0645 | 2.758 | 9.99e-56 | 27021 | 1213 | 25808 | 0.7354 | 0.03025 | Type 1 diabetes | endocrine/metabolic | UricA | |
| 250.11 | | | | | 16945 | 122 | 16823 | | | | | 25971 | 163 | 25808 | | | Type 1 diabetes with ketoacidosis | endocrine/metabolic | UricA | |
| 250.12 | | | | | 17020 | 197 | 16823 | | | | | 25984 | 176 | 25808 | | | Type 1 diabetes with renal manifestations | endocrine/metabolic | UricA | |
| 250.13 | | | | | 16957 | 134 | 16823 | | | | | 25957 | 149 | 25808 | | | Type 1 diabetes with ophthalmic manifestations | endocrine/metabolic | UricA | |
| 250.14 | | | | | 17123 | 300 | 16823 | | | | | 26082 | 274 | 25808 | 0.8299 | -0.03667 | Type 1 diabetes with neurological manifestations | endocrine/metabolic | UricA | |
| 250.15 | | | | | 16873 | 50 | 16823 | | | | | 25855 | 47 | 25808 | | | Diabetes type 1 with peripheral circulatory disorders | endocrine/metabolic | UricA | |
| 250.2 | 0.6946 | 0.03613 | 2.003 | 2.39e-82 | 22677 | 5854 | 16823 | 0.7321 | 0.03634 | 2.079 | 2.90e-90 | 31244 | 5436 | 25808 | 0.7315 | 0.01725 | Type 2 diabetes | endocrine/metabolic | UricA | |
| 250.21 | | | | | 16879 | 56 | 16823 | | | | | 25884 | 76 | 25808 | | | Type 2 diabetes with ketoacidosis | endocrine/metabolic | UricA | |
| 250.22 | 1.515 | 0.06112 | 4.549 | 1.27e-135 | 18226 | 1403 | 16823 | 1.601 | 0.07178 | 4.956 | 3.81e-110 | 26727 | 919 | 25808 | 0.4476 | 0.06995 | Type 2 diabetes with renal manifestations | endocrine/metabolic | UricA | |
| 250.23 | | | | | 17271 | 448 | 16823 | | | | | 26204 | 396 | 25808 | 0.2874 | 0.152 | Type 2 diabetes with ophthalmic manifestations | endocrine/metabolic | UricA | |

| phenotype | male_beta | male_SE | male_OR | male_p | male_n_total | male_n_cases | male_n_controls | female_beta | female_SE | female_OR | female_p | female_n_total | female_n_cases | female_n_controls | interaction_p | interaction_beta | description | group | lab | label |
|---|---|---|---|---|---|---|---|---|---|---|---|---|---|---|---|---|---|---|---|---|
| 250.24 | 1.036 | 0.05833 | 2.817 | 1.66e-70 | 18366 | 1543 | 16823 | 0.9958 | 0.0633 | 2.707 | 9.29e-56 | 27084 | 1276 | 25808 | 0.596 | -0.04451 | Type 2 diabetes with neurological manifestations | endocrine/metabolic | UricA | |
| 250.25 | | | | | 17307 | 484 | 16823 | | | | | 26109 | 301 | 25808 | 0.5954 | -0.081 | Diabetes type 2 with peripheral circulatory disorders | endocrine/metabolic | UricA | |
| 250.3 | 0.9843 | 0.05228 | 2.676 | 4.64e-79 | 18803 | 1980 | 16823 | 0.982 | 0.05426 | 2.67 | 3.28e-73 | 27587 | 1779 | 25808 | 0.6536 | -0.03301 | Insulin pump user | endocrine/metabolic | UricA | |
| 250.4 | 0.5865 | 0.04165 | 1.798 | 5.09e-45 | 20734 | 3911 | 16823 | 0.645 | 0.0423 | 1.906 | 1.64e-52 | 29488 | 3680 | 25808 | 0.2016 | 0.07418 | Abnormal glucose | endocrine/metabolic | UricA | |
| 250.41 | 0.5041 | 0.09315 | 1.655 | 6.25e-08 | 17402 | 579 | 16823 | 0.5227 | 0.0897 | 1.687 | 5.63e-09 | 26465 | 657 | 25808 | 0.5674 | -0.07178 | Impaired fasting glucose | endocrine/metabolic | UricA | |
| 250.42 | 0.5848 | 0.04352 | 1.795 | 3.67e-41 | 20295 | 3472 | 16823 | 0.6634 | 0.04499 | 1.941 | 3.38e-49 | 28947 | 3139 | 25808 | 0.08917 | 0.1039 | Other abnormal glucose | endocrine/metabolic | UricA | |
| 250.5 | | | | | 16841 | 18 | 16823 | | | | | 25844 | 36 | 25808 | | | Glycosuria or Acetonuria | endocrine/metabolic | UricA | |
| 250.6 | 1.023 | 0.06466 | 2.782 | 2.08e-56 | 18032 | 1209 | 16823 | 1.122 | 0.07092 | 3.07 | 2.41e-56 | 26769 | 961 | 25808 | 0.3107 | 0.09478 | Polyneuropathy in diabetes | endocrine/metabolic | UricA | |
| 250.7 | | | | | 24804 | 450 | 24354 | | | | | 33128 | 398 | 32730 | 0.1076 | 0.2264 | Diabetic retinopathy | endocrine/metabolic | UricA | |
| 251 | | | | | 23451 | 170 | 23281 | | | | | 31202 | 167 | 31035 | | | Other disorders of pancreatic internal secretion | endocrine/metabolic | UricA | |
| 251.1 | | | | | 15397 | 246 | 15151 | | | | | 23345 | 400 | 22945 | 0.3183 | -0.1696 | Hypoglycemia | endocrine/metabolic | UricA | |
| 251.8 | | | | | 23287 | 6 | 23281 | | | | | 31042 | 7 | 31035 | | | Abnormality of secretion of glucagon or gastrin | endocrine/metabolic | UricA | |
| 252 | | | | | 23548 | 267 | 23281 | 0.6191 | 0.08205 | 1.857 | 4.51e-14 | 31785 | 750 | 31035 | 0.004442 | -0.4195 | Disorders of parathyroid gland | endocrine/metabolic | UricA | |
| 252.1 | | | | | 23519 | 238 | 23281 | 0.6814 | 0.08653 | 1.977 | 3.40e-15 | 31693 | 658 | 31035 | 0.007815 | -0.4149 | Hyperparathyroidism | endocrine/metabolic | UricA | |
| 252.2 | | | | | 23316 | 35 | 23281 | | | | | 31163 | 128 | 31035 | | | Hypoparathyroidism | endocrine/metabolic | UricA | |
| 253 | | | | | 23665 | 384 | 23281 | | | | | 31431 | 396 | 31035 | 0.06519 | 0.3027 | Disorders of the pituitary gland and its hypothalamic control | endocrine/metabolic | UricA | |
| 253.1 | | | | | 23343 | 62 | 23281 | | | | | 31171 | 136 | 31035 | | | Pituitary hyperfunction | endocrine/metabolic | UricA | |
| 253.11 | | | | | 23302 | 21 | 23281 | | | | | 31046 | 11 | 31035 | | | Acromegaly and gigantism | endocrine/metabolic | UricA | |
| 253.2 | | | | | 23400 | 119 | 23281 | | | | | 31125 | 90 | 31035 | | | Pituitary hypofunction | endocrine/metabolic | UricA | |
| 253.3 | | | | | 23305 | 24 | 23281 | | | | | 31086 | 51 | 31035 | | | Diabetes insipidus | endocrine/metabolic | UricA | |
| 253.4 | | | | | 23423 | 142 | 23281 | | | | | 31110 | 75 | 31035 | | | Anterior pituitary disorders | endocrine/metabolic | UricA | |
| 253.5 | | | | | 23359 | 78 | 23281 | | | | | 31085 | 50 | 31035 | | | Pituitary dwarfism | endocrine/metabolic | UricA | |
| 253.7 | | | | | 23345 | 64 | 23281 | | | | | 31102 | 67 | 31035 | | | Other disorders of neurohypophysis | endocrine/metabolic | UricA | |
| 254 | | | | | 23286 | 5 | 23281 | | | | | 31042 | 7 | 31035 | | | Diseases of thymus gland | endocrine/metabolic | UricA | |
| phenotype | male_beta | male_SE | male_OR | male_p | male_n_total | male_n_cases | male_n_controls | female_beta | female_SE | female_OR | female_p | female_n_total | female_n_cases | female_n_controls | interaction_p | interaction_beta | description | group | lab | label |
| 255 | 0.8759 | 0.08339 | 2.401 | 8.32e-26 | 23957 | 676 | 23281 | 0.7107 | 0.07722 | 2.035 | 3.46e-20 | 31897 | 862 | 31035 | 0.3567 | -0.1015 | Disorders of adrenal glands | endocrine/metabolic | UricA | |
| 255.1 | | | | | 23371 | 90 | 23281 | | | | | 31211 | 176 | 31035 | | | Adrenal hyperfunction | endocrine/metabolic | UricA | |
| 255.11 | | | | | 23313 | 32 | 23281 | | | | | 31129 | 94 | 31035 | | | Cushing's syndrome | endocrine/metabolic | UricA | |
| 255.12 | | | | | 23315 | 34 | 23281 | | | | | 31064 | 29 | 31035 | | | Hyperaldosteronism | endocrine/metabolic | UricA | |

| phenotype | male_beta | male_SE | male_OR | male_p | male_n_total | male_n_cases | male_n_controls | female_beta | female_SE | female_OR | female_p | female_n_total | female_n_cases | female_n_controls | interaction_p | interaction_beta | description | group | lab | label |
|---|---|---|---|---|---|---|---|---|---|---|---|---|---|---|---|---|---|---|---|---|
| 255.13 | | | | | 23297 | 16 | 23281 | | | | | 31065 | 30 | 31035 | | | Medulloadrenal hyperfunction | endocrine/metabolic | UricA | |
| 255.2 | | | | | 23716 | 435 | 23281 | 0.7614 | 0.09981 | 2.141 | 2.38e-14 | 31535 | 500 | 31035 | 0.8969 | 0.01813 | Adrenal hypofunction | endocrine/metabolic | UricA | |
| 255.21 | | | | | 23674 | 393 | 23281 | | | | | 31474 | 439 | 31035 | 0.946 | 0.01001 | Glucocorticoid deficiency | endocrine/metabolic | UricA | |
| 255.22 | | | | | 23296 | 15 | 23281 | | | | | 31067 | 32 | 31035 | | | Mineralocorticoid deficiency | endocrine/metabolic | UricA | |
| 255.3 | | | | | 23289 | 8 | 23281 | | | | | 31065 | 30 | 31035 | | | Adrenogenital disorders | endocrine/metabolic | UricA | |
| 256 | | | | | 4 | | | 0.6782 | 0.09063 | 1.97 | 7.27e-14 | 31698 | 666 | 31032 | 0.9995 | 0.2153 | Ovarian dysfunction | endocrine/metabolic | UricA | |
| 256.1 | | | | | 23283 | 2 | 23281 | | | | | 31037 | 2 | 31035 | | | Hyperestrogenism | endocrine/metabolic | UricA | |
| 256.4 | | | | | 4 | | | 0.6221 | 0.1039 | 1.863 | 2.14e-09 | 31569 | 537 | 31032 | 0.9999 | -0.03953 | Polycystic ovaries | endocrine/metabolic | UricA | |
| 257 | 0.6777 | 0.07018 | 1.969 | 4.62e-22 | 24271 | 994 | 23277 | | | | | 3 | | | | | Testicular dysfunction | endocrine/metabolic | UricA | |
| 257.1 | 0.6725 | 0.07073 | 1.959 | 1.94e-21 | 24255 | 978 | 23277 | | | | | 3 | | | | | Testicular hypofunction | endocrine/metabolic | UricA | |
| 258 | | | | | 23304 | 23 | 23281 | | | | | 31068 | 33 | 31035 | | | Iatrogenic endocrine disorders | endocrine/metabolic | UricA | |
| 258.1 | | | | | 4 | | | | | | | 31087 | 55 | 31032 | | | Postablative ovarian failure | endocrine/metabolic | UricA | |
| 259 | | | | | 23461 | 180 | 23281 | | | | | 31392 | 357 | 31035 | 0.9636 | 0.009024 | Other endocrine disorders | endocrine/metabolic | UricA | |
| 259.1 | | | | | 23352 | 71 | 23281 | | | | | 31134 | 99 | 31035 | | | Nonspecific abnormal results of other endocrine function study | endocrine/metabolic | UricA | |
| 259.2 | | | | | 23320 | 39 | 23281 | | | | | 31092 | 57 | 31035 | | | Carcinoid syndrome | endocrine/metabolic | UricA | |
| 259.3 | | | | | 23324 | 43 | 23281 | | | | | 31063 | 28 | 31035 | | | Delay in sexual development and puberty NEC | endocrine/metabolic | UricA | |
| 259.4 | | | | | 23291 | 10 | 23281 | | | | | 31071 | 36 | 31035 | | | Precocious sexual development and puberty NEC | endocrine/metabolic | UricA | |
| 259.8 | | | | | 23297 | 16 | 23281 | | | | | 31067 | 32 | 31035 | | | Polyglandular activity in multiple endocrine adenomatosis | endocrine/metabolic | UricA | |
| 260 | 0.6255 | 0.04622 | 1.869 | 1.01e-41 | 22122 | 2837 | 19285 | 0.6993 | 0.04944 | 2.012 | 1.99e-45 | 27318 | 2532 | 24786 | 0.01045 | 0.1689 | Protein-calorie malnutrition | endocrine/metabolic | UricA | |
| 260.1 | | | | | 19435 | 150 | 19285 | | | | | 24879 | 93 | 24786 | | | Cachexia | endocrine/metabolic | UricA | |
| 260.2 | 0.7698 | 0.06924 | 2.159 | 1.04e-28 | 20371 | 1086 | 19285 | 0.7898 | 0.0741 | 2.203 | 1.59e-26 | 25774 | 988 | 24786 | 0.362 | 0.09015 | severe protein-calorie malnutrition | endocrine/metabolic | UricA | |
| 260.21 | | | | | 19301 | 16 | 19285 | | | | | 24805 | 19 | 24786 | | | Kwashiorkor | endocrine/metabolic | UricA | |
| 260.22 | | | | | 19399 | 114 | 19285 | | | | | 24929 | 143 | 24786 | | | Nutritional marasmus | endocrine/metabolic | UricA | |
| 260.3 | 0.8369 | 0.08782 | 2.309 | 1.58e-21 | 19914 | 629 | 19285 | 0.9457 | 0.09278 | 2.575 | 2.14e-24 | 25366 | 580 | 24786 | 0.08133 | 0.2166 | Adult failure to thrive | endocrine/metabolic | UricA | |
| 260.6 | 0.6214 | 0.09533 | 1.862 | 7.11e-11 | 19875 | 590 | 19285 | | | | | 25233 | 447 | 24786 | 1.30e-06 | 0.6622 | Anorexia | endocrine/metabolic | UricA | |
| 260.7 | | | | | 19286 | 1 | 19285 | | | | | 24804 | 18 | 24786 | | | Polyphagia | endocrine/metabolic | UricA | |
| 261 | 1.001 | 0.04755 | 2.72 | 2.63e-98 | 21716 | 2431 | 19285 | 0.5289 | 0.03709 | 1.697 | 3.84e-46 | 30360 | 5574 | 24786 | 7.73e-09 | -0.3398 | Vitamin deficiency | endocrine/metabolic | UricA | |
| 261.1 | | | | | 19312 | 27 | 19285 | | | | | 24829 | 43 | 24786 | | | Vitamin A deficiency | endocrine/metabolic | UricA | |
| 261.2 | 0.7261 | 0.08319 | 2.067 | 2.59e-18 | 19973 | 688 | 19285 | 0.5943 | 0.05988 | 1.812 | 3.21e-23 | 26376 | 1590 | 24786 | 0.2318 | -0.1193 | Vitamin B-complex deficiencies | endocrine/metabolic | UricA | |

| phenotype | male_beta | male_SE | male_OR | male_p | male_n_total | male_n_cases | male_n_controls | female_beta | female_SE | female_OR | female_p | female_n_total | female_n_cases | female_n_controls | interaction_p | interaction_beta | description | group | lab | label |
|---|---|---|---|---|---|---|---|---|---|---|---|---|---|---|---|---|---|---|---|---|
| 261.3 | | | | | 19292 | 7 | 19285 | | | | | 24807 | 21 | 24786 | | | Vitamin C deficiencies | endocrine/metabolic | UricA | |
| 261.4 | 1.089 | 0.05245 | 2.973 | 8.04e-96 | 21216 | 1931 | 19285 | 0.5615 | 0.03943 | 1.753 | 5.19e-46 | 29414 | 4628 | 24786 | 1.09e-08 | -0.3644 | Vitamin D deficiency | endocrine/metabolic | UricA | |
| 261.41 | | | | | 19297 | 12 | 19285 | | | | | 24819 | 33 | 24786 | | | Rickets or osteomalacia | endocrine/metabolic | UricA | |
| 262 | | | | | 19493 | 208 | 19285 | | | | | 25217 | 431 | 24786 | 0.2143 | -0.2168 | Mineral deficiency NEC | endocrine/metabolic | UricA | |
| 263 | | | | | 19569 | 284 | 19285 | | | | | 25175 | 389 | 24786 | 0.6256 | 0.08091 | Other nutritional deficiency | endocrine/metabolic | UricA | |
| 264 | | | | | 19689 | 404 | 19285 | | | | | 25083 | 297 | 24786 | 0.007133 | -0.7446 | Lack of normal physiological development | endocrine/metabolic | UricA | |
| 264.1 | | | | | 19424 | 139 | 19285 | | | | | 24879 | 93 | 24786 | | | Short stature | endocrine/metabolic | UricA | |
| 264.2 | | | | | 19409 | 124 | 19285 | | | | | 24883 | 97 | 24786 | | | Failure to thrive (childhood) | endocrine/metabolic | UricA | |
| 264.3 | | | | | 19388 | 103 | 19285 | | | | | 24857 | 71 | 24786 | | | Delayed milestones | endocrine/metabolic | UricA | |
| 264.9 | | | | | 19465 | 180 | 19285 | | | | | 24925 | 139 | 24786 | | | Lack of normal physiological development, unspecified | endocrine/metabolic | UricA | |
| 269 | 1.474 | 0.07234 | 4.365 | 3.04e-92 | 26340 | 917 | 25423 | 1.581 | 0.07813 | 4.859 | 4.95e-91 | 35408 | 726 | 34682 | 0.254 | 0.1174 | Proteinuria | endocrine/metabolic | UricA | |
| 270 | | | | | 25887 | 464 | 25423 | 1.087 | 0.09452 | 2.964 | 1.36e-30 | 35187 | 505 | 34682 | 0.8018 | -0.03302 | Disorders of protein plasma/amino-acid transport and metabolism | endocrine/metabolic | UricA | |
| 270.1 | | | | | 25452 | 29 | 25423 | | | | | 34732 | 50 | 34682 | | | Disturbances of amino-acid transport | endocrine/metabolic | UricA | |
| 270.11 | | | | | 25431 | 8 | 25423 | | | | | 34711 | 29 | 34682 | | | Disturbances of sulphur-bearing amino-acid metabolism | endocrine/metabolic | UricA | |
| 270.12 | | | | | 25434 | 11 | 25423 | | | | | 34696 | 14 | 34682 | | | Phenylketonuria [PKU] | endocrine/metabolic | UricA | |
| 270.2 | | | | | 25465 | 42 | 25423 | | | | | 34719 | 37 | 34682 | | | Disorders of amino-acid metabolism | endocrine/metabolic | UricA | |
| 270.21 | | | | | 25457 | 34 | 25423 | | | | | 34703 | 21 | 34682 | | | Disorders of urea cycle metabolism | endocrine/metabolic | UricA | |
| 270.3 | | | | | 25814 | 391 | 25423 | | | | | 35096 | 414 | 34682 | 0.9124 | -0.01579 | Disorders of plasma protein metabolism | endocrine/metabolic | UricA | |
| 270.31 | | | | | 25425 | 2 | 25423 | | | | | 34689 | 7 | 34682 | | | Polyclonal hypergammaglobulinemia | endocrine/metabolic | UricA | |
| 270.32 | | | | | 25665 | 242 | 25423 | | | | | 34920 | 238 | 34682 | | | Paraproteinemia | endocrine/metabolic | UricA | |
| 270.33 | | | | | 25484 | 61 | 25423 | | | | | 34731 | 49 | 34682 | | | Amyloidosis | endocrine/metabolic | UricA | |
| 270.34 | | | | | 25480 | 57 | 25423 | | | | | 34726 | 44 | 34682 | | | Alpha-1-antitrypsin deficiency | endocrine/metabolic | UricA | |
| 270.35 | | | | | 25440 | 17 | 25423 | | | | | 34692 | 10 | 34682 | | | Macroglobulinemia | endocrine/metabolic | UricA | |
| 270.38 | | | | | 25509 | 86 | 25423 | | | | | 34792 | 110 | 34682 | | | Other specified disorders of plasma protein metabolism | endocrine/metabolic | UricA | |
| 271 | | | | | 27474 | 374 | 27100 | 0.1647 | 0.08866 | 1.179 | 0.06319 | 36489 | 676 | 35813 | 0.5502 | -0.08225 | Disorders of carbohydrate transport and metabolism | endocrine/metabolic | UricA | |
| 271.3 | | | | | 27455 | 355 | 27100 | 0.151 | 0.08972 | 1.163 | 0.09236 | 36474 | 661 | 35813 | 0.761 | -0.04286 | Intestinal disaccharidase deficiencies and disaccharide malabsorption | endocrine/metabolic | UricA | |
| 271.9 | | | | | 27119 | 19 | 27100 | | | | | 35828 | 15 | 35813 | | | Other disorders of carbohydrate transport and metabolism | endocrine/metabolic | UricA | |

| phenotype | male_beta | male_SE | male_OR | male_p | male_n_total | male_n_cases | male_n_controls | female_beta | female_SE | female_OR | female_p | female_n_total | female_n_cases | female_n_controls | interaction_p | interaction_beta | description | group | lab | label |
|---|---|---|---|---|---|---|---|---|---|---|---|---|---|---|---|---|---|---|---|---|
| 272 | 0.8784 | 0.03664 | 2.407 | 4.73e-127 | 25396 | 11129 | 14267 | 0.7183 | 0.03483 | 2.051 | 1.67e-94 | 34085 | 10701 | 23384 | 3.64e-04 | -0.1791 | Disorders of lipoid metabolism | endocrine/metabolic | UricA | |
| 272.1 | 0.8788 | 0.03665 | 2.408 | 4.38e-127 | 25384 | 11117 | 14267 | 0.7185 | 0.03487 | 2.051 | 2.41e-94 | 34065 | 10681 | 23384 | 3.56e-04 | -0.1795 | Hyperlipidemia | endocrine/metabolic | UricA | |
| 272.11 | 1.02 | 0.05001 | 2.774 | 1.59e-92 | 18174 | 3907 | 14267 | 0.8186 | 0.04765 | 2.267 | 3.83e-66 | 27356 | 3972 | 23384 | 7.17e-04 | -0.2321 | Hypercholesterolemia | endocrine/metabolic | UricA | |
| 272.12 | | | | | 14667 | 400 | 14267 | | | | | 23678 | 294 | 23384 | 0.4437 | -0.1243 | Hyperglyceridemia | endocrine/metabolic | UricA | |
| 272.13 | 1.214 | 0.0439 | 3.367 | 2.78e-168 | 20087 | 5820 | 14267 | 0.9815 | 0.04214 | 2.668 | 5.30e-120 | 28806 | 5422 | 23384 | 2.15e-05 | -0.2574 | Mixed hyperlipidemia | endocrine/metabolic | UricA | |
| 272.14 | | | | | 14272 | 5 | 14267 | | | | | 23388 | 4 | 23384 | | | Hyperchylomicronemia | endocrine/metabolic | UricA | |
| 272.9 | | | | | 14286 | 19 | 14267 | | | | | 23403 | 19 | 23384 | | | Unspecified disorder of lipoid metabolism | endocrine/metabolic | UricA | |
| 274 | 2.99 | 0.06599 | 19.88 | 0 | 26980 | 1926 | 25054 | 3.017 | 0.09342 | 20.43 | 7.67e-229 | 36355 | 864 | 35491 | 0.8829 | 0.01657 | Gout and other crystal arthropathies | endocrine/metabolic | UricA | |
| 274.1 | 3.116 | 0.07005 | 22.55 | 0 | 26882 | 1828 | 25054 | 3.602 | 0.1214 | 36.68 | 2.37e-193 | 36217 | 726 | 35491 | 0.001507 | 0.4391 | Gout | endocrine/metabolic | UricA | Gout |
| 274.11 | 4.447 | 0.2058 | 85.38 | 1.42e-103 | 25702 | 648 | 25054 | | | | | 35744 | 253 | 35491 | 0.7427 | 0.1256 | Gouty arthropathy | endocrine/metabolic | UricA | |
| 274.2 | | | | | 25175 | 121 | 25054 | | | | | 35648 | 157 | 35491 | | | Crystal arthropathies | endocrine/metabolic | UricA | |
| 274.21 | | | | | 25165 | 111 | 25054 | | | | | 35627 | 136 | 35491 | | | Chondrocalcinosis | endocrine/metabolic | UricA | |
| 275 | 1.345 | 0.04847 | 3.837 | 2.19e-169 | 25825 | 2131 | 23694 | 1.115 | 0.04532 | 3.05 | 1.02e-133 | 34866 | 2480 | 32386 | 0.1535 | -0.09132 | Disorders of mineral metabolism | endocrine/metabolic | UricA | |
| 275.1 | | | | | 23853 | 159 | 23694 | | | | | 32511 | 125 | 32386 | | | Disorders of iron metabolism | hematopoietic | UricA | |
| 275.11 | | | | | 23755 | 61 | 23694 | | | | | 32420 | 34 | 32386 | | | Hereditary hemochromatosis | hematopoietic | UricA | |
| 275.2 | | | | | 23712 | 18 | 23694 | | | | | 32400 | 14 | 32386 | | | Disorders of copper metabolism | endocrine/metabolic | UricA | |
| 275.3 | 1.551 | 0.07414 | 4.716 | 3.57e-97 | 24542 | 848 | 23694 | 1.244 | 0.07073 | 3.468 | 3.36e-69 | 33312 | 926 | 32386 | 0.05341 | -0.1907 | Disorders of magnesium metabolism | endocrine/metabolic | UricA | |
| 275.5 | 1.552 | 0.07196 | 4.72 | 3.95e-103 | 24610 | 916 | 23694 | 1.357 | 0.07001 | 3.884 | 1.10e-83 | 33307 | 921 | 32386 | 0.5149 | -0.06285 | Disorders of calcium/phosphorus metabolism | endocrine/metabolic | UricA | |
| 275.51 | | | | | 23996 | 302 | 23694 | | | | | 32803 | 417 | 32386 | 0.6345 | 0.07412 | Hypocalcemia | endocrine/metabolic | UricA | |
| 275.53 | 1.827 | 0.09395 | 6.213 | 3.34e-84 | 24238 | 544 | 23694 | | | | | 32769 | 383 | 32386 | 0.4444 | 0.1049 | Disorders of phosphorus metabolism | endocrine/metabolic | UricA | |
| 275.6 | | | | | 24110 | 416 | 23694 | 0.7942 | 0.08328 | 2.213 | 1.47e-21 | 33089 | 703 | 32386 | 0.4082 | -0.1061 | Hypercalcemia | endocrine/metabolic | UricA | |
| 276 | 1.032 | 0.03284 | 2.805 | 1.30e-216 | 24148 | 8143 | 16005 | 1.083 | 0.03154 | 2.954 | 1.58e-258 | 32644 | 8450 | 24194 | 0.009763 | 0.115 | Disorders of fluid, electrolyte, and acid-base balance | endocrine/metabolic | UricA | |
| 276.1 | 1.141 | 0.03551 | 3.131 | 1.18e-226 | 22066 | 6061 | 16005 | 1.206 | 0.03451 | 3.339 | 2.10e-267 | 30513 | 6319 | 24194 | 0.008933 | 0.1262 | Electrolyte imbalance | endocrine/metabolic | UricA | |
| 276.11 | | | | | 16449 | 444 | 16005 | | | | | 24509 | 315 | 24194 | 0.8588 | -0.02802 | Hyperosmolality and/or hypernatremia | endocrine/metabolic | UricA | |
| 276.12 | 1.114 | 0.05119 | 3.046 | 6.06e-105 | 18173 | 2168 | 16005 | 1.273 | 0.05174 | 3.571 | 1.34e-133 | 26321 | 2127 | 24194 | 1.59e-04 | 0.266 | Hyposmolality and/or hyponatremia | endocrine/metabolic | UricA | |
| 276.13 | 1.553 | 0.05055 | 4.724 | 3.87e-207 | 18167 | 2162 | 16005 | 1.718 | 0.0561 | 5.575 | 4.63e-206 | 25804 | 1610 | 24194 | 8.19e-04 | 0.2453 | Hyperpotassemia | endocrine/metabolic | UricA | |
| 276.14 | 1.247 | 0.04678 | 3.478 | 1.87e-156 | 18672 | 2667 | 16005 | 1.248 | 0.04157 | 3.485 | 3.71e-198 | 27767 | 3573 | 24194 | 0.2333 | 0.0725 | Hypopotassemia | endocrine/metabolic | UricA | |
| 276.4 | 1.547 | 0.05104 | 4.7 | 6.13e-202 | 18109 | 2104 | 16005 | 1.62 | 0.05256 | 5.052 | 1.42e-208 | 26022 | 1828 | 24194 | 0.05794 | 0.1345 | Acid-base balance disorder | endocrine/metabolic | UricA | |
| 276.41 | 1.585 | 0.0535 | 4.878 | 7.79e-193 | 17879 | 1874 | 16005 | 1.644 | 0.05544 | 5.174 | 3.35e-193 | 25798 | 1604 | 24194 | 0.07954 | 0.1308 | Acidosis | endocrine/metabolic | UricA | |
| 276.42 | | | | | 16255 | 250 | 16005 | | | | | 24456 | 262 | 24194 | 0.05378 | 0.3565 | Alkalosis | endocrine/metabolic | UricA | |

| phenotype | male_beta | male_SE | male_OR | male_p | male_n_total | male_n_cases | male_n_controls | female_beta | female_SE | female_OR | female_p | female_n_total | female_n_cases | female_n_controls | interaction_p | interaction_beta | description | group | lab | label |
|---|---|---|---|---|---|---|---|---|---|---|---|---|---|---|---|---|---|---|---|---|
| 276.5 | 1.148 | 0.04418 | 3.152 | 7.09e-149 | 19178 | 3173 | 16005 | 1.153 | 0.04212 | 3.168 | 5.04e-165 | 27587 | 3393 | 24194 | 0.1437 | 0.08685 | Hypovolemia | endocrine/metabolic | UricA | |
| 276.6 | 1.669 | 0.05562 | 5.307 | 7.73e-198 | 17709 | 1704 | 16005 | 1.782 | 0.05974 | 5.942 | 1.43e-195 | 25588 | 1394 | 24194 | 0.1096 | 0.1266 | Fluid overload | endocrine/metabolic | UricA | |
| 276.8 | | | | | 16014 | 9 | 16005 | | | | | 24216 | 22 | 24194 | | | Polydipsia | endocrine/metabolic | UricA | |
| 277 | | | | | 26131 | 420 | 25711 | 1.028 | 0.09007 | 2.797 | 3.42e-30 | 35099 | 554 | 34545 | 0.08677 | -0.2278 | Other disorders of metabolism | endocrine/metabolic | UricA | |
| 277.1 | | | | | 25722 | 11 | 25711 | | | | | 34554 | 9 | 34545 | | | Disorders of porphyrin metabolism | endocrine/metabolic | UricA | |
| 277.2 | | | | | 25713 | 2 | 25711 | | | | | 34547 | 2 | 34545 | | | Other disorders of purine and pyrimidine metabolism | endocrine/metabolic | UricA | |
| 277.4 | | | | | 25910 | 199 | 25711 | | | | | 34665 | 120 | 34545 | | | Disorders of bilirubin excretion | endocrine/metabolic | UricA | |
| 277.5 | | | | | 25845 | 134 | 25711 | | | | | 34656 | 111 | 34545 | | | Other disorders of lipoid metabolism | endocrine/metabolic | UricA | |
| 277.51 | | | | | 25790 | 79 | 25711 | | | | | 34590 | 45 | 34545 | | | Lipoprotein disorders | endocrine/metabolic | UricA | |
| 277.6 | | | | | 25723 | 12 | 25711 | | | | | 34560 | 15 | 34545 | | | Other deficiencies of circulating enzymes | endocrine/metabolic | UricA | |
| 277.7 | | | | | 25918 | 207 | 25711 | | | | | 34828 | 283 | 34545 | | | Dysmetabolic syndrome X | endocrine/metabolic | UricA | |
| 277.8 | | | | | 25715 | 4 | 25711 | | | | | 34546 | 1 | 34545 | | | Carnitine deficiencies | endocrine/metabolic | UricA | |
| 278 | 0.7731 | 0.03907 | 2.166 | 3.87e-87 | 24739 | 3939 | 20800 | 0.6726 | 0.03334 | 1.959 | 1.67e-90 | 32627 | 6840 | 25787 | 0.04515 | -0.1001 | Overweight, obesity and other hyperalimentation | endocrine/metabolic | UricA | |
| 278.1 | 0.8129 | 0.04166 | 2.254 | 8.45e-85 | 24116 | 3316 | 20800 | 0.7285 | 0.03562 | 2.072 | 5.82e-93 | 31306 | 5519 | 25787 | 0.08418 | -0.09197 | Obesity | endocrine/metabolic | UricA | |
| 278.11 | 0.7764 | 0.05754 | 2.174 | 1.72e-41 | 22371 | 1571 | 20800 | 0.7149 | 0.04412 | 2.044 | 4.75e-59 | 28985 | 3198 | 25787 | 0.4325 | -0.05506 | Morbid obesity | endocrine/metabolic | UricA | |
| 278.3 | | | | | 20837 | 37 | 20800 | | | | | 25988 | 201 | 25787 | | | Localized adiposity | endocrine/metabolic | UricA | |
| 278.4 | | | | | 21218 | 418 | 20800 | 0.4094 | 0.06528 | 1.506 | 3.56e-10 | 27263 | 1476 | 25787 | 0.01518 | -0.2937 | Abnormal weight gain | endocrine/metabolic | UricA | |
| 279 | 1.473 | 0.06426 | 4.363 | 2.46e-116 | 25738 | 1171 | 24567 | 1.2 | 0.05603 | 3.322 | 7.67e-102 | 33388 | 1505 | 31883 | 0.03518 | -0.1742 | Disorders involving the immune mechanism | endocrine/metabolic | UricA | |
| 279.1 | 1.488 | 0.06624 | 4.43 | 8.46e-112 | 25668 | 1101 | 24567 | 1.201 | 0.05814 | 3.322 | 1.00e-94 | 33274 | 1391 | 31883 | 0.03276 | -0.1826 | Immunity deficiency | endocrine/metabolic | UricA | |
| 279.11 | | | | | 24666 | 99 | 24567 | | | | | 32052 | 169 | 31883 | | | Deficiency of humoral immunity | endocrine/metabolic | UricA | |
| 279.2 | | | | | 24578 | 11 | 24567 | | | | | 31914 | 31 | 31883 | | | Autoimmune disease NEC | endocrine/metabolic | UricA | |
| 279.7 | | | | | 24823 | 256 | 24567 | 0.5043 | 0.07625 | 1.656 | 3.74e-11 | 32824 | 941 | 31883 | 0.2939 | -0.1576 | Other immunological findings | endocrine/metabolic | UricA | |
| 279.8 | | | | | 24598 | 31 | 24567 | | | | | 31935 | 52 | 31883 | | | Other specified disorders involving the immune mechanism | endocrine/metabolic | UricA | |
| 280 | 1.265 | 0.05617 | 3.545 | 2.29e-112 | 18083 | 1688 | 16395 | 0.9974 | 0.04652 | 2.711 | 5.82e-102 | 25891 | 2832 | 23059 | 0.004825 | -0.1997 | Iron deficiency anemias | hematopoietic | UricA | |
| 280.1 | 1.331 | 0.06175 | 3.783 | 5.62e-103 | 17740 | 1345 | 16395 | 1.045 | 0.04942 | 2.843 | 3.36e-99 | 25454 | 2395 | 23059 | 0.003745 | -0.2225 | Iron deficiency anemias, unspecified or not due to blood loss | hematopoietic | UricA | |
| 280.2 | 1.344 | 0.09534 | 3.836 | 3.76e-45 | 16909 | 514 | 16395 | 0.9208 | 0.08283 | 2.511 | 1.04e-28 | 23821 | 762 | 23059 | 0.002317 | -0.372 | Iron deficiency anemia secondary to blood loss (chronic) | hematopoietic | UricA | |
| 281 | | | | | 16870 | 475 | 16395 | 1.055 | 0.07376 | 2.872 | 2.06e-46 | 24011 | 952 | 23059 | 0.9336 | -0.009995 | Other deficiency anemia | hematopoietic | UricA | |
| 281.1 | | | | | 16669 | 274 | 16395 | 0.897 | 0.08768 | 2.452 | 1.45e-24 | 23736 | 677 | 23059 | 0.9315 | -0.01303 | Megaloblastic anemia | hematopoietic | UricA | |
| 281.11 | | | | | 16464 | 69 | 16395 | | | | | 23278 | 219 | 23059 | | | Pernicious anemia | hematopoietic | UricA | |

| phenotype | male_beta | male_SE | male_OR | male_p | male_n_total | male_n_cases | male_n_controls | female_beta | female_SE | female_OR | female_p | female_n_total | female_n_cases | female_n_controls | interaction_p | interaction_beta | description | group | lab | label |
|---|---|---|---|---|---|---|---|---|---|---|---|---|---|---|---|---|---|---|---|---|
| 281.12 | | | | | 16590 | 195 | 16395 | 0.8448 | 0.1013 | 2.328 | 7.56e-17 | 23561 | 502 | 23059 | 0.7237 | -0.06271 | Other vitamin B12 deficiency anemia | hematopoietic | UricA | |
| 281.13 | | | | | 16415 | 20 | 16395 | | | | | 23107 | 48 | 23059 | | | Folate-deficiency anemia | hematopoietic | UricA | |
| 281.9 | | | | | 16581 | 186 | 16395 | | | | | 23292 | 233 | 23059 | | | Deficiency anemias | hematopoietic | UricA | |
| 282 | | | | | 16442 | 47 | 16395 | | | | | 23190 | 131 | 23059 | | | Hereditary hemolytic anemias | hematopoietic | UricA | |
| 282.5 | | | | | 16399 | 4 | 16395 | | | | | 23069 | 10 | 23059 | | | Sickle cell anemia | hematopoietic | UricA | |
| 282.8 | | | | | 16410 | 15 | 16395 | | | | | 23102 | 43 | 23059 | | | Other hemoglobinopathies | hematopoietic | UricA | |
| 282.9 | | | | | 16427 | 32 | 16395 | | | | | 23140 | 81 | 23059 | | | Other hereditary hemolytic anemias | hematopoietic | UricA | |
| 283 | | | | | 16455 | 60 | 16395 | | | | | 23168 | 109 | 23059 | | | Acquired hemolytic anemias | hematopoietic | UricA | |
| 283.1 | | | | | 16418 | 23 | 16395 | | | | | 23095 | 36 | 23059 | | | Autoimmune hemolytic anemias | hematopoietic | UricA | |
| 283.2 | | | | | 16419 | 24 | 16395 | | | | | 23100 | 41 | 23059 | | | Non-autoimmune hemolytic anemias | hematopoietic | UricA | |
| 283.21 | | | | | 16402 | 7 | 16395 | | | | | 23073 | 14 | 23059 | | | Hemolytic-uremic syndrome | hematopoietic | UricA | |
| 284 | 2.373 | 0.0781 | 10.73 | 7.77e-203 | 17250 | 855 | 16395 | 2.231 | 0.08042 | 9.311 | 2.10e-169 | 23771 | 712 | 23059 | 0.2937 | -0.114 | Aplastic anemia | hematopoietic | UricA | |
| 284.1 | 2.405 | 0.08288 | 11.08 | 4.15e-185 | 17146 | 751 | 16395 | 2.238 | 0.08445 | 9.371 | 1.05e-154 | 23702 | 643 | 23059 | 0.3244 | -0.113 | Pancytopenia | hematopoietic | UricA | |
| 284.2 | | | | | 16406 | 11 | 16395 | | | | | 23065 | 6 | 23059 | | | Constitutional aplastic anemia | hematopoietic | UricA | |
| 285 | 1.038 | 0.03353 | 2.824 | 1.72e-210 | 23977 | 7582 | 16395 | 1.117 | 0.03266 | 3.055 | 3.16e-256 | 30991 | 7932 | 23059 | 0.02055 | 0.106 | Other anemias | hematopoietic | UricA | |
| 285.1 | 0.8133 | 0.04738 | 2.255 | 4.98e-66 | 19293 | 2898 | 16395 | 1.066 | 0.05212 | 2.904 | 5.55e-93 | 25340 | 2281 | 23059 | 2.29e-04 | 0.2515 | Acute posthemorrhagic anemia | hematopoietic | UricA | |
| 285.2 | 1.748 | 0.04923 | 5.742 | 4.43e-276 | 18710 | 2315 | 16395 | 1.732 | 0.04967 | 5.653 | 2.05e-266 | 25252 | 2193 | 23059 | 0.7499 | -0.02155 | Anemia of chronic disease | hematopoietic | UricA | |
| 285.21 | 2.148 | 0.07864 | 8.571 | 2.51e-164 | 17283 | 888 | 16395 | 2.317 | 0.08347 | 10.15 | 1.36e-169 | 23775 | 716 | 23059 | 0.1206 | 0.1731 | Anemia in chronic kidney disease | hematopoietic | UricA | |
| 285.22 | 1.559 | 0.06737 | 4.752 | 2.12e-118 | 17510 | 1115 | 16395 | 1.51 | 0.06602 | 4.525 | 1.02e-115 | 24273 | 1214 | 23059 | 0.7972 | -0.0235 | Anemia in neoplastic disease | hematopoietic | UricA | |
| 285.3 | | | | | 16401 | 6 | 16395 | | | | | 23062 | 3 | 23059 | | | Sideroblastic anemia | hematopoietic | UricA | |
| 285.8 | | | | | 16399 | 4 | 16395 | | | | | 23072 | 13 | 23059 | | | Hemoglobinuria | hematopoietic | UricA | |
| 286 | 1.148 | 0.05334 | 3.151 | 1.12e-102 | 21087 | 1817 | 19270 | 1.118 | 0.05151 | 3.058 | 2.03e-104 | 30311 | 1937 | 28374 | 0.7442 | -0.02345 | Coagulation defects | hematopoietic | UricA | |
| 286.1 | | | | | 19382 | 112 | 19270 | | | | | 28488 | 114 | 28374 | | | Congenital coagulation defects | hematopoietic | UricA | |
| 286.11 | | | | | 19306 | 36 | 19270 | | | | | 28422 | 48 | 28374 | | | Von willebrand's disease | hematopoietic | UricA | |
| 286.12 | | | | | 19305 | 35 | 19270 | | | | | 28432 | 58 | 28374 | | | Congenital deficiency of other clotting factors (including factor VII) | hematopoietic | UricA | |
| 286.13 | | | | | 19324 | 54 | 19270 | | | | | 28385 | 11 | 28374 | | | Congenital factor VIII disorder | hematopoietic | UricA | |
| 286.2 | 0.8865 | 0.04392 | 2.427 | 1.40e-90 | 22410 | 3140 | 19270 | 0.9043 | 0.04651 | 2.47 | 3.48e-84 | 31115 | 2741 | 28374 | 0.7939 | 0.01628 | Encounter for long-term (current) use of anticoagulants | hematopoietic | UricA | |
| 286.3 | | | | | 6 | | | | | | | 28478 | 106 | 28372 | | | Coagulation defects complicating pregnancy or postpartum | hematopoietic | UricA | |
| 286.4 | | | | | 19353 | 83 | 19270 | | | | | 28429 | 55 | 28374 | | | Acquired coagulation factor deficiency | hematopoietic | UricA | |
| 286.5 | | | | | 19388 | 118 | 19270 | | | | | 28511 | 137 | 28374 | | | Hemorrhagic disorder due to intrinsic circulating anticoagulants | hematopoietic | UricA | |
| 286.6 | | | | | 19312 | 42 | 19270 | | | | | 28420 | 46 | 28374 | | | Defibrination syndrome | hematopoietic | UricA | |
| 286.7 | 1.177 | 0.07365 | 3.244 | 1.80e-57 | 20156 | 886 | 19270 | 1.239 | 0.07052 | 3.454 | 3.79e-69 | 29323 | 949 | 28374 | 0.4021 | 0.08261 | Other and unspecified coagulation defects | hematopoietic | UricA | |
| 286.8 | | | | | 19460 | 190 | 19270 | | | | | 28789 | 415 | 28374 | 0.5198 | 0.1205 | Hypercoagulable state | hematopoietic | UricA | |
| 286.81 | | | | | 19445 | 175 | 19270 | | | | | 28747 | 373 | 28374 | 0.4259 | 0.156 | Primary hypercoagulable state | hematopoietic | UricA | |
| 286.9 | | | | | 19741 | 471 | 19270 | | | | | 28784 | 410 | 28374 | 0.2634 | -0.1561 | Abnormal coagulation profile | hematopoietic | UricA | |
| 287 | 1.255 | 0.04769 | 3.507 | 1.35e-152 | 21610 | 2340 | 19270 | 1.303 | 0.05154 | 3.681 | 4.84e-141 | 30271 | 1897 | 28374 | 0.6899 | 0.0272 | Purpura and other hemorrhagic conditions | hematopoietic | UricA | |
| 287.1 | | | | | 19326 | 56 | 19270 | | | | | 28485 | 111 | 28374 | | | Spontaneous ecchymoses | hematopoietic | UricA | |

| phenotype | male_beta | male_SE | male_OR | male_p | male_n_total | male_n_cases | male_n_controls | female_beta | female_SE | female_OR | female_p | female_n_total | female_n_cases | female_n_controls | interaction_p | interaction_beta | description | group | lab | label |
|---|---|---|---|---|---|---|---|---|---|---|---|---|---|---|---|---|---|---|---|---|
| 287.2 | | | | | 19288 | 18 | 19270 | | | | | 28390 | 16 | 28374 | | | Allergic purpura | hematopoietic | UricA | |
| 287.3 | 1.299 | 0.04929 | 3.667 | 3.71e-153 | 21432 | 2162 | 19270 | 1.37 | 0.05359 | 3.934 | 4.49e-144 | 30102 | 1728 | 28374 | 0.4794 | 0.04995 | Thrombocytopenia | hematopoietic | UricA | |
| 287.31 | | | | | 19391 | 121 | 19270 | | | | | 28538 | 164 | 28374 | | | Primary thrombocytopenia | hematopoietic | UricA | |
| 287.32 | | | | | 19594 | 324 | 19270 | | | | | 28675 | 301 | 28374 | 0.2839 | 0.1745 | Secondary thrombocytopenia | hematopoietic | UricA | |
| 287.4 | | | | | 19290 | 20 | 19270 | | | | | 28393 | 19 | 28374 | | | Qualitative platelet defects | hematopoietic | UricA | |
| 288 | 1.027 | 0.04539 | 2.792 | 2.85e-113 | 21474 | 2790 | 18684 | 1.112 | 0.04473 | 3.04 | 2.15e-136 | 29549 | 2753 | 26796 | 0.007942 | 0.1633 | Diseases of white blood cells | hematopoietic | UricA | |
| 288.1 | 1.8 | 0.06671 | 6.047 | 2.95e-160 | 19788 | 1104 | 18684 | 1.277 | 0.05996 | 3.587 | 1.05e-100 | 28156 | 1360 | 26796 | 1.23e-05 | -0.3773 | Decreased white blood cell count | hematopoietic | UricA | |
| 288.11 | 1.872 | 0.0764 | 6.499 | 1.48e-132 | 19506 | 822 | 18684 | 1.27 | 0.069 | 3.56 | 1.25e-75 | 27815 | 1019 | 26796 | 5.63e-06 | -0.4501 | Neutropenia | hematopoietic | UricA | |
| 288.2 | 1.006 | 0.04912 | 2.733 | 3.98e-93 | 21008 | 2324 | 18684 | 1.066 | 0.04897 | 2.904 | 4.63e-105 | 29055 | 2259 | 26796 | 0.03025 | 0.1449 | Elevated white blood cell count | hematopoietic | UricA | |
| 288.3 | | | | | 18778 | 94 | 18684 | | | | | 26909 | 113 | 26796 | | | Eosinophilia | hematopoietic | UricA | |
| 289 | | | | | 19172 | 488 | 18684 | 1.439 | 0.08486 | 4.218 | 1.61e-64 | 27421 | 625 | 26796 | 0.003801 | -0.3655 | Other diseases of blood and blood-forming organs | hematopoietic | UricA | |
| 289.1 | | | | | 18755 | 71 | 18684 | | | | | 26856 | 60 | 26796 | | | Myelofibrosis | hematopoietic | UricA | |
| 289.3 | | | | | 18767 | 83 | 18684 | | | | | 26919 | 123 | 26796 | | | Personal history of diseases of blood and blood-forming organs | hematopoietic | UricA | |
| 289.4 | 0.8961 | 0.06024 | 2.45 | 4.78e-50 | 20171 | 1487 | 18684 | 0.7347 | 0.05858 | 2.085 | 4.39e-36 | 28443 | 1647 | 26796 | 0.7151 | -0.02961 | Lymphadenitis | hematopoietic | UricA | |
| 289.5 | | | | | 18840 | 156 | 18684 | | | | | 26948 | 152 | 26796 | | | Diseases of spleen | hematopoietic | UricA | |
| 289.8 | | | | | 18685 | 237 | 18448 | | | | | 26687 | 169 | 26518 | | | Polycythemia, secondary | hematopoietic | UricA | |
| 289.9 | | | | | 18788 | 104 | 18684 | | | | | 26914 | 118 | 26796 | | | Abnormality of red blood cells | hematopoietic | UricA | |
| 290 | 0.5574 | 0.0651 | 1.746 | 1.10e-17 | 21683 | 1272 | 20411 | 0.6388 | 0.06916 | 1.894 | 2.57e-20 | 30218 | 1222 | 28996 | 0.2085 | 0.1156 | Delirium dementia and amnestic and other cognitive disorders | mental disorders | UricA | |
| 290.1 | 0.3103 | 0.08409 | 1.364 | 2.24e-04 | 21205 | 794 | 20411 | 0.4075 | 0.08656 | 1.503 | 2.50e-06 | 29834 | 838 | 28996 | 0.5905 | 0.06272 | Dementias | mental disorders | UricA | |
| 290.11 | | | | | 20682 | 271 | 20411 | | | | | 29352 | 356 | 28996 | 0.5411 | 0.1139 | Alzheimer's disease | mental disorders | UricA | |
| 290.12 | | | | | 20506 | 95 | 20411 | | | | | 29043 | 47 | 28996 | | | Dementia with cerebral degenerations | mental disorders | UricA | |
| 290.13 | | | | | 20487 | 76 | 20411 | | | | | 29129 | 133 | 28996 | | | Senile dementia | mental disorders | UricA | |
| 290.16 | | | | | 20543 | 132 | 20411 | | | | | 29107 | 111 | 28996 | | | Vascular dementia | mental disorders | UricA | |
| 290.2 | | | | | 20877 | 466 | 20411 | | | | | 29341 | 345 | 28996 | 0.08189 | 0.2574 | Delirium due to conditions classified elsewhere | mental disorders | UricA | |
| 290.3 | | | | | 20762 | 351 | 20411 | | | | | 29366 | 370 | 28996 | 0.9064 | -0.01913 | Other persistent mental disorders due to conditions classified elsewhere | mental disorders | UricA | |
| 291 | 0.7228 | 0.06262 | 2.06 | 8.09e-31 | 21730 | 1319 | 20411 | 0.7156 | 0.06481 | 2.045 | 2.41e-28 | 30293 | 1297 | 28996 | 0.4538 | 0.06558 | Other specified nonpsychotic and/or transient mental disorders | mental disorders | UricA | |
| 291.1 | | | | | 20440 | 29 | 20411 | | | | | 29037 | 41 | 28996 | | | Transient mental disorders due to conditions classified elsewhere | mental disorders | UricA | |
| 291.4 | | | | | 20576 | 165 | 20411 | | | | | 29194 | 198 | 28996 | | | Specific nonpsychotic mental disorders due to brain damage | mental disorders | UricA | |
| 291.8 | 0.7787 | 0.0672 | 2.179 | 4.83e-31 | 21537 | 1126 | 20411 | 0.7509 | 0.07185 | 2.119 | 1.45e-25 | 30045 | 1049 | 28996 | 0.6885 | 0.03834 | Alteration of consciousness | mental disorders | UricA | |
| 292 | 0.6003 | 0.03943 | 1.823 | 2.40e-52 | 24322 | 3911 | 20411 | 0.5763 | 0.0396 | 1.779 | 5.55e-48 | 33173 | 4177 | 28996 | 0.6109 | 0.0277 | Neurological disorders | mental disorders | UricA | |
| 292.1 | 0.4075 | 0.08329 | 1.503 | 9.98e-07 | 21162 | 751 | 20411 | 0.693 | 0.08194 | 2 | 2.72e-17 | 29761 | 765 | 28996 | 0.001733 | 0.3554 | Aphasia/speech disturbance | mental disorders | UricA | |

| phenotype | male_beta | male_SE | male_OR | male_p | male_n_total | male_n_cases | male_n_controls | female_beta | female_SE | female_OR | female_p | female_n_total | female_n_cases | female_n_controls | interaction_p | interaction_beta | description | group | lab | label |
|---|---|---|---|---|---|---|---|---|---|---|---|---|---|---|---|---|---|---|---|---|
| 292.11 | | | | | 20684 | 273 | 20411 | | | | | 29270 | 274 | 28996 | 0.06454 | 0.3451 | Aphasia | mental disorders | UricA | |
| 292.12 | | | | | 20439 | 28 | 20411 | | | | | 29020 | 24 | 28996 | | | Symbolic dysfunction | mental disorders | UricA | |
| 292.2 | | | | | 20778 | 367 | 20411 | | | | | 29334 | 338 | 28996 | 0.6166 | -0.08246 | Mild cognitive impairment | mental disorders | UricA | |
| 292.3 | 0.2761 | 0.0755 | 1.318 | 2.55e-04 | 21327 | 916 | 20411 | 0.3206 | 0.07167 | 1.378 | 7.72e-06 | 30127 | 1131 | 28996 | 0.644 | 0.04672 | Memory loss | mental disorders | UricA | |
| 292.4 | 0.8334 | 0.04773 | 2.301 | 2.94e-68 | 22762 | 2351 | 20411 | 0.8329 | 0.04977 | 2.3 | 7.11e-63 | 31254 | 2258 | 28996 | 0.4389 | 0.05186 | Altered mental status | mental disorders | UricA | |
| 292.5 | | | | | 20547 | 136 | 20411 | | | | | 29184 | 188 | 28996 | | | Transient alteration of awareness | mental disorders | UricA | |
| 292.6 | | | | | 20491 | 80 | 20411 | | | | | 29129 | 133 | 28996 | | | Hallucinations | mental disorders | UricA | |
| 293 | | | | | 25542 | 80 | 25462 | | | | | 34286 | 108 | 34178 | | | Symptoms involving head and neck | mental disorders | UricA | |
| 293.1 | 0.4676 | 0.07586 | 1.596 | 7.11e-10 | 26351 | 889 | 25462 | 0.3353 | 0.08016 | 1.398 | 2.87e-05 | 35053 | 875 | 34178 | 0.8993 | 0.01351 | Swelling, mass, or lump in head and neck [Space-occupying lesion, intracranial NOS] | mental disorders | UricA | |
| 295 | | | | | 17073 | 479 | 16594 | 0.7161 | 0.1019 | 2.047 | 2.13e-12 | 19345 | 524 | 18821 | 0.2054 | 0.1782 | Schizophrenia and other psychotic disorders | mental disorders | UricA | |
| 295.1 | | | | | 16740 | 146 | 16594 | | | | | 18981 | 160 | 18821 | | | Schizophrenia | mental disorders | UricA | |
| 295.2 | | | | | 16700 | 106 | 16594 | | | | | 18968 | 147 | 18821 | | | Paranoid disorders | mental disorders | UricA | |
| 295.3 | | | | | 16878 | 284 | 16594 | | | | | 19094 | 273 | 18821 | 0.9262 | 0.01745 | Psychosis | mental disorders | UricA | |
| 296 | 0.7609 | 0.03931 | 2.14 | 1.92e-83 | 21203 | 4609 | 16594 | 0.6176 | 0.03488 | 1.855 | 3.59e-70 | 27169 | 8348 | 18821 | 0.8025 | -0.01282 | Mood disorders | mental disorders | UricA | |
| 296.1 | 0.9058 | 0.08332 | 2.474 | 1.59e-27 | 17404 | 810 | 16594 | 0.6469 | 0.0678 | 1.91 | 1.42e-21 | 20203 | 1382 | 18821 | 0.4494 | -0.07774 | Bipolar | mental disorders | UricA | |
| 296.2 | 0.7585 | 0.0414 | 2.135 | 5.46e-75 | 20515 | 3921 | 16594 | 0.6237 | 0.03604 | 1.866 | 4.25e-67 | 26233 | 7412 | 18821 | 0.7165 | -0.01945 | Depression | mental disorders | UricA | |
| 296.22 | 0.8179 | 0.05195 | 2.266 | 7.38e-56 | 18825 | 2231 | 16594 | 0.6606 | 0.04289 | 1.936 | 1.60e-53 | 23232 | 4411 | 18821 | 0.4294 | -0.05164 | Major depressive disorder | mental disorders | UricA | |
| 297 | | | | | 17005 | 411 | 16594 | 0.4365 | 0.1198 | 1.547 | 2.68e-04 | 19321 | 500 | 18821 | 0.4466 | -0.1285 | Suicidal ideation or attempt | mental disorders | UricA | |
| 297.1 | | | | | 16944 | 350 | 16594 | | | | | 19222 | 401 | 18821 | 0.224 | -0.2227 | Suicidal ideation | mental disorders | UricA | |
| 297.2 | | | | | 16666 | 72 | 16594 | | | | | 18942 | 121 | 18821 | | | Suicide or self-inflicted injury | mental disorders | UricA | |
| 300 | 0.7224 | 0.04228 | 2.059 | 1.82e-65 | 20603 | 4009 | 16594 | 0.6182 | 0.03544 | 1.856 | 3.87e-68 | 27035 | 8214 | 18821 | 0.5553 | 0.03164 | Anxiety disorders | mental disorders | UricA | |
| 300.1 | 0.7069 | 0.0446 | 2.028 | 1.43e-56 | 20057 | 3463 | 16594 | 0.6003 | 0.03686 | 1.823 | 1.24e-59 | 26048 | 7227 | 18821 | 0.4874 | 0.03904 | Anxiety disorder | mental disorders | UricA | |
| 300.11 | 0.6932 | 0.09082 | 2 | 2.31e-14 | 17284 | 690 | 16594 | 0.5909 | 0.06612 | 1.806 | 4.00e-19 | 20323 | 1502 | 18821 | 0.8193 | 0.02452 | Generalized anxiety disorder | mental disorders | UricA | |
| 300.12 | | | | | 16882 | 288 | 16594 | 0.8691 | 0.09593 | 2.385 | 1.31e-19 | 19405 | 584 | 18821 | 0.5021 | 0.1058 | Agorophobia, social phobia, and panic disorder | mental disorders | UricA | |
| 300.13 | | | | | 16650 | 56 | 16594 | | | | | 18930 | 109 | 18821 | | | Phobia | mental disorders | UricA | |
| 300.2 | | | | | 16594 | | | | | | | 18821 | | | | | Generalized anxiety & phobic disorders | mental disorders | UricA | |
| 300.3 | | | | | 16794 | 200 | 16594 | | | | | 19097 | 276 | 18821 | | | Obsessive-compulsive disorders | mental disorders | UricA | |
| 300.4 | | | | | 16995 | 401 | 16594 | 1.348 | 0.07751 | 3.851 | 8.70e-68 | 19615 | 794 | 18821 | 0.9679 | -0.005139 | Dysthymic disorder | mental disorders | UricA | |
| 300.8 | | | | | 16639 | 45 | 16594 | | | | | 18944 | 123 | 18821 | | | Acute reaction to stress | mental disorders | UricA | |

| phenotype | male_beta | male_SE | male_OR | male_p | male_n_total | male_n_cases | male_n_controls | female_beta | female_SE | female_OR | female_p | female_n_total | female_n_cases | female_n_controls | interaction_p | interaction_beta | description | group | lab | label |
|---|---|---|---|---|---|---|---|---|---|---|---|---|---|---|---|---|---|---|---|---|
| 300.9 | | | | | 16936 | 342 | 16594 | 0.6823 | 0.08201 | 1.978 | 8.86e-17 | 19768 | 947 | 18821 | 0.4784 | 0.1051 | Posttraumatic stress disorder | mental disorders | UricA | |
| 301 | | | | | 16724 | 130 | 16594 | | | | | 19101 | 280 | 18821 | | | Personality disorders | mental disorders | UricA | |
| 301.1 | | | | | 16605 | 11 | 16594 | | | | | 18829 | 8 | 18821 | | | Schizoid personality disorder | mental disorders | UricA | |
| 301.2 | | | | | 16645 | 51 | 16594 | | | | | 19000 | 179 | 18821 | | | Antisocial/borderline personality disorder | mental disorders | UricA | |
| 302 | | | | | 16787 | 193 | 16594 | | | | | 18953 | 132 | 18821 | | | Sexual and gender identity disorders | mental disorders | UricA | |
| 302.1 | | | | | 16728 | 134 | 16594 | | | | | 18905 | 84 | 18821 | | | Decreased libido | mental disorders | UricA | |
| 303 | | | | | 16730 | 136 | 16594 | | | | | 19170 | 349 | 18821 | | | Psychogenic and somatoform disorders | mental disorders | UricA | |
| 303.1 | | | | | 16603 | 9 | 16594 | | | | | 18849 | 28 | 18821 | | | Dissociative disorder | mental disorders | UricA | |
| 303.3 | | | | | 16631 | 37 | 16594 | | | | | 18918 | 97 | 18821 | | | Psychogenic disorder | mental disorders | UricA | |
| 303.31 | | | | | 16597 | 3 | 16594 | | | | | 18830 | 9 | 18821 | | | Gastrointestinal malfunction arising from mental factors | mental disorders | UricA | |
| 303.4 | | | | | 16689 | 95 | 16594 | | | | | 19064 | 243 | 18821 | | | Somatoform disorder | mental disorders | UricA | |
| 304 | 0.7662 | 0.07602 | 2.152 | 6.89e-24 | 17547 | 953 | 16594 | 0.5648 | 0.06108 | 1.759 | 2.31e-20 | 20609 | 1788 | 18821 | 0.9792 | -0.002455 | Adjustment reaction | mental disorders | UricA | |
| 305.2 | | | | | 16644 | 50 | 16594 | | | | | 19240 | 419 | 18821 | | | Eating disorder | mental disorders | UricA | |
| 305.21 | | | | | 16607 | 13 | 16594 | | | | | 18929 | 108 | 18821 | | | Anorexia nervosa | mental disorders | UricA | |
| 306 | 0.5179 | 0.07137 | 1.679 | 3.95e-13 | 17668 | 1074 | 16594 | 0.675 | 0.06134 | 1.964 | 3.65e-28 | 20481 | 1660 | 18821 | 0.05074 | 0.1785 | Other mental disorder | mental disorders | UricA | |
| 306.1 | | | | | 0 | | | | | | | 19216 | 395 | 18821 | | | Mental disorders durring/after pregnancy | mental disorders | UricA | |
| 306.9 | | | | | 16626 | 32 | 16594 | | | | | 18928 | 107 | 18821 | | | Tension headache | mental disorders | UricA | |
| 312 | | | | | 25880 | 213 | 25667 | | | | | 34967 | 157 | 34810 | | | Conduct disorders | mental disorders | UricA | |
| 312.3 | | | | | 25729 | 62 | 25667 | | | | | 34851 | 41 | 34810 | | | Impulse control disorder | mental disorders | UricA | |
| 313 | 0.01517 | 0.1215 | 1.015 | 0.9006 | 26378 | 711 | 25667 | -0.08331 | 0.1054 | 0.9201 | 0.4291 | 35536 | 726 | 34810 | 0.5239 | 0.09841 | Pervasive developmental disorders | mental disorders | UricA | |
| 313.1 | 0.09525 | 0.1376 | 1.1 | 0.4888 | 26178 | 511 | 25667 | -0.05515 | 0.1202 | 0.9463 | 0.6465 | 35352 | 542 | 34810 | 0.8815 | 0.02613 | Attention deficit hyperactivity disorder | mental disorders | UricA | |
| 313.2 | | | | | 25726 | 59 | 25667 | | | | | 34849 | 39 | 34810 | | | Tics and stuttering | mental disorders | UricA | |
| 313.3 | | | | | 25854 | 187 | 25667 | | | | | 34891 | 81 | 34810 | | | Autism | mental disorders | UricA | |
| 315 | -0.131 | 0.1146 | 0.8772 | 0.2528 | 26237 | 570 | 25667 | | | | | 35246 | 436 | 34810 | 0.1257 | 0.2499 | Develomental delays and disorders | mental disorders | UricA | |
| phenotype | male_beta | male_SE | male_OR | male_p | male_n_total | male_n_cases | male_n_controls | female_beta | female_SE | female_OR | female_p | female_n_total | female_n_cases | female_n_controls | interaction_p | interaction_beta | description | group | lab | label |
| 315.1 | | | | | 25693 | 26 | 25667 | | | | | 34830 | 20 | 34810 | | | Learning disorder | mental disorders | UricA | |
| 315.2 | | | | | 25753 | 86 | 25667 | | | | | 34881 | 71 | 34810 | | | Speech and language disorder | mental disorders | UricA | |
| 315.3 | | | | | 25849 | 182 | 25667 | | | | | 34981 | 171 | 34810 | | | Mental retardation | mental disorders | UricA | |
| 316 | 0.7256 | 0.06361 | 2.066 | 3.87e-30 | 19622 | 1399 | 18223 | 0.6283 | 0.05737 | 1.874 | 6.52e-28 | 29618 | 1741 | 27877 | 0.5561 | -0.04848 | Substance addiction and disorders | mental disorders | UricA | |

| phenotype | male_beta | male_SE | male_OR | male_p | male_n_total | male_n_cases | male_n_controls | female_beta | female_SE | female_OR | female_p | female_n_total | female_n_cases | female_n_controls | interaction_p | interaction_beta | description | group | lab | label |
|---|---|---|---|---|---|---|---|---|---|---|---|---|---|---|---|---|---|---|---|---|
| 316.1 | | | | | 18355 | 132 | 18223 | | | | | 28169 | 292 | 27877 | | | Polyneuropathy due to drugs | mental disorders | UricA | |
| 317 | 0.5732 | 0.0644 | 1.774 | 5.56e-19 | 19570 | 1347 | 18223 | 0.3375 | 0.0965 | 1.401 | 4.70e-04 | 28525 | 648 | 27877 | 0.1189 | -0.1757 | Alcohol-related disorders | mental disorders | UricA | |
| 317.1 | 0.5308 | 0.07562 | 1.7 | 2.22e-12 | 19193 | 970 | 18223 | | | | | 28321 | 444 | 27877 | 0.7384 | -0.04444 | Alcoholism | mental disorders | UricA | |
| 317.11 | | | | | 18678 | 455 | 18223 | | | | | 28037 | 160 | 27877 | 0.7174 | -0.08095 | Alcoholic liver damage | mental disorders | UricA | |
| 318 | 0.4555 | 0.03656 | 1.577 | 1.24e-35 | 23438 | 5215 | 18223 | 0.4928 | 0.03765 | 1.637 | 3.77e-39 | 32604 | 4727 | 27877 | 0.9914 | -5.54e-04 | Tobacco use disorder | mental disorders | UricA | |
| 320 | | | | | 27253 | 157 | 27096 | | | | | 36416 | 155 | 36261 | | | Meningitis | neurological | UricA | |
| 323 | | | | | 27190 | 94 | 27096 | | | | | 36377 | 116 | 36261 | | | Encephalitis | neurological | UricA | |
| 323.2 | | | | | 27111 | 15 | 27096 | | | | | 36289 | 28 | 36261 | | | Acute (transverse) myelitis | neurological | UricA | |
| 323.8 | | | | | 27165 | 69 | 27096 | | | | | 36347 | 86 | 36261 | | | Encephalitis, non-infectious | neurological | UricA | |
| 324 | | | | | 27283 | 187 | 27096 | | | | | 36381 | 120 | 36261 | | | Other CNS infection and poliomyelitis | neurological | UricA | |
| 324.1 | | | | | 27097 | 1 | 27096 | | | | | 36261 | | | | | Jakob-Creutzfeldt disease | neurological | UricA | |
| 325 | | | | | 27112 | 16 | 27096 | | | | | 36306 | 45 | 36261 | | | Phlebitis and thrombophlebitis of intracranial venous sinuses | neurological | UricA | |
| 327 | 0.7094 | 0.04342 | 2.033 | 5.41e-60 | 22679 | 3201 | 19478 | 0.4682 | 0.03857 | 1.597 | 6.47e-34 | 31789 | 4772 | 27017 | 0.01854 | -0.1336 | Sleep disorders | neurological | UricA | |
| 327.1 | | | | | 19663 | 185 | 19478 | | | | | 27358 | 341 | 27017 | 0.07268 | -0.3561 | Hypersomnia | neurological | UricA | |
| 327.3 | 0.6411 | 0.04029 | 1.899 | 5.16e-57 | 23346 | 3868 | 19478 | 0.5255 | 0.04438 | 1.691 | 2.42e-32 | 30225 | 3208 | 27017 | 0.421 | -0.04714 | Sleep apnea | neurological | UricA | |
| 327.31 | | | | | 19588 | 110 | 19478 | | | | | 27080 | 63 | 27017 | | | Central/nonobstructive sleep apnea | neurological | UricA | |
| 327.32 | 0.6655 | 0.04235 | 1.945 | 1.21e-55 | 22855 | 3377 | 19478 | 0.5733 | 0.04724 | 1.774 | 6.74e-34 | 29718 | 2701 | 27017 | 0.644 | -0.02865 | Obstructive sleep apnea | neurological | UricA | |
| 327.4 | 0.752 | 0.05269 | 2.121 | 3.23e-46 | 21429 | 1951 | 19478 | 0.4811 | 0.04524 | 1.618 | 2.07e-26 | 30146 | 3129 | 27017 | 0.05416 | -0.1304 | Insomnia | neurological | UricA | |
| 327.41 | | | | | 19695 | 217 | 19478 | | | | | 27448 | 431 | 27017 | 0.1705 | -0.2412 | Organic or persistent insomnia | neurological | UricA | |
| 327.5 | | | | | 19615 | 137 | 19478 | | | | | 27116 | 99 | 27017 | | | Parasomnia | neurological | UricA | |
| 327.6 | | | | | 19505 | 27 | 19478 | | | | | 27055 | 38 | 27017 | | | Circadian rhythm sleep disorder | neurological | UricA | |
| 327.7 | 0.8607 | 0.09451 | 2.365 | 8.49e-20 | 19996 | 518 | 19478 | 0.52 | 0.07356 | 1.682 | 1.56e-12 | 28011 | 994 | 27017 | 0.01563 | -0.2807 | Sleep related movement disorders | neurological | UricA | |
| 327.71 | | | | | 19923 | 445 | 19478 | 0.5276 | 0.0778 | 1.695 | 1.19e-11 | 27896 | 879 | 27017 | 0.03492 | -0.2615 | Restless legs syndrome | neurological | UricA | |
| 327.72 | | | | | 19495 | 17 | 19478 | | | | | 27043 | 26 | 27017 | | | Sleep related leg cramps | neurological | UricA | |
| 331 | 0.1439 | 0.1062 | 1.155 | 0.1756 | 20649 | 514 | 20135 | | | | | 28193 | 478 | 27715 | 0.5361 | 0.09443 | Other cerebral degenerations | neurological | UricA | |
| 331.1 | | | | | 20475 | 340 | 20135 | | | | | 28036 | 321 | 27715 | 0.888 | 0.02956 | Hydrocephalus | neurological | UricA | |
| 331.9 | | | | | 20281 | 146 | 20135 | | | | | 27836 | 121 | 27715 | | | Cerebral degeneration, unspecified | neurological | UricA | |
| 332 | | | | | 20573 | 438 | 20135 | | | | | 27974 | 259 | 27715 | 0.3558 | -0.1864 | Parkinson's disease | neurological | UricA | |
| 333 | 0.4579 | 0.0776 | 1.581 | 3.62e-09 | 20955 | 820 | 20135 | 0.3347 | 0.07827 | 1.398 | 1.90e-05 | 28634 | 919 | 27715 | 0.4898 | -0.07392 | Extrapyramidal disease and abnormal movement disorders | neurological | UricA | |
| 333.1 | | | | | 20501 | 366 | 20135 | | | | | 28134 | 419 | 27715 | 0.3904 | -0.1329 | Essential tremor | neurological | UricA | |
| 333.2 | | | | | 20258 | 123 | 20135 | | | | | 27820 | 105 | 27715 | | | Myoclonus | neurological | UricA | |
| 333.3 | | | | | 20164 | 29 | 20135 | | | | | 27748 | 33 | 27715 | | | Tics and choreas | neurological | UricA | |
| 333.4 | | | | | 20284 | 149 | 20135 | | | | | 28005 | 290 | 27715 | | | Torsion dystonia | neurological | UricA | |
| 333.8 | | | | | 20218 | 83 | 20135 | | | | | 27746 | 31 | 27715 | | | Other degenerative diseases of the basal ganglia | neurological | UricA | |
| 334 | | | | | 20526 | 391 | 20135 | | | | | 28145 | 430 | 27715 | 0.5119 | 0.1071 | Degenerative disease of the spinal cord | neurological | UricA | |
| 334.1 | | | | | 20181 | 46 | 20135 | | | | | 27786 | 71 | 27715 | | | Spinocerebellar disease | neurological | UricA | |
| 334.2 | | | | | 20201 | 66 | 20135 | | | | | 27768 | 53 | 27715 | | | Anterior horn cell disease | neurological | UricA | |
| 334.21 | | | | | 20183 | 48 | 20135 | | | | | 27759 | 44 | 27715 | | | Amyotrophic Lateral Sclerosis | neurological | UricA | |
| 335 | | | | | 20445 | 310 | 20135 | -0.987 | 0.1217 | 0.3727 | 5.05e-16 | 28694 | 979 | 27715 | 0.7766 | -0.06216 | Multiple sclerosis | neurological | UricA | |

| phenotype | male_beta | male_SE | male_OR | male_p | male_n_total | male_n_cases | male_n_controls | female_beta | female_SE | female_OR | female_p | female_n_total | female_n_cases | female_n_controls | interaction_p | interaction_beta | description | group | lab | label |
|---|---|---|---|---|---|---|---|---|---|---|---|---|---|---|---|---|---|---|---|---|
| 337 | | | | | 20390 | 255 | 20135 | | | | | 28028 | 313 | 27715 | 0.1024 | -0.2997 | Disorders of the autonomic nervous system | neurological | UricA | |
| 337.1 | | | | | 20203 | 68 | 20135 | | | | | 27793 | 78 | 27715 | | | Peripheral autonomic neuropathy | neurological | UricA | |
| 338 | 0.4269 | 0.03522 | 1.532 | 8.30e-34 | 24967 | 5863 | 19104 | 0.3387 | 0.03322 | 1.403 | 2.10e-24 | 33179 | 7691 | 25488 | 0.4136 | 0.03865 | Pain | neurological | UricA | |
| 338.1 | 0.373 | 0.0409 | 1.452 | 7.42e-20 | 23126 | 4022 | 19104 | 0.383 | 0.03899 | 1.467 | 8.91e-23 | 30282 | 4794 | 25488 | 0.008894 | 0.1439 | Acute pain | neurological | UricA | |
| 338.2 | 0.6133 | 0.04658 | 1.846 | 1.38e-39 | 21816 | 2712 | 19104 | 0.4582 | 0.04036 | 1.581 | 7.21e-30 | 29799 | 4311 | 25488 | 0.3245 | -0.05929 | Chronic pain | neurological | UricA | |
| 339 | 0.5912 | 0.08491 | 1.806 | 3.34e-12 | 25998 | 701 | 25297 | 0.6701 | 0.06005 | 1.955 | 6.40e-29 | 31803 | 1472 | 30331 | 0.05878 | 0.1881 | Other headache syndromes | neurological | UricA | |
| 340 | 0.3784 | 0.09042 | 1.46 | 2.85e-05 | 25980 | 683 | 25297 | 0.3652 | 0.04795 | 1.441 | 2.60e-14 | 33186 | 2855 | 30331 | 0.6387 | 0.04561 | Migraine | neurological | UricA | |
| 340.1 | | | | | 25434 | 137 | 25297 | 0.3674 | 0.08832 | 1.444 | 3.18e-05 | 31063 | 732 | 30331 | 0.8491 | 0.03855 | Migrain with aura | neurological | UricA | |
| 341 | | | | | 20276 | 141 | 20135 | | | | | 28206 | 491 | 27715 | 0.5453 | -0.1709 | Other demyelinating diseases of central nervous system | neurological | UricA | |
| 342 | | | | | 20493 | 358 | 20135 | | | | | 28011 | 296 | 27715 | 0.369 | 0.1563 | Hemiplegia | neurological | UricA | |
| 343 | | | | | 20279 | 144 | 20135 | | | | | 27844 | 129 | 27715 | | | Infantile cerebral palsy | neurological | UricA | |
| 344 | | | | | 20460 | 325 | 20135 | | | | | 27997 | 282 | 27715 | 3.02e-04 | 0.7168 | Other paralytic syndromes | neurological | UricA | |
| 345 | 0.2445 | 0.06419 | 1.277 | 1.40e-04 | 21729 | 1594 | 20135 | 0.3752 | 0.05814 | 1.455 | 1.10e-10 | 29581 | 1866 | 27715 | 0.01386 | 0.2062 | Epilepsy, recurrent seizures, convulsions | neurological | UricA | |
| 345.1 | 0.05654 | 0.101 | 1.058 | 0.5757 | 20847 | 712 | 20135 | 0.1127 | 0.09526 | 1.119 | 0.2367 | 28454 | 739 | 27715 | 0.1421 | 0.197 | Epilepsy | neurological | UricA | |
| 345.11 | | | | | 20451 | 316 | 20135 | | | | | 28000 | 285 | 27715 | 0.3689 | 0.1918 | Generalized convulsive epilepsy | neurological | UricA | |
| 345.12 | -0.1081 | 0.124 | 0.8975 | 0.383 | 20638 | 503 | 20135 | | | | | 28179 | 464 | 27715 | 0.05792 | 0.3173 | Partial epilepsy | neurological | UricA | |
| 345.3 | 0.3059 | 0.07047 | 1.358 | 1.42e-05 | 21418 | 1283 | 20135 | 0.4188 | 0.06404 | 1.52 | 6.21e-11 | 29200 | 1485 | 27715 | 0.03447 | 0.1946 | Convulsions | neurological | UricA | |

| phenotype | male_beta | male_SE | male_OR | male_p | male_n_total | male_n_cases | male_n_controls | female_beta | female_SE | female_OR | female_p | female_n_total | female_n_cases | female_n_controls | interaction_p | interaction_beta | description | group | lab | label |
|---|---|---|---|---|---|---|---|---|---|---|---|---|---|---|---|---|---|---|---|---|
| 346 | | | | | 20172 | 37 | 20135 | | | | | 27787 | 72 | 27715 | | | Abnormal findings on study of brain and/or nervous system | neurological | UricA | |
| 346.1 | | | | | 20298 | 163 | 20135 | | | | | 27984 | 269 | 27715 | | | Nonspecific abnormal findings on radiological and other examination of skull and head | neurological | UricA | |
| 346.2 | | | | | 20163 | 28 | 20135 | | | | | 27765 | 50 | 27715 | | | Nonspecific abnormal results of function study of brain and central nervous system | neurological | UricA | |
| 346.3 | | | | | 20173 | 38 | 20135 | | | | | 27758 | 43 | 27715 | | | Nonspecific abnormal findings in cerebrospinal fluid | neurological | UricA | |
| 347 | | | | | 20161 | 26 | 20135 | | | | | 27775 | 60 | 27715 | | | Cataplexy and narcolepsy | neurological | UricA | |
| 348 | 0.543 | 0.05967 | 1.721 | 9.06e-20 | 21644 | 1509 | 20135 | 0.6131 | 0.06077 | 1.846 | 6.22e-24 | 29223 | 1508 | 27715 | 0.1839 | 0.1099 | Other conditions of brain | neurological | UricA | |
| 348.2 | 0.3227 | 0.1068 | 1.381 | 0.002522 | 20647 | 512 | 20135 | 0.4188 | 0.1015 | 1.52 | 3.68e-05 | 28310 | 595 | 27715 | 0.3069 | 0.147 | Cerebral edema and compression of brain | neurological | UricA | |
| 348.4 | | | | | 20194 | 59 | 20135 | | | | | 27769 | 54 | 27715 | | | Cerebral cysts | neurological | UricA | |
| 348.7 | | | | | 20262 | 127 | 20135 | | | | | 27823 | 108 | 27715 | | | Coma | neurological | UricA | |
| 348.8 | 0.626 | 0.07229 | 1.87 | 4.71e-18 | 21082 | 947 | 20135 | 0.7781 | 0.07465 | 2.177 | 1.92e-25 | 28610 | 895 | 27715 | 0.05124 | 0.1963 | Encephalopathy, not elsewhere classified | neurological | UricA | |
| 348.9 | 0.3444 | 0.08586 | 1.411 | 6.04e-05 | 20916 | 781 | 20135 | 0.5439 | 0.0873 | 1.723 | 4.65e-10 | 28459 | 744 | 27715 | 0.02284 | 0.2713 | Other conditions of brain, NOS | neurological | UricA | |
| 349 | | | | | 20369 | 234 | 20135 | | | | | 27969 | 254 | 27715 | | | Other and unspecified disorders of the nervous system | neurological | UricA | |
| 350 | 0.3486 | 0.04589 | 1.417 | 3.04e-14 | 25783 | 2709 | 23074 | 0.3398 | 0.04288 | 1.405 | 2.26e-15 | 34411 | 3494 | 30917 | 0.4126 | 0.05009 | Abnormal movement | neurological | UricA | |
| 350.1 | 0.3685 | 0.07977 | 1.446 | 3.84e-06 | 23884 | 810 | 23074 | 0.3302 | 0.07145 | 1.391 | 3.80e-06 | 32013 | 1096 | 30917 | 0.5081 | 0.06874 | Abnormal involuntary movements | neurological | UricA | |

| phenotype | male_beta | male_SE | male_OR | male_p | male_n_total | male_n_cases | male_n_controls | female_beta | female_SE | female_OR | female_p | female_n_total | female_n_cases | female_n_controls | interaction_p | interaction_beta | description | group | lab | label |
|---|---|---|---|---|---|---|---|---|---|---|---|---|---|---|---|---|---|---|---|---|
| 350.2 | 0.3328 | 0.05474 | 1.395 | 1.20e-09 | 24868 | 1794 | 23074 | 0.3667 | 0.0509 | 1.443 | 5.86e-13 | 33227 | 2310 | 30917 | 0.2397 | 0.0855 | Abnormality of gait | neurological | UricA | |
| 350.3 | 0.05932 | 0.1027 | 1.061 | 0.5634 | 23605 | 531 | 23074 | -0.02175 | 0.08935 | 0.9785 | 0.8077 | 31754 | 837 | 30917 | 0.7723 | -0.03833 | Lack of coordination | neurological | UricA | |
| 350.5 | | | | | 23103 | 29 | 23074 | | | | | 30953 | 36 | 30917 | | | Abnormal reflex | neurological | UricA | |
| 350.6 | | | | | 23183 | 109 | 23074 | | | | | 31041 | 124 | 30917 | | | Disturbances of sensation of smell and taste | neurological | UricA | |
| 351 | 0.6823 | 0.06017 | 1.978 | 8.40e-30 | 25090 | 1362 | 23728 | 0.4665 | 0.05062 | 1.594 | 3.12e-20 | 32761 | 2244 | 30517 | 0.1504 | -0.1095 | Other peripheral nerve disorders | neurological | UricA | |
| 352 | | | | | 24026 | 298 | 23728 | 0.2644 | 0.1018 | 1.303 | 0.009376 | 31061 | 544 | 30517 | 0.05649 | 0.3134 | Disorders of other cranial nerves | neurological | UricA | |
| 352.1 | | | | | 23876 | 148 | 23728 | | | | | 30870 | 353 | 30517 | 0.08539 | 0.3983 | Trigeminal nerve disorders [CN5] | neurological | UricA | |
| 352.2 | | | | | 23861 | 133 | 23728 | | | | | 30696 | 179 | 30517 | | | Facial nerve disorders [CN7] | neurological | UricA | |
| 353 | | | | | 23928 | 200 | 23728 | | | | | 30746 | 229 | 30517 | | | Nerve root and plexus disorders | neurological | UricA | |
| 353.1 | | | | | 23805 | 77 | 23728 | | | | | 30605 | 88 | 30517 | | | Nerve plexus lesions | neurological | UricA | |
| 353.2 | | | | | 23800 | 72 | 23728 | | | | | 30603 | 86 | 30517 | | | Nerve root lesions | neurological | UricA | |
| 355 | | | | | 23869 | 141 | 23728 | | | | | 30825 | 308 | 30517 | | | Complex regional/central pain syndrome | neurological | UricA | |
| 355.1 | 0.4347 | 0.09084 | 1.545 | 1.70e-06 | 24316 | 588 | 23728 | 0.4556 | 0.07191 | 1.577 | 2.37e-10 | 31540 | 1023 | 30517 | 0.5001 | 0.07548 | Chronic pain syndrome | neurological | UricA | |
| 356 | 0.8038 | 0.06852 | 2.234 | 8.78e-32 | 24919 | 1003 | 23916 | 0.6309 | 0.07124 | 1.879 | 8.26e-19 | 33295 | 1002 | 32293 | 0.1645 | -0.1332 | Hereditary and idiopathic peripheral neuropathy | neurological | UricA | |
| 357 | 0.8655 | 0.0663 | 2.376 | 6.09e-39 | 24993 | 1077 | 23916 | 0.6481 | 0.063 | 1.912 | 8.07e-25 | 33576 | 1283 | 32293 | 0.008029 | -0.2352 | Inflammatory and toxic neuropathy | neurological | UricA | |
| 358 | | | | | 24025 | 109 | 23916 | | | | | 32462 | 169 | 32293 | | | Myoneural disorders | neurological | UricA | |
| 358.1 | | | | | 23987 | 71 | 23916 | | | | | 32384 | 91 | 32293 | | | Myasthenia gravis | neurological | UricA | |
| 359 | 0.7922 | 0.08492 | 2.208 | 1.07e-20 | 24562 | 646 | 23916 | 0.9459 | 0.08924 | 2.575 | 2.98e-26 | 32892 | 599 | 32293 | 0.1975 | 0.1538 | Muscular dystrophies and other myopathies | neurological | UricA | |
| 359.1 | | | | | 23949 | 33 | 23916 | | | | | 32328 | 35 | 32293 | | | Muscular dystrophies | neurological | UricA | |
| 359.2 | 0.8551 | 0.08617 | 2.352 | 3.28e-23 | 24538 | 622 | 23916 | 0.9906 | 0.09105 | 2.693 | 1.44e-27 | 32864 | 571 | 32293 | 0.2506 | 0.1395 | Myopathy | neurological | UricA | |
| 360 | | | | | 24749 | 89 | 24660 | | | | | 33075 | 87 | 32988 | | | Disorders of the globe | sense organs | UricA | |
| 360.2 | | | | | 24707 | 47 | 24660 | | | | | 33042 | 54 | 32988 | | | Progressive myopia | sense organs | UricA | |
| 360.3 | | | | | 24670 | 10 | 24660 | | | | | 33001 | 13 | 32988 | | | Hypotony of eye | sense organs | UricA | |
| 361 | | | | | 24928 | 268 | 24660 | | | | | 33163 | 175 | 32988 | | | Retinal detachments and defects | sense organs | UricA | |
| 361.1 | | | | | 24821 | 161 | 24660 | | | | | 33064 | 76 | 32988 | | | Retinal detachment with retinal defect | sense organs | UricA | |
| 361.2 | | | | | 24678 | 18 | 24660 | | | | | 32996 | 8 | 32988 | | | Retinoschisis and retinal cysts | sense organs | UricA | |
| 362 | 0.5083 | 0.06707 | 1.662 | 3.49e-14 | 25441 | 1087 | 24354 | 0.4888 | 0.06273 | 1.63 | 6.59e-15 | 34109 | 1379 | 32730 | 0.9505 | -0.005543 | Other retinal disorders | sense organs | UricA | |
| 362.1 | | | | | 24369 | 15 | 24354 | | | | | 32743 | 13 | 32730 | | | Retinopathy of prematurity | sense organs | UricA | |
| 362.2 | 0.3827 | 0.08876 | 1.466 | 1.63e-05 | 24974 | 620 | 24354 | 0.4113 | 0.08132 | 1.509 | 4.25e-07 | 33549 | 819 | 32730 | 0.8754 | 0.01828 | Degeneration of macula and posterior pole of retina | sense organs | UricA | |
| 362.21 | | | | | 24463 | 109 | 24354 | | | | | 32881 | 151 | 32730 | | | Macular degeneration, dry | sense organs | UricA | |
| 362.22 | | | | | 24410 | 56 | 24354 | | | | | 32819 | 89 | 32730 | | | Macular degeneration, wet | sense organs | UricA | |
| 362.23 | | | | | 24511 | 157 | 24354 | | | | | 32900 | 170 | 32730 | | | Cystoid macular degeneration of retina | sense organs | UricA | |
| 362.26 | | | | | 24590 | 236 | 24354 | | | | | 32963 | 233 | 32730 | | | Macular puckering of retina | sense organs | UricA | |
| 362.27 | | | | | 24433 | 79 | 24354 | | | | | 32875 | 145 | 32730 | | | Drusen (degenerative) of retina | sense organs | UricA | |
| 362.29 | | | | | 24520 | 166 | 24354 | | | | | 33004 | 274 | 32730 | | | Macular degeneration (senile) of retina NOS | sense organs | UricA | |
| 362.3 | | | | | 24535 | 181 | 24354 | | | | | 32906 | 176 | 32730 | | | Other nondiabetic retinopathy | sense organs | UricA | |
| 362.31 | | | | | 24393 | 39 | 24354 | | | | | 32753 | 23 | 32730 | | | Separation of retinal layers | sense organs | UricA | |
| 362.4 | | | | | 24528 | 174 | 24354 | | | | | 32939 | 209 | 32730 | | | Retinal vascular changes and abnormalities | sense organs | UricA | |
| 362.5 | | | | | 24369 | 15 | 24354 | | | | | 32823 | 93 | 32730 | | | Toxic maculopathy of retina | sense organs | UricA | |
| 362.6 | | | | | 24414 | 60 | 24354 | | | | | 32796 | 66 | 32730 | | | Peripheral retinal degenerations | sense organs | UricA | |

| phenotype | male_beta | male_SE | male_OR | male_p | male_n_total | male_n_cases | male_n_controls | female_beta | female_SE | female_OR | female_p | female_n_total | female_n_cases | female_n_controls | interaction_p | interaction_beta | description | group | lab | label |
|---|---|---|---|---|---|---|---|---|---|---|---|---|---|---|---|---|---|---|---|---|
| 362.7 | | | | | 24401 | 47 | 24354 | | | | | 32775 | 45 | 32730 | | | Hereditary retinal dystrophies | sense organs | UricA | |
| 362.8 | | | | | 24401 | 47 | 24354 | | | | | 32774 | 44 | 32730 | | | Retinal hemorrhage/ischemia | sense organs | UricA | |
| 362.9 | | | | | 24469 | 115 | 24354 | | | | | 32862 | 132 | 32730 | | | Retinal edema | sense organs | UricA | |
| 363 | | | | | 24770 | 110 | 24660 | | | | | 33134 | 146 | 32988 | | | Chorioretinal inflammations, scars, and other disorders of choroid | sense organs | UricA | |
| 363.3 | | | | | 24721 | 61 | 24660 | | | | | 33072 | 84 | 32988 | | | Chorioretinal scars | sense organs | UricA | |
| 363.4 | | | | | 24662 | 2 | 24660 | | | | | 32995 | 7 | 32988 | | | Choroidal degenerations | sense organs | UricA | |
| phenotype | male_beta | male_SE | male_OR | male_p | male_n_total | male_n_cases | male_n_controls | female_beta | female_SE | female_OR | female_p | female_n_total | female_n_cases | female_n_controls | interaction_p | interaction_beta | description | group | lab | label |
| 364 | | | | | 24841 | 181 | 24660 | | | | | 33258 | 270 | 32988 | | | Corneal opacity and other disorders of cornea | sense organs | UricA | |
| 364.1 | | | | | 24753 | 93 | 24660 | | | | | 33113 | 125 | 32988 | | | Corneal opacity | sense organs | UricA | |
| 364.2 | | | | | 24712 | 52 | 24660 | | | | | 33031 | 43 | 32988 | | | Corneal edema | sense organs | UricA | |
| 364.4 | | | | | 24715 | 55 | 24660 | | | | | 33057 | 69 | 32988 | | | Corneal degenerations | sense organs | UricA | |
| 364.41 | | | | | 24684 | 24 | 24660 | | | | | 33006 | 18 | 32988 | | | Keratoconus | sense organs | UricA | |
| 364.5 | | | | | 24720 | 60 | 24660 | | | | | 33111 | 123 | 32988 | | | Corneal dystrophy | sense organs | UricA | |
| 364.51 | | | | | 24699 | 39 | 24660 | | | | | 33061 | 73 | 32988 | | | Fuchs' dystrophy | sense organs | UricA | |
| 364.9 | | | | | 24688 | 28 | 24660 | | | | | 33020 | 32 | 32988 | | | Cornea replaced by transplant | sense organs | UricA | |
| 365 | 0.348 | 0.07338 | 1.416 | 2.12e-06 | 25576 | 916 | 24660 | 0.227 | 0.0706 | 1.255 | 0.001306 | 34125 | 1137 | 32988 | 0.2852 | -0.1057 | Glaucoma | sense organs | UricA | |
| 365.1 | 0.3667 | 0.09222 | 1.443 | 6.99e-05 | 25218 | 558 | 24660 | 0.183 | 0.08994 | 1.201 | 0.04185 | 33666 | 678 | 32988 | 0.1775 | -0.1686 | Open-angle glaucoma | sense organs | UricA | |
| 365.11 | | | | | 24878 | 218 | 24660 | | | | | 33217 | 229 | 32988 | | | Primary open angle glaucoma | sense organs | UricA | |
| 365.2 | | | | | 24753 | 93 | 24660 | | | | | 33132 | 144 | 32988 | | | Primary angle-closure glaucoma | sense organs | UricA | |
| 365.5 | | | | | 24671 | 11 | 24660 | | | | | 33004 | 16 | 32988 | | | Pseudoexfoliation glaucoma | sense organs | UricA | |
| 366 | 0.3648 | 0.04604 | 1.44 | 2.29e-15 | 26713 | 2883 | 23830 | 0.3522 | 0.04367 | 1.422 | 7.31e-16 | 35649 | 3722 | 31927 | 0.823 | -0.01392 | Cataract | sense organs | UricA | |
| 366.1 | | | | | 23845 | 15 | 23830 | | | | | 31936 | 9 | 31927 | | | Nonsenile Cataract | sense organs | UricA | |
| 366.2 | 0.3507 | 0.04894 | 1.42 | 7.68e-13 | 26293 | 2463 | 23830 | 0.3252 | 0.04649 | 1.384 | 2.64e-12 | 35061 | 3134 | 31927 | 0.723 | -0.02344 | Senile cataract | sense organs | UricA | |
| 366.3 | | | | | 23835 | 5 | 23830 | | | | | 31932 | 5 | 31927 | | | Traumatic cataract | sense organs | UricA | |
| 367 | 0.3592 | 0.06568 | 1.432 | 4.52e-08 | 26602 | 1195 | 25407 | 0.1971 | 0.05397 | 1.218 | 2.59e-04 | 35278 | 2096 | 33182 | 0.03979 | -0.1687 | Disorders of refraction and accommodation; blindness and low vision | sense organs | UricA | |
| 367.1 | 0.2911 | 0.09469 | 1.338 | 0.002108 | 25991 | 584 | 25407 | 0.1354 | 0.07 | 1.145 | 0.05307 | 34437 | 1255 | 33182 | 0.09649 | -0.1877 | Myopia | sense organs | UricA | |
| 367.2 | | | | | 25731 | 324 | 25407 | 0.07933 | 0.1042 | 1.083 | 0.4465 | 33701 | 519 | 33182 | 0.08407 | -0.266 | Astigmatism | sense organs | UricA | |
| 367.8 | | | | | 25506 | 99 | 25407 | | | | | 33431 | 249 | 33182 | | | Hypermetropia | sense organs | UricA | |
| 367.9 | | | | | 25707 | 300 | 25407 | | | | | 33524 | 342 | 33182 | 0.5273 | 0.1048 | Blindness and low vision | sense organs | UricA | |
| 368 | 0.4483 | 0.0705 | 1.566 | 2.04e-10 | 26249 | 990 | 25259 | 0.4069 | 0.06089 | 1.502 | 2.35e-11 | 34737 | 1481 | 33256 | 0.5475 | -0.05417 | Visual disturbances | sense organs | UricA | |
| 368.1 | | | | | 25311 | 52 | 25259 | | | | | 33326 | 70 | 33256 | | | Amblyopia | sense organs | UricA | |
| 368.2 | | | | | 25592 | 333 | 25259 | | | | | 33627 | 371 | 33256 | 0.06143 | 0.3043 | Diplopia and disorders of binocular vision | sense organs | UricA | |
| 368.3 | | | | | 25277 | 18 | 25259 | | | | | 33272 | 16 | 33256 | | | Anisometropia | sense organs | UricA | |
| 368.4 | | | | | 25410 | 151 | 25259 | | | | | 33464 | 208 | 33256 | | | Visual field defects | sense organs | UricA | |
| 368.5 | | | | | 25262 | 3 | 25259 | | | | | 33257 | 1 | 33256 | | | Color vision deficiencies | sense organs | UricA | |
| 368.7 | | | | | 25259 | | | | | | | 33257 | 1 | 33256 | | | Disorders of accommodation | sense organs | UricA | |
| 368.9 | | | | | 25451 | 192 | 25259 | | | | | 33633 | 377 | 33256 | 0.3774 | -0.165 | Subjective visual disturbances | sense organs | UricA | |
| 368.91 | | | | | 25299 | 40 | 25259 | | | | | 33323 | 67 | 33256 | | | Psychophysical visual disturbances | sense organs | UricA | |
| 369 | 0.2217 | 0.09847 | 1.248 | 0.02436 | 24317 | 512 | 23805 | 0.3305 | 0.0769 | 1.392 | 1.73e-05 | 31511 | 916 | 30595 | 0.3632 | 0.1099 | Infection of the eye | sense organs | UricA | |
| 369.2 | | | | | 23885 | 80 | 23805 | | | | | 30724 | 129 | 30595 | | | Eye infection, viral | sense organs | UricA | |
| 369.5 | | | | | 24187 | 382 | 23805 | 0.3679 | 0.08539 | 1.445 | 1.64e-05 | 31314 | 719 | 30595 | 0.3019 | 0.1414 | Conjunctivitis, infectious | sense organs | UricA | |

| phenotype | male_beta | male_SE | male_OR | male_p | male_n_total | male_n_cases | male_n_controls | female_beta | female_SE | female_OR | female_p | female_n_total | female_n_cases | female_n_controls | interaction_p | interaction_beta | description | group | lab | label |
|---|---|---|---|---|---|---|---|---|---|---|---|---|---|---|---|---|---|---|---|---|
| 370 | | | | | 24074 | 269 | 23805 | 0.4293 | 0.08717 | 1.536 | 8.46e-07 | 31271 | 676 | 30595 | 0.3139 | 0.1545 | Keratitis | sense organs | UricA | |
| 370.1 | | | | | 23880 | 75 | 23805 | | | | | 30694 | 99 | 30595 | | | Corneal ulcer | sense organs | UricA | |
| 370.2 | | | | | 23830 | 25 | 23805 | | | | | 30656 | 61 | 30595 | | | Superficial keratitis | sense organs | UricA | |
| 370.3 | | | | | 23964 | 159 | 23805 | | | | | 31063 | 468 | 30595 | 0.3807 | 0.1698 | Keratoconjunctivitis | sense organs | UricA | |
| 370.31 | | | | | 23923 | 118 | 23805 | | | | | 31009 | 414 | 30595 | 0.8434 | -0.04277 | Keratoconjunctivitis sicca | sense organs | UricA | |
| 371 | 0.2605 | 0.07238 | 1.298 | 3.19e-04 | 24762 | 957 | 23805 | 0.2827 | 0.06105 | 1.327 | 3.64e-06 | 32133 | 1538 | 30595 | 0.9971 | -3.29e-04 | Inflammation of the eye | sense organs | UricA | |
| 371.1 | | | | | 23925 | 120 | 23805 | | | | | 30815 | 220 | 30595 | | | Uveitis, noninfectious or NOS | sense organs | UricA | |
| 371.2 | | | | | 24253 | 448 | 23805 | 0.1842 | 0.08485 | 1.202 | 0.02993 | 31368 | 773 | 30595 | 0.3283 | -0.1266 | Conjunctivitis, noninfectious | sense organs | UricA | |
| 371.21 | | | | | 24201 | 396 | 23805 | 0.1728 | 0.09004 | 1.189 | 0.0549 | 31279 | 684 | 30595 | 0.2803 | -0.1482 | Allergic conjunctivitis | sense organs | UricA | |
| 371.3 | | | | | 24165 | 360 | 23805 | 0.4433 | 0.09442 | 1.558 | 2.67e-06 | 31154 | 559 | 30595 | 0.2031 | 0.1825 | Inflammation of eyelids | sense organs | UricA | |
| 371.33 | | | | | 23808 | 3 | 23805 | | | | | 30603 | 8 | 30595 | | | Noninfectious dermatoses of eyelid | sense organs | UricA | |
| 371.9 | | | | | 23817 | 12 | 23805 | | | | | 30607 | 12 | 30595 | | | Chronic inflammatory disorders of orbit | sense organs | UricA | |
| 372 | | | | | 23933 | 128 | 23805 | | | | | 30728 | 133 | 30595 | | | Disorders of conjunctiva | sense organs | UricA | |
| 374 | -0.01044 | 0.0826 | 0.9896 | 0.8994 | 24581 | 776 | 23805 | 0.1538 | 0.07426 | 1.166 | 0.03829 | 31646 | 1051 | 30595 | 0.1293 | 0.1635 | Other disorders of eyelids | sense organs | UricA | |
| 374.1 | | | | | 23920 | 115 | 23805 | | | | | 30673 | 78 | 30595 | | | Ectropion or entropion | sense organs | UricA | |
| 374.2 | | | | | 23850 | 45 | 23805 | | | | | 30619 | 24 | 30595 | | | Lagophthalmos | sense organs | UricA | |
| 374.3 | | | | | 24001 | 196 | 23805 | | | | | 30970 | 375 | 30595 | 0.243 | -0.2187 | Ptosis of eyelid | sense organs | UricA | |
| 374.6 | | | | | 24250 | 445 | 23805 | 0.1083 | 0.09555 | 1.114 | 0.257 | 31205 | 610 | 30595 | 0.1468 | 0.2033 | Dermatochalasis | sense organs | UricA | |
| 375 | | | | | 23884 | 86 | 23798 | | | | | 31096 | 146 | 30950 | | | Disorders of lacrimal system | sense organs | UricA | |
| 375.1 | 0.121 | 0.07029 | 1.129 | 0.08516 | 24860 | 1062 | 23798 | 0.1925 | 0.05577 | 1.212 | 5.59e-04 | 32921 | 1971 | 30950 | 0.5969 | 0.04613 | Dry eyes | sense organs | UricA | |
| 375.2 | | | | | 23848 | 50 | 23798 | | | | | 31036 | 86 | 30950 | | | Epiphora | sense organs | UricA | |
| 376 | | | | | 23842 | 44 | 23798 | | | | | 30982 | 32 | 30950 | | | Disorders of the orbit | sense organs | UricA | |
| 377 | | | | | 24004 | 206 | 23798 | | | | | 31323 | 373 | 30950 | 0.374 | -0.1725 | Disorders of optic nerve and visual pathways | sense organs | UricA | |
| 377.1 | | | | | 23872 | 74 | 23798 | | | | | 31057 | 107 | 30950 | | | Optic atrophy | sense organs | UricA | |
| 377.3 | | | | | 23899 | 101 | 23798 | | | | | 31129 | 179 | 30950 | | | Optic neuritis/neuropathy | sense organs | UricA | |
| 378 | | | | | 24117 | 319 | 23798 | | | | | 31299 | 349 | 30950 | 0.4027 | -0.1555 | Strabismus and other disorders of binocular eye movements | sense organs | UricA | |

| phenotype | male_beta | male_SE | male_OR | male_p | male_n_total | male_n_cases | male_n_controls | female_beta | female_SE | female_OR | female_p | female_n_total | female_n_cases | female_n_controls | interaction_p | interaction_beta | description | group | lab | label |
|---|---|---|---|---|---|---|---|---|---|---|---|---|---|---|---|---|---|---|---|---|
| 378.1 | | | | | 23973 | 175 | 23798 | | | | | 31136 | 186 | 30950 | | | Strabismus (not specified as paralytic) | sense organs | UricA | |
| 378.2 | | | | | 23835 | 37 | 23798 | | | | | 31001 | 51 | 30950 | | | Nystagmus and other irregular eye movements | sense organs | UricA | |
| 378.5 | | | | | 23889 | 91 | 23798 | | | | | 31030 | 80 | 30950 | | | Paralytic strabismus | sense organs | UricA | |
| 379 | 0.3466 | 0.06619 | 1.414 | 1.64e-07 | 24956 | 1158 | 23798 | 0.2656 | 0.06003 | 1.304 | 9.65e-06 | 32559 | 1609 | 30950 | 0.3573 | -0.07997 | Other disorders of eye | sense organs | UricA | |
| 379.1 | | | | | 23846 | 48 | 23798 | | | | | 31085 | 135 | 30950 | | | Scleritis and episcleritis | sense organs | UricA | |
| 379.2 | 0.3396 | 0.072 | 1.404 | 2.41e-06 | 24762 | 964 | 23798 | 0.218 | 0.06612 | 1.244 | 9.77e-04 | 32269 | 1319 | 30950 | 0.2567 | -0.1078 | Disorders of vitreous body | sense organs | UricA | |
| 379.3 | | | | | 23855 | 57 | 23798 | | | | | 30995 | 45 | 30950 | | | Aphakia and other disorders of lens | sense organs | UricA | |
| 379.4 | | | | | 23835 | 37 | 23798 | | | | | 31014 | 64 | 30950 | | | Anomalies of pupillary function | sense organs | UricA | |
| 379.5 | | | | | 23914 | 116 | 23798 | | | | | 31022 | 72 | 30950 | | | Disorders of iris and ciliary body | sense organs | UricA | |
| 379.51 | | | | | 23816 | 18 | 23798 | | | | | 30958 | 8 | 30950 | | | Pigmentary iris degeneration | sense organs | UricA | |
| 379.9 | | | | | 23893 | 95 | 23798 | | | | | 31121 | 171 | 30950 | | | Pain, swelling or discharge of eye | sense organs | UricA | |
| 380 | | | | | 25280 | 65 | 25215 | | | | | 34053 | 71 | 33982 | | | Disorders of external ear | sense organs | UricA | |
| 380.1 | | | | | 25546 | 331 | 25215 | | | | | 34343 | 361 | 33982 | 0.2593 | -0.1853 | Otitis externa | sense organs | UricA | |

| phenotype | male_beta | male_SE | male_OR | male_p | male_n_total | male_n_cases | male_n_controls | female_beta | female_SE | female_OR | female_p | female_n_total | female_n_cases | female_n_controls | interaction_p | interaction_beta | description | group | lab | label |
|---|---|---|---|---|---|---|---|---|---|---|---|---|---|---|---|---|---|---|---|---|
| 380.4 | 0.1446 | 0.07347 | 1.156 | 0.04904 | 26150 | 935 | 25215 | -0.01522 | 0.0783 | 0.9849 | 0.8459 | 34929 | 947 | 33982 | 0.1398 | -0.1537 | Impacted cerumen | sense organs | UricA | |
| 381 | 0.3182 | 0.06746 | 1.375 | 2.39e-06 | 25736 | 1209 | 24527 | 0.1067 | 0.05472 | 1.113 | 0.05114 | 33271 | 2155 | 31116 | 0.1052 | -0.1353 | Otitis media and Eustachian tube disorders | sense organs | UricA | |
| 381.1 | 0.2708 | 0.0952 | 1.311 | 0.004449 | 25160 | 633 | 24527 | 0.2677 | 0.07226 | 1.307 | 2.11e-04 | 32235 | 1119 | 31116 | 0.4483 | 0.0865 | Otitis media | sense organs | UricA | |
| 381.11 | | | | | 24939 | 412 | 24527 | 0.3357 | 0.08778 | 1.399 | 1.31e-04 | 31847 | 731 | 31116 | 0.1158 | 0.2243 | Suppurative and unspecified otitis media | sense organs | UricA | |
| 381.2 | 0.3296 | 0.08933 | 1.39 | 2.25e-04 | 25149 | 622 | 24527 | 0.0508 | 0.07333 | 1.052 | 0.4885 | 32238 | 1122 | 31116 | 0.07066 | -0.2016 | Eustachian tube disorders | sense organs | UricA | |
| 381.3 | | | | | 24579 | 52 | 24527 | | | | | 31179 | 63 | 31116 | | | Mastoiditis & related conditions | sense organs | UricA | |
| 381.9 | | | | | 24645 | 118 | 24527 | | | | | 31231 | 115 | 31116 | | | Otorrhea | sense organs | UricA | |
| 382 | | | | | 24849 | 322 | 24527 | 0.09364 | 0.08442 | 1.098 | 0.2674 | 31925 | 809 | 31116 | 0.04199 | -0.2882 | Otalgia | sense organs | UricA | |
| 383 | | | | | 24540 | 13 | 24527 | | | | | 31138 | 22 | 31116 | | | Otosclerosis | sense organs | UricA | |
| 384 | | | | | 24691 | 164 | 24527 | | | | | 31314 | 198 | 31116 | | | Other disorders of tympanic membrane | sense organs | UricA | |
| 384.1 | | | | | 24551 | 24 | 24527 | | | | | 31124 | 8 | 31116 | | | Myringitis | sense organs | UricA | |
| 384.4 | | | | | 24667 | 140 | 24527 | | | | | 31294 | 178 | 31116 | | | Perforation of tympanic membrane | sense organs | UricA | |
| 385 | | | | | 24625 | 98 | 24527 | | | | | 31245 | 129 | 31116 | | | Other disorders of middle ear and mastoid | sense organs | UricA | |
| 385.3 | | | | | 24606 | 79 | 24527 | | | | | 31204 | 88 | 31116 | | | Cholesteatoma | sense organs | UricA | |
| 385.5 | | | | | 24556 | 29 | 24527 | | | | | 31165 | 49 | 31116 | | | Tympanosclerosis and middle ear disease related to otitis media | sense organs | UricA | |
| 386 | | | | | 23483 | 489 | 22994 | 0.3811 | 0.07463 | 1.464 | 3.28e-07 | 30272 | 992 | 29280 | 0.204 | 0.154 | Vertiginous syndromes and other disorders of vestibular system | sense organs | UricA | |
| 386.1 | | | | | 23077 | 83 | 22994 | | | | | 29457 | 177 | 29280 | | | Meniere's disease | sense organs | UricA | |
| 386.2 | | | | | 23328 | 334 | 22994 | 0.3572 | 0.08917 | 1.429 | 6.17e-05 | 29955 | 675 | 29280 | 0.4857 | 0.1007 | Peripheral or central vertigo | sense organs | UricA | |
| 386.21 | | | | | 23041 | 47 | 22994 | | | | | 29385 | 105 | 29280 | | | Central origin vertigo | sense organs | UricA | |
| 386.3 | | | | | 23038 | 44 | 22994 | | | | | 29344 | 64 | 29280 | | | Labyrinthitis | sense organs | UricA | |
| 386.9 | 0.4876 | 0.04953 | 1.628 | 7.26e-23 | 25211 | 2217 | 22994 | 0.5204 | 0.04185 | 1.683 | 1.71e-35 | 32967 | 3687 | 29280 | 0.4496 | 0.04794 | Dizziness and giddiness (Light-headedness and vertigo) | sense organs | UricA | |
| 388 | | | | | 23411 | 34 | 23377 | | | | | 32030 | 47 | 31983 | | | Other disorders of ear | sense organs | UricA | |
| 389 | 0.2791 | 0.04693 | 1.322 | 2.74e-09 | 26031 | 2654 | 23377 | 0.1939 | 0.04668 | 1.214 | 3.26e-05 | 34938 | 2955 | 31983 | 0.173 | -0.08803 | Hearing loss | sense organs | UricA | |
| 389.1 | 0.2694 | 0.05785 | 1.309 | 3.22e-06 | 25007 | 1630 | 23377 | 0.2905 | 0.05854 | 1.337 | 6.99e-07 | 33675 | 1692 | 31983 | 0.9408 | 0.005961 | Sensorineural hearing loss | sense organs | UricA | |
| 389.2 | | | | | 23633 | 256 | 23377 | | | | | 32346 | 363 | 31983 | 0.6306 | 0.091 | Conductive hearing loss | sense organs | UricA | |
| 389.3 | | | | | 23398 | 21 | 23377 | | | | | 31995 | 12 | 31983 | | | Degenerative and vascular disorders of ear | sense organs | UricA | |
| 389.4 | 0.2775 | 0.0898 | 1.32 | 0.001998 | 23983 | 606 | 23377 | 0.09288 | 0.08822 | 1.097 | 0.2924 | 32714 | 731 | 31983 | 0.2331 | -0.146 | Tinnitus | sense organs | UricA | |
| 389.5 | | | | | 23394 | 17 | 23377 | | | | | 32002 | 19 | 31983 | | | Disorders of acoustic nerve | sense organs | UricA | |
| 394 | 0.7442 | 0.06075 | 2.105 | 1.65e-34 | 22330 | 1378 | 20952 | 0.8412 | 0.06018 | 2.319 | 2.14e-44 | 31197 | 1468 | 29729 | 0.4088 | 0.06881 | Rheumatic disease of the heart valves | circulatory system | UricA | |
| 394.1 | | | | | 21305 | 353 | 20952 | | | | | 30068 | 339 | 29729 | 0.6884 | -0.06533 | Mitral valve stenosis and aortic valve stenosis | circulatory system | UricA | |
| 394.2 | | | | | 21121 | 169 | 20952 | | | | | 29965 | 236 | 29729 | | | Mitral valve disease | circulatory system | UricA | |
| 394.3 | | | | | 20969 | 17 | 20952 | | | | | 29742 | 13 | 29729 | | | Aortic valve disease | circulatory system | UricA | |
| 394.4 | | | | | 20972 | 20 | 20952 | | | | | 29771 | 42 | 29729 | | | Acute rheumatic heart disease | circulatory system | UricA | |
| 394.7 | 1.14 | 0.09403 | 3.127 | 7.84e-34 | 21461 | 509 | 20952 | 1.027 | 0.0869 | 2.792 | 3.15e-32 | 30357 | 628 | 29729 | 0.5199 | -0.07992 | Disease of tricuspid valve | circulatory system | UricA | |
| 395 | 0.6287 | 0.04221 | 1.875 | 3.51e-50 | 24383 | 3431 | 20952 | 0.7218 | 0.04456 | 2.058 | 4.98e-59 | 32852 | 3123 | 29729 | 0.1852 | 0.0793 | Heart valve disorders | circulatory system | UricA | |

| phenotype | male_beta | male_SE | male_OR | male_p | male_n_total | male_n_cases | male_n_controls | female_beta | female_SE | female_OR | female_p | female_n_total | female_n_cases | female_n_controls | interaction_p | interaction_beta | description | group | lab | label |
|---|---|---|---|---|---|---|---|---|---|---|---|---|---|---|---|---|---|---|---|---|
| 395.1 | 0.8101 | 0.05372 | 2.248 | 2.21e-51 | 22776 | 1824 | 20952 | 0.7679 | 0.05425 | 2.155 | 1.74e-45 | 31647 | 1918 | 29729 | 0.5265 | -0.0471 | Nonrheumatic mitral valve disorders | circulatory system | UricA | |
| 395.2 | 0.4314 | 0.05616 | 1.539 | 1.56e-14 | 22773 | 1821 | 20952 | 0.6552 | 0.06327 | 1.926 | 3.93e-25 | 31147 | 1418 | 29729 | 0.02049 | 0.1902 | Nonrheumatic aortic valve disorders | circulatory system | UricA | |
| 395.3 | | | | | 21269 | 317 | 20952 | | | | | 30053 | 324 | 29729 | 0.6809 | 0.06615 | Nonrheumatic tricuspid valve disorders | circulatory system | UricA | |
| 395.4 | | | | | 21053 | 101 | 20952 | | | | | 29855 | 126 | 29729 | | | Nonrheumatic pulmonary valve disorders | circulatory system | UricA | |
| 395.6 | 0.3542 | 0.07802 | 1.425 | 5.62e-06 | 21815 | 863 | 20952 | 0.7493 | 0.09082 | 2.115 | 1.58e-16 | 30347 | 618 | 29729 | 0.009109 | 0.3029 | Heart valve replaced | circulatory system | UricA | |
| 396 | 0.5776 | 0.09092 | 1.782 | 2.11e-10 | 21534 | 582 | 20952 | 0.5936 | 0.07579 | 1.811 | 4.79e-15 | 30632 | 903 | 29729 | 0.5106 | 0.07539 | Abnormal heart sounds | circulatory system | UricA | |
| 401 | 0.8724 | 0.03905 | 2.393 | 1.61e-110 | 25158 | 14670 | 10488 | 0.99 | 0.03538 | 2.691 | 2.47e-172 | 33895 | 14782 | 19113 | 0.1074 | 0.08418 | Hypertension | circulatory system | UricA | |
| 401.1 | 0.877 | 0.03929 | 2.404 | 2.28e-110 | 24789 | 14301 | 10488 | 0.9901 | 0.03558 | 2.691 | 2.27e-170 | 33585 | 14472 | 19113 | 0.1274 | 0.08019 | Essential hypertension | circulatory system | UricA | |
| 401.2 | 1.703 | 0.05148 | 5.492 | 4.61e-240 | 14837 | 4349 | 10488 | 1.812 | 0.05295 | 6.12 | 1.46e-256 | 22464 | 3351 | 19113 | 0.7536 | 0.02276 | Hypertensive heart and/or renal disease | circulatory system | UricA | |
| 401.21 | 1.518 | 0.06444 | 4.561 | 1.29e-122 | 12798 | 2310 | 10488 | 1.635 | 0.06552 | 5.127 | 2.31e-137 | 21086 | 1973 | 19113 | 0.8065 | 0.02217 | Hypertensive heart disease | circulatory system | UricA | |
| 401.22 | 2.011 | 0.05827 | 7.47 | 5.72e-261 | 12900 | 2412 | 10488 | 2.16 | 0.06415 | 8.669 | 1.79e-248 | 20743 | 1630 | 19113 | 0.4113 | 0.06953 | Hypertensive chronic kidney disease | circulatory system | UricA | |
| 401.3 | 2.028 | 0.08332 | 7.597 | 7.61e-131 | 11324 | 836 | 10488 | 1.947 | 0.07659 | 7.007 | 1.59e-142 | 20074 | 961 | 19113 | 0.3279 | -0.1091 | Other hypertensive complications | circulatory system | UricA | |
| phenotype | male_beta | male_SE | male_OR | male_p | male_n_total | male_n_cases | male_n_controls | female_beta | female_SE | female_OR | female_p | female_n_total | female_n_cases | female_n_controls | interaction_p | interaction_beta | description | group | lab | label |
| 411 | 0.6068 | 0.03555 | 1.835 | 2.43e-65 | 24924 | 8227 | 16697 | 0.6806 | 0.03963 | 1.975 | 4.20e-66 | 33484 | 5060 | 28424 | 0.1908 | 0.06817 | Ischemic Heart Disease | circulatory system | UricA | |
| 411.1 | 0.7125 | 0.06273 | 2.039 | 6.79e-30 | 18153 | 1456 | 16697 | 0.8513 | 0.08141 | 2.343 | 1.35e-25 | 29191 | 767 | 28424 | 0.4131 | 0.0821 | Unstable angina (intermediate coronary syndrome) | circulatory system | UricA | |
| 411.2 | 0.7579 | 0.04731 | 2.134 | 9.50e-58 | 19713 | 3016 | 16697 | 0.8964 | 0.05834 | 2.451 | 2.84e-53 | 30089 | 1665 | 28424 | 0.1635 | 0.1021 | Myocardial infarction | circulatory system | UricA | |
| 411.3 | 0.6638 | 0.05958 | 1.942 | 7.97e-29 | 18373 | 1676 | 16697 | 0.6818 | 0.07262 | 1.977 | 6.08e-21 | 29445 | 1021 | 28424 | 0.7569 | -0.02838 | Angina pectoris | circulatory system | UricA | |
| 411.4 | 0.5989 | 0.0368 | 1.82 | 1.45e-59 | 24205 | 7508 | 16697 | 0.6745 | 0.04281 | 1.963 | 6.34e-56 | 32583 | 4159 | 28424 | 0.2541 | 0.06299 | Coronary atherosclerosis | circulatory system | UricA | |
| 411.41 | | | | | 16754 | 57 | 16697 | | | | | 28492 | 68 | 28424 | | | Aneurysm and dissection of heart | circulatory system | UricA | |
| 411.8 | 0.9656 | 0.04932 | 2.626 | 2.38e-85 | 19412 | 2715 | 16697 | 0.9943 | 0.07069 | 2.703 | 6.16e-45 | 29458 | 1034 | 28424 | 0.8523 | 0.0157 | Other chronic ischemic heart disease, unspecified | circulatory system | UricA | |
| 411.9 | | | | | 16791 | 94 | 16697 | | | | | 28492 | 68 | 28424 | | | Other acute and subacute forms of ischemic heart disease | circulatory system | UricA | |
| 414 | 0.8297 | 0.05249 | 2.293 | 2.76e-56 | 18860 | 2163 | 16697 | 0.9937 | 0.05981 | 2.701 | 5.55e-62 | 29887 | 1463 | 28424 | 0.067 | 0.1426 | Other forms of chronic heart disease | circulatory system | UricA | |
| 414.2 | | | | | 16930 | 233 | 16697 | | | | | 28541 | 117 | 28424 | | | ASCVD | circulatory system | UricA | |
| 415 | 1.092 | 0.05072 | 2.98 | 8.50e-103 | 22610 | 2006 | 20604 | 0.948 | 0.05209 | 2.581 | 5.13e-74 | 32928 | 1974 | 30954 | 0.09257 | -0.1184 | Pulmonary heart disease | circulatory system | UricA | |
| 415.1 | 0.7249 | 0.07649 | 2.064 | 2.62e-21 | 21476 | 872 | 20604 | 0.7586 | 0.08068 | 2.135 | 5.37e-21 | 31762 | 808 | 30954 | 0.6676 | 0.04615 | Acute pulmonary heart disease | circulatory system | UricA | |
| 415.11 | 0.6758 | 0.08228 | 1.966 | 2.15e-16 | 21366 | 762 | 20604 | 0.7074 | 0.08419 | 2.029 | 4.35e-17 | 31710 | 756 | 30954 | 0.6624 | 0.04967 | Pulmonary embolism and infarction, acute | circulatory system | UricA | |
| 415.2 | 1.536 | 0.06729 | 4.647 | 2.30e-115 | 21649 | 1045 | 20604 | 1.2 | 0.06621 | 3.32 | 1.97e-73 | 32069 | 1115 | 30954 | 5.80e-04 | -0.3141 | Chronic pulmonary heart disease | circulatory system | UricA | |
| 415.21 | | | | | 20763 | 159 | 20604 | | | | | 31201 | 247 | 30954 | | | Primary pulmonary hypertension | circulatory system | UricA | |
| 416 | 0.983 | 0.04352 | 2.673 | 5.68e-113 | 23564 | 2960 | 20604 | 1.112 | 0.05054 | 3.041 | 2.45e-107 | 32994 | 2040 | 30954 | 0.01375 | 0.1592 | Cardiomegaly | circulatory system | UricA | |

| phenotype | male_beta | male_SE | male_OR | male_p | male_n_total | male_n_cases | male_n_controls | female_beta | female_SE | female_OR | female_p | female_n_total | female_n_cases | female_n_controls | interaction_p | interaction_beta | description | group | lab | label |
|---|---|---|---|---|---|---|---|---|---|---|---|---|---|---|---|---|---|---|---|---|
| 418 | 0.7042 | 0.03424 | 2.022 | 5.67e-94 | 23574 | 7662 | 15912 | 0.6757 | 0.03309 | 1.965 | 1.18e-92 | 31720 | 9073 | 22647 | 0.6883 | 0.01888 | Nonspecific chest pain | circulatory system | UricA | |
| 418.1 | 1.078 | 0.0634 | 2.939 | 7.31e-65 | 17223 | 1311 | 15912 | 1.121 | 0.0614 | 3.067 | 1.93e-74 | 24061 | 1414 | 22647 | 0.3435 | 0.0823 | Precordial pain | circulatory system | UricA | |
| 420 | 0.9645 | 0.07143 | 2.623 | 1.53e-41 | 24630 | 934 | 23696 | 1.04 | 0.07243 | 2.829 | 9.68e-47 | 34594 | 895 | 33699 | 0.1672 | 0.1356 | Carditis | circulatory system | UricA | |
| 420.1 | | | | | 23727 | 31 | 23696 | | | | | 33725 | 26 | 33699 | | | Myocarditis | circulatory system | UricA | |
| 420.2 | 1.052 | 0.08683 | 2.864 | 8.54e-34 | 24311 | 615 | 23696 | 1.092 | 0.08402 | 2.98 | 1.29e-38 | 34349 | 650 | 33699 | 0.2663 | 0.1294 | Pericarditis | circulatory system | UricA | |
| 420.21 | | | | | 23792 | 96 | 23696 | | | | | 33781 | 82 | 33699 | | | Acute pericarditis | circulatory system | UricA | |
| 420.22 | | | | | 23800 | 104 | 23696 | | | | | 33807 | 108 | 33699 | | | Chronic pericarditis | circulatory system | UricA | |
| 420.3 | | | | | 23990 | 294 | 23696 | | | | | 33932 | 233 | 33699 | 0.4439 | 0.1406 | Endocarditis | circulatory system | UricA | |
| 425 | 0.9488 | 0.0495 | 2.583 | 6.89e-82 | 25754 | 2058 | 23696 | 0.9277 | 0.06236 | 2.529 | 4.71e-50 | 34958 | 1259 | 33699 | 0.9486 | 0.004977 | Cardiomyopathy | circulatory system | UricA | |
| 425.1 | 0.9861 | 0.05186 | 2.681 | 1.24e-80 | 25538 | 1842 | 23696 | 0.9942 | 0.06716 | 2.702 | 1.40e-49 | 34754 | 1055 | 33699 | 0.788 | 0.02214 | Primary/intrinsic cardiomyopathies | circulatory system | UricA | |
| 425.11 | | | | | 23809 | 113 | 23696 | | | | | 33810 | 111 | 33699 | | | Hypertrophic obstructive cardiomyopathy | circulatory system | UricA | |
| 425.12 | | | | | 23788 | 92 | 23696 | | | | | 33777 | 78 | 33699 | | | Other hypertrophic cardiomyopathy | circulatory system | UricA | |
| 425.2 | 1.099 | 0.08347 | 3 | 1.47e-39 | 24351 | 655 | 23696 | | | | | 34066 | 367 | 33699 | 0.2803 | -0.1464 | Secondary/extrinsic cardiomyopathies | circulatory system | UricA | |
| 425.8 | | | | | 23738 | 42 | 23696 | | | | | 33802 | 103 | 33699 | | | Other cardiomyopathy | circulatory system | UricA | |
| 426 | 0.8385 | 0.04398 | 2.313 | 4.73e-81 | 18055 | 3856 | 14199 | 0.9184 | 0.04886 | 2.505 | 8.26e-79 | 24927 | 2812 | 22115 | 0.09798 | 0.1069 | Cardiac conduction disorders | circulatory system | UricA | |
| 426.2 | 0.7604 | 0.07808 | 2.139 | 2.04e-22 | 15129 | 930 | 14199 | 0.758 | 0.1013 | 2.134 | 7.08e-14 | 22650 | 535 | 22115 | 0.4894 | -0.08614 | Atrioventricular [AV] block | circulatory system | UricA | |
| 426.21 | | | | | 14391 | 192 | 14199 | | | | | 22185 | 70 | 22115 | | | First degree AV block | circulatory system | UricA | |
| 426.22 | | | | | 14228 | 29 | 14199 | | | | | 22135 | 20 | 22115 | | | Mobitz II AV block | circulatory system | UricA | |
| 426.23 | | | | | 14388 | 189 | 14199 | | | | | 22218 | 103 | 22115 | | | Second degree AV block | circulatory system | UricA | |
| 426.24 | 0.6968 | 0.09438 | 2.007 | 1.55e-13 | 14798 | 599 | 14199 | | | | | 22478 | 363 | 22115 | 0.6917 | -0.05945 | Atrioventricular block, complete | circulatory system | UricA | |
| 426.25 | | | | | 14243 | 44 | 14199 | | | | | 22136 | 21 | 22115 | | | Other heart block | circulatory system | UricA | |
| 426.3 | 1.004 | 0.07763 | 2.729 | 2.96e-38 | 15070 | 871 | 14199 | 1.048 | 0.09421 | 2.851 | 9.94e-29 | 22678 | 563 | 22115 | 0.557 | 0.06987 | Bundle branch block | circulatory system | UricA | |
| 426.31 | | | | | 14547 | 348 | 14199 | | | | | 22262 | 147 | 22115 | | | Right bundle branch block | circulatory system | UricA | |
| 426.32 | | | | | 14638 | 439 | 14199 | | | | | 22500 | 385 | 22115 | 0.6416 | 0.07005 | Left bundle branch block | circulatory system | UricA | |
| 426.4 | | | | | 14250 | 51 | 14199 | | | | | 22148 | 33 | 22115 | | | Anomalous atrioventricular excitation | circulatory system | UricA | |
| 426.7 | 0.553 | 0.08841 | 1.738 | 3.97e-10 | 14888 | 689 | 14199 | 0.7196 | 0.08552 | 2.054 | 3.94e-17 | 22857 | 742 | 22115 | 0.01422 | 0.294 | Abnormal electrocardiogram [ECG] [EKG] | circulatory system | UricA | |
| 426.8 | | | | | 14301 | 102 | 14199 | | | | | 22230 | 115 | 22115 | | | Other cardiac conduction disorders | circulatory system | UricA | |
| 426.9 | 1.01 | 0.05497 | 2.747 | 1.87e-75 | 16283 | 2084 | 14199 | 0.9815 | 0.06874 | 2.668 | 3.02e-46 | 23352 | 1237 | 22115 | 0.8565 | -0.01555 | Cardiac pacemaker/device in situ | circulatory system | UricA | |
| 426.91 | 0.7974 | 0.06781 | 2.22 | 6.42e-32 | 15508 | 1309 | 14199 | 0.8267 | 0.07989 | 2.286 | 4.29e-25 | 23040 | 925 | 22115 | 0.9488 | 0.006568 | Cardiac pacemaker in situ | circulatory system | UricA | |

| phenotype | male_beta | male_SE | male_OR | male_p | male_n_total | male_n_cases | male_n_controls | female_beta | female_SE | female_OR | female_p | female_n_total | female_n_cases | female_n_controls | interaction_p | interaction_beta | description | group | lab | label |
|---|---|---|---|---|---|---|---|---|---|---|---|---|---|---|---|---|---|---|---|---|
| 426.92 | 1.347 | 0.06875 | 3.846 | 1.73e-85 | 15274 | 1075 | 14199 | | | | | 22521 | 406 | 22115 | 0.2535 | 0.1406 | Cardiac defibrillator in situ | circulatory system | UricA | |
| 427 | 0.6602 | 0.03372 | 1.935 | 2.40e-85 | 23930 | 9731 | 14199 | 0.6966 | 0.03191 | 2.007 | 1.15e-105 | 32189 | 10074 | 22115 | 0.2357 | 0.05424 | Cardiac dysrhythmias | circulatory system | UricA | |
| 427.1 | 0.9971 | 0.05579 | 2.71 | 1.99e-71 | 16046 | 1847 | 14199 | 0.9502 | 0.0623 | 2.586 | 1.64e-52 | 23534 | 1419 | 22115 | 0.9833 | 0.001718 | Paroxysmal tachycardia, unspecified | circulatory system | UricA | |
| 427.11 | 0.7487 | 0.08786 | 2.114 | 1.59e-17 | 14852 | 653 | 14199 | 0.8412 | 0.07834 | 2.319 | 6.75e-27 | 22978 | 863 | 22115 | 0.4565 | 0.08552 | Paroxysmal supraventricular tachycardia | circulatory system | UricA | |
| 427.12 | 1.136 | 0.06409 | 3.114 | 2.75e-70 | 15495 | 1296 | 14199 | 1.182 | 0.09127 | 3.26 | 2.40e-38 | 22690 | 575 | 22115 | 0.4625 | 0.08017 | Paroxysmal ventricular tachycardia | circulatory system | UricA | |
| 427.2 | 0.7077 | 0.04363 | 2.029 | 3.51e-59 | 18883 | 4684 | 14199 | 0.8325 | 0.05161 | 2.299 | 1.54e-58 | 25226 | 3111 | 22115 | 0.0417 | 0.1339 | Atrial fibrillation and flutter | circulatory system | UricA | |
| 427.21 | 0.7199 | 0.04404 | 2.054 | 4.61e-60 | 18758 | 4559 | 14199 | 0.8252 | 0.05248 | 2.282 | 1.06e-55 | 25130 | 3015 | 22115 | 0.09197 | 0.1123 | Atrial fibrillation | circulatory system | UricA | |
| 427.22 | 0.7467 | 0.06435 | 2.11 | 3.96e-31 | 15660 | 1461 | 14199 | 0.9161 | 0.08248 | 2.499 | 1.17e-28 | 22935 | 820 | 22115 | 0.0383 | 0.211 | Atrial flutter | circulatory system | UricA | |
| 427.3 | 0.7287 | 0.05648 | 2.072 | 4.39e-38 | 16128 | 1929 | 14199 | 0.8871 | 0.05849 | 2.428 | 5.79e-52 | 23822 | 1707 | 22115 | 0.09781 | 0.1318 | Other specified cardiac dysrhythmias | circulatory system | UricA | |
| 427.4 | | | | | 14682 | 483 | 14199 | | | | | 22371 | 256 | 22115 | 0.09454 | -0.2768 | Cardiac arrest and ventricular fibrillation | circulatory system | UricA | |
| 427.41 | | | | | 14406 | 207 | 14199 | | | | | 22188 | 73 | 22115 | | | Ventricular fibrillation and flutter | circulatory system | UricA | |
| 427.42 | | | | | 14543 | 344 | 14199 | | | | | 22324 | 209 | 22115 | 0.4774 | -0.1339 | Cardiac arrest | circulatory system | UricA | |
| 427.5 | 0.8792 | 0.04766 | 2.409 | 5.57e-76 | 17123 | 2924 | 14199 | 1.083 | 0.05116 | 2.953 | 1.97e-99 | 24375 | 2260 | 22115 | 0.001411 | 0.2195 | Arrhythmia (cardiac) NOS | circulatory system | UricA | |
| 427.6 | 0.6923 | 0.08083 | 1.998 | 1.08e-17 | 15021 | 822 | 14199 | 0.6718 | 0.09364 | 1.958 | 7.26e-13 | 22723 | 608 | 22115 | 0.7997 | -0.03066 | Premature beats | circulatory system | UricA | |
| 427.61 | | | | | 14305 | 106 | 14199 | | | | | 22243 | 128 | 22115 | | | Supraventricular premature beats | circulatory system | UricA | |

| phenotype | male_beta | male_SE | male_OR | male_p | male_n_total | male_n_cases | male_n_controls | female_beta | female_SE | female_OR | female_p | female_n_total | female_n_cases | female_n_controls | interaction_p | interaction_beta | description | group | lab | label |
|---|---|---|---|---|---|---|---|---|---|---|---|---|---|---|---|---|---|---|---|---|
| 427.7 | 0.9183 | 0.04793 | 2.505 | 7.70e-82 | 16928 | 2729 | 14199 | 0.9741 | 0.04427 | 2.649 | 2.57e-107 | 25298 | 3183 | 22115 | 0.1368 | 0.09485 | Tachycardia NOS | circulatory system | UricA | |
| 427.8 | 0.6939 | 0.07489 | 2.001 | 1.95e-20 | 15296 | 1097 | 14199 | 0.6811 | 0.08741 | 1.976 | 6.57e-15 | 22910 | 795 | 22115 | 0.6948 | -0.04405 | Sinoatrial node dysfunction (Bradycardia) | circulatory system | UricA | |
| 427.9 | 0.6018 | 0.05724 | 1.825 | 7.55e-26 | 16102 | 1903 | 14199 | 0.5954 | 0.04425 | 1.814 | 2.89e-41 | 25899 | 3784 | 22115 | 0.8032 | -0.01766 | Palpitations | circulatory system | UricA | |
| 428 | 0.943 | 0.03852 | 2.568 | 2.28e-132 | 24029 | 4495 | 19534 | 1.07 | 0.04174 | 2.915 | 7.40e-145 | 33140 | 3590 | 29550 | 0.03034 | 0.1198 | Congestive heart failure; nonhypertensive | circulatory system | UricA | |
| 428.1 | 1.102 | 0.0423 | 3.009 | 1.70e-149 | 22921 | 3387 | 19534 | 1.188 | 0.04707 | 3.28 | 1.71e-140 | 32151 | 2601 | 29550 | 0.2358 | 0.07282 | Congestive heart failure (CHF) NOS | circulatory system | UricA | |
| 428.2 | 1.558 | 0.06366 | 4.748 | 3.20e-132 | 20752 | 1218 | 19534 | 1.682 | 0.07882 | 5.377 | 4.67e-101 | 30286 | 736 | 29550 | 0.2322 | 0.1177 | Heart failure NOS | circulatory system | UricA | |
| 428.3 | 1.155 | 0.04903 | 3.175 | 9.18e-123 | 21811 | 2277 | 19534 | 1.268 | 0.0606 | 3.554 | 3.37e-97 | 30905 | 1355 | 29550 | 0.2523 | 0.08686 | Heart failure with reduced EF [Systolic or combined heart failure] | circulatory system | UricA | |
| 428.4 | 1.155 | 0.06276 | 3.175 | 1.11e-75 | 20846 | 1312 | 19534 | 1.15 | 0.05925 | 3.158 | 6.77e-84 | 31066 | 1516 | 29550 | 0.913 | -0.009171 | Heart failure with preserved EF [Diastolic heart failure] | circulatory system | UricA | |
| 429 | 0.8245 | 0.05812 | 2.281 | 1.11e-45 | 21072 | 1538 | 19534 | 0.8074 | 0.06168 | 2.242 | 3.76e-39 | 30921 | 1371 | 29550 | 0.9241 | 0.007868 | Ill-defined descriptions and complications of heart disease | circulatory system | UricA | |
| 429.1 | | | | | 19984 | 450 | 19534 | | | | | 29770 | 220 | 29550 | 0.802 | 0.04581 | Heart transplant/surgery | circulatory system | UricA | |
| 429.2 | 0.637 | 0.09719 | 1.891 | 5.61e-11 | 20041 | 507 | 19534 | | | | | 29943 | 393 | 29550 | 0.5723 | -0.08209 | Abnormal function study of cardiovascular system | circulatory system | UricA | |
| 429.3 | 0.9158 | 0.08926 | 2.499 | 1.06e-24 | 20117 | 583 | 19534 | 0.8509 | 0.08399 | 2.342 | 4.01e-24 | 30233 | 683 | 29550 | 0.739 | -0.03967 | Symptoms involving cardiovascular system | circulatory system | UricA | |
| 429.9 | | | | | 19772 | 238 | 19534 | | | | | 29698 | 148 | 29550 | | | Cardiac complications, not elsewhere classified | circulatory system | UricA | |

| phenotype | male_beta | male_SE | male_OR | male_p | male_n_total | male_n_cases | male_n_controls | female_beta | female_SE | female_OR | female_p | female_n_total | female_n_cases | female_n_controls | interaction_p | interaction_beta | description | group | lab | label |
|---|---|---|---|---|---|---|---|---|---|---|---|---|---|---|---|---|---|---|---|---|
| 430 | 0.5554 | 0.08781 | 1.743 | 2.53e-10 | 22249 | 653 | 21596 | 0.7228 | 0.09902 | 2.06 | 2.88e-13 | 31365 | 529 | 30836 | 0.1809 | 0.172 | Intracranial hemorrhage | circulatory system | UricA | |
| 430.1 | | | | | 21793 | 197 | 21596 | | | | | 31023 | 187 | 30836 | | | Subarachnoid hemorrhage | circulatory system | UricA | |
| 430.2 | | | | | 21911 | 315 | 21596 | | | | | 31093 | 257 | 30836 | 0.2952 | 0.1997 | Intracerebral hemorrhage | circulatory system | UricA | |
| 430.3 | | | | | 21798 | 202 | 21596 | | | | | 30969 | 133 | 30836 | | | Subdural hemorrhage | circulatory system | UricA | |
| 433 | 0.5147 | 0.04175 | 1.673 | 6.36e-35 | 25148 | 3552 | 21596 | 0.5957 | 0.04289 | 1.814 | 7.31e-44 | 34408 | 3572 | 30836 | 0.2066 | 0.07359 | Cerebrovascular disease | circulatory system | UricA | |
| 433.1 | 0.6603 | 0.06041 | 1.935 | 8.25e-28 | 23051 | 1455 | 21596 | 0.7459 | 0.06771 | 2.108 | 3.20e-28 | 32011 | 1175 | 30836 | 0.5126 | 0.05763 | Occlusion and stenosis of precerebral arteries | circulatory system | UricA | |
| 433.11 | | | | | 21806 | 210 | 21596 | | | | | 31009 | 173 | 30836 | | | Occlusion of cerebral arteries, with cerebral infarction | circulatory system | UricA | |
| 433.12 | | | | | 21766 | 170 | 21596 | | | | | 30990 | 154 | 30836 | | | Cerebral atherosclerosis | circulatory system | UricA | |
| 433.2 | 0.5242 | 0.06491 | 1.689 | 6.67e-16 | 22816 | 1220 | 21596 | 0.6943 | 0.0675 | 2.002 | 8.24e-25 | 32003 | 1167 | 30836 | 0.04776 | 0.1799 | Occlusion of cerebral arteries | circulatory system | UricA | |
| 433.21 | 0.5195 | 0.06626 | 1.681 | 4.46e-15 | 22764 | 1168 | 21596 | 0.6987 | 0.06904 | 2.011 | 4.53e-24 | 31945 | 1109 | 30836 | 0.03648 | 0.1942 | Cerebral artery occlusion, with cerebral infarction | circulatory system | UricA | |
| 433.3 | 0.6288 | 0.05533 | 1.875 | 6.24e-30 | 23331 | 1735 | 21596 | 0.6924 | 0.05631 | 1.999 | 9.49e-35 | 32595 | 1759 | 30836 | 0.4443 | 0.05871 | Cerebral ischemia | circulatory system | UricA | |
| 433.31 | 0.6016 | 0.05741 | 1.825 | 1.08e-25 | 23194 | 1598 | 21596 | 0.6965 | 0.05831 | 2.007 | 6.87e-33 | 32452 | 1616 | 30836 | 0.2524 | 0.091 | Transient cerebral ischemia | circulatory system | UricA | |
| 433.32 | | | | | 21602 | 6 | 21596 | | | | | 30850 | 14 | 30836 | | | Moyamoya disease | circulatory system | UricA | |
| 433.5 | | | | | 21737 | 141 | 21596 | | | | | 31155 | 319 | 30836 | | | Cerebral aneurysm | circulatory system | UricA | |
| 433.6 | | | | | 21926 | 330 | 21596 | | | | | 31171 | 335 | 30836 | 0.0364 | 0.3405 | Acute, but ill-defined cerebrovascular disease | circulatory system | UricA | |
| 433.8 | 0.5021 | 0.087 | 1.652 | 7.85e-09 | 22246 | 650 | 21596 | 0.5508 | 0.09142 | 1.735 | 1.69e-09 | 31470 | 634 | 30836 | 0.453 | 0.09189 | Late effects of cerebrovascular disease | circulatory system | UricA | |
| 440 | 0.8746 | 0.05881 | 2.398 | 5.15e-50 | 21894 | 1543 | 20351 | 0.9491 | 0.06455 | 2.583 | 6.16e-49 | 31649 | 1262 | 30387 | 0.5874 | 0.04603 | Atherosclerosis | circulatory system | UricA | |
| 440.1 | | | | | 20496 | 145 | 20351 | | | | | 30601 | 214 | 30387 | | | Atherosclerosis of renal artery | circulatory system | UricA | |
| 440.2 | 0.7706 | 0.0824 | 2.161 | 8.64e-21 | 21065 | 714 | 20351 | | | | | 30809 | 422 | 30387 | 0.04299 | 0.2623 | Atherosclerosis of the extremities | circulatory system | UricA | |
| 440.21 | | | | | 20544 | 193 | 20351 | | | | | 30472 | 85 | 30387 | | | Atherosclerosis of native arteries of the extremities with ulceration or gangrene | circulatory system | UricA | |
| 440.22 | | | | | 20765 | 414 | 20351 | | | | | 30616 | 229 | 30387 | 0.1076 | 0.2784 | Atherosclerosis of native arteries of the extremities with intermittent claudication | circulatory system | UricA | |
| 440.9 | | | | | 20820 | 469 | 20351 | | | | | 30841 | 454 | 30387 | 0.4631 | -0.1023 | Atherosclerosis of aorta | circulatory system | UricA | |
| 441 | | | | | 20539 | 188 | 20351 | | | | | 30626 | 239 | 30387 | | | Vascular insufficiency of intestine | circulatory system | UricA | |
| 441.1 | | | | | 20421 | 70 | 20351 | | | | | 30462 | 75 | 30387 | | | Acute vascular insufficiency of intestine | circulatory system | UricA | |
| 441.2 | | | | | 20385 | 34 | 20351 | | | | | 30454 | 67 | 30387 | | | Chronic vascular insufficiency of intestine | circulatory system | UricA | |
| 442 | 0.4271 | 0.06603 | 1.533 | 9.88e-11 | 21609 | 1258 | 20351 | 0.4757 | 0.09054 | 1.609 | 1.49e-07 | 31061 | 674 | 30387 | 0.9367 | -0.008672 | Other aneurysm | circulatory system | UricA | |
| 442.1 | 0.3733 | 0.07341 | 1.453 | 3.67e-07 | 21366 | 1015 | 20351 | | | | | 30810 | 423 | 30387 | 0.9823 | 0.002936 | Aortic aneurysm | circulatory system | UricA | |
| 442.11 | 0.4515 | 0.09323 | 1.571 | 1.28e-06 | 20965 | 614 | 20351 | | | | | 30570 | 183 | 30387 | 0.5577 | -0.1126 | Abdominal aortic aneurysm | circulatory system | UricA | |

| phenotype | male_beta | male_SE | male_OR | male_p | male_n_total | male_n_cases | male_n_controls | female_beta | female_SE | female_OR | female_p | female_n_total | female_n_cases | female_n_controls | interaction_p | interaction_beta | description | group | lab | label |
|---|---|---|---|---|---|---|---|---|---|---|---|---|---|---|---|---|---|---|---|---|
| 442.2 | | | | | 20421 | 70 | 20351 | | | | | 30397 | 10 | 30387 | | | Aneurysm of iliac artery | circulatory system | UricA | |
| 442.3 | | | | | 20442 | 91 | 20351 | | | | | 30441 | 54 | 30387 | | | Aneurysm of artery of lower extremity | circulatory system | UricA | |
| 442.4 | | | | | 20394 | 43 | 20351 | | | | | 30441 | 54 | 30387 | | | Arterial dissection | circulatory system | UricA | |
| 442.8 | | | | | 20454 | 103 | 20351 | | | | | 30512 | 125 | 30387 | | | Aneurysm of other specified artery | circulatory system | UricA | |
| 443 | 0.7798 | 0.05622 | 2.181 | 9.61e-44 | 22049 | 1698 | 20351 | 0.7129 | 0.05962 | 2.04 | 5.99e-33 | 31903 | 1516 | 30387 | 0.2589 | -0.09026 | Peripheral vascular disease | circulatory system | UricA | |
| 443.1 | | | | | 20444 | 93 | 20351 | | | | | 30852 | 465 | 30387 | 0.6811 | -0.1029 | Raynaud's syndrome | circulatory system | UricA | |
| 443.7 | | | | | 20611 | 260 | 20351 | | | | | 30517 | 130 | 30387 | | | Peripheral angiopathy in diseases classified elsewhere | circulatory system | UricA | |
| 443.8 | | | | | 20448 | 97 | 20351 | | | | | 30453 | 66 | 30387 | | | Other specified peripheral vascular diseases | circulatory system | UricA | |
| 443.9 | 0.8058 | 0.06049 | 2.238 | 1.73e-40 | 21791 | 1440 | 20351 | 0.8979 | 0.07265 | 2.454 | 4.33e-35 | 31349 | 962 | 30387 | 0.4896 | 0.06363 | Peripheral vascular disease, unspecified | circulatory system | UricA | |
| 444 | | | | | 20661 | 310 | 20351 | | | | | 30633 | 246 | 30387 | 0.9253 | -0.01693 | Arterial embolism and thrombosis | circulatory system | UricA | |
| 444.1 | | | | | 20465 | 114 | 20351 | | | | | 30462 | 75 | 30387 | | | Arterial embolism and thrombosis of lower extremity artery | circulatory system | UricA | |
| 444.2 | | | | | 20391 | 40 | 20351 | | | | | 30424 | 37 | 30387 | | | Embolism and thrombosis of abdominal aorta | circulatory system | UricA | |
| 444.5 | | | | | 20357 | 6 | 20351 | | | | | 30392 | 5 | 30387 | | | Atheroembolism | circulatory system | UricA | |
| 446 | | | | | 20562 | 211 | 20351 | | | | | 30776 | 389 | 30387 | 0.2272 | -0.2153 | Polyarteritis nodosa and allied conditions | circulatory system | UricA | |
| 446.1 | | | | | 20353 | 2 | 20351 | | | | | 30391 | 4 | 30387 | | | Thromboangiitis obliterans | circulatory system | UricA | |
| 446.2 | | | | | 20351 | | | | | | | 30393 | 6 | 30387 | | | Acute febrile mucocutaneous lymph node syndrome (Kawasaki disease) | circulatory system | UricA | |
| 446.3 | | | | | 20377 | 26 | 20351 | | | | | 30419 | 32 | 30387 | | | Hypersensitivity angiitis | circulatory system | UricA | |
| 446.4 | | | | | 20429 | 78 | 20351 | | | | | 30467 | 80 | 30387 | | | Wegener's granulomatosis | circulatory system | UricA | |
| 446.5 | | | | | 20386 | 35 | 20351 | | | | | 30500 | 113 | 30387 | | | Giant cell arteritis | circulatory system | UricA | |
| 446.6 | | | | | 20380 | 29 | 20351 | | | | | 30423 | 36 | 30387 | | | Polyarteritis nodosa | circulatory system | UricA | |
| 446.7 | | | | | 20353 | 2 | 20351 | | | | | 30403 | 16 | 30387 | | | Takayasu's disease | circulatory system | UricA | |
| 446.8 | | | | | 20362 | 11 | 20351 | | | | | 30409 | 22 | 30387 | | | Thrombotic microangiopathy | circulatory system | UricA | |
| 446.9 | | | | | 20443 | 92 | 20351 | | | | | 30554 | 167 | 30387 | | | Arteritis NOS | circulatory system | UricA | |
| 447 | 0.7695 | 0.06844 | 2.159 | 2.52e-29 | 21399 | 1048 | 20351 | 0.8547 | 0.09215 | 2.351 | 1.76e-20 | 30938 | 551 | 30387 | 0.5059 | 0.07449 | Other disorders of arteries and arterioles | circulatory system | UricA | |
| 447.1 | | | | | 20462 | 111 | 20351 | | | | | 30487 | 100 | 30387 | | | Stricture of artery | circulatory system | UricA | |
| 447.7 | 0.6207 | 0.08266 | 1.86 | 5.95e-14 | 21056 | 705 | 20351 | | | | | 30596 | 209 | 30387 | 0.08702 | -0.2965 | Aortic ectasia | circulatory system | UricA | |
| 448 | | | | | 20358 | 7 | 20351 | | | | | 30410 | 23 | 30387 | | | Disease of capillaries | circulatory system | UricA | |
| 450 | | | | | 27464 | 390 | 27074 | 0.1431 | 0.08883 | 1.154 | 0.1073 | 36525 | 739 | 35786 | 0.2771 | -0.1498 | Noninfectious disorders of lymphatic channels | circulatory system | UricA | |

| phenotype | male_beta | male_SE | male_OR | male_p | male_n_total | male_n_cases | male_n_controls | female_beta | female_SE | female_OR | female_p | female_n_total | female_n_cases | female_n_controls | interaction_p | interaction_beta | description | group | lab | label |
|---|---|---|---|---|---|---|---|---|---|---|---|---|---|---|---|---|---|---|---|---|
| 451 | | | | | 17675 | 229 | 17446 | | | | | 26197 | 265 | 25932 | | | Phlebitis and thrombophlebitis | circulatory system | UricA | |
| 451.2 | | | | | 17551 | 105 | 17446 | | | | | 26069 | 137 | 25932 | | | Phlebitis and thrombophlebitis of lower extremities | circulatory system | UricA | |
| 452 | 0.8605 | 0.05209 | 2.364 | 2.61e-61 | 19497 | 2051 | 17446 | 0.9296 | 0.05176 | 2.533 | 3.99e-72 | 28016 | 2084 | 25932 | 0.1262 | 0.1098 | Other venous embolism and thrombosis | circulatory system | UricA | |
| 452.1 | | | | | 17477 | 31 | 17446 | | | | | 25960 | 28 | 25932 | | | Iatrogenic pulmonary embolism and infarction | circulatory system | UricA | |
| 452.2 | 0.9187 | 0.06863 | 2.506 | 7.33e-41 | 18523 | 1077 | 17446 | 0.9252 | 0.06987 | 2.522 | 5.02e-40 | 26996 | 1064 | 25932 | 0.5238 | 0.06091 | Deep vein thrombosis [DVT] | circulatory system | UricA | |
| 452.8 | | | | | 17477 | 31 | 17446 | | | | | 25956 | 24 | 25932 | | | Postphlebitic syndrome | circulatory system | UricA | |
| 453 | | | | | 17473 | 27 | 17446 | | | | | 25945 | 13 | 25932 | | | Chronic venous hypertension | circulatory system | UricA | |
| 454 | | | | | 17905 | 459 | 17446 | 0.6516 | 0.09206 | 1.919 | 1.46e-12 | 26542 | 610 | 25932 | 0.7755 | 0.03811 | Varicose veins | circulatory system | UricA | |
| 454.1 | | | | | 17712 | 266 | 17446 | 0.5416 | 0.1018 | 1.719 | 1.03e-07 | 26438 | 506 | 25932 | 0.04009 | -0.3301 | Varicose veins of lower extremity | circulatory system | UricA | |
| 454.11 | | | | | 17657 | 211 | 17446 | | | | | 26297 | 365 | 25932 | 0.03195 | -0.3918 | Varicose veins of lower extremity, symptomtic | circulatory system | UricA | |
| 455 | 0.8001 | 0.0661 | 2.226 | 9.86e-34 | 18658 | 1212 | 17446 | 0.5422 | 0.06595 | 1.72 | 2.00e-16 | 27261 | 1329 | 25932 | 0.001796 | -0.2845 | Hemorrhoids | circulatory system | UricA | |
| 456 | | | | | 17849 | 403 | 17446 | | | | | 26402 | 470 | 25932 | 0.06553 | -0.264 | Chronic venous insufficiency [CVI] | circulatory system | UricA | |
| 457 | 0.9993 | 0.04912 | 2.716 | 5.21e-92 | 19943 | 2497 | 17446 | 1.152 | 0.05468 | 3.165 | 1.63e-98 | 27763 | 1831 | 25932 | 0.04602 | 0.1443 | Encounter for long-term (current) use of anticoagulants, antithrombotics, aspirin | circulatory system | UricA | |
| 457.2 | 0.9385 | 0.09415 | 2.556 | 2.09e-23 | 17977 | 531 | 17446 | | | | | 26262 | 330 | 25932 | 0.2086 | 0.1844 | Encounter for long-term (current) use of antiplatelets/antithrombotics | circulatory system | UricA | |
| 457.3 | 1.031 | 0.05063 | 2.805 | 3.01e-92 | 19733 | 2287 | 17446 | 1.178 | 0.05675 | 3.248 | 1.00e-95 | 27596 | 1664 | 25932 | 0.06606 | 0.1375 | Encounter for long-term (current) use of aspirin | circulatory system | UricA | |
| 458 | 0.9024 | 0.0404 | 2.465 | 1.65e-110 | 23987 | 3579 | 20408 | 0.9714 | 0.04384 | 2.642 | 8.61e-109 | 32936 | 2891 | 30045 | 0.1216 | 0.0898 | Hypotension | circulatory system | UricA | |
| 458.1 | 0.8841 | 0.08121 | 2.421 | 1.35e-27 | 21124 | 716 | 20408 | 0.8455 | 0.09321 | 2.329 | 1.18e-19 | 30599 | 554 | 30045 | 0.9136 | -0.01301 | Orthostatic hypotension | circulatory system | UricA | |
| 458.2 | | | | | 20542 | 134 | 20408 | | | | | 30168 | 123 | 30045 | | | Iatrogenic hypotension | circulatory system | UricA | |
| 458.9 | 0.9686 | 0.04437 | 2.634 | 1.22e-105 | 23196 | 2788 | 20408 | 1.061 | 0.04865 | 2.888 | 2.31e-105 | 32272 | 2227 | 30045 | 0.09391 | 0.1072 | Hypotension NOS | circulatory system | UricA | |
| 459 | 0.9284 | 0.07025 | 2.53 | 7.05e-40 | 21377 | 969 | 20408 | 1.008 | 0.07316 | 2.74 | 3.58e-43 | 30928 | 883 | 30045 | 0.2923 | 0.1035 | Other disorders of circulatory system | circulatory system | UricA | |
| 459.1 | | | | | 20541 | 133 | 20408 | | | | | 30175 | 130 | 30045 | | | Hemorrhage NOS | circulatory system | UricA | |
| 459.7 | | | | | 20566 | 158 | 20408 | | | | | 30162 | 117 | 30045 | | | Blood vessel replaced | circulatory system | UricA | |
| 459.9 | 0.9 | 0.08833 | 2.46 | 2.21e-24 | 21001 | 593 | 20408 | 0.9747 | 0.09439 | 2.65 | 5.33e-25 | 30563 | 518 | 30045 | 0.3801 | 0.1098 | Circulatory disease NEC | circulatory system | UricA | |
| 464 | 0.2574 | 0.06323 | 1.294 | 4.67e-05 | 22311 | 1470 | 20841 | 0.2361 | 0.04636 | 1.266 | 3.54e-07 | 28085 | 3592 | 24493 | 0.8977 | 0.009777 | Acute sinusitis | respiratory | UricA | |
| 465 | 0.3213 | 0.04866 | 1.379 | 4.05e-11 | 23812 | 2971 | 20841 | 0.2577 | 0.0384 | 1.294 | 1.93e-11 | 30856 | 6363 | 24493 | 0.697 | -0.02339 | Acute upper respiratory infections of multiple or unspecified sites | respiratory | UricA | |
| 465.2 | 0.2739 | 0.08129 | 1.315 | 7.52e-04 | 21831 | 990 | 20841 | 0.2212 | 0.054 | 1.248 | 4.20e-05 | 27189 | 2696 | 24493 | 0.6753 | 0.0392 | Acute pharyngitis | respiratory | UricA | |
| 465.4 | | | | | 20905 | 64 | 20841 | | | | | 24590 | 97 | 24493 | | | Acute laryngitis and tracheitis | respiratory | UricA | |
| 470 | 0.4368 | 0.07615 | 1.548 | 9.69e-09 | 19858 | 932 | 18926 | 0.2126 | 0.07342 | 1.237 | 0.003789 | 25321 | 1191 | 24130 | 0.03776 | -0.2138 | Septal Deviations/Turbinate Hypertrophy | respiratory | UricA | |
| 471 | | | | | 19079 | 153 | 18926 | | | | | 24245 | 115 | 24130 | | | Nasal polyps | respiratory | UricA | |

| phenotype | male_beta | male_SE | male_OR | male_p | male_n_total | male_n_cases | male_n_controls | female_beta | female_SE | female_OR | female_p | female_n_total | female_n_cases | female_n_controls | interaction_p | interaction_beta | description | group | lab | label |
|---|---|---|---|---|---|---|---|---|---|---|---|---|---|---|---|---|---|---|---|---|
| 472 | 0.3676 | 0.08659 | 1.444 | 2.18e-05 | 19621 | 695 | 18926 | 0.2739 | 0.07154 | 1.315 | 1.29e-04 | 25328 | 1198 | 24130 | 0.4585 | -0.08085 | Chronic pharyngitis and nasopharyngitis | respiratory | UricA | |
| 473 | 0.3356 | 0.0735 | 1.399 | 4.97e-06 | 19940 | 1014 | 18926 | 0.3078 | 0.06965 | 1.36 | 9.94e-06 | 25397 | 1267 | 24130 | 0.4655 | 0.07184 | Diseases of the larynx and vocal cords | respiratory | UricA | |
| 473.1 | | | | | 19013 | 87 | 18926 | | | | | 24250 | 120 | 24130 | | | Chronic laryngitis | respiratory | UricA | |
| 473.3 | | | | | 19140 | 214 | 18926 | | | | | 24368 | 238 | 24130 | | | Paralysis/spasm of vocal cords or larynx | respiratory | UricA | |
| 473.4 | 0.4495 | 0.08222 | 1.567 | 4.58e-08 | 19690 | 764 | 18926 | 0.3096 | 0.07745 | 1.363 | 6.42e-05 | 25123 | 993 | 24130 | 0.9137 | -0.01189 | Voice disturbance | respiratory | UricA | |
| 474 | | | | | 19193 | 267 | 18926 | | | | | 24615 | 485 | 24130 | 0.9459 | -0.01414 | Acute and chronic tonsillitis | respiratory | UricA | |
| 474.1 | | | | | 18953 | 27 | 18926 | | | | | 24211 | 81 | 24130 | | | Acute tonsillitis | respiratory | UricA | |
| 474.2 | | | | | 19156 | 230 | 18926 | | | | | 24527 | 397 | 24130 | 0.838 | -0.04689 | Chronic tonsillitis and adenoiditis | respiratory | UricA | |
| 475 | 0.5885 | 0.06277 | 1.801 | 6.89e-21 | 20308 | 1382 | 18926 | 0.3902 | 0.05423 | 1.477 | 6.24e-13 | 26319 | 2189 | 24130 | 0.1218 | -0.1248 | Chronic sinusitis | respiratory | UricA | |
| 475.9 | | | | | 19118 | 192 | 18926 | | | | | 24485 | 355 | 24130 | 0.5337 | 0.1192 | Postnasal drip | respiratory | UricA | |
| 476 | 0.4257 | 0.04917 | 1.531 | 4.79e-18 | 21597 | 2671 | 18926 | 0.233 | 0.04121 | 1.262 | 1.56e-08 | 28998 | 4868 | 24130 | 0.03245 | -0.1337 | Allergic rhinitis | respiratory | UricA | |
| 477 | | | | | 19327 | 401 | 18926 | | | | | 24561 | 431 | 24130 | 0.8576 | -0.02589 | Epistaxis or throat hemorrhage | respiratory | UricA | |
| 478 | | | | | 19150 | 224 | 18926 | | | | | 24407 | 277 | 24130 | 0.042 | 0.4131 | Throat pain | respiratory | UricA | |
| 479 | 0.6131 | 0.06386 | 1.846 | 7.97e-22 | 20272 | 1346 | 18926 | 0.4312 | 0.05728 | 1.539 | 5.12e-14 | 26030 | 1900 | 24130 | 0.5947 | -0.04432 | Other upper respiratory disease | respiratory | UricA | |
| 480 | 0.8589 | 0.03982 | 2.361 | 3.46e-103 | 23852 | 3704 | 20148 | 0.9366 | 0.04072 | 2.551 | 4.49e-117 | 31060 | 3539 | 27521 | 0.003155 | 0.1637 | Pneumonia | respiratory | UricA | |
| 480.1 | 0.8097 | 0.07206 | 2.247 | 2.71e-29 | 21107 | 959 | 20148 | 0.9995 | 0.076 | 2.717 | 1.66e-39 | 28349 | 828 | 27521 | 9.67e-04 | 0.3345 | Bacterial pneumonia | respiratory | UricA | |
| 480.11 | | | | | 20392 | 244 | 20148 | | | | | 27735 | 214 | 27521 | | | Pneumococcal pneumonia | respiratory | UricA | |
| 480.12 | | | | | 20313 | 165 | 20148 | | | | | 27625 | 104 | 27521 | | | Pseudomonal pneumonia | respiratory | UricA | |
| 480.13 | | | | | 20243 | 95 | 20148 | | | | | 27596 | 75 | 27521 | | | MRSA pneumonia | respiratory | UricA | |
| 480.2 | | | | | 20252 | 104 | 20148 | | | | | 27629 | 108 | 27521 | | | Viral pneumonia | respiratory | UricA | |
| 480.3 | | | | | 20261 | 113 | 20148 | | | | | 27595 | 74 | 27521 | | | Pneumonia due to fungus (mycoses) | respiratory | UricA | |
| 480.5 | | | | | 20273 | 125 | 20148 | | | | | 27647 | 126 | 27521 | | | Bronchopneumonia and lung abscess | respiratory | UricA | |

| phenotype | male_beta | male_SE | male_OR | male_p | male_n_total | male_n_cases | male_n_controls | female_beta | female_SE | female_OR | female_p | female_n_total | female_n_cases | female_n_controls | interaction_p | interaction_beta | description | group | lab | label |
|---|---|---|---|---|---|---|---|---|---|---|---|---|---|---|---|---|---|---|---|---|
| 481 | | | | | 20437 | 289 | 20148 | | | | | 27934 | 413 | 27521 | 0.695 | -0.06341 | Influenza | respiratory | UricA | |
| 483 | 0.4042 | 0.07145 | 1.498 | 1.55e-08 | 21180 | 1032 | 20148 | 0.4856 | 0.05663 | 1.625 | 9.96e-18 | 29347 | 1826 | 27521 | 0.25 | 0.1018 | Acute bronchitis and bronchiolitis | respiratory | UricA | |
| 495 | 0.396 | 0.06092 | 1.486 | 8.03e-11 | 22371 | 1521 | 20850 | 0.3877 | 0.0447 | 1.474 | 4.22e-18 | 31168 | 3223 | 27945 | 0.4385 | 0.05638 | Asthma | respiratory | UricA | |
| 495.1 | | | | | 21010 | 160 | 20850 | | | | | 28173 | 228 | 27945 | | | Chronic obstructive asthma | respiratory | UricA | |
| 495.11 | | | | | 20869 | 19 | 20850 | | | | | 27991 | 46 | 27945 | | | Chronic obstructive asthma with exacerbation | respiratory | UricA | |
| 495.2 | | | | | 21125 | 275 | 20850 | 0.4907 | 0.08414 | 1.633 | 5.49e-09 | 28693 | 748 | 27945 | 0.1074 | 0.2488 | Asthma with exacerbation | respiratory | UricA | |
| 496 | 0.4691 | 0.04369 | 1.599 | 6.70e-27 | 23991 | 3141 | 20850 | 0.5808 | 0.04645 | 1.787 | 7.10e-36 | 30852 | 2907 | 27945 | 0.02582 | 0.1381 | Chronic airway obstruction | respiratory | UricA | |
| 496.1 | 0.4464 | 0.07307 | 1.563 | 1.00e-09 | 21816 | 966 | 20850 | 0.8322 | 0.08287 | 2.298 | 9.98e-24 | 28671 | 726 | 27945 | 3.87e-04 | 0.3796 | Emphysema | respiratory | UricA | |
| 496.2 | 0.6479 | 0.0753 | 1.912 | 7.67e-18 | 21707 | 857 | 20850 | 0.6402 | 0.07484 | 1.897 | 1.19e-17 | 28880 | 935 | 27945 | 0.729 | 0.03565 | Chronic bronchitis | respiratory | UricA | |
| 496.21 | 0.7152 | 0.08295 | 2.045 | 6.57e-18 | 21544 | 694 | 20850 | 0.6896 | 0.08316 | 1.993 | 1.11e-16 | 28686 | 741 | 27945 | 0.9805 | 0.002782 | Obstructive chronic bronchitis | respiratory | UricA | |
| 496.3 | | | | | 21106 | 256 | 20850 | | | | | 28305 | 360 | 27945 | 0.06057 | 0.3326 | Bronchiectasis | respiratory | UricA | |
| 497 | | | | | 21118 | 268 | 20850 | | | | | 28430 | 485 | 27945 | 0.9527 | 0.009439 | Bronchitis | respiratory | UricA | |
| 498 | | | | | 20873 | 23 | 20850 | | | | | 27982 | 37 | 27945 | | | Acute bronchospasm | respiratory | UricA | |

| phenotype | male_beta | male_SE | male_OR | male_p | male_n_total | male_n_cases | male_n_controls | female_beta | female_SE | female_OR | female_p | female_n_total | female_n_cases | female_n_controls | interaction_p | interaction_beta | description | group | lab | label |
|---|---|---|---|---|---|---|---|---|---|---|---|---|---|---|---|---|---|---|---|---|
| 499 | | | | | 27770 | 89 | 27681 | | | | | 36921 | 77 | 36844 | | | Cystic fibrosis | respiratory | UricA | |
| 500 | | | | | 16503 | 145 | 16358 | | | | | 26637 | 93 | 26544 | | | Lung disease due to external agents | respiratory | UricA | |
| 500.1 | | | | | 16369 | 11 | 16358 | | | | | 26562 | 18 | 26544 | | | Extrinsic allergic alveolitis | respiratory | UricA | |
| 500.2 | | | | | 16384 | 26 | 16358 | | | | | 26547 | 3 | 26544 | | | Pneumoconiosis | respiratory | UricA | |
| 501 | 0.6012 | 0.0971 | 1.824 | 5.97e-10 | 16908 | 550 | 16358 | | | | | 26905 | 361 | 26544 | 0.02252 | 0.3347 | Pneumonitis due to inhalation of food or vomitus | respiratory | UricA | |
| 502 | | | | | 16775 | 417 | 16358 | | | | | 26948 | 404 | 26544 | 0.411 | -0.1226 | Postinflammatory pulmonary fibrosis | respiratory | UricA | |
| 503 | 1.328 | 0.06811 | 3.775 | 9.85e-85 | 17413 | 1055 | 16358 | 1.498 | 0.0745 | 4.472 | 6.96e-90 | 27379 | 835 | 26544 | 0.03127 | 0.2104 | Pulmonary congestion and hypostasis | respiratory | UricA | |
| 504 | | | | | 16686 | 328 | 16358 | | | | | 26850 | 306 | 26544 | 0.1421 | -0.246 | Other alveolar and parietoalveolar pneumonopathy | respiratory | UricA | |
| 504.1 | | | | | 16528 | 170 | 16358 | | | | | 26674 | 130 | 26544 | | | Idiopathic fibrosing alveolitis | respiratory | UricA | |
| 505 | | | | | 16583 | 225 | 16358 | | | | | 26786 | 242 | 26544 | | | Other pulmonary inflamation or edema | respiratory | UricA | |
| 506 | 0.4997 | 0.0659 | 1.648 | 3.36e-14 | 17754 | 1396 | 16358 | 0.6725 | 0.0746 | 1.959 | 1.98e-19 | 27619 | 1075 | 26544 | 0.09885 | 0.1599 | Empyema and pneumothorax | respiratory | UricA | |
| 507 | 0.8134 | 0.04096 | 2.256 | 9.05e-88 | 20407 | 4049 | 16358 | 0.9778 | 0.04339 | 2.659 | 1.93e-112 | 29890 | 3346 | 26544 | 0.005415 | 0.1608 | Pleurisy; pleural effusion | respiratory | UricA | |
| 508 | 0.7091 | 0.03647 | 2.032 | 3.25e-84 | 22186 | 5828 | 16358 | 0.8776 | 0.03952 | 2.405 | 3.16e-109 | 30908 | 4364 | 26544 | 0.005012 | 0.1463 | Pulmonary collapse; interstitial and compensatory emphysema | respiratory | UricA | |
| 509 | 0.7317 | 0.04023 | 2.079 | 6.16e-74 | 20689 | 4331 | 16358 | 0.8945 | 0.04209 | 2.446 | 3.12e-100 | 30174 | 3630 | 26544 | 0.001326 | 0.1813 | Respiratory failure, insufficiency, arrest | respiratory | UricA | |
| 509.1 | 0.8893 | 0.04558 | 2.433 | 9.07e-85 | 19312 | 2954 | 16358 | 0.9936 | 0.04738 | 2.701 | 1.14e-97 | 29134 | 2590 | 26544 | 0.0145 | 0.1557 | Respiratory failure | respiratory | UricA | |
| 509.2 | 0.8854 | 0.07089 | 2.424 | 8.48e-36 | 17397 | 1039 | 16358 | 1.08 | 0.08008 | 2.946 | 1.73e-41 | 27339 | 795 | 26544 | 0.1032 | 0.1691 | Respiratory insufficiency | respiratory | UricA | |
| 509.3 | 0.4384 | 0.07925 | 1.55 | 3.17e-08 | 17365 | 1007 | 16358 | 0.7395 | 0.09278 | 2.095 | 1.58e-15 | 27216 | 672 | 26544 | 0.007242 | 0.3196 | Pulmonary insufficiency or respiratory failure following trauma and surgery | respiratory | UricA | |
| 509.5 | | | | | 16395 | 37 | 16358 | | | | | 26570 | 26 | 26544 | | | Respiratory arrest | respiratory | UricA | |
| 509.8 | 0.757 | 0.08145 | 2.132 | 1.49e-20 | 17139 | 781 | 16358 | 1.015 | 0.08283 | 2.76 | 1.52e-34 | 27262 | 718 | 26544 | 0.001516 | 0.3566 | Dependence on respirator [Ventilator] or supplemental oxygen | respiratory | UricA | |
| 510 | 0.816 | 0.04526 | 2.261 | 1.17e-72 | 24918 | 2670 | 22248 | 0.9339 | 0.04854 | 2.544 | 1.80e-82 | 34185 | 2275 | 31910 | 0.01162 | 0.162 | Other diseases of lung | respiratory | UricA | |
| 510.2 | | | | | 22355 | 107 | 22248 | | | | | 32017 | 107 | 31910 | | | Lung transplant | respiratory | UricA | |
| 512 | 0.8026 | 0.03457 | 2.231 | 3.02e-119 | 23558 | 11275 | 12283 | 0.7664 | 0.03286 | 2.152 | 2.80e-120 | 31474 | 13478 | 17996 | 0.7754 | 0.01351 | Other symptoms of respiratory system | respiratory | UricA | |
| 512.1 | | | | | 12736 | 453 | 12283 | 1.001 | 0.08583 | 2.722 | 1.90e-31 | 18716 | 720 | 17996 | 0.05241 | 0.2589 | Wheezing | respiratory | UricA | |
| 512.2 | | | | | 12736 | 453 | 12283 | 1.186 | 0.09148 | 3.273 | 2.05e-38 | 18591 | 595 | 17996 | 0.2506 | 0.1546 | Painful respiration | respiratory | UricA | |
| 512.3 | | | | | 12290 | 7 | 12283 | | | | | 18006 | 10 | 17996 | | | Abnormal chest sounds | respiratory | UricA | |
| 512.7 | 0.9962 | 0.03844 | 2.708 | 4.14e-148 | 19002 | 6719 | 12283 | 1.003 | 0.03767 | 2.726 | 3.55e-156 | 25374 | 7378 | 17996 | 0.3841 | 0.04641 | Shortness of breath | respiratory | UricA | |
| 512.8 | 0.9751 | 0.04465 | 2.651 | 1.06e-105 | 16541 | 4258 | 12283 | 0.7682 | 0.04039 | 2.156 | 1.15e-80 | 24362 | 6366 | 17996 | 0.009792 | -0.1544 | Cough | respiratory | UricA | |
| 512.9 | 0.9932 | 0.04106 | 2.7 | 2.80e-129 | 17400 | 5117 | 12283 | 1.039 | 0.04075 | 2.826 | 2.42e-143 | 23359 | 5363 | 17996 | 0.1137 | 0.09028 | Other dyspnea | respiratory | UricA | |
| 513 | 0.8625 | 0.07206 | 2.369 | 5.10e-33 | 26022 | 945 | 25077 | 0.9512 | 0.07662 | 2.589 | 2.16e-35 | 35312 | 811 | 34501 | 0.03484 | 0.2135 | Respiratory abnormalities | respiratory | UricA | |
| 513.3 | | | | | 25286 | 209 | 25077 | | | | | 34707 | 206 | 34501 | | | Hypoventilation | respiratory | UricA | |
| 513.31 | | | | | 25126 | 49 | 25077 | | | | | 34541 | 40 | 34501 | | | Apnea | respiratory | UricA | |
| 513.32 | | | | | 25149 | 72 | 25077 | | | | | 34549 | 48 | 34501 | | | Orthopnea | respiratory | UricA | |
| 513.4 | | | | | 25445 | 368 | 25077 | | | | | 34837 | 336 | 34501 | 0.04622 | 0.3252 | Hyperventilation | respiratory | UricA | |
| 513.8 | | | | | 25261 | 184 | 25077 | | | | | 34580 | 79 | 34501 | | | Disorders of diaphragm | respiratory | UricA | |

| phenotype | male_beta | male_SE | male_OR | male_p | male_n_total | male_n_cases | male_n_controls | female_beta | female_SE | female_OR | female_p | female_n_total | female_n_cases | female_n_controls | interaction_p | interaction_beta | description | group | lab | label |
|---|---|---|---|---|---|---|---|---|---|---|---|---|---|---|---|---|---|---|---|---|
| 514 | 0.8016 | 0.04324 | 2.229 | 9.81e-77 | 24679 | 2921 | 21758 | 0.8261 | 0.04589 | 2.284 | 1.86e-72 | 33770 | 2584 | 31186 | 0.2172 | 0.07557 | Abnormal findings examination of lungs | respiratory | UricA | |
| 514.1 | | | | | 21848 | 90 | 21758 | | | | | 31294 | 108 | 31186 | | | Abnormal results of function study of pulmonary system | respiratory | UricA | |
| 514.2 | 0.5528 | 0.07003 | 1.738 | 2.94e-15 | 22786 | 1028 | 21758 | 0.4303 | 0.07439 | 1.538 | 7.27e-09 | 32192 | 1006 | 31186 | 0.5473 | -0.05958 | Solitary pulmonary nodule | respiratory | UricA | |
| 516 | 0.8062 | 0.09206 | 2.239 | 2.01e-18 | 27219 | 552 | 26667 | | | | | 36488 | 334 | 36154 | 0.3171 | 0.1447 | Abnormal sputum | respiratory | UricA | |
| 516.1 | | | | | 27145 | 478 | 26667 | | | | | 36446 | 292 | 36154 | 0.08751 | 0.2632 | Hemoptysis | respiratory | UricA | |
| 519 | 0.6912 | 0.06142 | 1.996 | 2.22e-29 | 25596 | 1340 | 24256 | 0.7111 | 0.06203 | 2.036 | 1.98e-30 | 34332 | 1351 | 32981 | 0.08253 | 0.1462 | Other diseases of respiratory system, not elsewhere classified | respiratory | UricA | |
| 519.1 | | | | | 24327 | 71 | 24256 | | | | | 33033 | 52 | 32981 | | | Tracheostomy complications | respiratory | UricA | |
| 519.2 | | | | | 24383 | 127 | 24256 | | | | | 33036 | 55 | 32981 | | | Respiratory complications | respiratory | UricA | |
| 519.8 | | | | | 24432 | 176 | 24256 | | | | | 33153 | 172 | 32981 | | | Other diseases of respiratory system, NEC | respiratory | UricA | |
| 519.9 | 0.7136 | 0.07243 | 2.041 | 6.68e-23 | 25196 | 940 | 24256 | 0.7135 | 0.071 | 2.041 | 9.30e-24 | 34006 | 1025 | 32981 | 0.1859 | 0.1295 | Symptoms involving respiratory system and other chest symptoms | respiratory | UricA | |
| 520 | | | | | 25518 | 159 | 25359 | | | | | 33516 | 257 | 33259 | | | Disorders of tooth development | digestive | UricA | |
| 520.1 | | | | | 25370 | 11 | 25359 | | | | | 33270 | 11 | 33259 | | | Hereditary disturbances in tooth structure | digestive | UricA | |
| 520.2 | | | | | 25503 | 144 | 25359 | | | | | 33503 | 244 | 33259 | | | Disturbances in tooth eruption | digestive | UricA | |
| 521 | 0.627 | 0.08423 | 1.872 | 9.79e-14 | 26066 | 707 | 25359 | 0.486 | 0.07726 | 1.626 | 3.17e-10 | 34146 | 887 | 33259 | 0.9884 | -0.001593 | Diseases of hard tissues of teeth | digestive | UricA | |
| 521.1 | 0.6432 | 0.0851 | 1.903 | 4.09e-14 | 26050 | 691 | 25359 | 0.4819 | 0.07835 | 1.619 | 7.72e-10 | 34122 | 863 | 33259 | 0.8595 | -0.01968 | Dental caries | digestive | UricA | |
| 521.2 | | | | | 25360 | 1 | 25359 | | | | | 33261 | 2 | 33259 | | | Dental abrasion, erosion and attrition | digestive | UricA | |
| 521.4 | | | | | 25359 | | | | | | | 33267 | 8 | 33259 | | | Tooth complications likely association with other diseases | digestive | UricA | |
| 522 | | | | | 25451 | 92 | 25359 | | | | | 33377 | 118 | 33259 | | | Diseases of pulp and periapical tissues | digestive | UricA | |
| 522.1 | | | | | 25361 | 2 | 25359 | | | | | 33263 | 4 | 33259 | | | Pulpitis and necrosis of tooth pulp | digestive | UricA | |
| 522.5 | | | | | 25445 | 86 | 25359 | | | | | 33365 | 106 | 33259 | | | Periapical abscess | digestive | UricA | |
| 523 | | | | | 25698 | 339 | 25359 | 0.2108 | 0.1011 | 1.235 | 0.03711 | 33819 | 560 | 33259 | 0.01059 | -0.3818 | Gingival and periodontal diseases | digestive | UricA | |
| 523.1 | | | | | 25536 | 177 | 25359 | | | | | 33688 | 429 | 33259 | 0.3033 | -0.2027 | Gingivitis | digestive | UricA | |
| 523.3 | | | | | 25492 | 133 | 25359 | | | | | 33407 | 148 | 33259 | | | Periodontitis (acute or chronic) | digestive | UricA | |
| 523.31 | | | | | 25454 | 95 | 25359 | | | | | 33368 | 109 | 33259 | | | Acute periodontitis | digestive | UricA | |
| 523.32 | | | | | 25382 | 23 | 25359 | | | | | 33285 | 26 | 33259 | | | Chronic periodontitis | digestive | UricA | |
| 524 | | | | | 25495 | 136 | 25359 | | | | | 33518 | 259 | 33259 | | | Dentofacial anomalies, including malocclusion | digestive | UricA | |
| 524.3 | | | | | 25481 | 122 | 25359 | | | | | 33493 | 234 | 33259 | | | Anomalies of tooth position/malocclusion | digestive | UricA | |
| 525 | | | | | 25595 | 236 | 25359 | | | | | 33641 | 382 | 33259 | 0.2601 | 0.1987 | Other diseases of the teeth and supporting structures | digestive | UricA | |
| 525.1 | | | | | 25495 | 136 | 25359 | | | | | 33466 | 207 | 33259 | | | Loss of teeth or edentulism | digestive | UricA | |
| 525.2 | | | | | 25366 | 7 | 25359 | | | | | 33279 | 20 | 33259 | | | Atrophy of edentulous alveolar ridge | digestive | UricA | |
| 526 | | | | | 25636 | 277 | 25359 | 0.3524 | 0.09653 | 1.422 | 2.62e-04 | 33838 | 579 | 33259 | 0.005422 | 0.4664 | Diseases of the jaws | digestive | UricA | |
| 526.1 | | | | | 25372 | 13 | 25359 | | | | | 33270 | 11 | 33259 | | | Cysts of the jaws | digestive | UricA | |
| 526.3 | | | | | 25393 | 34 | 25359 | | | | | 33336 | 77 | 33259 | | | Anomalies of jaw size/symmetry | digestive | UricA | |
| 526.4 | | | | | 25426 | 67 | 25359 | | | | | 33523 | 264 | 33259 | | | Temporomandibular joint disorders | digestive | UricA | |
| 526.41 | | | | | 25407 | 48 | 25359 | | | | | 33417 | 158 | 33259 | | | Temporomandibular joint disorder, unspecified | digestive | UricA | |

| phenotype | male_beta | male_SE | male_OR | male_p | male_n_total | male_n_cases | male_n_controls | female_beta | female_SE | female_OR | female_p | female_n_total | female_n_cases | female_n_controls | interaction_p | interaction_beta | description | group | lab | label |
|---|---|---|---|---|---|---|---|---|---|---|---|---|---|---|---|---|---|---|---|---|
| 526.42 | | | | | 25364 | 5 | 25359 | | | | | 33300 | 41 | 33259 | | | Arthralgia/ankylosis of temporomandibular joint | digestive | UricA | |
| 526.5 | | | | | 25416 | 57 | 25359 | | | | | 33320 | 61 | 33259 | | | Inflammatory conditions of jaw | digestive | UricA | |
| 526.8 | | | | | 25363 | 4 | 25359 | | | | | 33267 | 8 | 33259 | | | Exostosis of jaw | digestive | UricA | |
| 526.9 | | | | | 25414 | 55 | 25359 | | | | | 33304 | 45 | 33259 | | | Jaw disease NOS | digestive | UricA | |
| 527 | | | | | 25962 | 420 | 25542 | | | | | 34324 | 353 | 33971 | 0.001642 | 0.4999 | Diseases of the salivary glands | digestive | UricA | |
| 527.1 | | | | | 25545 | 3 | 25542 | | | | | 33976 | 5 | 33971 | | | Hypertrophy of salivary gland | digestive | UricA | |
| 527.2 | | | | | 25596 | 54 | 25542 | | | | | 34069 | 98 | 33971 | | | Sialoadenitis | digestive | UricA | |
| 527.7 | | | | | 25871 | 329 | 25542 | | | | | 34176 | 205 | 33971 | 0.001539 | 0.6478 | Disturbance of salivary secretion | digestive | UricA | |
| 527.8 | | | | | 25561 | 19 | 25542 | | | | | 34003 | 32 | 33971 | | | Other specified diseases of the salivary glands | digestive | UricA | |
| 528 | 0.8325 | 0.0784 | 2.299 | 2.46e-26 | 26317 | 775 | 25542 | 0.7069 | 0.07519 | 2.028 | 5.34e-21 | 34849 | 878 | 33971 | 0.6848 | 0.04249 | Diseases of the oral soft tissues, excluding lesions specific for gingiva and tongue | digestive | UricA | |
| 528.1 | | | | | 25938 | 396 | 25542 | | | | | 34341 | 370 | 33971 | 0.2851 | 0.1609 | Stomatitis and mucositis | digestive | UricA | |
| 528.11 | | | | | 25631 | 89 | 25542 | | | | | 34056 | 85 | 33971 | | | Stomatitis and mucositis (ulcerative) | digestive | UricA | |
| 528.12 | | | | | 25583 | 41 | 25542 | | | | | 34026 | 55 | 33971 | | | Oral aphthae | digestive | UricA | |
| 528.3 | | | | | 25579 | 37 | 25542 | | | | | 34020 | 49 | 33971 | | | Cellulitis and abscess of oral soft tissues | digestive | UricA | |
| 528.4 | | | | | 25551 | 9 | 25542 | | | | | 33984 | 13 | 33971 | | | Cysts of oral soft tissues | digestive | UricA | |
| 528.41 | | | | | 25551 | 9 | 25542 | | | | | 33982 | 11 | 33971 | | | Cyst of the salivary gland | digestive | UricA | |
| 528.5 | | | | | 25587 | 45 | 25542 | | | | | 34050 | 79 | 33971 | | | Diseases of lips | digestive | UricA | |
| 528.6 | | | | | 25578 | 36 | 25542 | | | | | 34015 | 44 | 33971 | | | Leukoplakia of oral mucosa | digestive | UricA | |
| 528.7 | | | | | 25551 | 9 | 25542 | | | | | 33984 | 13 | 33971 | | | Sialolithiasis | digestive | UricA | |
| 529 | | | | | 25647 | 105 | 25542 | | | | | 34108 | 137 | 33971 | | | Diseases and other conditions of the tongue | digestive | UricA | |
| 529.1 | | | | | 25565 | 23 | 25542 | | | | | 34012 | 41 | 33971 | | | Glossitis | digestive | UricA | |
| 529.6 | | | | | 25565 | 23 | 25542 | | | | | 34004 | 33 | 33971 | | | Glossodynia | digestive | UricA | |
| 530 | 0.5788 | 0.03466 | 1.784 | 1.38e-62 | 22713 | 7010 | 15703 | 0.5572 | 0.03301 | 1.746 | 6.06e-64 | 30970 | 8921 | 22049 | 0.2609 | 0.05291 | Diseases of esophagus | digestive | UricA | |
| 530.1 | 0.6204 | 0.03569 | 1.86 | 1.11e-67 | 22053 | 6350 | 15703 | 0.5678 | 0.03359 | 1.764 | 3.97e-64 | 30445 | 8396 | 22049 | 0.7415 | 0.0159 | Esophagitis, GERD and related diseases | digestive | UricA | |
| 530.11 | 0.6321 | 0.03661 | 1.882 | 8.78e-67 | 21565 | 5862 | 15703 | 0.5773 | 0.03406 | 1.781 | 1.94e-64 | 30082 | 8033 | 22049 | 0.9824 | 0.001087 | GERD | digestive | UricA | |
| 530.12 | | | | | 15778 | 75 | 15703 | | | | | 22113 | 64 | 22049 | | | Ulcer of esophagus | digestive | UricA | |
| 530.13 | | | | | 16021 | 318 | 15703 | | | | | 22193 | 144 | 22049 | | | Barrett's esophagus | digestive | UricA | |
| 530.14 | | | | | 16000 | 297 | 15703 | | | | | 22411 | 362 | 22049 | 0.0263 | 0.3857 | Reflux esophagitis | digestive | UricA | |
| 530.15 | | | | | 15777 | 74 | 15703 | | | | | 22097 | 48 | 22049 | | | Eosinophilic esophagitis | digestive | UricA | |
| 530.2 | 0.5372 | 0.09277 | 1.711 | 7.01e-09 | 16327 | 624 | 15703 | | | | | 22465 | 416 | 22049 | 0.202 | 0.185 | Esophageal bleeding (varices/hemorrhage) | digestive | UricA | |
| 530.3 | | | | | 16140 | 437 | 15703 | | | | | 22429 | 380 | 22049 | 0.06356 | 0.2924 | Stricture and stenosis of esophagus | digestive | UricA | |
| 530.5 | | | | | 15823 | 120 | 15703 | | | | | 22257 | 208 | 22049 | | | Disorders of esophageal motility | digestive | UricA | |
| 530.6 | | | | | 15740 | 37 | 15703 | | | | | 22084 | 35 | 22049 | | | Diverticulum of esophagus, acquired | digestive | UricA | |
| 530.7 | | | | | 15735 | 32 | 15703 | | | | | 22067 | 18 | 22049 | | | Gastroesophageal laceration-hemorrhage syndrome | digestive | UricA | |
| 530.9 | | | | | 15920 | 217 | 15703 | | | | | 22322 | 273 | 22049 | | | Heartburn | digestive | UricA | |
| 531 | 0.6047 | 0.07886 | 1.831 | 1.74e-14 | 26644 | 757 | 25887 | 0.6774 | 0.07924 | 1.969 | 1.25e-17 | 35675 | 773 | 34902 | 0.311 | 0.1094 | Peptic ulcer (excl. esophageal) | digestive | UricA | |
| 531.1 | | | | | 26046 | 159 | 25887 | | | | | 35004 | 102 | 34902 | | | Hemorrhage from gastrointestinal ulcer | digestive | UricA | |
| 531.2 | | | | | 26063 | 176 | 25887 | | | | | 35135 | 233 | 34902 | | | Gastric ulcer | digestive | UricA | |

| phenotype | male_beta | male_SE | male_OR | male_p | male_n_total | male_n_cases | male_n_controls | female_beta | female_SE | female_OR | female_p | female_n_total | female_n_cases | female_n_controls | interaction_p | interaction_beta | description | group | lab | label |
|---|---|---|---|---|---|---|---|---|---|---|---|---|---|---|---|---|---|---|---|---|
| 531.3 | | | | | 26049 | 162 | 25887 | | | | | 35017 | 115 | 34902 | | | Duodenal ulcer | digestive | UricA | |
| 531.4 | | | | | 26128 | 241 | 25887 | | | | | 35173 | 271 | 34902 | 0.2188 | 0.2238 | Peptic ulcer, site unspecified | digestive | UricA | |
| 531.5 | | | | | 25904 | 17 | 25887 | | | | | 34952 | 50 | 34902 | | | Gastrojejunal ulcer | digestive | UricA | |
| 532 | 0.4787 | 0.04657 | 1.614 | 8.68e-25 | 18697 | 2994 | 15703 | 0.6323 | 0.04709 | 1.882 | 4.14e-41 | 25127 | 3078 | 22049 | 3.51e-05 | 0.2682 | Dysphagia | digestive | UricA | |
| 535 | 0.5784 | 0.08858 | 1.783 | 6.60e-11 | 24211 | 698 | 23513 | 0.5 | 0.07854 | 1.649 | 1.93e-10 | 32091 | 888 | 31203 | 0.442 | 0.08755 | Gastritis and duodenitis | digestive | UricA | |
| 535.1 | | | | | 23554 | 41 | 23513 | | | | | 31256 | 53 | 31203 | | | Acute gastritis | digestive | UricA | |
| 535.2 | | | | | 23753 | 240 | 23513 | | | | | 31519 | 316 | 31203 | 0.1118 | 0.3332 | Atrophic gastritis | digestive | UricA | |
| 535.6 | | | | | 23634 | 121 | 23513 | | | | | 31294 | 91 | 31203 | | | Duodenitis | digestive | UricA | |
| 535.8 | | | | | 23673 | 160 | 23513 | | | | | 31440 | 237 | 31203 | | | Other specified gastritis | digestive | UricA | |
| 535.9 | | | | | 23653 | 140 | 23513 | | | | | 31383 | 180 | 31203 | | | Gastritis and duodenitis, NOS | digestive | UricA | |
| 536 | 0.5238 | 0.09274 | 1.688 | 1.62e-08 | 24111 | 598 | 23513 | 0.5027 | 0.07101 | 1.653 | 1.45e-12 | 32274 | 1071 | 31203 | 0.38 | 0.09873 | Disorders of function of stomach | digestive | UricA | |
| 536.3 | | | | | 23695 | 182 | 23513 | | | | | 31598 | 395 | 31203 | 0.5671 | -0.1061 | Gastroparesis | digestive | UricA | |
| 536.7 | | | | | 23722 | 209 | 23513 | | | | | 31426 | 223 | 31203 | | | Complications of gastrostomy, colostomy and enterostomy | digestive | UricA | |
| 536.8 | | | | | 23738 | 225 | 23513 | 0.4179 | 0.1028 | 1.519 | 4.83e-05 | 31704 | 501 | 31203 | 0.9917 | -0.001784 | Dyspepsia and other specified disorders of function of stomach | digestive | UricA | |
| 537 | 0.5545 | 0.08501 | 1.741 | 6.87e-11 | 24195 | 682 | 23513 | 0.2958 | 0.08998 | 1.344 | 0.00101 | 31926 | 723 | 31203 | 0.09477 | -0.2007 | Other disorders of stomach and duodenum | digestive | UricA | |
| 537.1 | | | | | 23574 | 61 | 23513 | | | | | 31261 | 58 | 31203 | | | Lesions of stomach and duodenum | digestive | UricA | |
| 539 | | | | | 27703 | 290 | 27413 | 0.07869 | 0.08225 | 1.082 | 0.3387 | 36747 | 945 | 35802 | 0.1206 | -0.233 | Bariatric surgery | digestive | UricA | |
| 540 | | | | | 27635 | 152 | 27483 | | | | | 36729 | 193 | 36536 | | | Appendiceal conditions | digestive | UricA | |
| 540.1 | | | | | 27611 | 128 | 27483 | | | | | 36714 | 178 | 36536 | | | Appendicitis | digestive | UricA | |
| 540.11 | | | | | 27592 | 109 | 27483 | | | | | 36692 | 156 | 36536 | | | Acute appendicitis | digestive | UricA | |
| 550 | 0.3745 | 0.04468 | 1.454 | 5.22e-17 | 25722 | 2883 | 22839 | 0.4161 | 0.04827 | 1.516 | 6.73e-18 | 34582 | 2531 | 32051 | 0.8954 | 0.008409 | Abdominal hernia | digestive | UricA | |
| 550.1 | 0.3723 | 0.07162 | 1.451 | 2.01e-07 | 23813 | 974 | 22839 | | | | | 32167 | 116 | 32051 | 0.2678 | -0.2459 | Inguinal hernia | digestive | UricA | |
| 550.2 | 0.4668 | 0.07184 | 1.595 | 8.16e-11 | 23793 | 954 | 22839 | 0.438 | 0.0621 | 1.55 | 1.75e-12 | 33474 | 1423 | 32051 | 0.7729 | -0.02656 | Diaphragmatic hernia | digestive | UricA | |
| 550.3 | | | | | 22846 | 7 | 22839 | | | | | 32067 | 16 | 32051 | | | Femoral hernia | digestive | UricA | |
| 550.4 | | | | | 23250 | 411 | 22839 | | | | | 32268 | 217 | 32051 | 0.3974 | -0.1532 | Umbilical hernia | digestive | UricA | |
| 550.5 | | | | | 23322 | 483 | 22839 | 0.5277 | 0.09814 | 1.695 | 7.56e-08 | 32584 | 533 | 32051 | 0.0555 | 0.2651 | Ventral hernia | digestive | UricA | |
| 550.6 | 0.1998 | 0.1009 | 1.221 | 0.04755 | 23364 | 525 | 22839 | 0.4465 | 0.09744 | 1.563 | 4.59e-06 | 32610 | 559 | 32051 | 0.03762 | 0.2832 | Incisional hernia | digestive | UricA | |
| 555 | 0.1338 | 0.08793 | 1.143 | 0.128 | 18550 | 1092 | 17458 | -0.1223 | 0.07968 | 0.8849 | 0.1248 | 24902 | 1438 | 23464 | 0.2001 | -0.149 | Inflammatory bowel disease and other gastroenteritis and colitis | digestive | UricA | |
| 555.1 | 0.1088 | 0.101 | 1.115 | 0.2815 | 18342 | 884 | 17458 | -0.3049 | 0.09301 | 0.7372 | 0.001047 | 24671 | 1207 | 23464 | 0.02935 | -0.2935 | Regional enteritis | digestive | UricA | |
| 555.2 | 0.09732 | 0.118 | 1.102 | 0.4094 | 18001 | 543 | 17458 | 0.07859 | 0.1063 | 1.082 | 0.4596 | 24116 | 652 | 23464 | 0.654 | 0.06984 | Ulcerative colitis | digestive | UricA | |
| 555.21 | | | | | 17689 | 231 | 17458 | | | | | 23710 | 246 | 23464 | | | Ulcerative colitis (chronic) | digestive | UricA | |
| 556 | | | | | 17670 | 212 | 17458 | | | | | 23667 | 203 | 23464 | | | Ulceration of the lower GI tract | digestive | UricA | |
| 556.1 | | | | | 17633 | 175 | 17458 | | | | | 23632 | 168 | 23464 | | | Ulceration of intestine | digestive | UricA | |
| 556.11 | | | | | 17498 | 40 | 17458 | | | | | 23504 | 40 | 23464 | | | Angiodysplasia of intestine (without mention of hemorrhage) | digestive | UricA | |
| 557 | | | | | 17645 | 187 | 17458 | | | | | 23820 | 356 | 23464 | 0.5321 | 0.1322 | Intestinal malabsorption (non-celiac) | digestive | UricA | |
| 557.1 | | | | | 17538 | 80 | 17458 | | | | | 23734 | 270 | 23464 | | | Celiac disease | digestive | UricA | |
| 558 | 0.7484 | 0.07488 | 2.114 | 1.60e-23 | 18499 | 1041 | 17458 | 0.5027 | 0.06253 | 1.653 | 9.06e-16 | 25010 | 1546 | 23464 | 0.4207 | -0.076 | Noninfectious gastroenteritis | digestive | UricA | |
| 559 | | | | | 17873 | 415 | 17458 | | | | | 23811 | 347 | 23464 | 0.8659 | 0.03056 | Ileostomy status | digestive | UricA | |
| 560 | 0.5191 | 0.06538 | 1.681 | 2.03e-15 | 18805 | 1347 | 17458 | 0.4962 | 0.07079 | 1.642 | 2.39e-12 | 24662 | 1198 | 23464 | 0.7283 | 0.03279 | Intestinal obstruction without mention of hernia | digestive | UricA | |

| phenotype | male_beta | male_SE | male_OR | male_p | male_n_total | male_n_cases | male_n_controls | female_beta | female_SE | female_OR | female_p | female_n_total | female_n_cases | female_n_controls | interaction_p | interaction_beta | description | group | lab | label |
|---|---|---|---|---|---|---|---|---|---|---|---|---|---|---|---|---|---|---|---|---|
| 560.1 | 0.6456 | 0.08603 | 1.907 | 6.17e-14 | 18153 | 695 | 17458 | | | | | 23849 | 385 | 23464 | 0.7426 | 0.04684 | Paralytic ileus | digestive | UricA | |
| 560.2 | | | | | 17514 | 56 | 17458 | | | | | 23568 | 104 | 23464 | | | Impaction of intestine | digestive | UricA | |
| 560.3 | | | | | 17538 | 80 | 17458 | | | | | 23554 | 90 | 23464 | | | Peritoneal or intestinal adhesions | digestive | UricA | |
| 560.4 | 0.4614 | 0.09097 | 1.586 | 3.92e-07 | 18149 | 691 | 17458 | 0.4181 | 0.08958 | 1.519 | 3.06e-06 | 24229 | 765 | 23464 | 0.991 | 0.001403 | Other intestinal obstruction | digestive | UricA | |
| 561 | 0.9258 | 0.05606 | 2.524 | 2.88e-61 | 19227 | 1769 | 17458 | 0.5854 | 0.04653 | 1.796 | 2.71e-36 | 26443 | 2979 | 23464 | 0.001726 | -0.2226 | Symptoms involving digestive system | digestive | UricA | |
| 562 | 0.7854 | 0.05079 | 2.193 | 6.13e-54 | 19778 | 2320 | 17458 | 0.6577 | 0.05415 | 1.93 | 6.09e-34 | 25685 | 2221 | 23464 | 0.1789 | -0.0983 | Diverticulosis and diverticulitis | digestive | UricA | |
| 562.1 | 0.7867 | 0.05281 | 2.196 | 3.50e-50 | 19570 | 2112 | 17458 | 0.6484 | 0.05733 | 1.913 | 1.16e-29 | 25403 | 1939 | 23464 | 0.1755 | -0.104 | Diverticulosis | digestive | UricA | |
| 562.2 | | | | | 17837 | 379 | 17458 | 0.9178 | 0.09788 | 2.504 | 6.83e-21 | 23966 | 502 | 23464 | 0.7602 | 0.04378 | Diverticulitis | digestive | UricA | |
| 564 | 0.5504 | 0.06451 | 1.734 | 1.43e-17 | 18857 | 1399 | 17458 | 0.5126 | 0.04989 | 1.67 | 9.09e-25 | 26038 | 2574 | 23464 | 0.4085 | 0.06555 | Functional digestive disorders | digestive | UricA | |
| 564.1 | | | | | 17914 | 456 | 17458 | 0.4802 | 0.0616 | 1.616 | 6.39e-15 | 25043 | 1579 | 23464 | 0.6865 | -0.04786 | Irritable Bowel Syndrome | digestive | UricA | |
| 564.9 | | | | | 17664 | 206 | 17458 | | | | | 23673 | 209 | 23464 | | | Personal history of diseases of digestive system | digestive | UricA | |
| 565 | 0.6103 | 0.07096 | 1.841 | 7.93e-18 | 23018 | 1093 | 21925 | 0.1702 | 0.07559 | 1.186 | 0.02436 | 31069 | 1106 | 29963 | 0.006832 | -0.271 | Anal and rectal conditions | digestive | UricA | |
| 565.1 | | | | | 22052 | 127 | 21925 | | | | | 30077 | 114 | 29963 | | | Anal and rectal polyp | digestive | UricA | |
| 567 | 0.482 | 0.09176 | 1.619 | 1.49e-07 | 22577 | 652 | 21925 | 0.6802 | 0.09446 | 1.974 | 6.00e-13 | 30541 | 578 | 29963 | 0.01204 | 0.3216 | Peritonitis and retroperitoneal infections | digestive | UricA | |
| 568 | | | | | 22412 | 487 | 21925 | 0.6178 | 0.1007 | 1.855 | 8.50e-10 | 30476 | 513 | 29963 | 0.002852 | 0.4294 | Other disorders of peritoneum | digestive | UricA | |
| 568.1 | | | | | 22164 | 239 | 21925 | | | | | 30277 | 314 | 29963 | 0.00404 | 0.5912 | Peritoneal adhesions (postoperative) (postinfection) | digestive | UricA | |
| 569 | 0.382 | 0.06254 | 1.465 | 1.01e-09 | 23419 | 1494 | 21925 | 0.353 | 0.05521 | 1.423 | 1.63e-10 | 31975 | 2012 | 29963 | 0.2845 | 0.08655 | Other disorders of intestine | digestive | UricA | |
| 569.1 | | | | | 21954 | 29 | 21925 | | | | | 30021 | 58 | 29963 | | | Toxic gastroenteritis and colitis | digestive | UricA | |
| 569.2 | 0.3226 | 0.09891 | 1.381 | 0.001108 | 22496 | 571 | 21925 | 0.2073 | 0.08045 | 1.23 | 0.009959 | 30942 | 979 | 29963 | 0.6748 | -0.05189 | Gastrointestinal complications | digestive | UricA | |
| 571 | 0.7214 | 0.0494 | 2.057 | 2.66e-48 | 21552 | 2387 | 19165 | 0.617 | 0.05124 | 1.853 | 2.12e-33 | 29934 | 2235 | 27699 | 0.7801 | 0.0192 | Chronic liver disease and cirrhosis | digestive | UricA | |
| 571.5 | 0.728 | 0.0509 | 2.071 | 2.14e-46 | 21382 | 2217 | 19165 | 0.6091 | 0.05317 | 1.839 | 2.25e-30 | 29754 | 2055 | 27699 | 0.8936 | 0.00951 | Other chronic nonalcoholic liver disease | digestive | UricA | |
| phenotype | male_beta | male_SE | male_OR | male_p | male_n_total | male_n_cases | male_n_controls | female_beta | female_SE | female_OR | female_p | female_n_total | female_n_cases | female_n_controls | interaction_p | interaction_beta | description | group | lab | label |
| 571.51 | 0.5624 | 0.06592 | 1.755 | 1.44e-17 | 20477 | 1312 | 19165 | 0.5557 | 0.07793 | 1.743 | 9.94e-13 | 28658 | 959 | 27699 | 0.5207 | 0.06351 | Cirrhosis of liver without mention of alcohol | digestive | UricA | |
| 571.6 | | | | | 19242 | 77 | 19165 | | | | | 27883 | 184 | 27699 | | | Primary biliary cirrhosis | digestive | UricA | |
| 571.8 | 0.5899 | 0.07485 | 1.804 | 3.23e-15 | 20161 | 996 | 19165 | 0.6379 | 0.08703 | 1.893 | 2.30e-13 | 28437 | 738 | 27699 | 0.3109 | 0.1129 | Liver abscess and sequelae of chronic liver disease | digestive | UricA | |
| 571.81 | 0.5913 | 0.08873 | 1.806 | 2.67e-11 | 19856 | 691 | 19165 | | | | | 28168 | 469 | 27699 | 0.3905 | 0.1165 | Portal hypertension | digestive | UricA | |
| 572 | 0.8015 | 0.06325 | 2.229 | 8.52e-37 | 20559 | 1394 | 19165 | 0.7606 | 0.06787 | 2.14 | 3.79e-29 | 28923 | 1224 | 27699 | 0.7085 | 0.03365 | Ascites (non malignant) | digestive | UricA | |
| 573 | 0.8594 | 0.07259 | 2.362 | 2.48e-32 | 20126 | 961 | 19165 | 0.8343 | 0.07336 | 2.303 | 5.68e-30 | 28655 | 956 | 27699 | 0.7027 | 0.03807 | Other disorders of liver | digestive | UricA | |
| 573.1 | | | | | 19195 | 30 | 19165 | | | | | 27731 | 32 | 27699 | | | Chronic passive congestion of liver | digestive | UricA | |
| 573.2 | | | | | 19596 | 431 | 19165 | | | | | 27945 | 246 | 27699 | 0.00795 | 0.4368 | Liver replaced by transplant | digestive | UricA | |
| 573.3 | | | | | 19292 | 127 | 19165 | | | | | 27860 | 161 | 27699 | | | Hepatomegaly | digestive | UricA | |
| 573.4 | | | | | 19356 | 191 | 19165 | | | | | 27881 | 182 | 27699 | | | Acute and subacute necrosis of liver | digestive | UricA | |
| 573.5 | | | | | 19606 | 441 | 19165 | | | | | 28006 | 307 | 27699 | 0.5268 | 0.1003 | Jaundice (not of newborn) | digestive | UricA | |

| phenotype | male_beta | male_SE | male_OR | male_p | male_n_total | male_n_cases | male_n_controls | female_beta | female_SE | female_OR | female_p | female_n_total | female_n_cases | female_n_controls | interaction_p | interaction_beta | description | group | lab | label |
|---|---|---|---|---|---|---|---|---|---|---|---|---|---|---|---|---|---|---|---|---|
| 573.6 | 1.359 | 0.08788 | 3.892 | 6.21e-54 | 19797 | 632 | 19165 | 1.02 | 0.09069 | 2.772 | 2.48e-29 | 28268 | 569 | 27699 | 0.1331 | -0.1817 | Nonspecific elevation of levels of transaminase or lactic acid dehydrogenase [LDH] | digestive | UricA | |
| 573.7 | 0.7938 | 0.07397 | 2.212 | 7.33e-27 | 20119 | 954 | 19165 | 0.6501 | 0.06858 | 1.916 | 2.56e-21 | 28828 | 1129 | 27699 | 0.9532 | -0.005681 | Abnormal results of function study of liver | digestive | UricA | |
| 573.9 | 0.7929 | 0.08017 | 2.21 | 4.61e-23 | 19944 | 779 | 19165 | 0.7628 | 0.07558 | 2.144 | 5.99e-24 | 28587 | 888 | 27699 | 0.5249 | 0.06755 | Abnormal serum enzyme levels | digestive | UricA | |
| 574 | 0.6684 | 0.06152 | 1.951 | 1.69e-27 | 25697 | 1283 | 24414 | 0.5815 | 0.05924 | 1.789 | 9.63e-23 | 34656 | 1503 | 33153 | 0.4279 | -0.06558 | Cholelithiasis and cholecystitis | digestive | UricA | |
| 574.1 | 0.708 | 0.06914 | 2.03 | 1.33e-24 | 25404 | 990 | 24414 | 0.5877 | 0.06656 | 1.8 | 1.06e-18 | 34320 | 1167 | 33153 | 0.2101 | -0.1164 | Cholelithiasis | digestive | UricA | |
| 574.11 | | | | | 24503 | 89 | 24414 | | | | | 33224 | 71 | 33153 | | | Cholelithiasis with acute cholecystitis | digestive | UricA | |
| 574.12 | | | | | 24528 | 114 | 24414 | | | | | 33340 | 187 | 33153 | | | Cholelithiasis with other cholecystitis | digestive | UricA | |
| 574.2 | | | | | 24625 | 211 | 24414 | | | | | 33383 | 230 | 33153 | | | Calculus of bile duct | digestive | UricA | |
| 574.3 | | | | | 24711 | 297 | 24414 | | | | | 33444 | 291 | 33153 | 0.7058 | 0.0659 | Cholecystitis without cholelithiasis | digestive | UricA | |
| 575 | 0.5218 | 0.07697 | 1.685 | 1.21e-11 | 25285 | 871 | 24414 | 0.7815 | 0.07267 | 2.185 | 5.67e-27 | 34100 | 947 | 33153 | 0.001643 | 0.3222 | Other biliary tract disease | digestive | UricA | |
| 575.1 | | | | | 24612 | 198 | 24414 | | | | | 33277 | 124 | 33153 | | | Cholangitis | digestive | UricA | |
| 575.2 | | | | | 24699 | 285 | 24414 | | | | | 33392 | 239 | 33153 | 0.001342 | 0.6101 | Obstruction of bile duct | digestive | UricA | |
| 575.6 | | | | | 24435 | 21 | 24414 | | | | | 33185 | 32 | 33153 | | | Cholesterolosis of gallbladder | digestive | UricA | |
| 575.7 | | | | | 24611 | 197 | 24414 | | | | | 33370 | 217 | 33153 | | | Other disorders of gallbladder | digestive | UricA | |
| 575.8 | | | | | 24783 | 369 | 24414 | | | | | 33584 | 431 | 33153 | 0.04769 | 0.301 | Other disorders of biliary tract | digestive | UricA | |
| 575.9 | | | | | 24488 | 74 | 24414 | | | | | 33244 | 91 | 33153 | | | Nonspecific abnormal findings on radiological and other examination of biliary tract | digestive | UricA | |
| 577 | 0.4433 | 0.0773 | 1.558 | 9.75e-09 | 27246 | 863 | 26383 | 0.5002 | 0.07977 | 1.649 | 3.59e-10 | 36384 | 840 | 35544 | 0.2662 | 0.1192 | Diseases of pancreas | digestive | UricA | |
| 577.1 | | | | | 26693 | 310 | 26383 | | | | | 35828 | 284 | 35544 | 0.3112 | 0.181 | Acute pancreatitis | digestive | UricA | |
| 577.2 | | | | | 26568 | 185 | 26383 | | | | | 35726 | 182 | 35544 | | | Chronic pancreatitis | digestive | UricA | |
| 577.3 | | | | | 26620 | 237 | 26383 | | | | | 35810 | 266 | 35544 | 0.3673 | 0.1777 | Cyst and pseudocyst of pancreas | digestive | UricA | |
| 578 | 0.6918 | 0.0479 | 1.997 | 2.81e-47 | 23398 | 2404 | 20994 | 0.5596 | 0.04807 | 1.75 | 2.53e-31 | 31110 | 2548 | 28562 | 0.316 | -0.06606 | Gastrointestinal hemorrhage | digestive | UricA | |
| 578.1 | | | | | 21273 | 279 | 20994 | | | | | 28819 | 257 | 28562 | 0.3281 | 0.1797 | Hematemesis | digestive | UricA | |
| 578.2 | 0.91 | 0.06654 | 2.484 | 1.43e-42 | 22097 | 1103 | 20994 | 0.6047 | 0.06714 | 1.831 | 2.13e-19 | 29747 | 1185 | 28562 | 0.01411 | -0.2246 | Blood in stool | digestive | UricA | |
| 578.8 | 0.5394 | 0.09037 | 1.715 | 2.40e-09 | 21622 | 628 | 20994 | 0.3773 | 0.08807 | 1.458 | 1.84e-05 | 29288 | 726 | 28562 | 0.4286 | -0.09652 | Hemorrhage of rectum and anus | digestive | UricA | |
| 578.9 | 0.8161 | 0.0684 | 2.262 | 8.19e-33 | 22034 | 1040 | 20994 | 0.9091 | 0.07736 | 2.482 | 6.91e-32 | 29380 | 818 | 28562 | 0.3391 | 0.09559 | Hemorrhage of gastrointestinal tract | digestive | UricA | |
| 579 | 0.8855 | 0.05834 | 2.424 | 4.91e-52 | 22498 | 1504 | 20994 | 0.6974 | 0.05236 | 2.009 | 1.77e-40 | 30586 | 2024 | 28562 | 0.2088 | -0.09538 | Other symptoms involving abdomen and pelvis | digestive | UricA | |
| 579.2 | 1.34 | 0.0899 | 3.821 | 2.84e-50 | 21574 | 580 | 20994 | | | | | 28950 | 388 | 28562 | 0.5235 | -0.08668 | Splenomegaly | digestive | UricA | |
| 579.8 | | | | | 21190 | 196 | 20994 | | | | | 28810 | 248 | 28562 | | | Nonspecific abnormal findings in stool contents | digestive | UricA | |
| 580 | 2.114 | 0.08745 | 8.28 | 4.36e-129 | 18292 | 656 | 17636 | 2.1 | 0.08779 | 8.164 | 1.95e-126 | 28613 | 588 | 28025 | 0.8203 | 0.02726 | Nephritis; nephrosis; renal sclerosis | genitourinary | UricA | |
| 580.1 | | | | | 17811 | 175 | 17636 | | | | | 28185 | 160 | 28025 | | | Glomerulonephritis | genitourinary | UricA | |
| 580.11 | | | | | 17690 | 54 | 17636 | | | | | 28073 | 48 | 28025 | | | Proliferative glomerulonephritis | genitourinary | UricA | |
| 580.12 | | | | | 17715 | 79 | 17636 | | | | | 28084 | 59 | 28025 | | | Non-proliferative glomerulonephritis | genitourinary | UricA | |
| 580.13 | | | | | 17654 | 18 | 17636 | | | | | 28048 | 23 | 28025 | | | Acute glomerulonephritis, NOS | genitourinary | UricA | |
| 580.14 | | | | | 17676 | 40 | 17636 | | | | | 28076 | 51 | 28025 | | | Chronic glomerulonephritis, NOS | genitourinary | UricA | |
| 580.2 | | | | | 17776 | 140 | 17636 | | | | | 28143 | 118 | 28025 | | | Nephrotic syndrome without mention of glomerulonephritis | genitourinary | UricA | |

| phenotype | male_beta | male_SE | male_OR | male_p | male_n_total | male_n_cases | male_n_controls | female_beta | female_SE | female_OR | female_p | female_n_total | female_n_cases | female_n_controls | interaction_p | interaction_beta | description | group | lab | label |
|---|---|---|---|---|---|---|---|---|---|---|---|---|---|---|---|---|---|---|---|---|
| 580.3 | | | | | 17973 | 337 | 17636 | | | | | 28368 | 343 | 28025 | 0.8337 | 0.03374 | Nephritis and nephropathy without mention of glomerulonephritis | genitourinary | UricA | |
| 580.31 | | | | | 17810 | 174 | 17636 | | | | | 28242 | 217 | 28025 | | | Nephritis and nephropathy in diseases classified elsewhere | genitourinary | UricA | |
| 580.32 | | | | | 17795 | 159 | 17636 | | | | | 28162 | 137 | 28025 | | | Nephritis and nephropathy with pathological lesion | genitourinary | UricA | |
| 580.4 | | | | | 17734 | 98 | 17636 | | | | | 28125 | 100 | 28025 | | | Renal sclerosis, NOS | genitourinary | UricA | |
| 585 | 1.365 | 0.03465 | 3.915 | 0 | 24023 | 6387 | 17636 | 1.394 | 0.03619 | 4.032 | 0 | 32893 | 4868 | 28025 | 0.5228 | 0.03127 | Renal failure | genitourinary | UricA | |
| 585.1 | 1.383 | 0.03792 | 3.987 | 3.44e-291 | 22356 | 4720 | 17636 | 1.395 | 0.04066 | 4.033 | 7.58e-258 | 31559 | 3534 | 28025 | 0.6469 | 0.02482 | Acute renal failure | genitourinary | UricA | |
| 585.2 | 1.911 | 0.09095 | 6.756 | 5.64e-98 | 18192 | 556 | 17636 | | | | | 28438 | 413 | 28025 | 0.1812 | 0.1789 | Renal failure NOS | genitourinary | UricA | |
| 585.3 | 1.719 | 0.04172 | 5.579 | 0 | 21314 | 3678 | 17636 | 1.776 | 0.04622 | 5.904 | 0 | 30569 | 2544 | 28025 | 0.3958 | 0.05153 | Chronic renal failure [CKD] | genitourinary | UricA | |
| 585.31 | 1.903 | 0.083 | 6.708 | 2.27e-116 | 18326 | 690 | 17636 | | | | | 28496 | 471 | 28025 | 0.0748 | 0.2217 | Renal dialysis | genitourinary | UricA | |
| 585.32 | 1.87 | 0.07018 | 6.487 | 2.19e-156 | 18620 | 984 | 17636 | 2.086 | 0.08207 | 8.051 | 1.66e-142 | 28699 | 674 | 28025 | 0.01865 | 0.2468 | End stage renal disease | genitourinary | UricA | |
| 585.33 | 2.021 | 0.05504 | 7.545 | 4.49e-295 | 19559 | 1923 | 17636 | 1.949 | 0.0614 | 7.019 | 4.57e-221 | 29378 | 1353 | 28025 | 0.3922 | -0.06881 | Chronic Kidney Disease, Stage III | genitourinary | UricA | |
| 585.34 | 2.208 | 0.07594 | 9.099 | 6.69e-186 | 18575 | 939 | 17636 | 2.353 | 0.08457 | 10.52 | 2.22e-170 | 28729 | 704 | 28025 | 0.2741 | 0.1211 | Chronic Kidney Disease, Stage IV | genitourinary | UricA | |
| 585.4 | 2.317 | 0.0825 | 10.14 | 1.48e-173 | 18446 | 810 | 17636 | 1.972 | 0.08516 | 7.182 | 1.42e-118 | 28650 | 625 | 28025 | 0.002217 | -0.3533 | Chronic kidney disease, Stage I or II | genitourinary | UricA | |
| 586 | 1.275 | 0.04258 | 3.579 | 5.48e-197 | 21065 | 3429 | 17636 | 1.328 | 0.04756 | 3.775 | 1.09e-171 | 30360 | 2335 | 28025 | 0.6903 | 0.02488 | Other disorders of the kidney and ureters | genitourinary | UricA | |
| phenotype | male_beta | male_SE | male_OR | male_p | male_n_total | male_n_cases | male_n_controls | female_beta | female_SE | female_OR | female_p | female_n_total | female_n_cases | female_n_controls | interaction_p | interaction_beta | description | group | lab | label |
| 586.1 | | | | | 17687 | 51 | 17636 | | | | | 28103 | 78 | 28025 | | | Anatomical abnormatilies of kidney and ureters | genitourinary | UricA | |
| 586.11 | | | | | 17638 | 2 | 17636 | | | | | 28033 | 8 | 28025 | | | Small kidney | genitourinary | UricA | |
| 586.12 | | | | | 17684 | 48 | 17636 | | | | | 28097 | 72 | 28025 | | | Vesicoureteral reflux | genitourinary | UricA | |
| 586.2 | 1.23 | 0.0758 | 3.423 | 2.96e-59 | 18502 | 866 | 17636 | 1.212 | 0.09431 | 3.359 | 9.01e-38 | 28545 | 520 | 28025 | 0.6617 | -0.05144 | Cyst of kidney, acquired | genitourinary | UricA | |
| 586.3 | | | | | 17685 | 49 | 17636 | | | | | 28057 | 32 | 28025 | | | Vascular disorders of kidney/hypertrophy | genitourinary | UricA | |
| 586.4 | 0.8655 | 0.08527 | 2.376 | 3.32e-24 | 18326 | 690 | 17636 | | | | | 28313 | 288 | 28025 | 0.1064 | -0.247 | Stricture/obstruction of ureter | genitourinary | UricA | |
| 587 | 2.58 | 0.09996 | 13.2 | 6.29e-147 | 18201 | 565 | 17636 | | | | | 28416 | 391 | 28025 | 0.3318 | 0.1437 | Kidney replaced by transpant | genitourinary | UricA | |
| 588 | 2.157 | 0.07583 | 8.643 | 5.93e-178 | 18557 | 921 | 17636 | 2.188 | 0.08307 | 8.919 | 6.49e-153 | 28718 | 693 | 28025 | 0.6113 | 0.05568 | Disorders resulting from impaired renal function | genitourinary | UricA | |
| 588.1 | 2.159 | 0.08345 | 8.658 | 1.60e-147 | 18381 | 745 | 17636 | 2.257 | 0.09509 | 9.557 | 1.48e-124 | 28555 | 530 | 28025 | 0.304 | 0.1267 | Renal osteodystrophy | genitourinary | UricA | |
| 588.2 | | | | | 17925 | 289 | 17636 | | | | | 28246 | 221 | 28025 | 0.2657 | 0.2166 | Secondary hyperparathyroidism (of renal origin) | genitourinary | UricA | |
| 589 | | | | | 17733 | 97 | 17636 | | | | | 28140 | 115 | 28025 | | | Abnormal results of function study of kidney | genitourinary | UricA | |
| 590 | | | | | 21698 | 323 | 21375 | 1.292 | 0.07925 | 3.641 | 8.94e-60 | 24682 | 746 | 23936 | 0.03391 | 0.2925 | Pyelonephritis | genitourinary | UricA | |
| 591 | 0.7323 | 0.04648 | 2.08 | 6.54e-56 | 23909 | 2534 | 21375 | 0.7902 | 0.03453 | 2.204 | 6.50e-116 | 30676 | 6740 | 23936 | 0.02853 | 0.1243 | Urinary tract infection | genitourinary | UricA | |
| 592 | | | | | 21768 | 393 | 21375 | 0.7632 | 0.06159 | 2.145 | 2.89e-35 | 25433 | 1497 | 23936 | 0.271 | 0.1333 | Cystitis and urethritis | genitourinary | UricA | |
| 592.1 | | | | | 21707 | 332 | 21375 | 0.7597 | 0.06267 | 2.138 | 8.18e-34 | 25375 | 1439 | 23936 | 0.2167 | 0.1599 | Cystitis | genitourinary | UricA | |

| phenotype | male_beta | male_SE | male_OR | male_p | male_n_total | male_n_cases | male_n_controls | female_beta | female_SE | female_OR | female_p | female_n_total | female_n_cases | female_n_controls | interaction_p | interaction_beta | description | group | lab | label |
|---|---|---|---|---|---|---|---|---|---|---|---|---|---|---|---|---|---|---|---|---|
| 592.11 | | | | | 21543 | 168 | 21375 | 0.8036 | 0.07654 | 2.234 | 8.65e-26 | 24834 | 898 | 23936 | 0.06462 | 0.3256 | Acute cystitis | genitourinary | UricA | |
| 592.12 | | | | | 21411 | 36 | 21375 | | | | | 23989 | 53 | 23936 | | | Chronic cystitis | genitourinary | UricA | |
| 592.13 | | | | | 21383 | 8 | 21375 | | | | | 24169 | 233 | 23936 | | | Chronic interstitial cystitis | genitourinary | UricA | |
| 592.2 | | | | | 21408 | 33 | 21375 | | | | | 23967 | 31 | 23936 | | | Urethritis and urethral syndrome | genitourinary | UricA | |
| 592.21 | | | | | 21378 | 3 | 21375 | | | | | 23938 | 2 | 23936 | | | Urethral syndrome | genitourinary | UricA | |
| 592.3 | | | | | 21378 | 3 | 21375 | | | | | 23938 | 2 | 23936 | | | Urethral stricture due to infecton | genitourinary | UricA | |
| 593 | 0.7083 | 0.05362 | 2.031 | 7.70e-40 | 23190 | 1815 | 21375 | 0.7664 | 0.0568 | 2.152 | 1.75e-41 | 25739 | 1803 | 23936 | 0.1227 | 0.118 | Hematuria | genitourinary | UricA | |
| 593.1 | 0.4566 | 0.08722 | 1.579 | 1.65e-07 | 22020 | 645 | 21375 | | | | | 24248 | 312 | 23936 | 8.31e-05 | 0.5728 | Gross hematuria | genitourinary | UricA | |
| 593.2 | | | | | 21568 | 193 | 21375 | | | | | 24188 | 252 | 23936 | | | Microscopic hematuria | genitourinary | UricA | |
| 594 | 0.9644 | 0.05206 | 2.623 | 1.30e-76 | 25965 | 1823 | 24142 | 1.073 | 0.05409 | 2.924 | 1.54e-87 | 35229 | 1634 | 33595 | 0.3797 | 0.06389 | Urinary calculus | genitourinary | UricA | |
| 594.1 | 1.109 | 0.05765 | 3.03 | 2.08e-82 | 25599 | 1457 | 24142 | 1.171 | 0.05878 | 3.225 | 2.65e-88 | 34942 | 1347 | 33595 | 0.7378 | 0.02664 | Calculus of kidney | genitourinary | UricA | |
| 594.2 | | | | | 24276 | 134 | 24142 | | | | | 33649 | 54 | 33595 | | | Calculus of lower urinary tract | genitourinary | UricA | |
| 594.3 | | | | | 24598 | 456 | 24142 | | | | | 33995 | 400 | 33595 | 0.3436 | -0.1327 | Calculus of ureter | genitourinary | UricA | |
| 594.8 | | | | | 24179 | 37 | 24142 | | | | | 33649 | 54 | 33595 | | | Renal colic | genitourinary | UricA | |
| 595 | 0.7856 | 0.07915 | 2.194 | 3.23e-23 | 24902 | 760 | 24142 | 0.7821 | 0.08234 | 2.186 | 2.15e-21 | 34327 | 732 | 33595 | 0.8949 | -0.01456 | Hydronephrosis | genitourinary | UricA | |
| 596 | 0.1683 | 0.06871 | 1.183 | 0.01431 | 25451 | 1131 | 24320 | 0.2352 | 0.07048 | 1.265 | 8.47e-04 | 34046 | 1171 | 32875 | 0.6483 | 0.04358 | Other disorders of bladder | genitourinary | UricA | |
| 596.1 | | | | | 24500 | 180 | 24320 | | | | | 32903 | 28 | 32875 | | | Bladder neck obstruction | genitourinary | UricA | |
| 596.5 | 0.04687 | 0.09656 | 1.048 | 0.6274 | 24918 | 598 | 24320 | 0.194 | 0.07953 | 1.214 | 0.01472 | 33798 | 923 | 32875 | 0.1517 | 0.1734 | Functional disorders of bladder | genitourinary | UricA | |
| 597 | | | | | 24709 | 389 | 24320 | | | | | 33264 | 389 | 32875 | 0.001793 | 0.4848 | Other disorders of urethra and urinary tract | genitourinary | UricA | |
| 597.1 | | | | | 24499 | 179 | 24320 | | | | | 32898 | 23 | 32875 | | | Urethral stricture (not specified as infectious) | genitourinary | UricA | |
| 597.2 | | | | | 24431 | 111 | 24320 | | | | | 32951 | 76 | 32875 | | | Urinary complications NEC | genitourinary | UricA | |
| 597.8 | | | | | 24446 | 126 | 24320 | | | | | 32996 | 121 | 32875 | | | Urethral hypermobility/ISD | genitourinary | UricA | |
| 598 | | | | | 27202 | 318 | 26884 | | | | | 36132 | 426 | 35706 | 0.06609 | 0.2778 | Abnormal findings on examination of urine | genitourinary | UricA | |
| 598.4 | | | | | 26884 | | | | | | | 35709 | 3 | 35706 | | | Other cells and casts in urine | genitourinary | UricA | |
| 599 | 0.671 | 0.04019 | 1.956 | 1.41e-62 | 23589 | 4209 | 19380 | 0.5825 | 0.03497 | 1.79 | 2.83e-62 | 31705 | 6851 | 24854 | 0.009345 | -0.1362 | Other symptoms/disorders or the urinary system | genitourinary | UricA | |
| 599.1 | | | | | 19615 | 235 | 19380 | | | | | 24927 | 73 | 24854 | | | Urinary obstruction | genitourinary | UricA | |
| 599.2 | 0.5574 | 0.05773 | 1.746 | 4.67e-22 | 21042 | 1662 | 19380 | 0.6253 | 0.07047 | 1.869 | 7.08e-19 | 25979 | 1125 | 24854 | 0.4885 | 0.06185 | Retention of urine | genitourinary | UricA | |
| 599.3 | 0.7878 | 0.08 | 2.199 | 6.98e-23 | 20130 | 750 | 19380 | 0.7373 | 0.04516 | 2.09 | 6.43e-60 | 27990 | 3136 | 24854 | 0.3138 | -0.09044 | Dysuria | genitourinary | UricA | |
| 599.4 | 0.3986 | 0.08367 | 1.49 | 1.89e-06 | 20122 | 742 | 19380 | 0.5511 | 0.05392 | 1.735 | 1.60e-24 | 27034 | 2180 | 24854 | 0.06576 | 0.179 | Urinary incontinence | genitourinary | UricA | |
| 599.5 | 0.7137 | 0.05548 | 2.042 | 7.10e-38 | 21290 | 1910 | 19380 | 0.6698 | 0.05431 | 1.954 | 6.06e-35 | 26913 | 2059 | 24854 | 0.09839 | -0.1262 | Frequency of urination and polyuria | genitourinary | UricA | |
| 599.6 | | | | | 19476 | 96 | 19380 | | | | | 24972 | 118 | 24854 | | | Oliguria and anuria | genitourinary | UricA | |
| 599.7 | | | | | 19390 | 10 | 19380 | | | | | 24855 | 1 | 24854 | | | Urethral discharge | genitourinary | UricA | |
| 599.8 | | | | | 19615 | 235 | 19380 | | | | | 25086 | 232 | 24854 | | | Other symptoms involving urinary system | genitourinary | UricA | |
| 599.9 | 0.8021 | 0.07869 | 2.23 | 2.14e-24 | 20173 | 793 | 19380 | 0.5323 | 0.06841 | 1.703 | 7.20e-15 | 26091 | 1237 | 24854 | 5.10e-04 | -0.3546 | Other abnormality of urination | genitourinary | UricA | |
| 600 | 0.6388 | 0.04535 | 1.894 | 4.74e-45 | 23028 | 3640 | 19388 | | | | | 4 | | | | | Hyperplasia of prostate | genitourinary | UricA | |
| 601 | 0.6524 | 0.08323 | 1.92 | 4.54e-15 | 20083 | 695 | 19388 | | | | | 4 | | | | | Inflammatory diseases of prostate | genitourinary | UricA | |

| phenotype | male_beta | male_SE | male_OR | male_p | male_n_total | male_n_cases | male_n_controls | female_beta | female_SE | female_OR | female_p | female_n_total | female_n_cases | female_n_controls | interaction_p | interaction_beta | description | group | lab | label |
|---|---|---|---|---|---|---|---|---|---|---|---|---|---|---|---|---|---|---|---|---|
| 601.1 | | | | | 19781 | 393 | 19388 | | | | | 4 | | | | | Prostatitis | genitourinary | UricA | |
| 601.11 | | | | | 19525 | 137 | 19388 | | | | | 4 | | | | | Acute prostatitis | genitourinary | UricA | |
| 601.12 | | | | | 19502 | 114 | 19388 | | | | | 4 | | | | | Chronic prostatitis | genitourinary | UricA | |
| 601.3 | | | | | 19577 | 189 | 19388 | | | | | 4 | | | | | Orchitis and epididymitis | genitourinary | UricA | |
| 601.4 | | | | | 19435 | 47 | 19388 | | | | | 4 | | | | | Balanoposthitis | genitourinary | UricA | |
| 601.8 | | | | | 19460 | 72 | 19388 | | | | | 4 | | | | | Other inflammatory disorders of male genital organs | genitourinary | UricA | |
| 602 | | | | | 19490 | 102 | 19388 | | | | | 4 | | | | | Other disorders of prostate | genitourinary | UricA | |
| 602.3 | | | | | 19390 | 2 | 19388 | | | | | 4 | | | | | Dysplasia of prostate | genitourinary | UricA | |
| 603 | | | | | 19102 | 243 | 18859 | | | | | 4 | | | | | Other disorders of testis | genitourinary | UricA | |
| phenotype | male_beta | male_SE | male_OR | male_p | male_n_total | male_n_cases | male_n_controls | female_beta | female_SE | female_OR | female_p | female_n_total | female_n_cases | female_n_controls | interaction_p | interaction_beta | description | group | lab | label |
| 603.1 | | | | | 19051 | 192 | 18859 | | | | | 4 | | | | | Hydrocele | genitourinary | UricA | |
| 603.2 | | | | | 18915 | 56 | 18859 | | | | | 4 | | | | | Spermatocele | genitourinary | UricA | |
| 604 | | | | | 19178 | 319 | 18859 | | | | | 4 | | | | | Disorders of penis | genitourinary | UricA | |
| 604.1 | | | | | 18955 | 96 | 18859 | | | | | 4 | | | | | Redundant prepuce and phimosis/BXO | genitourinary | UricA | |
| 604.2 | | | | | 18859 | | | | | | | 4 | | | | | Vascular disorders of penis | genitourinary | UricA | |
| 604.3 | | | | | 18985 | 126 | 18859 | | | | | 4 | | | | | Peyronie's disease | genitourinary | UricA | |
| 605 | 0.5271 | 0.05321 | 1.694 | 3.89e-23 | 21005 | 2146 | 18859 | | | | | 4 | | | | | Erectile dysfunction [ED] | genitourinary | UricA | |
| 608 | 0.7669 | 0.09471 | 2.153 | 5.63e-16 | 19398 | 539 | 18859 | | | | | 4 | | | | | Other disorders of male genital organs | genitourinary | UricA | |
| 609 | | | | | 18929 | 70 | 18859 | | | | | 4 | | | | | Male infertility and abnormal spermatozoa | genitourinary | UricA | |
| 609.1 | | | | | 18884 | 25 | 18859 | | | | | 4 | | | | | Infertility, male | genitourinary | UricA | |
| 609.11 | | | | | 18861 | 2 | 18859 | | | | | 4 | | | | | Azoospermia and oligospermia | genitourinary | UricA | |
| 609.2 | | | | | 18902 | 43 | 18859 | | | | | 4 | | | | | Abnormal spermatozoa | genitourinary | UricA | |
| 610 | | | | | 5 | | | 0.0422 | 0.06358 | 1.043 | 0.5068 | 29977 | 1611 | 28366 | 0.9984 | 0.5992 | Benign mammary dysplasias | genitourinary | UricA | |
| 610.1 | | | | | 5 | | | 0.02941 | 0.07174 | 1.03 | 0.6819 | 29598 | 1232 | 28366 | 0.9983 | 0.6153 | Cystic mastopathy | genitourinary | UricA | |
| 610.2 | | | | | 5 | | | | | | | 28425 | 59 | 28366 | | | Fibroadenosis of breast | genitourinary | UricA | |
| 610.3 | | | | | 5 | | | | | | | 28501 | 135 | 28366 | | | Fibrosclerosis of breast | genitourinary | UricA | |
| 610.4 | | | | | 5 | | | | | | | 28633 | 267 | 28366 | | | Benign neoplasm of breast | genitourinary | UricA | |
| 610.8 | | | | | 5 | | | | | | | 28599 | 233 | 28366 | | | Other specified benign mammary dysplasias | genitourinary | UricA | |
| 611 | | | | | 27275 | 62 | 27213 | 0.08354 | 0.03943 | 1.087 | 0.0341 | 33455 | 5088 | 28367 | 0.5006 | -0.1768 | Abnormal findings on mammogram or breast exam | genitourinary | UricA | |
| 611.1 | | | | | 27225 | 12 | 27213 | 0.04684 | 0.04281 | 1.048 | 0.274 | 32548 | 4181 | 28367 | 0.1256 | -0.8986 | Abnormal mammogram | genitourinary | UricA | |
| 611.11 | | | | | 27213 | | | 0.2118 | 0.0984 | 1.236 | 0.0314 | 28926 | 559 | 28367 | 0.9988 | 0.9712 | Mammographic microcalcification | genitourinary | UricA | |
| 611.3 | | | | | 27266 | 53 | 27213 | 0.2031 | 0.05998 | 1.225 | 7.09e-04 | 30081 | 1714 | 28367 | 0.9818 | 0.006629 | Lump or mass in breast | genitourinary | UricA | |
| 612 | | | | | 27326 | 113 | 27213 | | | | | 28757 | 390 | 28367 | 0.003287 | -0.663 | Breast conditions, congenital or relating to hormones | genitourinary | UricA | |
| 612.1 | | | | | 27213 | | | | | | | 28446 | 79 | 28367 | | | Galactorrhea | genitourinary | UricA | |
| 612.2 | | | | | 27325 | 112 | 27213 | | | | | 28664 | 297 | 28367 | | | Hypertrophy of breast (Gynecomastia) | genitourinary | UricA | |
| 612.3 | | | | | 27214 | 1 | 27213 | | | | | 28379 | 12 | 28367 | | | Congenital anomalies of breast | genitourinary | UricA | |
| 613 | | | | | 27667 | 60 | 27607 | 0.1059 | 0.05113 | 1.112 | 0.03842 | 34784 | 2435 | 32349 | 0.07858 | -0.4648 | Other nonmalignant breast conditions | genitourinary | UricA | |
| 613.1 | | | | | 27607 | | | | | | | 32616 | 267 | 32349 | | | Inflammatory disease of breast | genitourinary | UricA | |
| 613.5 | | | | | 27649 | 42 | 27607 | 0.3231 | 0.07387 | 1.381 | 1.22e-05 | 33336 | 987 | 32349 | 0.3181 | -0.3174 | Mastodynia | genitourinary | UricA | |
| 613.7 | | | | | 27616 | 9 | 27607 | | | | | 32785 | 436 | 32349 | | | Other signs and symptoms in breast | genitourinary | UricA | |
| 613.8 | | | | | 27608 | 1 | 27607 | -0.1324 | 0.09468 | 0.876 | 0.1619 | 33034 | 685 | 32349 | 0.9479 | -12.16 | Other specified disorders of breast | genitourinary | UricA | |

| phenotype | male_beta | male_SE | male_OR | male_p | male_n_total | male_n_cases | male_n_controls | female_beta | female_SE | female_OR | female_p | female_n_total | female_n_cases | female_n_controls | interaction_p | interaction_beta | description | group | lab | label |
|---|---|---|---|---|---|---|---|---|---|---|---|---|---|---|---|---|---|---|---|---|
| 613.9 | | | | | 27609 | 2 | 27607 | | | | | 32422 | 73 | 32349 | | | Breast disorder NOS | genitourinary | UricA | |
| 614 | | | | | 7 | | | 0.5657 | 0.05928 | 1.761 | 1.39e-21 | 33657 | 1625 | 32032 | 0.9989 | 0.3808 | Inflammatory diseases of female pelvic organs | genitourinary | UricA | |
| 614.1 | | | | | 7 | | | | | | | 32176 | 144 | 32032 | | | Pelvic peritoneal adhesions, female (postoperative) (postinfection) | genitourinary | UricA | |
| 614.3 | | | | | 7 | | | | | | | 32232 | 200 | 32032 | | | Pelvic inflammatory disease (PID) | genitourinary | UricA | |
| 614.31 | | | | | 7 | | | | | | | 32055 | 23 | 32032 | | | Acute inflammatory pelvic disease | genitourinary | UricA | |
| 614.32 | | | | | 7 | | | | | | | 32140 | 108 | 32032 | | | Chronic inflammatory pelvic disease | genitourinary | UricA | |
| 614.33 | | | | | 7 | | | | | | | 32100 | 68 | 32032 | | | Pelvic inflammatory disease, NOS | genitourinary | UricA | |
| 614.4 | | | | | 7 | | | | | | | 32069 | 37 | 32032 | | | Inflammatory diseases of uterus, except cervix | genitourinary | UricA | |
| 614.5 | | | | | 7 | | | 0.5177 | 0.06881 | 1.678 | 5.28e-14 | 33209 | 1177 | 32032 | 0.9989 | 0.3885 | Inflammatory disease of cervix, vagina, and vulva | genitourinary | UricA | |
| 614.51 | | | | | 7 | | | | | | | 32101 | 69 | 32032 | | | Cervicitis and endocervicitis | genitourinary | UricA | |
| 614.52 | | | | | 7 | | | 0.483 | 0.08101 | 1.621 | 2.48e-09 | 32877 | 845 | 32032 | 0.9994 | 0.3474 | Vaginitis and vulvovaginitis | genitourinary | UricA | |
| 614.53 | | | | | 7 | | | | | | | 32093 | 61 | 32032 | | | Cyst or abscess of Bartholin's gland | genitourinary | UricA | |
| 614.54 | | | | | 7 | | | | | | | 32097 | 65 | 32032 | | | Abscess or ulceration of vulva | genitourinary | UricA | |
| 615 | | | | | 7 | | | | | | | 32416 | 384 | 32032 | | | Endometriosis | genitourinary | UricA | |
| 618 | | | | | 7 | | | -0.07262 | 0.07348 | 0.93 | 0.323 | 36315 | 1140 | 35175 | 0.9985 | 0.5114 | Genital prolapse | genitourinary | UricA | |
| 618.1 | | | | | 7 | | | -0.0387 | 0.08153 | 0.962 | 0.635 | 36071 | 896 | 35175 | 0.999 | 0.5611 | Prolapse of vaginal walls | genitourinary | UricA | |
| 618.2 | | | | | 7 | | | | | | | 35515 | 340 | 35175 | | | Uterine/Uterovaginal prolapse | genitourinary | UricA | |
| 618.5 | | | | | 7 | | | | | | | 35494 | 319 | 35175 | | | Prolapse of vaginal vault after hysterectomy | genitourinary | UricA | |
| 618.6 | | | | | 7 | | | | | | | 35376 | 201 | 35175 | | | Vaginal enterocele, congenital or acquired | genitourinary | UricA | |
| 619 | | | | | 7 | | | 0.545 | 0.05268 | 1.725 | 4.40e-25 | 33857 | 2126 | 31731 | 0.9988 | 0.4237 | Noninflammatory female genital disorders | genitourinary | UricA | |
| 619.1 | | | | | 7 | | | | | | | 32002 | 271 | 31731 | | | Noninflammatory disorders of ovary, fallopian tube, and broad ligament | genitourinary | UricA | |
| 619.2 | | | | | 7 | | | | | | | 32192 | 461 | 31731 | | | Disorders of uterus, NEC | genitourinary | UricA | |
| 619.3 | | | | | 7 | | | | | | | 31864 | 133 | 31731 | | | Noninflammatory disorders of cervix | genitourinary | UricA | |
| 619.4 | | | | | 7 | | | 0.4005 | 0.1042 | 1.493 | 1.22e-04 | 32259 | 528 | 31731 | 0.9994 | 0.3274 | Noninflammatory disorders of vagina | genitourinary | UricA | |
| 619.5 | | | | | 7 | | | | | | | 31969 | 238 | 31731 | | | Noninflammatory disorders of vulva and perineum | genitourinary | UricA | |
| 620 | | | | | 7 | | | | | | | 35251 | 113 | 35138 | | | Dysplasia of female genital organs | genitourinary | UricA | |
| 620.1 | | | | | 7 | | | | | | | 35152 | 14 | 35138 | | | Dysplasia of cervix | genitourinary | UricA | |
| 621 | | | | | 7 | | | | | | | 35309 | 171 | 35138 | | | Endometrial hyperplasia | genitourinary | UricA | |
| 622 | | | | | 7 | | | | | | | 35534 | 396 | 35138 | | | Polyp of female genital organs | genitourinary | UricA | |
| phenotype | male_beta | male_SE | male_OR | male_p | male_n_total | male_n_cases | male_n_controls | female_beta | female_SE | female_OR | female_p | female_n_total | female_n_cases | female_n_controls | interaction_p | interaction_beta | description | group | lab | label |
| 622.1 | | | | | 7 | | | | | | | 35431 | 293 | 35138 | | | Polyp of corpus uteri | genitourinary | UricA | |
| 622.2 | | | | | 7 | | | | | | | 35215 | 77 | 35138 | | | Mucous polyp of cervix | genitourinary | UricA | |
| 623 | | | | | 7 | | | | | | | 35299 | 161 | 35138 | | | Hypertrophy of female genital organs | genitourinary | UricA | |
| 624 | | | | | 7 | | | | | | | 31709 | 227 | 31482 | | | Symptoms involving female genital tract | genitourinary | UricA | |
| 624.1 | | | | | 7 | | | | | | | 31539 | 57 | 31482 | | | Dystrophy of female genital tract | genitourinary | UricA | |
| 624.2 | | | | | 7 | | | | | | | 31503 | 21 | 31482 | | | Atrophy of female genital tract | genitourinary | UricA | |
| 624.9 | | | | | 7 | | | 0.2258 | 0.07529 | 1.253 | 0.002704 | 32466 | 984 | 31482 | 0.9988 | 0.6669 | stress incontinence, female | genitourinary | UricA | |

| phenotype | male_beta | male_SE | male_OR | male_p | male_n_total | male_n_cases | male_n_controls | female_beta | female_SE | female_OR | female_p | female_n_total | female_n_cases | female_n_controls | interaction_p | interaction_beta | description | group | lab | label |
|---|---|---|---|---|---|---|---|---|---|---|---|---|---|---|---|---|---|---|---|---|
| 625 | | | | | 7 | | | 0.4098 | 0.06488 | 1.507 | 2.66e-10 | 32899 | 1417 | 31482 | 0.9992 | 0.2777 | Pain and other symptoms associated with female genital organs | genitourinary | UricA | |
| 625.1 | | | | | 7 | | | | | | | 31749 | 267 | 31482 | | | Dyspareunia | genitourinary | UricA | |
| 626 | | | | | 5 | | | 0.41 | 0.04749 | 1.507 | 5.93e-18 | 27458 | 3874 | 23584 | 0.9998 | 0.03863 | Disorders of menstruation and other abnormal bleeding from female genital tract | genitourinary | UricA | |
| 626.1 | | | | | 5 | | | 0.4087 | 0.0515 | 1.505 | 2.08e-15 | 26685 | 3101 | 23584 | 0.9998 | 0.05978 | Irregular menstrual cycle/bleeding | genitourinary | UricA | |
| 626.11 | | | | | 5 | | | 0.6797 | 0.1027 | 1.973 | 3.65e-11 | 24169 | 585 | 23584 | 0.9999 | 0.08269 | Absent or infrequent menstruation | genitourinary | UricA | |
| 626.12 | | | | | 5 | | | 0.4602 | 0.07522 | 1.584 | 9.47e-10 | 24734 | 1150 | 23584 | 0.9992 | 0.2964 | Excessive or frequent menstruation | genitourinary | UricA | |
| 626.13 | | | | | 5 | | | 0.3805 | 0.07925 | 1.463 | 1.57e-06 | 24711 | 1127 | 23584 | 0.9998 | 0.07392 | Irregular menstrual cycle | genitourinary | UricA | |
| 626.14 | | | | | 5 | | | | | | | 23850 | 266 | 23584 | | | Irregular menstrual bleeding | genitourinary | UricA | |
| 626.15 | | | | | 5 | | | | | | | 23629 | 45 | 23584 | | | Infertility, female, associated with anovulation | genitourinary | UricA | |
| 626.2 | | | | | 5 | | | | | | | 24029 | 445 | 23584 | | | Dysmenorrhea | genitourinary | UricA | |
| 626.21 | | | | | 5 | | | | | | | 23591 | 7 | 23584 | | | Mittelschmerz | genitourinary | UricA | |
| 626.4 | | | | | 5 | | | | | | | 23636 | 52 | 23584 | | | Premenstrual tension syndromes | genitourinary | UricA | |
| 626.8 | | | | | 5 | | | | | | | 23771 | 187 | 23584 | | | Infertility, female | genitourinary | UricA | |
| 627 | | | | | 6 | | | 0.1854 | 0.041 | 1.204 | 6.14e-06 | 28947 | 5363 | 23584 | 0.9012 | 10.47 | Menopausal and postmenopausal disorders | genitourinary | UricA | |
| 627.1 | | | | | 5 | | | 0.1027 | 0.08846 | 1.108 | 0.2458 | 24382 | 798 | 23584 | 0.9982 | 1.073 | Postmenopausal bleeding | genitourinary | UricA | |
| 627.2 | | | | | 6 | | | 0.1283 | 0.04875 | 1.137 | 0.008493 | 26962 | 3378 | 23584 | 0.9347 | 11.39 | Symptomatic menopause | genitourinary | UricA | |
| 627.21 | | | | | 5 | | | | | | | 23934 | 350 | 23584 | | | Symptomatic artificial menopause | genitourinary | UricA | |
| 627.22 | | | | | 6 | | | 0.3706 | 0.08023 | 1.449 | 3.86e-06 | 24465 | 881 | 23584 | 0.956 | 12.65 | Need for Hormone replacement therapy (postmenopausal) | genitourinary | UricA | |
| 627.3 | | | | | 5 | | | 0.3267 | 0.06725 | 1.386 | 1.18e-06 | 24943 | 1359 | 23584 | 0.9958 | 1.568 | Postmenopausal atrophic vaginitis | genitourinary | UricA | |
| 627.4 | | | | | 5 | | | | | | | 23687 | 103 | 23584 | | | Premenopausal menorrhagia | genitourinary | UricA | |
| 627.5 | | | | | 5 | | | | | | | 23742 | 158 | 23584 | | | Premature menopause and other ovarian failure | genitourinary | UricA | |
| 628 | | | | | 5 | | | 0.5547 | 0.07766 | 1.741 | 9.14e-13 | 24586 | 1002 | 23584 | 0.9989 | 0.4114 | Ovarian cyst | genitourinary | UricA | |
| 634 | | | | | 7 | | | 1.681 | 0.1027 | 5.372 | 3.41e-60 | 34235 | 519 | 33716 | 0.9988 | 0.6881 | Miscarriage; stillbirth | pregnancy complications | UricA | |
| 634.1 | | | | | 7 | | | | | | | 33945 | 229 | 33716 | | | Missed abortion/Hydatidiform mole | pregnancy complications | UricA | |
| 634.3 | | | | | 7 | | | | | | | 33757 | 41 | 33716 | | | Ectopic pregnancy | pregnancy complications | UricA | |
| 635 | | | | | 7 | | | | | | | 34113 | 397 | 33716 | | | Hemorrhage during pregnancy; childbirth and postpartum | pregnancy complications | UricA | |
| 635.2 | | | | | 7 | | | | | | | 33989 | 273 | 33716 | | | Antepartum hemorrhage, abruptio placentae, and placenta previa | pregnancy complications | UricA | |
| 635.3 | | | | | 7 | | | | | | | 33851 | 135 | 33716 | | | Placenta previa and abruptio placenta | pregnancy complications | UricA | |
| 636 | | | | | 7 | | | 1.8 | 0.06183 | 6.047 | 3.04e-186 | 35417 | 1701 | 33716 | 0.9981 | 0.6347 | Early or threatened labor; hemorrhage in early pregnancy | pregnancy complications | UricA | |
| 636.1 | | | | | 7 | | | | | | | 34209 | 493 | 33716 | | | Threatened premature labor | pregnancy complications | UricA | |
| 636.2 | | | | | 7 | | | | | | | 34111 | 395 | 33716 | | | Early onset of delivery | pregnancy complications | UricA | |
| 636.3 | | | | | 7 | | | 1.625 | 0.1037 | 5.081 | 2.49e-55 | 34236 | 520 | 33716 | 0.9989 | 0.6327 | Hemorrhage in early pregnancy | pregnancy complications | UricA | |

| phenotype | male_beta | male_SE | male_OR | male_p | male_n_total | male_n_cases | male_n_controls | female_beta | female_SE | female_OR | female_p | female_n_total | female_n_cases | female_n_controls | interaction_p | interaction_beta | description | group | lab | label |
|---|---|---|---|---|---|---|---|---|---|---|---|---|---|---|---|---|---|---|---|---|
| 636.8 | | | | | 7 | | | | | | | 33814 | 98 | 33716 | | | Cervical incompetence | pregnancy complications | UricA | |
| 637 | | | | | 27759 | 8 | 27751 | | | | | 33709 | 19 | 33690 | | | Short gestation; low birth weight; and fetal growth retardation | pregnancy complications | UricA | |
| 642 | | | | | 7 | | | 4.755 | 0.1271 | 116.21 | 2.15e-306 | 36632 | 759 | 35873 | 0.9943 | 3.151 | Hypertension complicating pregnancy, childbirth, and the puerperium | pregnancy complications | UricA | |
| 642.1 | | | | | 7 | | | | | | | 36229 | 356 | 35873 | | | Preeclampsia and eclampsia | pregnancy complications | UricA | |
| 643 | | | | | 7 | | | | | | | 36714 | 156 | 36558 | | | Excessive vomiting in pregnancy | pregnancy complications | UricA | |
| 643.1 | | | | | 7 | | | | | | | 36608 | 50 | 36558 | | | Hyperemesis gravidarum | pregnancy complications | UricA | |
| 644 | | | | | 7 | | | | | | | 36546 | 186 | 36360 | | | Anemia during pregnancy | pregnancy complications | UricA | |
| 646 | | | | | 7 | | | 1.946 | 0.09499 | 7.003 | 2.66e-93 | 36367 | 578 | 35789 | 0.9988 | 0.6403 | Other complications of pregnancy NEC | pregnancy complications | UricA | |
| 647 | | | | | 7 | | | | | | | 36080 | 179 | 35901 | | | Infectious and parasitic complications affecting pregnancy | pregnancy complications | UricA | |
| 647.1 | | | | | 7 | | | | | | | 36102 | 201 | 35901 | | | Infections of genitourinary tract during pregnancy | pregnancy complications | UricA | |
| 647.3 | | | | | 7 | | | | | | | 35950 | 49 | 35901 | | | Major puerperal infection | pregnancy complications | UricA | |
| 649 | | | | | 7 | | | 2.152 | 0.0633 | 8.605 | 1.88e-253 | 35652 | 1664 | 33988 | 0.9974 | 0.8713 | Other conditions or status of the mother complicating pregnancy, childbirth, or the puerperium | pregnancy complications | UricA | |
| 649.1 | | | | | 7 | | | 2.498 | 0.0955 | 12.16 | 7.64e-151 | 34593 | 605 | 33988 | 0.9976 | 1.359 | Diabetes or abnormal glucose tolerance complicating pregnancy | pregnancy complications | UricA | |
| 653 | | | | | 7 | | | 1.801 | 0.08957 | 6.054 | 6.86e-90 | 35753 | 642 | 35111 | 0.9989 | 0.6284 | Problems associated with amniotic cavity and membranes | pregnancy complications | UricA | |
| 654 | | | | | 7 | | | | | | | 35408 | 297 | 35111 | | | Other and unspecified complications of birth; puerperium affecting management of mother | pregnancy complications | UricA | |
| 654.1 | | | | | 7 | | | | | | | 35312 | 201 | 35111 | | | Abnormality of organs and soft tissues of pelvis complicating pregnancy, childbirth, or the puerperium | pregnancy complications | UricA | |
| 654.2 | | | | | 7 | | | | | | | 35201 | 90 | 35111 | | | Rhesus isoimmunization in pregnancy | pregnancy complications | UricA | |
| 655 | | | | | 7 | | | 1.808 | 0.05586 | 6.1 | 6.76e-230 | 36061 | 2235 | 33826 | 0.9982 | 0.601 | Known or suspected fetal abnormality affecting management of mother | pregnancy complications | UricA | |
| 655.1 | | | | | 7 | | | 2.06 | 0.08696 | 7.846 | 4.78e-124 | 34615 | 789 | 33826 | 0.9986 | 0.78 | Abnormality in fetal heart rate or rhythm | pregnancy complications | UricA | |
| 656 | | | | | 27627 | 68 | 27559 | | | | | 36677 | 120 | 36557 | | | Other perinatal conditions of fetus or newborn | pregnancy complications | UricA | |
| 656.1 | | | | | 27562 | 3 | 27559 | | | | | 36558 | 1 | 36557 | | | Isoimmunization of fetus or newborn | pregnancy complications | UricA | |
| phenotype | male_beta | male_SE | male_OR | male_p | male_n_total | male_n_cases | male_n_controls | female_beta | female_SE | female_OR | female_p | female_n_total | female_n_cases | female_n_controls | interaction_p | interaction_beta | description | group | lab | label |
| 656.2 | | | | | 27597 | 38 | 27559 | | | | | 36585 | 28 | 36557 | | | Respiratory conditions of fetus and newborn | pregnancy complications | UricA | |
| 656.22 | | | | | 27562 | 3 | 27559 | | | | | 36559 | 2 | 36557 | | | Interstitial emphysema and related conditions of newborn | pregnancy complications | UricA | |
| 656.26 | | | | | 27567 | 8 | 27559 | | | | | 36561 | 4 | 36557 | | | Transitory tachypnea or apnea of newborn | pregnancy complications | UricA | |
| 656.3 | | | | | 27567 | 8 | 27559 | | | | | 36567 | 10 | 36557 | | | Endocrine and metabolic disturbances of fetus and newborn | pregnancy complications | UricA | |

| phenotype | male_beta | male_SE | male_OR | male_p | male_n_total | male_n_cases | male_n_controls | female_beta | female_SE | female_OR | female_p | female_n_total | female_n_cases | female_n_controls | interaction_p | interaction_beta | description | group | lab | label |
|---|---|---|---|---|---|---|---|---|---|---|---|---|---|---|---|---|---|---|---|---|
| 656.4 | | | | | 27568 | 9 | 27559 | | | | | 36563 | 6 | 36557 | | | Hemorrhage of fetus or newborn | pregnancy complications | UricA | |
| 656.5 | | | | | 27565 | 6 | 27559 | | | | | 36562 | 5 | 36557 | | | Hematological disorders of newborn | pregnancy complications | UricA | |
| 656.6 | | | | | 27567 | 8 | 27559 | | | | | 36562 | 5 | 36557 | | | Perinatal disorders of digestive system | pregnancy complications | UricA | |
| 656.7 | | | | | 27566 | 7 | 27559 | | | | | 36566 | 9 | 36557 | | | Conditions involving the integument and temperature regulation of fetus and newborn | pregnancy complications | UricA | |
| 656.8 | | | | | 27575 | 16 | 27559 | | | | | 36572 | 15 | 36557 | | | Perinatal jaundice | pregnancy complications | UricA | |
| 656.9 | | | | | 27559 | | | | | | | 36559 | 2 | 36557 | | | Neonatal bradycardia or tachycardia | pregnancy complications | UricA | |
| 657 | | | | | 27745 | 7 | 27738 | | | | | 36919 | 6 | 36913 | | | Infections specific to the perinatal period | pregnancy complications | UricA | |
| 669 | | | | | 7 | | | | | | | 36606 | 168 | 36438 | | | Complications of labor and delivery NEC | pregnancy complications | UricA | |
| 671 | | | | | 7 | | | | | | | 36547 | 117 | 36430 | | | Venous/cerebrovascular complications embolism in pregnancy and the puerperium | pregnancy complications | UricA | |
| 674 | | | | | 7 | | | | | | | 36509 | 79 | 36430 | | | Other complications of the puerperium NEC | pregnancy complications | UricA | |
| 676 | | | | | 7 | | | | | | | 36440 | 10 | 36430 | | | Other disorders of the breast associated with childbirth and disorders of lactation | pregnancy complications | UricA | |
| 681 | 0.7381 | 0.04487 | 2.092 | 8.39e-61 | 24732 | 2737 | 21995 | 0.7388 | 0.04311 | 2.093 | 7.69e-66 | 32934 | 3053 | 29881 | 0.3472 | 0.05684 | Superficial cellulitis and abscess | dermatologic | UricA | |
| 681.1 | | | | | 22394 | 399 | 21995 | | | | | 30304 | 423 | 29881 | 0.393 | -0.1249 | Cellulitis and abscess of fingers/toes | dermatologic | UricA | |
| 681.2 | | | | | 22204 | 209 | 21995 | | | | | 30123 | 242 | 29881 | | | Cellulitis and abscess of face/neck | dermatologic | UricA | |
| 681.3 | | | | | 22375 | 380 | 21995 | | | | | 30260 | 379 | 29881 | 0.8113 | 0.03584 | Cellulitis and abscess of arm/hand | dermatologic | UricA | |
| 681.5 | 0.9118 | 0.06953 | 2.489 | 2.75e-39 | 22978 | 983 | 21995 | 0.9842 | 0.07264 | 2.676 | 7.99e-42 | 30774 | 893 | 29881 | 0.1407 | 0.1433 | Cellulitis and abscess of leg, except foot | dermatologic | UricA | |
| 681.6 | | | | | 22233 | 238 | 21995 | | | | | 30029 | 148 | 29881 | | | Cellulitis and abscess of foot, toe | dermatologic | UricA | |
| 681.7 | | | | | 22491 | 496 | 21995 | 0.8206 | 0.08925 | 2.272 | 3.75e-20 | 30505 | 624 | 29881 | 0.1876 | 0.1714 | Cellulitis and abscess of trunk | dermatologic | UricA | |
| 686 | 0.9318 | 0.08772 | 2.539 | 2.37e-26 | 22609 | 614 | 21995 | 0.6865 | 0.08751 | 1.987 | 4.33e-15 | 30530 | 649 | 29881 | 0.3223 | -0.1183 | Other local infections of skin and subcutaneous tissue | dermatologic | UricA | |
| 686.1 | | | | | 22123 | 128 | 21995 | | | | | 30019 | 138 | 29881 | | | Carbuncle and furuncle | dermatologic | UricA | |
| 686.2 | | | | | 22049 | 54 | 21995 | | | | | 29945 | 64 | 29881 | | | Impetigo | dermatologic | UricA | |
| 686.3 | | | | | 22049 | 54 | 21995 | | | | | 29925 | 44 | 29881 | | | Pilonidal cyst | dermatologic | UricA | |
| 686.4 | | | | | 22031 | 36 | 21995 | | | | | 29904 | 23 | 29881 | | | Pyogenic granuloma | dermatologic | UricA | |
| 686.5 | | | | | 22021 | 26 | 21995 | | | | | 29916 | 35 | 29881 | | | Pyoderma | dermatologic | UricA | |
| 687 | | | | | 21481 | 302 | 21179 | 0.5198 | 0.09571 | 1.682 | 5.60e-08 | 26852 | 564 | 26288 | 0.007512 | -0.4016 | Symptoms affecting skin | dermatologic | UricA | |
| 687.1 | 0.9162 | 0.05514 | 2.5 | 5.39e-62 | 22861 | 1682 | 21179 | 0.6566 | 0.04614 | 1.928 | 6.02e-46 | 29124 | 2836 | 26288 | 0.05348 | -0.1351 | Rash and other nonspecific skin eruption | dermatologic | UricA | |
| 687.3 | | | | | 21208 | 29 | 21179 | | | | | 26359 | 71 | 26288 | | | Changes in skin texture | dermatologic | UricA | |
| 687.4 | 0.463 | 0.06237 | 1.589 | 1.13e-13 | 22559 | 1380 | 21179 | 0.4103 | 0.0506 | 1.507 | 5.08e-16 | 28776 | 2488 | 26288 | 0.8585 | -0.01392 | Disturbance of skin sensation | dermatologic | UricA | |
| 689 | 0.4745 | 0.07592 | 1.607 | 4.09e-10 | 26203 | 833 | 25370 | 0.3752 | 0.07047 | 1.455 | 1.01e-07 | 34664 | 1062 | 33602 | 0.4441 | -0.07663 | Disorder of skin and subcutaneous tissue NOS | dermatologic | UricA | |
| 690 | | | | | 23839 | 493 | 23346 | | | | | 29843 | 385 | 29458 | 0.4428 | -0.1125 | Erythematosquamous dermatosis | dermatologic | UricA | |
| 690.1 | | | | | 23835 | 489 | 23346 | | | | | 29839 | 381 | 29458 | 0.3176 | -0.1474 | Seborrheic dermatitis | dermatologic | UricA | |
| 691 | | | | | 23418 | 72 | 23346 | | | | | 29595 | 137 | 29458 | | | Congenital anomalies of skin | dermatologic | UricA | |
| 691.1 | | | | | 23351 | 5 | 23346 | | | | | 29460 | 2 | 29458 | | | Ichthyosis congenita | dermatologic | UricA | |

| phenotype | male_beta | male_SE | male_OR | male_p | male_n_total | male_n_cases | male_n_controls | female_beta | female_SE | female_OR | female_p | female_n_total | female_n_cases | female_n_controls | interaction_p | interaction_beta | description | group | lab | label |
|---|---|---|---|---|---|---|---|---|---|---|---|---|---|---|---|---|---|---|---|---|
| 691.3 | | | | | 23350 | 4 | 23346 | | | | | 29482 | 24 | 29458 | | | Congenital pigmentary anomalies of skin | dermatologic | UricA | |
| 694 | 0.4872 | 0.07656 | 1.628 | 1.97e-10 | 24181 | 835 | 23346 | 0.1729 | 0.06411 | 1.189 | 0.006988 | 30929 | 1471 | 29458 | 3.45e-04 | -0.3476 | Dyschromia and Vitiligo | dermatologic | UricA | |
| 694.1 | | | | | 23374 | 28 | 23346 | | | | | 29501 | 43 | 29458 | | | Vitiligo | dermatologic | UricA | |
| 694.2 | 0.5045 | 0.07793 | 1.656 | 9.52e-11 | 24148 | 802 | 23346 | 0.1508 | 0.0654 | 1.163 | 0.02116 | 30877 | 1419 | 29458 | 6.95e-05 | -0.3935 | Other dyschromia | dermatologic | UricA | |
| 694.3 | | | | | 23359 | 13 | 23346 | | | | | 29477 | 19 | 29458 | | | Vascular disorders of skin | dermatologic | UricA | |
| 695 | 0.6547 | 0.07572 | 1.925 | 5.29e-18 | 24180 | 834 | 23346 | 0.5154 | 0.05004 | 1.674 | 7.08e-25 | 31756 | 2298 | 29458 | 0.01646 | -0.2106 | Erythematous conditions | dermatologic | UricA | |
| 695.1 | | | | | 23358 | 12 | 23346 | | | | | 29480 | 22 | 29458 | | | Toxic erythema | dermatologic | UricA | |
| 695.2 | | | | | 23377 | 31 | 23346 | | | | | 29513 | 55 | 29458 | | | Bullous dermatoses | dermatologic | UricA | |
| 695.21 | | | | | 23356 | 10 | 23346 | | | | | 29480 | 22 | 29458 | | | Dermatitis herpetiformis | dermatologic | UricA | |
| 695.22 | | | | | 23363 | 17 | 23346 | | | | | 29486 | 28 | 29458 | | | Pemphigus and pemphigoid | dermatologic | UricA | |
| 695.3 | | | | | 23613 | 267 | 23346 | 0.1956 | 0.0918 | 1.216 | 0.03309 | 30121 | 663 | 29458 | 0.3683 | -0.1407 | Rosacea | dermatologic | UricA | |
| 695.4 | | | | | 23118 | 140 | 22978 | 0.5094 | 0.07673 | 1.664 | 3.15e-11 | 29377 | 925 | 28452 | 0.8037 | -0.04806 | Lupus (localized and systemic) | dermatologic | UricA | |
| 695.41 | | | | | 23008 | 30 | 22978 | | | | | 28621 | 169 | 28452 | | | Cutaneous lupus erythematosus | dermatologic | UricA | |
| 695.42 | | | | | 23104 | 126 | 22978 | 0.5032 | 0.07919 | 1.654 | 2.09e-10 | 29322 | 870 | 28452 | 0.7616 | -0.06165 | Systemic lupus erythematosus | dermatologic | UricA | |
| 695.7 | | | | | 23445 | 99 | 23346 | | | | | 29666 | 208 | 29458 | | | Prurigo and Lichen | dermatologic | UricA | |
| 695.8 | | | | | 23469 | 123 | 23346 | | | | | 29759 | 301 | 29458 | | | Other specified erythematous conditions | dermatologic | UricA | |
| 695.81 | | | | | 23357 | 11 | 23346 | | | | | 29501 | 43 | 29458 | | | Erythema nodosum | dermatologic | UricA | |
| 695.9 | | | | | 23478 | 132 | 23346 | | | | | 29596 | 138 | 29458 | | | Unspecified erythematous condition | dermatologic | UricA | |
| 696 | | | | | 23091 | 490 | 22601 | 0.5236 | 0.08728 | 1.688 | 1.99e-09 | 28469 | 689 | 27780 | 0.7369 | -0.04295 | Psoriasis and related disorders | dermatologic | UricA | |
| 696.2 | | | | | 23352 | 6 | 23346 | | | | | 29465 | 7 | 29458 | | | Parapsoriasis | dermatologic | UricA | |
| 696.3 | | | | | 23354 | 8 | 23346 | | | | | 29466 | 8 | 29458 | | | Pityriasis | dermatologic | UricA | |
| 696.4 | | | | | 23086 | 485 | 22601 | 0.5193 | 0.08837 | 1.681 | 4.20e-09 | 28453 | 673 | 27780 | 0.6976 | -0.05008 | Psoriasis | dermatologic | UricA | |
| 696.41 | | | | | 23001 | 400 | 22601 | 0.5268 | 0.09792 | 1.693 | 7.47e-08 | 28318 | 538 | 27780 | 0.5775 | -0.07887 | Psoriasis vulgaris | dermatologic | UricA | |
| 696.42 | | | | | 22782 | 181 | 22601 | | | | | 28051 | 271 | 27780 | | | Psoriatic arthropathy | dermatologic | UricA | |
| 697 | | | | | 23461 | 115 | 23346 | | | | | 29654 | 196 | 29458 | | | Sarcoidosis | dermatologic | UricA | |
| 698 | | | | | 27077 | 348 | 26729 | 0.5856 | 0.0797 | 1.796 | 2.02e-13 | 35552 | 771 | 34781 | 0.4265 | -0.1063 | Pruritus and related conditions | dermatologic | UricA | |
| 700 | | | | | 24764 | 207 | 24557 | | | | | 32043 | 272 | 31771 | | | Corns and callosities | dermatologic | UricA | |
| 701 | 0.5079 | 0.06514 | 1.662 | 6.32e-15 | 25713 | 1156 | 24557 | 0.2425 | 0.05326 | 1.274 | 5.31e-06 | 33879 | 2108 | 31771 | 0.001075 | -0.2669 | Other hypertrophic and atrophic conditions of skin | dermatologic | UricA | |
| 701.1 | | | | | 24638 | 81 | 24557 | | | | | 31856 | 85 | 31771 | | | Keratoderma, acquired | dermatologic | UricA | |
| 701.2 | | | | | 24996 | 439 | 24557 | 0.04252 | 0.08311 | 1.043 | 0.6089 | 32665 | 894 | 31771 | 0.207 | -0.1655 | Scar conditions and fibrosis of skin | dermatologic | UricA | |
| 701.3 | | | | | 24568 | 11 | 24557 | | | | | 31968 | 197 | 31771 | | | Circumscribed scleroderma | dermatologic | UricA | |
| 701.4 | | | | | 24632 | 75 | 24557 | | | | | 31928 | 157 | 31771 | | | Keloid scar | dermatologic | UricA | |
| 701.5 | | | | | 24606 | 49 | 24557 | | | | | 31836 | 65 | 31771 | | | Abnormal granulation tissue | dermatologic | UricA | |
| 701.6 | | | | | 24575 | 18 | 24557 | | | | | 31812 | 41 | 31771 | | | Acquired acanthosis nigricans | dermatologic | UricA | |
| 702 | 0.5707 | 0.0464 | 1.769 | 9.13e-35 | 24883 | 2887 | 21996 | 0.3034 | 0.04721 | 1.354 | 1.32e-10 | 32069 | 3086 | 28983 | 1.21e-04 | -0.2504 | Degenerative skin conditions and other dermatoses | dermatologic | UricA | |
| 702.1 | 0.5742 | 0.05199 | 1.776 | 2.34e-28 | 25602 | 2131 | 23471 | 0.2388 | 0.06293 | 1.27 | 1.48e-04 | 33264 | 1502 | 31762 | 4.00e-05 | -0.3285 | Actinic keratosis | dermatologic | UricA | |
| 702.2 | 0.5172 | 0.05239 | 1.677 | 5.48e-23 | 25554 | 2083 | 23471 | 0.2551 | 0.05013 | 1.291 | 3.59e-07 | 34321 | 2559 | 31762 | 8.04e-05 | -0.2803 | Seborrheic keratosis | dermatologic | UricA | |
| 702.4 | | | | | 21998 | 2 | 21996 | | | | | 28993 | 10 | 28983 | | | Degenerative skin disorders | dermatologic | UricA | |

| phenotype | male_beta | male_SE | male_OR | male_p | male_n_total | male_n_cases | male_n_controls | female_beta | female_SE | female_OR | female_p | female_n_total | female_n_cases | female_n_controls | interaction_p | interaction_beta | description | group | lab | label |
|---|---|---|---|---|---|---|---|---|---|---|---|---|---|---|---|---|---|---|---|---|
| 703 | | | | | 25804 | 185 | 25619 | | | | | 33096 | 363 | 32733 | 0.2514 | -0.2161 | Diseases of nail, NOS | dermatologic | UricA | |
| 703.1 | | | | | 25920 | 301 | 25619 | | | | | 33093 | 360 | 32733 | 0.3292 | 0.1569 | Ingrowing nail | dermatologic | UricA | |
| 704 | 0.7508 | 0.0967 | 2.119 | 8.24e-15 | 26130 | 511 | 25619 | 0.2393 | 0.07161 | 1.27 | 8.32e-04 | 33832 | 1099 | 32733 | 0.002216 | -0.3521 | Diseases of hair and hair follicles | dermatologic | UricA | |
| 704.1 | | | | | 25706 | 87 | 25619 | 0.246 | 0.08898 | 1.279 | 0.005704 | 33424 | 691 | 32733 | 0.5212 | 0.1579 | Alopecia | dermatologic | UricA | |
| 704.11 | | | | | 25639 | 20 | 25619 | | | | | 32796 | 63 | 32733 | | | Alopecia Areata | dermatologic | UricA | |
| 704.12 | | | | | 25622 | 3 | 25619 | | | | | 32824 | 91 | 32733 | | | Telogen effluvium | dermatologic | UricA | |
| 704.2 | | | | | 25621 | 2 | 25619 | | | | | 32916 | 183 | 32733 | | | Hirsutism | dermatologic | UricA | |
| 704.8 | | | | | 25903 | 284 | 25619 | | | | | 32966 | 233 | 32733 | 0.1571 | -0.2616 | Other specified diseases of hair and hair follicles | dermatologic | UricA | |
| 705 | | | | | 24635 | 110 | 24525 | | | | | 32184 | 194 | 31990 | | | Disorders of sweat glands | dermatologic | UricA | |
| 705.1 | | | | | 24588 | 63 | 24525 | | | | | 32070 | 80 | 31990 | | | Dyshidrosis | dermatologic | UricA | |
| 705.3 | | | | | 24563 | 38 | 24525 | | | | | 32095 | 105 | 31990 | | | Hidradenitis | dermatologic | UricA | |
| 705.8 | | | | | 24661 | 136 | 24525 | | | | | 32208 | 218 | 31990 | | | Hyperhidrosis | dermatologic | UricA | |
| 706 | 0.4465 | 0.06262 | 1.563 | 1.00e-12 | 25847 | 1322 | 24525 | 0.2182 | 0.05626 | 1.244 | 1.05e-04 | 33934 | 1944 | 31990 | 2.88e-05 | -0.3405 | Diseases of sebaceous glands | dermatologic | UricA | |
| 706.1 | | | | | 24797 | 272 | 24525 | 0.153 | 0.0916 | 1.165 | 0.09493 | 32819 | 829 | 31990 | 0.9607 | 0.008894 | Acne | dermatologic | UricA | |
| 706.2 | 0.3839 | 0.08181 | 1.468 | 2.69e-06 | 25246 | 721 | 24525 | 0.2427 | 0.08038 | 1.275 | 0.002537 | 32825 | 835 | 31990 | 0.1906 | -0.1456 | Sebaceous cyst | dermatologic | UricA | |
| 706.3 | | | | | 24526 | 1 | 24525 | | | | | 31991 | 1 | 31990 | | | Seborrhea | dermatologic | UricA | |
| 706.8 | | | | | 24813 | 288 | 24525 | | | | | 32225 | 235 | 31990 | 0.1759 | -0.2508 | Other specified diseases of sebaceous glands | dermatologic | UricA | |
| 707 | 0.6991 | 0.06202 | 2.012 | 1.82e-29 | 26784 | 1253 | 25531 | 0.9355 | 0.06882 | 2.548 | 4.42e-42 | 36008 | 999 | 35009 | 0.002803 | 0.2675 | Chronic ulcer of skin | dermatologic | UricA | |
| 707.1 | 0.5513 | 0.09188 | 1.736 | 1.97e-09 | 26112 | 581 | 25531 | | | | | 35488 | 479 | 35009 | 3.88e-04 | 0.4609 | Decubitus ulcer | dermatologic | UricA | |
| 707.2 | 0.8782 | 0.08115 | 2.407 | 2.68e-27 | 26220 | 689 | 25531 | | | | | 35483 | 474 | 35009 | 0.2125 | 0.153 | Chronic ulcer of leg or foot | dermatologic | UricA | |
| 707.3 | | | | | 25727 | 196 | 25531 | | | | | 35188 | 179 | 35009 | | | Chronic ulcer of unspecified site | dermatologic | UricA | |
| 709 | | | | | 23344 | 211 | 23133 | 0.2839 | 0.07187 | 1.328 | 7.80e-05 | 29862 | 1162 | 28700 | 0.6363 | 0.07935 | Diffuse diseases of connective tissue | dermatologic | UricA | |
| 709.2 | | | | | 23183 | 50 | 23133 | | | | | 29154 | 454 | 28700 | 0.3377 | -0.2965 | Sicca syndrome | dermatologic | UricA | |
| 709.3 | | | | | 23192 | 59 | 23133 | | | | | 28935 | 235 | 28700 | | | Systemic sclerosis | dermatologic | UricA | |
| 709.4 | | | | | 23156 | 23 | 23133 | | | | | 28760 | 60 | 28700 | | | Polymyositis | dermatologic | UricA | |
| 709.5 | | | | | 23161 | 28 | 23133 | | | | | 28793 | 93 | 28700 | | | Dermatomyositis | dermatologic | UricA | |
| 709.6 | | | | | 23165 | 32 | 23133 | | | | | 28946 | 246 | 28700 | | | Other specified diffuse diseases of connective tissue | dermatologic | UricA | |
| 709.7 | | | | | 23162 | 29 | 23133 | | | | | 28907 | 207 | 28700 | | | Unspecified diffuse connective tissue disease | dermatologic | UricA | |
| 710 | 0.4646 | 0.07899 | 1.591 | 4.07e-09 | 23512 | 830 | 22682 | 0.8179 | 0.08494 | 2.266 | 6.03e-22 | 30097 | 684 | 29413 | 4.37e-04 | 0.3969 | Osteomyelitis, periostitis, and other infections involving bone | musculoskeletal | UricA | |
| 710.1 | 0.4402 | 0.08358 | 1.553 | 1.38e-07 | 23427 | 745 | 22682 | 0.7577 | 0.09618 | 2.133 | 3.34e-15 | 29958 | 545 | 29413 | 0.007217 | 0.3336 | Osteomyelitis | musculoskeletal | UricA | |
| 710.11 | | | | | 22957 | 275 | 22682 | | | | | 29563 | 150 | 29413 | | | Acute osteomyelitis | musculoskeletal | UricA | |
| 710.12 | | | | | 23048 | 366 | 22682 | | | | | 29661 | 248 | 29413 | 0.1918 | 0.2393 | Chronic osteomyelitis | musculoskeletal | UricA | |
| 710.19 | 0.5658 | 0.09791 | 1.761 | 7.50e-09 | 23198 | 516 | 22682 | | | | | 29784 | 371 | 29413 | 0.1749 | 0.2006 | Unspecified osteomyelitis | musculoskeletal | UricA | |
| 710.2 | | | | | 22682 | | | | | | | 29416 | 3 | 29413 | | | Periostitis | musculoskeletal | UricA | |
| 710.3 | | | | | 22684 | 2 | 22682 | | | | | 29414 | 1 | 29413 | | | Osteopathy resulting from poliomyelitis | musculoskeletal | UricA | |
| 711 | | | | | 22939 | 257 | 22682 | | | | | 29633 | 220 | 29413 | | | Arthropathy associated with infections | musculoskeletal | UricA | |

| phenotype | male_beta | male_SE | male_OR | male_p | male_n_total | male_n_cases | male_n_controls | female_beta | female_SE | female_OR | female_p | female_n_total | female_n_cases | female_n_controls | interaction_p | interaction_beta | description | group | lab | label |
|---|---|---|---|---|---|---|---|---|---|---|---|---|---|---|---|---|---|---|---|---|
| 711.1 | | | | | 22893 | 211 | 22682 | | | | | 29566 | 153 | 29413 | | | Pyogenic arthritis | musculoskeletal | UricA | |
| 711.2 | | | | | 22696 | 14 | 22682 | | | | | 29424 | 11 | 29413 | | | Reiter's disease | musculoskeletal | UricA | |
| 711.3 | | | | | 22695 | 13 | 22682 | | | | | 29451 | 38 | 29413 | | | Behcet's syndrome | musculoskeletal | UricA | |
| 712 | | | | | 22798 | 116 | 22682 | | | | | 29481 | 68 | 29413 | | | Infective connective tissue disorders | musculoskeletal | UricA | |
| 713 | | | | | 22821 | 139 | 22682 | | | | | 29546 | 133 | 29413 | | | Arthropathy associated with other disorders classified elsewhere | musculoskeletal | UricA | |
| 713.5 | | | | | 22789 | 107 | 22682 | | | | | 29502 | 89 | 29413 | | | Arthropathy associated with neurological disorders | musculoskeletal | UricA | |
| 714 | 0.8455 | 0.07939 | 2.329 | 1.74e-26 | 24325 | 732 | 23593 | 0.6619 | 0.05356 | 1.938 | 4.42e-35 | 31860 | 1880 | 29980 | 0.219 | -0.1141 | Rheumatoid arthritis and other inflammatory polyarthropathies | musculoskeletal | UricA | |
| 714.1 | 0.718 | 0.09453 | 2.05 | 3.05e-14 | 24107 | 514 | 23593 | 0.6334 | 0.06134 | 1.884 | 5.40e-25 | 31387 | 1407 | 29980 | 0.945 | -0.007552 | Rheumatoid arthritis | musculoskeletal | UricA | |
| 714.2 | | | | | 23636 | 43 | 23593 | | | | | 30097 | 117 | 29980 | | | Juvenile rheumatoid arthritis | musculoskeletal | UricA | |
| 715 | | | | | 23907 | 314 | 23593 | 0.275 | 0.09304 | 1.317 | 0.003117 | 30615 | 635 | 29980 | 0.1967 | -0.1908 | Other inflammatory spondylopathies | musculoskeletal | UricA | |
| 715.1 | | | | | 23774 | 181 | 23593 | | | | | 30461 | 481 | 29980 | 0.04293 | -0.3718 | Sacroiliitis NEC | musculoskeletal | UricA | |
| 715.2 | | | | | 23683 | 90 | 23593 | | | | | 30051 | 71 | 29980 | | | Ankylosing spondylitis | musculoskeletal | UricA | |
| 715.3 | | | | | 23620 | 27 | 23593 | | | | | 30038 | 58 | 29980 | | | Spinal enthesopathy | musculoskeletal | UricA | |
| 716 | 0.6198 | 0.07362 | 1.858 | 3.83e-17 | 23562 | 880 | 22682 | 0.7298 | 0.06188 | 2.075 | 4.19e-32 | 30774 | 1361 | 29413 | 0.091 | 0.158 | Other arthropathies | musculoskeletal | UricA | |
| 716.1 | | | | | 22700 | 18 | 22682 | | | | | 29437 | 24 | 29413 | | | Unspecified polyarthropathy or polyarthritis | musculoskeletal | UricA | |
| 716.2 | | | | | 22688 | 6 | 22682 | | | | | 29420 | 7 | 29413 | | | Unspecified monoarthritis | musculoskeletal | UricA | |
| 716.3 | | | | | 22683 | 1 | 22682 | | | | | 29414 | 1 | 29413 | | | Kaschin-Beck disease | musculoskeletal | UricA | |
| 716.8 | | | | | 22688 | 6 | 22682 | | | | | 29426 | 13 | 29413 | | | Palindromic rheumatism | musculoskeletal | UricA | |
| 716.9 | 0.5902 | 0.07804 | 1.804 | 3.97e-14 | 23461 | 779 | 22682 | 0.6843 | 0.06528 | 1.982 | 1.05e-25 | 30638 | 1225 | 29413 | 0.1457 | 0.1438 | Arthropathy NOS | musculoskeletal | UricA | |
| 717 | | | | | 27682 | 166 | 27516 | | | | | 36769 | 263 | 36506 | | | Polymyalgia Rheumatica | musculoskeletal | UricA | |
| 720 | 0.4367 | 0.06102 | 1.548 | 8.24e-13 | 22346 | 1436 | 20910 | 0.3068 | 0.06061 | 1.359 | 4.17e-07 | 29466 | 1690 | 27776 | 0.2092 | -0.1054 | Spinal stenosis | musculoskeletal | UricA | |
| 720.1 | 0.4336 | 0.07379 | 1.543 | 4.21e-09 | 21846 | 936 | 20910 | 0.3051 | 0.0726 | 1.357 | 2.64e-05 | 28903 | 1127 | 27776 | 0.2674 | -0.1118 | Spinal stenosis of lumbar region | musculoskeletal | UricA | |
| 721 | 0.5241 | 0.05111 | 1.689 | 1.12e-24 | 23062 | 2152 | 20910 | 0.3522 | 0.04763 | 1.422 | 1.44e-13 | 30738 | 2962 | 27776 | 0.05073 | -0.1335 | Spondylosis and allied disorders | musculoskeletal | UricA | |
| 721.1 | 0.5333 | 0.05269 | 1.705 | 4.41e-24 | 22908 | 1998 | 20910 | 0.3682 | 0.04856 | 1.445 | 3.41e-14 | 30580 | 2804 | 27776 | 0.0598 | -0.1319 | Spondylosis without myelopathy | musculoskeletal | UricA | |
| 721.2 | | | | | 21155 | 245 | 20910 | | | | | 28026 | 250 | 27776 | | | Spondylosis with myelopathy | musculoskeletal | UricA | |
| 721.8 | | | | | 20961 | 51 | 20910 | | | | | 27797 | 21 | 27776 | | | Other allied disorders of spine | musculoskeletal | UricA | |
| 722 | 0.471 | 0.04834 | 1.602 | 1.95e-22 | 23387 | 2477 | 20910 | 0.2816 | 0.04527 | 1.325 | 4.98e-10 | 31200 | 3424 | 27776 | 0.01202 | -0.1627 | Intervertebral disc disorders | musculoskeletal | UricA | |
| 722.1 | 0.3798 | 0.09292 | 1.462 | 4.35e-05 | 21491 | 581 | 20910 | 0.1365 | 0.08815 | 1.146 | 0.1214 | 28557 | 781 | 27776 | 0.0781 | -0.2194 | Displacement of intervertebral disc | musculoskeletal | UricA | |

| phenotype | male_beta | male_SE | male_OR | male_p | male_n_total | male_n_cases | male_n_controls | female_beta | female_SE | female_OR | female_p | female_n_total | female_n_cases | female_n_controls | interaction_p | interaction_beta | description | group | lab | label |
|---|---|---|---|---|---|---|---|---|---|---|---|---|---|---|---|---|---|---|---|---|
| 722.3 | | | | | 20912 | 2 | 20910 | | | | | 27777 | 1 | 27776 | | | Schmorl's nodes | musculoskeletal | UricA | |
| 722.6 | 0.5368 | 0.05488 | 1.71 | 1.35e-22 | 22720 | 1810 | 20910 | 0.3505 | 0.04966 | 1.42 | 1.68e-12 | 30454 | 2678 | 27776 | 0.02039 | -0.1681 | Degeneration of intervertebral disc | musculoskeletal | UricA | |
| 722.7 | | | | | 21018 | 108 | 20910 | | | | | 27881 | 105 | 27776 | | | Intervertebral disc disorder with myelopathy | musculoskeletal | UricA | |
| 722.8 | | | | | 21238 | 328 | 20910 | | | | | 28155 | 379 | 27776 | 0.6946 | -0.06673 | Postlaminectomy syndrome | musculoskeletal | UricA | |
| 722.9 | | | | | 21012 | 102 | 20910 | | | | | 27883 | 107 | 27776 | | | Other and unspecified disc disorder | musculoskeletal | UricA | |
| 723 | | | | | 21252 | 342 | 20910 | | | | | 28186 | 410 | 27776 | 0.2163 | 0.2045 | Other disorders of cervical region | musculoskeletal | UricA | |
| 723.1 | | | | | 20943 | 33 | 20910 | | | | | 27812 | 36 | 27776 | | | Torticollis | musculoskeletal | UricA | |
| 724 | | | | | 21393 | 483 | 20910 | 0.3625 | 0.07932 | 1.437 | 4.86e-06 | 28648 | 872 | 27776 | 0.1708 | -0.1686 | Other and unspecified disorders of back | musculoskeletal | UricA | |
| 724.1 | | | | | 21038 | 128 | 20910 | | | | | 28127 | 351 | 27776 | | | Disorders of sacrum | musculoskeletal | UricA | |
| 724.2 | | | | | 21034 | 124 | 20910 | | | | | 28108 | 332 | 27776 | | | Disorders of coccyx | musculoskeletal | UricA | |
| 724.8 | | | | | 21013 | 103 | 20910 | | | | | 27951 | 175 | 27776 | | | Other symptoms referable to back | musculoskeletal | UricA | |
| 724.9 | | | | | 20981 | 71 | 20910 | | | | | 27892 | 116 | 27776 | | | Other unspecified back disorders | musculoskeletal | UricA | |
| 726 | 0.5906 | 0.04688 | 1.805 | 2.16e-36 | 23098 | 2817 | 20281 | 0.3524 | 0.04186 | 1.423 | 3.79e-17 | 30370 | 4595 | 25775 | 0.002835 | -0.1845 | Peripheral enthesopathies and allied syndromes | musculoskeletal | UricA | |
| 726.1 | 0.7071 | 0.0669 | 2.028 | 4.15e-26 | 21421 | 1140 | 20281 | 0.4009 | 0.05009 | 1.493 | 1.21e-15 | 28550 | 2775 | 25775 | 0.001421 | -0.262 | Enthesopathy | musculoskeletal | UricA | |
| 726.2 | | | | | 20365 | 84 | 20281 | | | | | 25894 | 119 | 25775 | | | Synoviopathy | musculoskeletal | UricA | |
| 726.3 | | | | | 20728 | 447 | 20281 | | | | | 26194 | 419 | 25775 | 0.05203 | -0.2822 | Bursitis | musculoskeletal | UricA | |
| 726.4 | | | | | 20361 | 80 | 20281 | | | | | 25949 | 174 | 25775 | | | Calcaneal spur; Exostosis NOS | musculoskeletal | UricA | |
| 727 | 0.6114 | 0.05748 | 1.843 | 2.01e-26 | 21945 | 1664 | 20281 | 0.4248 | 0.04941 | 1.529 | 8.21e-18 | 28564 | 2789 | 25775 | 0.07969 | -0.1301 | Other disorders of synovium, tendon, and bursa | musculoskeletal | UricA | |
| 727.1 | 0.7656 | 0.08144 | 2.15 | 5.41e-21 | 21009 | 728 | 20281 | 0.4945 | 0.06384 | 1.64 | 9.47e-15 | 27213 | 1438 | 25775 | 0.01863 | -0.2378 | Synovitis and tenosynovitis | musculoskeletal | UricA | |
| 727.2 | | | | | 20360 | 79 | 20281 | | | | | 25927 | 152 | 25775 | | | Bursitis disorders | musculoskeletal | UricA | |
| 727.4 | | | | | 20471 | 190 | 20281 | | | | | 26261 | 486 | 25775 | 0.4816 | -0.1308 | Ganglion and cyst of synovium, tendon, and bursa | musculoskeletal | UricA | |
| 727.5 | | | | | 20324 | 43 | 20281 | | | | | 25904 | 129 | 25775 | | | Rupture of synovium | musculoskeletal | UricA | |
| 727.6 | | | | | 20660 | 379 | 20281 | | | | | 26104 | 329 | 25775 | 0.1088 | 0.2623 | Rupture of tendon, nontraumatic | musculoskeletal | UricA | |
| 727.7 | | | | | 20347 | 66 | 20281 | | | | | 25864 | 89 | 25775 | | | Contracture of tendon (sheath) | musculoskeletal | UricA | |
| 727.8 | | | | | 20310 | 29 | 20281 | | | | | 25829 | 54 | 25775 | | | Plica syndrome | musculoskeletal | UricA | |
| 728 | | | | | 20364 | 83 | 20281 | | | | | 25899 | 124 | 25775 | | | Disorders of muscle, ligament, and fascia | musculoskeletal | UricA | |
| 728.1 | | | | | 20313 | 32 | 20281 | | | | | 25798 | 23 | 25775 | | | Muscular calcification and ossification | musculoskeletal | UricA | |
| 728.2 | | | | | 20304 | 23 | 20281 | | | | | 25866 | 91 | 25775 | | | Laxity of ligament or hypermobility syndrome | musculoskeletal | UricA | |
| 728.7 | | | | | 20721 | 440 | 20281 | 0.3838 | 0.08737 | 1.468 | 1.12e-05 | 26509 | 734 | 25775 | 0.3899 | -0.1142 | Fasciitis | musculoskeletal | UricA | |

| phenotype | male_beta | male_SE | male_OR | male_p | male_n_total | male_n_cases | male_n_controls | female_beta | female_SE | female_OR | female_p | female_n_total | female_n_cases | female_n_controls | interaction_p | interaction_beta | description | group | lab | label |
|---|---|---|---|---|---|---|---|---|---|---|---|---|---|---|---|---|---|---|---|---|
| 728.71 | | | | | 20411 | 130 | 20281 | | | | | 25848 | 73 | 25775 | | | Contracture of palmar fascia [Dupuytren's disease] | musculoskeletal | UricA | |
| 729 | 1.05 | 0.08273 | 2.858 | 6.33e-37 | 20964 | 683 | 20281 | 0.8717 | 0.07298 | 2.391 | 7.01e-33 | 26711 | 936 | 25775 | 0.2683 | -0.1191 | Other disorders of soft tissues | musculoskeletal | UricA | |
| 729.1 | | | | | 20284 | 3 | 20281 | | | | | 25782 | 7 | 25775 | | | Rheumatism, unspecified and fibrositis | musculoskeletal | UricA | |
| 729.3 | | | | | 20292 | 11 | 20281 | | | | | 25825 | 50 | 25775 | | | Panniculitis | musculoskeletal | UricA | |
| 729.7 | | | | | 20307 | 26 | 20281 | | | | | 25785 | 10 | 25775 | | | Nontraumatic compartment syndrome | musculoskeletal | UricA | |
| 731 | | | | | 22403 | 94 | 22309 | | | | | 27658 | 148 | 27510 | | | Osteitis deformans and osteopathies associated with other disorders classified elsewhere | musculoskeletal | UricA | |
| 731.1 | | | | | 22320 | 11 | 22309 | | | | | 27517 | 7 | 27510 | | | Osteitis deformans [Paget's disease of bone] | musculoskeletal | UricA | |
| 732 | | | | | 22384 | 75 | 22309 | | | | | 27583 | 73 | 27510 | | | Osteochondropathies | musculoskeletal | UricA | |
| 732.1 | | | | | 22341 | 32 | 22309 | | | | | 27549 | 39 | 27510 | | | Juvenile osteochondrosis | musculoskeletal | UricA | |
| 732.7 | | | | | 22348 | 39 | 22309 | | | | | 27539 | 29 | 27510 | | | Osteochondritis dissecans | musculoskeletal | UricA | |
| 733 | 0.8223 | 0.08758 | 2.276 | 6.03e-21 | 22918 | 609 | 22309 | 0.1844 | 0.05433 | 1.203 | 6.87e-04 | 29872 | 2362 | 27510 | 0.003421 | -0.2937 | Other disorders of bone and cartilage | musculoskeletal | UricA | |
| 733.2 | | | | | 22336 | 27 | 22309 | | | | | 27554 | 44 | 27510 | | | Cyst of bone | musculoskeletal | UricA | |

| phenotype | male_beta | male_SE | male_OR | male_p | male_n_total | male_n_cases | male_n_controls | female_beta | female_SE | female_OR | female_p | female_n_total | female_n_cases | female_n_controls | interaction_p | interaction_beta | description | group | lab | label |
|---|---|---|---|---|---|---|---|---|---|---|---|---|---|---|---|---|---|---|---|---|
| 733.4 | | | | | 22516 | 207 | 22309 | | | | | 27762 | 252 | 27510 | | | Aseptic necrosis of bone | musculoskeletal | UricA | |
| 733.6 | | | | | 22334 | 25 | 22309 | | | | | 27574 | 64 | 27510 | | | Costochondritis | musculoskeletal | UricA | |
| 733.8 | | | | | 22558 | 249 | 22309 | | | | | 27858 | 348 | 27510 | 5.31e-04 | 0.7047 | Malunion and nonunion of fracture | musculoskeletal | UricA | |
| 733.9 | | | | | 22350 | 41 | 22309 | | | | | 27565 | 55 | 27510 | | | Chondromalacia | musculoskeletal | UricA | |
| 735 | 0.6305 | 0.07538 | 1.879 | 6.05e-17 | 24919 | 827 | 24092 | 0.4533 | 0.06184 | 1.574 | 2.30e-13 | 32235 | 1405 | 30830 | 0.3626 | -0.08621 | Acquired foot deformities | musculoskeletal | UricA | |
| 735.1 | | | | | 24190 | 98 | 24092 | | | | | 30970 | 140 | 30830 | | | Flat foot | musculoskeletal | UricA | |
| 735.2 | | | | | 24498 | 406 | 24092 | 0.4968 | 0.08428 | 1.643 | 3.76e-09 | 31534 | 704 | 30830 | 0.3941 | -0.1116 | Acquired toe deformities | musculoskeletal | UricA | |
| 735.21 | | | | | 24374 | 282 | 24092 | 0.5334 | 0.09536 | 1.705 | 2.23e-08 | 31365 | 535 | 30830 | 0.3782 | -0.1348 | Hammer toe (acquired) | musculoskeletal | UricA | |
| 735.22 | | | | | 24107 | 15 | 24092 | | | | | 30844 | 14 | 30830 | | | Claw toe (acquired) | musculoskeletal | UricA | |
| 735.23 | | | | | 24184 | 92 | 24092 | | | | | 30968 | 138 | 30830 | | | Hallux rigidus | musculoskeletal | UricA | |
| 735.3 | | | | | 24207 | 115 | 24092 | 0.2955 | 0.1012 | 1.344 | 0.003518 | 31337 | 507 | 30830 | 0.6478 | -0.09782 | Hallux valgus (Bunion) | musculoskeletal | UricA | |
| 736 | | | | | 24293 | 201 | 24092 | | | | | 31078 | 248 | 30830 | | | Other acquired deformities of limbs | musculoskeletal | UricA | |
| 736.1 | | | | | 24103 | 11 | 24092 | | | | | 30840 | 10 | 30830 | | | Acquired deformities of forearm | musculoskeletal | UricA | |
| 736.2 | | | | | 24158 | 66 | 24092 | | | | | 30902 | 72 | 30830 | | | Acquired deformities of finger | musculoskeletal | UricA | |
| 736.3 | | | | | 24100 | 8 | 24092 | | | | | 30841 | 11 | 30830 | | | Acquired deformities of hip | musculoskeletal | UricA | |

| phenotype | male_beta | male_SE | male_OR | male_p | male_n_total | male_n_cases | male_n_controls | female_beta | female_SE | female_OR | female_p | female_n_total | female_n_cases | female_n_controls | interaction_p | interaction_beta | description | group | lab | label |
|---|---|---|---|---|---|---|---|---|---|---|---|---|---|---|---|---|---|---|---|---|
| 736.4 | | | | | 24104 | 12 | 24092 | | | | | 30857 | 27 | 30830 | | | Genu valgum or varum (acquired) | musculoskeletal | UricA | |
| 736.5 | | | | | 24095 | 3 | 24092 | | | | | 30840 | 10 | 30830 | | | Acquired deformities of knee | musculoskeletal | UricA | |
| 736.6 | | | | | 24114 | 22 | 24092 | | | | | 30876 | 46 | 30830 | | | Unequal leg length (acquired) | musculoskeletal | UricA | |
| 737 | | | | | 24514 | 422 | 24092 | 0.02915 | 0.08288 | 1.03 | 0.725 | 31760 | 930 | 30830 | 0.2438 | 0.1635 | Curvature of spine | musculoskeletal | UricA | |
| 737.1 | | | | | 24226 | 134 | 24092 | | | | | 31005 | 175 | 30830 | | | Kyphosis (acquired) | musculoskeletal | UricA | |
| 737.2 | | | | | 24093 | 1 | 24092 | | | | | 30832 | 2 | 30830 | | | Lordosis (acquired) | musculoskeletal | UricA | |
| 737.3 | | | | | 24413 | 321 | 24092 | -0.04641 | 0.09032 | 0.9547 | 0.6074 | 31641 | 811 | 30830 | 0.03657 | 0.3478 | Kyphoscoliosis and scoliosis | musculoskeletal | UricA | |
| 738 | 0.3813 | 0.094 | 1.464 | 4.99e-05 | 24637 | 545 | 24092 | 0.1589 | 0.0775 | 1.172 | 0.04037 | 31798 | 968 | 30830 | 0.3237 | -0.1164 | Other acquired musculoskeletal deformity | musculoskeletal | UricA | |
| 738.4 | | | | | 24467 | 375 | 24092 | 0.1777 | 0.08489 | 1.194 | 0.03631 | 31608 | 778 | 30830 | 0.1614 | -0.1897 | Acquired spondylolisthesis | musculoskeletal | UricA | |
| 739 | | | | | 24307 | 215 | 24092 | | | | | 31047 | 217 | 30830 | | | Contracture of joint | musculoskeletal | UricA | |
| 740 | 0.6596 | 0.03907 | 1.934 | 5.99e-64 | 25263 | 4641 | 20622 | 0.5298 | 0.03697 | 1.699 | 1.43e-46 | 33672 | 7527 | 26145 | 0.02923 | -0.1161 | Osteoarthrosis | musculoskeletal | UricA | |
| 740.1 | 0.6221 | 0.04595 | 1.863 | 9.49e-42 | 23687 | 3065 | 20622 | 0.5441 | 0.04333 | 1.723 | 3.62e-36 | 30839 | 4694 | 26145 | 0.3348 | -0.06027 | Osteoarthritis; localized | musculoskeletal | UricA | |
| 740.11 | 0.6032 | 0.04769 | 1.828 | 1.12e-36 | 23439 | 2817 | 20622 | 0.5277 | 0.04464 | 1.695 | 2.98e-32 | 30486 | 4341 | 26145 | 0.3319 | -0.0627 | Osteoarthrosis, localized, primary | musculoskeletal | UricA | |
| 740.12 | | | | | 20717 | 95 | 20622 | | | | | 26285 | 140 | 26145 | | | Osteoarthrosis, localized, secondary | musculoskeletal | UricA | |
| 740.2 | 1.036 | 0.08609 | 2.819 | 2.23e-33 | 21264 | 642 | 20622 | 0.8187 | 0.05754 | 2.267 | 6.25e-46 | 28076 | 1931 | 26145 | 0.02254 | -0.2329 | Osteoarthrosis, generalized | musculoskeletal | UricA | |
| 740.3 | | | | | 20669 | 47 | 20622 | | | | | 26282 | 137 | 26145 | | | Osteoarthrosis involving more than one site, but not specified as generalized | musculoskeletal | UricA | |
| 740.9 | 0.7901 | 0.04893 | 2.204 | 1.15e-58 | 23067 | 2445 | 20622 | 0.6103 | 0.04467 | 1.841 | 1.70e-42 | 30322 | 4177 | 26145 | 0.02017 | -0.1522 | Osteoarthrosis NOS | musculoskeletal | UricA | |
| 741 | 0.7527 | 0.04752 | 2.123 | 1.66e-56 | 24701 | 2405 | 22296 | 0.5202 | 0.04206 | 1.682 | 3.95e-35 | 32920 | 3637 | 29283 | 0.01052 | -0.1588 | Symptoms and disorders of the joints | musculoskeletal | UricA | |
| 741.1 | | | | | 22321 | 25 | 22296 | | | | | 29334 | 51 | 29283 | | | Ankylosis of joint | musculoskeletal | UricA | |
| 741.2 | 0.4748 | 0.09011 | 1.608 | 1.37e-07 | 22901 | 605 | 22296 | 0.3071 | 0.06888 | 1.359 | 8.26e-06 | 30510 | 1227 | 29283 | 0.3073 | -0.1123 | Stiffness of joint | musculoskeletal | UricA | |
| 741.3 | 0.7898 | 0.08479 | 2.203 | 1.23e-20 | 22942 | 646 | 22296 | 0.8161 | 0.07378 | 2.262 | 1.92e-28 | 30187 | 904 | 29283 | 0.5512 | 0.06531 | Difficulty in walking | musculoskeletal | UricA | |
| 741.4 | 1.223 | 0.08375 | 3.398 | 2.64e-48 | 22964 | 668 | 22296 | 0.7648 | 0.07171 | 2.149 | 1.49e-26 | 30241 | 958 | 29283 | 9.01e-04 | -0.3552 | Joint effusions | musculoskeletal | UricA | |
| 741.5 | | | | | 22328 | 32 | 22296 | | | | | 29301 | 18 | 29283 | | | Hemarthrosis | musculoskeletal | UricA | |
| 741.6 | | | | | 22307 | 11 | 22296 | | | | | 29300 | 17 | 29283 | | | Villonodular synovitis | musculoskeletal | UricA | |
| 742 | | | | | 22735 | 439 | 22296 | 0.4176 | 0.09109 | 1.518 | 4.55e-06 | 29934 | 651 | 29283 | 0.1082 | 0.2229 | Derangement of joint, non-traumatic | musculoskeletal | UricA | |
| 742.1 | | | | | 22319 | 23 | 22296 | | | | | 29301 | 18 | 29283 | | | Loose body in joint | musculoskeletal | UricA | |
| 742.2 | | | | | 22327 | 31 | 22296 | | | | | 29340 | 57 | 29283 | | | Pathological, developmental or recurrent dislocation | musculoskeletal | UricA | |
| 742.8 | | | | | 22322 | 26 | 22296 | | | | | 29329 | 46 | 29283 | | | Articular cartilage disorder | musculoskeletal | UricA | |
| 742.9 | | | | | 22643 | 347 | 22296 | 0.4001 | 0.101 | 1.492 | 7.41e-05 | 29808 | 525 | 29283 | 0.1311 | 0.2333 | Other derangement of joint | musculoskeletal | UricA | |

| phenotype | male_beta | male_SE | male_OR | male_p | male_n_total | male_n_cases | male_n_controls | female_beta | female_SE | female_OR | female_p | female_n_total | female_n_cases | female_n_controls | interaction_p | interaction_beta | description | group | lab | label |
|---|---|---|---|---|---|---|---|---|---|---|---|---|---|---|---|---|---|---|---|---|
| 743 | 0.9764 | 0.05042 | 2.655 | 1.58e-83 | 26032 | 1946 | 24086 | 0.6215 | 0.03726 | 1.862 | 1.83e-62 | 33952 | 6028 | 27924 | 0.07833 | -0.1076 | Osteoporosis, osteopenia and pathological fracture | musculoskeletal | UricA | |
| 743.1 | 1.186 | 0.06764 | 3.273 | 8.89e-69 | 25110 | 1024 | 24086 | 0.6684 | 0.04359 | 1.951 | 4.62e-53 | 31886 | 3962 | 27924 | 5.07e-05 | -0.3184 | Osteoporosis | musculoskeletal | UricA | |
| 743.11 | 1.074 | 0.07766 | 2.928 | 1.58e-43 | 24850 | 764 | 24086 | 0.6401 | 0.04721 | 1.897 | 6.88e-42 | 31204 | 3280 | 27924 | 0.006946 | -0.2396 | Osteoporosis NOS | musculoskeletal | UricA | |
| 743.12 | | | | | 24196 | 110 | 24086 | 0.7276 | 0.08053 | 2.07 | 1.63e-19 | 28758 | 834 | 27924 | 0.875 | -0.03267 | Senile osteoporosis | musculoskeletal | UricA | |
| 743.13 | | | | | 24546 | 460 | 24086 | 0.9726 | 0.06805 | 2.645 | 2.47e-46 | 29013 | 1089 | 27924 | 0.008537 | -0.311 | Other specified osteoporosis | musculoskeletal | UricA | |
| 743.2 | | | | | 24445 | 359 | 24086 | 0.7507 | 0.09113 | 2.118 | 1.77e-16 | 28551 | 627 | 27924 | 0.002989 | 0.4285 | Pathologic fracture | musculoskeletal | UricA | |
| 743.21 | | | | | 24187 | 101 | 24086 | | | | | 28089 | 165 | 27924 | | | Pathologic fracture of vertebrae | musculoskeletal | UricA | |
| 743.22 | | | | | 24160 | 74 | 24086 | | | | | 28032 | 108 | 27924 | | | Pathologic fracture of femur | musculoskeletal | UricA | |
| 743.4 | | | | | 24126 | 40 | 24086 | | | | | 28053 | 129 | 27924 | | | Stress fracture | musculoskeletal | UricA | |
| 743.9 | 0.8899 | 0.07966 | 2.435 | 5.67e-29 | 24819 | 733 | 24086 | 0.6392 | 0.05035 | 1.895 | 6.19e-37 | 30516 | 2592 | 27924 | 0.1805 | 0.1224 | Osteopenia or other disorder of bone and cartilage | musculoskeletal | UricA | |
| 745 | 0.7069 | 0.03538 | 2.028 | 7.88e-89 | 24606 | 7378 | 17228 | 0.453 | 0.03322 | 1.573 | 2.39e-42 | 32439 | 12064 | 20375 | 1.94e-04 | -0.1786 | Pain in joint | musculoskeletal | UricA | |
| 747 | 0.1612 | 0.06479 | 1.175 | 0.01284 | 26877 | 1323 | 25554 | 0.3483 | 0.06949 | 1.417 | 5.39e-07 | 36150 | 1183 | 34967 | 0.3599 | 0.08406 | Cardiac and circulatory congenital anomalies | congenital anomalies | UricA | |
| 747.1 | 0.1653 | 0.06722 | 1.18 | 0.0139 | 26767 | 1213 | 25554 | 0.3495 | 0.07372 | 1.418 | 2.13e-06 | 36008 | 1041 | 34967 | 0.4631 | 0.07073 | Cardiac congenital anomalies | congenital anomalies | UricA | |
| 747.11 | | | | | 25997 | 443 | 25554 | 0.2134 | 0.1032 | 1.238 | 0.03868 | 35539 | 572 | 34967 | 0.475 | -0.1043 | Cardiac shunt/ heart septal defect | congenital anomalies | UricA | |
| 747.12 | | | | | 25876 | 322 | 25554 | | | | | 35219 | 252 | 34967 | 0.1954 | 0.2507 | Valvular heart disease/ heart chambers | congenital anomalies | UricA | |
| 747.13 | | | | | 25697 | 143 | 25554 | | | | | 35131 | 164 | 34967 | | | Congenital anomalies of great vessels | congenital anomalies | UricA | |

| phenotype | male_beta | male_SE | male_OR | male_p | male_n_total | male_n_cases | male_n_controls | female_beta | female_SE | female_OR | female_p | female_n_total | female_n_cases | female_n_controls | interaction_p | interaction_beta | description | group | lab | label |
|---|---|---|---|---|---|---|---|---|---|---|---|---|---|---|---|---|---|---|---|---|
| 747.2 | | | | | 25638 | 84 | 25554 | | | | | 35084 | 117 | 34967 | | | Congenital anomalies of peripheral vascular system | congenital anomalies | UricA | |
| 748 | | | | | 27528 | 39 | 27489 | | | | | 36621 | 44 | 36577 | | | Anomalies of respiratory system, congenital | congenital anomalies | UricA | |
| 749 | | | | | 27561 | 72 | 27489 | | | | | 36671 | 94 | 36577 | | | Congenital anomalies of face and neck | congenital anomalies | UricA | |
| 749.1 | | | | | 22492 | 16 | 22476 | | | | | 31338 | 20 | 31318 | | | Cleft palate | congenital anomalies | UricA | |
| 749.2 | | | | | 27520 | 31 | 27489 | | | | | 36613 | 36 | 36577 | | | Congenital anomalies of skull and face bones | congenital anomalies | UricA | |
| 750 | | | | | 26052 | 143 | 25909 | | | | | 35186 | 190 | 34996 | | | Digestive congenital anomalies | congenital anomalies | UricA | |
| 750.1 | | | | | 25951 | 42 | 25909 | | | | | 35050 | 54 | 34996 | | | Upper gastrointestinal congenital anomalies | congenital anomalies | UricA | |
| 750.11 | | | | | 25920 | 11 | 25909 | | | | | 35005 | 9 | 34996 | | | Esophageal atresia/tracheoesophageal fistula | congenital anomalies | UricA | |
| 750.13 | | | | | 25928 | 19 | 25909 | | | | | 35009 | 13 | 34996 | | | Congenital anomalies of mouth/tongue | congenital anomalies | UricA | |
| 750.14 | | | | | 25914 | 5 | 25909 | | | | | 35012 | 16 | 34996 | | | Congenital anomalies of esophagus | congenital anomalies | UricA | |
| 750.15 | | | | | 25911 | 2 | 25909 | | | | | 34997 | 1 | 34996 | | | Congenital anomalies of stomach | congenital anomalies | UricA | |
| 750.2 | | | | | 26004 | 95 | 25909 | | | | | 35124 | 128 | 34996 | | | Lower gastrointestinal congenital anomalies | congenital anomalies | UricA | |

| phenotype | male_beta | male_SE | male_OR | male_p | male_n_total | male_n_cases | male_n_controls | female_beta | female_SE | female_OR | female_p | female_n_total | female_n_cases | female_n_controls | interaction_p | interaction_beta | description | group | lab | label |
|---|---|---|---|---|---|---|---|---|---|---|---|---|---|---|---|---|---|---|---|---|
| 750.21 | | | | | 25974 | 65 | 25909 | | | | | 35061 | 65 | 34996 | | | Congenital anomalies of intestine | congenital anomalies | UricA | |
| 750.22 | | | | | 25936 | 27 | 25909 | | | | | 35055 | 59 | 34996 | | | Congenital anomaly of gallbladder, bile ducts, liver, pancreas | congenital anomalies | UricA | |
| 750.5 | | | | | 25913 | 4 | 25909 | | | | | 34999 | 3 | 34996 | | | Congenital hypertrophic pyloric stenosis | congenital anomalies | UricA | |
| 751 | | | | | 26398 | 489 | 25909 | | | | | 35443 | 447 | 34996 | 0.635 | 0.06404 | Genitourinary congenital anomalies | congenital anomalies | UricA | |
| 751.1 | | | | | 26010 | 101 | 25909 | | | | | 35087 | 91 | 34996 | | | Congenital anomalies of genital organs | congenital anomalies | UricA | |
| 751.11 | | | | | 7 | | | | | | | 35071 | 79 | 34992 | | | Congenital anomalies of female genital organs | congenital anomalies | UricA | |
| 751.12 | | | | | 25992 | 90 | 25902 | | | | | 4 | | | | | Congenital anomalies of male genital organs | congenital anomalies | UricA | |
| 751.2 | | | | | 26267 | 358 | 25909 | | | | | 35304 | 308 | 34996 | 0.85 | 0.02983 | Congenital anomalies of urinary system | congenital anomalies | UricA | |
| 751.21 | | | | | 26140 | 231 | 25909 | | | | | 35199 | 203 | 34996 | | | Cystic kidney disease | congenital anomalies | UricA | |
| 751.22 | | | | | 26007 | 98 | 25909 | | | | | 35074 | 78 | 34996 | | | Other specified congenital anomalies of kidney | congenital anomalies | UricA | |
| 751.3 | | | | | 25949 | 40 | 25909 | | | | | 35047 | 51 | 34996 | | | Obstructive genitourinary defect | congenital anomalies | UricA | |
| 752 | | | | | 27335 | 160 | 27175 | | | | | 36299 | 223 | 36076 | | | Nervous system congenital anomalies | congenital anomalies | UricA | |
| 752.1 | | | | | 27234 | 59 | 27175 | | | | | 36188 | 112 | 36076 | | | Neural tube defects | congenital anomalies | UricA | |
| 752.11 | | | | | 27225 | 50 | 27175 | | | | | 36167 | 91 | 36076 | | | Spina bifida | congenital anomalies | UricA | |
| 752.2 | | | | | 27279 | 104 | 27175 | | | | | 36200 | 124 | 36076 | | | Other specified congenital anomalies of nervous system | congenital anomalies | UricA | |
| 753 | | | | | 27198 | 23 | 27175 | | | | | 36127 | 51 | 36076 | | | Congenital anomalies of the eye | congenital anomalies | UricA | |
| 753.1 | | | | | 27183 | 8 | 27175 | | | | | 36087 | 11 | 36076 | | | Congenital cataract and lens anomalies | congenital anomalies | UricA | |
| 753.2 | | | | | 27181 | 6 | 27175 | | | | | 36095 | 19 | 36076 | | | Congenital anomalies of posterior segment of eye | congenital anomalies | UricA | |
| 754 | | | | | 27083 | 58 | 27025 | | | | | 35886 | 111 | 35775 | | | Congenital musculoskeletal deformities of spine | congenital anomalies | UricA | |
| 754.1 | | | | | 27033 | 8 | 27025 | | | | | 35793 | 18 | 35775 | | | Lumbosacral spondylolysis, congenital | congenital anomalies | UricA | |
| 754.2 | | | | | 27059 | 34 | 27025 | | | | | 35837 | 62 | 35775 | | | Spondylolisthesis, congenital | congenital anomalies | UricA | |
| 755 | | | | | 27119 | 94 | 27025 | | | | | 35920 | 145 | 35775 | | | Congenital anomalies of limbs | congenital anomalies | UricA | |
| 755.1 | | | | | 27058 | 33 | 27025 | | | | | 35834 | 59 | 35775 | | | Congenital deformities of feet | congenital anomalies | UricA | |
| 755.3 | | | | | 27031 | 6 | 27025 | | | | | 35781 | 6 | 35775 | | | Congenital anomaly of fingers/toes | congenital anomalies | UricA | |
| 755.4 | | | | | 27034 | 9 | 27025 | | | | | 35795 | 20 | 35775 | | | Congenital anomalies of upper limb, including shoulder girdle | congenital anomalies | UricA | |
| 755.6 | | | | | 27065 | 40 | 27025 | | | | | 35837 | 62 | 35775 | | | Other congenital anomalies of lower limb, including pelvic girdle | congenital anomalies | UricA | |
| 755.61 | | | | | 27060 | 35 | 27025 | | | | | 35828 | 53 | 35775 | | | Congenital hip dysplasia and deformity | congenital anomalies | UricA | |
| 756 | | | | | 27125 | 100 | 27025 | | | | | 35939 | 164 | 35775 | | | Other congenital musculoskeletal anomalies | congenital anomalies | UricA | |
| 756.1 | | | | | 27036 | 11 | 27025 | | | | | 35791 | 16 | 35775 | | | Congenital anomalies of abdominal wall; diaphram | congenital anomalies | UricA | |

| phenotype | male_beta | male_SE | male_OR | male_p | male_n_total | male_n_cases | male_n_controls | female_beta | female_SE | female_OR | female_p | female_n_total | female_n_cases | female_n_controls | interaction_p | interaction_beta | description | group | lab | label |
|---|---|---|---|---|---|---|---|---|---|---|---|---|---|---|---|---|---|---|---|---|
| 756.2 | | | | | 27064 | 39 | 27025 | | | | | 35801 | 26 | 35775 | | | Pectus and other congenital anomalies of ribs/sternum | congenital anomalies | UricA | |
| 756.21 | | | | | 27054 | 29 | 27025 | | | | | 35798 | 23 | 35775 | | | Pectus excavatum | congenital anomalies | UricA | |
| 756.22 | | | | | 27036 | 11 | 27025 | | | | | 35776 | 1 | 35775 | | | Pectus carinatum | congenital anomalies | UricA | |
| 756.3 | | | | | 27041 | 16 | 27025 | | | | | 35840 | 65 | 35775 | | | Congenital anomalies of muscle, tendon, fascia, and connective tissue | congenital anomalies | UricA | |
| 756.5 | | | | | 27043 | 18 | 27025 | | | | | 35804 | 29 | 35775 | | | Congenital osteodystrophies | congenital anomalies | UricA | |
| 757 | | | | | 26746 | 13 | 26733 | | | | | 35965 | 21 | 35944 | | | Congenital anomalies of the integument | congenital anomalies | UricA | |
| 758 | | | | | 26835 | 102 | 26733 | | | | | 36032 | 88 | 35944 | | | Chromosomal anomalies and genetic disorders | congenital anomalies | UricA | |
| 758.1 | | | | | 26801 | 68 | 26733 | | | | | 36013 | 69 | 35944 | | | Chromosomal anomalies | congenital anomalies | UricA | |
| 759 | | | | | 27017 | 284 | 26733 | | | | | 36141 | 197 | 35944 | | | Other and unspecified congenital anomalies | congenital anomalies | UricA | |
| 759.1 | | | | | 26741 | 8 | 26733 | | | | | 35957 | 13 | 35944 | | | Anomalies of endocrine glands, congenital | congenital anomalies | UricA | |
| 760 | 0.6146 | 0.03758 | 1.849 | 3.95e-60 | 24911 | 4715 | 20196 | 0.4949 | 0.03369 | 1.64 | 7.64e-49 | 32596 | 7782 | 24814 | 0.312 | -0.05012 | Back pain | symptoms | UricA | |
| 761 | 0.4062 | 0.05225 | 1.501 | 7.60e-15 | 26084 | 1972 | 24112 | 0.3546 | 0.04191 | 1.426 | 2.65e-17 | 34098 | 3706 | 30392 | 0.7197 | 0.02336 | Cervicalgia | symptoms | UricA | |
| 763 | 0.3189 | 0.06496 | 1.376 | 9.15e-07 | 25089 | 1208 | 23881 | 0.2185 | 0.05682 | 1.244 | 1.20e-04 | 32717 | 1862 | 30855 | 0.5586 | -0.04909 | Thoracic or lumbosacral neuritis or radiculitis, unspecified | symptoms | UricA | |
| 764 | 0.5342 | 0.08181 | 1.706 | 6.58e-11 | 24586 | 705 | 23881 | 0.1662 | 0.06684 | 1.181 | 0.01289 | 32170 | 1315 | 30855 | 0.00331 | -0.3011 | Sciatica | symptoms | UricA | |
| 765 | 0.3343 | 0.08923 | 1.397 | 1.79e-04 | 24494 | 613 | 23881 | 0.2434 | 0.07998 | 1.276 | 0.002339 | 31724 | 869 | 30855 | 0.648 | -0.05295 | Cervical radiculitis | symptoms | UricA | |
| 766 | 0.5627 | 0.08331 | 1.755 | 1.44e-11 | 24563 | 682 | 23881 | 0.4692 | 0.07015 | 1.599 | 2.25e-11 | 31922 | 1067 | 30855 | 0.9486 | 0.006787 | Neuralgia, neuritis, and radiculitis NOS | symptoms | UricA | |
| 767 | | | | | 23904 | 23 | 23881 | | | | | 30882 | 27 | 30855 | | | Cervicocranial/Cervicobrachial syndrome | symptoms | UricA | |
| 769 | | | | | 27744 | 17 | 27727 | | | | | 36864 | 59 | 36805 | | | Nonallopathic lesions NEC | symptoms | UricA | |
| 770 | 0.7957 | 0.08213 | 2.216 | 3.39e-22 | 26643 | 686 | 25957 | 0.4018 | 0.04479 | 1.494 | 2.93e-19 | 34609 | 3011 | 31598 | 5.09e-05 | -0.3645 | Myalgia and myositis unspecified | symptoms | UricA | |
| 771 | 0.7317 | 0.08987 | 2.079 | 3.87e-16 | 23704 | 575 | 23129 | 0.7031 | 0.08852 | 2.02 | 1.97e-15 | 31912 | 628 | 31284 | 0.7729 | 0.03523 | Musculoskeletal symptoms referable to limbs | symptoms | UricA | |
| 771.1 | 1.093 | 0.06244 | 2.984 | 1.20e-68 | 24349 | 1220 | 23129 | 1.051 | 0.05815 | 2.861 | 4.77e-73 | 32708 | 1424 | 31284 | 0.4168 | 0.06706 | Swelling of limb | symptoms | UricA | |
| 771.2 | | | | | 23196 | 67 | 23129 | | | | | 31394 | 110 | 31284 | | | Cramp of limb | symptoms | UricA | |
| 772 | | | | | 23175 | 134 | 23041 | | | | | 30713 | 257 | 30456 | | | Symptoms of the muscles | symptoms | UricA | |
| 772.1 | | | | | 23094 | 53 | 23041 | | | | | 30516 | 60 | 30456 | | | Muscular wasting and disuse atrophy | symptoms | UricA | |
| 772.2 | 0.4935 | 0.08945 | 1.638 | 3.45e-08 | 23676 | 635 | 23041 | 0.2765 | 0.07337 | 1.319 | 1.64e-04 | 31533 | 1077 | 30456 | 0.3454 | -0.1055 | Spasm of muscle | symptoms | UricA | |
| 772.3 | 0.7482 | 0.05586 | 2.113 | 6.63e-41 | 24642 | 1601 | 23041 | 0.6149 | 0.04952 | 1.849 | 2.12e-35 | 32724 | 2268 | 30456 | 0.2044 | -0.09229 | Muscle weakness | symptoms | UricA | |
| 772.4 | | | | | 23175 | 134 | 23041 | | | | | 30527 | 71 | 30456 | | | Rhabdomyolysis | symptoms | UricA | |
| 772.6 | | | | | 23142 | 101 | 23041 | | | | | 30584 | 128 | 30456 | | | Facial weakness | symptoms | UricA | |
| 780 | | | | | 27429 | 77 | 27352 | | | | | 36521 | 68 | 36453 | | | Hypothermia/Chills | symptoms | UricA | |
| 781 | 0.6922 | 0.09298 | 1.998 | 9.72e-14 | 26628 | 527 | 26101 | 0.5493 | 0.08382 | 1.732 | 5.62e-11 | 35384 | 686 | 34698 | 0.539 | -0.07422 | Symptoms involving nervous and musculoskeletal systems | symptoms | UricA | |
| 781.1 | | | | | 26112 | 11 | 26101 | | | | | 34735 | 37 | 34698 | | | Loss of height | symptoms | UricA | |

| phenotype | male_beta | male_SE | male_OR | male_p | male_n_total | male_n_cases | male_n_controls | female_beta | female_SE | female_OR | female_p | female_n_total | female_n_cases | female_n_controls | interaction_p | interaction_beta | description | group | lab | label |
|---|---|---|---|---|---|---|---|---|---|---|---|---|---|---|---|---|---|---|---|---|
| 781.2 | | | | | 26298 | 197 | 26101 | | | | | 34933 | 235 | 34698 | | | Abnormal posture | symptoms | UricA | |
| 782 | | | | | 22002 | 106 | 21896 | | | | | 29294 | 158 | 29136 | | | Symptoms involving skin and other integumentary tissue | symptoms | UricA | |
| 782.3 | 1.166 | 0.04241 | 3.21 | 1.92e-166 | 24907 | 3011 | 21896 | 1.103 | 0.03924 | 3.012 | 1.14e-173 | 33039 | 3903 | 29136 | 0.9781 | 0.001551 | Edema | symptoms | UricA | |
| 782.6 | | | | | 21947 | 51 | 21896 | | | | | 29472 | 336 | 29136 | | | Pallor and flushing | symptoms | UricA | |
| 783 | 1.021 | 0.03833 | 2.777 | 1.94e-156 | 24799 | 4188 | 20611 | 0.9509 | 0.03744 | 2.588 | 2.77e-142 | 32997 | 4266 | 28731 | 0.6319 | 0.02481 | Fever of unknown origin | symptoms | UricA | |
| 783.1 | | | | | 20666 | 55 | 20611 | | | | | 28768 | 37 | 28731 | | | Postprocedural fever | symptoms | UricA | |
| 785 | 0.6717 | 0.03466 | 1.958 | 1.12e-83 | 24378 | 6486 | 17892 | 0.6419 | 0.03159 | 1.9 | 8.10e-92 | 32108 | 11436 | 20672 | 0.6783 | 0.019 | Abdominal pain | symptoms | UricA | |
| 788 | 0.5391 | 0.04903 | 1.714 | 4.06e-28 | 26167 | 2188 | 23979 | 0.5809 | 0.04665 | 1.788 | 1.36e-35 | 34916 | 2566 | 32350 | 0.5865 | 0.0358 | Syncope and collapse | symptoms | UricA | |
| 789 | 0.7066 | 0.03905 | 2.027 | 3.42e-73 | 24975 | 4146 | 20829 | 0.7615 | 0.033 | 2.142 | 7.36e-118 | 32311 | 6758 | 25553 | 7.95e-05 | 0.1949 | Nausea and vomiting | symptoms | UricA | |
| 789.1 | | | | | 20853 | 24 | 20829 | | | | | 25611 | 58 | 25553 | | | Persistent vomiting | symptoms | UricA | |
| 790 | | | | | 22915 | 183 | 22732 | | | | | 30897 | 312 | 30585 | | | Nonspecific findings on examination of blood | symptoms | UricA | |
| 790.1 | | | | | 22812 | 80 | 22732 | | | | | 30855 | 270 | 30585 | | | Elevated sedimentation rate | symptoms | UricA | |
| 790.6 | 1.115 | 0.04783 | 3.048 | 3.99e-120 | 25006 | 2274 | 22732 | 0.8388 | 0.04506 | 2.314 | 2.37e-77 | 33298 | 2713 | 30585 | 0.006272 | -0.1752 | Other abnormal blood chemistry | symptoms | UricA | |
| 790.8 | | | | | 22867 | 135 | 22732 | | | | | 30869 | 284 | 30585 | | | Elevated C-reactive protein (CRP) | symptoms | UricA | |
| 790.9 | | | | | 22737 | 5 | 22732 | | | | | 30591 | 6 | 30585 | | | Abnormal arterial blood gases | symptoms | UricA | |
| 791 | | | | | 27648 | 165 | 27483 | | | | | 36840 | 92 | 36748 | | | Gangrene | symptoms | UricA | |
| 792 | | | | | 7 | | | 0.5786 | 0.06038 | 1.783 | 9.56e-22 | 29446 | 1607 | 27839 | 0.9989 | 0.3779 | Abnormal Papanicolaou smear of cervix and cervical HPV | genitourinary | UricA | |
| 792.1 | | | | | 7 | | | 0.636 | 0.06548 | 1.889 | 2.63e-22 | 29183 | 1344 | 27839 | 0.9989 | 0.3612 | Papanicolaou smear of cervix or vagina with atypical squamous cells | genitourinary | UricA | |
| 794 | | | | | 27589 | 45 | 27544 | | | | | 36771 | 38 | 36733 | | | Abnormal results of other function studies (bladder, pancreas, placenta, spleen, etc) | symptoms | UricA | |
| 795 | | | | | 27272 | 12 | 27260 | | | | | 36232 | 106 | 36126 | | | Other and nonspecific abnormal cytological, histological and immunological findings | symptoms | UricA | |
| 795.8 | | | | | 27502 | 242 | 27260 | | | | | 36401 | 275 | 36126 | 0.5226 | 0.1322 | Abnormal tumor markers | symptoms | UricA | |
| 795.81 | | | | | 27386 | 126 | 27260 | | | | | 36227 | 101 | 36126 | | | Elevated carcinoembryonic antigen [CEA] | symptoms | UricA | |
| 795.82 | | | | | 27261 | 1 | 27260 | | | | | 36221 | 95 | 36126 | | | Elevated cancer antigen 125 [CA 125] | symptoms | UricA | |
| 796 | 0.4751 | 0.06357 | 1.608 | 7.81e-14 | 20872 | 1484 | 19388 | | | | | 4 | | | | | Elevated prostate specific antigen [PSA] | genitourinary | UricA | |
| 797 | 1.047 | 0.06745 | 2.848 | 2.54e-54 | 26901 | 1035 | 25866 | 1.048 | 0.08043 | 2.851 | 8.79e-39 | 36273 | 725 | 35548 | 0.4456 | 0.07736 | Shock | symptoms | UricA | |
| 797.1 | | | | | 26238 | 372 | 25866 | | | | | 35746 | 198 | 35548 | 0.5632 | -0.1035 | Cardiogenic shock | symptoms | UricA | |
| 798 | 0.9897 | 0.03398 | 2.691 | 1.47e-186 | 23475 | 7649 | 15826 | 0.6071 | 0.03218 | 1.835 | 2.28e-79 | 30980 | 11865 | 19115 | 2.31e-11 | -0.3098 | Malaise and fatigue | symptoms | UricA | Malaise and fatigue |
| 798.1 | | | | | 16119 | 293 | 15826 | 0.6806 | 0.08708 | 1.975 | 5.44e-15 | 19839 | 724 | 19115 | 0.002539 | -0.4471 | Chronic fatigue syndrome | symptoms | UricA | |
| 800 | 0.01899 | 0.07537 | 1.019 | 0.801 | 22951 | 1064 | 21887 | 0.4506 | 0.05808 | 1.569 | 8.62e-15 | 30522 | 1698 | 28824 | 1.56e-11 | 0.6211 | Fracture of lower limb | injuries & poisonings | UricA | Fracture of lower limb |
| 800.1 | | | | | 22264 | 377 | 21887 | 0.6273 | 0.09442 | 1.873 | 3.06e-11 | 29394 | 570 | 28824 | 2.19e-07 | 0.7782 | Fracture of neck of femur | injuries & poisonings | UricA | |
| 800.2 | | | | | 22107 | 220 | 21887 | | | | | 29062 | 238 | 28824 | | | Fracture of unspecified part of femur | injuries & poisonings | UricA | |

| phenotype | male_beta | male_SE | male_OR | male_p | male_n_total | male_n_cases | male_n_controls | female_beta | female_SE | female_OR | female_p | female_n_total | female_n_cases | female_n_controls | interaction_p | interaction_beta | description | group | lab | label |
|---|---|---|---|---|---|---|---|---|---|---|---|---|---|---|---|---|---|---|---|---|
| 800.3 | | | | | 22240 | 353 | 21887 | | | | | 29291 | 467 | 28824 | 0.001602 | 0.5357 | Fracture of tibia and fibula | injuries & poisonings | UricA | |
| 800.4 | | | | | 22024 | 137 | 21887 | | | | | 29015 | 191 | 28824 | | | Fracture of patella | injuries & poisonings | UricA | |
| 801 | 0.2373 | 0.09827 | 1.268 | 0.01577 | 22470 | 583 | 21887 | 0.3668 | 0.06785 | 1.443 | 6.43e-08 | 30057 | 1233 | 28824 | 3.01e-04 | 0.4163 | Fracture of ankle and foot | injuries & poisonings | UricA | |
| 801.1 | | | | | 22222 | 335 | 21887 | 0.2888 | 0.1038 | 1.335 | 0.005376 | 29349 | 525 | 28824 | 0.009953 | 0.4189 | Fracture of foot | injuries & poisonings | UricA | |
| 802 | | | | | 22131 | 244 | 21887 | | | | | 29104 | 280 | 28824 | 0.06974 | 0.3668 | Fracture of pelvis | injuries & poisonings | UricA | |
| 803 | 0.1999 | 0.07568 | 1.221 | 0.008264 | 22909 | 1022 | 21887 | 0.245 | 0.06181 | 1.278 | 7.40e-05 | 30429 | 1605 | 28824 | 0.001284 | 0.3031 | Fracture of upper limb | injuries & poisonings | UricA | |
| 803.1 | | | | | 22122 | 235 | 21887 | | | | | 29293 | 469 | 28824 | 0.05228 | 0.348 | Fracture of humerus | injuries & poisonings | UricA | |
| 803.2 | | | | | 22365 | 478 | 21887 | 0.2011 | 0.08302 | 1.223 | 0.0154 | 29684 | 860 | 28824 | 0.02843 | 0.2902 | Fracture of radius and ulna | injuries & poisonings | UricA | |
| 803.21 | | | | | 21931 | 44 | 21887 | | | | | 28974 | 150 | 28824 | | | Colles' fracture | injuries & poisonings | UricA | |
| 803.3 | | | | | 22141 | 254 | 21887 | | | | | 28997 | 173 | 28824 | | | Fracture of clavicle or scapula | injuries & poisonings | UricA | |
| 804 | | | | | 22374 | 487 | 21887 | | | | | 29317 | 493 | 28824 | 0.1882 | 0.195 | Fracture of hand or wrist | injuries & poisonings | UricA | |
| 805 | 0.1862 | 0.0806 | 1.205 | 0.02091 | 22729 | 842 | 21887 | 0.3405 | 0.07646 | 1.406 | 8.46e-06 | 29817 | 993 | 28824 | 0.007122 | 0.2897 | Fracture of vertebral column without mention of spinal cord injury | injuries & poisonings | UricA | |
| 807 | | | | | 22286 | 399 | 21887 | | | | | 29056 | 232 | 28824 | 0.7316 | 0.07177 | Fracture of ribs | injuries & poisonings | UricA | |
| 809 | 0.2092 | 0.06147 | 1.233 | 6.64e-04 | 23404 | 1517 | 21887 | 0.3883 | 0.05657 | 1.474 | 6.71e-12 | 30670 | 1846 | 28824 | 1.01e-04 | 0.3149 | Fracture of unspecified bones | injuries & poisonings | UricA | |
| 816 | | | | | 26400 | 43 | 26357 | | | | | 35690 | 21 | 35669 | | | Cerebral laceration and contusion | injuries & poisonings | UricA | |
| 817 | | | | | 26422 | 65 | 26357 | | | | | 35734 | 65 | 35669 | | | Concussion | injuries & poisonings | UricA | |
| 818 | | | | | 26606 | 249 | 26357 | | | | | 35816 | 147 | 35669 | | | Intracranial hemorrhage (injury) | injuries & poisonings | UricA | |
| phenotype | male_beta | male_SE | male_OR | male_p | male_n_total | male_n_cases | male_n_controls | female_beta | female_SE | female_OR | female_p | female_n_total | female_n_cases | female_n_controls | interaction_p | interaction_beta | description | group | lab | label |
| 818.1 | | | | | 26475 | 118 | 26357 | | | | | 35737 | 68 | 35669 | | | Subdural hemorrhage (injury) | injuries & poisonings | UricA | |
| 818.2 | | | | | 26451 | 94 | 26357 | | | | | 35727 | 58 | 35669 | | | Subarachnoid hemorrhage (injury) | injuries & poisonings | UricA | |
| 819 | -0.1437 | 0.1133 | 0.8662 | 0.2048 | 26908 | 551 | 26357 | | | | | 36031 | 362 | 35669 | 4.44e-04 | 0.5787 | Skull and face fracture and other intercranial injury | injuries & poisonings | UricA | |
| 823 | | | | | 21319 | | | | | | | 28267 | 2 | 28265 | | | Torus fracture | injuries & poisonings | UricA | |
| 830 | 0.005625 | 0.07864 | 1.006 | 0.943 | 26492 | 928 | 25564 | 0.1349 | 0.06943 | 1.144 | 0.05204 | 34859 | 1219 | 33640 | 0.014 | 0.249 | Dislocation | injuries & poisonings | UricA | |
| 835 | | | | | 26013 | 449 | 25564 | 0.1707 | 0.08153 | 1.186 | 0.03626 | 34495 | 855 | 33640 | 0.9067 | -0.01509 | Internal derangement of knee | injuries & poisonings | UricA | |
| 836 | | | | | 25643 | 79 | 25564 | | | | | 33724 | 84 | 33640 | | | Traumatic arthropathy | injuries & poisonings | UricA | |
| 840 | 0.2861 | 0.063 | 1.331 | 5.61e-06 | 25492 | 1396 | 24096 | 0.2791 | 0.05377 | 1.322 | 2.10e-07 | 33198 | 2125 | 31073 | 0.574 | 0.04499 | Sprains and strains | injuries & poisonings | UricA | |
| 840.1 | | | | | 24130 | 34 | 24096 | | | | | 31116 | 43 | 31073 | | | Muscle/tendon sprain | injuries & poisonings | UricA | |
| 840.2 | | | | | 24369 | 273 | 24096 | | | | | 31341 | 268 | 31073 | 0.4454 | 0.1455 | Rotator cuff (capsule) sprain | injuries & poisonings | UricA | |

| phenotype | male_beta | male_SE | male_OR | male_p | male_n_total | male_n_cases | male_n_controls | female_beta | female_SE | female_OR | female_p | female_n_total | female_n_cases | female_n_controls | interaction_p | interaction_beta | description | group | lab | label |
|---|---|---|---|---|---|---|---|---|---|---|---|---|---|---|---|---|---|---|---|---|
| 840.3 | | | | | 24387 | 291 | 24096 | | | | | 31485 | 412 | 31073 | 0.5816 | -0.09783 | Joint/ligament sprain | injuries & poisonings | UricA | |
| 841 | | | | | 24387 | 291 | 24096 | 0.6112 | 0.08731 | 1.843 | 2.55e-12 | 31714 | 641 | 31073 | 0.9709 | -0.005355 | Sprains and strains of back and neck | injuries & poisonings | UricA | |
| 842 | | | | | 24179 | 83 | 24096 | | | | | 31195 | 122 | 31073 | | | Other sprains and strains | injuries & poisonings | UricA | |
| 850 | | | | | 23223 | 456 | 22767 | | | | | 32260 | 465 | 31795 | 0.906 | -0.01654 | Hemorrhage or hematoma complicating a procedure | injuries & poisonings | UricA | |
| 851 | | | | | 23157 | 390 | 22767 | | | | | 32086 | 291 | 31795 | 0.9032 | -0.02244 | Complications of transplants and reattached limbs | injuries & poisonings | UricA | |
| 853 | | | | | 22918 | 151 | 22767 | | | | | 31956 | 161 | 31795 | | | Complication of colostomy or enterostomy | injuries & poisonings | UricA | |
| 854 | 1.091 | 0.08323 | 2.977 | 3.00e-39 | 23435 | 668 | 22767 | 1.284 | 0.09242 | 3.611 | 7.13e-44 | 32312 | 517 | 31795 | 0.08855 | 0.2043 | Complications of cardiac/vascular device, implant, and graft | injuries & poisonings | UricA | |
| 855 | | | | | 22967 | 200 | 22767 | | | | | 32019 | 224 | 31795 | | | Complication of nervous system device, implant, and graft | injuries & poisonings | UricA | |
| 856 | | | | | 22823 | 56 | 22767 | | | | | 31840 | 45 | 31795 | | | Vascular complications of surgery and medical procedures | injuries & poisonings | UricA | |
| 857 | | | | | 22852 | 85 | 22767 | | | | | 31886 | 91 | 31795 | | | Mechanical complication of unspecified genitourinary device, implant, and graft | injuries & poisonings | UricA | |
| 858 | | | | | 23191 | 424 | 22767 | 0.1846 | 0.09941 | 1.203 | 0.06331 | 32396 | 601 | 31795 | 0.04169 | 0.3055 | Complication of internal orthopedic device | injuries & poisonings | UricA | |
| 859 | | | | | 23165 | 398 | 22767 | | | | | 32227 | 432 | 31795 | 0.02955 | 0.3519 | Complication due to other implant and internal device | injuries & poisonings | UricA | |
| 860 | | | | | 27702 | 127 | 27575 | | | | | 36911 | 120 | 36791 | | | Bone marrow or stem cell transplant | neoplasms | UricA | |
| 870 | 0.1713 | 0.08825 | 1.187 | 0.05221 | 24342 | 736 | 23606 | 0.4516 | 0.08747 | 1.571 | 2.43e-07 | 33149 | 715 | 32434 | 0.002622 | 0.3623 | Open wounds of head; neck; and trunk | injuries & poisonings | UricA | |
| 870.1 | | | | | 23654 | 48 | 23606 | | | | | 32454 | 20 | 32434 | | | Open wound or laceration of eye or eyelid | injuries & poisonings | UricA | |
| 870.2 | | | | | 23622 | 16 | 23606 | | | | | 32443 | 9 | 32434 | | | Open wound of ear | injuries & poisonings | UricA | |
| 870.3 | | | | | 23791 | 185 | 23606 | | | | | 32553 | 119 | 32434 | | | Other open wound of head and face | injuries & poisonings | UricA | |
| 870.4 | | | | | 23626 | 20 | 23606 | | | | | 32447 | 13 | 32434 | | | Open wound of nose and sinus | injuries & poisonings | UricA | |
| 870.5 | | | | | 23652 | 46 | 23606 | | | | | 32464 | 30 | 32434 | | | Open wound of lip and mouth | injuries & poisonings | UricA | |
| 870.6 | | | | | 23625 | 19 | 23606 | | | | | 32440 | 6 | 32434 | | | Open wound of neck | injuries & poisonings | UricA | |
| 870.8 | | | | | 23613 | 7 | 23606 | | | | | 32442 | 8 | 32434 | | | Open wound of genital organs | injuries & poisonings | UricA | |
| 871 | 0.3258 | 0.07172 | 1.385 | 5.57e-06 | 24632 | 1026 | 23606 | 0.5284 | 0.07398 | 1.696 | 9.20e-13 | 33363 | 929 | 32434 | 6.10e-04 | 0.3408 | Open wounds of extremities | injuries & poisonings | UricA | |
| 871.1 | | | | | 23768 | 162 | 23606 | | | | | 32546 | 112 | 32434 | | | Open wound of hand except finger(s) | injuries & poisonings | UricA | |
| 871.2 | | | | | 23775 | 169 | 23606 | | | | | 32574 | 140 | 32434 | | | Open wound of finger(s) | injuries & poisonings | UricA | |
| 871.3 | | | | | 23720 | 114 | 23606 | | | | | 32510 | 76 | 32434 | | | Open wound of foot except toe(s) alone | injuries & poisonings | UricA | |
| 871.4 | | | | | 23638 | 32 | 23606 | | | | | 32453 | 19 | 32434 | | | Open wound of toe(s) | injuries & poisonings | UricA | |
| 872 | | | | | 23730 | 124 | 23606 | | | | | 32497 | 63 | 32434 | | | Traumatic amputation | injuries & poisonings | UricA | |
| 874 | | | | | 23727 | 121 | 23606 | | | | | 32492 | 58 | 32434 | | | Complication of amputation stump | injuries & poisonings | UricA | |
| 875 | | | | | 23705 | 99 | 23606 | | | | | 32557 | 123 | 32434 | | | Non-healing surgical wound | injuries & poisonings | UricA | |

| phenotype | male_beta | male_SE | male_OR | male_p | male_n_total | male_n_cases | male_n_controls | female_beta | female_SE | female_OR | female_p | female_n_total | female_n_cases | female_n_controls | interaction_p | interaction_beta | description | group | lab | label |
|---|---|---|---|---|---|---|---|---|---|---|---|---|---|---|---|---|---|---|---|---|
| 876 | | | | | 23630 | 24 | 23606 | | | | | 32475 | 41 | 32434 | | | Posttraumatic wound infection not elsewhere classified | injuries & poisonings | UricA | |
| 905 | | | | | 27300 | 4 | 27296 | | | | | 36496 | 6 | 36490 | | | Late effects of musculoskeletal and connective tissue injuries | injuries & poisonings | UricA | |
| 907 | | | | | 27494 | 198 | 27296 | | | | | 36638 | 148 | 36490 | | | Injuries to the nervous system | injuries & poisonings | UricA | |
| 910 | | | | | 27755 | 6 | 27749 | | | | | 36903 | 8 | 36895 | | | Superficial injury, infected | injuries & poisonings | UricA | |
| 911 | | | | | 27688 | 32 | 27656 | | | | | 36846 | 32 | 36814 | | | Blister | injuries & poisonings | UricA | |
| 912 | | | | | 27054 | 122 | 26932 | | | | | 35677 | 201 | 35476 | | | Insect bite | injuries & poisonings | UricA | |
| 913 | | | | | 26968 | 36 | 26932 | | | | | 35544 | 68 | 35476 | | | Toxic effect of venom | injuries & poisonings | UricA | |
| 916 | 0.647 | 0.09043 | 1.91 | 8.39e-13 | 26060 | 586 | 25474 | 0.4612 | 0.07065 | 1.586 | 6.72e-11 | 34442 | 1043 | 33399 | 0.9487 | -0.007082 | Contusion | injuries & poisonings | UricA | |
| 930 | | | | | 22109 | 280 | 21829 | 0.3998 | 0.09421 | 1.492 | 2.20e-05 | 27961 | 619 | 27342 | 0.5586 | 0.09285 | Allergic reaction to food | injuries & poisonings | UricA | |
| 931 | | | | | 21895 | 66 | 21829 | | | | | 27431 | 89 | 27342 | | | Contact dermatitis and other eczema due to plants [except food] | dermatologic | UricA | |
| 938 | 0.4338 | 0.07348 | 1.543 | 3.55e-09 | 22767 | 938 | 21829 | 0.1203 | 0.08494 | 1.128 | 0.1566 | 28152 | 810 | 27342 | 0.008743 | -0.2868 | Dermatitis due to solar radiation | dermatologic | UricA | |
| 938.1 | | | | | 21840 | 11 | 21829 | | | | | 27356 | 14 | 27342 | | | Acute dermatitis due to solar radiation | dermatologic | UricA | |
| 938.2 | 0.4335 | 0.07776 | 1.543 | 2.48e-08 | 22665 | 836 | 21829 | 0.1417 | 0.08886 | 1.152 | 0.1107 | 28065 | 723 | 27342 | 0.009007 | -0.3007 | Chronic dermatitis due to solar radiation | dermatologic | UricA | |
| 939 | 0.5188 | 0.06124 | 1.68 | 2.44e-17 | 23219 | 1390 | 21829 | 0.3997 | 0.05273 | 1.491 | 3.43e-14 | 29539 | 2197 | 27342 | 0.08323 | -0.1361 | Atopic/contact dermatitis due to other or unspecified | dermatologic | UricA | |
| 939.1 | | | | | 21831 | 2 | 21829 | | | | | 27354 | 12 | 27342 | | | Contact and allergic dermatitis of eyelid | dermatologic | UricA | |
| 941 | | | | | 21871 | 42 | 21829 | | | | | 27432 | 90 | 27342 | | | Adverse reaction to serum or vaccine | injuries & poisonings | UricA | |
| 942 | | | | | 21868 | 39 | 21829 | | | | | 27392 | 50 | 27342 | | | Infusion and transfusion reaction | injuries & poisonings | UricA | |
| 946 | | | | | 21895 | 66 | 21829 | | | | | 27494 | 152 | 27342 | | | Anaphylactic shock NOS | injuries & poisonings | UricA | |
| 947 | | | | | 21986 | 157 | 21829 | | | | | 27830 | 488 | 27342 | 0.5969 | -0.1052 | Urticaria | dermatologic | UricA | |
| 949 | 0.5369 | 0.09676 | 1.711 | 2.89e-08 | 22350 | 521 | 21829 | 0.5742 | 0.06674 | 1.776 | 7.71e-18 | 28540 | 1198 | 27342 | 0.6773 | 0.04718 | Allergies, other | injuries & poisonings | UricA | |
| 949.1 | | | | | 21836 | 7 | 21829 | | | | | 27350 | 8 | 27342 | | | Diaper or napkin rash | injuries & poisonings | UricA | |

| phenotype | male_beta | male_SE | male_OR | male_p | male_n_total | male_n_cases | male_n_controls | female_beta | female_SE | female_OR | female_p | female_n_total | female_n_cases | female_n_controls | interaction_p | interaction_beta | description | group | lab | label |
|---|---|---|---|---|---|---|---|---|---|---|---|---|---|---|---|---|---|---|---|---|
| 952 | | | | | 27725 | 66 | 27659 | | | | | 36923 | 32 | 36891 | | | Spinal cord injury without evidence of spinal bone injury | injuries & poisonings | UricA | |
| 957 | | | | | 27659 | | | | | | | 36893 | 2 | 36891 | | | Injury to other and unspecified nerves | injuries & poisonings | UricA | |
| 958 | | | | | 27212 | 374 | 26838 | | | | | 36518 | 232 | 36286 | 0.01764 | 0.4839 | Certain early complications of trauma or procedure | injuries & poisonings | UricA | |
| 958.1 | | | | | 26905 | 67 | 26838 | | | | | 36331 | 45 | 36286 | | | Postoperative shock | injuries & poisonings | UricA | |
| 958.2 | | | | | 26974 | 136 | 26838 | | | | | 36364 | 78 | 36286 | | | Traumatic and surgical subcutaneous emphysema | injuries & poisonings | UricA | |
| 960 | 0.9787 | 0.06424 | 2.661 | 2.04e-52 | 21000 | 1222 | 19778 | 0.8862 | 0.05099 | 2.426 | 1.19e-67 | 28211 | 2139 | 26072 | 0.6551 | 0.03552 | Poisoning by antibiotics | injuries & poisonings | UricA | |
| 960.1 | | | | | 19779 | 1 | 19778 | | | | | 26073 | 1 | 26072 | | | Adverse effects of antibacterials (not penicillins) | injuries & poisonings | UricA | |

| phenotype | male_beta | male_SE | male_OR | male_p | male_n_total | male_n_cases | male_n_controls | female_beta | female_SE | female_OR | female_p | female_n_total | female_n_cases | female_n_controls | interaction_p | interaction_beta | description | group | lab | label |
|---|---|---|---|---|---|---|---|---|---|---|---|---|---|---|---|---|---|---|---|---|
| 960.2 | 0.9801 | 0.0827 | 2.665 | 2.09e-32 | 20476 | 698 | 19778 | 0.8853 | 0.0655 | 2.424 | 1.25e-41 | 27268 | 1196 | 26072 | 0.7221 | 0.03629 | Allergy/adverse effect of penicillin | injuries & poisonings | UricA | |
| 960.3 | | | | | 19779 | 1 | 19778 | | | | | 26073 | 1 | 26072 | | | Poisoning by antifungal antibiotics | injuries & poisonings | UricA | |
| 961 | | | | | 19843 | 65 | 19778 | | | | | 26198 | 126 | 26072 | | | Poisoning by other anti-infectives | injuries & poisonings | UricA | |
| 961.1 | | | | | 20163 | 385 | 19778 | 0.8453 | 0.06897 | 2.329 | 1.56e-34 | 27149 | 1077 | 26072 | 0.2397 | 0.1477 | Poisoning/allergy of sulfonamides | injuries & poisonings | UricA | |
| 962 | | | | | 19885 | 107 | 19778 | | | | | 26193 | 121 | 26072 | | | Poisoning by hormones and synthetic substitutes | injuries & poisonings | UricA | |
| 962.1 | | | | | 19862 | 84 | 19778 | | | | | 26166 | 94 | 26072 | | | Adrenal cortical steroids causing adverse effects in therapeutic use | injuries & poisonings | UricA | |
| 962.2 | | | | | 19800 | 22 | 19778 | | | | | 26094 | 22 | 26072 | | | Insulins and antidiabetic agents causing adverse effects in therapeutic use | injuries & poisonings | UricA | |
| 962.3 | | | | | 19778 | | | | | | | 26078 | 6 | 26072 | | | Hormones and synthetic substitutes causing adverse effects in therapeutic use | injuries & poisonings | UricA | |
| 963 | 1.473 | 0.08991 | 4.362 | 2.56e-60 | 20349 | 571 | 19778 | 1.447 | 0.09008 | 4.249 | 4.99e-58 | 26631 | 559 | 26072 | 0.4798 | 0.08745 | Poisoning by primarily systemic agents | injuries & poisonings | UricA | |
| 963.1 | 1.472 | 0.09056 | 4.357 | 2.19e-59 | 20340 | 562 | 19778 | 1.479 | 0.09098 | 4.387 | 2.13e-59 | 26617 | 545 | 26072 | 0.3553 | 0.1154 | Antineoplastic and immunosuppressive drugs causing adverse effects | injuries & poisonings | UricA | |
| 964 | | | | | 19833 | 55 | 19778 | | | | | 26136 | 64 | 26072 | | | Poisoning by agents primarily affecting blood constituents | injuries & poisonings | UricA | |
| 964.1 | | | | | 19833 | 55 | 19778 | | | | | 26131 | 59 | 26072 | | | Anticoagulants causing adverse effects | injuries & poisonings | UricA | |
| 965 | | | | | 20187 | 409 | 19778 | 0.9992 | 0.07725 | 2.716 | 2.84e-38 | 26882 | 810 | 26072 | 0.4332 | -0.09921 | Poisoning by analgesics, antipyretics, and antirheumatics | injuries & poisonings | UricA | |
| 965.1 | 0.9543 | 0.09093 | 2.597 | 9.14e-26 | 20350 | 572 | 19778 | 0.8566 | 0.06308 | 2.355 | 5.34e-42 | 27386 | 1314 | 26072 | 0.9625 | 0.005031 | Opiates and related narcotics causing adverse effects in therapeutic use | injuries & poisonings | UricA | |
| 965.2 | | | | | 19780 | 2 | 19778 | | | | | 26073 | 1 | 26072 | | | Antirheumatics causing adverse effects in therapeutic use | injuries & poisonings | UricA | |
| 965.3 | | | | | 19783 | 5 | 19778 | | | | | 26085 | 13 | 26072 | | | Salicylates causing adverse effects in therapeutic use | injuries & poisonings | UricA | |
| 966 | | | | | 19797 | 19 | 19778 | | | | | 26089 | 17 | 26072 | | | Poisoning by anticonvulsants and anti-Parkinsonism drugs | injuries & poisonings | UricA | |
| 967 | | | | | 19831 | 53 | 19778 | | | | | 26222 | 150 | 26072 | | | Adverse effects of sedatives or other central nervous system depressants and anesthetics | injuries & poisonings | UricA | |
| 969 | | | | | 19838 | 60 | 19778 | | | | | 26169 | 97 | 26072 | | | Poisoning by psychotropic agents | injuries & poisonings | UricA | |
| 971 | | | | | 19780 | 2 | 19778 | | | | | 26075 | 3 | 26072 | | | Poisoning by drugs primarily affecting the autonomic nervous system | injuries & poisonings | UricA | |
| 972 | | | | | 19849 | 71 | 19778 | | | | | 26139 | 67 | 26072 | | | Poisoning by agents primarily affecting the cardiovascular system | injuries & poisonings | UricA | |
| 972.1 | | | | | 19792 | 14 | 19778 | | | | | 26083 | 11 | 26072 | | | Cardiac rhythm regulators causing adverse effects in therapeutic use | injuries & poisonings | UricA | |
| 972.2 | | | | | 19778 | | | | | | | 26072 | | | | | Antilipemic and antiarteriosclerotic drugs causing adverse effects in therapeutic use | injuries & poisonings | UricA | |
| 972.6 | | | | | 19783 | 5 | 19778 | | | | | 26077 | 5 | 26072 | | | Antihypertensive agents causing adverse effects | injuries & poisonings | UricA | |
| 973 | | | | | 19785 | 7 | 19778 | | | | | 26074 | 2 | 26072 | | | Poisoning by agents primarily affecting the gastrointestinal system | injuries & poisonings | UricA | |

| phenotype | male_beta | male_SE | male_OR | male_p | male_n_total | male_n_cases | male_n_controls | female_beta | female_SE | female_OR | female_p | female_n_total | female_n_cases | female_n_controls | interaction_p | interaction_beta | description | group | lab | label |
|---|---|---|---|---|---|---|---|---|---|---|---|---|---|---|---|---|---|---|---|---|
| 974 | | | | | 19842 | 64 | 19778 | | | | | 26094 | 22 | 26072 | | | Poisoning by water, mineral, and uric acid metabolism drugs | injuries & poisonings | UricA | |
| 975 | | | | | 19784 | 6 | 19778 | | | | | 26079 | 7 | 26072 | | | Poisoning by agents primarily acting on the smooth and skeletal muscles and respiratory system | injuries & poisonings | UricA | |
| 976 | | | | | 19780 | 2 | 19778 | | | | | 26075 | 3 | 26072 | | | Poisoning by agents primarily affecting skin & mucous membrane, ophthalmological, otorhinolaryngological, & dental drugs | injuries & poisonings | UricA | |
| 977 | 1.055 | 0.0605 | 2.873 | 3.88e-68 | 21166 | 1388 | 19778 | 0.9952 | 0.04992 | 2.705 | 2.09e-88 | 28281 | 2209 | 26072 | 0.94 | 0.005749 | Personal history of allergy to medicinal agents | injuries & poisonings | UricA | |
| 979 | 0.8055 | 0.0755 | 2.238 | 1.43e-26 | 20676 | 898 | 19778 | 0.9033 | 0.07394 | 2.468 | 2.51e-34 | 27006 | 934 | 26072 | 0.01584 | 0.247 | Adverse drug events and drug allergies | injuries & poisonings | UricA | |
| 980 | 0.7557 | 0.06474 | 2.129 | 1.77e-31 | 26597 | 1176 | 25421 | 0.9973 | 0.06836 | 2.711 | 3.33e-48 | 35862 | 1000 | 34862 | 4.10e-04 | 0.321 | Encounter for long-term (current) use of antibiotics | infectious diseases | UricA | |
| 981 | | | | | 27615 | 2 | 27613 | | | | | 36741 | 3 | 36738 | | | Toxic effect of (non-ethyl) alcohol and petroleum and other solvents | injuries & poisonings | UricA | |
| 983 | | | | | 27616 | 3 | 27613 | | | | | 36738 | | | | | Toxic effect of corrosive aromatics, acids, and caustic alkalis | injuries & poisonings | UricA | |
| 984 | | | | | 27616 | 3 | 27613 | | | | | 36739 | 1 | 36738 | | | Toxic effect of lead and its compounds (including fumes) | injuries & poisonings | UricA | |
| 985 | | | | | 27622 | 9 | 27613 | | | | | 36745 | 7 | 36738 | | | Toxic effect of other metals | injuries & poisonings | UricA | |
| 986 | | | | | 27616 | 3 | 27613 | | | | | 36740 | 2 | 36738 | | | Toxic effect of carbon monoxide | injuries & poisonings | UricA | |
| 987 | | | | | 27617 | 4 | 27613 | | | | | 36745 | 7 | 36738 | | | Toxic effect of other gases, fumes, or vapors | injuries & poisonings | UricA | |
| 988 | | | | | 27613 | | | | | | | 36740 | 2 | 36738 | | | Toxic effect of noxious substances eaten as food | injuries & poisonings | UricA | |
| 989 | | | | | 27632 | 19 | 27613 | | | | | 36758 | 20 | 36738 | | | Toxic effect of other substances, chiefly nonmedicinal as to source | injuries & poisonings | UricA | |
| 990 | 0.3667 | 0.07805 | 1.443 | 2.62e-06 | 23446 | 892 | 22554 | 0.3318 | 0.0881 | 1.393 | 1.66e-04 | 33242 | 782 | 32460 | 0.9896 | 0.001488 | Effects radiation NOS | injuries & poisonings | UricA | |
| 994 | 0.7363 | 0.04523 | 2.088 | 1.35e-59 | 26534 | 2690 | 23844 | 0.9267 | 0.04973 | 2.526 | 1.74e-77 | 35853 | 2103 | 33750 | 1.01e-04 | 0.2527 | Sepsis and SIRS | injuries & poisonings | UricA | |
| 994.1 | 0.5464 | 0.09804 | 1.727 | 2.51e-08 | 24423 | 579 | 23844 | | | | | 34199 | 449 | 33750 | 0.001597 | 0.4417 | Systemic inflammatory response syndrome (SIRS) | injuries & poisonings | UricA | |
| 994.2 | 0.7745 | 0.04758 | 2.17 | 1.39e-59 | 26190 | 2346 | 23844 | 0.9888 | 0.05201 | 2.688 | 1.34e-80 | 35609 | 1859 | 33750 | 5.87e-05 | 0.2737 | Sepsis | injuries & poisonings | UricA | |
| 994.21 | 0.9078 | 0.07354 | 2.479 | 5.19e-35 | 24740 | 896 | 23844 | 1.091 | 0.0835 | 2.977 | 5.05e-39 | 34424 | 674 | 33750 | 0.009987 | 0.2769 | Septic shock | injuries & poisonings | UricA | |
| 008 | 1.021 | 0.124 | 2.775 | 1.87e-16 | 27060 | 574 | 26486 | 1.387 | 0.1079 | 4.002 | 8.02e-38 | 35831 | 704 | 35127 | 0.004915 | 0.4311 | Intestinal infection | infectious diseases | RATE_C | |
| 008.5 | | | | | 26875 | 389 | 26486 | | | | | 35533 | 406 | 35127 | 0.001259 | 0.7057 | Bacterial enteritis | infectious diseases | RATE_C | |
| 008.51 | | | | | 26487 | 1 | 26486 | | | | | 35130 | 3 | 35127 | | | Intestinal e.coli | infectious diseases | RATE_C | |
| 008.52 | | | | | 26824 | 338 | 26486 | | | | | 35475 | 348 | 35127 | 0.01068 | 0.6502 | Intestinal infection due to C. difficile | infectious diseases | RATE_C | |
| 008.6 | | | | | 26592 | 106 | 26486 | | | | | 35263 | 136 | 35127 | | | Viral Enteritis | infectious diseases | RATE_C | |
| 008.7 | | | | | 26495 | 9 | 26486 | | | | | 35133 | 6 | 35127 | | | Intestinal infection due to protozoa | infectious diseases | RATE_C | |
| 010 | | | | | 21692 | 51 | 21641 | | | | | 30214 | 54 | 30160 | | | Tuberculosis | infectious diseases | RATE_C | |
| 031 | | | | | 21701 | 60 | 21641 | | | | | 30222 | 62 | 30160 | | | Diseases due to other mycobacteria | infectious diseases | RATE_C | |

| phenotype | male_beta | male_SE | male_OR | male_p | male_n_total | male_n_cases | male_n_controls | female_beta | female_SE | female_OR | female_p | female_n_total | female_n_cases | female_n_controls | interaction_p | interaction_beta | description | group | lab | label |
|---|---|---|---|---|---|---|---|---|---|---|---|---|---|---|---|---|---|---|---|---|
| 031.1 | | | | | 21641 | | | | | | | 30160 | | | | | Leprosy | infectious diseases | RATE_C | |
| phenotype | male_beta | male_SE | male_OR | male_p | male_n_total | male_n_cases | male_n_controls | female_beta | female_SE | female_OR | female_p | female_n_total | female_n_cases | female_n_controls | interaction_p | interaction_beta | description | group | lab | label |
| 038 | 1.809 | 0.06899 | 6.102 | 1.78e-151 | 24701 | 3060 | 21641 | 1.828 | 0.0652 | 6.221 | 5.58e-173 | 32656 | 2496 | 30160 | 0.329 | 0.08875 | Septicemia | infectious diseases | RATE_C | |
| 038.1 | | | | | 22136 | 495 | 21641 | | | | | 30550 | 390 | 30160 | 0.5698 | 0.146 | Gram negative septicemia | infectious diseases | RATE_C | |
| 038.2 | | | | | 22055 | 414 | 21641 | | | | | 30433 | 273 | 30160 | 0.04182 | 0.5362 | Gram positive septicemia | infectious diseases | RATE_C | |
| 038.3 | 1.944 | 0.1052 | 6.984 | 3.36e-76 | 23021 | 1380 | 21641 | 1.985 | 0.1056 | 7.277 | 7.88e-79 | 31182 | 1022 | 30160 | 0.3698 | 0.1284 | Bacteremia | infectious diseases | RATE_C | |
| 041 | 1.398 | 0.06061 | 4.049 | 8.72e-118 | 24639 | 2998 | 21641 | 1.485 | 0.05292 | 4.413 | 3.83e-173 | 33323 | 3163 | 30160 | 0.008358 | 0.1998 | Bacterial infection NOS | infectious diseases | RATE_C | |
| 041.1 | 1.444 | 0.08561 | 4.239 | 7.20e-64 | 22993 | 1352 | 21641 | 1.703 | 0.08905 | 5.491 | 1.56e-81 | 31259 | 1099 | 30160 | 0.002896 | 0.345 | Staphylococcus infections | infectious diseases | RATE_C | |
| 041.11 | 1.284 | 0.1199 | 3.61 | 9.94e-27 | 22231 | 590 | 21641 | | | | | 30598 | 438 | 30160 | 8.61e-04 | 0.5765 | Methicillin sensitive Staphylococcus aureus | infectious diseases | RATE_C | |
| 041.12 | 1.682 | 0.1389 | 5.375 | 9.49e-34 | 22233 | 592 | 21641 | | | | | 30654 | 494 | 30160 | 0.3553 | 0.1697 | Methicillin resistant Staphylococcus aureus | infectious diseases | RATE_C | |
| 041.2 | 1.546 | 0.1304 | 4.691 | 2.10e-32 | 22331 | 690 | 21641 | 1.772 | 0.1225 | 5.881 | 1.95e-47 | 30829 | 669 | 30160 | 0.08317 | 0.2956 | Streptococcus infection | infectious diseases | RATE_C | |
| 041.21 | | | | | 21646 | 5 | 21641 | | | | | 30170 | 10 | 30160 | | | Rheumatic fever / chorea | infectious diseases | RATE_C | |
| 041.4 | | | | | 22126 | 485 | 21641 | 1.607 | 0.1059 | 4.99 | 5.09e-52 | 31066 | 906 | 30160 | 0.8867 | 0.02768 | E. coli | infectious diseases | RATE_C | |
| 041.8 | | | | | 21691 | 50 | 21641 | | | | | 30238 | 78 | 30160 | | | H. pylori | infectious diseases | RATE_C | |
| 041.9 | | | | | 21990 | 349 | 21641 | | | | | 30526 | 366 | 30160 | 0.008832 | 0.5747 | Infection with drug-resistant microorganisms | infectious diseases | RATE_C | |
| 053 | | | | | 22267 | 352 | 21915 | 0.6441 | 0.1163 | 1.904 | 3.03e-08 | 29621 | 557 | 29064 | 0.0688 | 0.337 | Herpes zoster | infectious diseases | RATE_C | |
| 053.1 | | | | | 21986 | 71 | 21915 | | | | | 29192 | 128 | 29064 | | | Herpes zoster with nervous system complications | infectious diseases | RATE_C | |
| 054 | | | | | 22206 | 291 | 21915 | 0.347 | 0.09727 | 1.415 | 3.60e-04 | 29622 | 558 | 29064 | 0.376 | 0.1382 | Herpes simplex | infectious diseases | RATE_C | |
| 070 | 0.5018 | 0.08109 | 1.652 | 6.09e-10 | 22942 | 1027 | 21915 | 0.6775 | 0.09714 | 1.969 | 3.06e-12 | 29665 | 601 | 29064 | 0.05575 | 0.2223 | Viral hepatitis | infectious diseases | RATE_C | |
| 070.1 | | | | | 21931 | 16 | 21915 | | | | | 29069 | 5 | 29064 | | | Viral hepatitis A | infectious diseases | RATE_C | |
| 070.2 | | | | | 22087 | 172 | 21915 | | | | | 29138 | 74 | 29064 | | | Viral hepatitis B | infectious diseases | RATE_C | |
| 070.3 | 0.5495 | 0.08738 | 1.732 | 3.18e-10 | 22806 | 891 | 21915 | 0.6408 | 0.1038 | 1.898 | 6.70e-10 | 29579 | 515 | 29064 | 0.2784 | 0.135 | Viral hepatitis C | infectious diseases | RATE_C | |
| 070.4 | | | | | 22042 | 127 | 21915 | | | | | 29294 | 230 | 29064 | | | Chronic hepatitis | infectious diseases | RATE_C | |
| 070.9 | | | | | 22131 | 216 | 21915 | | | | | 29270 | 206 | 29064 | | | Hepatitis NOS | infectious diseases | RATE_C | |
| 071 | -0.5829 | 0.09302 | 0.5583 | 3.70e-10 | 22529 | 614 | 21915 | | | | | 29177 | 113 | 29064 | 2.38e-07 | 1.111 | Human immunodeficiency virus [HIV] disease | infectious diseases | RATE_C | |
| 071.1 | -0.5968 | 0.09361 | 0.5505 | 1.82e-10 | 22521 | 606 | 21915 | | | | | 29170 | 106 | 29064 | 2.81e-06 | 1.023 | HIV infection, symptomatic | infectious diseases | RATE_C | |
| 078 | -0.06928 | 0.09146 | 0.9331 | 0.4488 | 22638 | 723 | 21915 | 0.176 | 0.08211 | 1.192 | 0.03212 | 29855 | 791 | 29064 | 0.009757 | 0.2847 | Viral warts & HPV | infectious diseases | RATE_C | |
| 079 | 0.59 | 0.09872 | 1.804 | 2.28e-09 | 22615 | 700 | 21915 | 0.7122 | 0.07648 | 2.039 | 1.25e-20 | 30057 | 993 | 29064 | 0.3041 | 0.1134 | Viral infection | infectious diseases | RATE_C | |
| 079.1 | | | | | 21924 | 9 | 21915 | | | | | 29075 | 11 | 29064 | | | Varicella infection | infectious diseases | RATE_C | |

| phenotype | male_beta | male_SE | male_OR | male_p | male_n_total | male_n_cases | male_n_controls | female_beta | female_SE | female_OR | female_p | female_n_total | female_n_cases | female_n_controls | interaction_p | interaction_beta | description | group | lab | label |
|---|---|---|---|---|---|---|---|---|---|---|---|---|---|---|---|---|---|---|---|---|
| 079.2 | | | | | 21970 | 55 | 21915 | | | | | 29142 | 78 | 29064 | | | Infectious mononucleosis | infectious diseases | RATE_C | |
| 079.9 | | | | | 21977 | 62 | 21915 | | | | | 29164 | 100 | 29064 | | | Viremia, NOS | infectious diseases | RATE_C | |
| 080 | 1.284 | 0.1162 | 3.61 | 2.35e-28 | 26108 | 681 | 25427 | 1.67 | 0.1092 | 5.312 | 8.88e-53 | 35329 | 748 | 34581 | 6.37e-04 | 0.5106 | Postoperative infection | infectious diseases | RATE_C | |
| 081 | 1.328 | 0.1119 | 3.774 | 1.79e-32 | 26193 | 766 | 25427 | 1.687 | 0.1153 | 5.402 | 1.68e-48 | 35303 | 722 | 34581 | 1.71e-04 | 0.5681 | Infection/inflammation of internal prosthetic device; implant; and graft | infectious diseases | RATE_C | |
| 081.1 | | | | | 25468 | 41 | 25427 | | | | | 34621 | 40 | 34581 | | | Graft-versus-host disease | infectious diseases | RATE_C | |
| 081.11 | | | | | 25439 | 12 | 25427 | | | | | 34597 | 16 | 34581 | | | Acute graft-versus-host disease | infectious diseases | RATE_C | |
| 081.12 | | | | | 25458 | 31 | 25427 | | | | | 34611 | 30 | 34581 | | | Chronic graft-versus-host disease | infectious diseases | RATE_C | |
| 090 | | | | | 27678 | 201 | 27477 | | | | | 36786 | 64 | 36722 | | | Sexually transmitted infections (not HIV or hepatitis) | infectious diseases | RATE_C | |
| 090.2 | | | | | 27533 | 56 | 27477 | | | | | 36735 | 13 | 36722 | | | Gonococcal infections | infectious diseases | RATE_C | |
| 090.3 | | | | | 27479 | 2 | 27477 | | | | | 36731 | 9 | 36722 | | | Venereal diseases due to Chlamydia trachomatis | infectious diseases | RATE_C | |
| 110 | 0.3807 | 0.07993 | 1.463 | 1.90e-06 | 24822 | 1390 | 23432 | 0.6351 | 0.07714 | 1.887 | 1.83e-16 | 31711 | 1224 | 30487 | 0.01718 | 0.2515 | Dermatophytosis / Dermatomycosis | infectious diseases | RATE_C | |
| 110.1 | 0.3887 | 0.08321 | 1.475 | 2.99e-06 | 24733 | 1301 | 23432 | 0.6628 | 0.08081 | 1.94 | 2.36e-16 | 31638 | 1151 | 30487 | 0.01114 | 0.2808 | Dermatophytosis | infectious diseases | RATE_C | |
| 110.11 | 0.4796 | 0.09986 | 1.615 | 1.56e-06 | 24428 | 996 | 23432 | 0.6238 | 0.0914 | 1.866 | 8.79e-12 | 31407 | 920 | 30487 | 0.2837 | 0.1398 | Dermatophytosis of nail | infectious diseases | RATE_C | |
| 110.12 | | | | | 23675 | 243 | 23432 | | | | | 30615 | 128 | 30487 | | | Althete's foot | infectious diseases | RATE_C | |
| 110.13 | | | | | 23580 | 148 | 23432 | | | | | 30581 | 94 | 30487 | | | Dermatophytosis of the body | infectious diseases | RATE_C | |
| 110.2 | | | | | 23511 | 79 | 23432 | | | | | 30547 | 60 | 30487 | | | Dermatomycoses | infectious diseases | RATE_C | |
| 112 | 1.261 | 0.1113 | 3.529 | 8.87e-30 | 24237 | 805 | 23432 | 1.106 | 0.0664 | 3.022 | 2.85e-62 | 32100 | 1613 | 30487 | 0.5229 | -0.07742 | Candidiasis | infectious diseases | RATE_C | |
| 112.3 | | | | | 23511 | 79 | 23432 | | | | | 30667 | 180 | 30487 | | | Candidiasis of skin and nails | infectious diseases | RATE_C | |
| 117 | | | | | 23791 | 359 | 23432 | | | | | 30831 | 344 | 30487 | 0.4223 | 0.1709 | Mycoses | infectious diseases | RATE_C | |
| 117.1 | | | | | 23530 | 98 | 23432 | | | | | 30601 | 114 | 30487 | | | Histoplasmosis | infectious diseases | RATE_C | |
| 117.2 | | | | | 23434 | 2 | 23432 | | | | | 30487 | | | | | Coccidioidomycosis | infectious diseases | RATE_C | |
| 117.3 | | | | | 23448 | 16 | 23432 | | | | | 30500 | 13 | 30487 | | | Blastomycotic infection | infectious diseases | RATE_C | |
| 117.4 | | | | | 23501 | 69 | 23432 | | | | | 30544 | 57 | 30487 | | | Aspergillosis | infectious diseases | RATE_C | |
| 130 | | | | | 26354 | 10 | 26344 | | | | | 35233 | 9 | 35224 | | | Spirochetal infection | infectious diseases | RATE_C | |
| 130.1 | | | | | 26349 | 5 | 26344 | | | | | 35227 | 3 | 35224 | | | Lyme disease | infectious diseases | RATE_C | |
| 131 | | | | | 26353 | 9 | 26344 | | | | | 35256 | 32 | 35224 | | | Protozoan infection | infectious diseases | RATE_C | |
| 132 | | | | | 26380 | 36 | 26344 | | | | | 35274 | 50 | 35224 | | | Infestation (lice, mites) | infectious diseases | RATE_C | |
| 132.1 | | | | | 26348 | 4 | 26344 | | | | | 35245 | 21 | 35224 | | | Pediculosis and phthirus infestation | infectious diseases | RATE_C | |
| 133 | | | | | 26352 | 8 | 26344 | | | | | 35226 | 2 | 35224 | | | Arthropod-borne diseases | infectious diseases | RATE_C | |

| phenotype | male_beta | male_SE | male_OR | male_p | male_n_total | male_n_cases | male_n_controls | female_beta | female_SE | female_OR | female_p | female_n_total | female_n_cases | female_n_controls | interaction_p | interaction_beta | description | group | lab | label |
|---|---|---|---|---|---|---|---|---|---|---|---|---|---|---|---|---|---|---|---|---|
| 134 | | | | | 26349 | 5 | 26344 | | | | | 35248 | 24 | 35224 | | | Helminthiases | infectious diseases | RATE_C | |
| 134.1 | | | | | 26347 | 3 | 26344 | | | | | 35236 | 12 | 35224 | | | Intestinal helminthiases | infectious diseases | RATE_C | |
| 136 | | | | | 26733 | 389 | 26344 | | | | | 35599 | 375 | 35224 | 0.4273 | 0.1532 | Other infectious and parasitic diseases | infectious diseases | RATE_C | |
| 145 | 0.4507 | 0.1075 | 1.569 | 2.76e-05 | 26903 | 624 | 26279 | | | | | 36280 | 243 | 36037 | 0.003491 | 0.5766 | Cancer of mouth | neoplasms | RATE_C | |
| 145.1 | | | | | 26299 | 20 | 26279 | | | | | 36044 | 7 | 36037 | | | Cancer of lip | neoplasms | RATE_C | |
| 145.2 | | | | | 26660 | 381 | 26279 | | | | | 36136 | 99 | 36037 | | | Cancer of tongue | neoplasms | RATE_C | |
| 145.3 | | | | | 26384 | 105 | 26279 | | | | | 36087 | 50 | 36037 | | | Cancer of major salivary glands | neoplasms | RATE_C | |
| 145.4 | | | | | 26326 | 47 | 26279 | | | | | 36068 | 31 | 36037 | | | Cancer of the gums | neoplasms | RATE_C | |
| 145.5 | | | | | 26338 | 59 | 26279 | | | | | 36058 | 21 | 36037 | | | Cancer of the mouth floor | neoplasms | RATE_C | |
| 149 | 0.462 | 0.08918 | 1.587 | 2.22e-07 | 27197 | 918 | 26279 | | | | | 36384 | 347 | 36037 | 5.70e-04 | 0.5667 | Cancer of larynx, pharynx, nasal cavities | neoplasms | RATE_C | |
| 149.1 | | | | | 26708 | 429 | 26279 | | | | | 36147 | 110 | 36037 | 0.1432 | 0.372 | Cancer of oropharynx | neoplasms | RATE_C | |
| 149.2 | | | | | 26347 | 68 | 26279 | | | | | 36075 | 38 | 36037 | | | Cancer of nasopharynx | neoplasms | RATE_C | |
| 149.3 | | | | | 26333 | 54 | 26279 | | | | | 36064 | 27 | 36037 | | | Cancer of hypopharynx | neoplasms | RATE_C | |
| 149.4 | | | | | 26607 | 328 | 26279 | | | | | 36132 | 95 | 36037 | | | Cancer of larynx | neoplasms | RATE_C | |
| 149.5 | | | | | 26723 | 444 | 26279 | | | | | 36197 | 160 | 36037 | 0.1027 | 0.3859 | Hx of malignant neoplasm of oral cavity and pharynx | neoplasms | RATE_C | |
| 149.9 | | | | | 26377 | 98 | 26279 | | | | | 36109 | 72 | 36037 | | | Cancer of of nasal cavities | neoplasms | RATE_C | |
| 150 | | | | | 23056 | 399 | 22657 | | | | | 32735 | 70 | 32665 | | | Cancer of esophagus | neoplasms | RATE_C | |
| 151 | | | | | 23072 | 415 | 22657 | | | | | 32805 | 140 | 32665 | 0.002704 | 0.7767 | Cancer of stomach | neoplasms | RATE_C | |
| 153 | 0.2737 | 0.0702 | 1.315 | 9.64e-05 | 21472 | 1270 | 20202 | 0.7195 | 0.08091 | 2.053 | 5.97e-19 | 30905 | 948 | 29957 | 4.84e-09 | 0.5916 | Colorectal cancer | neoplasms | RATE_C | |
| 153.2 | 0.6557 | 0.09141 | 1.927 | 7.34e-13 | 21102 | 900 | 20202 | 0.7208 | 0.08982 | 2.056 | 1.02e-15 | 30731 | 774 | 29957 | 0.1777 | 0.1647 | Colon cancer | neoplasms | RATE_C | |
| 153.3 | 0.2162 | 0.08888 | 1.241 | 0.01499 | 20940 | 738 | 20202 | | | | | 30403 | 446 | 29957 | 4.16e-10 | 0.8759 | Malignant neoplasm of rectum, rectosigmoid junction, and anus | neoplasms | RATE_C | |
| 155 | | | | | 23097 | 440 | 22657 | | | | | 32892 | 227 | 32665 | 0.1292 | 0.3303 | Cancer of liver and intrahepatic bile duct | neoplasms | RATE_C | |
| 155.1 | | | | | 23033 | 376 | 22657 | | | | | 32838 | 173 | 32665 | 0.2534 | 0.2851 | Malignant neoplasm of liver, primary | neoplasms | RATE_C | |
| 157 | | | | | 23046 | 389 | 22657 | | | | | 32990 | 325 | 32665 | 0.218 | 0.244 | Pancreatic cancer | neoplasms | RATE_C | |
| 158 | | | | | 22882 | 225 | 22657 | | | | | 32904 | 239 | 32665 | | | Neoplasm of unspecified nature of digestive system | neoplasms | RATE_C | |
| 159 | 1.013 | 0.1299 | 2.754 | 6.14e-15 | 23177 | 520 | 22657 | 1.146 | 0.1229 | 3.145 | 1.11e-20 | 33167 | 502 | 32665 | 0.085 | 0.2941 | Malignant neoplasm of other and ill-defined sites within the digestive organs and peritoneum | neoplasms | RATE_C | |
| 159.2 | | | | | 22805 | 148 | 22657 | | | | | 32791 | 126 | 32665 | | | Malignant neoplasm of small intestine, including duodenum | neoplasms | RATE_C | |

| phenotype | male_beta | male_SE | male_OR | male_p | male_n_total | male_n_cases | male_n_controls | female_beta | female_SE | female_OR | female_p | female_n_total | female_n_cases | female_n_controls | interaction_p | interaction_beta | description | group | lab | label |
|---|---|---|---|---|---|---|---|---|---|---|---|---|---|---|---|---|---|---|---|---|
| 159.3 | | | | | 22758 | 101 | 22657 | | | | | 32760 | 95 | 32665 | | | Malignant neoplasm of gallbladder and extrahepatic bile ducts | neoplasms | RATE_C | |
| 159.4 | | | | | 22743 | 86 | 22657 | | | | | 32783 | 118 | 32665 | | | Malignant neoplasm of retroperitoneum and peritoneum | neoplasms | RATE_C | |
| 164 | | | | | 25778 | 71 | 25707 | | | | | 35298 | 55 | 35243 | | | Cancer of intrathoracic organs | neoplasms | RATE_C | |
| 165 | 0.6828 | 0.07324 | 1.98 | 1.12e-20 | 27233 | 1526 | 25707 | 0.8837 | 0.0739 | 2.42 | 5.85e-33 | 36502 | 1259 | 35243 | 0.105 | 0.1625 | Cancer within the respiratory system | neoplasms | RATE_C | |
| 165.1 | 0.6736 | 0.07358 | 1.961 | 5.42e-20 | 27209 | 1502 | 25707 | 0.8775 | 0.07408 | 2.405 | 2.29e-32 | 36492 | 1249 | 35243 | 0.09823 | 0.1663 | Cancer of bronchus; lung | neoplasms | RATE_C | |
| 170 | 1.06 | 0.1363 | 2.885 | 7.57e-15 | 27372 | 508 | 26864 | | | | | 36581 | 434 | 36147 | 0.3008 | 0.1877 | Cancer of bone and connective tissue | neoplasms | RATE_C | |
| 170.1 | | | | | 27106 | 242 | 26864 | | | | | 36353 | 206 | 36147 | | | Bone cancer | neoplasms | RATE_C | |
| 170.2 | | | | | 27193 | 329 | 26864 | | | | | 36433 | 286 | 36147 | 0.5905 | 0.1142 | Cancer of connective tissue | neoplasms | RATE_C | |
| 172 | 0.379 | 0.05292 | 1.461 | 7.94e-13 | 25415 | 3454 | 21961 | 0.5377 | 0.05253 | 1.712 | 1.37e-24 | 33247 | 2675 | 30572 | 0.1275 | 0.11 | Skin cancer | neoplasms | RATE_C | |
| 172.1 | 0.5839 | 0.09256 | 1.793 | 2.82e-10 | 22929 | 968 | 21961 | 0.7416 | 0.09379 | 2.099 | 2.64e-15 | 31295 | 723 | 30572 | 0.9512 | 0.007737 | Melanomas of skin, dx or hx | neoplasms | RATE_C | |
| 172.11 | 0.553 | 0.09833 | 1.738 | 1.87e-08 | 22766 | 805 | 21961 | 0.7425 | 0.1045 | 2.101 | 1.18e-12 | 31138 | 566 | 30572 | 0.6845 | 0.05579 | Melanomas of skin | neoplasms | RATE_C | |
| 172.2 | 0.3805 | 0.06031 | 1.463 | 2.80e-10 | 24804 | 2843 | 21961 | 0.4864 | 0.05982 | 1.627 | 4.22e-16 | 32709 | 2137 | 30572 | 0.3701 | 0.07402 | Other non-epithelial cancer of skin | neoplasms | RATE_C | |
| 172.21 | 0.2989 | 0.1009 | 1.348 | 0.003052 | 22933 | 972 | 21961 | 0.2997 | 0.09719 | 1.349 | 0.002048 | 31280 | 708 | 30572 | 0.4401 | -0.1047 | Basal cell carcinoma | neoplasms | RATE_C | |
| 172.22 | 0.5331 | 0.113 | 1.704 | 2.40e-06 | 22797 | 836 | 21961 | | | | | 30972 | 400 | 30572 | 0.2539 | -0.1919 | Squamous cell carcinoma | neoplasms | RATE_C | |
| 172.3 | | | | | 22359 | 398 | 21961 | | | | | 30807 | 235 | 30572 | 0.1077 | -0.4157 | Carcinoma in situ of skin | neoplasms | RATE_C | |
| 173 | 0.2474 | 0.06856 | 1.281 | 3.08e-04 | 23987 | 2026 | 21961 | 0.3111 | 0.05328 | 1.365 | 5.25e-09 | 32915 | 2343 | 30572 | 0.0888 | -0.1407 | Neoplasm of uncertain behavior of skin | neoplasms | RATE_C | |
| 174 | | | | | 27350 | 45 | 27305 | 0.5172 | 0.04582 | 1.677 | 1.54e-29 | 31827 | 3093 | 28734 | 0.6847 | 0.1594 | Breast cancer | neoplasms | RATE_C | |
| 174.1 | | | | | 7 | | | 0.517 | 0.04641 | 1.677 | 8.08e-29 | 31729 | 2998 | 28731 | 0.9979 | 0.6978 | Breast cancer [female] | neoplasms | RATE_C | |
| 174.11 | | | | | 7 | | | 0.5167 | 0.04664 | 1.676 | 1.58e-28 | 31698 | 2967 | 28731 | 0.9979 | 0.6981 | Malignant neoplasm of female breast | neoplasms | RATE_C | |
| 174.2 | | | | | 27313 | 15 | 27298 | | | | | 3 | | | | | Breast cancer [male] | neoplasms | RATE_C | |
| 174.3 | | | | | 27310 | 5 | 27305 | | | | | 28889 | 155 | 28734 | | | Neoplasm of uncertain behavior of breast | neoplasms | RATE_C | |
| 175 | | | | | 7 | | | 0.8735 | 0.07412 | 2.395 | 4.63e-32 | 27358 | 1237 | 26121 | 0.9973 | 0.9303 | Acquired absence of breast | neoplasms | RATE_C | |
| 180 | | | | | 7 | | | 0.5392 | 0.07788 | 1.715 | 4.43e-12 | 28736 | 897 | 27839 | 0.9995 | 0.2952 | Cervical cancer and dysplasia | neoplasms | RATE_C | |
| 180.1 | | | | | 7 | | | | | | | 28134 | 295 | 27839 | | | Cervical cancer | neoplasms | RATE_C | |
| 180.3 | | | | | 7 | | | 0.3405 | 0.09218 | 1.406 | 2.21e-04 | 28457 | 618 | 27839 | 1 | -0.02626 | Cervical intraepithelial neoplasia [CIN] [Cervical dysplasia] | neoplasms | RATE_C | |
| 182 | | | | | 7 | | | 1.042 | 0.1207 | 2.834 | 5.98e-18 | 29788 | 534 | 29254 | 0.9978 | 1.247 | Malignant neoplasm of uterus | neoplasms | RATE_C | |
| 184 | | | | | 7 | | | 0.7971 | 0.09116 | 2.219 | 2.24e-18 | 30877 | 771 | 30106 | 0.9983 | 0.9401 | Cancer of other female genital organs | neoplasms | RATE_C | |
| 184.1 | | | | | 7 | | | 0.9004 | 0.1083 | 2.46 | 9.33e-17 | 30679 | 573 | 30106 | 0.9981 | 1.072 | Malignant neoplasm of ovary and other uterine adnexa | neoplasms | RATE_C | |
| 184.11 | | | | | 7 | | | | | | | 30568 | 462 | 30106 | | | Malignant neoplasm of ovary | neoplasms | RATE_C | |

| phenotype | male_beta | male_SE | male_OR | male_p | male_n_total | male_n_cases | male_n_controls | female_beta | female_SE | female_OR | female_p | female_n_total | female_n_cases | female_n_controls | interaction_p | interaction_beta | description | group | lab | label |
|---|---|---|---|---|---|---|---|---|---|---|---|---|---|---|---|---|---|---|---|---|
| 184.2 | | | | | 7 | | | | | | | 30333 | 227 | 30106 | | | Cancer of other female genital organs (excluding uterus and ovary) | neoplasms | RATE_C | |
| 185 | 0.4097 | 0.06636 | 1.506 | 6.66e-10 | 21441 | 2053 | 19388 | | | | | 4 | | | 0.9919 | -3.175 | Cancer of prostate | neoplasms | RATE_C | |
| 187 | | | | | 25045 | 181 | 24864 | | | | | 4 | | | | | Cancer of other male genital organs | neoplasms | RATE_C | |
| 187.1 | | | | | 24907 | 43 | 24864 | | | | | 4 | | | | | Malignant neoplasm of unspecified male genital organ | neoplasms | RATE_C | |
| 187.2 | | | | | 24988 | 124 | 24864 | | | | | 4 | | | | | Malignant neoplasm of testis | neoplasms | RATE_C | |
| 187.8 | | | | | 24880 | 16 | 24864 | | | | | 4 | | | | | Neoplasm of uncertain behavior of male genital organs | neoplasms | RATE_C | |
| 189 | 0.7279 | 0.08399 | 2.071 | 4.46e-18 | 27414 | 1343 | 26071 | 1.183 | 0.1297 | 3.264 | 7.64e-20 | 36667 | 553 | 36114 | 0.005384 | 0.4172 | Cancer of urinary organs (incl. kidney and bladder) | neoplasms | RATE_C | |
| 189.1 | 0.8093 | 0.1124 | 2.246 | 5.95e-13 | 26801 | 730 | 26071 | | | | | 36472 | 358 | 36114 | 0.07415 | 0.3282 | Cancer of kidney and renal pelvis | neoplasms | RATE_C | |
| 189.11 | 0.8208 | 0.1194 | 2.272 | 6.35e-12 | 26706 | 635 | 26071 | | | | | 36422 | 308 | 36114 | 0.07946 | 0.3444 | Malignant neoplasm of kidney, except pelvis | neoplasms | RATE_C | |
| 189.12 | | | | | 26217 | 146 | 26071 | | | | | 36178 | 64 | 36114 | | | Malignant neoplasm of renal pelvis | neoplasms | RATE_C | |
| 189.2 | 0.751 | 0.1201 | 2.119 | 3.96e-10 | 26790 | 719 | 26071 | | | | | 36340 | 226 | 36114 | 0.007937 | 0.6972 | Cancer of bladder | neoplasms | RATE_C | |
| 189.21 | 0.7611 | 0.1214 | 2.141 | 3.59e-10 | 26780 | 709 | 26071 | | | | | 36336 | 222 | 36114 | 0.007218 | 0.7208 | Malignant neoplasm of bladder | neoplasms | RATE_C | |
| 189.4 | | | | | 26367 | 296 | 26071 | | | | | 36227 | 113 | 36114 | | | Malignant neoplasm of other urinary organs | neoplasms | RATE_C | |
| 190 | | | | | 26618 | 60 | 26558 | | | | | 35577 | 58 | 35519 | | | Cancer of eye | neoplasms | RATE_C | |
| 191 | | | | | 27023 | 465 | 26558 | | | | | 35973 | 454 | 35519 | 0.007081 | 0.4247 | Manlignant and unknown neoplasms of brain and nervous system | neoplasms | RATE_C | |
| 191.1 | | | | | 26929 | 371 | 26558 | | | | | 35854 | 335 | 35519 | 0.004805 | 0.5082 | Cancer of brain and nervous system | neoplasms | RATE_C | |
| 191.11 | | | | | 26893 | 335 | 26558 | | | | | 35801 | 282 | 35519 | 0.008576 | 0.5046 | Cancer of brain | neoplasms | RATE_C | |
| 193 | | | | | 27161 | 215 | 26946 | | | | | 35853 | 482 | 35371 | 0.2354 | -0.2454 | Thyroid cancer | neoplasms | RATE_C | |
| 194 | | | | | 26993 | 47 | 26946 | | | | | 35425 | 54 | 35371 | | | Cancer of other endocrine glands | neoplasms | RATE_C | |
| 195 | 0.8942 | 0.05944 | 2.445 | 3.81e-51 | 20735 | 2450 | 18285 | 1.292 | 0.06366 | 3.639 | 1.51e-91 | 28047 | 1903 | 26144 | 2.26e-06 | 0.3922 | Cancer, suspected or other | neoplasms | RATE_C | |
| 195.1 | 0.9661 | 0.08484 | 2.628 | 4.85e-30 | 19494 | 1209 | 18285 | 1.474 | 0.09328 | 4.365 | 3.20e-56 | 27102 | 958 | 26144 | 4.15e-05 | 0.4925 | Malignant neoplasm, other | neoplasms | RATE_C | |
| 195.3 | 0.5831 | 0.09045 | 1.792 | 1.14e-10 | 19155 | 870 | 18285 | | | | | 26453 | 309 | 26144 | 0.002813 | 0.4936 | Malignant neoplasm of head, face, and neck | neoplasms | RATE_C | |
| 196 | 0.7236 | 0.08838 | 2.062 | 2.66e-16 | 19459 | 1174 | 18285 | 0.8279 | 0.06982 | 2.289 | 1.97e-32 | 27555 | 1411 | 26144 | 0.8931 | 0.01434 | Radiotherapy | neoplasms | RATE_C | |
| 197 | 0.7237 | 0.04467 | 2.062 | 5.05e-59 | 22676 | 4391 | 18285 | 0.9089 | 0.03866 | 2.482 | 3.18e-122 | 30992 | 4848 | 26144 | 2.54e-04 | 0.2053 | Chemotherapy | neoplasms | RATE_C | |
| 198 | 0.7898 | 0.04789 | 2.203 | 4.09e-61 | 22039 | 3754 | 18285 | 1.171 | 0.04714 | 3.226 | 3.05e-136 | 29550 | 3406 | 26144 | 4.60e-09 | 0.3759 | Secondary malignant neoplasm | neoplasms | RATE_C | |
| 198.1 | 0.8085 | 0.06341 | 2.244 | 3.13e-37 | 20292 | 2007 | 18285 | 1.255 | 0.06645 | 3.509 | 1.34e-79 | 27825 | 1681 | 26144 | 5.55e-07 | 0.4375 | Secondary malignancy of lymph nodes | neoplasms | RATE_C | |
| 198.2 | 1.147 | 0.08222 | 3.149 | 3.02e-44 | 19626 | 1341 | 18285 | 1.439 | 0.08258 | 4.217 | 5.07e-68 | 27296 | 1152 | 26144 | 0.004535 | 0.3172 | Secondary malignancy of respiratory organs | neoplasms | RATE_C | |

| phenotype | male_beta | male_SE | male_OR | male_p | male_n_total | male_n_cases | male_n_controls | female_beta | female_SE | female_OR | female_p | female_n_total | female_n_cases | female_n_controls | interaction_p | interaction_beta | description | group | lab | label |
|---|---|---|---|---|---|---|---|---|---|---|---|---|---|---|---|---|---|---|---|---|
| 198.3 | 1.092 | 0.1207 | 2.979 | 1.48e-19 | 18851 | 566 | 18285 | 1.428 | 0.1153 | 4.169 | 3.43e-35 | 26726 | 582 | 26144 | 0.004992 | 0.447 | Secondary malignant neoplasm of digestive systems | neoplasms | RATE_C | |
| 198.4 | 0.9763 | 0.08599 | 2.655 | 7.06e-30 | 19347 | 1062 | 18285 | 1.305 | 0.08534 | 3.688 | 8.64e-53 | 27121 | 977 | 26144 | 8.99e-04 | 0.3835 | Secondary malignant neoplasm of liver | neoplasms | RATE_C | |
| 198.5 | 1.187 | 0.1306 | 3.277 | 9.76e-20 | 18799 | 514 | 18285 | 1.53 | 0.1144 | 4.616 | 8.86e-41 | 26737 | 593 | 26144 | 0.0427 | 0.3366 | Secondary malignancy of brain/spine | neoplasms | RATE_C | |
| 198.6 | 0.917 | 0.09119 | 2.502 | 8.67e-24 | 19284 | 999 | 18285 | 1.303 | 0.09534 | 3.68 | 1.61e-42 | 26962 | 818 | 26144 | 0.006027 | 0.3468 | Secondary malignancy of bone | neoplasms | RATE_C | |
| 198.7 | | | | | 18508 | 223 | 18285 | | | | | 26369 | 225 | 26144 | | | Secondary malignant neoplasm of skin | neoplasms | RATE_C | |
| 199 | 0.7236 | 0.08044 | 2.062 | 2.36e-19 | 19599 | 1314 | 18285 | 0.9906 | 0.06726 | 2.693 | 4.35e-49 | 27705 | 1561 | 26144 | 0.05049 | 0.1934 | Neoplasm of uncertain behavior | neoplasms | RATE_C | |
| 199.4 | | | | | 18315 | 30 | 18285 | | | | | 26186 | 42 | 26144 | | | Neurofibromatosis | neoplasms | RATE_C | |
| 200 | -0.4899 | 0.09072 | 0.6127 | 6.65e-08 | 26538 | 642 | 25896 | -0.002529 | 0.09483 | 0.9975 | 0.9787 | 35761 | 577 | 35184 | 7.22e-04 | 0.4199 | Myeloproliferative disease | neoplasms | RATE_C | |
| 200.1 | | | | | 21974 | 134 | 21840 | | | | | 30355 | 101 | 30254 | | | Polycythemia vera | neoplasms | RATE_C | |
| 201 | | | | | 25974 | 78 | 25896 | | | | | 35242 | 58 | 35184 | | | Hodgkin's disease | neoplasms | RATE_C | |
| 202 | | | | | 26283 | 387 | 25896 | | | | | 35476 | 292 | 35184 | 0.08365 | 0.3103 | Cancer of other lymphoid, histiocytic tissue | neoplasms | RATE_C | |
| 202.2 | | | | | 26223 | 327 | 25896 | | | | | 35425 | 241 | 35184 | 0.1506 | 0.2767 | Non-Hodgkins lymphoma | neoplasms | RATE_C | |
| 202.21 | | | | | 25957 | 61 | 25896 | | | | | 35235 | 51 | 35184 | | | Nodular lymphoma | neoplasms | RATE_C | |
| 202.22 | | | | | 25981 | 85 | 25896 | | | | | 35237 | 53 | 35184 | | | Reticulosarcoma | neoplasms | RATE_C | |
| 202.23 | | | | | 25947 | 51 | 25896 | | | | | 35208 | 24 | 35184 | | | Lymphosarcoma | neoplasms | RATE_C | |
| 202.24 | | | | | 25960 | 64 | 25896 | | | | | 35217 | 33 | 35184 | | | Large cell lymphoma | neoplasms | RATE_C | |
| 204 | -0.1734 | 0.08972 | 0.8408 | 0.05326 | 26591 | 695 | 25896 | 0.1674 | 0.1 | 1.182 | 0.09423 | 35705 | 521 | 35184 | 0.008054 | 0.3341 | Leukemia | neoplasms | RATE_C | |
| 204.1 | | | | | 26113 | 217 | 25896 | | | | | 35327 | 143 | 35184 | | | Lymphoid leukemia | neoplasms | RATE_C | |
| 204.11 | | | | | 25996 | 100 | 25896 | | | | | 35260 | 76 | 35184 | | | Lymphoid leukemia, acute | neoplasms | RATE_C | |
| 204.12 | | | | | 26033 | 137 | 25896 | | | | | 35270 | 86 | 35184 | | | Lymphoid leukemia, chronic | neoplasms | RATE_C | |
| 204.2 | | | | | 26331 | 435 | 25896 | | | | | 35537 | 353 | 35184 | 0.02242 | 0.351 | Myeloid leukemia | neoplasms | RATE_C | |
| 204.21 | | | | | 26263 | 367 | 25896 | | | | | 35482 | 298 | 35184 | 0.09426 | 0.2854 | Myeloid leukemia, acute | neoplasms | RATE_C | |
| 204.22 | | | | | 26069 | 173 | 25896 | | | | | 35327 | 143 | 35184 | | | Myeloid leukemia, chronic | neoplasms | RATE_C | |
| 204.3 | | | | | 25933 | 37 | 25896 | | | | | 35214 | 30 | 35184 | | | Monocytic leukemia | neoplasms | RATE_C | |
| 204.4 | | | | | 26060 | 164 | 25896 | | | | | 35291 | 107 | 35184 | | | Multiple myeloma | neoplasms | RATE_C | |
| 208 | 0.3015 | 0.05725 | 1.352 | 1.39e-07 | 24481 | 2858 | 21623 | 0.4405 | 0.05308 | 1.553 | 1.06e-16 | 33540 | 2573 | 30967 | 0.005588 | 0.2059 | Benign neoplasm of colon | neoplasms | RATE_C | |
| 209 | | | | | 27760 | 86 | 27674 | | | | | 36911 | 107 | 36804 | | | Neuroendocrine tumors | neoplasms | RATE_C | |
| 210 | | | | | 26386 | 107 | 26279 | | | | | 36167 | 130 | 36037 | | | Benign neoplasm of lip, oral cavity, and pharynx | neoplasms | RATE_C | |
| 211 | | | | | 24095 | 236 | 23859 | | | | | 33779 | 379 | 33400 | 0.0778 | 0.3984 | Benign neoplasm of other parts of digestive system | neoplasms | RATE_C | |

| phenotype | male_beta | male_SE | male_OR | male_p | male_n_total | male_n_cases | male_n_controls | female_beta | female_SE | female_OR | female_p | female_n_total | female_n_cases | female_n_controls | interaction_p | interaction_beta | description | group | lab | label |
|---|---|---|---|---|---|---|---|---|---|---|---|---|---|---|---|---|---|---|---|---|
| 212 | | | | | 27637 | 101 | 27536 | | | | | 36801 | 112 | 36689 | | | Benign neoplasm of respiratory and intrathoracic organs | neoplasms | RATE_C | |
| 213 | | | | | 21553 | 68 | 21485 | | | | | 26105 | 131 | 25974 | | | Benign neoplasm of bone and articular cartilage | neoplasms | RATE_C | |
| 214 | | | | | 27025 | 277 | 26748 | | | | | 35990 | 359 | 35631 | 0.8635 | -0.03376 | Lipoma | neoplasms | RATE_C | |
| 214.1 | | | | | 26919 | 171 | 26748 | | | | | 35867 | 236 | 35631 | | | Lipoma of skin and subcutaneous tissue | neoplasms | RATE_C | |
| 215 | | | | | 26820 | 72 | 26748 | | | | | 35793 | 162 | 35631 | | | Other benign neoplasm of connective and other soft tissue | neoplasms | RATE_C | |
| 216 | 0.04893 | 0.06862 | 1.05 | 0.4758 | 25936 | 1607 | 24329 | 0.2065 | 0.0488 | 1.229 | 2.31e-05 | 34007 | 2621 | 31386 | 0.8971 | 0.01007 | Benign neoplasm of skin | neoplasms | RATE_C | |
| 216.1 | | | | | 24408 | 79 | 24329 | | | | | 31484 | 98 | 31386 | | | Screening for malignant neoplasms of the skin | neoplasms | RATE_C | |
| 217 | | | | | 24477 | 148 | 24329 | | | | | 31698 | 312 | 31386 | | | Vascular hamartomas and non-neoplastic nevi | neoplasms | RATE_C | |
| 217.1 | | | | | 24444 | 115 | 24329 | | | | | 31637 | 251 | 31386 | | | Nevus, non-neoplastic | neoplasms | RATE_C | |
| 218 | | | | | 7 | | | 0.4907 | 0.08155 | 1.634 | 1.77e-09 | 31913 | 869 | 31044 | 0.9993 | 0.3658 | Benign neoplasm of uterus | neoplasms | RATE_C | |
| 218.1 | | | | | 7 | | | 0.4529 | 0.08326 | 1.573 | 5.36e-08 | 31868 | 824 | 31044 | 0.9994 | 0.3274 | Uterine leiomyoma | neoplasms | RATE_C | |
| 218.2 | | | | | 7 | | | | | | | 31064 | 20 | 31044 | | | Other benign neoplasm of uterus | neoplasms | RATE_C | |

| phenotype | male_beta | male_SE | male_OR | male_p | male_n_total | male_n_cases | male_n_controls | female_beta | female_SE | female_OR | female_p | female_n_total | female_n_cases | female_n_controls | interaction_p | interaction_beta | description | group | lab | label |
|---|---|---|---|---|---|---|---|---|---|---|---|---|---|---|---|---|---|---|---|---|
| 220 | | | | | 7 | | | | | | | 28308 | 209 | 28099 | | | Benign neoplasm of ovary | neoplasms | RATE_C | |
| 221 | | | | | 7 | | | | | | | 28086 | 59 | 28027 | | | Benign neoplasm of other female genital organs | neoplasms | RATE_C | |
| 222 | | | | | 24885 | 21 | 24864 | | | | | 4 | | | | | Benign neoplasm of male genital organs | neoplasms | RATE_C | |
| 223 | | | | | 21509 | 24 | 21485 | | | | | 26009 | 35 | 25974 | | | Benign neoplasm of kidney and other urinary organs | neoplasms | RATE_C | |
| 224 | | | | | 26668 | 110 | 26558 | | | | | 35659 | 140 | 35519 | | | Benign neoplasm of eye | neoplasms | RATE_C | |
| 224.1 | | | | | 26643 | 85 | 26558 | | | | | 35621 | 102 | 35519 | | | Benign neoplasm of eye, uveal | neoplasms | RATE_C | |
| 225 | | | | | 26749 | 191 | 26558 | | | | | 35909 | 390 | 35519 | 0.04606 | 0.4319 | Benign neoplasm of brain and other parts of nervous system | neoplasms | RATE_C | |
| 225.1 | | | | | 26730 | 172 | 26558 | | | | | 35878 | 359 | 35519 | 0.2157 | 0.2903 | Benign neoplasm of brain, cranial nerves, meninges | neoplasms | RATE_C | |
| 225.2 | | | | | 26574 | 16 | 26558 | | | | | 35551 | 32 | 35519 | | | Benign neoplasm of spinal cord, meninges | neoplasms | RATE_C | |
| 226 | | | | | 26971 | 25 | 26946 | | | | | 35500 | 129 | 35371 | | | Benign neoplasm of thyroid glands | neoplasms | RATE_C | |
| 227 | | | | | 27177 | 231 | 26946 | | | | | 35717 | 346 | 35371 | 0.005231 | 0.5761 | Benign neoplasm of other endocrine glands and related structures | neoplasms | RATE_C | |
| 227.1 | | | | | 27032 | 86 | 26946 | | | | | 35495 | 124 | 35371 | | | Benign neoplasm of adrenal gland | neoplasms | RATE_C | |
| 227.2 | | | | | 26972 | 26 | 26946 | | | | | 35433 | 62 | 35371 | | | Benign neoplasm of parathyroid gland | neoplasms | RATE_C | |
| 227.3 | | | | | 27062 | 116 | 26946 | | | | | 35525 | 154 | 35371 | | | Benign neoplasm of pituitary gland and craniopharyngeal duct (pouch) | neoplasms | RATE_C | |
| 228 | 0.1397 | 0.1232 | 1.15 | 0.2569 | 26936 | 519 | 26417 | 0.2053 | 0.08896 | 1.228 | 0.02102 | 35731 | 759 | 34972 | 0.7191 | -0.05153 | Hemangioma and lymphangioma, any site | neoplasms | RATE_C | |

| phenotype | male_beta | male_SE | male_OR | male_p | male_n_total | male_n_cases | male_n_controls | female_beta | female_SE | female_OR | female_p | female_n_total | female_n_cases | female_n_controls | interaction_p | interaction_beta | description | group | lab | label |
|---|---|---|---|---|---|---|---|---|---|---|---|---|---|---|---|---|---|---|---|---|
| 228.1 | | | | | 26853 | 436 | 26417 | 0.1238 | 0.0983 | 1.132 | 0.208 | 35591 | 619 | 34972 | 0.6211 | -0.07828 | Hemangioma of skin and subcutaneous tissue | neoplasms | RATE_C | |
| 229 | | | | | 27551 | 83 | 27468 | | | | | 36445 | 69 | 36376 | | | Benign neoplasm of unspecified sites | neoplasms | RATE_C | |
| 229.1 | | | | | 27477 | 9 | 27468 | | | | | 36386 | 10 | 36376 | | | Benign neoplasm of lymph nodes | neoplasms | RATE_C | |
| 230 | | | | | 27785 | 15 | 27770 | | | | | 36952 | 4 | 36948 | | | Kaposi's sarcoma | neoplasms | RATE_C | |
| 240 | | | | | 23255 | 80 | 23175 | | | | | 26401 | 346 | 26055 | | | Simple and unspecified goiter | endocrine/metabolic | RATE_C | |
| 241 | 0.2928 | 0.1227 | 1.34 | 0.01705 | 23679 | 504 | 23175 | 0.4795 | 0.05595 | 1.615 | 1.03e-17 | 28029 | 1974 | 26055 | 0.4654 | 0.09352 | Nontoxic nodular goiter | endocrine/metabolic | RATE_C | |
| 241.1 | | | | | 23492 | 317 | 23175 | 0.5057 | 0.07894 | 1.658 | 1.49e-10 | 27019 | 964 | 26055 | 0.9676 | -0.006896 | Nontoxic uninodular goiter | endocrine/metabolic | RATE_C | |
| 241.2 | | | | | 23428 | 253 | 23175 | 0.6111 | 0.07117 | 1.842 | 9.03e-18 | 27333 | 1278 | 26055 | 0.1521 | 0.256 | Nontoxic multinodular goiter | endocrine/metabolic | RATE_C | |
| 242 | | | | | 23461 | 286 | 23175 | 0.5641 | 0.08475 | 1.758 | 2.81e-11 | 26848 | 793 | 26055 | 0.3089 | 0.1684 | Thyrotoxicosis with or without goiter | endocrine/metabolic | RATE_C | |
| 242.1 | | | | | 23301 | 126 | 23175 | | | | | 26434 | 379 | 26055 | 0.4867 | 0.159 | Graves' disease | endocrine/metabolic | RATE_C | |
| 242.2 | | | | | 23196 | 21 | 23175 | | | | | 26178 | 123 | 26055 | | | Toxic multinodular goiter | endocrine/metabolic | RATE_C | |
| 242.3 | | | | | 23231 | 56 | 23175 | | | | | 26122 | 67 | 26055 | | | Exophthalmos | endocrine/metabolic | RATE_C | |
| 242.31 | | | | | 23186 | 11 | 23175 | | | | | 26071 | 16 | 26055 | | | Thyrotoxic exophthalmos | endocrine/metabolic | RATE_C | |
| 244 | 0.6725 | 0.06088 | 1.959 | 2.29e-28 | 25712 | 2537 | 23175 | 0.5783 | 0.03359 | 1.783 | 1.99e-66 | 32658 | 6603 | 26055 | 0.4791 | -0.04689 | Hypothyroidism | endocrine/metabolic | RATE_C | |
| 244.1 | 0.6628 | 0.1291 | 1.94 | 2.86e-07 | 23694 | 519 | 23175 | 0.5924 | 0.06764 | 1.808 | 2.00e-18 | 27402 | 1347 | 26055 | 0.7483 | -0.04438 | Secondary hypothyroidism | endocrine/metabolic | RATE_C | |
| 244.2 | | | | | 23522 | 347 | 23175 | 0.3865 | 0.08314 | 1.472 | 3.34e-06 | 26876 | 821 | 26055 | 0.9229 | 0.01522 | Acquired hypothyroidism | endocrine/metabolic | RATE_C | |
| 244.3 | | | | | 23175 | | | | | | | 26060 | 5 | 26055 | | | Iodine hypothyroidism | endocrine/metabolic | RATE_C | |
| 244.4 | 0.6844 | 0.06478 | 1.983 | 4.34e-26 | 25441 | 2266 | 23175 | 0.6144 | 0.03525 | 1.849 | 5.00e-68 | 32096 | 6041 | 26055 | 0.7588 | -0.02163 | Hypothyroidism NOS | endocrine/metabolic | RATE_C | |
| 244.5 | | | | | 23199 | 24 | 23175 | | | | | 26089 | 34 | 26055 | | | Congenital hypothyroidism | endocrine/metabolic | RATE_C | |
| 245 | | | | | 23436 | 261 | 23175 | 0.2457 | 0.07657 | 1.278 | 0.001334 | 26942 | 887 | 26055 | 0.341 | -0.1555 | Thyroiditis | endocrine/metabolic | RATE_C | |
| 245.1 | | | | | 23196 | 21 | 23175 | | | | | 26097 | 42 | 26055 | | | Thyroiditis, acute and subacute | endocrine/metabolic | RATE_C | |
| 245.2 | | | | | 23326 | 151 | 23175 | 0.1405 | 0.08789 | 1.151 | 0.1099 | 26707 | 652 | 26055 | 0.828 | -0.04336 | Chronic thyroiditis | endocrine/metabolic | RATE_C | |
| 245.21 | | | | | 23324 | 149 | 23175 | 0.1357 | 0.08823 | 1.145 | 0.1242 | 26701 | 646 | 26055 | 0.8617 | -0.03489 | Chronic lymphocytic thyroiditis | endocrine/metabolic | RATE_C | |
| 246 | | | | | 23388 | 213 | 23175 | | | | | 26539 | 484 | 26055 | 0.1678 | -0.319 | Other disorders of thyroid | endocrine/metabolic | RATE_C | |
| 246.2 | | | | | 23182 | 7 | 23175 | | | | | 26065 | 10 | 26055 | | | Thyroid cyst | endocrine/metabolic | RATE_C | |
| 246.7 | | | | | 23294 | 119 | 23175 | | | | | 26304 | 249 | 26055 | | | Abnormal results of function study of thyroid | endocrine/metabolic | RATE_C | |
| 249 | | | | | 17130 | 307 | 16823 | | | | | 26073 | 265 | 25808 | 0.7528 | 0.07854 | Secondary diabetes mellitus | endocrine/metabolic | RATE_C | |
| 250 | 1.087 | 0.0427 | 2.966 | 5.87e-143 | 22885 | 6062 | 16823 | 1.131 | 0.03775 | 3.1 | 2.18e-197 | 31483 | 5675 | 25808 | 0.8663 | -0.009191 | Diabetes mellitus | endocrine/metabolic | RATE_C | |
| 250.1 | 0.9963 | 0.08249 | 2.708 | 1.39e-33 | 18010 | 1187 | 16823 | 1.034 | 0.07123 | 2.813 | 9.09e-48 | 27021 | 1213 | 25808 | 0.3821 | -0.08795 | Type 1 diabetes | endocrine/metabolic | RATE_C | |

| phenotype | male_beta | male_SE | male_OR | male_p | male_n_total | male_n_cases | male_n_controls | female_beta | female_SE | female_OR | female_p | female_n_total | female_n_cases | female_n_controls | interaction_p | interaction_beta | description | group | lab | label |
|---|---|---|---|---|---|---|---|---|---|---|---|---|---|---|---|---|---|---|---|---|
| 250.11 | | | | | 16945 | 122 | 16823 | | | | | 25971 | 163 | 25808 | | | Type 1 diabetes with ketoacidosis | endocrine/metabolic | RATE_C | |
| 250.12 | | | | | 17020 | 197 | 16823 | | | | | 25984 | 176 | 25808 | | | Type 1 diabetes with renal manifestations | endocrine/metabolic | RATE_C | |
| 250.13 | | | | | 16957 | 134 | 16823 | | | | | 25957 | 149 | 25808 | | | Type 1 diabetes with ophthalmic manifestations | endocrine/metabolic | RATE_C | |
| 250.14 | | | | | 17123 | 300 | 16823 | | | | | 26082 | 274 | 25808 | 0.1089 | -0.4567 | Type 1 diabetes with neurological manifestations | endocrine/metabolic | RATE_C | |
| 250.15 | | | | | 16873 | 50 | 16823 | | | | | 25855 | 47 | 25808 | | | Diabetes type 1 with peripheral circulatory disorders | endocrine/metabolic | RATE_C | |
| 250.2 | 1.142 | 0.04421 | 3.133 | 4.39e-147 | 22677 | 5854 | 16823 | 1.187 | 0.03923 | 3.276 | 5.90e-201 | 31244 | 5436 | 25808 | 0.9731 | -0.00192 | Type 2 diabetes | endocrine/metabolic | RATE_C | |
| 250.21 | | | | | 16879 | 56 | 16823 | | | | | 25884 | 76 | 25808 | | | Type 2 diabetes with ketoacidosis | endocrine/metabolic | RATE_C | |
| 250.22 | 1.658 | 0.1091 | 5.251 | 3.68e-52 | 18226 | 1403 | 16823 | 1.73 | 0.1167 | 5.642 | 1.00e-49 | 26727 | 919 | 25808 | 0.4942 | 0.1065 | Type 2 diabetes with renal manifestations | endocrine/metabolic | RATE_C | |
| 250.23 | | | | | 17271 | 448 | 16823 | | | | | 26204 | 396 | 25808 | 0.8341 | 0.05157 | Type 2 diabetes with ophthalmic manifestations | endocrine/metabolic | RATE_C | |
| 250.24 | 1.496 | 0.0972 | 4.463 | 1.89e-53 | 18366 | 1543 | 16823 | 1.609 | 0.09405 | 4.997 | 1.33e-65 | 27084 | 1276 | 25808 | 0.3355 | 0.1268 | Type 2 diabetes with neurological manifestations | endocrine/metabolic | RATE_C | |
| 250.25 | | | | | 17307 | 484 | 16823 | | | | | 26109 | 301 | 25808 | 0.4091 | -0.2485 | Diabetes type 2 with peripheral circulatory disorders | endocrine/metabolic | RATE_C | |
| 250.3 | 1.417 | 0.07642 | 4.123 | 1.01e-76 | 18803 | 1980 | 16823 | 1.218 | 0.06418 | 3.381 | 2.48e-80 | 27587 | 1779 | 25808 | 0.00319 | -0.2787 | Insulin pump user | endocrine/metabolic | RATE_C | |
| 250.4 | 1.516 | 0.05723 | 4.554 | 1.22e-154 | 20734 | 3911 | 16823 | 1.393 | 0.049 | 4.028 | 6.90e-178 | 29488 | 3680 | 25808 | 0.1472 | -0.1049 | Abnormal glucose | endocrine/metabolic | RATE_C | |
| 250.41 | 0.5063 | 0.1192 | 1.659 | 2.17e-05 | 17402 | 579 | 16823 | 0.8255 | 0.1022 | 2.283 | 6.66e-16 | 26465 | 657 | 25808 | 0.2195 | 0.1823 | Impaired fasting glucose | endocrine/metabolic | RATE_C | |
| 250.42 | 1.686 | 0.06311 | 5.397 | 3.41e-157 | 20295 | 3472 | 16823 | 1.524 | 0.05439 | 4.593 | 6.93e-173 | 28947 | 3139 | 25808 | 0.1121 | -0.1275 | Other abnormal glucose | endocrine/metabolic | RATE_C | |
| 250.5 | | | | | 16841 | 18 | 16823 | | | | | 25844 | 36 | 25808 | | | Glycosuria or Acetonuria | endocrine/metabolic | RATE_C | |
| phenotype | male_beta | male_SE | male_OR | male_p | male_n_total | male_n_cases | male_n_controls | female_beta | female_SE | female_OR | female_p | female_n_total | female_n_cases | female_n_controls | interaction_p | interaction_beta | description | group | lab | label |
| 250.6 | 1.507 | 0.1091 | 4.511 | 2.36e-43 | 18032 | 1209 | 16823 | 1.492 | 0.1031 | 4.448 | 1.85e-47 | 26769 | 961 | 25808 | 0.9968 | 5.93e-04 | Polyneuropathy in diabetes | endocrine/metabolic | RATE_C | |
| 250.7 | | | | | 24804 | 450 | 24354 | | | | | 33128 | 398 | 32730 | 0.9897 | 0.003317 | Diabetic retinopathy | endocrine/metabolic | RATE_C | |
| 251 | | | | | 23451 | 170 | 23281 | | | | | 31202 | 167 | 31035 | | | Other disorders of pancreatic internal secretion | endocrine/metabolic | RATE_C | |
| 251.1 | | | | | 15397 | 246 | 15151 | | | | | 23345 | 400 | 22945 | 0.056 | -0.3991 | Hypoglycemia | endocrine/metabolic | RATE_C | |
| 251.8 | | | | | 23287 | 6 | 23281 | | | | | 31042 | 7 | 31035 | | | Abnormality of secretion of glucagon or gastrin | endocrine/metabolic | RATE_C | |
| 252 | | | | | 23548 | 267 | 23281 | 0.7645 | 0.09874 | 2.148 | 9.77e-15 | 31785 | 750 | 31035 | 0.9436 | -0.0148 | Disorders of parathyroid gland | endocrine/metabolic | RATE_C | |
| 252.1 | | | | | 23519 | 238 | 23281 | 0.7979 | 0.1074 | 2.221 | 1.08e-13 | 31693 | 658 | 31035 | 0.8419 | 0.04465 | Hyperparathyroidism | endocrine/metabolic | RATE_C | |
| 252.2 | | | | | 23316 | 35 | 23281 | | | | | 31163 | 128 | 31035 | | | Hypoparathyroidism | endocrine/metabolic | RATE_C | |
| 253 | | | | | 23665 | 384 | 23281 | | | | | 31431 | 396 | 31035 | 1.25e-04 | 0.5889 | Disorders of the pituitary gland and its hypothalamic control | endocrine/metabolic | RATE_C | |
| 253.1 | | | | | 23343 | 62 | 23281 | | | | | 31171 | 136 | 31035 | | | Pituitary hyperfunction | endocrine/metabolic | RATE_C | |
| 253.11 | | | | | 23302 | 21 | 23281 | | | | | 31046 | 11 | 31035 | | | Acromegaly and gigantism | endocrine/metabolic | RATE_C | |
| 253.2 | | | | | 23400 | 119 | 23281 | | | | | 31125 | 90 | 31035 | | | Pituitary hypofunction | endocrine/metabolic | RATE_C | |

| phenotype | male_beta | male_SE | male_OR | male_p | male_n_total | male_n_cases | male_n_controls | female_beta | female_SE | female_OR | female_p | female_n_total | female_n_cases | female_n_controls | interaction_p | interaction_beta | description | group | lab | label |
|---|---|---|---|---|---|---|---|---|---|---|---|---|---|---|---|---|---|---|---|---|
| 253.3 | | | | | 23305 | 24 | 23281 | | | | | 31086 | 51 | 31035 | | | Diabetes insipidus | endocrine/metabolic | RATE_C | |
| 253.4 | | | | | 23423 | 142 | 23281 | | | | | 31110 | 75 | 31035 | | | Anterior pituitary disorders | endocrine/metabolic | RATE_C | |
| 253.5 | | | | | 23359 | 78 | 23281 | | | | | 31085 | 50 | 31035 | | | Pituitary dwarfism | endocrine/metabolic | RATE_C | |
| 253.7 | | | | | 23345 | 64 | 23281 | | | | | 31102 | 67 | 31035 | | | Other disorders of neurohypophysis | endocrine/metabolic | RATE_C | |
| 254 | | | | | 23286 | 5 | 23281 | | | | | 31042 | 7 | 31035 | | | Diseases of thymus gland | endocrine/metabolic | RATE_C | |
| 255 | 1.007 | 0.1171 | 2.738 | 8.03e-18 | 23957 | 676 | 23281 | 1.22 | 0.09345 | 3.387 | 6.03e-39 | 31897 | 862 | 31035 | 0.2164 | 0.1742 | Disorders of adrenal glands | endocrine/metabolic | RATE_C | |
| 255.1 | | | | | 23371 | 90 | 23281 | | | | | 31211 | 176 | 31035 | | | Adrenal hyperfunction | endocrine/metabolic | RATE_C | |
| 255.11 | | | | | 23313 | 32 | 23281 | | | | | 31129 | 94 | 31035 | | | Cushing's syndrome | endocrine/metabolic | RATE_C | |
| 255.12 | | | | | 23315 | 34 | 23281 | | | | | 31064 | 29 | 31035 | | | Hyperaldosteronism | endocrine/metabolic | RATE_C | |
| 255.13 | | | | | 23297 | 16 | 23281 | | | | | 31065 | 30 | 31035 | | | Medulloadrenal hyperfunction | endocrine/metabolic | RATE_C | |
| 255.2 | | | | | 23716 | 435 | 23281 | 1.217 | 0.1204 | 3.376 | 5.36e-24 | 31535 | 500 | 31035 | 0.09691 | 0.2848 | Adrenal hypofunction | endocrine/metabolic | RATE_C | |
| 255.21 | | | | | 23674 | 393 | 23281 | | | | | 31474 | 439 | 31035 | 0.1021 | 0.2972 | Glucocorticoid deficiency | endocrine/metabolic | RATE_C | |
| 255.22 | | | | | 23296 | 15 | 23281 | | | | | 31067 | 32 | 31035 | | | Mineralocorticoid deficiency | endocrine/metabolic | RATE_C | |
| 255.3 | | | | | 23289 | 8 | 23281 | | | | | 31065 | 30 | 31035 | | | Adrenogenital disorders | endocrine/metabolic | RATE_C | |
| 256 | | | | | 4 | | | 0.3497 | 0.08783 | 1.419 | 6.85e-05 | 31698 | 666 | 31032 | 6.85e-05 | 0.3497 | Ovarian dysfunction | endocrine/metabolic | RATE_C | |
| 256.1 | | | | | 23283 | 2 | 23281 | | | | | 31037 | 2 | 31035 | | | Hyperestrogenism | endocrine/metabolic | RATE_C | |
| 256.4 | | | | | 4 | | | 0.4828 | 0.0979 | 1.621 | 8.18e-07 | 31569 | 537 | 31032 | 8.18e-07 | 0.4828 | Polycystic ovaries | endocrine/metabolic | RATE_C | |
| 257 | 0.2336 | 0.08329 | 1.263 | 0.005039 | 24271 | 994 | 23277 | | | | | 3 | | | 0.9995 | 0.2343 | Testicular dysfunction | endocrine/metabolic | RATE_C | |
| 257.1 | 0.2256 | 0.08389 | 1.253 | 0.007165 | 24255 | 978 | 23277 | | | | | 3 | | | 0.9995 | 0.2403 | Testicular hypofunction | endocrine/metabolic | RATE_C | |
| 258 | | | | | 23304 | 23 | 23281 | | | | | 31068 | 33 | 31035 | | | Iatrogenic endocrine disorders | endocrine/metabolic | RATE_C | |
| 258.1 | | | | | 4 | | | | | | | 31087 | 55 | 31032 | | | Postablative ovarian failure | endocrine/metabolic | RATE_C | |
| 259 | | | | | 23461 | 180 | 23281 | | | | | 31392 | 357 | 31035 | 0.5281 | -0.1268 | Other endocrine disorders | endocrine/metabolic | RATE_C | |
| 259.1 | | | | | 23352 | 71 | 23281 | | | | | 31134 | 99 | 31035 | | | Nonspecific abnormal results of other endocrine function study | endocrine/metabolic | RATE_C | |
| 259.2 | | | | | 23320 | 39 | 23281 | | | | | 31092 | 57 | 31035 | | | Carcinoid syndrome | endocrine/metabolic | RATE_C | |
| 259.3 | | | | | 23324 | 43 | 23281 | | | | | 31063 | 28 | 31035 | | | Delay in sexual development and puberty NEC | endocrine/metabolic | RATE_C | |
| 259.4 | | | | | 23291 | 10 | 23281 | | | | | 31071 | 36 | 31035 | | | Precocious sexual development and puberty NEC | endocrine/metabolic | RATE_C | |
| 259.8 | | | | | 23297 | 16 | 23281 | | | | | 31067 | 32 | 31035 | | | Polyglandular activity in multiple endocrine adenomatosis | endocrine/metabolic | RATE_C | |
| 260 | 1.436 | 0.06352 | 4.204 | 3.69e-113 | 22122 | 2837 | 19285 | 1.634 | 0.06164 | 5.127 | 6.10e-155 | 27318 | 2532 | 24786 | 4.62e-05 | 0.3445 | Protein-calorie malnutrition | endocrine/metabolic | RATE_C | |
| 260.1 | | | | | 19435 | 150 | 19285 | | | | | 24879 | 93 | 24786 | | | Cachexia | endocrine/metabolic | RATE_C | |

| phenotype | male_beta | male_SE | male_OR | male_p | male_n_total | male_n_cases | male_n_controls | female_beta | female_SE | female_OR | female_p | female_n_total | female_n_cases | female_n_controls | interaction_p | interaction_beta | description | group | lab | label |
|---|---|---|---|---|---|---|---|---|---|---|---|---|---|---|---|---|---|---|---|---|
| 260.2 | 1.563 | 0.1025 | 4.774 | 1.62e-52 | 20371 | 1086 | 19285 | 1.705 | 0.09603 | 5.503 | 1.48e-70 | 25774 | 988 | 24786 | 0.05186 | 0.2608 | severe protein-calorie malnutrition | endocrine/metabolic | RATE_C | |
| 260.21 | | | | | 19301 | 16 | 19285 | | | | | 24805 | 19 | 24786 | | | Kwashiorkor | endocrine/metabolic | RATE_C | |
| 260.22 | | | | | 19399 | 114 | 19285 | | | | | 24929 | 143 | 24786 | | | Nutritional marasmus | endocrine/metabolic | RATE_C | |
| 260.3 | 1.665 | 0.1533 | 5.287 | 1.68e-27 | 19914 | 629 | 19285 | 1.997 | 0.1565 | 7.367 | 2.68e-37 | 25366 | 580 | 24786 | 0.02437 | 0.4816 | Adult failure to thrive | endocrine/metabolic | RATE_C | |
| 260.6 | 1.156 | 0.123 | 3.178 | 5.42e-21 | 19875 | 590 | 19285 | | | | | 25233 | 447 | 24786 | 7.32e-06 | 0.8621 | Anorexia | endocrine/metabolic | RATE_C | |
| 260.7 | | | | | 19286 | 1 | 19285 | | | | | 24804 | 18 | 24786 | | | Polyphagia | endocrine/metabolic | RATE_C | |
| 261 | 0.4096 | 0.05659 | 1.506 | 4.54e-13 | 21716 | 2431 | 19285 | 0.4454 | 0.03614 | 1.561 | 6.77e-35 | 30360 | 5574 | 24786 | 2.76e-05 | 0.2617 | Vitamin deficiency | endocrine/metabolic | RATE_C | |
| 261.1 | | | | | 19312 | 27 | 19285 | | | | | 24829 | 43 | 24786 | | | Vitamin A deficiency | endocrine/metabolic | RATE_C | |
| 261.2 | 0.3364 | 0.1041 | 1.4 | 0.001235 | 19973 | 688 | 19285 | 0.7429 | 0.06589 | 2.102 | 1.75e-29 | 26376 | 1590 | 24786 | 0.001228 | 0.3765 | Vitamin B-complex deficiencies | endocrine/metabolic | RATE_C | |
| 261.3 | | | | | 19292 | 7 | 19285 | | | | | 24807 | 21 | 24786 | | | Vitamin C deficiencies | endocrine/metabolic | RATE_C | |
| 261.4 | 0.39 | 0.0621 | 1.477 | 3.37e-10 | 21216 | 1931 | 19285 | 0.4264 | 0.03894 | 1.532 | 6.66e-28 | 29414 | 4628 | 24786 | 1.96e-06 | 0.3224 | Vitamin D deficiency | endocrine/metabolic | RATE_C | |
| 261.41 | | | | | 19297 | 12 | 19285 | | | | | 24819 | 33 | 24786 | | | Rickets or osteomalacia | endocrine/metabolic | RATE_C | |
| 262 | | | | | 19493 | 208 | 19285 | | | | | 25217 | 431 | 24786 | 0.4421 | -0.1687 | Mineral deficiency NEC | endocrine/metabolic | RATE_C | |
| 263 | | | | | 19569 | 284 | 19285 | | | | | 25175 | 389 | 24786 | 0.1912 | -0.309 | Other nutritional deficiency | endocrine/metabolic | RATE_C | |
| 264 | | | | | 19689 | 404 | 19285 | | | | | 25083 | 297 | 24786 | 0.4276 | 0.1488 | Lack of normal physiological development | endocrine/metabolic | RATE_C | |
| 264.1 | | | | | 19424 | 139 | 19285 | | | | | 24879 | 93 | 24786 | | | Short stature | endocrine/metabolic | RATE_C | |
| 264.2 | | | | | 19409 | 124 | 19285 | | | | | 24883 | 97 | 24786 | | | Failure to thrive (childhood) | endocrine/metabolic | RATE_C | |
| 264.3 | | | | | 19388 | 103 | 19285 | | | | | 24857 | 71 | 24786 | | | Delayed milestones | endocrine/metabolic | RATE_C | |
| 264.9 | | | | | 19465 | 180 | 19285 | | | | | 24925 | 139 | 24786 | | | Lack of normal physiological development, unspecified | endocrine/metabolic | RATE_C | |
| phenotype | male_beta | male_SE | male_OR | male_p | male_n_total | male_n_cases | male_n_controls | female_beta | female_SE | female_OR | female_p | female_n_total | female_n_cases | female_n_controls | interaction_p | interaction_beta | description | group | lab | label |
| 269 | 0.5931 | 0.09415 | 1.81 | 2.99e-10 | 26340 | 917 | 25423 | 0.8199 | 0.09542 | 2.27 | 8.48e-18 | 35408 | 726 | 34682 | 0.1519 | 0.1779 | Proteinuria | endocrine/metabolic | RATE_C | |
| 270 | | | | | 25887 | 464 | 25423 | 0.7739 | 0.1141 | 2.168 | 1.17e-11 | 35187 | 505 | 34682 | 0.4123 | 0.134 | Disorders of protein plasma/amino-acid transport and metabolism | endocrine/metabolic | RATE_C | |
| 270.1 | | | | | 25452 | 29 | 25423 | | | | | 34732 | 50 | 34682 | | | Disturbances of amino-acid transport | endocrine/metabolic | RATE_C | |
| 270.11 | | | | | 25431 | 8 | 25423 | | | | | 34711 | 29 | 34682 | | | Disturbances of sulphur-bearing amino-acid metabolism | endocrine/metabolic | RATE_C | |
| 270.12 | | | | | 25434 | 11 | 25423 | | | | | 34696 | 14 | 34682 | | | Phenylketonuria [PKU] | endocrine/metabolic | RATE_C | |
| 270.2 | | | | | 25465 | 42 | 25423 | | | | | 34719 | 37 | 34682 | | | Disorders of amino-acid metabolism | endocrine/metabolic | RATE_C | |
| 270.21 | | | | | 25457 | 34 | 25423 | | | | | 34703 | 21 | 34682 | | | Disorders of urea cycle metabolism | endocrine/metabolic | RATE_C | |
| 270.3 | | | | | 25814 | 391 | 25423 | | | | | 35096 | 414 | 34682 | 0.3951 | 0.1626 | Disorders of plasma protein metabolism | endocrine/metabolic | RATE_C | |

| phenotype | male_beta | male_SE | male_OR | male_p | male_n_total | male_n_cases | male_n_controls | female_beta | female_SE | female_OR | female_p | female_n_total | female_n_cases | female_n_controls | interaction_p | interaction_beta | description | group | lab | label |
|---|---|---|---|---|---|---|---|---|---|---|---|---|---|---|---|---|---|---|---|---|
| 270.31 | | | | | 25425 | 2 | 25423 | | | | | 34689 | 7 | 34682 | | | Polyclonal hypergammaglobulinemia | endocrine/metabolic | RATE_C | |
| 270.32 | | | | | 25665 | 242 | 25423 | | | | | 34920 | 238 | 34682 | | | Paraproteinemia | endocrine/metabolic | RATE_C | |
| 270.33 | | | | | 25484 | 61 | 25423 | | | | | 34731 | 49 | 34682 | | | Amyloidosis | endocrine/metabolic | RATE_C | |
| 270.34 | | | | | 25480 | 57 | 25423 | | | | | 34726 | 44 | 34682 | | | Alpha-1-antitrypsin deficiency | endocrine/metabolic | RATE_C | |
| 270.35 | | | | | 25440 | 17 | 25423 | | | | | 34692 | 10 | 34682 | | | Macroglobulinemia | endocrine/metabolic | RATE_C | |
| 270.38 | | | | | 25509 | 86 | 25423 | | | | | 34792 | 110 | 34682 | | | Other specified disorders of plasma protein metabolism | endocrine/metabolic | RATE_C | |
| 271 | | | | | 27474 | 374 | 27100 | 0.4079 | 0.09717 | 1.504 | 2.70e-05 | 36489 | 676 | 35813 | 0.1598 | 0.2246 | Disorders of carbohydrate transport and metabolism | endocrine/metabolic | RATE_C | |
| 271.3 | | | | | 27455 | 355 | 27100 | 0.4175 | 0.09879 | 1.518 | 2.37e-05 | 36474 | 661 | 35813 | 0.1952 | 0.2136 | Intestinal disaccharidase deficiencies and disaccharide malabsorption | endocrine/metabolic | RATE_C | |
| 271.9 | | | | | 27119 | 19 | 27100 | | | | | 35828 | 15 | 35813 | | | Other disorders of carbohydrate transport and metabolism | endocrine/metabolic | RATE_C | |
| 272 | 1.103 | 0.03725 | 3.015 | 8.48e-193 | 25396 | 11129 | 14267 | 1.125 | 0.03224 | 3.082 | 4.44e-267 | 34085 | 10701 | 23384 | 0.09897 | 0.08019 | Disorders of lipoid metabolism | endocrine/metabolic | RATE_C | |
| 272.1 | 1.106 | 0.03729 | 3.022 | 2.46e-193 | 25384 | 11117 | 14267 | 1.127 | 0.03228 | 3.085 | 6.46e-267 | 34065 | 10681 | 23384 | 0.1032 | 0.0793 | Hyperlipidemia | endocrine/metabolic | RATE_C | |
| 272.11 | 1.096 | 0.05906 | 2.993 | 6.60e-77 | 18174 | 3907 | 14267 | 1.264 | 0.05197 | 3.54 | 1.16e-130 | 27356 | 3972 | 23384 | 0.002994 | 0.231 | Hypercholesterolemia | endocrine/metabolic | RATE_C | |
| 272.12 | | | | | 14667 | 400 | 14267 | | | | | 23678 | 294 | 23384 | 0.08615 | 0.3324 | Hyperglyceridemia | endocrine/metabolic | RATE_C | |
| 272.13 | 1.117 | 0.04958 | 3.056 | 2.13e-112 | 20087 | 5820 | 14267 | 1.146 | 0.04359 | 3.144 | 3.00e-152 | 28806 | 5422 | 23384 | 0.1087 | 0.1045 | Mixed hyperlipidemia | endocrine/metabolic | RATE_C | |
| 272.14 | | | | | 14272 | 5 | 14267 | | | | | 23388 | 4 | 23384 | | | Hyperchylomicronemia | endocrine/metabolic | RATE_C | |
| 272.9 | | | | | 14286 | 19 | 14267 | | | | | 23403 | 19 | 23384 | | | Unspecified disorder of lipoid metabolism | endocrine/metabolic | RATE_C | |
| 274 | 0.7752 | 0.07529 | 2.171 | 7.31e-25 | 26980 | 1926 | 25054 | 0.9395 | 0.1064 | 2.559 | 1.04e-18 | 36355 | 864 | 35491 | 0.005511 | 0.3514 | Gout and other crystal arthropathies | endocrine/metabolic | RATE_C | |
| 274.1 | 0.7668 | 0.07656 | 2.153 | 1.30e-23 | 26882 | 1828 | 25054 | 0.9572 | 0.1147 | 2.604 | 7.02e-17 | 36217 | 726 | 35491 | 0.008908 | 0.3499 | Gout | endocrine/metabolic | RATE_C | |
| 274.11 | 0.653 | 0.1252 | 1.921 | 1.81e-07 | 25702 | 648 | 25054 | | | | | 35744 | 253 | 35491 | 0.2999 | 0.2218 | Gouty arthropathy | endocrine/metabolic | RATE_C | |
| 274.2 | | | | | 25175 | 121 | 25054 | | | | | 35648 | 157 | 35491 | | | Crystal arthropathies | endocrine/metabolic | RATE_C | |
| 274.21 | | | | | 25165 | 111 | 25054 | | | | | 35627 | 136 | 35491 | | | Chondrocalcinosis | endocrine/metabolic | RATE_C | |
| 275 | 1.243 | 0.07062 | 3.468 | 2.11e-69 | 25825 | 2131 | 23694 | 1.16 | 0.05768 | 3.191 | 5.46e-90 | 34866 | 2480 | 32386 | 0.06016 | 0.1621 | Disorders of mineral metabolism | endocrine/metabolic | RATE_C | |
| 275.1 | | | | | 23853 | 159 | 23694 | | | | | 32511 | 125 | 32386 | | | Disorders of iron metabolism | hematopoietic | RATE_C | |
| 275.11 | | | | | 23755 | 61 | 23694 | | | | | 32420 | 34 | 32386 | | | Hereditary hemochromatosis | hematopoietic | RATE_C | |
| 275.2 | | | | | 23712 | 18 | 23694 | | | | | 32400 | 14 | 32386 | | | Disorders of copper metabolism | endocrine/metabolic | RATE_C | |
| 275.3 | 1.353 | 0.1107 | 3.869 | 2.26e-34 | 24542 | 848 | 23694 | 1.497 | 0.09718 | 4.469 | 1.49e-53 | 33312 | 926 | 32386 | 0.03783 | 0.2895 | Disorders of magnesium metabolism | endocrine/metabolic | RATE_C | |
| 275.5 | 1.618 | 0.1173 | 5.045 | 2.70e-43 | 24610 | 916 | 23694 | 1.433 | 0.09722 | 4.189 | 3.82e-49 | 33307 | 921 | 32386 | 0.989 | -0.001984 | Disorders of calcium/phosphorus metabolism | endocrine/metabolic | RATE_C | |
| 275.51 | | | | | 23996 | 302 | 23694 | | | | | 32803 | 417 | 32386 | 0.5513 | 0.1284 | Hypocalcemia | endocrine/metabolic | RATE_C | |
| 275.53 | 1.949 | 0.1658 | 7.025 | 6.27e-32 | 24238 | 544 | 23694 | | | | | 32769 | 383 | 32386 | 0.9325 | 0.01891 | Disorders of phosphorus metabolism | endocrine/metabolic | RATE_C | |

| phenotype | male_beta | male_SE | male_OR | male_p | male_n_total | male_n_cases | male_n_controls | female_beta | female_SE | female_OR | female_p | female_n_total | female_n_cases | female_n_controls | interaction_p | interaction_beta | description | group | lab | label |
|---|---|---|---|---|---|---|---|---|---|---|---|---|---|---|---|---|---|---|---|---|
| 275.6 | | | | | 24110 | 416 | 23694 | 0.8847 | 0.1081 | 2.422 | 2.68e-16 | 33089 | 703 | 32386 | 0.7196 | 0.06762 | Hypercalcemia | endocrine/metabolic | RATE_C | |
| 276 | 1.662 | 0.04063 | 5.268 | 0 | 24148 | 8143 | 16005 | 1.723 | 0.03508 | 5.604 | 0 | 32644 | 8450 | 24194 | 0.002866 | 0.153 | Disorders of fluid, electrolyte, and acid-base balance | endocrine/metabolic | RATE_C | |
| 276.1 | 1.824 | 0.04854 | 6.198 | 0 | 22066 | 6061 | 16005 | 1.849 | 0.04207 | 6.357 | 0 | 30513 | 6319 | 24194 | 0.00553 | 0.172 | Electrolyte imbalance | endocrine/metabolic | RATE_C | |
| 276.11 | | | | | 16449 | 444 | 16005 | | | | | 24509 | 315 | 24194 | 0.4891 | 0.158 | Hyperosmolality and/or hypernatremia | endocrine/metabolic | RATE_C | |
| 276.12 | 1.819 | 0.07653 | 6.163 | 8.42e-125 | 18173 | 2168 | 16005 | 1.943 | 0.07503 | 6.977 | 8.60e-148 | 26321 | 2127 | 24194 | 4.49e-05 | 0.4238 | Hyposmolality and/or hyponatremia | endocrine/metabolic | RATE_C | |
| 276.13 | 2.026 | 0.0871 | 7.583 | 1.10e-119 | 18167 | 2162 | 16005 | 1.944 | 0.086 | 6.986 | 3.91e-113 | 25804 | 1610 | 24194 | 0.4781 | 0.08454 | Hyperpotassemia | endocrine/metabolic | RATE_C | |
| 276.14 | 2.032 | 0.07428 | 7.627 | 1.05e-164 | 18672 | 2667 | 16005 | 1.991 | 0.05582 | 7.325 | 9.55e-279 | 27767 | 3573 | 24194 | 0.2558 | 0.1019 | Hypopotassemia | endocrine/metabolic | RATE_C | |
| 276.4 | 2.109 | 0.0852 | 8.239 | 3.09e-135 | 18109 | 2104 | 16005 | 2.088 | 0.07758 | 8.066 | 1.67e-159 | 26022 | 1828 | 24194 | 0.5899 | 0.05992 | Acid-base balance disorder | endocrine/metabolic | RATE_C | |
| 276.41 | 2.139 | 0.0913 | 8.487 | 2.41e-121 | 17879 | 1874 | 16005 | 2.013 | 0.08076 | 7.489 | 3.36e-137 | 25798 | 1604 | 24194 | 0.7104 | -0.04364 | Acidosis | endocrine/metabolic | RATE_C | |
| 276.42 | | | | | 16255 | 250 | 16005 | | | | | 24456 | 262 | 24194 | 0.07365 | 0.549 | Alkalosis | endocrine/metabolic | RATE_C | |
| 276.5 | 1.618 | 0.05865 | 5.043 | 1.53e-167 | 19178 | 3173 | 16005 | 1.768 | 0.0513 | 5.862 | 1.78e-260 | 27587 | 3393 | 24194 | 0.005507 | 0.2044 | Hypovolemia | endocrine/metabolic | RATE_C | |
| 276.6 | 2.447 | 0.1146 | 11.55 | 4.48e-101 | 17709 | 1704 | 16005 | 2.346 | 0.1036 | 10.44 | 1.38e-113 | 25588 | 1394 | 24194 | 0.9725 | -0.005217 | Fluid overload | endocrine/metabolic | RATE_C | |
| 276.8 | | | | | 16014 | 9 | 16005 | | | | | 24216 | 22 | 24194 | | | Polydipsia | endocrine/metabolic | RATE_C | |
| 277 | | | | | 26131 | 420 | 25711 | 0.9602 | 0.1107 | 2.612 | 4.10e-18 | 35099 | 554 | 34545 | 6.57e-04 | 0.5277 | Other disorders of metabolism | endocrine/metabolic | RATE_C | |
| 277.1 | | | | | 25722 | 11 | 25711 | | | | | 34554 | 9 | 34545 | | | Disorders of porphyrin metabolism | endocrine/metabolic | RATE_C | |
| 277.2 | | | | | 25713 | 2 | 25711 | | | | | 34547 | 2 | 34545 | | | Other disorders of purine and pyrimidine metabolism | endocrine/metabolic | RATE_C | |
| 277.4 | | | | | 25910 | 199 | 25711 | | | | | 34665 | 120 | 34545 | | | Disorders of bilirubin excretion | endocrine/metabolic | RATE_C | |
| 277.5 | | | | | 25845 | 134 | 25711 | | | | | 34656 | 111 | 34545 | | | Other disorders of lipoid metabolism | endocrine/metabolic | RATE_C | |
| 277.51 | | | | | 25790 | 79 | 25711 | | | | | 34590 | 45 | 34545 | | | Lipoprotein disorders | endocrine/metabolic | RATE_C | |
| 277.6 | | | | | 25723 | 12 | 25711 | | | | | 34560 | 15 | 34545 | | | Other deficiencies of circulating enzymes | endocrine/metabolic | RATE_C | |
| 277.7 | | | | | 25918 | 207 | 25711 | | | | | 34828 | 283 | 34545 | | | Dysmetabolic syndrome X | endocrine/metabolic | RATE_C | |
| 277.8 | | | | | 25715 | 4 | 25711 | | | | | 34546 | 1 | 34545 | | | Carnitine deficiencies | endocrine/metabolic | RATE_C | |
| 278 | 0.9342 | 0.04955 | 2.545 | 2.75e-79 | 24739 | 3939 | 20800 | 0.8809 | 0.03343 | 2.413 | 5.41e-153 | 32627 | 6840 | 25787 | 0.001728 | -0.1723 | Overweight, obesity and other hyperalimentation | endocrine/metabolic | RATE_C | |
| phenotype | male_beta | male_SE | male_OR | male_p | male_n_total | male_n_cases | male_n_controls | female_beta | female_SE | female_OR | female_p | female_n_total | female_n_cases | female_n_controls | interaction_p | interaction_beta | description | group | lab | label |
| 278.1 | 0.9833 | 0.05411 | 2.673 | 8.48e-74 | 24116 | 3316 | 20800 | 0.9465 | 0.03671 | 2.577 | 1.36e-146 | 31306 | 5519 | 25787 | 0.01174 | -0.1517 | Obesity | endocrine/metabolic | RATE_C | |
| 278.11 | 1.15 | 0.07801 | 3.16 | 3.21e-49 | 22371 | 1571 | 20800 | 1.101 | 0.04714 | 3.006 | 1.42e-120 | 28985 | 3198 | 25787 | 0.08526 | -0.145 | Morbid obesity | endocrine/metabolic | RATE_C | |
| 278.3 | | | | | 20837 | 37 | 20800 | | | | | 25988 | 201 | 25787 | | | Localized adiposity | endocrine/metabolic | RATE_C | |
| 278.4 | | | | | 21218 | 418 | 20800 | 0.6905 | 0.06355 | 1.995 | 1.69e-27 | 27263 | 1476 | 25787 | 0.3792 | 0.1121 | Abnormal weight gain | endocrine/metabolic | RATE_C | |
| 279 | 0.8596 | 0.09159 | 2.362 | 6.28e-21 | 25738 | 1171 | 24567 | 0.6524 | 0.06793 | 1.92 | 7.59e-22 | 33388 | 1505 | 31883 | 0.4554 | -0.07999 | Disorders involving the immune mechanism | endocrine/metabolic | RATE_C | |

| phenotype | male_beta | male_SE | male_OR | male_p | male_n_total | male_n_cases | male_n_controls | female_beta | female_SE | female_OR | female_p | female_n_total | female_n_cases | female_n_controls | interaction_p | interaction_beta | description | group | lab | label |
|---|---|---|---|---|---|---|---|---|---|---|---|---|---|---|---|---|---|---|---|---|
| 279.1 | 0.8675 | 0.09526 | 2.381 | 8.43e-20 | 25668 | 1101 | 24567 | 0.6933 | 0.07189 | 2 | 5.18e-22 | 33274 | 1391 | 31883 | 0.7881 | -0.03023 | Immunity deficiency | endocrine/metabolic | RATE_C | |
| 279.11 | | | | | 24666 | 99 | 24567 | | | | | 32052 | 169 | 31883 | | | Deficiency of humoral immunity | endocrine/metabolic | RATE_C | |
| 279.2 | | | | | 24578 | 11 | 24567 | | | | | 31914 | 31 | 31883 | | | Autoimmune disease NEC | endocrine/metabolic | RATE_C | |
| 279.7 | | | | | 24823 | 256 | 24567 | -0.1154 | 0.07348 | 0.891 | 0.1164 | 32824 | 941 | 31883 | 0.2154 | -0.1925 | Other immunological findings | endocrine/metabolic | RATE_C | |
| 279.8 | | | | | 24598 | 31 | 24567 | | | | | 31935 | 52 | 31883 | | | Other specified disorders involving the immune mechanism | endocrine/metabolic | RATE_C | |
| 280 | 1.204 | 0.07668 | 3.333 | 1.48e-55 | 18083 | 1688 | 16395 | 1.011 | 0.04971 | 2.747 | 7.19e-92 | 25891 | 2832 | 23059 | 0.01354 | -0.2139 | Iron deficiency anemias | hematopoietic | RATE_C | |
| 280.1 | 1.238 | 0.08638 | 3.45 | 1.29e-46 | 17740 | 1345 | 16395 | 1.028 | 0.05392 | 2.796 | 4.63e-81 | 25454 | 2395 | 23059 | 0.02063 | -0.2237 | Iron deficiency anemias, unspecified or not due to blood loss | hematopoietic | RATE_C | |
| 280.2 | 1.254 | 0.1425 | 3.504 | 1.36e-18 | 16909 | 514 | 16395 | 1.037 | 0.09298 | 2.821 | 6.82e-29 | 23821 | 762 | 23059 | 0.03155 | -0.3491 | Iron deficiency anemia secondary to blood loss (chronic) | hematopoietic | RATE_C | |
| 281 | | | | | 16870 | 475 | 16395 | 1.05 | 0.08918 | 2.858 | 5.33e-32 | 24011 | 952 | 23059 | 0.3023 | 0.1631 | Other deficiency anemia | hematopoietic | RATE_C | |
| 281.1 | | | | | 16669 | 274 | 16395 | 0.9588 | 0.1035 | 2.608 | 2.00e-20 | 23736 | 677 | 23059 | 0.1661 | 0.2664 | Megaloblastic anemia | hematopoietic | RATE_C | |
| 281.11 | | | | | 16464 | 69 | 16395 | | | | | 23278 | 219 | 23059 | | | Pernicious anemia | hematopoietic | RATE_C | |
| 281.12 | | | | | 16590 | 195 | 16395 | 1.036 | 0.1211 | 2.818 | 1.17e-17 | 23561 | 502 | 23059 | 0.6812 | 0.09812 | Other vitamin B12 deficiency anemia | hematopoietic | RATE_C | |
| 281.13 | | | | | 16415 | 20 | 16395 | | | | | 23107 | 48 | 23059 | | | Folate-deficiency anemia | hematopoietic | RATE_C | |
| 281.9 | | | | | 16581 | 186 | 16395 | | | | | 23292 | 233 | 23059 | | | Deficiency anemias | hematopoietic | RATE_C | |
| 282 | | | | | 16442 | 47 | 16395 | | | | | 23190 | 131 | 23059 | | | Hereditary hemolytic anemias | hematopoietic | RATE_C | |
| 282.5 | | | | | 16399 | 4 | 16395 | | | | | 23069 | 10 | 23059 | | | Sickle cell anemia | hematopoietic | RATE_C | |
| 282.8 | | | | | 16410 | 15 | 16395 | | | | | 23102 | 43 | 23059 | | | Other hemoglobinopathies | hematopoietic | RATE_C | |
| 282.9 | | | | | 16427 | 32 | 16395 | | | | | 23140 | 81 | 23059 | | | Other hereditary hemolytic anemias | hematopoietic | RATE_C | |
| 283 | | | | | 16455 | 60 | 16395 | | | | | 23168 | 109 | 23059 | | | Acquired hemolytic anemias | hematopoietic | RATE_C | |
| 283.1 | | | | | 16418 | 23 | 16395 | | | | | 23095 | 36 | 23059 | | | Autoimmune hemolytic anemias | hematopoietic | RATE_C | |
| 283.2 | | | | | 16419 | 24 | 16395 | | | | | 23100 | 41 | 23059 | | | Non-autoimmune hemolytic anemias | hematopoietic | RATE_C | |
| 283.21 | | | | | 16402 | 7 | 16395 | | | | | 23073 | 14 | 23059 | | | Hemolytic-uremic syndrome | hematopoietic | RATE_C | |
| 284 | 1.517 | 0.109 | 4.56 | 4.32e-44 | 17250 | 855 | 16395 | 1.557 | 0.1046 | 4.743 | 4.32e-50 | 23771 | 712 | 23059 | 0.6287 | 0.06985 | Aplastic anemia | hematopoietic | RATE_C | |
| 284.1 | 1.653 | 0.1216 | 5.221 | 4.34e-42 | 17146 | 751 | 16395 | 1.601 | 0.1121 | 4.956 | 3.12e-46 | 23702 | 643 | 23059 | 0.9848 | 0.003033 | Pancytopenia | hematopoietic | RATE_C | |
| 284.2 | | | | | 16406 | 11 | 16395 | | | | | 23065 | 6 | 23059 | | | Constitutional aplastic anemia | hematopoietic | RATE_C | |
| 285 | 1.414 | 0.0409 | 4.111 | 8.57e-262 | 23977 | 7582 | 16395 | 1.372 | 0.03403 | 3.942 | 0 | 30991 | 7932 | 23059 | 0.4258 | -0.04069 | Other anemias | hematopoietic | RATE_C | |
| 285.1 | 2.06 | 0.07775 | 7.843 | 1.33e-154 | 19293 | 2898 | 16395 | 2.037 | 0.07308 | 7.664 | 6.25e-171 | 25340 | 2281 | 23059 | 0.9467 | 0.006937 | Acute posthemorrhagic anemia | hematopoietic | RATE_C | |
| 285.2 | 1.474 | 0.07025 | 4.366 | 1.00e-97 | 18710 | 2315 | 16395 | 1.56 | 0.06313 | 4.759 | 8.44e-135 | 25252 | 2193 | 23059 | 0.3739 | 0.08107 | Anemia of chronic disease | hematopoietic | RATE_C | |
| 285.21 | 1.822 | 0.1359 | 6.183 | 5.44e-41 | 17283 | 888 | 16395 | 1.562 | 0.1162 | 4.769 | 3.32e-41 | 23775 | 716 | 23059 | 0.1905 | -0.2271 | Anemia in chronic kidney disease | hematopoietic | RATE_C | |

| phenotype | male_beta | male_SE | male_OR | male_p | male_n_total | male_n_cases | male_n_controls | female_beta | female_SE | female_OR | female_p | female_n_total | female_n_cases | female_n_controls | interaction_p | interaction_beta | description | group | lab | label |
|---|---|---|---|---|---|---|---|---|---|---|---|---|---|---|---|---|---|---|---|---|
| 285.22 | 1.159 | 0.08757 | 3.187 | 5.50e-40 | 17510 | 1115 | 16395 | 1.507 | 0.08071 | 4.515 | 7.67e-78 | 24273 | 1214 | 23059 | 0.003533 | 0.3353 | Anemia in neoplastic disease | hematopoietic | RATE_C | |
| 285.3 | | | | | 16401 | 6 | 16395 | | | | | 23062 | 3 | 23059 | | | Sideroblastic anemia | hematopoietic | RATE_C | |
| 285.8 | | | | | 16399 | 4 | 16395 | | | | | 23072 | 13 | 23059 | | | Hemoglobinuria | hematopoietic | RATE_C | |
| 286 | 1.665 | 0.08478 | 5.287 | 6.67e-86 | 21087 | 1817 | 19270 | 1.396 | 0.06392 | 4.039 | 1.03e-105 | 30311 | 1937 | 28374 | 0.00119 | -0.3297 | Coagulation defects | hematopoietic | RATE_C | |
| 286.1 | | | | | 19382 | 112 | 19270 | | | | | 28488 | 114 | 28374 | | | Congenital coagulation defects | hematopoietic | RATE_C | |
| 286.11 | | | | | 19306 | 36 | 19270 | | | | | 28422 | 48 | 28374 | | | Von willebrand's disease | hematopoietic | RATE_C | |
| 286.12 | | | | | 19305 | 35 | 19270 | | | | | 28432 | 58 | 28374 | | | Congenital deficiency of other clotting factors (including factor VII) | hematopoietic | RATE_C | |
| 286.13 | | | | | 19324 | 54 | 19270 | | | | | 28385 | 11 | 28374 | | | Congenital factor VIII disorder | hematopoietic | RATE_C | |
| 286.2 | 1.684 | 0.07528 | 5.385 | 8.82e-111 | 22410 | 3140 | 19270 | 1.782 | 0.07109 | 5.941 | 1.13e-138 | 31115 | 2741 | 28374 | 0.2549 | 0.1159 | Encounter for long-term (current) use of anticoagulants | hematopoietic | RATE_C | |
| 286.3 | | | | | 6 | | | | | | | 28478 | 106 | 28372 | | | Coagulation defects complicating pregnancy or postpartum | hematopoietic | RATE_C | |
| 286.4 | | | | | 19353 | 83 | 19270 | | | | | 28429 | 55 | 28374 | | | Acquired coagulation factor deficiency | hematopoietic | RATE_C | |
| 286.5 | | | | | 19388 | 118 | 19270 | | | | | 28511 | 137 | 28374 | | | Hemorrhagic disorder due to intrinsic circulating anticoagulants | hematopoietic | RATE_C | |
| 286.6 | | | | | 19312 | 42 | 19270 | | | | | 28420 | 46 | 28374 | | | Defibrination syndrome | hematopoietic | RATE_C | |
| 286.7 | 1.663 | 0.116 | 5.276 | 1.34e-46 | 20156 | 886 | 19270 | 1.531 | 0.0904 | 4.624 | 2.29e-64 | 29323 | 949 | 28374 | 0.07488 | -0.25 | Other and unspecified coagulation defects | hematopoietic | RATE_C | |
| 286.8 | | | | | 19460 | 190 | 19270 | | | | | 28789 | 415 | 28374 | 0.2369 | -0.2861 | Hypercoagulable state | hematopoietic | RATE_C | |
| 286.81 | | | | | 19445 | 175 | 19270 | | | | | 28747 | 373 | 28374 | 0.2565 | -0.2821 | Primary hypercoagulable state | hematopoietic | RATE_C | |
| 286.9 | | | | | 19741 | 471 | 19270 | | | | | 28784 | 410 | 28374 | 0.3253 | -0.2937 | Abnormal coagulation profile | hematopoietic | RATE_C | |
| 287 | 1.374 | 0.06795 | 3.95 | 6.82e-91 | 21610 | 2340 | 19270 | 1.353 | 0.06388 | 3.87 | 1.35e-99 | 30271 | 1897 | 28374 | 0.474 | -0.06396 | Purpura and other hemorrhagic conditions | hematopoietic | RATE_C | |
| 287.1 | | | | | 19326 | 56 | 19270 | | | | | 28485 | 111 | 28374 | | | Spontaneous ecchymoses | hematopoietic | RATE_C | |
| 287.2 | | | | | 19288 | 18 | 19270 | | | | | 28390 | 16 | 28374 | | | Allergic purpura | hematopoietic | RATE_C | |
| 287.3 | 1.409 | 0.07051 | 4.091 | 8.18e-89 | 21432 | 2162 | 19270 | 1.449 | 0.06805 | 4.257 | 1.45e-100 | 30102 | 1728 | 28374 | 0.976 | 0.002822 | Thrombocytopenia | hematopoietic | RATE_C | |
| 287.31 | | | | | 19391 | 121 | 19270 | | | | | 28538 | 164 | 28374 | | | Primary thrombocytopenia | hematopoietic | RATE_C | |
| 287.32 | | | | | 19594 | 324 | 19270 | | | | | 28675 | 301 | 28374 | 0.5197 | 0.133 | Secondary thrombocytopenia | hematopoietic | RATE_C | |
| 287.4 | | | | | 19290 | 20 | 19270 | | | | | 28393 | 19 | 28374 | | | Qualitative platelet defects | hematopoietic | RATE_C | |
| 288 | 1.596 | 0.06649 | 4.933 | 2.68e-127 | 21474 | 2790 | 18684 | 1.586 | 0.05785 | 4.884 | 1.87e-165 | 29549 | 2753 | 26796 | 0.2126 | 0.1041 | Diseases of white blood cells | hematopoietic | RATE_C | |
| 288.1 | 1.314 | 0.09279 | 3.719 | 1.72e-45 | 19788 | 1104 | 18684 | 1.257 | 0.07266 | 3.515 | 4.66e-67 | 28156 | 1360 | 26796 | 0.6467 | 0.05072 | Decreased white blood cell count | hematopoietic | RATE_C | |
| 288.11 | 1.373 | 0.1079 | 3.946 | 4.78e-37 | 19506 | 822 | 18684 | 1.419 | 0.08667 | 4.132 | 3.12e-60 | 27815 | 1019 | 26796 | 0.212 | 0.163 | Neutropenia | hematopoietic | RATE_C | |
| 288.2 | 1.709 | 0.07474 | 5.524 | 9.65e-116 | 21008 | 2324 | 18684 | 1.633 | 0.06425 | 5.118 | 1.97e-142 | 29055 | 2259 | 26796 | 0.5948 | 0.04985 | Elevated white blood cell count | hematopoietic | RATE_C | |

| phenotype | male_beta | male_SE | male_OR | male_p | male_n_total | male_n_cases | male_n_controls | female_beta | female_SE | female_OR | female_p | female_n_total | female_n_cases | female_n_controls | interaction_p | interaction_beta | description | group | lab | label |
|---|---|---|---|---|---|---|---|---|---|---|---|---|---|---|---|---|---|---|---|---|
| 288.3 | | | | | 18778 | 94 | 18684 | | | | | 26909 | 113 | 26796 | | | Eosinophilia | hematopoietic | RATE_C | |
| 289 | | | | | 19172 | 488 | 18684 | 0.5146 | 0.09526 | 1.673 | 6.59e-08 | 27421 | 625 | 26796 | 0.2508 | 0.1602 | Other diseases of blood and blood-forming organs | hematopoietic | RATE_C | |
| 289.1 | | | | | 18755 | 71 | 18684 | | | | | 26856 | 60 | 26796 | | | Myelofibrosis | hematopoietic | RATE_C | |
| 289.3 | | | | | 18767 | 83 | 18684 | | | | | 26919 | 123 | 26796 | | | Personal history of diseases of blood and blood-forming organs | hematopoietic | RATE_C | |
| 289.4 | 1.045 | 0.07633 | 2.843 | 1.20e-42 | 20171 | 1487 | 18684 | 1.072 | 0.0641 | 2.922 | 7.80e-63 | 28443 | 1647 | 26796 | 0.5302 | 0.05838 | Lymphadenitis | hematopoietic | RATE_C | |
| 289.5 | | | | | 18840 | 156 | 18684 | | | | | 26948 | 152 | 26796 | | | Diseases of spleen | hematopoietic | RATE_C | |
| 289.8 | | | | | 18685 | 237 | 18448 | | | | | 26687 | 169 | 26518 | | | Polycythemia, secondary | hematopoietic | RATE_C | |
| 289.9 | | | | | 18788 | 104 | 18684 | | | | | 26914 | 118 | 26796 | | | Abnormality of red blood cells | hematopoietic | RATE_C | |
| 290 | 0.9351 | 0.0959 | 2.547 | 1.84e-22 | 21683 | 1272 | 20411 | 1.07 | 0.09035 | 2.916 | 2.26e-32 | 30218 | 1222 | 28996 | 0.07629 | 0.2306 | Delirium dementia and amnestic and other cognitive disorders | mental disorders | RATE_C | |
| 290.1 | 0.412 | 0.1099 | 1.51 | 1.78e-04 | 21205 | 794 | 20411 | 0.7184 | 0.1031 | 2.051 | 3.16e-12 | 29834 | 838 | 28996 | 0.03333 | 0.3167 | Dementias | mental disorders | RATE_C | |
| 290.11 | | | | | 20682 | 271 | 20411 | | | | | 29352 | 356 | 28996 | 0.05872 | 0.396 | Alzheimer's disease | mental disorders | RATE_C | |
| 290.12 | | | | | 20506 | 95 | 20411 | | | | | 29043 | 47 | 28996 | | | Dementia with cerebral degenerations | mental disorders | RATE_C | |
| 290.13 | | | | | 20487 | 76 | 20411 | | | | | 29129 | 133 | 28996 | | | Senile dementia | mental disorders | RATE_C | |
| 290.16 | | | | | 20543 | 132 | 20411 | | | | | 29107 | 111 | 28996 | | | Vascular dementia | mental disorders | RATE_C | |
| 290.2 | | | | | 20877 | 466 | 20411 | | | | | 29341 | 345 | 28996 | 0.84 | 0.06574 | Delirium due to conditions classified elsewhere | mental disorders | RATE_C | |
| 290.3 | | | | | 20762 | 351 | 20411 | | | | | 29366 | 370 | 28996 | 0.108 | 0.4205 | Other persistent mental disorders due to conditions classified elsewhere | mental disorders | RATE_C | |
| 291 | 1.698 | 0.1023 | 5.464 | 7.37e-62 | 21730 | 1319 | 20411 | 1.599 | 0.08602 | 4.949 | 3.86e-77 | 30293 | 1297 | 28996 | 0.9036 | 0.01566 | Other specified nonpsychotic and/or transient mental disorders | mental disorders | RATE_C | |
| 291.1 | | | | | 20440 | 29 | 20411 | | | | | 29037 | 41 | 28996 | | | Transient mental disorders due to conditions classified elsewhere | mental disorders | RATE_C | |
| 291.4 | | | | | 20576 | 165 | 20411 | | | | | 29194 | 198 | 28996 | | | Specific nonpsychotic mental disorders due to brain damage | mental disorders | RATE_C | |
| 291.8 | 1.997 | 0.1255 | 7.365 | 5.62e-57 | 21537 | 1126 | 20411 | 2.016 | 0.112 | 7.508 | 1.83e-72 | 30045 | 1049 | 28996 | 0.4154 | 0.1339 | Alteration of consciousness | mental disorders | RATE_C | |
| 292 | 1.142 | 0.05374 | 3.133 | 3.17e-100 | 24322 | 3911 | 20411 | 1.228 | 0.04637 | 3.414 | 1.60e-154 | 33173 | 4177 | 28996 | 0.005122 | 0.1915 | Neurological disorders | mental disorders | RATE_C | |
| 292.1 | 1.22 | 0.1204 | 3.389 | 3.75e-24 | 21162 | 751 | 20411 | 1.554 | 0.1178 | 4.73 | 1.07e-39 | 29761 | 765 | 28996 | 0.001971 | 0.5042 | Aphasia/speech disturbance | mental disorders | RATE_C | |
| 292.11 | | | | | 20684 | 273 | 20411 | | | | | 29270 | 274 | 28996 | 0.01214 | 0.7513 | Aphasia | mental disorders | RATE_C | |
| 292.12 | | | | | 20439 | 28 | 20411 | | | | | 29020 | 24 | 28996 | | | Symbolic dysfunction | mental disorders | RATE_C | |
| 292.2 | | | | | 20778 | 367 | 20411 | | | | | 29334 | 338 | 28996 | 0.3404 | 0.2136 | Mild cognitive impairment | mental disorders | RATE_C | |
| 292.3 | 0.5071 | 0.09797 | 1.66 | 2.27e-07 | 21327 | 916 | 20411 | 0.5689 | 0.07826 | 1.766 | 3.61e-13 | 30127 | 1131 | 28996 | 0.3116 | 0.1229 | Memory loss | mental disorders | RATE_C | |
| 292.4 | 1.685 | 0.0808 | 5.393 | 1.35e-96 | 22762 | 2351 | 20411 | 1.812 | 0.07433 | 6.122 | 3.05e-131 | 31254 | 2258 | 28996 | 0.02071 | 0.2476 | Altered mental status | mental disorders | RATE_C | |
| 292.5 | | | | | 20547 | 136 | 20411 | | | | | 29184 | 188 | 28996 | | | Transient alteration of awareness | mental disorders | RATE_C | |
| 292.6 | | | | | 20491 | 80 | 20411 | | | | | 29129 | 133 | 28996 | | | Hallucinations | mental disorders | RATE_C | |

| phenotype | male_beta | male_SE | male_OR | male_p | male_n_total | male_n_cases | male_n_controls | female_beta | female_SE | female_OR | female_p | female_n_total | female_n_cases | female_n_controls | interaction_p | interaction_beta | description | group | lab | label |
|---|---|---|---|---|---|---|---|---|---|---|---|---|---|---|---|---|---|---|---|---|
| 293 | | | | | 25542 | 80 | 25462 | | | | | 34286 | 108 | 34178 | | | Symptoms involving head and neck | mental disorders | RATE_C | |
| 293.1 | 0.8772 | 0.09856 | 2.404 | 5.59e-19 | 26351 | 889 | 25462 | 1.035 | 0.09125 | 2.815 | 8.26e-30 | 35053 | 875 | 34178 | 0.05004 | 0.2468 | Swelling, mass, or lump in head and neck [Space-occupying lesion, intracranial NOS] | mental disorders | RATE_C | |
| 295 | | | | | 17073 | 479 | 16594 | 1.611 | 0.1221 | 5.006 | 9.31e-40 | 19345 | 524 | 18821 | 1.89e-04 | 0.6193 | Schizophrenia and other psychotic disorders | mental disorders | RATE_C | |
| 295.1 | | | | | 16740 | 146 | 16594 | | | | | 18981 | 160 | 18821 | | | Schizophrenia | mental disorders | RATE_C | |
| 295.2 | | | | | 16700 | 106 | 16594 | | | | | 18968 | 147 | 18821 | | | Paranoid disorders | mental disorders | RATE_C | |
| 295.3 | | | | | 16878 | 284 | 16594 | | | | | 19094 | 273 | 18821 | 0.03661 | 0.4704 | Psychosis | mental disorders | RATE_C | |
| 296 | 1.011 | 0.04613 | 2.747 | 2.30e-106 | 21203 | 4609 | 16594 | 1.125 | 0.03304 | 3.081 | 2.49e-254 | 27169 | 8348 | 18821 | 3.78e-07 | 0.2595 | Mood disorders | mental disorders | RATE_C | |
| 296.1 | 0.9588 | 0.09429 | 2.609 | 2.73e-24 | 17404 | 810 | 16594 | 1.372 | 0.0698 | 3.944 | 4.96e-86 | 20203 | 1382 | 18821 | 1.91e-06 | 0.4953 | Bipolar | mental disorders | RATE_C | |
| 296.2 | 1.05 | 0.05017 | 2.859 | 2.40e-97 | 20515 | 3921 | 16594 | 1.122 | 0.03461 | 3.069 | 2.32e-230 | 26233 | 7412 | 18821 | 3.48e-04 | 0.1974 | Depression | mental disorders | RATE_C | |
| 296.22 | 0.9434 | 0.06377 | 2.569 | 1.63e-49 | 18825 | 2231 | 16594 | 1.07 | 0.04269 | 2.916 | 1.02e-138 | 23232 | 4411 | 18821 | 0.002586 | 0.207 | Major depressive disorder | mental disorders | RATE_C | |
| 297 | | | | | 17005 | 411 | 16594 | 1.329 | 0.1067 | 3.778 | 1.30e-35 | 19321 | 500 | 18821 | 0.1922 | -0.1944 | Suicidal ideation or attempt | mental disorders | RATE_C | |
| 297.1 | | | | | 16944 | 350 | 16594 | | | | | 19222 | 401 | 18821 | 0.31 | -0.1627 | Suicidal ideation | mental disorders | RATE_C | |
| 297.2 | | | | | 16666 | 72 | 16594 | | | | | 18942 | 121 | 18821 | | | Suicide or self-inflicted injury | mental disorders | RATE_C | |
| 300 | 0.8601 | 0.04749 | 2.363 | 2.61e-73 | 20603 | 4009 | 16594 | 1.053 | 0.03309 | 2.865 | 5.05e-222 | 27035 | 8214 | 18821 | 6.76e-11 | 0.3358 | Anxiety disorders | mental disorders | RATE_C | Anxiety disorders |
| 300.1 | 0.8268 | 0.05016 | 2.286 | 4.85e-61 | 20057 | 3463 | 16594 | 1.062 | 0.03477 | 2.893 | 4.28e-205 | 26048 | 7227 | 18821 | 1.20e-12 | 0.3865 | Anxiety disorder | mental disorders | RATE_C | |
| 300.11 | 0.2965 | 0.09501 | 1.345 | 0.001806 | 17284 | 690 | 16594 | 0.8735 | 0.06389 | 2.395 | 1.51e-42 | 20323 | 1502 | 18821 | 4.21e-09 | 0.5933 | Generalized anxiety disorder | mental disorders | RATE_C | |
| 300.12 | | | | | 16882 | 288 | 16594 | 1.411 | 0.1039 | 4.102 | 4.65e-42 | 19405 | 584 | 18821 | 1.31e-04 | 0.6177 | Agorophobia, social phobia, and panic disorder | mental disorders | RATE_C | |
| 300.13 | | | | | 16650 | 56 | 16594 | | | | | 18930 | 109 | 18821 | | | Phobia | mental disorders | RATE_C | |
| 300.2 | | | | | 16594 | | | | | | | 18821 | | | | | Generalized anxiety & phobic disorders | mental disorders | RATE_C | |
| 300.3 | | | | | 16794 | 200 | 16594 | | | | | 19097 | 276 | 18821 | | | Obsessive-compulsive disorders | mental disorders | RATE_C | |
| 300.4 | | | | | 16995 | 401 | 16594 | 1.382 | 0.09507 | 3.982 | 7.27e-48 | 19615 | 794 | 18821 | 0.04371 | 0.3261 | Dysthymic disorder | mental disorders | RATE_C | |
| 300.8 | | | | | 16639 | 45 | 16594 | | | | | 18944 | 123 | 18821 | | | Acute reaction to stress | mental disorders | RATE_C | |
| 300.9 | | | | | 16936 | 342 | 16594 | 1.388 | 0.08109 | 4.008 | 1.07e-65 | 19768 | 947 | 18821 | 0.4025 | -0.1257 | Posttraumatic stress disorder | mental disorders | RATE_C | |
| 301 | | | | | 16724 | 130 | 16594 | | | | | 19101 | 280 | 18821 | | | Personality disorders | mental disorders | RATE_C | |
| 301.1 | | | | | 16605 | 11 | 16594 | | | | | 18829 | 8 | 18821 | | | Schizoid personality disorder | mental disorders | RATE_C | |
| 301.2 | | | | | 16645 | 51 | 16594 | | | | | 19000 | 179 | 18821 | | | Antisocial/borderline personality disorder | mental disorders | RATE_C | |
| 302 | | | | | 16787 | 193 | 16594 | | | | | 18953 | 132 | 18821 | | | Sexual and gender identity disorders | mental disorders | RATE_C | |
| 302.1 | | | | | 16728 | 134 | 16594 | | | | | 18905 | 84 | 18821 | | | Decreased libido | mental disorders | RATE_C | |

| phenotype | male_beta | male_SE | male_OR | male_p | male_n_total | male_n_cases | male_n_controls | female_beta | female_SE | female_OR | female_p | female_n_total | female_n_cases | female_n_controls | interaction_p | interaction_beta | description | group | lab | label |
|---|---|---|---|---|---|---|---|---|---|---|---|---|---|---|---|---|---|---|---|---|
| 303 | | | | | 16730 | 136 | 16594 | | | | | 19170 | 349 | 18821 | | | Psychogenic and somatoform disorders | mental disorders | RATE_C | |
| 303.1 | | | | | 16603 | 9 | 16594 | | | | | 18849 | 28 | 18821 | | | Dissociative disorder | mental disorders | RATE_C | |
| 303.3 | | | | | 16631 | 37 | 16594 | | | | | 18918 | 97 | 18821 | | | Psychogenic disorder | mental disorders | RATE_C | |
| 303.31 | | | | | 16597 | 3 | 16594 | | | | | 18830 | 9 | 18821 | | | Gastrointestinal malfunction arising from mental factors | mental disorders | RATE_C | |
| 303.4 | | | | | 16689 | 95 | 16594 | | | | | 19064 | 243 | 18821 | | | Somatoform disorder | mental disorders | RATE_C | |
| 304 | 0.9749 | 0.0895 | 2.651 | 1.25e-27 | 17547 | 953 | 16594 | 1.045 | 0.06062 | 2.845 | 1.18e-66 | 20609 | 1788 | 18821 | 0.02819 | 0.2125 | Adjustment reaction | mental disorders | RATE_C | |
| 305.2 | | | | | 16644 | 50 | 16594 | | | | | 19240 | 419 | 18821 | | | Eating disorder | mental disorders | RATE_C | |
| 305.21 | | | | | 16607 | 13 | 16594 | | | | | 18929 | 108 | 18821 | | | Anorexia nervosa | mental disorders | RATE_C | |
| 306 | 0.7655 | 0.0892 | 2.15 | 9.32e-18 | 17668 | 1074 | 16594 | 0.8796 | 0.06329 | 2.41 | 6.39e-44 | 20481 | 1660 | 18821 | 0.462 | 0.07346 | Other mental disorder | mental disorders | RATE_C | |
| 306.1 | | | | | 0 | | | | | | | 19216 | 395 | 18821 | | | Mental disorders durring/after pregnancy | mental disorders | RATE_C | |
| 306.9 | | | | | 16626 | 32 | 16594 | | | | | 18928 | 107 | 18821 | | | Tension headache | mental disorders | RATE_C | |
| 312 | | | | | 25880 | 213 | 25667 | | | | | 34967 | 157 | 34810 | | | Conduct disorders | mental disorders | RATE_C | |
| 312.3 | | | | | 25729 | 62 | 25667 | | | | | 34851 | 41 | 34810 | | | Impulse control disorder | mental disorders | RATE_C | |
| 313 | 0.1228 | 0.09669 | 1.131 | 0.2041 | 26378 | 711 | 25667 | 0.2245 | 0.08584 | 1.252 | 0.00892 | 35536 | 726 | 34810 | 5.90e-04 | 0.3888 | Pervasive developmental disorders | mental disorders | RATE_C | |
| 313.1 | 0.05331 | 0.1116 | 1.055 | 0.633 | 26178 | 511 | 25667 | 0.2369 | 0.09849 | 1.267 | 0.01615 | 35352 | 542 | 34810 | 0.001307 | 0.4196 | Attention deficit hyperactivity disorder | mental disorders | RATE_C | |
| 313.2 | | | | | 25726 | 59 | 25667 | | | | | 34849 | 39 | 34810 | | | Tics and stuttering | mental disorders | RATE_C | |
| 313.3 | | | | | 25854 | 187 | 25667 | | | | | 34891 | 81 | 34810 | | | Autism | mental disorders | RATE_C | |
| 315 | 0.4648 | 0.1038 | 1.592 | 7.56e-06 | 26237 | 570 | 25667 | | | | | 35246 | 436 | 34810 | 0.1118 | 0.2131 | Develomental delays and disorders | mental disorders | RATE_C | |
| 315.1 | | | | | 25693 | 26 | 25667 | | | | | 34830 | 20 | 34810 | | | Learning disorder | mental disorders | RATE_C | |
| 315.2 | | | | | 25753 | 86 | 25667 | | | | | 34881 | 71 | 34810 | | | Speech and language disorder | mental disorders | RATE_C | |
| 315.3 | | | | | 25849 | 182 | 25667 | | | | | 34981 | 171 | 34810 | | | Mental retardation | mental disorders | RATE_C | |
| 316 | 1.173 | 0.07614 | 3.231 | 1.57e-53 | 19622 | 1399 | 18223 | 1.342 | 0.06329 | 3.828 | 7.64e-100 | 29618 | 1741 | 27877 | 0.02403 | 0.2014 | Substance addiction and disorders | mental disorders | RATE_C | |
| 316.1 | | | | | 18355 | 132 | 18223 | | | | | 28169 | 292 | 27877 | | | Polyneuropathy due to drugs | mental disorders | RATE_C | |
| 317 | 1.12 | 0.07863 | 3.066 | 4.56e-46 | 19570 | 1347 | 18223 | 1.171 | 0.09947 | 3.224 | 5.67e-32 | 28525 | 648 | 27877 | 0.9813 | -0.002721 | Alcohol-related disorders | mental disorders | RATE_C | |
| 317.1 | 1.202 | 0.09321 | 3.326 | 4.98e-38 | 19193 | 970 | 18223 | | | | | 28321 | 444 | 27877 | 0.5378 | 0.08801 | Alcoholism | mental disorders | RATE_C | |
| 317.11 | | | | | 18678 | 455 | 18223 | | | | | 28037 | 160 | 27877 | 0.09081 | 0.4163 | Alcoholic liver damage | mental disorders | RATE_C | |
| 318 | 1.043 | 0.04465 | 2.836 | 1.34e-120 | 23438 | 5215 | 18223 | 1.097 | 0.03999 | 2.996 | 9.73e-166 | 32604 | 4727 | 27877 | 0.186 | -0.07372 | Tobacco use disorder | mental disorders | RATE_C | |
| 320 | | | | | 27253 | 157 | 27096 | | | | | 36416 | 155 | 36261 | | | Meningitis | neurological | RATE_C | |

| phenotype | male_beta | male_SE | male_OR | male_p | male_n_total | male_n_cases | male_n_controls | female_beta | female_SE | female_OR | female_p | female_n_total | female_n_cases | female_n_controls | interaction_p | interaction_beta | description | group | lab | label |
|---|---|---|---|---|---|---|---|---|---|---|---|---|---|---|---|---|---|---|---|---|
| 323 | | | | | 27190 | 94 | 27096 | | | | | 36377 | 116 | 36261 | | | Encephalitis | neurological | RATE_C | |
| 323.2 | | | | | 27111 | 15 | 27096 | | | | | 36289 | 28 | 36261 | | | Acute (transverse) myelitis | neurological | RATE_C | |
| 323.8 | | | | | 27165 | 69 | 27096 | | | | | 36347 | 86 | 36261 | | | Encephalitis, non-infectious | neurological | RATE_C | |
| 324 | | | | | 27283 | 187 | 27096 | | | | | 36381 | 120 | 36261 | | | Other CNS infection and poliomyelitis | neurological | RATE_C | |
| 324.1 | | | | | 27097 | 1 | 27096 | | | | | 36261 | | | | | Jakob-Creutzfeldt disease | neurological | RATE_C | |
| 325 | | | | | 27112 | 16 | 27096 | | | | | 36306 | 45 | 36261 | | | Phlebitis and thrombophlebitis of intracranial venous sinuses | neurological | RATE_C | |
| 327 | 0.6201 | 0.05188 | 1.859 | 6.22e-33 | 22679 | 3201 | 19478 | 0.6892 | 0.03933 | 1.992 | 9.78e-69 | 31789 | 4772 | 27017 | 1.88e-04 | 0.2255 | Sleep disorders | neurological | RATE_C | |
| 327.1 | | | | | 19663 | 185 | 19478 | | | | | 27358 | 341 | 27017 | 0.3776 | 0.1784 | Hypersomnia | neurological | RATE_C | |
| 327.3 | 0.8989 | 0.05129 | 2.457 | 9.14e-69 | 23346 | 3868 | 19478 | 1.08 | 0.0499 | 2.945 | 6.17e-104 | 30225 | 3208 | 27017 | 6.46e-04 | 0.2288 | Sleep apnea | neurological | RATE_C | |
| 327.31 | | | | | 19588 | 110 | 19478 | | | | | 27080 | 63 | 27017 | | | Central/nonobstructive sleep apnea | neurological | RATE_C | |
| 327.32 | 0.9172 | 0.05508 | 2.502 | 2.94e-62 | 22855 | 3377 | 19478 | 1.116 | 0.05479 | 3.053 | 3.33e-92 | 29718 | 2701 | 27017 | 5.69e-04 | 0.2517 | Obstructive sleep apnea | neurological | RATE_C | |
| 327.4 | 0.6692 | 0.06503 | 1.953 | 7.73e-25 | 21429 | 1951 | 19478 | 0.6818 | 0.04745 | 1.977 | 8.24e-47 | 30146 | 3129 | 27017 | 0.006457 | 0.2046 | Insomnia | neurological | RATE_C | |
| 327.41 | | | | | 19695 | 217 | 19478 | | | | | 27448 | 431 | 27017 | 0.05426 | 0.4017 | Organic or persistent insomnia | neurological | RATE_C | |
| 327.5 | | | | | 19615 | 137 | 19478 | | | | | 27116 | 99 | 27017 | | | Parasomnia | neurological | RATE_C | |
| 327.6 | | | | | 19505 | 27 | 19478 | | | | | 27055 | 38 | 27017 | | | Circadian rhythm sleep disorder | neurological | RATE_C | |
| 327.7 | 0.7728 | 0.1333 | 2.166 | 6.72e-09 | 19996 | 518 | 19478 | 0.7838 | 0.08332 | 2.19 | 5.10e-21 | 28011 | 994 | 27017 | 0.9059 | 0.01762 | Sleep related movement disorders | neurological | RATE_C | |
| 327.71 | | | | | 19923 | 445 | 19478 | 0.8227 | 0.0896 | 2.277 | 4.24e-20 | 27896 | 879 | 27017 | 0.7054 | -0.06297 | Restless legs syndrome | neurological | RATE_C | |
| 327.72 | | | | | 19495 | 17 | 19478 | | | | | 27043 | 26 | 27017 | | | Sleep related leg cramps | neurological | RATE_C | |
| 331 | 0.8385 | 0.122 | 2.313 | 6.39e-12 | 20649 | 514 | 20135 | | | | | 28193 | 478 | 27715 | 0.02247 | 0.3623 | Other cerebral degenerations | neurological | RATE_C | |
| 331.1 | | | | | 20475 | 340 | 20135 | | | | | 28036 | 321 | 27715 | 0.01119 | 0.4509 | Hydrocephalus | neurological | RATE_C | |
| 331.9 | | | | | 20281 | 146 | 20135 | | | | | 27836 | 121 | 27715 | | | Cerebral degeneration, unspecified | neurological | RATE_C | |
| 332 | | | | | 20573 | 438 | 20135 | | | | | 27974 | 259 | 27715 | 0.8253 | -0.0498 | Parkinson's disease | neurological | RATE_C | |
| 333 | 0.6582 | 0.1021 | 1.931 | 1.13e-10 | 20955 | 820 | 20135 | 0.5999 | 0.0843 | 1.822 | 1.11e-12 | 28634 | 919 | 27715 | 0.6212 | 0.06199 | Extrapyramidal disease and abnormal movement disorders | neurological | RATE_C | |
| 333.1 | | | | | 20501 | 366 | 20135 | | | | | 28134 | 419 | 27715 | 0.8824 | 0.02962 | Essential tremor | neurological | RATE_C | |
| 333.2 | | | | | 20258 | 123 | 20135 | | | | | 27820 | 105 | 27715 | | | Myoclonus | neurological | RATE_C | |
| 333.3 | | | | | 20164 | 29 | 20135 | | | | | 27748 | 33 | 27715 | | | Tics and choreas | neurological | RATE_C | |
| 333.4 | | | | | 20284 | 149 | 20135 | | | | | 28005 | 290 | 27715 | | | Torsion dystonia | neurological | RATE_C | |
| 333.8 | | | | | 20218 | 83 | 20135 | | | | | 27746 | 31 | 27715 | | | Other degenerative diseases of the basal ganglia | neurological | RATE_C | |
| 334 | | | | | 20526 | 391 | 20135 | | | | | 28145 | 430 | 27715 | 0.00321 | 0.476 | Degenerative disease of the spinal cord | neurological | RATE_C | |

| phenotype | male_beta | male_SE | male_OR | male_p | male_n_total | male_n_cases | male_n_controls | female_beta | female_SE | female_OR | female_p | female_n_total | female_n_cases | female_n_controls | interaction_p | interaction_beta | description | group | lab | label |
|---|---|---|---|---|---|---|---|---|---|---|---|---|---|---|---|---|---|---|---|---|
| 334.1 | | | | | 20181 | 46 | 20135 | | | | | 27786 | 71 | 27715 | | | Spinocerebellar disease | neurological | RATE_C | |
| 334.2 | | | | | 20201 | 66 | 20135 | | | | | 27768 | 53 | 27715 | | | Anterior horn cell disease | neurological | RATE_C | |
| 334.21 | | | | | 20183 | 48 | 20135 | | | | | 27759 | 44 | 27715 | | | Amyotrophic Lateral Sclerosis | neurological | RATE_C | |
| 335 | | | | | 20445 | 310 | 20135 | -0.4274 | 0.0712 | 0.6522 | 1.94e-09 | 28694 | 979 | 27715 | 5.10e-05 | 0.5379 | Multiple sclerosis | neurological | RATE_C | |
| 337 | | | | | 20390 | 255 | 20135 | | | | | 28028 | 313 | 27715 | 0.542 | -0.1697 | Disorders of the autonomic nervous system | neurological | RATE_C | |
| 337.1 | | | | | 20203 | 68 | 20135 | | | | | 27793 | 78 | 27715 | | | Peripheral autonomic neuropathy | neurological | RATE_C | |
| 338 | 0.8831 | 0.04076 | 2.418 | 4.52e-104 | 24967 | 5863 | 19104 | 0.9502 | 0.03265 | 2.586 | 3.93e-186 | 33179 | 7691 | 25488 | 2.36e-05 | 0.2053 | Pain | neurological | RATE_C | |
| 338.1 | 1.02 | 0.04846 | 2.773 | 2.51e-98 | 23126 | 4022 | 19104 | 1.174 | 0.04076 | 3.234 | 2.75e-182 | 30282 | 4794 | 25488 | 1.92e-06 | 0.2807 | Acute pain | neurological | RATE_C | |
| 338.2 | 0.8258 | 0.05854 | 2.284 | 3.50e-45 | 21816 | 2712 | 19104 | 0.9039 | 0.04252 | 2.469 | 2.75e-100 | 29799 | 4311 | 25488 | 6.23e-04 | 0.2294 | Chronic pain | neurological | RATE_C | |
| 339 | 0.8172 | 0.1051 | 2.264 | 7.60e-15 | 25998 | 701 | 25297 | 0.8994 | 0.06666 | 2.458 | 1.77e-41 | 31803 | 1472 | 30331 | 0.1425 | 0.164 | Other headache syndromes | neurological | RATE_C | |
| 340 | 0.669 | 0.1013 | 1.952 | 3.98e-11 | 25980 | 683 | 25297 | 0.7025 | 0.04728 | 2.019 | 6.05e-50 | 33186 | 2855 | 30331 | 0.2516 | 0.1123 | Migraine | neurological | RATE_C | |
| 340.1 | | | | | 25434 | 137 | 25297 | 0.6956 | 0.08903 | 2.005 | 5.58e-15 | 31063 | 732 | 30331 | 0.4851 | -0.1556 | Migrain with aura | neurological | RATE_C | |
| 341 | | | | | 20276 | 141 | 20135 | | | | | 28206 | 491 | 27715 | 0.1395 | 0.285 | Other demyelinating diseases of central nervous system | neurological | RATE_C | |
| 342 | | | | | 20493 | 358 | 20135 | | | | | 28011 | 296 | 27715 | 0.004779 | 0.6213 | Hemiplegia | neurological | RATE_C | |
| 343 | | | | | 20279 | 144 | 20135 | | | | | 27844 | 129 | 27715 | | | Infantile cerebral palsy | neurological | RATE_C | |
| 344 | | | | | 20460 | 325 | 20135 | | | | | 27997 | 282 | 27715 | 0.04468 | 0.3881 | Other paralytic syndromes | neurological | RATE_C | |
| 345 | 0.8449 | 0.06733 | 2.328 | 4.07e-36 | 21729 | 1594 | 20135 | 1.125 | 0.05876 | 3.081 | 9.13e-82 | 29581 | 1866 | 27715 | 3.05e-08 | 0.4433 | Epilepsy, recurrent seizures, convulsions | neurological | RATE_C | |
| 345.1 | 0.6311 | 0.0941 | 1.88 | 1.99e-11 | 20847 | 712 | 20135 | 0.8925 | 0.0878 | 2.441 | 2.82e-24 | 28454 | 739 | 27715 | 1.29e-06 | 0.5514 | Epilepsy | neurological | RATE_C | |
| 345.11 | | | | | 20451 | 316 | 20135 | | | | | 28000 | 285 | 27715 | 0.007632 | 0.452 | Generalized convulsive epilepsy | neurological | RATE_C | |
| 345.12 | 0.4901 | 0.1095 | 1.632 | 7.55e-06 | 20638 | 503 | 20135 | | | | | 28179 | 464 | 27715 | 1.62e-06 | 0.6613 | Partial epilepsy | neurological | RATE_C | |
| 345.3 | 0.9659 | 0.07549 | 2.627 | 1.75e-37 | 21418 | 1283 | 20135 | 1.205 | 0.06602 | 3.336 | 2.14e-74 | 29200 | 1485 | 27715 | 2.04e-06 | 0.427 | Convulsions | neurological | RATE_C | |
| 346 | | | | | 20172 | 37 | 20135 | | | | | 27787 | 72 | 27715 | | | Abnormal findings on study of brain and/or nervous system | neurological | RATE_C | |
| 346.1 | | | | | 20298 | 163 | 20135 | | | | | 27984 | 269 | 27715 | | | Nonspecific abnormal findings on radiological and other examination of skull and head | neurological | RATE_C | |
| 346.2 | | | | | 20163 | 28 | 20135 | | | | | 27765 | 50 | 27715 | | | Nonspecific abnormal results of function study of brain and central nervous system | neurological | RATE_C | |
| 346.3 | | | | | 20173 | 38 | 20135 | | | | | 27758 | 43 | 27715 | | | Nonspecific abnormal findings in cerebrospinal fluid | neurological | RATE_C | |
| 347 | | | | | 20161 | 26 | 20135 | | | | | 27775 | 60 | 27715 | | | Cataplexy and narcolepsy | neurological | RATE_C | |
| 348 | 1.221 | 0.08092 | 3.39 | 1.97e-51 | 21644 | 1509 | 20135 | 1.473 | 0.07435 | 4.361 | 2.48e-87 | 29223 | 1508 | 27715 | 0.004121 | 0.297 | Other conditions of brain | neurological | RATE_C | |

| phenotype | male_beta | male_SE | male_OR | male_p | male_n_total | male_n_cases | male_n_controls | female_beta | female_SE | female_OR | female_p | female_n_total | female_n_cases | female_n_controls | interaction_p | interaction_beta | description | group | lab | label |
|---|---|---|---|---|---|---|---|---|---|---|---|---|---|---|---|---|---|---|---|---|
| 348.2 | 1.325 | 0.1327 | 3.761 | 1.78e-23 | 20647 | 512 | 20135 | 1.721 | 0.1172 | 5.592 | 7.70e-49 | 28310 | 595 | 27715 | 0.01613 | 0.4012 | Cerebral edema and compression of brain | neurological | RATE_C | |
| 348.4 | | | | | 20194 | 59 | 20135 | | | | | 27769 | 54 | 27715 | | | Cerebral cysts | neurological | RATE_C | |
| 348.7 | | | | | 20262 | 127 | 20135 | | | | | 27823 | 108 | 27715 | | | Coma | neurological | RATE_C | |
| 348.8 | 1.192 | 0.1051 | 3.294 | 7.96e-30 | 21082 | 947 | 20135 | 1.448 | 0.1013 | 4.255 | 2.21e-46 | 28610 | 895 | 27715 | 0.00912 | 0.3621 | Encephalopathy, not elsewhere classified | neurological | RATE_C | |
| 348.9 | 1.259 | 0.107 | 3.523 | 5.33e-32 | 20916 | 781 | 20135 | 1.592 | 0.1078 | 4.915 | 2.36e-49 | 28459 | 744 | 27715 | 1.86e-04 | 0.5354 | Other conditions of brain, NOS | neurological | RATE_C | |
| 349 | | | | | 20369 | 234 | 20135 | | | | | 27969 | 254 | 27715 | | | Other and unspecified disorders of the nervous system | neurological | RATE_C | |
| 350 | 0.3999 | 0.05533 | 1.492 | 4.91e-13 | 25783 | 2709 | 23074 | 0.4838 | 0.04447 | 1.622 | 1.45e-27 | 34411 | 3494 | 30917 | 0.001238 | 0.2162 | Abnormal movement | neurological | RATE_C | |
| 350.1 | 0.4116 | 0.09244 | 1.509 | 8.48e-06 | 23884 | 810 | 23074 | 0.6109 | 0.07552 | 1.842 | 5.98e-16 | 32013 | 1096 | 30917 | 1.71e-04 | 0.4153 | Abnormal involuntary movements | neurological | RATE_C | |
| 350.2 | 0.4649 | 0.07072 | 1.592 | 4.91e-11 | 24868 | 1794 | 23074 | 0.535 | 0.0559 | 1.707 | 1.06e-21 | 33227 | 2310 | 30917 | 0.06436 | 0.1599 | Abnormality of gait | neurological | RATE_C | |
| 350.3 | -0.02549 | 0.1051 | 0.9748 | 0.8084 | 23605 | 531 | 23074 | -0.09439 | 0.07712 | 0.9099 | 0.221 | 31754 | 837 | 30917 | 0.6421 | 0.05579 | Lack of coordination | neurological | RATE_C | |
| 350.5 | | | | | 23103 | 29 | 23074 | | | | | 30953 | 36 | 30917 | | | Abnormal reflex | neurological | RATE_C | |
| 350.6 | | | | | 23183 | 109 | 23074 | | | | | 31041 | 124 | 30917 | | | Disturbances of sensation of smell and taste | neurological | RATE_C | |
| 351 | 0.7873 | 0.08166 | 2.197 | 5.37e-22 | 25090 | 1362 | 23728 | 0.8079 | 0.05661 | 2.243 | 3.34e-46 | 32761 | 2244 | 30517 | 0.1442 | 0.1361 | Other peripheral nerve disorders | neurological | RATE_C | |
| 352 | | | | | 24026 | 298 | 23728 | 0.7589 | 0.1094 | 2.136 | 4.04e-12 | 31061 | 544 | 30517 | 0.6108 | 0.09543 | Disorders of other cranial nerves | neurological | RATE_C | |
| 352.1 | | | | | 23876 | 148 | 23728 | | | | | 30870 | 353 | 30517 | 0.7974 | -0.0676 | Trigeminal nerve disorders [CN5] | neurological | RATE_C | |
| 352.2 | | | | | 23861 | 133 | 23728 | | | | | 30696 | 179 | 30517 | | | Facial nerve disorders [CN7] | neurological | RATE_C | |
| 353 | | | | | 23928 | 200 | 23728 | | | | | 30746 | 229 | 30517 | | | Nerve root and plexus disorders | neurological | RATE_C | |
| 353.1 | | | | | 23805 | 77 | 23728 | | | | | 30605 | 88 | 30517 | | | Nerve plexus lesions | neurological | RATE_C | |
| 353.2 | | | | | 23800 | 72 | 23728 | | | | | 30603 | 86 | 30517 | | | Nerve root lesions | neurological | RATE_C | |
| 355 | | | | | 23869 | 141 | 23728 | | | | | 30825 | 308 | 30517 | | | Complex regional/central pain syndrome | neurological | RATE_C | |
| 355.1 | 0.8879 | 0.1272 | 2.43 | 2.89e-12 | 24316 | 588 | 23728 | 0.99 | 0.08685 | 2.691 | 4.22e-30 | 31540 | 1023 | 30517 | 0.085 | 0.2484 | Chronic pain syndrome | neurological | RATE_C | |
| 356 | 0.5637 | 0.09577 | 1.757 | 3.96e-09 | 24919 | 1003 | 23916 | 0.9763 | 0.0935 | 2.655 | 1.59e-25 | 33295 | 1002 | 32293 | 3.76e-04 | 0.4586 | Hereditary and idiopathic peripheral neuropathy | neurological | RATE_C | |
| 357 | 0.5328 | 0.09044 | 1.704 | 3.83e-09 | 24993 | 1077 | 23916 | 0.6355 | 0.0733 | 1.888 | 4.34e-18 | 33576 | 1283 | 32293 | 0.2895 | 0.117 | Inflammatory and toxic neuropathy | neurological | RATE_C | |
| 358 | | | | | 24025 | 109 | 23916 | | | | | 32462 | 169 | 32293 | | | Myoneural disorders | neurological | RATE_C | |
| 358.1 | | | | | 23987 | 71 | 23916 | | | | | 32384 | 91 | 32293 | | | Myasthenia gravis | neurological | RATE_C | |
| 359 | 1.487 | 0.1572 | 4.423 | 3.08e-21 | 24562 | 646 | 23916 | 1.332 | 0.1295 | 3.788 | 8.20e-25 | 32892 | 599 | 32293 | 0.6336 | -0.09466 | Muscular dystrophies and other myopathies | neurological | RATE_C | |
| 359.1 | | | | | 23949 | 33 | 23916 | | | | | 32328 | 35 | 32293 | | | Muscular dystrophies | neurological | RATE_C | |
| 359.2 | 1.589 | 0.1692 | 4.899 | 5.94e-21 | 24538 | 622 | 23916 | 1.385 | 0.137 | 3.993 | 5.05e-24 | 32864 | 571 | 32293 | 0.5075 | -0.1412 | Myopathy | neurological | RATE_C | |
| 360 | | | | | 24749 | 89 | 24660 | | | | | 33075 | 87 | 32988 | | | Disorders of the globe | sense organs | RATE_C | |

| phenotype | male_beta | male_SE | male_OR | male_p | male_n_total | male_n_cases | male_n_controls | female_beta | female_SE | female_OR | female_p | female_n_total | female_n_cases | female_n_controls | interaction_p | interaction_beta | description | group | lab | label |
|---|---|---|---|---|---|---|---|---|---|---|---|---|---|---|---|---|---|---|---|---|
| 360.2 | | | | | 24707 | 47 | 24660 | | | | | 33042 | 54 | 32988 | | | Progressive myopia | sense organs | RATE_C | |
| 360.3 | | | | | 24670 | 10 | 24660 | | | | | 33001 | 13 | 32988 | | | Hypotony of eye | sense organs | RATE_C | |
| 361 | | | | | 24928 | 268 | 24660 | | | | | 33163 | 175 | 32988 | | | Retinal detachments and defects | sense organs | RATE_C | |
| 361.1 | | | | | 24821 | 161 | 24660 | | | | | 33064 | 76 | 32988 | | | Retinal detachment with retinal defect | sense organs | RATE_C | |
| 361.2 | | | | | 24678 | 18 | 24660 | | | | | 32996 | 8 | 32988 | | | Retinoschisis and retinal cysts | sense organs | RATE_C | |
| 362 | 0.6406 | 0.1008 | 1.898 | 2.08e-10 | 25441 | 1087 | 24354 | 0.7001 | 0.07864 | 2.014 | 5.42e-19 | 34109 | 1379 | 32730 | 0.2122 | 0.1553 | Other retinal disorders | sense organs | RATE_C | |
| 362.1 | | | | | 24369 | 15 | 24354 | | | | | 32743 | 13 | 32730 | | | Retinopathy of prematurity | sense organs | RATE_C | |
| 362.2 | 0.6345 | 0.1448 | 1.886 | 1.18e-05 | 24974 | 620 | 24354 | 0.766 | 0.112 | 2.151 | 8.05e-12 | 33549 | 819 | 32730 | 0.3137 | 0.1822 | Degeneration of macula and posterior pole of retina | sense organs | RATE_C | |
| phenotype | male_beta | male_SE | male_OR | male_p | male_n_total | male_n_cases | male_n_controls | female_beta | female_SE | female_OR | female_p | female_n_total | female_n_cases | female_n_controls | interaction_p | interaction_beta | description | group | lab | label |
| 362.21 | | | | | 24463 | 109 | 24354 | | | | | 32881 | 151 | 32730 | | | Macular degeneration, dry | sense organs | RATE_C | |
| 362.22 | | | | | 24410 | 56 | 24354 | | | | | 32819 | 89 | 32730 | | | Macular degeneration, wet | sense organs | RATE_C | |
| 362.23 | | | | | 24511 | 157 | 24354 | | | | | 32900 | 170 | 32730 | | | Cystoid macular degeneration of retina | sense organs | RATE_C | |
| 362.26 | | | | | 24590 | 236 | 24354 | | | | | 32963 | 233 | 32730 | | | Macular puckering of retina | sense organs | RATE_C | |
| 362.27 | | | | | 24433 | 79 | 24354 | | | | | 32875 | 145 | 32730 | | | Drusen (degenerative) of retina | sense organs | RATE_C | |
| 362.29 | | | | | 24520 | 166 | 24354 | | | | | 33004 | 274 | 32730 | | | Macular degeneration (senile) of retina NOS | sense organs | RATE_C | |
| 362.3 | | | | | 24535 | 181 | 24354 | | | | | 32906 | 176 | 32730 | | | Other nondiabetic retinopathy | sense organs | RATE_C | |
| 362.31 | | | | | 24393 | 39 | 24354 | | | | | 32753 | 23 | 32730 | | | Separation of retinal layers | sense organs | RATE_C | |
| 362.4 | | | | | 24528 | 174 | 24354 | | | | | 32939 | 209 | 32730 | | | Retinal vascular changes and abnomalities | sense organs | RATE_C | |
| 362.5 | | | | | 24369 | 15 | 24354 | | | | | 32823 | 93 | 32730 | | | Toxic maculopathy of retina | sense organs | RATE_C | |
| 362.6 | | | | | 24414 | 60 | 24354 | | | | | 32796 | 66 | 32730 | | | Peripheral retinal degenerations | sense organs | RATE_C | |
| 362.7 | | | | | 24401 | 47 | 24354 | | | | | 32775 | 45 | 32730 | | | Hereditary retinal dystrophies | sense organs | RATE_C | |
| 362.8 | | | | | 24401 | 47 | 24354 | | | | | 32774 | 44 | 32730 | | | Retinal hemorrhage/ischemia | sense organs | RATE_C | |
| 362.9 | | | | | 24469 | 115 | 24354 | | | | | 32862 | 132 | 32730 | | | Retinal edema | sense organs | RATE_C | |
| 363 | | | | | 24770 | 110 | 24660 | | | | | 33134 | 146 | 32988 | | | Chorioretinal inflammations, scars, and other disorders of choroid | sense organs | RATE_C | |
| 363.3 | | | | | 24721 | 61 | 24660 | | | | | 33072 | 84 | 32988 | | | Chorioretinal scars | sense organs | RATE_C | |
| 363.4 | | | | | 24662 | 2 | 24660 | | | | | 32995 | 7 | 32988 | | | Choroidal degenerations | sense organs | RATE_C | |
| 364 | | | | | 24841 | 181 | 24660 | | | | | 33258 | 270 | 32988 | | | Corneal opacity and other disorders of cornea | sense organs | RATE_C | |
| 364.1 | | | | | 24753 | 93 | 24660 | | | | | 33113 | 125 | 32988 | | | Corneal opacity | sense organs | RATE_C | |
| 364.2 | | | | | 24712 | 52 | 24660 | | | | | 33031 | 43 | 32988 | | | Corneal edema | sense organs | RATE_C | |

| phenotype | male_beta | male_SE | male_OR | male_p | male_n_total | male_n_cases | male_n_controls | female_beta | female_SE | female_OR | female_p | female_n_total | female_n_cases | female_n_controls | interaction_p | interaction_beta | description | group | lab | label |
|---|---|---|---|---|---|---|---|---|---|---|---|---|---|---|---|---|---|---|---|---|
| 364.4 | | | | | 24715 | 55 | 24660 | | | | | 33057 | 69 | 32988 | | | Corneal degenerations | sense organs | RATE_C | |
| 364.41 | | | | | 24684 | 24 | 24660 | | | | | 33006 | 18 | 32988 | | | Keratoconus | sense organs | RATE_C | |
| 364.5 | | | | | 24720 | 60 | 24660 | | | | | 33111 | 123 | 32988 | | | Corneal dystrophy | sense organs | RATE_C | |
| 364.51 | | | | | 24699 | 39 | 24660 | | | | | 33061 | 73 | 32988 | | | Fuchs' dystrophy | sense organs | RATE_C | |
| 364.9 | | | | | 24688 | 28 | 24660 | | | | | 33020 | 32 | 32988 | | | Cornea replaced by transplant | sense organs | RATE_C | |
| 365 | 0.2705 | 0.09917 | 1.311 | 0.006373 | 25576 | 916 | 24660 | 0.4073 | 0.07878 | 1.503 | 2.33e-07 | 34125 | 1137 | 32988 | 0.2447 | 0.1427 | Glaucoma | sense organs | RATE_C | |
| 365.1 | 0.3236 | 0.1311 | 1.382 | 0.01359 | 25218 | 558 | 24660 | 0.3146 | 0.1006 | 1.37 | 0.001768 | 33666 | 678 | 32988 | 0.9442 | -0.01126 | Open-angle glaucoma | sense organs | RATE_C | |
| 365.11 | | | | | 24878 | 218 | 24660 | | | | | 33217 | 229 | 32988 | | | Primary open angle glaucoma | sense organs | RATE_C | |
| 365.2 | | | | | 24753 | 93 | 24660 | | | | | 33132 | 144 | 32988 | | | Primary angle-closure glaucoma | sense organs | RATE_C | |
| 365.5 | | | | | 24671 | 11 | 24660 | | | | | 33004 | 16 | 32988 | | | Pseudoexfoliation glaucoma | sense organs | RATE_C | |
| 366 | 0.4512 | 0.06291 | 1.57 | 7.37e-13 | 26713 | 2883 | 23830 | 0.5214 | 0.04894 | 1.684 | 1.68e-26 | 35649 | 3722 | 31927 | 0.0804 | 0.1364 | Cataract | sense organs | RATE_C | |
| 366.1 | | | | | 23845 | 15 | 23830 | | | | | 31936 | 9 | 31927 | | | Nonsenile Cataract | sense organs | RATE_C | |
| 366.2 | 0.4832 | 0.06774 | 1.621 | 9.87e-13 | 26293 | 2463 | 23830 | 0.4978 | 0.05201 | 1.645 | 1.06e-21 | 35061 | 3134 | 31927 | 0.3592 | 0.07653 | Senile cataract | sense organs | RATE_C | |
| 366.3 | | | | | 23835 | 5 | 23830 | | | | | 31932 | 5 | 31927 | | | Traumatic cataract | sense organs | RATE_C | |
| 367 | 0.2188 | 0.0773 | 1.245 | 0.00464 | 26602 | 1195 | 25407 | 0.3467 | 0.05343 | 1.414 | 8.61e-11 | 35278 | 2096 | 33182 | 0.5705 | 0.04879 | Disorders of refraction and accommodation; blindness and low vision | sense organs | RATE_C | |
| 367.1 | 0.06282 | 0.1033 | 1.065 | 0.5429 | 25991 | 584 | 25407 | 0.2863 | 0.06627 | 1.331 | 1.56e-05 | 34437 | 1255 | 33182 | 0.2747 | 0.1204 | Myopia | sense organs | RATE_C | |
| 367.2 | | | | | 25731 | 324 | 25407 | 0.4429 | 0.1071 | 1.557 | 3.53e-05 | 33701 | 519 | 33182 | 0.1429 | 0.2432 | Astigmatism | sense organs | RATE_C | |
| 367.8 | | | | | 25506 | 99 | 25407 | | | | | 33431 | 249 | 33182 | | | Hypermetropia | sense organs | RATE_C | |
| 367.9 | | | | | 25707 | 300 | 25407 | | | | | 33524 | 342 | 33182 | 0.9523 | -0.01264 | Blindness and low vision | sense organs | RATE_C | |
| 368 | 0.7666 | 0.09622 | 2.152 | 1.62e-15 | 26249 | 990 | 25259 | 0.7313 | 0.06689 | 2.078 | 8.09e-28 | 34737 | 1481 | 33256 | 0.4919 | -0.07537 | Visual disturbances | sense organs | RATE_C | |
| 368.1 | | | | | 25311 | 52 | 25259 | | | | | 33326 | 70 | 33256 | | | Amblyopia | sense organs | RATE_C | |
| 368.2 | | | | | 25592 | 333 | 25259 | | | | | 33627 | 371 | 33256 | 0.9879 | 0.003384 | Diplopia and disorders of binocular vision | sense organs | RATE_C | |
| 368.3 | | | | | 25277 | 18 | 25259 | | | | | 33272 | 16 | 33256 | | | Anisometropia | sense organs | RATE_C | |
| 368.4 | | | | | 25410 | 151 | 25259 | | | | | 33464 | 208 | 33256 | | | Visual field defects | sense organs | RATE_C | |
| 368.5 | | | | | 25262 | 3 | 25259 | | | | | 33257 | 1 | 33256 | | | Color vision deficiencies | sense organs | RATE_C | |
| 368.7 | | | | | 25259 | | | | | | | 33257 | 1 | 33256 | | | Disorders of accommodation | sense organs | RATE_C | |
| 368.9 | | | | | 25451 | 192 | 25259 | | | | | 33633 | 377 | 33256 | 0.5606 | -0.1448 | Subjective visual disturbances | sense organs | RATE_C | |
| 368.91 | | | | | 25299 | 40 | 25259 | | | | | 33323 | 67 | 33256 | | | Psychophysical visual disturbances | sense organs | RATE_C | |
| 369 | 0.004142 | 0.1138 | 1.004 | 0.971 | 24317 | 512 | 23805 | 0.189 | 0.07835 | 1.208 | 0.01587 | 31511 | 916 | 30595 | 0.1866 | 0.1692 | Infection of the eye | sense organs | RATE_C | |

| phenotype | male_beta | male_SE | male_OR | male_p | male_n_total | male_n_cases | male_n_controls | female_beta | female_SE | female_OR | female_p | female_n_total | female_n_cases | female_n_controls | interaction_p | interaction_beta | description | group | lab | label |
|---|---|---|---|---|---|---|---|---|---|---|---|---|---|---|---|---|---|---|---|---|
| 369.2 | | | | | 23885 | 80 | 23805 | | | | | 30724 | 129 | 30595 | | | Eye infection, viral | sense organs | RATE_C | |
| 369.5 | | | | | 24187 | 382 | 23805 | 0.195 | 0.08829 | 1.215 | 0.02721 | 31314 | 719 | 30595 | 0.1761 | 0.1978 | Conjunctivitis, infectious | sense organs | RATE_C | |
| 370 | | | | | 24074 | 269 | 23805 | 0.5708 | 0.1001 | 1.77 | 1.19e-08 | 31271 | 676 | 30595 | 0.2995 | 0.1986 | Keratitis | sense organs | RATE_C | |
| 370.1 | | | | | 23880 | 75 | 23805 | | | | | 30694 | 99 | 30595 | | | Corneal ulcer | sense organs | RATE_C | |
| 370.2 | | | | | 23830 | 25 | 23805 | | | | | 30656 | 61 | 30595 | | | Superficial keratitis | sense organs | RATE_C | |
| 370.3 | | | | | 23964 | 159 | 23805 | | | | | 31063 | 468 | 30595 | 0.09776 | 0.4096 | Keratoconjunctivitis | sense organs | RATE_C | |
| 370.31 | | | | | 23923 | 118 | 23805 | | | | | 31009 | 414 | 30595 | 0.2373 | 0.3483 | Keratoconjunctivitis sicca | sense organs | RATE_C | |
| 371 | 0.06056 | 0.0896 | 1.062 | 0.4991 | 24762 | 957 | 23805 | 0.2847 | 0.06452 | 1.329 | 1.02e-05 | 32133 | 1538 | 30595 | 0.06016 | 0.1965 | Inflammation of the eye | sense organs | RATE_C | |
| 371.1 | | | | | 23925 | 120 | 23805 | | | | | 30815 | 220 | 30595 | | | Uveitis, noninfectious or NOS | sense organs | RATE_C | |
| 371.2 | | | | | 24253 | 448 | 23805 | 0.3128 | 0.09029 | 1.367 | 5.32e-04 | 31368 | 773 | 30595 | 0.5802 | 0.08571 | Conjunctivitis, noninfectious | sense organs | RATE_C | |
| 371.21 | | | | | 24201 | 396 | 23805 | 0.2953 | 0.09533 | 1.343 | 0.001954 | 31279 | 684 | 30595 | 0.6357 | 0.07761 | Allergic conjunctivitis | sense organs | RATE_C | |
| 371.3 | | | | | 24165 | 360 | 23805 | 0.4898 | 0.1122 | 1.632 | 1.26e-05 | 31154 | 559 | 30595 | 0.007011 | 0.4686 | Inflammation of eyelids | sense organs | RATE_C | |
| 371.33 | | | | | 23808 | 3 | 23805 | | | | | 30603 | 8 | 30595 | | | Noninfectious dermatoses of eyelid | sense organs | RATE_C | |
| 371.9 | | | | | 23817 | 12 | 23805 | | | | | 30607 | 12 | 30595 | | | Chronic inflammatory disorders of orbit | sense organs | RATE_C | |
| 372 | | | | | 23933 | 128 | 23805 | | | | | 30728 | 133 | 30595 | | | Disorders of conjunctiva | sense organs | RATE_C | |
| 374 | 0.2679 | 0.1086 | 1.307 | 0.01363 | 24581 | 776 | 23805 | 0.5919 | 0.08681 | 1.807 | 9.23e-12 | 31646 | 1051 | 30595 | 0.02149 | 0.3099 | Other disorders of eyelids | sense organs | RATE_C | |
| 374.1 | | | | | 23920 | 115 | 23805 | | | | | 30673 | 78 | 30595 | | | Ectropion or entropion | sense organs | RATE_C | |
| 374.2 | | | | | 23850 | 45 | 23805 | | | | | 30619 | 24 | 30595 | | | Lagophthalmos | sense organs | RATE_C | |
| 374.3 | | | | | 24001 | 196 | 23805 | | | | | 30970 | 375 | 30595 | 0.05039 | 0.51 | Ptosis of eyelid | sense organs | RATE_C | |
| 374.6 | | | | | 24250 | 445 | 23805 | 0.4124 | 0.1121 | 1.51 | 2.35e-04 | 31205 | 610 | 30595 | 0.3457 | 0.1703 | Dermatochalasis | sense organs | RATE_C | |
| 375 | | | | | 23884 | 86 | 23798 | | | | | 31096 | 146 | 30950 | | | Disorders of lacrimal system | sense organs | RATE_C | |
| 375.1 | 0.1106 | 0.09011 | 1.117 | 0.2198 | 24860 | 1062 | 23798 | 0.3225 | 0.06007 | 1.381 | 7.96e-08 | 32921 | 1971 | 30950 | 0.1239 | 0.1602 | Dry eyes | sense organs | RATE_C | |
| 375.2 | | | | | 23848 | 50 | 23798 | | | | | 31036 | 86 | 30950 | | | Epiphora | sense organs | RATE_C | |
| 376 | | | | | 23842 | 44 | 23798 | | | | | 30982 | 32 | 30950 | | | Disorders of the orbit | sense organs | RATE_C | |
| 377 | | | | | 24004 | 206 | 23798 | | | | | 31323 | 373 | 30950 | 0.4376 | 0.1472 | Disorders of optic nerve and visual pathways | sense organs | RATE_C | |
| 377.1 | | | | | 23872 | 74 | 23798 | | | | | 31057 | 107 | 30950 | | | Optic atrophy | sense organs | RATE_C | |
| 377.3 | | | | | 23899 | 101 | 23798 | | | | | 31129 | 179 | 30950 | | | Optic neuritis/neuropathy | sense organs | RATE_C | |
| 378 | | | | | 24117 | 319 | 23798 | | | | | 31299 | 349 | 30950 | 0.07352 | 0.2947 | Strabismus and other disorders of binocular eye movements | sense organs | RATE_C | |

| phenotype | male_beta | male_SE | male_OR | male_p | male_n_total | male_n_cases | male_n_controls | female_beta | female_SE | female_OR | female_p | female_n_total | female_n_cases | female_n_controls | interaction_p | interaction_beta | description | group | lab | label |
|---|---|---|---|---|---|---|---|---|---|---|---|---|---|---|---|---|---|---|---|---|
| 378.1 | | | | | 23973 | 175 | 23798 | | | | | 31136 | 186 | 30950 | | | Strabismus (not specified as paralytic) | sense organs | RATE_C | |
| 378.2 | | | | | 23835 | 37 | 23798 | | | | | 31001 | 51 | 30950 | | | Nystagmus and other irregular eye movements | sense organs | RATE_C | |
| 378.5 | | | | | 23889 | 91 | 23798 | | | | | 31030 | 80 | 30950 | | | Paralytic strabismus | sense organs | RATE_C | |
| 379 | 0.3652 | 0.09127 | 1.441 | 6.31e-05 | 24956 | 1158 | 23798 | 0.3594 | 0.06603 | 1.432 | 5.26e-08 | 32559 | 1609 | 30950 | 0.8805 | -0.01632 | Other disorders of eye | sense organs | RATE_C | |
| 379.1 | | | | | 23846 | 48 | 23798 | | | | | 31085 | 135 | 30950 | | | Scleritis and episcleritis | sense organs | RATE_C | |
| 379.2 | 0.3886 | 0.1028 | 1.475 | 1.57e-04 | 24762 | 964 | 23798 | 0.3538 | 0.07444 | 1.424 | 2.01e-06 | 32269 | 1319 | 30950 | 0.9097 | -0.01396 | Disorders of vitreous body | sense organs | RATE_C | |
| 379.3 | | | | | 23855 | 57 | 23798 | | | | | 30995 | 45 | 30950 | | | Aphakia and other disorders of lens | sense organs | RATE_C | |
| 379.4 | | | | | 23835 | 37 | 23798 | | | | | 31014 | 64 | 30950 | | | Anomalies of pupillary function | sense organs | RATE_C | |
| 379.5 | | | | | 23914 | 116 | 23798 | | | | | 31022 | 72 | 30950 | | | Disorders of iris and ciliary body | sense organs | RATE_C | |
| 379.51 | | | | | 23816 | 18 | 23798 | | | | | 30958 | 8 | 30950 | | | Pigmentary iris degeneration | sense organs | RATE_C | |
| 379.9 | | | | | 23893 | 95 | 23798 | | | | | 31121 | 171 | 30950 | | | Pain, swelling or discharge of eye | sense organs | RATE_C | |
| 380 | | | | | 25280 | 65 | 25215 | | | | | 34053 | 71 | 33982 | | | Disorders of external ear | sense organs | RATE_C | |
| 380.1 | | | | | 25546 | 331 | 25215 | | | | | 34343 | 361 | 33982 | 0.5798 | 0.09455 | Otitis externa | sense organs | RATE_C | |
| 380.4 | 0.0478 | 0.09153 | 1.049 | 0.6015 | 26150 | 935 | 25215 | 0.104 | 0.08125 | 1.11 | 0.2007 | 34929 | 947 | 33982 | 0.2265 | 0.1405 | Impacted cerumen | sense organs | RATE_C | |
| 381 | 0.04727 | 0.07405 | 1.048 | 0.5232 | 25736 | 1209 | 24527 | 0.1801 | 0.05202 | 1.197 | 5.35e-04 | 33271 | 2155 | 31116 | 0.004412 | 0.2312 | Otitis media and Eustachian tube disorders | sense organs | RATE_C | |
| 381.1 | -0.05927 | 0.09764 | 0.9424 | 0.5438 | 25160 | 633 | 24527 | 0.1961 | 0.0699 | 1.217 | 0.005015 | 32235 | 1119 | 31116 | 8.79e-04 | 0.3535 | Otitis media | sense organs | RATE_C | |
| 381.11 | | | | | 24939 | 412 | 24527 | 0.1505 | 0.08486 | 1.162 | 0.07615 | 31847 | 731 | 31116 | 3.89e-04 | 0.4543 | Suppurative and unspecified otitis media | sense organs | RATE_C | |
| 381.2 | 0.07116 | 0.1036 | 1.074 | 0.4921 | 25149 | 622 | 24527 | 0.2286 | 0.07182 | 1.257 | 0.001457 | 32238 | 1122 | 31116 | 0.0379 | 0.2397 | Eustachian tube disorders | sense organs | RATE_C | |
| 381.3 | | | | | 24579 | 52 | 24527 | | | | | 31179 | 63 | 31116 | | | Mastoiditis & related conditions | sense organs | RATE_C | |
| 381.9 | | | | | 24645 | 118 | 24527 | | | | | 31231 | 115 | 31116 | | | Otorrhea | sense organs | RATE_C | |
| 382 | | | | | 24849 | 322 | 24527 | 0.4644 | 0.08705 | 1.591 | 9.57e-08 | 31925 | 809 | 31116 | 0.03536 | 0.3286 | Otalgia | sense organs | RATE_C | |
| 383 | | | | | 24540 | 13 | 24527 | | | | | 31138 | 22 | 31116 | | | Otosclerosis | sense organs | RATE_C | |
| 384 | | | | | 24691 | 164 | 24527 | | | | | 31314 | 198 | 31116 | | | Other disorders of tympanic membrane | sense organs | RATE_C | |
| 384.1 | | | | | 24551 | 24 | 24527 | | | | | 31124 | 8 | 31116 | | | Myringitis | sense organs | RATE_C | |
| 384.4 | | | | | 24667 | 140 | 24527 | | | | | 31294 | 178 | 31116 | | | Perforation of tympanic membrane | sense organs | RATE_C | |
| 385 | | | | | 24625 | 98 | 24527 | | | | | 31245 | 129 | 31116 | | | Other disorders of middle ear and mastoid | sense organs | RATE_C | |
| 385.3 | | | | | 24606 | 79 | 24527 | | | | | 31204 | 88 | 31116 | | | Cholesteatoma | sense organs | RATE_C | |
| 385.5 | | | | | 24556 | 29 | 24527 | | | | | 31165 | 49 | 31116 | | | Tympanosclerosis and middle ear disease related to otitis media | sense organs | RATE_C | |
| 386 | | | | | 23483 | 489 | 22994 | 0.6644 | 0.08443 | 1.943 | 3.56e-15 | 30272 | 992 | 29280 | 0.07376 | 0.2676 | Vertiginous syndromes and other disorders of vestibular system | sense organs | RATE_C | |

| phenotype | male_beta | male_SE | male_OR | male_p | male_n_total | male_n_cases | male_n_controls | female_beta | female_SE | female_OR | female_p | female_n_total | female_n_cases | female_n_controls | interaction_p | interaction_beta | description | group | lab | label |
|---|---|---|---|---|---|---|---|---|---|---|---|---|---|---|---|---|---|---|---|---|
| 386.1 | | | | | 23077 | 83 | 22994 | | | | | 29457 | 177 | 29280 | | | Meniere's disease | sense organs | RATE_C | |
| 386.2 | | | | | 23328 | 334 | 22994 | 0.5468 | 0.1001 | 1.728 | 4.63e-08 | 29955 | 675 | 29280 | 0.04281 | 0.3523 | Peripheral or central vertigo | sense organs | RATE_C | |
| 386.21 | | | | | 23041 | 47 | 22994 | | | | | 29385 | 105 | 29280 | | | Central origin vertigo | sense organs | RATE_C | |
| 386.3 | | | | | 23038 | 44 | 22994 | | | | | 29344 | 64 | 29280 | | | Labyrinthitis | sense organs | RATE_C | |
| 386.9 | 0.9006 | 0.07208 | 2.461 | 8.00e-36 | 25211 | 2217 | 22994 | 1.129 | 0.04959 | 3.092 | 1.08e-114 | 32967 | 3687 | 29280 | 0.05553 | 0.1612 | Dizziness and giddiness (Light-headedness and vertigo) | sense organs | RATE_C | |
| 388 | | | | | 23411 | 34 | 23377 | | | | | 32030 | 47 | 31983 | | | Other disorders of ear | sense organs | RATE_C | |
| 389 | 0.2364 | 0.05771 | 1.267 | 4.19e-05 | 26031 | 2654 | 23377 | 0.4288 | 0.04911 | 1.535 | 2.54e-18 | 34938 | 2955 | 31983 | 0.005905 | 0.1979 | Hearing loss | sense organs | RATE_C | |
| 389.1 | 0.3248 | 0.07685 | 1.384 | 2.38e-05 | 25007 | 1630 | 23377 | 0.5458 | 0.06819 | 1.726 | 1.20e-15 | 33675 | 1692 | 31983 | 0.01479 | 0.2422 | Sensorineural hearing loss | sense organs | RATE_C | |
| 389.2 | | | | | 23633 | 256 | 23377 | | | | | 32346 | 363 | 31983 | 0.0537 | 0.326 | Conductive hearing loss | sense organs | RATE_C | |
| 389.3 | | | | | 23398 | 21 | 23377 | | | | | 31995 | 12 | 31983 | | | Degenerative and vascular disorders of ear | sense organs | RATE_C | |
| 389.4 | 0.3144 | 0.1124 | 1.369 | 0.005164 | 23983 | 606 | 23377 | 0.512 | 0.09444 | 1.669 | 5.93e-08 | 32714 | 731 | 31983 | 0.08178 | 0.2421 | Tinnitus | sense organs | RATE_C | |
| 389.5 | | | | | 23394 | 17 | 23377 | | | | | 32002 | 19 | 31983 | | | Disorders of acoustic nerve | sense organs | RATE_C | |
| 394 | 2.036 | 0.1345 | 7.656 | 1.04e-51 | 22330 | 1378 | 20952 | 2.01 | 0.1073 | 7.461 | 3.28e-78 | 31197 | 1468 | 29729 | 0.9484 | -0.01105 | Rheumatic disease of the heart valves | circulatory system | RATE_C | |
| 394.1 | | | | | 21305 | 353 | 20952 | | | | | 30068 | 339 | 29729 | 0.3441 | -0.3438 | Mitral valve stenosis and aortic valve stenosis | circulatory system | RATE_C | |
| phenotype | male_beta | male_SE | male_OR | male_p | male_n_total | male_n_cases | male_n_controls | female_beta | female_SE | female_OR | female_p | female_n_total | female_n_cases | female_n_controls | interaction_p | interaction_beta | description | group | lab | label |
| 394.2 | | | | | 21121 | 169 | 20952 | | | | | 29965 | 236 | 29729 | | | Mitral valve disease | circulatory system | RATE_C | |
| 394.3 | | | | | 20969 | 17 | 20952 | | | | | 29742 | 13 | 29729 | | | Aortic valve disease | circulatory system | RATE_C | |
| 394.4 | | | | | 20972 | 20 | 20952 | | | | | 29771 | 42 | 29729 | | | Acute rheumatic heart disease | circulatory system | RATE_C | |
| 394.7 | 2.311 | 0.2259 | 10.09 | 1.46e-24 | 21461 | 509 | 20952 | 2.36 | 0.1787 | 10.59 | 8.30e-40 | 30357 | 628 | 29729 | 0.5809 | 0.1573 | Disease of tricuspid valve | circulatory system | RATE_C | |
| 395 | 2.031 | 0.08366 | 7.618 | 3.90e-130 | 24383 | 3431 | 20952 | 1.933 | 0.07015 | 6.908 | 4.40e-167 | 32852 | 3123 | 29729 | 0.4561 | -0.08061 | Heart valve disorders | circulatory system | RATE_C | |
| 395.1 | 2.199 | 0.1233 | 9.015 | 3.81e-71 | 22776 | 1824 | 20952 | 2.111 | 0.09646 | 8.255 | 3.72e-106 | 31647 | 1918 | 29729 | 0.6312 | -0.07458 | Nonrheumatic mitral valve disorders | circulatory system | RATE_C | |
| 395.2 | 2.093 | 0.1218 | 8.113 | 3.62e-66 | 22773 | 1821 | 20952 | 1.967 | 0.1095 | 7.15 | 3.91e-72 | 31147 | 1418 | 29729 | 0.5346 | -0.1011 | Nonrheumatic aortic valve disorders | circulatory system | RATE_C | |
| 395.3 | | | | | 21269 | 317 | 20952 | | | | | 30053 | 324 | 29729 | 0.2089 | 0.5586 | Nonrheumatic tricuspid valve disorders | circulatory system | RATE_C | |
| 395.4 | | | | | 21053 | 101 | 20952 | | | | | 29855 | 126 | 29729 | | | Nonrheumatic pulmonary valve disorders | circulatory system | RATE_C | |
| 395.6 | 2.266 | 0.1799 | 9.645 | 2.21e-36 | 21815 | 863 | 20952 | 2.618 | 0.2025 | 13.71 | 3.02e-38 | 30347 | 618 | 29729 | 0.2054 | 0.3403 | Heart valve replaced | circulatory system | RATE_C | |
| 396 | 1.022 | 0.1253 | 2.778 | 3.50e-16 | 21534 | 582 | 20952 | 1.177 | 0.09614 | 3.246 | 1.76e-34 | 30632 | 903 | 29729 | 0.03038 | 0.329 | Abnormal heart sounds | circulatory system | RATE_C | |
| 401 | 1.496 | 0.03452 | 4.464 | 0 | 25158 | 14670 | 10488 | 1.497 | 0.02922 | 4.469 | 0 | 33895 | 14782 | 19113 | 0.2892 | 0.04767 | Hypertension | circulatory system | RATE_C | |
| 401.1 | 1.503 | 0.03492 | 4.496 | 0 | 24789 | 14301 | 10488 | 1.505 | 0.02951 | 4.503 | 0 | 33585 | 14472 | 19113 | 0.3125 | 0.04594 | Essential hypertension | circulatory system | RATE_C | |
| 401.2 | 2.277 | 0.07098 | 9.748 | 7.76e-226 | 14837 | 4349 | 10488 | 2.187 | 0.0666 | 8.908 | 1.58e-236 | 22464 | 3351 | 19113 | 0.4328 | -0.07641 | Hypertensive heart and/or renal disease | circulatory system | RATE_C | |

| phenotype | male_beta | male_SE | male_OR | male_p | male_n_total | male_n_cases | male_n_controls | female_beta | female_SE | female_OR | female_p | female_n_total | female_n_cases | female_n_controls | interaction_p | interaction_beta | description | group | lab | label |
|---|---|---|---|---|---|---|---|---|---|---|---|---|---|---|---|---|---|---|---|---|
| 401.21 | 2.424 | 0.1044 | 11.29 | 2.59e-119 | 12798 | 2310 | 10488 | 2.25 | 0.0903 | 9.483 | 5.60e-137 | 21086 | 1973 | 19113 | 0.2428 | -0.1614 | Hypertensive heart disease | circulatory system | RATE_C | |
| 401.22 | 2.35 | 0.08986 | 10.49 | 9.52e-151 | 12900 | 2412 | 10488 | 2.361 | 0.09392 | 10.6 | 1.83e-139 | 20743 | 1630 | 19113 | 0.7738 | 0.03733 | Hypertensive chronic kidney disease | circulatory system | RATE_C | |
| 401.3 | 1.461 | 0.1052 | 4.308 | 7.58e-44 | 11324 | 836 | 10488 | 1.688 | 0.09596 | 5.407 | 2.99e-69 | 20074 | 961 | 19113 | 0.006638 | 0.3811 | Other hypertensive complications | circulatory system | RATE_C | |
| 411 | 2.005 | 0.0505 | 7.427 | 0 | 24924 | 8227 | 16697 | 2.113 | 0.05857 | 8.27 | 6.78e-285 | 33484 | 5060 | 28424 | 0.2173 | 0.09522 | Ischemic Heart Disease | circulatory system | RATE_C | |
| 411.1 | 2.803 | 0.1646 | 16.49 | 4.94e-65 | 18153 | 1456 | 16697 | 2.895 | 0.2089 | 18.09 | 1.15e-43 | 29191 | 767 | 28424 | 0.8309 | 0.05662 | Unstable angina (intermediate coronary syndrome) | circulatory system | RATE_C | |
| 411.2 | 2.61 | 0.102 | 13.6 | 2.21e-144 | 19713 | 3016 | 16697 | 2.632 | 0.123 | 13.9 | 1.54e-101 | 30089 | 1665 | 28424 | 0.9739 | -0.005207 | Myocardial infarction | circulatory system | RATE_C | |
| 411.3 | 2.415 | 0.1294 | 11.19 | 8.74e-78 | 18373 | 1676 | 16697 | 2.516 | 0.1528 | 12.37 | 6.15e-61 | 29445 | 1021 | 28424 | 0.7462 | 0.0645 | Angina pectoris | circulatory system | RATE_C | |
| 411.4 | 2.012 | 0.05331 | 7.482 | 0 | 24205 | 7508 | 16697 | 2.095 | 0.06447 | 8.122 | 1.57e-231 | 32583 | 4159 | 28424 | 0.4177 | 0.06765 | Coronary atherosclerosis | circulatory system | RATE_C | |
| 411.41 | | | | | 16754 | 57 | 16697 | | | | | 28492 | 68 | 28424 | | | Aneurysm and dissection of heart | circulatory system | RATE_C | |
| 411.8 | 2.757 | 0.1184 | 15.76 | 5.04e-120 | 19412 | 2715 | 16697 | 3.074 | 0.1964 | 21.63 | 3.20e-55 | 29458 | 1034 | 28424 | 0.21 | 0.2869 | Other chronic ischemic heart disease, unspecified | circulatory system | RATE_C | |
| 411.9 | | | | | 16791 | 94 | 16697 | | | | | 28492 | 68 | 28424 | | | Other acute and subacute forms of ischemic heart disease | circulatory system | RATE_C | |
| 414 | 2.524 | 0.1144 | 12.48 | 7.56e-108 | 18860 | 2163 | 16697 | 2.275 | 0.1097 | 9.731 | 1.59e-95 | 29887 | 1463 | 28424 | 0.06637 | -0.2892 | Other forms of chronic heart disease | circulatory system | RATE_C | |
| 414.2 | | | | | 16930 | 233 | 16697 | | | | | 28541 | 117 | 28424 | | | ASCVD | circulatory system | RATE_C | |
| 415 | 2.019 | 0.09183 | 7.527 | 4.42e-107 | 22610 | 2006 | 20604 | 1.941 | 0.07952 | 6.969 | 1.21e-131 | 32928 | 1974 | 30954 | 0.9154 | 0.01261 | Pulmonary heart disease | circulatory system | RATE_C | |
| 415.1 | 1.847 | 0.1295 | 6.338 | 3.85e-46 | 21476 | 872 | 20604 | 1.721 | 0.1114 | 5.592 | 7.51e-54 | 31762 | 808 | 30954 | 0.492 | -0.1142 | Acute pulmonary heart disease | circulatory system | RATE_C | |
| 415.11 | 1.742 | 0.1308 | 5.708 | 1.78e-40 | 21366 | 762 | 20604 | 1.685 | 0.1131 | 5.39 | 3.30e-50 | 31710 | 756 | 30954 | 0.9021 | -0.02062 | Pulmonary embolism and infarction, acute | circulatory system | RATE_C | |
| 415.2 | 2.391 | 0.1481 | 10.93 | 1.18e-58 | 21649 | 1045 | 20604 | 2.3 | 0.1237 | 9.977 | 3.27e-77 | 32069 | 1115 | 30954 | 0.8091 | 0.04588 | Chronic pulmonary heart disease | circulatory system | RATE_C | |
| 415.21 | | | | | 20763 | 159 | 20604 | | | | | 31201 | 247 | 30954 | | | Primary pulmonary hypertension | circulatory system | RATE_C | |
| 416 | 2.5 | 0.1004 | 12.18 | 5.93e-137 | 23564 | 2960 | 20604 | 2.43 | 0.1028 | 11.35 | 1.48e-123 | 32994 | 2040 | 30954 | 0.9149 | 0.01518 | Cardiomegaly | circulatory system | RATE_C | |
| 418 | 2.147 | 0.04873 | 8.557 | 0 | 23574 | 7662 | 15912 | 2.23 | 0.03882 | 9.299 | 0 | 31720 | 9073 | 22647 | 0.00417 | 0.1709 | Nonspecific chest pain | circulatory system | RATE_C | |
| 418.1 | 1.99 | 0.1068 | 7.312 | 2.08e-77 | 17223 | 1311 | 15912 | 1.964 | 0.08525 | 7.127 | 2.03e-117 | 24061 | 1414 | 22647 | 0.5565 | 0.07742 | Precordial pain | circulatory system | RATE_C | |
| 420 | 1.982 | 0.134 | 7.255 | 1.72e-49 | 24630 | 934 | 23696 | 1.865 | 0.1146 | 6.455 | 1.67e-59 | 34594 | 895 | 33699 | 0.9872 | 0.002733 | Carditis | circulatory system | RATE_C | |
| 420.1 | | | | | 23727 | 31 | 23696 | | | | | 33725 | 26 | 33699 | | | Myocarditis | circulatory system | RATE_C | |
| 420.2 | 1.975 | 0.1629 | 7.204 | 8.16e-34 | 24311 | 615 | 23696 | 1.779 | 0.1325 | 5.925 | 3.89e-41 | 34349 | 650 | 33699 | 0.9915 | -0.002149 | Pericarditis | circulatory system | RATE_C | |
| 420.21 | | | | | 23792 | 96 | 23696 | | | | | 33781 | 82 | 33699 | | | Acute pericarditis | circulatory system | RATE_C | |
| 420.22 | | | | | 23800 | 104 | 23696 | | | | | 33807 | 108 | 33699 | | | Chronic pericarditis | circulatory system | RATE_C | |
| 420.3 | | | | | 23990 | 294 | 23696 | | | | | 33932 | 233 | 33699 | 0.9621 | 0.01554 | Endocarditis | circulatory system | RATE_C | |
| 425 | 2.289 | 0.1181 | 9.869 | 9.19e-84 | 25754 | 2058 | 23696 | 2.077 | 0.1142 | 7.978 | 7.33e-74 | 34958 | 1259 | 33699 | 0.2052 | -0.2044 | Cardiomyopathy | circulatory system | RATE_C | |
| 425.1 | 2.299 | 0.1254 | 9.964 | 4.75e-75 | 25538 | 1842 | 23696 | 2.19 | 0.1295 | 8.938 | 3.88e-64 | 34754 | 1055 | 33699 | 0.4854 | -0.1236 | Primary/intrinsic cardiomyopathies | circulatory system | RATE_C | |

| phenotype | male_beta | male_SE | male_OR | male_p | male_n_total | male_n_cases | male_n_controls | female_beta | female_SE | female_OR | female_p | female_n_total | female_n_cases | female_n_controls | interaction_p | interaction_beta | description | group | lab | label |
|---|---|---|---|---|---|---|---|---|---|---|---|---|---|---|---|---|---|---|---|---|
| 425.11 | | | | | 23809 | 113 | 23696 | | | | | 33810 | 111 | 33699 | | | Hypertrophic obstructive cardiomyopathy | circulatory system | RATE_C | |
| 425.12 | | | | | 23788 | 92 | 23696 | | | | | 33777 | 78 | 33699 | | | Other hypertrophic cardiomyopathy | circulatory system | RATE_C | |
| 425.2 | 2.856 | 0.2728 | 17.4 | 1.18e-25 | 24351 | 655 | 23696 | | | | | 34066 | 367 | 33699 | 0.6769 | -0.1601 | Secondary/extrinsic cardiomyopathies | circulatory system | RATE_C | |
| 425.8 | | | | | 23738 | 42 | 23696 | | | | | 33802 | 103 | 33699 | | | Other cardiomyopathy | circulatory system | RATE_C | |
| 426 | 2.417 | 0.07588 | 11.21 | 1.33e-222 | 18055 | 3856 | 14199 | 2.385 | 0.0746 | 10.86 | 2.28e-224 | 24927 | 2812 | 22115 | 0.8355 | -0.02187 | Cardiac conduction disorders | circulatory system | RATE_C | |
| 426.2 | 2.319 | 0.1593 | 10.17 | 4.98e-48 | 15129 | 930 | 14199 | 2.618 | 0.1932 | 13.71 | 8.00e-42 | 22650 | 535 | 22115 | 0.4192 | 0.2013 | Atrioventricular [AV] block | circulatory system | RATE_C | |
| 426.21 | | | | | 14391 | 192 | 14199 | | | | | 22185 | 70 | 22115 | | | First degree AV block | circulatory system | RATE_C | |
| 426.22 | | | | | 14228 | 29 | 14199 | | | | | 22135 | 20 | 22115 | | | Mobitz II AV block | circulatory system | RATE_C | |
| 426.23 | | | | | 14388 | 189 | 14199 | | | | | 22218 | 103 | 22115 | | | Second degree AV block | circulatory system | RATE_C | |
| 426.24 | 2.2 | 0.1875 | 9.022 | 8.56e-32 | 14798 | 599 | 14199 | | | | | 22478 | 363 | 22115 | 0.4055 | 0.2438 | Atrioventricular block, complete | circulatory system | RATE_C | |
| 426.25 | | | | | 14243 | 44 | 14199 | | | | | 22136 | 21 | 22115 | | | Other heart block | circulatory system | RATE_C | |
| 426.3 | 2.751 | 0.1913 | 15.67 | 6.26e-47 | 15070 | 871 | 14199 | 2.692 | 0.1963 | 14.75 | 8.92e-43 | 22678 | 563 | 22115 | 0.8004 | -0.06896 | Bundle branch block | circulatory system | RATE_C | |
| 426.31 | | | | | 14547 | 348 | 14199 | | | | | 22262 | 147 | 22115 | | | Right bundle branch block | circulatory system | RATE_C | |
| 426.32 | | | | | 14638 | 439 | 14199 | | | | | 22500 | 385 | 22115 | 0.2801 | -0.5072 | Left bundle branch block | circulatory system | RATE_C | |
| 426.4 | | | | | 14250 | 51 | 14199 | | | | | 22148 | 33 | 22115 | | | Anomalous atrioventricular excitation | circulatory system | RATE_C | |
| 426.7 | 2.12 | 0.1414 | 8.331 | 7.59e-51 | 14888 | 689 | 14199 | 1.961 | 0.119 | 7.108 | 5.04e-61 | 22857 | 742 | 22115 | 0.8875 | 0.02546 | Abnormal electrocardiogram [ECG] [EKG] | circulatory system | RATE_C | |
| 426.8 | | | | | 14301 | 102 | 14199 | | | | | 22230 | 115 | 22115 | | | Other cardiac conduction disorders | circulatory system | RATE_C | |
| 426.9 | 2.881 | 0.1294 | 17.82 | 9.80e-110 | 16283 | 2084 | 14199 | 2.851 | 0.1425 | 17.31 | 4.02e-89 | 23352 | 1237 | 22115 | 0.9741 | -0.006224 | Cardiac pacemaker/device in situ | circulatory system | RATE_C | |
| 426.91 | 2.771 | 0.1613 | 15.97 | 3.71e-66 | 15508 | 1309 | 14199 | 2.845 | 0.1698 | 17.2 | 5.52e-63 | 23040 | 925 | 22115 | 0.7426 | 0.07674 | Cardiac pacemaker in situ | circulatory system | RATE_C | |
| 426.92 | 3.341 | 0.2082 | 28.26 | 6.09e-58 | 15274 | 1075 | 14199 | | | | | 22521 | 406 | 22115 | 0.4629 | 0.2802 | Cardiac defibrillator in situ | circulatory system | RATE_C | |
| 427 | 2.155 | 0.04305 | 8.625 | 0 | 23930 | 9731 | 14199 | 2.185 | 0.03561 | 8.894 | 0 | 32189 | 10074 | 22115 | 0.2702 | -0.05988 | Cardiac dysrhythmias | circulatory system | RATE_C | |
| 427.1 | 2.771 | 0.1222 | 15.97 | 8.02e-114 | 16046 | 1847 | 14199 | 2.727 | 0.1115 | 15.29 | 4.34e-132 | 23534 | 1419 | 22115 | 0.4264 | -0.1299 | Paroxysmal tachycardia, unspecified | circulatory system | RATE_C | |
| 427.11 | 2.501 | 0.1757 | 12.19 | 5.52e-46 | 14852 | 653 | 14199 | 2.626 | 0.1357 | 13.81 | 1.95e-83 | 22978 | 863 | 22115 | 0.6681 | 0.09345 | Paroxysmal supraventricular tachycardia | circulatory system | RATE_C | |
| 427.12 | 3.076 | 0.1689 | 21.68 | 4.20e-74 | 15495 | 1296 | 14199 | 3.08 | 0.2037 | 21.76 | 1.22e-51 | 22690 | 575 | 22115 | 0.7245 | -0.09242 | Paroxysmal ventricular tachycardia | circulatory system | RATE_C | |
| 427.2 | 2.468 | 0.07357 | 11.8 | 8.64e-247 | 18883 | 4684 | 14199 | 2.667 | 0.08617 | 14.4 | 2.20e-210 | 25226 | 3111 | 22115 | 0.06085 | 0.2121 | Atrial fibrillation and flutter | circulatory system | RATE_C | |
| 427.21 | 2.456 | 0.07428 | 11.65 | 1.17e-239 | 18758 | 4559 | 14199 | 2.676 | 0.0882 | 14.53 | 3.01e-202 | 25130 | 3015 | 22115 | 0.04555 | 0.2302 | Atrial fibrillation | circulatory system | RATE_C | |
| 427.22 | 2.905 | 0.1589 | 18.27 | 1.19e-74 | 15660 | 1461 | 14199 | 3.207 | 0.2132 | 24.71 | 3.94e-51 | 22935 | 820 | 22115 | 0.1843 | 0.3524 | Atrial flutter | circulatory system | RATE_C | |
| 427.3 | 2.202 | 0.09679 | 9.047 | 1.30e-114 | 16128 | 1929 | 14199 | 2.38 | 0.08939 | 10.81 | 3.48e-156 | 23822 | 1707 | 22115 | 0.6357 | 0.06137 | Other specified cardiac dysrhythmias | circulatory system | RATE_C | |

| phenotype | male_beta | male_SE | male_OR | male_p | male_n_total | male_n_cases | male_n_controls | female_beta | female_SE | female_OR | female_p | female_n_total | female_n_cases | female_n_controls | interaction_p | interaction_beta | description | group | lab | label |
|---|---|---|---|---|---|---|---|---|---|---|---|---|---|---|---|---|---|---|---|---|
| 427.4 | | | | | 14682 | 483 | 14199 | | | | | 22371 | 256 | 22115 | 0.9055 | -0.03777 | Cardiac arrest and ventricular fibrillation | circulatory system | RATE_C | |
| 427.41 | | | | | 14406 | 207 | 14199 | | | | | 22188 | 73 | 22115 | | | Ventricular fibrillation and flutter | circulatory system | RATE_C | |
| 427.42 | | | | | 14543 | 344 | 14199 | | | | | 22324 | 209 | 22115 | 0.8355 | 0.06883 | Cardiac arrest | circulatory system | RATE_C | |
| 427.5 | 2.422 | 0.08493 | 11.27 | 6.29e-179 | 17123 | 2924 | 14199 | 2.534 | 0.08421 | 12.61 | 6.17e-199 | 24375 | 2260 | 22115 | 0.5099 | 0.07766 | Arrhythmia (cardiac) NOS | circulatory system | RATE_C | |
| 427.6 | 2.159 | 0.151 | 8.661 | 2.29e-46 | 15021 | 822 | 14199 | 2.284 | 0.145 | 9.812 | 6.47e-56 | 22723 | 608 | 22115 | 0.657 | -0.09142 | Premature beats | circulatory system | RATE_C | |
| 427.61 | | | | | 14305 | 106 | 14199 | | | | | 22243 | 128 | 22115 | | | Supraventricular premature beats | circulatory system | RATE_C | |
| 427.7 | 2.387 | 0.07422 | 10.88 | 5.63e-227 | 16928 | 2729 | 14199 | 2.453 | 0.05963 | 11.62 | 0 | 25298 | 3183 | 22115 | 0.9492 | 0.005823 | Tachycardia NOS | circulatory system | RATE_C | |
| 427.8 | 2.748 | 0.1825 | 15.61 | 3.21e-51 | 15296 | 1097 | 14199 | 3.102 | 0.2136 | 22.24 | 9.14e-48 | 22910 | 795 | 22115 | 0.2235 | 0.3416 | Sinoatrial node dysfunction (Bradycardia) | circulatory system | RATE_C | |
| 427.9 | 2.259 | 0.09283 | 9.576 | 7.85e-131 | 16102 | 1903 | 14199 | 2.319 | 0.05755 | 10.17 | 0 | 25899 | 3784 | 22115 | 0.9261 | -0.009794 | Palpitations | circulatory system | RATE_C | |
| 428 | 2.242 | 0.07587 | 9.409 | 7.47e-192 | 24029 | 4495 | 19534 | 2.197 | 0.07167 | 8.997 | 2.40e-206 | 33140 | 3590 | 29550 | 0.8655 | -0.01756 | Congestive heart failure; nonhypertensive | circulatory system | RATE_C | |
| 428.1 | 2.36 | 0.09043 | 10.59 | 4.08e-150 | 22921 | 3387 | 19534 | 2.439 | 0.0919 | 11.46 | 3.24e-155 | 32151 | 2601 | 29550 | 0.3943 | 0.1092 | Congestive heart failure (CHF) NOS | circulatory system | RATE_C | |
| 428.2 | 3.232 | 0.2215 | 25.34 | 3.20e-48 | 20752 | 1218 | 19534 | 3.144 | 0.2284 | 23.2 | 3.93e-43 | 30286 | 736 | 29550 | 0.7922 | -0.08348 | Heart failure NOS | circulatory system | RATE_C | |
| 428.3 | 2.473 | 0.1166 | 11.86 | 7.73e-100 | 21811 | 2277 | 19534 | 2.373 | 0.1208 | 10.73 | 7.66e-86 | 30905 | 1355 | 29550 | 0.4702 | -0.1204 | Heart failure with reduced EF [Systolic or combined heart failure] | circulatory system | RATE_C | |
| 428.4 | 2.493 | 0.1667 | 12.09 | 1.57e-50 | 20846 | 1312 | 19534 | 2.526 | 0.1358 | 12.51 | 2.73e-77 | 31066 | 1516 | 29550 | 0.7678 | 0.06327 | Heart failure with preserved EF [Diastolic heart failure] | circulatory system | RATE_C | |
| 429 | 1.911 | 0.1102 | 6.763 | 1.94e-67 | 21072 | 1538 | 19534 | 1.838 | 0.09622 | 6.281 | 2.63e-81 | 30921 | 1371 | 29550 | 0.5102 | -0.09473 | Ill-defined descriptions and complications of heart disease | circulatory system | RATE_C | |
| 429.1 | | | | | 19984 | 450 | 19534 | | | | | 29770 | 220 | 29550 | 0.2509 | -0.3128 | Heart transplant/surgery | circulatory system | RATE_C | |
| 429.2 | 1.592 | 0.1706 | 4.916 | 1.04e-20 | 20041 | 507 | 19534 | | | | | 29943 | 393 | 29550 | 0.2266 | 0.3021 | Abnormal function study of cardiovascular system | circulatory system | RATE_C | |
| 429.3 | 2.039 | 0.1859 | 7.685 | 5.24e-28 | 20117 | 583 | 19534 | 1.549 | 0.1204 | 4.708 | 6.42e-38 | 30233 | 683 | 29550 | 0.01513 | -0.5273 | Symptoms involving cardiovascular system | circulatory system | RATE_C | |
| 429.9 | | | | | 19772 | 238 | 19534 | | | | | 29698 | 148 | 29550 | | | Cardiac complications, not elsewhere classified | circulatory system | RATE_C | |
| 430 | 1.504 | 0.1388 | 4.498 | 2.44e-27 | 22249 | 653 | 21596 | 1.803 | 0.1485 | 6.07 | 5.94e-34 | 31365 | 529 | 30836 | 0.05653 | 0.3787 | Intracranial hemorrhage | circulatory system | RATE_C | |
| 430.1 | | | | | 21793 | 197 | 21596 | | | | | 31023 | 187 | 30836 | | | Subarachnoid hemorrhage | circulatory system | RATE_C | |
| 430.2 | | | | | 21911 | 315 | 21596 | | | | | 31093 | 257 | 30836 | 0.01318 | 0.7144 | Intracerebral hemorrhage | circulatory system | RATE_C | |
| 430.3 | | | | | 21798 | 202 | 21596 | | | | | 30969 | 133 | 30836 | | | Subdural hemorrhage | circulatory system | RATE_C | |
| 433 | 1.314 | 0.06438 | 3.72 | 1.48e-92 | 25148 | 3552 | 21596 | 1.467 | 0.0574 | 4.334 | 5.47e-144 | 34408 | 3572 | 30836 | 0.03472 | 0.1798 | Cerebrovascular disease | circulatory system | RATE_C | |
| 433.1 | 1.882 | 0.1334 | 6.566 | 3.32e-45 | 23051 | 1455 | 21596 | 1.654 | 0.1106 | 5.23 | 1.49e-50 | 32011 | 1175 | 30836 | 0.1762 | -0.233 | Occlusion and stenosis of precerebral arteries | circulatory system | RATE_C | |
| 433.11 | | | | | 21806 | 210 | 21596 | | | | | 31009 | 173 | 30836 | | | Occlusion of cerebral arteries, with cerebral infarction | circulatory system | RATE_C | |
| 433.12 | | | | | 21766 | 170 | 21596 | | | | | 30990 | 154 | 30836 | | | Cerebral atherosclerosis | circulatory system | RATE_C | |
| 433.2 | 1.607 | 0.1168 | 4.989 | 4.08e-43 | 22816 | 1220 | 21596 | 1.811 | 0.1101 | 6.115 | 8.05e-61 | 32003 | 1167 | 30836 | 0.08647 | 0.2712 | Occlusion of cerebral arteries | circulatory system | RATE_C | |
| 433.21 | 1.612 | 0.1196 | 5.014 | 1.95e-41 | 22764 | 1168 | 21596 | 1.835 | 0.1142 | 6.265 | 3.85e-58 | 31945 | 1109 | 30836 | 0.07188 | 0.2934 | Cerebral artery occlusion, with cerebral infarction | circulatory system | RATE_C | |

| phenotype | male_beta | male_SE | male_OR | male_p | male_n_total | male_n_cases | male_n_controls | female_beta | female_SE | female_OR | female_p | female_n_total | female_n_cases | female_n_controls | interaction_p | interaction_beta | description | group | lab | label |
|---|---|---|---|---|---|---|---|---|---|---|---|---|---|---|---|---|---|---|---|---|
| 433.3 | 1.745 | 0.1099 | 5.728 | 8.43e-57 | 23331 | 1735 | 21596 | 1.794 | 0.09262 | 6.014 | 1.38e-83 | 32595 | 1759 | 30836 | 0.6449 | 0.06563 | Cerebral ischemia | circulatory system | RATE_C | |
| 433.31 | 1.745 | 0.1142 | 5.724 | 1.11e-52 | 23194 | 1598 | 21596 | 1.842 | 0.09837 | 6.309 | 3.16e-78 | 32452 | 1616 | 30836 | 0.4414 | 0.115 | Transient cerebral ischemia | circulatory system | RATE_C | |
| 433.32 | | | | | 21602 | 6 | 21596 | | | | | 30850 | 14 | 30836 | | | Moyamoya disease | circulatory system | RATE_C | |
| 433.5 | | | | | 21737 | 141 | 21596 | | | | | 31155 | 319 | 30836 | | | Cerebral aneurysm | circulatory system | RATE_C | |
| 433.6 | | | | | 21926 | 330 | 21596 | | | | | 31171 | 335 | 30836 | 0.1821 | 0.5676 | Acute, but ill-defined cerebrovascular disease | circulatory system | RATE_C | |
| 433.8 | 1.075 | 0.1293 | 2.929 | 9.41e-17 | 22246 | 650 | 21596 | 1.47 | 0.13 | 4.35 | 1.21e-29 | 31470 | 634 | 30836 | 0.009473 | 0.4657 | Late effects of cerebrovascular disease | circulatory system | RATE_C | |
| 440 | 2.133 | 0.1373 | 8.439 | 2.09e-54 | 21894 | 1543 | 20351 | 1.788 | 0.1136 | 5.976 | 8.11e-56 | 31649 | 1262 | 30387 | 0.05015 | -0.3473 | Atherosclerosis | circulatory system | RATE_C | |
| 440.1 | | | | | 20496 | 145 | 20351 | | | | | 30601 | 214 | 30387 | | | Atherosclerosis of renal artery | circulatory system | RATE_C | |
| 440.2 | 2.401 | 0.2185 | 11.03 | 4.31e-28 | 21065 | 714 | 20351 | | | | | 30809 | 422 | 30387 | 0.06189 | -0.5395 | Atherosclerosis of the extremities | circulatory system | RATE_C | |
| 440.21 | | | | | 20544 | 193 | 20351 | | | | | 30472 | 85 | 30387 | | | Atherosclerosis of native arteries of the extremities with ulceration or gangrene | circulatory system | RATE_C | |
| 440.22 | | | | | 20765 | 414 | 20351 | | | | | 30616 | 229 | 30387 | 0.7082 | -0.141 | Atherosclerosis of native arteries of the extremities with intermittent claudication | circulatory system | RATE_C | |
| 440.9 | | | | | 20820 | 469 | 20351 | | | | | 30841 | 454 | 30387 | 0.04376 | -0.681 | Atherosclerosis of aorta | circulatory system | RATE_C | |
| 441 | | | | | 20539 | 188 | 20351 | | | | | 30626 | 239 | 30387 | | | Vascular insufficiency of intestine | circulatory system | RATE_C | |
| 441.1 | | | | | 20421 | 70 | 20351 | | | | | 30462 | 75 | 30387 | | | Acute vascular insufficiency of intestine | circulatory system | RATE_C | |
| 441.2 | | | | | 20385 | 34 | 20351 | | | | | 30454 | 67 | 30387 | | | Chronic vascular insufficiency of intestine | circulatory system | RATE_C | |
| 442 | 1.685 | 0.1211 | 5.394 | 5.16e-44 | 21609 | 1258 | 20351 | 1.445 | 0.1252 | 4.241 | 8.63e-31 | 31061 | 674 | 30387 | 0.04056 | -0.3517 | Other aneurysm | circulatory system | RATE_C | |
| 442.1 | 1.743 | 0.1401 | 5.717 | 1.47e-35 | 21366 | 1015 | 20351 | | | | | 30810 | 423 | 30387 | 0.2876 | -0.2327 | Aortic aneurysm | circulatory system | RATE_C | |
| 442.11 | 1.804 | 0.193 | 6.076 | 8.71e-21 | 20965 | 614 | 20351 | | | | | 30570 | 183 | 30387 | 0.6621 | -0.1515 | Abdominal aortic aneurysm | circulatory system | RATE_C | |
| 442.2 | | | | | 20421 | 70 | 20351 | | | | | 30397 | 10 | 30387 | | | Aneurysm of iliac artery | circulatory system | RATE_C | |
| 442.3 | | | | | 20442 | 91 | 20351 | | | | | 30441 | 54 | 30387 | | | Aneurysm of artery of lower extremity | circulatory system | RATE_C | |

| phenotype | male_beta | male_SE | male_OR | male_p | male_n_total | male_n_cases | male_n_controls | female_beta | female_SE | female_OR | female_p | female_n_total | female_n_cases | female_n_controls | interaction_p | interaction_beta | description | group | lab | label |
|---|---|---|---|---|---|---|---|---|---|---|---|---|---|---|---|---|---|---|---|---|
| 442.4 | | | | | 20394 | 43 | 20351 | | | | | 30441 | 54 | 30387 | | | Arterial dissection | circulatory system | RATE_C | |
| 442.8 | | | | | 20454 | 103 | 20351 | | | | | 30512 | 125 | 30387 | | | Aneurysm of other specified artery | circulatory system | RATE_C | |
| 443 | 1.637 | 0.1042 | 5.139 | 1.36e-55 | 22049 | 1698 | 20351 | 0.8993 | 0.07176 | 2.458 | 4.95e-36 | 31903 | 1516 | 30387 | 3.45e-11 | -0.826 | Peripheral vascular disease | circulatory system | RATE_C | Peripheral vascular disease |
| 443.1 | | | | | 20444 | 93 | 20351 | | | | | 30852 | 465 | 30387 | 0.2995 | -0.2658 | Raynaud's syndrome | circulatory system | RATE_C | |
| 443.7 | | | | | 20611 | 260 | 20351 | | | | | 30517 | 130 | 30387 | | | Peripheral angiopathy in diseases classified elsewhere | circulatory system | RATE_C | |
| 443.8 | | | | | 20448 | 97 | 20351 | | | | | 30453 | 66 | 30387 | | | Other specified peripheral vascular diseases | circulatory system | RATE_C | |

| phenotype | male_beta | male_SE | male_OR | male_p | male_n_total | male_n_cases | male_n_controls | female_beta | female_SE | female_OR | female_p | female_n_total | female_n_cases | female_n_controls | interaction_p | interaction_beta | description | group | lab | label |
|---|---|---|---|---|---|---|---|---|---|---|---|---|---|---|---|---|---|---|---|---|
| 443.9 | 1.854 | 0.1252 | 6.387 | 1.27e-49 | 21791 | 1440 | 20351 | 1.751 | 0.1267 | 5.759 | 1.86e-43 | 31349 | 962 | 30387 | 0.5319 | -0.1105 | Peripheral vascular disease, unspecified | circulatory system | RATE_C | |
| 444 | | | | | 20661 | 310 | 20351 | | | | | 30633 | 246 | 30387 | 0.2353 | -0.3832 | Arterial embolism and thrombosis | circulatory system | RATE_C | |
| 444.1 | | | | | 20465 | 114 | 20351 | | | | | 30462 | 75 | 30387 | | | Arterial embolism and thrombosis of lower extremity artery | circulatory system | RATE_C | |
| 444.2 | | | | | 20391 | 40 | 20351 | | | | | 30424 | 37 | 30387 | | | Embolism and thrombosis of abdominal aorta | circulatory system | RATE_C | |
| 444.5 | | | | | 20357 | 6 | 20351 | | | | | 30392 | 5 | 30387 | | | Atheroembolism | circulatory system | RATE_C | |
| 446 | | | | | 20562 | 211 | 20351 | | | | | 30776 | 389 | 30387 | 0.1603 | -0.2684 | Polyarteritis nodosa and allied conditions | circulatory system | RATE_C | |
| 446.1 | | | | | 20353 | 2 | 20351 | | | | | 30391 | 4 | 30387 | | | Thromboangiitis obliterans | circulatory system | RATE_C | |
| 446.2 | | | | | 20351 | | | | | | | 30393 | 6 | 30387 | | | Acute febrile mucocutaneous lymph node syndrome (Kawasaki disease) | circulatory system | RATE_C | |
| 446.3 | | | | | 20377 | 26 | 20351 | | | | | 30419 | 32 | 30387 | | | Hypersensitivity angiitis | circulatory system | RATE_C | |
| 446.4 | | | | | 20429 | 78 | 20351 | | | | | 30467 | 80 | 30387 | | | Wegener's granulomatosis | circulatory system | RATE_C | |
| 446.5 | | | | | 20386 | 35 | 20351 | | | | | 30500 | 113 | 30387 | | | Giant cell arteritis | circulatory system | RATE_C | |
| 446.6 | | | | | 20380 | 29 | 20351 | | | | | 30423 | 36 | 30387 | | | Polyarteritis nodosa | circulatory system | RATE_C | |
| 446.7 | | | | | 20353 | 2 | 20351 | | | | | 30403 | 16 | 30387 | | | Takayasu's disease | circulatory system | RATE_C | |
| 446.8 | | | | | 20362 | 11 | 20351 | | | | | 30409 | 22 | 30387 | | | Thrombotic microangiopathy | circulatory system | RATE_C | |
| 446.9 | | | | | 20443 | 92 | 20351 | | | | | 30554 | 167 | 30387 | | | Arteritis NOS | circulatory system | RATE_C | |
| 447 | 1.677 | 0.1308 | 5.347 | 1.29e-37 | 21399 | 1048 | 20351 | 1.304 | 0.1347 | 3.685 | 3.63e-22 | 30938 | 551 | 30387 | 0.03557 | -0.3871 | Other disorders of arteries and arterioles | circulatory system | RATE_C | |
| 447.1 | | | | | 20462 | 111 | 20351 | | | | | 30487 | 100 | 30387 | | | Stricture of artery | circulatory system | RATE_C | |
| 447.7 | 1.742 | 0.1665 | 5.706 | 1.36e-25 | 21056 | 705 | 20351 | | | | | 30596 | 209 | 30387 | 0.06818 | -0.4922 | Aortic ectasia | circulatory system | RATE_C | |
| 448 | | | | | 20358 | 7 | 20351 | | | | | 30410 | 23 | 30387 | | | Disease of capillaries | circulatory system | RATE_C | |
| 450 | | | | | 27464 | 390 | 27074 | 0.8287 | 0.09919 | 2.29 | 6.59e-17 | 36525 | 739 | 35786 | 0.0207 | 0.3786 | Noninfectious disorders of lymphatic channels | circulatory system | RATE_C | |
| 451 | | | | | 17675 | 229 | 17446 | | | | | 26197 | 265 | 25932 | | | Phlebitis and thrombophlebitis | circulatory system | RATE_C | |
| 451.2 | | | | | 17551 | 105 | 17446 | | | | | 26069 | 137 | 25932 | | | Phlebitis and thrombophlebitis of lower extremities | circulatory system | RATE_C | |
| 452 | 1.511 | 0.07415 | 4.532 | 2.58e-92 | 19497 | 2051 | 17446 | 1.698 | 0.06817 | 5.462 | 6.43e-137 | 28016 | 2084 | 25932 | 0.006041 | 0.2667 | Other venous embolism and thrombosis | circulatory system | RATE_C | |
| 452.1 | | | | | 17477 | 31 | 17446 | | | | | 25960 | 28 | 25932 | | | Iatrogenic pulmonary embolism and infarction | circulatory system | RATE_C | |
| 452.2 | 1.389 | 0.0969 | 4.01 | 1.39e-46 | 18523 | 1077 | 17446 | 1.714 | 0.09423 | 5.554 | 5.76e-74 | 26996 | 1064 | 25932 | 0.004319 | 0.3718 | Deep vein thrombosis [DVT] | circulatory system | RATE_C | |
| 452.8 | | | | | 17477 | 31 | 17446 | | | | | 25956 | 24 | 25932 | | | Postphlebitic syndrome | circulatory system | RATE_C | |
| 453 | | | | | 17473 | 27 | 17446 | | | | | 25945 | 13 | 25932 | | | Chronic venous hypertension | circulatory system | RATE_C | |
| 454 | | | | | 17905 | 459 | 17446 | 0.7847 | 0.105 | 2.192 | 7.90e-14 | 26542 | 610 | 25932 | 0.1517 | 0.2283 | Varicose veins | circulatory system | RATE_C | |
| 454.1 | | | | | 17712 | 266 | 17446 | 0.6627 | 0.1144 | 1.94 | 6.89e-09 | 26438 | 506 | 25932 | 0.7301 | 0.06957 | Varicose veins of lower extremity | circulatory system | RATE_C | |

| phenotype | male_beta | male_SE | male_OR | male_p | male_n_total | male_n_cases | male_n_controls | female_beta | female_SE | female_OR | female_p | female_n_total | female_n_cases | female_n_controls | interaction_p | interaction_beta | description | group | lab | label |
|---|---|---|---|---|---|---|---|---|---|---|---|---|---|---|---|---|---|---|---|---|
| 454.11 | | | | | 17657 | 211 | 17446 | | | | | 26297 | 365 | 25932 | 0.964 | -0.0104 | Varicose veins of lower extremity, symptomtic | circulatory system | RATE_C | |
| 455 | 0.7764 | 0.08305 | 2.174 | 8.84e-21 | 18658 | 1212 | 17446 | 0.8248 | 0.0702 | 2.281 | 7.15e-32 | 27261 | 1329 | 25932 | 0.4865 | 0.07166 | Hemorrhoids | circulatory system | RATE_C | |
| 456 | | | | | 17849 | 403 | 17446 | | | | | 26402 | 470 | 25932 | 0.7569 | -0.06975 | Chronic venous insufficiency [CVI] | circulatory system | RATE_C | |
| 457 | 1.963 | 0.09575 | 7.122 | 2.05e-93 | 19943 | 2497 | 17446 | 1.878 | 0.09009 | 6.543 | 1.54e-96 | 27763 | 1831 | 25932 | 0.6034 | -0.06742 | Encounter for long-term (current) use of anticoagulants, antithrombotics, aspirin | circulatory system | RATE_C | |
| 457.2 | 2.29 | 0.2369 | 9.874 | 4.16e-22 | 17977 | 531 | 17446 | | | | | 26262 | 330 | 25932 | 0.4801 | -0.2281 | Encounter for long-term (current) use of antiplatelets/antithrombotics | circulatory system | RATE_C | |
| 457.3 | 1.993 | 0.1008 | 7.339 | 4.94e-87 | 19733 | 2287 | 17446 | 1.888 | 0.0946 | 6.604 | 1.40e-88 | 27596 | 1664 | 25932 | 0.5465 | -0.08229 | Encounter for long-term (current) use of aspirin | circulatory system | RATE_C | |
| 458 | 1.989 | 0.07252 | 7.308 | 1.42e-165 | 23987 | 3579 | 20408 | 2.024 | 0.06856 | 7.566 | 1.69e-191 | 32936 | 2891 | 30045 | 0.8477 | 0.01874 | Hypotension | circulatory system | RATE_C | |
| 458.1 | 1.667 | 0.153 | 5.295 | 1.18e-27 | 21124 | 716 | 20408 | 1.82 | 0.1503 | 6.174 | 8.96e-34 | 30599 | 554 | 30045 | 0.875 | 0.03301 | Orthostatic hypotension | circulatory system | RATE_C | |
| 458.2 | | | | | 20542 | 134 | 20408 | | | | | 30168 | 123 | 30045 | | | Iatrogenic hypotension | circulatory system | RATE_C | |
| 458.9 | 2.098 | 0.08397 | 8.147 | 9.99e-138 | 23196 | 2788 | 20408 | 2.101 | 0.07948 | 8.174 | 5.68e-154 | 32272 | 2227 | 30045 | 0.9262 | 0.01048 | Hypotension NOS | circulatory system | RATE_C | |
| 459 | 1.562 | 0.1176 | 4.771 | 2.59e-40 | 21377 | 969 | 20408 | 1.693 | 0.1112 | 5.434 | 2.45e-52 | 30928 | 883 | 30045 | 0.3515 | 0.1457 | Other disorders of circulatory system | circulatory system | RATE_C | |
| 459.1 | | | | | 20541 | 133 | 20408 | | | | | 30175 | 130 | 30045 | | | Hemorrhage NOS | circulatory system | RATE_C | |
| 459.7 | | | | | 20566 | 158 | 20408 | | | | | 30162 | 117 | 30045 | | | Blood vessel replaced | circulatory system | RATE_C | |
| 459.9 | 1.853 | 0.1727 | 6.378 | 7.31e-27 | 21001 | 593 | 20408 | 1.827 | 0.1578 | 6.212 | 5.50e-31 | 30563 | 518 | 30045 | 0.9389 | 0.01748 | Circulatory disease NEC | circulatory system | RATE_C | |
| 464 | 0.1034 | 0.0713 | 1.109 | 0.1469 | 22311 | 1470 | 20841 | 0.3361 | 0.04427 | 1.4 | 3.12e-14 | 28085 | 3592 | 24493 | 0.00191 | 0.2383 | Acute sinusitis | respiratory | RATE_C | |
| 465 | -0.03889 | 0.05169 | 0.9619 | 0.4519 | 23812 | 2971 | 20841 | 0.2286 | 0.03532 | 1.257 | 9.67e-11 | 30856 | 6363 | 24493 | 5.83e-06 | 0.2539 | Acute upper respiratory infections of multiple or unspecified sites | respiratory | RATE_C | |
| 465.2 | -0.04731 | 0.08085 | 0.9538 | 0.5584 | 21831 | 990 | 20841 | 0.2071 | 0.04901 | 1.23 | 2.39e-05 | 27189 | 2696 | 24493 | 6.40e-04 | 0.2855 | Acute pharyngitis | respiratory | RATE_C | |
| 465.4 | | | | | 20905 | 64 | 20841 | | | | | 24590 | 97 | 24493 | | | Acute laryngitis and tracheitis | respiratory | RATE_C | |
| 470 | 0.374 | 0.088 | 1.453 | 2.14e-05 | 19858 | 932 | 18926 | 0.4597 | 0.06918 | 1.584 | 3.03e-11 | 25321 | 1191 | 24130 | 0.4524 | 0.07719 | Septal Deviations/Turbinate Hypertrophy | respiratory | RATE_C | |
| 471 | | | | | 19079 | 153 | 18926 | | | | | 24245 | 115 | 24130 | | | Nasal polyps | respiratory | RATE_C | |
| 472 | 0.2912 | 0.1024 | 1.338 | 0.004464 | 19621 | 695 | 18926 | 0.5215 | 0.07175 | 1.684 | 3.65e-13 | 25328 | 1198 | 24130 | 0.03098 | 0.2503 | Chronic pharyngitis and nasopharyngitis | respiratory | RATE_C | |
| 473 | 0.5848 | 0.0904 | 1.795 | 9.89e-11 | 19940 | 1014 | 18926 | 0.8979 | 0.07492 | 2.454 | 4.30e-33 | 25397 | 1267 | 24130 | 0.00602 | 0.3063 | Diseases of the larynx and vocal cords | respiratory | RATE_C | |
| 473.1 | | | | | 19013 | 87 | 18926 | | | | | 24250 | 120 | 24130 | | | Chronic laryngitis | respiratory | RATE_C | |
| 473.3 | | | | | 19140 | 214 | 18926 | | | | | 24368 | 238 | 24130 | | | Paralysis/spasm of vocal cords or larynx | respiratory | RATE_C | |
| 473.4 | 0.572 | 0.1043 | 1.772 | 4.13e-08 | 19690 | 764 | 18926 | 0.8643 | 0.08451 | 2.373 | 1.50e-24 | 25123 | 993 | 24130 | 0.01754 | 0.3035 | Voice disturbance | respiratory | RATE_C | |

| phenotype | male_beta | male_SE | male_OR | male_p | male_n_total | male_n_cases | male_n_controls | female_beta | female_SE | female_OR | female_p | female_n_total | female_n_cases | female_n_controls | interaction_p | interaction_beta | description | group | lab | label |
|---|---|---|---|---|---|---|---|---|---|---|---|---|---|---|---|---|---|---|---|---|
| 474 | | | | | 19193 | 267 | 18926 | | | | | 24615 | 485 | 24130 | 0.3586 | 0.1487 | Acute and chronic tonsillitis | respiratory | RATE_C | |
| 474.1 | | | | | 18953 | 27 | 18926 | | | | | 24211 | 81 | 24130 | | | Acute tonsillitis | respiratory | RATE_C | |

| phenotype | male_beta | male_SE | male_OR | male_p | male_n_total | male_n_cases | male_n_controls | female_beta | female_SE | female_OR | female_p | female_n_total | female_n_cases | female_n_controls | interaction_p | interaction_beta | description | group | lab | label |
|---|---|---|---|---|---|---|---|---|---|---|---|---|---|---|---|---|---|---|---|---|
| 474.2 | | | | | 19156 | 230 | 18926 | | | | | 24527 | 397 | 24130 | 0.2461 | 0.2053 | Chronic tonsillitis and adenoiditis | respiratory | RATE_C | |
| 475 | 0.4509 | 0.07475 | 1.57 | 1.62e-09 | 20308 | 1382 | 18926 | 0.6725 | 0.05517 | 1.959 | 3.60e-34 | 26319 | 2189 | 24130 | 3.13e-04 | 0.3093 | Chronic sinusitis | respiratory | RATE_C | |
| 475.9 | | | | | 19118 | 192 | 18926 | | | | | 24485 | 355 | 24130 | 0.7929 | 0.05887 | Postnasal drip | respiratory | RATE_C | |
| 476 | 0.07659 | 0.05369 | 1.08 | 0.1537 | 21597 | 2671 | 18926 | 0.3216 | 0.03818 | 1.379 | 3.68e-17 | 28998 | 4868 | 24130 | 1.41e-07 | 0.3162 | Allergic rhinitis | respiratory | RATE_C | |
| 477 | | | | | 19327 | 401 | 18926 | | | | | 24561 | 431 | 24130 | 0.6107 | 0.0946 | Epistaxis or throat hemorrhage | respiratory | RATE_C | |
| 478 | | | | | 19150 | 224 | 18926 | | | | | 24407 | 277 | 24130 | 0.3765 | 0.1768 | Throat pain | respiratory | RATE_C | |
| 479 | 0.5464 | 0.07439 | 1.727 | 2.06e-13 | 20272 | 1346 | 18926 | 0.654 | 0.05775 | 1.923 | 9.86e-30 | 26030 | 1900 | 24130 | 0.002455 | 0.2619 | Other upper respiratory disease | respiratory | RATE_C | |
| 480 | 1.451 | 0.05839 | 4.268 | 2.61e-136 | 23852 | 3704 | 20148 | 1.525 | 0.05327 | 4.595 | 3.18e-180 | 31060 | 3539 | 27521 | 0.002146 | 0.2324 | Pneumonia | respiratory | RATE_C | |
| 480.1 | 1.23 | 0.1013 | 3.42 | 6.44e-34 | 21107 | 959 | 20148 | 1.45 | 0.1047 | 4.263 | 1.34e-43 | 28349 | 828 | 27521 | 0.001128 | 0.4508 | Bacterial pneumonia | respiratory | RATE_C | |
| 480.11 | | | | | 20392 | 244 | 20148 | | | | | 27735 | 214 | 27521 | | | Pneumococcal pneumonia | respiratory | RATE_C | |
| 480.12 | | | | | 20313 | 165 | 20148 | | | | | 27625 | 104 | 27521 | | | Pseudomonal pneumonia | respiratory | RATE_C | |
| 480.13 | | | | | 20243 | 95 | 20148 | | | | | 27596 | 75 | 27521 | | | MRSA pneumonia | respiratory | RATE_C | |
| 480.2 | | | | | 20252 | 104 | 20148 | | | | | 27629 | 108 | 27521 | | | Viral pneumonia | respiratory | RATE_C | |
| 480.3 | | | | | 20261 | 113 | 20148 | | | | | 27595 | 74 | 27521 | | | Pneumonia due to fungus (mycoses) | respiratory | RATE_C | |
| 480.5 | | | | | 20273 | 125 | 20148 | | | | | 27647 | 126 | 27521 | | | Bronchopneumonia and lung abscess | respiratory | RATE_C | |
| 481 | | | | | 20437 | 289 | 20148 | | | | | 27934 | 413 | 27521 | 0.4033 | 0.1475 | Influenza | respiratory | RATE_C | |
| 483 | 0.6009 | 0.08974 | 1.824 | 2.15e-11 | 21180 | 1032 | 20148 | 0.6754 | 0.06097 | 1.965 | 1.62e-28 | 29347 | 1826 | 27521 | 0.1084 | 0.1629 | Acute bronchitis and bronchiolitis | respiratory | RATE_C | |
| 495 | 0.5558 | 0.06849 | 1.743 | 4.87e-16 | 22371 | 1521 | 20850 | 0.7786 | 0.04511 | 2.178 | 9.40e-67 | 31168 | 3223 | 27945 | 1.01e-06 | 0.3653 | Asthma | respiratory | RATE_C | |
| 495.1 | | | | | 21010 | 160 | 20850 | | | | | 28173 | 228 | 27945 | | | Chronic obstructive asthma | respiratory | RATE_C | |
| 495.11 | | | | | 20869 | 19 | 20850 | | | | | 27991 | 46 | 27945 | | | Chronic obstructive asthma with exacerbation | respiratory | RATE_C | |
| 495.2 | | | | | 21125 | 275 | 20850 | 0.8118 | 0.09033 | 2.252 | 2.56e-19 | 28693 | 748 | 27945 | 7.81e-05 | 0.6231 | Asthma with exacerbation | respiratory | RATE_C | |
| 496 | 1.317 | 0.0658 | 3.734 | 3.57e-89 | 23991 | 3141 | 20850 | 1.549 | 0.06241 | 4.708 | 4.93e-136 | 30852 | 2907 | 27945 | 0.001839 | 0.2762 | Chronic airway obstruction | respiratory | RATE_C | |
| 496.1 | 1.337 | 0.1197 | 3.807 | 6.05e-29 | 21816 | 966 | 20850 | 1.802 | 0.1381 | 6.064 | 6.21e-39 | 28671 | 726 | 27945 | 0.005227 | 0.5009 | Emphysema | respiratory | RATE_C | |
| 496.2 | 1.349 | 0.1319 | 3.853 | 1.49e-24 | 21707 | 857 | 20850 | 1.725 | 0.1203 | 5.613 | 1.22e-46 | 28880 | 935 | 27945 | 0.02433 | 0.3942 | Chronic bronchitis | respiratory | RATE_C | |
| 496.21 | 1.547 | 0.1608 | 4.697 | 6.51e-22 | 21544 | 694 | 20850 | 1.885 | 0.1446 | 6.589 | 6.99e-39 | 28686 | 741 | 27945 | 0.1226 | 0.3289 | Obstructive chronic bronchitis | respiratory | RATE_C | |
| 496.3 | | | | | 21106 | 256 | 20850 | | | | | 28305 | 360 | 27945 | 3.68e-04 | 0.7001 | Bronchiectasis | respiratory | RATE_C | |
| 497 | | | | | 21118 | 268 | 20850 | | | | | 28430 | 485 | 27945 | 0.07162 | 0.3704 | Bronchitis | respiratory | RATE_C | |
| 498 | | | | | 20873 | 23 | 20850 | | | | | 27982 | 37 | 27945 | | | Acute bronchospasm | respiratory | RATE_C | |
| 499 | | | | | 27770 | 89 | 27681 | | | | | 36921 | 77 | 36844 | | | Cystic fibrosis | respiratory | RATE_C | |

| phenotype | male_beta | male_SE | male_OR | male_p | male_n_total | male_n_cases | male_n_controls | female_beta | female_SE | female_OR | female_p | female_n_total | female_n_cases | female_n_controls | interaction_p | interaction_beta | description | group | lab | label |
|---|---|---|---|---|---|---|---|---|---|---|---|---|---|---|---|---|---|---|---|---|
| 500 | | | | | 16503 | 145 | 16358 | | | | | 26637 | 93 | 26544 | | | Lung disease due to external agents | respiratory | RATE_C | |
| 500.1 | | | | | 16369 | 11 | 16358 | | | | | 26562 | 18 | 26544 | | | Extrinsic allergic alveolitis | respiratory | RATE_C | |
| 500.2 | | | | | 16384 | 26 | 16358 | | | | | 26547 | 3 | 26544 | | | Pneumoconiosis | respiratory | RATE_C | |
| 501 | 1.809 | 0.1492 | 6.106 | 7.43e-34 | 16908 | 550 | 16358 | | | | | 26905 | 361 | 26544 | 0.2261 | 0.2717 | Pneumonitis due to inhalation of food or vomitus | respiratory | RATE_C | |
| 502 | | | | | 16775 | 417 | 16358 | | | | | 26948 | 404 | 26544 | 0.2524 | -0.2314 | Postinflammatory pulmonary fibrosis | respiratory | RATE_C | |
| 503 | 2.475 | 0.1378 | 11.88 | 3.68e-72 | 17413 | 1055 | 16358 | 2.606 | 0.144 | 13.54 | 3.45e-73 | 27379 | 835 | 26544 | 0.2923 | 0.2058 | Pulmonary congestion and hypostasis | respiratory | RATE_C | |
| 504 | | | | | 16686 | 328 | 16358 | | | | | 26850 | 306 | 26544 | 0.01067 | -0.5594 | Other alveolar and parietoalveolar pneumonopathy | respiratory | RATE_C | |
| 504.1 | | | | | 16528 | 170 | 16358 | | | | | 26674 | 130 | 26544 | | | Idiopathic fibrosing alveolitis | respiratory | RATE_C | |
| 505 | | | | | 16583 | 225 | 16358 | | | | | 26786 | 242 | 26544 | | | Other pulmonary inflamation or edema | respiratory | RATE_C | |
| 506 | 1.983 | 0.09641 | 7.261 | 5.79e-94 | 17754 | 1396 | 16358 | 1.979 | 0.09585 | 7.236 | 1.02e-94 | 27619 | 1075 | 26544 | 0.9471 | 0.008738 | Empyema and pneumothorax | respiratory | RATE_C | |
| 507 | 2.245 | 0.06662 | 9.443 | 5.28e-249 | 20407 | 4049 | 16358 | 2.235 | 0.06377 | 9.344 | 5.28e-269 | 29890 | 3346 | 26544 | 0.7269 | 0.03147 | Pleurisy; pleural effusion | respiratory | RATE_C | |
| 508 | 2.365 | 0.05861 | 10.64 | 0 | 22186 | 5828 | 16358 | 2.404 | 0.05902 | 11.07 | 0 | 30908 | 4364 | 26544 | 0.4345 | 0.06344 | Pulmonary collapse; interstitial and compensatory emphysema | respiratory | RATE_C | |
| 509 | 2.144 | 0.06068 | 8.535 | 1.63e-273 | 20689 | 4331 | 16358 | 2.198 | 0.05972 | 9.01 | 1.18e-296 | 30174 | 3630 | 26544 | 0.08283 | 0.1435 | Respiratory failure, insufficiency, arrest | respiratory | RATE_C | |
| 509.1 | 2.143 | 0.07249 | 8.528 | 4.13e-192 | 19312 | 2954 | 16358 | 2.186 | 0.07042 | 8.903 | 1.27e-211 | 29134 | 2590 | 26544 | 0.1147 | 0.1551 | Respiratory failure | respiratory | RATE_C | |
| 509.2 | 3.339 | 0.1976 | 28.2 | 4.75e-64 | 17397 | 1039 | 16358 | 3.022 | 0.1712 | 20.53 | 9.59e-70 | 27339 | 795 | 26544 | 0.3345 | -0.2494 | Respiratory insufficiency | respiratory | RATE_C | |
| 509.3 | 2.099 | 0.111 | 8.156 | 1.06e-79 | 17365 | 1007 | 16358 | 2.306 | 0.1308 | 10.03 | 1.41e-69 | 27216 | 672 | 26544 | 0.07032 | 0.3006 | Pulmonary insufficiency or respiratory failure following trauma and surgery | respiratory | RATE_C | |
| 509.5 | | | | | 16395 | 37 | 16358 | | | | | 26570 | 26 | 26544 | | | Respiratory arrest | respiratory | RATE_C | |
| 509.8 | 1.875 | 0.1252 | 6.523 | 1.02e-50 | 17139 | 781 | 16358 | 2.032 | 0.1298 | 7.628 | 3.46e-55 | 27262 | 718 | 26544 | 0.03811 | 0.3632 | Dependence on respirator [Ventilator] or supplemental oxygen | respiratory | RATE_C | |
| 510 | 1.626 | 0.07173 | 5.081 | 1.07e-113 | 24918 | 2670 | 22248 | 1.746 | 0.06991 | 5.732 | 1.08e-137 | 34185 | 2275 | 31910 | 0.02466 | 0.2172 | Other diseases of lung | respiratory | RATE_C | |
| 510.2 | | | | | 22355 | 107 | 22248 | | | | | 32017 | 107 | 31910 | | | Lung transplant | respiratory | RATE_C | |
| 512 | 1.566 | 0.03614 | 4.787 | 0 | 23558 | 11275 | 12283 | 1.513 | 0.02902 | 4.54 | 0 | 31474 | 13478 | 17996 | 0.7875 | -0.01184 | Other symptoms of respiratory system | respiratory | RATE_C | |
| 512.1 | | | | | 12736 | 453 | 12283 | 1.581 | 0.1012 | 4.858 | 4.84e-55 | 18716 | 720 | 17996 | 0.03104 | 0.3445 | Wheezing | respiratory | RATE_C | |
| 512.2 | | | | | 12736 | 453 | 12283 | 2.472 | 0.1407 | 11.84 | 4.81e-69 | 18591 | 595 | 17996 | 0.116 | 0.3349 | Painful respiration | respiratory | RATE_C | |
| 512.3 | | | | | 12290 | 7 | 12283 | | | | | 18006 | 10 | 17996 | | | Abnormal chest sounds | respiratory | RATE_C | |
| 512.7 | 2.031 | 0.05085 | 7.625 | 0 | 19002 | 6719 | 12283 | 2.076 | 0.0418 | 7.972 | 0 | 25374 | 7378 | 17996 | 0.1824 | 0.08563 | Shortness of breath | respiratory | RATE_C | |
| 512.8 | 1.146 | 0.0494 | 3.146 | 4.33e-119 | 16541 | 4258 | 12283 | 1.214 | 0.03724 | 3.367 | 4.55e-233 | 24362 | 6366 | 17996 | 0.02023 | 0.1348 | Cough | respiratory | RATE_C | |
| 512.9 | 1.973 | 0.05519 | 7.194 | 5.77e-280 | 17400 | 5117 | 12283 | 2.032 | 0.04703 | 7.628 | 0 | 23359 | 5363 | 17996 | 0.027 | 0.1555 | Other dyspnea | respiratory | RATE_C | |
| 513 | 1.458 | 0.106 | 4.296 | 5.15e-43 | 26022 | 945 | 25077 | 1.748 | 0.1119 | 5.745 | 5.06e-55 | 35312 | 811 | 34501 | 0.001302 | 0.4695 | Respiratory abnormalities | respiratory | RATE_C | |
| 513.3 | | | | | 25286 | 209 | 25077 | | | | | 34707 | 206 | 34501 | | | Hypoventilation | respiratory | RATE_C | |

| phenotype | male_beta | male_SE | male_OR | male_p | male_n_total | male_n_cases | male_n_controls | female_beta | female_SE | female_OR | female_p | female_n_total | female_n_cases | female_n_controls | interaction_p | interaction_beta | description | group | lab | label |
|---|---|---|---|---|---|---|---|---|---|---|---|---|---|---|---|---|---|---|---|---|
| 513.31 | | | | | 25126 | 49 | 25077 | | | | | 34541 | 40 | 34501 | | | Apnea | respiratory | RATE_C | |
| 513.32 | | | | | 25149 | 72 | 25077 | | | | | 34549 | 48 | 34501 | | | Orthopnea | respiratory | RATE_C | |
| 513.4 | | | | | 25445 | 368 | 25077 | | | | | 34837 | 336 | 34501 | 0.002272 | 0.6753 | Hyperventilation | respiratory | RATE_C | |
| 513.8 | | | | | 25261 | 184 | 25077 | | | | | 34580 | 79 | 34501 | | | Disorders of diaphragm | respiratory | RATE_C | |
| 514 | 1.252 | 0.06274 | 3.496 | 1.48e-88 | 24679 | 2921 | 21758 | 1.32 | 0.0593 | 3.742 | 1.03e-109 | 33770 | 2584 | 31186 | 0.02774 | 0.1818 | Abnormal findings examination of lungs | respiratory | RATE_C | |
| 514.1 | | | | | 21848 | 90 | 21758 | | | | | 31294 | 108 | 31186 | | | Abnormal results of function study of pulmonary system | respiratory | RATE_C | |
| 514.2 | 0.8418 | 0.09588 | 2.321 | 1.63e-18 | 22786 | 1028 | 21758 | 1.005 | 0.08809 | 2.732 | 3.72e-30 | 32192 | 1006 | 31186 | 0.1106 | 0.1989 | Solitary pulmonary nodule | respiratory | RATE_C | |
| 516 | 1.613 | 0.158 | 5.017 | 1.82e-24 | 27219 | 552 | 26667 | | | | | 36488 | 334 | 36154 | 0.308 | 0.2374 | Abnormal sputum | respiratory | RATE_C | |
| 516.1 | | | | | 27145 | 478 | 26667 | | | | | 36446 | 292 | 36154 | 0.7037 | 0.09801 | Hemoptysis | respiratory | RATE_C | |
| 519 | 1.401 | 0.09293 | 4.057 | 2.55e-51 | 25596 | 1340 | 24256 | 1.441 | 0.08307 | 4.227 | 1.93e-67 | 34332 | 1351 | 32981 | 0.07406 | 0.2124 | Other diseases of respiratory system, not elsewhere classified | respiratory | RATE_C | |
| 519.1 | | | | | 24327 | 71 | 24256 | | | | | 33033 | 52 | 32981 | | | Tracheostomy complications | respiratory | RATE_C | |
| 519.2 | | | | | 24383 | 127 | 24256 | | | | | 33036 | 55 | 32981 | | | Respiratory complications | respiratory | RATE_C | |
| 519.8 | | | | | 24432 | 176 | 24256 | | | | | 33153 | 172 | 32981 | | | Other diseases of respiratory system, NEC | respiratory | RATE_C | |
| 519.9 | 1.539 | 0.1189 | 4.661 | 2.44e-38 | 25196 | 940 | 24256 | 1.403 | 0.09482 | 4.066 | 1.62e-49 | 34006 | 1025 | 32981 | 0.9844 | -0.002868 | Symptoms involving respiratory system and other chest symptoms | respiratory | RATE_C | |
| 520 | | | | | 25518 | 159 | 25359 | | | | | 33516 | 257 | 33259 | | | Disorders of tooth development | digestive | RATE_C | |
| 520.1 | | | | | 25370 | 11 | 25359 | | | | | 33270 | 11 | 33259 | | | Hereditary disturbances in tooth structure | digestive | RATE_C | |
| 520.2 | | | | | 25503 | 144 | 25359 | | | | | 33503 | 244 | 33259 | | | Disturbances in tooth eruption | digestive | RATE_C | |
| 521 | 0.7607 | 0.1036 | 2.14 | 2.15e-13 | 26066 | 707 | 25359 | 0.8715 | 0.08433 | 2.391 | 4.90e-25 | 34146 | 887 | 33259 | 0.1494 | 0.1749 | Diseases of hard tissues of teeth | digestive | RATE_C | |
| 521.1 | 0.7666 | 0.1048 | 2.152 | 2.59e-13 | 26050 | 691 | 25359 | 0.8664 | 0.08534 | 2.378 | 3.21e-24 | 34122 | 863 | 33259 | 0.1677 | 0.1691 | Dental caries | digestive | RATE_C | |
| 521.2 | | | | | 25360 | 1 | 25359 | | | | | 33261 | 2 | 33259 | | | Dental abrasion, erosion and attrition | digestive | RATE_C | |
| 521.4 | | | | | 25359 | | | | | | | 33267 | 8 | 33259 | | | Tooth complications likely association with other diseases | digestive | RATE_C | |
| 522 | | | | | 25451 | 92 | 25359 | | | | | 33377 | 118 | 33259 | | | Diseases of pulp and periapical tissues | digestive | RATE_C | |
| 522.1 | | | | | 25361 | 2 | 25359 | | | | | 33263 | 4 | 33259 | | | Pulpitis and necrosis of tooth pulp | digestive | RATE_C | |
| 522.5 | | | | | 25445 | 86 | 25359 | | | | | 33365 | 106 | 33259 | | | Periapical abscess | digestive | RATE_C | |
| 523 | | | | | 25698 | 339 | 25359 | 0.3993 | 0.09901 | 1.491 | 5.50e-05 | 33819 | 560 | 33259 | 0.833 | -0.03306 | Gingival and periodontal diseases | digestive | RATE_C | |
| 523.1 | | | | | 25536 | 177 | 25359 | | | | | 33688 | 429 | 33259 | 0.186 | 0.2524 | Gingivitis | digestive | RATE_C | |
| 523.3 | | | | | 25492 | 133 | 25359 | | | | | 33407 | 148 | 33259 | | | Periodontitis (acute or chronic) | digestive | RATE_C | |
| 523.31 | | | | | 25454 | 95 | 25359 | | | | | 33368 | 109 | 33259 | | | Acute periodontitis | digestive | RATE_C | |
| 523.32 | | | | | 25382 | 23 | 25359 | | | | | 33285 | 26 | 33259 | | | Chronic periodontitis | digestive | RATE_C | |

| phenotype | male_beta | male_SE | male_OR | male_p | male_n_total | male_n_cases | male_n_controls | female_beta | female_SE | female_OR | female_p | female_n_total | female_n_cases | female_n_controls | interaction_p | interaction_beta | description | group | lab | label |
|---|---|---|---|---|---|---|---|---|---|---|---|---|---|---|---|---|---|---|---|---|
| 524 | | | | | 25495 | 136 | 25359 | | | | | 33518 | 259 | 33259 | | | Dentofacial anomalies, including malocclusion | digestive | RATE_C | |
| 524.3 | | | | | 25481 | 122 | 25359 | | | | | 33493 | 234 | 33259 | | | Anomalies of tooth position/malocclusion | digestive | RATE_C | |
| 525 | | | | | 25595 | 236 | 25359 | | | | | 33641 | 382 | 33259 | 0.03405 | 0.443 | Other diseases of the teeth and supporting structures | digestive | RATE_C | |
| 525.1 | | | | | 25495 | 136 | 25359 | | | | | 33466 | 207 | 33259 | | | Loss of teeth or edentulism | digestive | RATE_C | |
| 525.2 | | | | | 25366 | 7 | 25359 | | | | | 33279 | 20 | 33259 | | | Atrophy of edentulous alveolar ridge | digestive | RATE_C | |
| 526 | | | | | 25636 | 277 | 25359 | 0.5658 | 0.09964 | 1.761 | 1.36e-08 | 33838 | 579 | 33259 | 0.7319 | -0.05947 | Diseases of the jaws | digestive | RATE_C | |
| 526.1 | | | | | 25372 | 13 | 25359 | | | | | 33270 | 11 | 33259 | | | Cysts of the jaws | digestive | RATE_C | |
| 526.3 | | | | | 25393 | 34 | 25359 | | | | | 33336 | 77 | 33259 | | | Anomalies of jaw size/symmetry | digestive | RATE_C | |
| 526.4 | | | | | 25426 | 67 | 25359 | | | | | 33523 | 264 | 33259 | | | Temporomandibular joint disorders | digestive | RATE_C | |
| 526.41 | | | | | 25407 | 48 | 25359 | | | | | 33417 | 158 | 33259 | | | Temporomandibular joint disorder, unspecified | digestive | RATE_C | |
| 526.42 | | | | | 25364 | 5 | 25359 | | | | | 33300 | 41 | 33259 | | | Arthralgia/ankylosis of temporomandibular joint | digestive | RATE_C | |
| 526.5 | | | | | 25416 | 57 | 25359 | | | | | 33320 | 61 | 33259 | | | Inflammatory conditions of jaw | digestive | RATE_C | |
| 526.8 | | | | | 25363 | 4 | 25359 | | | | | 33267 | 8 | 33259 | | | Exostosis of jaw | digestive | RATE_C | |
| 526.9 | | | | | 25414 | 55 | 25359 | | | | | 33304 | 45 | 33259 | | | Jaw disease NOS | digestive | RATE_C | |
| 527 | | | | | 25962 | 420 | 25542 | | | | | 34324 | 353 | 33971 | 0.0112 | 0.433 | Diseases of the salivary glands | digestive | RATE_C | |
| 527.1 | | | | | 25545 | 3 | 25542 | | | | | 33976 | 5 | 33971 | | | Hypertrophy of salivary gland | digestive | RATE_C | |
| 527.2 | | | | | 25596 | 54 | 25542 | | | | | 34069 | 98 | 33971 | | | Sialoadenitis | digestive | RATE_C | |
| 527.7 | | | | | 25871 | 329 | 25542 | | | | | 34176 | 205 | 33971 | 0.1224 | 0.3132 | Disturbance of salivary secretion | digestive | RATE_C | |
| 527.8 | | | | | 25561 | 19 | 25542 | | | | | 34003 | 32 | 33971 | | | Other specified diseases of the salivary glands | digestive | RATE_C | |
| 528 | 0.4947 | 0.09535 | 1.64 | 2.13e-07 | 26317 | 775 | 25542 | 0.9344 | 0.08865 | 2.546 | 5.65e-26 | 34849 | 878 | 33971 | 2.73e-06 | 0.5653 | Diseases of the oral soft tissues, excluding lesions specific for gingiva and tongue | digestive | RATE_C | |
| 528.1 | | | | | 25938 | 396 | 25542 | | | | | 34341 | 370 | 33971 | 1.45e-04 | 0.6829 | Stomatitis and mucositis | digestive | RATE_C | |
| 528.11 | | | | | 25631 | 89 | 25542 | | | | | 34056 | 85 | 33971 | | | Stomatitis and mucositis (ulcerative) | digestive | RATE_C | |
| 528.12 | | | | | 25583 | 41 | 25542 | | | | | 34026 | 55 | 33971 | | | Oral aphthae | digestive | RATE_C | |
| 528.3 | | | | | 25579 | 37 | 25542 | | | | | 34020 | 49 | 33971 | | | Cellulitis and abscess of oral soft tissues | digestive | RATE_C | |
| 528.4 | | | | | 25551 | 9 | 25542 | | | | | 33984 | 13 | 33971 | | | Cysts of oral soft tissues | digestive | RATE_C | |
| 528.41 | | | | | 25551 | 9 | 25542 | | | | | 33982 | 11 | 33971 | | | Cyst of the salivary gland | digestive | RATE_C | |
| 528.5 | | | | | 25587 | 45 | 25542 | | | | | 34050 | 79 | 33971 | | | Diseases of lips | digestive | RATE_C | |
| 528.6 | | | | | 25578 | 36 | 25542 | | | | | 34015 | 44 | 33971 | | | Leukoplakia of oral mucosa | digestive | RATE_C | |
| 528.7 | | | | | 25551 | 9 | 25542 | | | | | 33984 | 13 | 33971 | | | Sialolithiasis | digestive | RATE_C | |

| phenotype | male_beta | male_SE | male_OR | male_p | male_n_total | male_n_cases | male_n_controls | female_beta | female_SE | female_OR | female_p | female_n_total | female_n_cases | female_n_controls | interaction_p | interaction_beta | description | group | lab | label |
|---|---|---|---|---|---|---|---|---|---|---|---|---|---|---|---|---|---|---|---|---|
| 529 | | | | | 25647 | 105 | 25542 | | | | | 34108 | 137 | 33971 | | | Diseases and other conditions of the tongue | digestive | RATE_C | |
| 529.1 | | | | | 25565 | 23 | 25542 | | | | | 34012 | 41 | 33971 | | | Glossitis | digestive | RATE_C | |

| phenotype | male_beta | male_SE | male_OR | male_p | male_n_total | male_n_cases | male_n_controls | female_beta | female_SE | female_OR | female_p | female_n_total | female_n_cases | female_n_controls | interaction_p | interaction_beta | description | group | lab | label |
|---|---|---|---|---|---|---|---|---|---|---|---|---|---|---|---|---|---|---|---|---|
| 529.6 | | | | | 25565 | 23 | 25542 | | | | | 34004 | 33 | 33971 | | | Glossodynia | digestive | RATE_C | |
| 530 | 0.8889 | 0.03882 | 2.433 | 4.53e-116 | 22713 | 7010 | 15703 | 1.049 | 0.03154 | 2.856 | 1.25e-242 | 30970 | 8921 | 22049 | 7.09e-12 | 0.3227 | Diseases of esophagus | digestive | RATE_C | |
| 530.1 | 0.8789 | 0.04059 | 2.408 | 5.47e-104 | 22053 | 6350 | 15703 | 1.051 | 0.03241 | 2.859 | 1.61e-230 | 30445 | 8396 | 22049 | 5.93e-12 | 0.3356 | Esophagitis, GERD and related diseases | digestive | RATE_C | |
| 530.11 | 0.9033 | 0.04238 | 2.468 | 8.07e-101 | 21565 | 5862 | 15703 | 1.072 | 0.0332 | 2.921 | 8.40e-229 | 30082 | 8033 | 22049 | 4.80e-10 | 0.3156 | GERD | digestive | RATE_C | GERD |
| 530.12 | | | | | 15778 | 75 | 15703 | | | | | 22113 | 64 | 22049 | | | Ulcer of esophagus | digestive | RATE_C | |
| 530.13 | | | | | 16021 | 318 | 15703 | | | | | 22193 | 144 | 22049 | | | Barrett's esophagus | digestive | RATE_C | |
| 530.14 | | | | | 16000 | 297 | 15703 | | | | | 22411 | 362 | 22049 | 9.30e-07 | 0.8658 | Reflux esophagitis | digestive | RATE_C | |
| 530.15 | | | | | 15777 | 74 | 15703 | | | | | 22097 | 48 | 22049 | | | Eosinophilic esophagitis | digestive | RATE_C | |
| 530.2 | 1.226 | 0.1185 | 3.408 | 4.44e-25 | 16327 | 624 | 15703 | | | | | 22465 | 416 | 22049 | 0.006686 | 0.4674 | Esophageal bleeding (varices/hemorrhage) | digestive | RATE_C | |
| 530.3 | | | | | 16140 | 437 | 15703 | | | | | 22429 | 380 | 22049 | 0.003346 | 0.5665 | Stricture and stenosis of esophagus | digestive | RATE_C | |
| 530.5 | | | | | 15823 | 120 | 15703 | | | | | 22257 | 208 | 22049 | | | Disorders of esophageal motility | digestive | RATE_C | |
| 530.6 | | | | | 15740 | 37 | 15703 | | | | | 22084 | 35 | 22049 | | | Diverticulum of esophagus, acquired | digestive | RATE_C | |
| 530.7 | | | | | 15735 | 32 | 15703 | | | | | 22067 | 18 | 22049 | | | Gastroesophageal laceration-hemorrhage syndrome | digestive | RATE_C | |
| 530.9 | | | | | 15920 | 217 | 15703 | | | | | 22322 | 273 | 22049 | | | Heartburn | digestive | RATE_C | |
| 531 | 1.093 | 0.1223 | 2.983 | 4.11e-19 | 26644 | 757 | 25887 | 1.398 | 0.1134 | 4.046 | 6.68e-35 | 35675 | 773 | 34902 | 0.02856 | 0.3493 | Peptic ulcer (excl. esophageal) | digestive | RATE_C | |
| 531.1 | | | | | 26046 | 159 | 25887 | | | | | 35004 | 102 | 34902 | | | Hemorrhage from gastrointestinal ulcer | digestive | RATE_C | |
| 531.2 | | | | | 26063 | 176 | 25887 | | | | | 35135 | 233 | 34902 | | | Gastric ulcer | digestive | RATE_C | |
| 531.3 | | | | | 26049 | 162 | 25887 | | | | | 35017 | 115 | 34902 | | | Duodenal ulcer | digestive | RATE_C | |
| 531.4 | | | | | 26128 | 241 | 25887 | | | | | 35173 | 271 | 34902 | 0.9352 | 0.03038 | Peptic ulcer, site unspecified | digestive | RATE_C | |
| 531.5 | | | | | 25904 | 17 | 25887 | | | | | 34952 | 50 | 34902 | | | Gastrojejunal ulcer | digestive | RATE_C | |
| 532 | 1.05 | 0.05691 | 2.859 | 4.56e-76 | 18697 | 2994 | 15703 | 1.253 | 0.05151 | 3.501 | 1.01e-130 | 25127 | 3078 | 22049 | 3.01e-06 | 0.344 | Dysphagia | digestive | RATE_C | |
| 535 | 0.2137 | 0.09247 | 1.238 | 0.02083 | 24211 | 698 | 23513 | 0.4848 | 0.07804 | 1.624 | 5.21e-10 | 32091 | 888 | 31203 | 2.57e-05 | 0.4535 | Gastritis and duodenitis | digestive | RATE_C | |
| 535.1 | | | | | 23554 | 41 | 23513 | | | | | 31256 | 53 | 31203 | | | Acute gastritis | digestive | RATE_C | |
| 535.2 | | | | | 23753 | 240 | 23513 | | | | | 31519 | 316 | 31203 | 2.72e-04 | 0.6321 | Atrophic gastritis | digestive | RATE_C | |
| 535.6 | | | | | 23634 | 121 | 23513 | | | | | 31294 | 91 | 31203 | | | Duodenitis | digestive | RATE_C | |
| 535.8 | | | | | 23673 | 160 | 23513 | | | | | 31440 | 237 | 31203 | | | Other specified gastritis | digestive | RATE_C | |

| phenotype | male_beta | male_SE | male_OR | male_p | male_n_total | male_n_cases | male_n_controls | female_beta | female_SE | female_OR | female_p | female_n_total | female_n_cases | female_n_controls | interaction_p | interaction_beta | description | group | lab | label |
|---|---|---|---|---|---|---|---|---|---|---|---|---|---|---|---|---|---|---|---|---|
| 535.9 | | | | | 23653 | 140 | 23513 | | | | | 31383 | 180 | 31203 | | | Gastritis and duodenitis, NOS | digestive | RATE_C | |
| 536 | 1.081 | 0.1183 | 2.949 | 6.36e-20 | 24111 | 598 | 23513 | 1.144 | 0.08092 | 3.141 | 2.07e-45 | 32274 | 1071 | 31203 | 0.06027 | 0.2489 | Disorders of function of stomach | digestive | RATE_C | |
| 536.3 | | | | | 23695 | 182 | 23513 | | | | | 31598 | 395 | 31203 | 0.7071 | 0.09314 | Gastroparesis | digestive | RATE_C | |
| 536.7 | | | | | 23722 | 209 | 23513 | | | | | 31426 | 223 | 31203 | | | Complications of gastrostomy, colostomy and enterostomy | digestive | RATE_C | |
| 536.8 | | | | | 23738 | 225 | 23513 | 0.8209 | 0.112 | 2.272 | 2.33e-13 | 31704 | 501 | 31203 | 0.2074 | 0.2475 | Dyspepsia and other specified disorders of function of stomach | digestive | RATE_C | |
| 537 | 0.9374 | 0.1134 | 2.553 | 1.37e-16 | 24195 | 682 | 23513 | 1.196 | 0.1023 | 3.306 | 1.44e-31 | 31926 | 723 | 31203 | 0.01499 | 0.3487 | Other disorders of stomach and duodenum | digestive | RATE_C | |
| 537.1 | | | | | 23574 | 61 | 23513 | | | | | 31261 | 58 | 31203 | | | Lesions of stomach and duodenum | digestive | RATE_C | |
| 539 | | | | | 27703 | 290 | 27413 | 1.169 | 0.08651 | 3.22 | 1.24e-41 | 36747 | 945 | 35802 | 0.8799 | 0.02786 | Bariatric surgery | digestive | RATE_C | |
| 540 | | | | | 27635 | 152 | 27483 | | | | | 36729 | 193 | 36536 | | | Appendiceal conditions | digestive | RATE_C | |
| 540.1 | | | | | 27611 | 128 | 27483 | | | | | 36714 | 178 | 36536 | | | Appendicitis | digestive | RATE_C | |
| 540.11 | | | | | 27592 | 109 | 27483 | | | | | 36692 | 156 | 36536 | | | Acute appendicitis | digestive | RATE_C | |
| 550 | 0.8286 | 0.05965 | 2.29 | 7.20e-44 | 25722 | 2883 | 22839 | 1.14 | 0.05912 | 3.125 | 8.81e-83 | 34582 | 2531 | 32051 | 3.02e-06 | 0.3742 | Abdominal hernia | digestive | RATE_C | |
| 550.1 | 0.4611 | 0.09386 | 1.586 | 9.01e-07 | 23813 | 974 | 22839 | | | | | 32167 | 116 | 32051 | 0.3867 | 0.2132 | Inguinal hernia | digestive | RATE_C | |
| 550.2 | 0.8617 | 0.1048 | 2.367 | 1.96e-16 | 23793 | 954 | 22839 | 1.181 | 0.0827 | 3.256 | 3.12e-46 | 33474 | 1423 | 32051 | 0.001222 | 0.4161 | Diaphragmatic hernia | digestive | RATE_C | |
| 550.3 | | | | | 22846 | 7 | 22839 | | | | | 32067 | 16 | 32051 | | | Femoral hernia | digestive | RATE_C | |
| 550.4 | | | | | 23250 | 411 | 22839 | | | | | 32268 | 217 | 32051 | 0.7342 | -0.07265 | Umbilical hernia | digestive | RATE_C | |
| 550.5 | | | | | 23322 | 483 | 22839 | 1.372 | 0.1282 | 3.942 | 1.06e-26 | 32584 | 533 | 32051 | 0.4495 | 0.1445 | Ventral hernia | digestive | RATE_C | |
| 550.6 | 1.377 | 0.1536 | 3.963 | 3.07e-19 | 23364 | 525 | 22839 | 1.51 | 0.1328 | 4.528 | 5.61e-30 | 32610 | 559 | 32051 | 0.2559 | 0.2211 | Incisional hernia | digestive | RATE_C | |
| 555 | -0.3792 | 0.0719 | 0.6844 | 1.33e-07 | 18550 | 1092 | 17458 | -0.0968 | 0.05962 | 0.9077 | 0.1044 | 24902 | 1438 | 23464 | 2.22e-08 | 0.4722 | Inflammatory bowel disease and other gastroenteritis and colitis | digestive | RATE_C | |
| 555.1 | -0.4867 | 0.08046 | 0.6147 | 1.46e-09 | 18342 | 884 | 17458 | -0.1558 | 0.06535 | 0.8557 | 0.01713 | 24671 | 1207 | 23464 | 2.28e-08 | 0.5242 | Regional enteritis | digestive | RATE_C | |
| 555.2 | -0.1595 | 0.09957 | 0.8526 | 0.1092 | 18001 | 543 | 17458 | 0.1224 | 0.08668 | 1.13 | 0.158 | 24116 | 652 | 23464 | 2.57e-04 | 0.4358 | Ulcerative colitis | digestive | RATE_C | |
| 555.21 | | | | | 17689 | 231 | 17458 | | | | | 23710 | 246 | 23464 | | | Ulcerative colitis (chronic) | digestive | RATE_C | |
| 556 | | | | | 17670 | 212 | 17458 | | | | | 23667 | 203 | 23464 | | | Ulceration of the lower GI tract | digestive | RATE_C | |
| 556.1 | | | | | 17633 | 175 | 17458 | | | | | 23632 | 168 | 23464 | | | Ulceration of intestine | digestive | RATE_C | |
| 556.11 | | | | | 17498 | 40 | 17458 | | | | | 23504 | 40 | 23464 | | | Angiodysplasia of intestine (without mention of hemorrhage) | digestive | RATE_C | |
| 557 | | | | | 17645 | 187 | 17458 | | | | | 23820 | 356 | 23464 | 0.06264 | 0.3874 | Intestinal malabsorption (non-celiac) | digestive | RATE_C | |
| 557.1 | | | | | 17538 | 80 | 17458 | | | | | 23734 | 270 | 23464 | | | Celiac disease | digestive | RATE_C | |
| 558 | 0.1278 | 0.07562 | 1.136 | 0.09108 | 18499 | 1041 | 17458 | 0.4658 | 0.0591 | 1.593 | 3.26e-15 | 25010 | 1546 | 23464 | 1.31e-10 | 0.5553 | Noninfectious gastroenteritis | digestive | RATE_C | |
| 559 | | | | | 17873 | 415 | 17458 | | | | | 23811 | 347 | 23464 | 0.04682 | 0.3497 | Ileostomy status | digestive | RATE_C | |

| phenotype | male_beta | male_SE | male_OR | male_p | male_n_total | male_n_cases | male_n_controls | female_beta | female_SE | female_OR | female_p | female_n_total | female_n_cases | female_n_controls | interaction_p | interaction_beta | description | group | lab | label |
|---|---|---|---|---|---|---|---|---|---|---|---|---|---|---|---|---|---|---|---|---|
| 560 | 1.049 | 0.07959 | 2.854 | 1.17e-39 | 18805 | 1347 | 17458 | 1.185 | 0.07548 | 3.269 | 1.68e-55 | 24662 | 1198 | 23464 | 0.01492 | 0.2513 | Intestinal obstruction without mention of hernia | digestive | RATE_C | |
| 560.1 | 1.391 | 0.1251 | 4.017 | 1.09e-28 | 18153 | 695 | 17458 | | | | | 23849 | 385 | 23464 | 0.08345 | 0.3291 | Paralytic ileus | digestive | RATE_C | |
| 560.2 | | | | | 17514 | 56 | 17458 | | | | | 23568 | 104 | 23464 | | | Impaction of intestine | digestive | RATE_C | |
| 560.3 | | | | | 17538 | 80 | 17458 | | | | | 23554 | 90 | 23464 | | | Peritoneal or intestinal adhesions | digestive | RATE_C | |
| 560.4 | 0.9414 | 0.1043 | 2.564 | 1.76e-19 | 18149 | 691 | 17458 | 1.127 | 0.09173 | 3.085 | 1.14e-34 | 24229 | 765 | 23464 | 0.004082 | 0.3725 | Other intestinal obstruction | digestive | RATE_C | |
| phenotype | male_beta | male_SE | male_OR | male_p | male_n_total | male_n_cases | male_n_controls | female_beta | female_SE | female_OR | female_p | female_n_total | female_n_cases | female_n_controls | interaction_p | interaction_beta | description | group | lab | label |
| 561 | 0.4894 | 0.06522 | 1.631 | 6.18e-14 | 19227 | 1769 | 17458 | 0.6627 | 0.04697 | 1.94 | 3.37e-45 | 26443 | 2979 | 23464 | 6.94e-07 | 0.3651 | Symptoms involving digestive system | digestive | RATE_C | |
| 562 | 0.7838 | 0.07059 | 2.19 | 1.20e-28 | 19778 | 2320 | 17458 | 0.8917 | 0.06367 | 2.439 | 1.47e-44 | 25685 | 2221 | 23464 | 0.01541 | 0.2246 | Diverticulosis and diverticulitis | digestive | RATE_C | |
| 562.1 | 0.8099 | 0.07497 | 2.248 | 3.29e-27 | 19570 | 2112 | 17458 | 0.9408 | 0.06931 | 2.562 | 5.72e-42 | 25403 | 1939 | 23464 | 0.01274 | 0.2484 | Diverticulosis | digestive | RATE_C | |
| 562.2 | | | | | 17837 | 379 | 17458 | 0.9341 | 0.1268 | 2.545 | 1.77e-13 | 23966 | 502 | 23464 | 0.0111 | 0.4854 | Diverticulitis | digestive | RATE_C | |
| 564 | 0.2541 | 0.06885 | 1.289 | 2.24e-04 | 18857 | 1399 | 17458 | 0.7546 | 0.05033 | 2.127 | 8.24e-51 | 26038 | 2574 | 23464 | 4.74e-16 | 0.6344 | Functional digestive disorders | digestive | RATE_C | |
| 564.1 | | | | | 17914 | 456 | 17458 | 0.7231 | 0.06257 | 2.061 | 6.84e-31 | 25043 | 1579 | 23464 | 7.83e-13 | 0.8378 | Irritable Bowel Syndrome | digestive | RATE_C | |
| 564.9 | | | | | 17664 | 206 | 17458 | | | | | 23673 | 209 | 23464 | | | Personal history of diseases of digestive system | digestive | RATE_C | |
| 565 | 0.07879 | 0.07375 | 1.082 | 0.2854 | 23018 | 1093 | 21925 | 0.4789 | 0.07062 | 1.614 | 1.19e-11 | 31069 | 1106 | 29963 | 5.13e-10 | 0.5717 | Anal and rectal conditions | digestive | RATE_C | |
| 565.1 | | | | | 22052 | 127 | 21925 | | | | | 30077 | 114 | 29963 | | | Anal and rectal polyp | digestive | RATE_C | |
| 567 | 1.218 | 0.1154 | 3.382 | 4.48e-26 | 22577 | 652 | 21925 | 1.302 | 0.1103 | 3.675 | 3.90e-32 | 30541 | 578 | 29963 | 0.1214 | 0.2307 | Peritonitis and retroperitoneal infections | digestive | RATE_C | |
| 568 | | | | | 22412 | 487 | 21925 | 1.685 | 0.1311 | 5.394 | 8.28e-38 | 30476 | 513 | 29963 | 0.3578 | 0.1765 | Other disorders of peritoneum | digestive | RATE_C | |
| 568.1 | | | | | 22164 | 239 | 21925 | | | | | 30277 | 314 | 29963 | 0.3151 | 0.2623 | Peritoneal adhesions (postoperative) (postinfection) | digestive | RATE_C | |
| 569 | 0.7685 | 0.07076 | 2.157 | 1.78e-27 | 23419 | 1494 | 21925 | 0.9982 | 0.05742 | 2.713 | 1.11e-67 | 31975 | 2012 | 29963 | 2.29e-06 | 0.3935 | Other disorders of intestine | digestive | RATE_C | |
| 569.1 | | | | | 21954 | 29 | 21925 | | | | | 30021 | 58 | 29963 | | | Toxic gastroenteritis and colitis | digestive | RATE_C | |
| 569.2 | 1.059 | 0.1205 | 2.883 | 1.51e-18 | 22496 | 571 | 21925 | 1.218 | 0.08376 | 3.381 | 6.44e-48 | 30942 | 979 | 29963 | 0.08532 | 0.2334 | Gastrointestinal complications | digestive | RATE_C | |
| 571 | 0.7797 | 0.05743 | 2.181 | 5.56e-42 | 21552 | 2387 | 19165 | 0.9675 | 0.05515 | 2.631 | 6.68e-69 | 29934 | 2235 | 27699 | 3.97e-07 | 0.3739 | Chronic liver disease and cirrhosis | digestive | RATE_C | |
| 571.5 | 0.788 | 0.05961 | 2.199 | 6.81e-40 | 21382 | 2217 | 19165 | 1.008 | 0.05803 | 2.739 | 1.46e-67 | 29754 | 2055 | 27699 | 1.09e-07 | 0.4093 | Other chronic nonalcoholic liver disease | digestive | RATE_C | |
| 571.51 | 0.825 | 0.07679 | 2.282 | 6.35e-27 | 20477 | 1312 | 19165 | 0.9751 | 0.08328 | 2.651 | 1.16e-31 | 28658 | 959 | 27699 | 0.00126 | 0.3418 | Cirrhosis of liver without mention of alcohol | digestive | RATE_C | |
| 571.6 | | | | | 19242 | 77 | 19165 | | | | | 27883 | 184 | 27699 | | | Primary biliary cirrhosis | digestive | RATE_C | |
| 571.8 | 1.328 | 0.09918 | 3.775 | 6.54e-41 | 20161 | 996 | 19165 | 1.629 | 0.1097 | 5.099 | 7.25e-50 | 28437 | 738 | 27699 | 0.001128 | 0.4549 | Liver abscess and sequelae of chronic liver disease | digestive | RATE_C | |
| 571.81 | 1.226 | 0.1154 | 3.407 | 2.44e-26 | 19856 | 691 | 19165 | | | | | 28168 | 469 | 27699 | 0.006593 | 0.4512 | Portal hypertension | digestive | RATE_C | |
| 572 | 1.7 | 0.09082 | 5.477 | 3.15e-78 | 20559 | 1394 | 19165 | 1.716 | 0.08397 | 5.564 | 7.42e-93 | 28923 | 1224 | 27699 | 0.1884 | 0.1536 | Ascites (non malignant) | digestive | RATE_C | |
| 573 | 1.499 | 0.1086 | 4.479 | 2.44e-43 | 20126 | 961 | 19165 | 1.464 | 0.09216 | 4.323 | 7.91e-57 | 28655 | 956 | 27699 | 0.6936 | 0.05301 | Other disorders of liver | digestive | RATE_C | |

| phenotype | male_beta | male_SE | male_OR | male_p | male_n_total | male_n_cases | male_n_controls | female_beta | female_SE | female_OR | female_p | female_n_total | female_n_cases | female_n_controls | interaction_p | interaction_beta | description | group | lab | label |
|---|---|---|---|---|---|---|---|---|---|---|---|---|---|---|---|---|---|---|---|---|
| 573.1 | | | | | 19195 | 30 | 19165 | | | | | 27731 | 32 | 27699 | | | Chronic passive congestion of liver | digestive | RATE_C | |
| 573.2 | | | | | 19596 | 431 | 19165 | | | | | 27945 | 246 | 27699 | 0.6121 | 0.1353 | Liver replaced by transplant | digestive | RATE_C | |
| 573.3 | | | | | 19292 | 127 | 19165 | | | | | 27860 | 161 | 27699 | | | Hepatomegaly | digestive | RATE_C | |
| 573.4 | | | | | 19356 | 191 | 19165 | | | | | 27881 | 182 | 27699 | | | Acute and subacute necrosis of liver | digestive | RATE_C | |
| 573.5 | | | | | 19606 | 441 | 19165 | | | | | 28006 | 307 | 27699 | 0.9143 | -0.02009 | Jaundice (not of newborn) | digestive | RATE_C | |
| 573.6 | 0.928 | 0.1099 | 2.529 | 2.98e-17 | 19797 | 632 | 19165 | 1.026 | 0.1052 | 2.791 | 1.72e-22 | 28268 | 569 | 27699 | 0.107 | 0.2233 | Nonspecific elevation of levels of transaminase or lactic acid dehydrogenase [LDH] | digestive | RATE_C | |
| 573.7 | 0.4798 | 0.08436 | 1.616 | 1.29e-08 | 20119 | 954 | 19165 | 0.7588 | 0.07471 | 2.136 | 3.09e-24 | 28828 | 1129 | 27699 | 1.89e-07 | 0.5348 | Abnormal results of function study of liver | digestive | RATE_C | |
| 573.9 | 0.681 | 0.09834 | 1.976 | 4.36e-12 | 19944 | 779 | 19165 | 0.8812 | 0.08636 | 2.414 | 1.91e-24 | 28587 | 888 | 27699 | 0.002395 | 0.3671 | Abnormal serum enzyme levels | digestive | RATE_C | |
| 574 | 0.9786 | 0.09004 | 2.661 | 1.63e-27 | 25697 | 1283 | 24414 | 1.085 | 0.07059 | 2.959 | 2.73e-53 | 34656 | 1503 | 33153 | 0.5413 | -0.06608 | Cholelithiasis and cholecystitis | digestive | RATE_C | |
| 574.1 | 0.9208 | 0.1013 | 2.511 | 9.93e-20 | 25404 | 990 | 24414 | 1.188 | 0.08148 | 3.281 | 3.71e-48 | 34320 | 1167 | 33153 | 0.7751 | 0.03514 | Cholelithiasis | digestive | RATE_C | |
| 574.11 | | | | | 24503 | 89 | 24414 | | | | | 33224 | 71 | 33153 | | | Cholelithiasis with acute cholecystitis | digestive | RATE_C | |
| 574.12 | | | | | 24528 | 114 | 24414 | | | | | 33340 | 187 | 33153 | | | Cholelithiasis with other cholecystitis | digestive | RATE_C | |
| 574.2 | | | | | 24625 | 211 | 24414 | | | | | 33383 | 230 | 33153 | | | Calculus of bile duct | digestive | RATE_C | |
| 574.3 | | | | | 24711 | 297 | 24414 | | | | | 33444 | 291 | 33153 | 0.05015 | -0.4689 | Cholecystitis without cholelithiasis | digestive | RATE_C | |
| 575 | 1.214 | 0.1057 | 3.368 | 1.48e-30 | 25285 | 871 | 24414 | 0.9592 | 0.08359 | 2.61 | 1.75e-30 | 34100 | 947 | 33153 | 0.2034 | -0.1606 | Other biliary tract disease | digestive | RATE_C | |
| 575.1 | | | | | 24612 | 198 | 24414 | | | | | 33277 | 124 | 33153 | | | Cholangitis | digestive | RATE_C | |
| 575.2 | | | | | 24699 | 285 | 24414 | | | | | 33392 | 239 | 33153 | 0.188 | -0.3153 | Obstruction of bile duct | digestive | RATE_C | |
| 575.6 | | | | | 24435 | 21 | 24414 | | | | | 33185 | 32 | 33153 | | | Cholesterolosis of gallbladder | digestive | RATE_C | |
| 575.7 | | | | | 24611 | 197 | 24414 | | | | | 33370 | 217 | 33153 | | | Other disorders of gallbladder | digestive | RATE_C | |
| 575.8 | | | | | 24783 | 369 | 24414 | | | | | 33584 | 431 | 33153 | 0.5 | -0.1295 | Other disorders of biliary tract | digestive | RATE_C | |
| 575.9 | | | | | 24488 | 74 | 24414 | | | | | 33244 | 91 | 33153 | | | Nonspecific abnormal findings on radiological and other examination of biliary tract | digestive | RATE_C | |
| 577 | 0.8649 | 0.09804 | 2.375 | 1.12e-18 | 27246 | 863 | 26383 | 0.9851 | 0.09126 | 2.678 | 3.63e-27 | 36384 | 840 | 35544 | 0.03869 | 0.2577 | Diseases of pancreas | digestive | RATE_C | |
| 577.1 | | | | | 26693 | 310 | 26383 | | | | | 35828 | 284 | 35544 | 0.97 | -0.009231 | Acute pancreatitis | digestive | RATE_C | |
| 577.2 | | | | | 26568 | 185 | 26383 | | | | | 35726 | 182 | 35544 | | | Chronic pancreatitis | digestive | RATE_C | |
| 577.3 | | | | | 26620 | 237 | 26383 | | | | | 35810 | 266 | 35544 | 0.4079 | 0.2001 | Cyst and pseudocyst of pancreas | digestive | RATE_C | |
| 578 | 0.7575 | 0.05929 | 2.133 | 2.20e-37 | 23398 | 2404 | 20994 | 0.9737 | 0.05266 | 2.648 | 2.42e-76 | 31110 | 2548 | 28562 | 5.08e-04 | 0.2564 | Gastrointestinal hemorrhage | digestive | RATE_C | |
| 578.1 | | | | | 21273 | 279 | 20994 | | | | | 28819 | 257 | 28562 | 0.01884 | 0.5898 | Hematemesis | digestive | RATE_C | |
| 578.2 | 0.9139 | 0.08923 | 2.494 | 1.28e-24 | 22097 | 1103 | 20994 | 1.026 | 0.07643 | 2.791 | 4.15e-41 | 29747 | 1185 | 28562 | 0.196 | 0.1419 | Blood in stool | digestive | RATE_C | |

| phenotype | male_beta | male_SE | male_OR | male_p | male_n_total | male_n_cases | male_n_controls | female_beta | female_SE | female_OR | female_p | female_n_total | female_n_cases | female_n_controls | interaction_p | interaction_beta | description | group | lab | label |
|---|---|---|---|---|---|---|---|---|---|---|---|---|---|---|---|---|---|---|---|---|
| 578.8 | 0.3271 | 0.1012 | 1.387 | 0.001227 | 21622 | 628 | 20994 | 0.6143 | 0.08892 | 1.848 | 4.88e-12 | 29288 | 726 | 28562 | 0.00483 | 0.347 | Hemorrhage of rectum and anus | digestive | RATE_C | |
| 578.9 | 1.393 | 0.1139 | 4.028 | 2.16e-34 | 22034 | 1040 | 20994 | 1.742 | 0.1218 | 5.706 | 2.25e-46 | 29380 | 818 | 28562 | 0.01112 | 0.411 | Hemorrhage of gastrointestinal tract | digestive | RATE_C | |
| 579 | 1.397 | 0.08295 | 4.043 | 1.22e-63 | 22498 | 1504 | 20994 | 1.383 | 0.06234 | 3.988 | 4.39e-109 | 30586 | 2024 | 28562 | 0.1983 | 0.1247 | Other symptoms involving abdomen and pelvis | digestive | RATE_C | |
| 579.2 | 1.369 | 0.1252 | 3.933 | 7.92e-28 | 21574 | 580 | 20994 | | | | | 28950 | 388 | 28562 | 0.7402 | -0.05507 | Splenomegaly | digestive | RATE_C | |
| 579.8 | | | | | 21190 | 196 | 20994 | | | | | 28810 | 248 | 28562 | | | Nonspecific abnormal findings in stool contents | digestive | RATE_C | |
| 580 | 1.278 | 0.1175 | 3.588 | 1.61e-27 | 18292 | 656 | 17636 | 1.251 | 0.1094 | 3.493 | 3.03e-30 | 28613 | 588 | 28025 | 0.8411 | -0.03028 | Nephritis; nephrosis; renal sclerosis | genitourinary | RATE_C | |
| 580.1 | | | | | 17811 | 175 | 17636 | | | | | 28185 | 160 | 28025 | | | Glomerulonephritis | genitourinary | RATE_C | |
| 580.11 | | | | | 17690 | 54 | 17636 | | | | | 28073 | 48 | 28025 | | | Proliferative glomerulonephritis | genitourinary | RATE_C | |
| 580.12 | | | | | 17715 | 79 | 17636 | | | | | 28084 | 59 | 28025 | | | Non-proliferative glomerulonephritis | genitourinary | RATE_C | |
| 580.13 | | | | | 17654 | 18 | 17636 | | | | | 28048 | 23 | 28025 | | | Acute glomerulonephritis, NOS | genitourinary | RATE_C | |
| 580.14 | | | | | 17676 | 40 | 17636 | | | | | 28076 | 51 | 28025 | | | Chronic glomerulonephritis, NOS | genitourinary | RATE_C | |
| 580.2 | | | | | 17776 | 140 | 17636 | | | | | 28143 | 118 | 28025 | | | Nephrotic syndrome without mention of glomerulonephritis | genitourinary | RATE_C | |
| 580.3 | | | | | 17973 | 337 | 17636 | | | | | 28368 | 343 | 28025 | 0.3471 | -0.1888 | Nephritis and nephropathy without mention of glomerulonephritis | genitourinary | RATE_C | |
| 580.31 | | | | | 17810 | 174 | 17636 | | | | | 28242 | 217 | 28025 | | | Nephritis and nephropathy in diseases classified elsewhere | genitourinary | RATE_C | |
| 580.32 | | | | | 17795 | 159 | 17636 | | | | | 28162 | 137 | 28025 | | | Nephritis and nephropathy with pathological lesion | genitourinary | RATE_C | |
| 580.4 | | | | | 17734 | 98 | 17636 | | | | | 28125 | 100 | 28025 | | | Renal sclerosis, NOS | genitourinary | RATE_C | |
| 585 | 1.542 | 0.04685 | 4.674 | 1.25e-237 | 24023 | 6387 | 17636 | 1.553 | 0.04653 | 4.724 | 4.11e-244 | 32893 | 4868 | 28025 | 0.623 | 0.0316 | Renal failure | genitourinary | RATE_C | |
| 585.1 | 1.899 | 0.05988 | 6.678 | 1.24e-220 | 22356 | 4720 | 17636 | 1.886 | 0.05975 | 6.593 | 1.14e-218 | 31559 | 3534 | 28025 | 0.8447 | 0.01619 | Acute renal failure | genitourinary | RATE_C | |
| 585.2 | 2.536 | 0.205 | 12.62 | 3.82e-35 | 18192 | 556 | 17636 | | | | | 28438 | 413 | 28025 | 0.6702 | -0.1179 | Renal failure NOS | genitourinary | RATE_C | |
| 585.3 | 1.534 | 0.06264 | 4.637 | 1.87e-132 | 21314 | 3678 | 17636 | 1.59 | 0.06646 | 4.902 | 1.92e-126 | 30569 | 2544 | 28025 | 0.4337 | 0.06999 | Chronic renal failure [CKD] | genitourinary | RATE_C | |
| 585.31 | 2.115 | 0.1529 | 8.287 | 1.73e-43 | 18326 | 690 | 17636 | | | | | 28496 | 471 | 28025 | 0.7768 | 0.06151 | Renal dialysis | genitourinary | RATE_C | |
| 585.32 | 2.088 | 0.1295 | 8.07 | 1.82e-58 | 18620 | 984 | 17636 | 2.189 | 0.1427 | 8.922 | 4.48e-53 | 28699 | 674 | 28025 | 0.4417 | 0.1443 | End stage renal disease | genitourinary | RATE_C | |
| 585.33 | 1.466 | 0.08735 | 4.332 | 3.23e-63 | 19559 | 1923 | 17636 | 1.397 | 0.08951 | 4.044 | 6.31e-55 | 29378 | 1353 | 28025 | 0.9447 | -0.008505 | Chronic Kidney Disease, Stage III | genitourinary | RATE_C | |
| 585.34 | 1.821 | 0.1383 | 6.177 | 1.34e-39 | 18575 | 939 | 17636 | 1.528 | 0.1246 | 4.609 | 1.50e-34 | 28729 | 704 | 28025 | 0.1624 | -0.2549 | Chronic Kidney Disease, Stage IV | genitourinary | RATE_C | |
| 585.4 | 1.041 | 0.1059 | 2.831 | 8.97e-23 | 18446 | 810 | 17636 | 1.032 | 0.1052 | 2.808 | 9.78e-23 | 28650 | 625 | 28025 | 0.7219 | -0.0504 | Chronic kidney disease, Stage I or II | genitourinary | RATE_C | |
| 586 | 1.344 | 0.05972 | 3.836 | 3.38e-112 | 21065 | 3429 | 17636 | 1.303 | 0.06022 | 3.682 | 6.84e-104 | 30360 | 2335 | 28025 | 0.1199 | -0.1273 | Other disorders of the kidney and ureters | genitourinary | RATE_C | |
| 586.1 | | | | | 17687 | 51 | 17636 | | | | | 28103 | 78 | 28025 | | | Anatomical abnormalities of kidney and ureters | genitourinary | RATE_C | |
| 586.11 | | | | | 17638 | 2 | 17636 | | | | | 28033 | 8 | 28025 | | | Small kidney | genitourinary | RATE_C | |

| phenotype | male_beta | male_SE | male_OR | male_p | male_n_total | male_n_cases | male_n_controls | female_beta | female_SE | female_OR | female_p | female_n_total | female_n_cases | female_n_controls | interaction_p | interaction_beta | description | group | lab | label |
|---|---|---|---|---|---|---|---|---|---|---|---|---|---|---|---|---|---|---|---|---|
| 586.12 | | | | | 17684 | 48 | 17636 | | | | | 28097 | 72 | 28025 | | | Vesicoureteral reflux | genitourinary | RATE_C | |
| 586.2 | 1.38 | 0.1272 | 3.977 | 1.94e-27 | 18502 | 866 | 17636 | 1.021 | 0.1223 | 2.776 | 6.80e-17 | 28545 | 520 | 28025 | 0.006521 | -0.4657 | Cyst of kidney, acquired | genitourinary | RATE_C | |
| 586.3 | | | | | 17685 | 49 | 17636 | | | | | 28057 | 32 | 28025 | | | Vascular disorders of kidney/hypertrophy | genitourinary | RATE_C | |
| 586.4 | 0.7016 | 0.1139 | 2.017 | 7.30e-10 | 18326 | 690 | 17636 | | | | | 28313 | 288 | 28025 | 0.715 | -0.0669 | Stricture/obstruction of ureter | genitourinary | RATE_C | |
| 587 | 2.254 | 0.167 | 9.522 | 1.76e-41 | 18201 | 565 | 17636 | | | | | 28416 | 391 | 28025 | 0.1613 | -0.3079 | Kidney replaced by transpant | genitourinary | RATE_C | |
| 588 | 1.646 | 0.1219 | 5.188 | 1.50e-41 | 18557 | 921 | 17636 | 1.481 | 0.1165 | 4.395 | 5.28e-37 | 28718 | 693 | 28025 | 0.4222 | -0.1307 | Disorders resulting from impaired renal function | genitourinary | RATE_C | |
| 588.1 | 1.795 | 0.1433 | 6.022 | 4.95e-36 | 18381 | 745 | 17636 | 1.633 | 0.1404 | 5.119 | 2.82e-31 | 28555 | 530 | 28025 | 0.525 | -0.1236 | Renal osteodystrophy | genitourinary | RATE_C | |
| 588.2 | | | | | 17925 | 289 | 17636 | | | | | 28246 | 221 | 28025 | 0.9052 | 0.03204 | Secondary hyperparathyroidism (of renal origin) | genitourinary | RATE_C | |
| 589 | | | | | 17733 | 97 | 17636 | | | | | 28140 | 115 | 28025 | | | Abnormal results of function study of kidney | genitourinary | RATE_C | |
| 590 | | | | | 21698 | 323 | 21375 | 1.321 | 0.09565 | 3.748 | 2.13e-43 | 24682 | 746 | 23936 | 0.2934 | -0.2033 | Pyelonephritis | genitourinary | RATE_C | |
| 591 | 1.177 | 0.07002 | 3.244 | 2.19e-63 | 23909 | 2534 | 21375 | 1.062 | 0.0356 | 2.892 | 1.84e-195 | 30676 | 6740 | 23936 | 0.0025 | -0.2293 | Urinary tract infection | genitourinary | RATE_C | |
| 592 | | | | | 21768 | 393 | 21375 | 0.8581 | 0.06842 | 2.359 | 4.47e-36 | 25433 | 1497 | 23936 | 0.3904 | -0.1459 | Cystitis and urethritis | genitourinary | RATE_C | |
| 592.1 | | | | | 21707 | 332 | 21375 | 0.8519 | 0.0699 | 2.344 | 3.68e-34 | 25375 | 1439 | 23936 | 0.02177 | -0.474 | Cystitis | genitourinary | RATE_C | |
| 592.11 | | | | | 21543 | 168 | 21375 | 0.7844 | 0.08761 | 2.191 | 3.45e-19 | 24834 | 898 | 23936 | 0.6474 | -0.1161 | Acute cystitis | genitourinary | RATE_C | |
| 592.12 | | | | | 21411 | 36 | 21375 | | | | | 23989 | 53 | 23936 | | | Chronic cystitis | genitourinary | RATE_C | |
| 592.13 | | | | | 21383 | 8 | 21375 | | | | | 24169 | 233 | 23936 | | | Chronic interstitial cystitis | genitourinary | RATE_C | |
| 592.2 | | | | | 21408 | 33 | 21375 | | | | | 23967 | 31 | 23936 | | | Urethritis and urethral syndrome | genitourinary | RATE_C | |
| 592.21 | | | | | 21378 | 3 | 21375 | | | | | 23938 | 2 | 23936 | | | Urethral syndrome | genitourinary | RATE_C | |
| 592.3 | | | | | 21378 | 3 | 21375 | | | | | 23938 | 2 | 23936 | | | Urethral stricture due to infecton | genitourinary | RATE_C | |
| 593 | 0.7243 | 0.0731 | 2.063 | 3.83e-23 | 23190 | 1815 | 21375 | 0.9683 | 0.0637 | 2.633 | 3.48e-52 | 25739 | 1803 | 23936 | 0.157 | 0.1313 | Hematuria | genitourinary | RATE_C | |
| 593.1 | 0.8473 | 0.1312 | 2.333 | 1.04e-10 | 22020 | 645 | 21375 | | | | | 24248 | 312 | 23936 | 0.5369 | 0.1225 | Gross hematuria | genitourinary | RATE_C | |
| 593.2 | | | | | 21568 | 193 | 21375 | | | | | 24188 | 252 | 23936 | | | Microscopic hematuria | genitourinary | RATE_C | |
| 594 | 0.6082 | 0.06812 | 1.837 | 4.33e-19 | 25965 | 1823 | 24142 | 0.849 | 0.06382 | 2.337 | 2.22e-40 | 35229 | 1634 | 33595 | 0.4699 | 0.0626 | Urinary calculus | genitourinary | RATE_C | |
| 594.1 | 0.68 | 0.07656 | 1.974 | 6.58e-19 | 25599 | 1457 | 24142 | 0.7964 | 0.06916 | 2.218 | 1.10e-30 | 34942 | 1347 | 33595 | 0.8013 | -0.02405 | Calculus of kidney | genitourinary | RATE_C | |
| 594.2 | | | | | 24276 | 134 | 24142 | | | | | 33649 | 54 | 33595 | | | Calculus of lower urinary tract | genitourinary | RATE_C | |
| 594.3 | | | | | 24598 | 456 | 24142 | | | | | 33995 | 400 | 33595 | 0.9786 | 0.004495 | Calculus of ureter | genitourinary | RATE_C | |
| 594.8 | | | | | 24179 | 37 | 24142 | | | | | 33649 | 54 | 33595 | | | Renal colic | genitourinary | RATE_C | |
| 595 | 0.6393 | 0.1011 | 1.895 | 2.60e-10 | 24902 | 760 | 24142 | 0.9589 | 0.09572 | 2.609 | 1.28e-23 | 34327 | 732 | 33595 | 0.0737 | 0.2318 | Hydronephrosis | genitourinary | RATE_C | |
| 596 | 0.6989 | 0.08956 | 2.011 | 6.04e-15 | 25451 | 1131 | 24320 | 0.5913 | 0.07315 | 1.806 | 6.29e-16 | 34046 | 1171 | 32875 | 0.07967 | -0.1908 | Other disorders of bladder | genitourinary | RATE_C | |

| phenotype | male_beta | male_SE | male_OR | male_p | male_n_total | male_n_cases | male_n_controls | female_beta | female_SE | female_OR | female_p | female_n_total | female_n_cases | female_n_controls | interaction_p | interaction_beta | description | group | lab | label |
|---|---|---|---|---|---|---|---|---|---|---|---|---|---|---|---|---|---|---|---|---|
| 596.1 | | | | | 24500 | 180 | 24320 | | | | | 32903 | 28 | 32875 | | | Bladder neck obstruction | genitourinary | RATE_C | |
| 596.5 | 0.6041 | 0.1122 | 1.83 | 7.29e-08 | 24918 | 598 | 24320 | 0.5289 | 0.08059 | 1.697 | 5.26e-11 | 33798 | 923 | 32875 | 0.659 | 0.05616 | Functional disorders of bladder | genitourinary | RATE_C | |
| 597 | | | | | 24709 | 389 | 24320 | | | | | 33264 | 389 | 32875 | 0.8559 | 0.03956 | Other disorders of urethra and urinary tract | genitourinary | RATE_C | |
| 597.1 | | | | | 24499 | 179 | 24320 | | | | | 32898 | 23 | 32875 | | | Urethral stricture (not specified as infectious) | genitourinary | RATE_C | |
| 597.2 | | | | | 24431 | 111 | 24320 | | | | | 32951 | 76 | 32875 | | | Urinary complications NEC | genitourinary | RATE_C | |
| 597.8 | | | | | 24446 | 126 | 24320 | | | | | 32996 | 121 | 32875 | | | Urethral hypermobility/ISD | genitourinary | RATE_C | |
| 598 | | | | | 27202 | 318 | 26884 | | | | | 36132 | 426 | 35706 | 0.9087 | -0.02518 | Abnormal findings on examination of urine | genitourinary | RATE_C | |
| 598.4 | | | | | 26884 | | | | | | | 35709 | 3 | 35706 | | | Other cells and casts in urine | genitourinary | RATE_C | |
| 599 | 0.6176 | 0.04991 | 1.854 | 3.61e-35 | 23589 | 4209 | 19380 | 0.7948 | 0.0347 | 2.214 | 4.46e-116 | 31705 | 6851 | 24854 | 0.5362 | -0.03563 | Other symptoms/disorders or the urinary system | genitourinary | RATE_C | |
| 599.1 | | | | | 19615 | 235 | 19380 | | | | | 24927 | 73 | 24854 | | | Urinary obstruction | genitourinary | RATE_C | |
| 599.2 | 0.9765 | 0.08282 | 2.655 | 4.41e-32 | 21042 | 1662 | 19380 | 1.296 | 0.08815 | 3.655 | 6.10e-49 | 25979 | 1125 | 24854 | 0.1899 | 0.1538 | Retention of urine | genitourinary | RATE_C | |
| 599.3 | 0.7322 | 0.109 | 2.08 | 1.87e-11 | 20130 | 750 | 19380 | 0.7743 | 0.04783 | 2.169 | 6.08e-59 | 27990 | 3136 | 24854 | 0.2851 | -0.1208 | Dysuria | genitourinary | RATE_C | |

| phenotype | male_beta | male_SE | male_OR | male_p | male_n_total | male_n_cases | male_n_controls | female_beta | female_SE | female_OR | female_p | female_n_total | female_n_cases | female_n_controls | interaction_p | interaction_beta | description | group | lab | label |
|---|---|---|---|---|---|---|---|---|---|---|---|---|---|---|---|---|---|---|---|---|
| 599.4 | 0.7083 | 0.1122 | 2.03 | 2.70e-10 | 20122 | 742 | 19380 | 0.9327 | 0.06112 | 2.541 | 1.38e-52 | 27034 | 2180 | 24854 | 0.0685 | 0.2249 | Urinary incontinence | genitourinary | RATE_C | |
| 599.5 | 0.5289 | 0.0743 | 1.697 | 1.09e-12 | 21290 | 1910 | 19380 | 0.9721 | 0.06358 | 2.643 | 9.02e-53 | 26913 | 2059 | 24854 | 0.008573 | 0.2475 | Frequency of urination and polyuria | genitourinary | RATE_C | |
| 599.6 | | | | | 19476 | 96 | 19380 | | | | | 24972 | 118 | 24854 | | | Oliguria and anuria | genitourinary | RATE_C | |
| 599.7 | | | | | 19390 | 10 | 19380 | | | | | 24855 | 1 | 24854 | | | Urethral discharge | genitourinary | RATE_C | |
| 599.8 | | | | | 19615 | 235 | 19380 | | | | | 25086 | 232 | 24854 | | | Other symptoms involving urinary system | genitourinary | RATE_C | |
| 599.9 | 0.7648 | 0.1168 | 2.149 | 5.77e-11 | 20173 | 793 | 19380 | 0.9893 | 0.08025 | 2.689 | 6.37e-35 | 26091 | 1237 | 24854 | 0.5611 | 0.07958 | Other abnormality of urination | genitourinary | RATE_C | |
| 600 | 0.7535 | 0.06216 | 2.125 | 7.93e-34 | 23028 | 3640 | 19388 | | | | | 4 | | | 0.9915 | -3.251 | Hyperplasia of prostate | genitourinary | RATE_C | |
| 601 | 0.7199 | 0.111 | 2.054 | 8.73e-11 | 20083 | 695 | 19388 | | | | | 4 | | | 0.9979 | -0.8188 | Inflammatory diseases of prostate | genitourinary | RATE_C | |
| 601.1 | | | | | 19781 | 393 | 19388 | | | | | 4 | | | | | Prostatitis | genitourinary | RATE_C | |
| 601.11 | | | | | 19525 | 137 | 19388 | | | | | 4 | | | | | Acute prostatitis | genitourinary | RATE_C | |
| 601.12 | | | | | 19502 | 114 | 19388 | | | | | 4 | | | | | Chronic prostatitis | genitourinary | RATE_C | |
| 601.3 | | | | | 19577 | 189 | 19388 | | | | | 4 | | | | | Orchitis and epididymitis | genitourinary | RATE_C | |
| 601.4 | | | | | 19435 | 47 | 19388 | | | | | 4 | | | | | Balanoposthitis | genitourinary | RATE_C | |
| 601.8 | | | | | 19460 | 72 | 19388 | | | | | 4 | | | | | Other inflammatory disorders of male genital organs | genitourinary | RATE_C | |
| 602 | | | | | 19490 | 102 | 19388 | | | | | 4 | | | | | Other disorders of prostate | genitourinary | RATE_C | |
| 602.3 | | | | | 19390 | 2 | 19388 | | | | | 4 | | | | | Dysplasia of prostate | genitourinary | RATE_C | |

| phenotype | male_beta | male_SE | male_OR | male_p | male_n_total | male_n_cases | male_n_controls | female_beta | female_SE | female_OR | female_p | female_n_total | female_n_cases | female_n_controls | interaction_p | interaction_beta | description | group | lab | label |
|---|---|---|---|---|---|---|---|---|---|---|---|---|---|---|---|---|---|---|---|---|
| 603 | | | | | 19102 | 243 | 18859 | | | | | 4 | | | | | Other disorders of testis | genitourinary | RATE_C | |
| 603.1 | | | | | 19051 | 192 | 18859 | | | | | 4 | | | | | Hydrocele | genitourinary | RATE_C | |
| 603.2 | | | | | 18915 | 56 | 18859 | | | | | 4 | | | | | Spermatocele | genitourinary | RATE_C | |
| 604 | | | | | 19178 | 319 | 18859 | | | | | 4 | | | | | Disorders of penis | genitourinary | RATE_C | |
| 604.1 | | | | | 18955 | 96 | 18859 | | | | | 4 | | | | | Redundant prepuce and phimosis/BXO | genitourinary | RATE_C | |
| 604.2 | | | | | 18859 | | | | | | | 4 | | | | | Vascular disorders of penis | genitourinary | RATE_C | |
| 604.3 | | | | | 18985 | 126 | 18859 | | | | | 4 | | | | | Peyronie's disease | genitourinary | RATE_C | |
| 605 | 0.4955 | 0.06549 | 1.641 | 3.83e-14 | 21005 | 2146 | 18859 | | | | | 4 | | | 0.9976 | -0.9226 | Erectile dysfunction [ED] | genitourinary | RATE_C | |
| 608 | 0.5595 | 0.1134 | 1.75 | 8.14e-07 | 19398 | 539 | 18859 | | | | | 4 | | | 0.9999 | -0.09813 | Other disorders of male genital organs | genitourinary | RATE_C | |
| 609 | | | | | 18929 | 70 | 18859 | | | | | 4 | | | | | Male infertility and abnormal spermatozoa | genitourinary | RATE_C | |
| 609.1 | | | | | 18884 | 25 | 18859 | | | | | 4 | | | | | Infertility, male | genitourinary | RATE_C | |
| 609.11 | | | | | 18861 | 2 | 18859 | | | | | 4 | | | | | Azoospermia and oligospermia | genitourinary | RATE_C | |
| 609.2 | | | | | 18902 | 43 | 18859 | | | | | 4 | | | | | Abnormal spermatozoa | genitourinary | RATE_C | |
| 610 | | | | | 5 | | | 0.3896 | 0.06202 | 1.476 | 3.36e-10 | 29977 | 1611 | 28366 | 0.9997 | -0.1315 | Benign mammary dysplasias | genitourinary | RATE_C | |
| 610.1 | | | | | 5 | | | 0.382 | 0.07044 | 1.465 | 5.87e-08 | 29598 | 1232 | 28366 | 0.9996 | -0.1814 | Cystic mastopathy | genitourinary | RATE_C | |
| 610.2 | | | | | 5 | | | | | | | 28425 | 59 | 28366 | | | Fibroadenosis of breast | genitourinary | RATE_C | |
| 610.3 | | | | | 5 | | | | | | | 28501 | 135 | 28366 | | | Fibrosclerosis of breast | genitourinary | RATE_C | |
| 610.4 | | | | | 5 | | | | | | | 28633 | 267 | 28366 | | | Benign neoplasm of breast | genitourinary | RATE_C | |
| 610.8 | | | | | 5 | | | | | | | 28599 | 233 | 28366 | | | Other specified benign mammary dysplasias | genitourinary | RATE_C | |
| 611 | | | | | 27275 | 62 | 27213 | 0.4063 | 0.03782 | 1.501 | 6.39e-27 | 33455 | 5088 | 28367 | 0.1682 | -0.5939 | Abnormal findings on mammogram or breast exam | genitourinary | RATE_C | |
| 611.1 | | | | | 27225 | 12 | 27213 | 0.3997 | 0.04145 | 1.491 | 5.29e-22 | 32548 | 4181 | 28367 | 0.9179 | -11.18 | Abnormal mammogram | genitourinary | RATE_C | |
| 611.11 | | | | | 27213 | | | 0.7995 | 0.1193 | 2.224 | 2.10e-11 | 28926 | 559 | 28367 | 0.998 | 1.476 | Mammographic microcalcification | genitourinary | RATE_C | |
| 611.3 | | | | | 27266 | 53 | 27213 | 0.6811 | 0.06297 | 1.976 | 2.88e-27 | 30081 | 1714 | 28367 | 0.5894 | -0.2361 | Lump or mass in breast | genitourinary | RATE_C | |
| 612 | | | | | 27326 | 113 | 27213 | | | | | 28757 | 390 | 28367 | 0.4093 | -0.2155 | Breast conditions, congenital or relating to hormones | genitourinary | RATE_C | |
| 612.1 | | | | | 27213 | | | | | | | 28446 | 79 | 28367 | | | Galactorrhea | genitourinary | RATE_C | |
| 612.2 | | | | | 27325 | 112 | 27213 | | | | | 28664 | 297 | 28367 | | | Hypertrophy of breast (Gynecomastia) | genitourinary | RATE_C | |
| 612.3 | | | | | 27214 | 1 | 27213 | | | | | 28379 | 12 | 28367 | | | Congenital anomalies of breast | genitourinary | RATE_C | |
| 613 | | | | | 27667 | 60 | 27607 | 0.4686 | 0.05183 | 1.598 | 1.53e-19 | 34784 | 2435 | 32349 | 0.8151 | 0.07893 | Other nonmalignant breast conditions | genitourinary | RATE_C | |
| 613.1 | | | | | 27607 | | | | | | | 32616 | 267 | 32349 | | | Inflammatory disease of breast | genitourinary | RATE_C | |

| phenotype | male_beta | male_SE | male_OR | male_p | male_n_total | male_n_cases | male_n_controls | female_beta | female_SE | female_OR | female_p | female_n_total | female_n_cases | female_n_controls | interaction_p | interaction_beta | description | group | lab | label |
|---|---|---|---|---|---|---|---|---|---|---|---|---|---|---|---|---|---|---|---|---|
| 613.5 | | | | | 27649 | 42 | 27607 | 0.4398 | 0.07801 | 1.552 | 1.72e-08 | 33336 | 987 | 32349 | 0.6388 | 0.18 | Mastodynia | genitourinary | RATE_C | |
| 613.7 | | | | | 27616 | 9 | 27607 | | | | | 32785 | 436 | 32349 | | | Other signs and symptoms in breast | genitourinary | RATE_C | |
| 613.8 | | | | | 27608 | 1 | 27607 | 0.4392 | 0.09627 | 1.551 | 5.08e-06 | 33034 | 685 | 32349 | 0.9567 | -9.678 | Other specified disorders of breast | genitourinary | RATE_C | |
| 613.9 | | | | | 27609 | 2 | 27607 | | | | | 32422 | 73 | 32349 | | | Breast disorder NOS | genitourinary | RATE_C | |
| 614 | | | | | 7 | | | 0.5181 | 0.05941 | 1.679 | 2.79e-18 | 33657 | 1625 | 32032 | 0.9995 | 0.1845 | Inflammatory diseases of female pelvic organs | genitourinary | RATE_C | |
| 614.1 | | | | | 7 | | | | | | | 32176 | 144 | 32032 | | | Pelvic peritoneal adhesions, female (postoperative) (postinfection) | genitourinary | RATE_C | |
| 614.3 | | | | | 7 | | | | | | | 32232 | 200 | 32032 | | | Pelvic inflammatory disease (PID) | genitourinary | RATE_C | |
| 614.31 | | | | | 7 | | | | | | | 32055 | 23 | 32032 | | | Acute inflammatory pelvic disease | genitourinary | RATE_C | |
| 614.32 | | | | | 7 | | | | | | | 32140 | 108 | 32032 | | | Chronic inflammatory pelvic disease | genitourinary | RATE_C | |
| 614.33 | | | | | 7 | | | | | | | 32100 | 68 | 32032 | | | Pelvic inflammatory disease, NOS | genitourinary | RATE_C | |
| 614.4 | | | | | 7 | | | | | | | 32069 | 37 | 32032 | | | Inflammatory diseases of uterus, except cervix | genitourinary | RATE_C | |
| 614.5 | | | | | 7 | | | 0.4623 | 0.06921 | 1.588 | 2.40e-11 | 33209 | 1177 | 32032 | 0.9995 | 0.1561 | Inflammatory disease of cervix, vagina, and vulva | genitourinary | RATE_C | |
| 614.51 | | | | | 7 | | | | | | | 32101 | 69 | 32032 | | | Cervicitis and endocervicitis | genitourinary | RATE_C | |
| 614.52 | | | | | 7 | | | 0.4321 | 0.08067 | 1.54 | 8.51e-08 | 32877 | 845 | 32032 | 0.9998 | 0.1199 | Vaginitis and vulvovaginitis | genitourinary | RATE_C | |
| 614.53 | | | | | 7 | | | | | | | 32093 | 61 | 32032 | | | Cyst or abscess of Bartholin's gland | genitourinary | RATE_C | |
| 614.54 | | | | | 7 | | | | | | | 32097 | 65 | 32032 | | | Abscess or ulceration of vulva | genitourinary | RATE_C | |
| 615 | | | | | 7 | | | | | | | 32416 | 384 | 32032 | | | Endometriosis | genitourinary | RATE_C | |
| 618 | | | | | 7 | | | 0.6278 | 0.08234 | 1.873 | 2.45e-14 | 36315 | 1140 | 35175 | 0.9975 | 0.8574 | Genital prolapse | genitourinary | RATE_C | |
| 618.1 | | | | | 7 | | | 0.7449 | 0.0964 | 2.106 | 1.10e-14 | 36071 | 896 | 35175 | 0.9982 | 0.9858 | Prolapse of vaginal walls | genitourinary | RATE_C | |
| 618.2 | | | | | 7 | | | | | | | 35515 | 340 | 35175 | | | Uterine/Uterovaginal prolapse | genitourinary | RATE_C | |
| 618.5 | | | | | 7 | | | | | | | 35494 | 319 | 35175 | | | Prolapse of vaginal vault after hysterectomy | genitourinary | RATE_C | |
| 618.6 | | | | | 7 | | | | | | | 35376 | 201 | 35175 | | | Vaginal enterocele, congenital or acquired | genitourinary | RATE_C | |
| 619 | | | | | 7 | | | 0.4824 | 0.0525 | 1.62 | 4.00e-20 | 33857 | 2126 | 31731 | 0.9994 | 0.1989 | Noninflammatory female genital disorders | genitourinary | RATE_C | |
| 619.1 | | | | | 7 | | | | | | | 32002 | 271 | 31731 | | | Noninflammatory disorders of ovary, fallopian tube, and broad ligament | genitourinary | RATE_C | |
| 619.2 | | | | | 7 | | | | | | | 32192 | 461 | 31731 | | | Disorders of uterus, NEC | genitourinary | RATE_C | |
| 619.3 | | | | | 7 | | | | | | | 31864 | 133 | 31731 | | | Noninflammatory disorders of cervix | genitourinary | RATE_C | |
| 619.4 | | | | | 7 | | | 0.8123 | 0.1059 | 2.253 | 1.70e-14 | 32259 | 528 | 31731 | 0.9991 | 0.5289 | Noninflammatory disorders of vagina | genitourinary | RATE_C | |

| phenotype | male_beta | male_SE | male_OR | male_p | male_n_total | male_n_cases | male_n_controls | female_beta | female_SE | female_OR | female_p | female_n_total | female_n_cases | female_n_controls | interaction_p | interaction_beta | description | group | lab | label |
|---|---|---|---|---|---|---|---|---|---|---|---|---|---|---|---|---|---|---|---|---|
| 619.5 | | | | | 7 | | | | | | | 31969 | 238 | 31731 | | | Noninflammatory disorders of vulva and perineum | genitourinary | RATE_C | |
| 620 | | | | | 7 | | | | | | | 35251 | 113 | 35138 | | | Dysplasia of female genital organs | genitourinary | RATE_C | |
| 620.1 | | | | | 7 | | | | | | | 35152 | 14 | 35138 | | | Dysplasia of cervix | genitourinary | RATE_C | |
| 621 | | | | | 7 | | | | | | | 35309 | 171 | 35138 | | | Endometrial hyperplasia | genitourinary | RATE_C | |
| 622 | | | | | 7 | | | | | | | 35534 | 396 | 35138 | | | Polyp of female genital organs | genitourinary | RATE_C | |
| 622.1 | | | | | 7 | | | | | | | 35431 | 293 | 35138 | | | Polyp of corpus uteri | genitourinary | RATE_C | |
| 622.2 | | | | | 7 | | | | | | | 35215 | 77 | 35138 | | | Mucous polyp of cervix | genitourinary | RATE_C | |
| 623 | | | | | 7 | | | | | | | 35299 | 161 | 35138 | | | Hypertrophy of female genital organs | genitourinary | RATE_C | |
| 624 | | | | | 7 | | | | | | | 31709 | 227 | 31482 | | | Symptoms involving female genital tract | genitourinary | RATE_C | |
| 624.1 | | | | | 7 | | | | | | | 31539 | 57 | 31482 | | | Dystrophy of female genital tract | genitourinary | RATE_C | |
| 624.2 | | | | | 7 | | | | | | | 31503 | 21 | 31482 | | | Atrophy of female genital tract | genitourinary | RATE_C | |
| 624.9 | | | | | 7 | | | 0.9289 | 0.09135 | 2.532 | 2.73e-24 | 32466 | 984 | 31482 | 0.9982 | 1.022 | stress incontinence, female | genitourinary | RATE_C | |
| 625 | | | | | 7 | | | 0.7792 | 0.06463 | 2.18 | 1.79e-33 | 32899 | 1417 | 31482 | 0.9986 | 0.4631 | Pain and other symptoms associated with female genital organs | genitourinary | RATE_C | |
| 625.1 | | | | | 7 | | | | | | | 31749 | 267 | 31482 | | | Dyspareunia | genitourinary | RATE_C | |
| 626 | | | | | 5 | | | 0.3969 | 0.04253 | 1.487 | 1.04e-20 | 27458 | 3874 | 23584 | 0.9994 | -0.1718 | Disorders of menstruation and other abnormal bleeding from female genital tract | genitourinary | RATE_C | |
| 626.1 | | | | | 5 | | | 0.4703 | 0.04652 | 1.6 | 5.07e-24 | 26685 | 3101 | 23584 | 0.9998 | -0.1077 | Irregular menstrual cycle/bleeding | genitourinary | RATE_C | |
| 626.11 | | | | | 5 | | | 0.5388 | 0.09667 | 1.714 | 2.50e-08 | 24169 | 585 | 23584 | 1 | -0.007313 | Absent or infrequent menstruation | genitourinary | RATE_C | |
| 626.12 | | | | | 5 | | | 0.7833 | 0.07223 | 2.189 | 2.12e-27 | 24734 | 1150 | 23584 | 0.9995 | 0.2254 | Excessive or frequent menstruation | genitourinary | RATE_C | |
| 626.13 | | | | | 5 | | | 0.3955 | 0.07183 | 1.485 | 3.65e-08 | 24711 | 1127 | 23584 | 0.9995 | -0.2173 | Irregular menstrual cycle | genitourinary | RATE_C | |
| 626.14 | | | | | 5 | | | | | | | 23850 | 266 | 23584 | | | Irregular menstrual bleeding | genitourinary | RATE_C | |
| 626.15 | | | | | 5 | | | | | | | 23629 | 45 | 23584 | | | Infertility, female, associated with anovulation | genitourinary | RATE_C | |
| 626.2 | | | | | 5 | | | | | | | 24029 | 445 | 23584 | | | Dysmenorrhea | genitourinary | RATE_C | |
| 626.21 | | | | | 5 | | | | | | | 23591 | 7 | 23584 | | | Mittelschmerz | genitourinary | RATE_C | |
| 626.4 | | | | | 5 | | | | | | | 23636 | 52 | 23584 | | | Premenstrual tension syndromes | genitourinary | RATE_C | |
| 626.8 | | | | | 5 | | | | | | | 23771 | 187 | 23584 | | | Infertility, female | genitourinary | RATE_C | |
| 627 | | | | | 6 | | | 0.6025 | 0.04074 | 1.827 | 1.69e-49 | 28947 | 5363 | 23584 | 0.9384 | 9.238 | Menopausal and postmenopausal disorders | genitourinary | RATE_C | |
| 627.1 | | | | | 5 | | | 0.5738 | 0.09154 | 1.775 | 3.65e-10 | 24382 | 798 | 23584 | 1 | -0.002511 | Postmenopausal bleeding | genitourinary | RATE_C | |
| 627.2 | | | | | 6 | | | 0.6087 | 0.04919 | 1.838 | 3.60e-35 | 26962 | 3378 | 23584 | 0.9386 | 9.205 | Symptomatic menopause | genitourinary | RATE_C | |

| phenotype | male_beta | male_SE | male_OR | male_p | male_n_total | male_n_cases | male_n_controls | female_beta | female_SE | female_OR | female_p | female_n_total | female_n_cases | female_n_controls | interaction_p | interaction_beta | description | group | lab | label |
|---|---|---|---|---|---|---|---|---|---|---|---|---|---|---|---|---|---|---|---|---|
| 627.21 | | | | | 5 | | | | | | | 23934 | 350 | 23584 | | | Symptomatic artificial menopause | genitourinary | RATE_C | |
| 627.22 | | | | | 6 | | | 0.5741 | 0.08875 | 1.775 | 9.89e-11 | 24465 | 881 | 23584 | 0.9588 | 10.18 | Need for Hormone replacement therapy (postmenopausal) | genitourinary | RATE_C | |
| 627.3 | | | | | 5 | | | 0.7386 | 0.07883 | 2.093 | 7.29e-21 | 24943 | 1359 | 23584 | 0.9997 | 0.1263 | Postmenopausal atrophic vaginitis | genitourinary | RATE_C | |
| 627.4 | | | | | 5 | | | | | | | 23687 | 103 | 23584 | | | Premenopausal menorrhagia | genitourinary | RATE_C | |
| 627.5 | | | | | 5 | | | | | | | 23742 | 158 | 23584 | | | Premature menopause and other ovarian failure | genitourinary | RATE_C | |
| 628 | | | | | 5 | | | 0.8121 | 0.07631 | 2.253 | 1.89e-26 | 24586 | 1002 | 23584 | 0.9992 | 0.3765 | Ovarian cyst | genitourinary | RATE_C | |
| 634 | | | | | 7 | | | -0.15 | 0.1054 | 0.8607 | 0.1548 | 34235 | 519 | 33716 | 0.9986 | -0.7798 | Miscarriage; stillbirth | pregnancy complications | RATE_C | |
| 634.1 | | | | | 7 | | | | | | | 33945 | 229 | 33716 | | | Missed abortion/Hydatidiform mole | pregnancy complications | RATE_C | |
| 634.3 | | | | | 7 | | | | | | | 33757 | 41 | 33716 | | | Ectopic pregnancy | pregnancy complications | RATE_C | |
| 635 | | | | | 7 | | | | | | | 34113 | 397 | 33716 | | | Hemorrhage during pregnancy; childbirth and postpartum | pregnancy complications | RATE_C | |
| 635.2 | | | | | 7 | | | | | | | 33989 | 273 | 33716 | | | Antepartum hemorrhage, abruptio placentae, and placenta previa | pregnancy complications | RATE_C | |
| 635.3 | | | | | 7 | | | | | | | 33851 | 135 | 33716 | | | Placenta previa and abruptio placenta | pregnancy complications | RATE_C | |
| 636 | | | | | 7 | | | -0.01551 | 0.05978 | 0.9846 | 0.7954 | 35417 | 1701 | 33716 | 0.9977 | -0.7695 | Early or threatened labor; hemorrhage in early pregnancy | pregnancy complications | RATE_C | |
| 636.1 | | | | | 7 | | | | | | | 34209 | 493 | 33716 | | | Threatened premature labor | pregnancy complications | RATE_C | |
| 636.2 | | | | | 7 | | | | | | | 34111 | 395 | 33716 | | | Early onset of delivery | pregnancy complications | RATE_C | |
| 636.3 | | | | | 7 | | | -0.02831 | 0.1048 | 0.9721 | 0.7871 | 34236 | 520 | 33716 | 0.9987 | -0.7137 | Hemorrhage in early pregnancy | pregnancy complications | RATE_C | |
| 636.8 | | | | | 7 | | | | | | | 33814 | 98 | 33716 | | | Cervical incompetence | pregnancy complications | RATE_C | |
| 637 | | | | | 27759 | 8 | 27751 | | | | | 33709 | 19 | 33690 | | | Short gestation; low birth weight; and fetal growth retardation | pregnancy complications | RATE_C | |
| 642 | | | | | 7 | | | 0.4709 | 0.08362 | 1.601 | 1.79e-08 | 36632 | 759 | 35873 | 0.9995 | -0.26 | Hypertension complicating pregnancy, childbirth, and the puerperium | pregnancy complications | RATE_C | |
| 642.1 | | | | | 7 | | | | | | | 36229 | 356 | 35873 | | | Preeclampsia and eclampsia | pregnancy complications | RATE_C | |
| phenotype | male_beta | male_SE | male_OR | male_p | male_n_total | male_n_cases | male_n_controls | female_beta | female_SE | female_OR | female_p | female_n_total | female_n_cases | female_n_controls | interaction_p | interaction_beta | description | group | lab | label |
| 643 | | | | | 7 | | | | | | | 36714 | 156 | 36558 | | | Excessive vomiting in pregnancy | pregnancy complications | RATE_C | |
| 643.1 | | | | | 7 | | | | | | | 36608 | 50 | 36558 | | | Hyperemesis gravidarum | pregnancy complications | RATE_C | |
| 644 | | | | | 7 | | | | | | | 36546 | 186 | 36360 | | | Anemia during pregnancy | pregnancy complications | RATE_C | |
| 646 | | | | | 7 | | | 0.5827 | 0.09739 | 1.791 | 2.18e-09 | 36367 | 578 | 35789 | 0.9994 | -0.3581 | Other complications of pregnancy NEC | pregnancy complications | RATE_C | |
| 647 | | | | | 7 | | | | | | | 36080 | 179 | 35901 | | | Infectious and parasitic complications affecting pregnancy | pregnancy complications | RATE_C | |
| 647.1 | | | | | 7 | | | | | | | 36102 | 201 | 35901 | | | Infections of genitourinary tract during pregnancy | pregnancy complications | RATE_C | |
| 647.3 | | | | | 7 | | | | | | | 35950 | 49 | 35901 | | | Major puerperal infection | pregnancy complications | RATE_C | |

| phenotype | male_beta | male_SE | male_OR | male_p | male_n_total | male_n_cases | male_n_controls | female_beta | female_SE | female_OR | female_p | female_n_total | female_n_cases | female_n_controls | interaction_p | interaction_beta | description | group | lab | label |
|---|---|---|---|---|---|---|---|---|---|---|---|---|---|---|---|---|---|---|---|---|
| 649 | | | | | 7 | | | 0.2239 | 0.06007 | 1.251 | 1.94e-04 | 35652 | 1664 | 33988 | 0.9983 | -0.5757 | Other conditions or status of the mother complicating pregnancy, childbirth, or the puerperium | pregnancy complications | RATE_C | |
| 649.1 | | | | | 7 | | | 0.4498 | 0.09461 | 1.568 | 1.99e-06 | 34593 | 605 | 33988 | 0.9995 | -0.2652 | Diabetes or abnormal glucose tolerance complicating pregnancy | pregnancy complications | RATE_C | |
| 653 | | | | | 7 | | | 0.05502 | 0.09366 | 1.057 | 0.5569 | 35753 | 642 | 35111 | 0.9988 | -0.6528 | Problems associated with amniotic cavity and membranes | pregnancy complications | RATE_C | |
| 654 | | | | | 7 | | | | | | | 35408 | 297 | 35111 | | | Other and unspecified complications of birth; puerperium affecting management of mother | pregnancy complications | RATE_C | |
| 654.1 | | | | | 7 | | | | | | | 35312 | 201 | 35111 | | | Abnormality of organs and soft tissues of pelvis complicating pregnancy, childbirth, or the puerperium | pregnancy complications | RATE_C | |
| 654.2 | | | | | 7 | | | | | | | 35201 | 90 | 35111 | | | Rhesus isoimmunization in pregnancy | pregnancy complications | RATE_C | |
| 655 | | | | | 7 | | | -0.1319 | 0.05311 | 0.8764 | 0.01298 | 36061 | 2235 | 33826 | 0.9974 | -0.8748 | Known or suspected fetal abnormality affecting management of mother | pregnancy complications | RATE_C | |
| 655.1 | | | | | 7 | | | -0.2131 | 0.08805 | 0.8081 | 0.0155 | 34615 | 789 | 33826 | 0.9984 | -0.9102 | Abnormality in fetal heart rate or rhythm | pregnancy complications | RATE_C | |
| 656 | | | | | 27627 | 68 | 27559 | | | | | 36677 | 120 | 36557 | | | Other perinatal conditions of fetus or newborn | pregnancy complications | RATE_C | |
| 656.1 | | | | | 27562 | 3 | 27559 | | | | | 36558 | 1 | 36557 | | | Isoimmunization of fetus or newborn | pregnancy complications | RATE_C | |
| 656.2 | | | | | 27597 | 38 | 27559 | | | | | 36585 | 28 | 36557 | | | Respiratory conditions of fetus and newborn | pregnancy complications | RATE_C | |
| 656.22 | | | | | 27562 | 3 | 27559 | | | | | 36559 | 2 | 36557 | | | Interstitial emphysema and related conditions of newborn | pregnancy complications | RATE_C | |
| 656.26 | | | | | 27567 | 8 | 27559 | | | | | 36561 | 4 | 36557 | | | Transitory tachypnea or apnea of newborn | pregnancy complications | RATE_C | |
| 656.3 | | | | | 27567 | 8 | 27559 | | | | | 36567 | 10 | 36557 | | | Endocrine and metabolic disturbances of fetus and newborn | pregnancy complications | RATE_C | |
| 656.4 | | | | | 27568 | 9 | 27559 | | | | | 36563 | 6 | 36557 | | | Hemorrhage of fetus or newborn | pregnancy complications | RATE_C | |
| 656.5 | | | | | 27565 | 6 | 27559 | | | | | 36562 | 5 | 36557 | | | Hematological disorders of newborn | pregnancy complications | RATE_C | |
| 656.6 | | | | | 27567 | 8 | 27559 | | | | | 36562 | 5 | 36557 | | | Perinatal disorders of digestive system | pregnancy complications | RATE_C | |
| 656.7 | | | | | 27566 | 7 | 27559 | | | | | 36566 | 9 | 36557 | | | Conditions involving the integument and temperature regulation of fetus and newborn | pregnancy complications | RATE_C | |
| 656.8 | | | | | 27575 | 16 | 27559 | | | | | 36572 | 15 | 36557 | | | Perinatal jaundice | pregnancy complications | RATE_C | |
| 656.9 | | | | | 27559 | | | | | | | 36559 | 2 | 36557 | | | Neonatal bradycardia or tachycardia | pregnancy complications | RATE_C | |
| 657 | | | | | 27745 | 7 | 27738 | | | | | 36919 | 6 | 36913 | | | Infections specific to the perinatal period | pregnancy complications | RATE_C | |
| 669 | | | | | 7 | | | | | | | 36606 | 168 | 36438 | | | Complications of labor and delivery NEC | pregnancy complications | RATE_C | |
| 671 | | | | | 7 | | | | | | | 36547 | 117 | 36430 | | | Venous/cerebrovascular complications embolism in pregnancy and the puerperium | pregnancy complications | RATE_C | |
| 674 | | | | | 7 | | | | | | | 36509 | 79 | 36430 | | | Other complications of the puerperium NEC | pregnancy complications | RATE_C | |
| 676 | | | | | 7 | | | | | | | 36440 | 10 | 36430 | | | Other disorders of the breast associated with childbirth and disorders of lactation | pregnancy complications | RATE_C | |

| phenotype | male_beta | male_SE | male_OR | male_p | male_n_total | male_n_cases | male_n_controls | female_beta | female_SE | female_OR | female_p | female_n_total | female_n_cases | female_n_controls | interaction_p | interaction_beta | description | group | lab | label |
|---|---|---|---|---|---|---|---|---|---|---|---|---|---|---|---|---|---|---|---|---|
| 681 | 0.8501 | 0.05861 | 2.34 | 1.16e-47 | 24732 | 2737 | 21995 | 0.9791 | 0.04957 | 2.662 | 7.73e-87 | 32934 | 3053 | 29881 | 0.003633 | 0.2077 | Superficial cellulitis and abscess | dermatologic | RATE_C | |
| 681.1 | | | | | 22394 | 399 | 21995 | | | | | 30304 | 423 | 29881 | 0.2671 | 0.2013 | Cellulitis and abscess of fingers/toes | dermatologic | RATE_C | |
| 681.2 | | | | | 22204 | 209 | 21995 | | | | | 30123 | 242 | 29881 | | | Cellulitis and abscess of face/neck | dermatologic | RATE_C | |
| 681.3 | | | | | 22375 | 380 | 21995 | | | | | 30260 | 379 | 29881 | 0.1652 | 0.2664 | Cellulitis and abscess of arm/hand | dermatologic | RATE_C | |
| 681.5 | 1.157 | 0.1063 | 3.182 | 1.34e-27 | 22978 | 983 | 21995 | 1.159 | 0.09801 | 3.185 | 3.05e-32 | 30774 | 893 | 29881 | 0.1317 | 0.2068 | Cellulitis and abscess of leg, except foot | dermatologic | RATE_C | |
| 681.6 | | | | | 22233 | 238 | 21995 | | | | | 30029 | 148 | 29881 | | | Cellulitis and abscess of foot, toe | dermatologic | RATE_C | |
| 681.7 | | | | | 22491 | 496 | 21995 | 1.562 | 0.1126 | 4.769 | 9.47e-44 | 30505 | 624 | 29881 | 0.1533 | 0.2373 | Cellulitis and abscess of trunk | dermatologic | RATE_C | |
| 686 | 1.076 | 0.123 | 2.933 | 2.12e-18 | 22609 | 614 | 21995 | 1.219 | 0.1099 | 3.384 | 1.38e-28 | 30530 | 649 | 29881 | 0.05142 | 0.3002 | Other local infections of skin and subcutaneous tissue | dermatologic | RATE_C | |
| 686.1 | | | | | 22123 | 128 | 21995 | | | | | 30019 | 138 | 29881 | | | Carbuncle and furuncle | dermatologic | RATE_C | |
| 686.2 | | | | | 22049 | 54 | 21995 | | | | | 29945 | 64 | 29881 | | | Impetigo | dermatologic | RATE_C | |
| 686.3 | | | | | 22049 | 54 | 21995 | | | | | 29925 | 44 | 29881 | | | Pilonidal cyst | dermatologic | RATE_C | |
| 686.4 | | | | | 22031 | 36 | 21995 | | | | | 29904 | 23 | 29881 | | | Pyogenic granuloma | dermatologic | RATE_C | |
| 686.5 | | | | | 22021 | 26 | 21995 | | | | | 29916 | 35 | 29881 | | | Pyoderma | dermatologic | RATE_C | |
| 687 | | | | | 21481 | 302 | 21179 | 0.7228 | 0.1068 | 2.06 | 1.29e-11 | 26852 | 564 | 26288 | 0.04919 | 0.3446 | Symptoms affecting skin | dermatologic | RATE_C | |
| 687.1 | 0.5176 | 0.0676 | 1.678 | 1.90e-14 | 22861 | 1682 | 21179 | 0.8251 | 0.05027 | 2.282 | 1.53e-60 | 29124 | 2836 | 26288 | 3.25e-08 | 0.43 | Rash and other nonspecific skin eruption | dermatologic | RATE_C | |
| 687.3 | | | | | 21208 | 29 | 21179 | | | | | 26359 | 71 | 26288 | | | Changes in skin texture | dermatologic | RATE_C | |
| 687.4 | 0.6146 | 0.07732 | 1.849 | 1.89e-15 | 22559 | 1380 | 21179 | 0.7652 | 0.05224 | 2.149 | 1.39e-48 | 28776 | 2488 | 26288 | 0.05433 | 0.1664 | Disturbance of skin sensation | dermatologic | RATE_C | |
| 689 | 0.2827 | 0.09581 | 1.327 | 0.003176 | 26203 | 833 | 25370 | 0.5525 | 0.07887 | 1.738 | 2.46e-12 | 34664 | 1062 | 33602 | 0.01833 | 0.2739 | Disorder of skin and subcutaneous tissue NOS | dermatologic | RATE_C | |
| 690 | | | | | 23839 | 493 | 23346 | | | | | 29843 | 385 | 29458 | 0.9413 | -0.01201 | Erythematosquamous dermatosis | dermatologic | RATE_C | |
| 690.1 | | | | | 23835 | 489 | 23346 | | | | | 29839 | 381 | 29458 | 0.7956 | -0.0425 | Seborrheic dermatitis | dermatologic | RATE_C | |
| 691 | | | | | 23418 | 72 | 23346 | | | | | 29595 | 137 | 29458 | | | Congenital anomalies of skin | dermatologic | RATE_C | |
| 691.1 | | | | | 23351 | 5 | 23346 | | | | | 29460 | 2 | 29458 | | | Ichthyosis congenita | dermatologic | RATE_C | |
| 691.3 | | | | | 23350 | 4 | 23346 | | | | | 29482 | 24 | 29458 | | | Congenital pigmentary anomalies of skin | dermatologic | RATE_C | |
| 694 | 0.05636 | 0.09574 | 1.058 | 0.556 | 24181 | 835 | 23346 | 0.1828 | 0.06491 | 1.201 | 0.004865 | 30929 | 1471 | 29458 | 0.9483 | 0.007041 | Dyschromia and Vitiligo | dermatologic | RATE_C | |
| 694.1 | | | | | 23374 | 28 | 23346 | | | | | 29501 | 43 | 29458 | | | Vitiligo | dermatologic | RATE_C | |
| 694.2 | 0.05538 | 0.09794 | 1.057 | 0.5717 | 24148 | 802 | 23346 | 0.1804 | 0.06596 | 1.198 | 0.006239 | 30877 | 1419 | 29458 | 0.9692 | -0.004278 | Other dyschromia | dermatologic | RATE_C | |
| 694.3 | | | | | 23359 | 13 | 23346 | | | | | 29477 | 19 | 29458 | | | Vascular disorders of skin | dermatologic | RATE_C | |
| 695 | 0.2662 | 0.0954 | 1.305 | 0.005271 | 24180 | 834 | 23346 | 0.1925 | 0.05044 | 1.212 | 1.36e-04 | 31756 | 2298 | 29458 | 0.02045 | -0.2335 | Erythematous conditions | dermatologic | RATE_C | |

| phenotype | male_beta | male_SE | male_OR | male_p | male_n_total | male_n_cases | male_n_controls | female_beta | female_SE | female_OR | female_p | female_n_total | female_n_cases | female_n_controls | interaction_p | interaction_beta | description | group | lab | label |
|---|---|---|---|---|---|---|---|---|---|---|---|---|---|---|---|---|---|---|---|---|
| 695.1 | | | | | 23358 | 12 | 23346 | | | | | 29480 | 22 | 29458 | | | Toxic erythema | dermatologic | RATE_C | |
| 695.2 | | | | | 23377 | 31 | 23346 | | | | | 29513 | 55 | 29458 | | | Bullous dermatoses | dermatologic | RATE_C | |
| 695.21 | | | | | 23356 | 10 | 23346 | | | | | 29480 | 22 | 29458 | | | Dermatitis herpetiformis | dermatologic | RATE_C | |
| 695.22 | | | | | 23363 | 17 | 23346 | | | | | 29486 | 28 | 29458 | | | Pemphigus and pemphigoid | dermatologic | RATE_C | |
| 695.3 | | | | | 23613 | 267 | 23346 | 0.2421 | 0.09403 | 1.274 | 0.01003 | 30121 | 663 | 29458 | 0.5207 | 0.1118 | Rosacea | dermatologic | RATE_C | |
| 695.4 | | | | | 23118 | 140 | 22978 | -0.2121 | 0.07378 | 0.8089 | 0.004042 | 29377 | 925 | 28452 | 0.8612 | -0.03372 | Lupus (localized and systemic) | dermatologic | RATE_C | |
| 695.41 | | | | | 23008 | 30 | 22978 | | | | | 28621 | 169 | 28452 | | | Cutaneous lupus erythematosus | dermatologic | RATE_C | |
| 695.42 | | | | | 23104 | 126 | 22978 | -0.1941 | 0.07604 | 0.8236 | 0.01071 | 29322 | 870 | 28452 | 0.8153 | -0.04744 | Systemic lupus erythematosus | dermatologic | RATE_C | |
| 695.7 | | | | | 23445 | 99 | 23346 | | | | | 29666 | 208 | 29458 | | | Prurigo and Lichen | dermatologic | RATE_C | |
| 695.8 | | | | | 23469 | 123 | 23346 | | | | | 29759 | 301 | 29458 | | | Other specified erythematous conditions | dermatologic | RATE_C | |
| 695.81 | | | | | 23357 | 11 | 23346 | | | | | 29501 | 43 | 29458 | | | Erythema nodosum | dermatologic | RATE_C | |
| 695.9 | | | | | 23478 | 132 | 23346 | | | | | 29596 | 138 | 29458 | | | Unspecified erythematous condition | dermatologic | RATE_C | |
| 696 | | | | | 23091 | 490 | 22601 | -0.2596 | 0.08601 | 0.7714 | 0.002544 | 28469 | 689 | 27780 | 0.9557 | 0.00712 | Psoriasis and related disorders | dermatologic | RATE_C | |
| 696.2 | | | | | 23352 | 6 | 23346 | | | | | 29465 | 7 | 29458 | | | Parapsoriasis | dermatologic | RATE_C | |
| 696.3 | | | | | 23354 | 8 | 23346 | | | | | 29466 | 8 | 29458 | | | Pityriasis | dermatologic | RATE_C | |
| 696.4 | | | | | 23086 | 485 | 22601 | -0.2725 | 0.08685 | 0.7615 | 0.001702 | 28453 | 673 | 27780 | 0.9206 | -0.01287 | Psoriasis | dermatologic | RATE_C | |
| 696.41 | | | | | 23001 | 400 | 22601 | -0.1651 | 0.0977 | 0.8478 | 0.09113 | 28318 | 538 | 27780 | 0.8492 | -0.02756 | Psoriasis vulgaris | dermatologic | RATE_C | |
| 696.42 | | | | | 22782 | 181 | 22601 | | | | | 28051 | 271 | 27780 | | | Psoriatic arthropathy | dermatologic | RATE_C | |
| 697 | | | | | 23461 | 115 | 23346 | | | | | 29654 | 196 | 29458 | | | Sarcoidosis | dermatologic | RATE_C | |
| 698 | | | | | 27077 | 348 | 26729 | 0.5501 | 0.09097 | 1.733 | 1.48e-09 | 35552 | 771 | 34781 | 0.5051 | -0.1156 | Pruritus and related conditions | dermatologic | RATE_C | |
| 700 | | | | | 24764 | 207 | 24557 | | | | | 32043 | 272 | 31771 | | | Corns and callosities | dermatologic | RATE_C | |
| 701 | 0.3248 | 0.08318 | 1.384 | 9.44e-05 | 25713 | 1156 | 24557 | 0.5652 | 0.05582 | 1.76 | 4.20e-24 | 33879 | 2108 | 31771 | 0.1145 | 0.1475 | Other hypertrophic and atrophic conditions of skin | dermatologic | RATE_C | |
| 701.1 | | | | | 24638 | 81 | 24557 | | | | | 31856 | 85 | 31771 | | | Keratoderma, acquired | dermatologic | RATE_C | |
| 701.2 | | | | | 24996 | 439 | 24557 | 0.651 | 0.08485 | 1.917 | 1.69e-14 | 32665 | 894 | 31771 | 0.08364 | 0.2527 | Scar conditions and fibrosis of skin | dermatologic | RATE_C | |
| 701.3 | | | | | 24568 | 11 | 24557 | | | | | 31968 | 197 | 31771 | | | Circumscribed scleroderma | dermatologic | RATE_C | |
| 701.4 | | | | | 24632 | 75 | 24557 | | | | | 31928 | 157 | 31771 | | | Keloid scar | dermatologic | RATE_C | |
| 701.5 | | | | | 24606 | 49 | 24557 | | | | | 31836 | 65 | 31771 | | | Abnormal granulation tissue | dermatologic | RATE_C | |
| 701.6 | | | | | 24575 | 18 | 24557 | | | | | 31812 | 41 | 31771 | | | Acquired acanthosis nigricans | dermatologic | RATE_C | |
| 702 | 0.2301 | 0.06039 | 1.259 | 1.39e-04 | 24883 | 2887 | 21996 | 0.4426 | 0.05098 | 1.557 | 3.91e-18 | 32069 | 3086 | 28983 | 0.03258 | 0.1628 | Degenerative skin conditions and other dermatoses | dermatologic | RATE_C | |

| phenotype | male_beta | male_SE | male_OR | male_p | male_n_total | male_n_cases | male_n_controls | female_beta | female_SE | female_OR | female_p | female_n_total | female_n_cases | female_n_controls | interaction_p | interaction_beta | description | group | lab | label |
|---|---|---|---|---|---|---|---|---|---|---|---|---|---|---|---|---|---|---|---|---|
| 702.1 | 0.2043 | 0.0702 | 1.227 | 0.003618 | 25602 | 2131 | 23471 | 0.4014 | 0.07189 | 1.494 | 2.35e-08 | 33264 | 1502 | 31762 | 0.1051 | 0.1582 | Actinic keratosis | dermatologic | RATE_C | |
| 702.2 | 0.1783 | 0.06946 | 1.195 | 0.01024 | 25554 | 2083 | 23471 | 0.3633 | 0.05471 | 1.438 | 3.13e-11 | 34321 | 2559 | 31762 | 0.164 | 0.1185 | Seborrheic keratosis | dermatologic | RATE_C | |
| 702.4 | | | | | 21998 | 2 | 21996 | | | | | 28993 | 10 | 28983 | | | Degenerative skin disorders | dermatologic | RATE_C | |
| 703 | | | | | 25804 | 185 | 25619 | | | | | 33096 | 363 | 32733 | 0.504 | -0.1599 | Diseases of nail, NOS | dermatologic | RATE_C | |
| 703.1 | | | | | 25920 | 301 | 25619 | | | | | 33093 | 360 | 32733 | 0.2943 | 0.2134 | Ingrowing nail | dermatologic | RATE_C | |
| 704 | 0.3418 | 0.116 | 1.407 | 0.003224 | 26130 | 511 | 25619 | 0.3295 | 0.07324 | 1.39 | 6.84e-06 | 33832 | 1099 | 32733 | 0.05949 | 0.2344 | Diseases of hair and hair follicles | dermatologic | RATE_C | |
| 704.1 | | | | | 25706 | 87 | 25619 | 0.2944 | 0.09156 | 1.342 | 0.0013 | 33424 | 691 | 32733 | 2.90e-05 | 0.9841 | Alopecia | dermatologic | RATE_C | |
| 704.11 | | | | | 25639 | 20 | 25619 | | | | | 32796 | 63 | 32733 | | | Alopecia Areata | dermatologic | RATE_C | |
| 704.12 | | | | | 25622 | 3 | 25619 | | | | | 32824 | 91 | 32733 | | | Telogen effluvium | dermatologic | RATE_C | |
| 704.2 | | | | | 25621 | 2 | 25619 | | | | | 32916 | 183 | 32733 | | | Hirsutism | dermatologic | RATE_C | |
| 704.8 | | | | | 25903 | 284 | 25619 | | | | | 32966 | 233 | 32733 | 0.6957 | -0.08169 | Other specified diseases of hair and hair follicles | dermatologic | RATE_C | |
| 705 | | | | | 24635 | 110 | 24525 | | | | | 32184 | 194 | 31990 | | | Disorders of sweat glands | dermatologic | RATE_C | |
| 705.1 | | | | | 24588 | 63 | 24525 | | | | | 32070 | 80 | 31990 | | | Dyshidrosis | dermatologic | RATE_C | |
| 705.3 | | | | | 24563 | 38 | 24525 | | | | | 32095 | 105 | 31990 | | | Hidradenitis | dermatologic | RATE_C | |
| 705.8 | | | | | 24661 | 136 | 24525 | | | | | 32208 | 218 | 31990 | | | Hyperhidrosis | dermatologic | RATE_C | |
| 706 | 0.1512 | 0.07441 | 1.163 | 0.04221 | 25847 | 1322 | 24525 | 0.1918 | 0.0541 | 1.211 | 3.92e-04 | 33934 | 1944 | 31990 | 0.03543 | -0.1765 | Diseases of sebaceous glands | dermatologic | RATE_C | |
| 706.1 | | | | | 24797 | 272 | 24525 | 0.232 | 0.07989 | 1.261 | 0.003677 | 32819 | 829 | 31990 | 0.1997 | 0.1845 | Acne | dermatologic | RATE_C | |
| 706.2 | 0.2572 | 0.1048 | 1.293 | 0.01415 | 25246 | 721 | 24525 | 0.2244 | 0.0847 | 1.252 | 0.008069 | 32825 | 835 | 31990 | 0.8835 | -0.01854 | Sebaceous cyst | dermatologic | RATE_C | |
| 706.3 | | | | | 24526 | 1 | 24525 | | | | | 31991 | 1 | 31990 | | | Seborrhea | dermatologic | RATE_C | |
| 706.8 | | | | | 24813 | 288 | 24525 | | | | | 32225 | 235 | 31990 | 0.4024 | 0.2079 | Other specified diseases of sebaceous glands | dermatologic | RATE_C | |
| 707 | 1.392 | 0.1048 | 4.024 | 2.68e-40 | 26784 | 1253 | 25531 | 1.37 | 0.1023 | 3.933 | 6.63e-41 | 36008 | 999 | 35009 | 0.2935 | 0.1483 | Chronic ulcer of skin | dermatologic | RATE_C | |
| 707.1 | 1.429 | 0.1447 | 4.175 | 5.40e-23 | 26112 | 581 | 25531 | | | | | 35488 | 479 | 35009 | 0.2541 | 0.2223 | Decubitus ulcer | dermatologic | RATE_C | |
| 707.2 | 1.454 | 0.1532 | 4.282 | 2.25e-21 | 26220 | 689 | 25531 | | | | | 35483 | 474 | 35009 | 0.2254 | 0.2686 | Chronic ulcer of leg or foot | dermatologic | RATE_C | |
| 707.3 | | | | | 25727 | 196 | 25531 | | | | | 35188 | 179 | 35009 | | | Chronic ulcer of unspecified site | dermatologic | RATE_C | |
| 709 | | | | | 23344 | 211 | 23133 | -0.2884 | 0.06596 | 0.7494 | 1.23e-05 | 29862 | 1162 | 28700 | 0.01021 | 0.3961 | Diffuse diseases of connective tissue | dermatologic | RATE_C | |
| 709.2 | | | | | 23183 | 50 | 23133 | | | | | 29154 | 454 | 28700 | 0.5423 | 0.2018 | Sicca syndrome | dermatologic | RATE_C | |
| 709.3 | | | | | 23192 | 59 | 23133 | | | | | 28935 | 235 | 28700 | | | Systemic sclerosis | dermatologic | RATE_C | |
| 709.4 | | | | | 23156 | 23 | 23133 | | | | | 28760 | 60 | 28700 | | | Polymyositis | dermatologic | RATE_C | |
| 709.5 | | | | | 23161 | 28 | 23133 | | | | | 28793 | 93 | 28700 | | | Dermatomyositis | dermatologic | RATE_C | |

| phenotype | male_beta | male_SE | male_OR | male_p | male_n_total | male_n_cases | male_n_controls | female_beta | female_SE | female_OR | female_p | female_n_total | female_n_cases | female_n_controls | interaction_p | interaction_beta | description | group | lab | label |
|---|---|---|---|---|---|---|---|---|---|---|---|---|---|---|---|---|---|---|---|---|
| 709.6 | | | | | 23165 | 32 | 23133 | | | | | 28946 | 246 | 28700 | | | Other specified diffuse diseases of connective tissue | dermatologic | RATE_C | |
| 709.7 | | | | | 23162 | 29 | 23133 | | | | | 28907 | 207 | 28700 | | | Unspecified diffuse connective tissue disease | dermatologic | RATE_C | |
| phenotype | male_beta | male_SE | male_OR | male_p | male_n_total | male_n_cases | male_n_controls | female_beta | female_SE | female_OR | female_p | female_n_total | female_n_cases | female_n_controls | interaction_p | interaction_beta | description | group | lab | label |
| 710 | 1.136 | 0.1088 | 3.116 | 1.60e-25 | 23512 | 830 | 22682 | 1.203 | 0.1054 | 3.329 | 3.95e-30 | 30097 | 684 | 29413 | 0.2211 | 0.1754 | Osteomyelitis, periostitis, and other infections involving bone | musculoskeletal | RATE_C | |
| 710.1 | 1.146 | 0.115 | 3.147 | 2.01e-23 | 23427 | 745 | 22682 | 1.263 | 0.1191 | 3.536 | 2.78e-26 | 29958 | 545 | 29413 | 0.186 | 0.2073 | Osteomyelitis | musculoskeletal | RATE_C | |
| 710.11 | | | | | 22957 | 275 | 22682 | | | | | 29563 | 150 | 29413 | | | Acute osteomyelitis | musculoskeletal | RATE_C | |
| 710.12 | | | | | 23048 | 366 | 22682 | | | | | 29661 | 248 | 29413 | 0.1882 | 0.2996 | Chronic osteomyelitis | musculoskeletal | RATE_C | |
| 710.19 | 1.289 | 0.143 | 3.628 | 2.06e-19 | 23198 | 516 | 22682 | | | | | 29784 | 371 | 29413 | 0.3575 | 0.1796 | Unspecified osteomyelitis | musculoskeletal | RATE_C | |
| 710.2 | | | | | 22682 | | | | | | | 29416 | 3 | 29413 | | | Periostitis | musculoskeletal | RATE_C | |
| 710.3 | | | | | 22684 | 2 | 22682 | | | | | 29414 | 1 | 29413 | | | Osteopathy resulting from poliomyelitis | musculoskeletal | RATE_C | |
| 711 | | | | | 22939 | 257 | 22682 | | | | | 29633 | 220 | 29413 | | | Arthropathy associated with infections | musculoskeletal | RATE_C | |
| 711.1 | | | | | 22893 | 211 | 22682 | | | | | 29566 | 153 | 29413 | | | Pyogenic arthritis | musculoskeletal | RATE_C | |
| 711.2 | | | | | 22696 | 14 | 22682 | | | | | 29424 | 11 | 29413 | | | Reiter's disease | musculoskeletal | RATE_C | |
| 711.3 | | | | | 22695 | 13 | 22682 | | | | | 29451 | 38 | 29413 | | | Behcet's syndrome | musculoskeletal | RATE_C | |
| 712 | | | | | 22798 | 116 | 22682 | | | | | 29481 | 68 | 29413 | | | Infective connective tissue disorders | musculoskeletal | RATE_C | |
| 713 | | | | | 22821 | 139 | 22682 | | | | | 29546 | 133 | 29413 | | | Arthropathy associated with other disorders classified elsewhere | musculoskeletal | RATE_C | |
| 713.5 | | | | | 22789 | 107 | 22682 | | | | | 29502 | 89 | 29413 | | | Arthropathy associated with neurological disorders | musculoskeletal | RATE_C | |
| 714 | -0.393 | 0.08644 | 0.675 | 5.45e-06 | 24325 | 732 | 23593 | -0.2355 | 0.05247 | 0.7902 | 7.20e-06 | 31860 | 1880 | 29980 | 0.01988 | 0.2194 | Rheumatoid arthritis and other inflammatory polyarthropathies | musculoskeletal | RATE_C | |
| 714.1 | -0.4027 | 0.1028 | 0.6685 | 8.99e-05 | 24107 | 514 | 23593 | -0.2135 | 0.06032 | 0.8078 | 4.02e-04 | 31387 | 1407 | 29980 | 0.05208 | 0.218 | Rheumatoid arthritis | musculoskeletal | RATE_C | |
| 714.2 | | | | | 23636 | 43 | 23593 | | | | | 30097 | 117 | 29980 | | | Juvenile rheumatoid arthritis | musculoskeletal | RATE_C | |
| 715 | | | | | 23907 | 314 | 23593 | 0.7946 | 0.1071 | 2.214 | 1.20e-13 | 30615 | 635 | 29980 | 0.06176 | 0.3434 | Other inflammatory spondylopathies | musculoskeletal | RATE_C | |
| 715.1 | | | | | 23774 | 181 | 23593 | | | | | 30461 | 481 | 29980 | 0.5888 | 0.1401 | Sacroiliitis NEC | musculoskeletal | RATE_C | |
| 715.2 | | | | | 23683 | 90 | 23593 | | | | | 30051 | 71 | 29980 | | | Ankylosing spondylitis | musculoskeletal | RATE_C | |
| 715.3 | | | | | 23620 | 27 | 23593 | | | | | 30038 | 58 | 29980 | | | Spinal enthesopathy | musculoskeletal | RATE_C | |
| 716 | 1.069 | 0.119 | 2.912 | 2.65e-19 | 23562 | 880 | 22682 | 1.15 | 0.08519 | 3.159 | 1.56e-41 | 30774 | 1361 | 29413 | 0.1495 | 0.2041 | Other arthropathies | musculoskeletal | RATE_C | |
| 716.1 | | | | | 22700 | 18 | 22682 | | | | | 29437 | 24 | 29413 | | | Unspecified polyarthropathy or polyarthritis | musculoskeletal | RATE_C | |
| 716.2 | | | | | 22688 | 6 | 22682 | | | | | 29420 | 7 | 29413 | | | Unspecified monoarthritis | musculoskeletal | RATE_C | |
| 716.3 | | | | | 22683 | 1 | 22682 | | | | | 29414 | 1 | 29413 | | | Kaschin-Beck disease | musculoskeletal | RATE_C | |
| 716.8 | | | | | 22688 | 6 | 22682 | | | | | 29426 | 13 | 29413 | | | Palindromic rheumatism | musculoskeletal | RATE_C | |

| phenotype | male_beta | male_SE | male_OR | male_p | male_n_total | male_n_cases | male_n_controls | female_beta | female_SE | female_OR | female_p | female_n_total | female_n_cases | female_n_controls | interaction_p | interaction_beta | description | group | lab | label |
|---|---|---|---|---|---|---|---|---|---|---|---|---|---|---|---|---|---|---|---|---|
| 716.9 | 1.192 | 0.133 | 3.294 | 3.14e-19 | 23461 | 779 | 22682 | 1.24 | 0.09324 | 3.454 | 2.48e-40 | 30638 | 1225 | 29413 | 0.2616 | 0.1772 | Arthropathy NOS | musculoskeletal | RATE_C | |
| 717 | | | | | 27682 | 166 | 27516 | | | | | 36769 | 263 | 36506 | | | Polymyalgia Rheumatica | musculoskeletal | RATE_C | |
| 720 | 0.9027 | 0.09041 | 2.466 | 1.78e-23 | 22346 | 1436 | 20910 | 1.28 | 0.07955 | 3.597 | 2.94e-58 | 29466 | 1690 | 27776 | 1.71e-04 | 0.4421 | Spinal stenosis | musculoskeletal | RATE_C | |
| 720.1 | 0.9383 | 0.1152 | 2.556 | 3.70e-16 | 21846 | 936 | 20910 | 1.304 | 0.1004 | 3.683 | 1.37e-38 | 28903 | 1127 | 27776 | 0.004126 | 0.43 | Spinal stenosis of lumbar region | musculoskeletal | RATE_C | |
| 721 | 0.9106 | 0.07356 | 2.486 | 3.35e-35 | 23062 | 2152 | 20910 | 1.051 | 0.05611 | 2.861 | 2.50e-78 | 30738 | 2962 | 27776 | 0.0197 | 0.2094 | Spondylosis and allied disorders | musculoskeletal | RATE_C | |
| 721.1 | 0.9312 | 0.07705 | 2.538 | 1.26e-33 | 22908 | 1998 | 20910 | 1.045 | 0.0576 | 2.844 | 1.40e-73 | 30580 | 2804 | 27776 | 0.05428 | 0.1798 | Spondylosis without myelopathy | musculoskeletal | RATE_C | |
| 721.2 | | | | | 21155 | 245 | 20910 | | | | | 28026 | 250 | 27776 | | | Spondylosis with myelopathy | musculoskeletal | RATE_C | |
| 721.8 | | | | | 20961 | 51 | 20910 | | | | | 27797 | 21 | 27776 | | | Other allied disorders of spine | musculoskeletal | RATE_C | |
| 722 | 0.724 | 0.06393 | 2.063 | 9.75e-30 | 23387 | 2477 | 20910 | 0.8497 | 0.04876 | 2.339 | 5.43e-68 | 31200 | 3424 | 27776 | 0.004427 | 0.2199 | Intervertebral disc disorders | musculoskeletal | RATE_C | |
| 722.1 | 0.9195 | 0.1257 | 2.508 | 2.59e-13 | 21491 | 581 | 20910 | 1.05 | 0.09657 | 2.857 | 1.58e-27 | 28557 | 781 | 27776 | 0.173 | 0.2045 | Displacement of intervertebral disc | musculoskeletal | RATE_C | |
| 722.3 | | | | | 20912 | 2 | 20910 | | | | | 27777 | 1 | 27776 | | | Schmorl's nodes | musculoskeletal | RATE_C | |
| 722.6 | 0.69 | 0.07561 | 1.994 | 7.09e-20 | 22720 | 1810 | 20910 | 0.7982 | 0.05529 | 2.221 | 3.07e-47 | 30454 | 2678 | 27776 | 0.0363 | 0.1895 | Degeneration of intervertebral disc | musculoskeletal | RATE_C | |
| 722.7 | | | | | 21018 | 108 | 20910 | | | | | 27881 | 105 | 27776 | | | Intervertebral disc disorder with myelopathy | musculoskeletal | RATE_C | |
| 722.8 | | | | | 21238 | 328 | 20910 | | | | | 28155 | 379 | 27776 | 0.2269 | 0.2961 | Postlaminectomy syndrome | musculoskeletal | RATE_C | |
| 722.9 | | | | | 21012 | 102 | 20910 | | | | | 27883 | 107 | 27776 | | | Other and unspecified disc disorder | musculoskeletal | RATE_C | |
| 723 | | | | | 21252 | 342 | 20910 | | | | | 28186 | 410 | 27776 | 0.0698 | 0.3503 | Other disorders of cervical region | musculoskeletal | RATE_C | |
| 723.1 | | | | | 20943 | 33 | 20910 | | | | | 27812 | 36 | 27776 | | | Torticollis | musculoskeletal | RATE_C | |
| 724 | | | | | 21393 | 483 | 20910 | 0.7009 | 0.08841 | 2.016 | 2.23e-15 | 28648 | 872 | 27776 | 0.2193 | 0.1854 | Other and unspecified disorders of back | musculoskeletal | RATE_C | |
| 724.1 | | | | | 21038 | 128 | 20910 | | | | | 28127 | 351 | 27776 | | | Disorders of sacrum | musculoskeletal | RATE_C | |
| 724.2 | | | | | 21034 | 124 | 20910 | | | | | 28108 | 332 | 27776 | | | Disorders of coccyx | musculoskeletal | RATE_C | |
| 724.8 | | | | | 21013 | 103 | 20910 | | | | | 27951 | 175 | 27776 | | | Other symptoms referable to back | musculoskeletal | RATE_C | |
| 724.9 | | | | | 20981 | 71 | 20910 | | | | | 27892 | 116 | 27776 | | | Other unspecified back disorders | musculoskeletal | RATE_C | |
| 726 | 0.3451 | 0.05708 | 1.412 | 1.49e-09 | 23098 | 2817 | 20281 | 0.4992 | 0.04152 | 1.647 | 2.70e-33 | 30370 | 4595 | 25775 | 8.00e-05 | 0.2613 | Peripheral enthesopathies and allied syndromes | musculoskeletal | RATE_C | |
| 726.1 | 0.3758 | 0.08691 | 1.456 | 1.53e-05 | 21421 | 1140 | 20281 | 0.5049 | 0.05169 | 1.657 | 1.54e-22 | 28550 | 2775 | 25775 | 0.04992 | 0.1874 | Enthesopathy | musculoskeletal | RATE_C | |
| 726.2 | | | | | 20365 | 84 | 20281 | | | | | 25894 | 119 | 25775 | | | Synoviopathy | musculoskeletal | RATE_C | |
| 726.3 | | | | | 20728 | 447 | 20281 | | | | | 26194 | 419 | 25775 | 0.04765 | 0.3416 | Bursitis | musculoskeletal | RATE_C | |
| 726.4 | | | | | 20361 | 80 | 20281 | | | | | 25949 | 174 | 25775 | | | Calcaneal spur; Exostosis NOS | musculoskeletal | RATE_C | |
| 727 | 0.4641 | 0.07259 | 1.591 | 1.63e-10 | 21945 | 1664 | 20281 | 0.6218 | 0.05134 | 1.862 | 9.07e-34 | 28564 | 2789 | 25775 | 0.002264 | 0.2542 | Other disorders of synovium, tendon, and bursa | musculoskeletal | RATE_C | |
| 727.1 | 0.4608 | 0.1066 | 1.585 | 1.53e-05 | 21009 | 728 | 20281 | 0.6444 | 0.06872 | 1.905 | 6.79e-21 | 27213 | 1438 | 25775 | 0.05548 | 0.2283 | Synovitis and tenosynovitis | musculoskeletal | RATE_C | |

| phenotype | male_beta | male_SE | male_OR | male_p | male_n_total | male_n_cases | male_n_controls | female_beta | female_SE | female_OR | female_p | female_n_total | female_n_cases | female_n_controls | interaction_p | interaction_beta | description | group | lab | label |
|---|---|---|---|---|---|---|---|---|---|---|---|---|---|---|---|---|---|---|---|---|
| 727.2 | | | | | 20360 | 79 | 20281 | | | | | 25927 | 152 | 25775 | | | Bursitis disorders | musculoskeletal | RATE_C | |
| 727.4 | | | | | 20471 | 190 | 20281 | | | | | 26261 | 486 | 25775 | 0.02803 | 0.4468 | Ganglion and cyst of synovium, tendon, and bursa | musculoskeletal | RATE_C | |
| 727.5 | | | | | 20324 | 43 | 20281 | | | | | 25904 | 129 | 25775 | | | Rupture of synovium | musculoskeletal | RATE_C | |
| 727.6 | | | | | 20660 | 379 | 20281 | | | | | 26104 | 329 | 25775 | 0.5663 | 0.1258 | Rupture of tendon, nontraumatic | musculoskeletal | RATE_C | |
| 727.7 | | | | | 20347 | 66 | 20281 | | | | | 25864 | 89 | 25775 | | | Contracture of tendon (sheath) | musculoskeletal | RATE_C | |
| phenotype | male_beta | male_SE | male_OR | male_p | male_n_total | male_n_cases | male_n_controls | female_beta | female_SE | female_OR | female_p | female_n_total | female_n_cases | female_n_controls | interaction_p | interaction_beta | description | group | lab | label |
| 727.8 | | | | | 20310 | 29 | 20281 | | | | | 25829 | 54 | 25775 | | | Plica syndrome | musculoskeletal | RATE_C | |
| 728 | | | | | 20364 | 83 | 20281 | | | | | 25899 | 124 | 25775 | | | Disorders of muscle, ligament, and fascia | musculoskeletal | RATE_C | |
| 728.1 | | | | | 20313 | 32 | 20281 | | | | | 25798 | 23 | 25775 | | | Muscular calcification and ossification | musculoskeletal | RATE_C | |
| 728.2 | | | | | 20304 | 23 | 20281 | | | | | 25866 | 91 | 25775 | | | Laxity of ligament or hypermobility syndrome | musculoskeletal | RATE_C | |
| 728.7 | | | | | 20721 | 440 | 20281 | 0.504 | 0.09145 | 1.655 | 3.57e-08 | 26509 | 734 | 25775 | 0.05189 | 0.2879 | Fasciitis | musculoskeletal | RATE_C | |
| 728.71 | | | | | 20411 | 130 | 20281 | | | | | 25848 | 73 | 25775 | | | Contracture of palmar fascia [Dupuytren's disease] | musculoskeletal | RATE_C | |
| 729 | 1.061 | 0.1278 | 2.89 | 9.96e-17 | 20964 | 683 | 20281 | 1.036 | 0.09112 | 2.818 | 5.99e-30 | 26711 | 936 | 25775 | 0.6074 | 0.07656 | Other disorders of soft tissues | musculoskeletal | RATE_C | |
| 729.1 | | | | | 20284 | 3 | 20281 | | | | | 25782 | 7 | 25775 | | | Rheumatism, unspecified and fibrositis | musculoskeletal | RATE_C | |
| 729.3 | | | | | 20292 | 11 | 20281 | | | | | 25825 | 50 | 25775 | | | Panniculitis | musculoskeletal | RATE_C | |
| 729.7 | | | | | 20307 | 26 | 20281 | | | | | 25785 | 10 | 25775 | | | Nontraumatic compartment syndrome | musculoskeletal | RATE_C | |
| 731 | | | | | 22403 | 94 | 22309 | | | | | 27658 | 148 | 27510 | | | Osteitis deformans and osteopathies associated with other disorders classified elsewhere | musculoskeletal | RATE_C | |
| 731.1 | | | | | 22320 | 11 | 22309 | | | | | 27517 | 7 | 27510 | | | Osteitis deformans [Paget's disease of bone] | musculoskeletal | RATE_C | |
| 732 | | | | | 22384 | 75 | 22309 | | | | | 27583 | 73 | 27510 | | | Osteochondropathies | musculoskeletal | RATE_C | |
| 732.1 | | | | | 22341 | 32 | 22309 | | | | | 27549 | 39 | 27510 | | | Juvenile osteochondrosis | musculoskeletal | RATE_C | |
| 732.7 | | | | | 22348 | 39 | 22309 | | | | | 27539 | 29 | 27510 | | | Osteochondritis dissecans | musculoskeletal | RATE_C | |
| 733 | 0.3941 | 0.1087 | 1.483 | 2.90e-04 | 22918 | 609 | 22309 | 0.3552 | 0.05548 | 1.426 | 1.52e-10 | 29872 | 2362 | 27510 | 1.27e-06 | 0.5523 | Other disorders of bone and cartilage | musculoskeletal | RATE_C | |
| 733.2 | | | | | 22336 | 27 | 22309 | | | | | 27554 | 44 | 27510 | | | Cyst of bone | musculoskeletal | RATE_C | |
| 733.4 | | | | | 22516 | 207 | 22309 | | | | | 27762 | 252 | 27510 | | | Aseptic necrosis of bone | musculoskeletal | RATE_C | |
| 733.6 | | | | | 22334 | 25 | 22309 | | | | | 27574 | 64 | 27510 | | | Costochondritis | musculoskeletal | RATE_C | |
| 733.8 | | | | | 22558 | 249 | 22309 | | | | | 27858 | 348 | 27510 | 1.90e-08 | 1.188 | Malunion and nonunion of fracture | musculoskeletal | RATE_C | |
| 733.9 | | | | | 22350 | 41 | 22309 | | | | | 27565 | 55 | 27510 | | | Chondromalacia | musculoskeletal | RATE_C | |
| 735 | 0.4401 | 0.09958 | 1.553 | 9.90e-06 | 24919 | 827 | 24092 | 0.6079 | 0.07076 | 1.837 | 8.64e-18 | 32235 | 1405 | 30830 | 0.002536 | 0.3485 | Acquired foot deformities | musculoskeletal | RATE_C | |

| phenotype | male_beta | male_SE | male_OR | male_p | male_n_total | male_n_cases | male_n_controls | female_beta | female_SE | female_OR | female_p | female_n_total | female_n_cases | female_n_controls | interaction_p | interaction_beta | description | group | lab | label |
|---|---|---|---|---|---|---|---|---|---|---|---|---|---|---|---|---|---|---|---|---|
| 735.1 | | | | | 24190 | 98 | 24092 | | | | | 30970 | 140 | 30830 | | | Flat foot | musculoskeletal | RATE_C | |
| 735.2 | | | | | 24498 | 406 | 24092 | 0.8006 | 0.1087 | 2.227 | 1.72e-13 | 31534 | 704 | 30830 | 0.5531 | 0.115 | Acquired toe deformities | musculoskeletal | RATE_C | |
| 735.21 | | | | | 24374 | 282 | 24092 | 0.9752 | 0.1356 | 2.652 | 6.35e-13 | 31365 | 535 | 30830 | 0.1033 | 0.3836 | Hammer toe (acquired) | musculoskeletal | RATE_C | |
| 735.22 | | | | | 24107 | 15 | 24092 | | | | | 30844 | 14 | 30830 | | | Claw toe (acquired) | musculoskeletal | RATE_C | |
| 735.23 | | | | | 24184 | 92 | 24092 | | | | | 30968 | 138 | 30830 | | | Hallux rigidus | musculoskeletal | RATE_C | |
| 735.3 | | | | | 24207 | 115 | 24092 | 0.573 | 0.1165 | 1.774 | 8.78e-07 | 31337 | 507 | 30830 | 0.4974 | -0.2209 | Hallux valgus (Bunion) | musculoskeletal | RATE_C | |
| 736 | | | | | 24293 | 201 | 24092 | | | | | 31078 | 248 | 30830 | | | Other acquired deformities of limbs | musculoskeletal | RATE_C | |
| 736.1 | | | | | 24103 | 11 | 24092 | | | | | 30840 | 10 | 30830 | | | Acquired deformities of forearm | musculoskeletal | RATE_C | |
| 736.2 | | | | | 24158 | 66 | 24092 | | | | | 30902 | 72 | 30830 | | | Acquired deformities of finger | musculoskeletal | RATE_C | |
| 736.3 | | | | | 24100 | 8 | 24092 | | | | | 30841 | 11 | 30830 | | | Acquired deformities of hip | musculoskeletal | RATE_C | |
| 736.4 | | | | | 24104 | 12 | 24092 | | | | | 30857 | 27 | 30830 | | | Genu valgum or varum (acquired) | musculoskeletal | RATE_C | |
| 736.5 | | | | | 24095 | 3 | 24092 | | | | | 30840 | 10 | 30830 | | | Acquired deformities of knee | musculoskeletal | RATE_C | |
| 736.6 | | | | | 24114 | 22 | 24092 | | | | | 30876 | 46 | 30830 | | | Unequal leg length (acquired) | musculoskeletal | RATE_C | |
| 737 | | | | | 24514 | 422 | 24092 | 0.6543 | 0.08004 | 1.924 | 2.94e-16 | 31760 | 930 | 30830 | 1.14e-11 | 0.8666 | Curvature of spine | musculoskeletal | RATE_C | |
| 737.1 | | | | | 24226 | 134 | 24092 | | | | | 31005 | 175 | 30830 | | | Kyphosis (acquired) | musculoskeletal | RATE_C | |
| 737.2 | | | | | 24093 | 1 | 24092 | | | | | 30832 | 2 | 30830 | | | Lordosis (acquired) | musculoskeletal | RATE_C | |
| 737.3 | | | | | 24413 | 321 | 24092 | 0.6032 | 0.08421 | 1.828 | 7.91e-13 | 31641 | 811 | 30830 | 9.89e-13 | 0.9944 | Kyphoscoliosis and scoliosis | musculoskeletal | RATE_C | |
| 738 | 0.6546 | 0.1261 | 1.924 | 2.11e-07 | 24637 | 545 | 24092 | 0.9715 | 0.09338 | 2.642 | 2.40e-25 | 31798 | 968 | 30830 | 8.06e-05 | 0.5882 | Other acquired musculoskeletal deformity | musculoskeletal | RATE_C | |
| 738.4 | | | | | 24467 | 375 | 24092 | 0.9833 | 0.1092 | 2.673 | 2.14e-19 | 31608 | 778 | 30830 | 0.0525 | 0.378 | Acquired spondylolisthesis | musculoskeletal | RATE_C | |
| 739 | | | | | 24307 | 215 | 24092 | | | | | 31047 | 217 | 30830 | | | Contracture of joint | musculoskeletal | RATE_C | |
| 740 | 0.5456 | 0.04912 | 1.726 | 1.16e-28 | 25263 | 4641 | 20622 | 0.4801 | 0.03538 | 1.616 | 6.06e-42 | 33672 | 7527 | 26145 | 0.1227 | 0.09114 | Osteoarthrosis | musculoskeletal | RATE_C | |
| 740.1 | 0.5418 | 0.0601 | 1.719 | 1.98e-19 | 23687 | 3065 | 20622 | 0.5822 | 0.04403 | 1.79 | 6.57e-40 | 30839 | 4694 | 26145 | 0.009762 | 0.1875 | Osteoarthritis; localized | musculoskeletal | RATE_C | |
| 740.11 | 0.5293 | 0.06265 | 1.698 | 2.92e-17 | 23439 | 2817 | 20622 | 0.569 | 0.04546 | 1.767 | 6.02e-36 | 30486 | 4341 | 26145 | 0.02247 | 0.1721 | Osteoarthrosis, localized, primary | musculoskeletal | RATE_C | |
| 740.12 | | | | | 20717 | 95 | 20622 | | | | | 26285 | 140 | 26145 | | | Osteoarthrosis, localized, secondary | musculoskeletal | RATE_C | |
| 740.2 | -0.09457 | 0.1052 | 0.9098 | 0.3688 | 21264 | 642 | 20622 | 0.0381 | 0.05851 | 1.039 | 0.5149 | 28076 | 1931 | 26145 | 0.06564 | 0.215 | Osteoarthrosis, generalized | musculoskeletal | RATE_C | |
| 740.3 | | | | | 20669 | 47 | 20622 | | | | | 26282 | 137 | 26145 | | | Osteoarthrosis involving more than one site, but not specified as generalized | musculoskeletal | RATE_C | |
| 740.9 | 1.015 | 0.07507 | 2.76 | 1.09e-41 | 23067 | 2445 | 20622 | 0.9348 | 0.04993 | 2.547 | 3.41e-78 | 30322 | 4177 | 26145 | 0.4678 | 0.06446 | Osteoarthrosis NOS | musculoskeletal | RATE_C | |
| 741 | 0.5646 | 0.06137 | 1.759 | 3.57e-20 | 24701 | 2405 | 22296 | 0.653 | 0.04547 | 1.921 | 8.98e-47 | 32920 | 3637 | 29283 | 0.00163 | 0.2267 | Symptoms and disorders of the joints | musculoskeletal | RATE_C | |
| 741.1 | | | | | 22321 | 25 | 22296 | | | | | 29334 | 51 | 29283 | | | Ankylosis of joint | musculoskeletal | RATE_C | |

| phenotype | male_beta | male_SE | male_OR | male_p | male_n_total | male_n_cases | male_n_controls | female_beta | female_SE | female_OR | female_p | female_n_total | female_n_cases | female_n_controls | interaction_p | interaction_beta | description | group | lab | label |
|---|---|---|---|---|---|---|---|---|---|---|---|---|---|---|---|---|---|---|---|---|
| 741.2 | 0.4901 | 0.1128 | 1.632 | 1.40e-05 | 22901 | 605 | 22296 | 0.4328 | 0.07068 | 1.542 | 9.21e-10 | 30510 | 1227 | 29283 | 0.3877 | 0.1076 | Stiffness of joint | musculoskeletal | RATE_C | |
| 741.3 | 0.7068 | 0.1276 | 2.028 | 3.03e-08 | 22942 | 646 | 22296 | 1.142 | 0.1052 | 3.133 | 1.82e-27 | 30187 | 904 | 29283 | 0.002313 | 0.489 | Difficulty in walking | musculoskeletal | RATE_C | |
| 741.4 | 0.5026 | 0.1074 | 1.653 | 2.90e-06 | 22964 | 668 | 22296 | 0.609 | 0.08183 | 1.839 | 9.89e-14 | 30241 | 958 | 29283 | 0.0539 | 0.2444 | Joint effusions | musculoskeletal | RATE_C | |
| 741.5 | | | | | 22328 | 32 | 22296 | | | | | 29301 | 18 | 29283 | | | Hemarthrosis | musculoskeletal | RATE_C | |
| 741.6 | | | | | 22307 | 11 | 22296 | | | | | 29300 | 17 | 29283 | | | Villonodular synovitis | musculoskeletal | RATE_C | |
| 742 | | | | | 22735 | 439 | 22296 | 0.6766 | 0.09577 | 1.967 | 1.60e-12 | 29934 | 651 | 29283 | 0.00101 | 0.4668 | Derangement of joint, non-traumatic | musculoskeletal | RATE_C | |
| 742.1 | | | | | 22319 | 23 | 22296 | | | | | 29301 | 18 | 29283 | | | Loose body in joint | musculoskeletal | RATE_C | |
| 742.2 | | | | | 22327 | 31 | 22296 | | | | | 29340 | 57 | 29283 | | | Pathological, developmental or recurrent dislocation | musculoskeletal | RATE_C | |
| 742.8 | | | | | 22322 | 26 | 22296 | | | | | 29329 | 46 | 29283 | | | Articular cartilage disorder | musculoskeletal | RATE_C | |
| 742.9 | | | | | 22643 | 347 | 22296 | 0.7061 | 0.1078 | 2.026 | 5.65e-11 | 29808 | 525 | 29283 | 0.006615 | 0.4404 | Other derangement of joint | musculoskeletal | RATE_C | |
| 743 | 0.8982 | 0.07076 | 2.455 | 6.42e-37 | 26032 | 1946 | 24086 | 0.566 | 0.03777 | 1.761 | 9.29e-51 | 33952 | 6028 | 27924 | 0.004062 | 0.2213 | Osteoporosis, osteopenia and pathological fracture | musculoskeletal | RATE_C | |
| 743.1 | 0.9165 | 0.1003 | 2.501 | 6.55e-20 | 25110 | 1024 | 24086 | 0.5102 | 0.04595 | 1.666 | 1.22e-28 | 31886 | 3962 | 27924 | 0.1203 | 0.1648 | Osteoporosis | musculoskeletal | RATE_C | |
| 743.11 | 0.8984 | 0.1177 | 2.456 | 2.33e-14 | 24850 | 764 | 24086 | 0.5902 | 0.0517 | 1.804 | 3.46e-30 | 31204 | 3280 | 27924 | 0.06861 | 0.2262 | Osteoporosis NOS | musculoskeletal | RATE_C | |
| 743.12 | | | | | 24196 | 110 | 24086 | 0.1191 | 0.08762 | 1.126 | 0.174 | 28758 | 834 | 27924 | 0.07086 | 0.4599 | Senile osteoporosis | musculoskeletal | RATE_C | |
| 743.13 | | | | | 24546 | 460 | 24086 | 0.6796 | 0.08197 | 1.973 | 1.13e-16 | 29013 | 1089 | 27924 | 0.0186 | 0.3672 | Other specified osteoporosis | musculoskeletal | RATE_C | |
| 743.2 | | | | | 24445 | 359 | 24086 | 1.034 | 0.1109 | 2.812 | 1.19e-20 | 28551 | 627 | 27924 | 0.1274 | 0.2935 | Pathologic fracture | musculoskeletal | RATE_C | |
| 743.21 | | | | | 24187 | 101 | 24086 | | | | | 28089 | 165 | 27924 | | | Pathologic fracture of vertebrae | musculoskeletal | RATE_C | |
| 743.22 | | | | | 24160 | 74 | 24086 | | | | | 28032 | 108 | 27924 | | | Pathologic fracture of femur | musculoskeletal | RATE_C | |
| 743.4 | | | | | 24126 | 40 | 24086 | | | | | 28053 | 129 | 27924 | | | Stress fracture | musculoskeletal | RATE_C | |
| 743.9 | 0.9278 | 0.1139 | 2.529 | 3.68e-16 | 24819 | 733 | 24086 | 0.6442 | 0.05639 | 1.904 | 3.15e-30 | 30516 | 2592 | 27924 | 0.00509 | 0.3413 | Osteopenia or other disorder of bone and cartilage | musculoskeletal | RATE_C | |
| 745 | 0.4512 | 0.03864 | 1.57 | 1.67e-31 | 24606 | 7378 | 17228 | 0.5267 | 0.02912 | 1.693 | 4.10e-73 | 32439 | 12064 | 20375 | 5.17e-08 | 0.2424 | Pain in joint | musculoskeletal | RATE_C | |
| 747 | 1.435 | 0.09331 | 4.2 | 2.18e-53 | 26877 | 1323 | 25554 | 1.605 | 0.08518 | 4.98 | 3.18e-79 | 36150 | 1183 | 34967 | 0.7923 | 0.03113 | Cardiac and circulatory congenital anomalies | congenital anomalies | RATE_C | |
| 747.1 | 1.493 | 0.09966 | 4.449 | 1.01e-50 | 26767 | 1213 | 25554 | 1.781 | 0.09536 | 5.938 | 7.29e-78 | 36008 | 1041 | 34967 | 0.3552 | 0.1199 | Cardiac congenital anomalies | congenital anomalies | RATE_C | |
| 747.11 | | | | | 25997 | 443 | 25554 | 1.97 | 0.1292 | 7.168 | 1.86e-52 | 35539 | 572 | 34967 | 0.03017 | 0.3993 | Cardiac shunt/ heart septal defect | congenital anomalies | RATE_C | |
| 747.12 | | | | | 25876 | 322 | 25554 | | | | | 35219 | 252 | 34967 | 0.4055 | -0.18 | Valvular heart disease/ heart chambers | congenital anomalies | RATE_C | |
| 747.13 | | | | | 25697 | 143 | 25554 | | | | | 35131 | 164 | 34967 | | | Congenital anomalies of great vessels | congenital anomalies | RATE_C | |
| 747.2 | | | | | 25638 | 84 | 25554 | | | | | 35084 | 117 | 34967 | | | Congenital anomalies of peripheral vascular system | congenital anomalies | RATE_C | |
| 748 | | | | | 27528 | 39 | 27489 | | | | | 36621 | 44 | 36577 | | | Anomalies of respiratory system, congenital | congenital anomalies | RATE_C | |

| phenotype | male_beta | male_SE | male_OR | male_p | male_n_total | male_n_cases | male_n_controls | female_beta | female_SE | female_OR | female_p | female_n_total | female_n_cases | female_n_controls | interaction_p | interaction_beta | description | group | lab | label |
|---|---|---|---|---|---|---|---|---|---|---|---|---|---|---|---|---|---|---|---|---|
| 749 | | | | | 27561 | 72 | 27489 | | | | | 36671 | 94 | 36577 | | | Congenital anomalies of face and neck | congenital anomalies | RATE_C | |
| 749.1 | | | | | 22492 | 16 | 22476 | | | | | 31338 | 20 | 31318 | | | Cleft palate | congenital anomalies | RATE_C | |
| 749.2 | | | | | 27520 | 31 | 27489 | | | | | 36613 | 36 | 36577 | | | Congenital anomalies of skull and face bones | congenital anomalies | RATE_C | |
| 750 | | | | | 26052 | 143 | 25909 | | | | | 35186 | 190 | 34996 | | | Digestive congenital anomalies | congenital anomalies | RATE_C | |
| 750.1 | | | | | 25951 | 42 | 25909 | | | | | 35050 | 54 | 34996 | | | Upper gastrointestinal congenital anomalies | congenital anomalies | RATE_C | |
| 750.11 | | | | | 25920 | 11 | 25909 | | | | | 35005 | 9 | 34996 | | | Esophageal atresia/tracheoesophageal fistula | congenital anomalies | RATE_C | |
| 750.13 | | | | | 25928 | 19 | 25909 | | | | | 35009 | 13 | 34996 | | | Congenital anomalies of mouth/tongue | congenital anomalies | RATE_C | |
| 750.14 | | | | | 25914 | 5 | 25909 | | | | | 35012 | 16 | 34996 | | | Congenital anomalies of esophagus | congenital anomalies | RATE_C | |
| 750.15 | | | | | 25911 | 2 | 25909 | | | | | 34997 | 1 | 34996 | | | Congenital anomalies of stomach | congenital anomalies | RATE_C | |
| 750.2 | | | | | 26004 | 95 | 25909 | | | | | 35124 | 128 | 34996 | | | Lower gastrointestinal congenital anomalies | congenital anomalies | RATE_C | |
| 750.21 | | | | | 25974 | 65 | 25909 | | | | | 35061 | 65 | 34996 | | | Congenital anomalies of intestine | congenital anomalies | RATE_C | |
| 750.22 | | | | | 25936 | 27 | 25909 | | | | | 35055 | 59 | 34996 | | | Congenital anomaly of gallbladder, bile ducts, liver, pancreas | congenital anomalies | RATE_C | |
| 750.5 | | | | | 25913 | 4 | 25909 | | | | | 34999 | 3 | 34996 | | | Congenital hypertrophic pyloric stenosis | congenital anomalies | RATE_C | |
| 751 | | | | | 26398 | 489 | 25909 | | | | | 35443 | 447 | 34996 | 0.7545 | 0.04602 | Genitourinary congenital anomalies | congenital anomalies | RATE_C | |
| 751.1 | | | | | 26010 | 101 | 25909 | | | | | 35087 | 91 | 34996 | | | Congenital anomalies of genital organs | congenital anomalies | RATE_C | |
| 751.11 | | | | | 7 | | | | | | | 35071 | 79 | 34992 | | | Congenital anomalies of female genital organs | congenital anomalies | RATE_C | |
| 751.12 | | | | | 25992 | 90 | 25902 | | | | | 4 | | | | | Congenital anomalies of male genital organs | congenital anomalies | RATE_C | |
| 751.2 | | | | | 26267 | 358 | 25909 | | | | | 35304 | 308 | 34996 | 0.3321 | -0.1819 | Congenital anomalies of urinary system | congenital anomalies | RATE_C | |
| 751.21 | | | | | 26140 | 231 | 25909 | | | | | 35199 | 203 | 34996 | | | Cystic kidney disease | congenital anomalies | RATE_C | |
| 751.22 | | | | | 26007 | 98 | 25909 | | | | | 35074 | 78 | 34996 | | | Other specified congenital anomalies of kidney | congenital anomalies | RATE_C | |
| 751.3 | | | | | 25949 | 40 | 25909 | | | | | 35047 | 51 | 34996 | | | Obstructive genitourinary defect | congenital anomalies | RATE_C | |
| 752 | | | | | 27335 | 160 | 27175 | | | | | 36299 | 223 | 36076 | | | Nervous system congenital anomalies | congenital anomalies | RATE_C | |
| 752.1 | | | | | 27234 | 59 | 27175 | | | | | 36188 | 112 | 36076 | | | Neural tube defects | congenital anomalies | RATE_C | |
| 752.11 | | | | | 27225 | 50 | 27175 | | | | | 36167 | 91 | 36076 | | | Spina bifida | congenital anomalies | RATE_C | |
| 752.2 | | | | | 27279 | 104 | 27175 | | | | | 36200 | 124 | 36076 | | | Other specified congenital anomalies of nervous system | congenital anomalies | RATE_C | |
| 753 | | | | | 27198 | 23 | 27175 | | | | | 36127 | 51 | 36076 | | | Congenital anomalies of the eye | congenital anomalies | RATE_C | |
| 753.1 | | | | | 27183 | 8 | 27175 | | | | | 36087 | 11 | 36076 | | | Congenital cataract and lens anomalies | congenital anomalies | RATE_C | |
| 753.2 | | | | | 27181 | 6 | 27175 | | | | | 36095 | 19 | 36076 | | | Congenital anomalies of posterior segment of eye | congenital anomalies | RATE_C | |
| 754 | | | | | 27083 | 58 | 27025 | | | | | 35886 | 111 | 35775 | | | Congenital musculoskeletal deformities of spine | congenital anomalies | RATE_C | |

| phenotype | male_beta | male_SE | male_OR | male_p | male_n_total | male_n_cases | male_n_controls | female_beta | female_SE | female_OR | female_p | female_n_total | female_n_cases | female_n_controls | interaction_p | interaction_beta | description | group | lab | label |
|---|---|---|---|---|---|---|---|---|---|---|---|---|---|---|---|---|---|---|---|---|
| 754.1 | | | | | 27033 | 8 | 27025 | | | | | 35793 | 18 | 35775 | | | Lumbosacral spondylolysis, congenital | congenital anomalies | RATE_C | |
| 754.2 | | | | | 27059 | 34 | 27025 | | | | | 35837 | 62 | 35775 | | | Spondylolisthesis, congenital | congenital anomalies | RATE_C | |
| 755 | | | | | 27119 | 94 | 27025 | | | | | 35920 | 145 | 35775 | | | Congenital anomalies of limbs | congenital anomalies | RATE_C | |
| 755.1 | | | | | 27058 | 33 | 27025 | | | | | 35834 | 59 | 35775 | | | Congenital deformities of feet | congenital anomalies | RATE_C | |
| 755.3 | | | | | 27031 | 6 | 27025 | | | | | 35781 | 6 | 35775 | | | Congenital anomaly of fingers/toes | congenital anomalies | RATE_C | |
| 755.4 | | | | | 27034 | 9 | 27025 | | | | | 35795 | 20 | 35775 | | | Congenital anomalies of upper limb, including shoulder girdle | congenital anomalies | RATE_C | |
| 755.6 | | | | | 27065 | 40 | 27025 | | | | | 35837 | 62 | 35775 | | | Other congenital anomalies of lower limb, including pelvic girdle | congenital anomalies | RATE_C | |
| 755.61 | | | | | 27060 | 35 | 27025 | | | | | 35828 | 53 | 35775 | | | Congenital hip dysplasia and deformity | congenital anomalies | RATE_C | |
| 756 | | | | | 27125 | 100 | 27025 | | | | | 35939 | 164 | 35775 | | | Other congenital musculoskeletal anomalies | congenital anomalies | RATE_C | |
| 756.1 | | | | | 27036 | 11 | 27025 | | | | | 35791 | 16 | 35775 | | | Congenital anomalies of abdominal wall; diaphram | congenital anomalies | RATE_C | |
| 756.2 | | | | | 27064 | 39 | 27025 | | | | | 35801 | 26 | 35775 | | | Pectus and other congenital anomalies of ribs/sternum | congenital anomalies | RATE_C | |
| 756.21 | | | | | 27054 | 29 | 27025 | | | | | 35798 | 23 | 35775 | | | Pectus excavatum | congenital anomalies | RATE_C | |
| phenotype | male_beta | male_SE | male_OR | male_p | male_n_total | male_n_cases | male_n_controls | female_beta | female_SE | female_OR | female_p | female_n_total | female_n_cases | female_n_controls | interaction_p | interaction_beta | description | group | lab | label |
| 756.22 | | | | | 27036 | 11 | 27025 | | | | | 35776 | 1 | 35775 | | | Pectus carinatum | congenital anomalies | RATE_C | |
| 756.3 | | | | | 27041 | 16 | 27025 | | | | | 35840 | 65 | 35775 | | | Congenital anomalies of muscle, tendon, fascia, and connective tissue | congenital anomalies | RATE_C | |
| 756.5 | | | | | 27043 | 18 | 27025 | | | | | 35804 | 29 | 35775 | | | Congenital osteodystrophies | congenital anomalies | RATE_C | |
| 757 | | | | | 26746 | 13 | 26733 | | | | | 35965 | 21 | 35944 | | | Congenital anomalies of the integument | congenital anomalies | RATE_C | |
| 758 | | | | | 26835 | 102 | 26733 | | | | | 36032 | 88 | 35944 | | | Chromosomal anomalies and genetic disorders | congenital anomalies | RATE_C | |
| 758.1 | | | | | 26801 | 68 | 26733 | | | | | 36013 | 69 | 35944 | | | Chromosomal anomalies | congenital anomalies | RATE_C | |
| 759 | | | | | 27017 | 284 | 26733 | | | | | 36141 | 197 | 35944 | | | Other and unspecified congenital anomalies | congenital anomalies | RATE_C | |
| 759.1 | | | | | 26741 | 8 | 26733 | | | | | 35957 | 13 | 35944 | | | Anomalies of endocrine glands, congenital | congenital anomalies | RATE_C | |
| 760 | 0.7315 | 0.04591 | 2.078 | 3.66e-57 | 24911 | 4715 | 20196 | 0.8008 | 0.03295 | 2.227 | 1.73e-130 | 32596 | 7782 | 24814 | 0.007963 | 0.1402 | Back pain | symptoms | RATE_C | |
| 761 | 0.5765 | 0.06626 | 1.78 | 3.32e-18 | 26084 | 1972 | 24112 | 0.66 | 0.04409 | 1.935 | 1.15e-50 | 34098 | 3706 | 30392 | 0.05463 | 0.142 | Cervicalgia | symptoms | RATE_C | |
| 763 | 0.6213 | 0.08904 | 1.861 | 2.99e-12 | 25089 | 1208 | 23881 | 0.7096 | 0.06366 | 2.033 | 7.47e-29 | 32717 | 1862 | 30855 | 0.05809 | 0.1968 | Thoracic or lumbosacral neuritis or radiculitis, unspecified | symptoms | RATE_C | |
| 764 | 0.3446 | 0.1071 | 1.411 | 0.0013 | 24586 | 705 | 23881 | 0.6116 | 0.07329 | 1.843 | 7.10e-17 | 32170 | 1315 | 30855 | 0.003593 | 0.3566 | Sciatica | symptoms | RATE_C | |
| 765 | 0.5007 | 0.1159 | 1.65 | 1.55e-05 | 24494 | 613 | 23881 | 0.8347 | 0.0904 | 2.304 | 2.60e-20 | 31724 | 869 | 30855 | 0.008389 | 0.3628 | Cervical radiculitis | symptoms | RATE_C | |
| 766 | 0.7403 | 0.1165 | 2.097 | 2.09e-10 | 24563 | 682 | 23881 | 1.082 | 0.08873 | 2.951 | 3.21e-34 | 31922 | 1067 | 30855 | 0.001287 | 0.4463 | Neuralgia, neuritis, and radiculitis NOS | symptoms | RATE_C | |
| 767 | | | | | 23904 | 23 | 23881 | | | | | 30882 | 27 | 30855 | | | Cervicocranial/Cervicobrachial syndrome | symptoms | RATE_C | |

| phenotype | male_beta | male_SE | male_OR | male_p | male_n_total | male_n_cases | male_n_controls | female_beta | female_SE | female_OR | female_p | female_n_total | female_n_cases | female_n_controls | interaction_p | interaction_beta | description | group | lab | label |
|---|---|---|---|---|---|---|---|---|---|---|---|---|---|---|---|---|---|---|---|---|
| 769 | | | | | 27744 | 17 | 27727 | | | | | 36864 | 59 | 36805 | | | Nonallopathic lesions NEC | symptoms | RATE_C | |
| 770 | 0.4934 | 0.1066 | 1.638 | 3.68e-06 | 26643 | 686 | 25957 | 0.3853 | 0.04525 | 1.47 | 1.65e-17 | 34609 | 3011 | 31598 | 0.6337 | -0.05096 | Myalgia and myositis unspecified | symptoms | RATE_C | |
| 771 | 0.9366 | 0.133 | 2.551 | 1.87e-12 | 23704 | 575 | 23129 | 1.125 | 0.1132 | 3.079 | 2.99e-23 | 31912 | 628 | 31284 | 0.2822 | 0.1796 | Musculoskeletal symptoms referable to limbs | symptoms | RATE_C | |
| 771.1 | 1.463 | 0.1022 | 4.317 | 1.94e-46 | 24349 | 1220 | 23129 | 1.462 | 0.08297 | 4.316 | 1.61e-69 | 32708 | 1424 | 31284 | 0.1421 | 0.1848 | Swelling of limb | symptoms | RATE_C | |
| 771.2 | | | | | 23196 | 67 | 23129 | | | | | 31394 | 110 | 31284 | | | Cramp of limb | symptoms | RATE_C | |
| 772 | | | | | 23175 | 134 | 23041 | | | | | 30713 | 257 | 30456 | | | Symptoms of the muscles | symptoms | RATE_C | |
| 772.1 | | | | | 23094 | 53 | 23041 | | | | | 30516 | 60 | 30456 | | | Muscular wasting and disuse atrophy | symptoms | RATE_C | |
| 772.2 | 0.6365 | 0.1076 | 1.89 | 3.27e-09 | 23676 | 635 | 23041 | 0.8969 | 0.07896 | 2.452 | 6.70e-30 | 31533 | 1077 | 30456 | 4.48e-04 | 0.4309 | Spasm of muscle | symptoms | RATE_C | |
| 772.3 | 0.6184 | 0.07535 | 1.856 | 2.27e-16 | 24642 | 1601 | 23041 | 0.7608 | 0.05673 | 2.14 | 5.30e-41 | 32724 | 2268 | 30456 | 0.04273 | 0.1807 | Muscle weakness | symptoms | RATE_C | |
| 772.4 | | | | | 23175 | 134 | 23041 | | | | | 30527 | 71 | 30456 | | | Rhabdomyolysis | symptoms | RATE_C | |
| 772.6 | | | | | 23142 | 101 | 23041 | | | | | 30584 | 128 | 30456 | | | Facial weakness | symptoms | RATE_C | |
| 780 | | | | | 27429 | 77 | 27352 | | | | | 36521 | 68 | 36453 | | | Hypothermia/Chills | symptoms | RATE_C | |
| 781 | 0.47 | 0.1272 | 1.6 | 2.20e-04 | 26628 | 527 | 26101 | 0.9059 | 0.1126 | 2.474 | 8.53e-16 | 35384 | 686 | 34698 | 2.36e-04 | 0.5935 | Symptoms involving nervous and musculoskeletal systems | symptoms | RATE_C | |
| 781.1 | | | | | 26112 | 11 | 26101 | | | | | 34735 | 37 | 34698 | | | Loss of height | symptoms | RATE_C | |
| 781.2 | | | | | 26298 | 197 | 26101 | | | | | 34933 | 235 | 34698 | | | Abnormal posture | symptoms | RATE_C | |
| 782 | | | | | 22002 | 106 | 21896 | | | | | 29294 | 158 | 29136 | | | Symptoms involving skin and other integumentary tissue | symptoms | RATE_C | |
| 782.3 | 1.347 | 0.06785 | 3.845 | 1.10e-87 | 24907 | 3011 | 21896 | 1.497 | 0.05299 | 4.466 | 1.80e-175 | 33039 | 3903 | 29136 | 0.002785 | 0.25 | Edema | symptoms | RATE_C | |
| 782.6 | | | | | 21947 | 51 | 21896 | | | | | 29472 | 336 | 29136 | | | Pallor and flushing | symptoms | RATE_C | |
| 783 | 1.245 | 0.04906 | 3.472 | 5.24e-142 | 24799 | 4188 | 20611 | 1.256 | 0.0423 | 3.512 | 8.88e-194 | 32997 | 4266 | 28731 | 0.144 | 0.08623 | Fever of unknown origin | symptoms | RATE_C | |
| 783.1 | | | | | 20666 | 55 | 20611 | | | | | 28768 | 37 | 28731 | | | Postprocedural fever | symptoms | RATE_C | |
| 785 | 0.8684 | 0.03914 | 2.383 | 4.50e-109 | 24378 | 6486 | 17892 | 0.9014 | 0.02884 | 2.463 | 2.00e-214 | 32108 | 11436 | 20672 | 0.3907 | -0.03776 | Abdominal pain | symptoms | RATE_C | |
| 788 | 1.594 | 0.08747 | 4.923 | 3.49e-74 | 26167 | 2188 | 23979 | 1.613 | 0.06483 | 5.017 | 1.36e-136 | 34916 | 2566 | 32350 | 0.3518 | -0.09807 | Syncope and collapse | symptoms | RATE_C | |
| 789 | 1.104 | 0.04731 | 3.015 | 2.22e-120 | 24975 | 4146 | 20829 | 1.234 | 0.03437 | 3.435 | 3.37e-282 | 32311 | 6758 | 25553 | 3.96e-06 | 0.2455 | Nausea and vomiting | symptoms | RATE_C | |
| 789.1 | | | | | 20853 | 24 | 20829 | | | | | 25611 | 58 | 25553 | | | Persistent vomiting | symptoms | RATE_C | |
| 790 | | | | | 22915 | 183 | 22732 | | | | | 30897 | 312 | 30585 | | | Nonspecific findings on examination of blood | symptoms | RATE_C | |
| 790.1 | | | | | 22812 | 80 | 22732 | | | | | 30855 | 270 | 30585 | | | Elevated sedimentation rate | symptoms | RATE_C | |
| 790.6 | 0.5486 | 0.06102 | 1.731 | 2.47e-19 | 25006 | 2274 | 22732 | 0.716 | 0.05189 | 2.046 | 2.63e-43 | 33298 | 2713 | 30585 | 1.28e-05 | 0.3268 | Other abnormal blood chemistry | symptoms | RATE_C | |
| 790.8 | | | | | 22867 | 135 | 22732 | | | | | 30869 | 284 | 30585 | | | Elevated C-reactive protein (CRP) | symptoms | RATE_C | |
| 790.9 | | | | | 22737 | 5 | 22732 | | | | | 30591 | 6 | 30585 | | | Abnormal arterial blood gases | symptoms | RATE_C | |

| phenotype | male_beta | male_SE | male_OR | male_p | male_n_total | male_n_cases | male_n_controls | female_beta | female_SE | female_OR | female_p | female_n_total | female_n_cases | female_n_controls | interaction_p | interaction_beta | description | group | lab | label |
|---|---|---|---|---|---|---|---|---|---|---|---|---|---|---|---|---|---|---|---|---|
| 791 | | | | | 27648 | 165 | 27483 | | | | | 36840 | 92 | 36748 | | | Gangrene | symptoms | RATE_C | |
| 792 | | | | | 7 | | | 0.2588 | 0.05852 | 1.295 | 9.77e-06 | 29446 | 1607 | 27839 | 0.9998 | -0.0624 | Abnormal Papanicolaou smear of cervix and cervical HPV | genitourinary | RATE_C | |
| 792.1 | | | | | 7 | | | 0.2638 | 0.06352 | 1.302 | 3.28e-05 | 29183 | 1344 | 27839 | 0.9997 | -0.09353 | Papanicolaou smear of cervix or vagina with atypical squamous cells | genitourinary | RATE_C | |
| 794 | | | | | 27589 | 45 | 27544 | | | | | 36771 | 38 | 36733 | | | Abnormal results of other function studies (bladder, pancreas, placenta, spleen, etc) | symptoms | RATE_C | |
| 795 | | | | | 27272 | 12 | 27260 | | | | | 36232 | 106 | 36126 | | | Other and nonspecific abnormal cytological, histological and immunological findings | symptoms | RATE_C | |
| 795.8 | | | | | 27502 | 242 | 27260 | | | | | 36401 | 275 | 36126 | 0.05336 | 0.4436 | Abnormal tumor markers | symptoms | RATE_C | |
| 795.81 | | | | | 27386 | 126 | 27260 | | | | | 36227 | 101 | 36126 | | | Elevated carcinoembryonic antigen [CEA] | symptoms | RATE_C | |
| 795.82 | | | | | 27261 | 1 | 27260 | | | | | 36221 | 95 | 36126 | | | Elevated cancer antigen 125 [CA 125] | symptoms | RATE_C | |
| 796 | 0.3074 | 0.08155 | 1.36 | 1.63e-04 | 20872 | 1484 | 19388 | | | | | 4 | | | 0.9936 | -2.443 | Elevated prostate specific antigen [PSA] | genitourinary | RATE_C | |
| 797 | 2.161 | 0.1426 | 8.679 | 7.03e-52 | 26901 | 1035 | 25866 | 2.079 | 0.1403 | 8 | 1.10e-49 | 36273 | 725 | 35548 | 0.9549 | 0.01103 | Shock | symptoms | RATE_C | |
| 797.1 | | | | | 26238 | 372 | 25866 | | | | | 35746 | 198 | 35548 | 0.336 | 0.4595 | Cardiogenic shock | symptoms | RATE_C | |
| 798 | 0.831 | 0.03755 | 2.296 | 1.56e-108 | 23475 | 7649 | 15826 | 0.7892 | 0.02846 | 2.202 | 3.42e-169 | 30980 | 11865 | 19115 | 0.5983 | -0.0233 | Malaise and fatigue | symptoms | RATE_C | |
| 798.1 | | | | | 16119 | 293 | 15826 | 0.4995 | 0.08955 | 1.648 | 2.44e-08 | 19839 | 724 | 19115 | 0.8155 | 0.03886 | Chronic fatigue syndrome | symptoms | RATE_C | |
| 800 | 0.4199 | 0.08006 | 1.522 | 1.56e-07 | 22951 | 1064 | 21887 | 0.698 | 0.06372 | 2.01 | 6.33e-28 | 30522 | 1698 | 28824 | 1.59e-13 | 0.7026 | Fracture of lower limb | injuries & poisonings | RATE_C | |
| 800.1 | | | | | 22264 | 377 | 21887 | 1.013 | 0.128 | 2.753 | 2.53e-15 | 29394 | 570 | 28824 | 6.32e-07 | 0.9204 | Fracture of neck of femur | injuries & poisonings | RATE_C | |
| 800.2 | | | | | 22107 | 220 | 21887 | | | | | 29062 | 238 | 28824 | | | Fracture of unspecified part of femur | injuries & poisonings | RATE_C | |
| phenotype | male_beta | male_SE | male_OR | male_p | male_n_total | male_n_cases | male_n_controls | female_beta | female_SE | female_OR | female_p | female_n_total | female_n_cases | female_n_controls | interaction_p | interaction_beta | description | group | lab | label |
| 800.3 | | | | | 22240 | 353 | 21887 | | | | | 29291 | 467 | 28824 | 1.60e-05 | 0.6883 | Fracture of tibia and fibula | injuries & poisonings | RATE_C | |
| 800.4 | | | | | 22024 | 137 | 21887 | | | | | 29015 | 191 | 28824 | | | Fracture of patella | injuries & poisonings | RATE_C | |
| 801 | 0.7529 | 0.1107 | 2.123 | 1.06e-11 | 22470 | 583 | 21887 | 0.6521 | 0.07134 | 1.92 | 6.19e-20 | 30057 | 1233 | 28824 | 0.001218 | 0.3896 | Fracture of ankle and foot | injuries & poisonings | RATE_C | |
| 801.1 | | | | | 22222 | 335 | 21887 | 0.9395 | 0.113 | 2.559 | 9.53e-17 | 29349 | 525 | 28824 | 0.002635 | 0.5153 | Fracture of foot | injuries & poisonings | RATE_C | |
| 802 | | | | | 22131 | 244 | 21887 | | | | | 29104 | 280 | 28824 | 0.003571 | 0.6171 | Fracture of pelvis | injuries & poisonings | RATE_C | |
| 803 | 0.4717 | 0.08098 | 1.603 | 5.73e-09 | 22909 | 1022 | 21887 | 0.5183 | 0.0625 | 1.679 | 1.11e-16 | 30429 | 1605 | 28824 | 6.16e-11 | 0.6102 | Fracture of upper limb | injuries & poisonings | RATE_C | |
| 803.1 | | | | | 22122 | 235 | 21887 | | | | | 29293 | 469 | 28824 | 1.16e-06 | 0.9251 | Fracture of humerus | injuries & poisonings | RATE_C | |
| 803.2 | | | | | 22365 | 478 | 21887 | 0.3808 | 0.08129 | 1.463 | 2.81e-06 | 29684 | 860 | 28824 | 7.45e-06 | 0.5667 | Fracture of radius and ulna | injuries & poisonings | RATE_C | |
| 803.21 | | | | | 21931 | 44 | 21887 | | | | | 28974 | 150 | 28824 | | | Colles' fracture | injuries & poisonings | RATE_C | |
| 803.3 | | | | | 22141 | 254 | 21887 | | | | | 28997 | 173 | 28824 | | | Fracture of clavicle or scapula | injuries & poisonings | RATE_C | |
| 804 | | | | | 22374 | 487 | 21887 | | | | | 29317 | 493 | 28824 | 4.81e-05 | 0.569 | Fracture of hand or wrist | injuries & poisonings | RATE_C | |

| phenotype | male_beta | male_SE | male_OR | male_p | male_n_total | male_n_cases | male_n_controls | female_beta | female_SE | female_OR | female_p | female_n_total | female_n_cases | female_n_controls | interaction_p | interaction_beta | description | group | lab | label |
|---|---|---|---|---|---|---|---|---|---|---|---|---|---|---|---|---|---|---|---|---|
| 805 | 0.7516 | 0.09935 | 2.12 | 3.88e-14 | 22729 | 842 | 21887 | 0.8672 | 0.08813 | 2.38 | 7.50e-23 | 29817 | 993 | 28824 | 4.31e-05 | 0.5174 | Fracture of vertebral column without mention of spinal cord injury | injuries & poisonings | RATE_C | |
| 807 | | | | | 22286 | 399 | 21887 | | | | | 29056 | 232 | 28824 | 0.9231 | -0.02243 | Fracture of ribs | injuries & poisonings | RATE_C | |
| 809 | 0.6332 | 0.0715 | 1.884 | 8.36e-19 | 23404 | 1517 | 21887 | 0.6968 | 0.05969 | 2.007 | 1.75e-31 | 30670 | 1846 | 28824 | 4.69e-05 | 0.3526 | Fracture of unspecified bones | injuries & poisonings | RATE_C | |
| 816 | | | | | 26400 | 43 | 26357 | | | | | 35690 | 21 | 35669 | | | Cerebral laceration and contusion | injuries & poisonings | RATE_C | |
| 817 | | | | | 26422 | 65 | 26357 | | | | | 35734 | 65 | 35669 | | | Concussion | injuries & poisonings | RATE_C | |
| 818 | | | | | 26606 | 249 | 26357 | | | | | 35816 | 147 | 35669 | | | Intracranial hemorrhage (injury) | injuries & poisonings | RATE_C | |
| 818.1 | | | | | 26475 | 118 | 26357 | | | | | 35737 | 68 | 35669 | | | Subdural hemorrhage (injury) | injuries & poisonings | RATE_C | |
| 818.2 | | | | | 26451 | 94 | 26357 | | | | | 35727 | 58 | 35669 | | | Subarachnoid hemorrhage (injury) | injuries & poisonings | RATE_C | |
| 819 | 0.8925 | 0.113 | 2.441 | 2.85e-15 | 26908 | 551 | 26357 | | | | | 36031 | 362 | 35669 | 0.02526 | 0.3514 | Skull and face fracture and other intercranial injury | injuries & poisonings | RATE_C | |
| 823 | | | | | 21319 | | | | | | | 28267 | 2 | 28265 | | | Torus fracture | injuries & poisonings | RATE_C | |
| 830 | 0.2521 | 0.08503 | 1.287 | 0.003031 | 26492 | 928 | 25564 | 0.4707 | 0.07068 | 1.601 | 2.76e-11 | 34859 | 1219 | 33640 | 3.31e-05 | 0.4186 | Dislocation | injuries & poisonings | RATE_C | |
| 835 | | | | | 26013 | 449 | 25564 | 0.3579 | 0.0816 | 1.43 | 1.16e-05 | 34495 | 855 | 33640 | 0.8976 | 0.01765 | Internal derangement of knee | injuries & poisonings | RATE_C | |
| 836 | | | | | 25643 | 79 | 25564 | | | | | 33724 | 84 | 33640 | | | Traumatic arthropathy | injuries & poisonings | RATE_C | |
| 840 | 0.295 | 0.07161 | 1.343 | 3.80e-05 | 25492 | 1396 | 24096 | 0.4914 | 0.05387 | 1.635 | 7.45e-20 | 33198 | 2125 | 31073 | 0.005782 | 0.224 | Sprains and strains | injuries & poisonings | RATE_C | |
| 840.1 | | | | | 24130 | 34 | 24096 | | | | | 31116 | 43 | 31073 | | | Muscle/tendon sprain | injuries & poisonings | RATE_C | |
| 840.2 | | | | | 24369 | 273 | 24096 | | | | | 31341 | 268 | 31073 | 0.4676 | 0.163 | Rotator cuff (capsule) sprain | injuries & poisonings | RATE_C | |
| 840.3 | | | | | 24387 | 291 | 24096 | | | | | 31485 | 412 | 31073 | 0.00371 | 0.4686 | Joint/ligament sprain | injuries & poisonings | RATE_C | |
| 841 | | | | | 24387 | 291 | 24096 | 0.9362 | 0.106 | 2.55 | 1.06e-18 | 31714 | 641 | 31073 | 0.0903 | 0.3145 | Sprains and strains of back and neck | injuries & poisonings | RATE_C | |
| 842 | | | | | 24179 | 83 | 24096 | | | | | 31195 | 122 | 31073 | | | Other sprains and strains | injuries & poisonings | RATE_C | |
| 850 | | | | | 23223 | 456 | 22767 | | | | | 32260 | 465 | 31795 | 0.07318 | 0.3955 | Hemorrhage or hematoma complicating a procedure | injuries & poisonings | RATE_C | |
| 851 | | | | | 23157 | 390 | 22767 | | | | | 32086 | 291 | 31795 | 0.03792 | -0.6979 | Complications of transplants and reattached limbs | injuries & poisonings | RATE_C | |
| 853 | | | | | 22918 | 151 | 22767 | | | | | 31956 | 161 | 31795 | | | Complication of colostomy or enterostomy | injuries & poisonings | RATE_C | |
| 854 | 2.325 | 0.1755 | 10.22 | 4.73e-40 | 23435 | 668 | 22767 | 2.121 | 0.1549 | 8.341 | 1.08e-42 | 32312 | 517 | 31795 | 0.506 | -0.1504 | Complications of cardiac/vascular device, implant, and graft | injuries & poisonings | RATE_C | |
| 855 | | | | | 22967 | 200 | 22767 | | | | | 32019 | 224 | 31795 | | | Complication of nervous system device, implant, and graft | injuries & poisonings | RATE_C | |
| 856 | | | | | 22823 | 56 | 22767 | | | | | 31840 | 45 | 31795 | | | Vascular complications of surgery and medical procedures | injuries & poisonings | RATE_C | |
| 857 | | | | | 22852 | 85 | 22767 | | | | | 31886 | 91 | 31795 | | | Mechanical complication of unspecified genitourinary device, implant, and graft | injuries & poisonings | RATE_C | |
| 858 | | | | | 23191 | 424 | 22767 | 1.183 | 0.116 | 3.264 | 2.08e-24 | 32396 | 601 | 31795 | 3.06e-04 | 0.6033 | Complication of internal orthopedic device | injuries & poisonings | RATE_C | |
| 859 | | | | | 23165 | 398 | 22767 | | | | | 32227 | 432 | 31795 | 0.3865 | 0.1676 | Complication due to other implant and internal device | injuries & poisonings | RATE_C | |

| phenotype | male_beta | male_SE | male_OR | male_p | male_n_total | male_n_cases | male_n_controls | female_beta | female_SE | female_OR | female_p | female_n_total | female_n_cases | female_n_controls | interaction_p | interaction_beta | description | group | lab | label |
|---|---|---|---|---|---|---|---|---|---|---|---|---|---|---|---|---|---|---|---|---|
| 860 | | | | | 27702 | 127 | 27575 | | | | | 36911 | 120 | 36791 | | | Bone marrow or stem cell transplant | neoplasms | RATE_C | |
| 870 | 1.128 | 0.1094 | 3.091 | 6.15e-25 | 24342 | 736 | 23606 | 1.359 | 0.1086 | 3.892 | 6.59e-36 | 33149 | 715 | 32434 | 1.98e-04 | 0.5371 | Open wounds of head; neck; and trunk | injuries & poisonings | RATE_C | |
| 870.1 | | | | | 23654 | 48 | 23606 | | | | | 32454 | 20 | 32434 | | | Open wound or laceration of eye or eyelid | injuries & poisonings | RATE_C | |
| 870.2 | | | | | 23622 | 16 | 23606 | | | | | 32443 | 9 | 32434 | | | Open wound of ear | injuries & poisonings | RATE_C | |
| 870.3 | | | | | 23791 | 185 | 23606 | | | | | 32553 | 119 | 32434 | | | Other open wound of head and face | injuries & poisonings | RATE_C | |
| 870.4 | | | | | 23626 | 20 | 23606 | | | | | 32447 | 13 | 32434 | | | Open wound of nose and sinus | injuries & poisonings | RATE_C | |
| 870.5 | | | | | 23652 | 46 | 23606 | | | | | 32464 | 30 | 32434 | | | Open wound of lip and mouth | injuries & poisonings | RATE_C | |
| 870.6 | | | | | 23625 | 19 | 23606 | | | | | 32440 | 6 | 32434 | | | Open wound of neck | injuries & poisonings | RATE_C | |
| 870.8 | | | | | 23613 | 7 | 23606 | | | | | 32442 | 8 | 32434 | | | Open wound of genital organs | injuries & poisonings | RATE_C | |
| 871 | 0.642 | 0.08665 | 1.9 | 1.27e-13 | 24632 | 1026 | 23606 | 0.9635 | 0.09012 | 2.621 | 1.12e-26 | 33363 | 929 | 32434 | 3.13e-07 | 0.5958 | Open wounds of extremities | injuries & poisonings | RATE_C | |
| 871.1 | | | | | 23768 | 162 | 23606 | | | | | 32546 | 112 | 32434 | | | Open wound of hand except finger(s) | injuries & poisonings | RATE_C | |
| 871.2 | | | | | 23775 | 169 | 23606 | | | | | 32574 | 140 | 32434 | | | Open wound of finger(s) | injuries & poisonings | RATE_C | |
| 871.3 | | | | | 23720 | 114 | 23606 | | | | | 32510 | 76 | 32434 | | | Open wound of foot except toe(s) alone | injuries & poisonings | RATE_C | |
| 871.4 | | | | | 23638 | 32 | 23606 | | | | | 32453 | 19 | 32434 | | | Open wound of toe(s) | injuries & poisonings | RATE_C | |
| 872 | | | | | 23730 | 124 | 23606 | | | | | 32497 | 63 | 32434 | | | Traumatic amputation | injuries & poisonings | RATE_C | |
| 874 | | | | | 23727 | 121 | 23606 | | | | | 32492 | 58 | 32434 | | | Complication of amputation stump | injuries & poisonings | RATE_C | |
| 875 | | | | | 23705 | 99 | 23606 | | | | | 32557 | 123 | 32434 | | | Non-healing surgical wound | injuries & poisonings | RATE_C | |
| 876 | | | | | 23630 | 24 | 23606 | | | | | 32475 | 41 | 32434 | | | Posttraumatic wound infection not elsewhere classified | injuries & poisonings | RATE_C | |
| 905 | | | | | 27300 | 4 | 27296 | | | | | 36496 | 6 | 36490 | | | Late effects of musculoskeletal and connective tissue injuries | injuries & poisonings | RATE_C | |
| 907 | | | | | 27494 | 198 | 27296 | | | | | 36638 | 148 | 36490 | | | Injuries to the nervous system | injuries & poisonings | RATE_C | |
| 910 | | | | | 27755 | 6 | 27749 | | | | | 36903 | 8 | 36895 | | | Superficial injury, infected | injuries & poisonings | RATE_C | |
| 911 | | | | | 27688 | 32 | 27656 | | | | | 36846 | 32 | 36814 | | | Blister | injuries & poisonings | RATE_C | |
| 912 | | | | | 27054 | 122 | 26932 | | | | | 35677 | 201 | 35476 | | | Insect bite | injuries & poisonings | RATE_C | |
| 913 | | | | | 26968 | 36 | 26932 | | | | | 35544 | 68 | 35476 | | | Toxic effect of venom | injuries & poisonings | RATE_C | |
| 916 | 0.8126 | 0.1185 | 2.254 | 7.09e-12 | 26060 | 586 | 25474 | 1.07 | 0.08754 | 2.915 | 2.38e-34 | 34442 | 1043 | 33399 | 5.41e-05 | 0.5516 | Contusion | injuries & poisonings | RATE_C | |
| 930 | | | | | 22109 | 280 | 21829 | 0.6753 | 0.0975 | 1.965 | 4.33e-12 | 27961 | 619 | 27342 | 0.02738 | 0.3622 | Allergic reaction to food | injuries & poisonings | RATE_C | |
| 931 | | | | | 21895 | 66 | 21829 | | | | | 27431 | 89 | 27342 | | | Contact dermatitis and other eczema due to plants [except food] | dermatologic | RATE_C | |
| 938 | 0.1613 | 0.0978 | 1.175 | 0.09902 | 22767 | 938 | 21829 | 0.3768 | 0.09129 | 1.458 | 3.67e-05 | 28152 | 810 | 27342 | 0.5339 | 0.0801 | Dermatitis due to solar radiation | dermatologic | RATE_C | |

| phenotype | male_beta | male_SE | male_OR | male_p | male_n_total | male_n_cases | male_n_controls | female_beta | female_SE | female_OR | female_p | female_n_total | female_n_cases | female_n_controls | interaction_p | interaction_beta | description | group | lab | label |
|---|---|---|---|---|---|---|---|---|---|---|---|---|---|---|---|---|---|---|---|---|
| 938.1 | | | | | 21840 | 11 | 21829 | | | | | 27356 | 14 | 27342 | | | Acute dermatitis due to solar radiation | dermatologic | RATE_C | |
| 938.2 | 0.2015 | 0.1071 | 1.223 | 0.06006 | 22665 | 836 | 21829 | 0.3234 | 0.09612 | 1.382 | 7.67e-04 | 28065 | 723 | 27342 | 0.7546 | -0.04345 | Chronic dermatitis due to solar radiation | dermatologic | RATE_C | |
| 939 | 0.2253 | 0.07411 | 1.253 | 0.002369 | 23219 | 1390 | 21829 | 0.3661 | 0.05328 | 1.442 | 6.38e-12 | 29539 | 2197 | 27342 | 0.3012 | 0.08768 | Atopic/contact dermatitis due to other or unspecified | dermatologic | RATE_C | |
| 939.1 | | | | | 21831 | 2 | 21829 | | | | | 27354 | 12 | 27342 | | | Contact and allergic dermatitis of eyelid | dermatologic | RATE_C | |
| 941 | | | | | 21871 | 42 | 21829 | | | | | 27432 | 90 | 27342 | | | Adverse reaction to serum or vaccine | injuries & poisonings | RATE_C | |
| 942 | | | | | 21868 | 39 | 21829 | | | | | 27392 | 50 | 27342 | | | Infusion and transfusion reaction | injuries & poisonings | RATE_C | |
| 946 | | | | | 21895 | 66 | 21829 | | | | | 27494 | 152 | 27342 | | | Anaphylactic shock NOS | injuries & poisonings | RATE_C | |
| 947 | | | | | 21986 | 157 | 21829 | | | | | 27830 | 488 | 27342 | 0.2922 | 0.2054 | Urticaria | dermatologic | RATE_C | |
| 949 | 0.4792 | 0.119 | 1.615 | 5.64e-05 | 22350 | 521 | 21829 | 0.8463 | 0.07546 | 2.331 | 3.42e-29 | 28540 | 1198 | 27342 | 0.001364 | 0.4162 | Allergies, other | injuries & poisonings | RATE_C | |
| 949.1 | | | | | 21836 | 7 | 21829 | | | | | 27350 | 8 | 27342 | | | Diaper or napkin rash | injuries & poisonings | RATE_C | |
| 952 | | | | | 27725 | 66 | 27659 | | | | | 36923 | 32 | 36891 | | | Spinal cord injury without evidence of spinal bone injury | injuries & poisonings | RATE_C | |
| 957 | | | | | 27659 | | | | | | | 36893 | 2 | 36891 | | | Injury to other and unspecified nerves | injuries & poisonings | RATE_C | |
| 958 | | | | | 27212 | 374 | 26838 | | | | | 36518 | 232 | 36286 | 0.09612 | 0.5453 | Certain early complications of trauma or procedure | injuries & poisonings | RATE_C | |
| 958.1 | | | | | 26905 | 67 | 26838 | | | | | 36331 | 45 | 36286 | | | Postoperative shock | injuries & poisonings | RATE_C | |
| 958.2 | | | | | 26974 | 136 | 26838 | | | | | 36364 | 78 | 36286 | | | Traumatic and surgical subcutaneous emphysema | injuries & poisonings | RATE_C | |
| 960 | 0.9755 | 0.08508 | 2.652 | 1.97e-30 | 21000 | 1222 | 19778 | 1.267 | 0.06137 | 3.549 | 1.24e-94 | 28211 | 2139 | 26072 | 2.01e-05 | 0.4186 | Poisoning by antibiotics | injuries & poisonings | RATE_C | |
| 960.1 | | | | | 19779 | 1 | 19778 | | | | | 26073 | 1 | 26072 | | | Adverse effects of antibacterials (not penicillins) | injuries & poisonings | RATE_C | |
| 960.2 | 1.007 | 0.1135 | 2.737 | 7.58e-19 | 20476 | 698 | 19778 | 1.22 | 0.08017 | 3.388 | 2.65e-52 | 27268 | 1196 | 26072 | 0.01498 | 0.3186 | Allergy/adverse effect of penicillin | injuries & poisonings | RATE_C | |
| 960.3 | | | | | 19779 | 1 | 19778 | | | | | 26073 | 1 | 26072 | | | Poisoning by antifungal antibiotics | injuries & poisonings | RATE_C | |
| 961 | | | | | 19843 | 65 | 19778 | | | | | 26198 | 126 | 26072 | | | Poisoning by other anti-infectives | injuries & poisonings | RATE_C | |
| 961.1 | | | | | 20163 | 385 | 19778 | 1.147 | 0.08476 | 3.149 | 9.71e-42 | 27149 | 1077 | 26072 | 0.4403 | 0.1344 | Poisoning/allergy of sulfonamides | injuries & poisonings | RATE_C | |
| 962 | | | | | 19885 | 107 | 19778 | | | | | 26193 | 121 | 26072 | | | Poisoning by hormones and synthetic substitutes | injuries & poisonings | RATE_C | |
| 962.1 | | | | | 19862 | 84 | 19778 | | | | | 26166 | 94 | 26072 | | | Adrenal cortical steroids causing adverse effects in therapeutic use | injuries & poisonings | RATE_C | |
| 962.2 | | | | | 19800 | 22 | 19778 | | | | | 26094 | 22 | 26072 | | | Insulins and antidiabetic agents causing adverse effects in therapeutic use | injuries & poisonings | RATE_C | |
| 962.3 | | | | | 19778 | | | | | | | 26078 | 6 | 26072 | | | Hormones and synthetic substitutes causing adverse effects in therapeutic use | injuries & poisonings | RATE_C | |
| 963 | 1.091 | 0.1174 | 2.977 | 1.49e-20 | 20349 | 571 | 19778 | 1.87 | 0.127 | 6.486 | 4.44e-49 | 26631 | 559 | 26072 | 4.02e-07 | 0.83 | Poisoning by primarily systemic agents | injuries & poisonings | RATE_C | |
| 963.1 | 1.083 | 0.1184 | 2.954 | 5.70e-20 | 20340 | 562 | 19778 | 1.861 | 0.1285 | 6.428 | 1.72e-47 | 26617 | 545 | 26072 | 6.05e-07 | 0.8262 | Antineoplastic and immunosuppressive drugs causing adverse effects | injuries & poisonings | RATE_C | |
| 964 | | | | | 19833 | 55 | 19778 | | | | | 26136 | 64 | 26072 | | | Poisoning by agents primarily affecting blood constituents | injuries & poisonings | RATE_C | |

| phenotype | male_beta | male_SE | male_OR | male_p | male_n_total | male_n_cases | male_n_controls | female_beta | female_SE | female_OR | female_p | female_n_total | female_n_cases | female_n_controls | interaction_p | interaction_beta | description | group | lab | label |
|---|---|---|---|---|---|---|---|---|---|---|---|---|---|---|---|---|---|---|---|---|
| 964.1 | | | | | 19833 | 55 | 19778 | | | | | 26131 | 59 | 26072 | | | Anticoagulants causing adverse effects | injuries & poisonings | RATE_C | |
| 965 | | | | | 20187 | 409 | 19778 | 1.228 | 0.09362 | 3.414 | 2.69e-39 | 26882 | 810 | 26072 | 0.234 | 0.1934 | Poisoning by analgesics, antipyretics, and antirheumatics | injuries & poisonings | RATE_C | |
| 965.1 | 1.57 | 0.1505 | 4.808 | 1.77e-25 | 20350 | 572 | 19778 | 1.435 | 0.08235 | 4.2 | 5.34e-68 | 27386 | 1314 | 26072 | 0.8397 | 0.03307 | Opiates and related narcotics causing adverse effects in therapeutic use | injuries & poisonings | RATE_C | |
| 965.2 | | | | | 19780 | 2 | 19778 | | | | | 26073 | 1 | 26072 | | | Antirheumatics causing adverse effects in therapeutic use | injuries & poisonings | RATE_C | |
| 965.3 | | | | | 19783 | 5 | 19778 | | | | | 26085 | 13 | 26072 | | | Salicylates causing adverse effects in therapeutic use | injuries & poisonings | RATE_C | |
| 966 | | | | | 19797 | 19 | 19778 | | | | | 26089 | 17 | 26072 | | | Poisoning by anticonvulsants and anti-Parkinsonism drugs | injuries & poisonings | RATE_C | |
| 967 | | | | | 19831 | 53 | 19778 | | | | | 26222 | 150 | 26072 | | | Adverse effects of sedatives or other central nervous system depressants and anesthetics | injuries & poisonings | RATE_C | |
| 969 | | | | | 19838 | 60 | 19778 | | | | | 26169 | 97 | 26072 | | | Poisoning by psychotropic agents | injuries & poisonings | RATE_C | |
| 971 | | | | | 19780 | 2 | 19778 | | | | | 26075 | 3 | 26072 | | | Poisoning by drugs primarily affecting the autonomic nervous system | injuries & poisonings | RATE_C | |
| 972 | | | | | 19849 | 71 | 19778 | | | | | 26139 | 67 | 26072 | | | Poisoning by agents primarily affecting the cardiovascular system | injuries & poisonings | RATE_C | |
| 972.1 | | | | | 19792 | 14 | 19778 | | | | | 26083 | 11 | 26072 | | | Cardiac rhythm regulators causing adverse effects in therapeutic use | injuries & poisonings | RATE_C | |
| 972.2 | | | | | 19778 | | | | | | | 26072 | | | | | Antilipemic and antiarteriosclerotic drugs causing adverse effects in therapeutic use | injuries & poisonings | RATE_C | |
| 972.6 | | | | | 19783 | 5 | 19778 | | | | | 26077 | 5 | 26072 | | | Antihypertensive agents causing adverse effects | injuries & poisonings | RATE_C | |
| 973 | | | | | 19785 | 7 | 19778 | | | | | 26074 | 2 | 26072 | | | Poisoning by agents primarily affecting the gastrointestinal system | injuries & poisonings | RATE_C | |
| 974 | | | | | 19842 | 64 | 19778 | | | | | 26094 | 22 | 26072 | | | Poisoning by water, mineral, and uric acid metabolism drugs | injuries & poisonings | RATE_C | |
| 975 | | | | | 19784 | 6 | 19778 | | | | | 26079 | 7 | 26072 | | | Poisoning by agents primarily acting on the smooth and skeletal muscles and respiratory system | injuries & poisonings | RATE_C | |
| 976 | | | | | 19780 | 2 | 19778 | | | | | 26075 | 3 | 26072 | | | Poisoning by agents primarily affecting skin & mucous membrane, ophthalmological, otorhinolaryngological, & dental drugs | injuries & poisonings | RATE_C | |
| 977 | 1.331 | 0.09345 | 3.783 | 5.22e-46 | 21166 | 1388 | 19778 | 1.47 | 0.06534 | 4.351 | 3.86e-112 | 28281 | 2209 | 26072 | 0.04795 | 0.2139 | Personal history of allergy to medicinal agents | injuries & poisonings | RATE_C | |
| 979 | 2.134 | 0.1401 | 8.448 | 2.18e-52 | 20676 | 898 | 19778 | 1.772 | 0.102 | 5.882 | 1.27e-67 | 27006 | 934 | 26072 | 0.1801 | -0.224 | Adverse drug events and drug allergies | injuries & poisonings | RATE_C | |
| 980 | 1.432 | 0.0982 | 4.187 | 3.70e-48 | 26597 | 1176 | 25421 | 1.52 | 0.09447 | 4.57 | 3.24e-58 | 35862 | 1000 | 34862 | 0.1674 | 0.1771 | Encounter for long-term (current) use of antibiotics | infectious diseases | RATE_C | |
| 981 | | | | | 27615 | 2 | 27613 | | | | | 36741 | 3 | 36738 | | | Toxic effect of (non-ethyl) alcohol and petroleum and other solvents | injuries & poisonings | RATE_C | |
| 983 | | | | | 27616 | 3 | 27613 | | | | | 36738 | | | | | Toxic effect of corrosive aromatics, acids, and caustic alkalis | injuries & poisonings | RATE_C | |
| 984 | | | | | 27616 | 3 | 27613 | | | | | 36739 | 1 | 36738 | | | Toxic effect of lead and its compounds (including fumes) | injuries & poisonings | RATE_C | |
| 985 | | | | | 27622 | 9 | 27613 | | | | | 36745 | 7 | 36738 | | | Toxic effect of other metals | injuries & poisonings | RATE_C | |
| 986 | | | | | 27616 | 3 | 27613 | | | | | 36740 | 2 | 36738 | | | Toxic effect of carbon monoxide | injuries & poisonings | RATE_C | |

| phenotype | male_beta | male_SE | male_OR | male_p | male_n_total | male_n_cases | male_n_controls | female_beta | female_SE | female_OR | female_p | female_n_total | female_n_cases | female_n_controls | interaction_p | interaction_beta | description | group | lab | label |
|---|---|---|---|---|---|---|---|---|---|---|---|---|---|---|---|---|---|---|---|---|
| 987 | | | | | 27617 | 4 | 27613 | | | | | 36745 | 7 | 36738 | | | Toxic effect of other gases, fumes, or vapors | injuries & poisonings | RATE_C | |
| 988 | | | | | 27613 | | | | | | | 36740 | 2 | 36738 | | | Toxic effect of noxious substances eaten as food | injuries & poisonings | RATE_C | |
| 989 | | | | | 27632 | 19 | 27613 | | | | | 36758 | 20 | 36738 | | | Toxic effect of other substances, chiefly nonmedicinal as to source | injuries & poisonings | RATE_C | |
| 990 | 0.8148 | 0.09844 | 2.259 | 1.26e-16 | 23446 | 892 | 22554 | 1.094 | 0.09767 | 2.986 | 4.08e-29 | 33242 | 782 | 32460 | 0.06409 | 0.244 | Effects radiation NOS | injuries & poisonings | RATE_C | |
| 994 | 1.809 | 0.07362 | 6.106 | 2.26e-133 | 26534 | 2690 | 23844 | 1.882 | 0.0729 | 6.566 | 5.80e-147 | 35853 | 2103 | 33750 | 0.1595 | 0.1395 | Sepsis and SIRS | injuries & poisonings | RATE_C | |
| 994.1 | 1.988 | 0.1487 | 7.302 | 9.02e-41 | 24423 | 579 | 23844 | | | | | 34199 | 449 | 33750 | 0.001102 | 0.75 | Systemic inflammatory response syndrome (SIRS) | injuries & poisonings | RATE_C | |
| 994.2 | 1.893 | 0.0825 | 6.639 | 1.63e-116 | 26190 | 2346 | 23844 | 1.845 | 0.07735 | 6.325 | 1.09e-125 | 35609 | 1859 | 33750 | 0.9607 | 0.005349 | Sepsis | injuries & poisonings | RATE_C | |
| 994.21 | 2.383 | 0.159 | 10.84 | 7.96e-51 | 24740 | 896 | 23844 | 2.389 | 0.1558 | 10.9 | 4.91e-53 | 34424 | 674 | 33750 | 0.7046 | 0.08229 | Septic shock | injuries & poisonings | RATE_C | |

#### STable16: Sex stratified regression estimate results from reweighting via inverse probability for ascertainment weighting for vitamin D, LDL cholesterol, heart rate from EKG and uric acid

24 rows × 11 columns

| lab | snp | sex | term | estimate | std.error | statistic | p.value | conf.low | conf.high | weight_type |
|---|---|---|---|---|---|---|---|---|---|---|
| ldlc_median | rs7412_T_binary | Female | rs7412_T_binary | -15.99 | 0.602 | 705.24 | 0 | -17.17 | -14.81 | Unweighted |
| ldlc_median | rs7412_T_binary | Female | rs7412_T_binary | -16.96 | 0.9279 | 333.96 | 0 | -18.78 | -15.14 | Reweighted to BioVU genetic cohort |
| ldlc_median | rs7412_T_binary | Female | rs7412_T_binary | -17.93 | 1.178 | 231.50 | 0 | -20.24 | -15.62 | Reweighted to general US population estimate |
| urica_median | rs1014290_G_binary | Female | rs1014290_G_binary | -0.5323 | 0.04464 | 142.17 | 0 | -0.6198 | -0.4448 | Unweighted |
| urica_median | rs1014290_G_binary | Female | rs1014290_G_binary | -0.49 | 0.04754 | 106.24 | 0 | -0.5832 | -0.3968 | Reweighted to BioVU genetic cohort |
| urica_median | rs1014290_G_binary | Female | rs1014290_G_binary | -0.5025 | 0.05172 | 94.4 | 0 | -0.6039 | -0.4011 | Reweighted to general US population estimate |
| ratec_median | rs79346577_T_binary | Female | rs79346577_T_binary | 0.5661 | 0.2264 | 6.253 | 0.0124 | 0.1224 | 1.01 | Unweighted |
| ratec_median | rs79346577_T_binary | Female | rs79346577_T_binary | 0.7524 | 0.2866 | 6.891 | 0.008663 | 0.1906 | 1.314 | Reweighted to BioVU genetic cohort |
| ratec_median | rs79346577_T_binary | Female | rs79346577_T_binary | 0.6225 | 0.3432 | 3.29 | 0.06971 | -0.05016 | 1.295 | Reweighted to general US population estimate |
| vitd_median | rs731844_C_binary | Female | rs731844_C_binary | -0.8549 | 0.3175 | 7.251 | 0.007085 | -1.477 | -0.2327 | Unweighted |
| vitd_median | rs731844_C_binary | Female | rs731844_C_binary | -0.6437 | 0.5168 | 1.552 | 0.2129 | -1.656 | 0.3692 | Reweighted to BioVU genetic cohort |
| vitd_median | rs731844_C_binary | Female | rs731844_C_binary | -0.6184 | 0.6097 | 1.029 | 0.3104 | -1.813 | 0.5765 | Reweighted to general US population estimate |
| ldlc_median | rs7412_T_binary | Male | rs7412_T_binary | -11.34 | 0.7041 | 259.47 | 0 | -12.72 | -9.962 | Unweighted |
| ldlc_median | rs7412_T_binary | Male | rs7412_T_binary | -11.51 | 1.056 | 118.78 | 0 | -13.58 | -9.44 | Reweighted to BioVU genetic cohort |
| ldlc_median | rs7412_T_binary | Male | rs7412_T_binary | -12.04 | 1.446 | 69.38 | 1.11e-16 | -14.88 | -9.209 | Reweighted to general US population estimate |
| urica_median | rs1014290_G_binary | Male | rs1014290_G_binary | -0.2872 | 0.04672 | 37.79 | 7.87e-10 | -0.3788 | -0.1957 | Unweighted |
| urica_median | rs1014290_G_binary | Male | rs1014290_G_binary | -0.2302 | 0.06479 | 12.63 | 3.81e-04 | -0.3572 | -0.1032 | Reweighted to BioVU genetic cohort |
| urica_median | rs1014290_G_binary | Male | rs1014290_G_binary | -0.2355 | 0.07009 | 11.29 | 7.81e-04 | -0.3728 | -0.0981 | Reweighted to general US population estimate |
| ratec_median | rs79346577_T_binary | Male | rs79346577_T_binary | -1.16 | 0.245 | 22.43 | 2.18e-06 | -1.641 | -0.6802 | Unweighted |
| ratec_median | rs79346577_T_binary | Male | rs79346577_T_binary | -1.298 | 0.2927 | 19.67 | 9.20e-06 | -1.872 | -0.7245 | Reweighted to BioVU genetic cohort |
| ratec_median | rs79346577_T_binary | Male | rs79346577_T_binary | -1.344 | 0.3477 | 14.95 | 1.11e-04 | -2.026 | -0.6627 | Reweighted to general US population estimate |
| vitd_median | rs731844_C_binary | Male | rs731844_C_binary | 1.559 | 0.4459 | 12.22 | 4.73e-04 | 0.6848 | 2.433 | Unweighted |
| vitd_median | rs731844_C_binary | Male | rs731844_C_binary | 1.511 | 0.7124 | 4.5 | 0.0339 | 0.1149 | 2.908 | Reweighted to BioVU genetic cohort |
| vitd_median | rs731844_C_binary | Male | rs731844_C_binary | 1.003 | 0.8388 | 1.429 | 0.2319 | -0.6413 | 2.647 | Reweighted to general US population estimate |

#### STable17: SNP and SNP*sex interaction regression estimate results from reweighting via inverse probability for ascertainment weighting for vitamin D, LDL cholesterol, heart rate from EKG and uric acid in sex combined models

24 rows × 11 columns

| lab | snp | term | estimate | std.error | statistic | p.value | conf.low | conf.high | weight_type | interaction_term |
|---|---|---|---|---|---|---|---|---|---|---|
| ldlc_median | rs7412_T_binary | rs7412_T_binary | -15.99 | 0.602 | 705.24 | 0 | -17.17 | -14.81 | Unweighted | main SNP effect |
| ldlc_median | rs7412_T_binary | rs7412_T_binary:genetic_sexMale | 4.645 | 0.9264 | 25.14 | 5.32e-07 | 2.83 | 6.461 | Unweighted | sex interaction |
| ldlc_median | rs7412_T_binary | rs7412_T_binary | -17.17 | 0.9631 | 317.83 | 0 | -19.06 | -15.28 | Reweighted to BioVU genetic cohort | main SNP effect |
| ldlc_median | rs7412_T_binary | rs7412_T_binary:genetic_sexMale | 5.756 | 1.41 | 16.68 | 4.43e-05 | 2.994 | 8.519 | Reweighted to BioVU genetic cohort | sex interaction |
| ldlc_median | rs7412_T_binary | rs7412_T_binary | -18.12 | 1.239 | 214.10 | 0 | -20.55 | -15.7 | Reweighted to general US population estimate | main SNP effect |
| ldlc_median | rs7412_T_binary | rs7412_T_binary:genetic_sexMale | 6.381 | 1.853 | 11.86 | 5.73e-04 | 2.75 | 10.01 | Reweighted to general US population estimate | sex interaction |
| urica_median | rs1014290_G_binary | rs1014290_G_binary | -0.5323 | 0.04464 | 142.17 | 0 | -0.6198 | -0.4448 | Unweighted | main SNP effect |
| urica_median | rs1014290_G_binary | rs1014290_G_binary:genetic_sexMale | 0.245 | 0.06462 | 14.38 | 1.49e-04 | 0.1184 | 0.3717 | Unweighted | sex interaction |
| urica_median | rs1014290_G_binary | rs1014290_G_binary | -0.4828 | 0.04737 | 103.89 | 0 | -0.5757 | -0.39 | Reweighted to BioVU genetic cohort | main SNP effect |
| urica_median | rs1014290_G_binary | rs1014290_G_binary:genetic_sexMale | 0.266 | 0.07524 | 12.5 | 4.06e-04 | 0.1186 | 0.4135 | Reweighted to BioVU genetic cohort | sex interaction |
| urica_median | rs1014290_G_binary | rs1014290_G_binary | -0.493 | 0.05136 | 92.15 | 0 | -0.5937 | -0.3924 | Reweighted to general US population estimate | main SNP effect |
| urica_median | rs1014290_G_binary | rs1014290_G_binary:genetic_sexMale | 0.2741 | 0.0813 | 11.37 | 7.46e-04 | 0.1148 | 0.4335 | Reweighted to general US population estimate | sex interaction |
| ratec_median | rs79346577_T_binary | rs79346577_T_binary | 0.5661 | 0.2264 | 6.253 | 0.0124 | 0.1224 | 1.01 | Unweighted | main SNP effect |
| ratec_median | rs79346577_T_binary | rs79346577_T_binary:genetic_sexMale | -1.726 | 0.3336 | 26.79 | 2.27e-07 | -2.38 | -1.073 | Unweighted | sex interaction |
| ratec_median | rs79346577_T_binary | rs79346577_T_binary | 0.7578 | 0.2881 | 6.918 | 0.008532 | 0.1931 | 1.322 | Reweighted to BioVU genetic cohort | main SNP effect |
| ratec_median | rs79346577_T_binary | rs79346577_T_binary:genetic_sexMale | -2.107 | 0.4101 | 26.39 | 2.80e-07 | -2.911 | -1.303 | Reweighted to BioVU genetic cohort | sex interaction |
| ratec_median | rs79346577_T_binary | rs79346577_T_binary | 0.6508 | 0.3441 | 3.576 | 0.05861 | -0.0237 | 1.325 | Reweighted to general US population estimate | main SNP effect |
| ratec_median | rs79346577_T_binary | rs79346577_T_binary:genetic_sexMale | -2.046 | 0.4887 | 17.52 | 2.84e-05 | -3.004 | -1.088 | Reweighted to general US population estimate | sex interaction |
| vitd_median | rs731844_C_binary | rs731844_C_binary | -0.8549 | 0.3175 | 7.251 | 0.007085 | -1.477 | -0.2327 | Unweighted | main SNP effect |
| vitd_median | rs731844_C_binary | rs731844_C_binary:genetic_sexMale | 2.414 | 0.5474 | 19.44 | 1.04e-05 | 1.341 | 3.487 | Unweighted | sex interaction |
| vitd_median | rs731844_C_binary | rs731844_C_binary | -0.6314 | 0.5105 | 1.53 | 0.2161 | -1.632 | 0.3691 | Reweighted to BioVU genetic cohort | main SNP effect |
| vitd_median | rs731844_C_binary | rs731844_C_binary:genetic_sexMale | 2.301 | 0.8553 | 7.241 | 0.007126 | 0.6251 | 3.978 | Reweighted to BioVU genetic cohort | sex interaction |
| vitd_median | rs731844_C_binary | rs731844_C_binary | -0.6014 | 0.5998 | 1.005 | 0.3161 | -1.777 | 0.5743 | Reweighted to general US population estimate | main SNP effect |

| lab | snp | term | estimate | std.error | statistic | p.value | conf.low | conf.high | weight_type | interaction_term |
|---|---|---|---|---|---|---|---|---|---|---|
| vitd_median | rs731844_C_binary | rs731844_C_binary:genetic_sexMale | 1.81 | 0.9993 | 3.28 | 0.07012 | -0.1487 | 3.769 | Reweighted to general US population estimate | sex interaction |

#### STable18: Colocalization statistics from loci with PP4>0.5 for colocalization between sex-stratified and sex-biased eQTL data and sex-stratifiied GWAS statistics from BioVU

391 rows × 10 columns

| trait | | gene_name | Locus_id | coloc_data_default_nsnps | coloc_data_default_PP.H0.abf | coloc_data_default_PP.H1.abf | coloc_data_default_PP.H2.abf | coloc_data_default_PP.H3.abf | coloc_data_default_PP.H4.abf |
|---|---|---|---|---|---|---|---|---|---|
| A.2Gg__Breast_Mammary_Tissue | males | NUDT14 | ENSG00000183828.14 | 1233 | 0.04059 | 0.3815 | 0.003252 | 0.03001 | 0.5447 |
| A.2Gg__Breast_Mammary_Tissue | males | DYNC2LI1 | ENSG00000138036.18 | 2850 | 0.07675 | 0.102 | 0.1107 | 0.1466 | 0.5639 |
| A1Anti_Breast_Mammary_Tissue | males | STEAP3 | ENSG00000115107.19 | 2111 | 0.1459 | 0.2295 | 0.03737 | 0.05826 | 0.529 |
| A1Anti_Breast_Mammary_Tissue | males | DEFB1 | ENSG00000164825.3 | 2575 | 0.08603 | 0.2966 | 0.0176 | 0.06013 | 0.5397 |
| A1Anti_Breast_Mammary_Tissue | males | PBX3 | ENSG00000167081.16 | 1814 | 0.003181 | 0.1589 | 6.70e-04 | 0.03266 | 0.8046 |
| ABS_BASO_CT_Breast_Mammary_Tissue | females | HAUS4 | ENSG00000092036.18 | 2224 | 3.02e-31 | 0.2205 | 6.35e-32 | 0.04561 | 0.7339 |
| ABS_BASO_CT_Breast_Mammary_Tissue | females | MESP1 | ENSG00000166823.5 | 2120 | 3.39e-06 | 0.1006 | 2.66e-06 | 0.07804 | 0.8214 |
| ABS_BASO_CT_Breast_Mammary_Tissue | males | ZNF507 | ENSG00000168813.16 | 2142 | 0.04533 | 0.3024 | 0.01135 | 0.07514 | 0.5658 |
| ABS_BASO_CT_Breast_Mammary_Tissue | males | LINC01806 | ENSG00000227403.1 | 2079 | 6.41e-07 | 0.4215 | 1.08e-07 | 0.07067 | 0.5079 |
| ACAIgG_Breast_Mammary_Tissue | females | MCM8 | ENSG00000125885.13 | 2226 | 3.18e-10 | 0.3934 | 3.85e-11 | 0.04706 | 0.5596 |
| ACAIgG_Breast_Mammary_Tissue | females | CSTB | ENSG00000160213.6 | 1516 | 0.005827 | 0.3971 | 4.61e-04 | 0.03085 | 0.5658 |
| ACAIgG_Breast_Mammary_Tissue | females | RP11-529F4.1 | ENSG00000244932.2 | 1794 | 0.04215 | 0.05783 | 0.0282 | 0.03786 | 0.834 |
| ACAIgG_Breast_Mammary_Tissue | males | C19orf33 | ENSG00000167644.11 | 2053 | 2.71e-05 | 0.3677 | 9.00e-06 | 0.1218 | 0.5105 |
| ACAIgG_Breast_Mammary_Tissue | males | ODF2 | ENSG00000136811.16 | 831 | 0.002058 | 0.2335 | 3.12e-04 | 0.03472 | 0.7294 |
| ACTH_Breast_Mammary_Tissue | males | CNIH1 | ENSG00000100528.11 | 1966 | 1.46e-06 | 0.08126 | 1.39e-06 | 0.07682 | 0.8419 |
| ACTH_Breast_Mammary_Tissue | males | RP3-394A18.1 | ENSG00000279159.1 | 2169 | 2.23e-06 | 0.1303 | 1.35e-06 | 0.07843 | 0.7913 |

| trait | | gene_name | Locus_id | coloc_data_default_nsnps | coloc_data_default_PP.H0.abf | coloc_data_default_PP.H1.abf | coloc_data_default_PP.H2.abf | coloc_data_default_PP.H3.abf | coloc_data_default_PP.H4.abf |
|---|---|---|---|---|---|---|---|---|---|
| AFPTum_Breast_Mammary_Tissue | males | PITRM1-AS1 | ENSG00000237399.7 | 3243 | 1.34e-05 | 0.187 | 5.63e-06 | 0.07801 | 0.735 |
| AFPTum_Breast_Mammary_Tissue | males | SPATA13 | ENSG00000182957.15 | 3902 | 0.07971 | 0.2386 | 0.026 | 0.07725 | 0.5784 |
| AFPTum_Breast_Mammary_Tissue | males | HLA-DQB2 | ENSG00000232629.8 | 7068 | 4.07e-42 | 0.2268 | 2.93e-42 | 0.1627 | 0.6105 |
| Alb_Breast_Mammary_Tissue | females | LINC01137 | ENSG00000233621.1 | 1950 | 3.07e-05 | 0.2817 | 1.74e-06 | 0.01528 | 0.703 |
| Alb_Breast_Mammary_Tissue | females | RP11-254F7.2 | ENSG00000260077.1 | 2834 | 0.002355 | 0.1409 | 2.93e-04 | 0.01669 | 0.8398 |
| Alb_Breast_Mammary_Tissue | males | RP11-54A4.2 | ENSG00000237781.3 | 1306 | 4.11e-05 | 0.008265 | 0.001154 | 0.2314 | 0.7591 |
| AlkP_Breast_Mammary_Tissue | females | SMG1P7 | ENSG00000261556.9 | 1203 | 7.17e-17 | 0.1139 | 7.37e-18 | 0.01084 | 0.8753 |
| Amyl_Breast_Mammary_Tissue | males | SNX27 | ENSG00000143376.12 | 1565 | 8.28e-07 | 0.4503 | 7.98e-08 | 0.04291 | 0.5068 |
| AN.GAP_Breast_Mammary_Tissue | females | LINC01504 | ENSG00000225434.2 | 2174 | 1.99e-33 | 0.08625 | 1.06e-34 | 0.003659 | 0.9101 |
| BAbs_Breast_Mammary_Tissue | males | C17orf97 | ENSG00000187624.8 | 1306 | 1.81e-57 | 0.4081 | 1.05e-58 | 0.02325 | 0.5686 |
| BAbs_Breast_Mammary_Tissue | males | TBC1D25 | ENSG00000068354.15 | 1055 | 0.03342 | 0.1265 | 0.02907 | 0.1094 | 0.7016 |
| BASOAB_Breast_Mammary_Tissue | females | GCNT1P1 | ENSG00000236474.1 | 2791 | 0.004226 | 0.4276 | 3.86e-04 | 0.03855 | 0.5293 |
| BASOAB_Breast_Mammary_Tissue | females | ZKSCAN3 | ENSG00000189298.13 | 3084 | 0.004748 | 0.005766 | 0.1123 | 0.1356 | 0.7416 |
| BBTAb_Breast_Mammary_Tissue | males | CDC37P1 | ENSG00000259982.1 | 831 | 6.45e-04 | 0.163 | 3.21e-04 | 0.08049 | 0.7555 |
| BBTAb_Breast_Mammary_Tissue | males | KRT16 | ENSG00000186832.8 | 2260 | 0.004515 | 0.1117 | 0.001216 | 0.02922 | 0.8534 |
| BBTAb_Breast_Mammary_Tissue | males | AP001062.7 | ENSG00000184441.4 | 1603 | 1.20e-05 | 0.3088 | 1.37e-06 | 0.03481 | 0.6564 |
| Blasts_Breast_Mammary_Tissue | females | LINC00839 | ENSG00000185904.11 | 2396 | 6.69e-15 | 0.3732 | 1.63e-15 | 0.09034 | 0.5365 |
| BLOOD.PRESSURE_Breast_Mammary_Tissue | females | TRIM33 | ENSG00000197323.11 | 2091 | 0.01988 | 0.3387 | 0.001621 | 0.02702 | 0.6128 |

| trait | | gene_name | Locus_id | coloc_data_default_nsnps | coloc_data_default_PP.H0.abf | coloc_data_default_PP.H1.abf | coloc_data_default_PP.H2.abf | coloc_data_default_PP.H3.abf | coloc_data_default_PP.H4.abf |
|---|---|---|---|---|---|---|---|---|---|
| BLOOD.PRESSURE_Breast_Mammary_Tissue | females | LRRC37A15P | ENSG00000230069.3 | 2236 | 0.03282 | 0.06486 | 0.02499 | 0.04856 | 0.8288 |
| BLOOD.PRESSURE_Breast_Mammary_Tissue | males | TUBG2 | ENSG00000037042.8 | 1347 | 0.002802 | 0.3528 | 3.94e-04 | 0.04902 | 0.595 |
| BLOOD.PRESSURE_Breast_Mammary_Tissue | males | CYP4F11 | ENSG00000171903.16 | 2507 | 1.58e-05 | 0.4194 | 1.47e-06 | 0.03844 | 0.5421 |
| BLOOD.PRESSURE_Breast_Mammary_Tissue | males | PLA2G12A | ENSG00000123739.10 | 2103 | 2.18e-05 | 0.06597 | 9.06e-06 | 0.02658 | 0.9074 |
| BMI_CLEAN_Breast_Mammary_Tissue | males | CDC37P1 | ENSG00000259982.1 | 831 | 2.60e-05 | 0.006559 | 3.13e-04 | 0.07826 | 0.9148 |
| BMI_CLEAN_Breast_Mammary_Tissue | males | LMOD1 | ENSG00000163431.12 | 2033 | 0.1013 | 0.3015 | 0.01632 | 0.04804 | 0.5328 |
| BUN_Breast_Mammary_Tissue | males | RGCC | ENSG00000102760.12 | 2164 | 0.02042 | 0.2509 | 0.001233 | 0.01444 | 0.713 |
| BUN_Breast_Mammary_Tissue | males | RP11-331F4.5 | ENSG00000280152.1 | 1798 | 0.005729 | 0.004948 | 0.0265 | 0.02194 | 0.9409 |
| C.Gluc_Breast_Mammary_Tissue | females | IGHV3-64D | ENSG00000282639.1 | 623 | 5.23e-10 | 0.4624 | 2.76e-11 | 0.0239 | 0.5137 |
| C.Gluc_Breast_Mammary_Tissue | females | IGHV5-10-1 | ENSG00000282651.2 | 591 | 8.06e-15 | 0.3947 | 4.18e-16 | 0.01988 | 0.5854 |
| C.Gluc_Breast_Mammary_Tissue | females | URB1 | ENSG00000142207.6 | 2430 | 0.04928 | 0.2062 | 0.03709 | 0.1546 | 0.5528 |
| C4Quan_Breast_Mammary_Tissue | males | SOHLH2 | ENSG00000120669.15 | 2367 | 4.41e-04 | 0.1561 | 4.20e-04 | 0.1479 | 0.6951 |
| C4Quan_Breast_Mammary_Tissue | males | ZKSCAN3 | ENSG00000189298.13 | 3084 | 4.55e-05 | 5.74e-04 | 0.02587 | 0.3257 | 0.6478 |
| Ca_Breast_Mammary_Tissue | females | TSTD1 | ENSG00000215845.10 | 1786 | 5.02e-04 | 0.4463 | 3.56e-05 | 0.03112 | 0.522 |
| Ca_Breast_Mammary_Tissue | males | ZKSCAN3 | ENSG00000189298.13 | 3084 | 0.007173 | 0.09051 | 0.01963 | 0.247 | 0.6357 |
| CBil_Breast_Mammary_Tissue | females | ZNF208 | ENSG00000160321.14 | 3056 | 0.008347 | 0.3719 | 0.00228 | 0.101 | 0.5165 |
| CBil_Breast_Mammary_Tissue | males | ZNF660 | ENSG00000144792.9 | 1981 | 4.96e-06 | 0.2917 | 8.72e-07 | 0.05059 | 0.6577 |

| trait | | gene_name | Locus_id | coloc_data_default_nsnps | coloc_data_default_PP.H0.abf | coloc_data_default_PP.H1.abf | coloc_data_default_PP.H2.abf | coloc_data_default_PP.H3.abf | coloc_data_default_PP.H4.abf |
|---|---|---|---|---|---|---|---|---|---|
| CBil_Breast_Mammary_Tissue | males | PBX3 | ENSG00000167081.16 | 1814 | 0.004486 | 0.2241 | 4.78e-04 | 0.02313 | 0.7478 |
| Chol_Breast_Mammary_Tissue | females | TBKBP1 | ENSG00000198933.9 | 2461 | 3.08e-05 | 0.02471 | 2.86e-04 | 0.2285 | 0.7465 |
| Chol_Breast_Mammary_Tissue | males | GGA3 | ENSG00000125447.16 | 1354 | 0.01202 | 0.442 | 8.92e-04 | 0.03228 | 0.5129 |
| Cl_Breast_Mammary_Tissue | females | CDCA7L | ENSG00000164649.19 | 3266 | 0.006396 | 0.006435 | 0.05667 | 0.05615 | 0.8743 |
| Cl_Breast_Mammary_Tissue | males | CTD-3080P12.3 | ENSG00000249201.2 | 2752 | 1.67e-15 | 0.4577 | 7.88e-17 | 0.02104 | 0.5212 |
| CLymph_Breast_Mammary_Tissue | females | LINC01481 | ENSG00000257815.5 | 3130 | 5.19e-05 | 0.1138 | 4.49e-05 | 0.09762 | 0.7885 |
| CLymph_Breast_Mammary_Tissue | females | CAPN12 | ENSG00000182472.8 | 2192 | 0.001507 | 0.251 | 4.29e-04 | 0.07082 | 0.6762 |
| CLymph_Breast_Mammary_Tissue | males | JAK3 | ENSG00000105639.18 | 1740 | 0.06522 | 0.3222 | 0.01195 | 0.05851 | 0.5421 |
| CNeut_Breast_Mammary_Tissue | females | AF011889.2 | ENSG00000238039.1 | 1158 | 6.19e-23 | 0.4497 | 6.21e-24 | 0.04466 | 0.5056 |
| trait | | gene_name | Locus_id | coloc_data_default_nsnps | coloc_data_default_PP.H0.abf | coloc_data_default_PP.H1.abf | coloc_data_default_PP.H2.abf | coloc_data_default_PP.H3.abf | coloc_data_default_PP.H4.abf |
| CNeut_Breast_Mammary_Tissue | males | CHRAC1 | ENSG00000104472.9 | 2056 | 9.84e-07 | 0.4415 | 1.20e-07 | 0.05334 | 0.5051 |
| CNucCl_Breast_Mammary_Tissue | females | CAPN12 | ENSG00000182472.8 | 2192 | 0.002464 | 0.4103 | 5.07e-04 | 0.08398 | 0.5028 |
| CNucCl_Breast_Mammary_Tissue | females | HAUS6 | ENSG00000147874.10 | 2796 | 0.00291 | 0.05437 | 0.02136 | 0.3985 | 0.5228 |
| CNucCl_Breast_Mammary_Tissue | males | AMZ2P1 | ENSG00000214174.8 | 1043 | 4.48e-04 | 0.3097 | 2.69e-05 | 0.01792 | 0.6719 |
| CO2_Breast_Mammary_Tissue | females | RP11-1099M24.6 | ENSG00000280046.1 | 1427 | 0.01679 | 0.02299 | 0.01437 | 0.01875 | 0.9271 |
| CPepti_Breast_Mammary_Tissue | females | LINC01806 | ENSG00000227403.1 | 2079 | 1.91e-14 | 0.3806 | 5.59e-15 | 0.1106 | 0.5088 |
| CPepti_Breast_Mammary_Tissue | males | ASAH2 | ENSG00000188611.14 | 1670 | 1.40e-05 | 0.2017 | 3.71e-06 | 0.05258 | 0.7458 |
| CPepti_Breast_Mammary_Tissue | males | ANKRD36 | ENSG00000135976.17 | 693 | 8.34e-06 | 0.1216 | 4.21e-06 | 0.06064 | 0.8178 |
| CPepti_Breast_Mammary_Tissue | males | ZAP70 | ENSG00000115085.13 | 1104 | 0.1788 | 0.118 | 0.0965 | 0.06312 | 0.5436 |

| trait | | gene_name | Locus_id | coloc_data_default_nsnps | coloc_data_default_PP.H0.abf | coloc_data_default_PP.H1.abf | coloc_data_default_PP.H2.abf | coloc_data_default_PP.H3.abf | coloc_data_default_PP.H4.abf |
|---|---|---|---|---|---|---|---|---|---|
| CPK_Breast_Mammary_Tissue | females | MRE11 | ENSG00000020922.12 | 1966 | 3.54e-61 | 0.4003 | 3.93e-62 | 0.04389 | 0.5559 |
| CPK_Breast_Mammary_Tissue | females | PGPEP1 | ENSG00000130517.13 | 1515 | 1.67e-04 | 0.07508 | 1.01e-05 | 0.003614 | 0.9211 |
| CPK_Breast_Mammary_Tissue | females | ALMS1P1 | ENSG00000163016.9 | 1310 | 5.80e-11 | 0.39 | 1.27e-11 | 0.08459 | 0.5254 |
| CPK_Breast_Mammary_Tissue | females | PIGG | ENSG00000174227.15 | 1797 | 0.01682 | 0.2612 | 0.004604 | 0.07083 | 0.6466 |
| CPK_Breast_Mammary_Tissue | males | LDHC | ENSG00000166796.11 | 2464 | 3.38e-07 | 0.4358 | 3.75e-08 | 0.04783 | 0.5164 |
| CPK_Breast_Mammary_Tissue | males | RP11-507B12.1 | ENSG00000259675.1 | 2421 | 2.87e-07 | 0.2611 | 3.56e-08 | 0.03171 | 0.7072 |
| CRBC_Breast_Mammary_Tissue | females | RP5-1017F8.2 | ENSG00000276093.1 | 1667 | 7.99e-10 | 0.1589 | 9.73e-11 | 0.01852 | 0.8226 |
| CRBC_Breast_Mammary_Tissue | males | ASAH2 | ENSG00000188611.14 | 1670 | 9.56e-06 | 0.1374 | 2.37e-06 | 0.03324 | 0.8293 |
| Creat_Breast_Mammary_Tissue | females | CTD-2651B20.3 | ENSG00000259520.5 | 1308 | 2.94e-15 | 0.3318 | 1.01e-15 | 0.1131 | 0.5552 |
| Creat_Breast_Mammary_Tissue | males | FAM174B | ENSG00000185442.12 | 2700 | 4.42e-06 | 0.05764 | 2.03e-06 | 0.02553 | 0.9168 |
| Creat_Breast_Mammary_Tissue | males | LINC01060 | ENSG00000249378.5 | 4336 | 7.49e-12 | 0.2537 | 2.78e-12 | 0.09371 | 0.6526 |
| CRP_Breast_Mammary_Tissue | females | NKTR | ENSG00000114857.17 | 1970 | 1.34e-12 | 0.4473 | 1.23e-13 | 0.04041 | 0.5123 |
| D25OHT_Breast_Mammary_Tissue | females | LY75 | ENSG00000054219.10 | 2863 | 1.13e-07 | 0.301 | 6.22e-08 | 0.1646 | 0.5344 |
| DirLDL_Breast_Mammary_Tissue | males | RP11-365P13.5 | ENSG00000277767.1 | 2862 | 5.42e-05 | 0.02426 | 4.97e-05 | 0.02127 | 0.9544 |
| DirLDL_Breast_Mammary_Tissue | males | RGCC | ENSG00000102760.12 | 2164 | 0.01396 | 0.1715 | 0.001709 | 0.0202 | 0.7926 |
| DirLDL_Breast_Mammary_Tissue | males | LMOD1 | ENSG00000163431.12 | 2033 | 0.06556 | 0.195 | 0.01479 | 0.04332 | 0.6813 |
| DLCOOVA_PRD_Breast_Mammary_Tissue | females | GSTM4 | ENSG00000168765.16 | 2007 | 0.03399 | 0.1763 | 0.003554 | 0.01767 | 0.7685 |
| EGFR_Breast_Mammary_Tissue | females | CTD-2651B20.3 | ENSG00000259520.5 | 1308 | 1.68e-17 | 0.001894 | 1.69e-15 | 0.1902 | 0.8079 |

| trait | | gene_name | Locus_id | coloc_data_default_nsnps | coloc_data_default_PP.H0.abf | coloc_data_default_PP.H1.abf | coloc_data_default_PP.H2.abf | coloc_data_default_PP.H3.abf | coloc_data_default_PP.H4.abf |
|---|---|---|---|---|---|---|---|---|---|
| EGFR_Breast_Mammary_Tissue | males | FAM174B | ENSG00000185442.12 | 2700 | 1.34e-05 | 0.1747 | 4.31e-06 | 0.05543 | 0.7698 |
| EGFR_Breast_Mammary_Tissue | males | SPAG7 | ENSG00000091640.7 | 2929 | 0.004844 | 0.4307 | 5.31e-04 | 0.04667 | 0.5173 |
| EGFR_Breast_Mammary_Tissue | males | LINC01060 | ENSG00000249378.5 | 4336 | 7.29e-12 | 0.2469 | 1.91e-12 | 0.06409 | 0.689 |
| EGFR_Breast_Mammary_Tissue | males | PTPN12 | ENSG00000127947.15 | 2756 | 0.03641 | 0.04413 | 0.1498 | 0.1809 | 0.5888 |
| EoAbs_Breast_Mammary_Tissue | females | GOLGA2P10 | ENSG00000255769.7 | 1590 | 0.001958 | 0.3607 | 4.89e-04 | 0.08953 | 0.5473 |
| EoAbs_Breast_Mammary_Tissue | males | LINC00639 | ENSG00000259070.5 | 2646 | 0.05264 | 0.1993 | 0.01159 | 0.04317 | 0.6933 |
| Eos_Breast_Mammary_Tissue | females | GOLGA2P10 | ENSG00000255769.7 | 1590 | 0.001193 | 0.2199 | 0.001243 | 0.2285 | 0.5492 |
| Eos_Breast_Mammary_Tissue | females | MAATS1 | ENSG00000183833.16 | 2830 | 6.57e-11 | 0.3601 | 1.10e-11 | 0.0597 | 0.5802 |
| Eos_Breast_Mammary_Tissue | males | LINC00639 | ENSG00000259070.5 | 2646 | 0.05716 | 0.2164 | 0.01164 | 0.04339 | 0.6714 |
| EOSIAB_Breast_Mammary_Tissue | females | WDR6 | ENSG00000178252.17 | 1255 | 1.96e-11 | 0.191 | 6.26e-12 | 0.06032 | 0.7487 |
| EOSIAB_Breast_Mammary_Tissue | females | LINC01504 | ENSG00000225434.2 | 2174 | 1.02e-32 | 0.4396 | 1.39e-33 | 0.0596 | 0.5008 |
| EOSIM_Breast_Mammary_Tissue | females | KRT6C | ENSG00000170465.9 | 2329 | 0.00132 | 0.1186 | 0.001842 | 0.1648 | 0.7135 |
| EOSIM_Breast_Mammary_Tissue | females | KRT6A | ENSG00000205420.10 | 2308 | 5.96e-05 | 0.09768 | 8.32e-05 | 0.1355 | 0.7666 |
| EOSIM_Breast_Mammary_Tissue | males | WDR6 | ENSG00000178252.17 | 1255 | 7.27e-16 | 0.04913 | 1.96e-15 | 0.1317 | 0.8192 |
| EOSIRE_Breast_Mammary_Tissue | females | WDR6 | ENSG00000178252.17 | 1255 | 4.14e-11 | 0.4043 | 5.57e-12 | 0.0538 | 0.5419 |
| FEF25_75_PPR_Breast_Mammary_Tissue | females | PLCB2 | ENSG00000137841.11 | 1865 | 0.004188 | 0.4376 | 4.22e-04 | 0.04362 | 0.5142 |
| FEF25_75_PPR_Breast_Mammary_Tissue | females | SPAG7 | ENSG00000091640.7 | 2929 | 0.001302 | 0.2577 | 4.61e-04 | 0.09057 | 0.65 |
| FEF25_75_PPR_Breast_Mammary_Tissue | females | NAPSB | ENSG00000131401.11 | 1314 | 3.35e-04 | 0.3648 | 5.23e-05 | 0.05629 | 0.5785 |

| trait | | gene_name | Locus_id | coloc_data_default_nsnps | coloc_data_default_PP.H0.abf | coloc_data_default_PP.H1.abf | coloc_data_default_PP.H2.abf | coloc_data_default_PP.H3.abf | coloc_data_default_PP.H4.abf |
|---|---|---|---|---|---|---|---|---|---|
| FEF25_75_PPR_Breast_Mammary_Tissue | females | SPC25 | ENSG00000152253.8 | 2460 | 2.38e-06 | 0.2296 | 5.66e-07 | 0.05395 | 0.7164 |
| FEF25_75_PPR_Breast_Mammary_Tissue | males | PDZD8 | ENSG00000165650.11 | 1359 | 1.10e-08 | 0.4479 | 1.05e-09 | 0.04214 | 0.5099 |
| FEF25_75_PRD_Breast_Mammary_Tissue | females | FAM53B | ENSG00000189319.13 | 2284 | 0.04466 | 0.2598 | 0.006859 | 0.03927 | 0.6494 |
| FEF25_75_PRD_Breast_Mammary_Tissue | males | ATP8A1 | ENSG00000124406.16 | 2891 | 2.23e-04 | 0.3737 | 4.52e-05 | 0.07528 | 0.5508 |
| FEF25_75_PRL_Breast_Mammary_Tissue | females | TAS2R20 | ENSG00000255837.1 | 2349 | 1.52e-11 | 0.2044 | 3.73e-12 | 0.04952 | 0.7461 |
| FEF25_75_PRL_Breast_Mammary_Tissue | females | CRYBB3 | ENSG00000100053.9 | 1319 | 0.01028 | 0.06876 | 0.005947 | 0.0389 | 0.8761 |
| FEF25_75_PRL_Breast_Mammary_Tissue | males | LSR | ENSG00000105699.16 | 1748 | 0.001073 | 0.2233 | 1.54e-04 | 0.03121 | 0.7442 |
| FEF25_75_PRL_Breast_Mammary_Tissue | males | ATP8A1 | ENSG00000124406.16 | 2891 | 1.52e-05 | 0.02554 | 6.85e-06 | 0.01054 | 0.9639 |
| FEV1_PRD_Breast_Mammary_Tissue | females | LDHC | ENSG00000166796.11 | 2464 | 1.63e-07 | 0.4094 | 2.83e-08 | 0.07071 | 0.5199 |
| FEV1_PRD_Breast_Mammary_Tissue | females | HEATR6 | ENSG00000068097.14 | 725 | 3.32e-04 | 0.1239 | 6.26e-05 | 0.02248 | 0.8533 |
| FEV1_PRD_Breast_Mammary_Tissue | females | RP3-465N24.5 | ENSG00000261349.1 | 1535 | 0.005479 | 0.2314 | 6.65e-04 | 0.02734 | 0.7351 |
| FEV1_PRD_Breast_Mammary_Tissue | females | SDHDP6 | ENSG00000224183.1 | 1568 | 3.44e-04 | 0.1868 | 4.31e-05 | 0.02258 | 0.7902 |
| FEV1_PRD_Breast_Mammary_Tissue | females | WDR6 | ENSG00000178252.17 | 1255 | 2.06e-11 | 0.2014 | 1.08e-11 | 0.1046 | 0.694 |
| FEV1_PRD_Breast_Mammary_Tissue | females | CTC-487M23.6 | ENSG00000279522.1 | 3325 | 5.70e-05 | 0.07434 | 1.02e-04 | 0.1324 | 0.7931 |
| FEV1_PRD_Breast_Mammary_Tissue | males | RPL8 | ENSG00000161016.17 | 442 | 8.81e-33 | 0.4645 | 5.96e-34 | 0.03092 | 0.5045 |
| trait | | gene_name | Locus_id | coloc_data_default_nsnps | coloc_data_default_PP.H0.abf | coloc_data_default_PP.H1.abf | coloc_data_default_PP.H2.abf | coloc_data_default_PP.H3.abf | coloc_data_default_PP.H4.abf |
| FEV1OFVC_AFD_Breast_Mammary_Tissue | females | RP11-44F14.7 | ENSG00000279344.1 | 1900 | 4.93e-18 | 0.1628 | 1.13e-18 | 0.03645 | 0.8008 |
| FEV1OFVC_AFD_Breast_Mammary_Tissue | males | HAUS4 | ENSG00000092036.18 | 2224 | 5.79e-21 | 0.2563 | 1.37e-21 | 0.05983 | 0.6839 |

| trait | | gene_name | Locus_id | coloc_data_default_nsnps | coloc_data_default_PP.H0.abf | coloc_data_default_PP.H1.abf | coloc_data_default_PP.H2.abf | coloc_data_default_PP.H3.abf | coloc_data_default_PP.H4.abf |
|---|---|---|---|---|---|---|---|---|---|
| FEV1OFVC_PPR_Breast_Mammary_Tissue | females | RBM26-AS1 | ENSG00000227354.6 | 2033 | 6.62e-17 | 0.2483 | 2.23e-17 | 0.08317 | 0.6685 |
| FEV1OFVC_PRD_Breast_Mammary_Tissue | males | ATP8A1 | ENSG00000124406.16 | 2891 | 2.02e-04 | 0.3387 | 6.78e-05 | 0.1134 | 0.5476 |
| FEV1OFVC_PRL_Breast_Mammary_Tissue | females | GTF2A1L | ENSG00000242441.7 | 3325 | 5.31e-58 | 0.3088 | 1.14e-58 | 0.06549 | 0.6257 |
| FEV1OFVC_PRL_Breast_Mammary_Tissue | females | QRSL1 | ENSG00000130348.11 | 2326 | 1.19e-20 | 0.1125 | 4.91e-21 | 0.04544 | 0.842 |
| FEV1OFVC_PRL_Breast_Mammary_Tissue | females | CHRAC1 | ENSG00000104472.9 | 2056 | 2.50e-12 | 0.0803 | 3.70e-13 | 0.011 | 0.9087 |
| FEV1OFVC_PRL_Breast_Mammary_Tissue | males | MRI1 | ENSG00000037757.13 | 1197 | 0.07146 | 0.1351 | 0.007657 | 0.01371 | 0.772 |
| FEV1OFVC_PRL_Breast_Mammary_Tissue | males | LSR | ENSG00000105699.16 | 1748 | 0.002059 | 0.4287 | 2.95e-04 | 0.061 | 0.508 |
| FEV1OFVC_PRL_Breast_Mammary_Tissue | males | ATP8A1 | ENSG00000124406.16 | 2891 | 2.30e-04 | 0.3855 | 6.16e-05 | 0.1029 | 0.5113 |
| FEV1OFVC_PRL_Breast_Mammary_Tissue | males | ELP3 | ENSG00000134014.16 | 2448 | 3.70e-07 | 0.3962 | 7.02e-08 | 0.07473 | 0.5291 |
| FEV1OFVC_ZBD_Breast_Mammary_Tissue | females | RBM26-AS1 | ENSG00000227354.6 | 2033 | 6.26e-17 | 0.2348 | 2.35e-17 | 0.08732 | 0.6779 |
| FProt1_Breast_Mammary_Tissue | females | IGKV2-29 | ENSG00000253998.3 | 635 | 0.002167 | 0.2841 | 1.92e-04 | 0.02444 | 0.6891 |
| FProt1_Breast_Mammary_Tissue | females | POMT1 | ENSG00000130714.15 | 1514 | 1.38e-04 | 0.1455 | 4.02e-05 | 0.04153 | 0.8128 |
| FProt1_Breast_Mammary_Tissue | males | GTF2A1L | ENSG00000242441.7 | 3325 | 5.67e-60 | 0.2923 | 1.44e-60 | 0.07369 | 0.634 |
| FProt1_Breast_Mammary_Tissue | males | TBC1D25 | ENSG00000068354.15 | 1055 | 0.09492 | 0.3594 | 0.008211 | 0.03058 | 0.5069 |
| FRONTAL.AXIS.P_Breast_Mammary_Tissue | females | PLEC | ENSG00000178209.14 | 1326 | 3.20e-17 | 0.08261 | 1.29e-17 | 0.03235 | 0.885 |
| FRONTAL.AXIS.P_Breast_Mammary_Tissue | males | ANKRD36 | ENSG00000135976.17 | 693 | 1.06e-06 | 0.01553 | 3.34e-06 | 0.04778 | 0.9367 |
| FRONTAL.AXIS.P_Breast_Mammary_Tissue | males | MUC20 | ENSG00000176945.16 | 2192 | 0.003697 | 0.4108 | 7.09e-04 | 0.07828 | 0.5065 |

| trait | | gene_name | Locus_id | coloc_data_default_nsnps | coloc_data_default_PP.H0.abf | coloc_data_default_PP.H1.abf | coloc_data_default_PP.H2.abf | coloc_data_default_PP.H3.abf | coloc_data_default_PP.H4.abf |
|---|---|---|---|---|---|---|---|---|---|
| FRONTAL.AXIS.TERMINAL.40.MS_Breast_Mammary_Tissue | females | PPIL2 | ENSG00000100023.18 | 1419 | 1.72e-05 | 0.4374 | 1.45e-06 | 0.03618 | 0.5264 |
| FRONTAL.AXIS.TERMINAL.40.MS_Breast_Mammary_Tissue | females | AF011889.2 | ENSG00000238039.1 | 1158 | 4.08e-23 | 0.2967 | 4.41e-24 | 0.03141 | 0.6719 |
| FRONTAL.AXIS.TERMINAL.40.MS_Breast_Mammary_Tissue | males | CNST | ENSG00000162852.13 | 2941 | 0.002566 | 0.3162 | 8.23e-04 | 0.1008 | 0.5796 |
| FT4_Breast_Mammary_Tissue | males | SCAND2P | ENSG00000176700.19 | 2394 | 0.003852 | 0.3066 | 5.79e-04 | 0.04546 | 0.6435 |
| FVC_PRD_Breast_Mammary_Tissue | females | LDHC | ENSG00000166796.11 | 2464 | 1.70e-07 | 0.4284 | 2.85e-08 | 0.07114 | 0.5005 |
| FVC_PRD_Breast_Mammary_Tissue | females | HEATR6 | ENSG00000068097.14 | 725 | 2.70e-04 | 0.1008 | 5.80e-05 | 0.02074 | 0.8782 |
| FVC_PRD_Breast_Mammary_Tissue | females | RP3-465N24.5 | ENSG00000261349.1 | 1535 | 0.006245 | 0.2637 | 7.39e-04 | 0.03051 | 0.6988 |
| FVC_PRD_Breast_Mammary_Tissue | females | SDHDP6 | ENSG00000224183.1 | 1568 | 3.99e-04 | 0.2165 | 4.86e-05 | 0.02564 | 0.7575 |
| FVC_PRD_Breast_Mammary_Tissue | females | WDR6 | ENSG00000178252.17 | 1255 | 2.17e-11 | 0.2115 | 1.07e-11 | 0.1034 | 0.6851 |
| FVC_PRD_Breast_Mammary_Tissue | females | CTC-487M23.6 | ENSG00000279522.1 | 3325 | 4.82e-05 | 0.06288 | 9.55e-05 | 0.1238 | 0.8131 |
| GluBed_Breast_Mammary_Tissue | females | HLA-DQB2 | ENSG00000232629.8 | 7068 | 9.49e-26 | 0.05127 | 1.46e-25 | 0.07808 | 0.8706 |
| GluBed_Breast_Mammary_Tissue | females | IL1RAPL2 | ENSG00000189108.12 | 890 | 0.002373 | 0.1127 | 6.79e-04 | 0.03139 | 0.8529 |
| GluBed_Breast_Mammary_Tissue | males | SHANK3 | ENSG00000251322.7 | 1269 | 0.1125 | 0.1794 | 0.07549 | 0.1199 | 0.5127 |
| Gluc_Breast_Mammary_Tissue | females | SPTBN5 | ENSG00000137877.9 | 1790 | 7.53e-05 | 0.2139 | 2.05e-05 | 0.05758 | 0.7284 |
| Haptgl_Breast_Mammary_Tissue | females | RP11-643A5.3 | ENSG00000280362.1 | 3468 | 0.001658 | 0.161 | 8.29e-04 | 0.07974 | 0.7567 |
| Haptgl_Breast_Mammary_Tissue | females | MAG | ENSG00000105695.14 | 1752 | 4.33e-08 | 0.3007 | 4.18e-09 | 0.02834 | 0.671 |

| trait | | gene_name | Locus_id | coloc_data_default_nsnps | coloc_data_default_PP.H0.abf | coloc_data_default_PP.H1.abf | coloc_data_default_PP.H2.abf | coloc_data_default_PP.H3.abf | coloc_data_default_PP.H4.abf |
|---|---|---|---|---|---|---|---|---|---|
| Haptgl_Breast_Mammary_Tissue | females | CD22 | ENSG00000012124.16 | 1738 | 1.54e-07 | 0.301 | 1.48e-08 | 0.02812 | 0.6708 |
| Haptgl_Breast_Mammary_Tissue | females | FFAR1 | ENSG00000126266.3 | 1748 | 0.1065 | 0.3101 | 0.0102 | 0.02916 | 0.5441 |
| Haptgl_Breast_Mammary_Tissue | males | RNF183 | ENSG00000165188.13 | 2637 | 0.02427 | 0.1278 | 0.007655 | 0.03951 | 0.8008 |
| HBSA._Breast_Mammary_Tissue | females | IGHV5-10-1 | ENSG00000282651.2 | 591 | 9.69e-15 | 0.4746 | 4.50e-16 | 0.02151 | 0.5039 |
| HBSA._Breast_Mammary_Tissue | females | HLA-DQA2 | ENSG00000237541.3 | 7023 | 1.70e-24 | 0.119 | 2.40e-24 | 0.1673 | 0.7137 |
| HBSA._Breast_Mammary_Tissue | females | POMT1 | ENSG00000130714.15 | 1514 | 2.78e-04 | 0.2927 | 7.94e-05 | 0.083 | 0.6239 |
| HDL.C_Breast_Mammary_Tissue | females | ASB4 | ENSG00000005981.12 | 2149 | 4.23e-05 | 0.06696 | 1.85e-05 | 0.02836 | 0.9046 |
| Height_Breast_Mammary_Tissue | females | FGFR3 | ENSG00000068078.17 | 2034 | 0.001951 | 0.009658 | 0.01121 | 0.05457 | 0.9226 |
| HEIGHT_Breast_Mammary_Tissue | females | FAM53B | ENSG00000189319.13 | 2284 | 0.04813 | 0.2801 | 0.01886 | 0.1092 | 0.5437 |
| Height_Breast_Mammary_Tissue | males | AP001062.7 | ENSG00000184441.4 | 1603 | 1.13e-05 | 0.2915 | 3.14e-07 | 0.007412 | 0.7011 |
| Height_Breast_Mammary_Tissue | males | TMEM165 | ENSG00000134851.12 | 2439 | 0.002478 | 0.1969 | 0.002113 | 0.1673 | 0.6312 |
| Height_Breast_Mammary_Tissue | males | MTHFD1P1 | ENSG00000231831.1 | 1055 | 1.06e-04 | 0.05166 | 8.15e-04 | 0.3964 | 0.551 |
| HgbA1C_Breast_Mammary_Tissue | females | RP11-274B21.13 | ENSG00000281896.1 | 2208 | 2.14e-23 | 0.4219 | 3.51e-24 | 0.06886 | 0.5093 |
| HgbA1C_Breast_Mammary_Tissue | females | RP11-274B21.2 | ENSG00000243302.3 | 2233 | 1.14e-22 | 0.4088 | 1.88e-23 | 0.06687 | 0.5243 |
| HgbA1C_Breast_Mammary_Tissue | females | RP11-274B21.4 | ENSG00000230715.3 | 2233 | 1.36e-38 | 0.4009 | 2.25e-39 | 0.06554 | 0.5336 |
| HORIZONTAL.AXIS..INITIAL.40.MS_Breast_Mammary_Tissue | males | AC005682.5 | ENSG00000228649.8 | 2697 | 5.68e-06 | 0.4466 | 6.24e-07 | 0.0485 | 0.5049 |

| trait | | gene_name | Locus_id | coloc_data_default_nsnps | coloc_data_default_PP.H0.abf | coloc_data_default_PP.H1.abf | coloc_data_default_PP.H2.abf | coloc_data_default_PP.H3.abf | coloc_data_default_PP.H4.abf |
|---|---|---|---|---|---|---|---|---|---|
| HORIZONTAL.AXIS..TERMINAL.40.MS_Breast_Mammary_Tissue | males | LINC00639 | ENSG00000259070.5 | 2646 | 0.07479 | 0.2831 | 0.0277 | 0.1043 | 0.51 |
| HR_INTERVAL_Breast_Mammary_Tissue | females | RP11-254F7.2 | ENSG00000260077.1 | 2834 | 0.002357 | 0.141 | 0.001691 | 0.1004 | 0.7545 |
| HSCRP_Breast_Mammary_Tissue | females | LYNX1 | ENSG00000180155.19 | 2535 | 3.11e-31 | 0.2259 | 8.46e-32 | 0.06071 | 0.7134 |
| HSCRP_Breast_Mammary_Tissue | males | DYNLL2 | ENSG00000264364.2 | 1819 | 0.174 | 0.1659 | 0.02689 | 0.02503 | 0.6081 |
| IctIdx_Breast_Mammary_Tissue | males | SCAND2P | ENSG00000176700.19 | 2394 | 5.59e-04 | 0.04445 | 2.80e-04 | 0.02133 | 0.9334 |
| IGAB_Breast_Mammary_Tissue | females | IGKV2-29 | ENSG00000253998.3 | 635 | 0.002567 | 0.3366 | 1.04e-04 | 0.013 | 0.6477 |
| IGAB_Breast_Mammary_Tissue | males | ZKSCAN3 | ENSG00000189298.13 | 3084 | 0.004308 | 0.05436 | 0.02271 | 0.2859 | 0.6327 |
| IgM.Q_Breast_Mammary_Tissue | females | PRKCB | ENSG00000166501.12 | 1836 | 0.08988 | 0.2076 | 0.01948 | 0.04434 | 0.6387 |
| IgM.Q_Breast_Mammary_Tissue | females | FAM8A1 | ENSG00000137414.5 | 2626 | 0.04092 | 0.3359 | 0.008657 | 0.07051 | 0.544 |
| IgM.Q_Breast_Mammary_Tissue | females | LINC00265 | ENSG00000188185.11 | 2245 | 0.01206 | 0.3908 | 0.002067 | 0.06643 | 0.5286 |
| IGRE_Breast_Mammary_Tissue | females | IGKV2-29 | ENSG00000253998.3 | 635 | 0.002505 | 0.3285 | 1.58e-04 | 0.0201 | 0.6488 |
| IGRE_Breast_Mammary_Tissue | females | RPL8 | ENSG00000161016.17 | 442 | 4.28e-07 | 0.4178 | 3.19e-08 | 0.03059 | 0.5516 |
| IRF_Breast_Mammary_Tissue | females | ASB4 | ENSG00000005981.12 | 2149 | 1.28e-04 | 0.2017 | 3.80e-05 | 0.0594 | 0.7388 |
| IRF_Breast_Mammary_Tissue | males | RP11-831A10.1 | ENSG00000255216.1 | 2109 | 0.0602 | 0.2269 | 0.01017 | 0.03765 | 0.6651 |
| KapLCQ_Breast_Mammary_Tissue | males | DNAH6 | ENSG00000115423.18 | 2319 | 5.06e-07 | 0.2284 | 1.14e-07 | 0.05065 | 0.721 |
| LamLCQ_Breast_Mammary_Tissue | males | URB1 | ENSG00000142207.6 | 2430 | 0.04791 | 0.1397 | 0.03957 | 0.1147 | 0.6581 |
| LamLCQ_Breast_Mammary_Tissue | males | SMARCB1 | ENSG00000099956.18 | 1300 | 1.27e-06 | 0.05489 | 2.83e-06 | 0.1213 | 0.8238 |

| trait | | gene_name | Locus_id | coloc_data_default_nsnps | coloc_data_default_PP.H0.abf | coloc_data_default_PP.H1.abf | coloc_data_default_PP.H2.abf | coloc_data_default_PP.H3.abf | coloc_data_default_PP.H4.abf |
|---|---|---|---|---|---|---|---|---|---|
| LDL.C_Breast_Mammary_Tissue | females | SMG1P7 | ENSG00000261556.9 | 1203 | 2.38e-16 | 0.3784 | 1.07e-17 | 0.01642 | 0.6052 |
| LDL.C_Breast_Mammary_Tissue | females | TBKBP1 | ENSG00000198933.9 | 2461 | 1.28e-04 | 0.1023 | 1.78e-04 | 0.1422 | 0.7552 |
| LDL.C_Breast_Mammary_Tissue | females | HLA-DQB2 | ENSG00000232629.8 | 7068 | 1.06e-25 | 0.05705 | 7.27e-25 | 0.3924 | 0.5506 |
| LDL.C_Breast_Mammary_Tissue | males | RP11-54A4.2 | ENSG00000237781.3 | 1306 | 3.80e-04 | 0.07641 | 0.001015 | 0.2036 | 0.7186 |
| LipIdx_Breast_Mammary_Tissue | males | SOHLH2 | ENSG00000120669.15 | 2367 | 3.03e-04 | 0.1071 | 4.00e-04 | 0.1409 | 0.7513 |
| LipIdx_Breast_Mammary_Tissue | males | C14orf37 | ENSG00000139971.15 | 2673 | 0.001043 | 0.3728 | 2.95e-04 | 0.1048 | 0.521 |
| LipIdx_Breast_Mammary_Tissue | males | CTSH | ENSG00000103811.15 | 2708 | 5.00e-08 | 0.2771 | 7.24e-09 | 0.03941 | 0.6835 |
| Luc_percent__Breast_Mammary_Tissue | females | GXYLT1P5 | ENSG00000276591.1 | 2 | 2.92e-04 | 0.4444 | 3.77e-07 | 1.74e-05 | 0.5553 |
| Luc_percent__Breast_Mammary_Tissue | females | RP11-459O16.8 | ENSG00000276128.1 | 2 | 1.82e-09 | 0.4445 | 2.35e-12 | 1.74e-05 | 0.5555 |
| LucAbs_Breast_Mammary_Tissue | females | TUBBP5 | ENSG00000159247.13 | 320 | 3.54e-11 | 0.3119 | 1.09e-12 | 0.008951 | 0.6791 |
| LucAbs_Breast_Mammary_Tissue | males | HIBCH | ENSG00000198130.15 | 1823 | 4.00e-13 | 0.3645 | 5.51e-14 | 0.04967 | 0.5858 |
| LymAbs_Breast_Mammary_Tissue | males | C17orf97 | ENSG00000187624.8 | 1306 | 1.38e-57 | 0.3122 | 7.79e-59 | 0.01694 | 0.6708 |
| LymAbs_Breast_Mammary_Tissue | males | ZKSCAN3 | ENSG00000189298.13 | 3084 | 0.01315 | 0.1659 | 0.0234 | 0.2948 | 0.5027 |
| LYMPAB_Breast_Mammary_Tissue | males | ZKSCAN3 | ENSG00000189298.13 | 3084 | 6.19e-05 | 7.81e-04 | 0.03608 | 0.4547 | 0.5084 |
| MBRat_Breast_Mammary_Tissue | females | RPS26 | ENSG00000197728.9 | 1470 | 7.04e-160 | 0.2698 | 7.80e-161 | 0.02922 | 0.7009 |
| MBRat_Breast_Mammary_Tissue | males | NPC1 | ENSG00000141458.12 | 1630 | 2.12e-05 | 0.2217 | 7.23e-06 | 0.07498 | 0.7033 |
| MBRat_Breast_Mammary_Tissue | males | NKTR | ENSG00000114857.17 | 1970 | 8.29e-14 | 0.3926 | 1.19e-14 | 0.05567 | 0.5517 |
| MCH_Breast_Mammary_Tissue | females | OXCT2P1 | ENSG00000237624.1 | 1482 | 0.03593 | 0.04581 | 0.1288 | 0.1636 | 0.6258 |

| trait | | gene_name | Locus_id | coloc_data_default_nsnps | coloc_data_default_PP.H0.abf | coloc_data_default_PP.H1.abf | coloc_data_default_PP.H2.abf | coloc_data_default_PP.H3.abf | coloc_data_default_PP.H4.abf |
|---|---|---|---|---|---|---|---|---|---|
| MCH_Breast_Mammary_Tissue | females | PLEC | ENSG00000178209.14 | 1326 | 9.44e-17 | 0.2434 | 2.39e-17 | 0.06097 | 0.6956 |
| MCH_Breast_Mammary_Tissue | males | RP11-161M6.2 | ENSG00000260807.6 | 2142 | 2.44e-04 | 8.61e-05 | 0.3472 | 0.122 | 0.5304 |
| MCHC_Breast_Mammary_Tissue | females | AMZ2P1 | ENSG00000214174.8 | 1043 | 6.94e-08 | 0.3632 | 1.50e-09 | 0.007222 | 0.6296 |
| MCHC_Breast_Mammary_Tissue | females | RP3-465N24.5 | ENSG00000261349.1 | 1535 | 0.003616 | 0.1527 | 3.07e-04 | 0.01215 | 0.8312 |
| MCHC_Breast_Mammary_Tissue | females | SDHDP6 | ENSG00000224183.1 | 1568 | 2.06e-04 | 0.1117 | 1.76e-05 | 0.008652 | 0.8794 |
| MCHC_Breast_Mammary_Tissue | females | HLA-DQB2 | ENSG00000232629.8 | 7068 | 1.97e-25 | 0.1064 | 2.32e-25 | 0.1247 | 0.7689 |
| MCHC_Breast_Mammary_Tissue | males | CDC37P1 | ENSG00000259982.1 | 831 | 5.50e-04 | 0.139 | 8.12e-04 | 0.2046 | 0.6551 |
| MCHC_Breast_Mammary_Tissue | males | NUAK2 | ENSG00000163545.8 | 1735 | 0.001462 | 0.001067 | 0.2352 | 0.171 | 0.5912 |
| MCHC_Breast_Mammary_Tissue | males | ZNF34 | ENSG00000196378.11 | 442 | 0.002068 | 0.1225 | 0.001002 | 0.05849 | 0.816 |
| Meta_Breast_Mammary_Tissue | males | RP11-757G1.6 | ENSG00000250508.1 | 1910 | 0.02975 | 0.2145 | 0.01292 | 0.0925 | 0.6503 |
| Mg_Breast_Mammary_Tissue | females | C1QL4 | ENSG00000186897.4 | 1440 | 7.75e-04 | 0.02428 | 6.55e-04 | 0.01958 | 0.9547 |
| Mg_Breast_Mammary_Tissue | males | LONP2 | ENSG00000102910.13 | 1186 | 0.1004 | 0.1137 | 0.003416 | 0.00309 | 0.7793 |
| MmAcid_Breast_Mammary_Tissue | females | HIBCH | ENSG00000198130.15 | 1823 | 8.15e-12 | 2.47e-10 | 8.40e-04 | 0.02449 | 0.9747 |
| MmAcid_Breast_Mammary_Tissue | males | HIBCH | ENSG00000198130.15 | 1823 | 1.46e-22 | 1.33e-10 | 3.21e-14 | 0.02826 | 0.9717 |
| MonAbs_Breast_Mammary_Tissue | females | ZNF837 | ENSG00000152475.6 | 1203 | 0.04527 | 0.4197 | 0.002612 | 0.0237 | 0.5087 |
| MONOA_Breast_Mammary_Tissue | males | GGH | ENSG00000137563.11 | 2771 | 1.57e-07 | 0.2171 | 9.47e-08 | 0.1302 | 0.6527 |
| MONOAB_Breast_Mammary_Tissue | females | SMG5 | ENSG00000198952.8 | 1223 | 0.1145 | 0.1826 | 0.03498 | 0.05515 | 0.6127 |
| MONOAB_Breast_Mammary_Tissue | males | ZKSCAN3 | ENSG00000189298.13 | 3084 | 2.31e-06 | 2.91e-05 | 0.01301 | 0.1634 | 0.8236 |

| trait | | gene_name | Locus_id | coloc_data_default_nsnps | coloc_data_default_PP.H0.abf | coloc_data_default_PP.H1.abf | coloc_data_default_PP.H2.abf | coloc_data_default_PP.H3.abf | coloc_data_default_PP.H4.abf |
|---|---|---|---|---|---|---|---|---|---|
| Monocy_Breast_Mammary_Tissue | males | NELL2 | ENSG00000184613.10 | 1559 | 1.55e-13 | 0.3293 | 1.63e-14 | 0.03395 | 0.6368 |
| Monocy_Breast_Mammary_Tissue | males | ORMDL3 | ENSG00000172057.9 | 1445 | 0.1016 | 0.06697 | 0.1297 | 0.08482 | 0.6169 |
| MPV_Breast_Mammary_Tissue | females | SPAG7 | ENSG00000091640.7 | 2929 | 2.77e-04 | 0.05492 | 6.37e-04 | 0.1253 | 0.8189 |
| MPV_Breast_Mammary_Tissue | females | YWHAB | ENSG00000166913.12 | 2297 | 0.002943 | 0.03098 | 0.003723 | 0.03827 | 0.9241 |
| MPV_Breast_Mammary_Tissue | females | PLEC | ENSG00000178209.14 | 1326 | 3.31e-17 | 0.0854 | 1.56e-17 | 0.0394 | 0.8752 |
| MPV_Breast_Mammary_Tissue | females | FAM221B | ENSG00000204930.9 | 1570 | 1.57e-04 | 0.09331 | 9.78e-05 | 0.05721 | 0.8492 |
| MPV_Breast_Mammary_Tissue | males | HAUS4 | ENSG00000092036.18 | 2224 | 3.90e-21 | 0.1727 | 8.04e-22 | 0.03483 | 0.7924 |
| MPV_Breast_Mammary_Tissue | males | PIF1 | ENSG00000140451.12 | 1389 | 0.01126 | 0.001981 | 0.3146 | 0.05475 | 0.6174 |
| MPV_Breast_Mammary_Tissue | males | PLEC | ENSG00000178209.14 | 1326 | 2.21e-07 | 3.11e-04 | 2.93e-05 | 0.04024 | 0.9594 |
| MPV_Breast_Mammary_Tissue | males | MTHFD1P1 | ENSG00000231831.1 | 1055 | 1.45e-09 | 7.08e-07 | 7.82e-04 | 0.3803 | 0.6189 |
| Na_Breast_Mammary_Tissue | females | LINC01482 | ENSG00000267659.5 | 1893 | 1.32e-07 | 0.4174 | 1.83e-08 | 0.05737 | 0.5252 |
| NEUTAB_Breast_Mammary_Tissue | males | ZKSCAN3 | ENSG00000189298.13 | 3084 | 0.009608 | 0.1212 | 0.02376 | 0.2993 | 0.5461 |
| NEUTRE_Breast_Mammary_Tissue | males | C14orf37 | ENSG00000139971.15 | 2673 | 0.001057 | 0.3776 | 7.98e-05 | 0.02794 | 0.5933 |
| NRBC._Breast_Mammary_Tissue | males | LINC01291 | ENSG00000204792.2 | 1768 | 2.85e-70 | 0.3682 | 4.66e-71 | 0.05971 | 0.5721 |
| NRBC._Breast_Mammary_Tissue | males | RPL8 | ENSG00000161016.17 | 442 | 7.68e-33 | 0.4047 | 4.26e-34 | 0.02188 | 0.5734 |
| NtAbs_Breast_Mammary_Tissue | females | ASB4 | ENSG00000005981.12 | 2149 | 2.65e-04 | 0.4184 | 2.23e-05 | 0.03479 | 0.5466 |
| trait | | gene_name | Locus_id | coloc_data_default_nsnps | coloc_data_default_PP.H0.abf | coloc_data_default_PP.H1.abf | coloc_data_default_PP.H2.abf | coloc_data_default_PP.H3.abf | coloc_data_default_PP.H4.abf |
| NtAbs_Breast_Mammary_Tissue | males | DYNC1I2 | ENSG00000077380.15 | 2331 | 0.0117 | 0.2415 | 0.003647 | 0.07463 | 0.6685 |

| trait | | gene_name | Locus_id | coloc_data_default_nsnps | coloc_data_default_PP.H0.abf | coloc_data_default_PP.H1.abf | coloc_data_default_PP.H2.abf | coloc_data_default_PP.H3.abf | coloc_data_default_PP.H4.abf |
|---|---|---|---|---|---|---|---|---|---|
| NTAuto_Breast_Mammary_Tissue | males | ADORA2B | ENSG00000170425.3 | 1657 | 2.60e-13 | 0.0663 | 3.46e-13 | 0.08725 | 0.8464 |
| P.HORIZONTAL.AXIS_Breast_Mammary_Tissue | females | TBKBP1 | ENSG00000198933.9 | 2461 | 1.11e-04 | 0.089 | 4.33e-05 | 0.03388 | 0.877 |
| Pat.PT_Breast_Mammary_Tissue | males | RP1-178F15.4 | ENSG00000272030.1 | 1927 | 1.95e-28 | 0.437 | 1.97e-29 | 0.04348 | 0.5195 |
| PCAct_Breast_Mammary_Tissue | females | NAPSB | ENSG00000131401.11 | 1314 | 1.06e-04 | 0.1153 | 2.87e-05 | 0.0304 | 0.8541 |
| PCAct_Breast_Mammary_Tissue | males | NELL2 | ENSG00000184613.10 | 1559 | 1.73e-13 | 0.3669 | 4.34e-14 | 0.09161 | 0.5414 |
| PCAct_Breast_Mammary_Tissue | males | RP11-156E8.1 | ENSG00000272195.1 | 1763 | 1.15e-10 | 0.325 | 2.81e-11 | 0.07868 | 0.5964 |
| PCAct_Breast_Mammary_Tissue | males | RP11-477D19.2 | ENSG00000235381.1 | 2250 | 0.001079 | 0.106 | 8.31e-04 | 0.08082 | 0.8112 |
| pCO2_Breast_Mammary_Tissue | females | ZNF208 | ENSG00000160321.14 | 3056 | 0.009354 | 0.4167 | 0.001537 | 0.06795 | 0.5044 |
| PCR_Breast_Mammary_Tissue | males | CKAP2 | ENSG00000136108.14 | 1629 | 9.44e-27 | 0.2022 | 2.39e-27 | 0.05041 | 0.7474 |
| PCR_Breast_Mammary_Tissue | males | MBTPS1 | ENSG00000140943.16 | 3987 | 0.07662 | 0.2627 | 0.02015 | 0.06853 | 0.572 |
| PCR_Breast_Mammary_Tissue | males | PEX6 | ENSG00000124587.13 | 1455 | 1.65e-36 | 0.3851 | 1.21e-37 | 0.02766 | 0.5873 |
| Plt.Ct_Breast_Mammary_Tissue | females | USP30 | ENSG00000135093.12 | 2353 | 2.63e-05 | 1.34e-05 | 0.01226 | 0.005271 | 0.9824 |
| Plt.Ct_Breast_Mammary_Tissue | females | RP3-465N24.5 | ENSG00000261349.1 | 1535 | 3.90e-04 | 0.01645 | 9.74e-05 | 0.003134 | 0.9799 |
| Plt.Ct_Breast_Mammary_Tissue | females | SDHDP6 | ENSG00000224183.1 | 1568 | 2.02e-05 | 0.01095 | 5.25e-06 | 0.001862 | 0.9872 |
| Plt.Ct_Breast_Mammary_Tissue | females | GCNT1P1 | ENSG00000236474.1 | 2791 | 0.00269 | 0.2722 | 2.23e-04 | 0.02186 | 0.7031 |
| Plt.Ct_Breast_Mammary_Tissue | females | PLEC | ENSG00000178209.14 | 1326 | 9.78e-17 | 0.2522 | 1.03e-17 | 0.02589 | 0.7219 |
| POCA1C_Breast_Mammary_Tissue | females | PIGB | ENSG00000069943.9 | 3373 | 0.1152 | 0.1713 | 0.04983 | 0.07354 | 0.5901 |

| trait | | gene_name | Locus_id | coloc_data_default_nsnps | coloc_data_default_PP.H0.abf | coloc_data_default_PP.H1.abf | coloc_data_default_PP.H2.abf | coloc_data_default_PP.H3.abf | coloc_data_default_PP.H4.abf |
|---|---|---|---|---|---|---|---|---|---|
| POCA1C_Breast_Mammary_Tissue | females | POGZ | ENSG00000143442.21 | 1604 | 4.21e-04 | 0.1082 | 1.59e-04 | 0.03999 | 0.8512 |
| POCA1C_Breast_Mammary_Tissue | females | ANKRD36 | ENSG00000135976.17 | 693 | 5.83e-05 | 0.1458 | 3.83e-05 | 0.09504 | 0.7591 |
| POCA1C_Breast_Mammary_Tissue | females | CYP21A2 | ENSG00000231852.6 | 9452 | 9.83e-04 | 0.02413 | 0.01746 | 0.4281 | 0.5293 |
| POCA1C_Breast_Mammary_Tissue | males | UFSP2 | ENSG00000109775.10 | 2838 | 7.70e-19 | 0.3453 | 1.46e-19 | 0.06499 | 0.5897 |
| PR.INTERVAL_Breast_Mammary_Tissue | females | RRP15 | ENSG00000067533.5 | 2842 | 0.07082 | 0.06685 | 0.02025 | 0.01829 | 0.8238 |
| PR.INTERVAL_Breast_Mammary_Tissue | males | ZNF98 | ENSG00000197360.9 | 2924 | 7.20e-09 | 0.05688 | 2.96e-09 | 0.02246 | 0.9207 |
| Prolac_Breast_Mammary_Tissue | females | ARHGEF19 | ENSG00000142632.16 | 1881 | 5.77e-04 | 0.1151 | 1.13e-04 | 0.02164 | 0.8626 |
| PT.inr_Breast_Mammary_Tissue | males | MMP24-AS1 | ENSG00000126005.16 | 1628 | 0.002553 | 0.001183 | 0.3196 | 0.1475 | 0.5291 |
| PTT.pt_Breast_Mammary_Tissue | males | TCTN2 | ENSG00000168778.11 | 1838 | 1.23e-07 | 0.3071 | 4.64e-08 | 0.1149 | 0.578 |
| PULSE_Breast_Mammary_Tissue | females | RP11-15I11.3 | ENSG00000226251.5 | 2314 | 4.43e-16 | 0.3675 | 4.87e-17 | 0.03984 | 0.5927 |
| Pulse_Breast_Mammary_Tissue | males | ODF2 | ENSG00000136811.16 | 831 | 0.003984 | 0.4521 | 1.91e-04 | 0.02117 | 0.5226 |
| PULSE_Breast_Mammary_Tissue | males | RAB18 | ENSG00000099246.16 | 3089 | 0.001372 | 0.1652 | 4.49e-04 | 0.05326 | 0.7798 |
| QRS_DURATION_Breast_Mammary_Tissue | females | RP11-81A1.6 | ENSG00000278600.1 | 2866 | 1.90e-04 | 0.2697 | 2.02e-05 | 0.02798 | 0.7022 |
| QRS_DURATION_Breast_Mammary_Tissue | females | RP11-545I5.3 | ENSG00000235652.7 | 2149 | 4.62e-11 | 0.2094 | 3.27e-11 | 0.1475 | 0.643 |
| QRS_DURATION_Breast_Mammary_Tissue | males | CNIH1 | ENSG00000100528.11 | 1966 | 4.08e-06 | 0.2276 | 9.28e-07 | 0.05109 | 0.7213 |
| QT_INTERVAL_Breast_Mammary_Tissue | males | HOTAIRM1 | ENSG00000233429.9 | 2527 | 4.05e-10 | 0.3632 | 2.71e-11 | 0.0237 | 0.6131 |
| QT_INTERVAL_C_Breast_Mammary_Tissue | males | ARHGEF19 | ENSG00000142632.16 | 1881 | 1.70e-13 | 0.3203 | 1.40e-14 | 0.02564 | 0.654 |
| QT.INTERVAL_Breast_Mammary_Tissue | females | CLHC1 | ENSG00000162994.15 | 3096 | 3.69e-32 | 0.3735 | 4.94e-33 | 0.04946 | 0.5771 |

| trait | | gene_name | Locus_id | coloc_data_default_nsnps | coloc_data_default_PP.H0.abf | coloc_data_default_PP.H1.abf | coloc_data_default_PP.H2.abf | coloc_data_default_PP.H3.abf | coloc_data_default_PP.H4.abf |
|---|---|---|---|---|---|---|---|---|---|
| QT.INTERVAL_Breast_Mammary_Tissue | males | IFI27L1 | ENSG00000165948.10 | 2603 | 0.02099 | 0.3373 | 0.003181 | 0.05053 | 0.588 |
| QT.INTERVAL_Breast_Mammary_Tissue | males | BIRC7 | ENSG00000101197.12 | 2137 | 0.001085 | 0.2848 | 1.84e-04 | 0.04771 | 0.6663 |
| QT.INTERVAL_Breast_Mammary_Tissue | males | TXNDC5 | ENSG00000239264.8 | 3068 | 1.80e-04 | 0.09495 | 5.19e-05 | 0.02658 | 0.8782 |
| QT3_Breast_Mammary_Tissue | females | BMS1P7 | ENSG00000270025.2 | 1493 | 0.08977 | 0.3052 | 0.009433 | 0.03151 | 0.564 |
| QT3_Breast_Mammary_Tissue | females | RPL8 | ENSG00000161016.17 | 442 | 2.00e-07 | 0.1951 | 6.00e-08 | 0.05782 | 0.7471 |
| QTB.NL_Breast_Mammary_Tissue | females | LDHC | ENSG00000166796.11 | 2464 | 1.06e-07 | 0.2654 | 2.95e-08 | 0.07349 | 0.6611 |
| QTB.NL_Breast_Mammary_Tissue | females | NUP85 | ENSG00000125450.10 | 1340 | 4.42e-11 | 0.2513 | 2.99e-11 | 0.1695 | 0.5791 |
| QTB.NL_Breast_Mammary_Tissue | females | CSTB | ENSG00000160213.6 | 1516 | 0.006314 | 0.4303 | 6.59e-04 | 0.04441 | 0.5183 |
| QTCF_Breast_Mammary_Tissue | males | TXNDC5 | ENSG00000239264.8 | 3068 | 2.82e-04 | 0.1491 | 1.91e-04 | 0.1004 | 0.75 |
| R_F_Breast_Mammary_Tissue | females | HTR7P1 | ENSG00000183935.5 | 2602 | 0.002037 | 0.2168 | 4.66e-04 | 0.04889 | 0.7318 |
| R_F_Breast_Mammary_Tissue | females | ADORA2B | ENSG00000170425.3 | 1657 | 2.47e-06 | 0.2739 | 9.23e-07 | 0.1017 | 0.6243 |
| R.AN.HEART.RATE.MONITOR_Breast_Mammary_Tissue | males | LMOD1 | ENSG00000163431.12 | 2033 | 0.05786 | 0.1722 | 0.01694 | 0.04969 | 0.7034 |
| R.AN.MEAN.ARTERIAL.BLOOD.PRESSURE_Breast_Mammary_Tissue | females | UCA1 | ENSG00000214049.6 | 2682 | 9.82e-14 | 0.3448 | 2.48e-14 | 0.0864 | 0.5688 |
| R.AN.MEAN.ARTERIAL.BLOOD.PRESSURE_Breast_Mammary_Tissue | females | ZC3H3 | ENSG00000014164.6 | 1817 | 1.42e-42 | 0.1875 | 2.78e-43 | 0.03601 | 0.7765 |
| R.AN.PULSE.SPO2_Breast_Mammary_Tissue | males | LMOD1 | ENSG00000163431.12 | 2033 | 0.1038 | 0.3088 | 0.01573 | 0.04628 | 0.5254 |

| trait | | gene_name | Locus_id | coloc_data_default_nsnps | coloc_data_default_PP.H0.abf | coloc_data_default_PP.H1.abf | coloc_data_default_PP.H2.abf | coloc_data_default_PP.H3.abf | coloc_data_default_PP.H4.abf |
|---|---|---|---|---|---|---|---|---|---|
| R.ED.PRE.ARRIVAL.EMS.O2.SAT_Breast_Mammary_Tissue | females | L1TD1 | ENSG00000240563.1 | 2205 | 1.58e-04 | 0.07857 | 3.10e-05 | 0.0145 | 0.9067 |
| R.ED.PRE.ARRIVAL.EMS.O2.SAT_Breast_Mammary_Tissue | females | UBA6 | ENSG00000033178.12 | 2596 | 3.02e-04 | 0.3395 | 9.52e-05 | 0.1066 | 0.5535 |
| R.ED.PRE.ARRIVAL.EMS.O2.SAT_Breast_Mammary_Tissue | males | LINC01806 | ENSG00000227403.1 | 2079 | 5.09e-07 | 0.3346 | 1.28e-07 | 0.08345 | 0.582 |
| R.ED.PRE.ARRIVAL.EMS.O2.SAT_Breast_Mammary_Tissue | males | ATP8A1 | ENSG00000124406.16 | 2891 | 3.94e-05 | 0.06624 | 1.79e-05 | 0.02923 | 0.9045 |
| R.ED.PRE.ARRIVAL.RESP.RATE_Breast_Mammary_Tissue | females | LMOD1 | ENSG00000163431.12 | 2033 | 0.005175 | 0.2424 | 0.001393 | 0.06459 | 0.6864 |
| R.IP.IDEAL.BODY.WEIGHT_Breast_Mammary_Tissue | females | FAM53B | ENSG00000189319.13 | 2284 | 0.04881 | 0.284 | 0.0161 | 0.0931 | 0.5579 |
| R.RT..FEMALE.ADULT..IDEAL.BODY.WEIGHT_Breast_Mammary_Tissue | females | FAM53B | ENSG00000189319.13 | 2284 | 0.04865 | 0.2831 | 0.01711 | 0.099 | 0.5521 |
| R.RT..MALE.ADULT..IDEAL.BODY.WEIGHT_Breast_Mammary_Tissue | females | FAM53B | ENSG00000189319.13 | 2284 | 0.04865 | 0.2831 | 0.01711 | 0.099 | 0.5521 |
| R.VUMC.AN.RESPIRATION.RATE_Breast_Mammary_Tissue | females | PGPEP1 | ENSG00000130517.13 | 1515 | 6.30e-04 | 0.2836 | 4.45e-05 | 0.01936 | 0.6964 |
| trait | | gene_name | Locus_id | coloc_data_default_nsnps | coloc_data_default_PP.H0.abf | coloc_data_default_PP.H1.abf | coloc_data_default_PP.H2.abf | coloc_data_default_PP.H3.abf | coloc_data_default_PP.H4.abf |
| R.VUMC.AN.RESPIRATION.RATE_Breast_Mammary_Tissue | males | SHANK3 | ENSG00000251322.7 | 1269 | 0.1594 | 0.2544 | 0.01118 | 0.01728 | 0.5578 |
| RDW_Breast_Mammary_Tissue | females | CDC37P1 | ENSG00000259982.1 | 831 | 1.37e-07 | 0.34 | 3.47e-08 | 0.08551 | 0.5745 |

| trait | | gene_name | Locus_id | coloc_data_default_nsnps | coloc_data_default_PP.H0.abf | coloc_data_default_PP.H1.abf | coloc_data_default_PP.H2.abf | coloc_data_default_PP.H3.abf | coloc_data_default_PP.H4.abf |
|---|---|---|---|---|---|---|---|---|---|
| RDW_Breast_Mammary_Tissue | females | AMZ2P1 | ENSG00000214174.8 | 1043 | 1.26e-09 | 0.006607 | 2.09e-10 | 9.75e-05 | 0.9933 |
| RDW_Breast_Mammary_Tissue | females | RP3-465N24.5 | ENSG00000261349.1 | 1535 | 0.004798 | 0.2026 | 0.006034 | 0.2543 | 0.5323 |
| RDW_Breast_Mammary_Tissue | females | SDHDP6 | ENSG00000224183.1 | 1568 | 3.96e-04 | 0.2152 | 4.99e-04 | 0.2702 | 0.5137 |
| RDWSD_Breast_Mammary_Tissue | females | CDC37P1 | ENSG00000259982.1 | 831 | 4.55e-08 | 0.1129 | 7.07e-08 | 0.1749 | 0.7122 |
| RDWSD_Breast_Mammary_Tissue | females | TRIM33 | ENSG00000197323.11 | 2091 | 0.008726 | 0.1487 | 8.45e-04 | 0.01357 | 0.8282 |
| RDWSD_Breast_Mammary_Tissue | females | RP11-54A4.2 | ENSG00000237781.3 | 1306 | 0.01748 | 0.09354 | 0.00502 | 0.02601 | 0.858 |
| RDWSD_Breast_Mammary_Tissue | females | CCDC125 | ENSG00000183323.12 | 963 | 1.10e-10 | 0.2489 | 8.63e-12 | 0.01876 | 0.7323 |
| RDWSD_Breast_Mammary_Tissue | males | LSG1 | ENSG00000041802.10 | 2455 | 1.27e-09 | 0.3341 | 1.57e-10 | 0.04075 | 0.6252 |
| RESPIRATIONS_Breast_Mammary_Tissue | males | YWHAB | ENSG00000166913.12 | 2297 | 0.1728 | 0.154 | 0.05155 | 0.04538 | 0.5763 |
| RR.INTERVAL_Breast_Mammary_Tissue | males | PEX7 | ENSG00000112357.12 | 1500 | 0.05219 | 0.3379 | 0.004807 | 0.03055 | 0.5745 |
| SedRat_Breast_Mammary_Tissue | females | RP3-465N24.5 | ENSG00000261349.1 | 1535 | 4.03e-12 | 1.70e-10 | 1.10e-04 | 0.003644 | 0.9962 |
| SedRat_Breast_Mammary_Tissue | females | SDHDP6 | ENSG00000224183.1 | 1568 | 2.27e-13 | 1.23e-10 | 6.20e-06 | 0.002365 | 0.9976 |
| SedRat_Breast_Mammary_Tissue | males | RP3-465N24.5 | ENSG00000261349.1 | 1535 | 6.64e-09 | 7.06e-05 | 2.51e-07 | 0.001668 | 0.9983 |
| SedRat_Breast_Mammary_Tissue | males | SDHDP6 | ENSG00000224183.1 | 1568 | 8.27e-06 | 8.93e-05 | 3.12e-04 | 0.002374 | 0.9972 |
| SGPT_Breast_Mammary_Tissue | males | HTR7P1 | ENSG00000183935.5 | 2602 | 0.06216 | 0.2852 | 0.01199 | 0.05444 | 0.5862 |
| SGPT_Breast_Mammary_Tissue | males | RPL8 | ENSG00000161016.17 | 442 | 4.57e-33 | 0.241 | 2.59e-34 | 0.01289 | 0.7461 |
| SSAIgG_Breast_Mammary_Tissue | males | SNHG5 | ENSG00000203875.10 | 3507 | 5.06e-114 | 0.1338 | 2.37e-114 | 0.06187 | 0.8043 |
| SU.Pro_Breast_Mammary_Tissue | females | NKTR | ENSG00000114857.17 | 1970 | 1.27e-13 | 0.04226 | 5.64e-14 | 0.01783 | 0.9399 |

| trait | | gene_name | Locus_id | coloc_data_default_nsnps | coloc_data_default_PP.H0.abf | coloc_data_default_PP.H1.abf | coloc_data_default_PP.H2.abf | coloc_data_default_PP.H3.abf | coloc_data_default_PP.H4.abf |
|---|---|---|---|---|---|---|---|---|---|
| SU.Pro_Breast_Mammary_Tissue | females | DFNA5 | ENSG00000105928.13 | 3616 | 2.56e-06 | 0.3382 | 3.91e-07 | 0.05111 | 0.6107 |
| SU.Pro_Breast_Mammary_Tissue | males | MRI1 | ENSG00000037757.13 | 1197 | 0.005479 | 0.01036 | 8.42e-04 | 6.09e-04 | 0.9827 |
| SUCrea_Breast_Mammary_Tissue | females | MRPS9 | ENSG00000135972.8 | 2536 | 2.03e-07 | 0.4044 | 4.23e-08 | 0.08362 | 0.512 |
| SUCrea_Breast_Mammary_Tissue | males | LDHC | ENSG00000166796.11 | 2464 | 2.31e-07 | 0.298 | 5.49e-08 | 0.0701 | 0.6319 |
| SUCrea_Breast_Mammary_Tissue | males | RPS26 | ENSG00000197728.9 | 1470 | 2.49e-59 | 0.3289 | 4.25e-60 | 0.0555 | 0.6156 |
| SUCrea_Breast_Mammary_Tissue | males | RP11-1191J2.2 | ENSG00000242686.4 | 2035 | 6.75e-08 | 0.3784 | 8.48e-09 | 0.04694 | 0.5747 |
| SUCrea_Breast_Mammary_Tissue | males | SNHG5 | ENSG00000203875.10 | 3507 | 1.06e-113 | 0.2794 | 3.03e-114 | 0.0796 | 0.641 |
| TC.HDL_Breast_Mammary_Tissue | females | LINC00445 | ENSG00000236036.1 | 2506 | 0.003001 | 0.3956 | 5.41e-04 | 0.07082 | 0.53 |
| TC.HDL_Breast_Mammary_Tissue | males | TSTD1 | ENSG00000215845.10 | 1786 | 0.008306 | 0.2668 | 0.001244 | 0.03929 | 0.6843 |
| TC.HDL_Breast_Mammary_Tissue | males | RP11-273G15.2 | ENSG00000247317.3 | 2463 | 0.03043 | 0.2365 | 0.003483 | 0.02636 | 0.7032 |
| TERMINAL_40_MS_Breast_Mammary_Tissue | females | AC079922.3 | ENSG00000237753.1 | 1908 | 4.53e-04 | 0.1417 | 4.98e-05 | 0.01474 | 0.843 |
| TIBC_Breast_Mammary_Tissue | males | COPG2 | ENSG00000158623.14 | 1950 | 2.63e-05 | 0.0111 | 1.16e-04 | 0.04814 | 0.9406 |
| TIBC_Breast_Mammary_Tissue | males | LYNX1 | ENSG00000180155.19 | 2535 | 7.98e-08 | 0.1261 | 9.45e-09 | 0.01408 | 0.8598 |
| TLC_PPR_Breast_Mammary_Tissue | males | MUC20 | ENSG00000176945.16 | 2192 | 0.00232 | 0.2578 | 4.77e-04 | 0.05237 | 0.687 |
| TProSe_Breast_Mammary_Tissue | females | CBLB | ENSG00000114423.18 | 2938 | 0.1087 | 0.1458 | 0.04896 | 0.06502 | 0.6315 |
| TProSe_Breast_Mammary_Tissue | males | ZKSCAN3 | ENSG00000189298.13 | 3084 | 0.001112 | 0.01403 | 0.02448 | 0.3082 | 0.6521 |
| TProSe_Breast_Mammary_Tissue | males | FBXL6 | ENSG00000182325.10 | 869 | 5.93e-11 | 0.4266 | 2.16e-12 | 0.01495 | 0.5585 |

| trait | | gene_name | Locus_id | coloc_data_default_nsnps | coloc_data_default_PP.H0.abf | coloc_data_default_PP.H1.abf | coloc_data_default_PP.H2.abf | coloc_data_default_PP.H3.abf | coloc_data_default_PP.H4.abf |
|---|---|---|---|---|---|---|---|---|---|
| TProt_Breast_Mammary_Tissue | females | TPM3P6 | ENSG00000250731.1 | 1501 | 0.007288 | 0.4329 | 2.88e-04 | 0.01658 | 0.5429 |
| TProt_Breast_Mammary_Tissue | females | XAB2 | ENSG00000076924.11 | 1628 | 2.46e-13 | 0.3728 | 1.50e-14 | 0.02213 | 0.605 |
| TProt_Breast_Mammary_Tissue | males | ZKSCAN3 | ENSG00000189298.13 | 3084 | 0.001489 | 0.01879 | 0.03115 | 0.3925 | 0.556 |
| Trigs_Breast_Mammary_Tissue | males | VPS11 | ENSG00000160695.14 | 2134 | 1.96e-07 | 0.2498 | 3.22e-08 | 0.04027 | 0.7099 |
| TSH_Breast_Mammary_Tissue | females | HLA-DQA2 | ENSG00000237541.3 | 7023 | 5.14e-25 | 0.036 | 4.53e-24 | 0.3167 | 0.6473 |
| TSH_Breast_Mammary_Tissue | females | HLA-DQB2 | ENSG00000232629.8 | 7068 | 4.36e-26 | 0.02356 | 3.84e-25 | 0.2069 | 0.7695 |
| TSH_Breast_Mammary_Tissue | males | SULT1C2 | ENSG00000198203.9 | 1688 | 0.12 | 0.2316 | 0.02352 | 0.04483 | 0.5801 |
| TSH_Breast_Mammary_Tissue | males | ZKSCAN3 | ENSG00000189298.13 | 3084 | 0.01173 | 0.148 | 0.02489 | 0.3136 | 0.5018 |
| TSH_Breast_Mammary_Tissue | males | HLA-DQB2 | ENSG00000232629.8 | 7068 | 1.96e-42 | 0.1089 | 5.23e-42 | 0.2909 | 0.6002 |
| TT4_Breast_Mammary_Tissue | males | SFXN2 | ENSG00000156398.12 | 1837 | 0.07344 | 0.1022 | 0.03626 | 0.04972 | 0.7384 |
| TT4_Breast_Mammary_Tissue | males | LRCH1 | ENSG00000136141.14 | 2321 | 0.0191 | 0.2481 | 0.004969 | 0.06389 | 0.6639 |
| TU.Pro_Breast_Mammary_Tissue | males | RPL8 | ENSG00000161016.17 | 442 | 8.82e-33 | 0.4649 | 6.57e-34 | 0.03414 | 0.5009 |
| TUCrea_Breast_Mammary_Tissue | females | PLEKHB1 | ENSG00000021300.13 | 1886 | 2.87e-04 | 0.3874 | 6.66e-05 | 0.08956 | 0.5227 |
| UAlb_Breast_Mammary_Tissue | females | MESP1 | ENSG00000166823.5 | 2120 | 1.09e-06 | 0.03239 | 2.67e-06 | 0.07834 | 0.8893 |
| UAlb_Breast_Mammary_Tissue | females | COPG2 | ENSG00000158623.14 | 1950 | 0.2072 | 0.1818 | 0.05694 | 0.04946 | 0.5046 |
| UGluc_Breast_Mammary_Tissue | males | PXMP2 | ENSG00000176894.9 | 1570 | 0.09288 | 0.03422 | 0.1912 | 0.06982 | 0.6119 |
| UGluc_Breast_Mammary_Tissue | males | EPCAM | ENSG00000119888.10 | 2538 | 0.167 | 0.1128 | 0.09249 | 0.06195 | 0.5658 |
| UricA_Breast_Mammary_Tissue | females | DAB1 | ENSG00000173406.15 | 2423 | 0.05997 | 0.3405 | 0.01363 | 0.07689 | 0.509 |

| trait | | gene_name | Locus_id | coloc_data_default_nsnps | coloc_data_default_PP.H0.abf | coloc_data_default_PP.H1.abf | coloc_data_default_PP.H2.abf | coloc_data_default_PP.H3.abf | coloc_data_default_PP.H4.abf |
|---|---|---|---|---|---|---|---|---|---|
| UricA_Breast_Mammary_Tissue | males | MRI1 | ENSG00000037757.13 | 1197 | 0.1464 | 0.2768 | 0.005544 | 0.009922 | 0.5614 |
| VC_PPR_Breast_Mammary_Tissue | females | DOC2GP | ENSG00000231793.5 | 921 | 0.05353 | 0.3818 | 0.007005 | 0.04946 | 0.5082 |
| VC_PPR_Breast_Mammary_Tissue | females | ASB4 | ENSG00000005981.12 | 2149 | 2.68e-04 | 0.4233 | 4.66e-05 | 0.07311 | 0.5033 |
| VC_PPR_Breast_Mammary_Tissue | females | GXYLT1P5 | ENSG00000276591.1 | 2 | 2.04e-04 | 0.31 | 4.66e-07 | 1.91e-05 | 0.6898 |
| VC_PPR_Breast_Mammary_Tissue | females | RP11-459O16.8 | ENSG00000276128.1 | 2 | 1.27e-09 | 0.31 | 2.90e-12 | 1.91e-05 | 0.6899 |
| trait | | gene_name | Locus_id | coloc_data_default_nsnps | coloc_data_default_PP.H0.abf | coloc_data_default_PP.H1.abf | coloc_data_default_PP.H2.abf | coloc_data_default_PP.H3.abf | coloc_data_default_PP.H4.abf |
| VC_PPR_Breast_Mammary_Tissue | males | RP11-479G22.8 | ENSG00000273038.2 | 3388 | 8.11e-06 | 0.21 | 8.76e-06 | 0.2263 | 0.5637 |
| VC_PPR_Breast_Mammary_Tissue | males | YWHAB | ENSG00000166913.12 | 2297 | 0.154 | 0.1373 | 0.06029 | 0.05315 | 0.5953 |
| VC_PPR_Breast_Mammary_Tissue | males | LINC01504 | ENSG00000225434.2 | 2174 | 1.27e-36 | 0.3651 | 2.43e-37 | 0.06914 | 0.5658 |
| VC_ZBD_Breast_Mammary_Tissue | females | RP11-817J15.3 | ENSG00000281655.1 | 3506 | 8.42e-07 | 0.2623 | 3.56e-07 | 0.1103 | 0.6274 |
| VC_ZBD_Breast_Mammary_Tissue | females | PIGG | ENSG00000174227.15 | 1797 | 0.01676 | 0.2602 | 0.007195 | 0.1111 | 0.6048 |
| VC_ZBD_Breast_Mammary_Tissue | males | LINC01504 | ENSG00000225434.2 | 2174 | 4.77e-37 | 0.1369 | 1.31e-37 | 0.03676 | 0.8263 |
| VD25_Breast_Mammary_Tissue | females | ZNF98 | ENSG00000197360.9 | 2924 | 0.1878 | 0.1608 | 0.07397 | 0.06285 | 0.5146 |
| Vi.B12_Breast_Mammary_Tissue | females | TRIM23 | ENSG00000113595.14 | 2177 | 1.12e-04 | 0.2624 | 4.87e-05 | 0.1131 | 0.6244 |
| Vi.B12_Breast_Mammary_Tissue | males | LINC01481 | ENSG00000257815.5 | 3130 | 5.31e-04 | 0.2937 | 3.50e-04 | 0.193 | 0.5125 |
| ViARet_Breast_Mammary_Tissue | females | FAM86C1 | ENSG00000158483.15 | 1245 | 0.08633 | 0.1763 | 0.007055 | 0.01369 | 0.7166 |
| ViARet_Breast_Mammary_Tissue | females | C17orf97 | ENSG00000187624.8 | 1306 | 1.09e-70 | 0.3396 | 1.70e-71 | 0.05266 | 0.6078 |
| ViARet_Breast_Mammary_Tissue | males | C19orf33 | ENSG00000167644.11 | 2053 | 1.47e-05 | 0.2001 | 7.37e-06 | 0.09945 | 0.7004 |

| trait | | gene_name | Locus_id | coloc_data_default_nsnps | coloc_data_default_PP.H0.abf | coloc_data_default_PP.H1.abf | coloc_data_default_PP.H2.abf | coloc_data_default_PP.H3.abf | coloc_data_default_PP.H4.abf |
|---|---|---|---|---|---|---|---|---|---|
| WBC_Breast_Mammary_Tissue | females | FAM53B | ENSG00000189319.13 | 2284 | 0.03164 | 0.1841 | 0.00497 | 0.02817 | 0.7511 |
| WBC_Breast_Mammary_Tissue | males | ADORA2B | ENSG00000170425.3 | 1657 | 7.08e-13 | 0.1806 | 4.75e-13 | 0.1205 | 0.699 |
| WBC_Breast_Mammary_Tissue | males | ZKSCAN3 | ENSG00000189298.13 | 3084 | 6.66e-07 | 8.41e-06 | 0.02809 | 0.3539 | 0.618 |
| WBC_Breast_Mammary_Tissue | males | TAP1 | ENSG00000168394.10 | 7236 | 5.63e-07 | 1.47e-04 | 0.001193 | 0.3116 | 0.6871 |
| WBCrt_Breast_Mammary_Tissue | females | CD8BP | ENSG00000254126.6 | 3440 | 0.003402 | 0.08798 | 5.24e-04 | 0.01267 | 0.8954 |
| WBCrt_Breast_Mammary_Tissue | females | MUC20 | ENSG00000176945.16 | 2192 | 0.0426 | 0.1586 | 0.02233 | 0.08246 | 0.694 |
| WBCrt_Breast_Mammary_Tissue | males | SMYD4 | ENSG00000186532.11 | 1809 | 3.83e-04 | 0.2714 | 1.06e-04 | 0.07416 | 0.654 |
| Weight_Breast_Mammary_Tissue | females | NPC1 | ENSG00000141458.12 | 1630 | 3.56e-08 | 0.4229 | 6.46e-09 | 0.0762 | 0.5009 |
| Weight_Breast_Mammary_Tissue | females | RPRD1A | ENSG00000141425.17 | 2069 | 0.1092 | 0.1509 | 0.0195 | 0.02623 | 0.6941 |
| Weight_Breast_Mammary_Tissue | males | CDC37P1 | ENSG00000259982.1 | 831 | 0.001562 | 0.3949 | 2.83e-04 | 0.07103 | 0.5323 |
| Weight_Breast_Mammary_Tissue | males | LY75 | ENSG00000054219.10 | 2863 | 6.62e-30 | 0.1544 | 1.21e-29 | 0.2819 | 0.5637 |
| Weight_Breast_Mammary_Tissue | males | CAST | ENSG00000153113.23 | 2601 | 1.67e-12 | 0.1788 | 3.07e-13 | 0.03194 | 0.7893 |
| WEIGHT_SCALE_Breast_Mammary_Tissue | females | FGFR3 | ENSG00000068078.17 | 2034 | 0.04821 | 0.2387 | 0.01324 | 0.06492 | 0.635 |
| X_percent_SAT_Breast_Mammary_Tissue | females | MUC20 | ENSG00000176945.16 | 2192 | 0.02693 | 0.1003 | 0.01343 | 0.04919 | 0.8102 |
| X_percent_SAT_Breast_Mammary_Tissue | males | KCNC4 | ENSG00000116396.14 | 1983 | 6.92e-06 | 0.3984 | 5.96e-07 | 0.03378 | 0.5678 |
| X.WBC_Breast_Mammary_Tissue | females | CCDC88C | ENSG00000015133.18 | 2302 | 0.01631 | 0.1862 | 0.003237 | 0.0362 | 0.758 |
| X.WBC_Breast_Mammary_Tissue | females | RABL2A | ENSG00000144134.18 | 2284 | 2.17e-04 | 0.3369 | 4.12e-05 | 0.06347 | 0.5993 |
| X.WBC_Breast_Mammary_Tissue | males | CCDC125 | ENSG00000183323.12 | 963 | 1.69e-07 | 0.2814 | 5.69e-08 | 0.09434 | 0.6242 |

| trait | | gene_name | Locus_id | coloc_data_default_nsnps | coloc_data_default_PP.H0.abf | coloc_data_default_PP.H1.abf | coloc_data_default_PP.H2.abf | coloc_data_default_PP.H3.abf | coloc_data_default_PP.H4.abf |
|---|---|---|---|---|---|---|---|---|---|
| X25DTot_Breast_Mammary_Tissue | females | C17orf97 | ENSG00000187624.8 | 1306 | 2.67e-71 | 0.08356 | 3.35e-72 | 0.009568 | 0.9069 |
